# Genome-wide analysis of 102,084 migraine cases identifies 123 risk loci and subtype-specific risk alleles

**DOI:** 10.1101/2021.01.20.21249647

**Authors:** Heidi Hautakangas, Bendik S. Winsvold, Sanni E. Ruotsalainen, Gyda Bjornsdottir, Aster V. E. Harder, Lisette J. A. Kogelman, Laurent F. Thomas, Raymond Noordam, Christian Benner, Padhraig Gormley, Ville Artto, Karina Banasik, Anna Bjornsdottir, Dorret I. Boomsma, Ben M. Brumpton, Kristoffer Sølvsten Burgdorf, Julie E. Buring, Mona Ameri Chalmer, Irene de Boer, Martin Dichgans, Christian Erikstrup, Markus Färkkilä, Maiken Elvestad Garbrielsen, Mohsen Ghanbari, Knut Hagen, Paavo Häppölä, Jouke-Jan Hottenga, Maria G. Hrafnsdottir, Kristian Hveem, Marianne Bakke Johnsen, Mika Kähönen, Espen S. Kristoffersen, Tobias Kurth, Terho Lehtimäki, Lannie Lighart, Sigurdur H. Magnusson, Rainer Malik, Ole Birger Pedersen, Nadine Pelzer, Brenda W. J. H. Penninx, Caroline Ran, Paul M. Ridker, Frits R. Rosendaal, Gudrun R. Sigurdardottir, Anne Heidi Skogholt, Olafur A. Sveinsson, Thorgeir E. Thorgeirsson, Henrik Ullum, Lisanne S. Vijfhuizen, Elisabeth Widén, Ko Willems van Dijk, International Headache Genetics Consortium, HUNT All-in Headache, Danish Blood Donor Study Genomic Cohort, Arpo Aromaa, Andrea Carmine Belin, Tobias Freilinger, M. Arfan Ikram, Marjo-Riitta Järvelin, Olli T. Raitakari, Gisela M. Terwindt, Mikko Kallela, Maija Wessman, Jes Olesen, Daniel I. Chasman, Dale R. Nyholt, Hreinn Stefánsson, Kari Stefansson, Arn M. J. M. van den Maagdenberg, Thomas Folkmann Hansen, Samuli Ripatti, John-Anker Zwart, Aarno Palotie, Matti Pirinen

## Abstract

Migraine affects over a billion individuals worldwide but its genetic underpinning remains largely unknown. This genome-wide association study (GWAS) of 102,084 migraine cases and 771,257 controls identified 123 loci of which 86 are novel. The loci provide an opportunity to evaluate shared and distinct genetic components in the two main migraine subtypes: migraine with aura and migraine without aura. A stratification of the risk loci using 29,679 cases with subtype information, of which approximately half have never been used in a GWAS before, indicated three risk variants that appear specific for migraine with aura (in *HMOX2, CACNA1A* and *MPPED2*), two that appear specific for migraine without aura (near *SPINK2* and near *FECH*), and nine that increase susceptibility for migraine regardless of subtype. The new risk loci include genes encoding recent migraine-specific drug targets, namely calcitonin gene-related peptide (*CALCA/CALCB*) and serotonin 1F receptor (*HTR1F*). Overall, genomic annotations among migraine-associated variants were enriched in both vascular and central nervous system tissue/cell types supporting unequivocally that neurovascular mechanisms underlie migraine pathophysiology.

## Introduction

Migraine is a highly prevalent brain disorder characterized by disabling attacks of moderate to severe pulsating and usually one-sided headache that may be aggravated by physical activity and can be associated with symptoms such as a hypersensitivity to light and sound, nausea and vomiting (Headache Classification Committee of the International Headache Society (IHS), 2018). Migraine has a lifetime prevalence of 15-20% and is ranked as the second most disabling condition in terms of years lived with disability (Vos et al., 2020), (Steiner et al., 2020). Migraine is three times more prevalent in females than in males. For about one-third of patients, migraine attacks often include an aura phase (Russell et al., 1995) characterized by transient neurological symptoms such as scintillations. Hence, the two main migraine subtypes are defined as migraine with aura (MA) and migraine without aura (MO).

It has been debated for decades whether or not the migraine subtypes are in fact two separate disorders (Russell and Olesen, 1995),(Kallela et al., 2001),(de Boer et al., 2019) and if so, what the underlying causes are. Prevailing theories about migraine pathophysiology emphasize neuronal and/or vascular dysfunction (Tfelt-Hansen and Koehler, 2011), (Anttila et al., 2018). Current knowledge on disease mechanisms largely comes from studies of a rare monogenic sub-form of MA, familial hemiplegic migraine, for which three ion transporter genes (*CACNA1A, ATP1A2* and *SCN1A*) have been identified (Ferrari et al., 2015). The common forms of migraine, MA and MO, instead have a complex polygenic architecture with an increased familial relative risk (Russell and Olesen, 1995), increased concordance in monozygotic twins (Ulrich et al., 1999) and a heritability of 40-60% (Gervil et al., 1999). The largest GWAS thus far with 59,674 cases and 316,078 controls reported 38 genomic loci that confer migraine risk (Gormley et al., 2016). Subsequent analyses of these GWAS data (Finucane et al., 2018) showed enrichment of migraine signals near activating histone marks specific to cardiovascular and central nervous system tissues, as well as for genes expressed in vascular and smooth muscle tissues (Gormley et al., 2016). Other smaller GWAS (Anttila et al., 2010), (Chasman et al., 2011), (Freilinger et al., 2012), (Anttila et al., 2013), (Pickrell et al., 2016), (Chen et al., 2018), (Chang et al., 2018) have suggested 10 additional loci. Of note, the previous datasets were too small to perform a meaningful comparison of the genetic background between migraine subtypes.

As migraine is globally the second largest contributor to years lived with disability (Vos et al., 2020), (Steiner et al., 2020) there clearly is a large need for new treatments. Triptans, i.e., serotonin 5-HT_1B/1D_ receptor agonists, are migraine-specific acute treatments for the headache phase but are not effective in every patient, whereas preventive medication is far from satisfactory all together (Tfelt-Hansen and Olesen, 2012). Recent promising alternatives for acute treatment are serotonin 5-HT_1F_ receptor agonists (‘ditans’) (Kuca et al., 2018) and small-molecule calcitonin-gene related peptide (CGRP) receptor antagonists (’gepants’) (Lipton et al., 2019), (Dodick, 2018). For preventive treatment, monoclonal antibodies (mAbs) targeting CGRP or its receptor have recently proven effective (Charles and Pozo-Rosich, 2019) and new gepants are under development for migraine prevention (Goadsby et al., 2020). Still, there remains an urgent need for treatment options for patients who do not respond to the existing treatments. Genetics has proven a promising way to develop novel therapeutic hypotheses in other prevalent complex diseases, such as cardiovascular disease (Cohen et al., 2005) and type 2 diabetes (Flannick et al., 2014), and we anticipate that large genetic studies of migraine could also yield to similar insights.

We conducted a GWAS meta-analysis of migraine by adding to the previous meta-analysis of Gormley et al. (2016) 42,410 new migraine cases from four study collections (Table 1). This increased the number of migraine cases by 71% for a total sample of 102,084 cases and 771,257 controls. Furthermore, we assessed the subtype specificity of the risk loci in 8,292 new MA and 6,707 new MO cases in addition to the 6,332 MA and 8,348 MO cases used previously (Gormley et al. 2016) (Table 2). Here we report 123 genomic loci, of which 86 are novel, and include the first four loci that reach genome-wide significance (*P* < 5 × 10^− 8^) in MA. Our subtype data compellingly show that migraine risk is conferred both by risk loci that appear specific for only one subtype as well as by loci that are shared by both subtypes. Our findings also include new risk loci containing target genes of recent migraine drugs acting on the CGRP pathway and the serotonin 5-HT_1F_ receptor. Finally, our data support the concept that migraine is brought about by both neuronal and vascular genetic factors, strengthening the view that migraine truly is a neurovascular disorder.

**Table 1.** Five migraine study collections included in the meta-analysis.

| Abbreviation | Full Name | Ethnicity | Cases | Controls | Case % | Migraine Definition |
| --- | --- | --- | --- | --- | --- | --- |
| IHGC2016* | Gormley et al. 2016<br>(no 23andMe) | European<br>descent | 29,209 | 172,931 | 14.4 | Self-reported and ICHD-II |
| 23andMe | 23andMe, Inc.<br>(23andMe.com) | European<br>descent | 53,109 | 230,876 | 18.7 | Self-reported |
| UKBB | UK Biobank<br>(ukbiobank.ac.uk) | European,<br>British | 10,881 | 330,170 | 3.2 | Self-reported |
| GeneRISK | GeneRISK<br>(generisk.fi) | European,<br>Finnish | 1,084 | 4,857 | 18.2 | Self-reported |
| HUNT | Nord-Trøndelag<br>Health Study<br>(ntnu.edu/hunt) | European,<br>Norwegian | 7,801 | 32,423 | 19.4 | Self-reported migraine or<br>fulfilling modified ICHD-II criteria |

\*IHGC2016 is a meta-analysis of 21 studies listed in Supplementary Table 1. Some studies of IHGC2016 determined migraine status through clinical phenotyping while migraine status in other studies is based on self-reported information. ICHD-II = the International Classification of Headache Disorders 2<sup>nd</sup> edition.

**Table 2.**
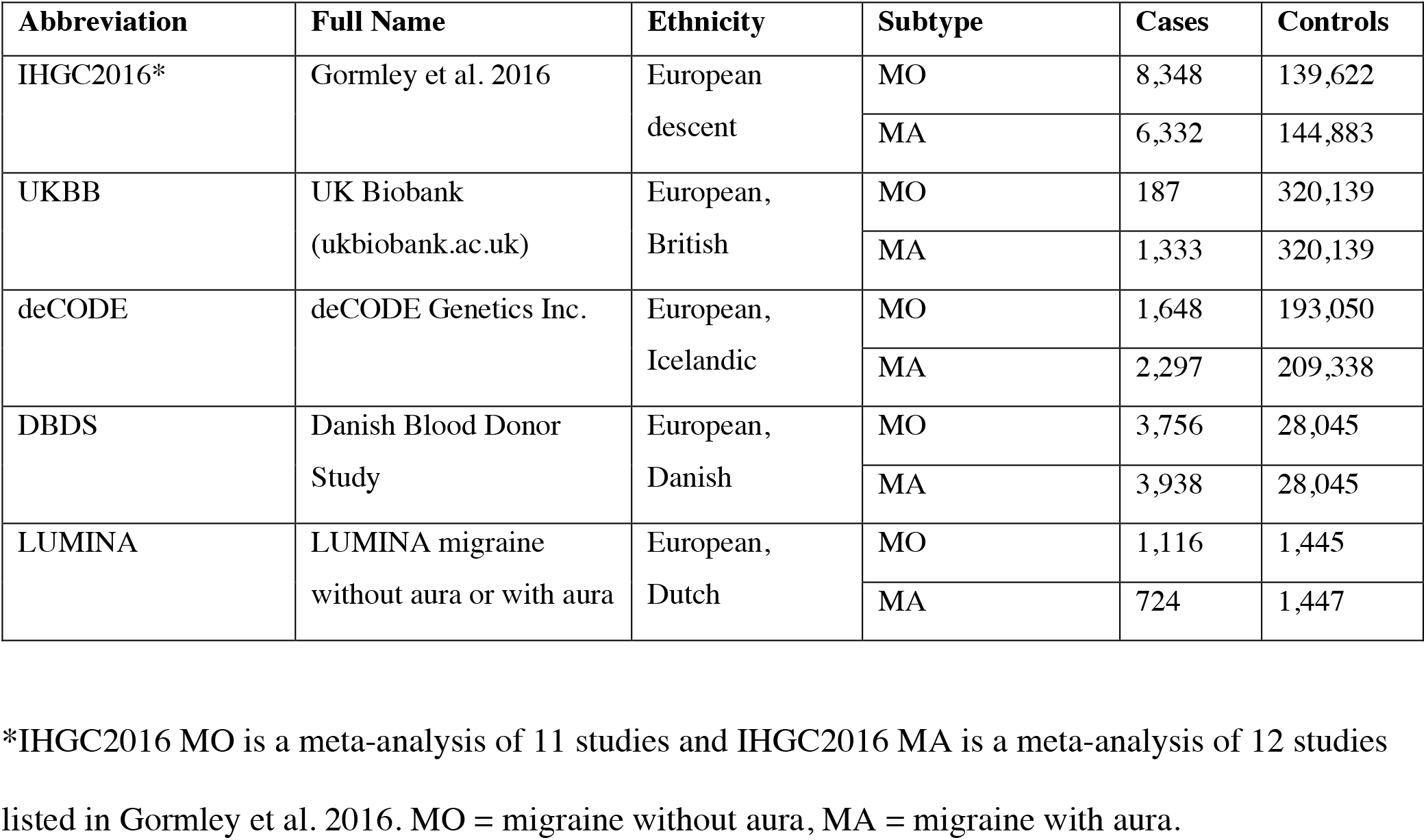
Study collections included in MO and MA subtype analyses.

| Abbreviation | Full Name | Ethnicity | Subtype | Cases | Controls |
| --- | --- | --- | --- | --- | --- |
| IHGC2016* | Gormley et al. 2016 | European descent | MO | 8,348 | 139,622 |
|  |  |  | MA | 6,332 | 144,883 |
| UKBB | UK Biobank<br>(ukbiobank.ac.uk) | European,<br>British | MO | 187 | 320,139 |
|  |  |  | MA | 1,333 | 320,139 |
| deCODE | deCODE Genetics Inc. | European,<br>Icelandic | MO | 1,648 | 193,050 |
|  |  |  | MA | 2,297 | 209,338 |
| DBDS | Danish Blood Donor<br>Study | European,<br>Danish | MO | 3,756 | 28,045 |
|  |  |  | MA | 3,938 | 28,045 |
| LUMINA | LUMINA migraine<br>without aura or with aura | European,<br>Dutch | MO | 1,116 | 1,445 |
|  |  |  | MA | 724 | 1,447 |

\*IHGC2016 MO is a meta-analysis of 11 studies and IHGC2016 MA is a meta-analysis of 12 studies
listed in Gormley et al. 2016. MO = migraine without aura, MA = migraine with aura.

## Results

### Genome-wide meta-analysis

We combined data on 873,341 individuals of European ancestry (102,084 cases and 771,257 controls) from 5 study collections (Table 1 and Supplementary Table 1) and analyzed 10,843,197 common variants (Methods). In spite of different approaches to the ascertainment of migraine cases across the studies, the pairwise genetic correlations were all near 1 (Supplementary Table 2), as determined by LD Score (LDSC) regression (Bulik-Sullivan et al., 2015a), showing high genetic and phenotypic similarity across the studies justifying their meta-analysis. Pairwise LDSC intercepts were all near 0, indicating little or no sample overlap (Supplementary Table 2).

The genomic inflation factor (λ_GC_) of the fixed-effect meta-analysis results was 1.33 (Supplementary Fig. 1), which is in line with other large meta-analyses (Nagel et al., 2018), (Pardiñ as et al., 2018), (Howard et al., 2019) and is as expected for a polygenic trait (Yang et al., 2011). The univariate LDSC (Bulik-Sullivan et al., 2015b) intercept was 1.05 (s.e. 0.01), which, being close to 1.0, suggests that most of the genome-wide elevation of the association statistics comes from true additive polygenic effects rather than from a confounding bias such as population stratification. The LDSC analysis showed a linear trend between the variant’s LD-score and its association with migraine as expected from a highly polygenic phenotype such as migraine (Supplementary Fig. 2). The SNP-heritability estimate from LDSC was 11.2% (95%CI 10.8-11.6%) on a liability scale when assuming a population prevalence of 16%.

We identified 8,117 genome-wide significant (GWS; *P* < 5 × 10^− 8^) variants represented by 170 LD-independent index variants (*r*^2^ < 0.1). We defined the risk loci by including all variants in high LD (*r*^2^ > 0.6) with the index variants and merged loci that were closer than 250 kb (Methods). This resulted in 123 independent risk loci (Fig. 1, Supplementary Table 3a, Supplementary Fig. 3 and 4). Of the 123 loci, 86 are novel whereas 36 overlap with the previously reported 47 autosomal risk loci (Supplementary Table 4) and one with the previously reported X chromosome risk locus. When we represented each risk locus by its lead variant, i.e., the variant with the smallest GWAS *P*-value, 47 GWS variants were LD-independent (*r*^2^< 0.1) of the 123 lead variants, and with a more stringent threshold (*r*^2^< 0.01), 15 GWS variants remained LD independent of the 123 lead variants (Supplementary Table 5).

**Fig. 1:**
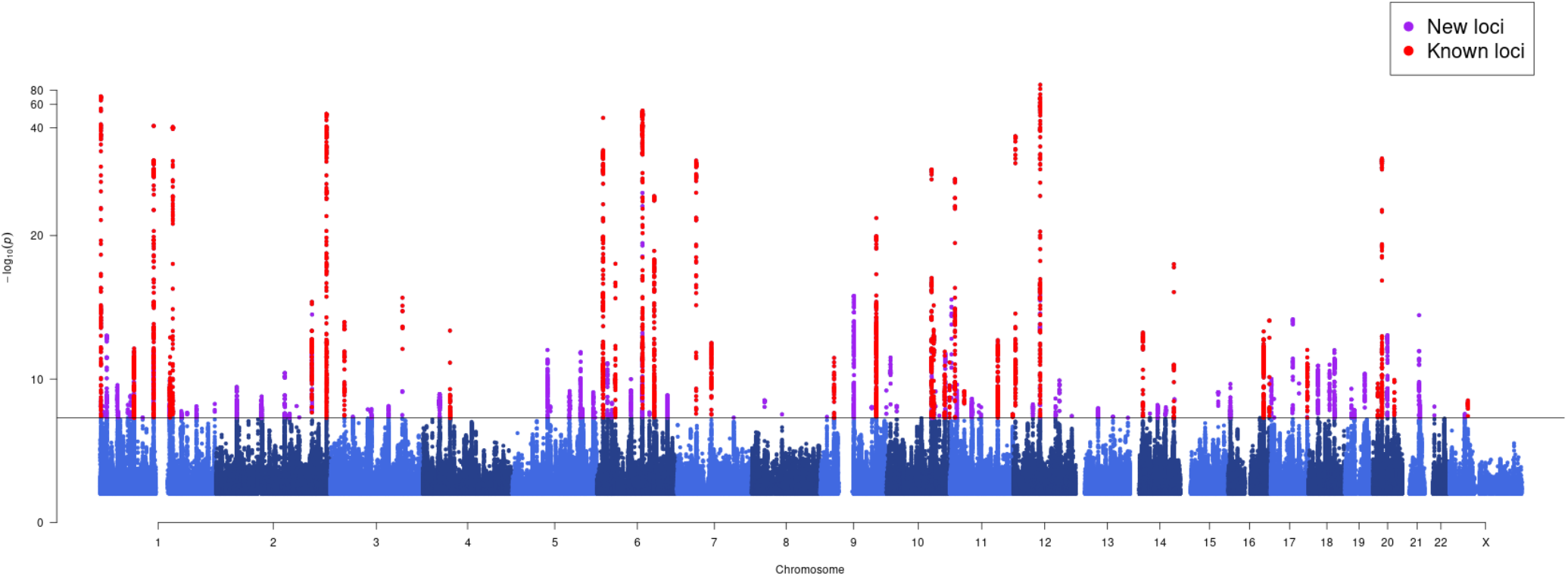
Manhattan plot of migraine GWAS meta-analysis (N = 873,341; 102,084 cases and 771,257 controls). On the X-axis, variants are plotted along the 22 autosomes and the X chromosome. Y-axis shows the statistical strength of the association from the fixed-effect meta-analysis as the negative log10 of the *P*-value. Horizontal line is the genome-wide significance threshold (*P* = 5 × 10^− 8^). The 123 risk loci passing the threshold are divided into 86 new loci (purple) and 37 previously known loci (red).

In addition, we conducted an approximate stepwise conditional analysis for the 123 risk loci (Methods). Since sample sizes per variant varied considerably, we restricted the conditional analysis to variants with similar effective sample sizes to the lead variant, which excluded approximately 17% of all GWS variants. The conditional analysis returned 6 SNPs within the 123 risk loci that remained GWS after conditioning on the lead variants (Supplementary Tables 6a,b).

### Characterization of migraine risk loci

We mapped the 123 risk loci to genes by their physical location using the Ensembl Variant Effect Predictor (VEP) (McLaren et al., 2016). In total, transcripts of 390 protein coding and 329 non-protein coding genes were identified within the genomic region of 20 kb around each locus. The number of transcripts increased to 1,036 protein coding and 1,236 non-protein coding genes when the genomic region was expanded to 250 kb around each locus (Supplementary Table 3). Of the lead variants, 59% (72 of 123) were within a transcript of a protein coding gene, and 80% (99 of 123) of the loci contained at least one protein coding gene within 20 kb; and 93% (114 of 123) within 250 kb. Five of the 123 lead variants were missense variants (in genes *PLCE1, MRGPRE, SERPINA1, ZBTB4* and *ZNF462*), and 40 more missense variants were in high LD (*r*^2^ > 0.6) with the lead variants (Supplementary Table 7a). Of note, three variants with a predicted high impact consequence on protein function were in high LD with the lead variants: (1) a stop gained variant (rs34358) with lead variant rs42854 (*r*^2^ = 0.85) in gene *ANKDD1B*, (2) a splice donor variant (rs66880209) with lead variant rs1472662 (*r*^2^ = 0.71) in *RP11-420K8*.*1*, and (3) a splice acceptor variant (rs11042902) with lead variant rs4910165 (*r*^2^ = 0.69) in *MRVI1* (Supplementary Table 7b).

We further mapped the 123 lead variants to genes via expression quantitative trait locus (eQTL) association using the GTEx V8 data (Aguet et al., 2019) and data repositories included in FUMA (Watanabe et al., 2017) at a false discovery rate (FDR) of 5% (Methods). The lead variants were *cis*-eQTLs for 589 genes (Supplementary Table 8). Variants in high LD with the lead variants were *cis*-eQTLs for an additional 624 genes (Supplementary Table 9). In total, 84% (103/123) of lead variants were *cis*-eQTLs for at least one gene, and 67% (84/123) were *cis*-eQTLs in the GTEx data. All 49 GTEx tissues had at least one lead variant as a significant *cis*-eQTL. Tibial artery had the highest number (47/123) of lead variants as *cis*-eQTLs, and it was the only GTEx tissue type where the enrichment was statistically higher (*P* = 6.37 × 10^− 6^) than expected based on the overall number of *cis*-eQTLs per tissue reported by GTEx (Supplementary Fig. 5, Supplementary Note 1).

We used the stratified LDSC (S-LDSC) to partition migraine heritability by 24 functional genomic annotations (Finucane et al., 2015). Enrichment was observed for 14 functional categories at an FDR of 5% (Supplementary Table 10a), of which 11 remained statistically significant after Bonferroni correction for the 24 hypotheses tested (Supplementary Fig. 6). Conserved regions showed the highest enrichment, namely 15.43x (s.e. 1.66; *P* =3.32 × 10^− 16^), followed by coding regions at 9.97x (s.e. 2.53; *P* = 6.74 × 10^− 4^), and transcription start sites (TSS) at 6.70x (s.e. 1.83; *P* = 1.18 × 10^− 3^) (Supplementary Table 10b).

Notably, two of the new risk loci contain genes (*CALCA*/*CALCB* and *HTR1F*) whose protein products are closely related to targets of two migraine-specific drug therapies (Do et al., 2019). We observe a convincing association at the chromosome 11 locus that contains the *CALCA* and *CALCB* genes encoding CGRP itself (lead SNP rs1003194, *P* = 2.43 × 10^− 10^; Fig. 2a), while none of the genes encoding CGRP receptor proteins (*CALCRL, RAMP1* or *RCP*) show a statistically comparable association (all *P* > 10^− 4^; Supplementary Fig. 7). Variant rs1003194 is a *cis*-eQTL for *CALCB*, but also for *COPB1, PDE3B* and *INSC* (Supplementary Table 8). In addition, a new locus on chromosome 3 contains *HTR1F* (lead SNP rs6795209, *P* = 1.23 × 10^− 8^; Fig. 2b), which encodes the serotonin 5-HT_1F_ receptor. Variant rs6795209 is a significant *cis*-eQTL for *HTR1F*, as well as for three other genes (*CGGBP1, ZNF654, C3orf38*) in the same locus (Supplementary Table 8).

**Fig. 2:**
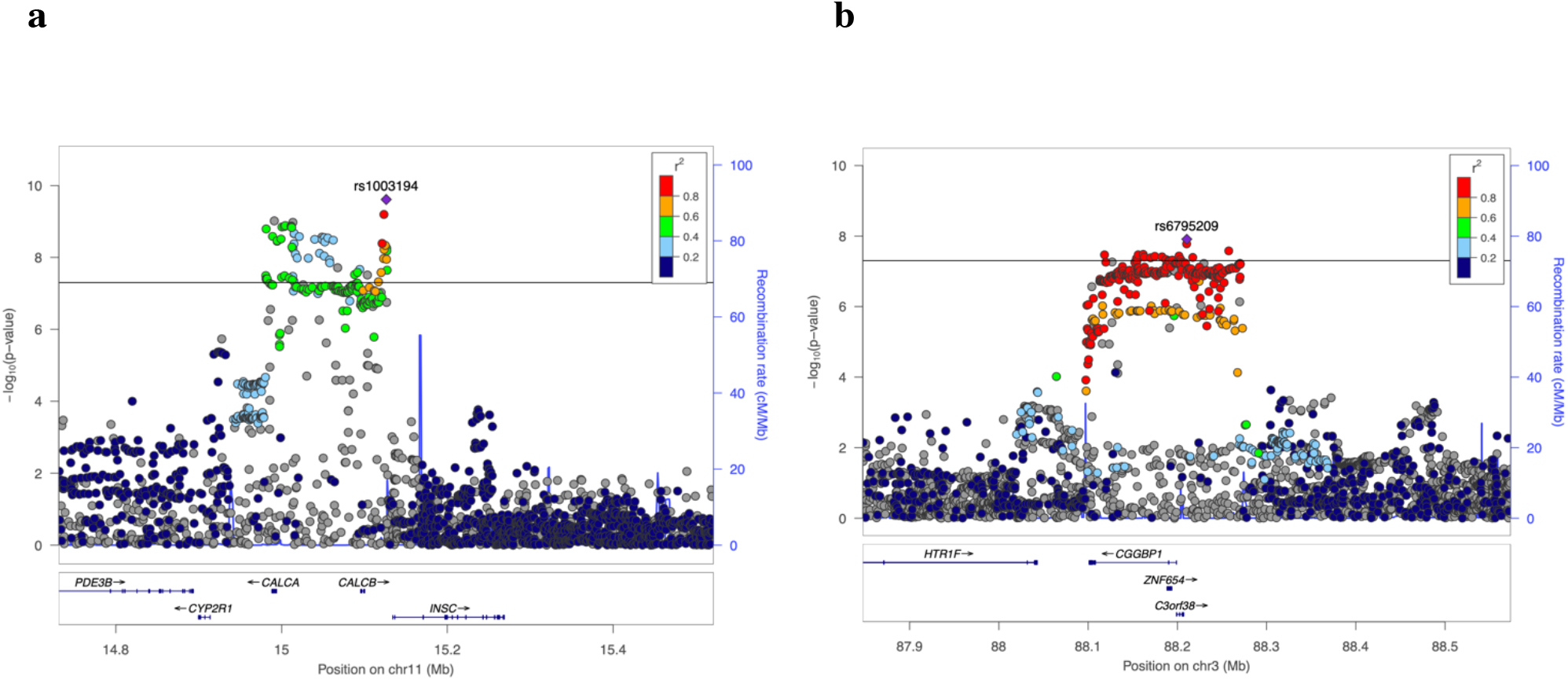
Locuszoom-plots of two novel migraine loci with genes that are targets of recent migraine specific drugs. **a**, Locus containing *CALCA* and *CALCB* genes which encode CGRP, that is the target of preventive and acute therapies via monoclonal antibodies and gepants. **b**, Locus containing *HTR1F* gene that encodes a serotonin 5-HT_1F_ receptor that is the target of acute therapies via ditans. The squared correlation to the lead variant is shown by colors based on the UK Biobank data for variants that have an effective sample size +/-20% of the lead variant’s effective sample size. Horizontal line corresponds to *P* = 5 × 10^− 8^. Blue graph shows the recombination rate.

### Migraine subtypes with aura and without aura

Previously, Gormley et al. (2016) conducted subtype-specific GWAS with 6,332 MA cases against 144,883 controls and 8,348 MO cases against 139,622 controls, and reported that 7 loci were GWS in MO but none was GWS in MA. Here, we added to the previous data 8,292 new MA and 6,707 new MO cases from headache specialist centers in Denmark and the Netherlands as well as from study collections in Iceland and UK Biobank (Table 2), for a total sample sizes of 14,624 MA cases and 703,852 controls, and 15,055 MO cases and 682,301 controls. We estimated the effect size for each subtype at the 123 lead variants of the migraine GWAS (Supplementary Tables 3b,c, Supplementary Fig. 8 and 9) and detected four GWS variants in the MA meta-analysis and 15 GWS variants in the MO meta-analysis. We also estimated a probability that the lead variant is either subtype-specific (i.e., associated only with MO or with MA but not with both), shared by both subtypes, or not associated with either subtype (Methods; Supplementary Table 11a, Supplementary Fig. 10). With a probability above 95%, three lead variants, i.e., rs12598836 in the *HMOX2* locus, rs10405121 in the *CACNA1A* locus and rs11031122 in the *MPPED2* locus, are MA-specific, while two lead variants, i.e., rs7684253 in the locus near *SPINK2*, rs8087942 in the locus near *FECH*, are MO-specific at a similar threshold. Nine lead variants were shared by MA and MO with > 95% probability (Fig. 3a). In addition to the five subtype-specific lead variants, also four other lead variants showed differences in effect size between the subtypes (*P* < 0.05/123) (Fig. 3b).

**Fig. 3:**
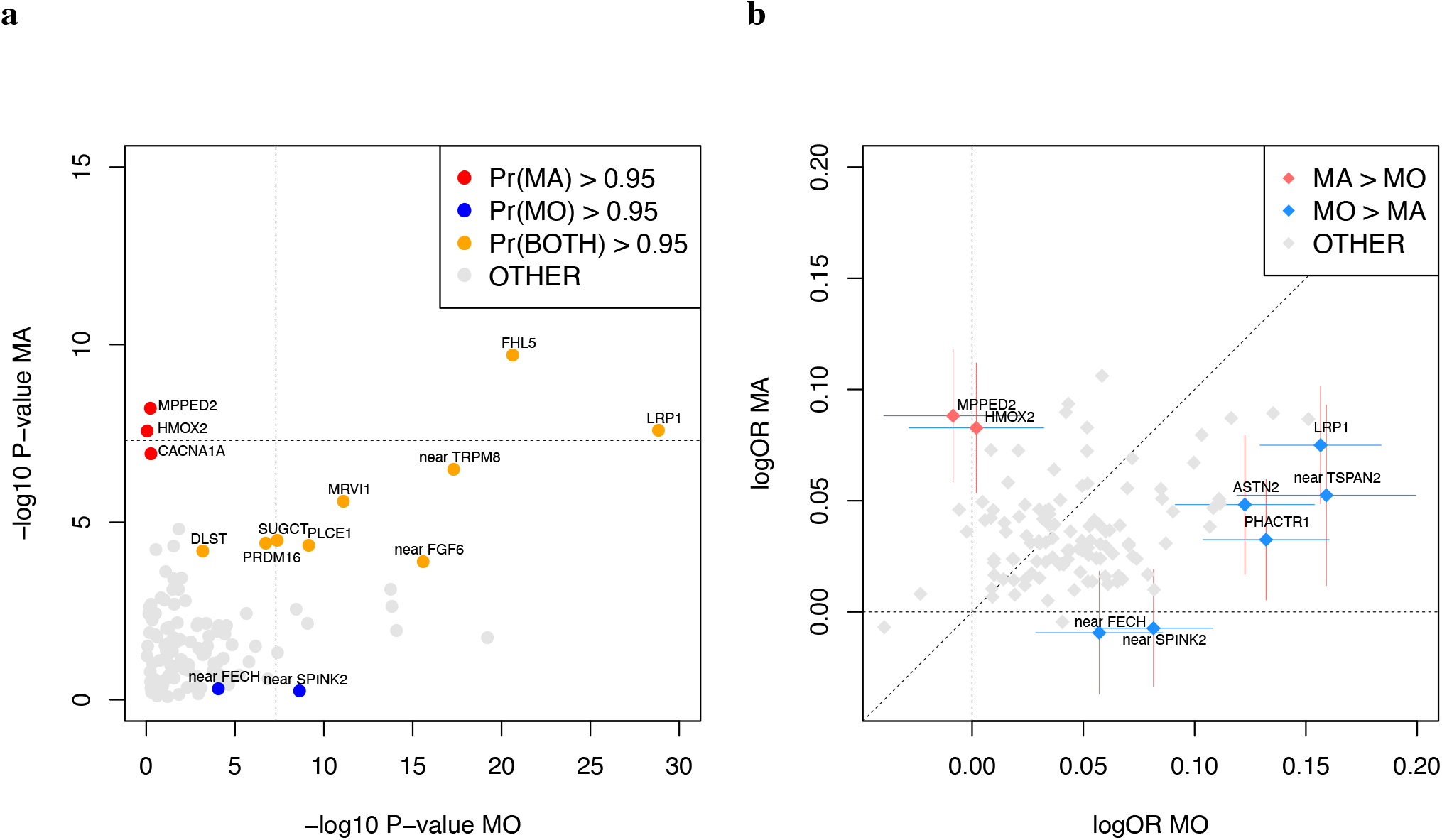
Lead variants stratified by migraine subtype for risk loci with minor allele frequency > 5%. **a**, Axes show the negative log10 *P*-value of MO (X-axis) and MA (Y-axis) analyses. Symbols that are colored and annotated indicate > 95% posterior probability that a non-zero effect is present in both MO and MA (model BOTH), or that the effect is present only in MO or only in MA but not both (models MO and MA, respectively). Variants with a probability less than 95% for each of the three models are shown as gray. **b**, Axes show logarithm of odds ratios for MO (X-axis) and MA (Y-axis) calculated for the migraine risk allele. The effects at variants that have been colored and annotated differ between the subtypes at significance level of 0.0004 = 0.05/123. The 95% confidence intervals are shown for the annotated variants. MO = migraine without aura, MA = migraine with aura.

### PheWAS with NHGRI GWAS Catalog and FinnGen R4

Next, we conducted phenome-wide association scans (PheWAS) for the lead variants for 4,314 traits with reported associations in the NHGRI GWAS Catalog (https://www.ebi.ac.uk/gwas/) and for the GWAS summary statistics of 2,263 disease traits in the FinnGen release 4 data. We identified 25 lead variants that were reported to be associated with 23 different phenotype categories (Methods) in the GWAS Catalog, and 17 lead variants with 26 defined disease categories in FinnGen at *P* < 1 × 10− 5. The categories with the highest number of reported associations were cardiovascular disease (7 lead variants) and blood pressure (6 lead variants) in GWAS Catalog, and diseases of the circulatory system (11 lead variants) in FinnGen. When we performed PheWAS for all variants in high LD (*r*^2^> 0.6) with the lead variants, we observed associations for 79 loci with 54 different phenotype categories in the GWAS Catalog, and for 41 loci with 26 disease categories in FinnGen (Supplementary Table 12a and Supplementary Fig. 11).

These findings are consistent with previous results that migraine is a risk factor for multiple cardiovascular traits, (Kurth et al., 2016), (Hippisley-Cox et al., 2017), (Adelborg et al., 2018), and genetically correlated with blood pressure (Siewert et al., 2020), (Guo et al., 2020). However, we did not observe a trend in the direction of the allelic effects between migraine and coronary artery disease (CAD) or migraine and blood pressure traits (Supplementary Table 12d), using the latest meta-analysis of CARDIoGRAMplusCD4 Consortium (Nelson et al., 2017), (N = 336,924), and blood pressure GWAS from UK Biobank (Loh et al., 2018), (N = 422,771).

### Enrichment in tissue or cell types and gene sets

We used LDSC applied to specifically expressed genes (LDSC-SEG) (Finucane et al., 2018) (Methods) to evaluate whether the polygenic migraine signal was enriched near genes that were particularly active in certain tissue or cell types as determined by gene expression or activating histone marks. Using multi-tissue gene expression data, we found enrichment at FDR 5% in three cardiovascular tissue/cell types, i.e., aorta artery (*P* = 1.78 × 10^− 4^), tibial artery (*P* = 3.60 × 10^− 4^) and coronary artery (*P* = 4.29 × 10^− 4^) (Table 3 and Supplementary Table 13a), all of which have previously been reported enriched in migraine without aura (Finucane et al., 2018). The fine-scale brain expression data from GTEx, since recently including 13 brain regions, showed enrichment in the caudate nucleus of striatum, a component of basal ganglia (*P* = 6.02 × 10^− 4^; Table 3 and Supplementary Table 13b). With chromatin-based annotations, we found enrichment at FDR 5% in five central nervous system (CNS) cell types, in three cardiovascular cell types, one cell type of the digestive system, one musculoskeletal/connective cell type, and in ovary tissue (Table 3 and Supplementary Table 13c). In addition to replicating previous findings (Gormley et al., 2016), (Finucane et al., 2018), the signal linking to ovary tissue has not been reported before.

**Table 3.** LDSC-SEG results that are significant at FDR 5%.

| Tissue/Cell type and histone mark | Tissue category | *P*-value | FDR |
| --- | --- | --- | --- |
| **Multi-tissue gene expression data** |  |  |  |
| Aorta | Cardiovascular | 1.78E-04 | 0.029 |
| Tibial Artery | Cardiovascular | 3.60E-04 | 0.029 |
| Coronary Artery | Cardiovascular | 4.29E-04 | 0.029 |
| **Gene expression data of 13 brain regions from GTEx** |  |  |  |
| Caudate (basal ganglia) | Central nervous system | 6.00E-04 | 0.008 |
| **Multi-tissue chromatin annotation data** |  |  |  |
| Fetal Brain Female, H3K4me3 | Central nervous system | 2.49E-05 | 0.012 |
| Brain Dorsolateral Prefrontal Cortex, H3K27ac | Central nervous system | 8.43E-05 | 0.018 |
| Brain Dorsolateral Prefrontal Cortex, H3K4me3 | Central nervous system | 1.11E-04 | 0.018 |
| Aorta, H3K4me1 | Cardiovascular | 2.57E-04 | 0.031 |
| Stomach Mucosa, H3K36me3 | Digestive | 3.36E-04 | 0.032 |
| Aorta, H3K27ac | Cardiovascular | 4.40E-04 | 0.032 |
| Artery-Tibial ENTEX, H3K4me1 | Cardiovascular | 4.53E-04 | 0.032 |
| Ganglion Eminence derived primary cultured neurospheres, H3K4me3 | Central nervous system | 6.53E-04 | 0.04 |
| Brain Germinal Matrix, H3K4me3 | Central nervous system | 8.42E-04 | 0.043 |
| Aorta ENTEX, H3K27ac | Cardiovascular | 1.11E-03 | 0.043 |
| Artery-Coronary ENTEX, H3K4me3 | Cardiovascular | 1.13E-03 | 0.043 |
| Cortex derived primary cultured neurospheres, H3K36me3 | Central nervous system | 1.14E-03 | 0.043 |
| Ovary, H3K27ac | Other | 1.15E-03 | 0.043 |
| Cortex derived primary cultured neurospheres, H3K4me3 | Central nervous system | 1.29E-03 | 0.045 |
| Aorta ENTEX, H3K4me1 | Cardiovascular | 1.39E-03 | 0.045 |
| Stomach Smooth Muscle, H3K4me3 | Musculoskeletal/Connective | 1.55E-03 | 0.047 |

*P*-value from testing whether the regression coefficient is positive. FDR = False discovery rate based on Benjamini-Hochberg method. Full results are in Supplementary Tables 12a-f.

Next, we used DEPICT (Pers et al., 2015) to identify tissues whose eQTLs were enriched for migraine-associated variants. The tissue enrichment analysis replicated three previously reported tissues (Gormley et al., 2016): arteries (nominal *P* = 1.03 × 10^− 3^), stomach (nominal *P* = 1.04 × 10^− 3^) and upper gastrointestinal tract (nominal *P* = 1.29 × 10^− 3^) (Supplementary Table 14a). Results of gene set analyses using DEPICT (Pers et al., 2015) and MAGMA (de Leeuw et al., 2015) are presented in Supplementary Tables 14 and 15.

## Discussion

We conducted the largest GWAS meta-analysis on migraine thus far by combining genetic data on 102,084 cases and 771,257 controls. We identified 123 migraine risk loci of which 86 are novel since the previous migraine meta-analysis that yielded 38 loci (Gormley et al., 2016). This shows that we have now reached the statistical power for rapid accumulation of new risk loci for migraine, in line with the progress of GWAS seen with other common diseases (Need and Goldstein, 2014), and as expected for a highly polygenic disorder like migraine (Gormley et al., 2018).

Migraine subtypes MO and MA were defined as separate disease entities some 30 years ago, and since then, there has been a continuous debate to what extent they are biologically similar. Over the years arguments in favour (Kallela et al., 2001) and against (Russell and Olesen, 1995) have been presented but convincing genetic evidence to support subtype-specific risk alleles has been lacking in previous genetic studies with smaller sample sizes (Anttila et al., 2013), (Chasman et al., 2014), (Nyholt et al., 2014). Here we increased considerably the evidence for subtype specificity of some risk alleles by including new migraine subtype data at the 123 migraine risk variants. We observed that, with a probability of > 95%, three lead variants (in *HMOX2*, in *CACNA1A* and in *MPPED2*) are associated with MA but not MO. Of them, *CACNA1A* is a well-known gene linked to familial hemiplegic migraine, a rare subform of MA (Ophoff et al., 1996), (de Vries et al., 2009). The observation that *CACNA1A* seems involved in both monogenic and polygenic forms of migraine provides first gene-based support for the increased sharing of common variants between the two disorders (Gormley et al., 2018). We do not find evidence that any of the seven loci, that were earlier reported GWS in MO but not in MA by Gormley et al. (2016), would be specific for MO, while four of them (*LRP1, FHL5*, near *FGF6* and near *TRPM8*) are among the nine loci shared by both subtypes with a probability over 95%. Loci (e.g., *LRP1* and *FHL5*) that are strongly associated with both subtypes provide convincing evidence for a previous hypothesis that the subtypes partly share a genetic background (Gormley et al., 2016), (Zhao et al., 2016). In accordance with our analysis, effects in both subtypes were suggested before at the *TRPM8* and *TSPAN2* loci while, in contrast to our results, the *LRP1* locus was earlier reported specific for MO (Chasman et al., 2014). Finally, we also detected four lead variants (including *LRP1*) that do not appear specific to MO but do confer a higher risk for MO than for MA.

It has been long debated whether migraine has a vascular or a neuronal origin, or whether it is a combination of both (Tfelt-Hansen and Koehler, 2011), (Jacobs and Dussor, 2016), (Anttila et al., 2018), (Hoffmann et al., 2019). Here, we found genetic evidence for the role of both vascular and central nervous tissue types in migraine from several tissue enrichment analyses, which refined earlier analyses on smaller sample sizes (Gormley et al., 2016), (Finucane et al., 2018).

With respect to a vascular involvement in the pathophysiology of migraine, both gene expression and chromatin annotation data from LDSC-SEG showed that migraine signals are enriched for genes and cell type-specific annotations that are highly expressed in aorta and tibial and coronary arteries. The involvement of arteries was also proposed by the DEPICT tissue enrichment analysis. In addition, cardiovascular disease and blood pressure phenotypes were among the top categories in the PheWAS analyses. These results are consistent with previous studies that have reported a shared etiology and some genetic correlation between migraine and cardiovascular and cerebrovascular endpoints (Bigal et al., 2009), (Malik et al., 2015), (Winsvold et al., 2015), (Adelborg et al., 2018), (Mahmoud et al., 2018), (Siewert et al., 2020), (Guo et al., 2020), (Daghlas et al., 2020). However, in our analysis, the migraine risk alleles did neither consistently increase nor consistently decrease the risk of coronary artery disease or the risk of hypertension.

A key role of the central nervous system (CNS) in migraine pathophysiology has emerged from animal models, human imaging, and neurophysiological studies (Ferrari et al., 2015), (Charles, 2018) while support for CNS involment from genetic studies has been more difficult to obtain. A likely reason is the paucity of gene expression data from CNS tissue types, but recently more data has become available making such studies feasible. Our LDSC-SEG analysis using gene expression data from 13 brain regions showed an enrichment for caudate nucleus in the basal ganglia, and with chromatin-based annotations for five CNS tissue types: dorsolateral prefrontal cortex, neurospheres derived from cortex, fetal brain, germinal matrix and neurospheres derived from ganglion eminence. Genes related to neurogenesis were also enriched for migraine-associated genes in the gene set enrichment analysis performed by MAGMA. Alterations in the structure and/or function of several brain regions (Burstein et al., 2015), (Charles, 2018), (Andreou and Edvinsson, 2019) including basal ganglia, cortex, hypothalamus, thalamus, brainstem, amygdala and cerebellum have been reported for individuals who suffer from migraine, but the cause of these changes is not known.

In addition to the support for the hypothesis that both vascular and CNS are important in migraine pathogenesis (Olesen et al., 2009), (Tfelt-Hansen and Koehler, 2011), (Charles, 2018), the tissue enrichment analyses also reported some tissue types of the digestive system as well as ovary at FDR 5%. Given the female preponderance and suggested influence of sex hormones (e.g. menstrual related migraine) in migraine (Brandes, 2006), (Borsook et al., 2014), (Delaruelle et al., 2018), the involvement of the ovary is an interesting finding, although the statistical evidence for it currently remains weaker compared to that for the vascular and central nervous systems.

A particularly interesting finding in our GWAS was the identification of risk loci containing genes that encode targets for migraine-specific therapeutics. One new locus contains *CALCA* and *CALCB* genes on chromosome 11 that encode calcitonin gene-related peptide (CGRP). CGRP is a highly potent vasodilator with a potential role in migraine headache pain (Edvinsson et al., 2018), (Hargreaves and Olesen, 2019). Several biological migraine therapeutics have recently been developed against CGRP or its receptor. In particular, CGRP-related monoclonal antibodies have been successful for the preventive treatment of migraine (Diener, 2019), and they are considered a major breakthrough in migraine-specific treatments since the development of the triptans for acute migraine over two decades ago. Another new locus contains the *HTR1F* gene that encodes serotonin 5-HT_1F_ receptor that is the target of another recent migraine drug class called ditans (de Vries et al., 2020). The 5-HT_1F_ receptors are expressed in the trigeminal ganglion, the trigeminal nucleus caudalis and cephalic blood vessels (Mitsikostas and Tfelt-Hansen, 2012). Ditans provide a promising acute treatment especially for those migraine patients that cannot use triptans because of cardiovascular risk factors (Kuca et al., 2018). These two new GWAS associations near genes that are already targeted by effective migraine drugs suggest that there could be other potential drug targets among the new loci and provide a clear rationale for future GWAS efforts to increase the number of loci by increasing sample sizes further. In addition, GWAS data with migraine subtype information can help prioritize treatment targets for particular migraine symptomatology, such as aura symptons, that currently lack treatment options. More generally, utilizing genetic evidence when selecting new drug targets is estimated to double the success rate in clinical development (Nelson et al., 2015), (King et al., 2019).

A limitation of our study is that a large proportion of migraine diagnoses is self-reported. Therefore, we cannot rule out misdiagnosis, such as, e.g., tension headache being reported as migraine, which could overemphasize genetic factors related to general pain mechanisms and not migraine *per se*. Regardless, the high genetic correlation that we observed supports a strong phenotypic concordance between the study collections that include also deeply phenotypes clinical cohorts from headache specialist centers, which were instrumental for the migraine subtype analyses. While the subtype data provided convincing evidence of both loci with genetic differences and other loci with genetic overlap between subtype, larger samples are still needed to achieve a more accurate picture of the similarities and differences in genetic architecture behind the subtypes.

To conclude, we report the largest GWAS meta-analysis on migraine to date detecting 123 risk loci. We demonstrated that both vascular and central nervous systems are involved in migraine pathophysiology, supporting the notion that migraine is a neurovascular disease. Our subtype analysis of migraine with aura and migraine without aura shows that these migraine subtypes have both shared risk alleles and risk alleles that appear specific to one subtype. In addition, new loci include two targets of recently developed and effective migraine treatments. Therefore, we expect that these and future GWAS data will reveal more of the heterogeneous biology of migraine and potentially point to new therapies against migraine that currently is a leading burden for population health throughout the world.

## Methods

### Cohorts and phenotyping

First, we performed a genome-wide meta-analysis on migraine including 5 study collections listed in Table 1 and Supplementary Table 1. Second, we performed subtype-specific meta-analyses on MA and on MO, both including 5 study collections listed in Table 2, for the 123 independent risk variants identified in the migraine analysis. A description of IHGC2016 (including 21 cohorts in the migraine meta-analysis, 12 cohorts in the MA meta-analysis and 11 cohorts in the MO meta-analysis) have been previously provided (Gormley et al., 2016). Details of the four other cohorts in the migraine meta-analysis as well as the four new cohorts that were only used in the migraine subtype analyses are shown in Supplementary Note 1. In particular, the migraine phenotype has been self-reported in other cohorts except in IHGC2016, where a subset of patients were phenotyped in specialized headache centers, as previously explained (Gormley et al., 2016).

### Quality control

Before conducting the meta-analysis, a standard quality control protocol was applied to each individual GWAS. Related individuals were removed from all other cohorts except HUNT by using an IBD cut-off of 0.185 or smaller. Multiallelic variants were excluded from all studies and only variants that satisfied the following thresholds were kept for further analysis: minor allele frequency (MAF) > 0.01, IMPUTE2 info or MACH *r*^2^ > 0.6, and, when available, Hardy-Weinberg equilibrium (HWE) *P*-value > 1×10^− 6^ and missingness < 0.05. Variants were matched by chromosome, position and alleles to the UK Biobank data. Indels were recoded as insertions (I) and deletions (D). For each study, the SNPs with an effect allele frequency (EAF) discrepancy of > 0.30 and indels with EAF discrepancy of > 0.20 to UK Biobank were excluded. MAF and EAF plots of cohorts against the reference cohort are shown in Supplementary Fig. 12. We conducted a sensitivity analysis on strand-ambiguous SNPs, i.e. A/T and G/C variants, by counting, for each pair of studies, how often the same allele of A/T or G/C variant was coded as the minor allele in both cohorts, as a function of MAF thresholds (Supplementary Table 16). Minor alleles were same at least in 97.39% of the variants without MAF threshold and the corresponding proportions were 99.96% and 79.58% when MAF < 0.25 and when MAF > 0.4, respectively. The very high concordance for variants with MAF < 0.25 suggests that the strand-ambiguous variants were consistently labelled for almost every variant. Therefore, we did not exclude any variants based on possible labelling mismatches due to strand ambiguity.

### Statistical analysis

The GWAS for the individual study cohorts were performed by logistic regression with an additive model of imputed dosage of the effect allele on the log-odds of migraine. The analyses for IHGC2016 (Gormley et al., 2016) and 23andMe (Pickrell et al., 2016) have been described before. For UKBB data and GeneRISK data, we used PLINK v2.0 (Chang et al., 2015). For HUNT data, we used a generalized logistic mixed model with the saddlepoint approximation as implemented in SAIGE v0.20 (Zhou et al., 2018) that accounts for the genetic relatedness. All models were adjusted for sex and at least for the four leading principal components of the genetic population structure (Supplementary Table 17). Age was used as a covariate when available. A detailed description is provided in Supplementary Note 1. For the chromosome X meta-analysis, male genotypes were coded as {0,2} in all cohorts, and the GWAS were conducted with an X chromosome inactivation model that treats hemizygous males as equivalent to homozygous females (Tukiainen et al., 2014).

We performed an inverse-variance weighted fixed-effect meta-analysis on the five study collections by using GWAMA (Mägi and Morris, 2010). After the meta-analysis, we excluded the variants with effective sample size N_eff_ < 2500 to remove results with very low precision compared to the majority of variants and were left with 10,843,197 variants surpassing the QC thresholds. We estimated the effective sample size for variant *i* as 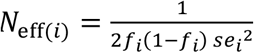, where *f*_*i*_ is the effect allele frequency for variant *i* and *se*_*i*_ is the standard error estimated by the GWAS software. This quantity approximates the value *N t (1-t) I*, where *N* is the total sample size (cases + controls), *t* is the proportion of cases and *I* is the imputation info (derivation in Supplementary Note 1).

### Risk loci

There were 8,117 genome-wide significant (GWS) variants with the meta-analysis *P*-value < 5 × 10^− 8^. For 8,067 of them, that were available in UK Biobank, an LD matrix was obtained from UK Biobank using a random sample of 10,000 individuals included in the UKBB GWAS. We defined the index variants as the LD-independent GWS variants at LD threshold of *r*^2^< 0.1 in the following way. First, the GWS variant with the lowest *P*-value was chosen, and subsequently all GWS variants that were in LD with the chosen variant (*r*^2^> 0.1) were excluded. Next, out of the remaining GWS variants, the variant with the lowest *P*-value was chosen and the GWS variants in LD with that variant were excluded. This procedure was repeated until there were no GWS variants left. Out of the 8,067 variants with LD information, 170 were LD-independent (at *r*^2^< 0.1). For 18/50 variants that were not found in UK Biobank, LD information was available from the 23andMe data, and all 18 variants were in LD (*r*^2^> 0.1) with some index variant. Two of the 18 variants (rs111404218 and rs12149936) had lower *P*-value than the original index variant they were in LD with and hence they replaced the original index variants. For 32 GWS variants, LD remained unknown. Thus, at this stage, the GWS associations were represented by 202 = 168 + 2 + 32 index variants.

Next, to define the risk loci and their lead variants, an LD block around each index variant was formed by the interval spanning all GWS variants that were in high LD (*r*^2^ > 0.6) with the index variant. Sizes of these regions ranged from 1 base pair (only the variant itself, e.g., the variants with unknown LD) to 1,089 kb. Sets of regions that were less than 250 kb away from each other were merged (distance from the end of the first region to the beginning of the second region). This definition resulted in 126 loci. All other GWS variants were included in their nearest locus based on their position and the locus boundaries were updated, and finally loci within 250 kb from each other were merged. This resulted in our final list of 123 risk loci. Each risk locus was represented by its lead variant defined as the variant with the lowest *P*-value and named by the nearest protein-coding gene to the lead variant or by the nearest non-coding gene if there was no protein-coding gene within 250 kb. The term “*Near”* was added to the locus name if the lead variant did not overlap with a gene transcript. We note that the nearest gene to the lead variant need not be a causal gene. None of the 32 variants without LD information became a lead variant of a risk locus because all had a variant in the vicinity with a smaller *P*-value.

We annotated and mapped these loci by their physical position to genes by using the Ensembl Variant Effect Predictor (VEP, GRCh37) (McLaren et al., 2016). We used two different thresholds for annotating the nearest genes: a distance of 20 kb and 250 kb to the nearest transcript of a gene. The filtered results including all variants within a gene or a regulatory element are in Supplementary Table 7b.

### Stepwise conditional analysis

We performed a stepwise conditional analysis (CA) on each risk locus by using FINEMAP v1.4. (Benner et al., 2016). FINEMAP uses GWAS summary statistics together with an LD reference panel and does not require individual-level data. The reference LD needs to accurately match the GWAS data for reliable fine-mapping (Benner et al., 2017) but a simpler stepwise CA is more robust to mismatches between the reference LD and the true in-sample LD of the GWAS data than the full fine-mapping. Since we did not have the in-sample LD from our GWAS data, we only carried out the CA and not the full fine-mapping. For the CA, we included only the SNPs, so no indels, and we used the same reference LD from the UK Biobank data as we used to define the risk loci. We restricted the CA only to the variants with a similar effective sample size (N_eff_) by using a threshold of ±10% of the N_eff_ of the lead SNP of the risk locus, because our summary statistics came from the meta-analysis where sample sizes per variant vary greatly. This was necessary since without this restriction, CA led to spurious conditional *P*-values, such as *P* < 10^− 250^, for some loci. As a consequence, for two of the loci where the lead variant was an indel, the lead variant was not included in the CA. For such regions, we checked that the new lead variant from the CA output was in LD (*r*^2^ > 0.3) with the original lead variant. For one locus (rs111404218) where the lead variant does not have LD information in the UK Biobank data, there were no GWS variants left in the CA after filtering by N_eff_. We used the standard GWS (*P* < 5 × 10^− 8^) threshold to define the secondary variants that were conditionally independent from the lead variant. The CA results are in Supplementary Tables 6a,b.

### eQTL mapping to genes and tissues

We used two data sources to map the risk variants to genes via eQTL associations. From GTEx v8 database (https://gtexportal.org), we downloaded the data of 49 tissues. We first mapped all 123 lead variants to all significant *cis*-eQTLs across tissues using the FDR cut-off of 5% as provided by the GTEx project (Aguet et al., 2019). Next, we also mapped the variants in high LD (*r*^2^ > 0.6) with the lead variants to all significant *cis*-eQTLs. Finally, we filtered the results to include only the new significant gene-tissue pairs that were not implicated by the lead variants. Results are shown in Supplementary Tables 8 and 9.

With FUMA v1.3.6 (Watanabe et al., 2017), we mapped the 123 lead variants, and the variants in high LD (*r*^2^ > 0.6) with the lead variants, to the other eQTL data repositories provided by FUMA except GTEx, i.e., Blood eQTL Browser (Westra et al., 2013), BIOS QTL browser (Zhernakova et al., 2017), BRAINEAC (Ramasamy et al., 2014), MuTHER (Grundberg et al., 2012), xQTLServer (Ng et al., 2017), CommonMind Consortium (Fromer et al., 2016), eQTLGen (Võsa et al., 2018), eQTL Cataloque (Kerimov et al., 2020), DICE (Schmiedel et al., 2018), scRNA eQTLs (van der Wijst et al., 2018) and PsychENCODE (Wang et al., 2018). Results are shown in Supplementary Tables 8 and 9.

To study whether the lead variants were enriched in any of the 49 tissues from GTEx v8, we fitted a linear regression model where the number of lead variants that are significant *cis*-eQTLs for a specific tissue was used as the outcome, and the overall number of genes with at least one significant *cis*-eQTL reported by GTEx for the tissue was the predictor (Aguet et al. 2019). We did a separate regression model for each tissue type by leaving the tissue of interest out from the model, and we used the model fitted on the other tissues for predicting the outcome variable for the tissue type of interest. Finally, we checked in which tissues the true observed number of migraine lead variants was outside of the 95% prediction intervals as given by the function ‘predict.lm(, interval=”prediction”)’ in R software. Details of the procedure are in Supplementary Note 1.

### LD Score regression

We estimated both the SNP-heritability (*h*^2^_*SNP*_) of migraine and pairwise genetic correlations (*r*_*G*_) between each pair of study collections using LDSC v1.0.0 (Bulik-Sullivan et al., 2015b) (Bulik-Sullivan et al., 2015a). SNP-heritability and genetic correlations were estimated using European LD scores from the 1000 Genomes Project Phase 3 data for the HapMap3 SNPs, downloaded from https://data.broadinstitute.org/alkesgroup/LDSCORE/. From the meta-analysis association statistics, we excluded variants that did not match with the HapMap3 SNPs, that had strand ambiguity (i.e., A/T or G/C SNPs), MAF < 0.01, INFO < 0.6 or missingness more than two-thirds of the 90^th^ percentile of the total sample size, that resided in long-range LD regions (Price et al., 2008), in centromere regions or in the major histocompatibility locus (MHC) of chromosome 6, leaving 1,165,201 SNPs for the LDSC analyses. We used a migraine population prevalence of 16% and a sample proportion of cases of 11.7% = 102,084/(102,084 + 771,257) to turn the LDSC slope into the estimate of *h*^2^_*SNP*_ on the liability scale (Lee et al., 2011). Pairwise genetic correlation results are listed in Supplementary Table 2. We note that in the previous migraine meta-analysis (Gormley et al. 2016), LDSC reported *h*^2^_*SNP*_ value of 14.6% (13.8 – 15.5%), which was considerably larger than the value 11.2% (10.8 – 11.6%) that we report in our analysis. When we ran our LDSC pipeline on the data of Gormley et al. (2016), we estimated *h*^2^_*SNP*_ value of 10.6% (10.1 – 11.1%). Thus, it seems that our liability transformation estimates lower values of heritability than the transformation used by Gormley et al. (2016).

### Stratified LD Score regression

We used stratified LD Score regression (S-LDSC) to partition the SNP heritability by functional genomic annotations (Finucane et al., 2015). We used the baseline model that contains 24 main functional categories including conserved, coding and regulatory regions of the genome and different histone modifications. We used the same QC as with the univariate LDSC, and the baseline European LD scores estimated from the 1000 Genomes Project Phase 3, downloaded from https://data.broadinstitute.org/alkesgroup/LDSCORE/. Results are listed in Supplementary Tables 10a,b.

### Subtype analyses of migraine with and without aura

First, we combined new MA and MO data (Table 2) with the previously used migraine subtype-specific meta-analysis data (Gormley et al., 2016), and estimated migraine subtype-specific effect sizes for the 123 lead variants from the migraine meta-analysis. We tested how often the direction of allelic effects was similar between the IHGC MA/MO and the new cohorts using a binomial test (Supplementary Table 11b). Next, we stratified the lead variants by using the information from the migraine subtype-specific analyses. For each of the variants, we estimated probabilities between four possible explanations of the observed data that we call ‘NULL’, ‘MO’, ‘MA’ and ‘BOTH’. Under model NULL the effect is not present in either of the migraine subtypes (i.e., the effect is zero); under model MO or MA the effect is present only in MO or only in MA but not in both; and under model BOTH, a non-zero effect is shared by both MO and MA. We used a Bayesian approach for model comparison that combines a bivariate Gaussian prior distribution on the two effect sizes with a bivariate Gaussian approximation to the likelihood using GWAS summary statistics (Trochet et al., 2019). Across all models, the prior standard deviation for the effect is 0.2 on the log-odds scale for non-zero effects and 0 for a zero effect. The bivariate priors for the four models are as follows: NULL assumes a zero effect in both migraine subtypes, MO and MA assume a non-zero effect for one subtype and a zero effect for the other subtype, and BOTH combines the fixed-effect model (exactly the same effect in both subtypes) with the independent-effects model (the two effect sizes are non-zero but uncorrelated with each other) with equal weights. Finally, we assumed that each of the four models (NULL, MO, MA, BOTH) is equally probable *a priori*, which we considered an appropriate assumption since all these variants show a convincing association to overall migraine (*P* < 5 × 10^− 8^). Then we used the Bayes formula to work out the posterior probability on each model. The results are shown in Fig. 3a, thresholded by a probability cut-off of 95% and in Supplementary Table 11a. The correlation parameter between MO and MA GWAS statistics needed in the bivariate likelihood approximation was estimated to be 0.148 using the empirical Pearson correlation of the effect size estimates of the common variants (MAF > 0.05) that did not show a strong association to either of the migraine subtypes (*P* > 1 × 10^− 4^) (Cichonska et al., 2016). We tested whether the effect sizes between MA and MO were equal at a Bonferroni corrected significance threshold of *α* = 0.05/123 by using a normal approximation and accounting for the correlation in effect size estimators.

We note that the amount of information in the data (“statistical power”) is taken automatically into account in this model comparison, which we consider an advantage compared to a comparison of the raw *P*-values between the subtype analyses that does not automatically account for statistical power. We want to point out that the inference in the model comparison approach is conditional on the particular set of models being included in the comparison as well as on the particular choice of the prior distributions.

### PheWAS with NHGRI GWAS Catalog and FinnGen R4

We performed phenome-wide association studies (PheWAS) for the 123 lead variants using the NHGRI GWAS Catalog and the FinnGen R4 GWAS summary statistics. In addition, we performed the same lookups for the 123 risk loci including all variants in high LD (*r*^2^ > 0.6) with the lead variants. With the GWAS Catalog, we first downloaded all the available results (4,314 traits) from the GWAS Catalog webpage (accessed 6.4.2020). Next, we obtained all the associations for the 123 risk loci with all the high LD variants included using *P*-value thresholds of *P* < 1 × 10^− 5^, *P* < 1 × 10^− 6^ and *P* < 1 × 10^− 4^ (Supplementary Tables 12a-c). Because the GWAS Catalog includes results from several different GWAS for the same phenotype or for a very similar phenotype with a different name, we divided the phenotype associations into broader categories. The new categories are listed in Supplementary Table 18. The same approach was used for the PheWAS of FinnGen R4. We first downloaded all the available summary statistics (2,263 endpoints), and next, obtained all the associations for the 123 risk loci using the same three *P*-value thresholds as with the GWAS Catalog (Supplementary Tables 12a-c). We also divided similar endpoints into broader categories that are listed in Supplementary Table 19.

We tested the direction of allelic effects between migraine and the following three traits that shared multiple associated variants with migraine: coronary artery disease (CAD) (Nelson et al., 2017), diastolic blood pressure (Loh et al., 2018) and systolic blood pressure (Loh et al., 2018). We first took all migraine lead variants that were available also in the summary statistics of the other trait without any *P*-value threshold, and used a binomial test to test whether the proportion of variants with same direction of effects was 0.5. Next, we used a *P*-value threshold of 1 × 10^− 5^ for the association with the other trait. Results are in the Supplementary Table 12d.

### LD Score regression applied to specifically expressed genes (LDSC-SEG)

We used LD Score regression applied to specifically expressed genes (LDSC-SEG) (Finucane et al., 2018) to identify tissues and cell types implicated by the migraine GWAS results. LDSC-SEG uses gene expression data and GWAS results from all variants together with an LD reference panel. For our analyses, we used the same QC as for the other LDSC analyses and six different sets of readily constructed annotation-specific LD scores downloaded from https://data.broadinstitute.org/alkesgroup/LDSCORE/LDSC_SEG_ldscores/: multi-tissue gene expression, multi-tissue chromatin, GTEx brain, Cahoy, Corces ATAC and ImmGen LD Scores. FDR was controlled by the Benjamini-Hochberg method. The results are in Supplementary Tables 13a-f. There were no significant results with the Cahoy, Corces ATAC and ImmGen data at FDR 5%.

### MAGMA (Multi-marker Analysis of GenoMic Annotation)

We applied MAGMA v1.07 (de Leeuw et al., 2015) to identify genes and gene sets associated with the migraine meta-analysis results. First, we mapped the meta-analysis SNPs to 18,985 protein-coding genes based on their physical position in the NCBI 37 build by using default settings of MAGMA. Next, we performed a gene-based analysis using the default SNPwise-mean model and the same UK Biobank LD reference as for the other analyses. We applied a Bonferroni correction (*α* = 0.05/18,985) to identify significantly associated genes for migraine with the results listed in Supplementary Table 15a. Finally, we used the results from the gene-based analysis to perform a gene-set analysis by using two different gene-set collections from the Molecular Signature Database v.7.0 (Liberzon et al., 2011), (Subramanian et al., 2005): the curated gene sets containing 5,500 gene sets and the GO gene sets containing 9,988 gene sets. The gene-set analysis was performed using the competitive gene set model that tests whether the genes in the gene-set are more strongly associated with the phenotype compared to the other genes. To correct for multiple testing, we used a Bonferroni correction (*α* = 0.05/(5,500+9,988)). Results are in Supplementary Tables 15b,c and in Supplementary Fig. 13.

### DEPICT (Data-driven Expression Prioritized Integration for Complex Traits)

DEPICT (Pers et al., 2015) is an integrative tool to identify the most likely causal genes at associated loci, and enriched pathways and tissues or cell types in which the genes from the associated loci are highly expressed. As an input, DEPICT takes a set of trait-associated SNPs. First, DEPICT uses co-regulation data from 77,840 microarrays to predict biological functions of genes and to construct 14,461 reconstituted gene sets. Next, information of similar predicted gene functions is used to identify and prioritize gene sets that are enriched for genes in the associated loci. For the tissue and cell type enrichment analysis, DEPICT uses a set of 37,427 human gene expression microarrays. We used DEPICT v1.194, and ran the analyses twice for each of the *P*-value thresholds for clumping, as recommended in (Pers et al., 2015), and using the default settings of 500 permutations for bias adjustment and 50 replications for the FDR estimation and for the *P*-value calculation. As an input, we used only the autosomal SNPs and the same UK Biobank LD reference data as for the other analyses. First, we ran the analysis using a clumping *P*-value threshold of 5 × 10^− 8^ that resulted in 165 clumps formed from 7,672 variants (Supplementary Tables 14d-f). Second, we used a *P*-value threshold of 1 × 10^− 5^ leading to 612 clumps formed from 22,480 variants (Supplementary Tables 14a-c).

## Data Availability

Results for 8,117 genome-wide significant SNP associations (P < 5x10-8) from the meta-analysis including 23andMe data will be available on the International Headache Genetics Consortium (IHGC) website (http://www.headachegenetics.org/). Genome-wide summary statistics for the four other study collections except 23andMe will be available as described on the IHGC website. The full GWAS summary statistics for the 23andMe discovery data set will be made available through 23andMe to qualified researchers under an agreement with 23andMe that protects the privacy of the 23andMe participants. Please visit research.23andme.com/collaborate/#publication for more information and to apply to access the data.

https://research.23andme.com/collaborate/#publication

**Supplementary Tables 1-19**. MANU_SUPP_TABLES_all6f.xlsx

**Supplementary Figures**: Supplementary_Figures_a4.docx

## Data access

Results for 8,117 genome-wide significant SNP associations (*P* < 5 × 10^− 8^) from the meta-analysis including 23andMe data are available on the International Headache Genetics Consortium (IHGC) website (http://www.headachegenetics.org/). Genome-wide summary statistics for the four other study collections except 23andMe are available as described on the IHGC website. The full GWAS summary statistics for the 23andMe discovery data set will be made available through 23andMe to qualified researchers under an agreement with 23andMe that protects the privacy of the 23andMe participants. Please visit research.23andme.com/collaborate/#publication for more information and to apply to access the data.

## Acknowledgements

We thank the study participants for their contribution to this research. We also thank the numerous individuals who contributed to sample collection, storage, handling, phenotyping, and genotyping for each of the individual cohorts. We acknowledge the participants and investigators of the FinnGen study. This research has been conducted using the UK Biobank Resource under Application Number 22627. A list of study-specific acknowledgements and funding information can be found in the Supplementary Note 1.

