## Supplementary Note. for "Genome-wide analysis of 102,084 migraine cases identifies 123 risk loci and subtype-specific risk alleles"

### Supplementary Note 1

#### Study descriptions

Our primary genome-wide meta-analysis on migraine included 5 study collections: IHGC2016, 23andMe, UKBB, GeneRISK and HUNT All-in Headache. In addition, we performed subtype-specific analyses for migraine with aura (MA) and migraine without aura (MO) for the 123 lead variants of the migraine risk loci identified in the primary meta-analysis by using data from 5 study collections: IHGC2016, UKBB, deCODE, DBDS, and LUMINA. All these study collections are described below.

#### 23andMe

23andMe cohort includes 53,109 migraine cases and 220,876 controls. 23andMe migraine GWAS was performed by a personal genomics company 23andMe, Inc. (https://www.23andme.com/) and detailed description of the migraine GWAS is provided elsewhere (Pickrell et al., 2016). All participants have provided informed consent and filled an online survey according to 23andMe's human subjects protocol, which was reviewed and approved by Ethical & Independent Review Services, a private institutional review board. Briefly, migraine cases were assessed from the participants that had reported migraine or answered "yes" to any of the questions related to migraine, and controls from participants that did not report having migraine or answered "No" to all of the questions related to migraine, excluding participants with discordant answers. The GWAS was conducted for a set of unrelated individuals with European ancestry by logistic regression assuming additive allelic effects and controlled for age, sex and five principal components of genetic ancestry. The relatedness exclusion was based on an analysis of a segmental identity-by-descent algorithm, and the cut-off was chosen to be the minimal expected amount of sharing between first cousins. Besides the variant QC reported in (Pickrell et al., 2016), we excluded also duplicated variants and variants with MAF $\leq 0.01$ or average Minimac2 $r^2 \leq 0.6$.

#### GeneRISK

GeneRISK cohort includes 1,084 migraine cases and 4,857 controls from the GeneRISK Study. The GeneRISK Study is a prospective observational study that focuses on genetic risk factors of cardiovascular diseases. Participants for the recruitment were randomly selected from 45 to 65 years old individuals from Southern Finland, and the data were collected during 2015-2018.

Exclusion criteria for the participation were diagnosed stroke, myocardial infarction, cerebral hemorrhage, cerebral infarction, coronary heart disease, or if the participant had undergone coronary artery bypass surgery or coronary angioplasty, or if the participant was pregnant, or had a guardian of interest. All study participants attended to a clinical health check-up and filled an electronic questionnaire that included several topics related to their health status and lifestyle factors. Migraine cases were assessed from participants who answered "Yes" to the question of "Have you been diagnosed migraine by doctor? Yes/No", and controls from participants who answered "No". The samples were genotyped by using Illumina HumanCoreExome arrays (HumanCoreExome24 v1.0 or HumanCoreExome24 v1.1) and ancestry were inferred by principal component analysis using PLINK v2.0. Genotype data were imputed using a population specific reference panel SISu v3 with Beagle 4.1 version 08Jun17.d8b (Browning and Browning, 2007) as described here: dx.doi.org/10.17504/protocols.io.nmndc5e. Related individuals were removed using a threshold of IBD > 0.1 such that individuals who had many relatives were preferred for exclusion. Genetic analysis was run by PLINK v2.0 by applying logistic regression with firth-fallback -mode and controlling for sex, age and first 10 principal components of genetic ancestry. For the meta-analysis, we included only biallelic variants with MAF > 0.01, INFO > 0.6, HWE $P$-value > $1\times10^{-6}$ and missingness < 0.05

All study participants have given their informed consent, and the study protocol has been approved by the Ethical Committee of the Helsinki and Uusimaa Hospital district.

#### HUNT All-in Headache

Sample ascertainment and phenotype definition. The Trøndelag Health Study (HUNT) consists of three different population-based health surveys conducted in the county of Nord-Trøndelag, Norway over approximately 20 years (HUNT1 [1984-1986], HUNT2 [1995-1997] and HUNT3 [2006-2008]) (Krokstad et al., 2012). At each survey, the entire adult population (≥ 20 years) was invited to participate by completing questionnaires, attending clinical examinations and interviews. Participation rates in HUNT1, HUNT2 and HUNT3 were 89.4% (n=77,212), 69.5% (n=65 237) and 54.1% (n=50 807), respectively (Krokstad et al., 2012). Taken together, the study included more than 120,000 different individuals from Nord-Trøndelag County. Biological samples including DNA have been collected for approximately 70,000 participants. The HUNT Study has been described in more detail elsewhere (Krokstad et al., 2012). For the present study, we included participants from HUNT2 and HUNT3. Migraine was assessed using questionnaires, and based on a modified version of the International Classification of Headache Disorders (ICHD II) (Headache Classification Subcommittee of the International Headache, 2004). Subjects who answered "yes" to the question "Have you suffered from headache during the last 12 months?" were classified as headache sufferers. Those who answered "no" in one or both studies, and "yes" in neither, constitute the control group. Based on the subsequent headache questions (Hagen et al., 2000), (Hagen et al., 2010) headache sufferers were classified as having migraine if they fulfilled the following 3 criteria: (1) headache attacks lasting 4 to 72 hours, (< 4 hours was accepted for those who reported commonly occurring visual disturbances before headache); (2) headache with at least one of the following characteristics: pulsating quality, unilateral location, or aggravation by physical activity; (3) during headache, at least one of the following occurred: nausea, photophobia and phonophobia. In addition, the participants were asked if they suffered from migraine; those who responded positively to this question were also included in the migraine group. For the present study, subjects were classified as having migraine if they fulfilled the criteria for migraine in either HUNT2 or HUNT3. The migraine diagnoses have been validated by clinical interviews performed by neurologists. In HUNT2, the sensitivity was 69% and specificity 89% ($\kappa$= 0.59, 95% CI 0.47–0.71) (Hagen et al., 2000). In HUNT3 the sensitivity was 49% and specificity 96% ($\kappa$ = 0.51, 95% CI 0.34–0.68) (Hagen et al., 2010).

Genotyping, quality control and imputation. In total, DNA from 71,860 HUNT samples was genotyped using one of three different Illumina HumanCoreExome arrays (HumanCoreExome12 v1.0, HumanCoreExome12 v1.1 and UM HUNT Biobank v1.0). Samples that failed to reach a 99% call rate, had contamination > 2.5% as estimated with BAF Regress (Jun et al., 2012), large chromosomal copy number variants, lower call rate of a technical duplicate pair and twins, gonosomal constellations other than XX and XY, or whose inferred sex contradicted the reported gender, were excluded. Samples that passed quality control were analyzed in a second round of genotype calling following the Genome Studio quality control protocol described elsewhere (Guo et al., 2014). Genomic position, strand orientation and the reference allele of genotyped variants were determined by aligning their probe sequences against the human genome (Genome Reference Consortium Human genome build 37 and revised Cambridge Reference Sequence of the human mitochondrial DNA; http://genome.ucsc.edu) using BLAT (Consortium, 2012). PLINK v1.90 (Chang et al., 2015) was then used to exclude variants if their probe sequences could not be perfectly mapped, cluster separation was < 0.3, Gentrain score < 0.15, showed deviations from Hardy Weinberg equilibrium in unrelated samples of European ancestry with $P$-value < 0.0001), had a call rate < 99%, or another assay with higher call rate genotyped the same variant. Ancestry of all samples was inferred by projecting all genotyped samples into the space of the principal components of the Human Genome Diversity Project (HGDP) reference panel (938 unrelated individuals; downloaded from http://csg.sph.umich.edu/chaolong/LASER/) (Wang et al. 2014), (Li et al., 2008) using PLINK. Recent European ancestry was defined as samples that fell into an ellipsoid spanning exclusively European populations of the HGDP panel. The different arrays were harmonized by reducing to a set of overlapping variants and excluding variants that showed frequency differences > 15% between data sets, or that were monomorphic in one and had MAF > 1% in another data set. The resulting genotype data were phased using Eagle2 v2.3 47.

Imputation was performed on the 69,716 samples of recent European ancestry using Minimac3 (v2.0.1, http://genome.sph.umich.edu/wiki/Minimac3) (Das et al., 2016) with default settings (2.5 Mb reference based chunking with 500kb windows) and a customized Haplotype Reference consortium release 1.1 (HRC v1.1) for autosomal variants and HRC v1.1 for chromosome X variants (McCarthy et al., 2016). The customized reference panel represented the merged panel of two reciprocally imputed reference panels: (1) 2,201 low-coverage whole-genome sequences samples from the HUNT study and (2) HRC v1.1 with 1,023 HUNT WGS samples removed before merging. We excluded imputed variants with $r^2 < 0.3$ resulting in over 24.9 million well-imputed variants. After restricting to those with available phenotype information, and after excluding 2,697 HUNT participants that were genotyped and included in a previous GWAS meta-analysis of migraine (Gormley et al., 2016), 40,224 individuals (7,801 cases and 32,423 controls) were included in the analysis.

<u>Association analysis.</u> Association analyses were conducted using SAIGE [Efficiently controlling for case-control imbalance and sample relatedness in large-scale genetic association studies (Zhou et al., 2018)], a generalized mixed effects model approach, to account for cryptic population structure and relatedness when modelling the association between genotype probabilities (dosages) and migraine. Models were adjusted for sex, age, genotyping batch and four principal components (PCs). PCs were computed using PLINK. Additional filters applied to the analysis included minor allele count ≥ 1 and imputation $r^2 \geq 0.3$. For the meta-analysis, we included only biallelic variants with MAF > 0.01, imputation $r^2 > 0.6$ and HWE $P$-value > $1\times10^{-6}$.

Written informed consent was obtained from all participants, and The Regional Committee for Medical and Health Research Ethics approved the study (ref. 2015/576).

#### IHGC2016

IHGC2016 study collection includes 21 cohorts from (Gormley et al., 2016) meta-analysis excluding 23andMe cohort. IHGC2016 MA includes 12 cohorts, and IHGC2016 MO includes 11 cohorts. A detailed cohort and phenotype description of the included 21 cohorts are provided in the supplementary material of (Gormley et al., 2016).

#### UKBB

UKBB cohort includes 10,881 migraine cases and 330,170 controls from UK Biobank. UKBB MA cohort includes 1,333 MA cases and 320,139 controls, and UKBB MO cohort includes 187 MO cases and 320,139 controls from UK Biobank. The UK Biobank project is a population-based prospective cohort study, that consist of over 500,000 participants aged 40-69 at recruitment collected from several regions across the United Kingdom. Participants completed questionnaires and attended interviews and clinal examinations by a trained staff member. Detailed description of UK Biobank is provided elsewhere (Bycroft et al., 2018), and all detailed genotyping, quality control and imputation procedures are described at the UK Biobank website (https://www.ukbiobank.ac.uk/). Migraine cases were selected from the self-reported non-cancer illness code (Data field 20002, code 1265). This field includes participants who had self-reported migraine on the questionnaire and participants who had been uncertain about the illness they had had, but have been classified as having migraine by a trained nurse during the verbal interview after describing their symptoms. The control group was selected from the participants without self-reported migraine and without self-reported headache (Data field 20002, code 1436). MA and MO cases were compiled from the following data fields: GP clinical event records (42040, codes 14740, F260., .F351, X007J, X007K, X007N, XaXkv, F26y0, X007L, X007M for MA, and code F261.. for MO), ICD-10 main summary diagnoses (41270, code G431 for MA and code G430 for MO) and ICD-9 main summary diagnoses (41203, code 3460 for MA and code 3461 for MO). Shared control group for MA and MO were selected from the participants who did not have any of the migraine diagnoses listed above, other migraine diagnosis in the Data Field 42040 or self-reported migraine or headache in Data field 20002. We restricted the sample to include only individuals who were of European ancestry and were classified as 'White British' (Bycroft et al., 2018), and we excluded related individuals using a kinship value of 0.0442 corresponding to excluding 3rd degree relatives.

The kinship values were obtained from UK Biobank and had been computed using KING (Manichaikul et al., 2010). We ran the GWAS by PLINK v2.0 by applying logistic regression with firth-fallback -mode and controlling for sex, age and first 10 principal components of genetic ancestry. We included only biallelic variants with MAF > 0.01, INFO > 0.6, HWE *P*-value > $1\times10^{-6}$ and missingness < 0.05.

UK Biobank received ethical approval from the North West Multi-centre Research Ethics Committee (MREC) and informed consent has been obtained from participants.

#### DBDS

The DBDS Genomic Cohort contains 7,694 migraine cases (3,756 migraine without aura and 3,938 migraine with aura) and 28,045 controls from the Danish Blood Donor Study. The DBDS is a population-based study that consists of over 100K blood donors comprising extensive phenotype data based on questionnaires and whole genome genotyping data (Hansen et al., 2019). Migraine diagnosis was based on a self-reported migraine-questionnaire enabling diagnosis of migraine with and without aura in accordance with the ICHD 3 criteria. Genotyping was performed at deCODE genetics using the Global Screening Array by Illumina and imputed using a pan-Scandinavian whole genome sequencing sample of size > 8, 000 as reference including > 6,000 samples of Danish descent. Non-European participants were excluded based on principal component analysis comparing to 1000G data. Related individuals were excluded based on proportion IBD of 0.125. Genetic variants were removed using PLINK v2.0 based on MAF < 0.05, HWE *P*-value < $1x10^{-5}$, missingness per variant > 0.05 and per individual > 0.1. Of the 123 independent risk loci, 104 were present in the DBDS data. The 104 loci were tested for association with MO and MA using PLINK v2.0 by applying logistic regression (MO vs controls, and MA vs controls) and controlling for sex, age and the first ten principal components.

Written informed consent was obtained from all participants in DBDS. DBDS has secured necessary permissions and approval from the Danish Data Protection Agency (2007-58-0015) and the Scientific Ethical Committee system (M-20090237).

#### deCODE

The deCODE cohorts of migraine without aura (MO, N = 1,736) and migraine with aura (MA, N = 3,376) were established from participants in an ongoing study on the genetics of migraine in Iceland who were (A) diagnosed by neurologists (with ICD10 G43.0 (MO) or ICD10 G43.1 (MA), respectively), (B) or identified through a survey of the larger deCODE migraine cohort that also includes unsubtyped migraine cases (ICD10 G43), migraineurs diagnosed by primary care physicians (ICD10 G43 or ICPC-2 N89), cases identified through prescription data from the National Drug Database (with at least two prescriptions for anti-migraine drugs, Triptans) and cases defined by answers to the third edition of the deCODE Migraine Questionnaire (DMQ3) designed for use in genetic studies (Kirchmann et al., 2006). From these, we included in the MO/MA case groups individuals who responded "yes" to either of recent (2020) survey questions "Has a physician diagnosed you with migraine without aura? " or "Has a physician diagnosed you with migraine with aura?". For both MO and MA, we compared cases to population controls, excluding from controls all other migraineurs as defined above (in B).

The MO/MA GWAS were performed using 37.6 million high-quality sequence variants by whole-genome sequencing of Icelanders by methods that have been described in detail elsewhere (Gudbjartsson et al., 2015a), (Jónsson et al., 2017). In short, the 37.6 million variants were identified by sequencing 49,962 Icelanders using GAIIx, HiSeq, HiSeqX, and NovaSeq Illumina technology to a mean depth of at least 17.8×. SNPs and indels were identified and their genotypes called using joint calling with Graphtyper (Eggertsson et al., 2019). Additionally, over 165,000 Icelanders (including all sequenced Icelanders) have been genotyped using various Illumina SNP chips and phased using long-range phasing (Kong et al., 2008), which allows for improving genotype calls using the information about haplotype sharing. The genotypes of the high-quality sequence variants were imputed into the chip-typed Icelanders (Gudbjartsson et al., 2015b). To increase the sample size and power to detect associations, the sequence variants were also imputed into relatives of the chip-typed using genealogic information. All tested variants had imputation information over 0.8. Association analyses were performed using LD score regression to account for distribution inflation due to cryptic relatedness and population stratification. They were also adjusted for gender, age, county of origin, current age or age at death (first and second order term included), blood sample availability for the individual, and an indicator function for the overlap of the lifetime of the individual with the time span of phenotype collection.

Approval for these studies was provided by the National Bioethics Committee (approval no. VSN 19-158). All participants who donated blood signed informed consent. The personal identities of participants were encrypted using a third-party system approved and monitored by the Icelandic Data Protection Authority.

#### LUMINA

Selection of Dutch cases from LUMINA and controls from NEO.

For the follow-up association study to validate the 123 top SNPs of the meta-analysis, additional migraine cases, that is the Dutch MO and Dutch MA studies described below, were recruited and genotyped as part of the Leiden University Migraine Neuro Analysis (LUMINA) cohort (Gormley et al., 2016). The Dutch MO study contains 1,220 Dutch MO patients that were recruited through our specialized headache clinic. After genotyping and standard GWA study quality control procedures 1,116 cases remained for the analysis. Of the 1,116 Dutch MO patients, 136 (12.2%) were male and 980 (87.8%) were female. The Dutch MA study contains 766 Dutch MA patients that were recruited through our specialized headache clinic. After genotyping and standard GWA study quality control procedures, 724 cases remained for the analysis. Of the 724 MA cases, 130 (18.2%) were male and 594 (82.0%) were female.

Self-reported migraineurs were recruited via the project's website (www.lumc.nl/hoofdpijn). A set of screening questions validated previously in a population-based study was used (Launer et al., 1999). Participants fulfilling the screening criteria then completed an extended questionnaire that focuses on signs and symptoms of migraine headache and aura (aura symptoms were absent in the selected MO patient group) as outlined in formerly ICHD-II, now ICHD-III (Headache Classification Subcommittee of the International Headache, 2004), (Headache Classification Committee of the International Headache Society (IHS), 2018). Individual diagnoses were made using an algorithm based on these criteria, validated by a semi-structured telephone interview performed by experienced physicians or by well-trained medical students, when necessary in consultation with a neurologist specialized in headache (GMT) (van Oosterhout et al., 2011). A subset of patients was asked to participate upon visiting our outpatient clinic. The LUMINA project was approved by the medical ethics committee of the Leiden University Medical Center. All respondents provided written informed consent.

For the association study to validate the 123 top SNPs of the meta-analysis, population-matched controls were obtained from the Netherlands Epidemiology of Obesity Study (NEO) (N = 5,644) (de Mutsert et al., 2013). A subset of this cohort (N = 1,671 before QC) not selected on any clinical parameter (except age that ranged from 45 to 65 years) living in a nearby municipality (Leiderdorp, The Netherlands) was used for the final analysis. Written informed consent was obtained from all participants, and the local ethics committee approved the study.

Genotyping, quality control, and imputation: Genomic DNA was extracted from peripheral blood leukocytes according to standard protocols and genotyping of both cases and controls was performed on InfiniumCoreExome-24v1-1_A beadchip. Cases were genotyped at FIMM Technology Centre (Helsinki, Finland) and controls at the Centre National de Génotypage (Paris, France). For the cases, variant calling was performed with Genome Studio v.2011.1 following a standard quality protocol. For quality control (QC), markers with high missingness rates (≥ 2%), and those failing the Hardy-Weinberg equilibrium were excluded. Individuals were excluded if they had a high proportion of missing genotype data (≥ 2%), inconsistent sex information, were related (PI-HAT ≥ 0.2), were ancestry outliers, or heterozygosity outliers. Principal component analysis (PCA) was performed on the pruned data set (with a 50-kb sliding window, $r^2 > 0.2$) using PLINK and population outliers were excluded. After combining the genotyped SNP information from cases and controls imputation was performed on the Michigan Imputation Server using Haplotype Reference Consortium (HRC v1.1 2016) as a reference panel after phasing by Eagle (v2.3), (Das et al., 2016), (Loh et al., 2016) using the default parameters.

For the MO dataset a total of 239,606 SNPs from 6,794 individuals (1,150 cases and 5,644 controls) was available for imputation. All cases (N = 1,116) and the NEO Leiderdorp control subset (N = 1,445; 630 male and 815 female), whom had passed previous QC steps were extracted for analysis. The analysis was based on a total of 16,180,654 SNPs.

For the MA dataset a total of 316,010 SNPs from 6,394 individuals (741 cases and 5,653 controls) were available for imputation. All cases (N = 724) and the NEO Leiderdorp control subset (N = 1,447; 629 male and 818 female), whom had passed previous QC steps were extracted for analysis. The analysis was based on a total of 15,336,357 SNPs.

For both the MO and the MA sample an association analysis was performed using a logistic regression model implemented in SNPTEST (version 2.5.2) for autosomal variants,(Marchini et al., 2007) with case-control status as outcome and assuming additive allelic effects. The model was adjusted for sex and the first ten principal components.

#### Study-specific acknowledgements

**23andMe:** We would like to thank the research participants and employees of 23andMe for making this work possible.

**DeCODE**: The financial support from the European Commission to the NeuroPain project (FP7#HEALTH-2013-602891-2) and painFACT project (H2020-2020-848099), and the National Institutes of Health (R01DE022905) is acknowledged.

**GeneRISK**: We thank the study participants for their contribution to this research. The GeneRISK study was funded by Business Finland through the Personalized Diagnostics and Care program coordinated by SalWe Ltd (Grant No 3986/31/2013).

**HUNT**: The Trøndelag Health Study (HUNT) is a collaboration between the HUNT Research Center (Faculty of Medicine and Health Sciences, NTNU, Norwegian University of Science and Technology), Trøndelag County Council, Central Norway Regional Health Authority, and the Norwegian Institute of Public Health. The genotyping was financed by the National Institute of health (NIH), University of Michigan, The Norwegian Research council, and Central Norway Regional Health Authority and the Faculty of Medicine and Health Sciences, Norwegian University of Science and Technology (NTNU). The genotype quality control and imputation has been conducted by the K.G. Jebsen center for genetic epidemiology, Department of public health and nursing, Faculty of medicine and health sciences, Norwegian University of Science and Technology (NTNU).

**IHGC2016**: Study-specific acknowledgements for the 21 cohorts included in the IHGC2016 can be found from (Gormley et al., 2016).

**NEO:** We express our gratitude to all individuals who participate in the Netherlands Epidemiology in Obesity study. We are grateful to all participating general practitioners for inviting eligible participants. We also thank P. van Beelen and all research nurses for collecting the data and P. Noordijk and her team for sample handling and storage and I. de Jonge, MSc for data management of the NEO study. The NEO study is supported by the participating Departments, the Division and the Board of Directors of the Leiden University Medical Centre, and by the Leiden University, Research Profile Area 'Vascular and Regenerative Medicine'.

**UKBB**: We thank the study participants for their contribution to this research. This research has been conducted using the UK Biobank Resource under Application Number 22627.

#### Ethics statement

All participating studies were approved by local research ethics committees and written informed consent was obtained from all study participants. For all the participating studies, an approval was received to use the data in the present work.

#### Derivation of effective sample size $N_{eff}$

We define $N_{eff} = N\phi(1 - \phi)I_i$ , where N = total sample size (cases + controls), $\phi = \frac{N_{cases}}{N}$, and $I_i$ is an imputation info for a variant $i$. The standard error (se) from logistic regression for a variant $i$ is approximately: $se_i = \frac{1}{\sqrt{2N\phi(1-\phi)I_i f_i(1-f_i)}}$ , where $f_i$ is a frequency of variant $i$. Then

$$se_i^2 \approx \frac{1}{2N\phi(1-\phi)I_i f_i(1-f_i)} = \frac{1}{2N_{eff(i)} f_i(1-f_i)}$$

and

$$N_{eff(i)} \approx \frac{1}{2f_i(1-f_i)\, se_i^2}.$$

#### Enrichment of migraine associated variants for the GTEx v8 tissues

GTEx v8 data (Aguet et al., 2019) downloaded from (gtexportal.org) contain 49 tissues with sample size over 70 and 23,268 genes that have at least one significant *cis*-eQTL at FDR 5%. The number of genes that have at least one significant *cis*-eQTL found per tissue in GTEx v8 and the tissue sample size are highly correlated, and GWAS associated variants are enriched for *cis*-eQTLs compared to all variants tested (1.46-fold) (Aguet et al., 2019).

To study whether migraine associated variants are enriched in any of the 49 tissues from GTEx v8, we first fitted a linear model where the number of migraine lead variants that overlap with significant *cis*-eQTLs for a specific tissue were used as an outcome, and the number of genes with at least one significant *cis*-eQTL for a specific tissue as a predictor (Supplementary Figure 14) using R software. The adjusted $R^2$ of the model was 0.90, and the predictor was statistically significant ($P$-value < $2{\times}10^{-16}$). Next, for each tissue type we did a separate regression model by leaving the tissue of interest out from the model, and then predicted the outcome by using the model fitted on the other tissues together with its 95% prediction intervals (Supplementary Figure 5). A Z-score was formed by dividing the difference between the observed and the predicted value by an estimate of its standard error that was estimated as one quarter of the length of the 95% prediction interval. $P$-values for the squared Z-scores were estimated from the $\chi^2$-distribution with one degree of freedom, and we applied Bonferroni correction ($\alpha$=0.05/49) to identify tissues that were enriched for migraine associated variants.

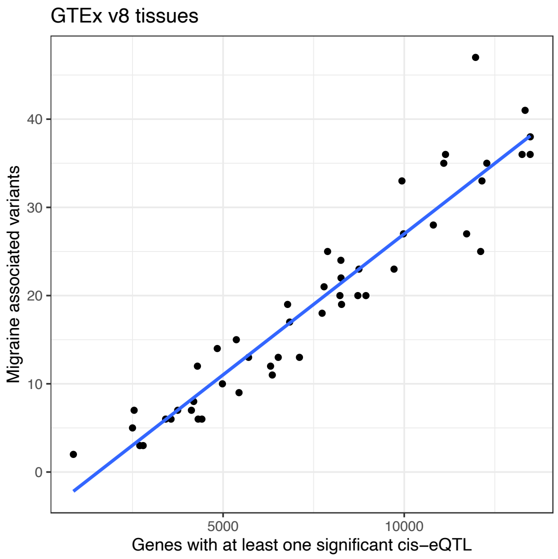


**Supplementary Figure 14**. A Scatter plot of GTEx v8 tissues, where the X-axis shows the number of genes with at least one significant *cis*-eQTL for a given tissue and Y-axis shows the number of migraine lead variants that overlap with significant *cis*-eQTLs for that tissue. Blue line is the regression line from the fitted linear model.

#### Funding

DIC is supported in part by the US National Institute of Neurological Disorders and Stroke (NINDS) of the US National Institutes of Health (NIH) (grant numbers R21NS09296 and R21NS104398).
EW was supported by the Finnish innovation fund Sitra and Finska Läkaresällskapet.
MP was supported by the Academy of Finland (Grant Nos 288509, 312076, 336825) and the Sigrid Juselius Foundation.

SR was supported by the Academy of Finland Center of Excellence in Complex Disease Genetics (Grant No 312062), the Finnish Foundation for Cardiovascular Research, the Sigrid Juselius Foundation and University of Helsinki HiLIFE Fellow and Grand Challenge grants.
TFH and KB are supported by The Novo Nordisk Foundation (NNF14CC0001 and NNF17OC0027594). TFH is supported by CANDY foundation (CEHEAD).

#### International Headache Genetics Consortium

Verneri Anttila[1], Ville Artto[2], Andrea Carmine Belin[3], Anna Bjornsdottir[4], Gyda Bjornsdottir[5], Dorret I. Boomsma[6], Sigrid Børte[7], Mona Ameri Chalmer[8], Daniel I. Chasman[9], Bru Cormand[10], Ester Cuenca[11], George Davey Smith[12], Irene de Boer[13], Martin Dichgans[14], Tonu Esko[15], Tobias Freilinger[16], Nick Furlotte[17], Padhraig Gormley[1], Lyn R. Griffiths[18], Thomas Folkmann Hansen[8], Aster V.E. Harder[13], Heidi Hautakangas[20], Marjo Hiekkala[21], Maria G. Hrafnsdottir[22], Eija Hämäläinen[20], M. Arfan Ikram[19], Marjo-Riitta Järvelin[23], Risto Kajanne[20], Mikko Kallela[2], Jaakko Kaprio[20], Mari Kaunisto[20], Lisette J. A. Kogelman[8], Espen Saxhaug Kristoffersen[24], Christian Kubisch[25], Mitja Kurki[1], Tobias Kurth[26], Lenore Launer[27], Terho Lehtimäki[28], Davor Lessel[25], Lannie Ligthart[6], Nadia Litterman[17], Sigurdur H. Magnusson[5], Rainer Malik[14], Bertram Müller-Myhsok[29], Benjamin M. Neale[30], Carrie Northover[17], Dale R. Nyholt[18], Jes Olesen[8], Priit Palta[20], Linda Pedersen[31], Nancy Pedersen[32], Matti Pirinen[20], Danielle Posthuma[33], Patricia Pozo-Rosich[34], Alice Pressman[35], Olli Raitakari[36], Caroline Ran[3], Gudrun R. Sigurdardottir[4], Hreinn Stefánsson[5], Kari Stefansson[5], Olafur A. Sveinsson[22], Gisela M. Terwindt[19], Thorgeir E. Thorgeirsson[5,] Arn M. J. M. van den Maagdenberg[13], Maija Wessman[21], Bendik S. Winsvold[31], John-Anker Zwart[31]

1. Psychiatric and Neurodevelopmental Genetics Unit, Department of Medicine, Massachusetts General Hospital, Boston, Massachusetts, USA. 2. Department of Neurology, Helsinki University Central Hospital, Helsinki, Finland. 3. Department of Neuroscience, Karolinska Institutet, Stockholm, Sweden. 4. Neurology private practice, Laeknasetrid, Reykjavik, Iceland. 5. deCODE genetics/Amgen Inc., Reykjavik, Iceland. 6. Department of Biological Psychology, VU University, Amsterdam, the Netherlands. 7. K. G. Jebsen Center for Genetic Epidemiology, Department of Public Health and Nursing, Faculty of Medicine and Health Sciences, Norwegian University of Science and Technology, Trondheim, Norway. 8. Danish Headache Center, Department of Neurology, Copenhagen University Hospital, Denmark. 9. Department of Medicine, Division of Preventive Medicine, Brigham and Women's Hospital, Harvard Medical School, Boston, Massachusetts, USA. 10. Department of Genetics, University of Barcelona, Spain Centre for Biomedical Network Research on Rare Diseases (CIBERER), Barcelona, Spain. 11. Pediatric Neurology Research

Group, Vall d'Hebron Research Institute, Barcelona, Spain. 12. University of Bristol/Medical Research Council (MRC) Integrative Epidemiology Unit, University of Bristol, Bristol, UK. 13. Department of Human Genetics, Leiden University Medical Center, Leiden, the Netherlands. 14. Institute for Stroke and Dementia Research, Klinikum der Universität München, Munich, Germany. 15. Estonian Biobank Registry, the Estonian Genome Center, University of Tartu, Tartu, Estonia. 16. Department of Neurology and Epileptology, Hertie Institute for Clinical Brain Research, University of Tuebingen, Tuebingen, Germany. 17. 23&Me Inc., Mountain View, California, USA. 18. School of Biomedical Sciences and Centre for Genomics and Personalised Health, Faculty of Health, Queensland University of Technology, Brisbane, QLD, Australia. 19. Department of Neurology, Leiden University Medical Center, Leiden, the Netherlands. 20. Institute for Molecular Medicine Finland (FIMM), Helsinki Institute of Life Science (HiLIFE), University of Helsinki, Helsinki, Finland. 21. Folkhälsan Research Center, Helsinki, Finland. 22. Landspitali University Hospital, Reykjavik, Iceland. 23. Institute of Health Sciences, University of Oulu, Oulu, Finland. 24. Department of Neurology, Akershus University Hospital and University of Oslo, Oslo, Norway. 25. Institute of Human Genetics, University Medical Center Hamburg-Eppendorf, Hamburg, Germany. 26. Institute of Public Health, Charité – Universitätsmedizin Berlin, Berlin, Germany. 27. Laboratory of Epidemiology and Population Sciences, Intramural Research Program, National Institute on Aging, Bethesda, Maryland, USA. 28. Department of Clinical Chemistry, University of Tampere School of Medicine, Tampere, Finland. 29. Max Planck Institute of Psychiatry, Munich, Germany. 30. Analytical and Translational Genetics Unit, Department of Medicine, Massachusetts General Hospital, Boston, Massachusetts, USA. 31. Department of Research, Innovation and Education, Division of Clinical Neuroscience, Oslo University Hospital, Oslo, Norway 32. Department of Medical Epidemiology and Biostatistics, Karolinska Institutet, Stockholm, Sweden. 33. Department of Complex Trait Genetics, Center for Neurogenomics and Cognitive Research, Neuroscience Campus Amsterdam, VU University, Amsterdam, The Netherlands. 34. Headache Unit, Neurology Department, Vall d'Hebron University Hospital, Barcelona, Spain. 35. Sutter Health, Sacramento, California, USA. 36. Research Centre of Applied and Preventive Cardiovascular Medicine,University of Turku, Turku University Hospital, Turku, Finland.

#### HUNT All-in Headache

Lars G. Fritsche[37], Oddgeir Lingaas Holmen[38], Marie Udnesseter Lie[39,40], Amy E. Martinsen[39,40], Jonas Bille Nielsen[7,41,42], Kristian Bernhard Nilsen[39,43], Linda M. Pedersen[31], Lars Jacob Stovner[44,45], Erling Tronvik[44,45,46], Cristen Willer[41], Wei Zhou[47,48]

37. Center for Statistical Genetics, Department of Biostatistics, University of Michigan, Ann Arbor, MI, USA. 38. HUNT Research Center, Department of Public Health and Nursing, Faculty of Medicine and Health Sciences, Norwegian University of Science and Technology, Trondheim, Norway. 39. Research and Communication Unit for Musculoskeletal Health (FORMI), Department of Research, Innovation and Education, Division of Clinical Neuroscience, Oslo University Hospital, Oslo, Norway. 40. Institute of Clinical Medicine, Faculty of Medicine, University of Oslo, Oslo, Norway. 41. Department of Internal Medicine, Division of Cardiovascular Medicine, University of Michigan, Ann Arbor, MI, USA. 42. Department of Epidemiology Research, Statens Serum Institut, Copenhagen, Denmark. 43. Department of Neurology, Oslo University Hospital, Oslo, Norway. 44. Department of

Neuromedicine and Movement Science, Faculty of Medicine and Health Sciences, Norwegian University of Science and Technology (NTNU), Trondheim, Norway. 45. Norwegian Advisory Unit on Headaches, St. Olavs Hospital, Trondheim University Hospital, Trondheim, Norway. 46. Department of Neurology, St. Olavs Hospital, Trondheim, Norway. 47. Department of Computational Medicine and Bioinformatics, University of Michigan, Ann Arbor, MI, USA. 48. Analytic and Translational Genetics Unit, Massachusetts General Hospital, Boston, Massachusetts, USA.

#### Danish Blood Donor Study Genomic Cohort

Steffen Andersen49, Karina Banasik[50], Søren Brunak[51], Kristoffer Sølvsten Burgdorf[52], Daniel Gudbjartsson[5], Henrik Hjalgrim[42], Gregor Jemec[34], Poul Jennum[54], Per Ingemar Johansson[55], Topholm Bruun Mie[56], Kasper Rene Nielsen[57], Mette Nyegaard[58], Mikkel Petersen[59], Erik Sørensen[55], Kari Stefansson[5], Hreinn Stefánsson[5], Henrik Ullum[52], Thomas Werge[60], Unnur Þorsteinsdóttir[5]

49. Department of Finance, Copenhagen Business School, Copenhagen, Denmark. 50. Novo Nordisk Foundation Center for Basic Metabolic Research, University of Copenhagen, Copenhagen, Denmark. 51. Novo Nordisk Foundation Center for Protein Research, Faculty of Health and Medical Sciences, University of Copenhagen, Copenhagen, Denmark. 52. Department of Clinical Immunology, Copenhagen University Hospital, Rigshospitalet, Copenhagen, Denmark. 53. Department of Clinical Medicine, Sealand University hospital, Roskilde, Denmark. 54. Department of clinical neurophysiology at University of Copenhagen, Copenhagen, Denmark. 55. Department of Clinical Immunology, Copenhagen University Hospital, Copenhagen, Denmark. 56. Department of Clinical Immunology, Odense University Hospital, Odense, Denmark. 57. Department of Clinical Immunology, Aalborg University Hospital, Aalborg, Denmark. 58. Department of Biomedicine, Aarhus University, Aarhus, Denmark. 59. Department of Clinical Immunology, Aarhus University Hospital, Aarhus, Denmark. 60. Institute of Biological Psychiatry, Mental Health Centre Sct. Hans, Copenhagen University Hospital, Roskilde, Denmark.
