## Supplementary Figures 1,2, 5-7 and 11. for "Genome-wide analysis of 102,084 migraine cases identifies 123 risk loci and subtype-specific risk alleles"

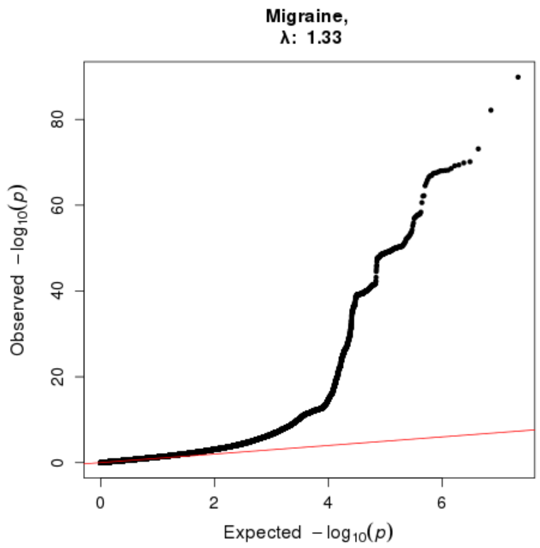


**Supplementary Figure 1**. QQ-plot of the migraine meta-analysis test statistics with 102,084 cases and 771,257 controls. A genomic inflation factor ($\lambda_{GC}$) is 1.33. X-axis shows the expected -log10 *P*-value for each quantile, and Y-axis shows the observed -log10 *P*-Value for the corresponding quantile.

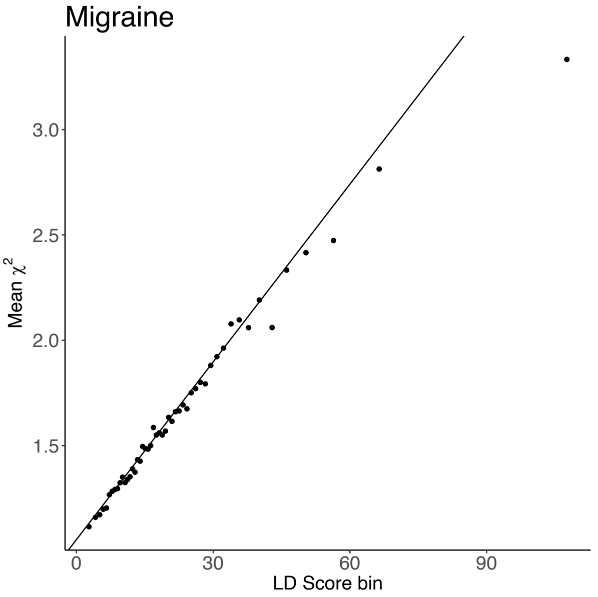


**Supplementary Figure 2**. Plot shows linear trend between LD Score bins and mean migraine GWAS summary statistics of each bin. Heritability is estimated by regressing GWAS summary statistics against LD Scores. A deviation of the intercept term from 1 would indicate confounding inflation of the association statistics, such as population stratification or model misspecification.

**Supplementary Figure 3**.
Regional Locuszoom-plots of the 123 independent migraine risk loci identified from the meta-analysis. The squared correlation to the lead variant is shown by colors based on the UK Biobank data for variants that have an effective sample size ±20% of the lead variant´s effective sample size. Black horizontal line corresponds to $P = 5 \times 10^{-8}$ and blue line shows the recombination rate.

**Supplementary Figure 4**.
Forest plots of the 123 lead migraine variants. For each variant we plot the beta estimate with its 95%-confidence intervals (green) from each five GWAS study included in the meta-analysis and a combined beta estimate from the fixed-effect meta-analysis (blue diamond). Grey squares indicate the sample sizes of each study. We annotate each plot with the lead variant and effect allele, $P$-value by the inverse-variance weighted fixed-effect meta-analysis, and the heterogeneity index ($I^2$).

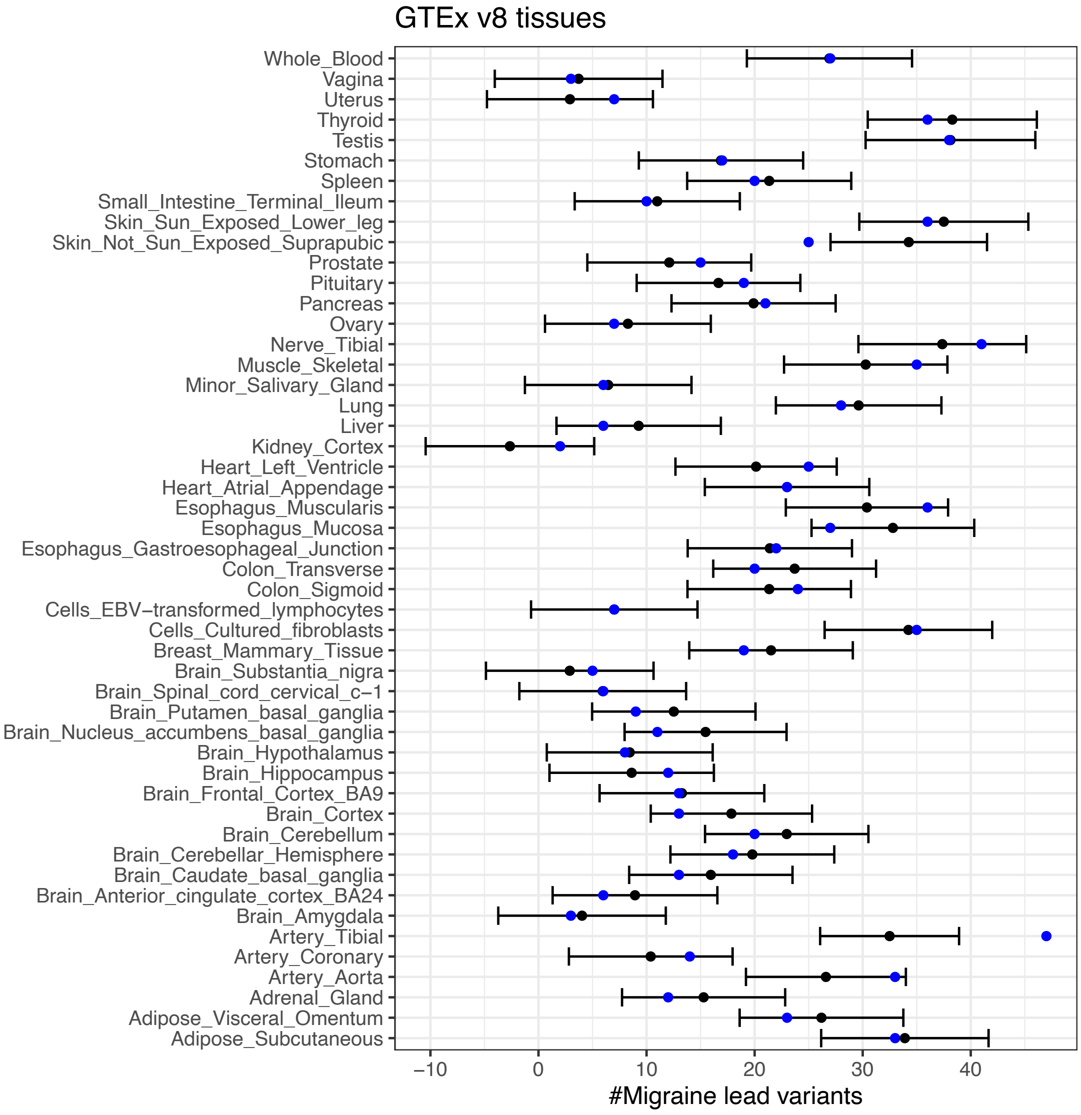


**Supplementary Figure 5**. Results from a prediction model of enriched tissues in GTEx v8 data for migraine lead variants. Y-axis shows each of the 49 tissues, and X-axis shows the number of migraine lead variants that are also significant *cis*-eQTLs for each tissue. Black dot is the predicted value from a linear regression trained on the other 48 tissues, where the number of migraine lead variants that are significant *cis*-eQTLs for each tissue was used as the outcome, and the overall number of genes with at least one significant *cis*-eQTL reported by GTEx for each tissue was the predictor. Black lines are the 95% prediction intervals. Blue dots are the true observed values. When the observed value is far from the prediction interval, the corresponding tissue has an exceptional enrichment or depletion of eQTLs among the migraine lead variants compared to the other tissues.

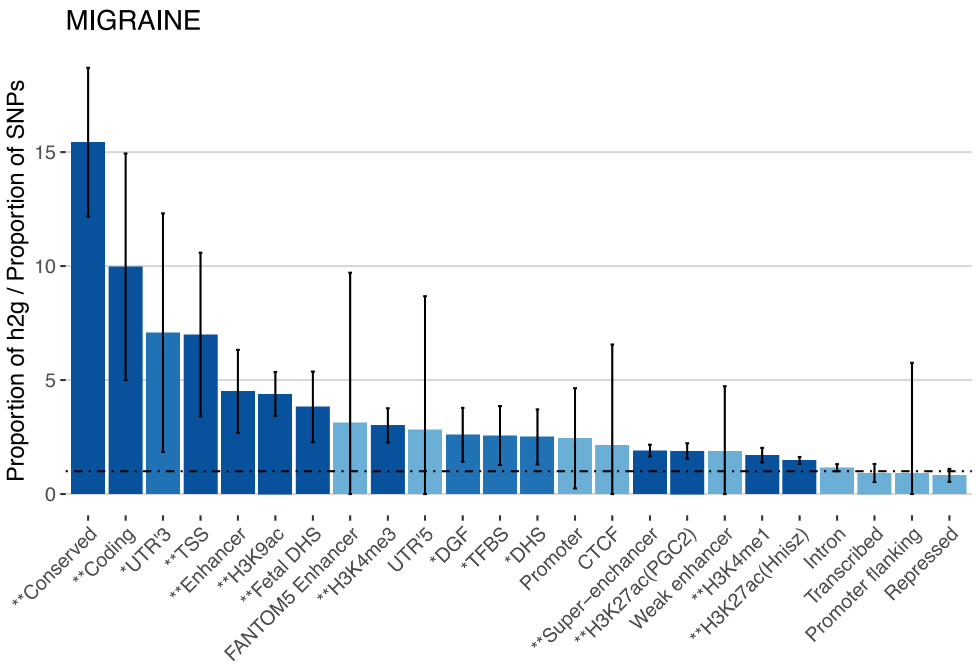


**Supplementary Figure 6.** Enrichment estimates for the 24 main functional annotations for migraine with 95%-confidence intervals from the stratified LD Score regression (S-LDSC). The black dash-dotted line at 1 denotes no enrichment, and values above 1 indicate enriched heritability for a given category and values below 1 depleted heritability for the given category. One asterisk and middle shade of the bar coloring indicate significance at FDR $< 0.05$ and two asterisks and the darkest shade of the bar coloring indicate significance at $P < 0.05$ after Bonferroni correction for the 24 hypotheses tested. Confidence intervals are truncated from below at 0.

a)

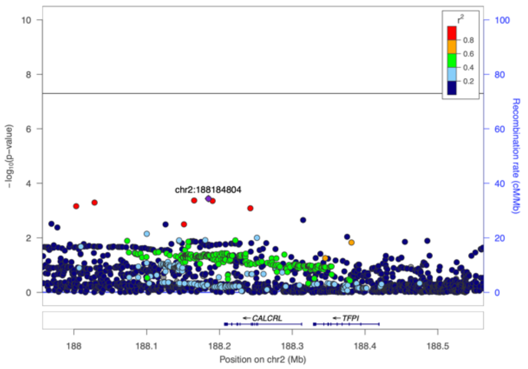
chr2:188184804
-log10(p-value)
Recombination rate (cM/Mb)
r2
CALCRL
TFPI
Position on chr2 (Mb)

b)

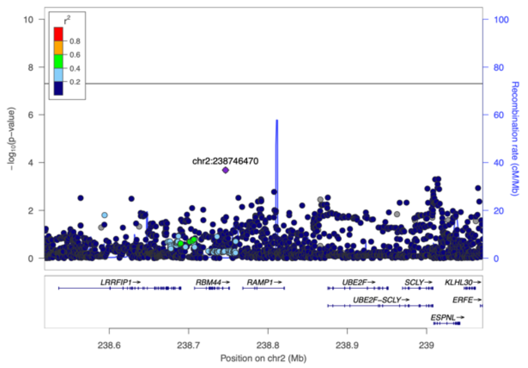
chr2:238746470
-log10(p-value)
Recombination rate (cM/Mb)
r2
LRRFIP1
RBM44
RAMP1
UBE2F
SCLY
KLHL30
UBE2F-SCLY
ERFE
ESPNL
Position on chr2 (Mb)

c)

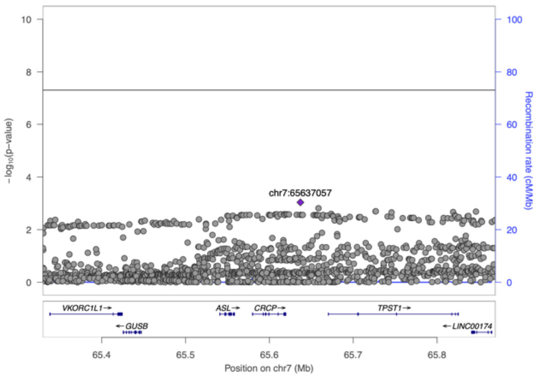
chr7:65637057
-log10(p-value)
Recombination rate (cM/Mb)
VKORC1L1
ASL
CRCP
TPST1
GUSB
LINC00174
Position on chr7 (Mb)

d)

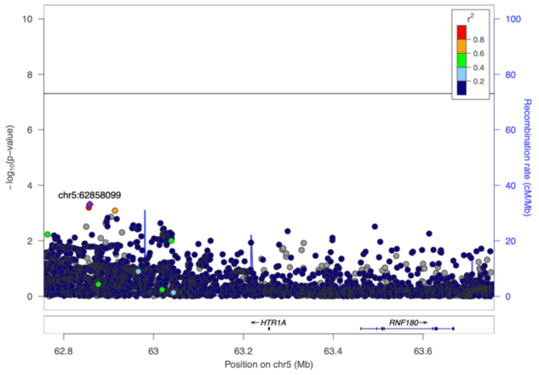
chr5:62858099
-log10(p-value)
Recombination rate (cM/Mb)
r2
HTR1A
RNF180
Position on chr5 (Mb)

e)

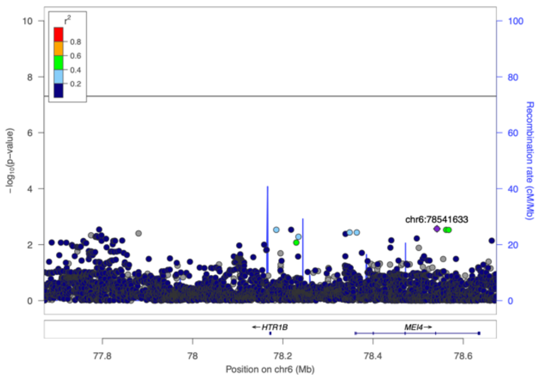
chr6:78541633
-log10(p-value)
Recombination rate (cM/Mb)
r2
HTR1B
MEI4
Position on chr6 (Mb)

f)

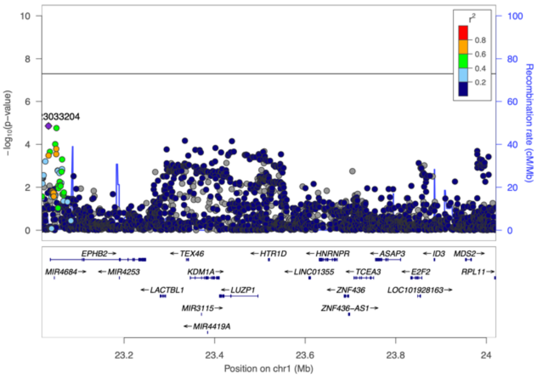
3033204
-log10(p-value)
Recombination rate (cM/Mb)
r2
EPHB2
TEX46
HTR1D
HNRNPR
ASAP3
ID3
MDS2
MIR4684
MIR4253
KDM1A
LINC01355
TCEA3
E2F2
RPL11
LACTBL1
LUZP1
ZNF436
LOC101928163
MIR3115
ZNF436-AS1
MIR4419A
Position on chr1 (Mb)

g)

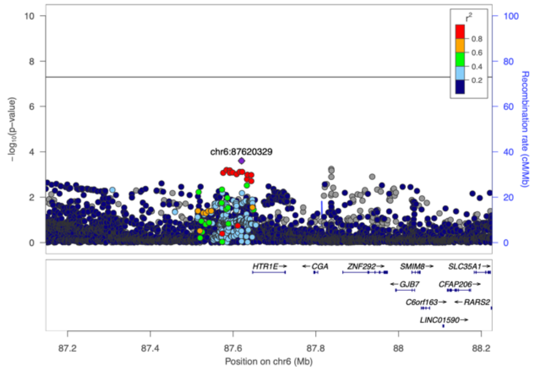
chr6:87620329
-log10(p-value)
Recombination rate (cM/Mb)
r2
HTR1E
CGA
ZNF292
SMIM8
SLC35A1
GJB7
CFAP206
C6orf163
RARS2
LINC01590
Position on chr6 (Mb)

**Supplementary Figure 7**. Locuszoom-plots of three genes, a) *CALCRL*, b) *RAMP1* and c) *CRCP*, that encode central proteins of CGRP receptor complex, and four genes d) *HTR1A*, e) *HTR1B*, f) *HTR1D* and g) *HTR1E* that encode serotonin 5-$HT_1$ receptors. None of the seven regions show a clear association with migraine even though effective therapies targeting CGRP receptor and serotonin 5-$HT_{1B/1D}$ receptors exist. The squared correlation to the lead variant is shown by colors based on the UK Biobank data. Black horizontal line corresponds to $P = 5 \times 10^{-8}$. Blue line shows the recombination rate.

**Supplementary Figure 8**.
Forest plots of the 123 lead migraine variants from the MO meta-analysis. For each variant we plot the beta estimate with its 95%-confidence intervals (green) from each five GWAS study included in the MO meta-analysis and a combined beta estimate from the fixed-effect meta-analysis (blue diamond). Grey squares indicate the sample sizes of each study. We annotate each plot with the lead variant and effect allele, *P*-value by the inverse-variance weighted fixed-effect meta-analysis, and the heterogeneity index ($I^2$).

**Supplementary Figure 9**.
Forest plots of the 123 lead migraine variants from the MA meta-analysis. For each variant we plot the beta estimate with its 95%-confidence intervals (green) from each five GWAS study included in the MA meta-analysis and a combined beta estimate from the fixed-effect meta-analysis (blue diamond). Grey squares indicate the sample sizes of each study. We annotate each plot with the lead variant and effect allele, *P*-value by the inverse-variance weighted fixed-effect meta-analysis, and the heterogeneity index ($I^2$).

**Supplementary Figure 10**.
Plotted are subtype-specific combined beta estimates from the fixed-effect meta-analysis (red dot MA, blue square MO) with their 95%-confidence intervals for the identified 123 lead variants from the primary meta-analysis. For each plot, we annotate *P*-values from the subtype-specific meta-analyses, *P*-value from testing whether the effects are equal in both subtypes ($P_{\text{DIFF}}$) and posterior probabilities for the four models (NULL, MA, MO, BOTH) from the subtype-specificity analysis (Methods).

a)

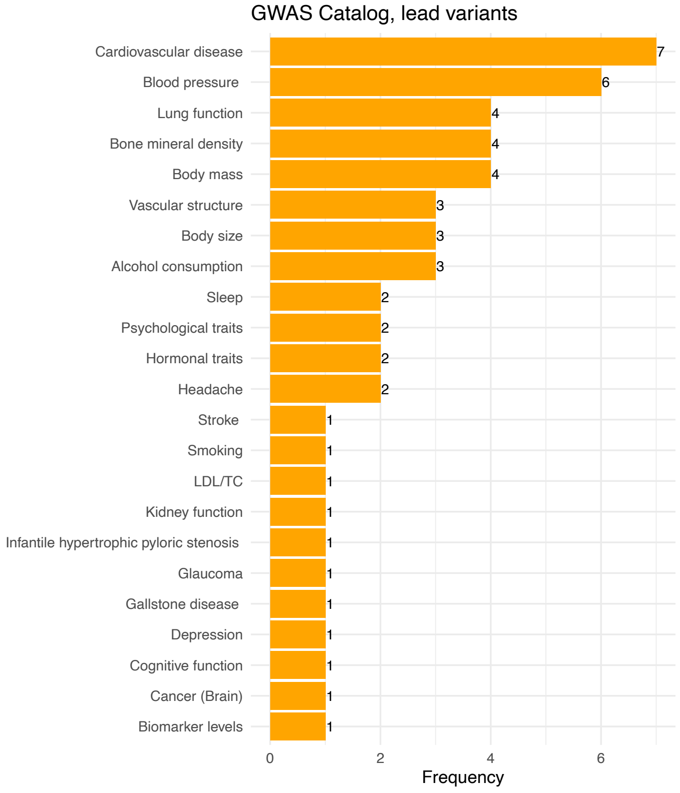
GWAS Catalog, lead variants
Cardiovascular disease 7
Blood pressure 6
Lung function 4
Bone mineral density 4
Body mass 4
Vascular structure 3
Body size 3
Alcohol consumption 3
Sleep 2
Psychological traits 2
Hormonal traits 2
Headache 2
Stroke 1
Smoking 1
LDL/TC 1
Kidney function 1
Infantile hypertrophic pyloric stenosis 1
Glaucoma 1
Gallstone disease 1
Depression 1
Cognitive function 1
Cancer (Brain) 1
Biomarker levels 1
0 2 4 6
Frequency

b)

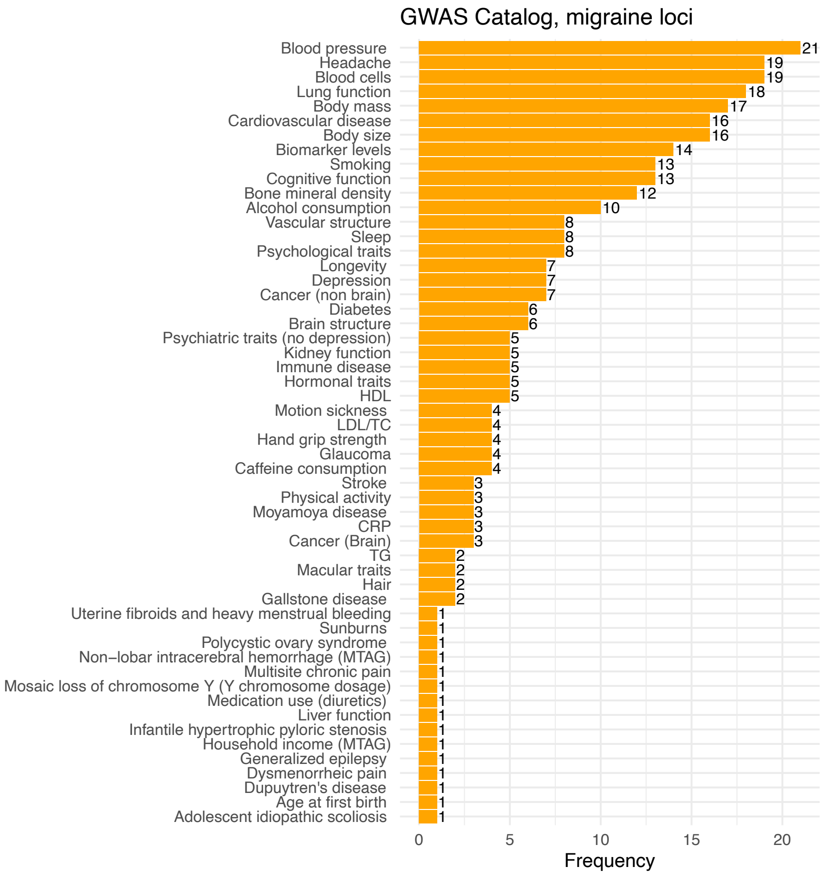
GWAS Catalog, migraine loci
Blood pressure 21
Headache 19
Blood cells 19
Lung function 18
Body mass 17
Cardiovascular disease 16
Body size 16
Biomarker levels 14
Smoking 13
Cognitive function 13
Bone mineral density 12
Alcohol consumption 10
Vascular structure 8
Sleep 8
Psychological traits 8
Longevity 7
Depression 7
Cancer (non brain) 7
Diabetes 6
Brain structure 6
Psychiatric traits (no depression) 5
Kidney function 5
Immune disease 5
Hormonal traits 5
HDL 5
Motion sickness 4
LDL/TC 4
Hand grip strength 4
Glaucoma 4
Caffeine consumption 4
Stroke 3
Physical activity 3
Moyamoya disease 3
CRP 3
Cancer (Brain) 3
TG 2
Macular traits 2
Hair 2
Gallstone disease 2
Uterine fibroids and heavy menstrual bleeding 1
Sunburns 1
Polycystic ovary syndrome 1
Non−lobar intracerebral hemorrhage (MTAG) 1
Multisite chronic pain 1
Mosaic loss of chromosome Y (Y chromosome dosage) 1
Medication use (diuretics) 1
Liver function 1
Infantile hypertrophic pyloric stenosis 1
Household income (MTAG) 1
Generalized epilepsy 1
Dysmenorrheic pain 1
Dupuytren's disease 1
Age at first birth 1
Adolescent idiopathic scoliosis 1
0 5 10 15 20
Frequency

c)

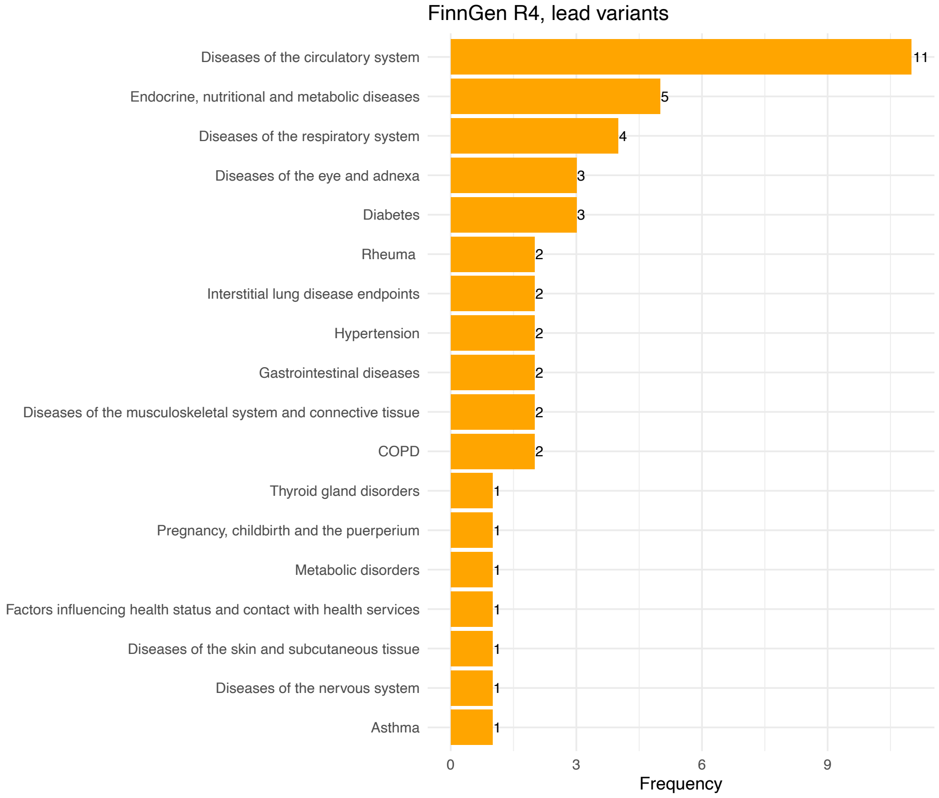
FinnGen R4, lead variants
Diseases of the circulatory system
Endocrine, nutritional and metabolic diseases
Diseases of the respiratory system
Diseases of the eye and adnexa
Diabetes
Rheuma
Interstitial lung disease endpoints
Hypertension
Gastrointestinal diseases
Diseases of the musculoskeletal system and connective tissue
COPD
Thyroid gland disorders
Pregnancy, childbirth and the puerperium
Metabolic disorders
Factors influencing health status and contact with health services
Diseases of the skin and subcutaneous tissue
Diseases of the nervous system
Asthma
11
5
4
3
3
2
2
2
2
2
2
1
1
1
1
1
1
1
0
3
6
9
Frequency

d)

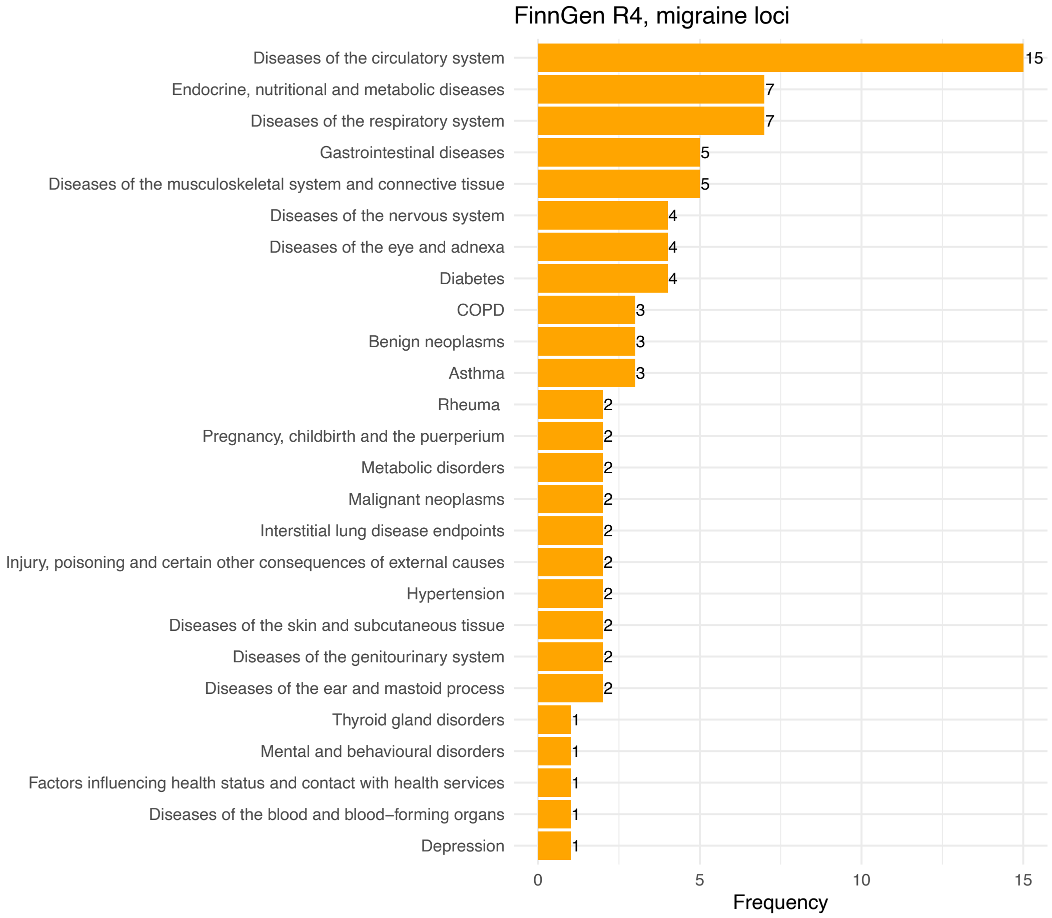
FinnGen R4, migraine loci
Diseases of the circulatory system
Endocrine, nutritional and metabolic diseases
Diseases of the respiratory system
Gastrointestinal diseases
Diseases of the musculoskeletal system and connective tissue
Diseases of the nervous system
Diseases of the eye and adnexa
Diabetes
COPD
Benign neoplasms
Asthma
Rheuma
Pregnancy, childbirth and the puerperium
Metabolic disorders
Malignant neoplasms
Interstitial lung disease endpoints
Injury, poisoning and certain other consequences of external causes
Hypertension
Diseases of the skin and subcutaneous tissue
Diseases of the genitourinary system
Diseases of the ear and mastoid process
Thyroid gland disorders
Mental and behavioural disorders
Factors influencing health status and contact with health services
Diseases of the blood and blood–forming organs
Depression
15
7
7
5
5
4
4
4
3
3
3
2
2
2
2
2
2
2
2
2
2
1
1
1
1
1
0
5
10
15
Frequency

**Supplementary Figure 11**. Results from the PheWAS of NHGRI GWAS Catalog and FinnGen R4. All reported associations for 123 migraine lead variants, and 123 loci with high LD variants included (migraine loci) at $P < 1 \times 10^{-5}$ grouped into broad phenotype categories (Methods). X-axis shows the frequency of lead variants or loci for each category. a) Lead variants in GWAS Catalog; b) Lead variants and variants in high LD in GWAS Catalog; c) Lead variants in FinnGen R4; d) Lead variants and variants in high LD in FinnGen R4.

**Supplementary Figure 12**.

MAF and EAF plots of 4 GWAS against the reference cohort (UKBB). For each GWAS pair, four pairwise plots are presented: MAF and EAF plots before allele flipping and variant exclusions, and MAF and EAF plots after allele flipping and after excluding variants with non-matching alleles and variants with EAF discrepancy over 0.3 (SNPs) or over 0.2 (indels).

**Supplementary Figure 13**.

Gene-set specific QQ-plots of the residual Z-scores from the MAGMA gene-set analysis for the gene sets that are significantly enriched for migraine associated genes after Bonferroni correction using either curated gene sets or GO gene sets. X-axis shows the excepted residual Z-scores from the null model based on the quantiles across all genes in the data and Y-axis shows the observed residual Z-scores. The black points indicate the 25$^{th}$, 50$^{th}$ and 75$^{th}$ percentile. A one-sided 95% confidence band is the dashed black line. The red dots are quantiles that deviate more than would be expected by chance. One gene set (GO_SCHWANN_CELL_MIGRATION) was discarded based on the small size.
