## Supplementary Figure 3. for "Genome-wide analysis of 102,084 migraine cases identifies 123 risk loci and subtype-specific risk alleles"

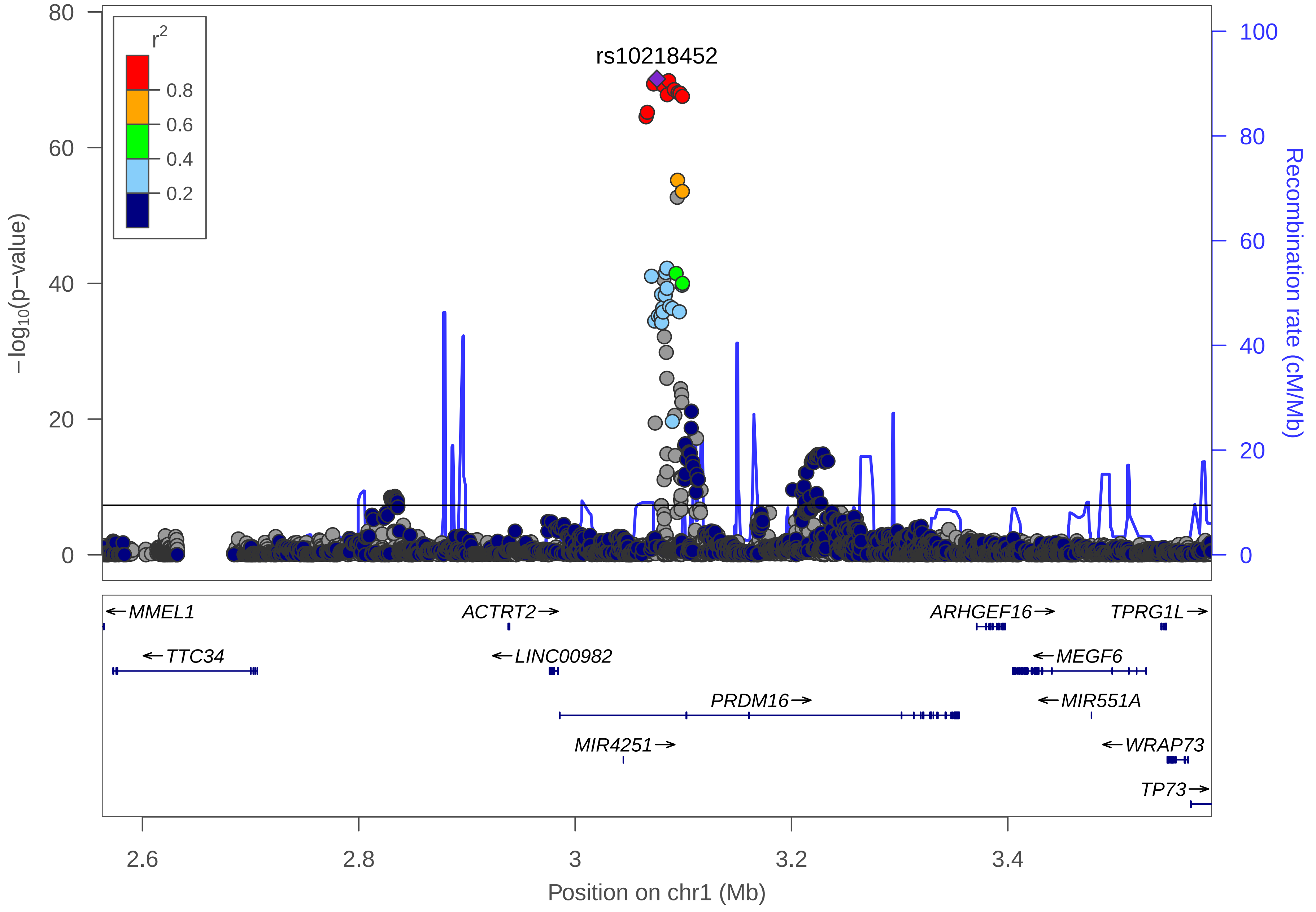

rs10218452
r2
0.8
0.6
0.4
0.2
−log10(p−value)
80
60
40
20
0
Recombination rate (cM/Mb)
100
80
60
40
20
0
MMEL1
TTC34
ACTRT2
LINC00982
PRDM16
MIR4251
ARHGEF16
MEGF6
MIR551A
TPRG1L
WRAP73
TP73
2.6
2.8
3
3.2
3.4
Position on chr1 (Mb)

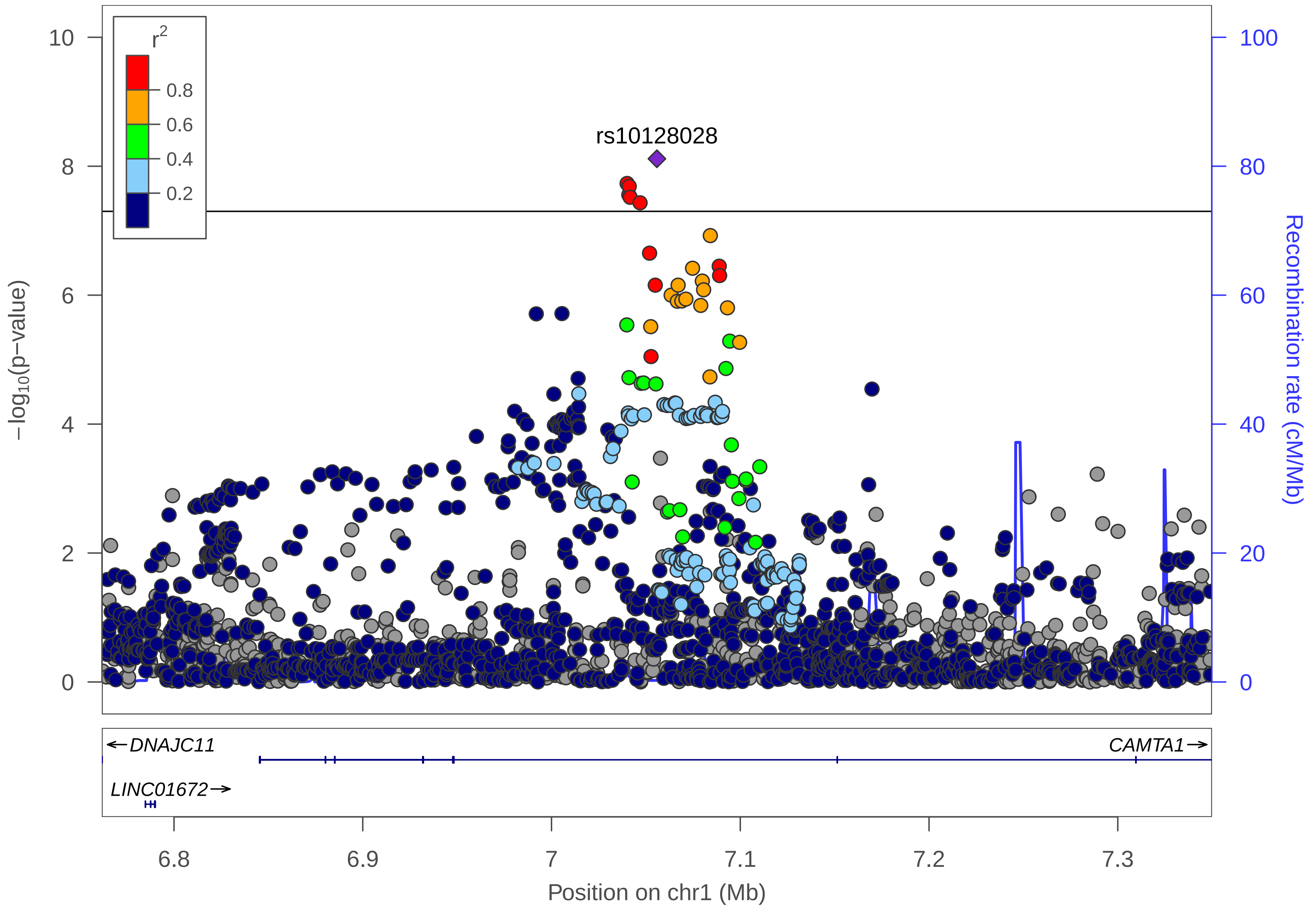

r2
0.8
0.6
0.4
0.2
rs10128028
10
8
6
4
2
0
−log10(p−value)
100
80
60
40
20
0
Recombination rate (cM/Mb)
←DNAJC11
CAMTA1→
LINC01672→
6.8
6.9
7
7.1
7.2
7.3
Position on chr1 (Mb)

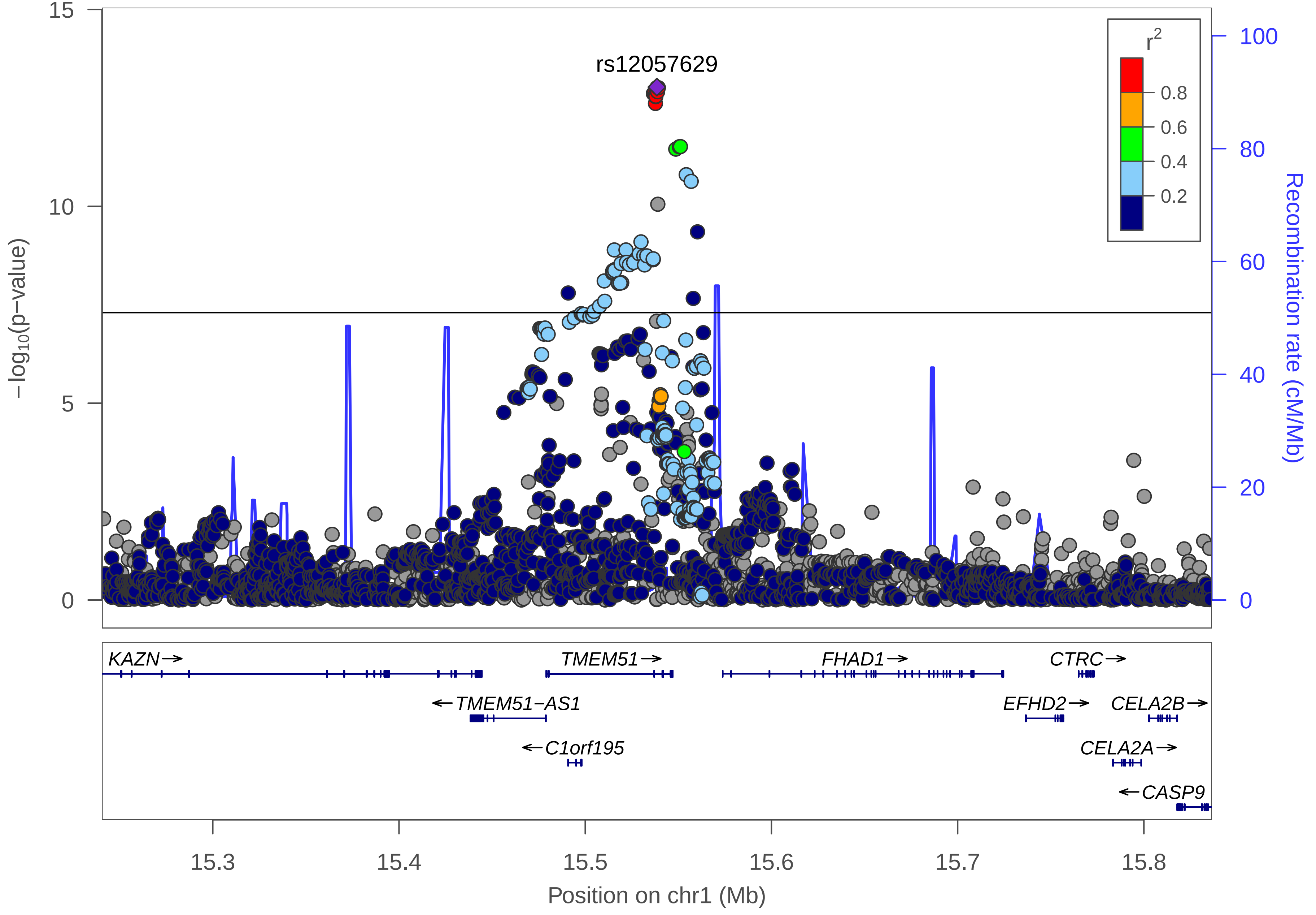

rs12057629
r2
0.8
0.6
0.4
0.2
15
10
5
0
−log10(p−value)
100
80
60
40
20
0
Recombination rate (cM/Mb)
KAZN→
TMEM51→
FHAD1→
CTRC→
←TMEM51−AS1
EFHD2→
CELA2B→
←C1orf195
CELA2A→
←CASP9
15.3
15.4
15.5
15.6
15.7
15.8
Position on chr1 (Mb)

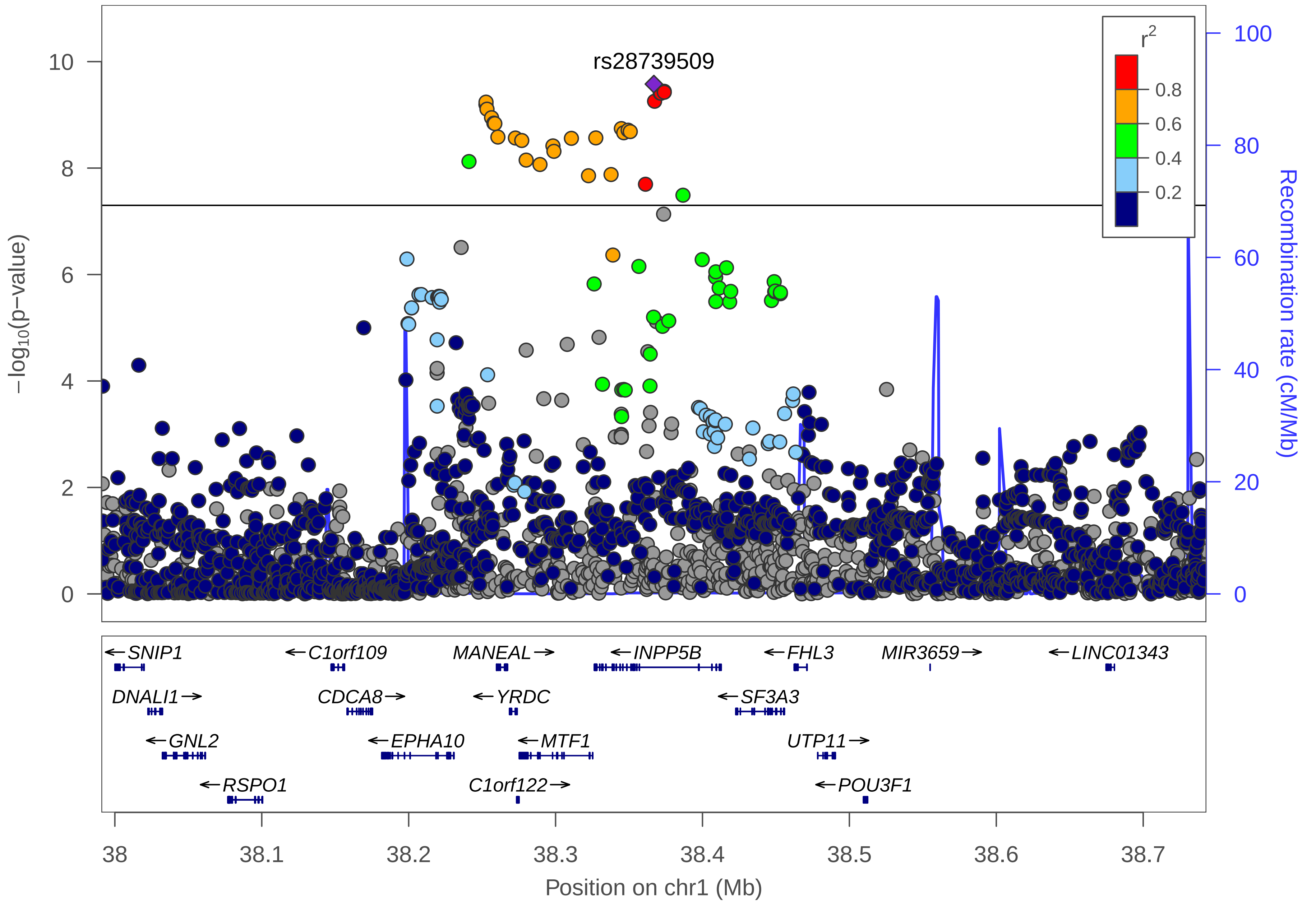

rs28739509
r2
0.8
0.6
0.4
0.2
−log10(p−value)
Recombination rate (cM/Mb)
SNIP1
DNALI1
GNL2
RSPO1
C1orf109
CDCA8
EPHA10
MANEAL
YRDC
MTF1
C1orf122
INPP5B
SF3A3
FHL3
UTP11
POU3F1
MIR3659
LINC01343
Position on chr1 (Mb)

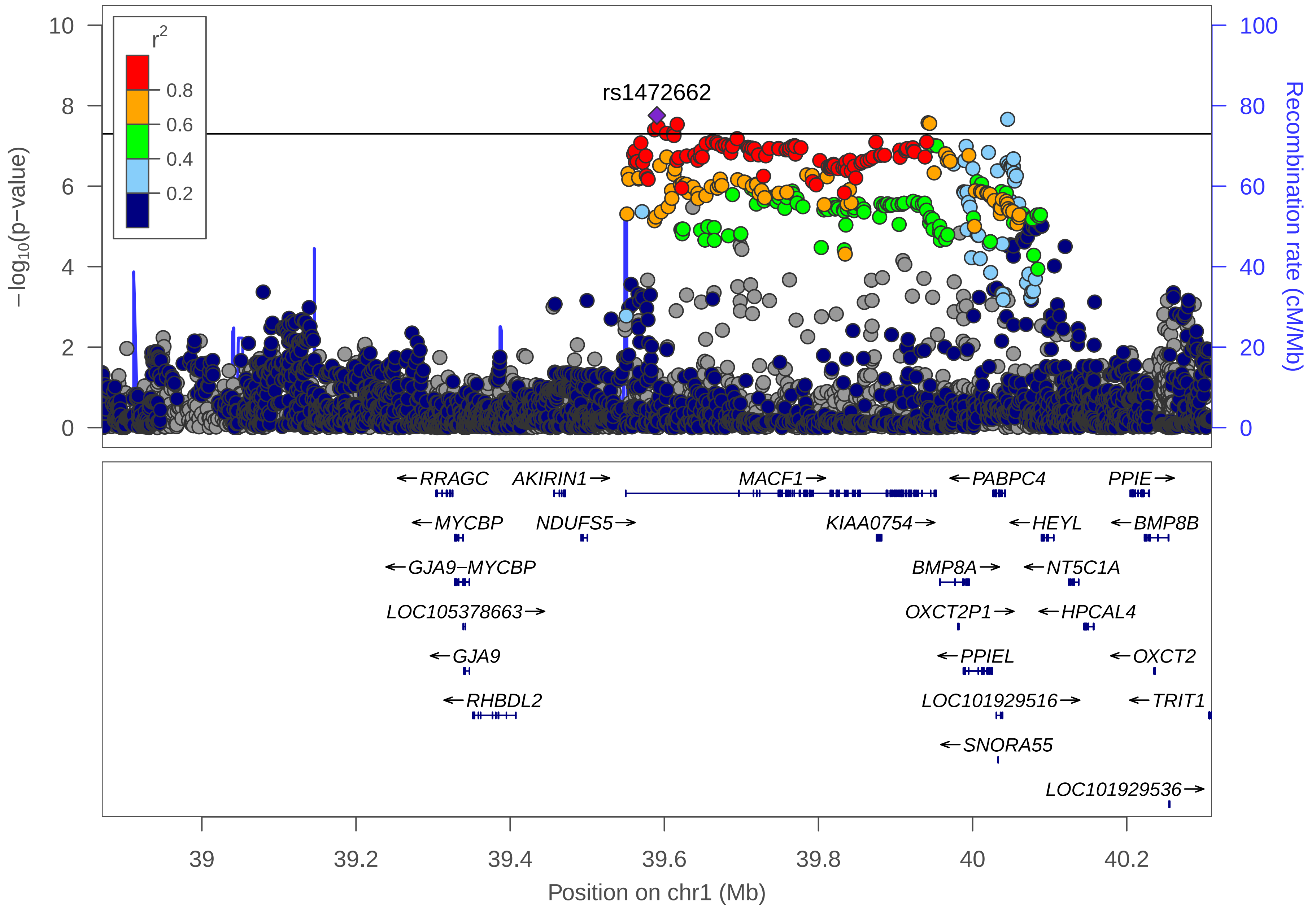
r2
0.8
0.6
0.4
0.2
rs1472662
−log10(p−value)
Recombination rate (cM/Mb)
10
8
6
4
2
0
100
80
60
40
20
0
←RRAGC
AKIRIN1→
MACF1→
←PABPC4
PPIE→
←MYCBP
NDUFS5→
KIAA0754→
←HEYL
←BMP8B
←GJA9−MYCBP
BMP8A→
←NT5C1A
LOC105378663→
OXCT2P1→
←HPCAL4
←GJA9
←PPIEL
←OXCT2
←RHBDL2
LOC101929516→
←TRIT1
←SNORA55
LOC101929536→
39
39.2
39.4
39.6
39.8
40
40.2
Position on chr1 (Mb)

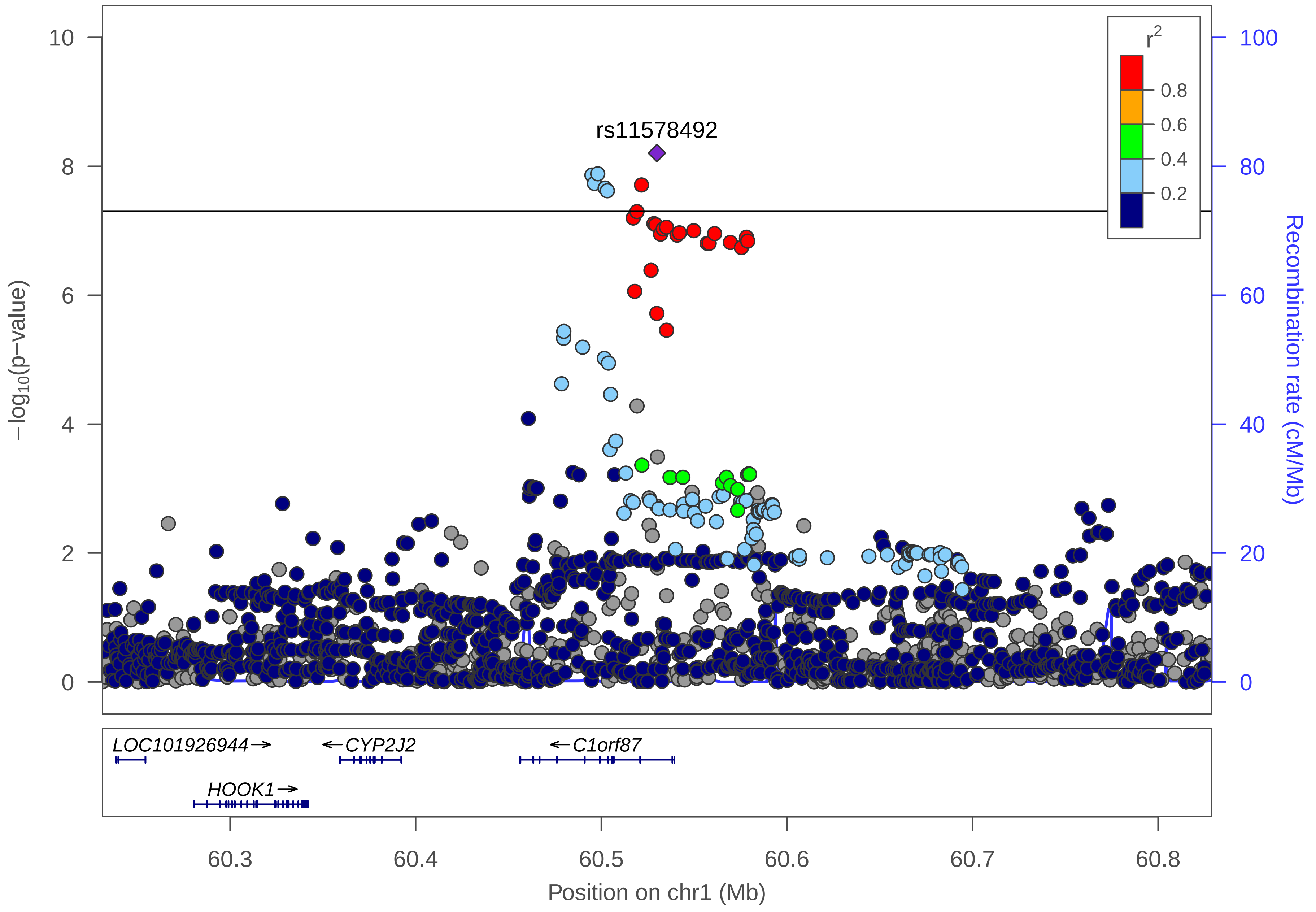

rs11578492
r2
0.8
0.6
0.4
0.2
−log10(p−value)
Recombination rate (cM/Mb)
10
8
6
4
2
0
100
80
60
40
20
0
LOC101926944→
←CYP2J2
←C1orf87
HOOK1→
60.3
60.4
60.5
60.6
60.7
60.8
Position on chr1 (Mb)

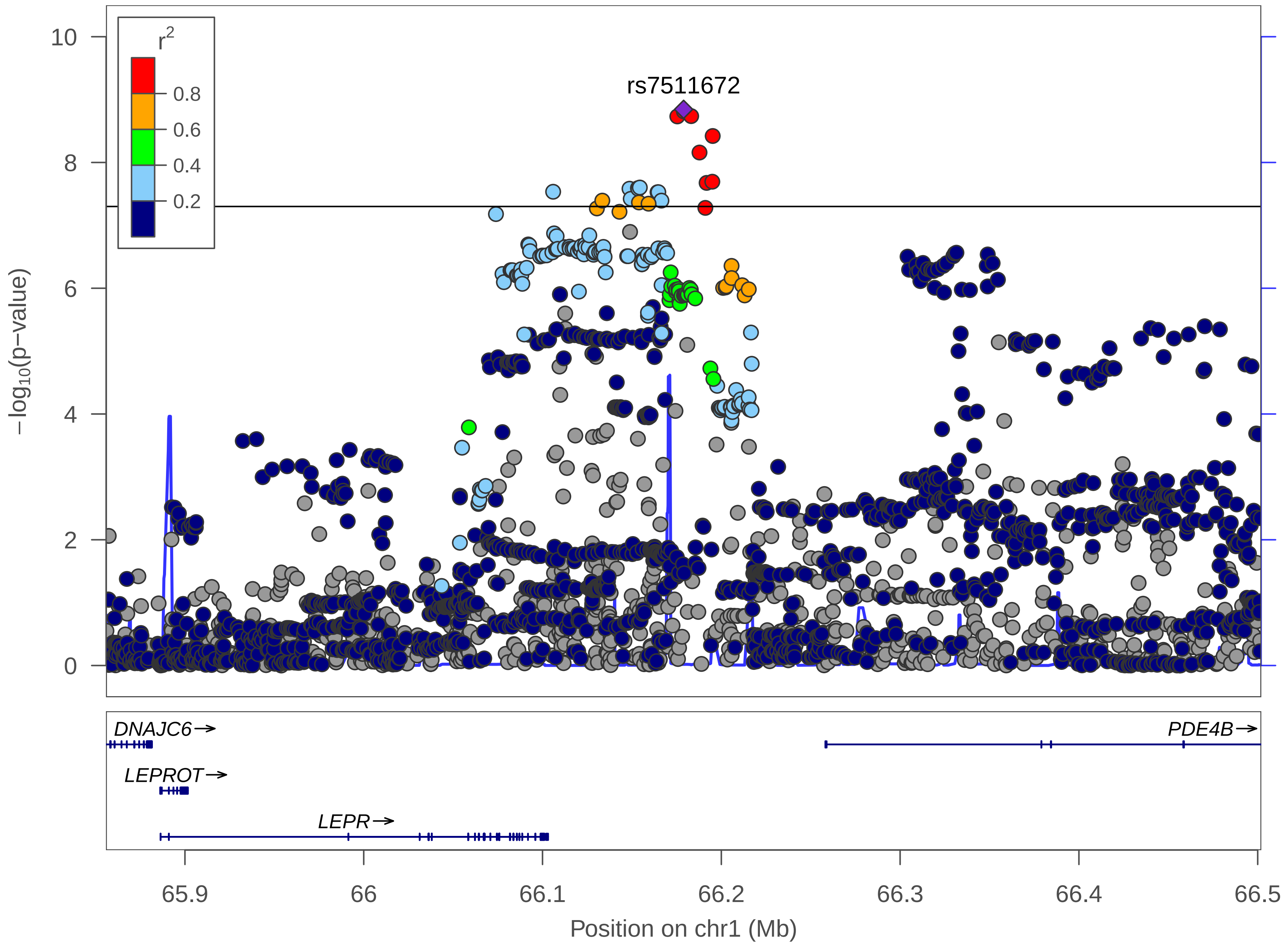

rs7511672
r2
0.8
0.6
0.4
0.2
−log10(p−value)
Recombination rate (cM/Mb)
DNAJC6→
LEPROT→
LEPR→
PDE4B→
Position on chr1 (Mb)

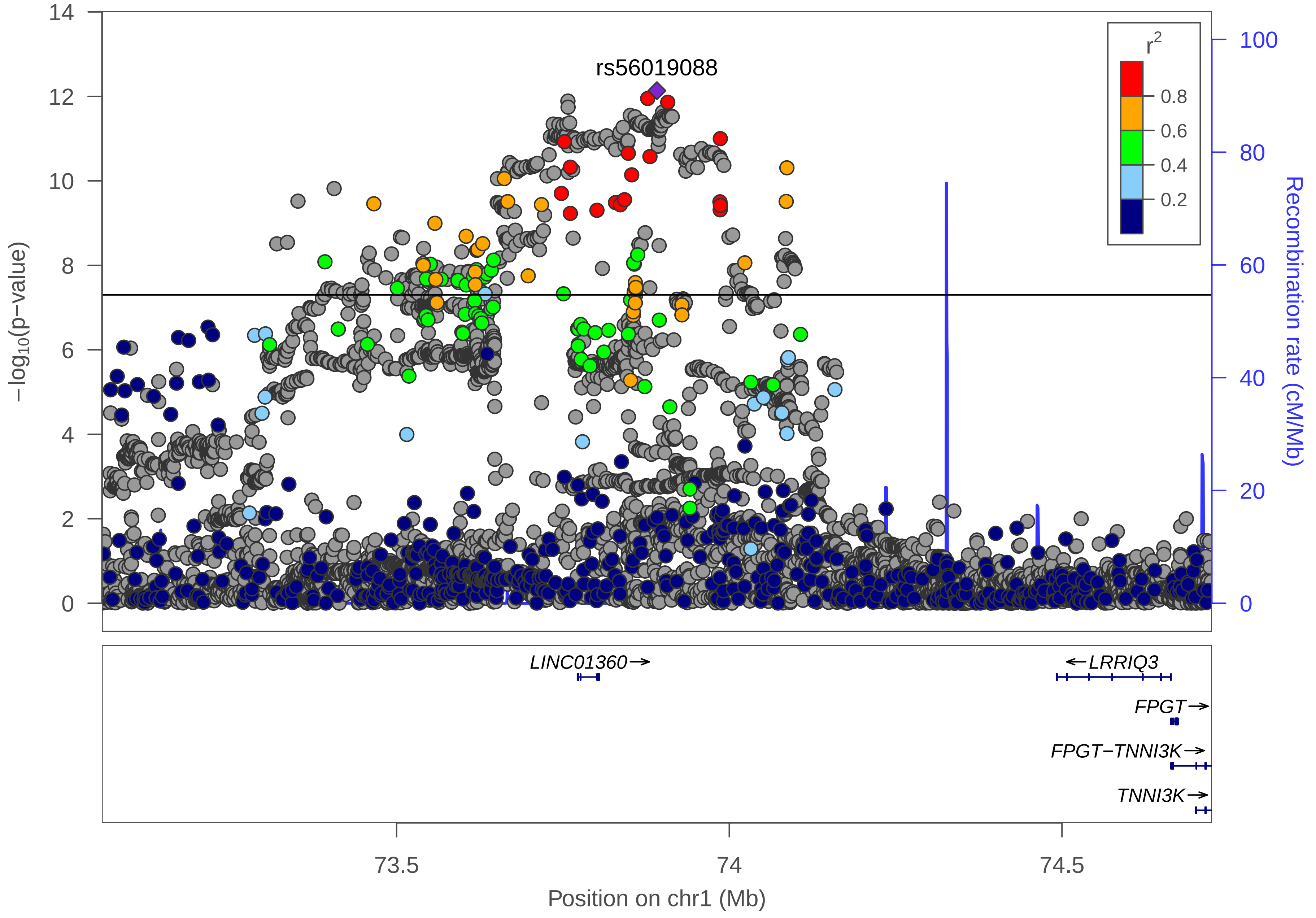

rs56019088
r2
0.8
0.6
0.4
0.2
−log10(p−value)
Recombination rate (cM/Mb)
LINC01360→
←LRRIQ3
FPGT→
FPGT−TNNI3K→
TNNI3K→
73.5
74
74.5
Position on chr1 (Mb)

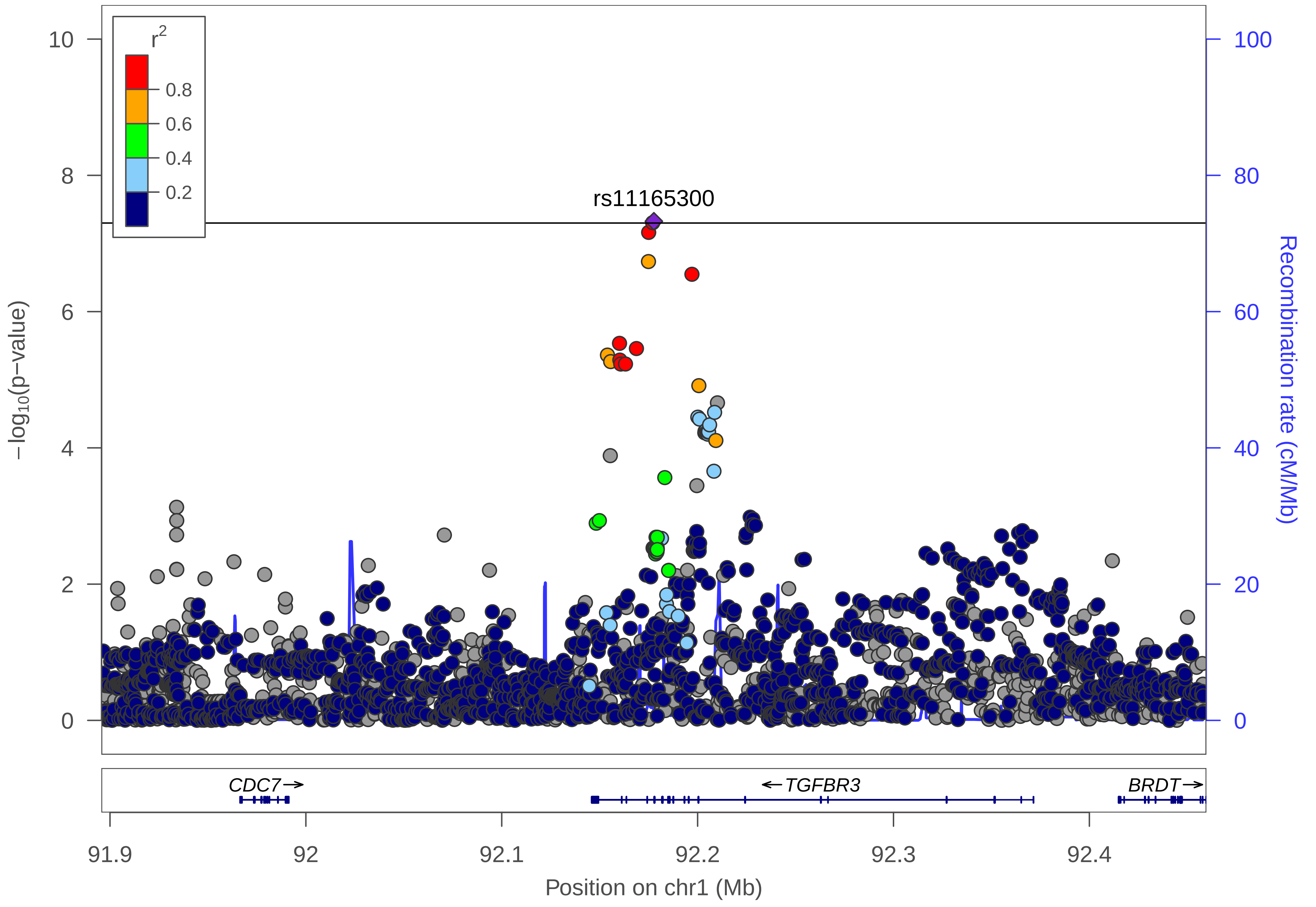

r2
0.8
0.6
0.4
0.2
rs11165300
−log10(p−value)
Recombination rate (cM/Mb)
CDC7
TGFBR3
BRDT
91.9
92
92.1
92.2
92.3
92.4
Position on chr1 (Mb)

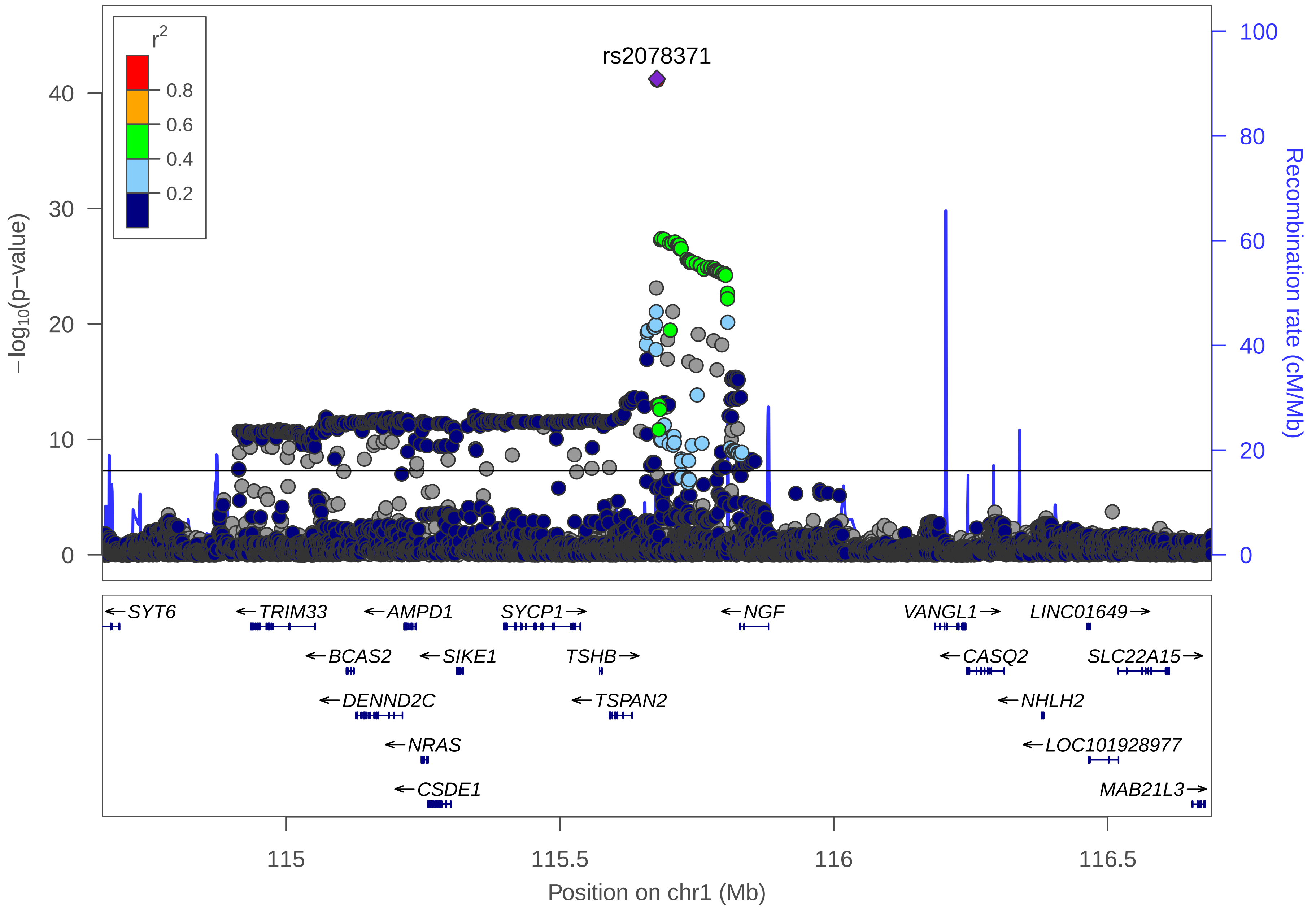

rs2078371
r2
0.8
0.6
0.4
0.2
−log10(p−value)
Recombination rate (cM/Mb)
SYT6
TRIM33
AMPD1
SYCP1
NGF
VANGL1
LINC01649
BCAS2
SIKE1
TSHB
CASQ2
SLC22A15
DENND2C
TSPAN2
NHLH2
NRAS
LOC101928977
CSDE1
MAB21L3
Position on chr1 (Mb)

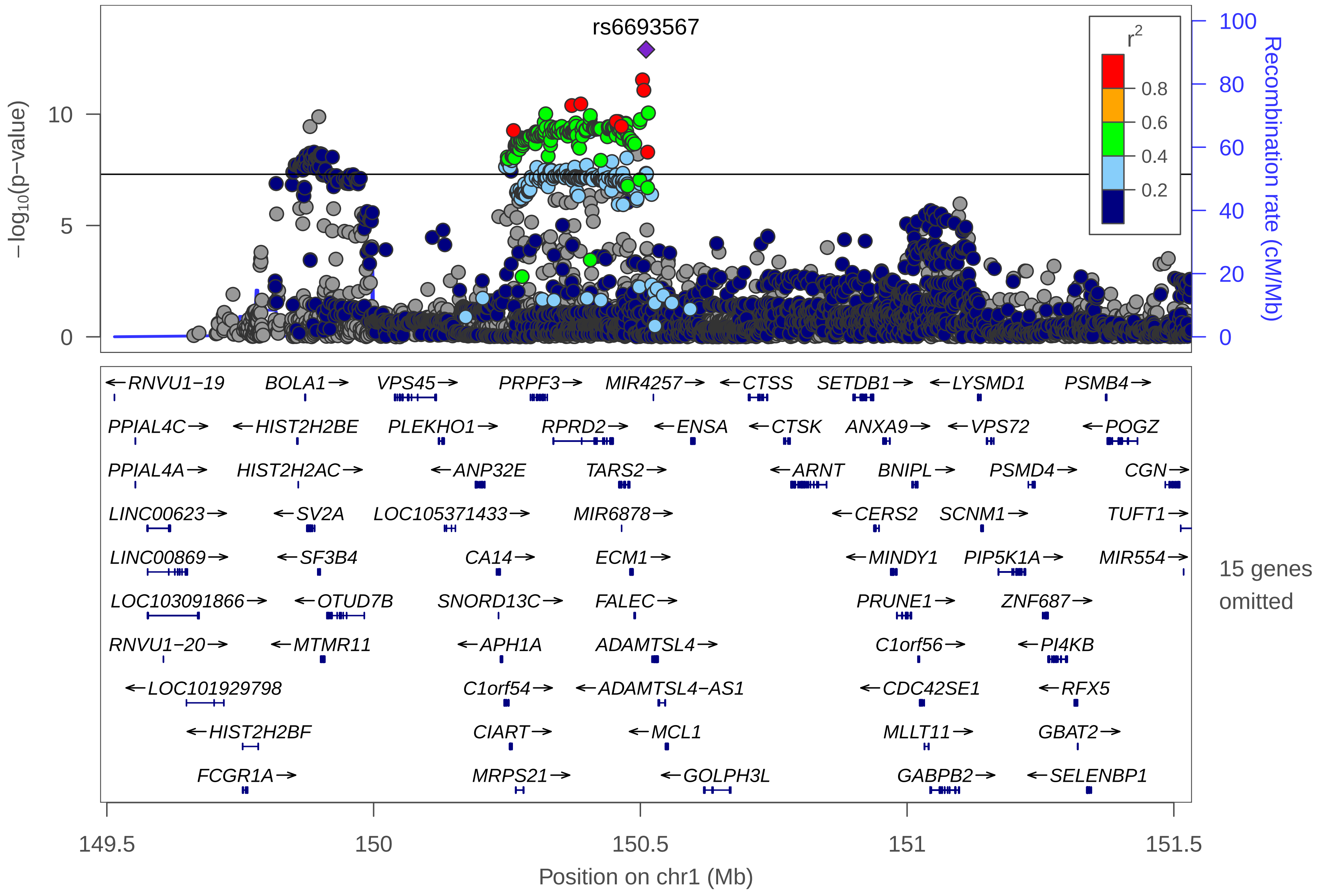
rs6693567
−log10(p−value)
Recombination rate (cM/Mb)
r2
0.8
0.6
0.4
0.2
RNVU1−19
BOLA1
VPS45
PRPF3
MIR4257
CTSS
SETDB1
LYSMD1
PSMB4
PPIAL4C
HIST2H2BE
PLEKHO1
RPRD2
ENSA
CTSK
ANXA9
VPS72
POGZ
PPIAL4A
HIST2H2AC
ANP32E
TARS2
ARNT
BNIPL
PSMD4
CGN
LINC00623
SV2A
LOC105371433
MIR6878
CERS2
SCNM1
TUFT1
LINC00869
SF3B4
CA14
ECM1
MINDY1
PIP5K1A
MIR554
LOC103091866
OTUD7B
SNORD13C
FALEC
PRUNE1
ZNF687
RNVU1−20
MTMR11
APH1A
ADAMTSL4
C1orf56
PI4KB
LOC101929798
C1orf54
ADAMTSL4−AS1
CDC42SE1
RFX5
HIST2H2BF
CIART
MCL1
MLLT11
GBAT2
FCGR1A
MRPS21
GOLPH3L
GABPB2
SELENBP1
15 genes omitted
149.5
150
150.5
151
151.5
Position on chr1 (Mb)

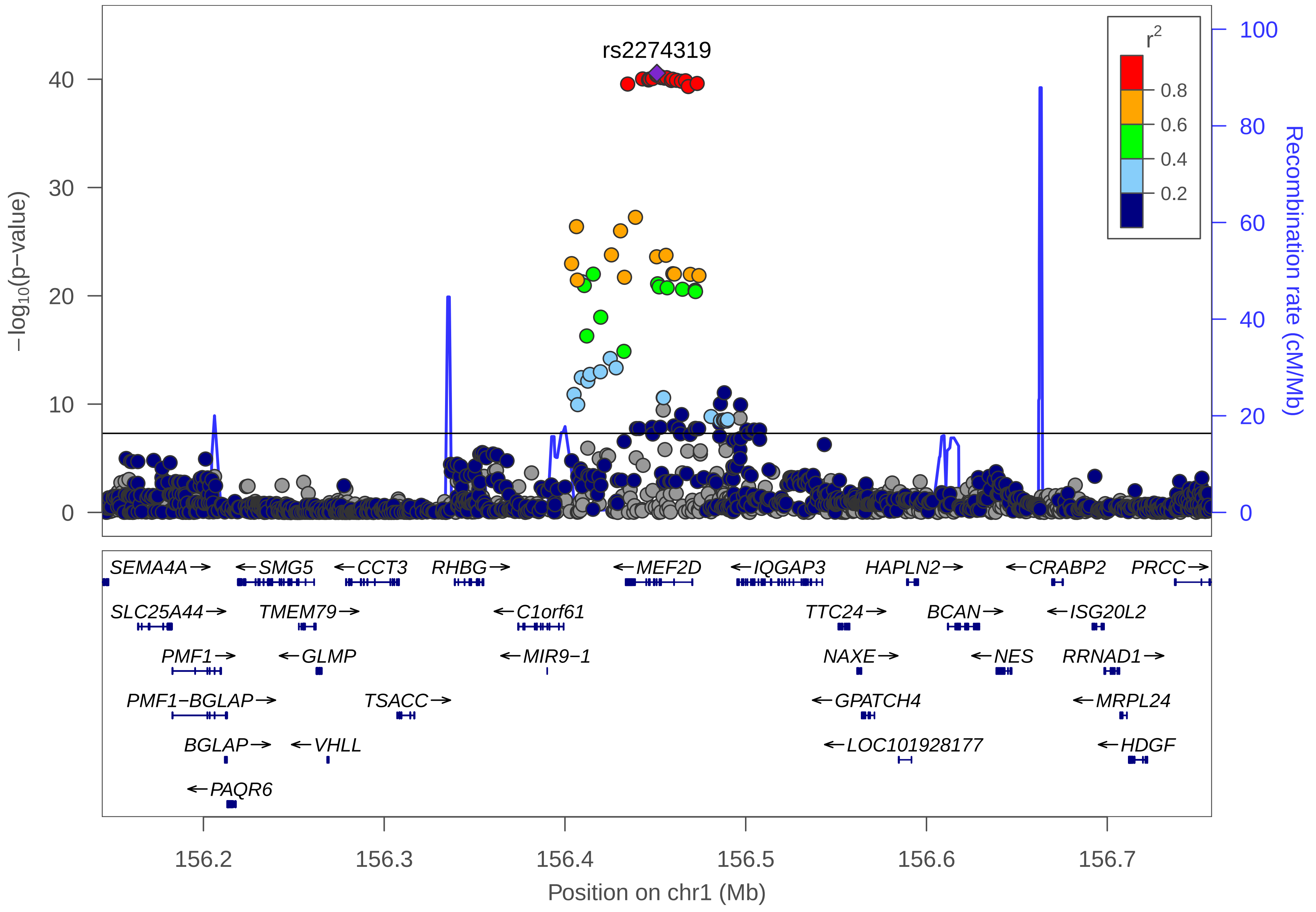

rs2274319
r2
0.8
0.6
0.4
0.2
−log10(p−value)
0
10
20
30
40
Recombination rate (cM/Mb)
0
20
40
60
80
100
SEMA4A→
←SMG5
←CCT3
RHBG→
←MEF2D
←IQGAP3
HAPLN2→
←CRABP2
PRCC→
SLC25A44→
TMEM79→
←C1orf61
TTC24→
BCAN→
←ISG20L2
PMF1→
←GLMP
←MIR9−1
NAXE→
←NES
RRNAD1→
PMF1−BGLAP→
TSACC→
←GPATCH4
←MRPL24
BGLAP→
←VHLL
←LOC101928177
←HDGF
←PAQR6
156.2
156.3
156.4
156.5
156.6
156.7
Position on chr1 (Mb)

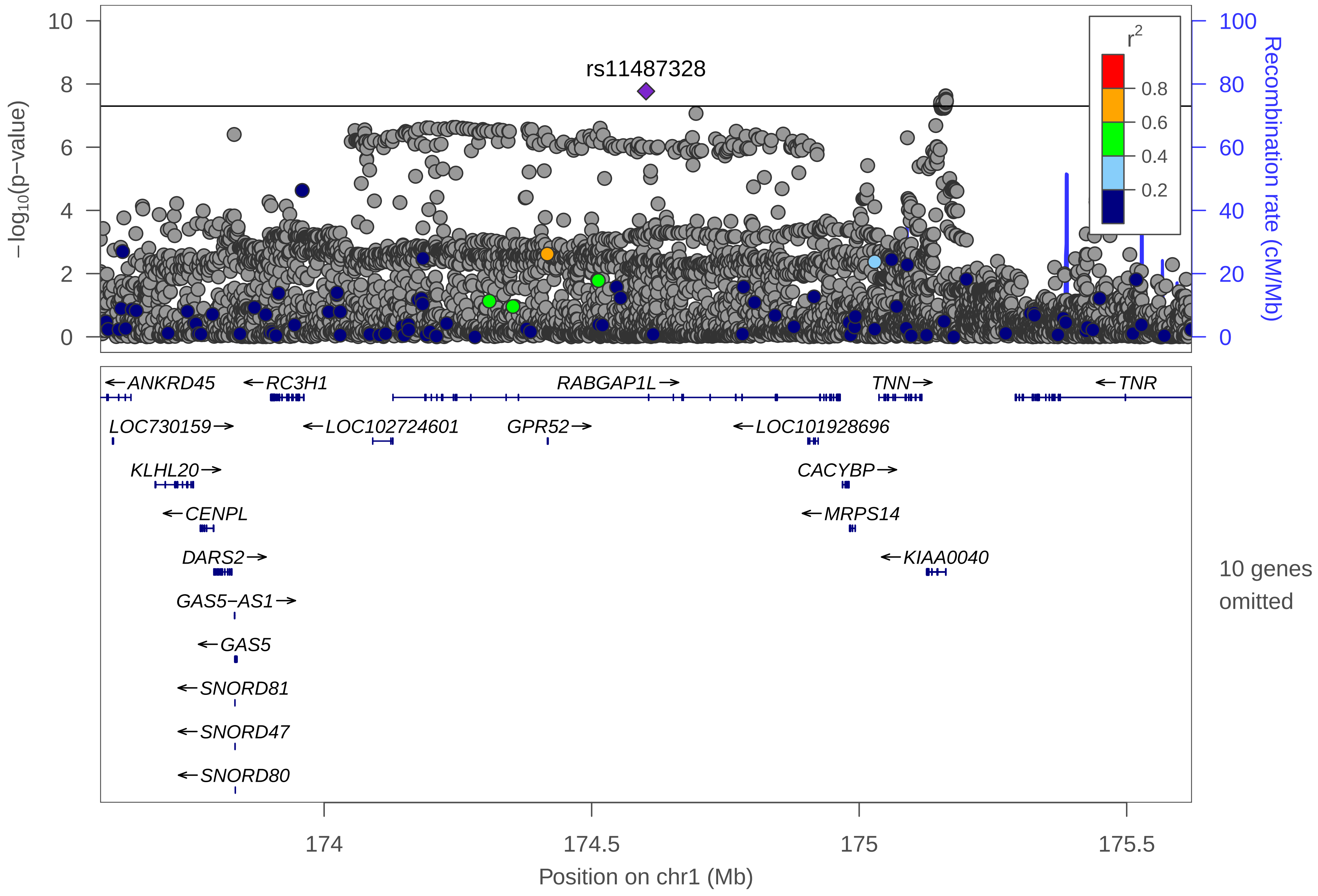

rs11487328
−log10(p−value)
Recombination rate (cM/Mb)
r2
0.8
0.6
0.4
0.2
0
2
4
6
8
10
20
40
60
80
100
ANKRD45
RC3H1
RABGAP1L
TNN
TNR
LOC730159
LOC102724601
GPR52
LOC101928696
KLHL20
CACYBP
CENPL
MRPS14
DARS2
KIAA0040
GAS5−AS1
GAS5
SNORD81
SNORD47
SNORD80
10 genes omitted
174
174.5
175
175.5
Position on chr1 (Mb)

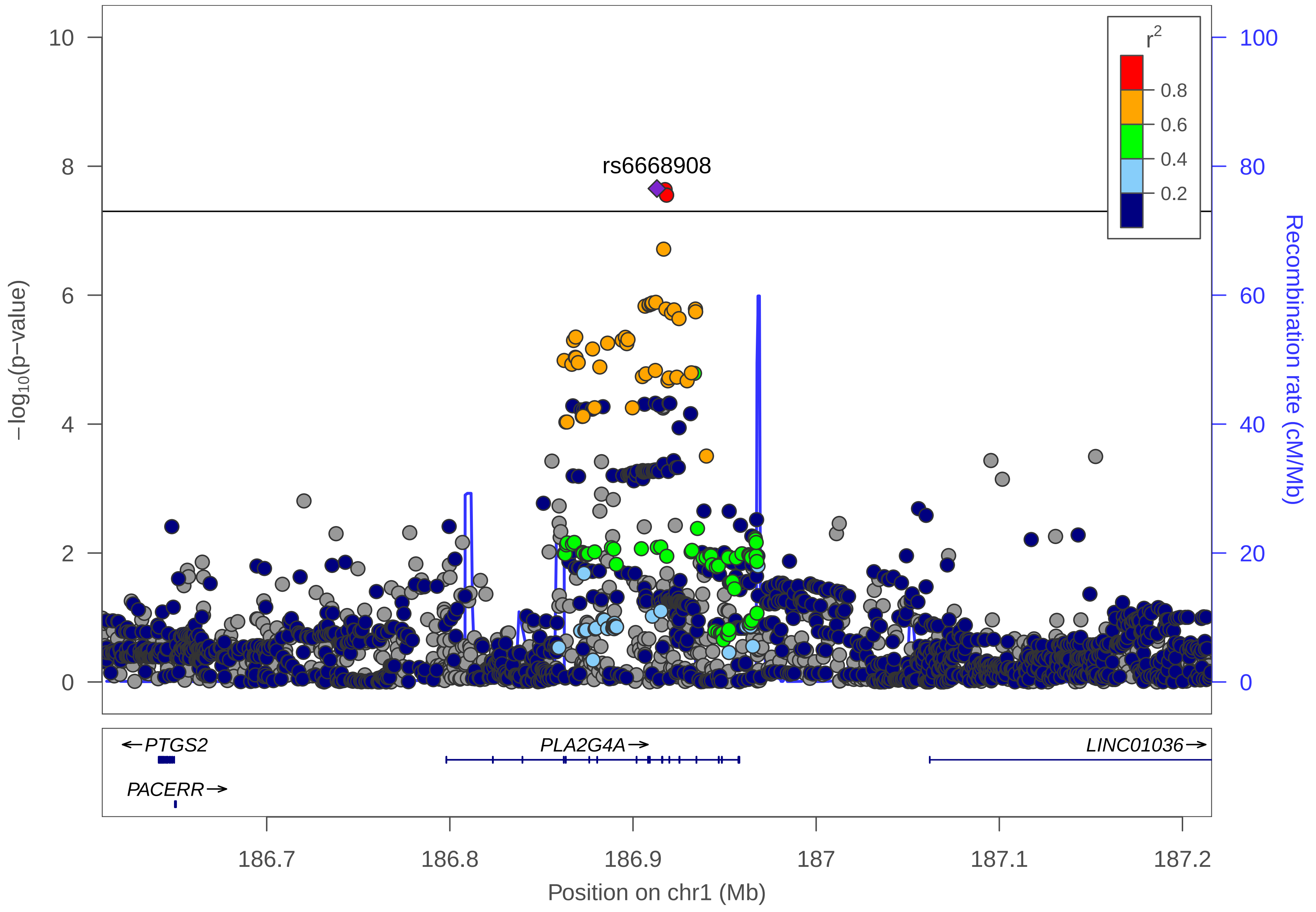

rs6668908
r2
0.8
0.6
0.4
0.2
10
8
6
4
2
0
100
80
60
40
20
0
−log10(p−value)
Recombination rate (cM/Mb)
←PTGS2
PLA2G4A→
LINC01036→
PACERR→
186.7
186.8
186.9
187
187.1
187.2
Position on chr1 (Mb)

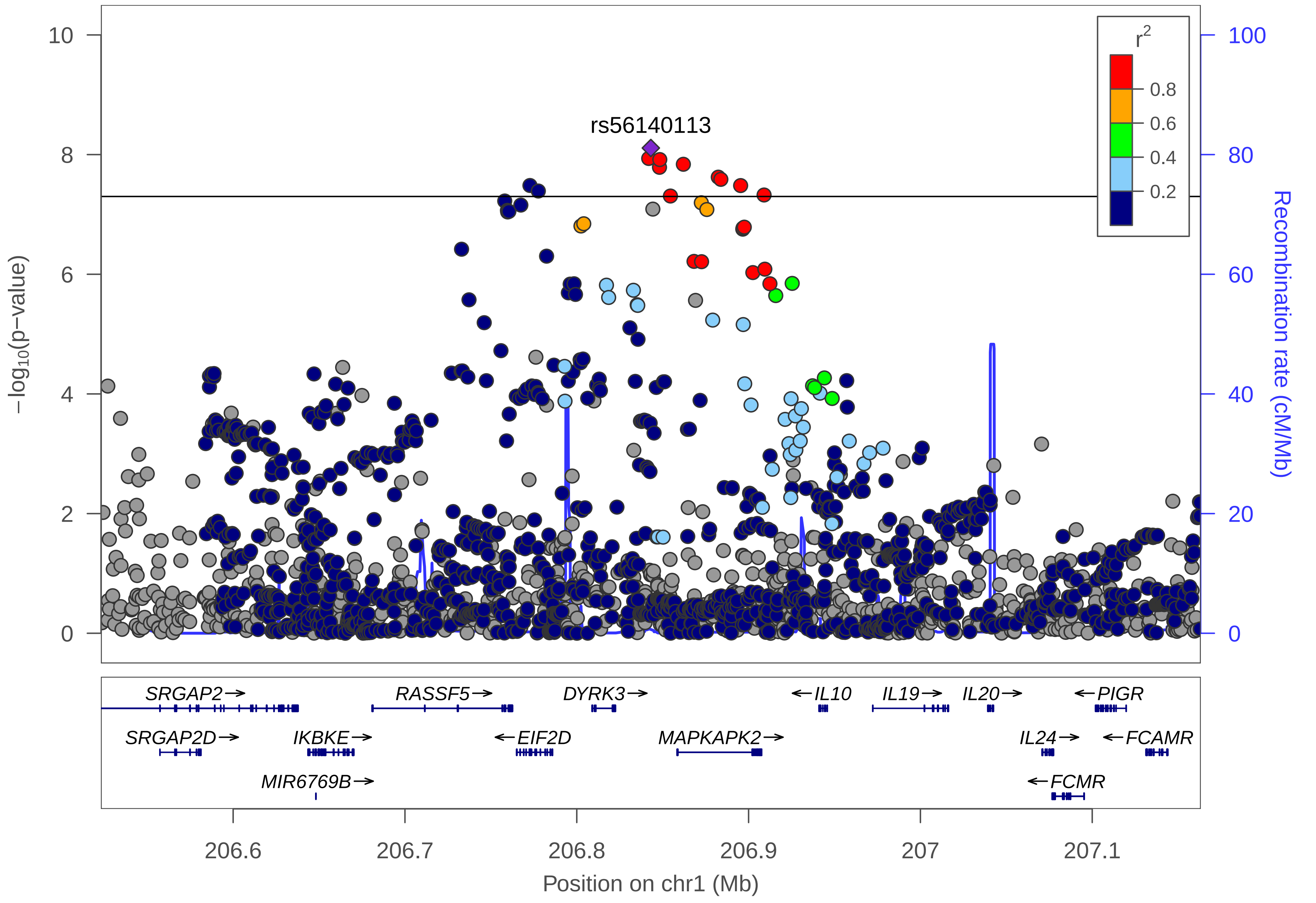
rs56140113
r2
0.8
0.6
0.4
0.2
−log10(p−value)
Recombination rate (cM/Mb)
0
2
4
6
8
10
0
20
40
60
80
100
SRGAP2→
RASSF5→
DYRK3→
←IL10
IL19→
IL20→
←PIGR
SRGAP2D→
IKBKE→
←EIF2D
MAPKAPK2→
IL24→
←FCAMR
MIR6769B→
←FCMR
206.6
206.7
206.8
206.9
207
207.1
Position on chr1 (Mb)

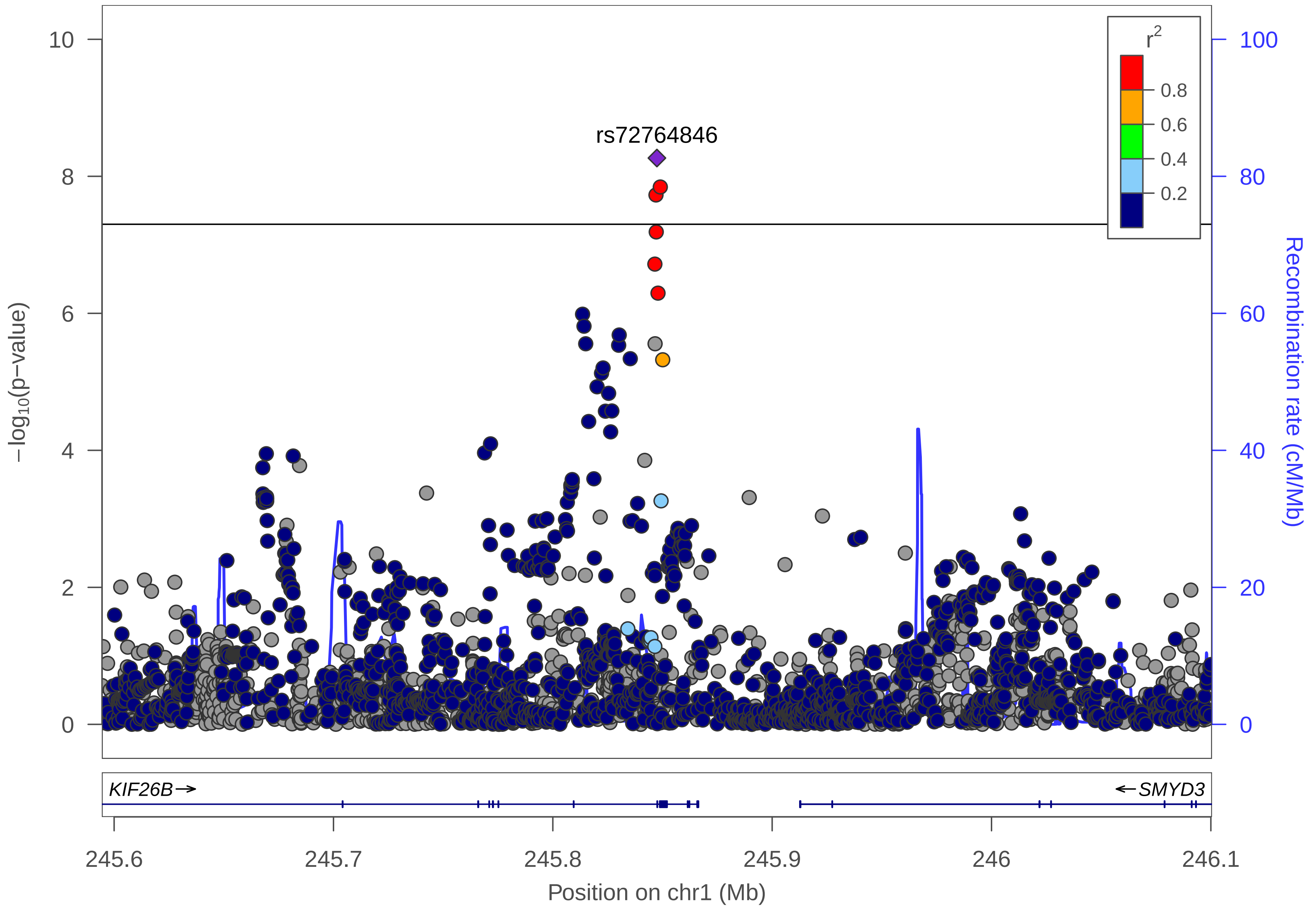
rs72764846
r2
0.8
0.6
0.4
0.2
−log10(p−value)
Recombination rate (cM/Mb)
KIF26B→
←SMYD3
245.6
245.7
245.8
245.9
246
246.1
Position on chr1 (Mb)

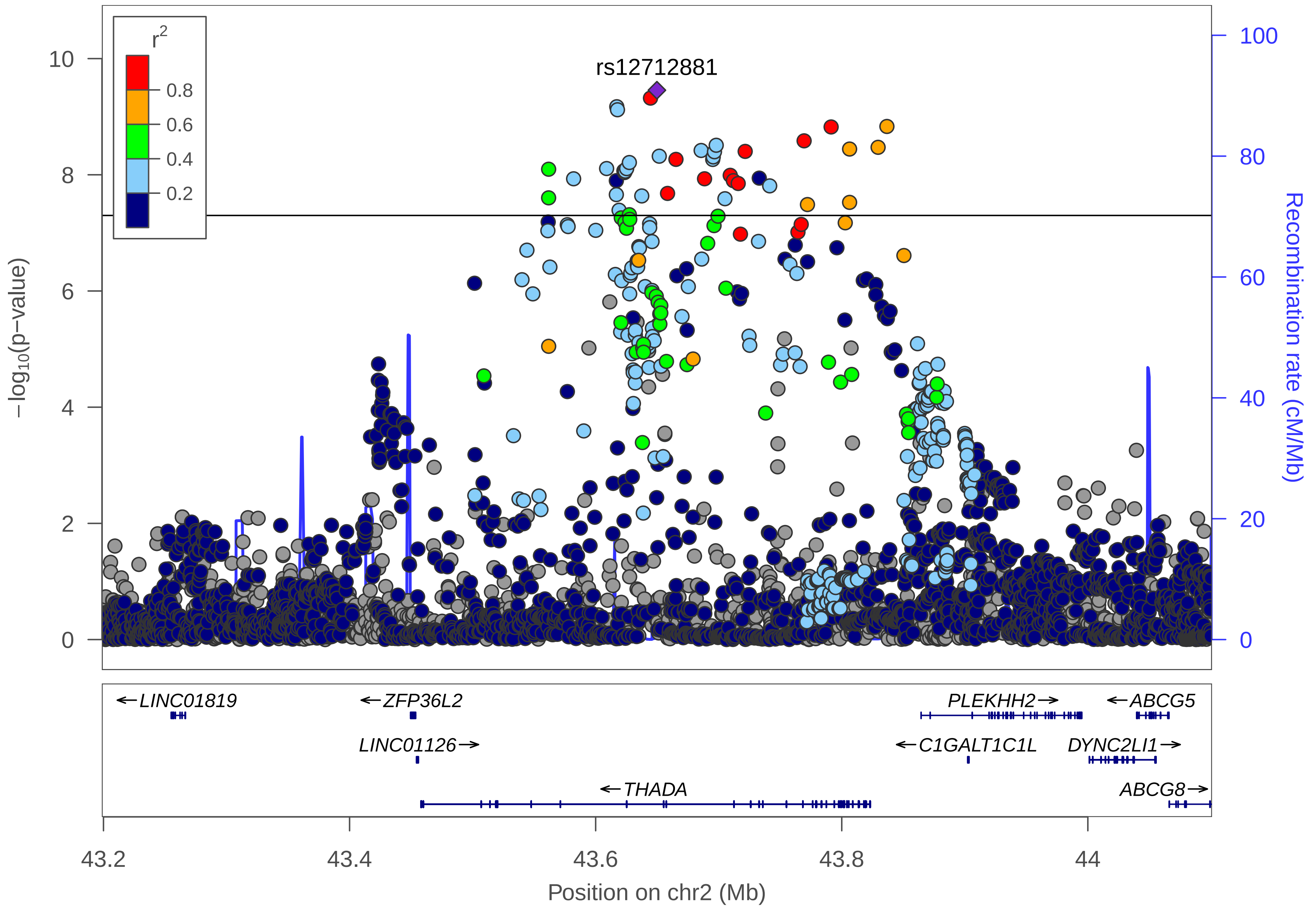

rs12712881
$r^2$
0.8
0.6
0.4
0.2
10
8
6
4
2
0
100
80
60
40
20
0
$-\log_{10}$(p-value)
Recombination rate (cM/Mb)
←LINC01819
←ZFP36L2
PLEKHH2→
←ABCG5
LINC01126→
←C1GALT1C1L
DYNC2LI1→
←THADA
ABCG8→
43.2
43.4
43.6
43.8
44
Position on chr2 (Mb)

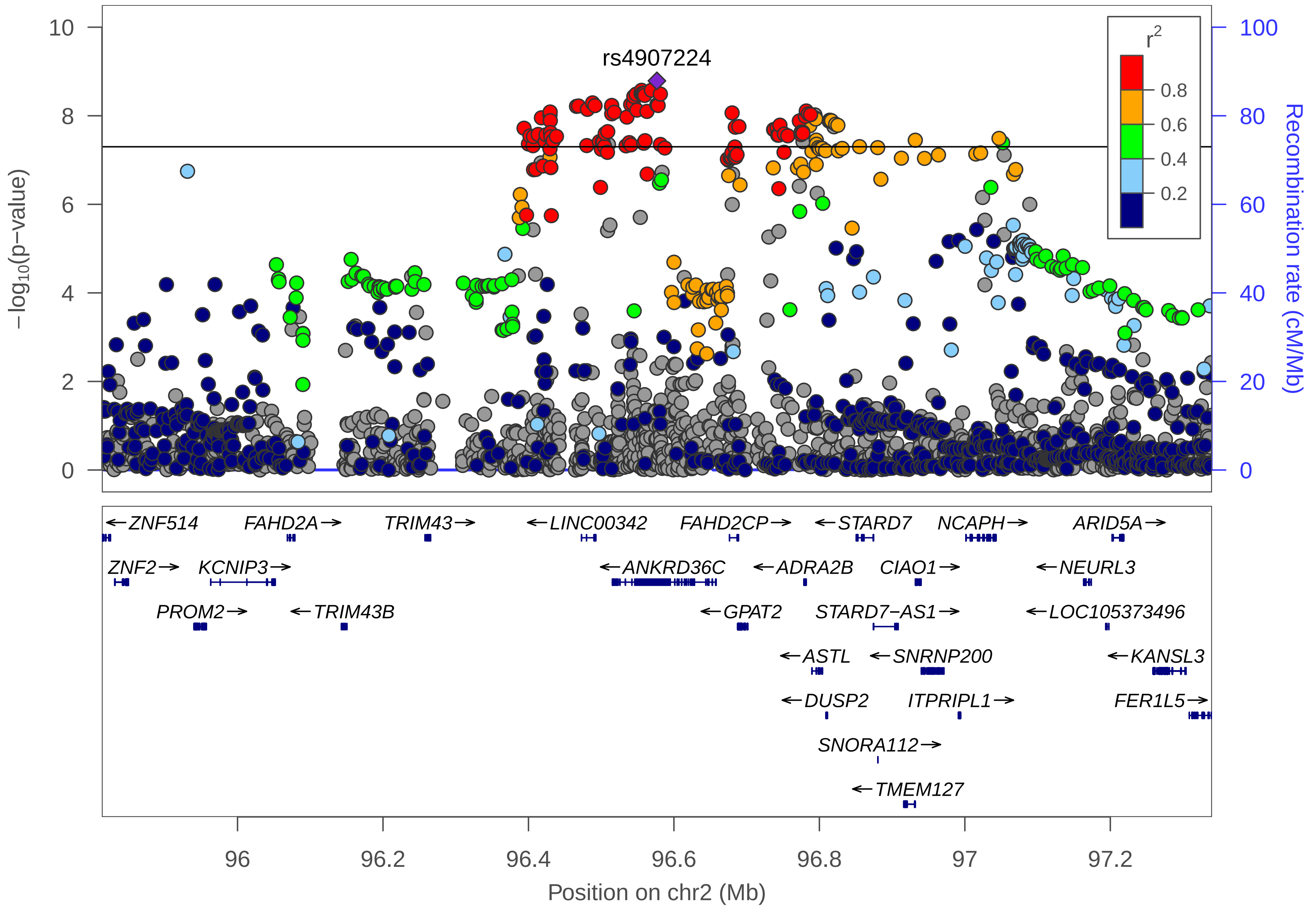
rs4907224
r2
0.8
0.6
0.4
0.2
−log10(p−value)
Recombination rate (cM/Mb)
10
8
6
4
2
0
100
80
60
40
20
0
ZNF514
FAHD2A
TRIM43
LINC00342
FAHD2CP
STARD7
NCAPH
ARID5A
ZNF2
KCNIP3
ANKRD36C
ADRA2B
CIAO1
NEURL3
PROM2
TRIM43B
GPAT2
STARD7−AS1
LOC105373496
ASTL
SNRNP200
KANSL3
DUSP2
ITPRIPL1
FER1L5
SNORA112
TMEM127
96
96.2
96.4
96.6
96.8
97
97.2
Position on chr2 (Mb)

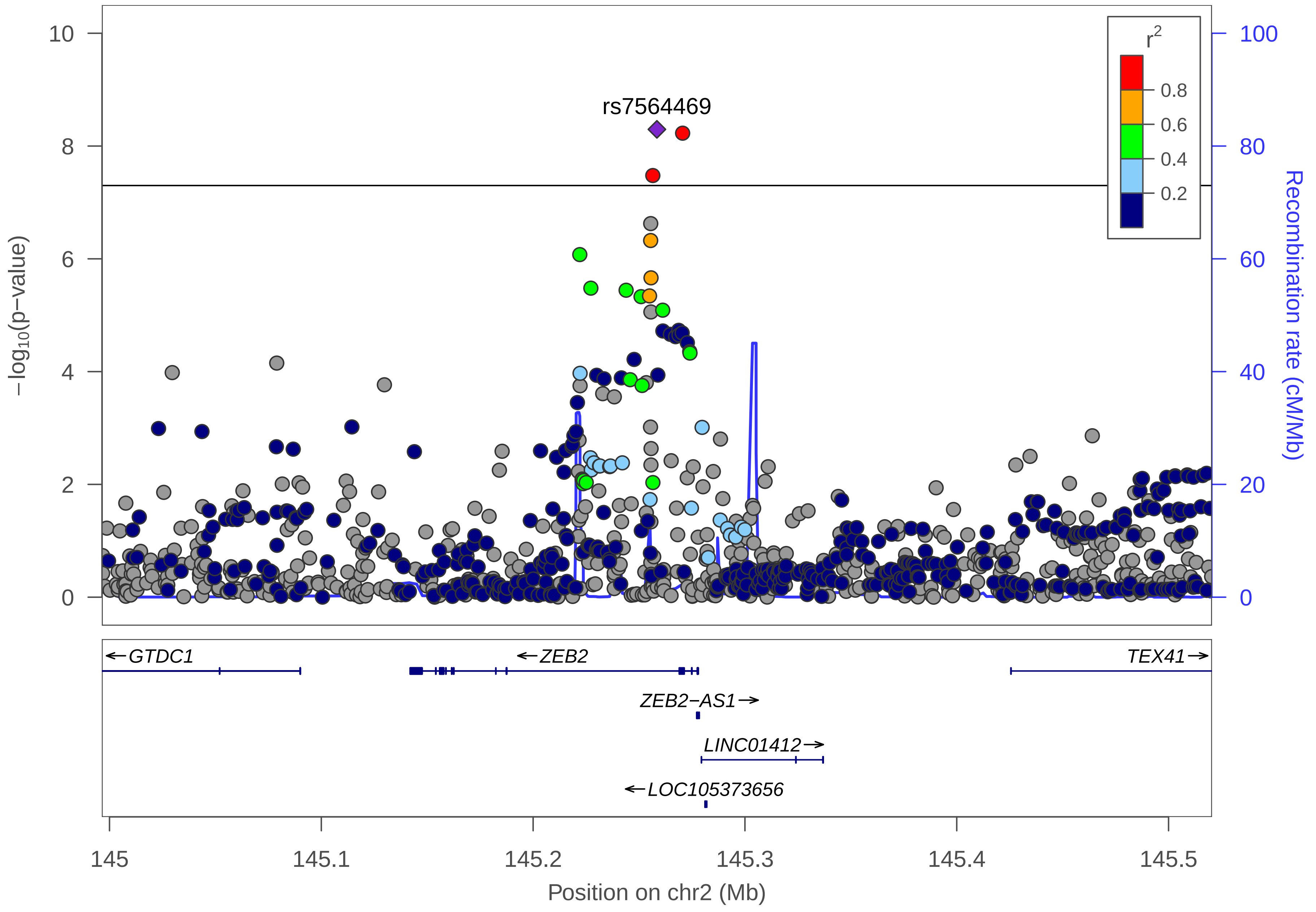
rs7564469
r2
0.8
0.6
0.4
0.2
−log10(p−value)
Recombination rate (cM/Mb)
10
8
6
4
2
0
100
80
60
40
20
0
←GTDC1
←ZEB2
TEX41→
ZEB2−AS1→
LINC01412→
←LOC105373656
145
145.1
145.2
145.3
145.4
145.5
Position on chr2 (Mb)

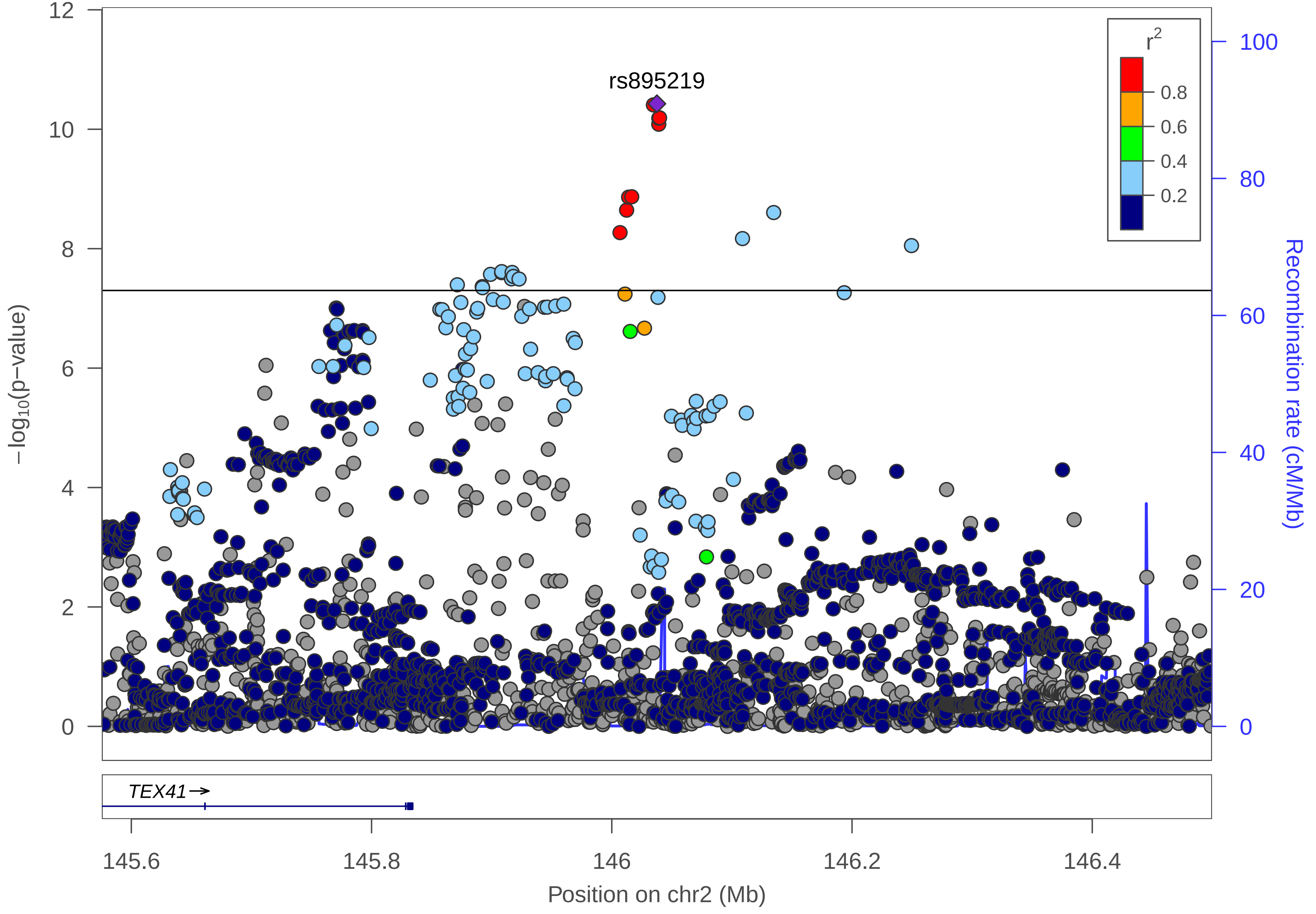

rs895219
r2
0.8
0.6
0.4
0.2
12
10
8
6
4
2
0
−log10(p−value)
100
80
60
40
20
0
Recombination rate (cM/Mb)
TEX41
145.6
145.8
146
146.2
146.4
Position on chr2 (Mb)

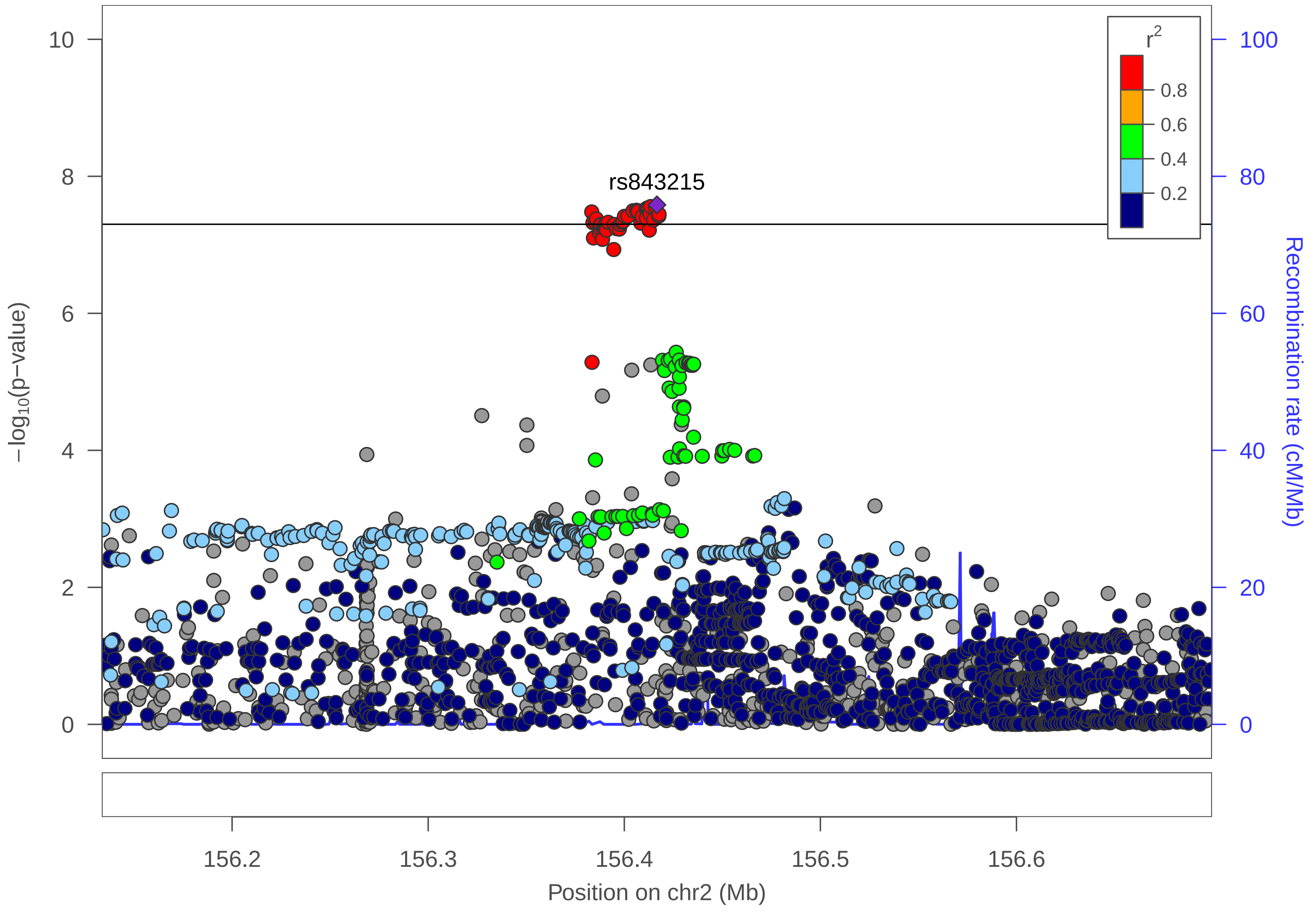

rs843215
r²
0.8
0.6
0.4
0.2
10
8
6
4
2
0
−log10(p−value)
100
80
60
40
20
0
Recombination rate (cM/Mb)
156.2
156.3
156.4
156.5
156.6
Position on chr2 (Mb)

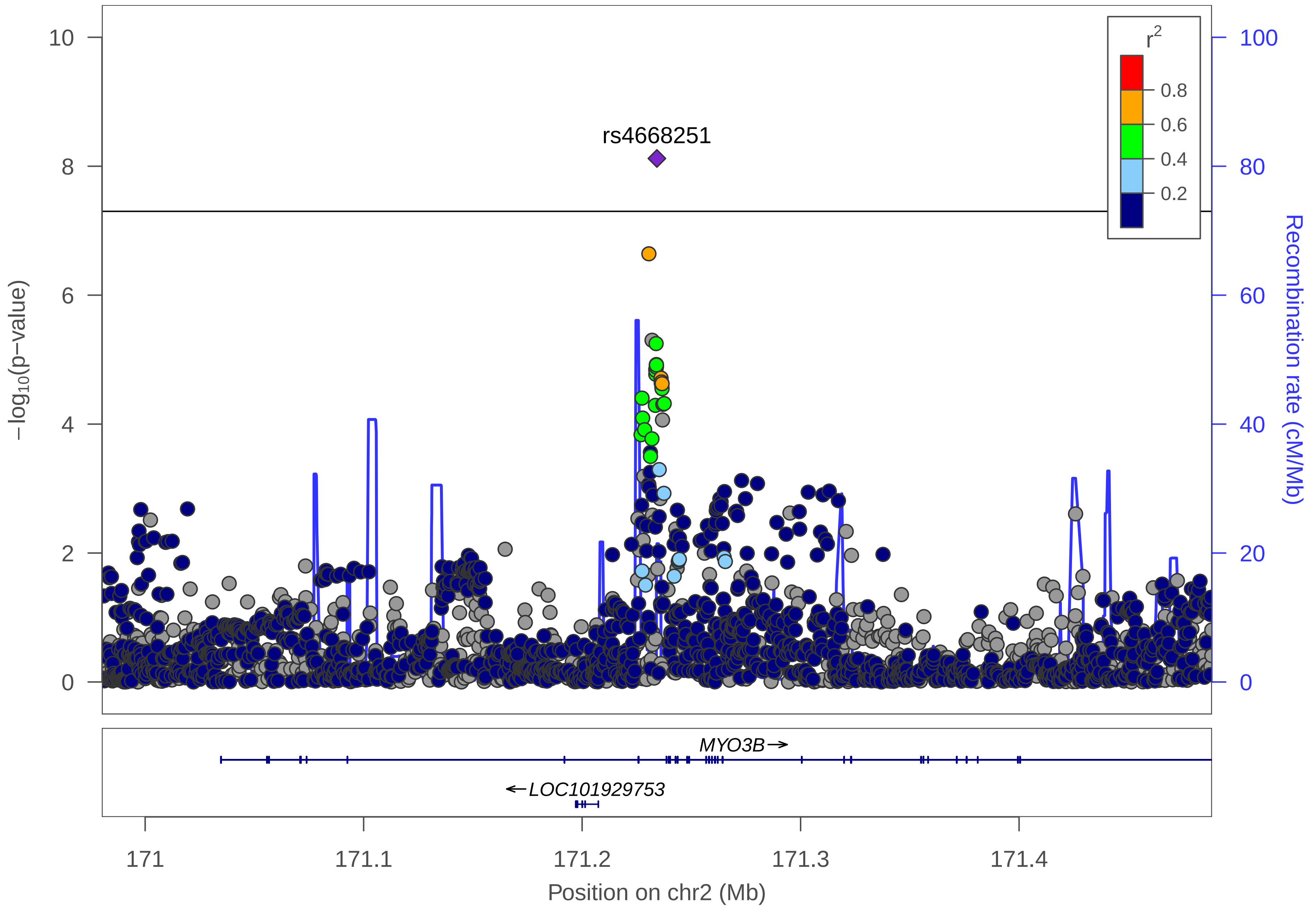

rs4668251
r2
0.8
0.6
0.4
0.2
10
8
6
4
2
0
100
80
60
40
20
0
−log10(p−value)
Recombination rate (cM/Mb)
MYO3B→
←LOC101929753
171
171.1
171.2
171.3
171.4
Position on chr2 (Mb)

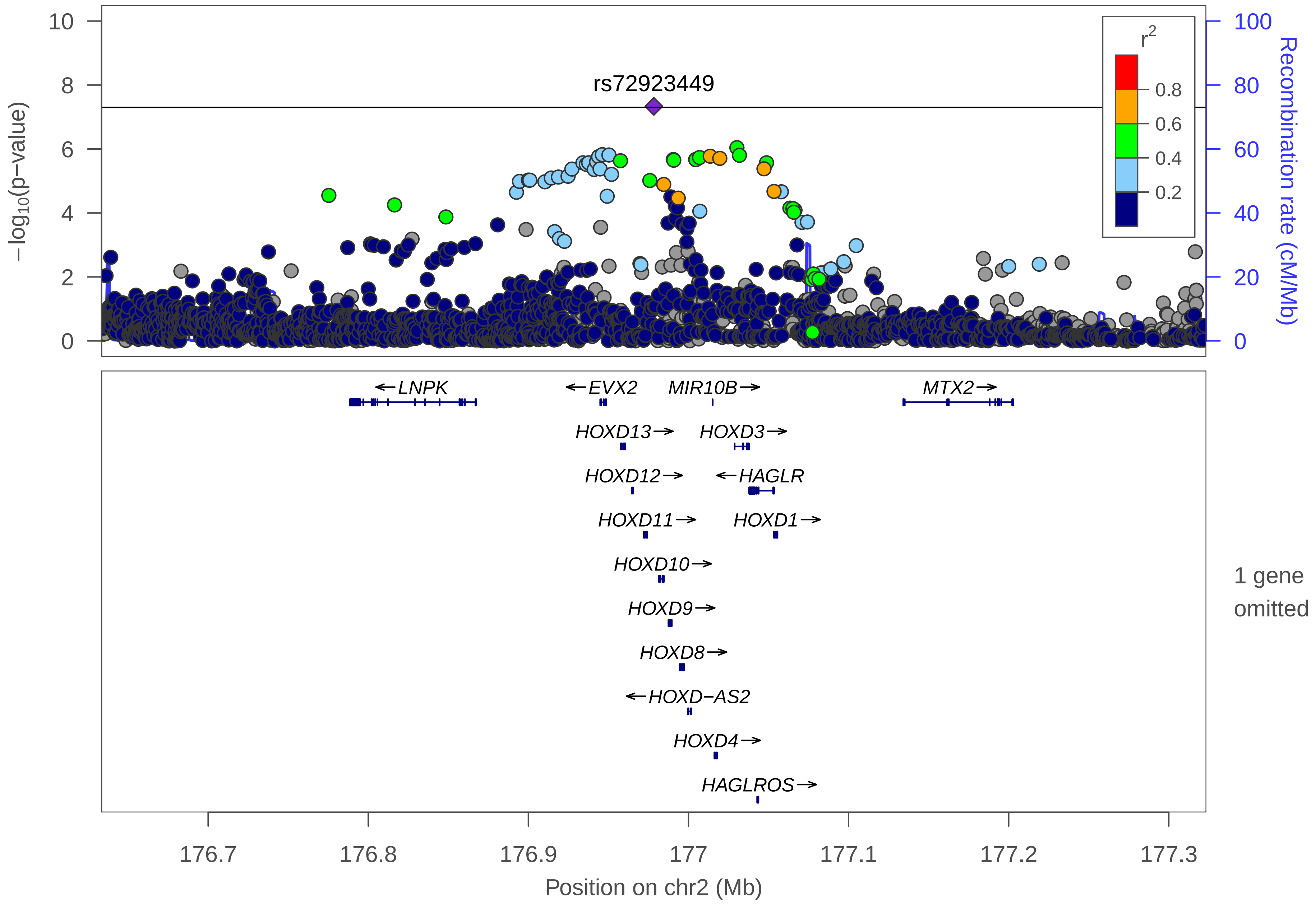
rs72923449
r2
0.8
0.6
0.4
0.2
−log10(p−value)
Recombination rate (cM/Mb)
LNPK
EVX2
MIR10B
MTX2
HOXD13
HOXD3
HOXD12
HAGLR
HOXD11
HOXD1
HOXD10
HOXD9
HOXD8
HOXD−AS2
HOXD4
HAGLROS
1 gene omitted
176.7
176.8
176.9
177
177.1
177.2
177.3
Position on chr2 (Mb)

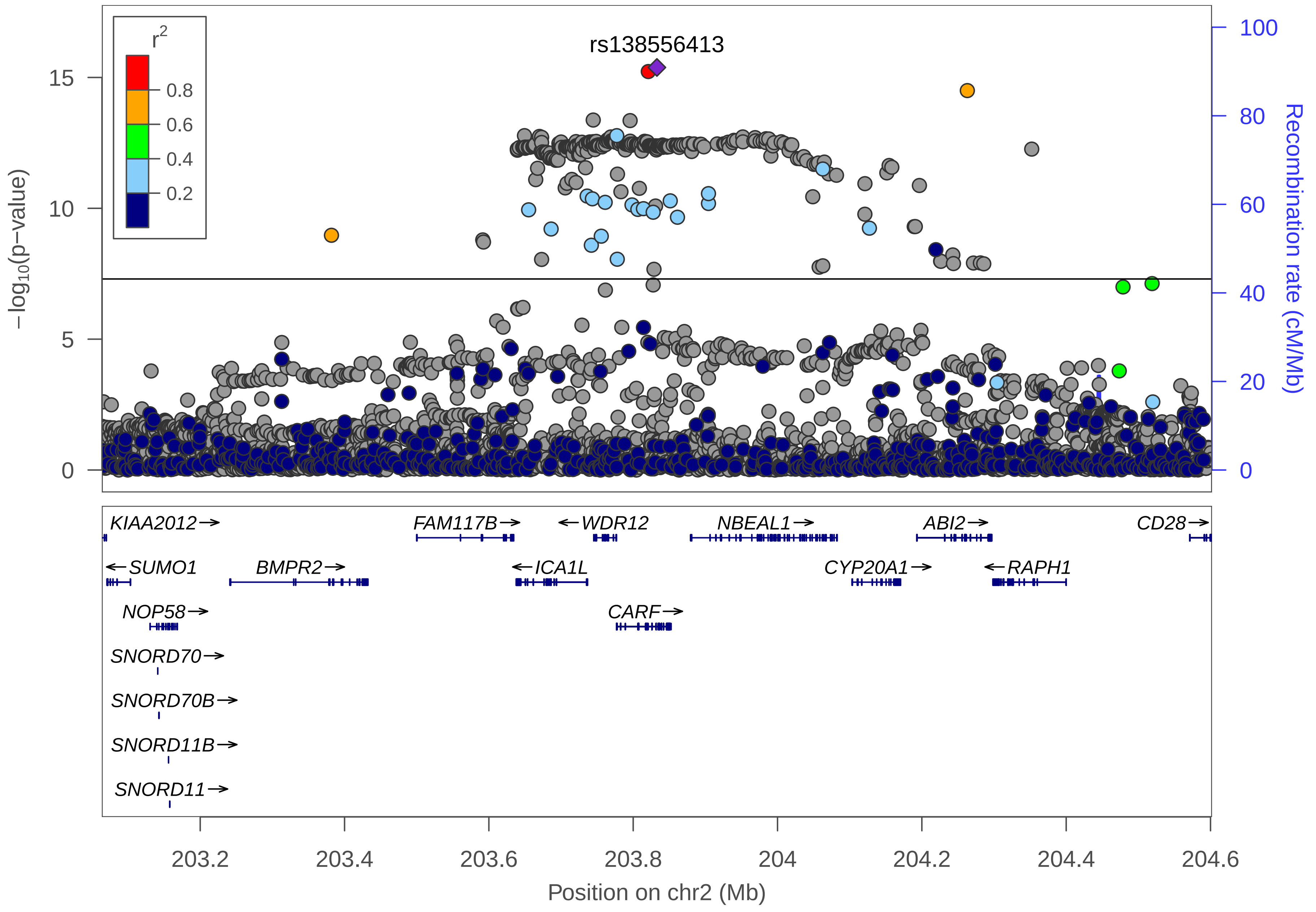
rs138556413
r2
0.8
0.6
0.4
0.2
−log10(p−value)
Recombination rate (cM/Mb)
KIAA2012→
FAM117B→
←WDR12
NBEAL1→
ABI2→
CD28→
←SUMO1
BMPR2→
←ICA1L
CYP20A1→
←RAPH1
NOP58→
CARF→
SNORD70→
SNORD70B→
SNORD11B→
SNORD11→
203.2
203.4
203.6
203.8
204
204.2
204.4
204.6
Position on chr2 (Mb)

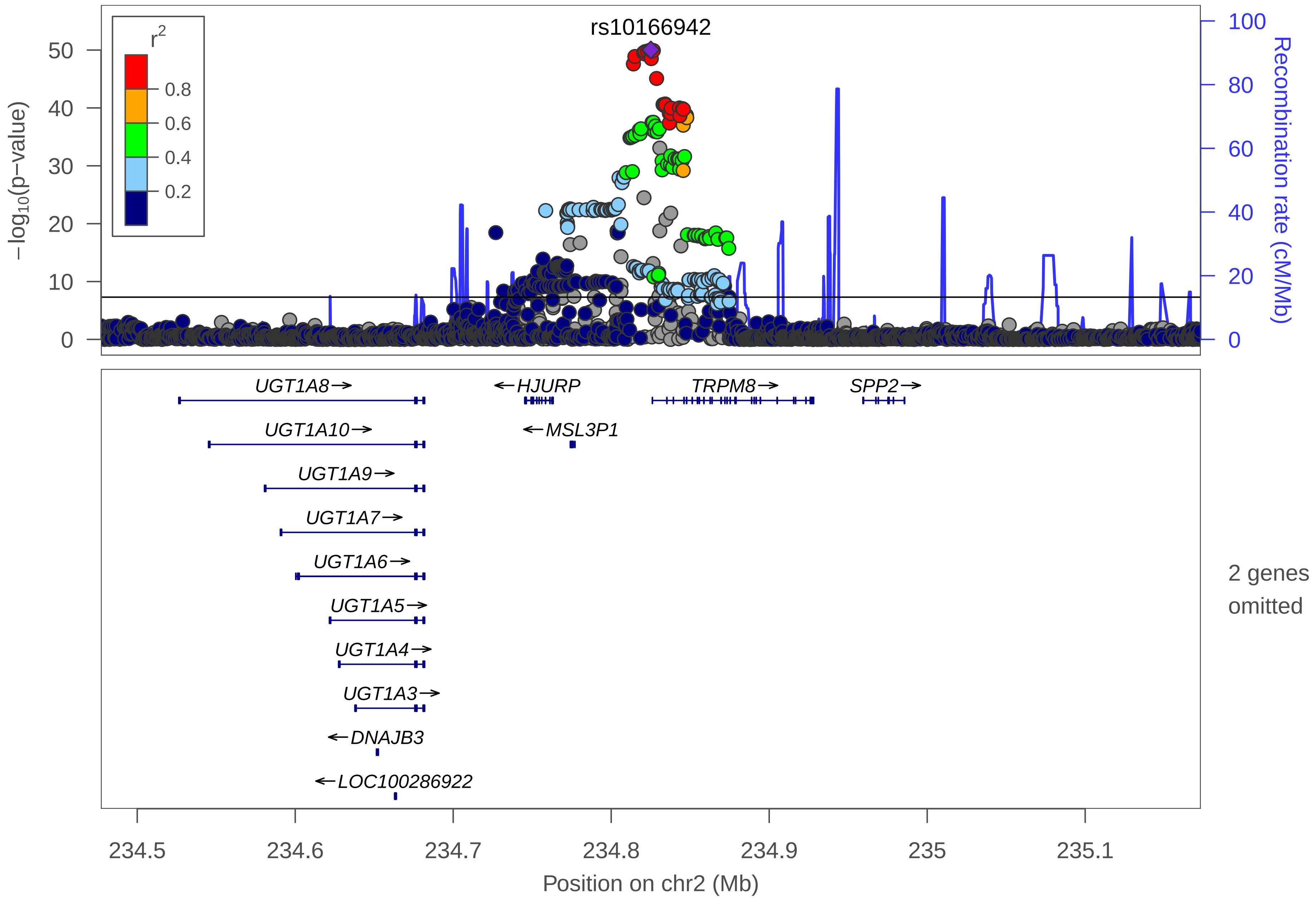

rs10166942
r2
0.8
0.6
0.4
0.2
−log10(p−value)
Recombination rate (cM/Mb)
UGT1A8→
←HJURP
TRPM8→
SPP2→
UGT1A10→
←MSL3P1
UGT1A9→
UGT1A7→
UGT1A6→
UGT1A5→
UGT1A4→
UGT1A3→
←DNAJB3
←LOC100286922
2 genes omitted
234.5
234.6
234.7
234.8
234.9
235
235.1
Position on chr2 (Mb)

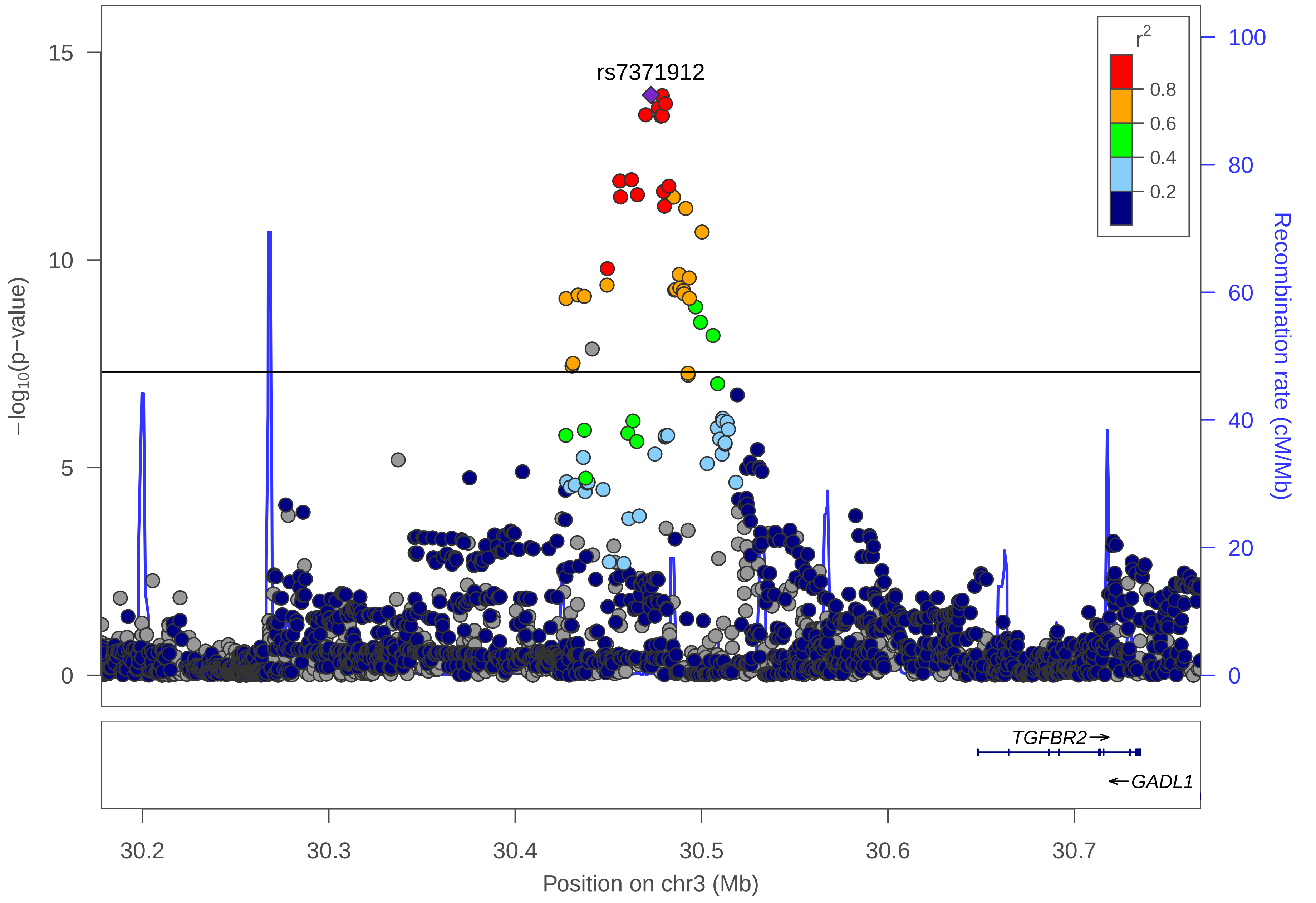

rs7371912
r²
0.8
0.6
0.4
0.2
15
10
5
0
−log10(p−value)
100
80
60
40
20
0
Recombination rate (cM/Mb)
TGFBR2→
←GADL1
30.2
30.3
30.4
30.5
30.6
30.7
Position on chr3 (Mb)

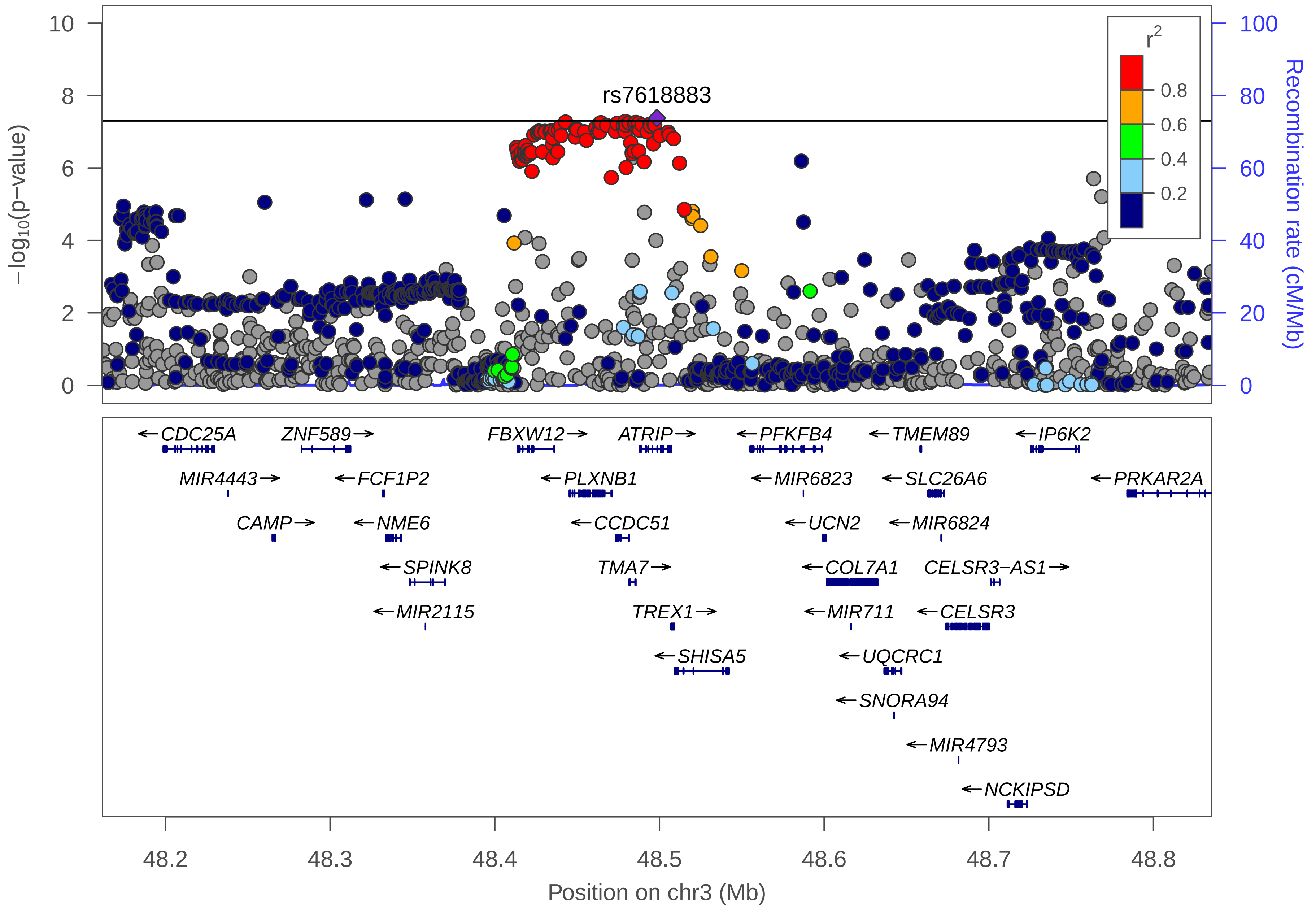

rs7618883
r2
0.8
0.6
0.4
0.2
−log10(p−value)
Recombination rate (cM/Mb)
CDC25A
ZNF589
FBXW12
ATRIP
PFKFB4
TMEM89
IP6K2
MIR4443
FCF1P2
PLXNB1
MIR6823
SLC26A6
PRKAR2A
CAMP
NME6
CCDC51
UCN2
MIR6824
SPINK8
TMA7
COL7A1
CELSR3−AS1
MIR2115
TREX1
MIR711
CELSR3
SHISA5
UQCRC1
SNORA94
MIR4793
NCKIPSD
Position on chr3 (Mb)

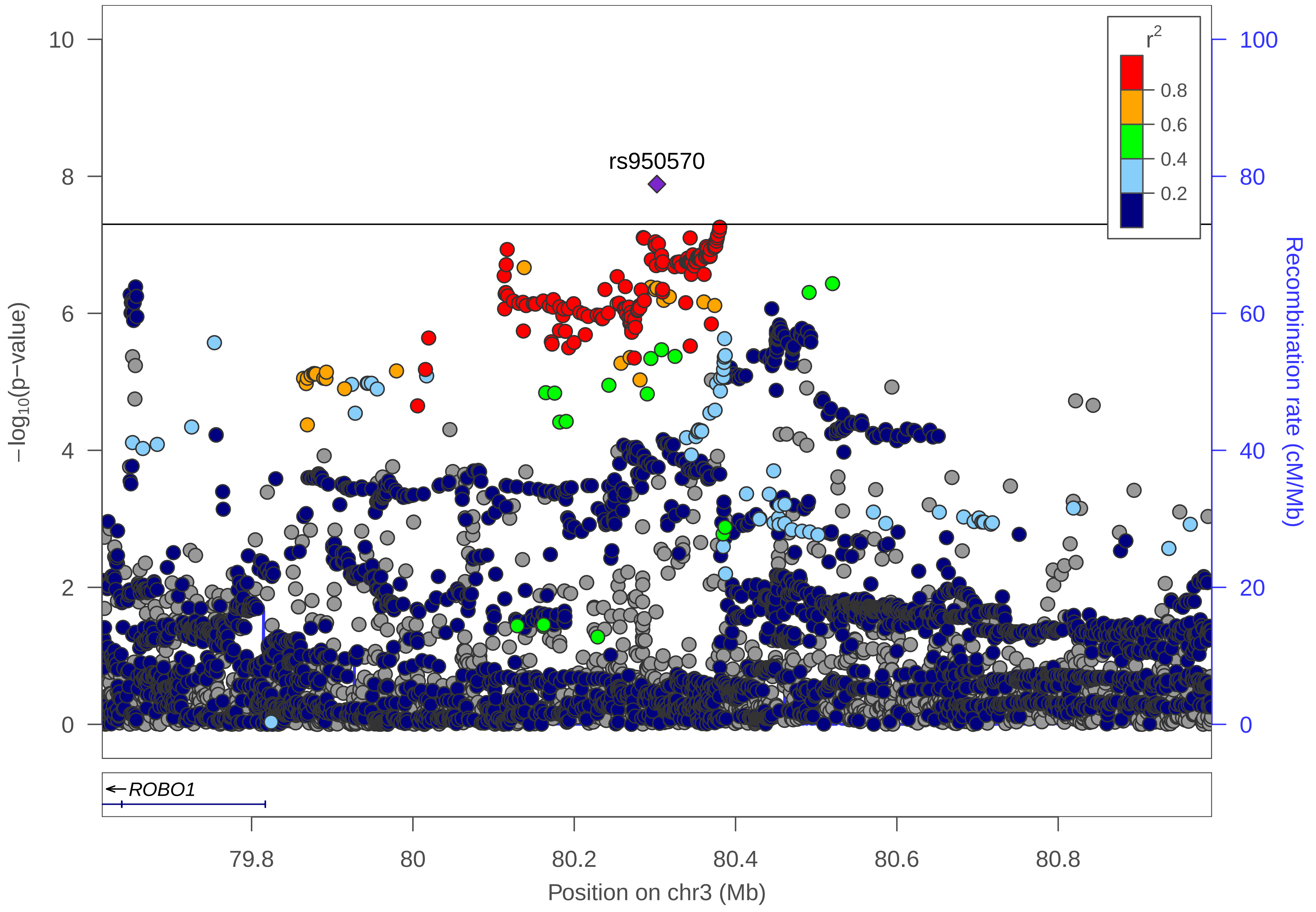

rs950570
r^2
0.8
0.6
0.4
0.2
−log10(p−value)
Recombination rate (cM/Mb)
0
2
4
6
8
10
0
20
40
60
80
100
ROBO1
79.8
80
80.2
80.4
80.6
80.8
Position on chr3 (Mb)

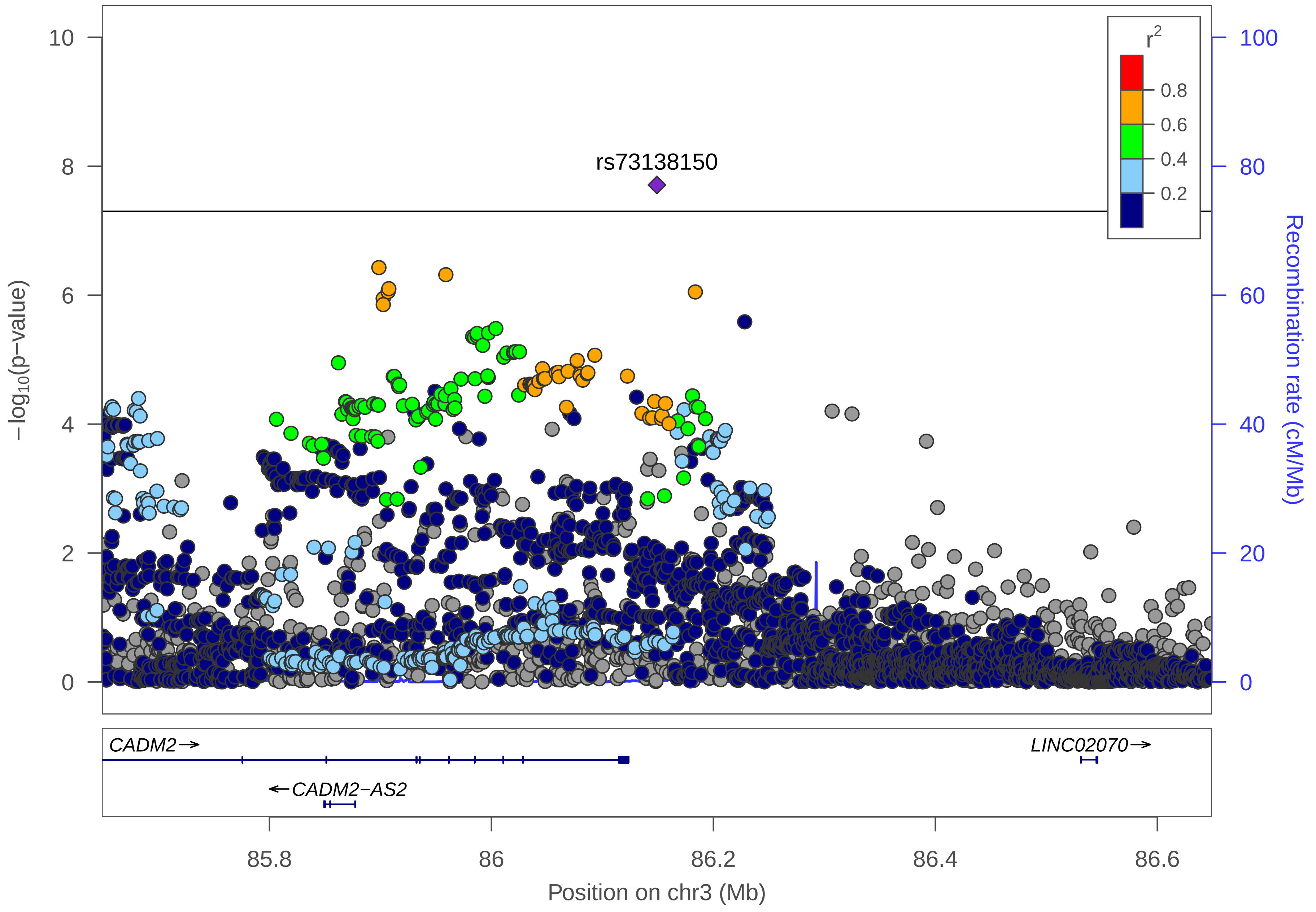

rs73138150
r2
0.8
0.6
0.4
0.2
−log10(p−value)
Recombination rate (cM/Mb)
10
8
6
4
2
0
100
80
60
40
20
0
CADM2→
←CADM2−AS2
LINC02070→
85.8
86
86.2
86.4
86.6
Position on chr3 (Mb)

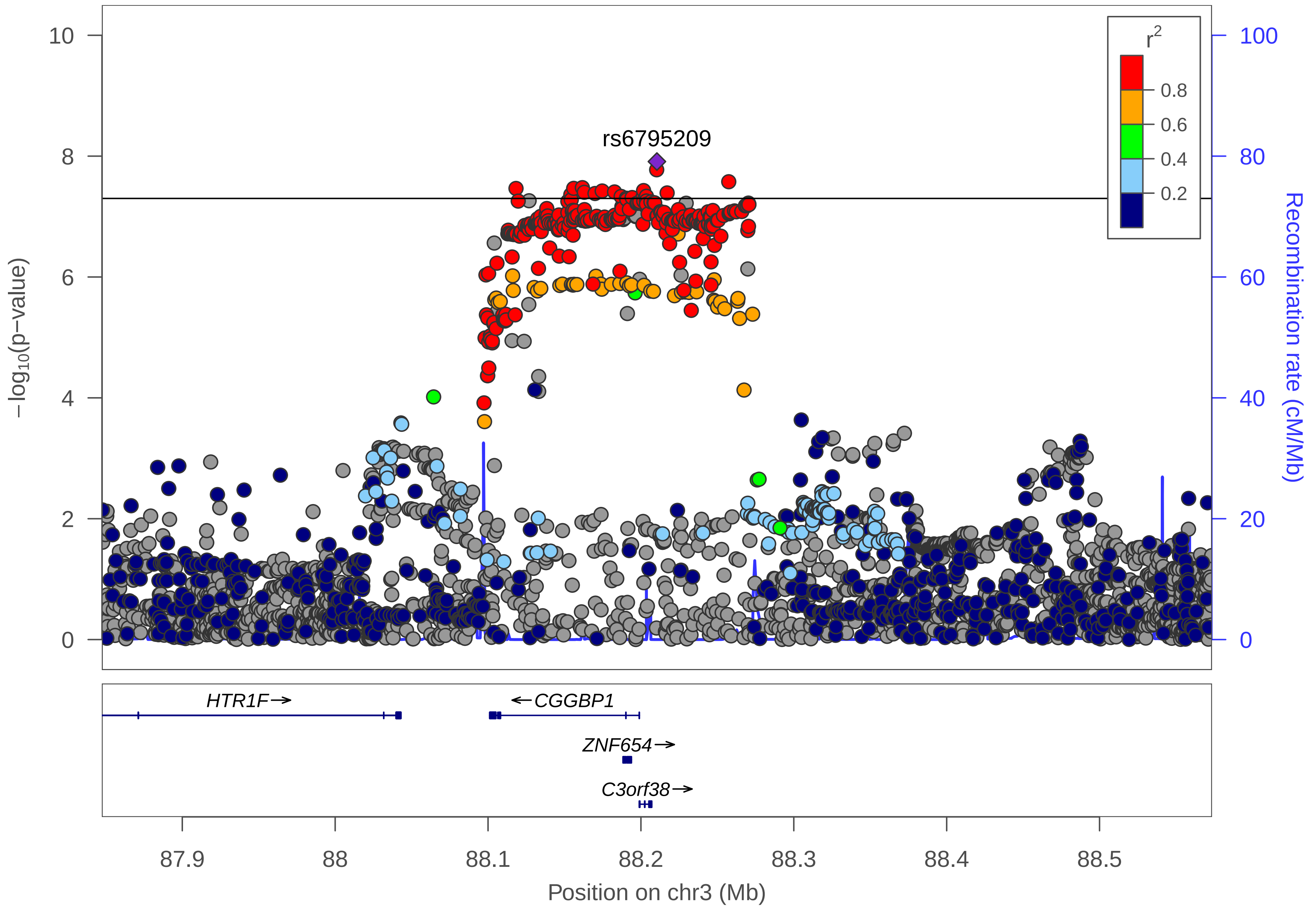
rs6795209
r2
0.8
0.6
0.4
0.2
−log10(p−value)
Recombination rate (cM/Mb)
10
8
6
4
2
0
100
80
60
40
20
0
HTR1F→
←CGGBP1
ZNF654→
C3orf38→
87.9
88
88.1
88.2
88.3
88.4
88.5
Position on chr3 (Mb)

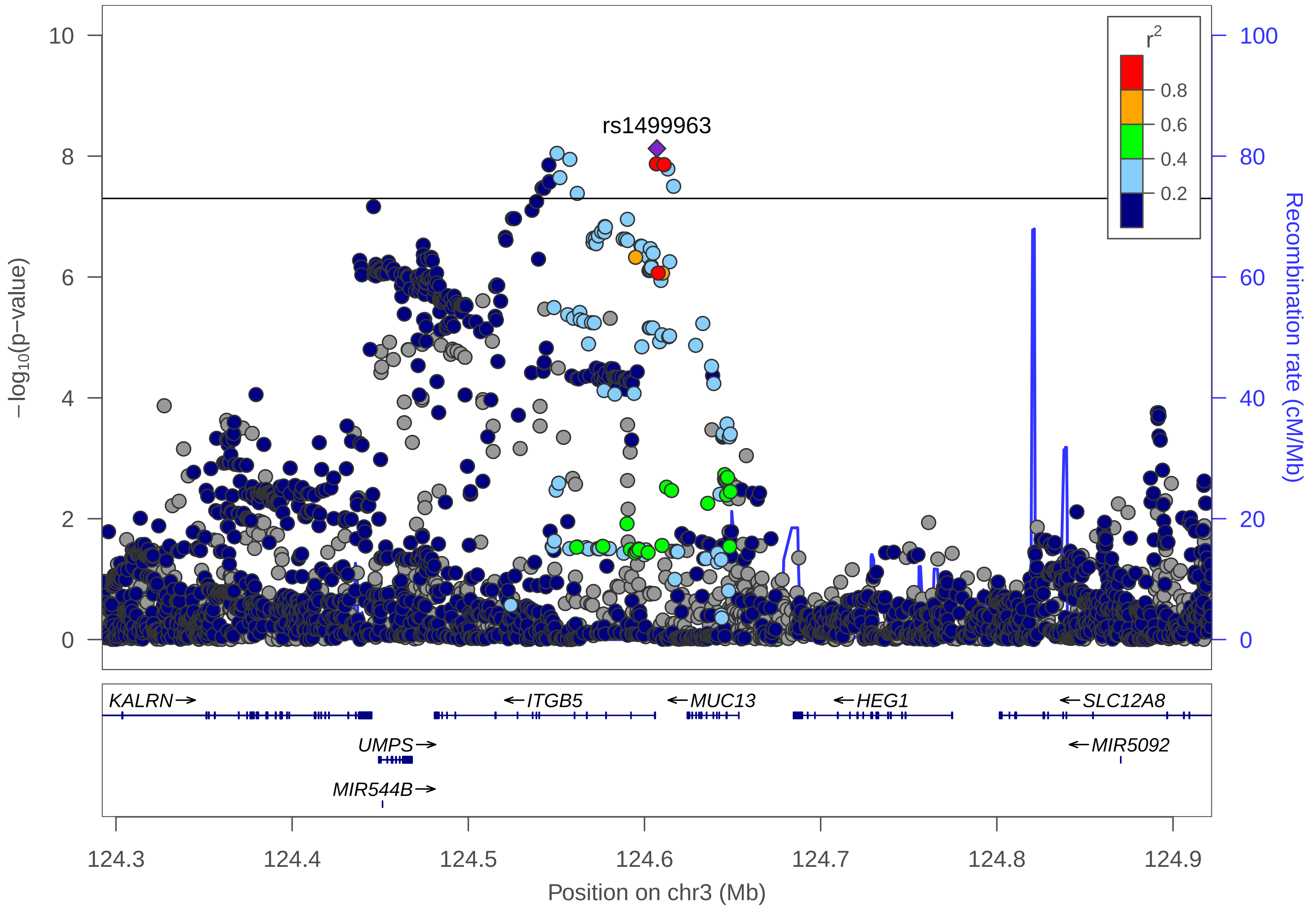

rs1499963
r²
0.8
0.6
0.4
0.2
−log10(p−value)
Recombination rate (cM/Mb)
10
8
6
4
2
0
100
80
60
40
20
0
KALRN→
←ITGB5
←MUC13
←HEG1
←SLC12A8
UMPS→
←MIR5092
MIR544B→
124.3
124.4
124.5
124.6
124.7
124.8
124.9
Position on chr3 (Mb)

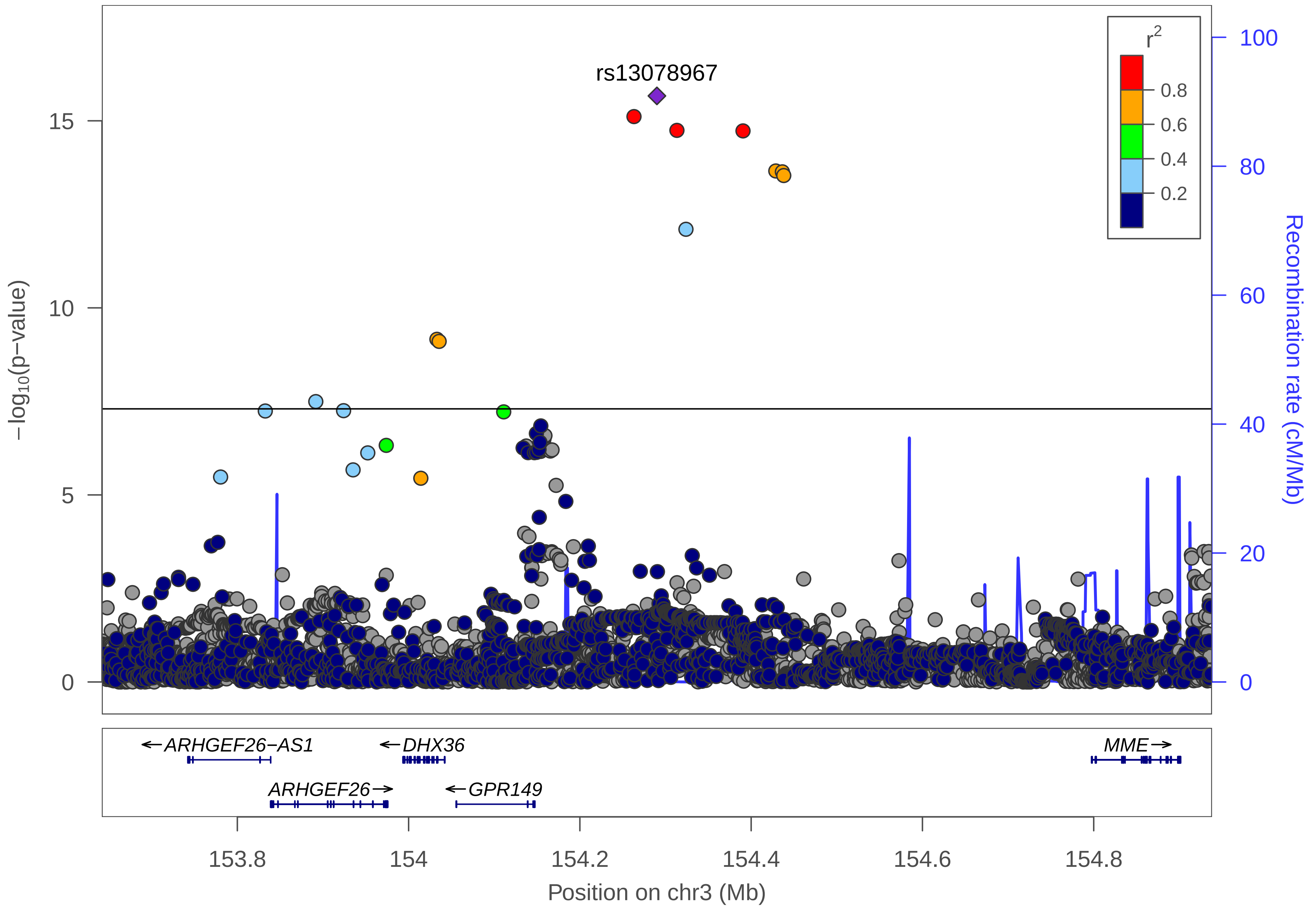

rs13078967
r2
0.8
0.6
0.4
0.2
−log10(p−value)
15
10
5
0
Recombination rate (cM/Mb)
100
80
60
40
20
0
←ARHGEF26−AS1
←DHX36
MME→
ARHGEF26→
←GPR149
153.8
154
154.2
154.4
154.6
154.8
Position on chr3 (Mb)

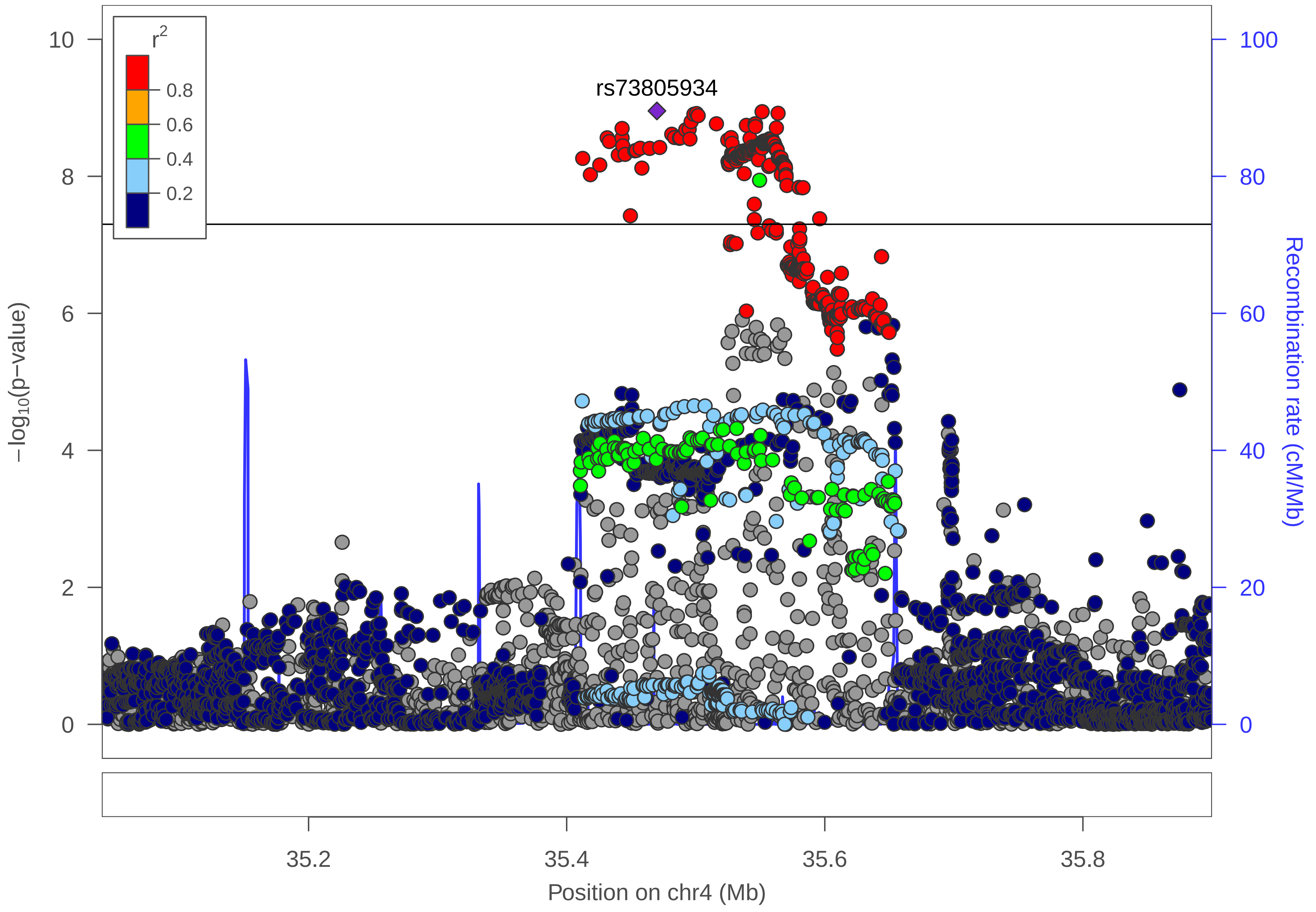

r2
0.8
0.6
0.4
0.2
rs73805934
10
8
6
4
2
0
−log10(p−value)
100
80
60
40
20
0
Recombination rate (cM/Mb)
35.2
35.4
35.6
35.8
Position on chr4 (Mb)

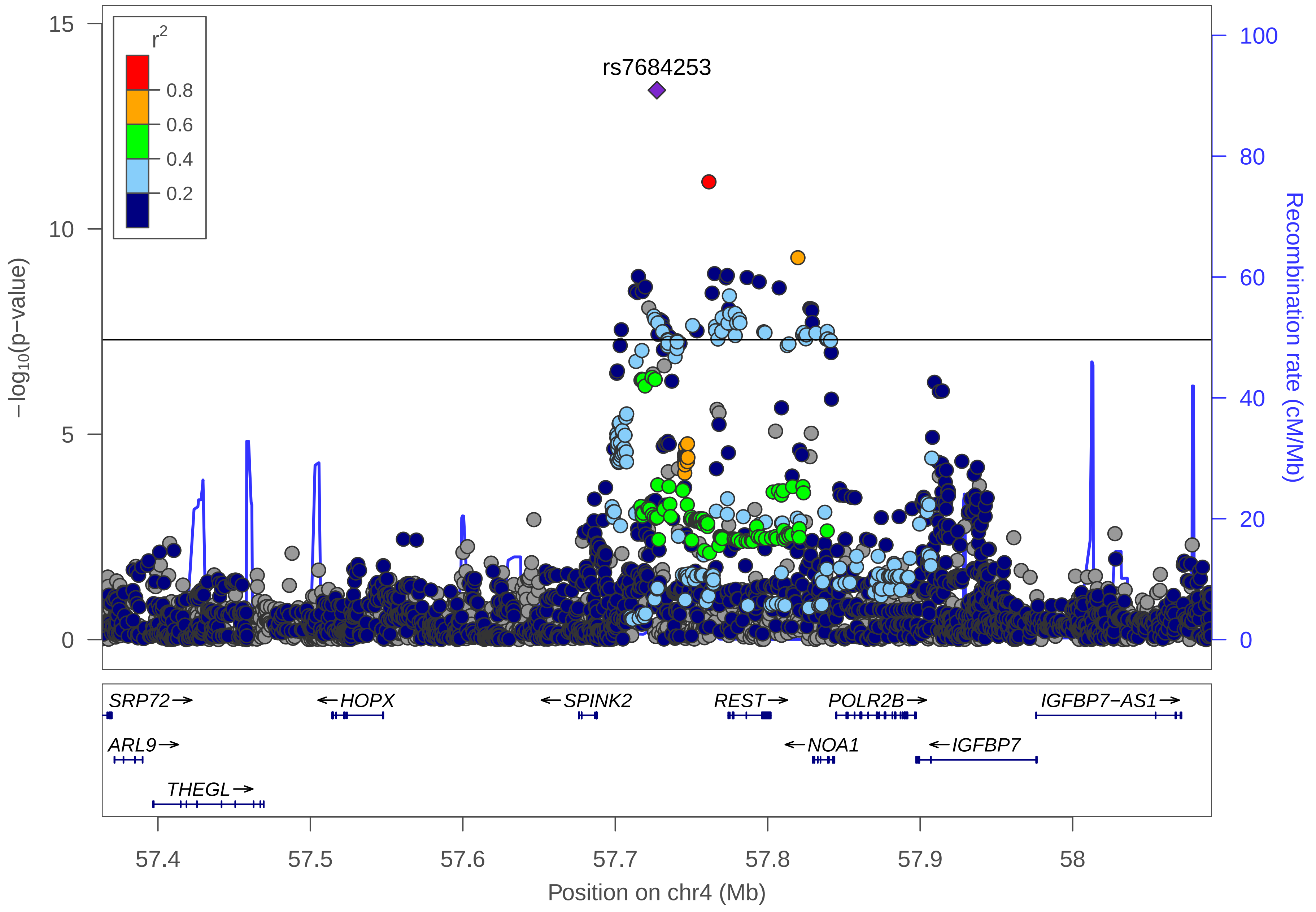

r2
0.8
0.6
0.4
0.2
rs7684253
15
10
5
0
−log10(p−value)
100
80
60
40
20
0
Recombination rate (cM/Mb)
SRP72→
←HOPX
←SPINK2
REST→
POLR2B→
IGFBP7−AS1→
ARL9→
←NOA1
←IGFBP7
THEGL→
57.4
57.5
57.6
57.7
57.8
57.9
58
Position on chr4 (Mb)

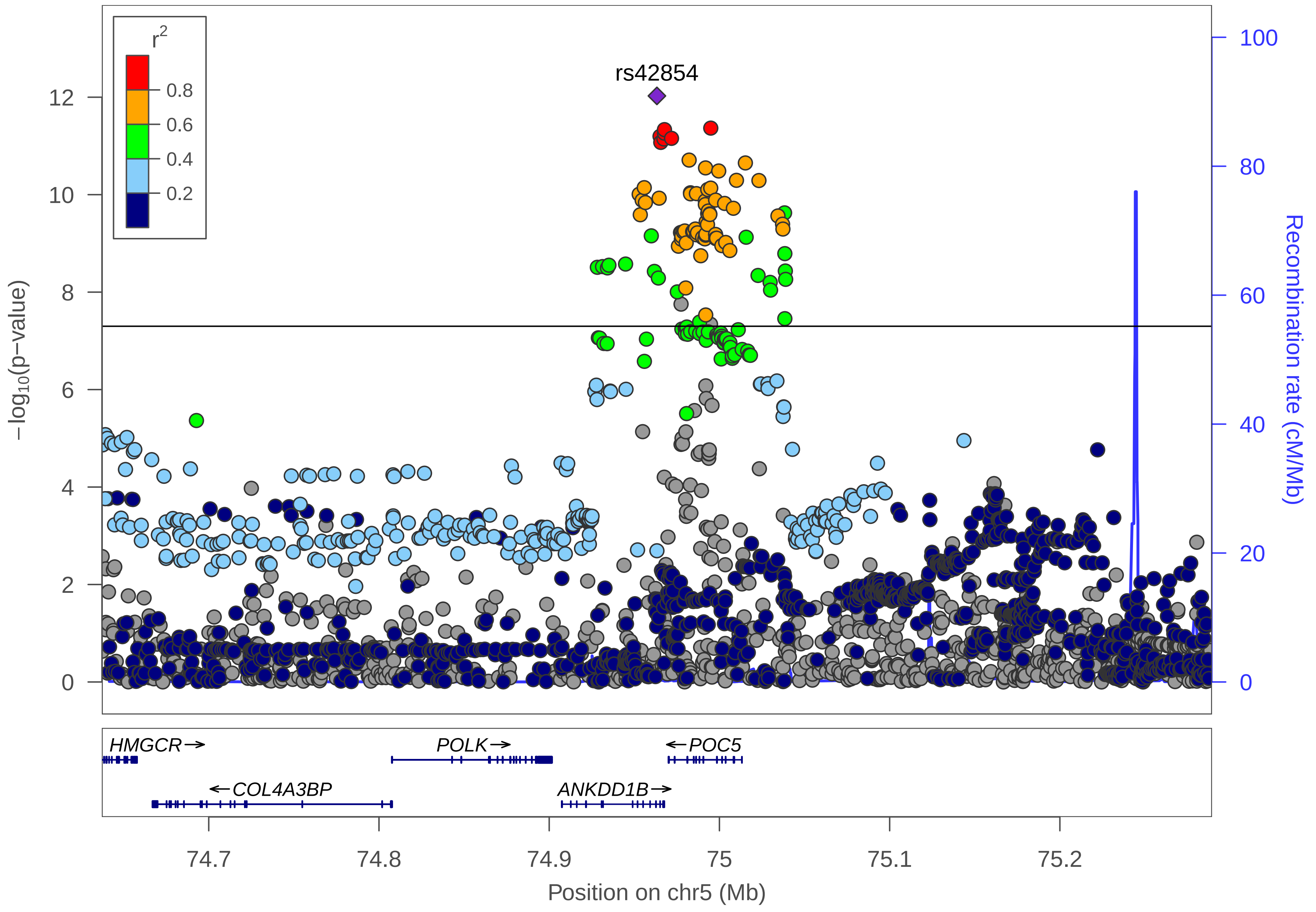

r²
0.8
0.6
0.4
0.2
rs42854
12
10
8
6
4
2
0
−log10(p−value)
100
80
60
40
20
0
Recombination rate (cM/Mb)
HMGCR→
POLK→
←POC5
←COL4A3BP
ANKDD1B→
74.7
74.8
74.9
75
75.1
75.2
Position on chr5 (Mb)

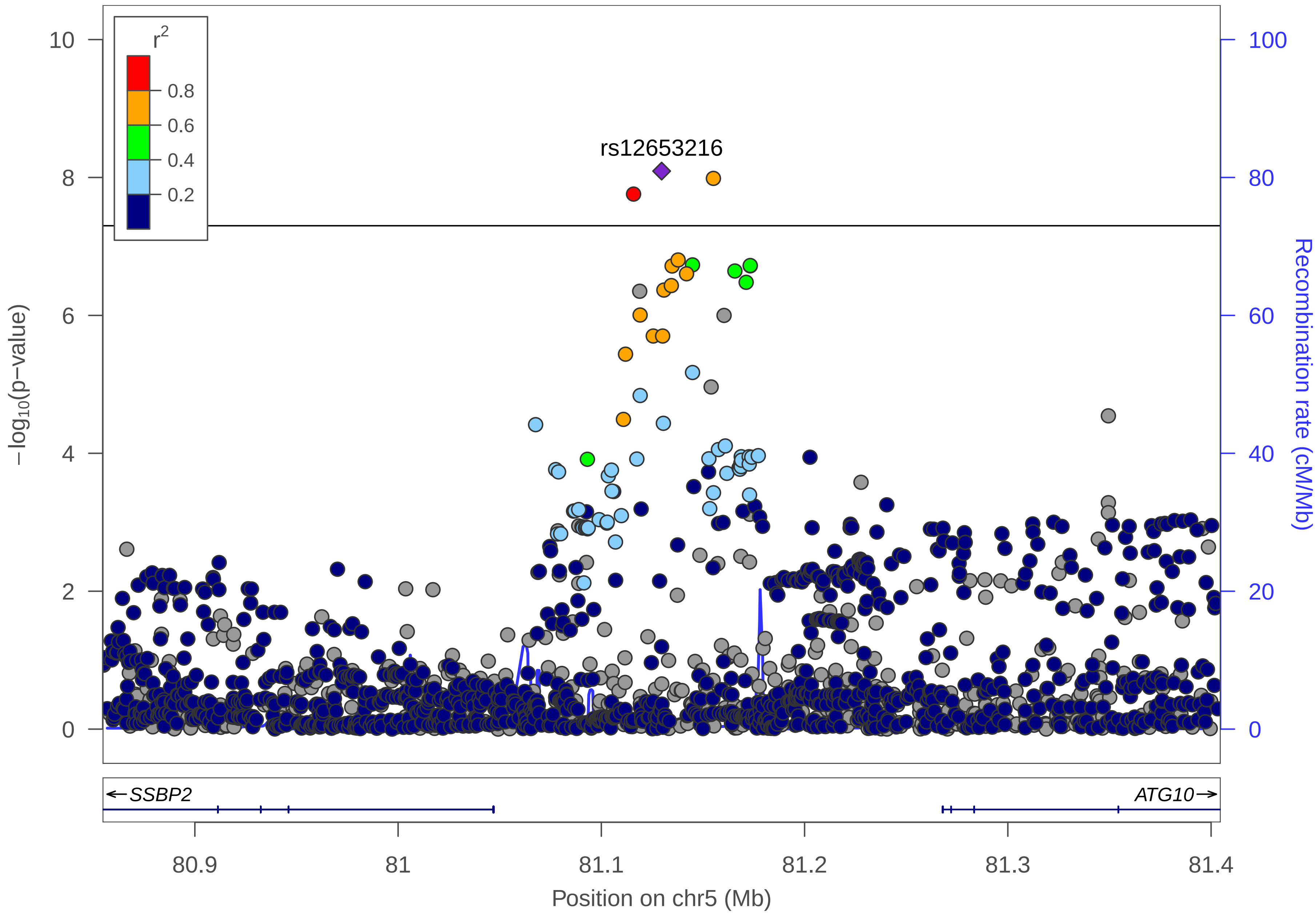

r2
0.8
0.6
0.4
0.2
rs12653216
−log10(p−value)
Recombination rate (cM/Mb)
SSBP2
ATG10
80.9
81
81.1
81.2
81.3
81.4
Position on chr5 (Mb)

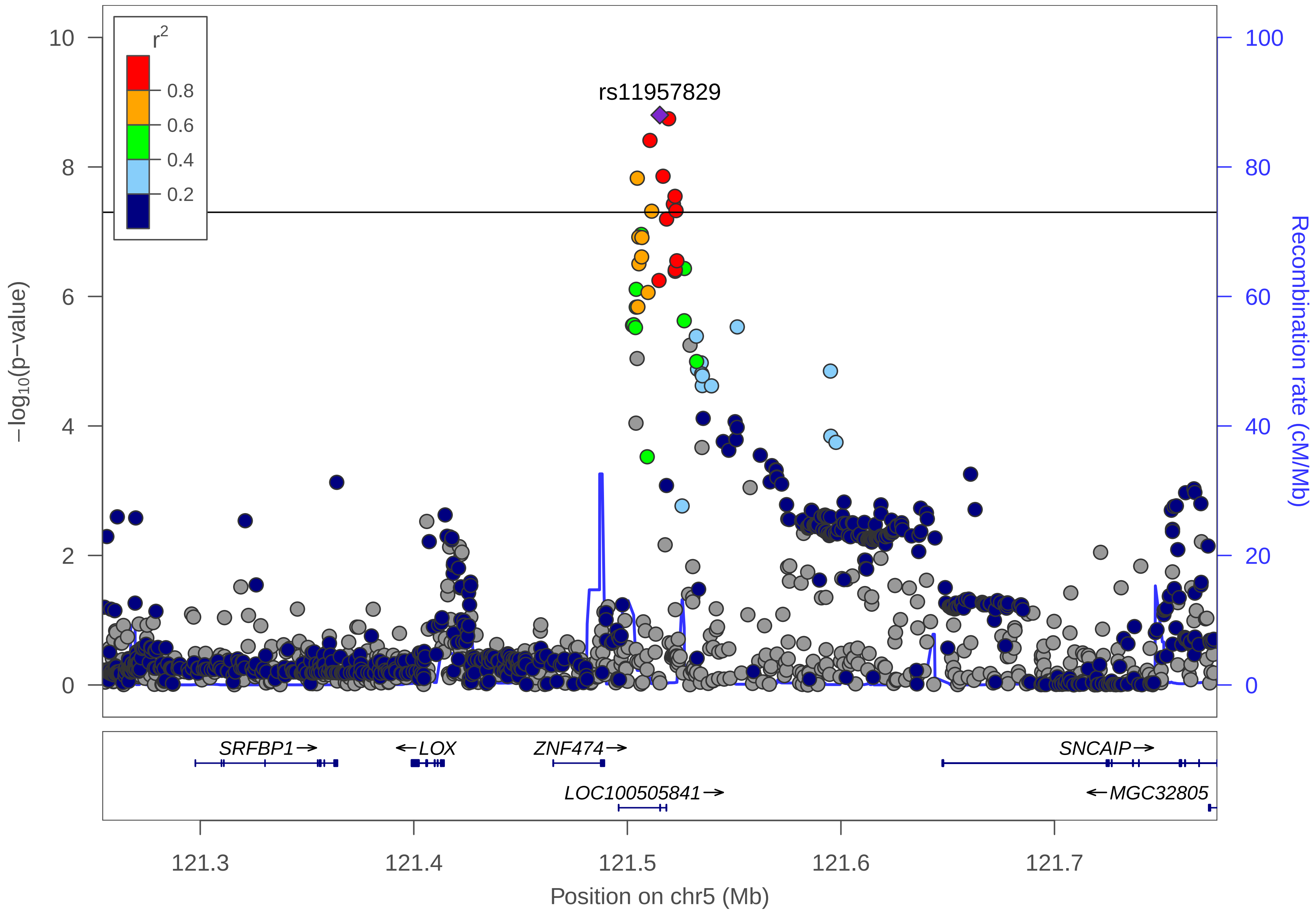

r2
0.8
0.6
0.4
0.2
rs11957829
−log10(p−value)
Recombination rate (cM/Mb)
10
8
6
4
2
0
100
80
60
40
20
0
SRFBP1→
←LOX
ZNF474→
SNCAIP→
LOC100505841→
←MGC32805
121.3
121.4
121.5
121.6
121.7
Position on chr5 (Mb)

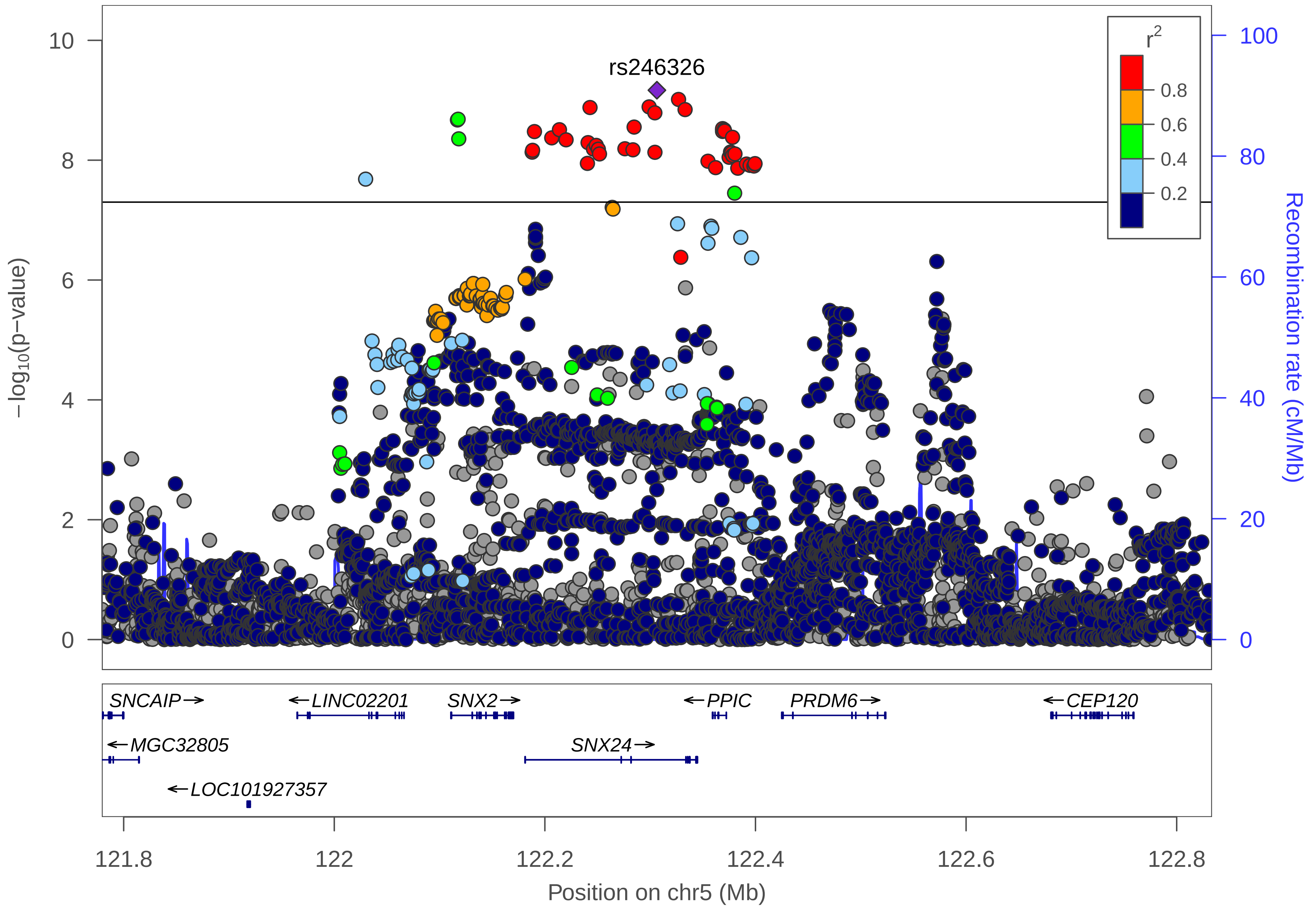

rs246326
r2
0.8
0.6
0.4
0.2
10
8
6
4
2
0
−log10(p−value)
100
80
60
40
20
0
Recombination rate (cM/Mb)
SNCAIP→
←LINC02201
SNX2→
←PPIC
PRDM6→
←CEP120
←MGC32805
SNX24→
←LOC101927357
121.8
122
122.2
122.4
122.6
122.8
Position on chr5 (Mb)

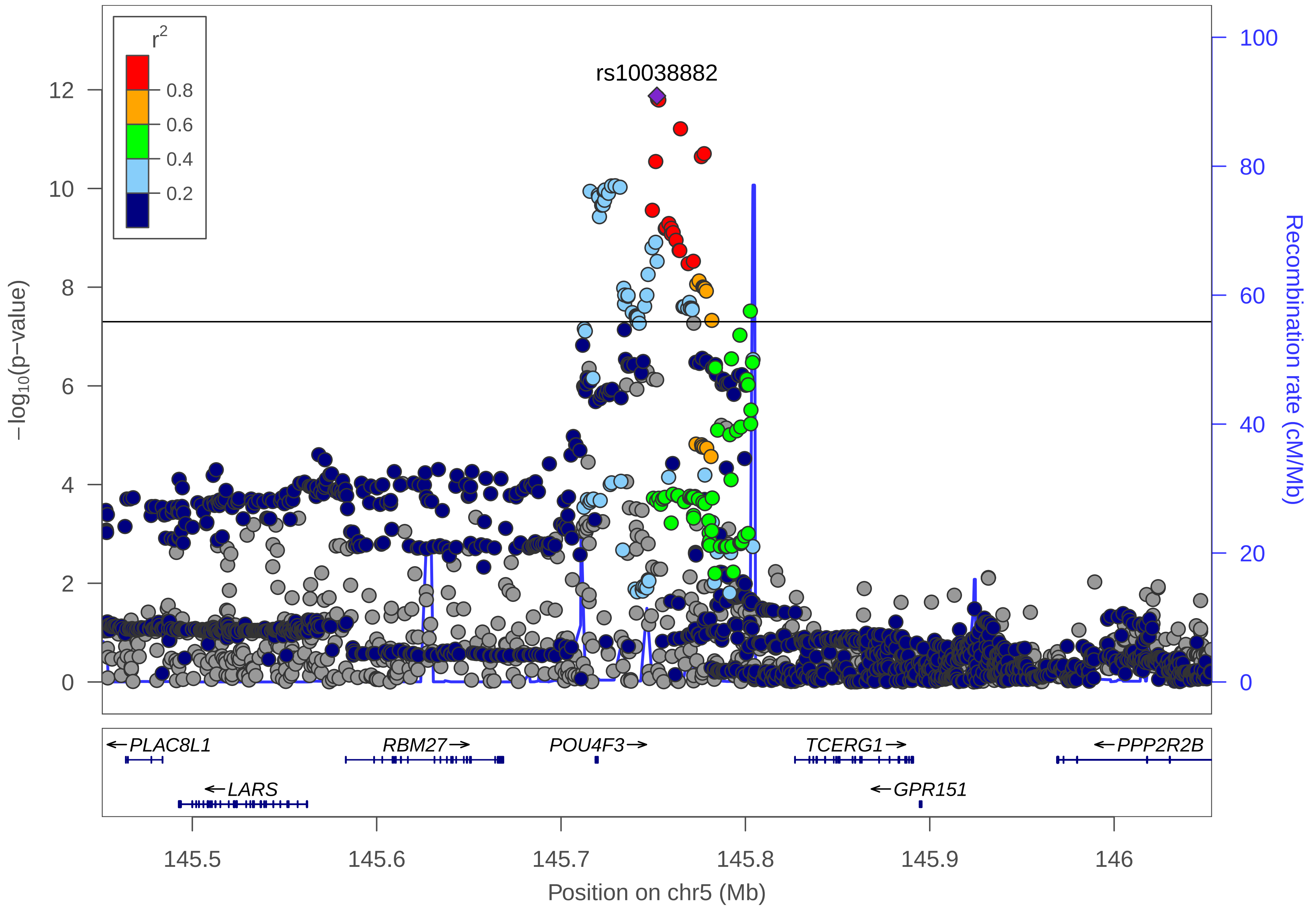

rs10038882
r2
0.8
0.6
0.4
0.2
−log10(p−value)
0
2
4
6
8
10
12
Recombination rate (cM/Mb)
0
20
40
60
80
100
PLAC8L1
LARS
RBM27
POU4F3
TCERG1
GPR151
PPP2R2B
145.5
145.6
145.7
145.8
145.9
146
Position on chr5 (Mb)

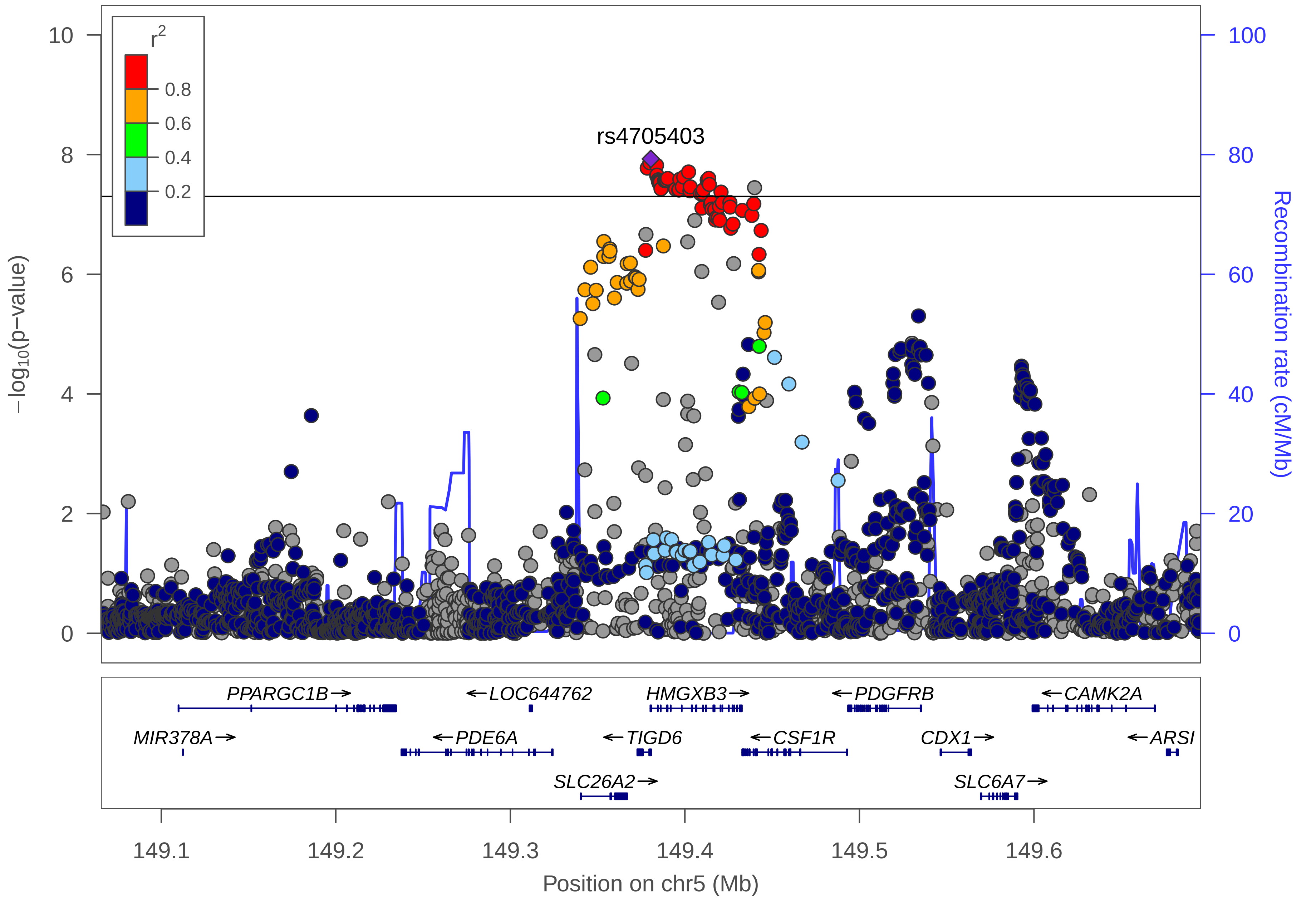
r2
0.8
0.6
0.4
0.2
rs4705403
10
8
6
4
2
0
100
80
60
40
20
0
−log10(p−value)
Recombination rate (cM/Mb)
PPARGC1B→
←LOC644762
HMGXB3→
←PDGFRB
←CAMK2A
MIR378A→
←PDE6A
←TIGD6
←CSF1R
CDX1→
←ARSI
SLC26A2→
SLC6A7→
149.1
149.2
149.3
149.4
149.5
149.6
Position on chr5 (Mb)

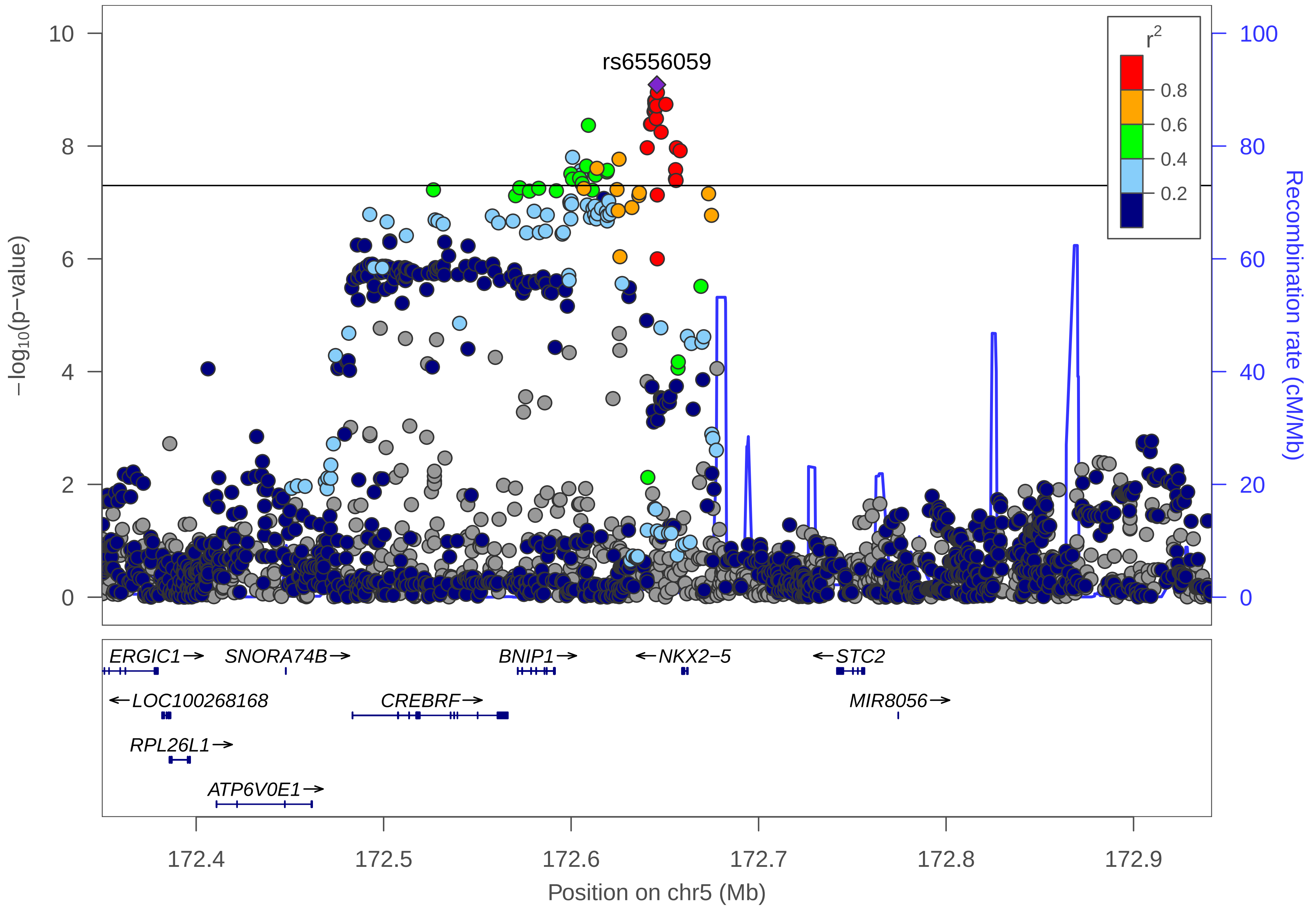

rs6556059
r2
0.8
0.6
0.4
0.2
10
8
6
4
2
0
100
80
60
40
20
0
−log10(p−value)
Recombination rate (cM/Mb)
ERGIC1
SNORA74B
BNIP1
NKX2−5
STC2
LOC100268168
CREBRF
MIR8056
RPL26L1
ATP6V0E1
172.4
172.5
172.6
172.7
172.8
172.9
Position on chr5 (Mb)

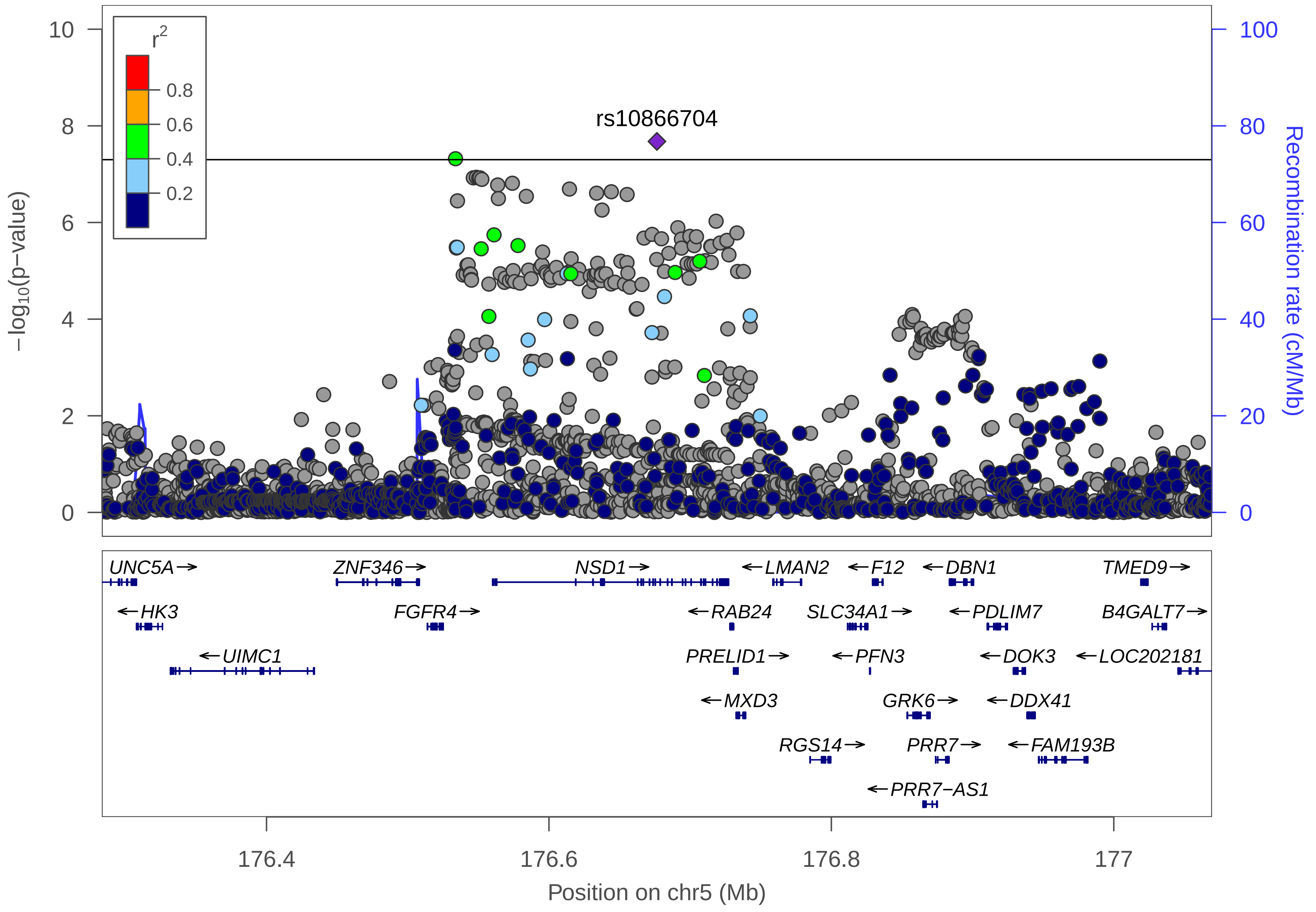

rs10866704
r2
0.8
0.6
0.4
0.2
−log10(p−value)
Recombination rate (cM/Mb)
UNC5A→
ZNF346→
NSD1→
←LMAN2
←F12
←DBN1
TMED9→
←HK3
FGFR4→
←RAB24
SLC34A1→
←PDLIM7
B4GALT7→
←UIMC1
PRELID1→
←PFN3
←DOK3
←LOC202181
←MXD3
GRK6→
←DDX41
RGS14→
PRR7→
←FAM193B
←PRR7−AS1
176.4
176.6
176.8
177
Position on chr5 (Mb)

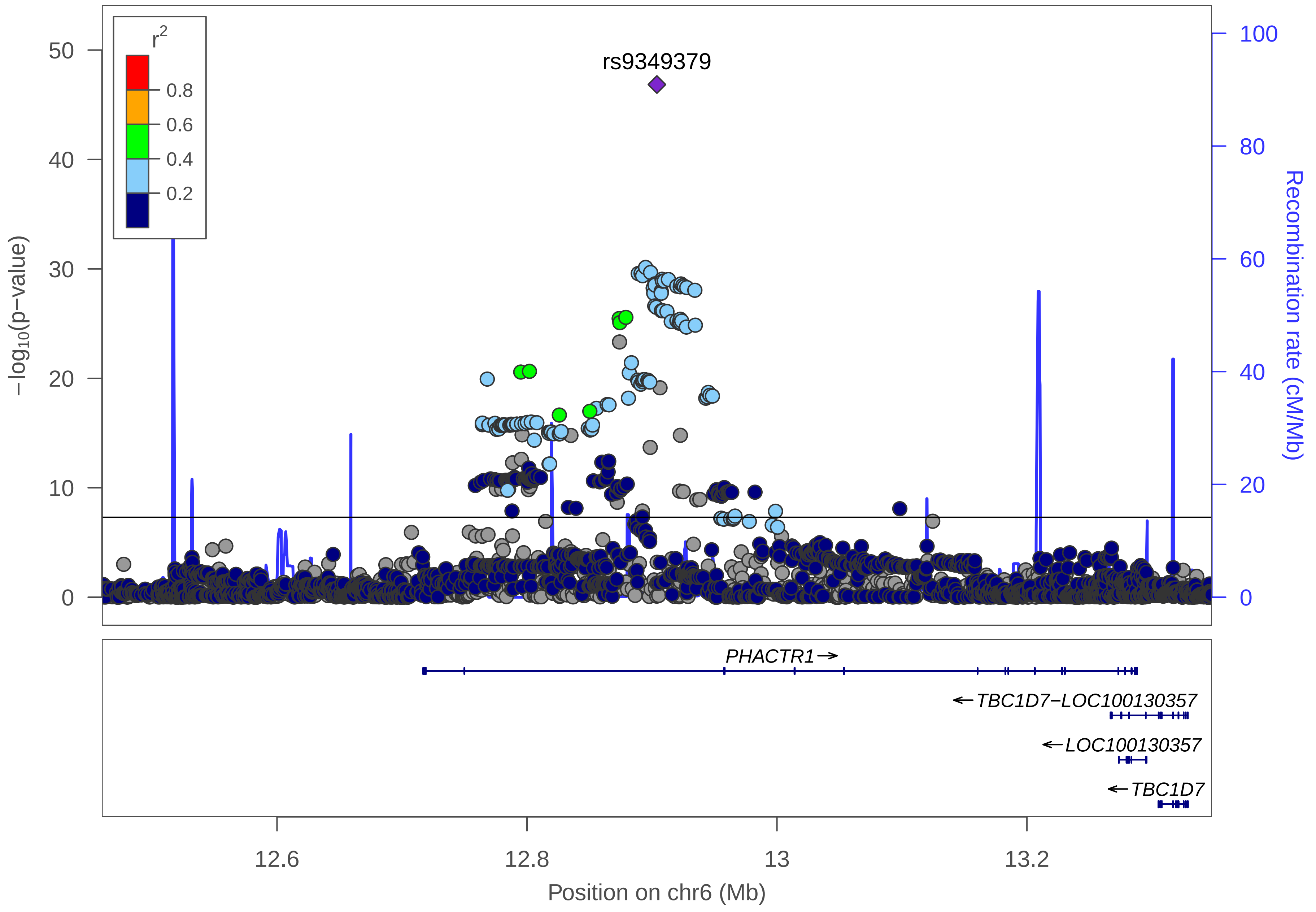

rs9349379
r2
0.8
0.6
0.4
0.2
50
40
30
20
10
0
−log10(p−value)
100
80
60
40
20
0
Recombination rate (cM/Mb)
PHACTR1→
←TBC1D7−LOC100130357
←LOC100130357
←TBC1D7
12.6
12.8
13
13.2
Position on chr6 (Mb)

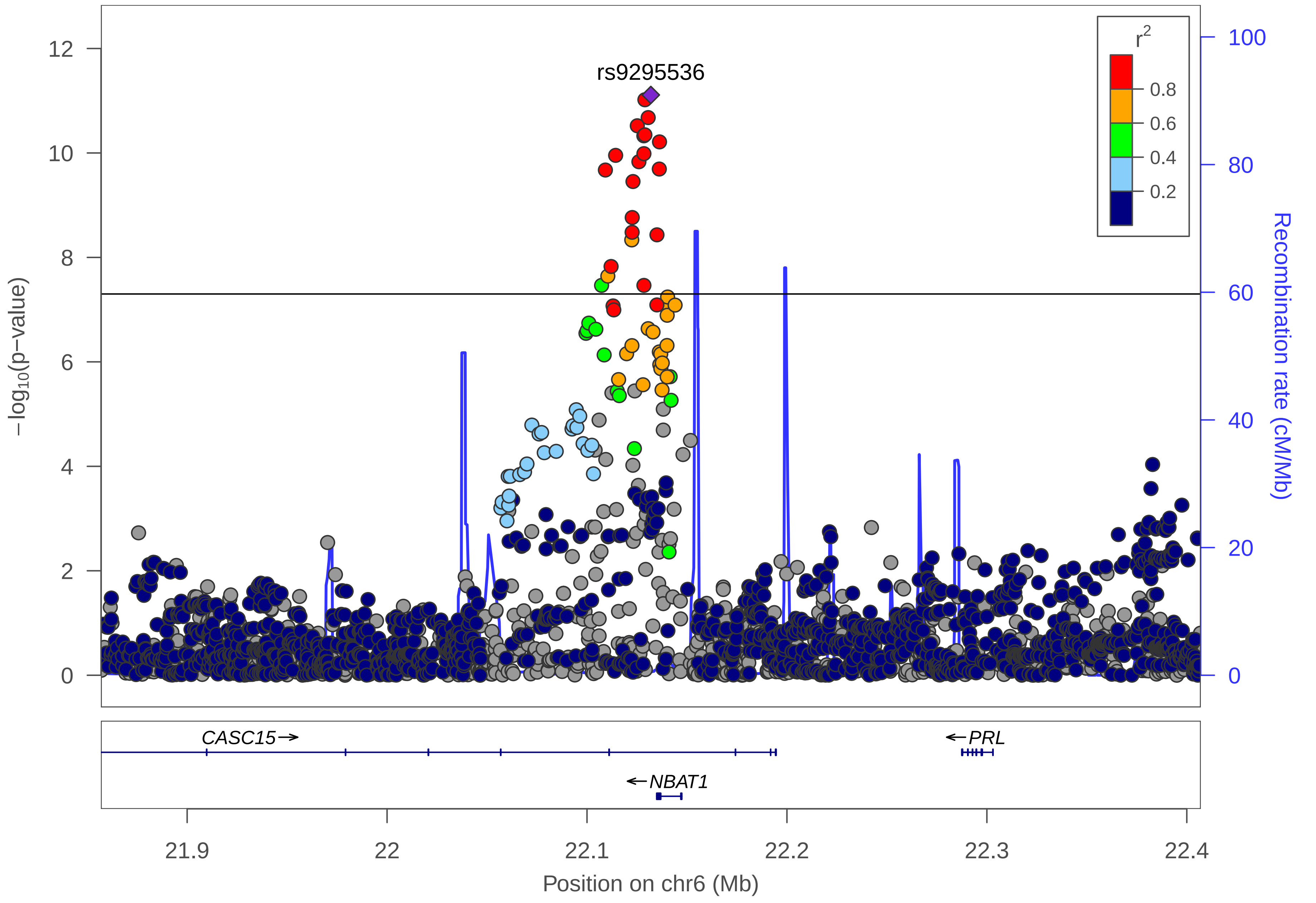
rs9295536
r2
0.8
0.6
0.4
0.2
−log10(p−value)
Recombination rate (cM/Mb)
0
2
4
6
8
10
12
20
40
60
80
100
CASC15→
←PRL
←NBAT1
21.9
22
22.1
22.2
22.3
22.4
Position on chr6 (Mb)

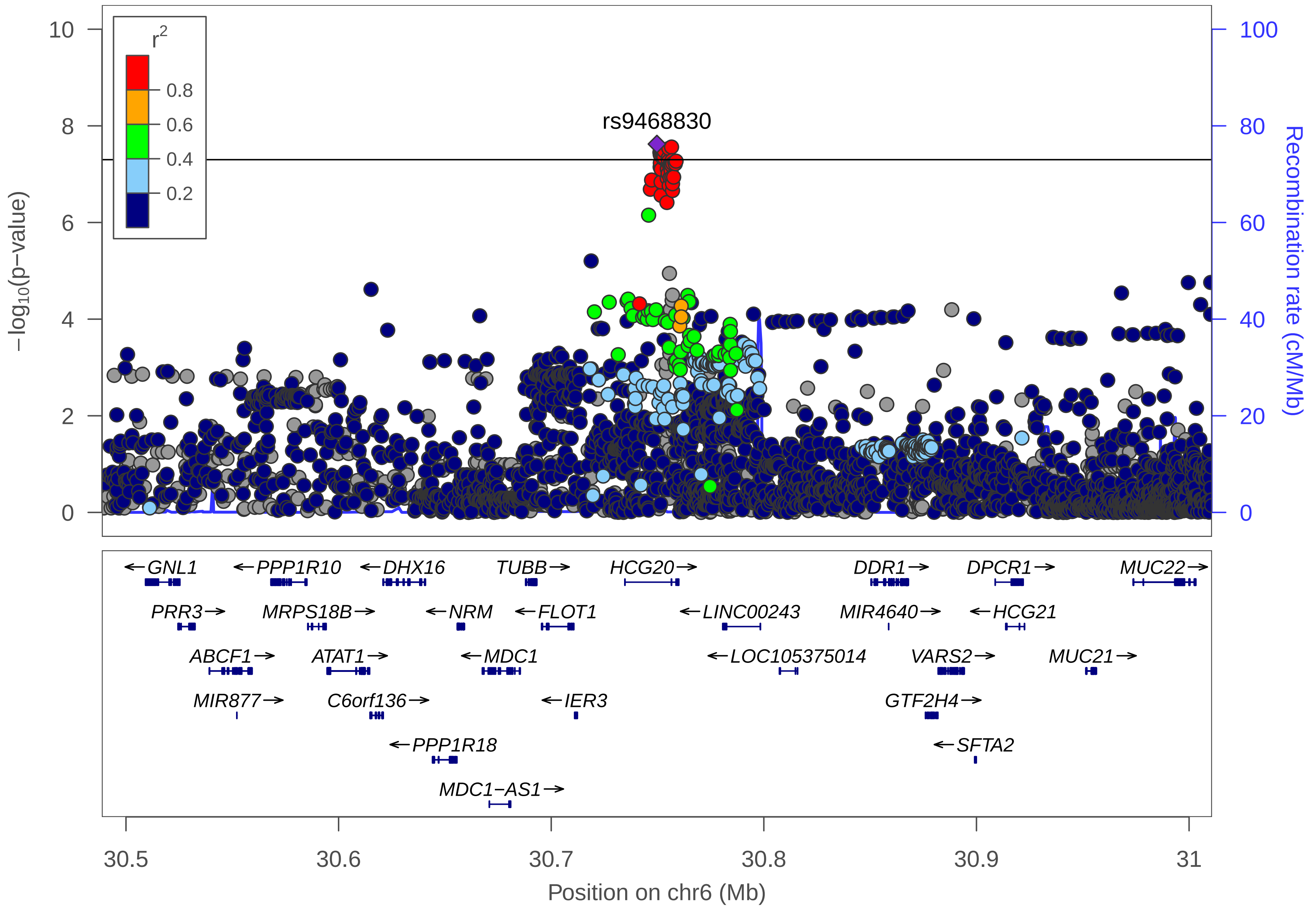

rs9468830
r2
0.8
0.6
0.4
0.2
−log10(p−value)
Recombination rate (cM/Mb)
GNL1
PPP1R10
DHX16
TUBB
HCG20
DDR1
DPCR1
MUC22
PRR3
MRPS18B
NRM
FLOT1
LINC00243
MIR4640
HCG21
ABCF1
ATAT1
MDC1
LOC105375014
VARS2
MUC21
MIR877
C6orf136
IER3
GTF2H4
PPP1R18
SFTA2
MDC1−AS1
30.5
30.6
30.7
30.8
30.9
31
Position on chr6 (Mb)

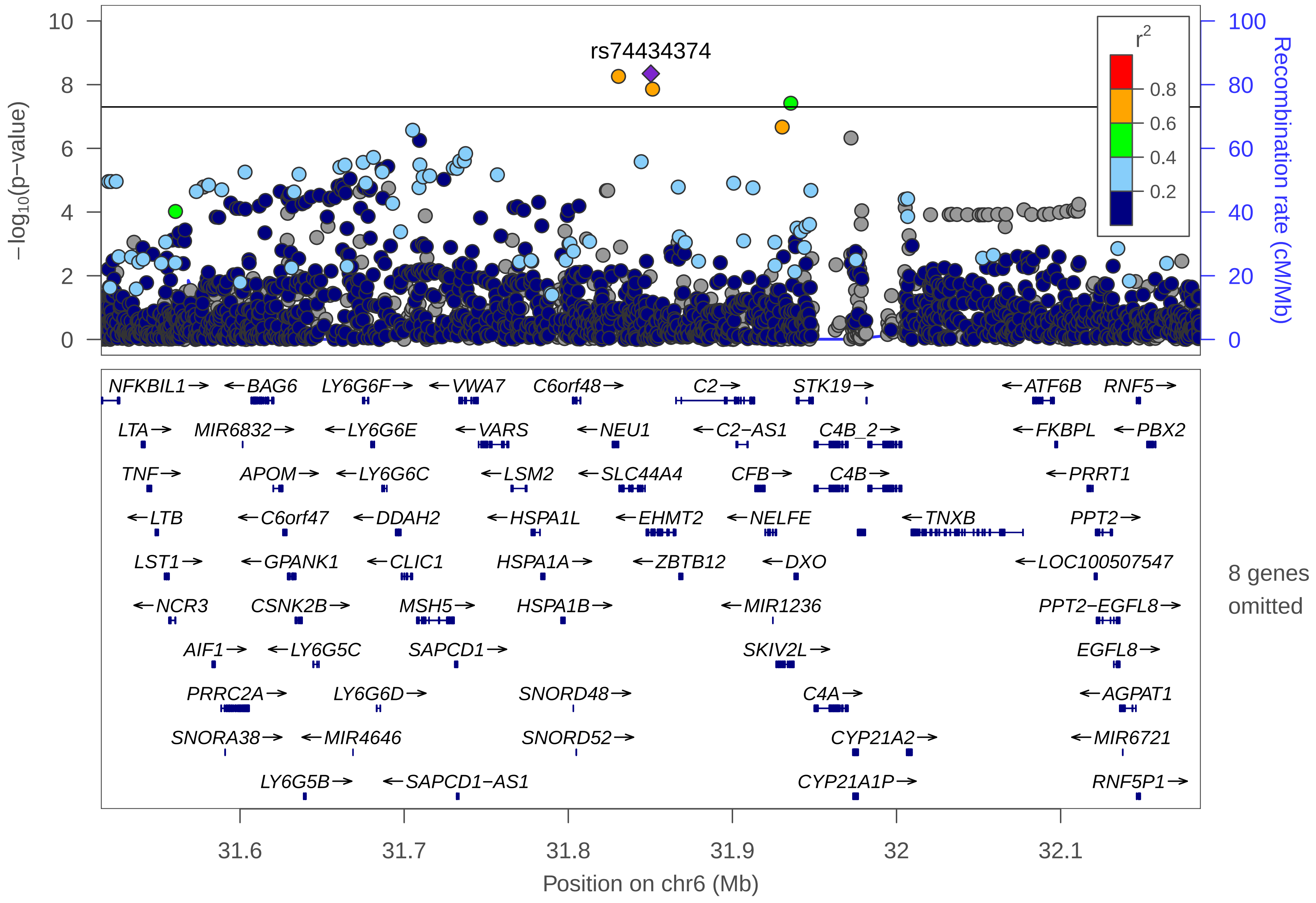
rs74434374
$-\log_{10}$(p−value)
Recombination rate (cM/Mb)
$r^2$
0.8
0.6
0.4
0.2
10
8
6
4
2
0
100
80
60
40
20
0
NFKBIL1→
←BAG6
LY6G6F→
←VWA7
C6orf48→
C2→
STK19→
←ATF6B
RNF5→
LTA→
MIR6832→
←LY6G6E
←VARS
←NEU1
←C2−AS1
C4B_2→
←FKBPL
←PBX2
TNF→
APOM→
←LY6G6C
←LSM2
←SLC44A4
CFB→
C4B→
←PRRT1
←LTB
←C6orf47
←DDAH2
←HSPA1L
←EHMT2
←NELFE
←TNXB
PPT2→
LST1→
←GPANK1
←CLIC1
HSPA1A→
←ZBTB12
←DXO
←LOC100507547
←NCR3
CSNK2B→
MSH5→
HSPA1B→
←MIR1236
PPT2−EGFL8→
AIF1→
←LY6G5C
SAPCD1→
SKIV2L→
EGFL8→
PRRC2A→
LY6G6D→
SNORD48→
C4A→
←AGPAT1
SNORA38→
←MIR4646
SNORD52→
CYP21A2→
←MIR6721
LY6G5B→
←SAPCD1−AS1
CYP21A1P→
RNF5P1→
8 genes omitted
31.6
31.7
31.8
31.9
32
32.1
Position on chr6 (Mb)

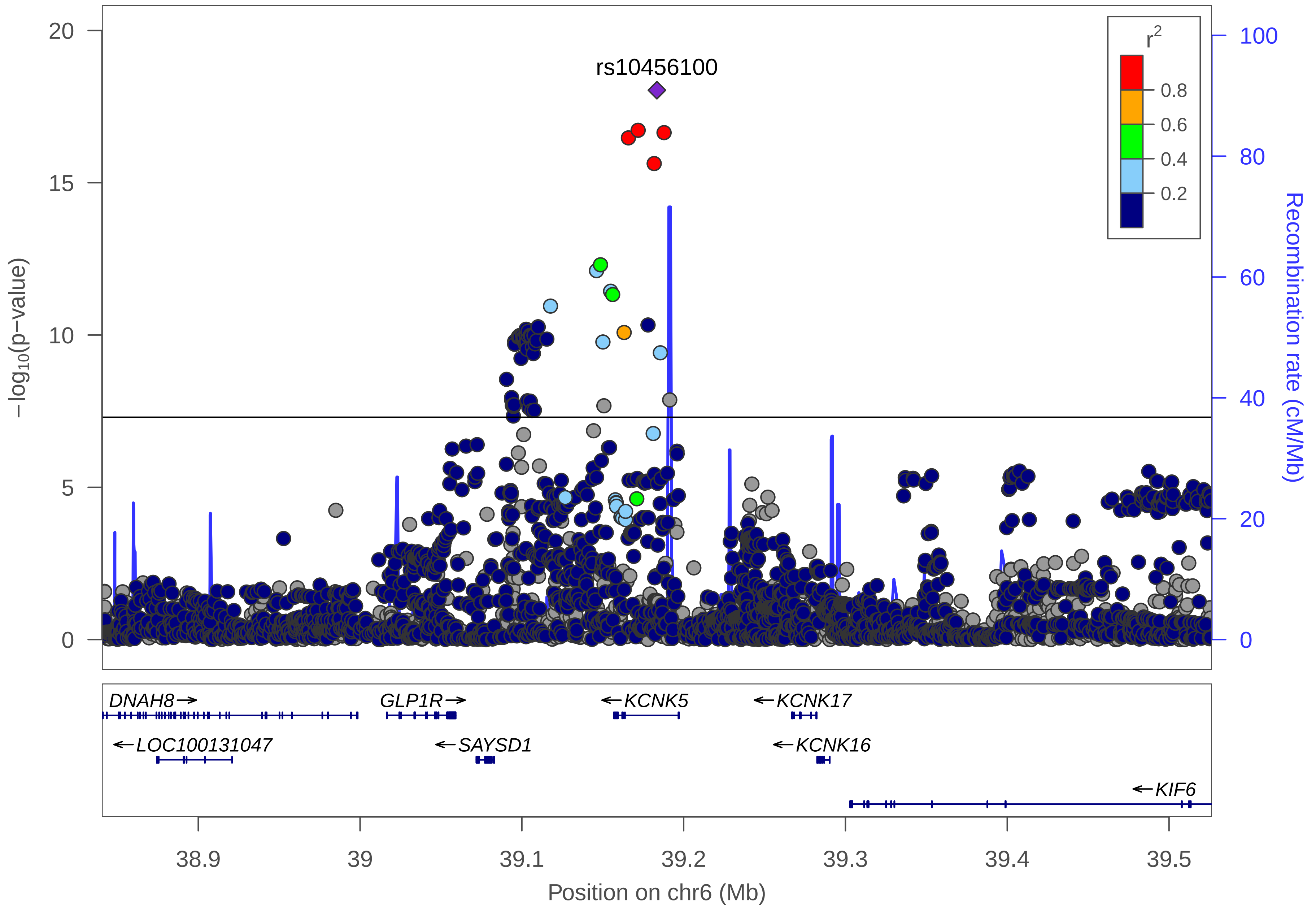

rs10456100
r2
0.8
0.6
0.4
0.2
−log10(p−value)
Recombination rate (cM/Mb)
20
15
10
5
0
100
80
60
40
20
0
DNAH8→
GLP1R→
←KCNK5
←KCNK17
←LOC100131047
←SAYSD1
←KCNK16
←KIF6
38.9
39
39.1
39.2
39.3
39.4
39.5
Position on chr6 (Mb)

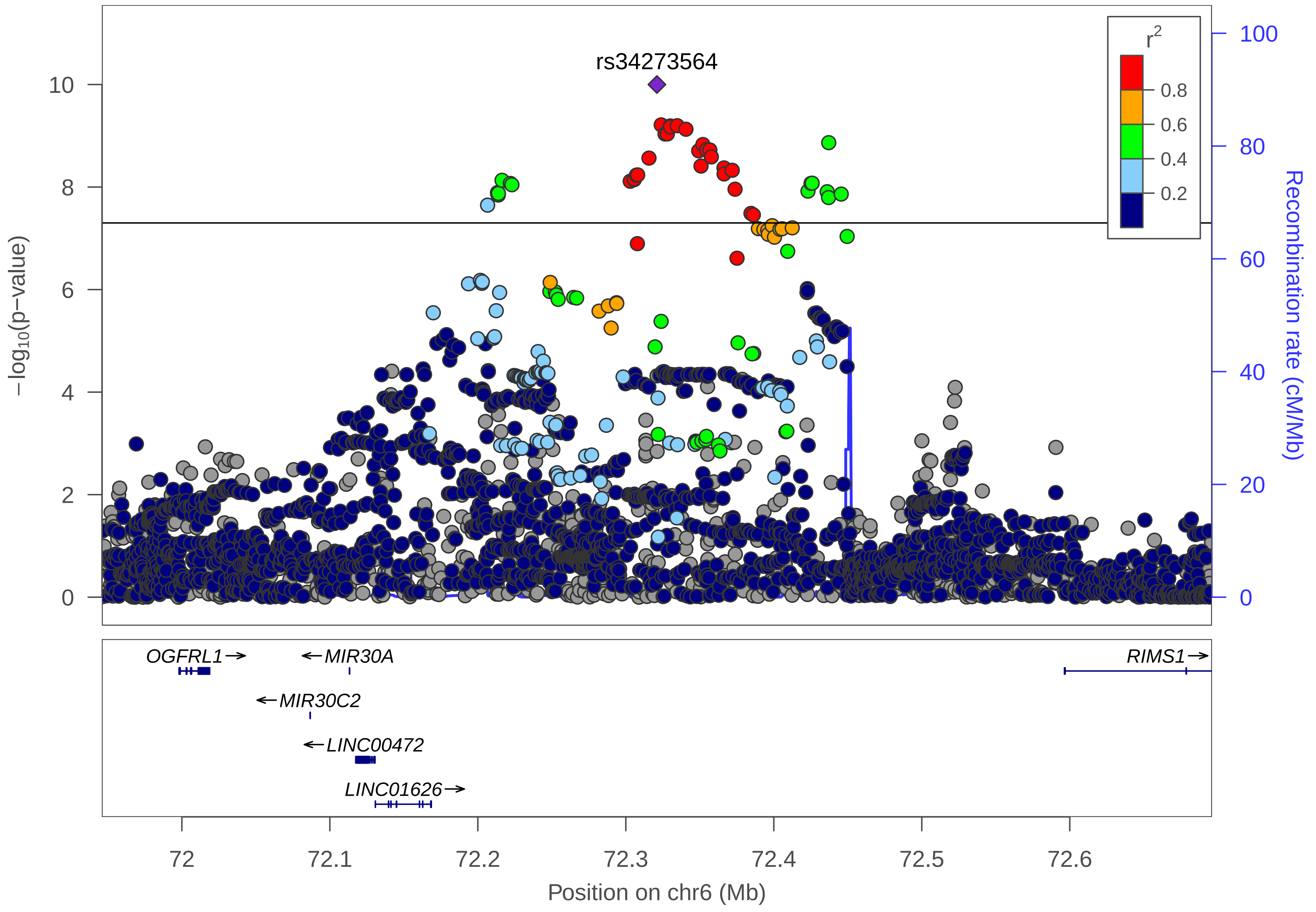

rs34273564
r2
0.8
0.6
0.4
0.2
10
8
6
4
2
0
−log10(p−value)
100
80
60
40
20
0
Recombination rate (cM/Mb)
OGFRL1→
←MIR30A
←MIR30C2
←LINC00472
LINC01626→
RIMS1→
72
72.1
72.2
72.3
72.4
72.5
72.6
Position on chr6 (Mb)

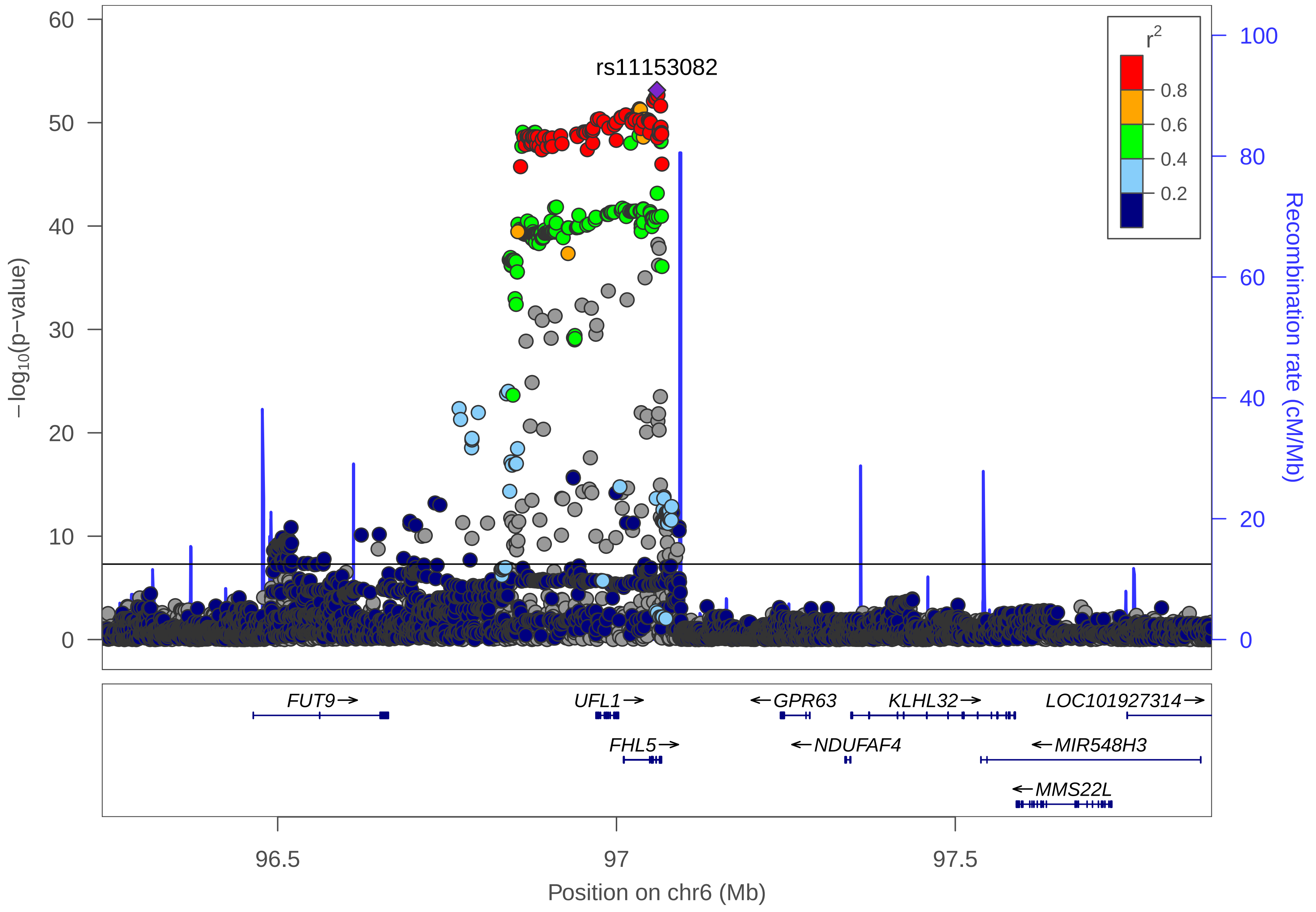

rs11153082
r2
0.8
0.6
0.4
0.2
60
50
40
30
20
10
0
100
80
60
40
20
0
−log10(p−value)
Recombination rate (cM/Mb)
FUT9→
UFL1→
←GPR63
KLHL32→
LOC101927314→
FHL5→
←NDUFAF4
←MIR548H3
←MMS22L
96.5
97
97.5
Position on chr6 (Mb)

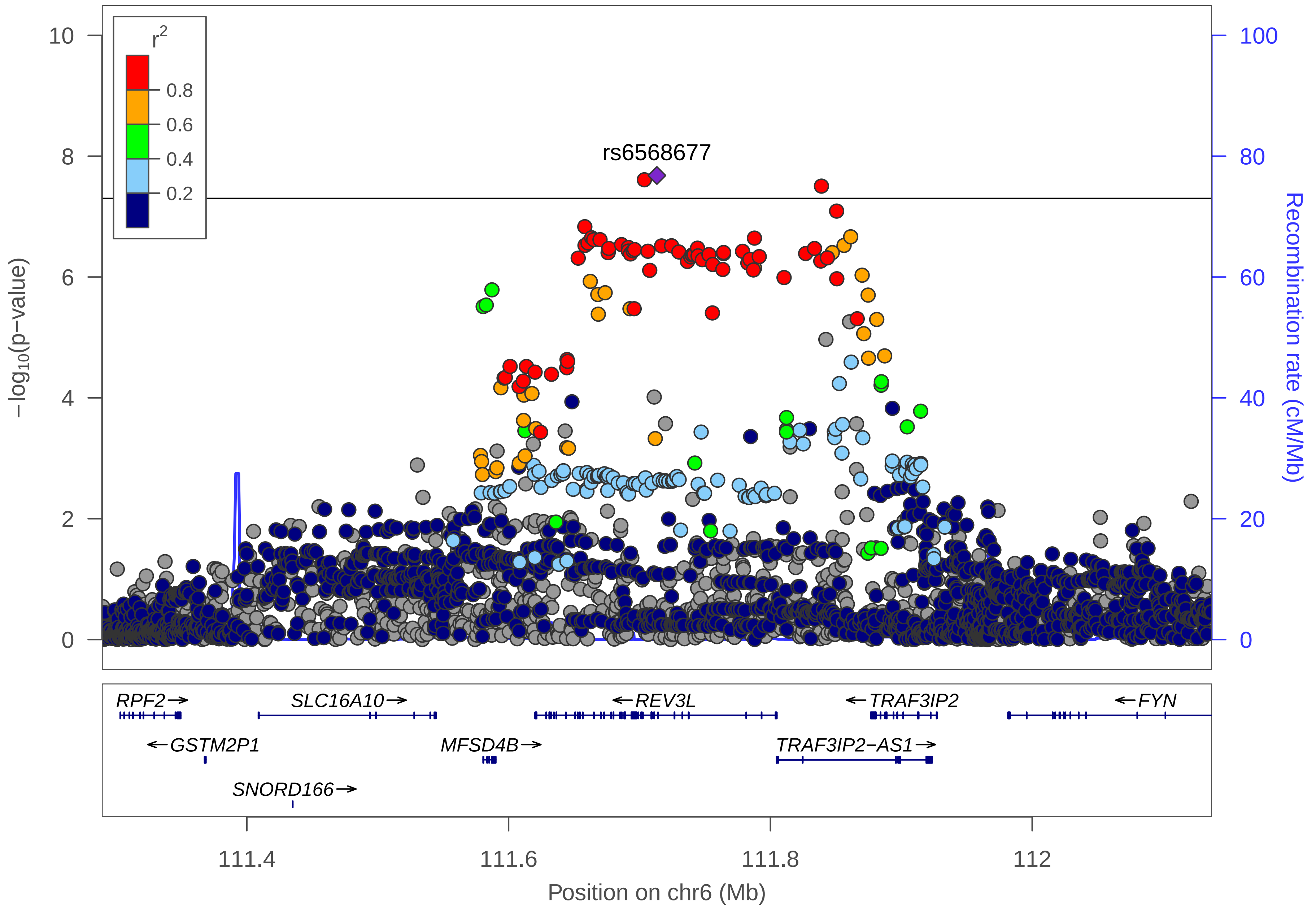
r2
0.8
0.6
0.4
0.2
rs6568677
−log10(p−value)
Recombination rate (cM/Mb)
10
8
6
4
2
0
100
80
60
40
20
0
RPF2→
SLC16A10→
←REV3L
←TRAF3IP2
←FYN
←GSTM2P1
MFSD4B→
TRAF3IP2−AS1→
SNORD166→
111.4
111.6
111.8
112
Position on chr6 (Mb)

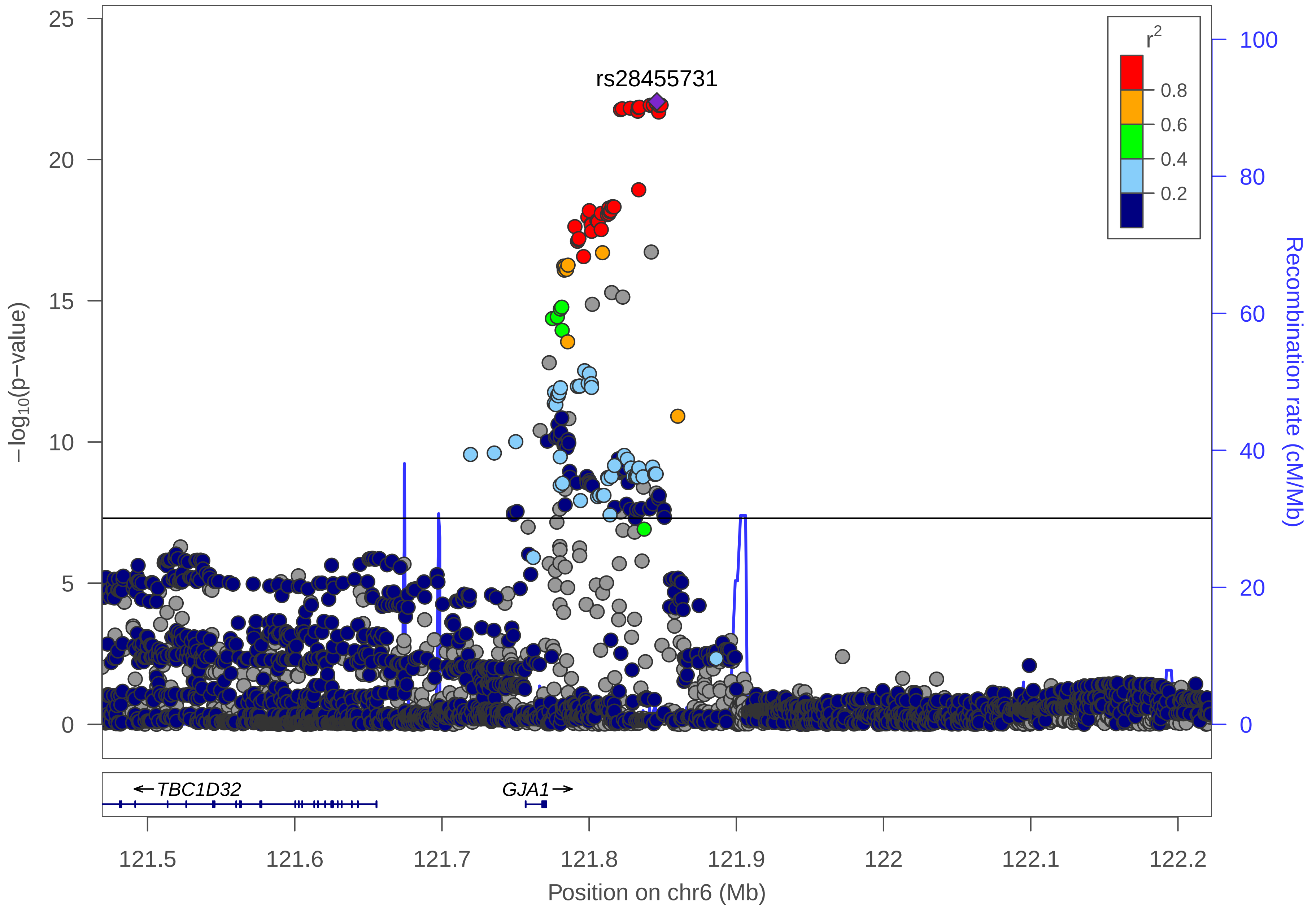

rs28455731
r2
0.8
0.6
0.4
0.2
−log10(p−value)
0
5
10
15
20
25
Recombination rate (cM/Mb)
0
20
40
60
80
100
TBC1D32
GJA1
121.5
121.6
121.7
121.8
121.9
122
122.1
122.2
Position on chr6 (Mb)

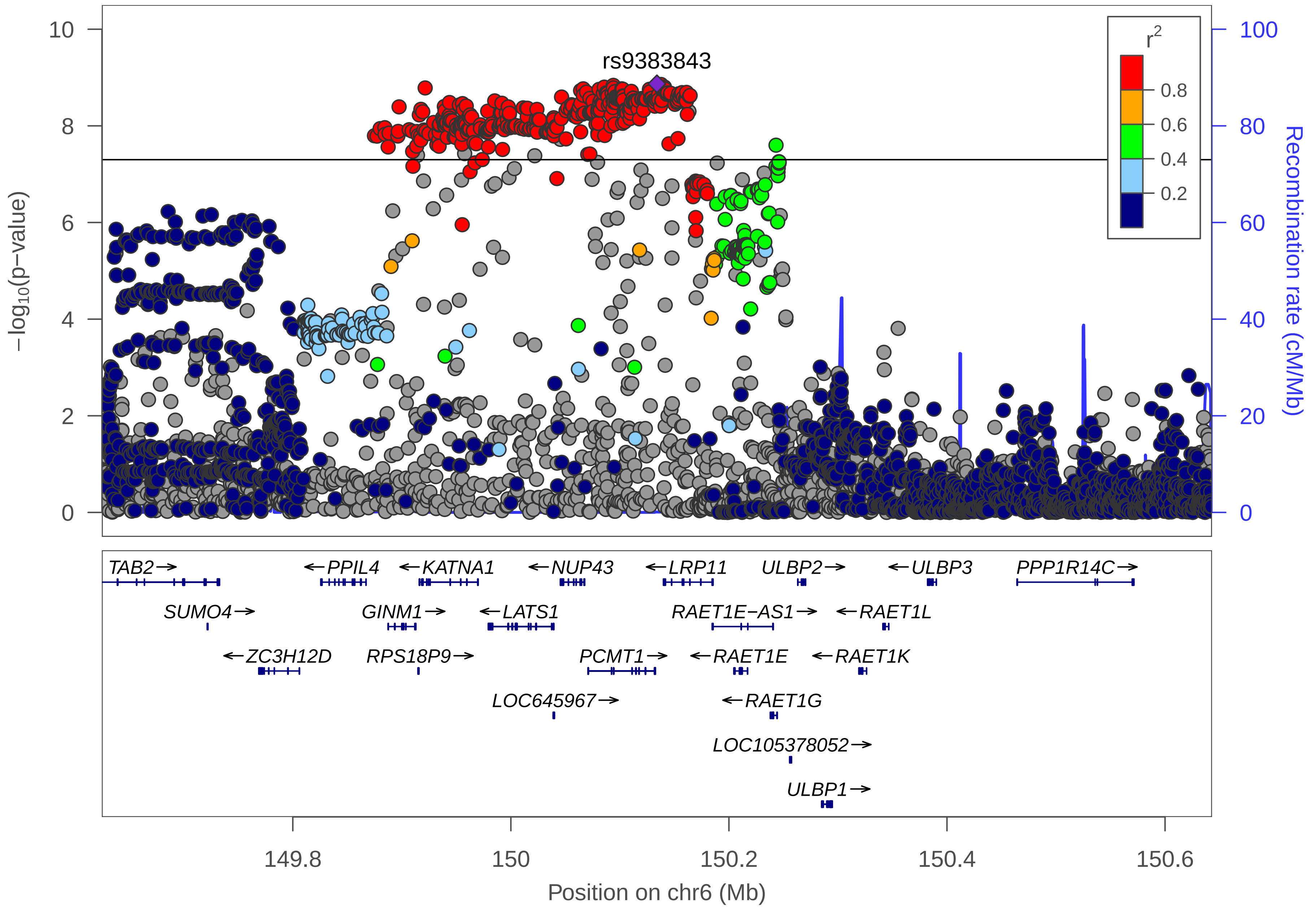
rs9383843
r2
0.8
0.6
0.4
0.2
−log10(p−value)
Recombination rate (cM/Mb)
10
8
6
4
2
0
100
80
60
40
20
0
TAB2→
←PPIL4
←KATNA1
←NUP43
←LRP11
ULBP2→
←ULBP3
PPP1R14C→
SUMO4→
GINM1→
←LATS1
RAET1E−AS1→
←RAET1L
←ZC3H12D
RPS18P9→
PCMT1→
←RAET1E
←RAET1K
LOC645967→
←RAET1G
LOC105378052→
ULBP1→
149.8
150
150.2
150.4
150.6
Position on chr6 (Mb)

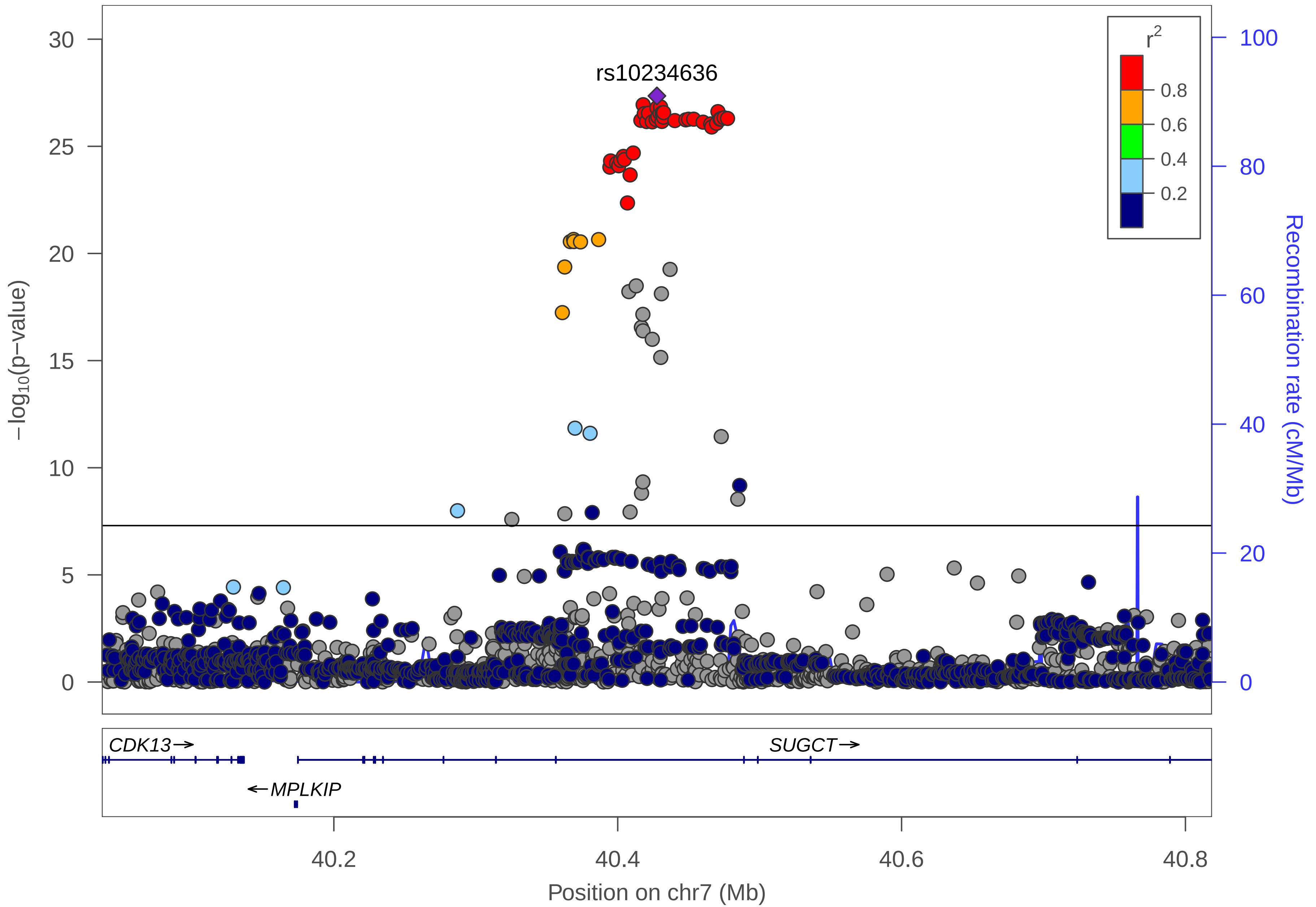

rs10234636
r^2
0.8
0.6
0.4
0.2
−log10(p−value)
Recombination rate (cM/Mb)
0
5
10
15
20
25
30
0
20
40
60
80
100
CDK13→
SUGCT→
←MPLKIP
40.2
40.4
40.6
40.8
Position on chr7 (Mb)

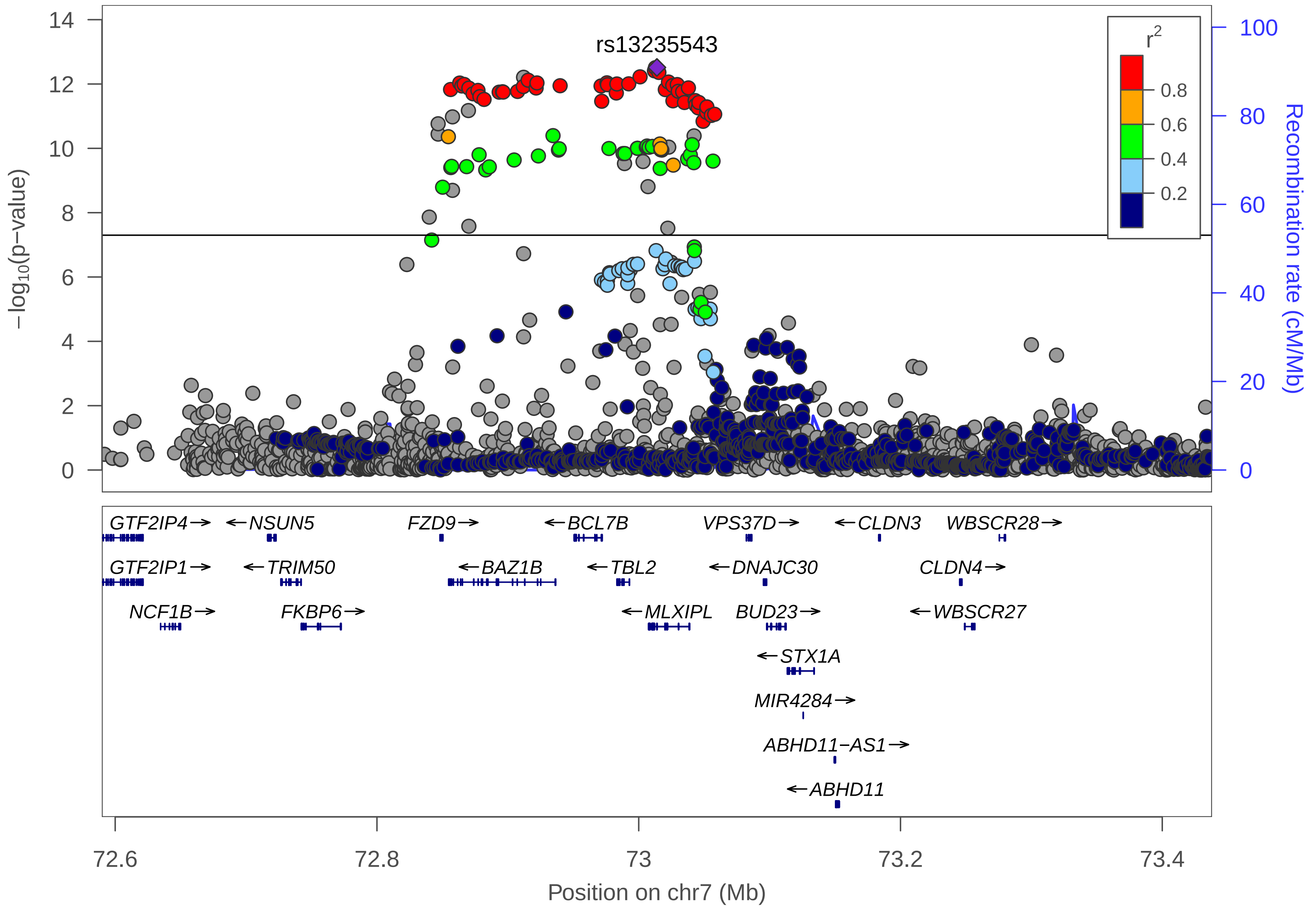
rs13235543
r²
0.8
0.6
0.4
0.2
−log10(p−value)
Recombination rate (cM/Mb)
GTF2IP4→
←NSUN5
FZD9→
←BCL7B
VPS37D→
←CLDN3
WBSCR28→
GTF2IP1→
←TRIM50
←BAZ1B
←TBL2
←DNAJC30
CLDN4→
NCF1B→
FKBP6→
←MLXIPL
BUD23→
←WBSCR27
←STX1A
MIR4284→
ABHD11−AS1→
←ABHD11
72.6
72.8
73
73.2
73.4
Position on chr7 (Mb)

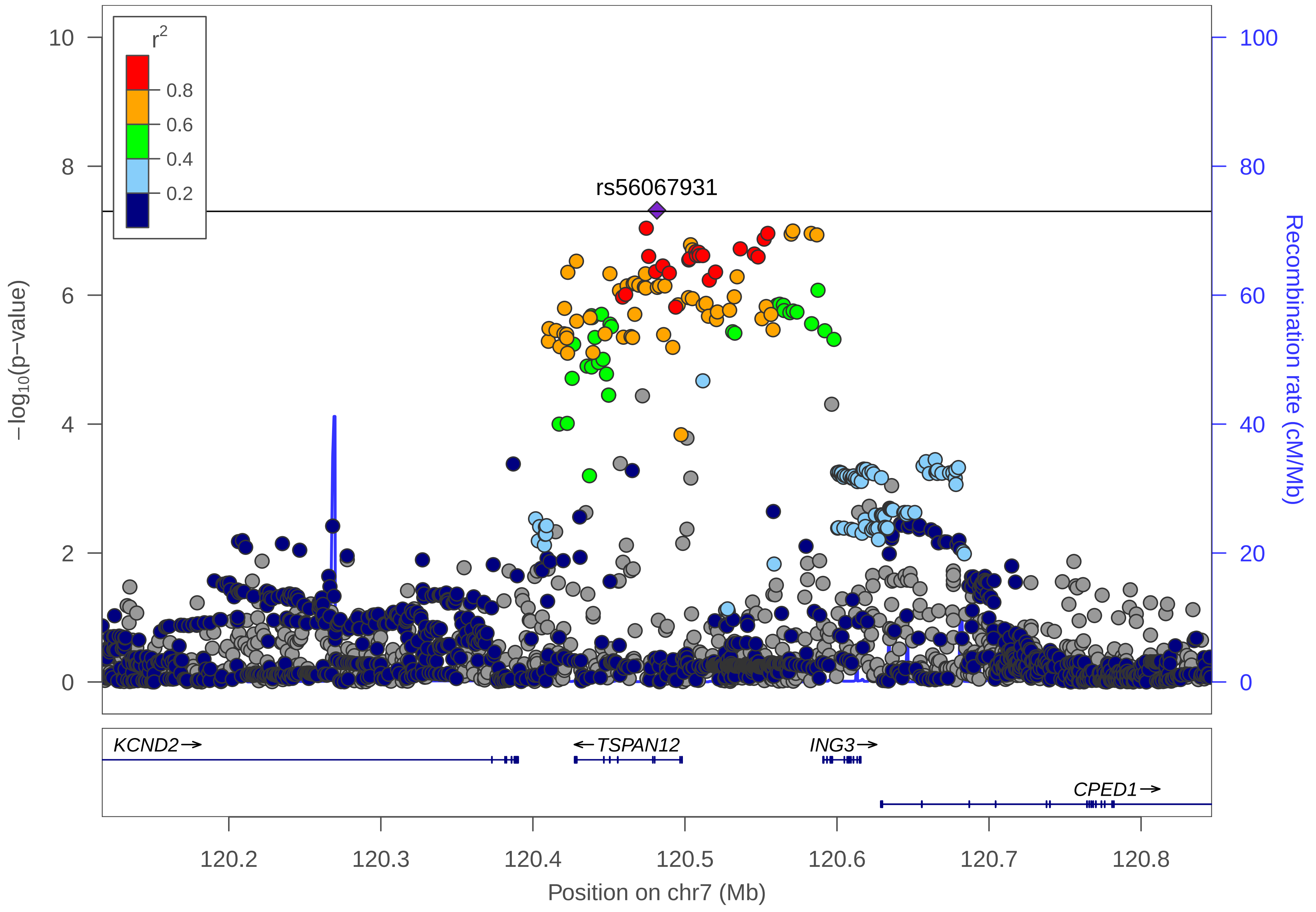

r2
0.8
0.6
0.4
0.2
rs56067931
−log10(p−value)
Recombination rate (cM/Mb)
10
8
6
4
2
0
100
80
60
40
20
0
KCND2→
←TSPAN12
ING3→
CPED1→
120.2
120.3
120.4
120.5
120.6
120.7
120.8
Position on chr7 (Mb)

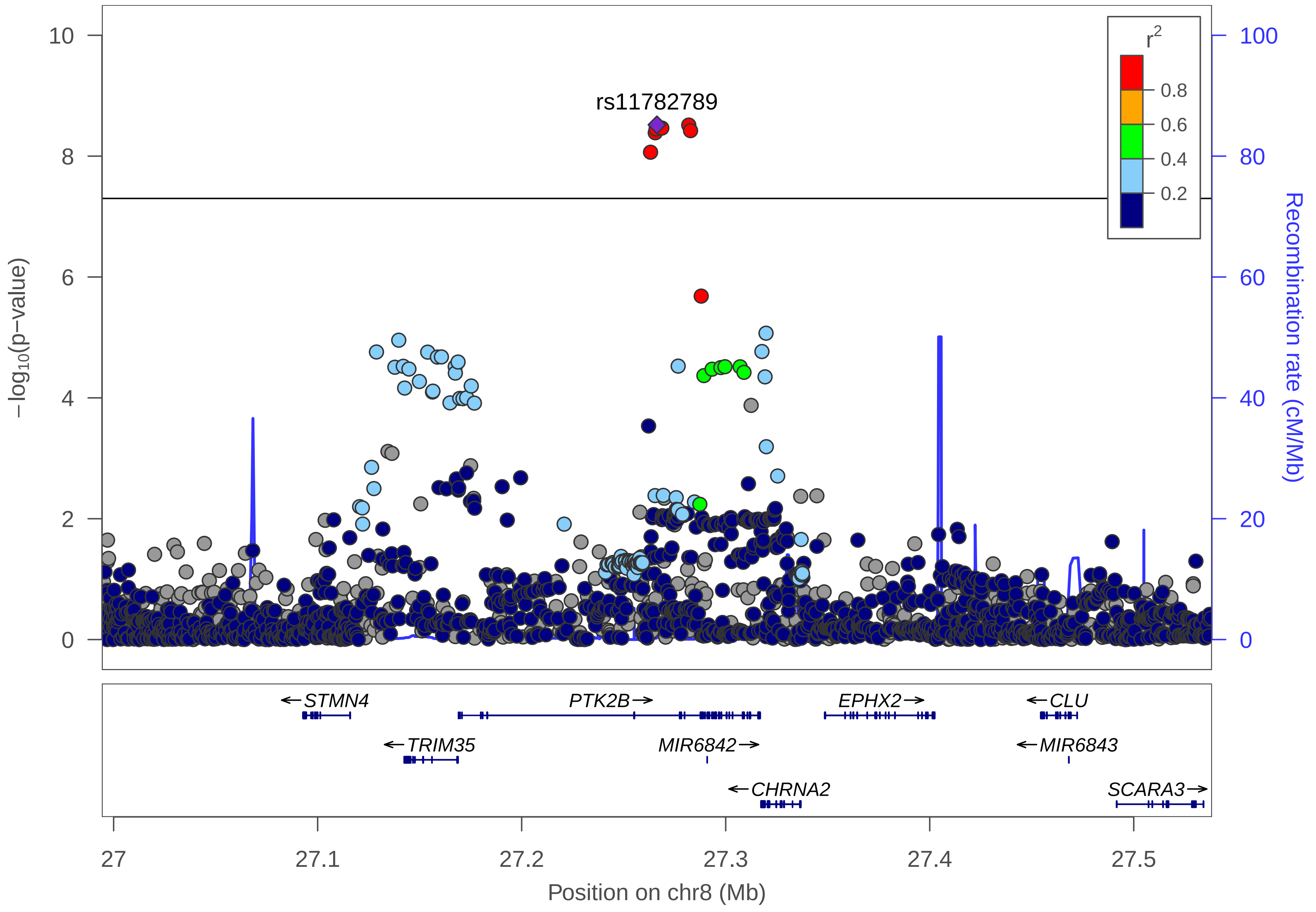

rs11782789
−log10(p−value)
Recombination rate (cM/Mb)
r2
0.8
0.6
0.4
0.2
10
8
6
4
2
0
100
80
60
40
20
0
STMN4
PTK2B
EPHX2
CLU
TRIM35
MIR6842
MIR6843
CHRNA2
SCARA3
27
27.1
27.2
27.3
27.4
27.5
Position on chr8 (Mb)

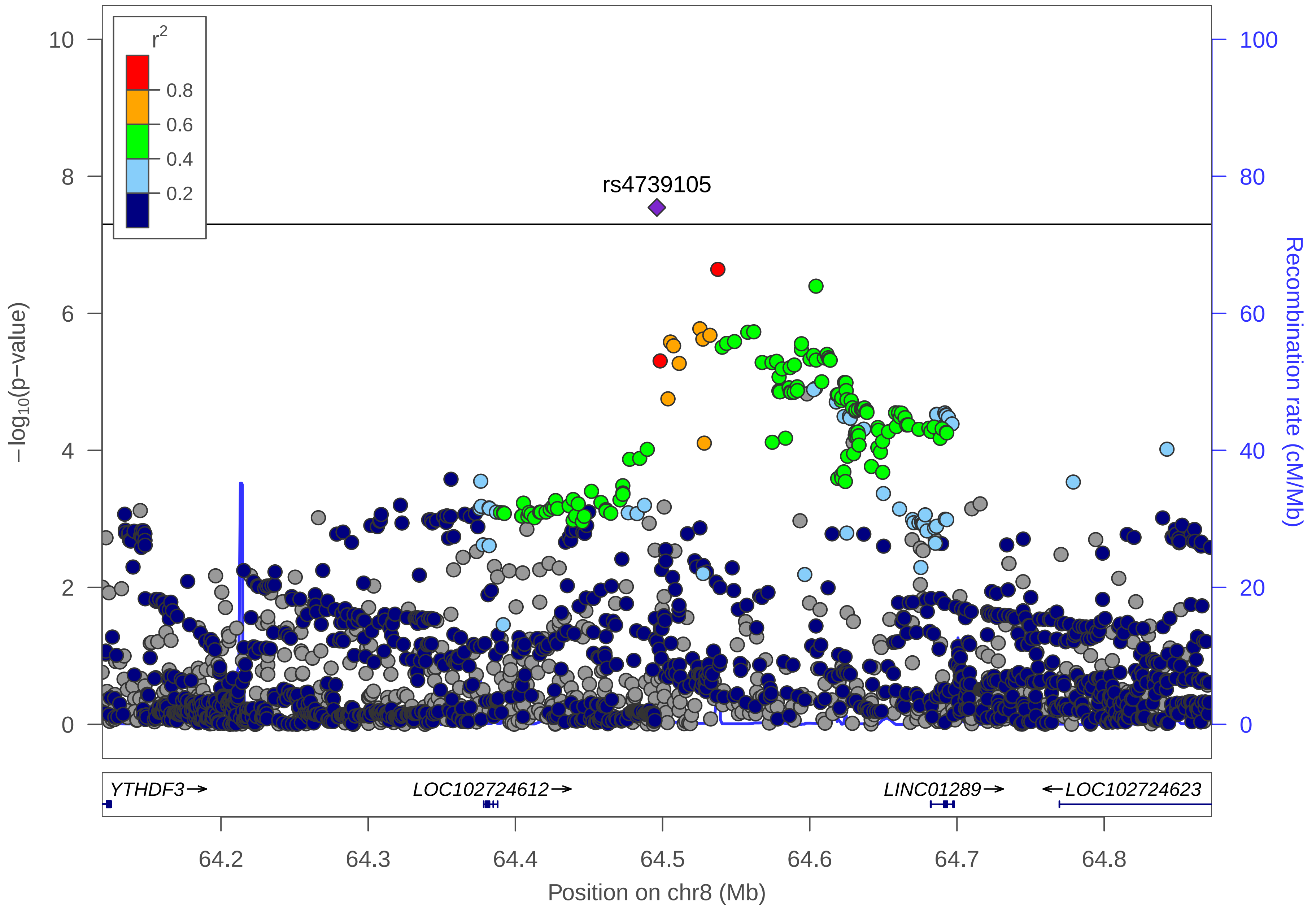

r2
0.8
0.6
0.4
0.2
rs4739105
−log10(p−value)
Recombination rate (cM/Mb)
0
2
4
6
8
10
0
20
40
60
80
100
YTHDF3→
LOC102724612→
LINC01289→
←LOC102724623
64.2
64.3
64.4
64.5
64.6
64.7
64.8
Position on chr8 (Mb)

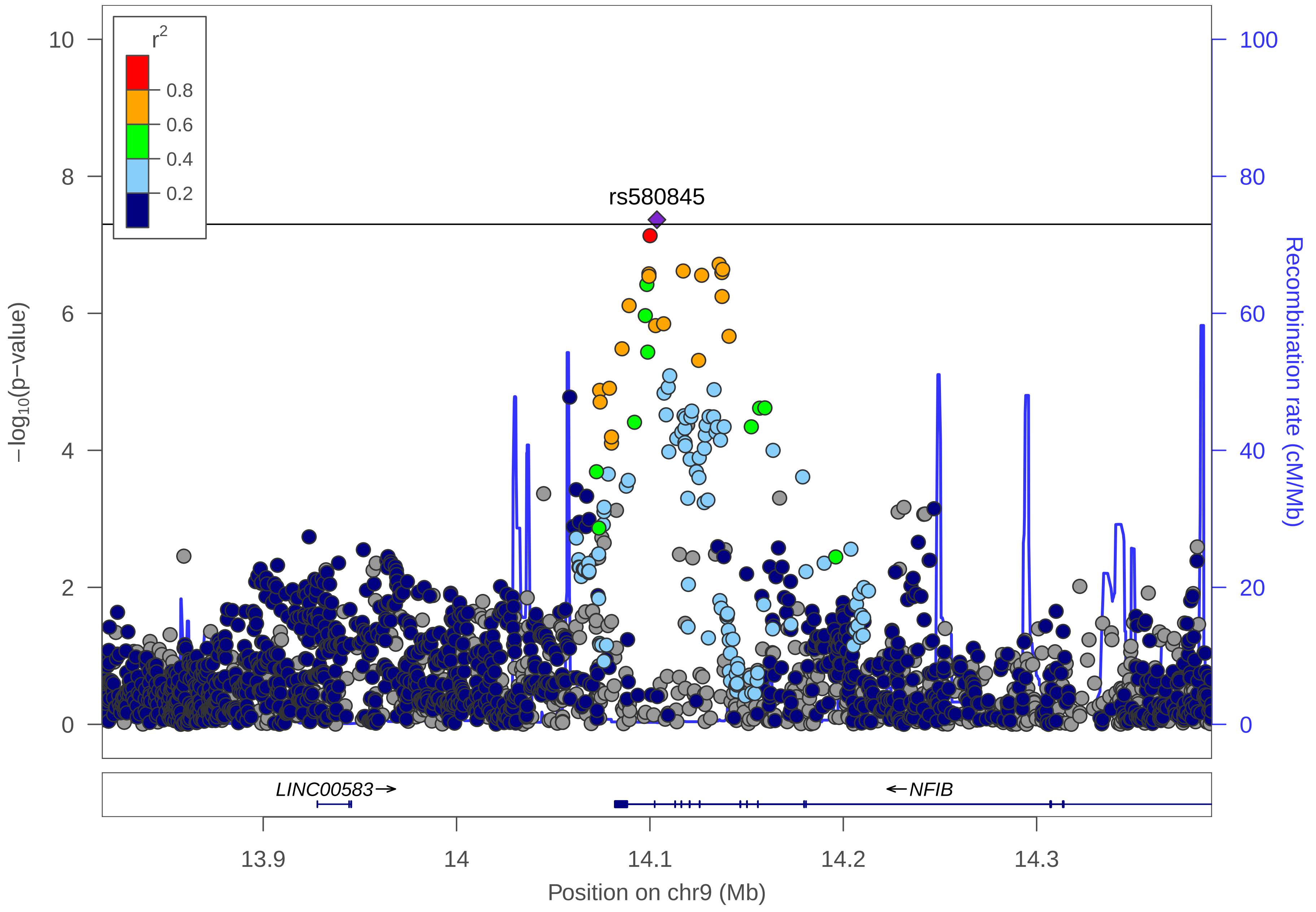

r²
0.8
0.6
0.4
0.2
rs580845
−log10(p−value)
Recombination rate (cM/Mb)
LINC00583→
←NFIB
13.9
14
14.1
14.2
14.3
Position on chr9 (Mb)

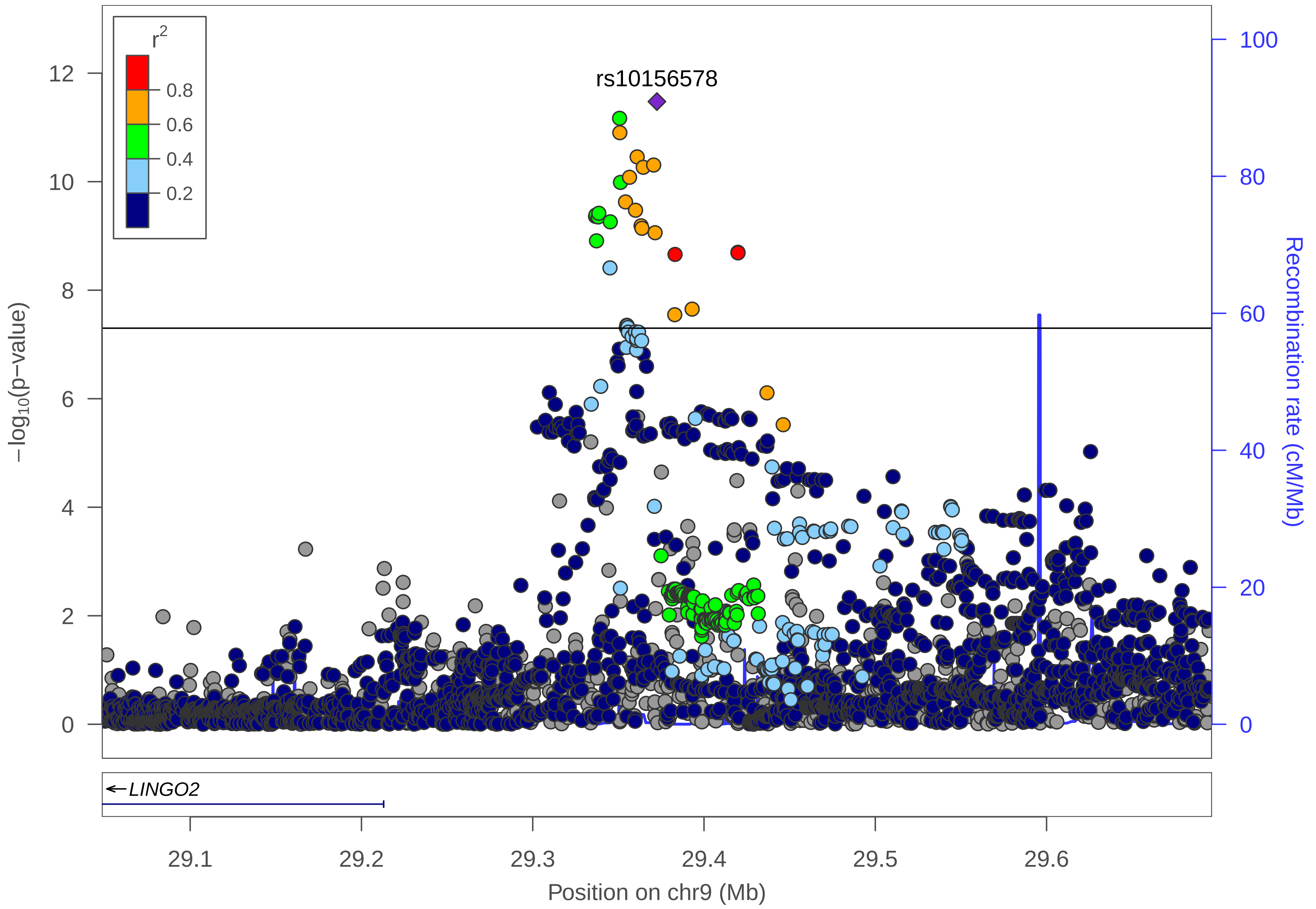
r2
0.8
0.6
0.4
0.2
rs10156578
12
10
8
6
4
2
0
−log10(p−value)
100
80
60
40
20
0
Recombination rate (cM/Mb)
←LINGO2
29.1
29.2
29.3
29.4
29.5
29.6
Position on chr9 (Mb)

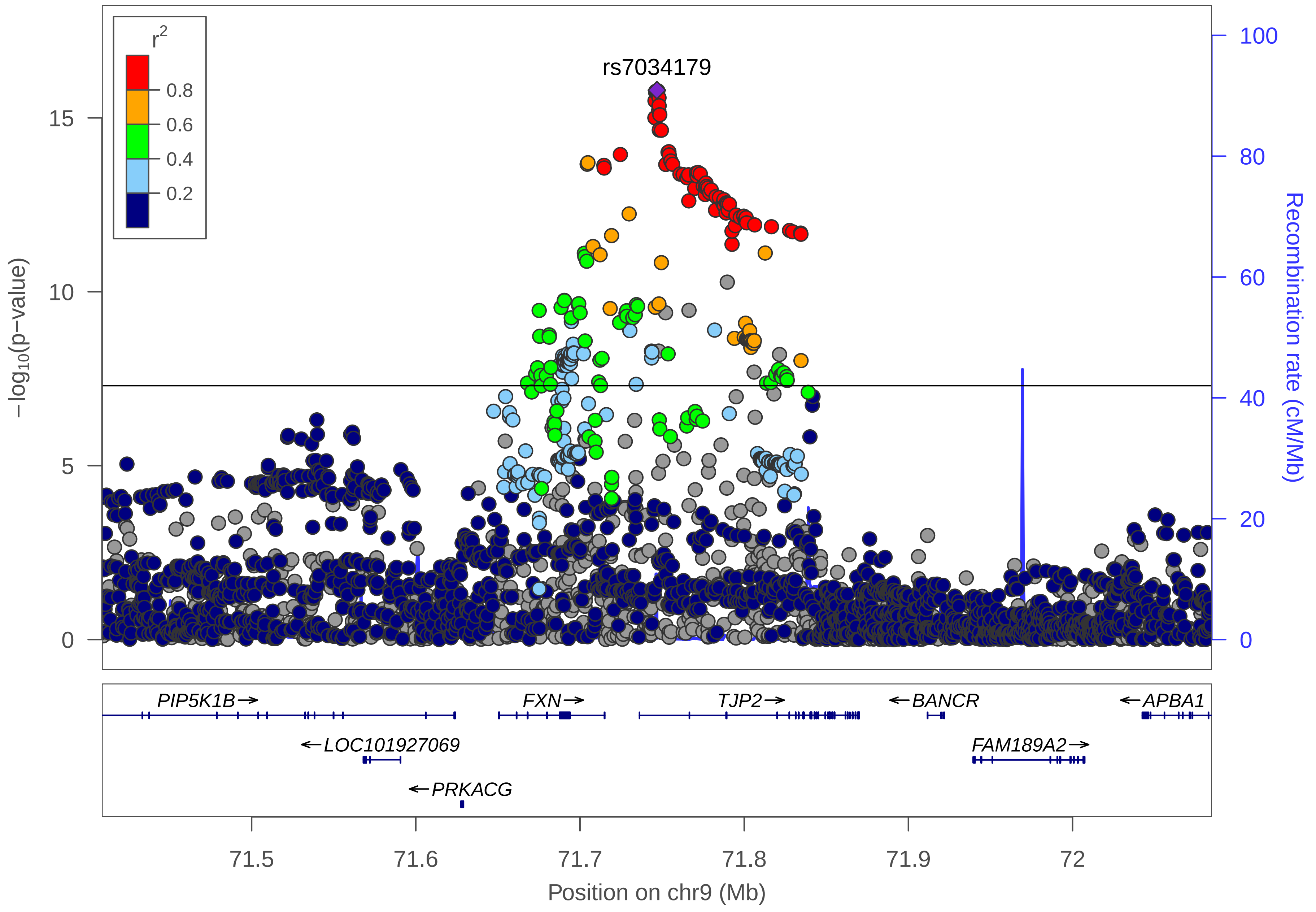
r2
0.8
0.6
0.4
0.2
rs7034179
15
10
5
0
−log10(p−value)
100
80
60
40
20
0
Recombination rate (cM/Mb)
PIP5K1B→
FXN→
TJP2→
←BANCR
←APBA1
←LOC101927069
FAM189A2→
←PRKACG
71.5
71.6
71.7
71.8
71.9
72
Position on chr9 (Mb)

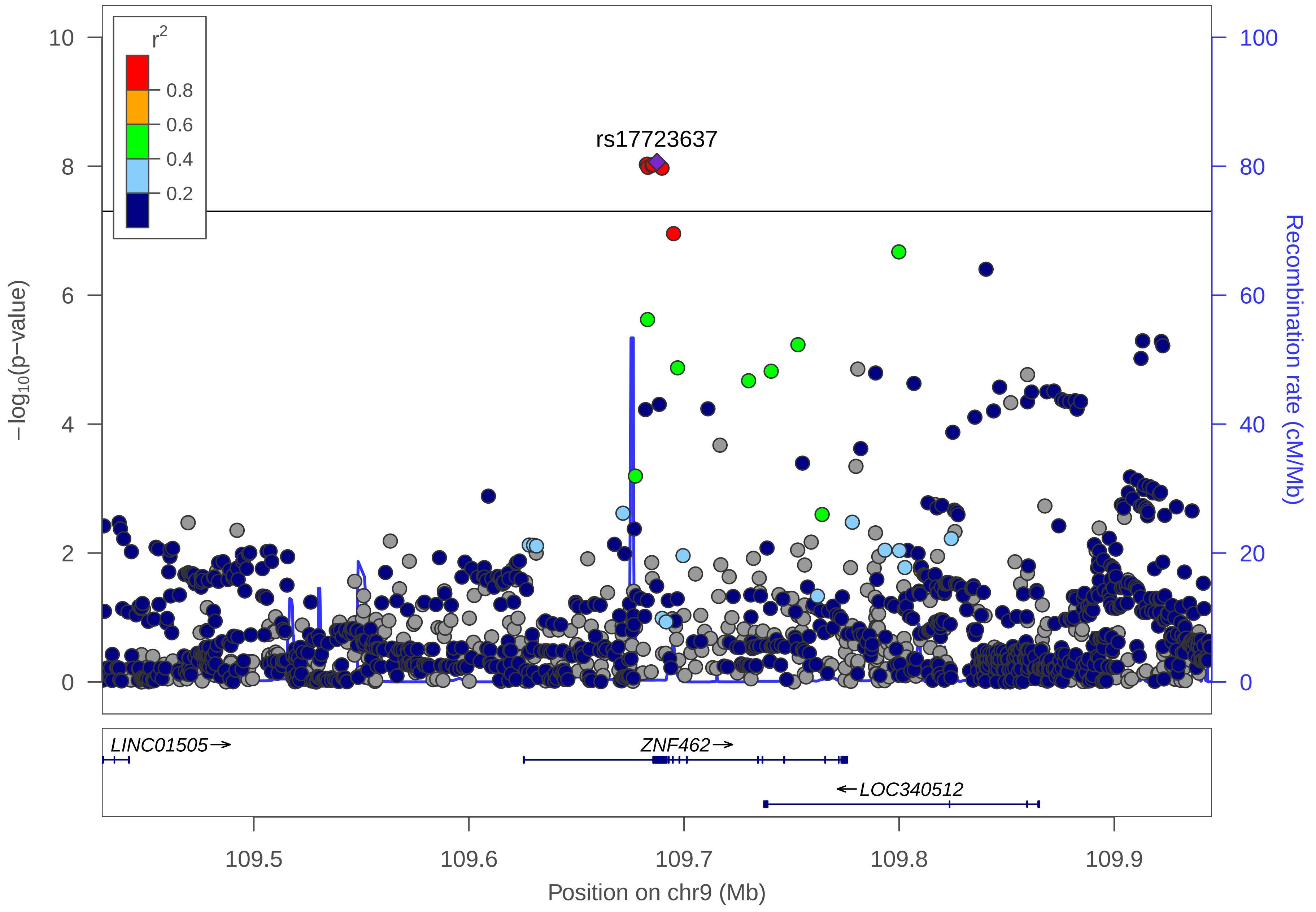

r2
0.8
0.6
0.4
0.2
rs17723637
10
8
6
4
2
0
−log10(p−value)
100
80
60
40
20
0
Recombination rate (cM/Mb)
LINC01505→
ZNF462→
←LOC340512
109.5
109.6
109.7
109.8
109.9
Position on chr9 (Mb)

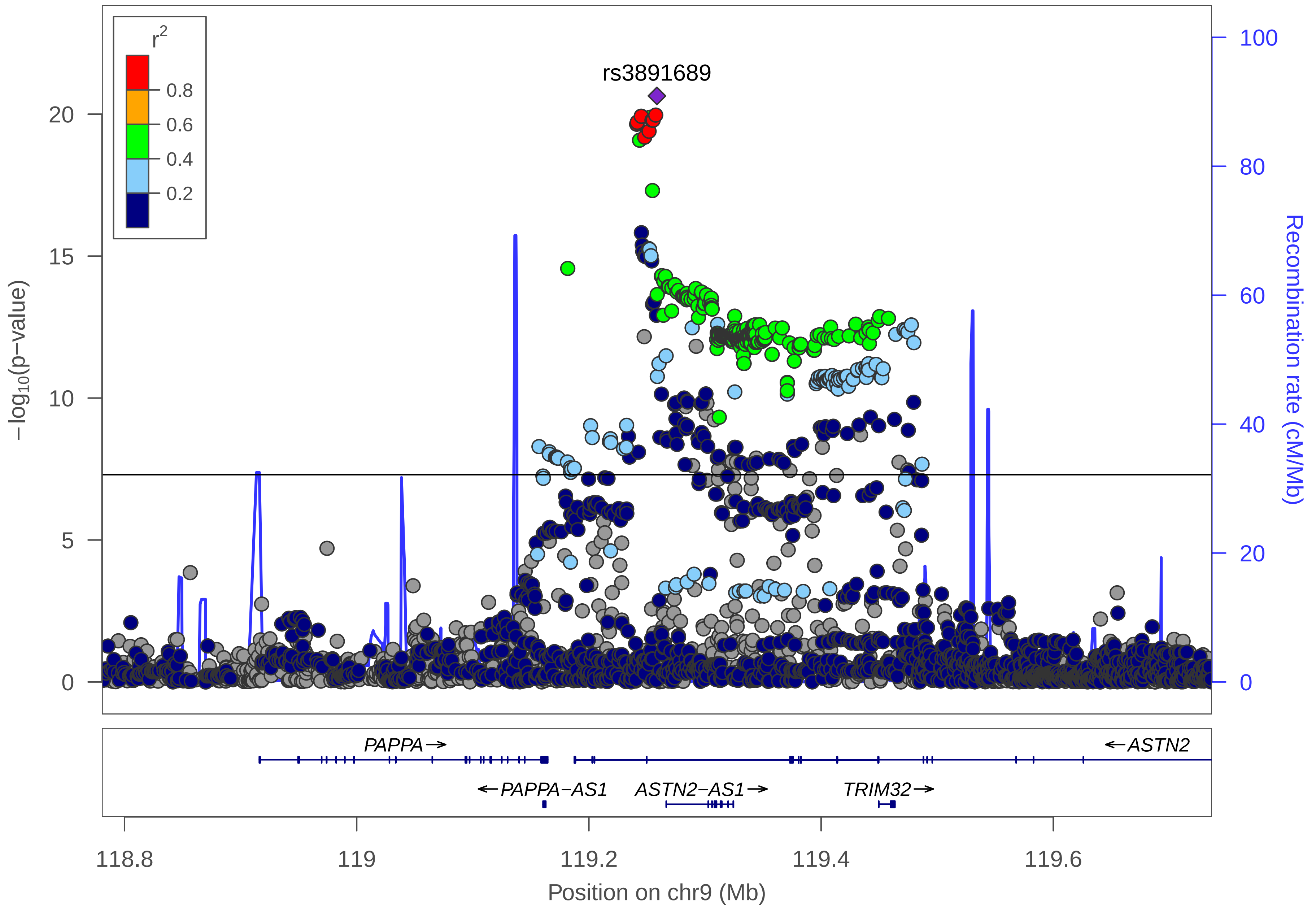

r2
0.8
0.6
0.4
0.2
rs3891689
20
15
10
5
0
−log10(p−value)
100
80
60
40
20
0
Recombination rate (cM/Mb)
PAPPA→
←ASTN2
←PAPPA−AS1
ASTN2−AS1→
TRIM32→
118.8
119
119.2
119.4
119.6
Position on chr9 (Mb)

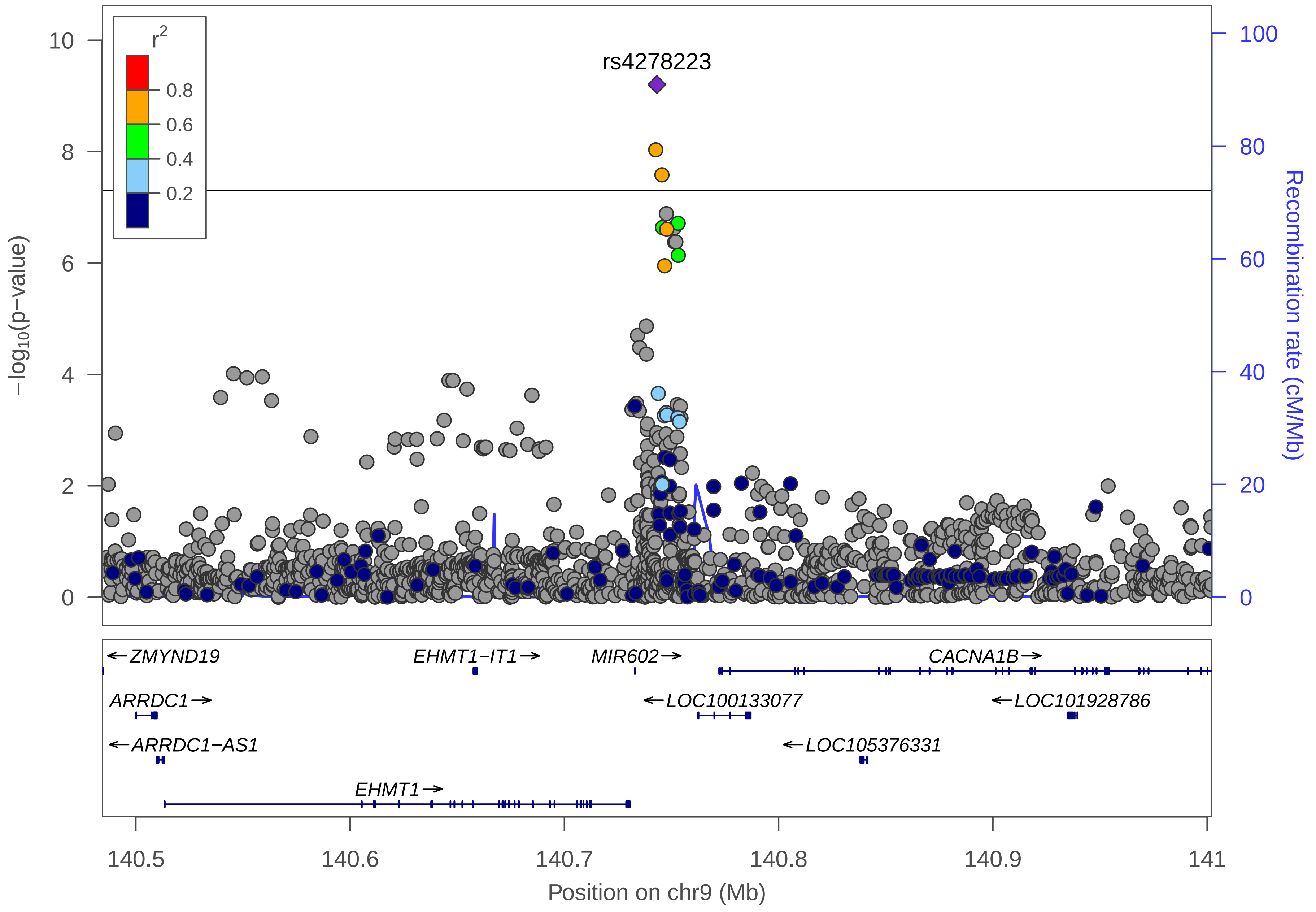

r2
0.8
0.6
0.4
0.2
rs4278223
10
8
6
4
2
0
100
80
60
40
20
0
−log10(p−value)
Recombination rate (cM/Mb)
←ZMYND19
EHMT1−IT1→
MIR602→
CACNA1B→
ARRDC1→
←LOC100133077
←LOC101928786
←ARRDC1−AS1
←LOC105376331
EHMT1→
140.5
140.6
140.7
140.8
140.9
141
Position on chr9 (Mb)

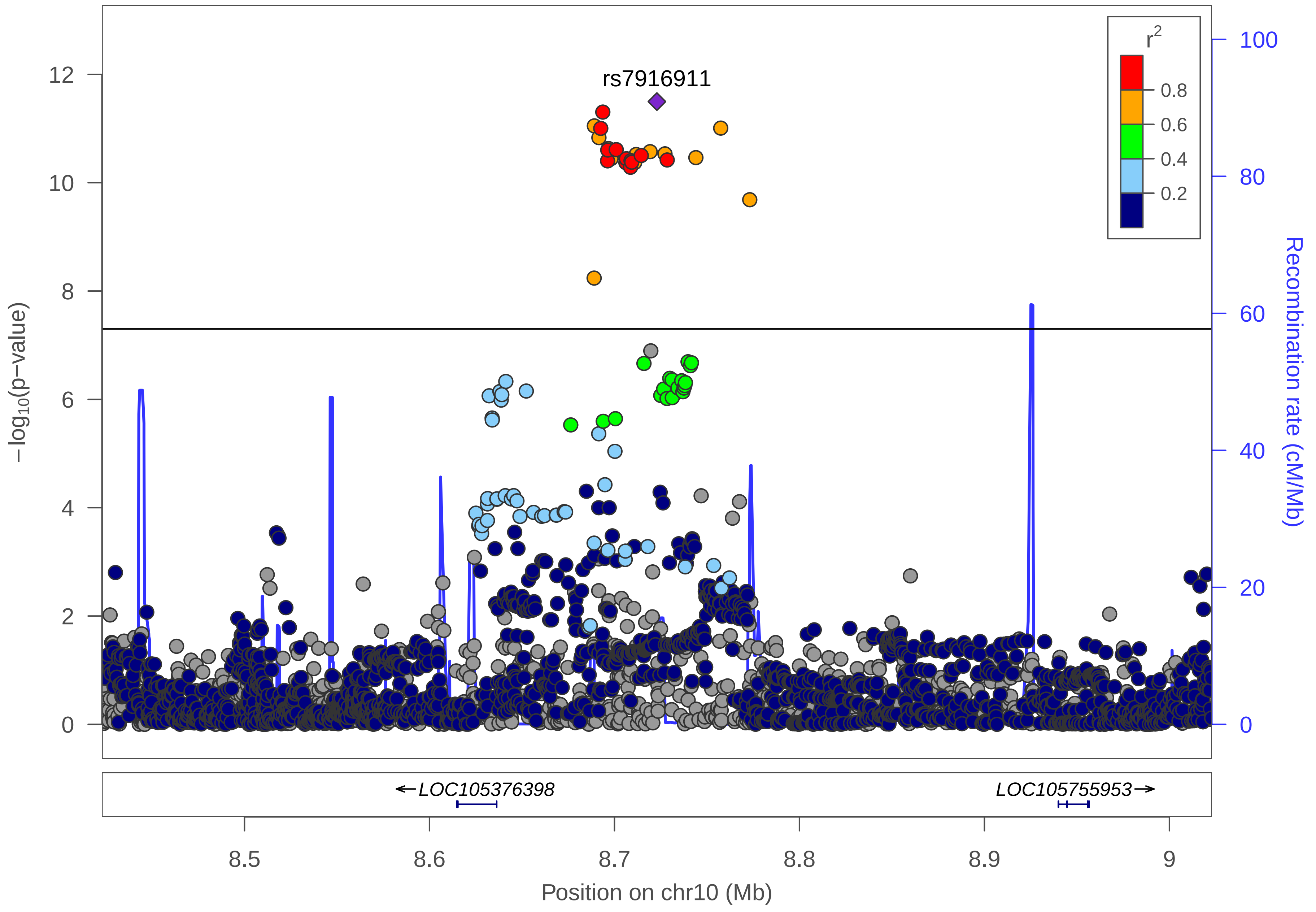

rs7916911
r2
0.8
0.6
0.4
0.2
12
10
8
6
4
2
0
−log10(p−value)
100
80
60
40
20
0
Recombination rate (cM/Mb)
←LOC105376398
LOC105755953→
8.5
8.6
8.7
8.8
8.9
9
Position on chr10 (Mb)

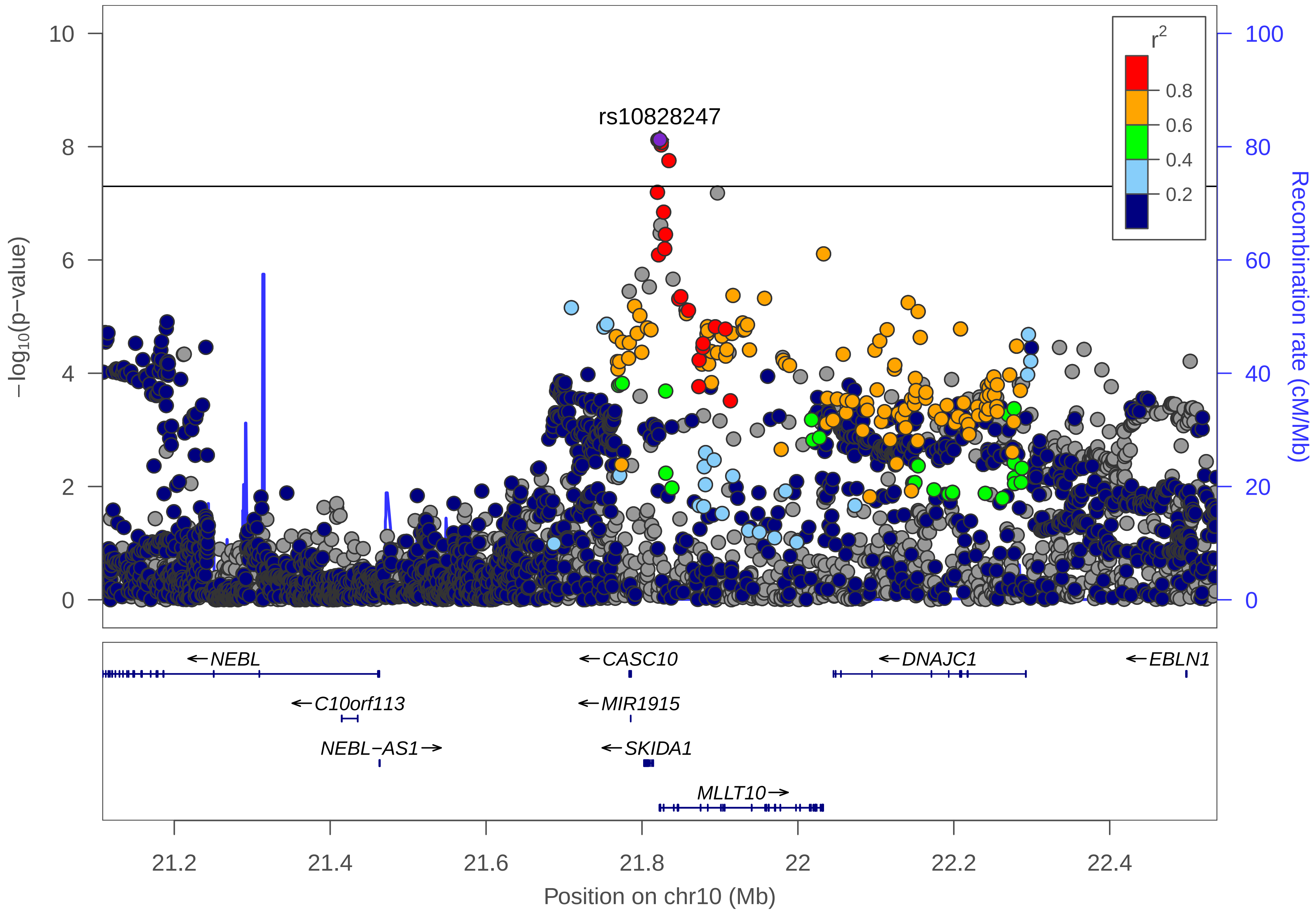

rs10828247
r2
0.8
0.6
0.4
0.2
10
8
6
4
2
0
100
80
60
40
20
0
−log10(p−value)
Recombination rate (cM/Mb)
←NEBL
←CASC10
←DNAJC1
←EBLN1
←C10orf113
←MIR1915
NEBL−AS1→
←SKIDA1
MLLT10→
21.2
21.4
21.6
21.8
22
22.2
22.4
Position on chr10 (Mb)

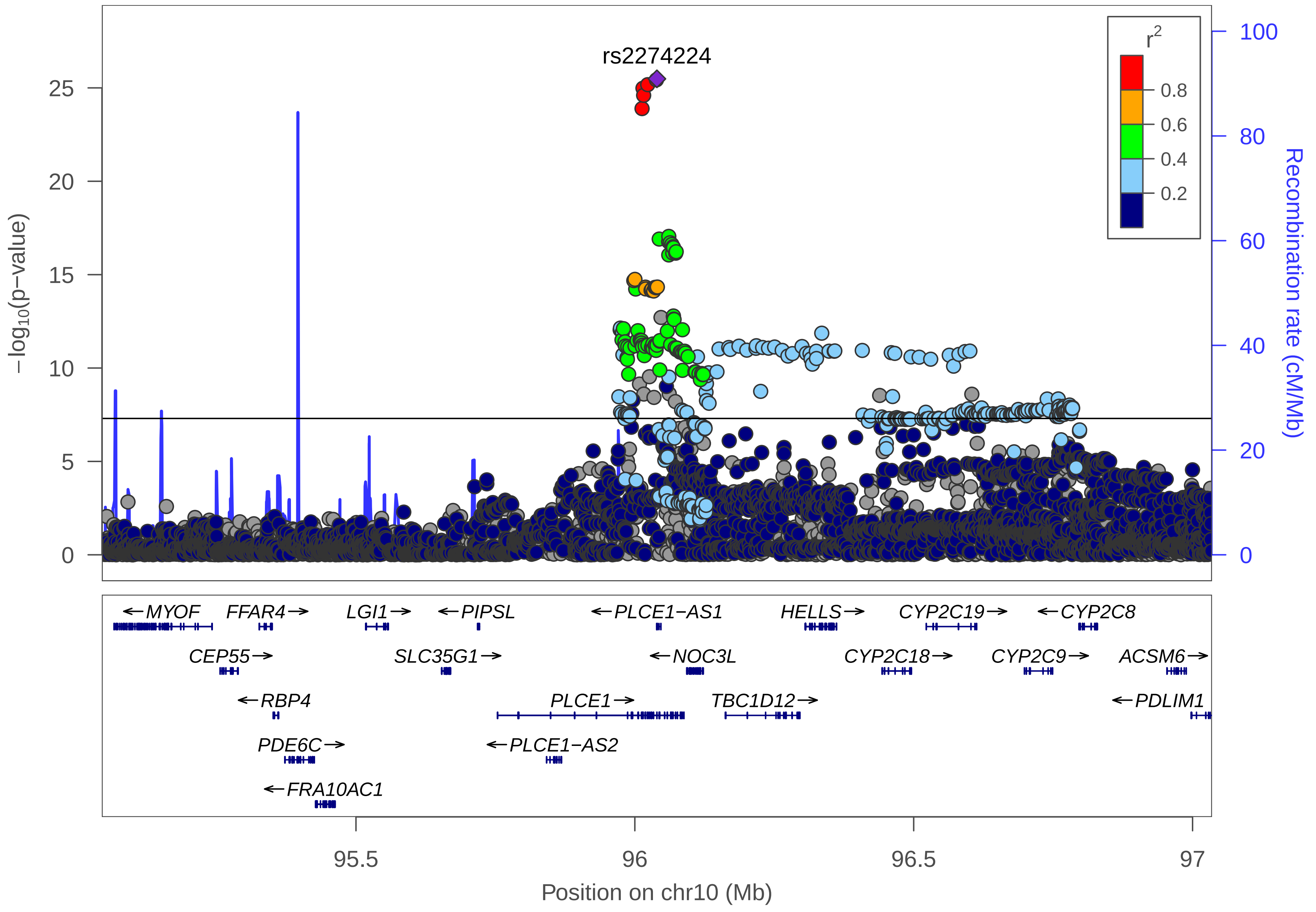

rs2274224
r2
0.8
0.6
0.4
0.2
−log10(p−value)
Recombination rate (cM/Mb)
0
5
10
15
20
25
0
20
40
60
80
100
←MYOF
FFAR4→
LGI1→
←PIPSL
←PLCE1−AS1
HELLS→
CYP2C19→
←CYP2C8
CEP55→
SLC35G1→
←NOC3L
CYP2C18→
CYP2C9→
ACSM6→
←RBP4
PLCE1→
TBC1D12→
←PDLIM1
PDE6C→
←PLCE1−AS2
←FRA10AC1
95.5
96
96.5
97
Position on chr10 (Mb)

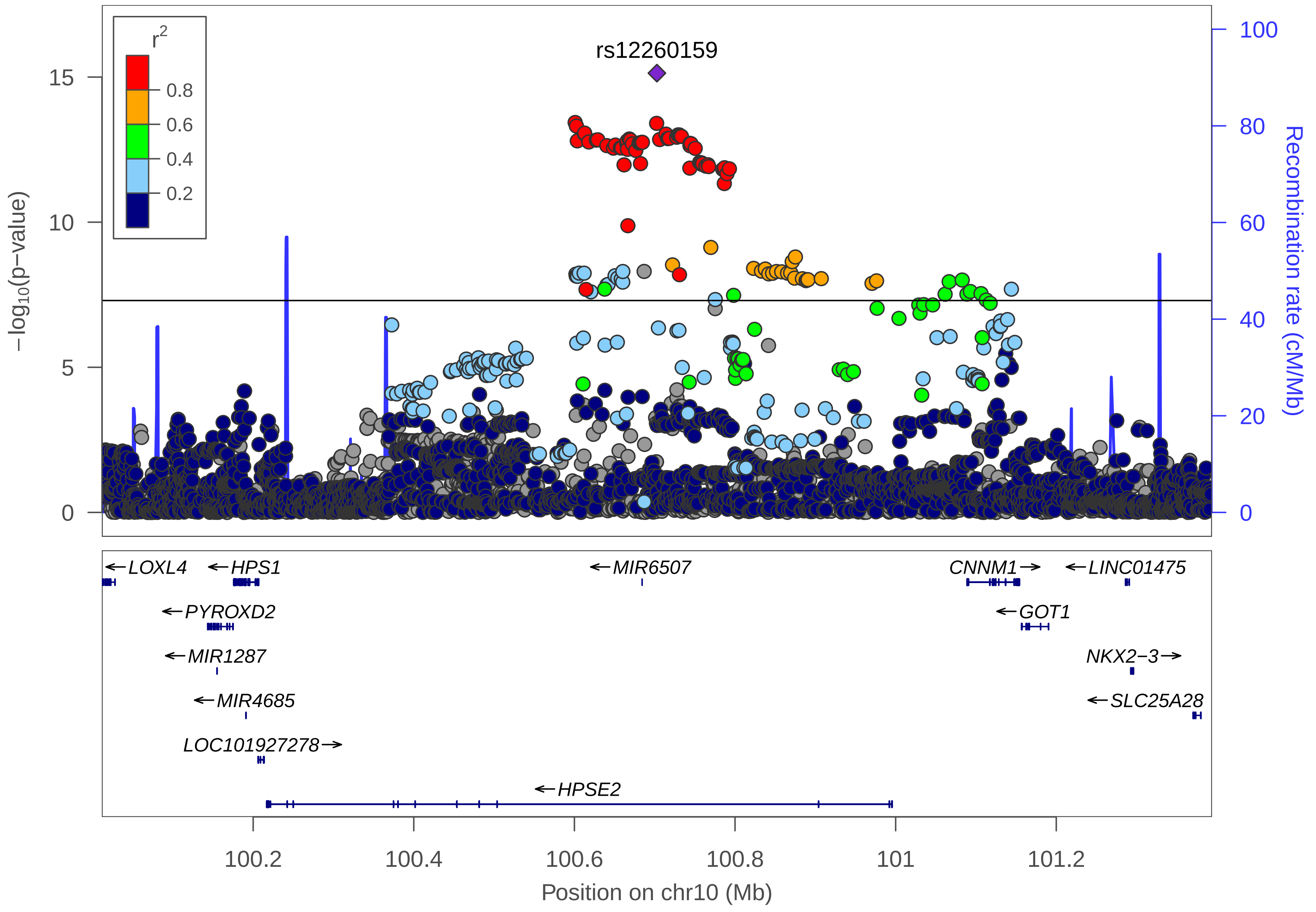

rs12260159
r2
0.8
0.6
0.4
0.2
−log10(p−value)
0
5
10
15
Recombination rate (cM/Mb)
0
20
40
60
80
100
LOXL4
HPS1
MIR6507
CNNM1
LINC01475
PYROXD2
GOT1
MIR1287
NKX2−3
MIR4685
SLC25A28
LOC101927278
HPSE2
100.2
100.4
100.6
100.8
101
101.2
Position on chr10 (Mb)

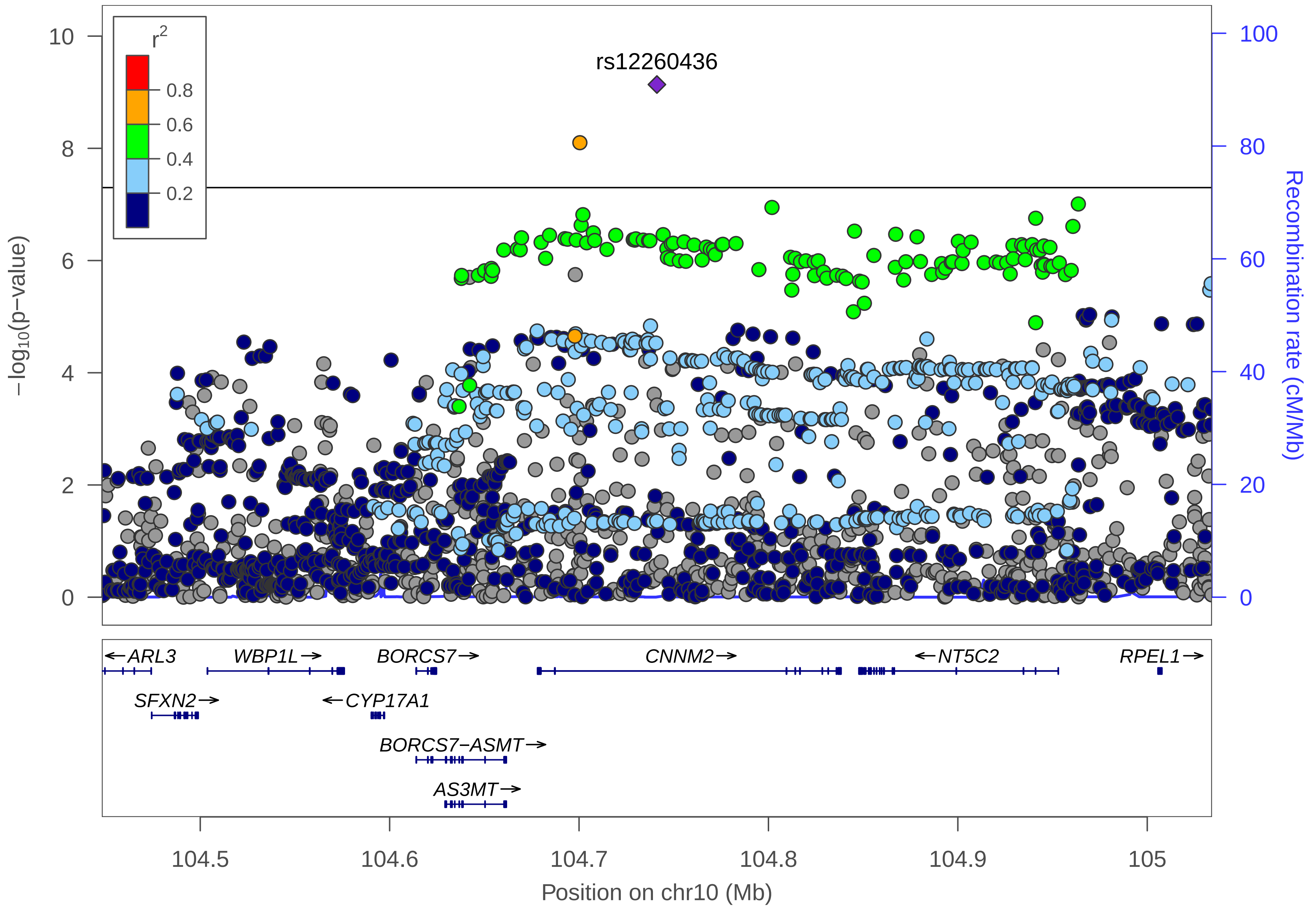

r2
0.8
0.6
0.4
0.2
rs12260436
−log10(p−value)
Recombination rate (cM/Mb)
10
8
6
4
2
0
100
80
60
40
20
0
←ARL3
WBP1L→
BORCS7→
CNNM2→
←NT5C2
RPEL1→
SFXN2→
←CYP17A1
BORCS7−ASMT→
AS3MT→
104.5
104.6
104.7
104.8
104.9
105
Position on chr10 (Mb)

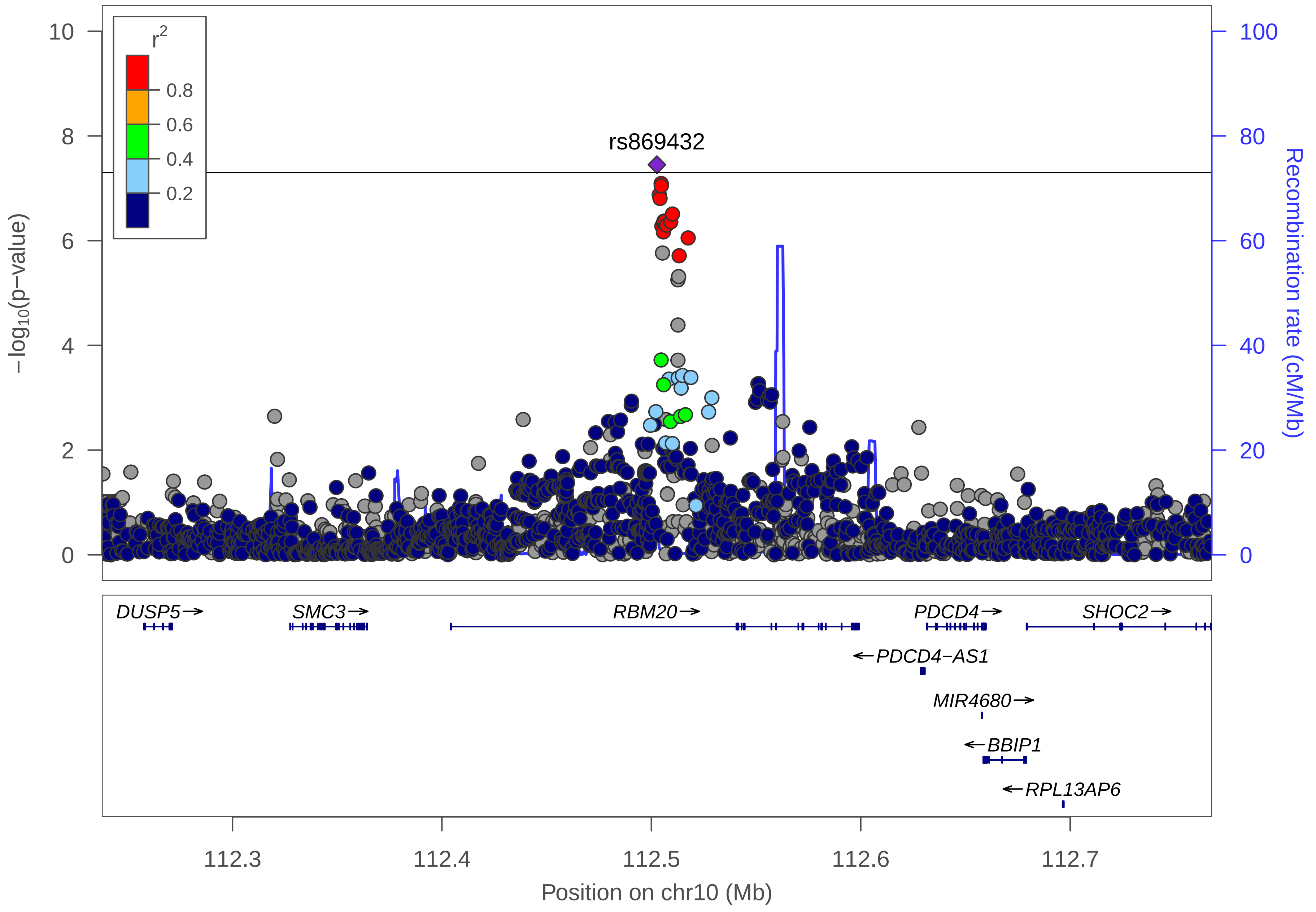

r2
0.8
0.6
0.4
0.2
rs869432
−log10(p−value)
Recombination rate (cM/Mb)
0
2
4
6
8
10
0
20
40
60
80
100
DUSP5→
SMC3→
RBM20→
PDCD4→
SHOC2→
←PDCD4−AS1
MIR4680→
←BBIP1
←RPL13AP6
112.3
112.4
112.5
112.6
112.7
Position on chr10 (Mb)

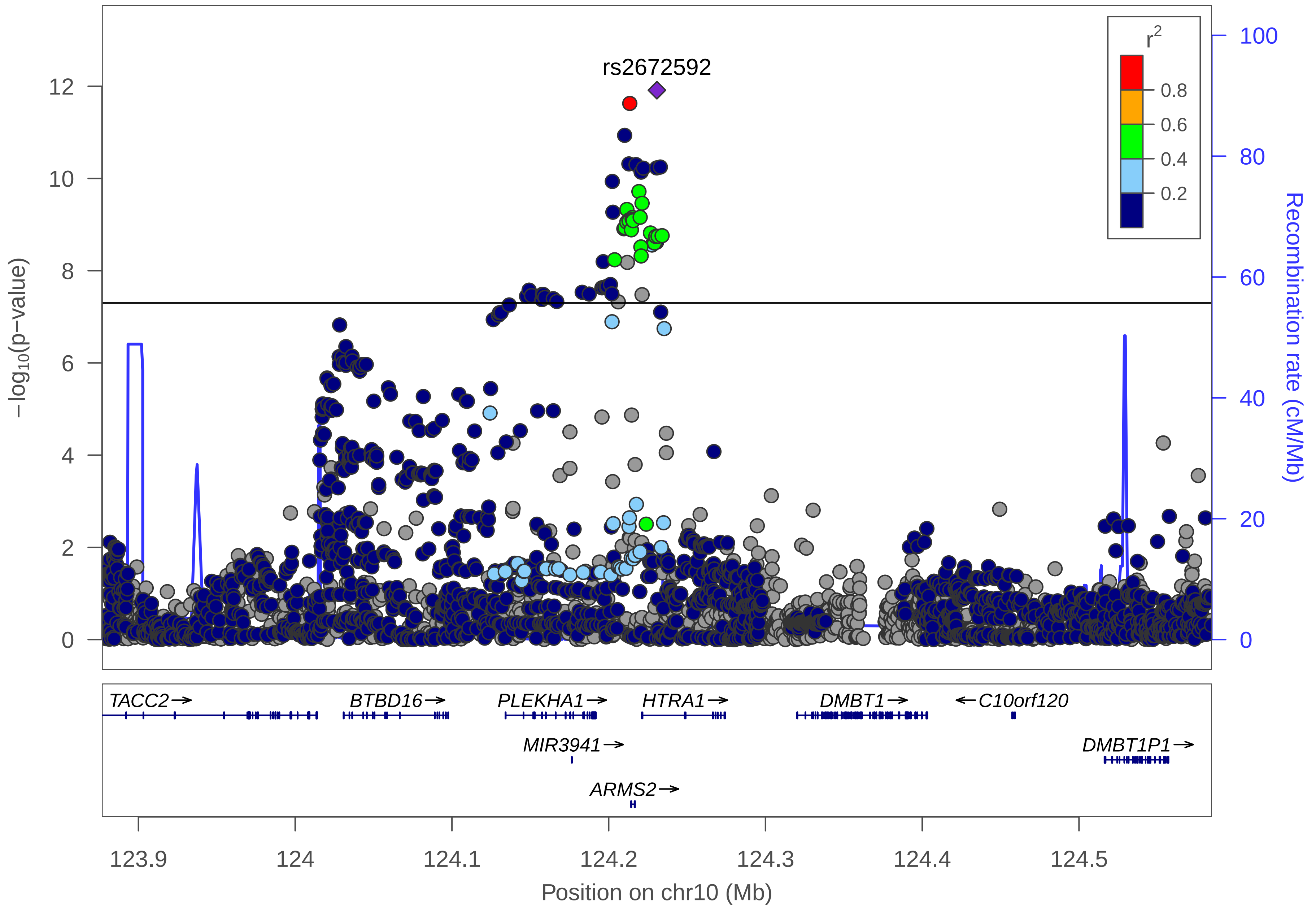
rs2672592
r2
0.8
0.6
0.4
0.2
12
10
8
6
4
2
0
−log10(p−value)
100
80
60
40
20
0
Recombination rate (cM/Mb)
TACC2→
BTBD16→
PLEKHA1→
HTRA1→
DMBT1→
←C10orf120
MIR3941→
DMBT1P1→
ARMS2→
123.9
124
124.1
124.2
124.3
124.4
124.5
Position on chr10 (Mb)

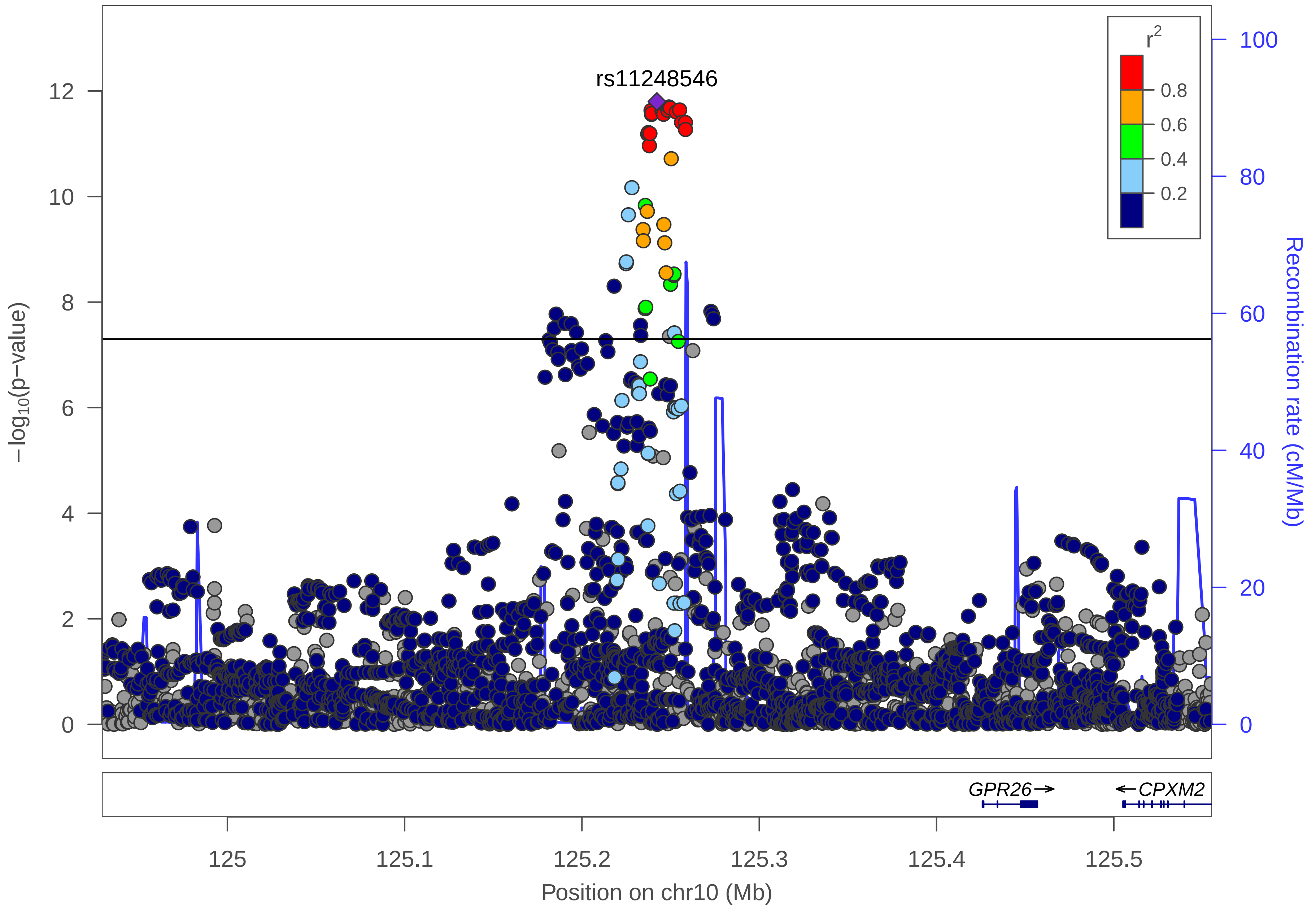

rs11248546
r2
0.8
0.6
0.4
0.2
−log10(p−value)
Recombination rate (cM/Mb)
0
2
4
6
8
10
12
0
20
40
60
80
100
GPR26→
←CPXM2
125
125.1
125.2
125.3
125.4
125.5
Position on chr10 (Mb)

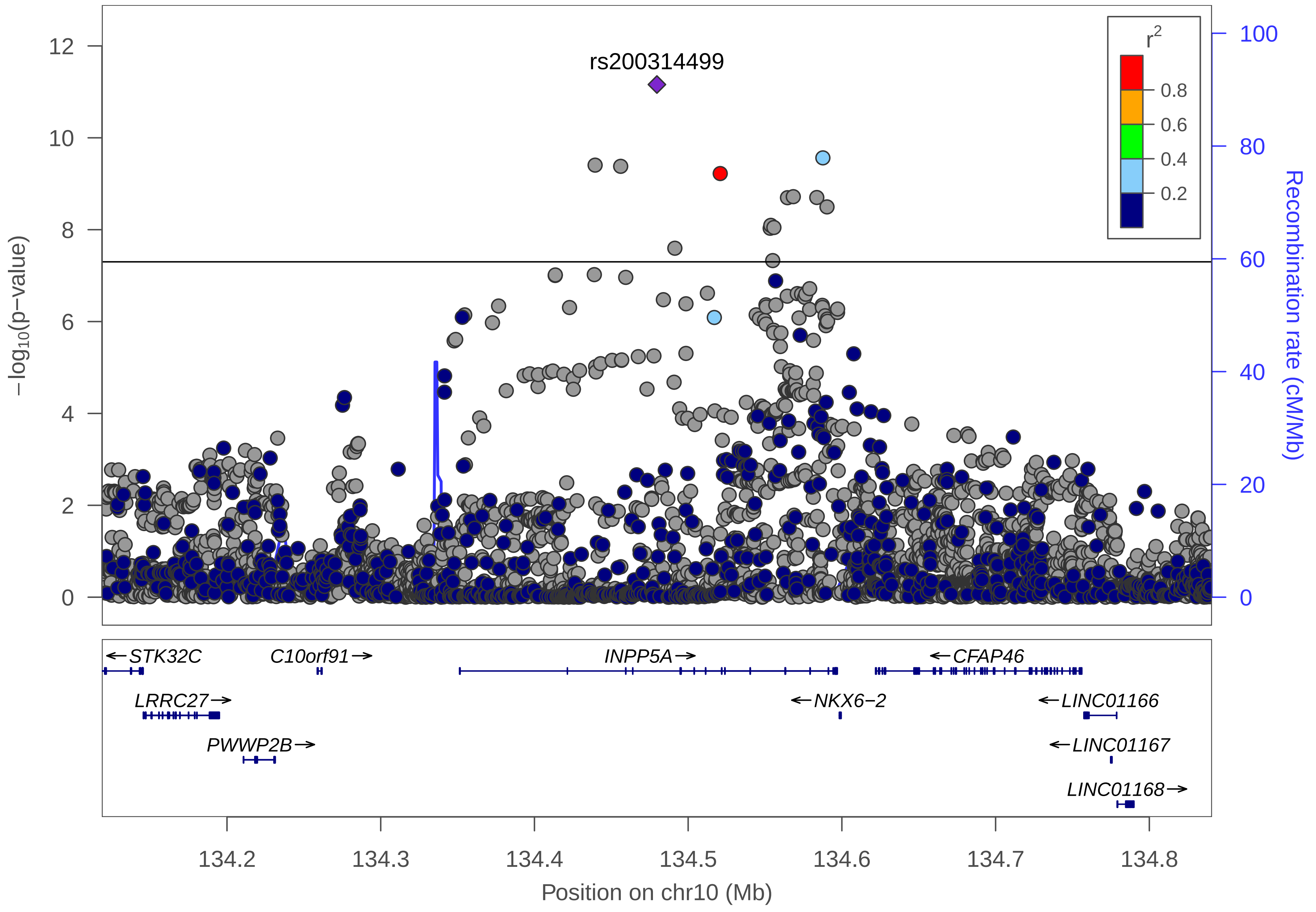

rs200314499
r2
0.8
0.6
0.4
0.2
12
10
8
6
4
2
0
−log10(p−value)
100
80
60
40
20
0
Recombination rate (cM/Mb)
STK32C
C10orf91
INPP5A
CFAP46
LRRC27
NKX6−2
LINC01166
PWWP2B
LINC01167
LINC01168
134.2
134.3
134.4
134.5
134.6
134.7
134.8
Position on chr10 (Mb)

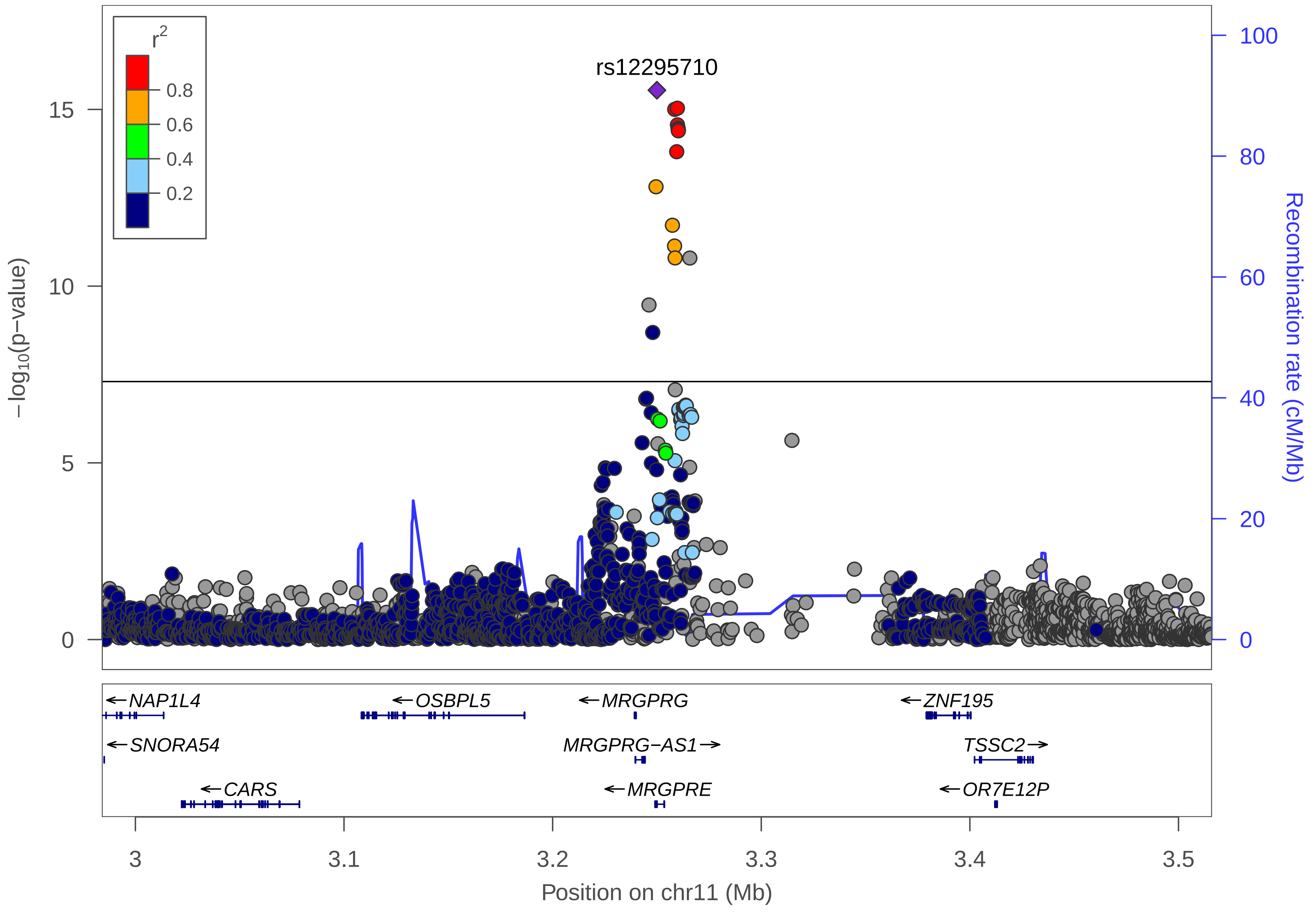

r2
0.8
0.6
0.4
0.2
rs12295710
15
10
5
0
100
80
60
40
20
0
−log10(p−value)
Recombination rate (cM/Mb)
NAP1L4
OSBPL5
MRGPRG
ZNF195
SNORA54
MRGPRG−AS1
TSSC2
CARS
MRGPRE
OR7E12P
3
3.1
3.2
3.3
3.4
3.5
Position on chr11 (Mb)

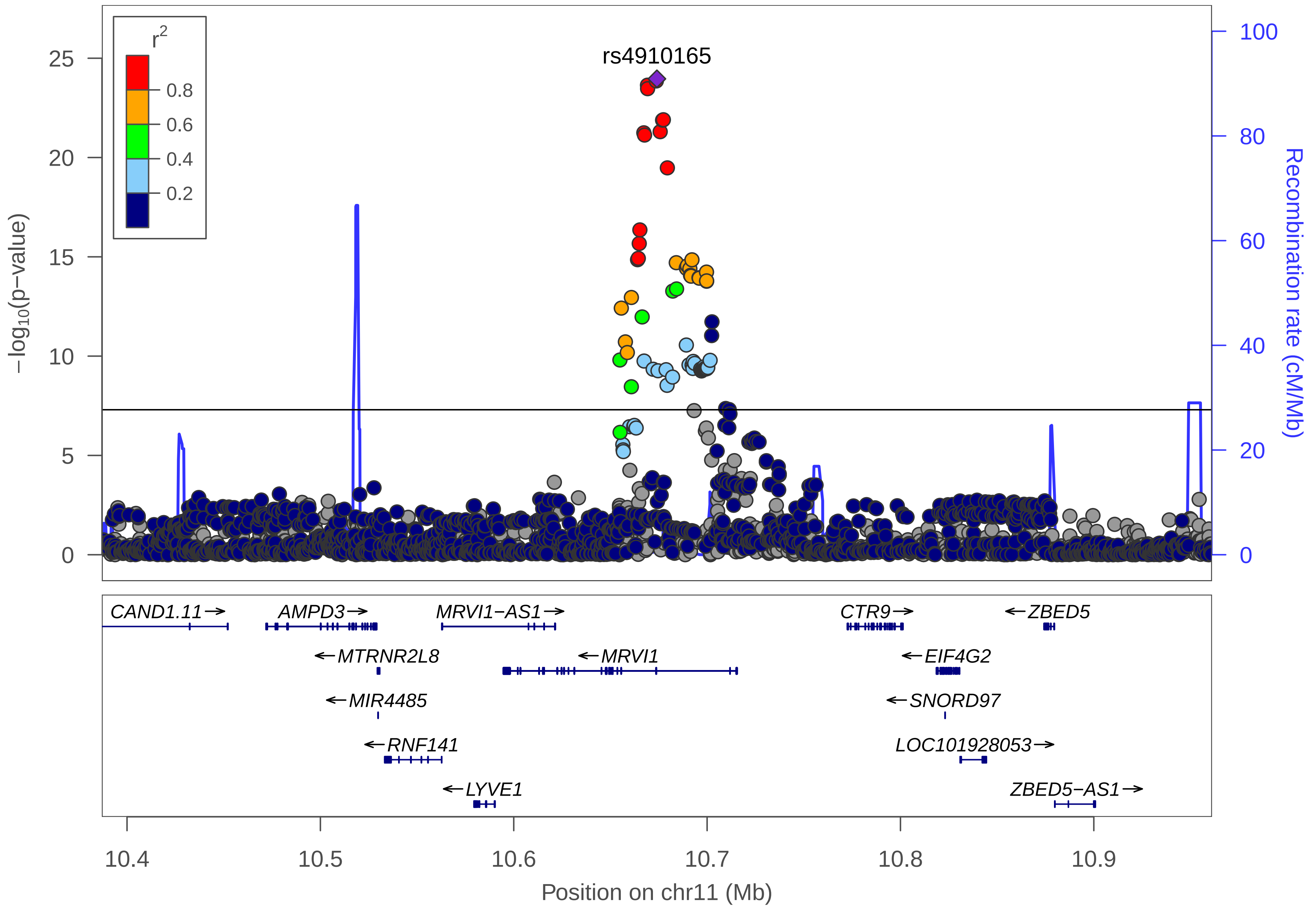

rs4910165
r2
0.8
0.6
0.4
0.2
−log10(p−value)
Recombination rate (cM/Mb)
CAND1.11
AMPD3
MRVI1−AS1
CTR9
ZBED5
MTRNR2L8
MRVI1
EIF4G2
MIR4485
SNORD97
RNF141
LOC101928053
LYVE1
ZBED5−AS1
10.4
10.5
10.6
10.7
10.8
10.9
Position on chr11 (Mb)

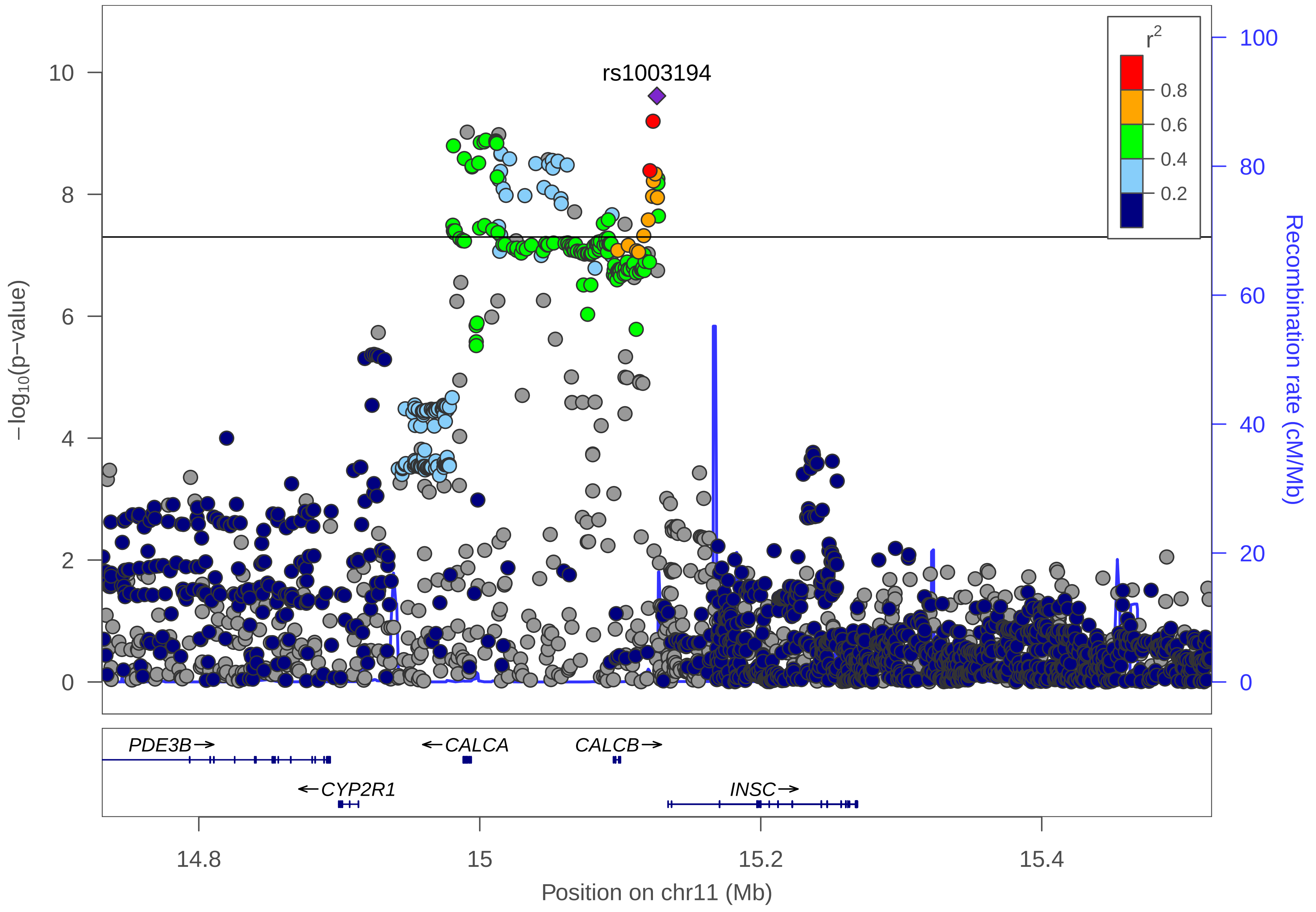

rs1003194
r2
0.8
0.6
0.4
0.2
−log10(p−value)
Recombination rate (cM/Mb)
PDE3B→
←CALCA
CALCB→
←CYP2R1
INSC→
14.8
15
15.2
15.4
Position on chr11 (Mb)

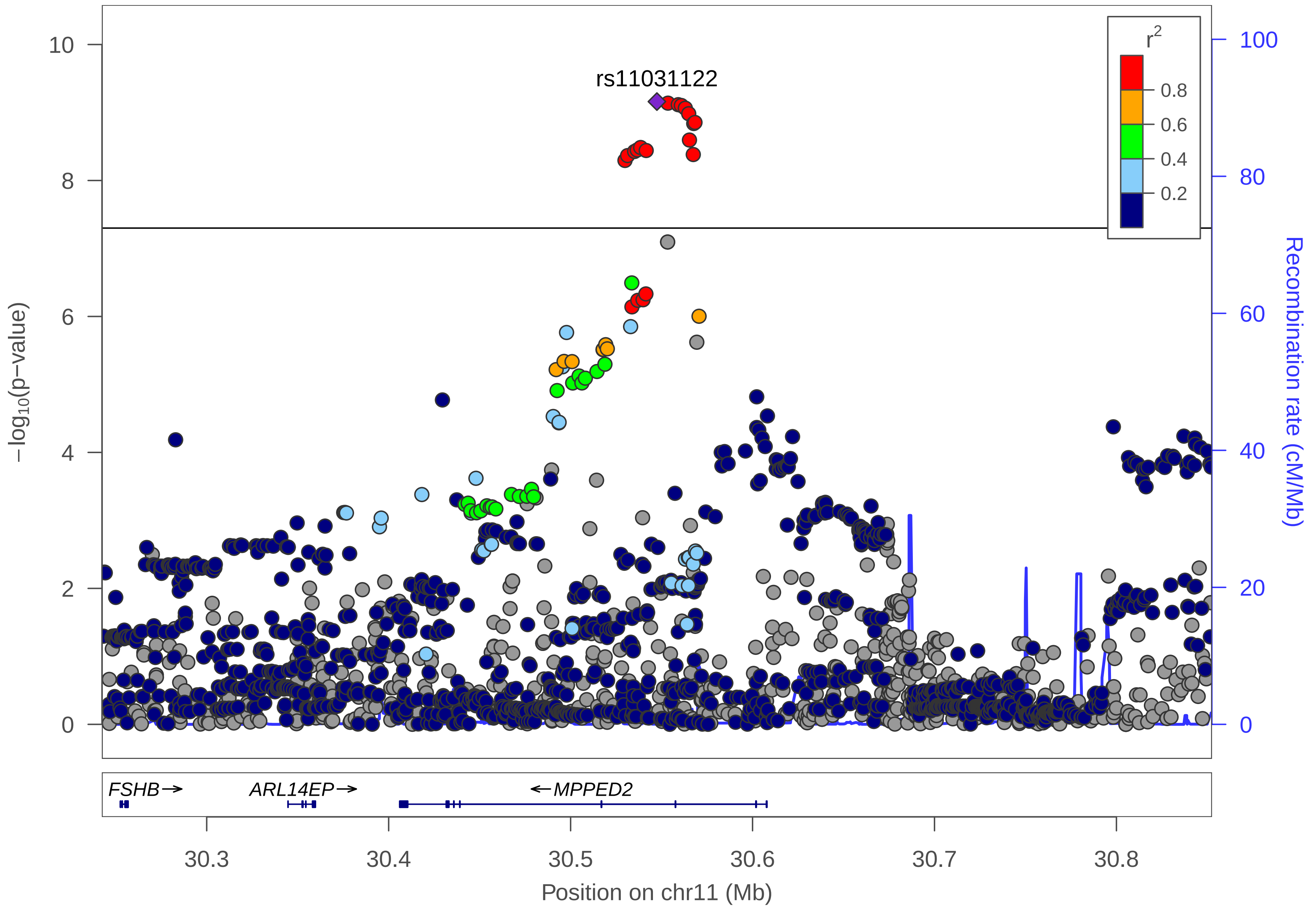

rs11031122
r2
0.8
0.6
0.4
0.2
10
8
6
4
2
0
−log10(p−value)
100
80
60
40
20
0
Recombination rate (cM/Mb)
FSHB→
ARL14EP→
←MPPED2
30.3
30.4
30.5
30.6
30.7
30.8
Position on chr11 (Mb)

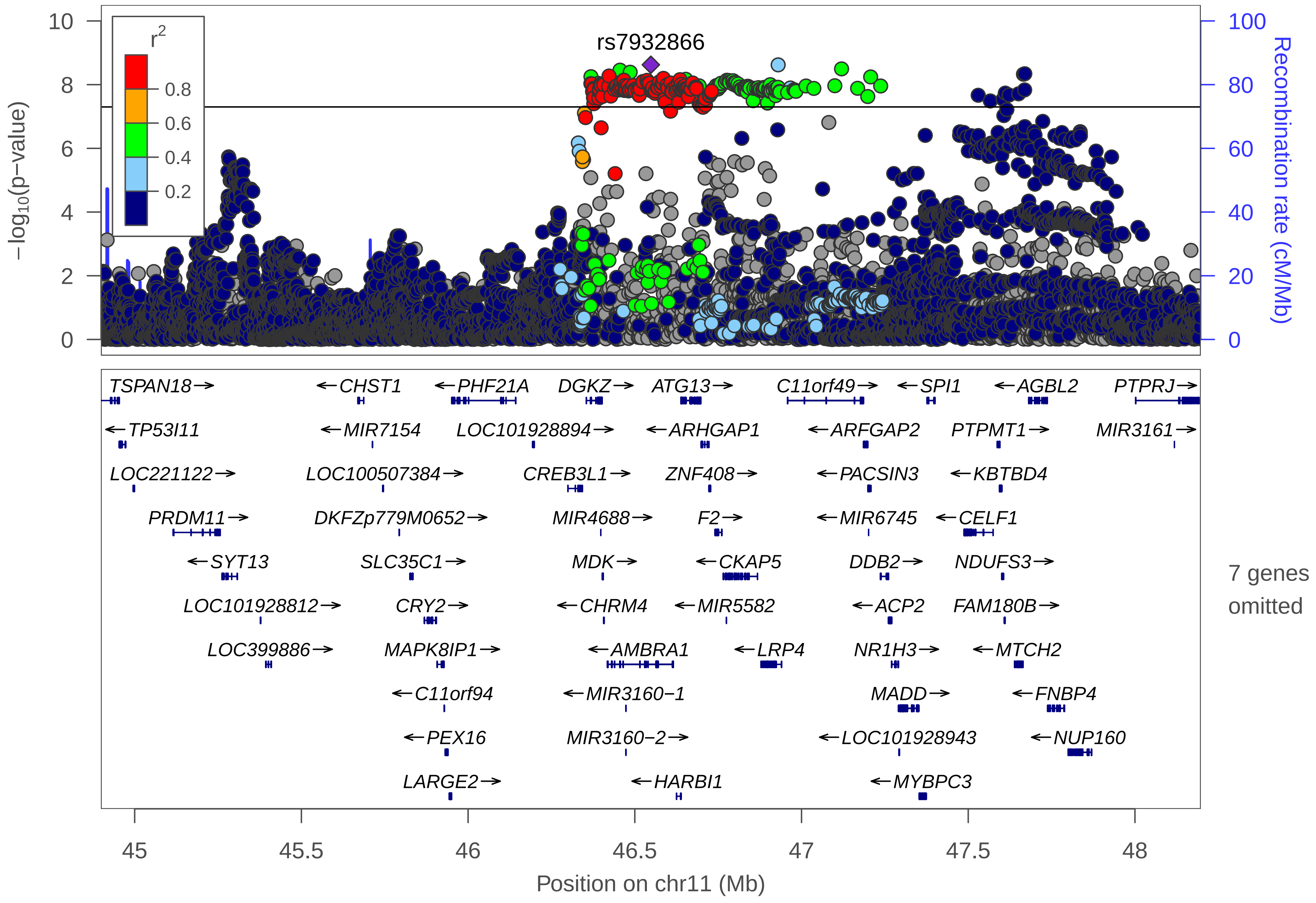
rs7932866
r2
0.8
0.6
0.4
0.2
−log10(p−value)
Recombination rate (cM/Mb)
TSPAN18→
←TP53I11
LOC221122→
PRDM11→
←SYT13
LOC101928812→
LOC399886→
←CHST1
←MIR7154
LOC100507384→
DKFZp779M0652→
SLC35C1→
CRY2→
MAPK8IP1→
←C11orf94
←PEX16
LARGE2→
←PHF21A
LOC101928894→
DGKZ→
CREB3L1→
MIR4688→
MDK→
←CHRM4
←AMBRA1
←MIR3160−1
MIR3160−2→
ATG13→
←ARHGAP1
ZNF408→
F2→
←CKAP5
←MIR5582
←LRP4
←HARBI1
C11orf49→
←ARFGAP2
←PACSIN3
←MIR6745
DDB2→
←ACP2
NR1H3→
MADD→
←LOC101928943
←MYBPC3
←SPI1
PTPMT1→
←KBTBD4
←CELF1
NDUFS3→
FAM180B→
←MTCH2
←FNBP4
←NUP160
←AGBL2
PTPRJ→
MIR3161→
7 genes omitted
Position on chr11 (Mb)

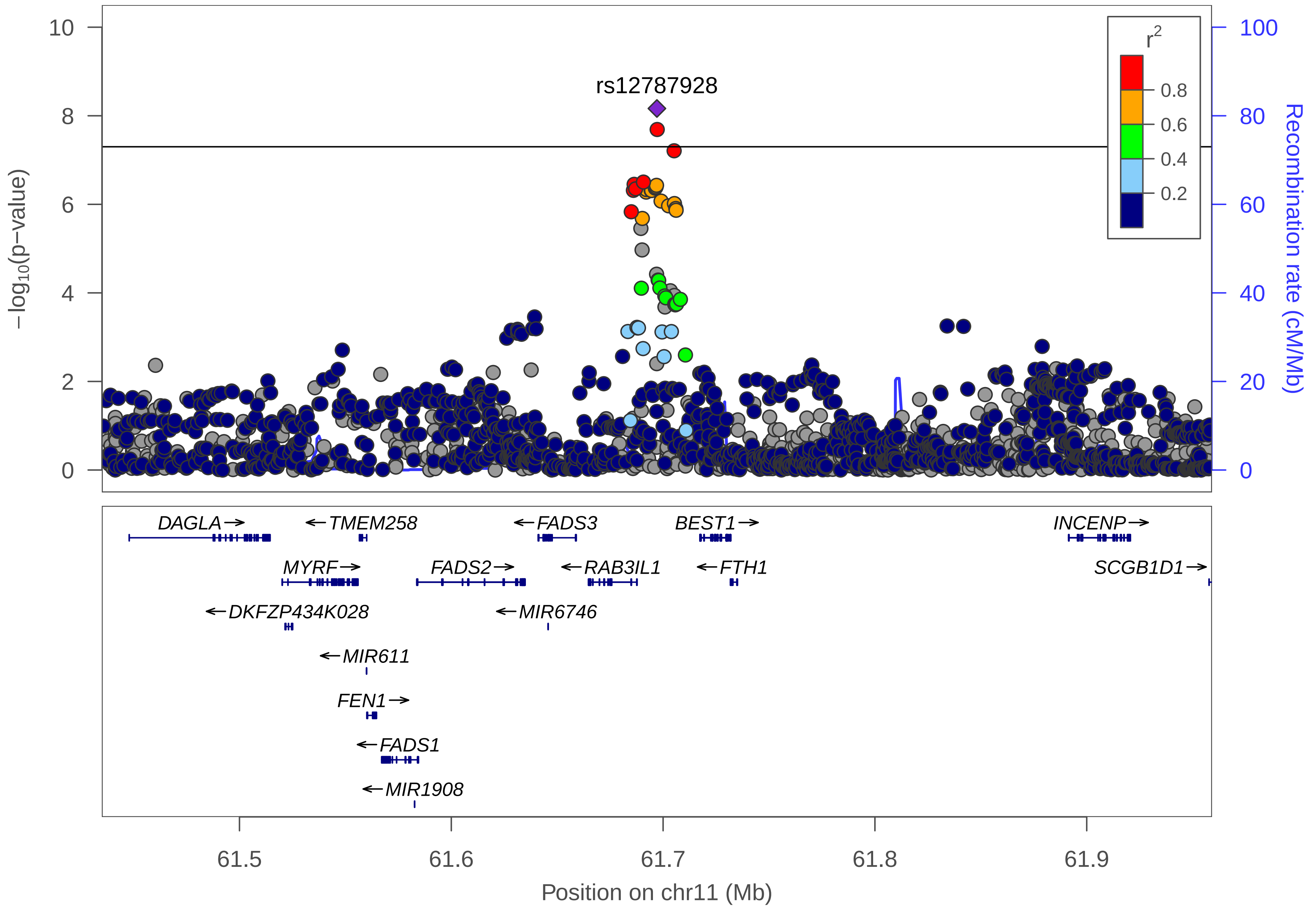

rs12787928
r2
0.8
0.6
0.4
0.2
−log10(p−value)
Recombination rate (cM/Mb)
0
2
4
6
8
10
0
20
40
60
80
100
DAGLA
TMEM258
FADS3
BEST1
INCENP
MYRF
FADS2
RAB3IL1
FTH1
SCGB1D1
DKFZP434K028
MIR6746
MIR611
FEN1
FADS1
MIR1908
61.5
61.6
61.7
61.8
61.9
Position on chr11 (Mb)

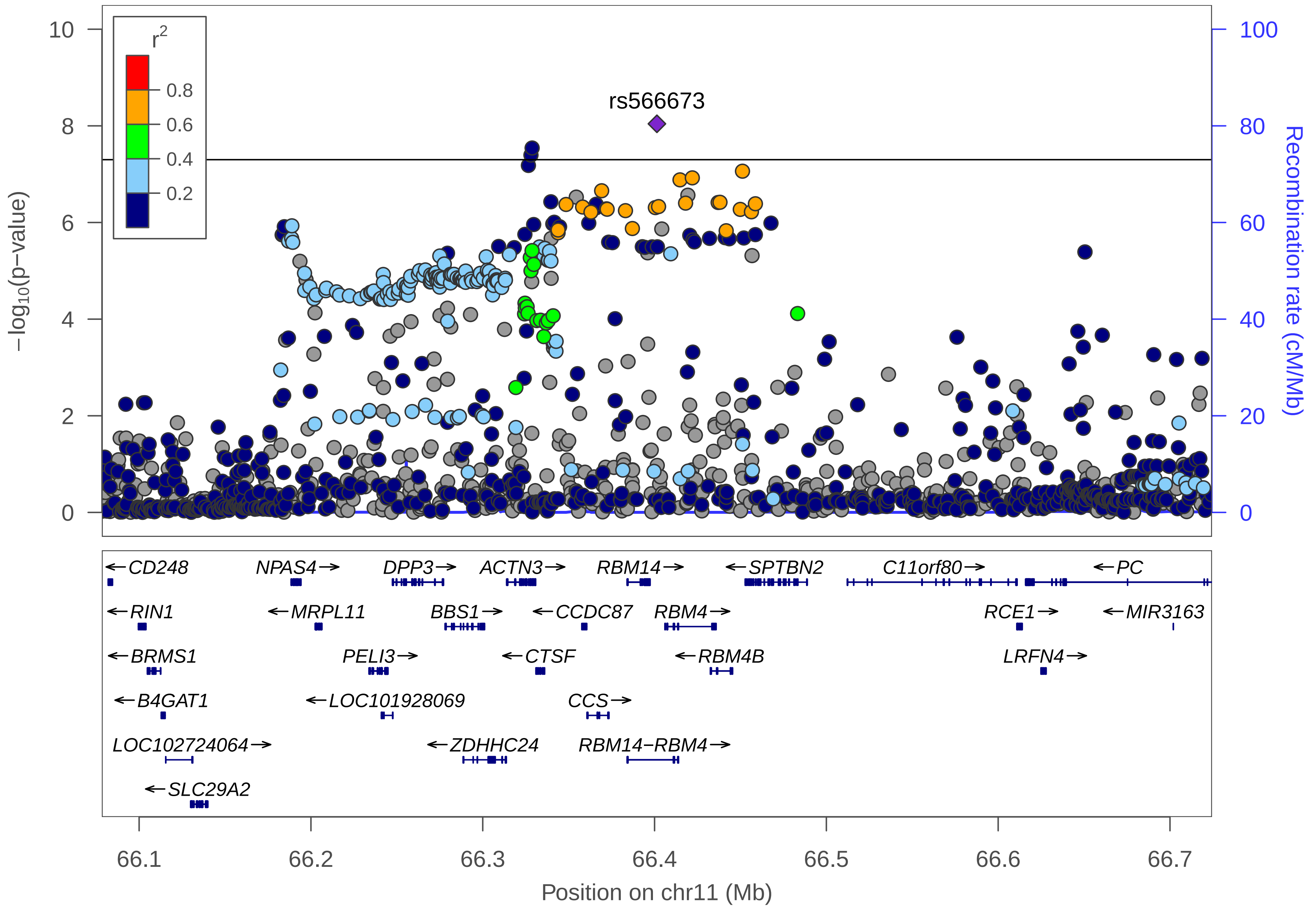

r2
0.8
0.6
0.4
0.2
rs566673
−log10(p−value)
Recombination rate (cM/Mb)
Position on chr11 (Mb)
66.1
66.2
66.3
66.4
66.5
66.6
66.7
CD248
NPAS4
DPP3
ACTN3
RBM14
SPTBN2
C11orf80
PC
RIN1
MRPL11
BBS1
CCDC87
RBM4
RCE1
MIR3163
BRMS1
PELI3
CTSF
RBM4B
LRFN4
B4GAT1
LOC101928069
CCS
LOC102724064
ZDHHC24
RBM14−RBM4
SLC29A2

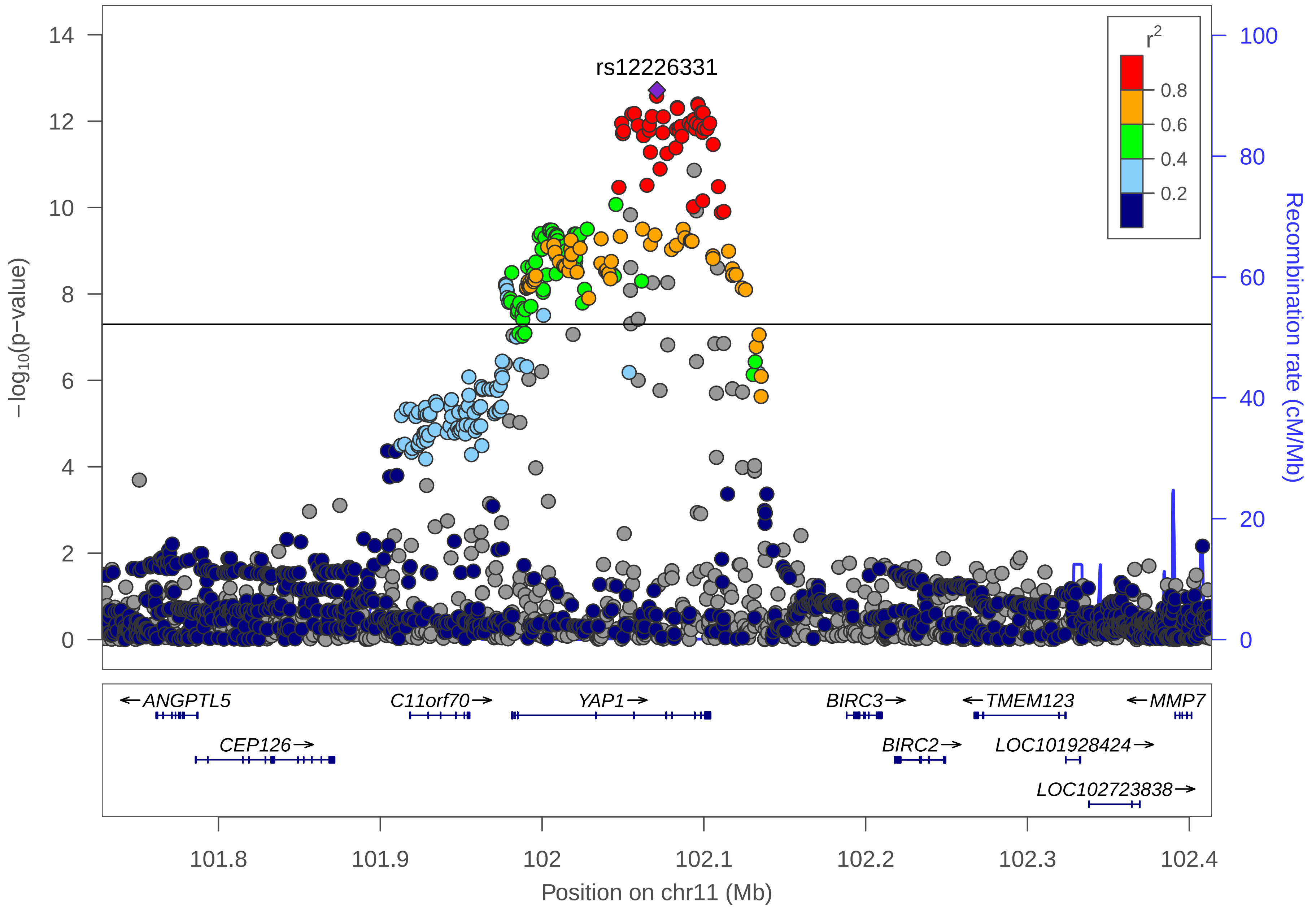

rs12226331
−log10(p−value)
Recombination rate (cM/Mb)
r2
0.8
0.6
0.4
0.2
0
2
4
6
8
10
12
14
0
20
40
60
80
100
ANGPTL5
CEP126
C11orf70
YAP1
BIRC3
BIRC2
TMEM123
LOC101928424
LOC102723838
MMP7
101.8
101.9
102
102.1
102.2
102.3
102.4
Position on chr11 (Mb)

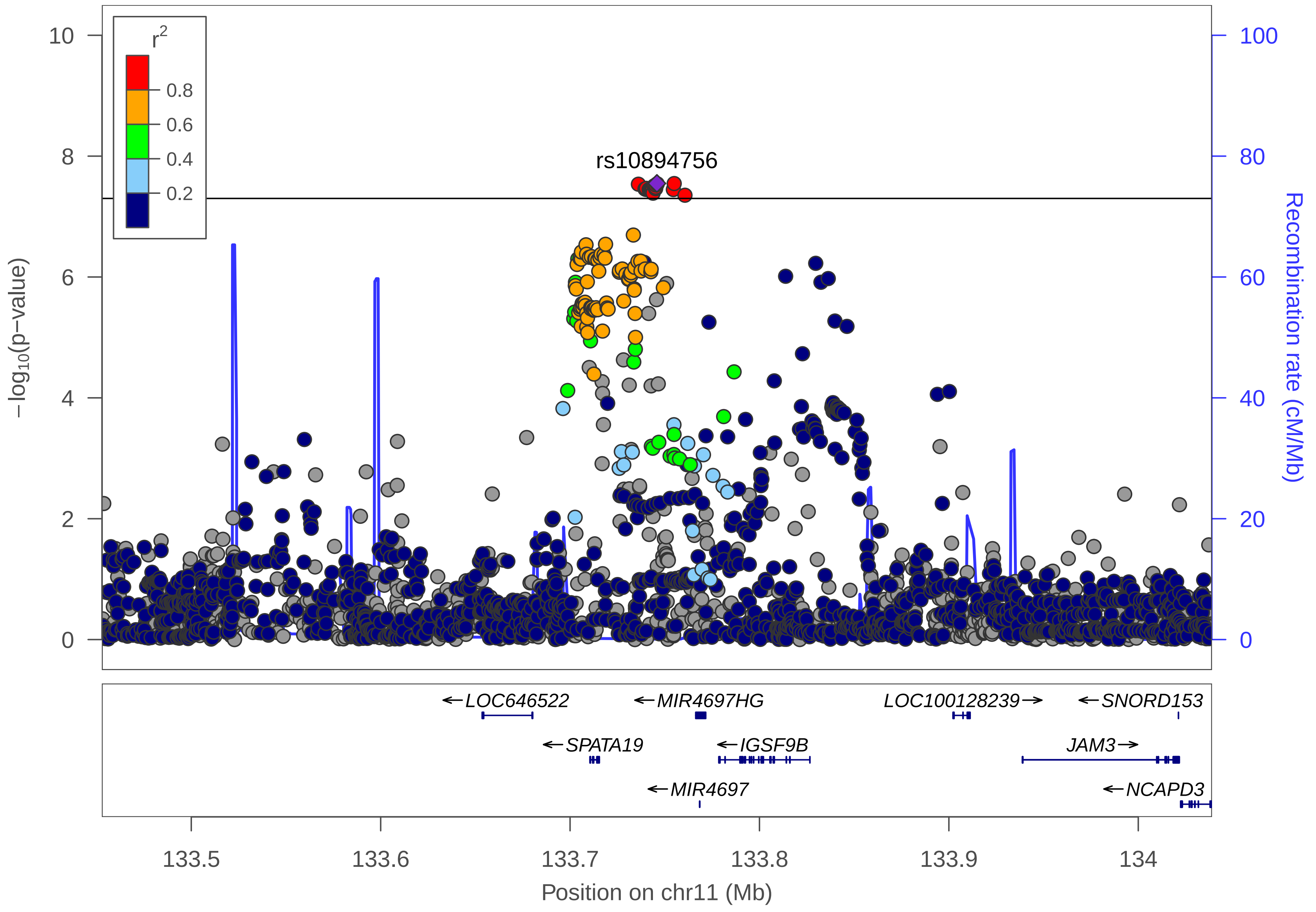
r2
0.8
0.6
0.4
0.2
rs10894756
−log10(p−value)
Recombination rate (cM/Mb)
LOC646522
MIR4697HG
LOC100128239
SNORD153
SPATA19
IGSF9B
JAM3
MIR4697
NCAPD3
Position on chr11 (Mb)
133.5
133.6
133.7
133.8
133.9
134

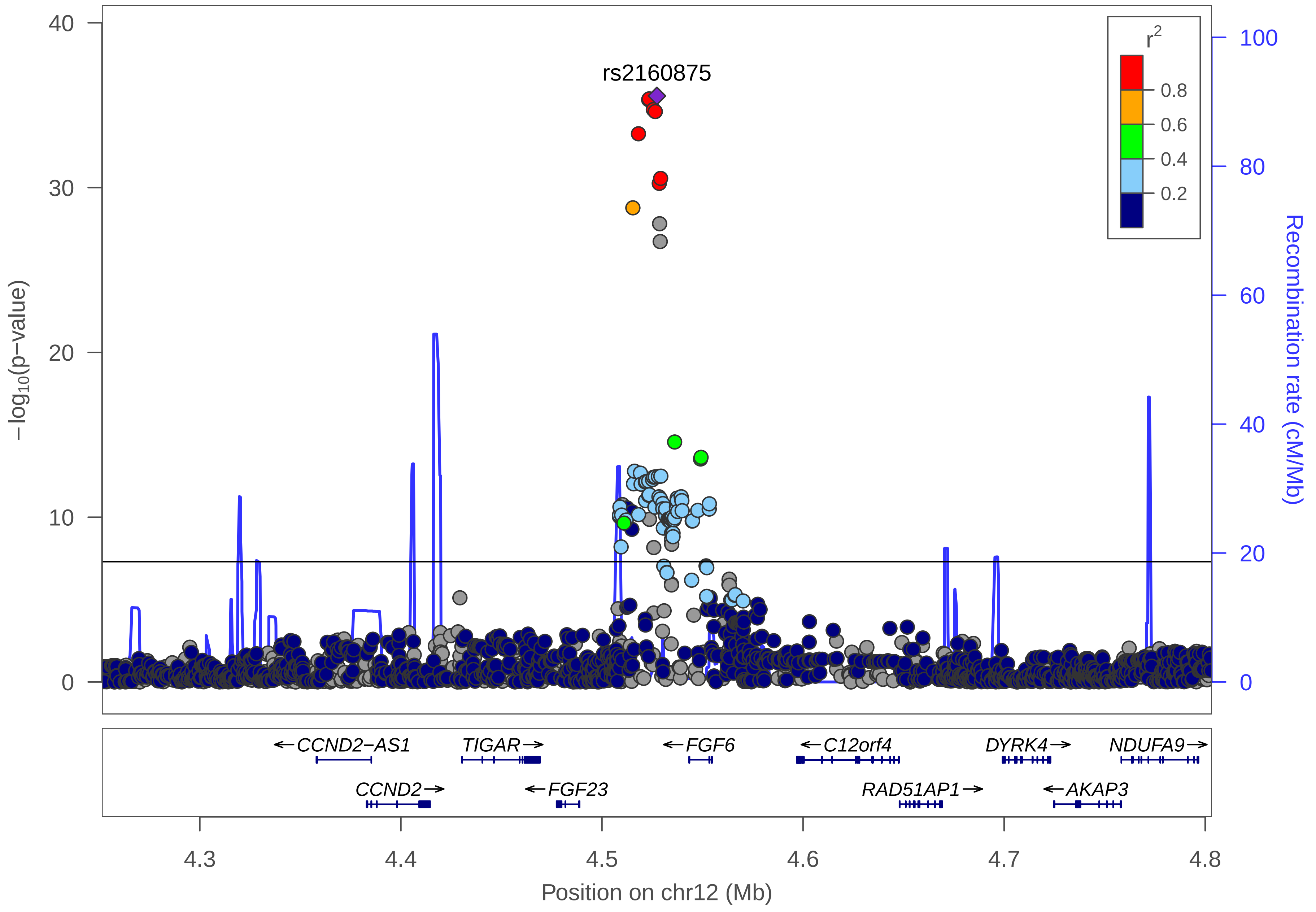

rs2160875
r2
0.8
0.6
0.4
0.2
−log10(p−value)
Recombination rate (cM/Mb)
CCND2−AS1
TIGAR
FGF6
C12orf4
DYRK4
NDUFA9
CCND2
FGF23
RAD51AP1
AKAP3
4.3
4.4
4.5
4.6
4.7
4.8
Position on chr12 (Mb)

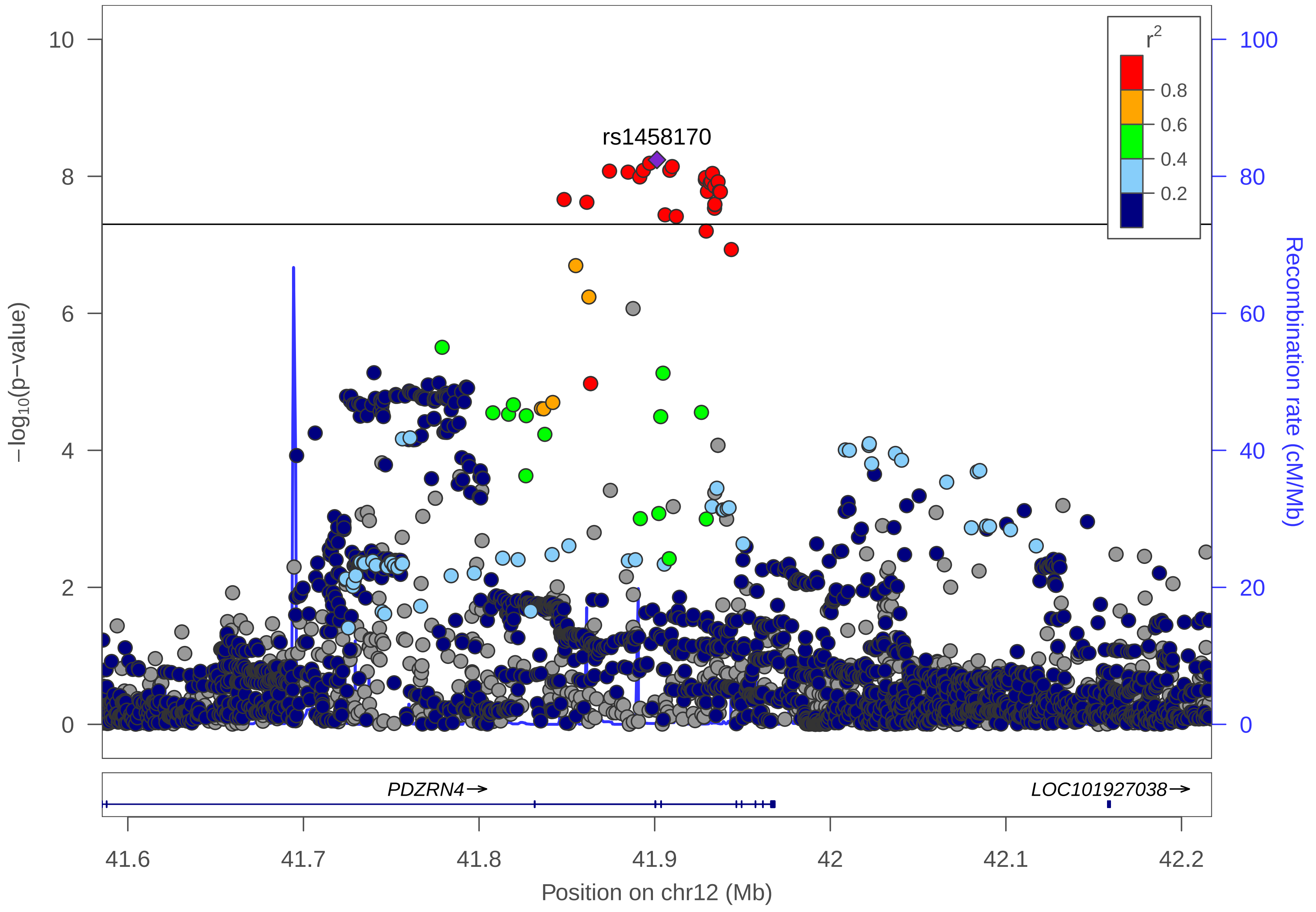

rs1458170
r2
0.8
0.6
0.4
0.2
10
8
6
4
2
0
100
80
60
40
20
0
−log10(p−value)
Recombination rate (cM/Mb)
PDZRN4→
LOC101927038→
41.6
41.7
41.8
41.9
42
42.1
42.2
Position on chr12 (Mb)

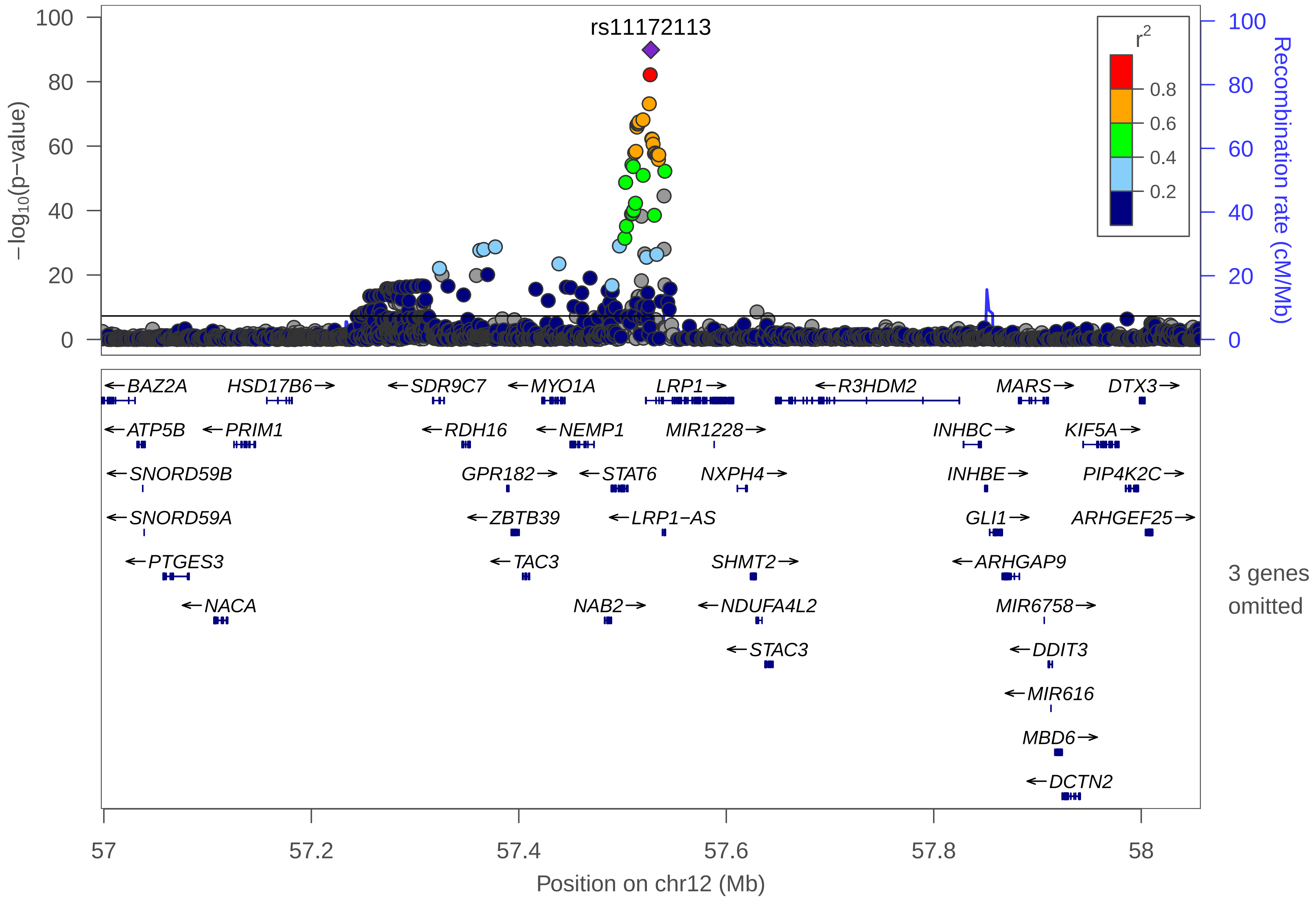

rs11172113
r2
0.8
0.6
0.4
0.2
−log10(p−value)
Recombination rate (cM/Mb)
100
80
60
40
20
0
BAZ2A
HSD17B6
SDR9C7
MYO1A
LRP1
R3HDM2
MARS
DTX3
ATP5B
PRIM1
RDH16
NEMP1
MIR1228
INHBC
KIF5A
SNORD59B
GPR182
STAT6
NXPH4
INHBE
PIP4K2C
SNORD59A
ZBTB39
LRP1−AS
GLI1
ARHGEF25
PTGES3
TAC3
SHMT2
ARHGAP9
NACA
NAB2
NDUFA4L2
MIR6758
STAC3
DDIT3
MIR616
MBD6
DCTN2
3 genes omitted
57
57.2
57.4
57.6
57.8
58
Position on chr12 (Mb)

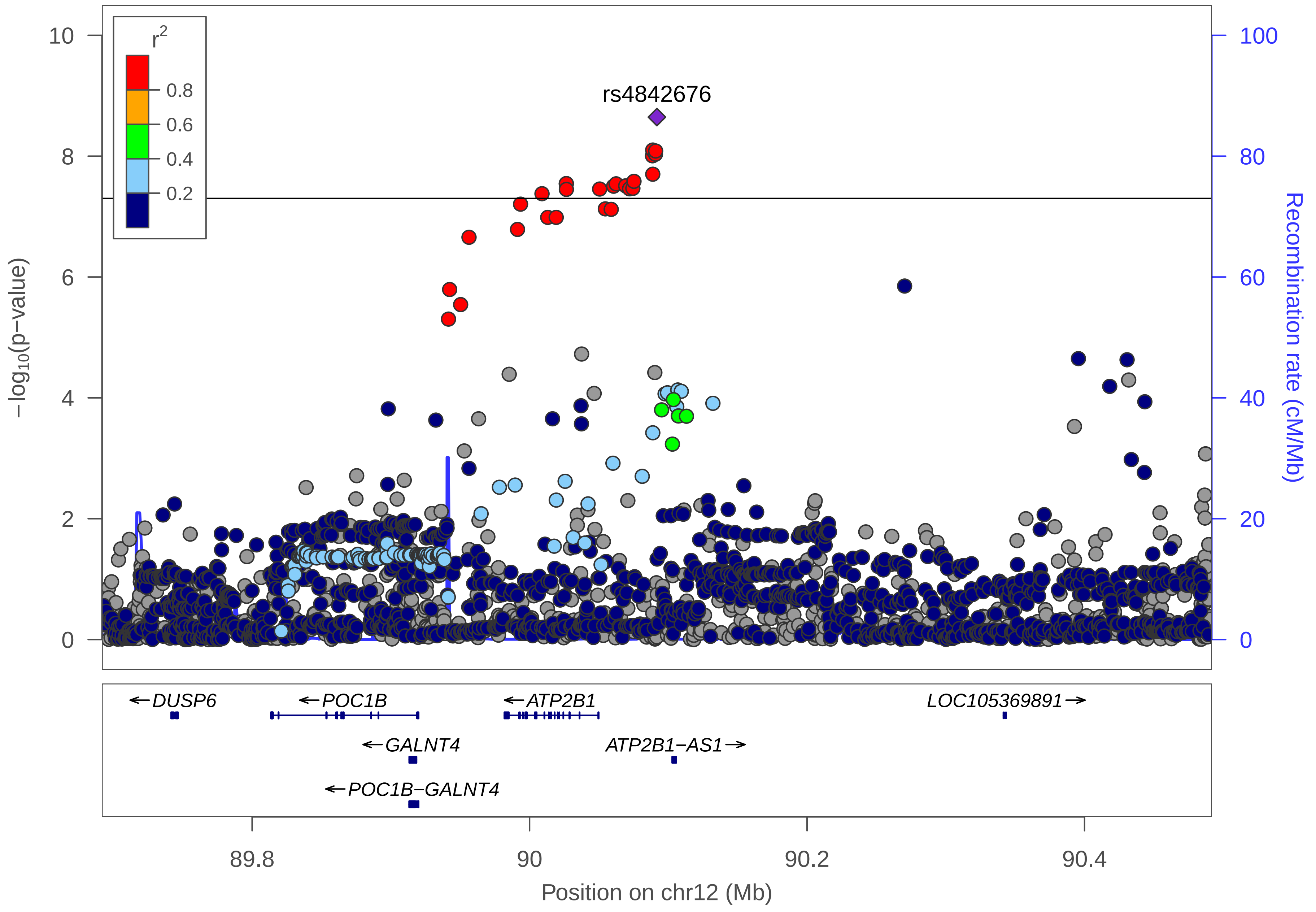

r2
0.8
0.6
0.4
0.2
rs4842676
10
8
6
4
2
0
100
80
60
40
20
0
−log10(p−value)
Recombination rate (cM/Mb)
←DUSP6
←POC1B
←ATP2B1
LOC105369891→
←GALNT4
ATP2B1−AS1→
←POC1B−GALNT4
89.8
90
90.2
90.4
Position on chr12 (Mb)

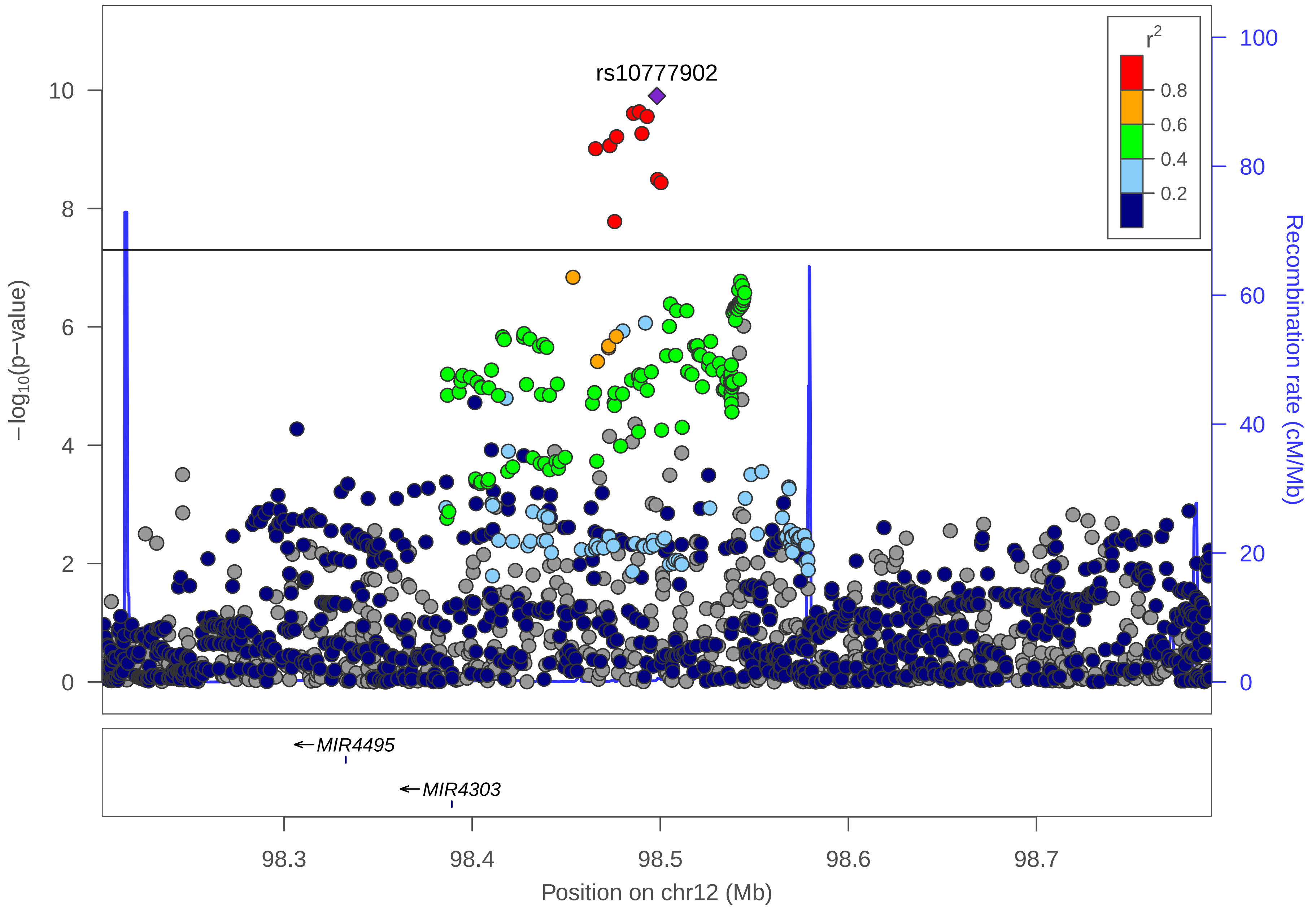

rs10777902
r2
0.8
0.6
0.4
0.2
−log10(p−value)
Recombination rate (cM/Mb)
10
8
6
4
2
0
100
80
60
40
20
0
MIR4495
MIR4303
98.3
98.4
98.5
98.6
98.7
Position on chr12 (Mb)

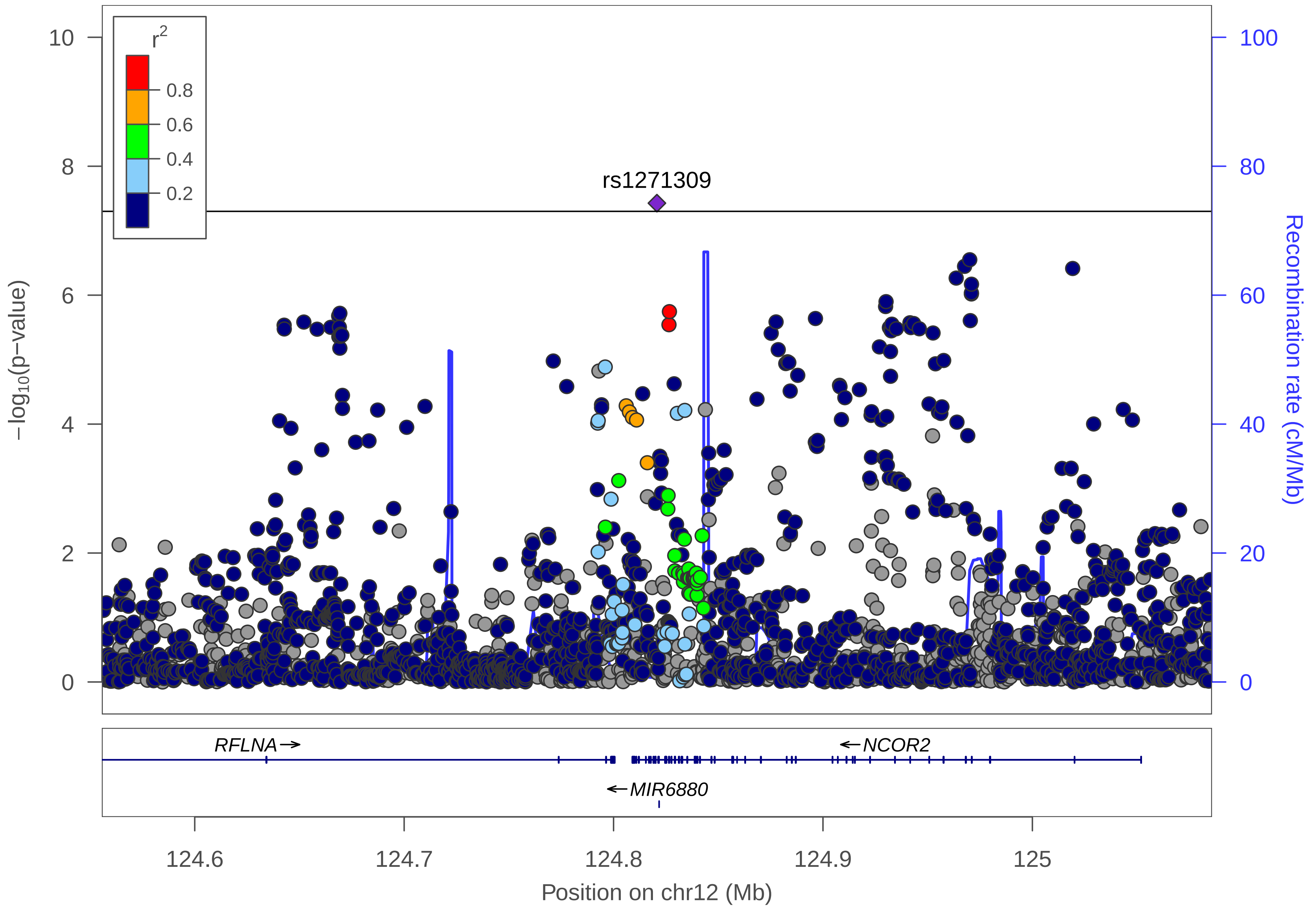

r2
0.8
0.6
0.4
0.2
10
8
6
4
2
0
−log10(p−value)
rs1271309
100
80
60
40
20
0
Recombination rate (cM/Mb)
RFLNA→
←NCOR2
←MIR6880
124.6
124.7
124.8
124.9
125
Position on chr12 (Mb)

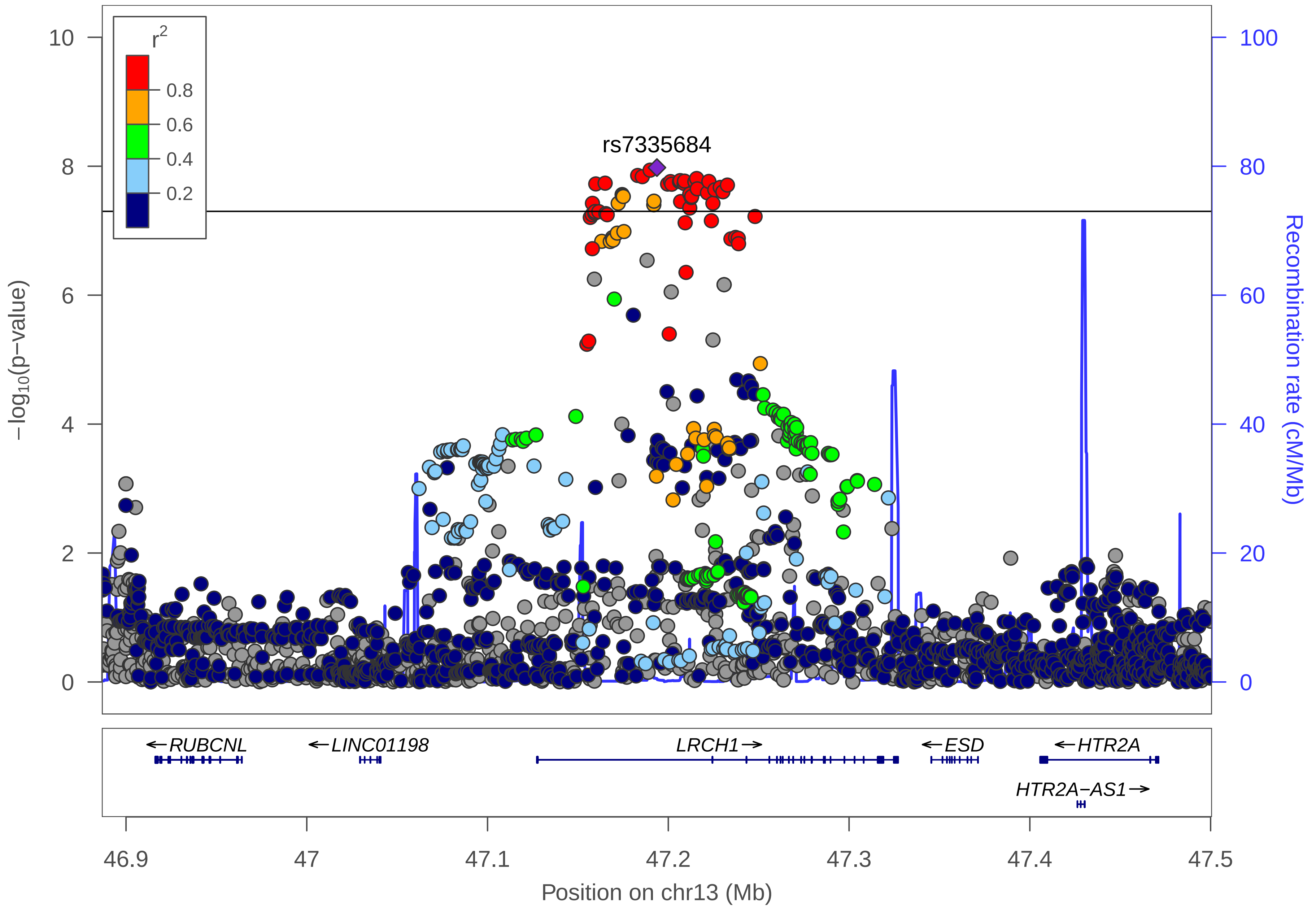

r2
0.8
0.6
0.4
0.2
rs7335684
10
8
6
4
2
0
−log10(p−value)
100
80
60
40
20
0
Recombination rate (cM/Mb)
←RUBCNL
←LINC01198
LRCH1→
←ESD
←HTR2A
HTR2A−AS1→
46.9
47
47.1
47.2
47.3
47.4
47.5
Position on chr13 (Mb)

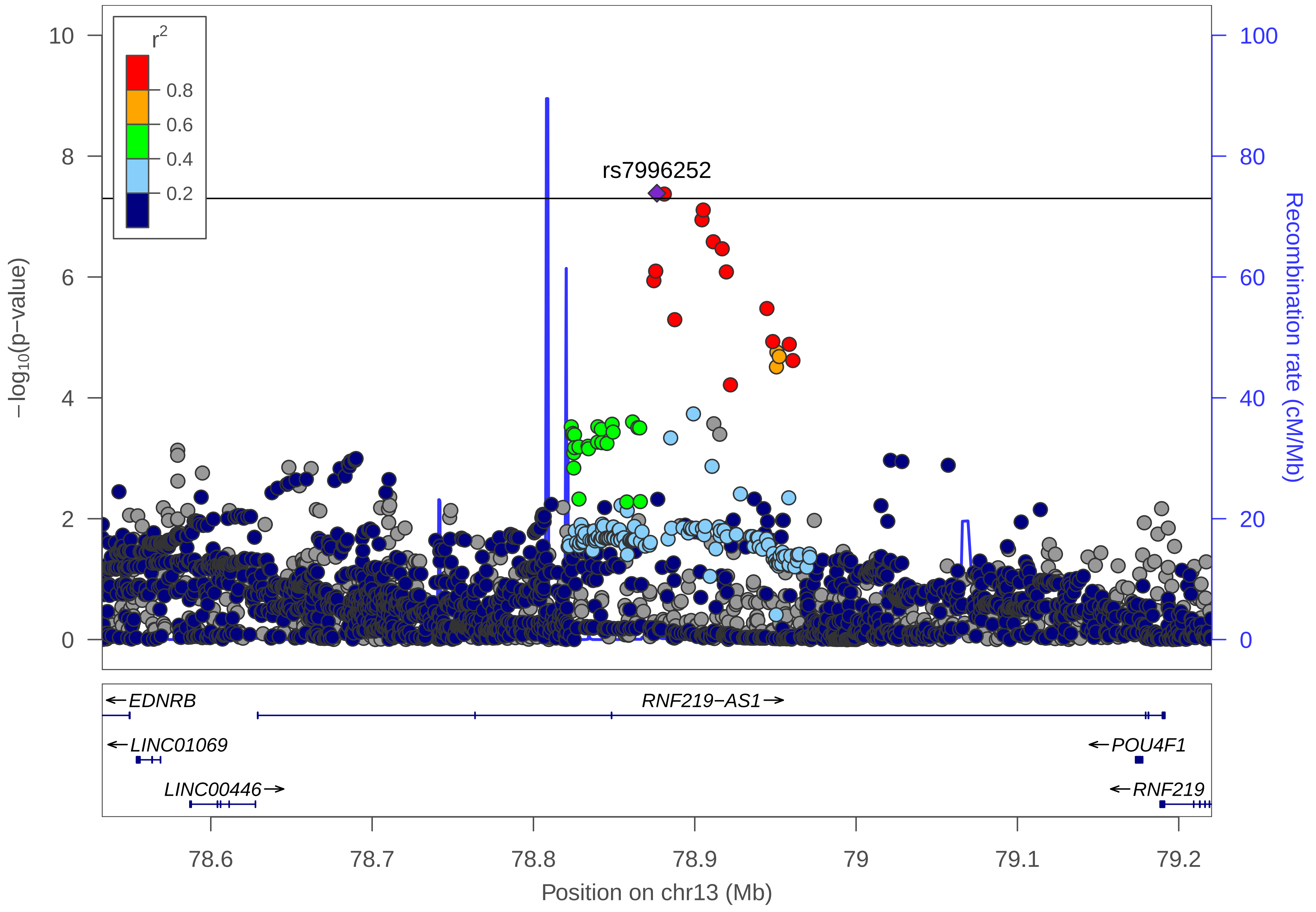
r2
0.8
0.6
0.4
0.2
rs7996252
−log10(p−value)
Recombination rate (cM/Mb)
10
8
6
4
2
0
100
80
60
40
20
0
EDNRB
RNF219−AS1
LINC01069
POU4F1
LINC00446
RNF219
78.6
78.7
78.8
78.9
79
79.1
79.2
Position on chr13 (Mb)

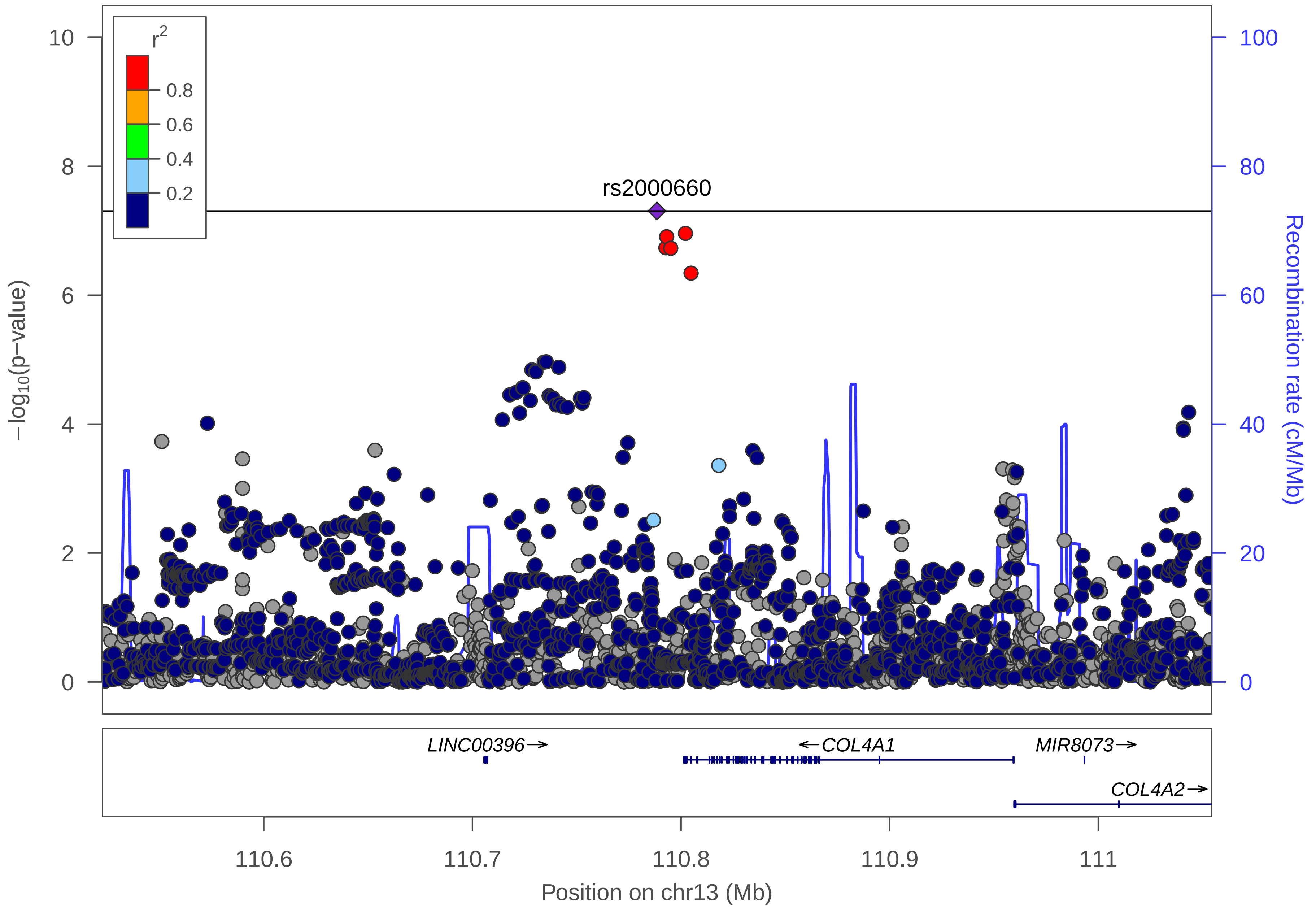

r2
0.8
0.6
0.4
0.2
rs2000660
−log10(p−value)
Recombination rate (cM/Mb)
10
8
6
4
2
0
100
80
60
40
20
0
LINC00396→
←COL4A1
MIR8073→
COL4A2→
110.6
110.7
110.8
110.9
111
Position on chr13 (Mb)

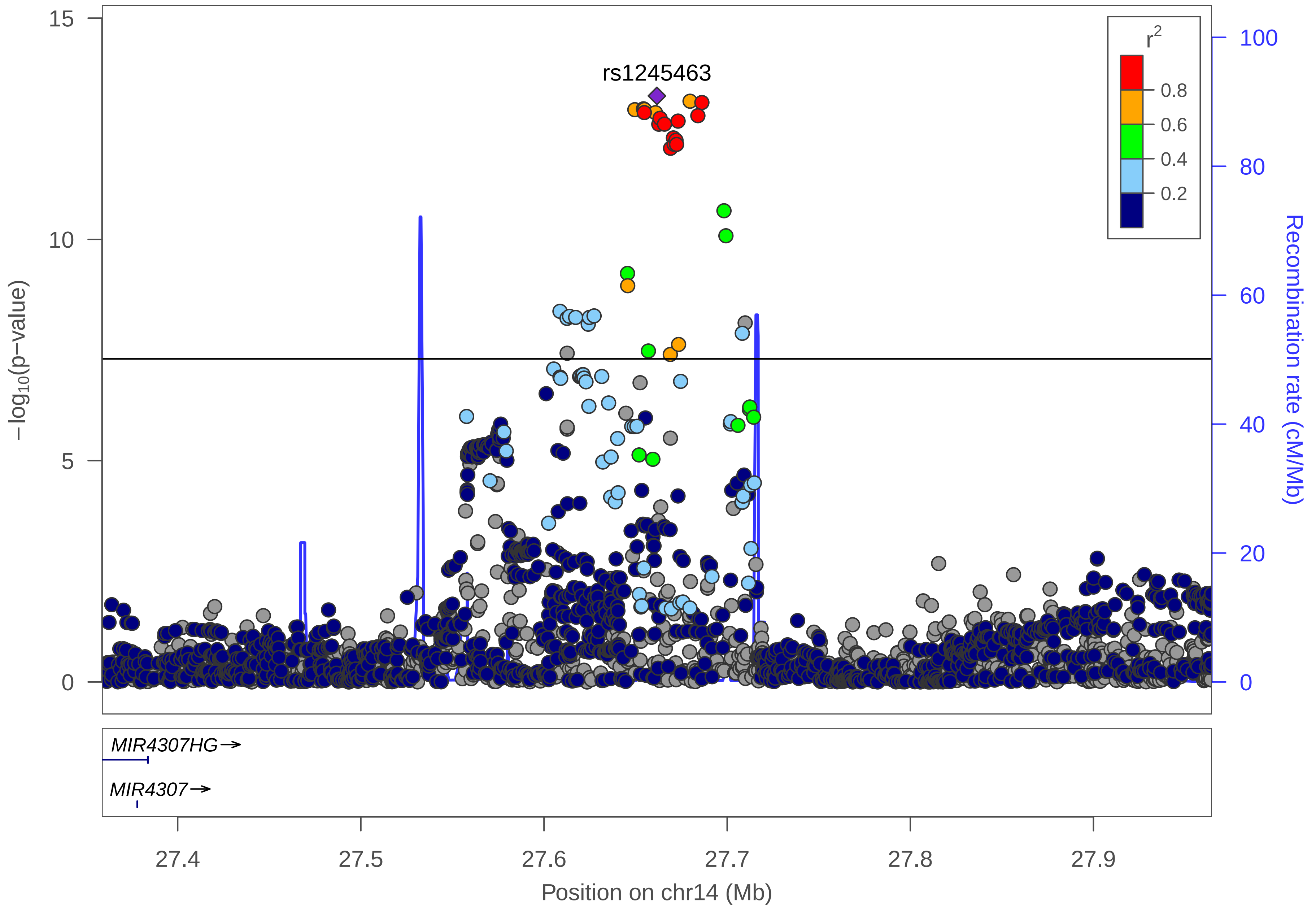
rs1245463
r2
0.8
0.6
0.4
0.2
15
10
5
0
−log10(p−value)
100
80
60
40
20
0
Recombination rate (cM/Mb)
MIR4307HG→
MIR4307→
27.4
27.5
27.6
27.7
27.8
27.9
Position on chr14 (Mb)

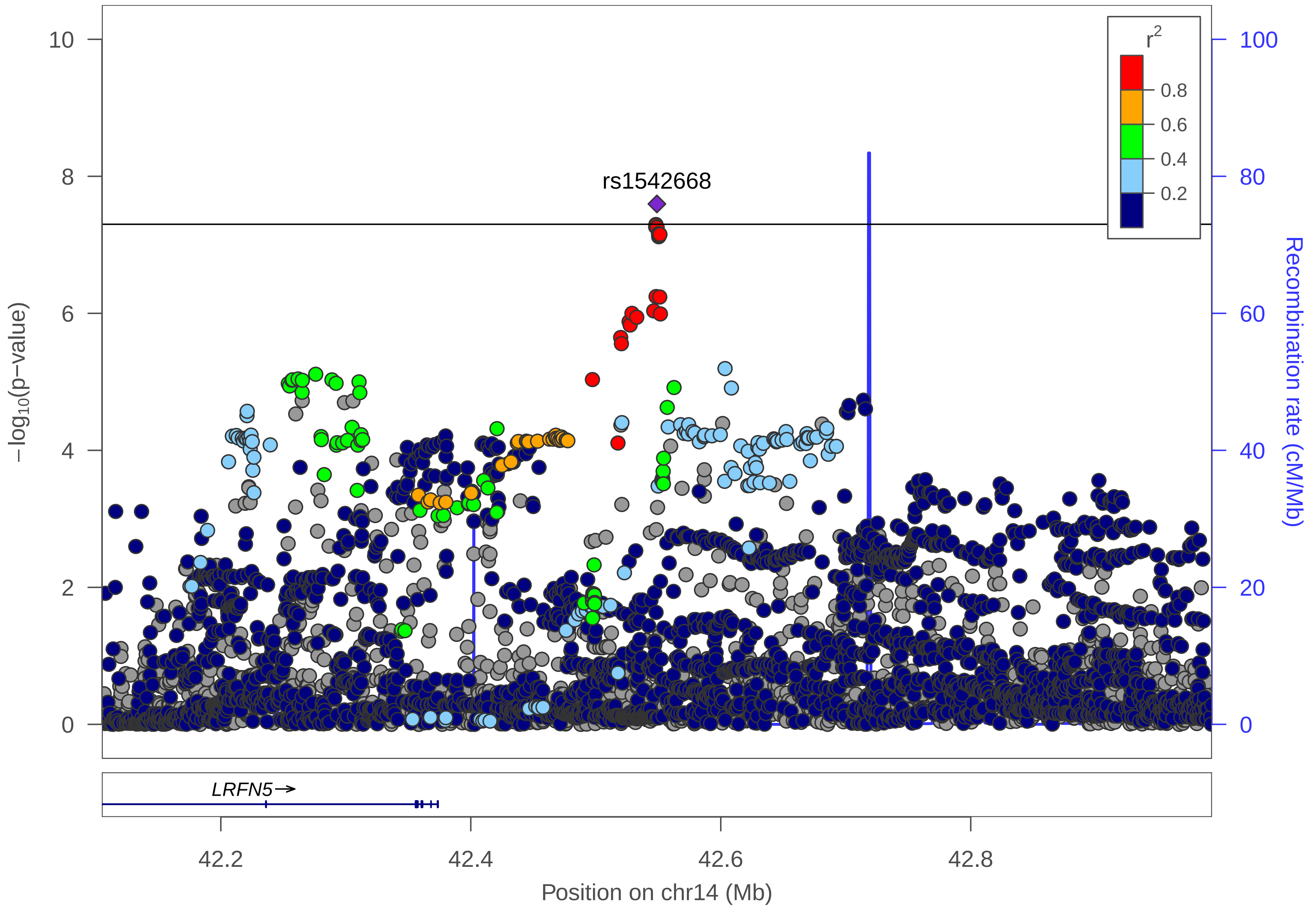

r2
0.8
0.6
0.4
0.2
rs1542668
−log10(p−value)
Recombination rate (cM/Mb)
LRFN5→
Position on chr14 (Mb)
42.2
42.4
42.6
42.8
0
2
4
6
8
10
20
40
60
80
100

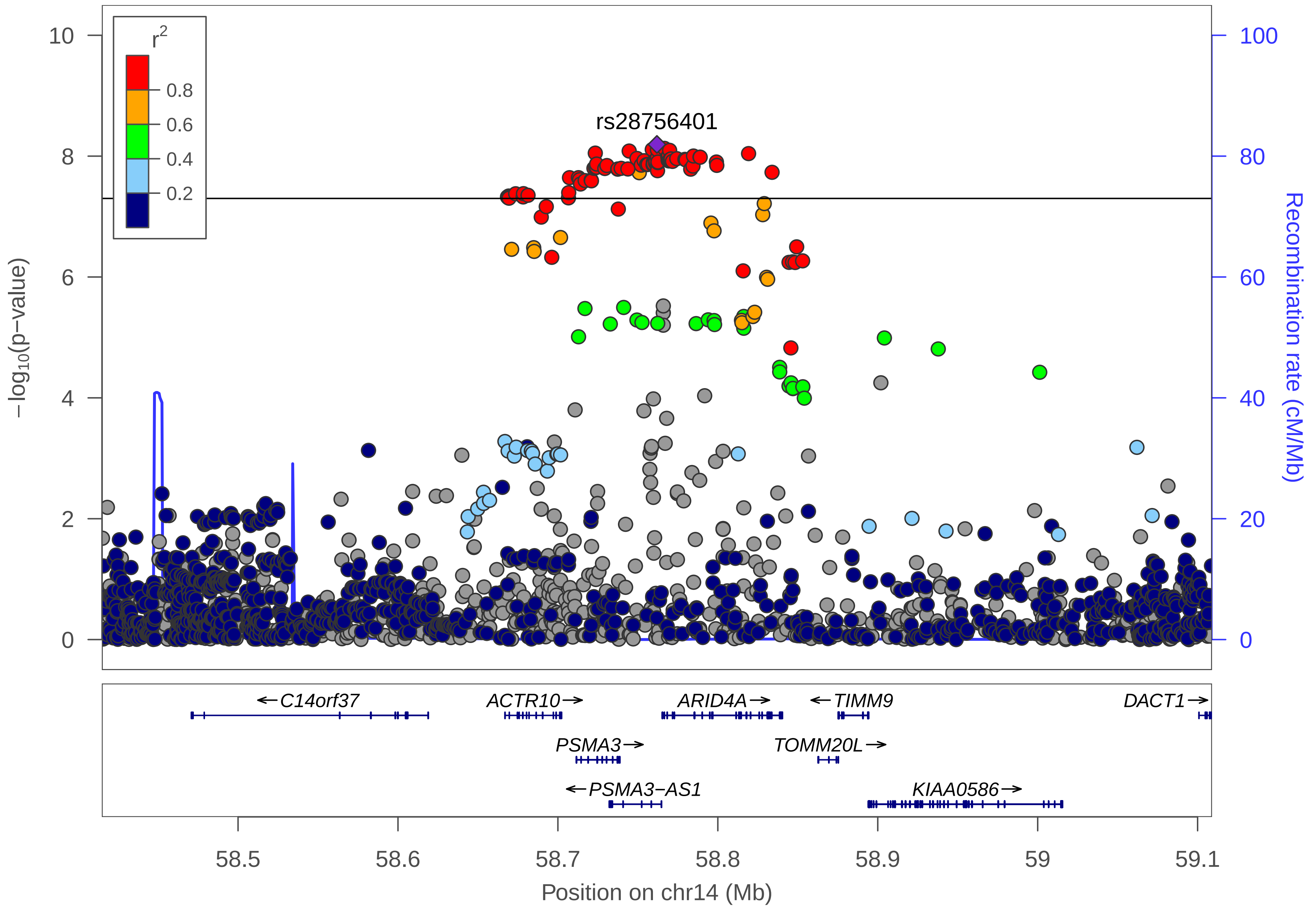

r2
0.8
0.6
0.4
0.2
rs28756401
10
8
6
4
2
0
−log10(p−value)
100
80
60
40
20
0
Recombination rate (cM/Mb)
←C14orf37
ACTR10→
ARID4A→
←TIMM9
DACT1→
PSMA3→
TOMM20L→
←PSMA3−AS1
KIAA0586→
58.5
58.6
58.7
58.8
58.9
59
59.1
Position on chr14 (Mb)

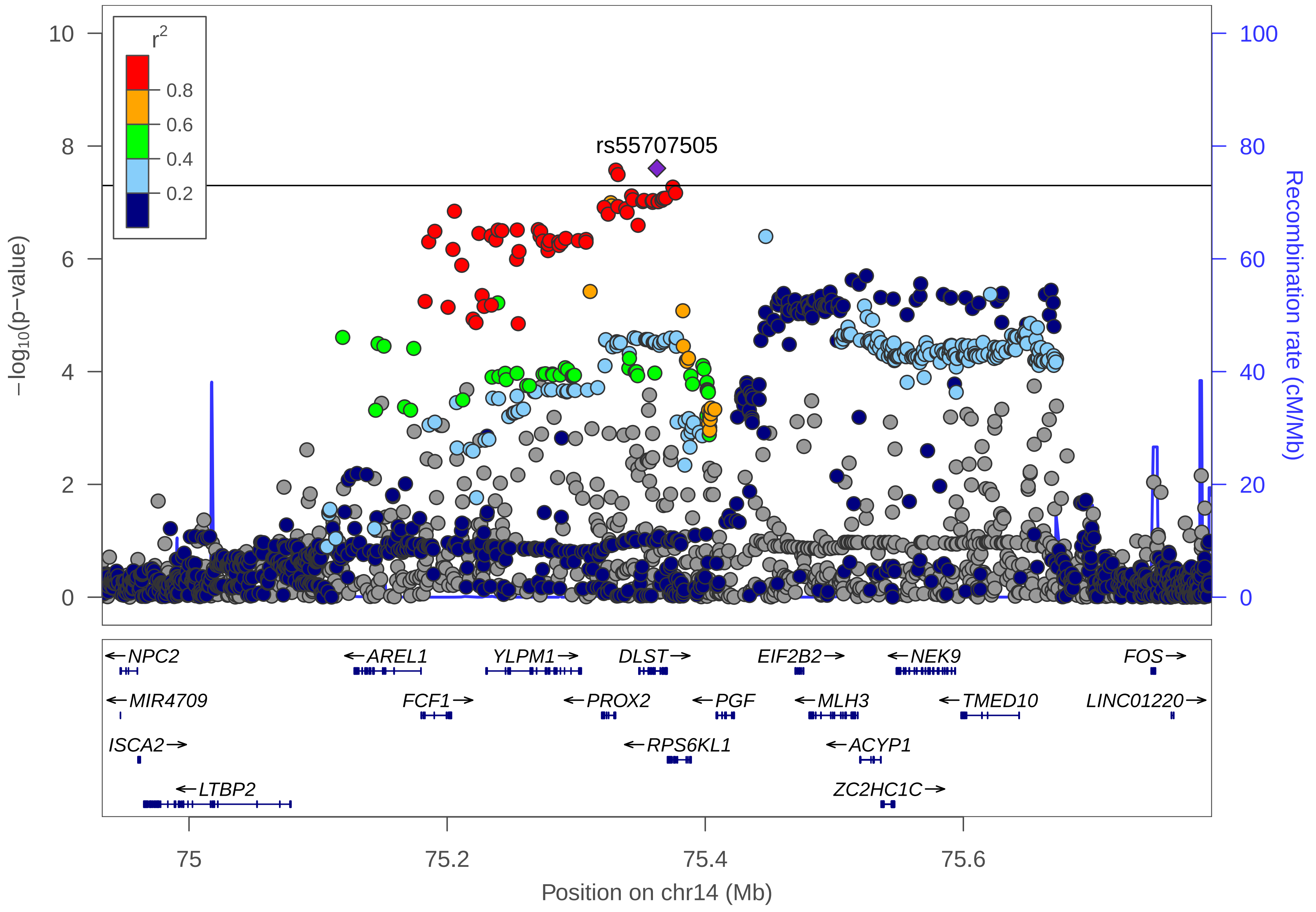

r2
0.8
0.6
0.4
0.2
rs55707505
−log10(p−value)
Recombination rate (cM/Mb)
0
2
4
6
8
10
0
20
40
60
80
100
NPC2
MIR4709
ISCA2
LTBP2
AREL1
FCF1
YLPM1
PROX2
DLST
RPS6KL1
PGF
EIF2B2
MLH3
ACYP1
ZC2HC1C
NEK9
TMED10
FOS
LINC01220
75
75.2
75.4
75.6
Position on chr14 (Mb)

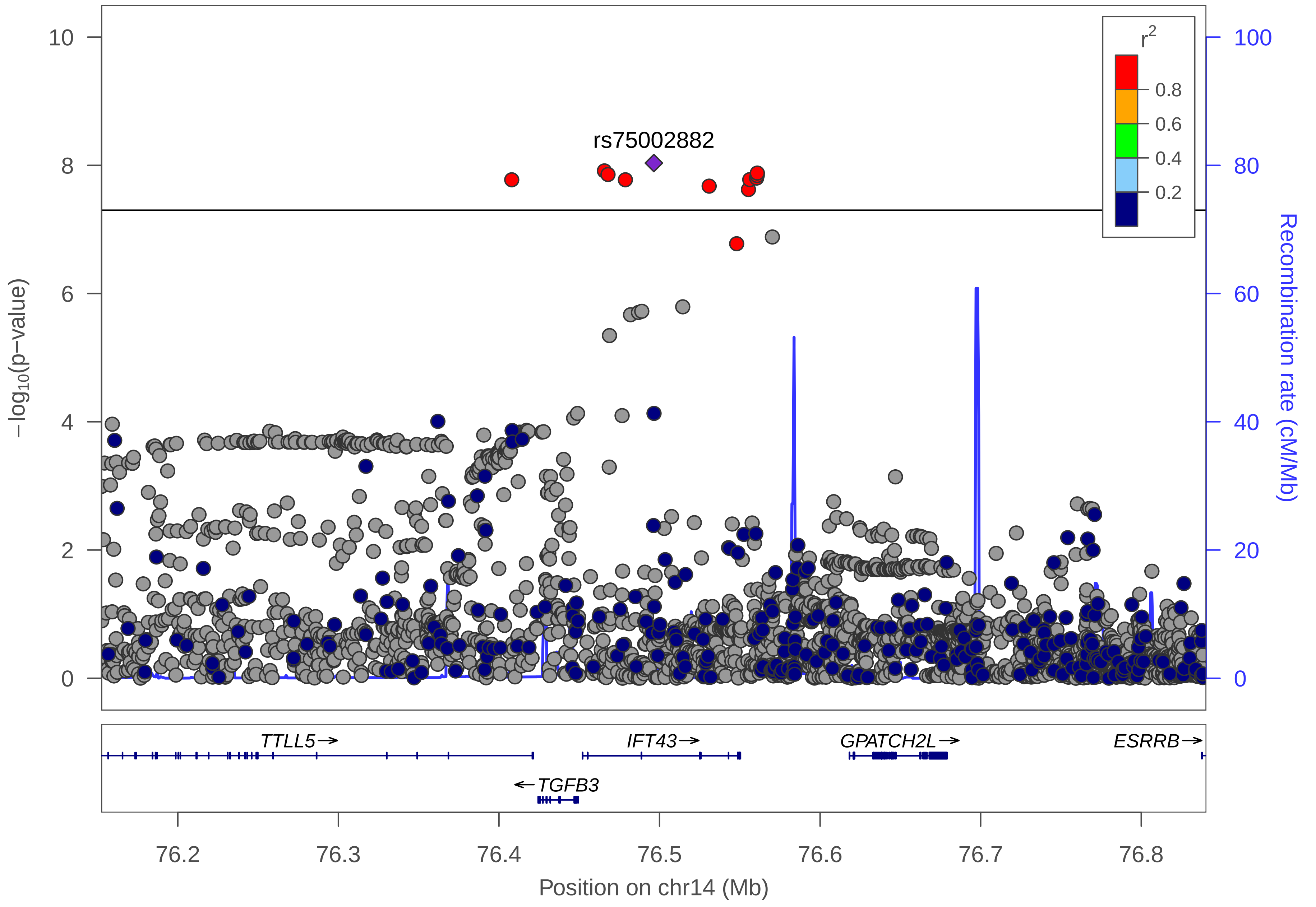

rs75002882
r2
0.8
0.6
0.4
0.2
−log10(p−value)
Recombination rate (cM/Mb)
TTLL5→
IFT43→
GPATCH2L→
ESRRB→
←TGFB3
Position on chr14 (Mb)
76.2
76.3
76.4
76.5
76.6
76.7
76.8
0
2
4
6
8
10
20
40
60
80
100

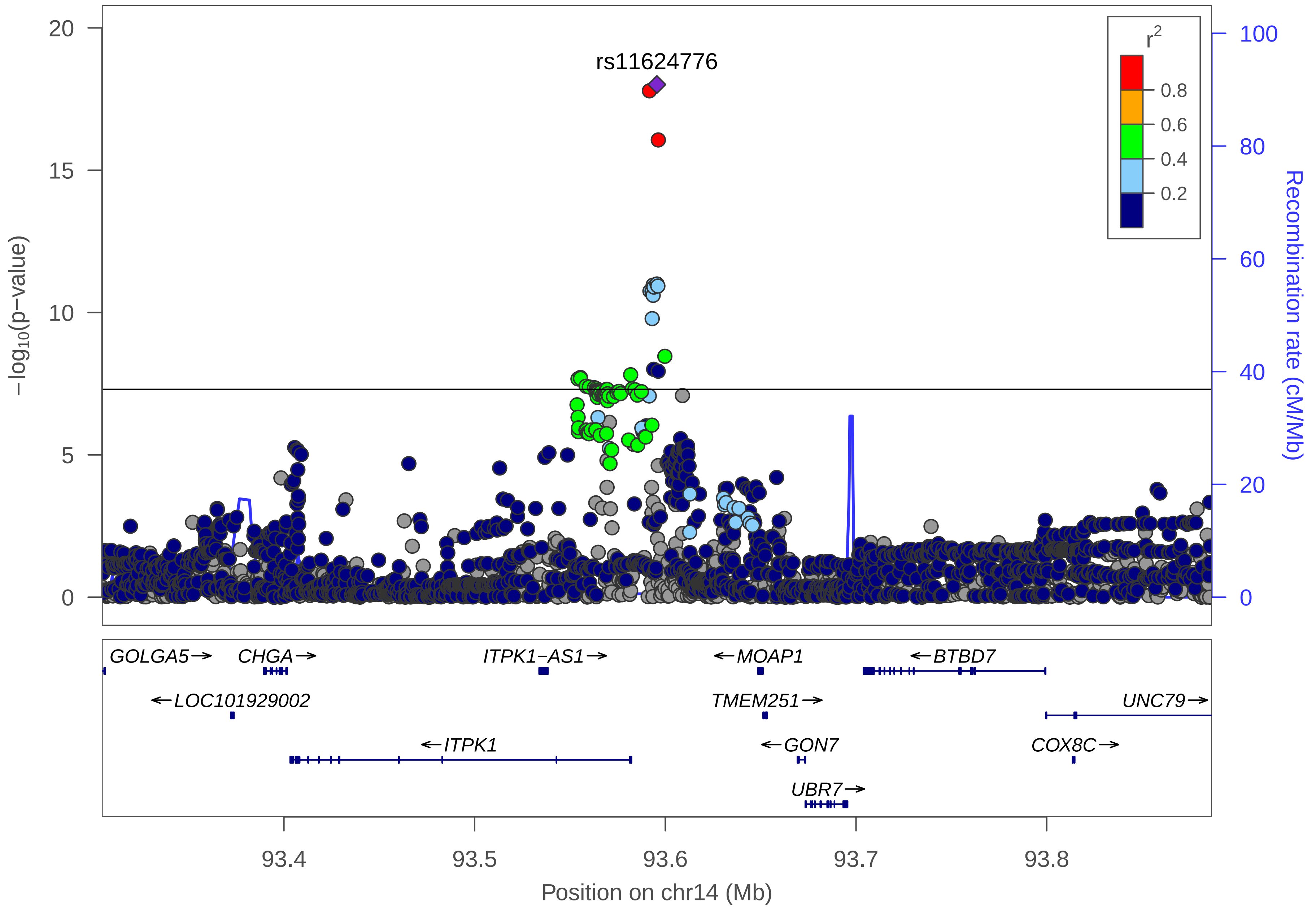

rs11624776
r^2
0.8
0.6
0.4
0.2
20
15
10
5
0
−log10(p−value)
100
80
60
40
20
0
Recombination rate (cM/Mb)
GOLGA5→
CHGA→
ITPK1−AS1→
←MOAP1
←BTBD7
←LOC101929002
TMEM251→
UNC79→
←ITPK1
←GON7
COX8C→
UBR7→
93.4
93.5
93.6
93.7
93.8
Position on chr14 (Mb)

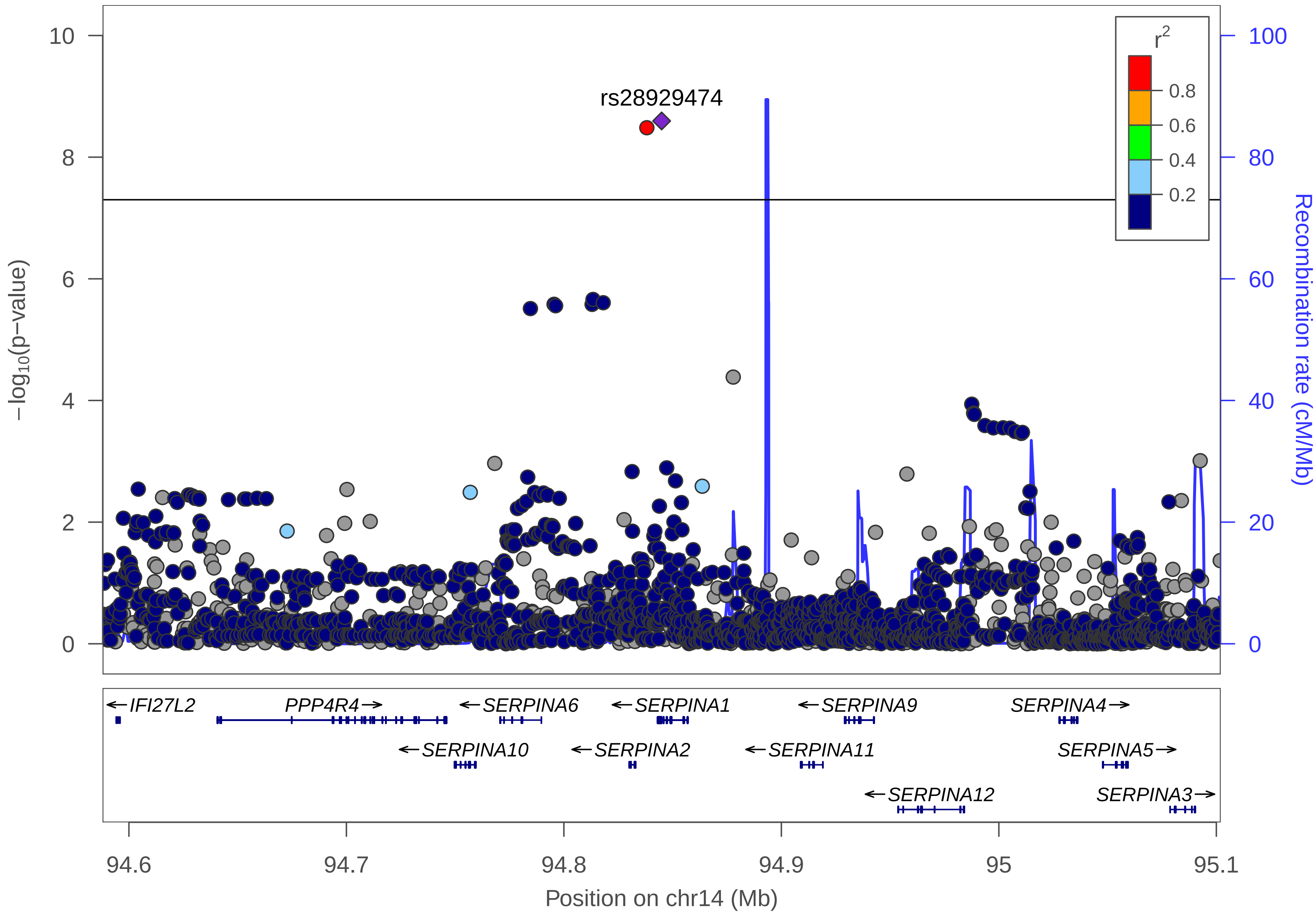

rs28929474
r2
0.8
0.6
0.4
0.2
10
8
6
4
2
0
100
80
60
40
20
0
−log10(p−value)
Recombination rate (cM/Mb)
IFI27L2
PPP4R4
SERPINA6
SERPINA1
SERPINA9
SERPINA4
SERPINA10
SERPINA2
SERPINA11
SERPINA5
SERPINA12
SERPINA3
94.6
94.7
94.8
94.9
95
95.1
Position on chr14 (Mb)

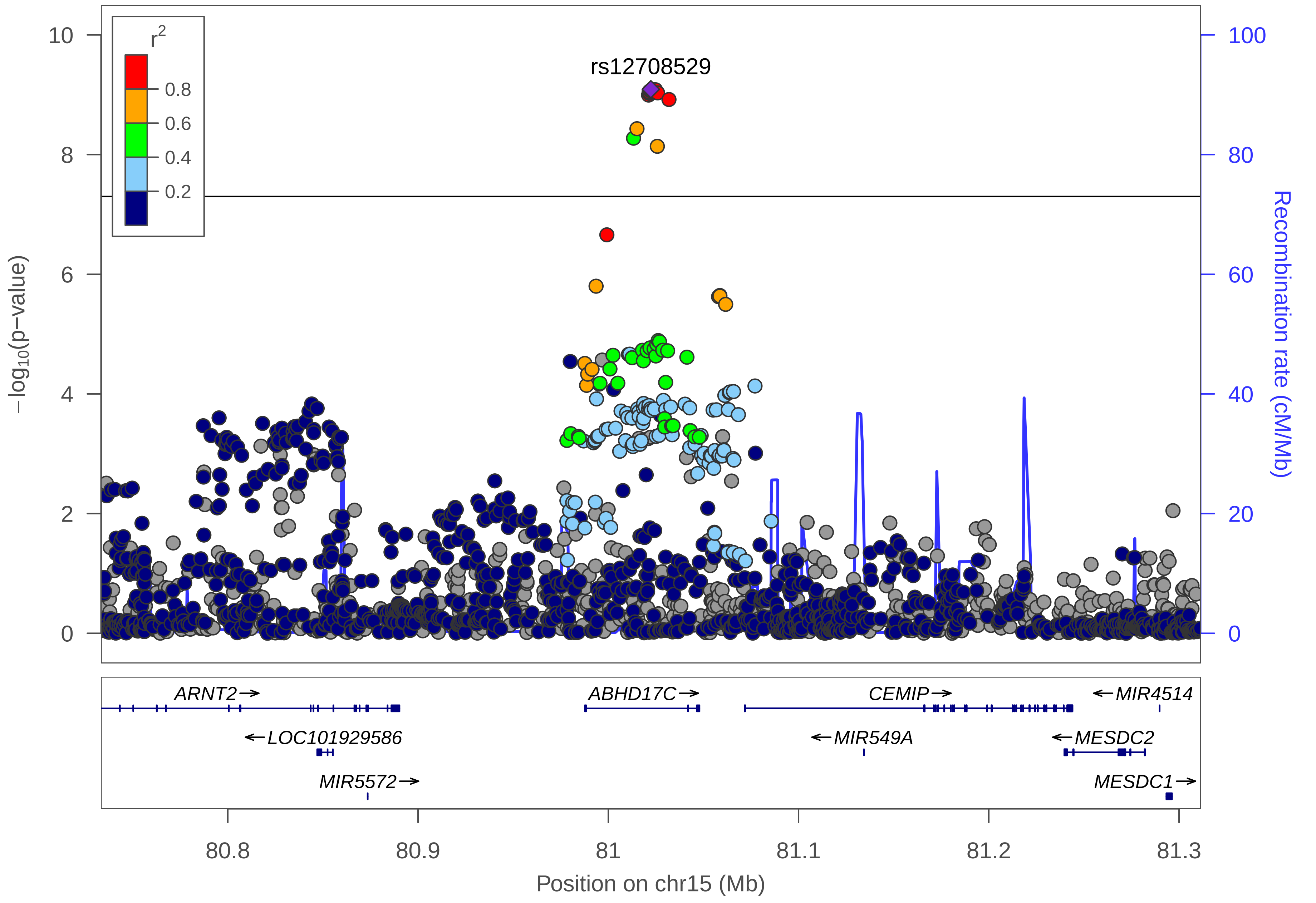

r2
0.8
0.6
0.4
0.2
rs12708529
10
8
6
4
2
0
−log10(p−value)
100
80
60
40
20
0
Recombination rate (cM/Mb)
ARNT2→
ABHD17C→
CEMIP→
←MIR4514
←LOC101929586
←MIR549A
←MESDC2
MIR5572→
MESDC1→
80.8
80.9
81
81.1
81.2
81.3
Position on chr15 (Mb)

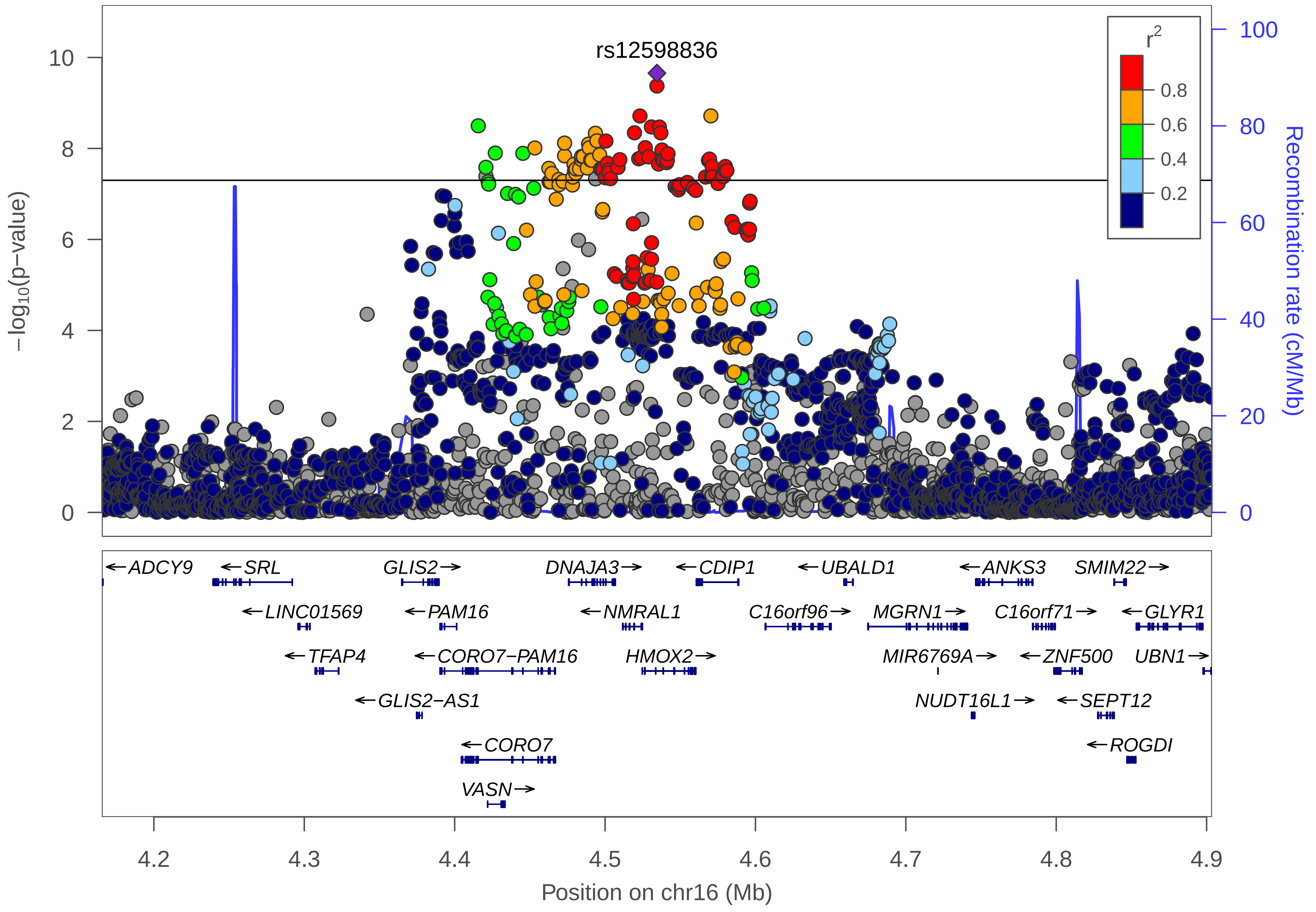
rs12598836
r2
0.8
0.6
0.4
0.2
−log10(p−value)
Recombination rate (cM/Mb)
ADCY9
SRL
GLIS2
DNAJA3
CDIP1
UBALD1
ANKS3
SMIM22
LINC01569
PAM16
NMRAL1
C16orf96
MGRN1
C16orf71
GLYR1
TFAP4
CORO7−PAM16
HMOX2
MIR6769A
ZNF500
UBN1
GLIS2−AS1
NUDT16L1
SEPT12
CORO7
ROGDI
VASN
Position on chr16 (Mb)

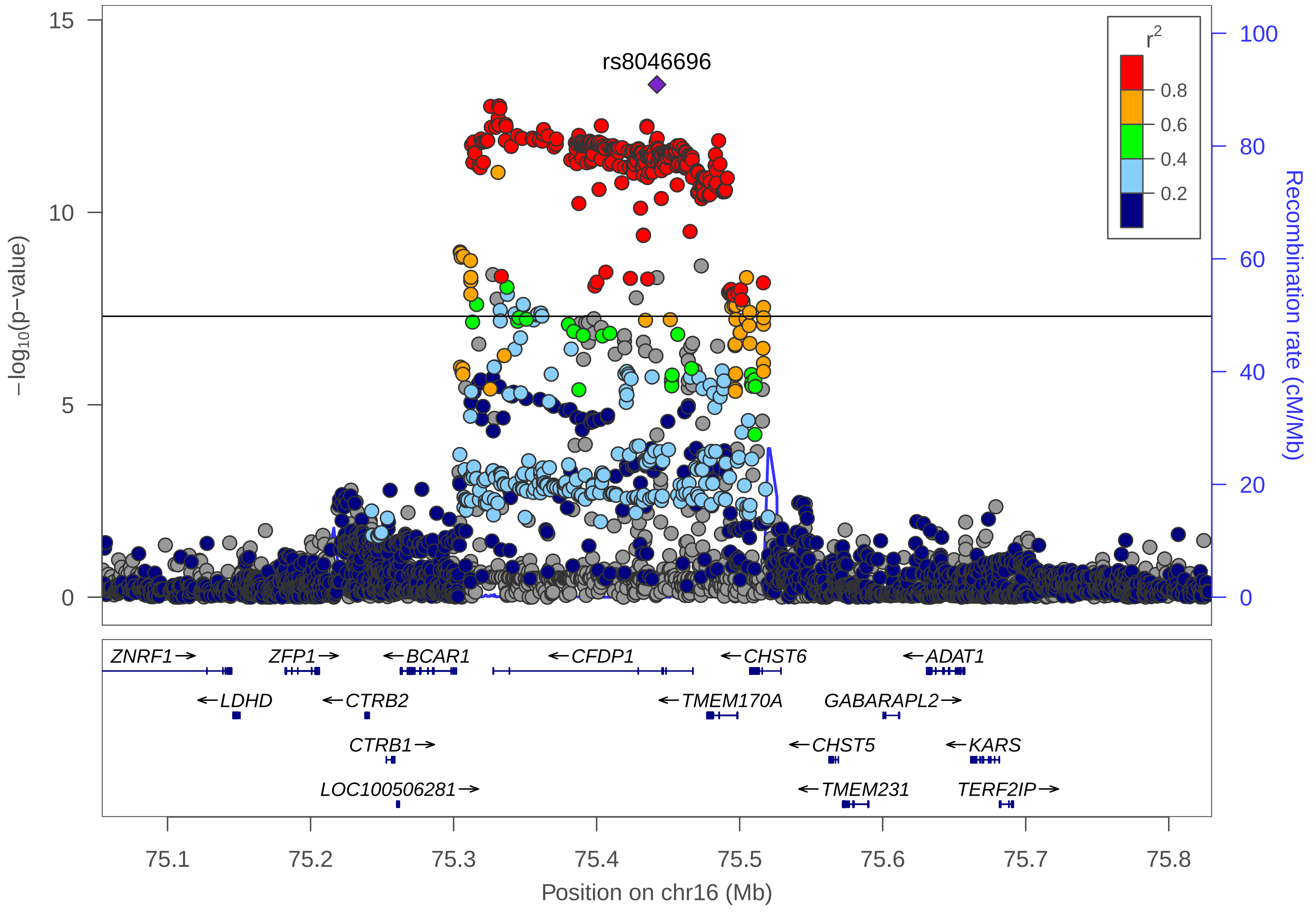
rs8046696
r2
0.8
0.6
0.4
0.2
15
10
5
0
−log10(p−value)
100
80
60
40
20
0
Recombination rate (cM/Mb)
ZNRF1→
ZFP1→
←BCAR1
←CFDP1
←CHST6
←ADAT1
←LDHD
←CTRB2
←TMEM170A
GABARAPL2→
CTRB1→
←CHST5
←KARS
LOC100506281→
←TMEM231
TERF2IP→
75.1
75.2
75.3
75.4
75.5
75.6
75.7
75.8
Position on chr16 (Mb)

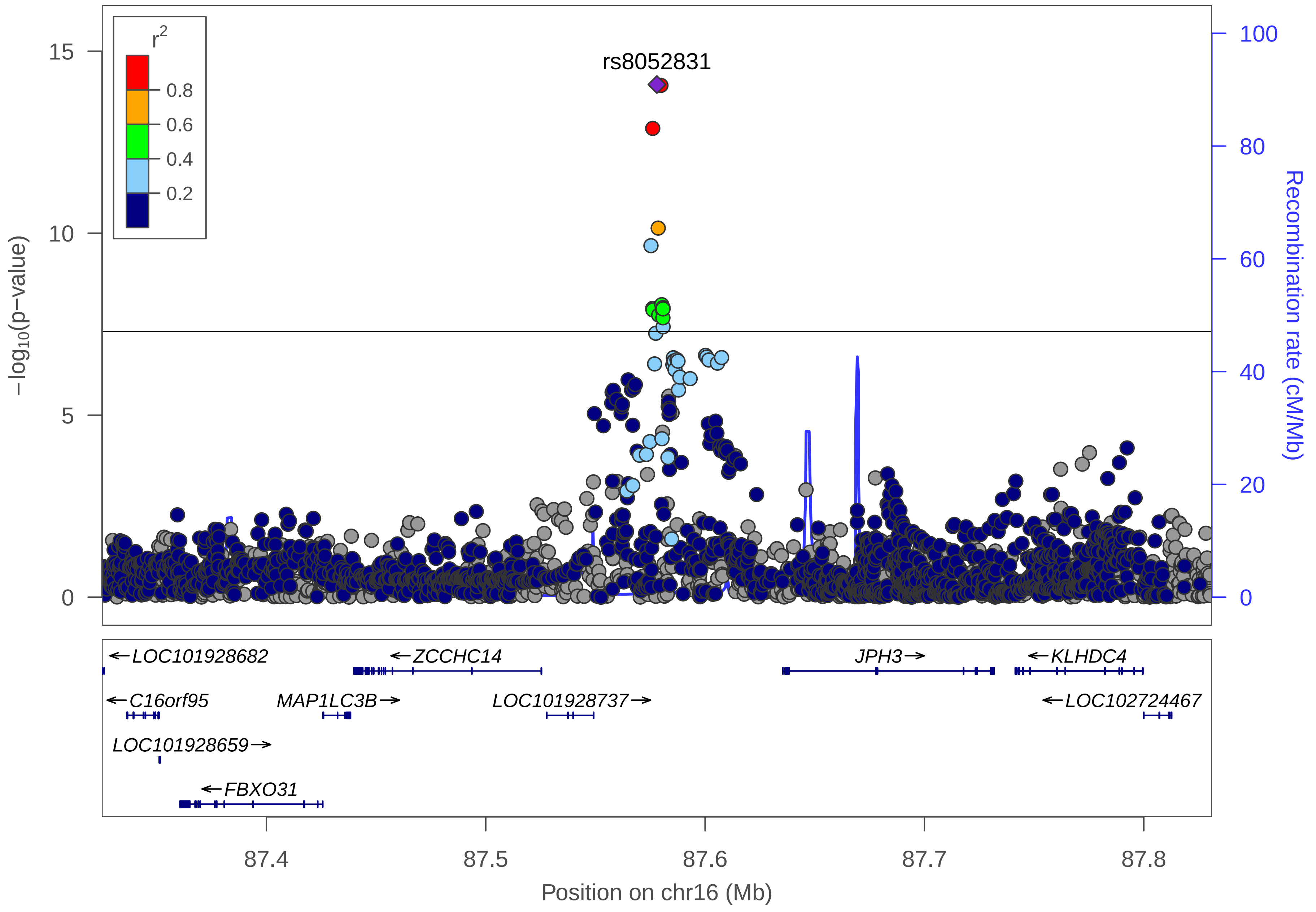

r2
0.8
0.6
0.4
0.2
rs8052831
15
10
5
0
−log10(p−value)
100
80
60
40
20
0
Recombination rate (cM/Mb)
←LOC101928682
←ZCCHC14
JPH3→
←KLHDC4
←C16orf95
MAP1LC3B→
LOC101928737→
←LOC102724467
LOC101928659→
←FBXO31
87.4
87.5
87.6
87.7
87.8
Position on chr16 (Mb)

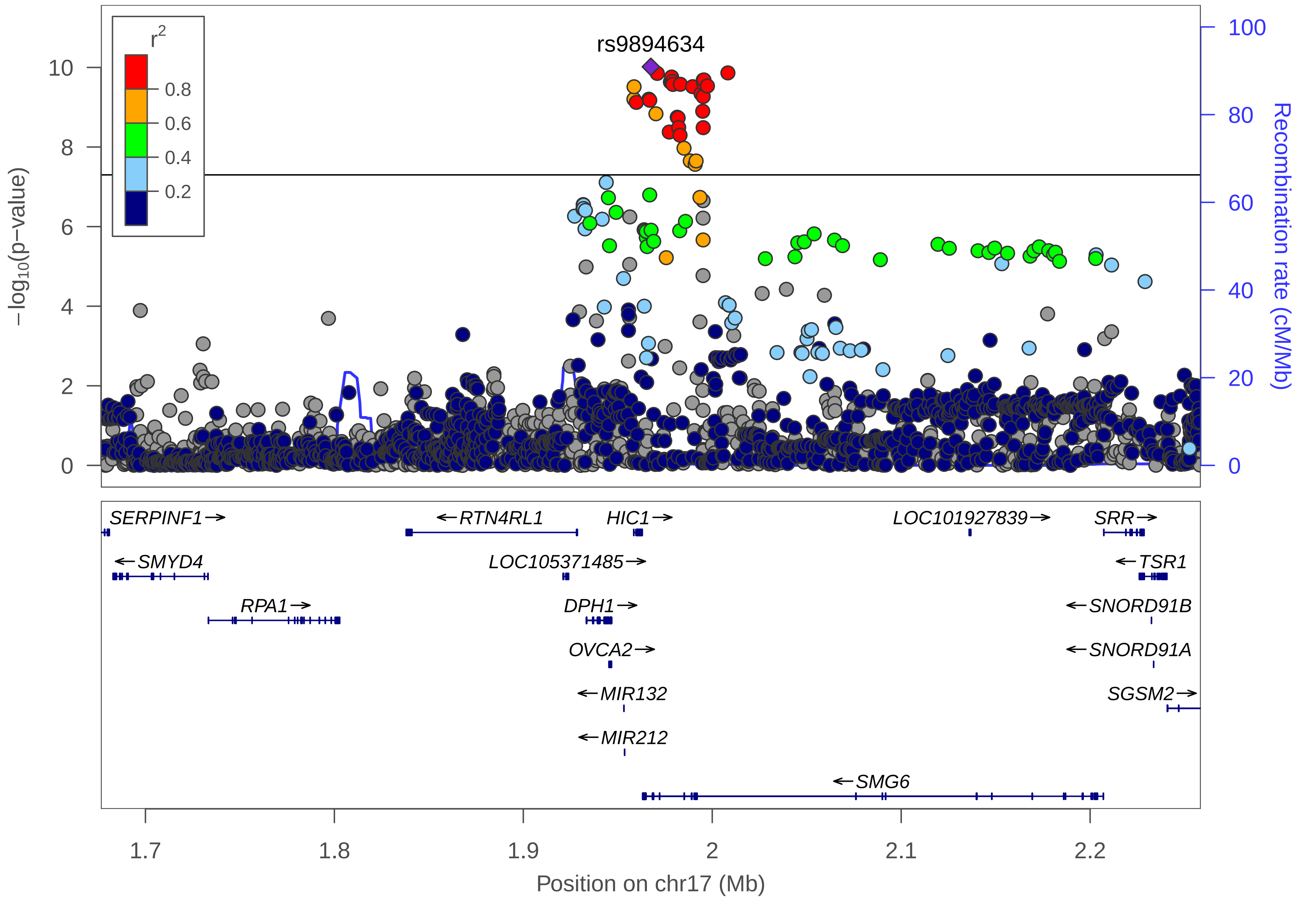

r2
0.8
0.6
0.4
0.2
rs9894634
10
8
6
4
2
0
$-\log_{10}$(p−value)
100
80
60
40
20
0
Recombination rate (cM/Mb)
SERPINF1→
←RTN4RL1
HIC1→
LOC101927839→
SRR→
←SMYD4
LOC105371485→
←TSR1
RPA1→
DPH1→
←SNORD91B
OVCA2→
←SNORD91A
←MIR132
SGSM2→
←MIR212
←SMG6
1.7
1.8
1.9
2
2.1
2.2
Position on chr17 (Mb)

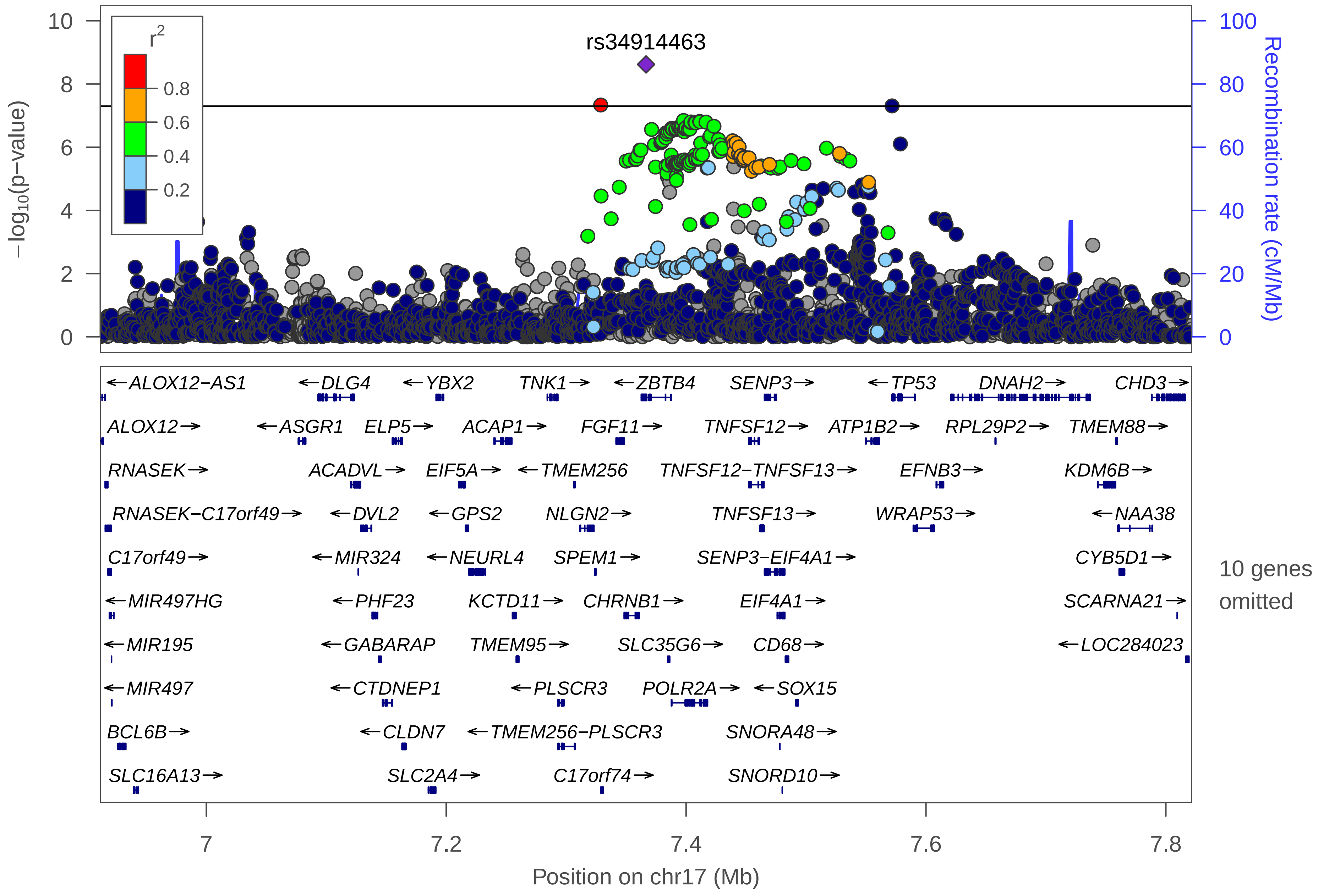

rs34914463
r2
0.8
0.6
0.4
0.2
−log10(p−value)
Recombination rate (cM/Mb)
10
8
6
4
2
0
100
80
60
40
20
0
ALOX12−AS1
DLG4
YBX2
TNK1
ZBTB4
SENP3
TP53
DNAH2
CHD3
ALOX12
ASGR1
ELP5
ACAP1
FGF11
TNFSF12
ATP1B2
RPL29P2
TMEM88
RNASEK
ACADVL
EIF5A
TMEM256
TNFSF12−TNFSF13
EFNB3
KDM6B
RNASEK−C17orf49
DVL2
GPS2
NLGN2
TNFSF13
WRAP53
NAA38
C17orf49
MIR324
NEURL4
SPEM1
SENP3−EIF4A1
CYB5D1
MIR497HG
PHF23
KCTD11
CHRNB1
EIF4A1
SCARNA21
MIR195
GABARAP
TMEM95
SLC35G6
CD68
LOC284023
MIR497
CTDNEP1
PLSCR3
POLR2A
SOX15
BCL6B
CLDN7
TMEM256−PLSCR3
SNORA48
SLC16A13
SLC2A4
C17orf74
SNORD10
10 genes omitted
7
7.2
7.4
7.6
7.8
Position on chr17 (Mb)

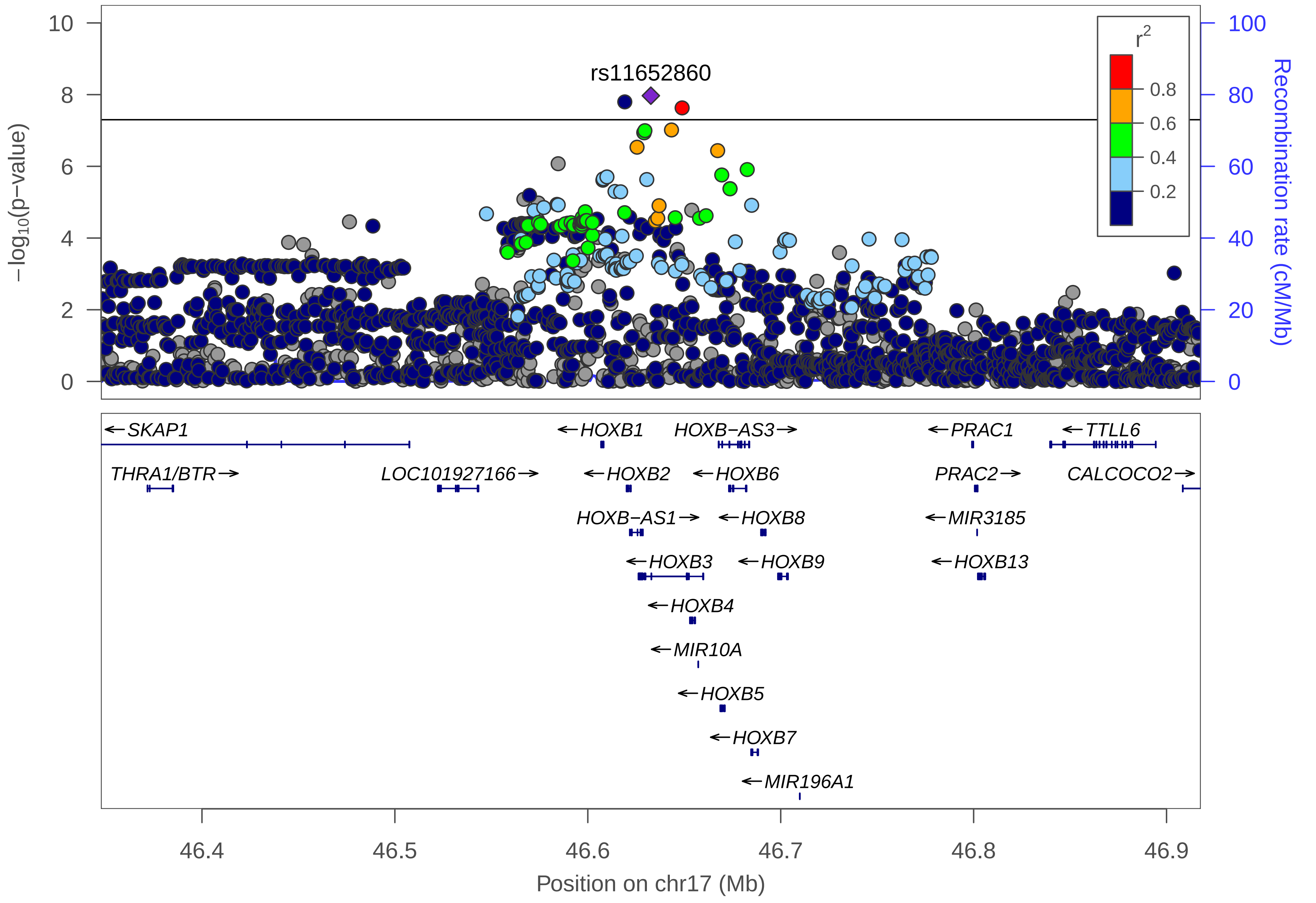

rs11652860
−log10(p−value)
Recombination rate (cM/Mb)
r2
0.8
0.6
0.4
0.2
0
2
4
6
8
10
0
20
40
60
80
100
←SKAP1
THRA1/BTR→
LOC101927166→
←HOXB1
←HOXB2
HOXB−AS1→
←HOXB3
←HOXB4
←MIR10A
←HOXB5
←HOXB7
←MIR196A1
HOXB−AS3→
←HOXB6
←HOXB8
←HOXB9
←PRAC1
PRAC2→
←MIR3185
←HOXB13
←TTLL6
CALCOCO2→
46.4
46.5
46.6
46.7
46.8
46.9
Position on chr17 (Mb)

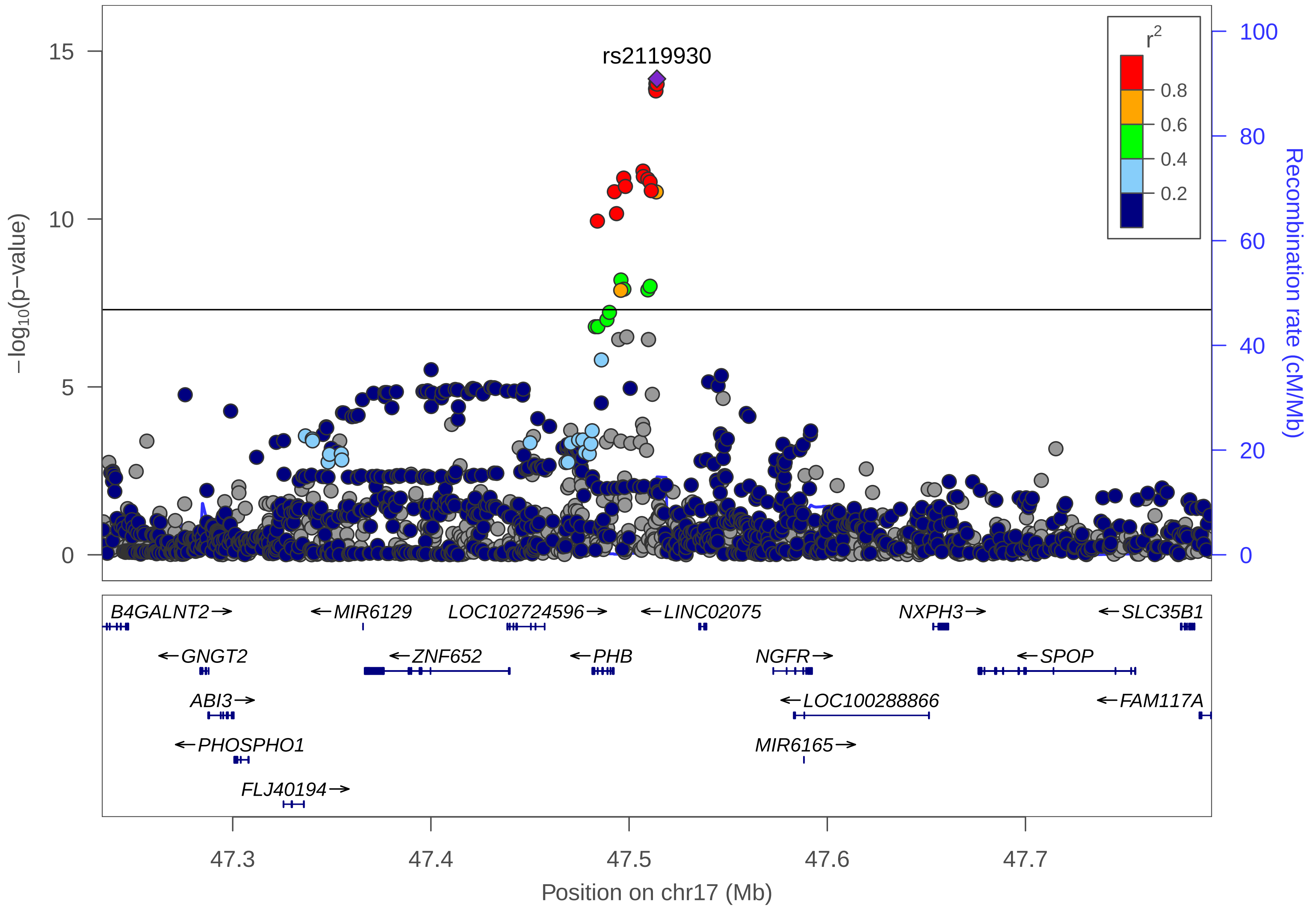

rs2119930
r2
0.8
0.6
0.4
0.2
−log10(p−value)
Recombination rate (cM/Mb)
B4GALNT2→
←MIR6129
LOC102724596→
←LINC02075
NXPH3→
←SLC35B1
←GNGT2
←ZNF652
←PHB
NGFR→
←SPOP
ABI3→
←LOC100288866
←FAM117A
←PHOSPHO1
MIR6165→
FLJ40194→
Position on chr17 (Mb)

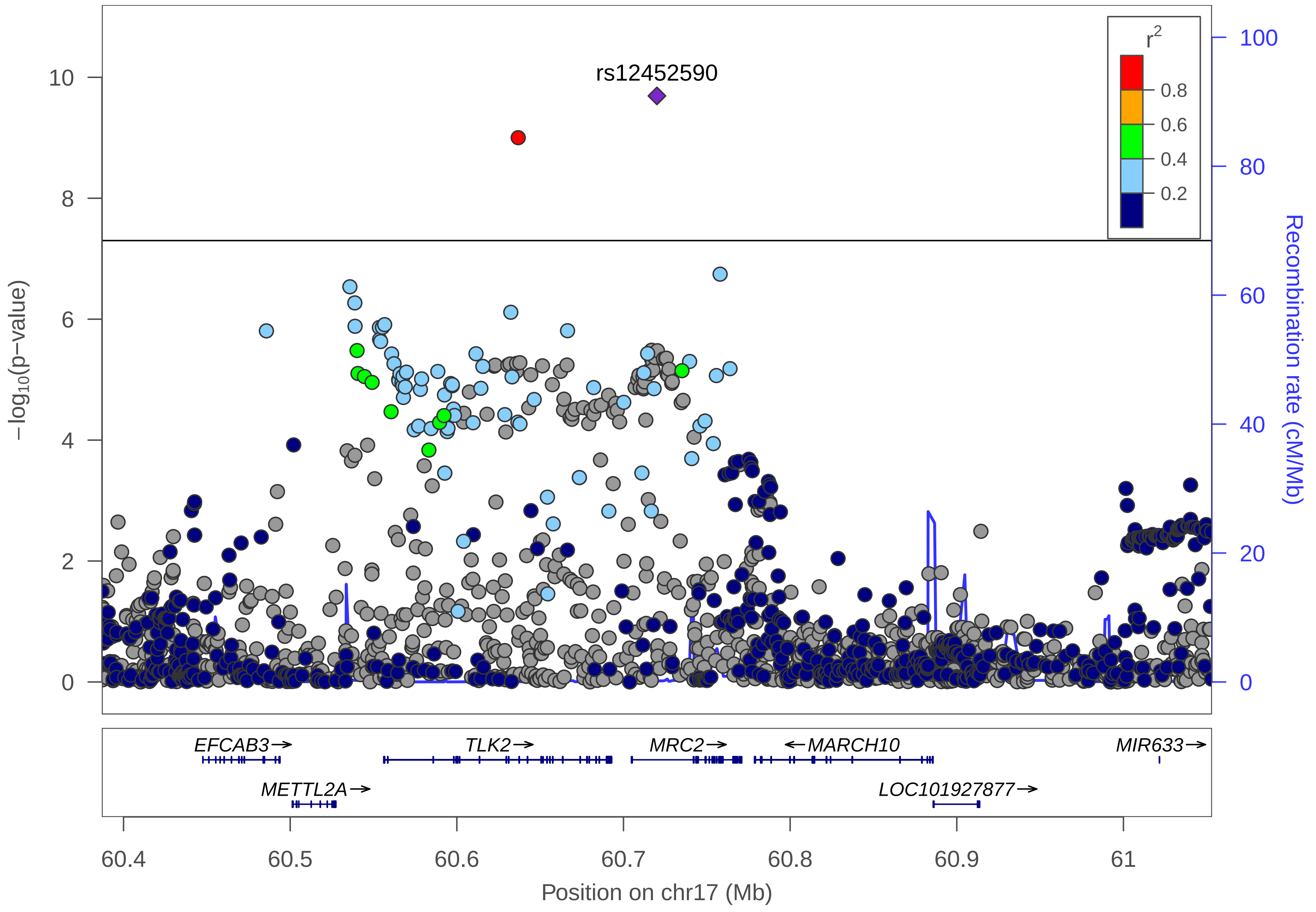

rs12452590
r^2
0.8
0.6
0.4
0.2
−log10(p−value)
Recombination rate (cM/Mb)
10
8
6
4
2
0
100
80
60
40
20
0
EFCAB3→
TLK2→
MRC2→
←MARCH10
MIR633→
METTL2A→
LOC101927877→
60.4
60.5
60.6
60.7
60.8
60.9
61
Position on chr17 (Mb)

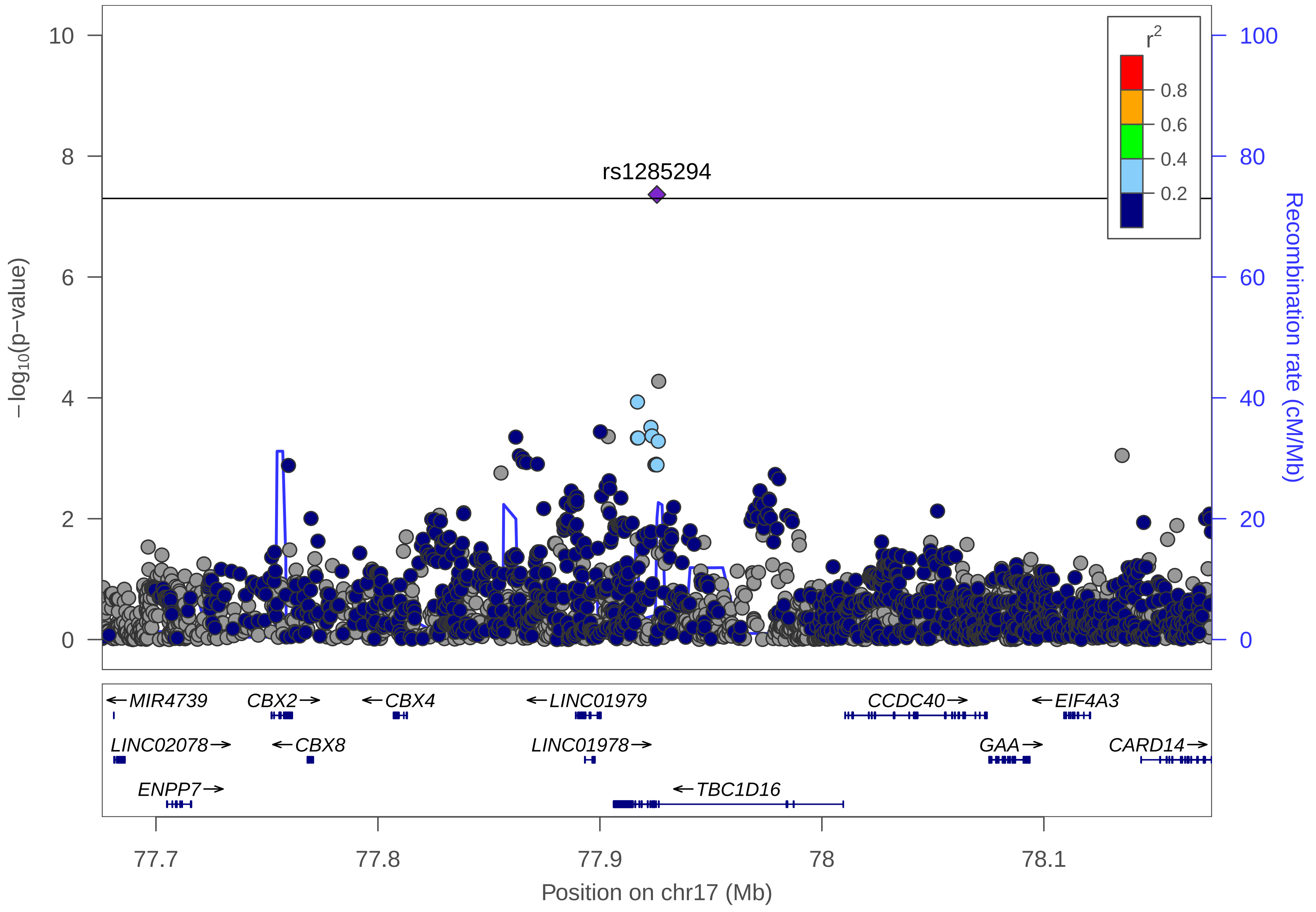

rs1285294
r2
0.8
0.6
0.4
0.2
−log10(p−value)
Recombination rate (cM/Mb)
0
2
4
6
8
10
0
20
40
60
80
100
← MIR4739
CBX2 →
← CBX4
← LINC01979
CCDC40 →
← EIF4A3
LINC02078 →
← CBX8
LINC01978 →
GAA →
CARD14 →
ENPP7 →
← TBC1D16
77.7
77.8
77.9
78
78.1
Position on chr17 (Mb)

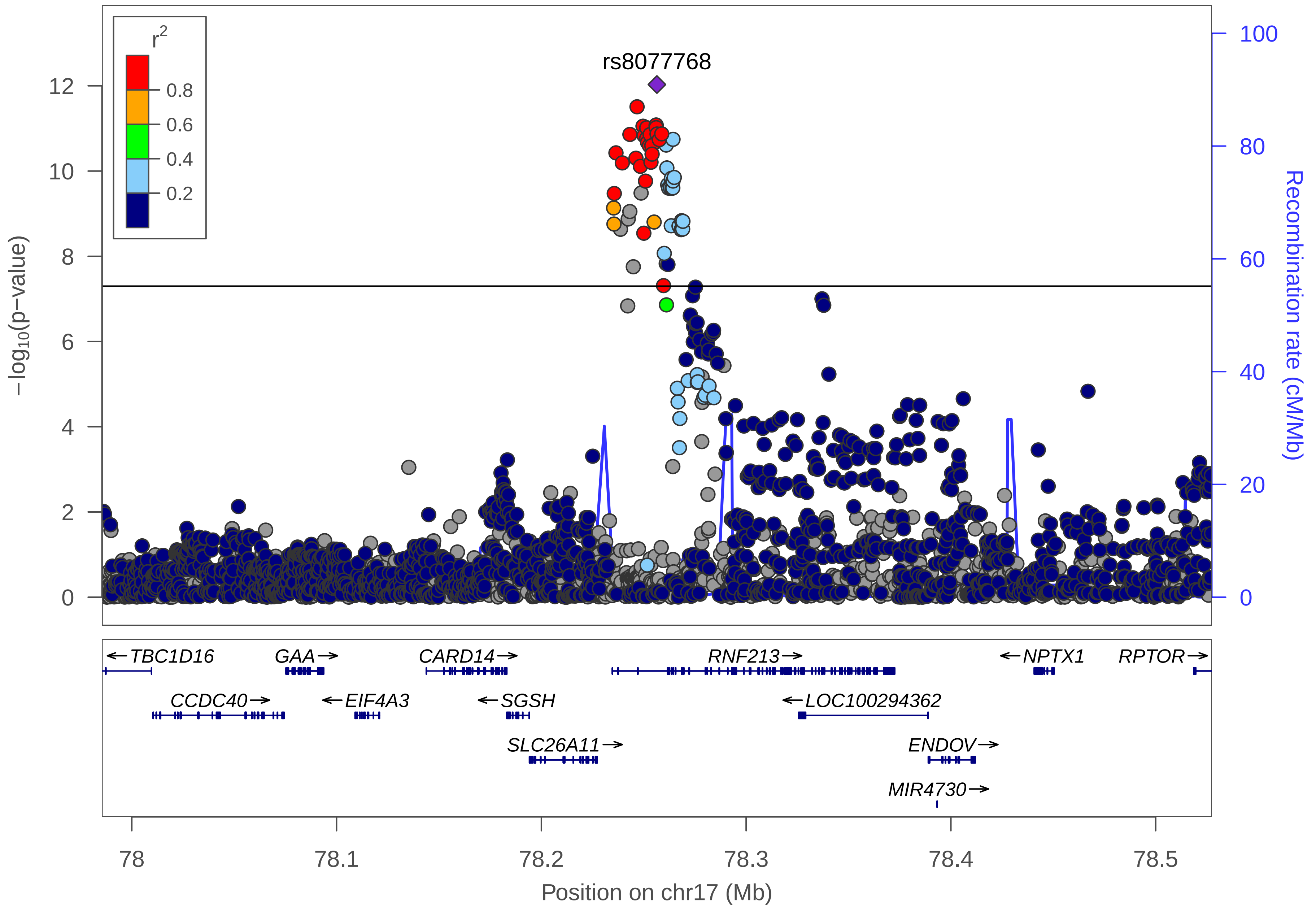

r2
0.8
0.6
0.4
0.2
rs8077768
−log10(p−value)
Recombination rate (cM/Mb)
TBC1D16
GAA
CARD14
RNF213
NPTX1
RPTOR
CCDC40
EIF4A3
SGSH
LOC100294362
SLC26A11
ENDOV
MIR4730
78
78.1
78.2
78.3
78.4
78.5
Position on chr17 (Mb)

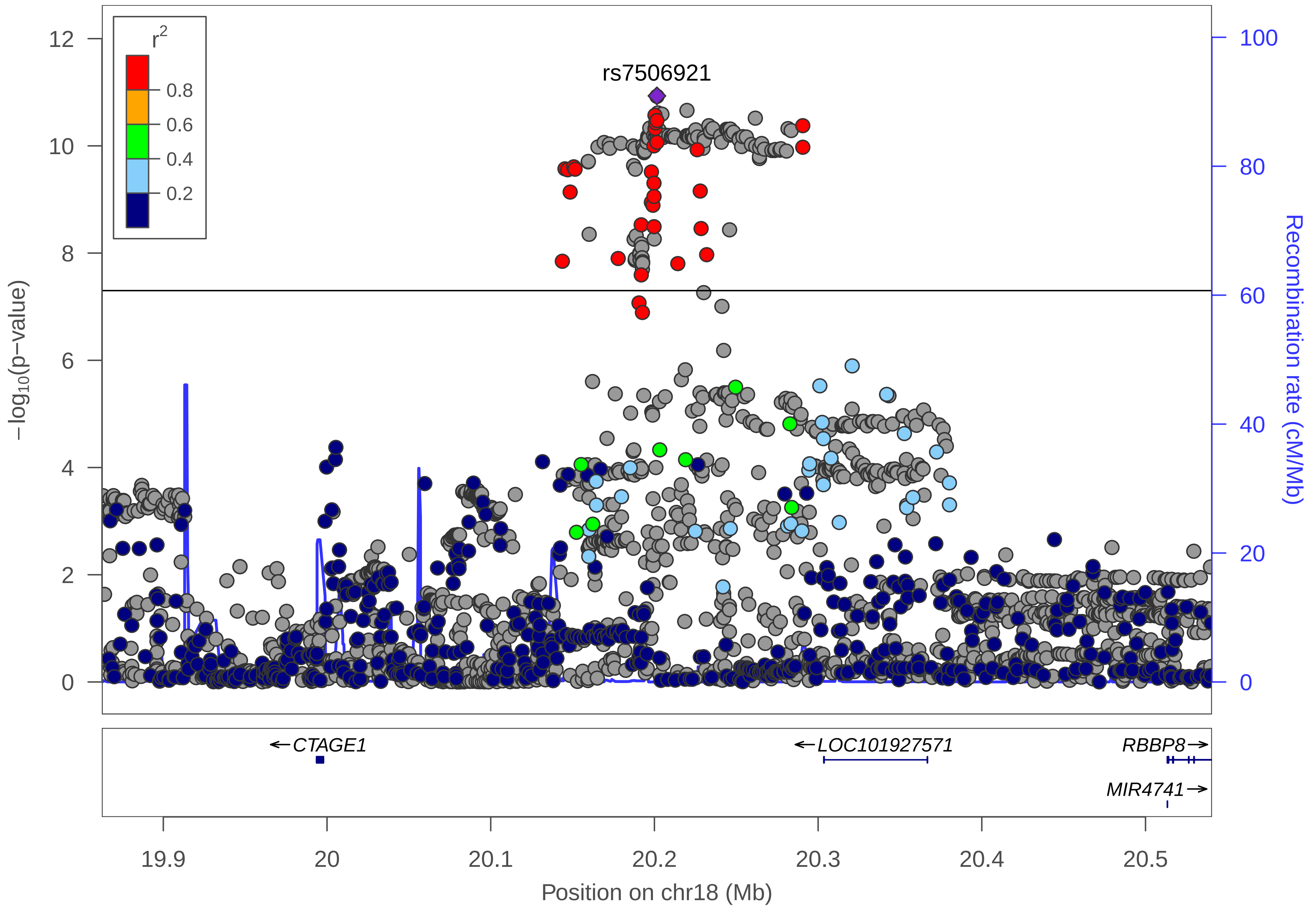

r2
0.8
0.6
0.4
0.2
rs7506921
12
10
8
6
4
2
0
−log10(p−value)
100
80
60
40
20
0
Recombination rate (cM/Mb)
← CTAGE1
← LOC101927571
RBBP8 →
MIR4741 →
19.9
20
20.1
20.2
20.3
20.4
20.5
Position on chr18 (Mb)

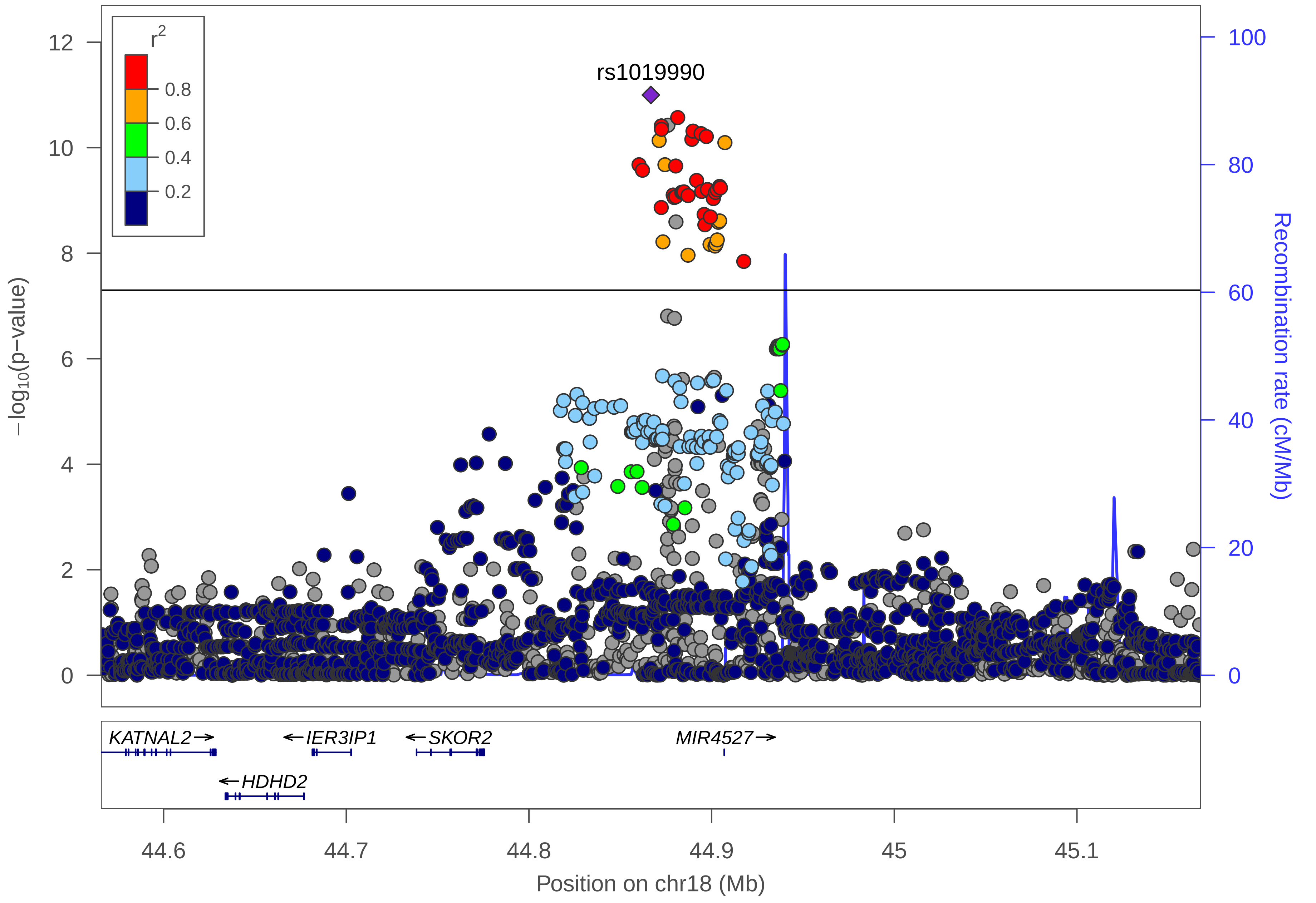

r2
0.8
0.6
0.4
0.2
rs1019990
12
10
8
6
4
2
0
−log10(p−value)
100
80
60
40
20
0
Recombination rate (cM/Mb)
KATNAL2→
←IER3IP1
←SKOR2
MIR4527→
←HDHD2
44.6
44.7
44.8
44.9
45
45.1
Position on chr18 (Mb)

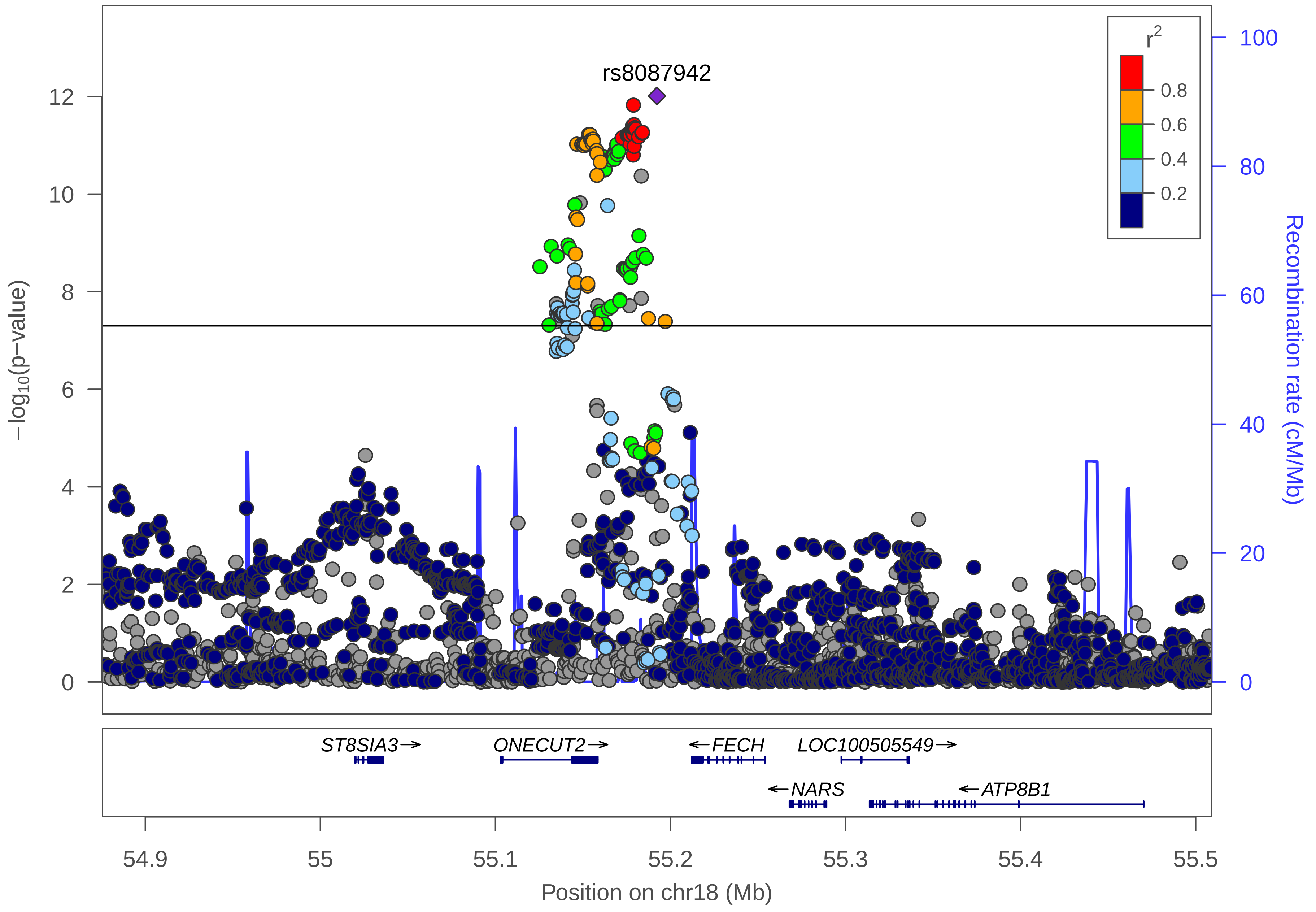
rs8087942
r²
0.8
0.6
0.4
0.2
−log10(p−value)
12
10
8
6
4
2
0
100
80
60
40
20
0
Recombination rate (cM/Mb)
ST8SIA3→
ONECUT2→
←FECH
LOC100505549→
←NARS
←ATP8B1
54.9
55
55.1
55.2
55.3
55.4
55.5
Position on chr18 (Mb)

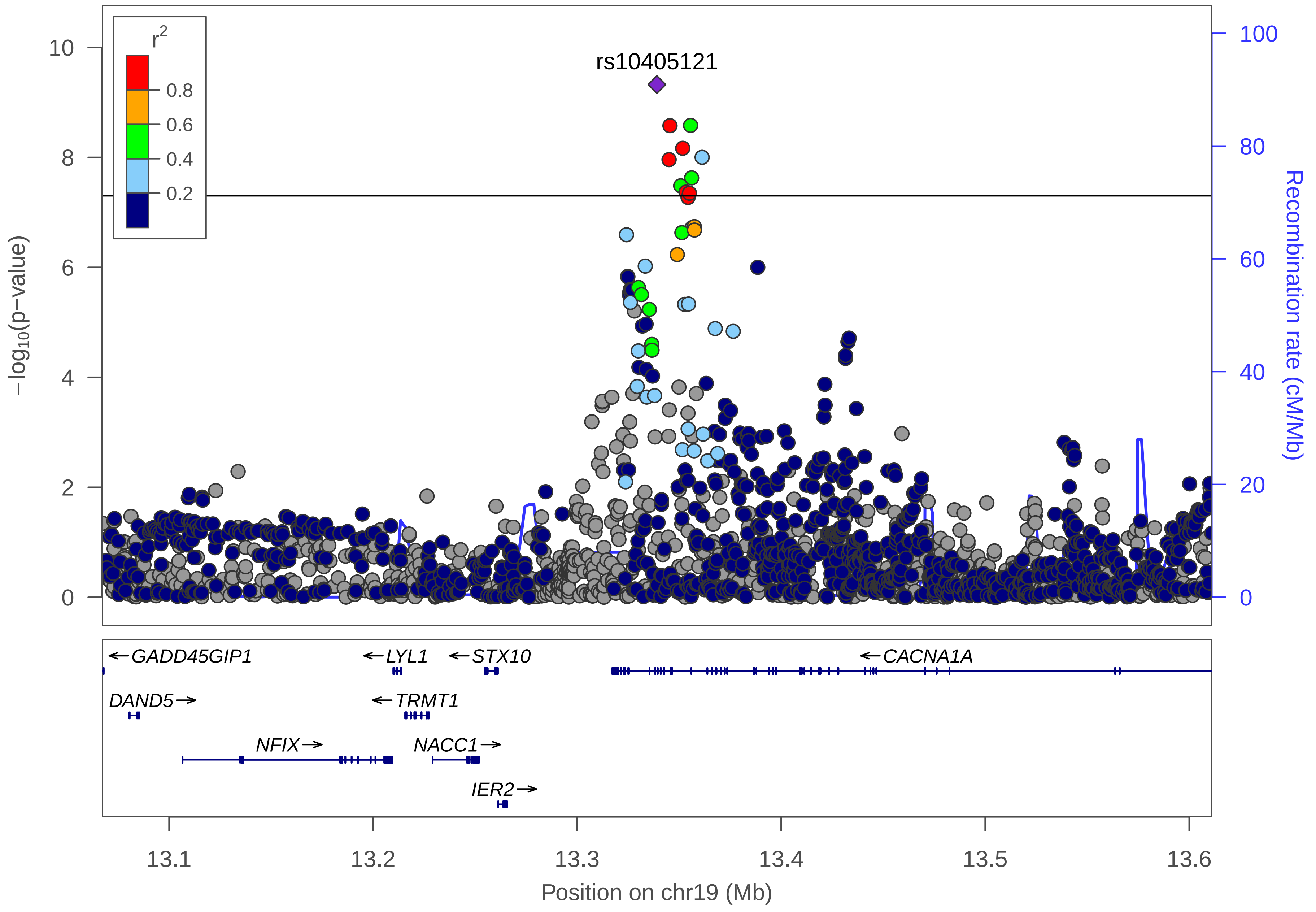

rs10405121
r2
0.8
0.6
0.4
0.2
10
8
6
4
2
0
−log10(p−value)
100
80
60
40
20
0
Recombination rate (cM/Mb)
GADD45GIP1
LYL1
STX10
CACNA1A
DAND5
TRMT1
NFIX
NACC1
IER2
13.1
13.2
13.3
13.4
13.5
13.6
Position on chr19 (Mb)

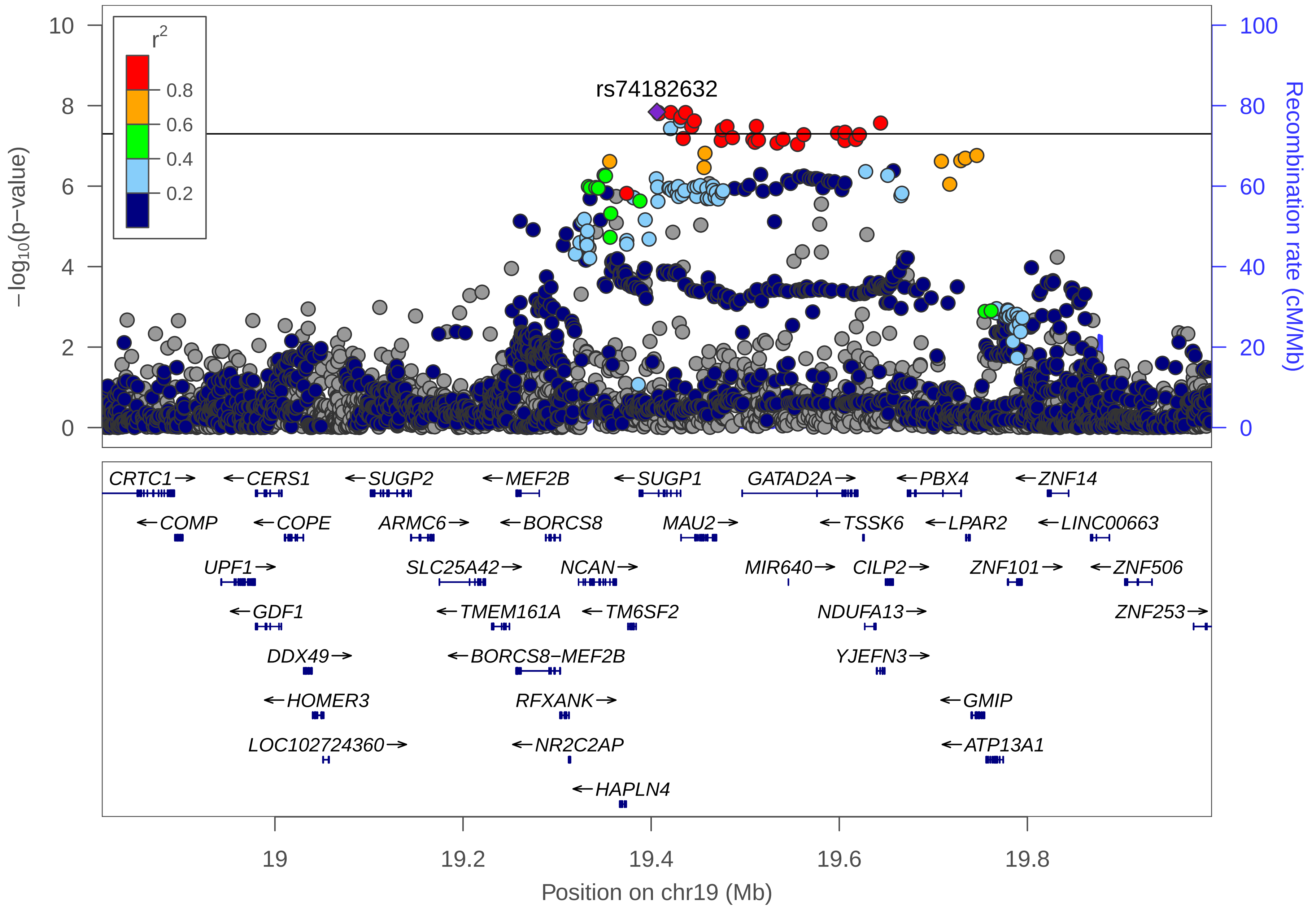
r2
0.8
0.6
0.4
0.2
rs74182632
−log10(p−value)
Recombination rate (cM/Mb)
10
8
6
4
2
0
100
80
60
40
20
0
CRTC1→
←CERS1
←SUGP2
←MEF2B
←SUGP1
GATAD2A→
←PBX4
←ZNF14
←COMP
←COPE
ARMC6→
←BORCS8
MAU2→
←TSSK6
←LPAR2
←LINC00663
UPF1→
SLC25A42→
NCAN→
MIR640→
CILP2→
ZNF101→
←ZNF506
←GDF1
←TMEM161A
←TM6SF2
NDUFA13→
ZNF253→
DDX49→
←BORCS8−MEF2B
YJEFN3→
←HOMER3
RFXANK→
←GMIP
LOC102724360→
←NR2C2AP
←ATP13A1
←HAPLN4
19
19.2
19.4
19.6
19.8
Position on chr19 (Mb)

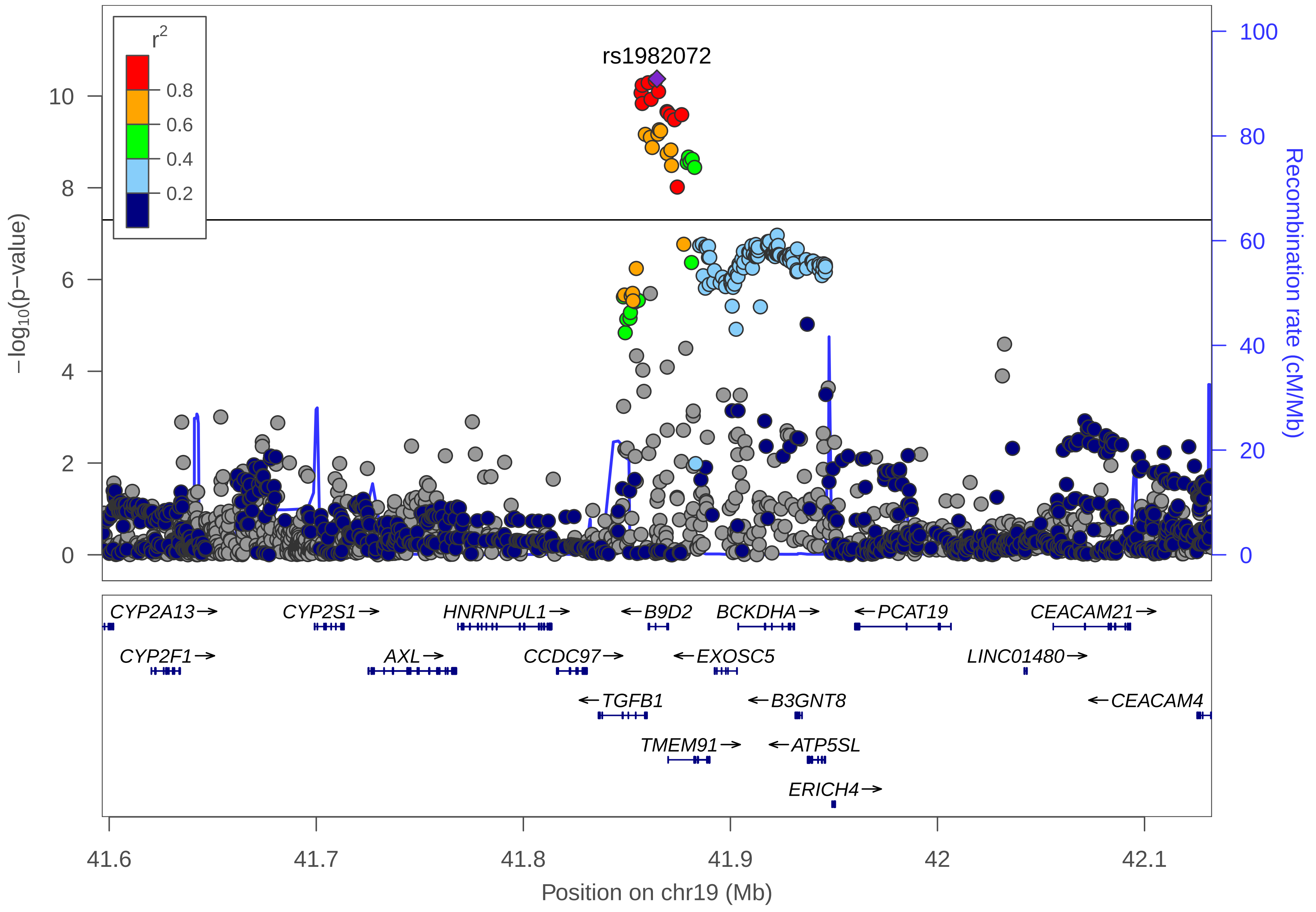

r2
0.8
0.6
0.4
0.2
rs1982072
−log10(p−value)
Recombination rate (cM/Mb)
0
2
4
6
8
10
0
20
40
60
80
100
CYP2A13→
CYP2S1→
HNRNPUL1→
←B9D2
BCKDHA→
←PCAT19
CEACAM21→
CYP2F1→
AXL→
CCDC97→
←EXOSC5
LINC01480→
←TGFB1
←B3GNT8
←CEACAM4
TMEM91→
←ATP5SL
ERICH4→
41.6
41.7
41.8
41.9
42
42.1
Position on chr19 (Mb)

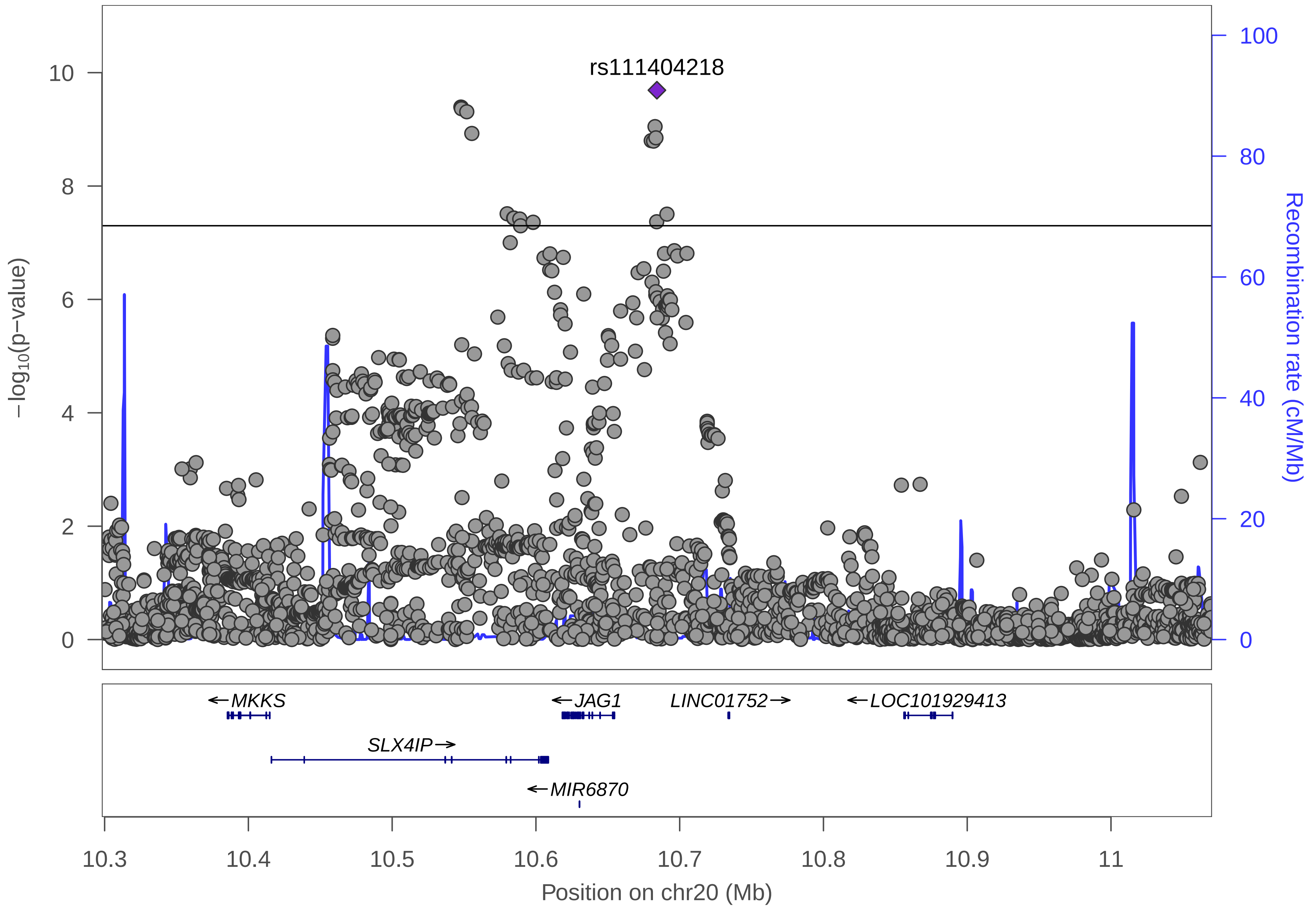
rs111404218
−log10(p−value)
Recombination rate (cM/Mb)
0
2
4
6
8
10
0
20
40
60
80
100
MKKS
JAG1
LINC01752
LOC101929413
SLX4IP
MIR6870
10.3
10.4
10.5
10.6
10.7
10.8
10.9
11
Position on chr20 (Mb)

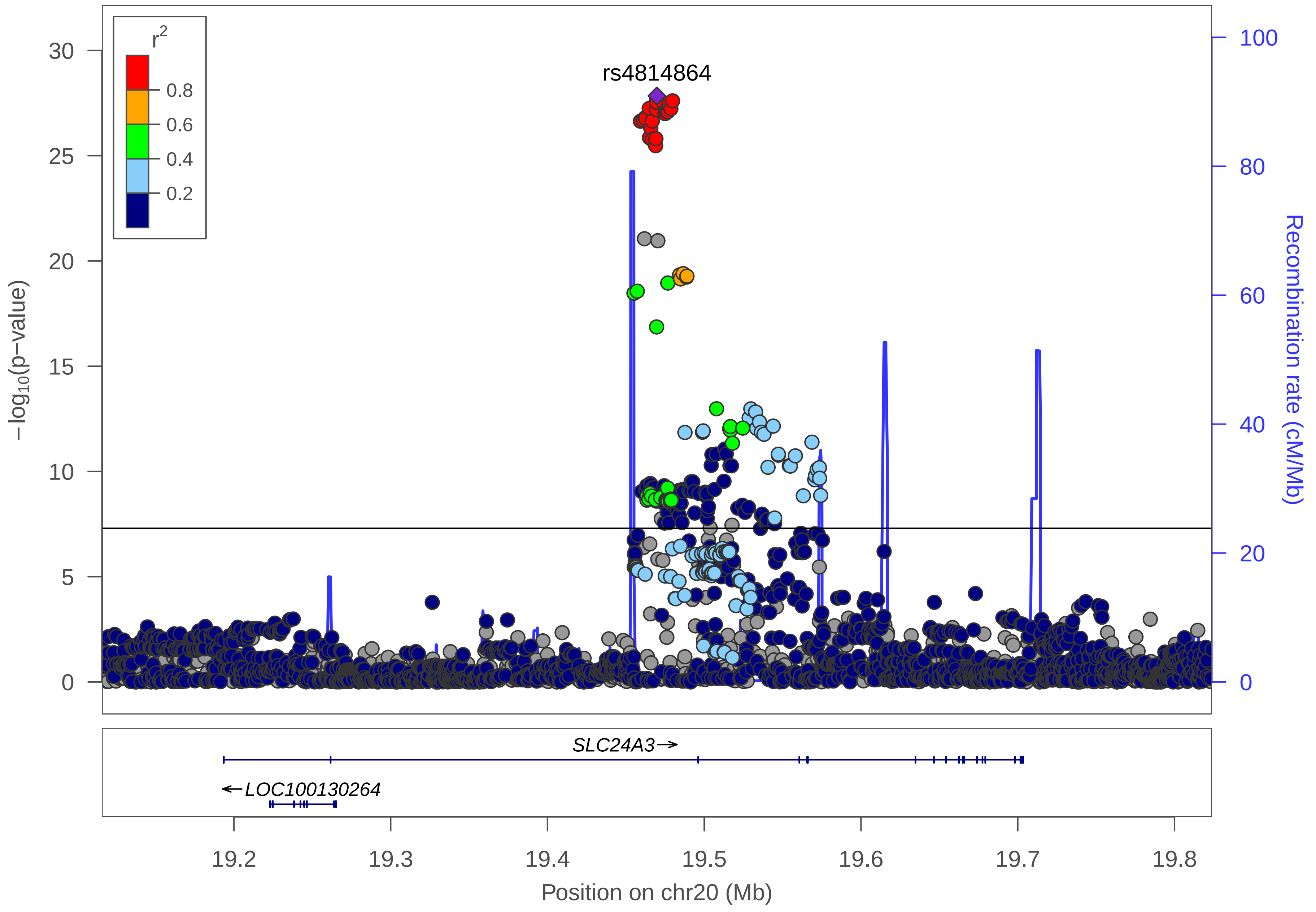
r2
0.8
0.6
0.4
0.2
rs4814864
30
25
20
15
10
5
0
−log10(p−value)
100
80
60
40
20
0
Recombination rate (cM/Mb)
SLC24A3→
←LOC100130264
19.2
19.3
19.4
19.5
19.6
19.7
19.8
Position on chr20 (Mb)

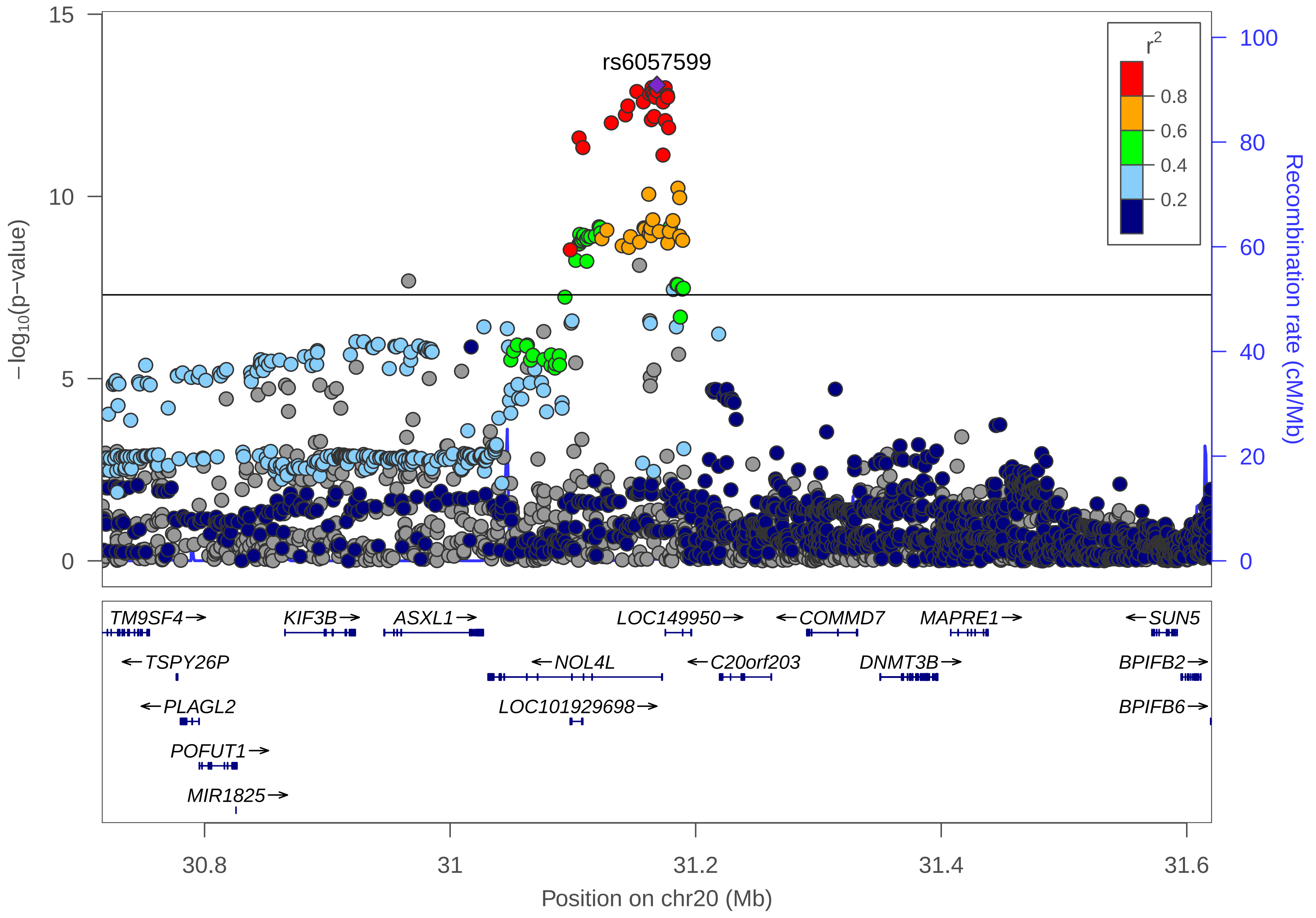

rs6057599
r^2
0.8
0.6
0.4
0.2
−log10(p−value)
Recombination rate (cM/Mb)
15
10
5
0
100
80
60
40
20
0
TM9SF4→
KIF3B→
ASXL1→
LOC149950→
←COMMD7
MAPRE1→
←SUN5
←TSPY26P
←NOL4L
←C20orf203
DNMT3B→
BPIFB2→
←PLAGL2
LOC101929698→
BPIFB6→
POFUT1→
MIR1825→
30.8
31
31.2
31.4
31.6
Position on chr20 (Mb)

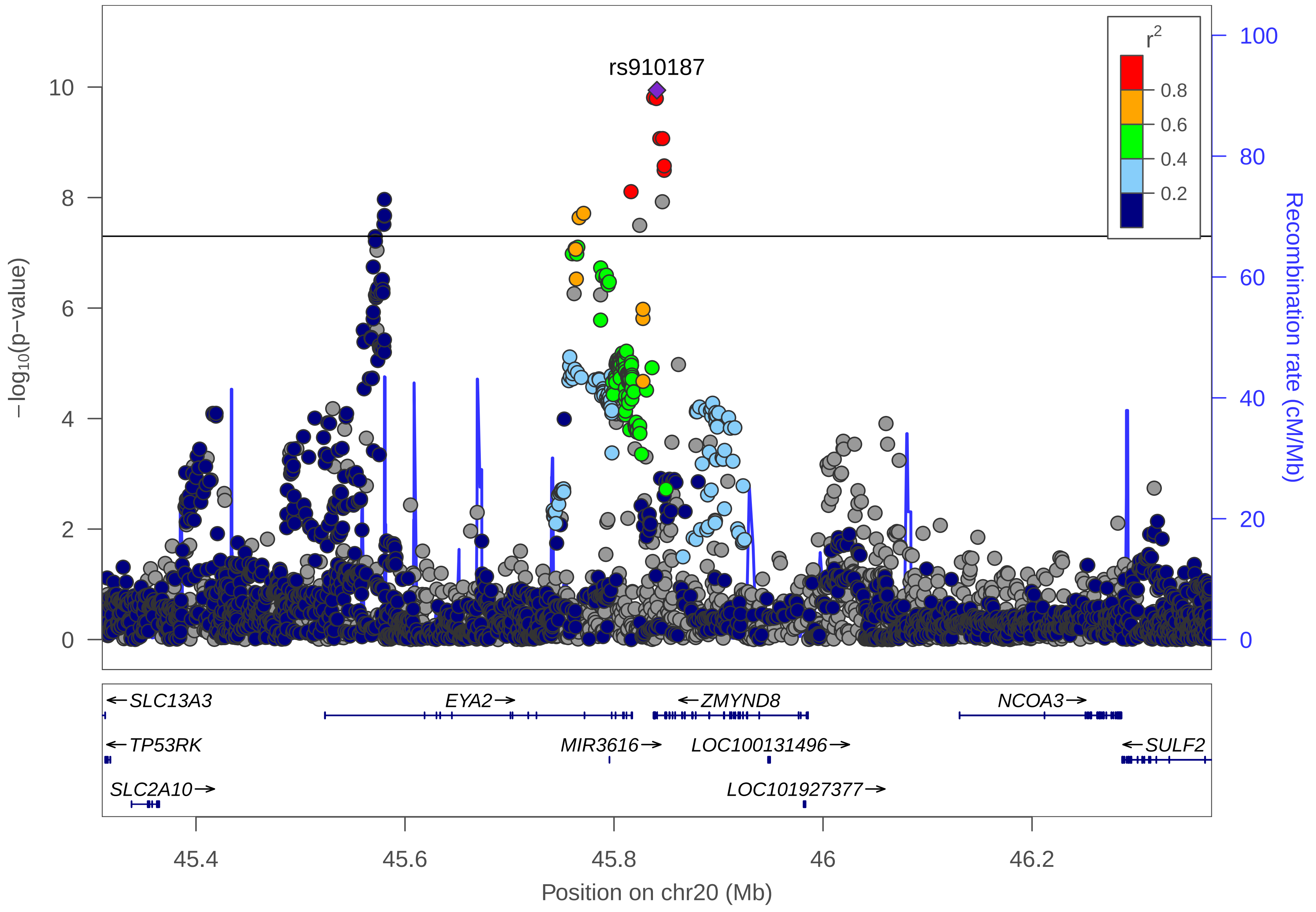

rs910187
r2
0.8
0.6
0.4
0.2
10
8
6
4
2
0
100
80
60
40
20
0
−log10(p−value)
Recombination rate (cM/Mb)
SLC13A3
EYA2
ZMYND8
NCOA3
TP53RK
MIR3616
LOC100131496
SULF2
SLC2A10
LOC101927377
45.4
45.6
45.8
46
46.2
Position on chr20 (Mb)

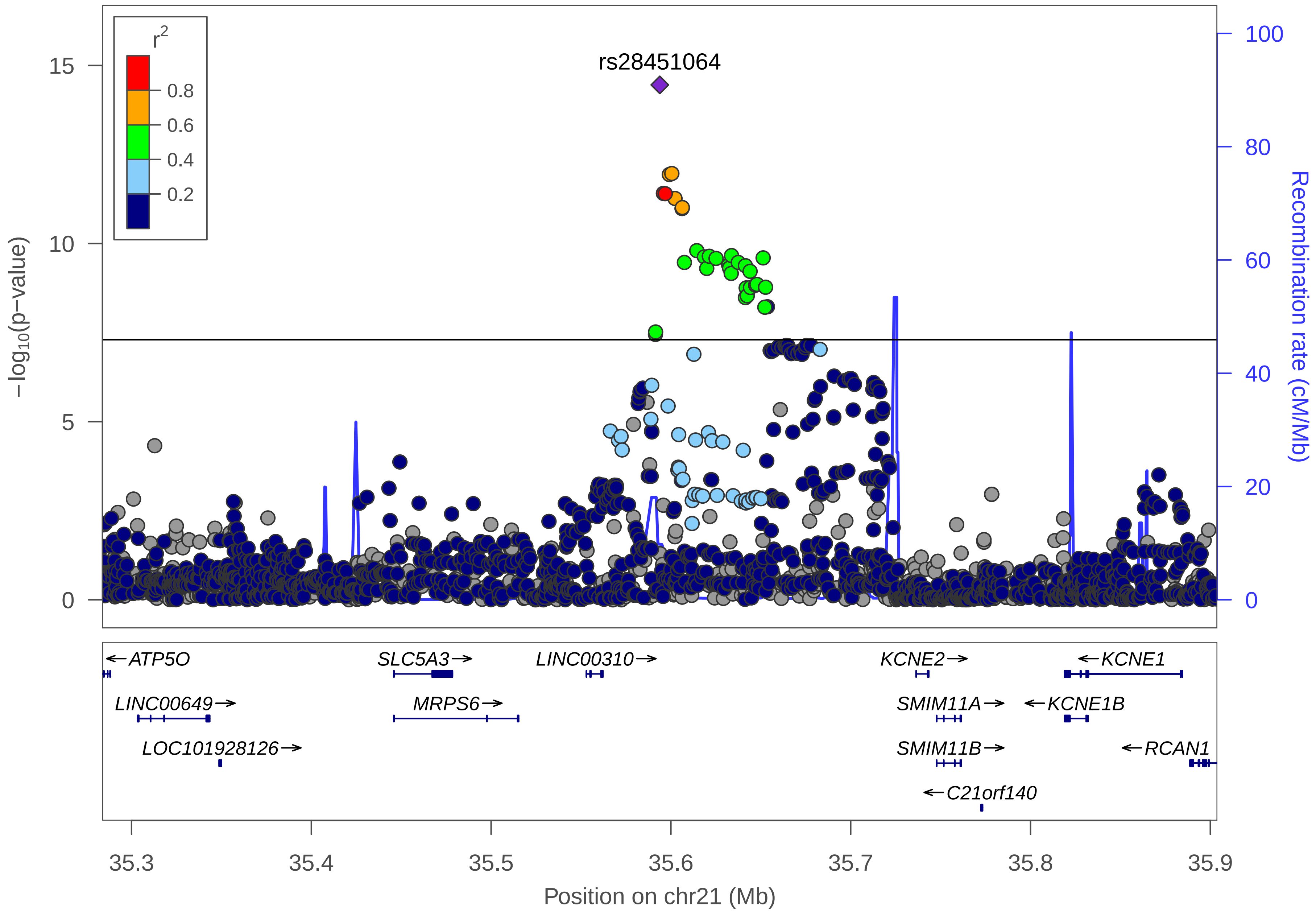

rs28451064
r2
0.8
0.6
0.4
0.2
15
10
5
0
−log10(p−value)
100
80
60
40
20
0
Recombination rate (cM/Mb)
←ATP5O
SLC5A3→
LINC00310→
KCNE2→
←KCNE1
LINC00649→
MRPS6→
SMIM11A→
←KCNE1B
LOC101928126→
SMIM11B→
←RCAN1
←C21orf140
35.3
35.4
35.5
35.6
35.7
35.8
35.9
Position on chr21 (Mb)

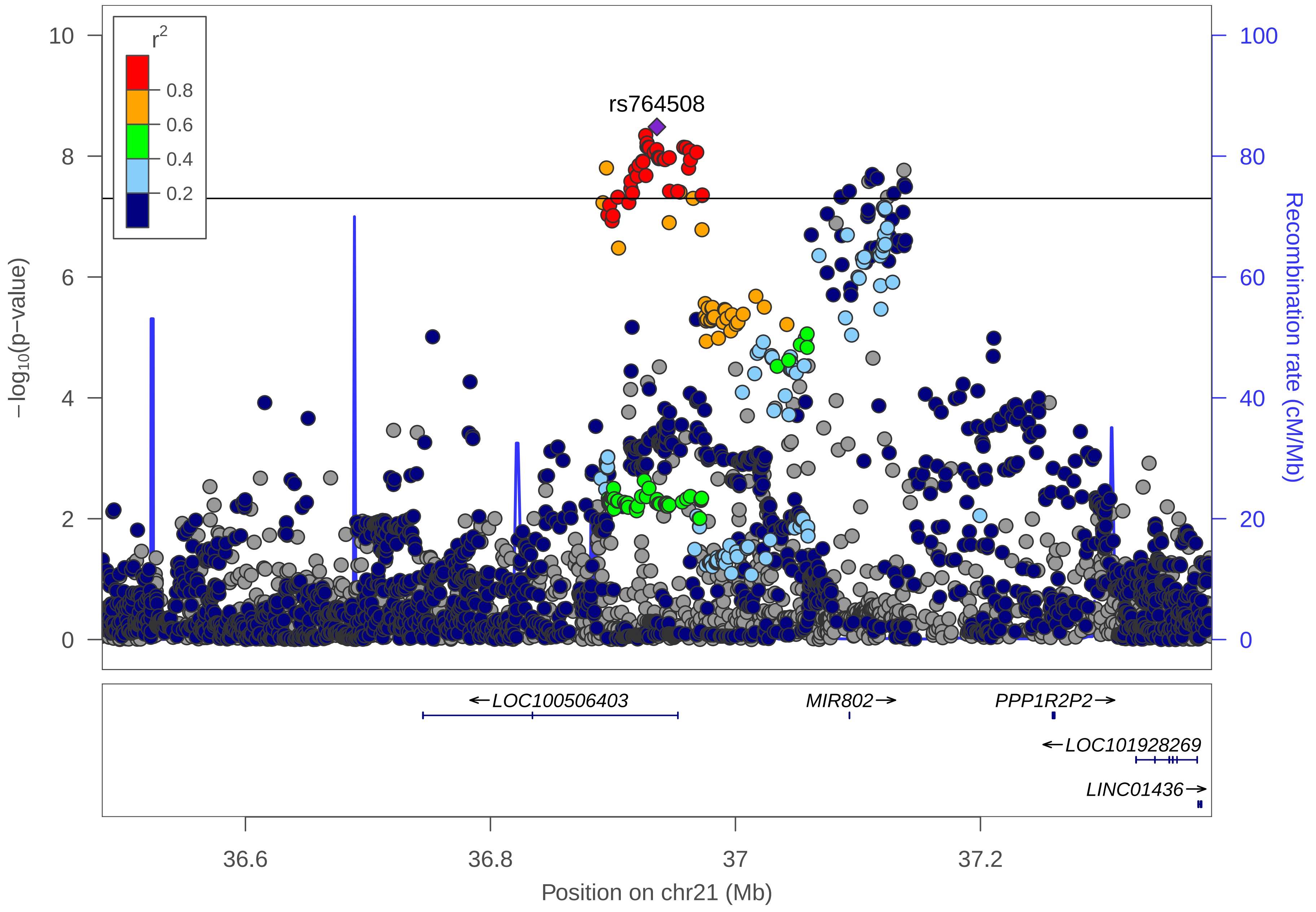
r2
0.8
0.6
0.4
0.2
rs764508
10
8
6
4
2
0
−log10(p−value)
100
80
60
40
20
0
Recombination rate (cM/Mb)
←LOC100506403
MIR802→
PPP1R2P2→
←LOC101928269
LINC01436→
36.6
36.8
37
37.2
Position on chr21 (Mb)

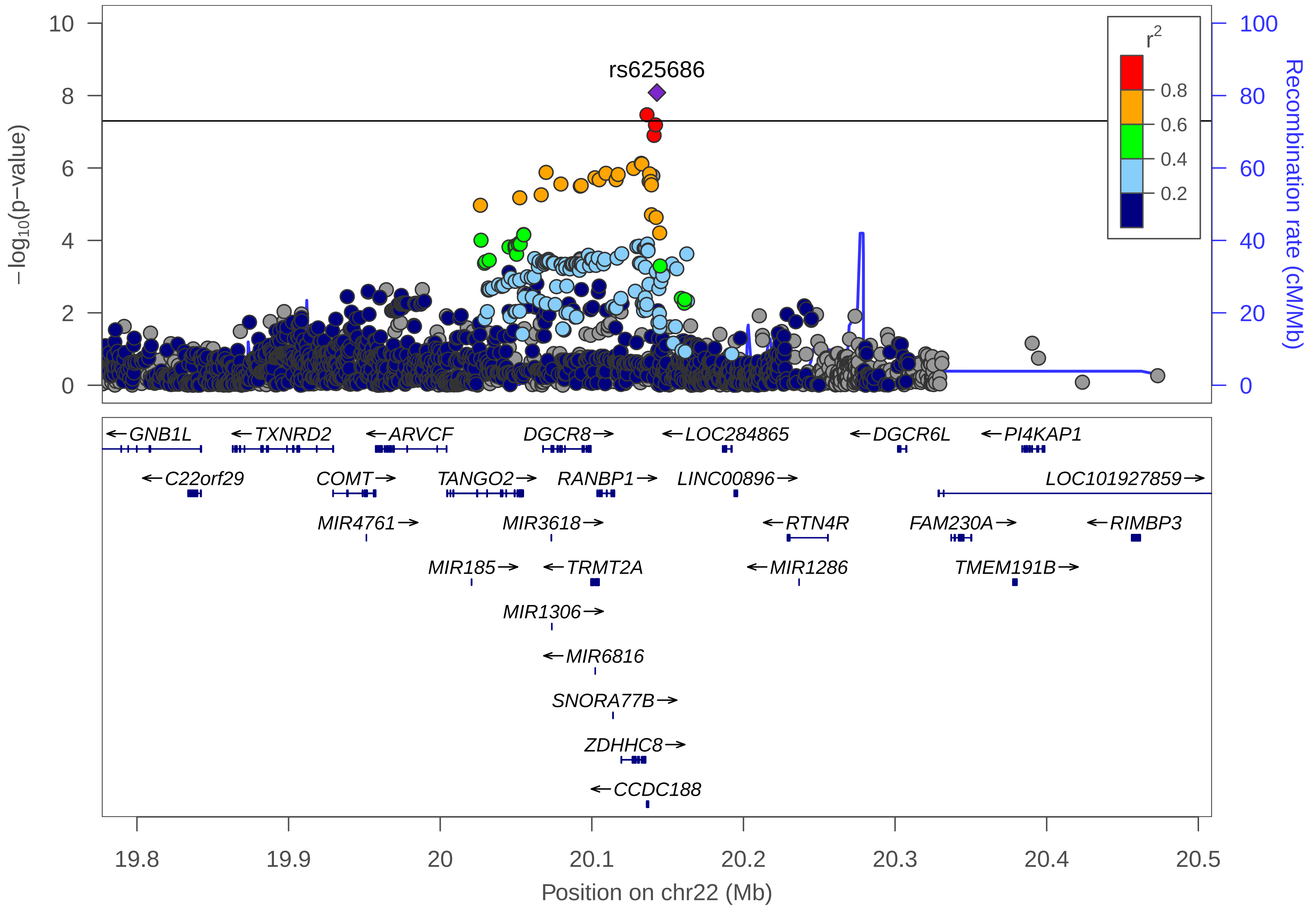

rs625686
r2
0.8
0.6
0.4
0.2
−log10(p−value)
Recombination rate (cM/Mb)
GNB1L
TXNRD2
ARVCF
DGCR8
LOC284865
DGCR6L
PI4KAP1
C22orf29
COMT
TANGO2
RANBP1
LINC00896
LOC101927859
MIR4761
MIR3618
RTN4R
FAM230A
RIMBP3
MIR185
TRMT2A
MIR1286
TMEM191B
MIR1306
MIR6816
SNORA77B
ZDHHC8
CCDC188
Position on chr22 (Mb)

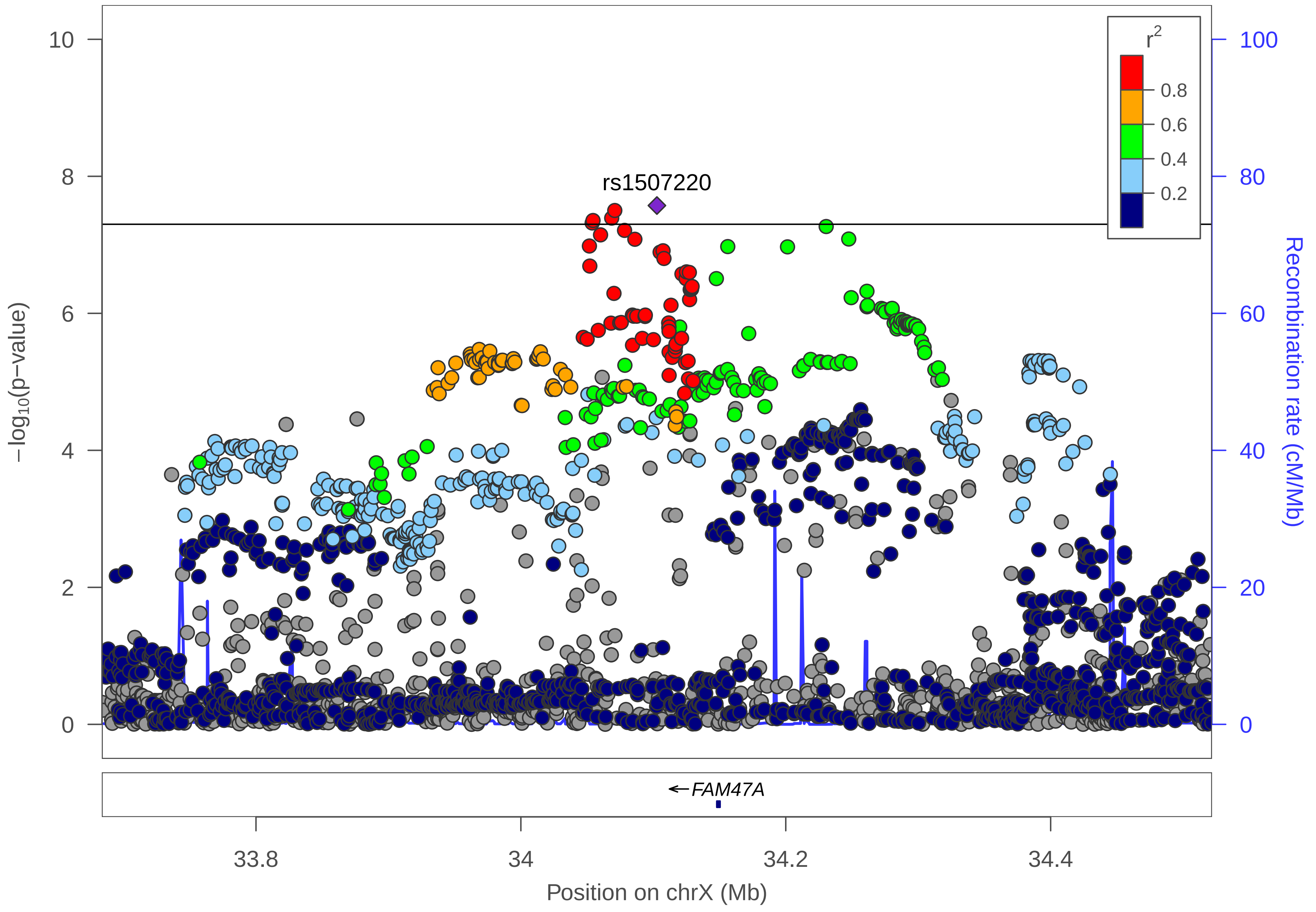

rs1507220
r2
0.8
0.6
0.4
0.2
10
8
6
4
2
0
100
80
60
40
20
0
−log10(p−value)
Recombination rate (cM/Mb)
FAM47A
33.8
34
34.2
34.4
Position on chrX (Mb)

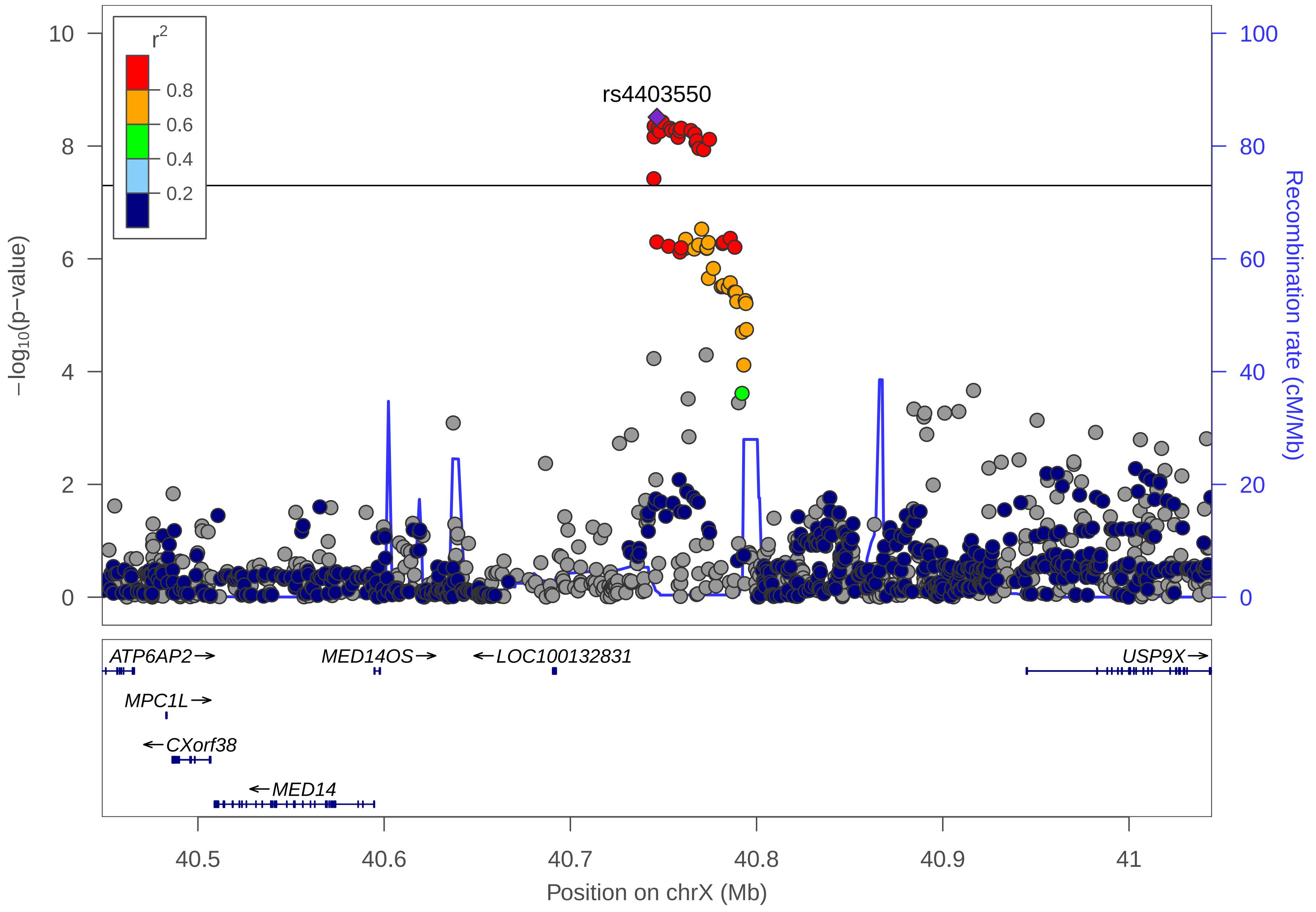
r2
0.8
0.6
0.4
0.2
rs4403550
−log10(p−value)
Recombination rate (cM/Mb)
0
2
4
6
8
10
0
20
40
60
80
100
ATP6AP2→
MED14OS→
←LOC100132831
USP9X→
MPC1L→
←CXorf38
←MED14
40.5
40.6
40.7
40.8
40.9
41
Position on chrX (Mb)
