## Supplementary Figure 4. for "Genome-wide analysis of 102,084 migraine cases identifies 123 risk loci and subtype-specific risk alleles"

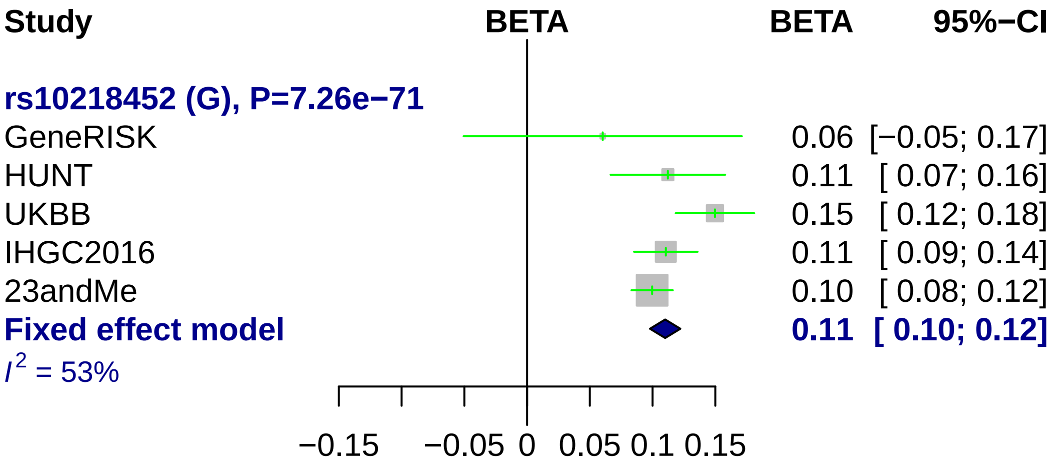
Study
BETA
BETA
95%−CI
rs10218452 (G), P=7.26e−71
GeneRISK
0.06 [−0.05; 0.17]
HUNT
0.11 [ 0.07; 0.16]
UKBB
0.15 [ 0.12; 0.18]
IHGC2016
0.11 [ 0.09; 0.14]
23andMe
0.10 [ 0.08; 0.12]
Fixed effect model
0.11 [ 0.10; 0.12]
$I^2$ = 53%
−0.15
−0.05
0
0.05
0.1
0.15

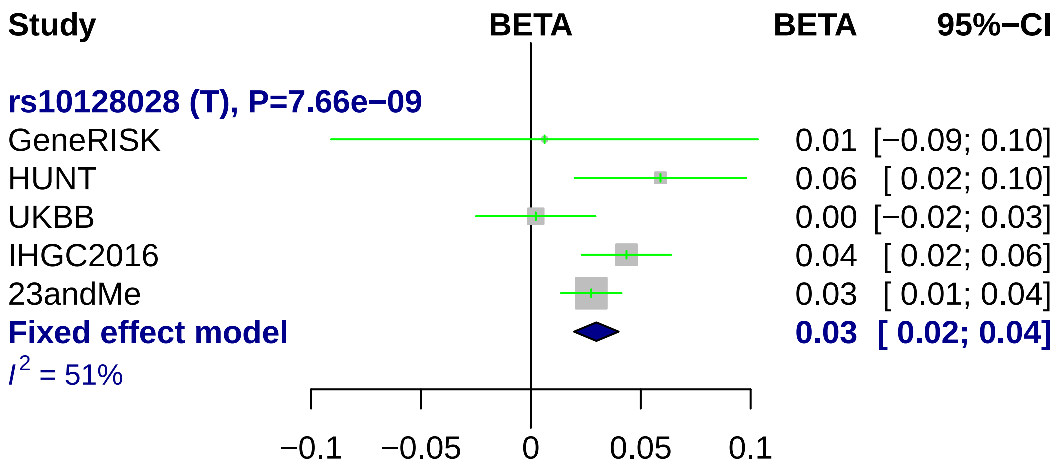
Study
BETA
BETA
95%−CI
rs10128028 (T), P=7.66e−09
GeneRISK
0.01 [−0.09; 0.10]
HUNT
0.06 [ 0.02; 0.10]
UKBB
0.00 [−0.02; 0.03]
IHGC2016
0.04 [ 0.02; 0.06]
23andMe
0.03 [ 0.01; 0.04]
Fixed effect model
0.03 [ 0.02; 0.04]
I2 = 51%
−0.1
−0.05
0
0.05
0.1

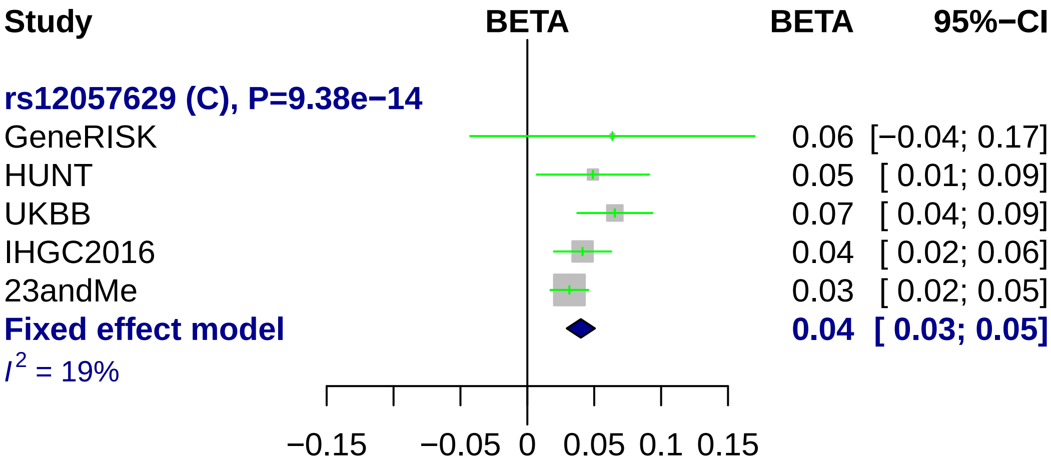
Study
BETA
BETA
95%−CI
rs12057629 (C), P=9.38e−14
GeneRISK 0.06 [−0.04; 0.17]
HUNT 0.05 [ 0.01; 0.09]
UKBB 0.07 [ 0.04; 0.09]
IHGC2016 0.04 [ 0.02; 0.06]
23andMe 0.03 [ 0.02; 0.05]
Fixed effect model 0.04 [ 0.03; 0.05]
I^2 = 19%
−0.15
−0.05
0
0.05
0.1
0.15

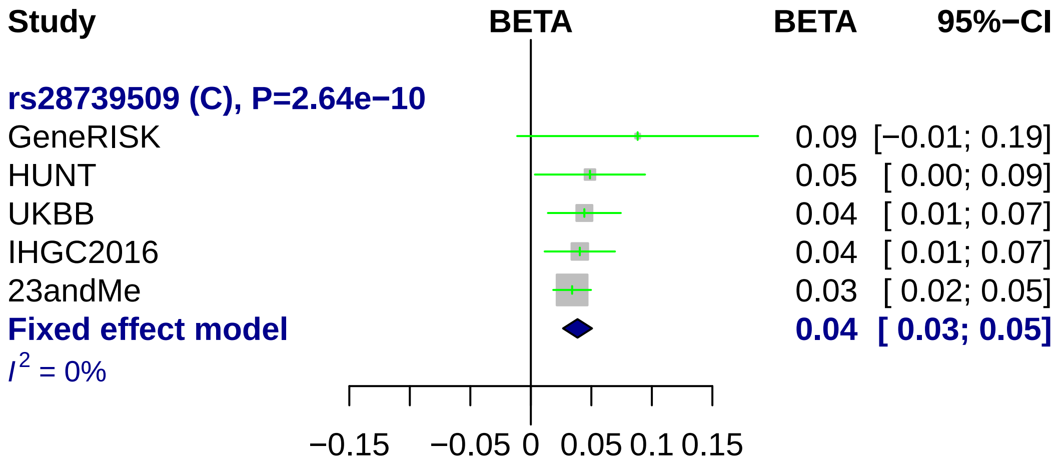
Study
BETA
BETA
95%−CI
rs28739509 (C), P=2.64e−10
GeneRISK 0.09 [−0.01; 0.19]
HUNT 0.05 [ 0.00; 0.09]
UKBB 0.04 [ 0.01; 0.07]
IHGC2016 0.04 [ 0.01; 0.07]
23andMe 0.03 [ 0.02; 0.05]
Fixed effect model 0.04 [ 0.03; 0.05]
I2 = 0%
−0.15
−0.05
0
0.05
0.1
0.15

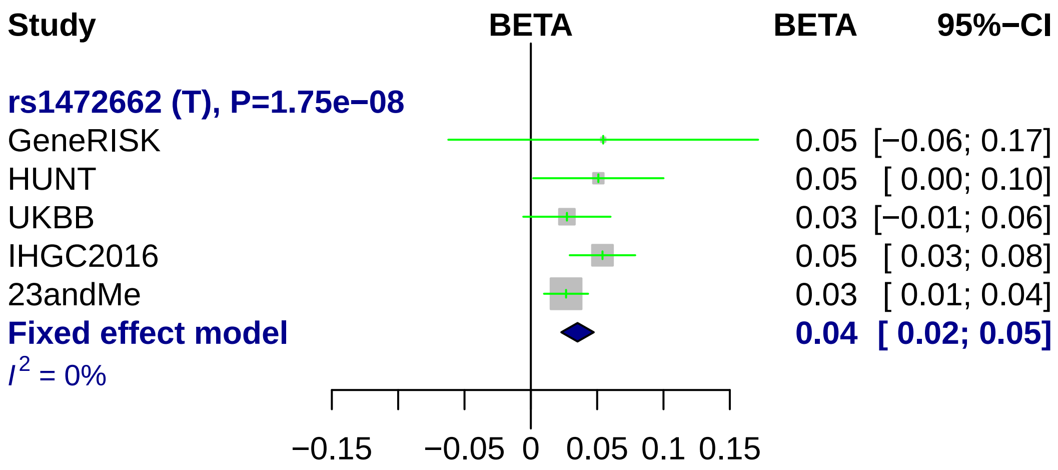
Study
BETA
BETA
95%−CI
rs1472662 (T), P=1.75e−08
GeneRISK 0.05 [−0.06; 0.17]
HUNT 0.05 [ 0.00; 0.10]
UKBB 0.03 [−0.01; 0.06]
IHGC2016 0.05 [ 0.03; 0.08]
23andMe 0.03 [ 0.01; 0.04]
Fixed effect model 0.04 [ 0.02; 0.05]
I^2 = 0%
−0.15
−0.05
0
0.05
0.1
0.15

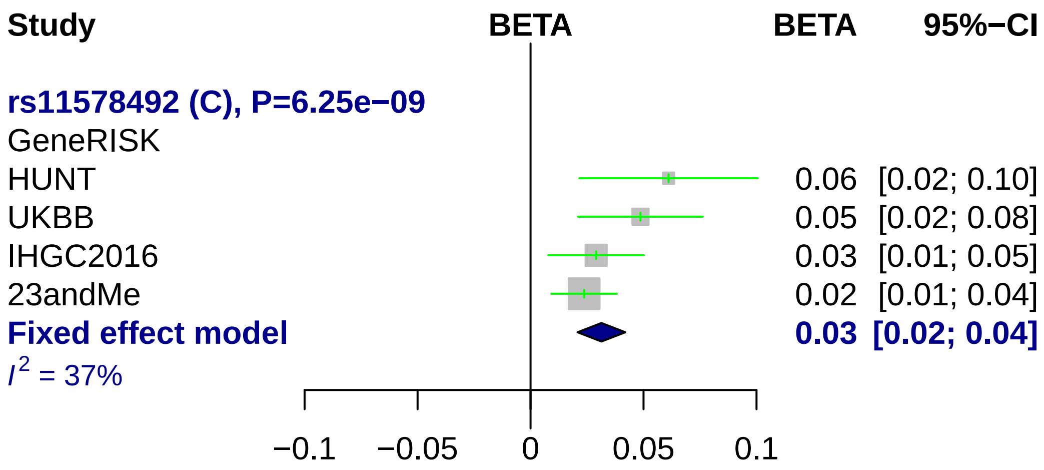
Study
BETA
BETA
95%−CI
rs11578492 (C), P=6.25e−09
GeneRISK
HUNT 0.06 [0.02; 0.10]
UKBB 0.05 [0.02; 0.08]
IHGC2016 0.03 [0.01; 0.05]
23andMe 0.02 [0.01; 0.04]
Fixed effect model 0.03 [0.02; 0.04]
I2 = 37%
−0.1
−0.05
0
0.05
0.1

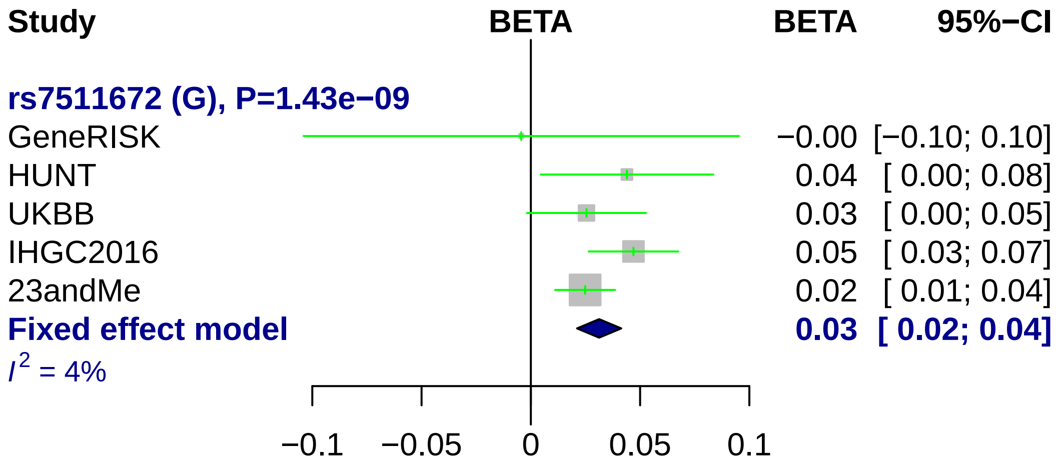

Study
BETA
BETA
95%−CI
rs7511672 (G), P=1.43e−09
GeneRISK
HUNT
UKBB
IHGC2016
23andMe
Fixed effect model
$I^2$ = 4%
−0.00 [−0.10; 0.10]
0.04 [ 0.00; 0.08]
0.03 [ 0.00; 0.05]
0.05 [ 0.03; 0.07]
0.02 [ 0.01; 0.04]
0.03 [ 0.02; 0.04]
−0.1
−0.05
0
0.05
0.1

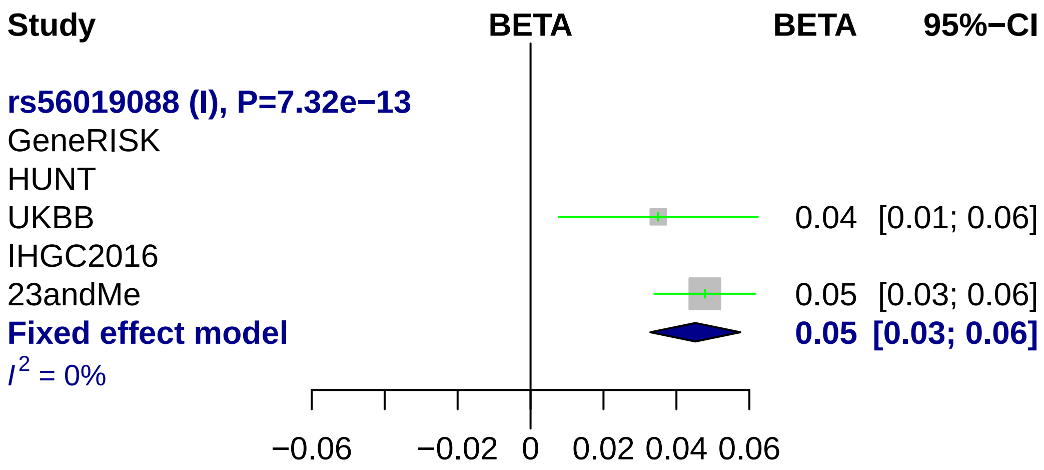
Study
BETA
BETA
95%−CI
rs56019088 (I), P=7.32e−13
GeneRISK
HUNT
UKBB
0.04 [0.01; 0.06]
IHGC2016
23andMe
0.05 [0.03; 0.06]
Fixed effect model
0.05 [0.03; 0.06]
I^2 = 0%
−0.06
−0.02
0
0.02
0.04
0.06

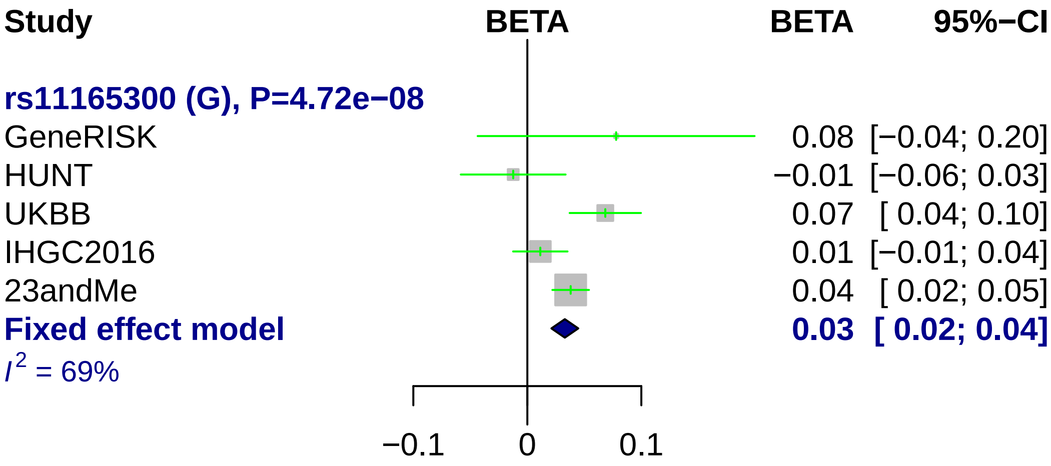

Study
BETA
BETA
95%−CI
rs11165300 (G), P=4.72e−08
GeneRISK 0.08 [−0.04; 0.20]
HUNT −0.01 [−0.06; 0.03]
UKBB 0.07 [ 0.04; 0.10]
IHGC2016 0.01 [−0.01; 0.04]
23andMe 0.04 [ 0.02; 0.05]
Fixed effect model 0.03 [ 0.02; 0.04]
I2 = 69%
−0.1
0
0.1

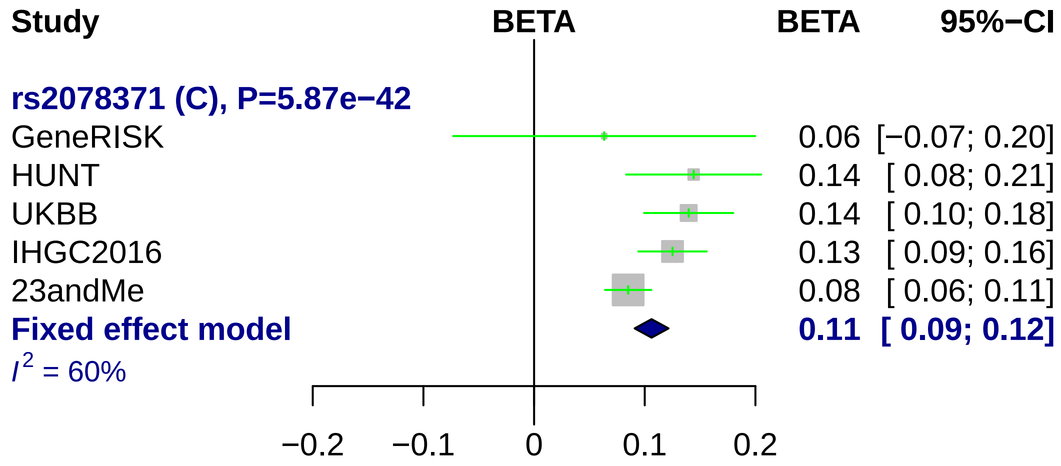
Study
BETA
BETA
95%−CI
rs2078371 (C), P=5.87e−42
GeneRISK
0.06 [−0.07; 0.20]
HUNT
0.14 [ 0.08; 0.21]
UKBB
0.14 [ 0.10; 0.18]
IHGC2016
0.13 [ 0.09; 0.16]
23andMe
0.08 [ 0.06; 0.11]
Fixed effect model
0.11 [ 0.09; 0.12]
I2 = 60%
−0.2
−0.1
0
0.1
0.2

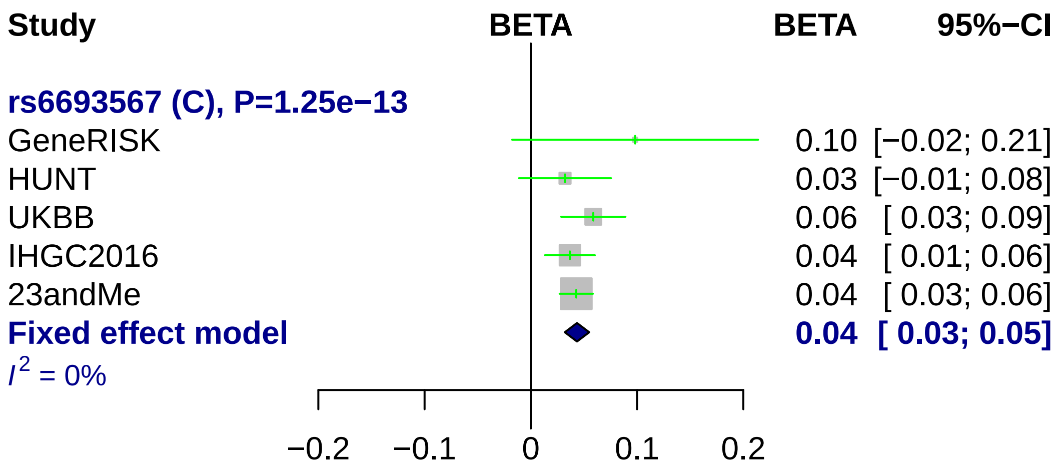
Study
BETA
BETA
95%−CI
rs6693567 (C), P=1.25e−13
GeneRISK 0.10 [−0.02; 0.21]
HUNT 0.03 [−0.01; 0.08]
UKBB 0.06 [ 0.03; 0.09]
IHGC2016 0.04 [ 0.01; 0.06]
23andMe 0.04 [ 0.03; 0.06]
Fixed effect model 0.04 [ 0.03; 0.05]
$I^2$ = 0%
−0.2
−0.1
0
0.1
0.2

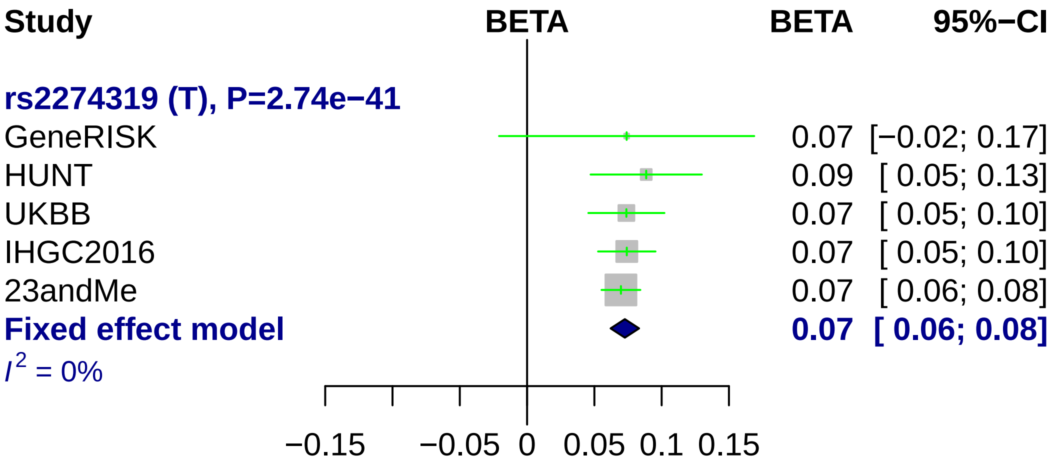
Study
BETA
BETA
95%−CI
rs2274319 (T), P=2.74e−41
GeneRISK
0.07 [−0.02; 0.17]
HUNT
0.09 [ 0.05; 0.13]
UKBB
0.07 [ 0.05; 0.10]
IHGC2016
0.07 [ 0.05; 0.10]
23andMe
0.07 [ 0.06; 0.08]
Fixed effect model
0.07 [ 0.06; 0.08]
I2 = 0%
−0.15
−0.05
0
0.05
0.1
0.15

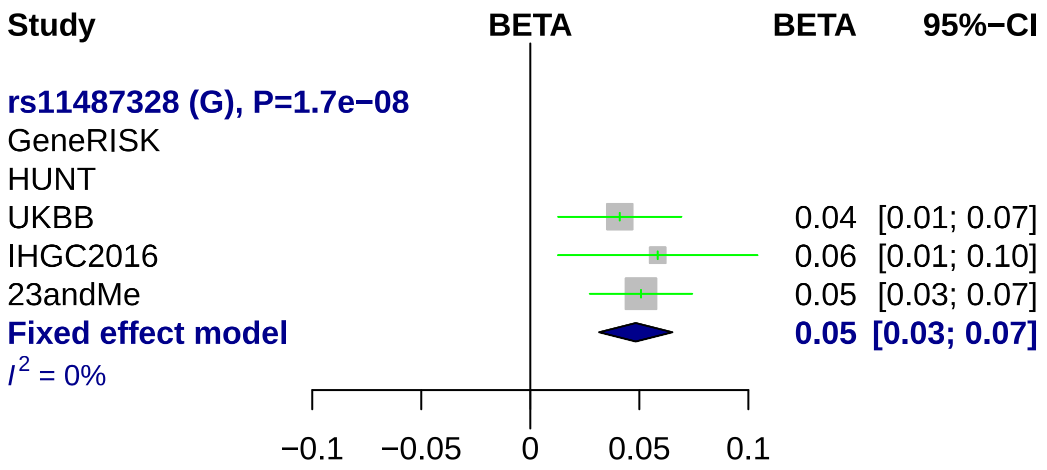
Study
BETA
BETA
95%−CI
rs11487328 (G), P=1.7e−08
GeneRISK
HUNT
UKBB
0.04 [0.01; 0.07]
IHGC2016
0.06 [0.01; 0.10]
23andMe
0.05 [0.03; 0.07]
Fixed effect model
0.05 [0.03; 0.07]
I^2 = 0%
−0.1
−0.05
0
0.05
0.1

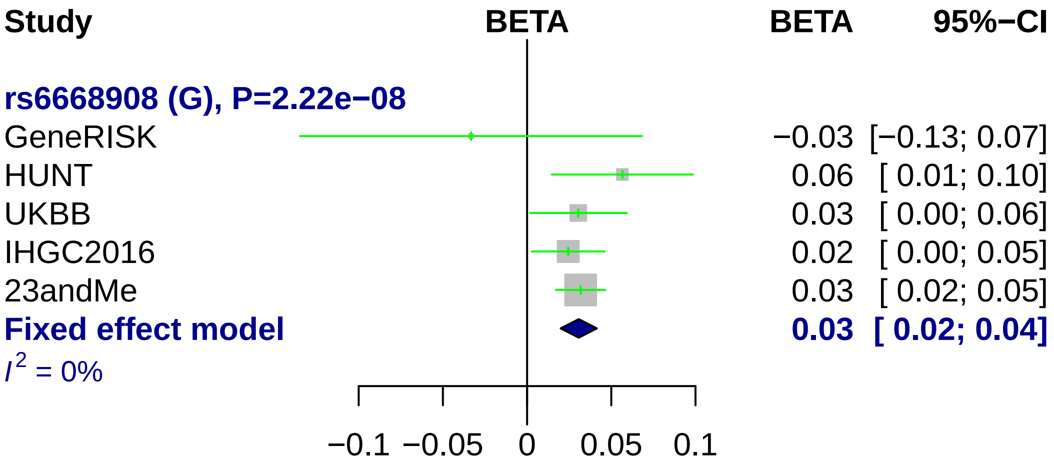
Study
BETA
BETA
95%−CI
rs6668908 (G), P=2.22e−08
GeneRISK
−0.03 [−0.13; 0.07]
HUNT
0.06 [ 0.01; 0.10]
UKBB
0.03 [ 0.00; 0.06]
IHGC2016
0.02 [ 0.00; 0.05]
23andMe
0.03 [ 0.02; 0.05]
Fixed effect model
0.03 [ 0.02; 0.04]
$I^2$ = 0%
−0.1
−0.05
0
0.05
0.1

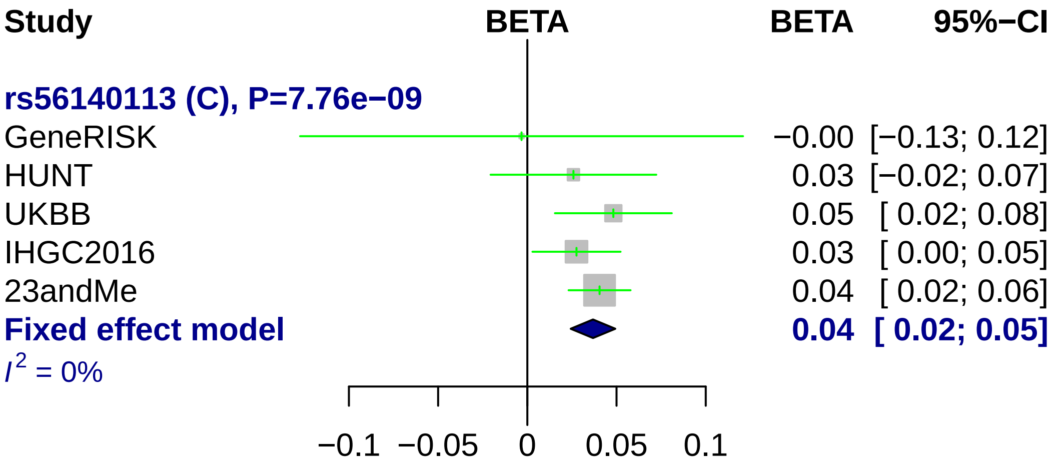

Study
BETA
BETA
95%−CI
rs56140113 (C), P=7.76e−09
GeneRISK
−0.00 [−0.13; 0.12]
HUNT
0.03 [−0.02; 0.07]
UKBB
0.05 [ 0.02; 0.08]
IHGC2016
0.03 [ 0.00; 0.05]
23andMe
0.04 [ 0.02; 0.06]
Fixed effect model
0.04 [ 0.02; 0.05]
I2 = 0%
−0.1
−0.05
0
0.05
0.1

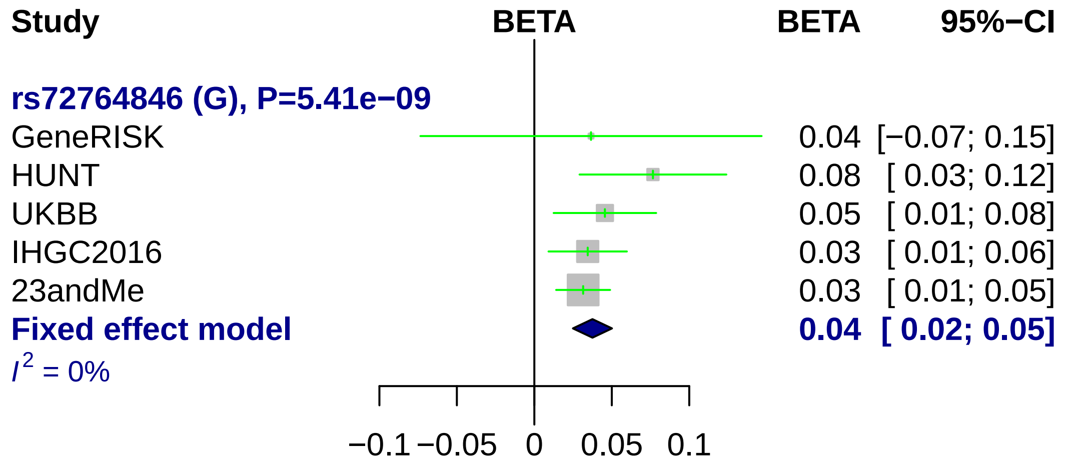

Study
BETA
BETA
95%−CI
rs72764846 (G), P=5.41e−09
GeneRISK
0.04 [−0.07; 0.15]
HUNT
0.08 [ 0.03; 0.12]
UKBB
0.05 [ 0.01; 0.08]
IHGC2016
0.03 [ 0.01; 0.06]
23andMe
0.03 [ 0.01; 0.05]
Fixed effect model
0.04 [ 0.02; 0.05]
I^2 = 0%
−0.1
−0.05
0
0.05
0.1

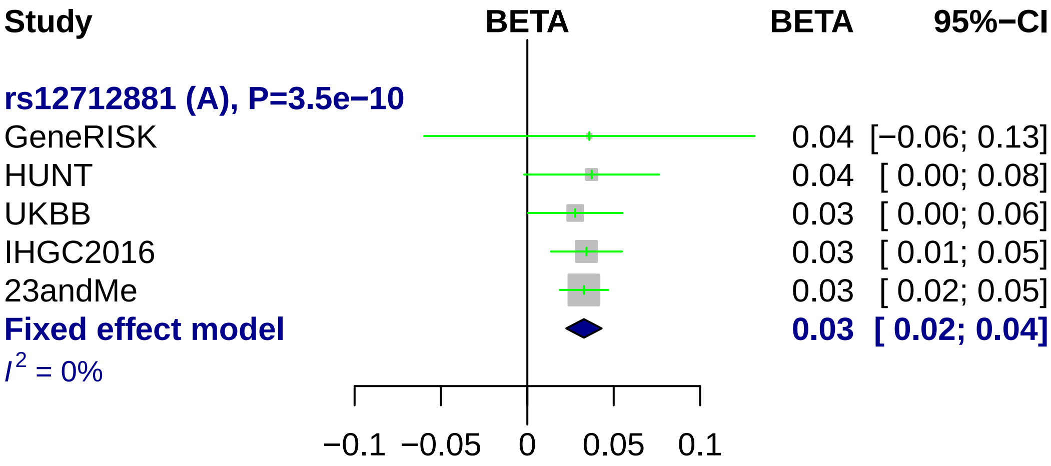
Study
BETA
BETA
95%−CI
rs12712881 (A), P=3.5e−10
GeneRISK
0.04 [−0.06; 0.13]
HUNT
0.04 [ 0.00; 0.08]
UKBB
0.03 [ 0.00; 0.06]
IHGC2016
0.03 [ 0.01; 0.05]
23andMe
0.03 [ 0.02; 0.05]
Fixed effect model
0.03 [ 0.02; 0.04]
I^2 = 0%
−0.1
−0.05
0
0.05
0.1

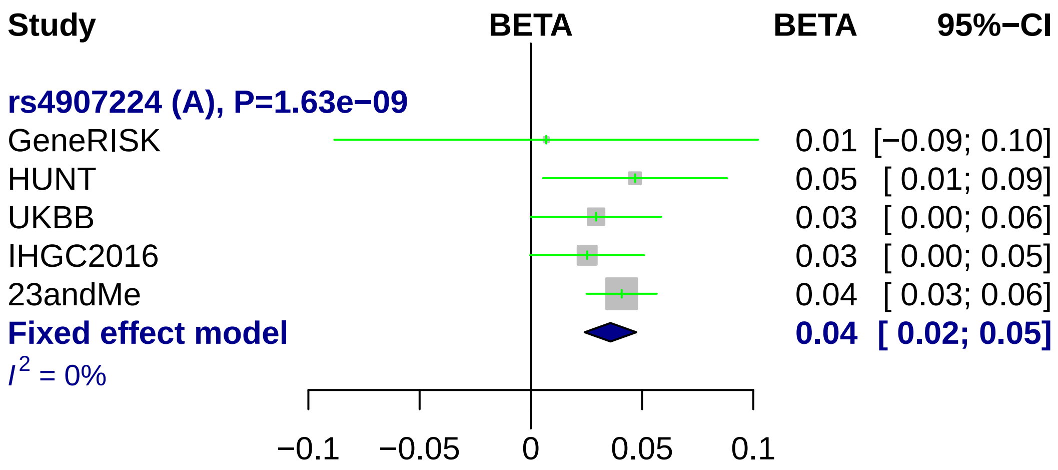
Study
BETA
BETA
95%−CI
rs4907224 (A), P=1.63e−09
GeneRISK
0.01
[−0.09; 0.10]
HUNT
0.05
[ 0.01; 0.09]
UKBB
0.03
[ 0.00; 0.06]
IHGC2016
0.03
[ 0.00; 0.05]
23andMe
0.04
[ 0.03; 0.06]
Fixed effect model
0.04
[ 0.02; 0.05]
I2 = 0%
−0.1
−0.05
0
0.05
0.1

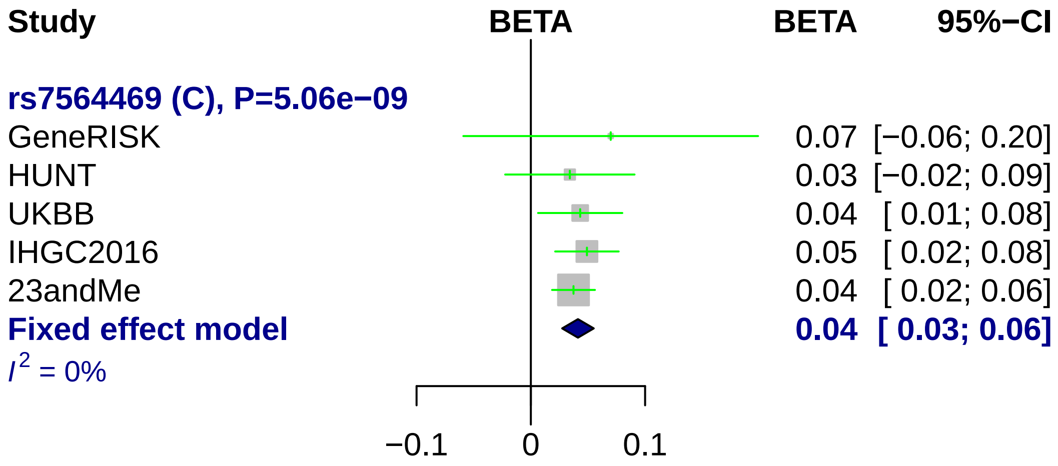
Study
BETA
BETA
95%−CI
rs7564469 (C), P=5.06e−09
GeneRISK
0.07 [−0.06; 0.20]
HUNT
0.03 [−0.02; 0.09]
UKBB
0.04 [ 0.01; 0.08]
IHGC2016
0.05 [ 0.02; 0.08]
23andMe
0.04 [ 0.02; 0.06]
Fixed effect model
0.04 [ 0.03; 0.06]
$I^2$ = 0%
−0.1
0
0.1

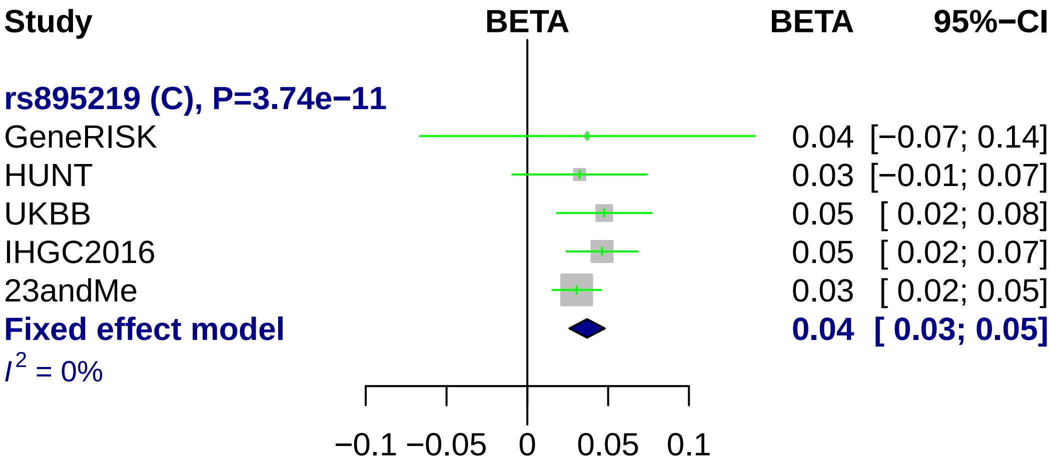

Study
BETA
BETA
95%−CI
rs895219 (C), P=3.74e−11
GeneRISK 0.04 [−0.07; 0.14]
HUNT 0.03 [−0.01; 0.07]
UKBB 0.05 [ 0.02; 0.08]
IHGC2016 0.05 [ 0.02; 0.07]
23andMe 0.03 [ 0.02; 0.05]
Fixed effect model 0.04 [ 0.03; 0.05]
I2 = 0%
−0.1 −0.05 0 0.05 0.1

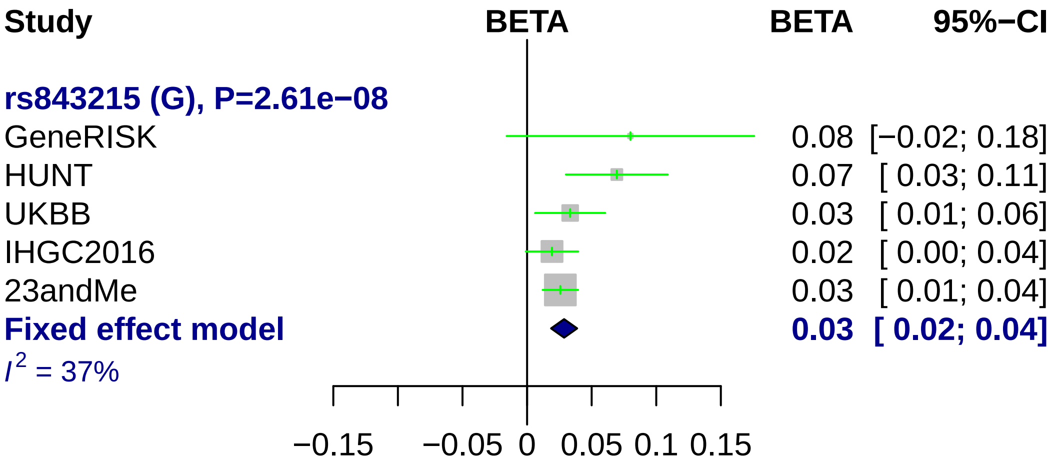

Study
BETA
BETA
95%−CI
rs843215 (G), P=2.61e−08
GeneRISK
0.08 [−0.02; 0.18]
HUNT
0.07 [ 0.03; 0.11]
UKBB
0.03 [ 0.01; 0.06]
IHGC2016
0.02 [ 0.00; 0.04]
23andMe
0.03 [ 0.01; 0.04]
Fixed effect model
0.03 [ 0.02; 0.04]
I² = 37%
−0.15
−0.05
0
0.05
0.1
0.15

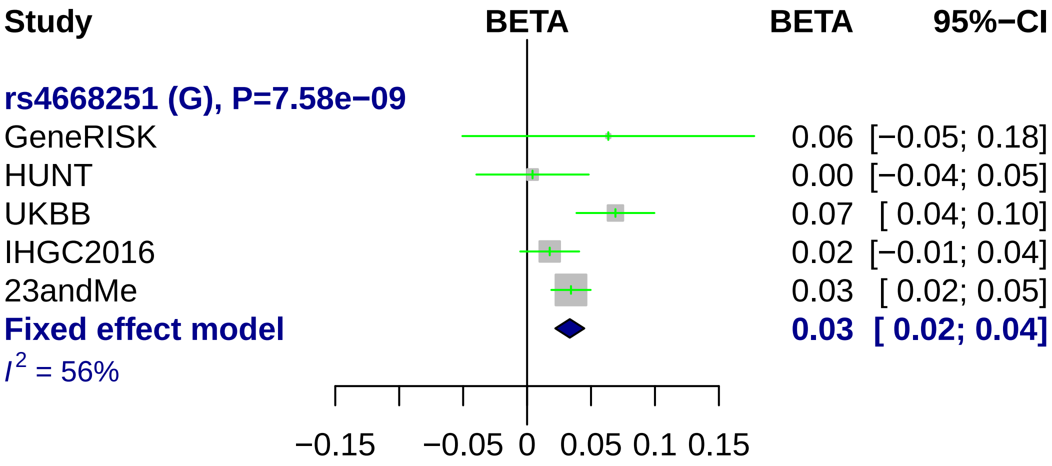
Study
BETA
BETA
95%−CI
rs4668251 (G), P=7.58e−09
GeneRISK
0.06 [−0.05; 0.18]
HUNT
0.00 [−0.04; 0.05]
UKBB
0.07 [ 0.04; 0.10]
IHGC2016
0.02 [−0.01; 0.04]
23andMe
0.03 [ 0.02; 0.05]
Fixed effect model
0.03 [ 0.02; 0.04]
I^2 = 56%
−0.15
−0.05
0
0.05
0.1
0.15

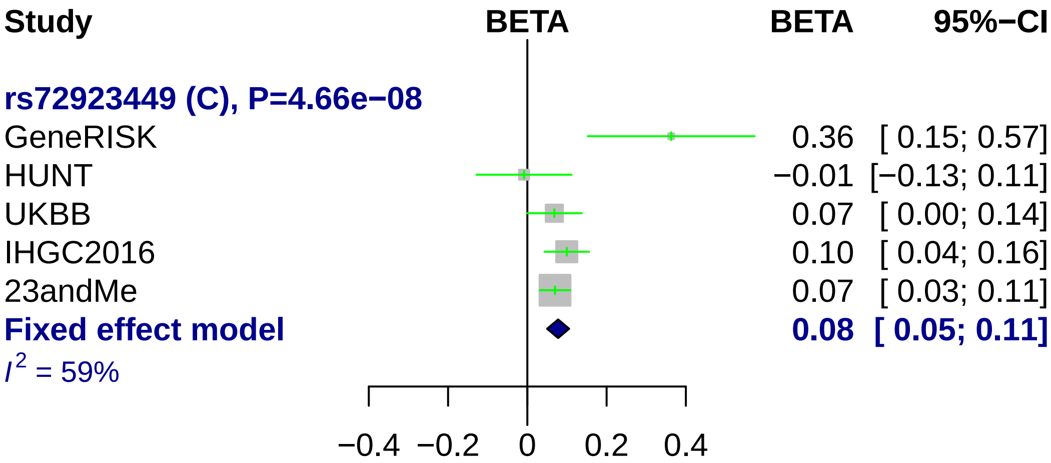
Study
BETA
BETA
95%−CI
rs72923449 (C), P=4.66e−08
GeneRISK
0.36 [ 0.15; 0.57]
HUNT
−0.01 [−0.13; 0.11]
UKBB
0.07 [ 0.00; 0.14]
IHGC2016
0.10 [ 0.04; 0.16]
23andMe
0.07 [ 0.03; 0.11]
Fixed effect model
0.08 [ 0.05; 0.11]
$I^2$ = 59%
−0.4
−0.2
0
0.2
0.4

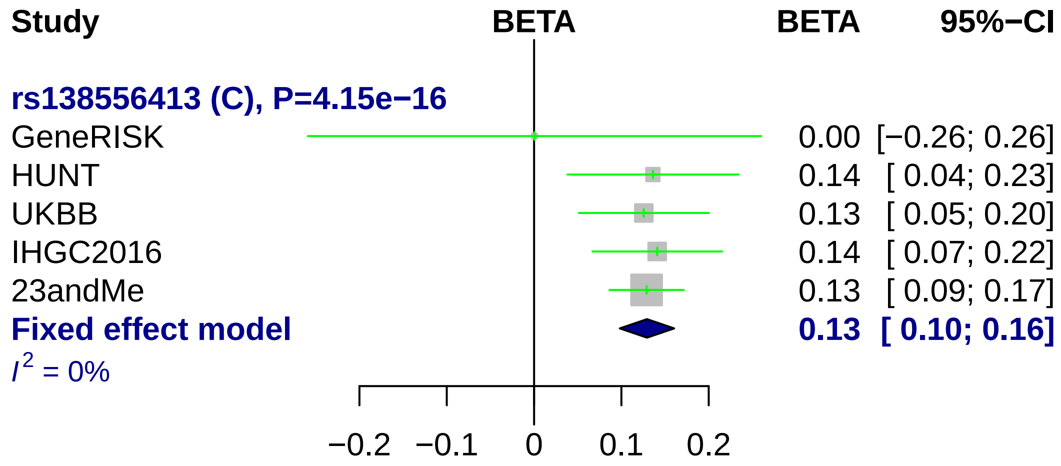
Study
BETA
BETA
95%−CI
rs138556413 (C), P=4.15e−16
GeneRISK 0.00 [−0.26; 0.26]
HUNT 0.14 [ 0.04; 0.23]
UKBB 0.13 [ 0.05; 0.20]
IHGC2016 0.14 [ 0.07; 0.22]
23andMe 0.13 [ 0.09; 0.17]
Fixed effect model 0.13 [ 0.10; 0.16]
I2 = 0%
−0.2
−0.1
0
0.1
0.2

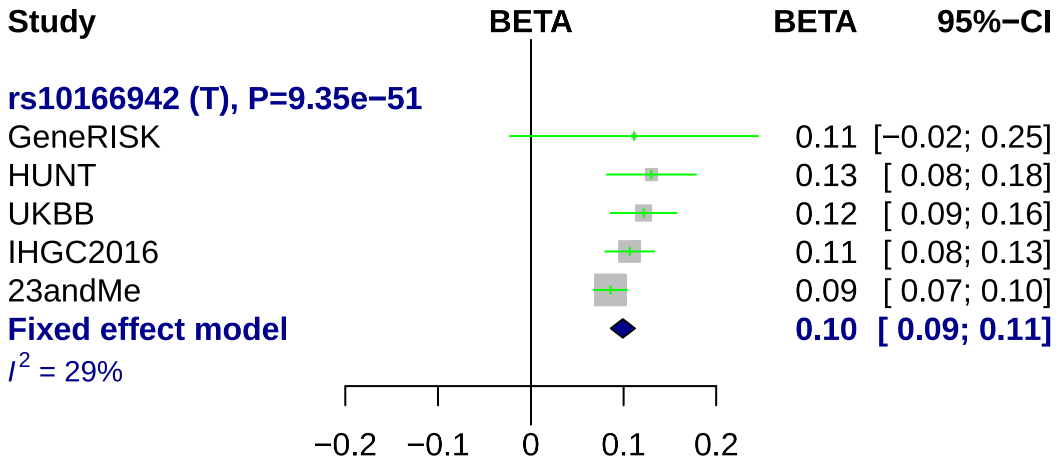
Study
BETA
BETA
95%−CI
rs10166942 (T), P=9.35e−51
GeneRISK
HUNT
UKBB
IHGC2016
23andMe
Fixed effect model
$I^2$ = 29%
0.11 [−0.02; 0.25]
0.13 [ 0.08; 0.18]
0.12 [ 0.09; 0.16]
0.11 [ 0.08; 0.13]
0.09 [ 0.07; 0.10]
0.10 [ 0.09; 0.11]
−0.2
−0.1
0
0.1
0.2

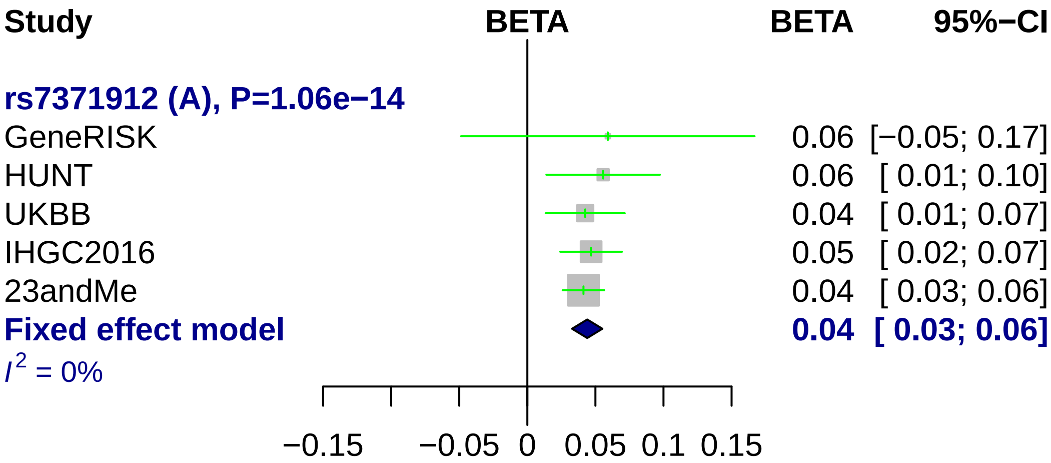

Study
BETA
BETA
95%−CI
rs7371912 (A), P=1.06e−14
GeneRISK 0.06 [−0.05; 0.17]
HUNT 0.06 [ 0.01; 0.10]
UKBB 0.04 [ 0.01; 0.07]
IHGC2016 0.05 [ 0.02; 0.07]
23andMe 0.04 [ 0.03; 0.06]
Fixed effect model 0.04 [ 0.03; 0.06]
I² = 0%
−0.15
−0.05
0
0.05
0.1
0.15

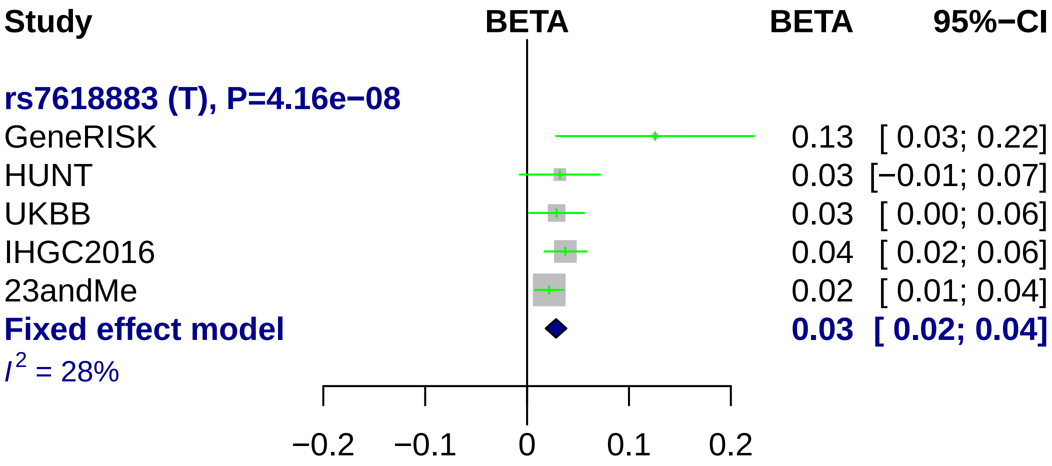

Study
BETA
BETA
95%−CI
rs7618883 (T), P=4.16e−08
GeneRISK
0.13 [ 0.03; 0.22]
HUNT
0.03 [−0.01; 0.07]
UKBB
0.03 [ 0.00; 0.06]
IHGC2016
0.04 [ 0.02; 0.06]
23andMe
0.02 [ 0.01; 0.04]
Fixed effect model
0.03 [ 0.02; 0.04]
I2 = 28%
−0.2
−0.1
0
0.1
0.2

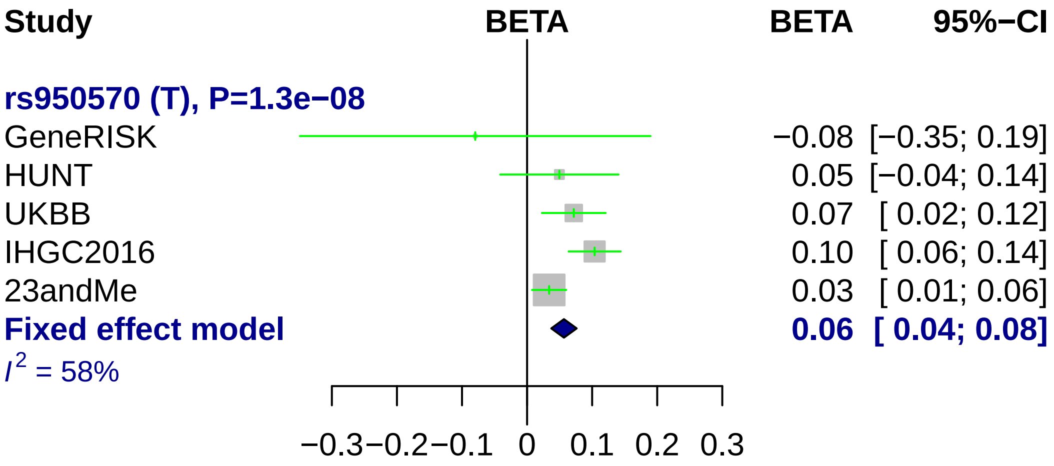
Study
BETA
BETA
95%−CI
rs950570 (T), P=1.3e−08
GeneRISK
−0.08 [−0.35; 0.19]
HUNT
0.05 [−0.04; 0.14]
UKBB
0.07 [ 0.02; 0.12]
IHGC2016
0.10 [ 0.06; 0.14]
23andMe
0.03 [ 0.01; 0.06]
Fixed effect model
0.06 [ 0.04; 0.08]
I2 = 58%
−0.3
−0.2
−0.1
0
0.1
0.2
0.3

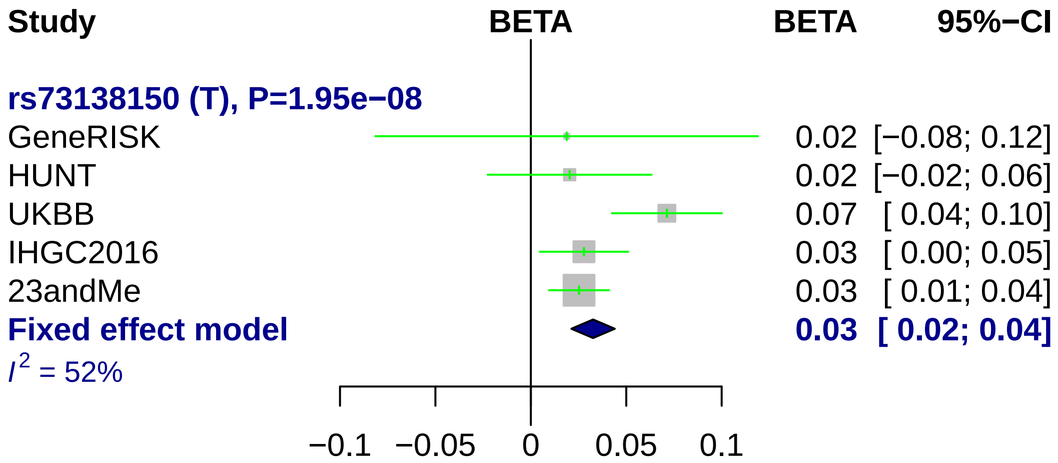

Study
BETA
BETA
95%−CI
rs73138150 (T), P=1.95e−08
GeneRISK 0.02 [−0.08; 0.12]
HUNT 0.02 [−0.02; 0.06]
UKBB 0.07 [ 0.04; 0.10]
IHGC2016 0.03 [ 0.00; 0.05]
23andMe 0.03 [ 0.01; 0.04]
Fixed effect model 0.03 [ 0.02; 0.04]
I^2 = 52%
−0.1
−0.05
0
0.05
0.1

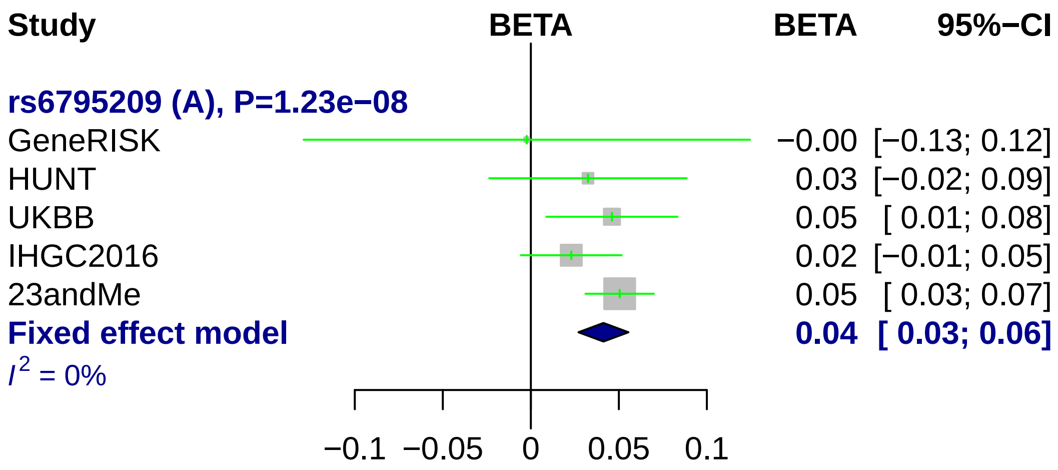
Study
BETA
BETA
95%−CI
rs6795209 (A), P=1.23e−08
GeneRISK
−0.00 [−0.13; 0.12]
HUNT
0.03 [−0.02; 0.09]
UKBB
0.05 [ 0.01; 0.08]
IHGC2016
0.02 [−0.01; 0.05]
23andMe
0.05 [ 0.03; 0.07]
Fixed effect model
0.04 [ 0.03; 0.06]
$I^2$ = 0%
−0.1
−0.05
0
0.05
0.1

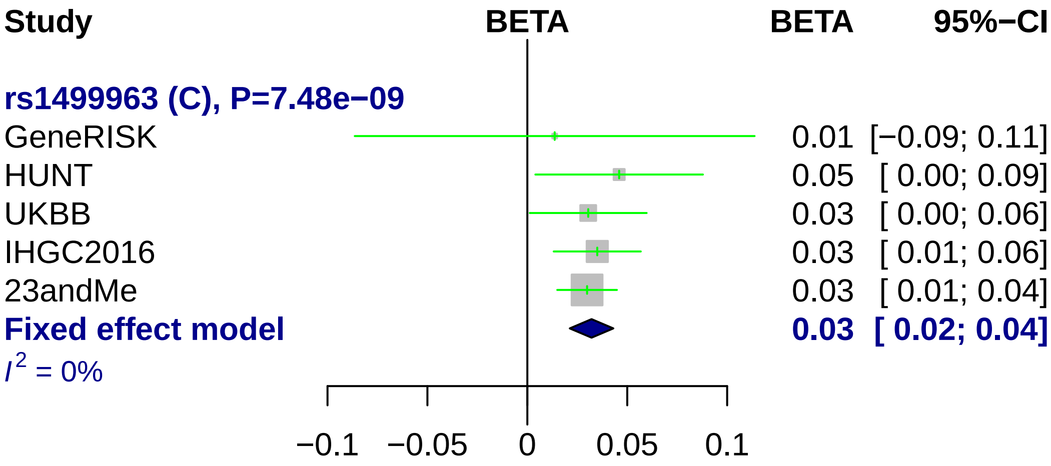
Study
BETA
BETA
95%−CI
rs1499963 (C), P=7.48e−09
GeneRISK
0.01 [−0.09; 0.11]
HUNT
0.05 [ 0.00; 0.09]
UKBB
0.03 [ 0.00; 0.06]
IHGC2016
0.03 [ 0.01; 0.06]
23andMe
0.03 [ 0.01; 0.04]
Fixed effect model
0.03 [ 0.02; 0.04]
I^2 = 0%
−0.1
−0.05
0
0.05
0.1

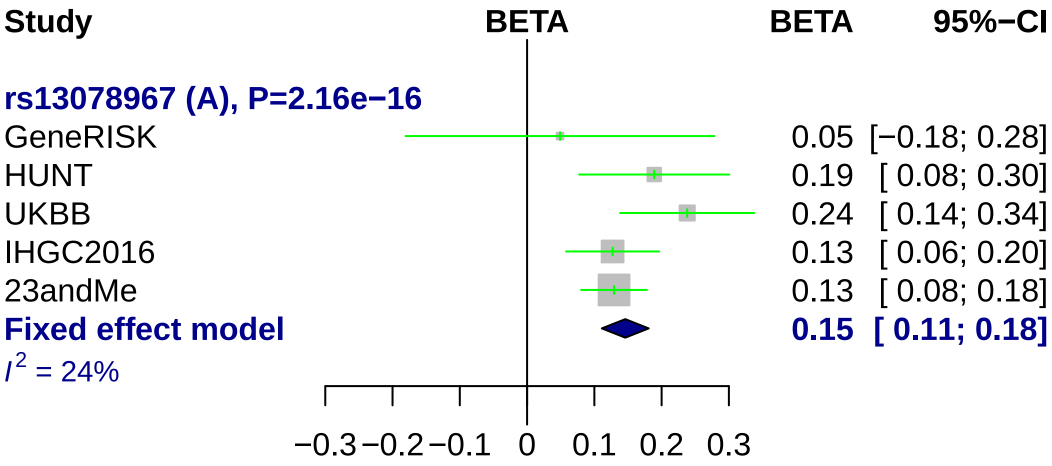

Study
BETA
BETA
95%−CI
rs13078967 (A), P=2.16e−16
GeneRISK
0.05 [−0.18; 0.28]
HUNT
0.19 [ 0.08; 0.30]
UKBB
0.24 [ 0.14; 0.34]
IHGC2016
0.13 [ 0.06; 0.20]
23andMe
0.13 [ 0.08; 0.18]
Fixed effect model
0.15 [ 0.11; 0.18]
$I^2$ = 24%
−0.3 −0.2 −0.1 0 0.1 0.2 0.3

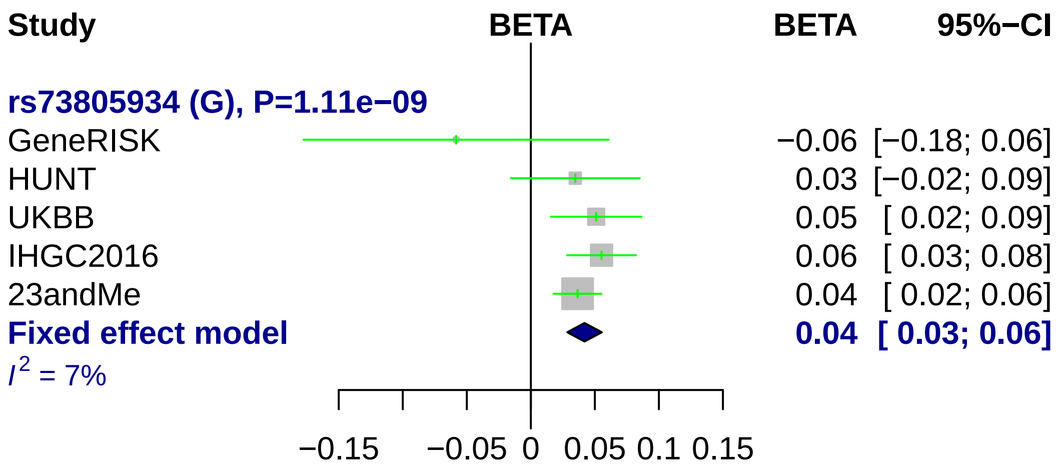
Study
BETA
BETA
95%−CI
rs73805934 (G), P=1.11e−09
GeneRISK −0.06 [−0.18; 0.06]
HUNT 0.03 [−0.02; 0.09]
UKBB 0.05 [ 0.02; 0.09]
IHGC2016 0.06 [ 0.03; 0.08]
23andMe 0.04 [ 0.02; 0.06]
Fixed effect model 0.04 [ 0.03; 0.06]
I^2 = 7%
−0.15 −0.05 0 0.05 0.1 0.15

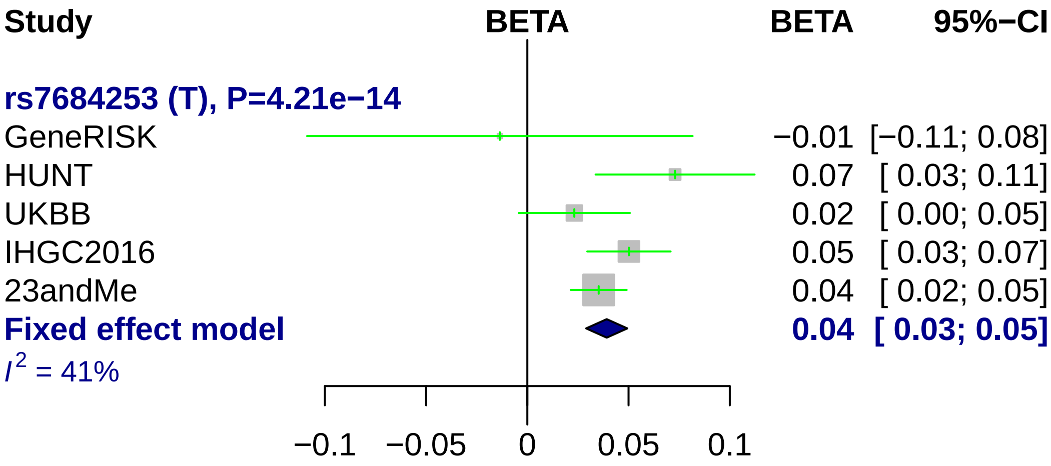
Study
BETA
BETA
95%−CI
rs7684253 (T), P=4.21e−14
GeneRISK −0.01 [−0.11; 0.08]
HUNT 0.07 [ 0.03; 0.11]
UKBB 0.02 [ 0.00; 0.05]
IHGC2016 0.05 [ 0.03; 0.07]
23andMe 0.04 [ 0.02; 0.05]
Fixed effect model 0.04 [ 0.03; 0.05]
I2 = 41%
−0.1
−0.05
0
0.05
0.1

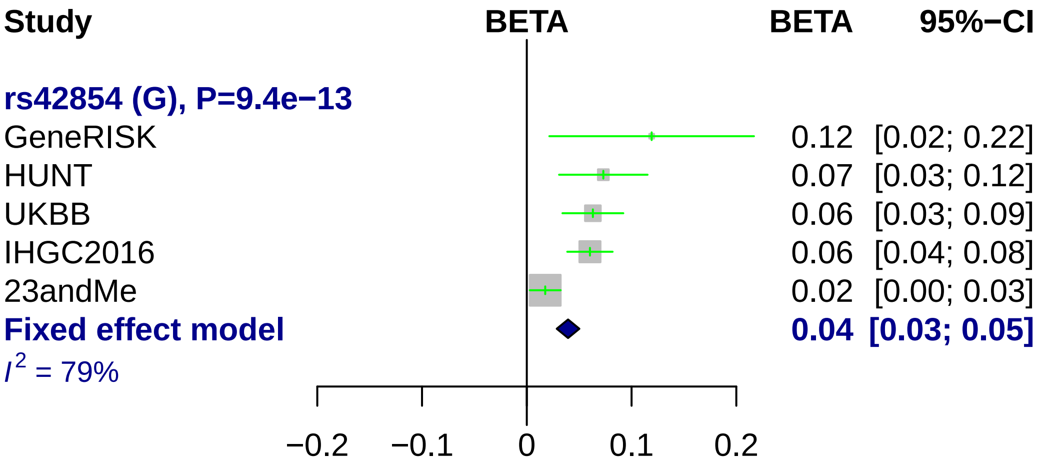
Study
BETA
BETA
95%−CI
rs42854 (G), P=9.4e−13
GeneRISK
0.12 [0.02; 0.22]
HUNT
0.07 [0.03; 0.12]
UKBB
0.06 [0.03; 0.09]
IHGC2016
0.06 [0.04; 0.08]
23andMe
0.02 [0.00; 0.03]
Fixed effect model
0.04 [0.03; 0.05]
$I^2$ = 79%
−0.2
−0.1
0
0.1
0.2

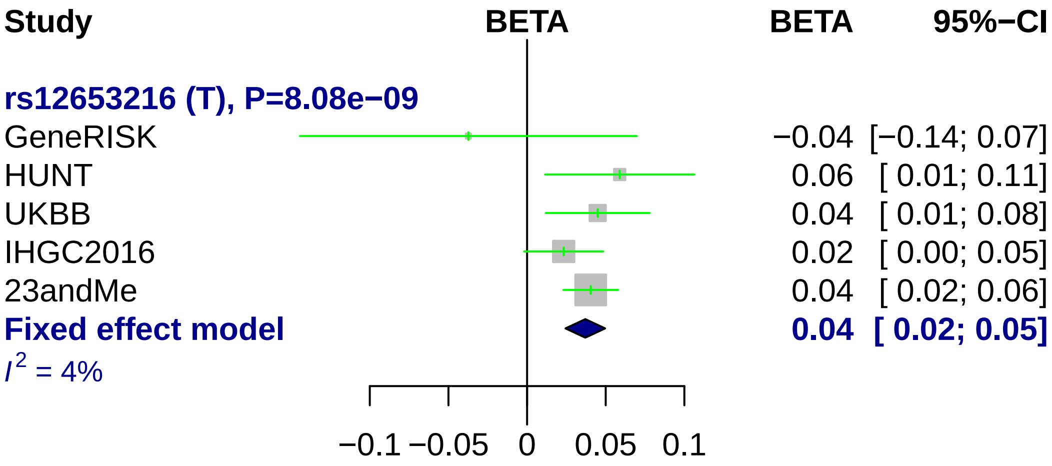
Study
BETA
BETA
95%−CI
rs12653216 (T), P=8.08e−09
GeneRISK −0.04 [−0.14; 0.07]
HUNT 0.06 [ 0.01; 0.11]
UKBB 0.04 [ 0.01; 0.08]
IHGC2016 0.02 [ 0.00; 0.05]
23andMe 0.04 [ 0.02; 0.06]
Fixed effect model 0.04 [ 0.02; 0.05]
$I^2$ = 4%
−0.1 −0.05 0 0.05 0.1

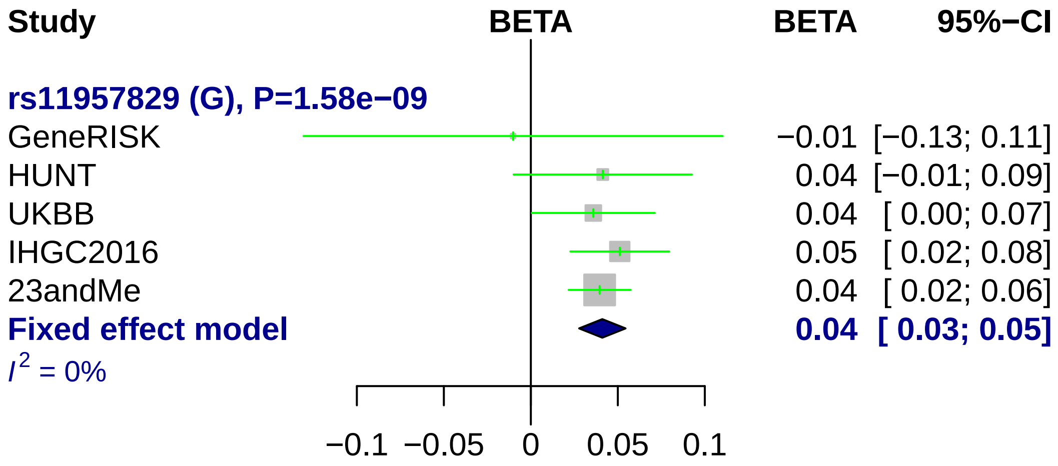
Study
BETA
BETA
95%−CI
rs11957829 (G), P=1.58e−09
GeneRISK −0.01 [−0.13; 0.11]
HUNT 0.04 [−0.01; 0.09]
UKBB 0.04 [ 0.00; 0.07]
IHGC2016 0.05 [ 0.02; 0.08]
23andMe 0.04 [ 0.02; 0.06]
Fixed effect model 0.04 [ 0.03; 0.05]
I2 = 0%
−0.1
−0.05
0
0.05
0.1

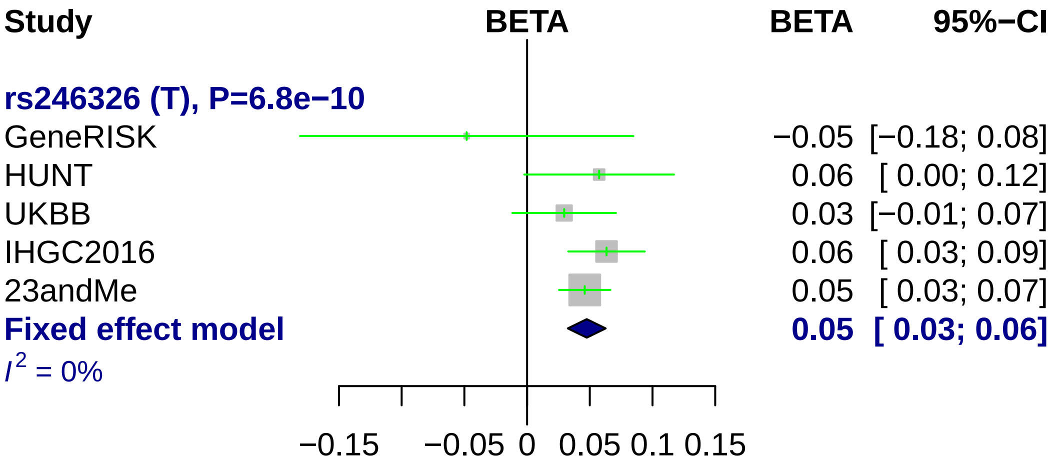
Study
BETA
BETA
95%−CI
rs246326 (T), P=6.8e−10
GeneRISK −0.05 [−0.18; 0.08]
HUNT 0.06 [ 0.00; 0.12]
UKBB 0.03 [−0.01; 0.07]
IHGC2016 0.06 [ 0.03; 0.09]
23andMe 0.05 [ 0.03; 0.07]
Fixed effect model 0.05 [ 0.03; 0.06]
I^2 = 0%
−0.15
−0.05
0
0.05
0.1
0.15

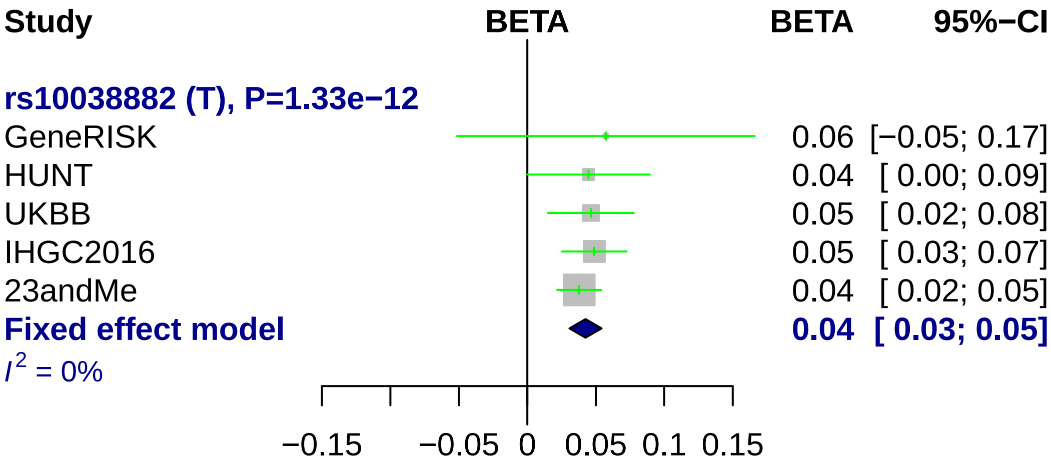
Study
BETA
BETA
95%−CI
rs10038882 (T), P=1.33e−12
GeneRISK
0.06 [−0.05; 0.17]
HUNT
0.04 [ 0.00; 0.09]
UKBB
0.05 [ 0.02; 0.08]
IHGC2016
0.05 [ 0.03; 0.07]
23andMe
0.04 [ 0.02; 0.05]
Fixed effect model
0.04 [ 0.03; 0.05]
I2 = 0%
−0.15
−0.05
0
0.05
0.1
0.15

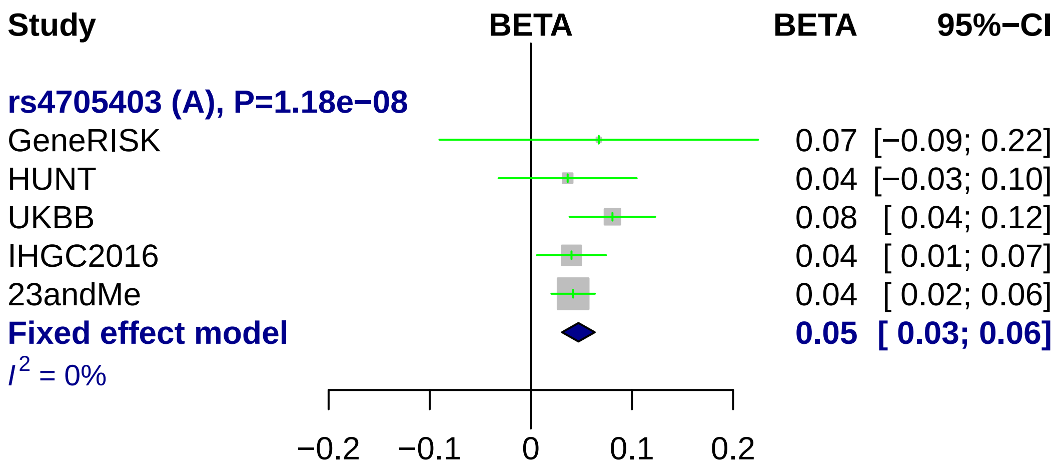
Study
BETA
BETA
95%−CI
rs4705403 (A), P=1.18e−08
GeneRISK
0.07 [−0.09; 0.22]
HUNT
0.04 [−0.03; 0.10]
UKBB
0.08 [ 0.04; 0.12]
IHGC2016
0.04 [ 0.01; 0.07]
23andMe
0.04 [ 0.02; 0.06]
Fixed effect model
0.05 [ 0.03; 0.06]
I 2 = 0%
−0.2
−0.1
0
0.1
0.2

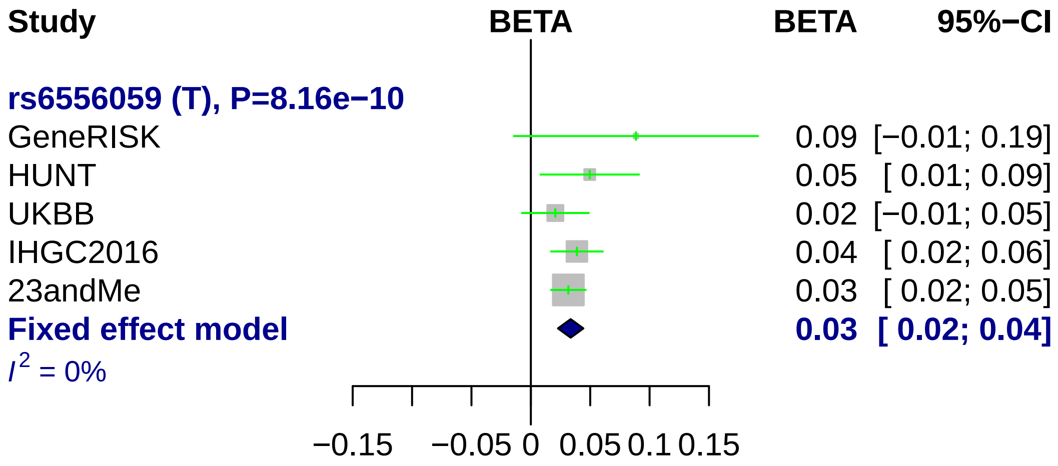
Study
BETA
BETA
95%−CI
rs6556059 (T), P=8.16e−10
GeneRISK
0.09 [−0.01; 0.19]
HUNT
0.05 [ 0.01; 0.09]
UKBB
0.02 [−0.01; 0.05]
IHGC2016
0.04 [ 0.02; 0.06]
23andMe
0.03 [ 0.02; 0.05]
Fixed effect model
0.03 [ 0.02; 0.04]
I2 = 0%
−0.15
−0.05
0
0.05
0.1
0.15

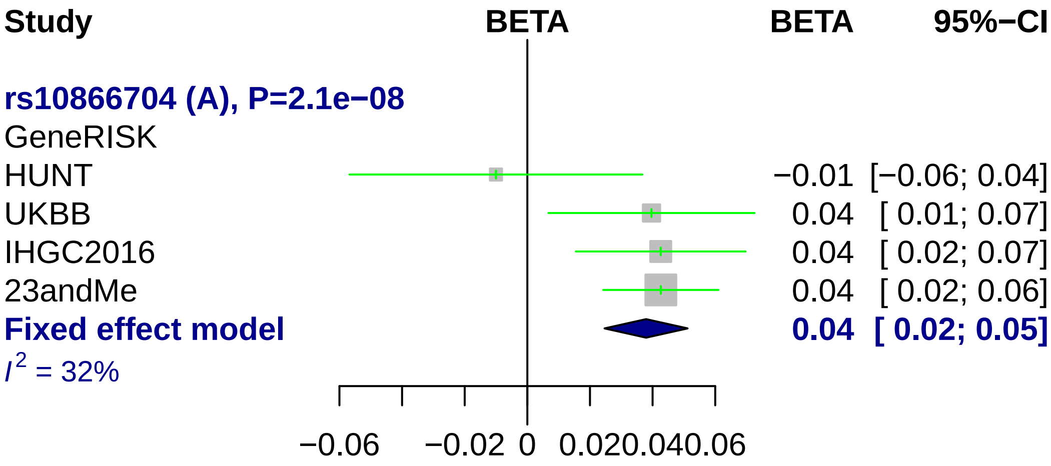

Study
BETA
BETA
95%−CI
rs10866704 (A), P=2.1e−08
GeneRISK
HUNT
−0.01 [−0.06; 0.04]
UKBB
0.04 [ 0.01; 0.07]
IHGC2016
0.04 [ 0.02; 0.07]
23andMe
0.04 [ 0.02; 0.06]
Fixed effect model
0.04 [ 0.02; 0.05]
I2 = 32%
−0.06
−0.02
0
0.02
0.04
0.06

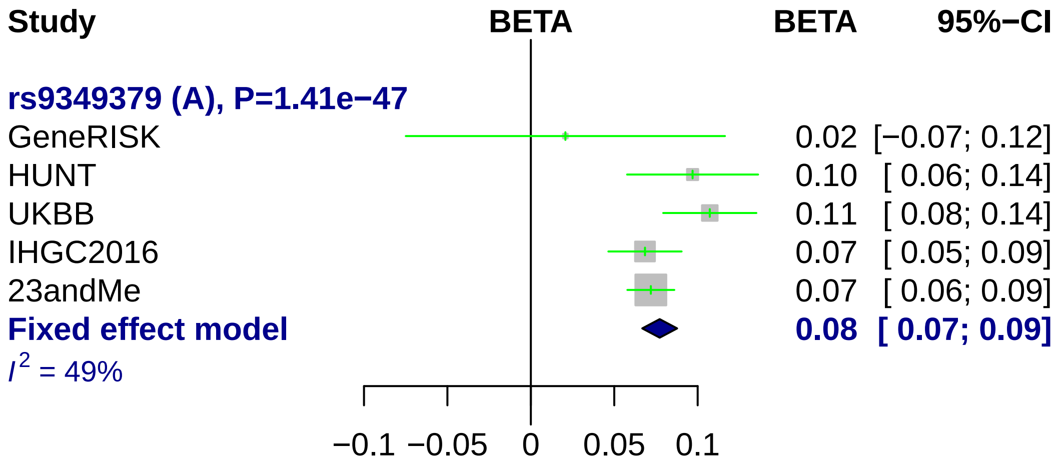
Study
BETA
BETA
95%−CI
rs9349379 (A), P=1.41e−47
GeneRISK 0.02 [−0.07; 0.12]
HUNT 0.10 [ 0.06; 0.14]
UKBB 0.11 [ 0.08; 0.14]
IHGC2016 0.07 [ 0.05; 0.09]
23andMe 0.07 [ 0.06; 0.09]
Fixed effect model 0.08 [ 0.07; 0.09]
$I^2$ = 49%
−0.1 −0.05 0 0.05 0.1

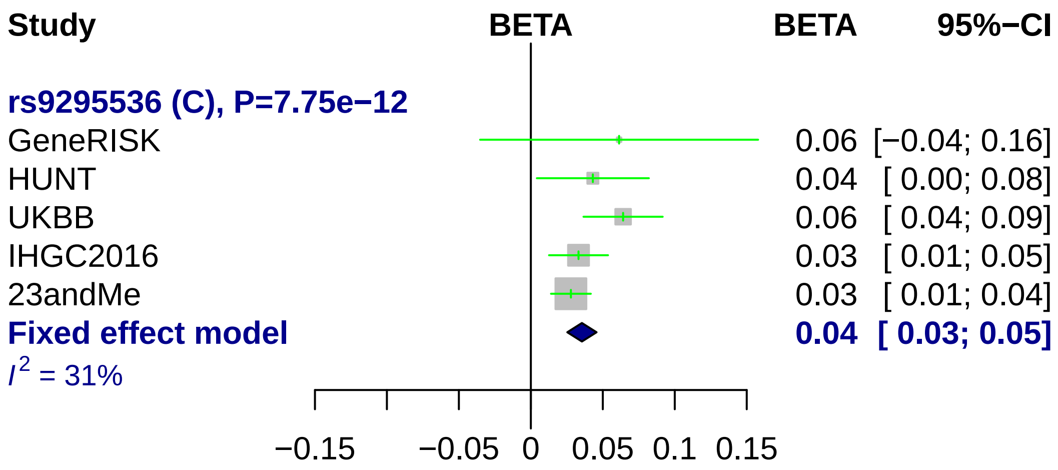
Study
BETA
BETA
95%−CI
rs9295536 (C), P=7.75e−12
GeneRISK
0.06 [−0.04; 0.16]
HUNT
0.04 [ 0.00; 0.08]
UKBB
0.06 [ 0.04; 0.09]
IHGC2016
0.03 [ 0.01; 0.05]
23andMe
0.03 [ 0.01; 0.04]
Fixed effect model
0.04 [ 0.03; 0.05]
I2 = 31%
−0.15
−0.05
0
0.05
0.1
0.15

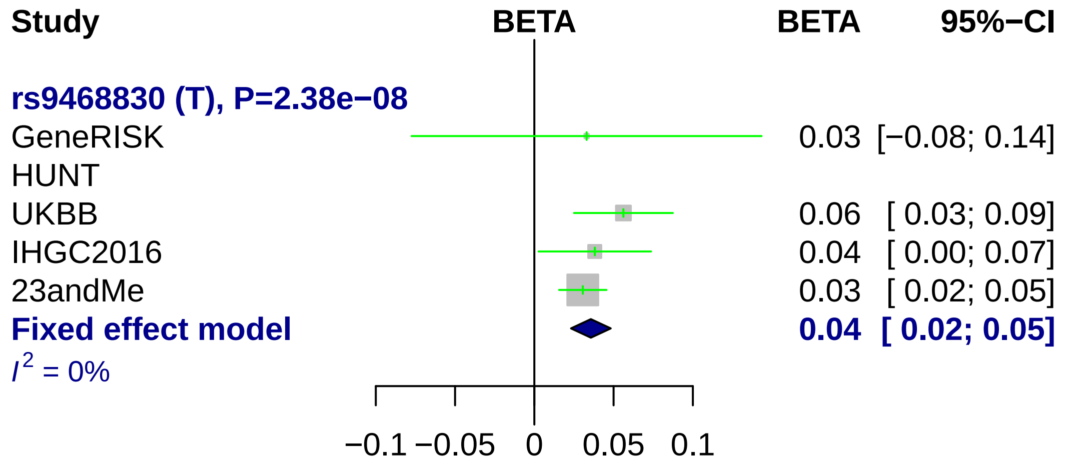
Study
BETA
BETA
95%−CI
rs9468830 (T), P=2.38e−08
GeneRISK 0.03 [−0.08; 0.14]
HUNT
UKBB 0.06 [ 0.03; 0.09]
IHGC2016 0.04 [ 0.00; 0.07]
23andMe 0.03 [ 0.02; 0.05]
Fixed effect model 0.04 [ 0.02; 0.05]
I2 = 0%
−0.1 −0.05 0 0.05 0.1

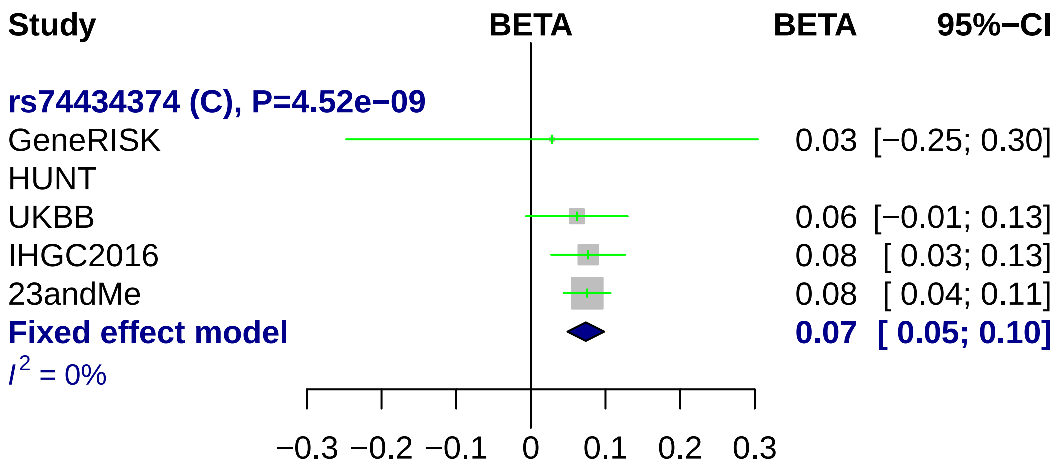
Study
BETA
BETA
95%−CI
rs74434374 (C), P=4.52e−09
GeneRISK
0.03 [−0.25; 0.30]
HUNT
UKBB
0.06 [−0.01; 0.13]
IHGC2016
0.08 [ 0.03; 0.13]
23andMe
0.08 [ 0.04; 0.11]
Fixed effect model
0.07 [ 0.05; 0.10]
I2 = 0%
−0.3 −0.2 −0.1 0 0.1 0.2 0.3

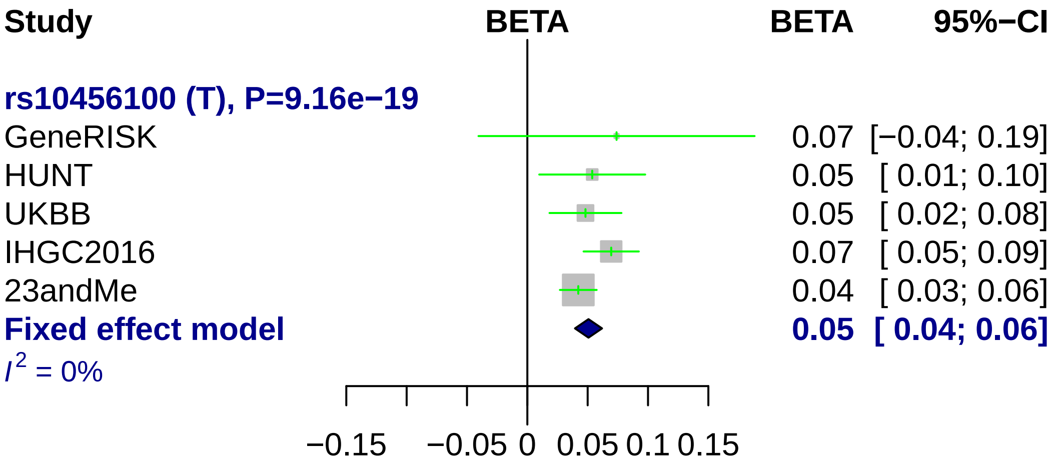

Study
BETA
BETA
95%−CI
rs10456100 (T), P=9.16e−19
GeneRISK
0.07 [−0.04; 0.19]
HUNT
0.05 [ 0.01; 0.10]
UKBB
0.05 [ 0.02; 0.08]
IHGC2016
0.07 [ 0.05; 0.09]
23andMe
0.04 [ 0.03; 0.06]
Fixed effect model
0.05 [ 0.04; 0.06]
I2 = 0%
−0.15
−0.05
0
0.05
0.1
0.15

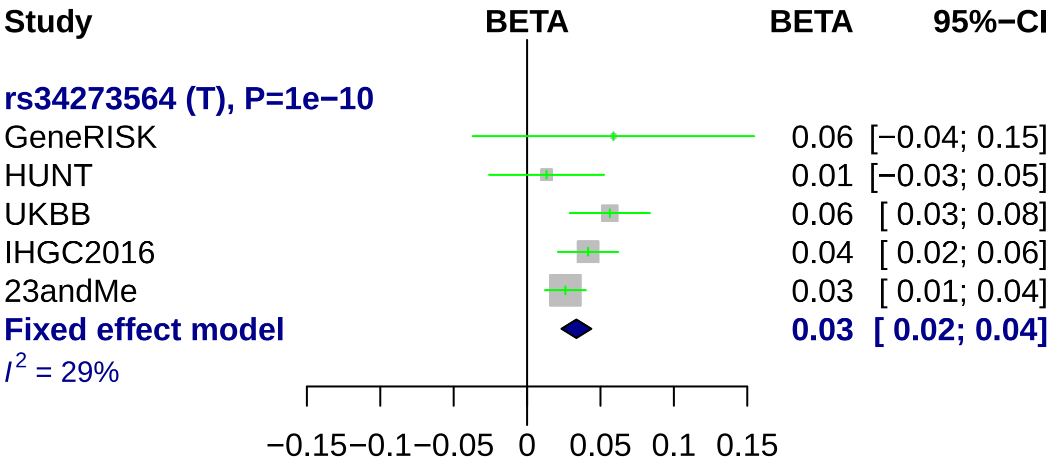
Study
BETA
BETA
95%−CI
rs34273564 (T), P=1e−10
GeneRISK
0.06 [−0.04; 0.15]
HUNT
0.01 [−0.03; 0.05]
UKBB
0.06 [ 0.03; 0.08]
IHGC2016
0.04 [ 0.02; 0.06]
23andMe
0.03 [ 0.01; 0.04]
Fixed effect model
0.03 [ 0.02; 0.04]
I2 = 29%
−0.15
−0.1
−0.05
0
0.05
0.1
0.15

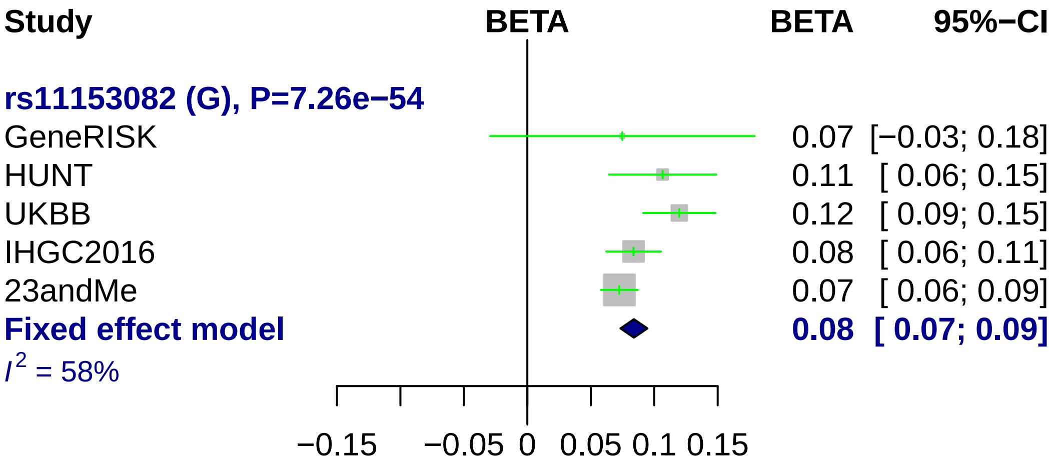
Study
BETA
BETA
95%−CI
rs11153082 (G), P=7.26e−54
GeneRISK 0.07 [−0.03; 0.18]
HUNT 0.11 [ 0.06; 0.15]
UKBB 0.12 [ 0.09; 0.15]
IHGC2016 0.08 [ 0.06; 0.11]
23andMe 0.07 [ 0.06; 0.09]
Fixed effect model 0.08 [ 0.07; 0.09]
I2 = 58%
−0.15
−0.05
0
0.05
0.1
0.15

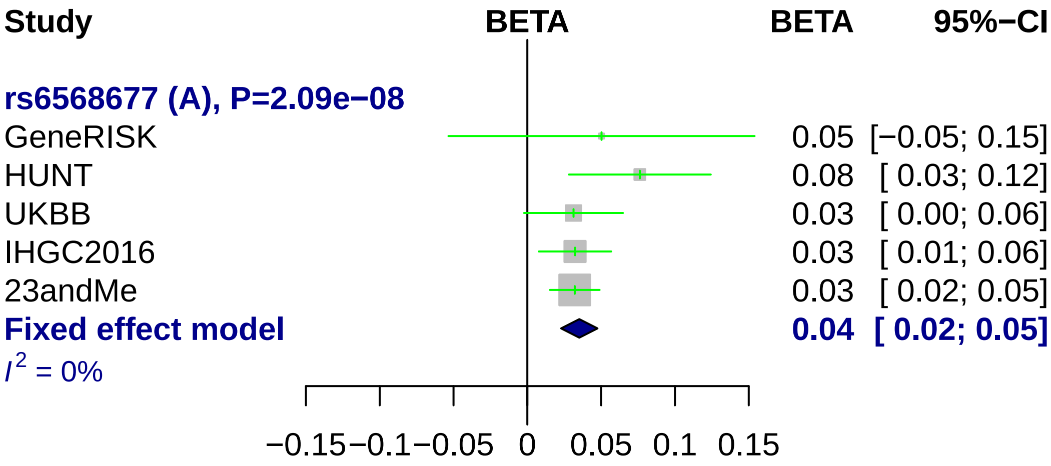
Study
BETA
BETA
95%−CI
rs6568677 (A), P=2.09e−08
GeneRISK
0.05 [−0.05; 0.15]
HUNT
0.08 [ 0.03; 0.12]
UKBB
0.03 [ 0.00; 0.06]
IHGC2016
0.03 [ 0.01; 0.06]
23andMe
0.03 [ 0.02; 0.05]
Fixed effect model
0.04 [ 0.02; 0.05]
$I^2$ = 0%
−0.15 −0.1 −0.05 0 0.05 0.1 0.15

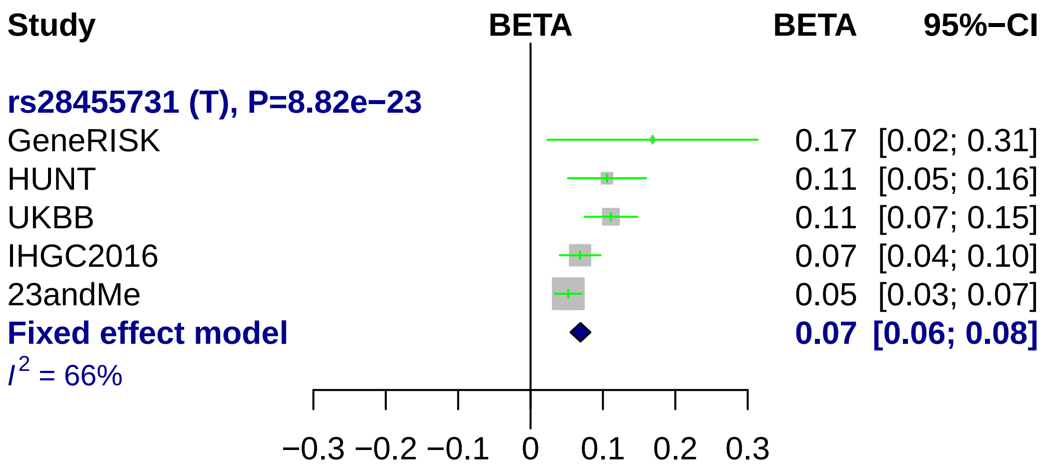
Study
BETA
BETA
95%−CI
rs28455731 (T), P=8.82e−23
GeneRISK 0.17 [0.02; 0.31]
HUNT 0.11 [0.05; 0.16]
UKBB 0.11 [0.07; 0.15]
IHGC2016 0.07 [0.04; 0.10]
23andMe 0.05 [0.03; 0.07]
Fixed effect model 0.07 [0.06; 0.08]
I2 = 66%
−0.3 −0.2 −0.1 0 0.1 0.2 0.3

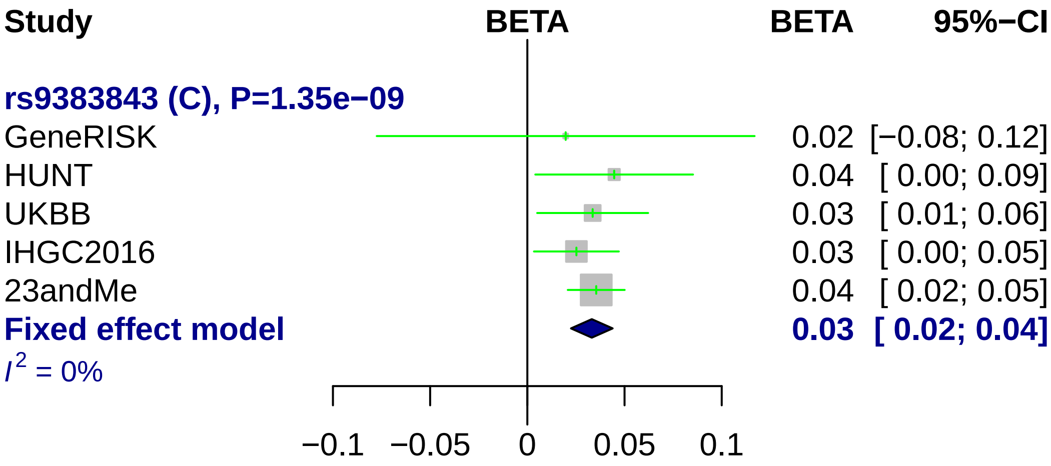
Study
BETA
BETA
95%−CI
rs9383843 (C), P=1.35e−09
GeneRISK
HUNT
UKBB
IHGC2016
23andMe
Fixed effect model
$I^2$ = 0%
0.02 [−0.08; 0.12]
0.04 [ 0.00; 0.09]
0.03 [ 0.01; 0.06]
0.03 [ 0.00; 0.05]
0.04 [ 0.02; 0.05]
0.03 [ 0.02; 0.04]
−0.1
−0.05
0
0.05
0.1

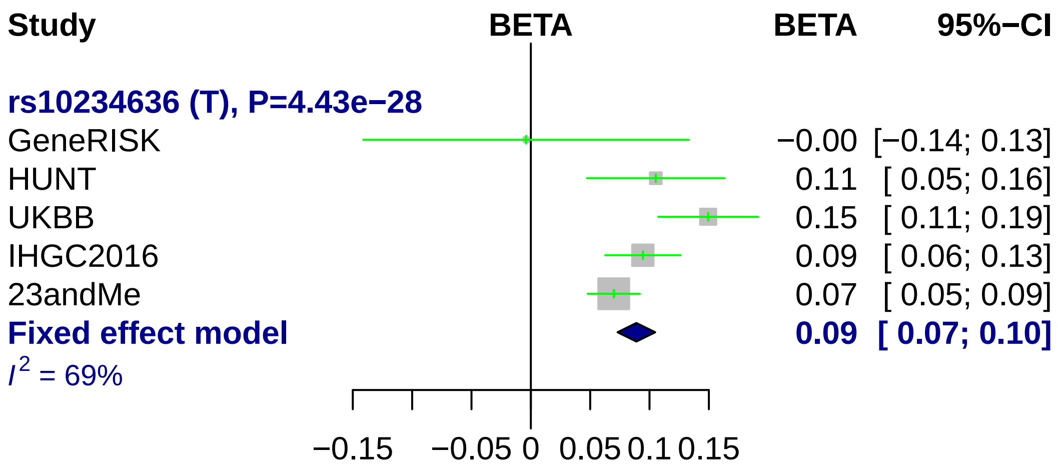
Study
BETA
BETA
95%−CI
rs10234636 (T), P=4.43e−28
GeneRISK −0.00 [−0.14; 0.13]
HUNT 0.11 [ 0.05; 0.16]
UKBB 0.15 [ 0.11; 0.19]
IHGC2016 0.09 [ 0.06; 0.13]
23andMe 0.07 [ 0.05; 0.09]
Fixed effect model 0.09 [ 0.07; 0.10]
I2 = 69%
−0.15
−0.05
0
0.05
0.1
0.15

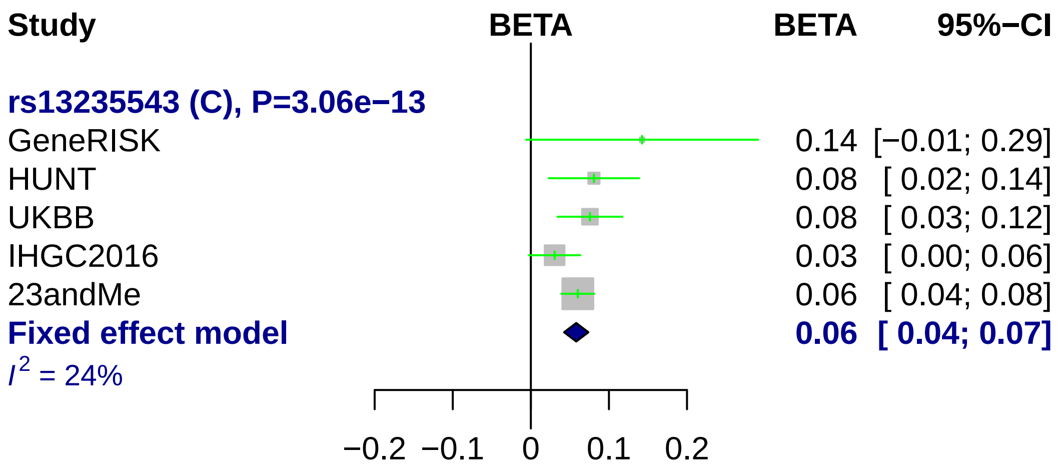
Study
BETA
BETA
95%−CI
rs13235543 (C), P=3.06e−13
GeneRISK
0.14 [−0.01; 0.29]
HUNT
0.08 [ 0.02; 0.14]
UKBB
0.08 [ 0.03; 0.12]
IHGC2016
0.03 [ 0.00; 0.06]
23andMe
0.06 [ 0.04; 0.08]
Fixed effect model
0.06 [ 0.04; 0.07]
I2 = 24%
−0.2
−0.1
0
0.1
0.2

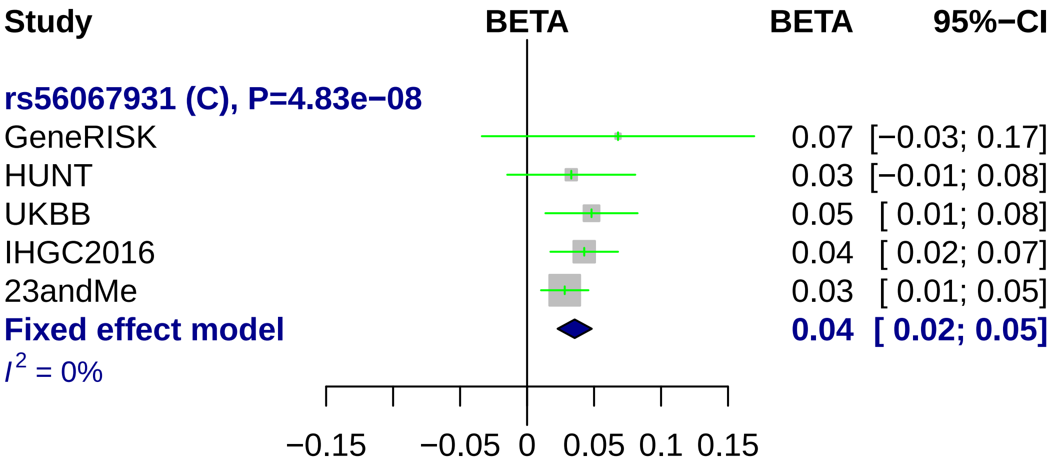
Study
BETA
BETA
95%−CI
rs56067931 (C), P=4.83e−08
GeneRISK
0.07 [−0.03; 0.17]
HUNT
0.03 [−0.01; 0.08]
UKBB
0.05 [ 0.01; 0.08]
IHGC2016
0.04 [ 0.02; 0.07]
23andMe
0.03 [ 0.01; 0.05]
Fixed effect model
0.04 [ 0.02; 0.05]
I 2 = 0%
−0.15
−0.05
0
0.05
0.1
0.15

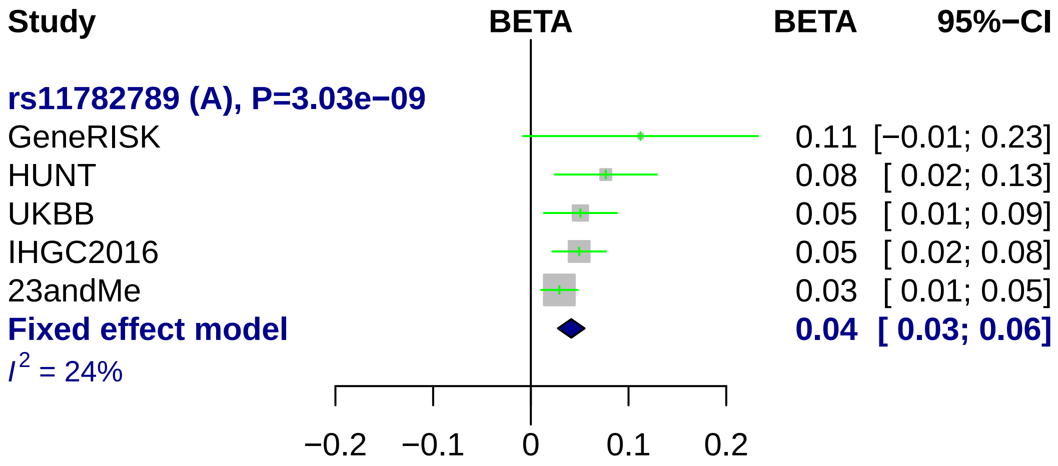
Study
BETA
BETA
95%−CI
rs11782789 (A), P=3.03e−09
GeneRISK 0.11 [−0.01; 0.23]
HUNT 0.08 [ 0.02; 0.13]
UKBB 0.05 [ 0.01; 0.09]
IHGC2016 0.05 [ 0.02; 0.08]
23andMe 0.03 [ 0.01; 0.05]
Fixed effect model 0.04 [ 0.03; 0.06]
I2 = 24%
−0.2
−0.1
0
0.1
0.2

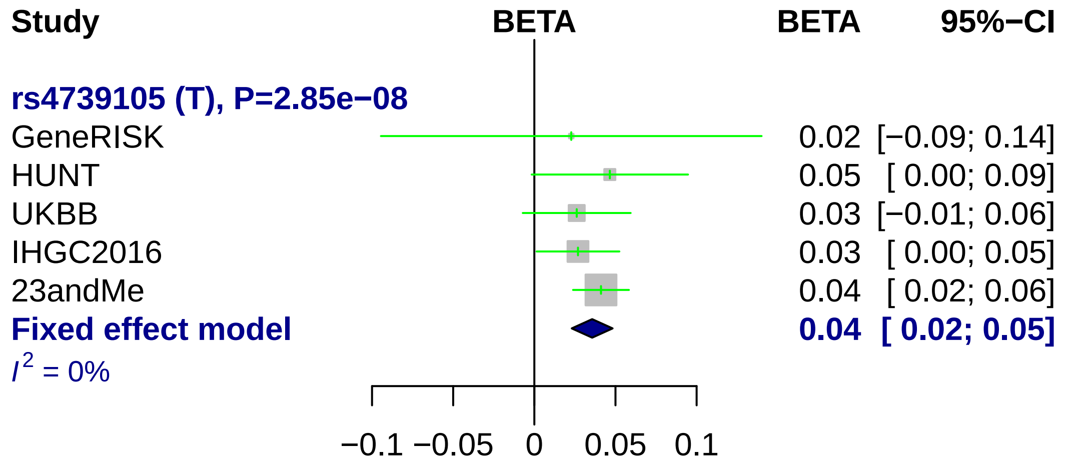
Study
BETA
BETA
95%−CI
rs4739105 (T), P=2.85e−08
GeneRISK
0.02 [−0.09; 0.14]
HUNT
0.05 [ 0.00; 0.09]
UKBB
0.03 [−0.01; 0.06]
IHGC2016
0.03 [ 0.00; 0.05]
23andMe
0.04 [ 0.02; 0.06]
Fixed effect model
0.04 [ 0.02; 0.05]
I^2 = 0%
−0.1
−0.05
0
0.05
0.1

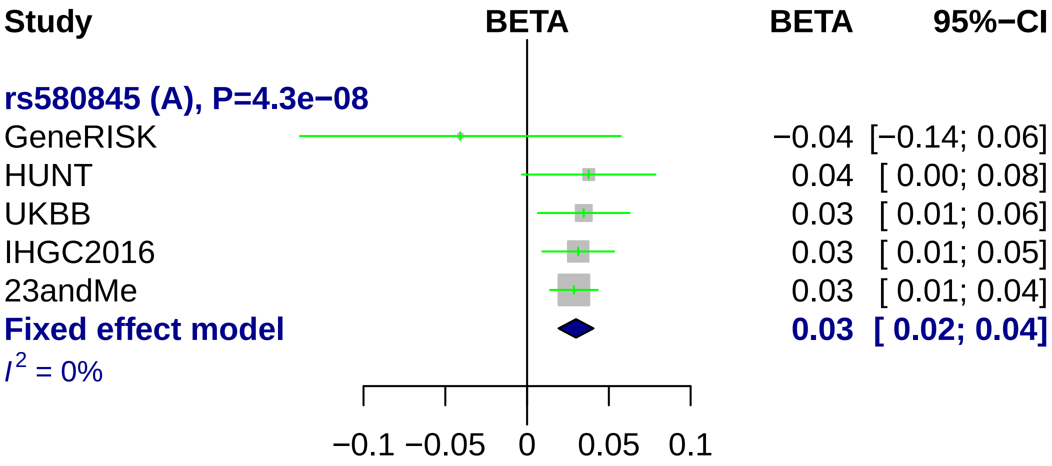
Study
BETA
BETA
95%−CI
rs580845 (A), P=4.3e−08
GeneRISK −0.04 [−0.14; 0.06]
HUNT 0.04 [ 0.00; 0.08]
UKBB 0.03 [ 0.01; 0.06]
IHGC2016 0.03 [ 0.01; 0.05]
23andMe 0.03 [ 0.01; 0.04]
Fixed effect model 0.03 [ 0.02; 0.04]
I2 = 0%
−0.1
−0.05
0
0.05
0.1

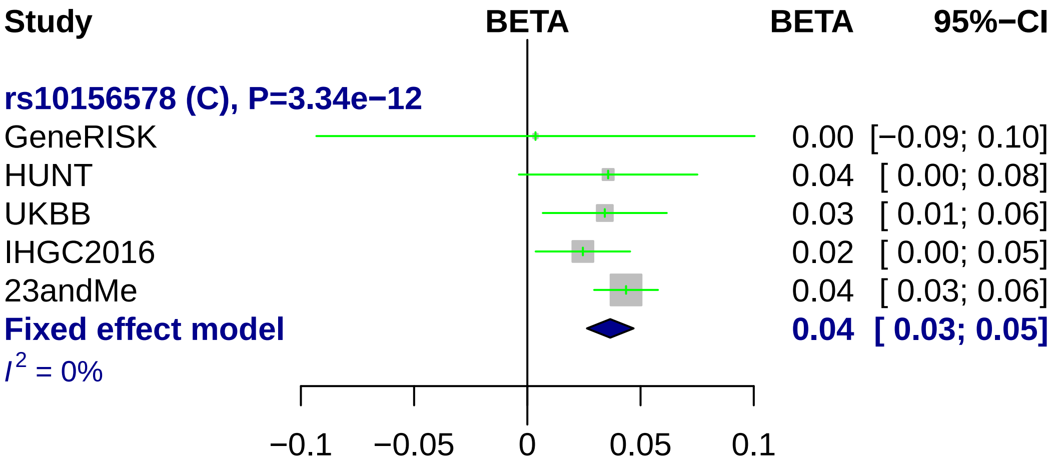
Study
BETA
BETA
95%−CI
rs10156578 (C), P=3.34e−12
GeneRISK
0.00 [−0.09; 0.10]
HUNT
0.04 [ 0.00; 0.08]
UKBB
0.03 [ 0.01; 0.06]
IHGC2016
0.02 [ 0.00; 0.05]
23andMe
0.04 [ 0.03; 0.06]
Fixed effect model
0.04 [ 0.03; 0.05]
I2 = 0%
−0.1
−0.05
0
0.05
0.1

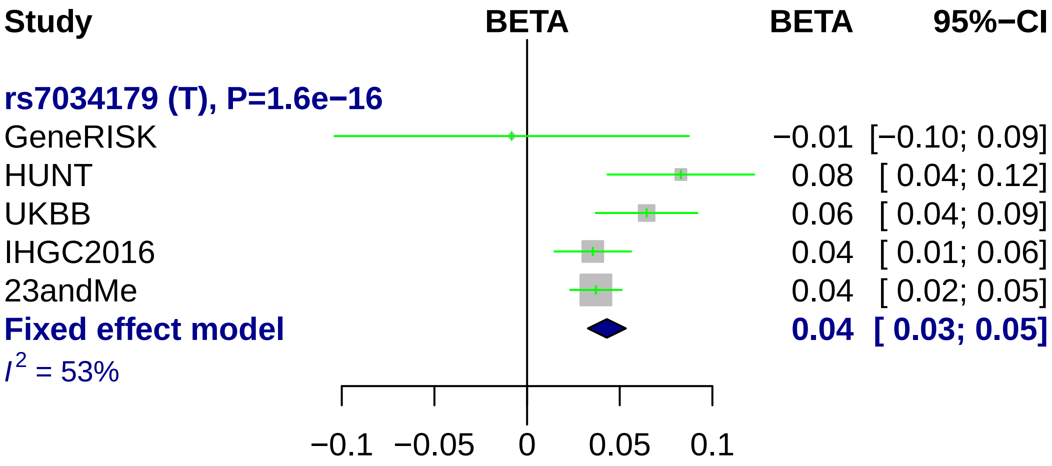

Study
BETA
BETA
95%−CI
rs7034179 (T), P=1.6e−16
GeneRISK −0.01 [−0.10; 0.09]
HUNT 0.08 [ 0.04; 0.12]
UKBB 0.06 [ 0.04; 0.09]
IHGC2016 0.04 [ 0.01; 0.06]
23andMe 0.04 [ 0.02; 0.05]
Fixed effect model 0.04 [ 0.03; 0.05]
I2 = 53%
−0.1
−0.05
0
0.05
0.1

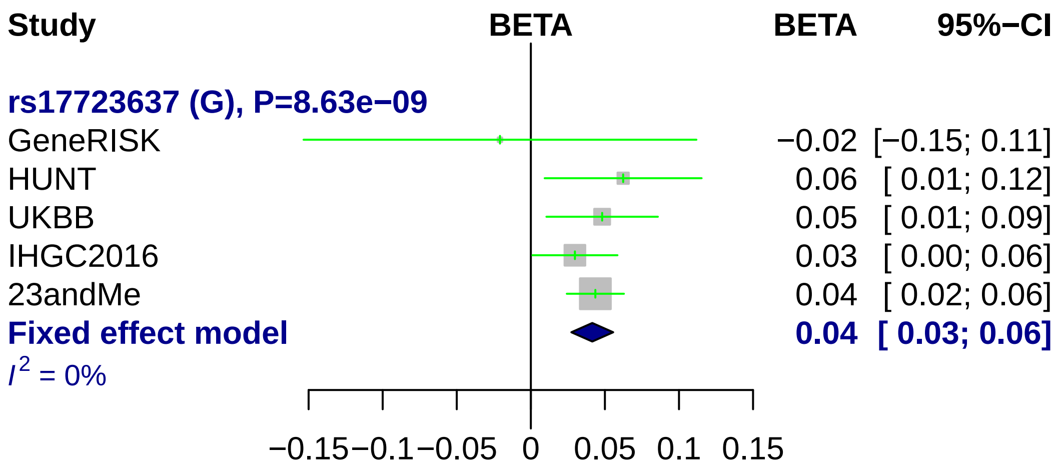
Study
BETA
BETA
95%−CI
rs17723637 (G), P=8.63e−09
GeneRISK
−0.02 [−0.15; 0.11]
HUNT
0.06 [ 0.01; 0.12]
UKBB
0.05 [ 0.01; 0.09]
IHGC2016
0.03 [ 0.00; 0.06]
23andMe
0.04 [ 0.02; 0.06]
Fixed effect model
0.04 [ 0.03; 0.06]
I2 = 0%
−0.15
−0.1
−0.05
0
0.05
0.1
0.15

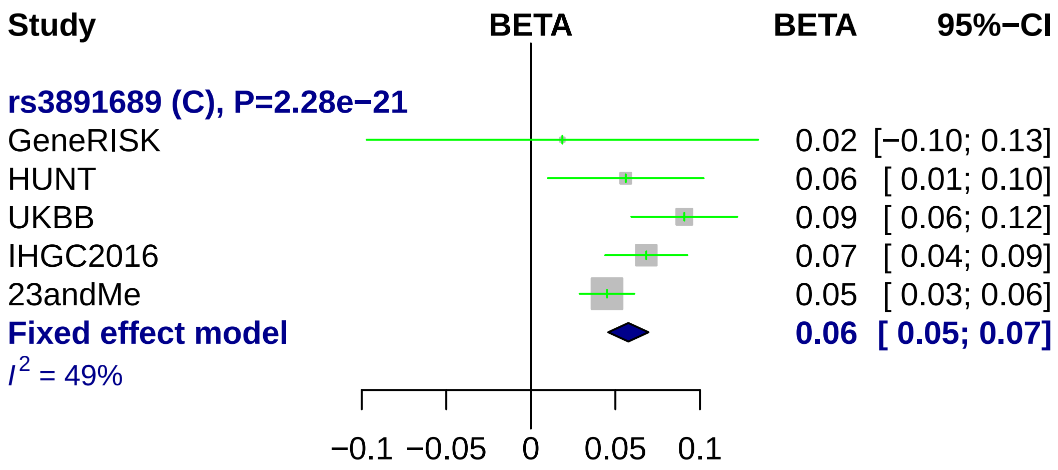
Study
BETA
BETA
95%−CI
rs3891689 (C), P=2.28e−21
GeneRISK
0.02 [−0.10; 0.13]
HUNT
0.06 [ 0.01; 0.10]
UKBB
0.09 [ 0.06; 0.12]
IHGC2016
0.07 [ 0.04; 0.09]
23andMe
0.05 [ 0.03; 0.06]
Fixed effect model
0.06 [ 0.05; 0.07]
I² = 49%
−0.1
−0.05
0
0.05
0.1

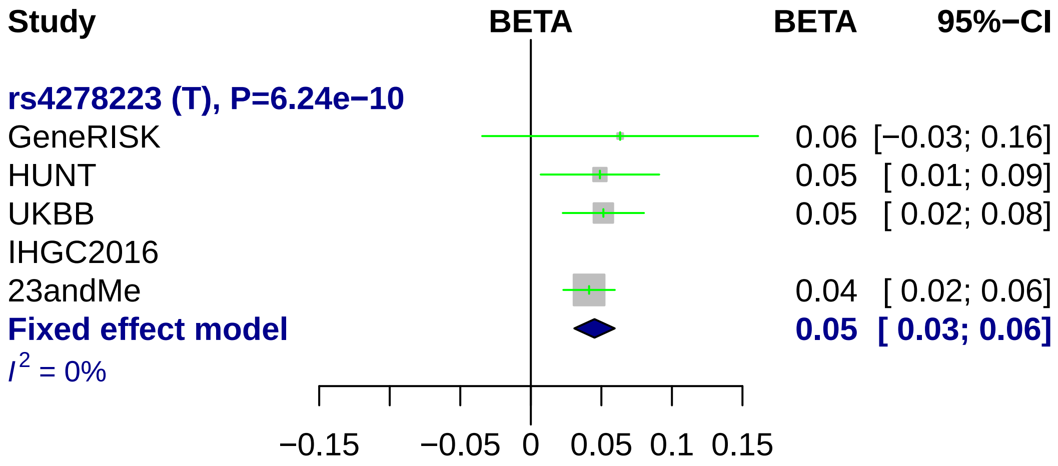
Study
BETA
BETA
95%−CI
rs4278223 (T), P=6.24e−10
GeneRISK
0.06 [−0.03; 0.16]
HUNT
0.05 [ 0.01; 0.09]
UKBB
0.05 [ 0.02; 0.08]
IHGC2016
23andMe
0.04 [ 0.02; 0.06]
Fixed effect model
0.05 [ 0.03; 0.06]
I2 = 0%
−0.15
−0.05
0
0.05
0.1
0.15

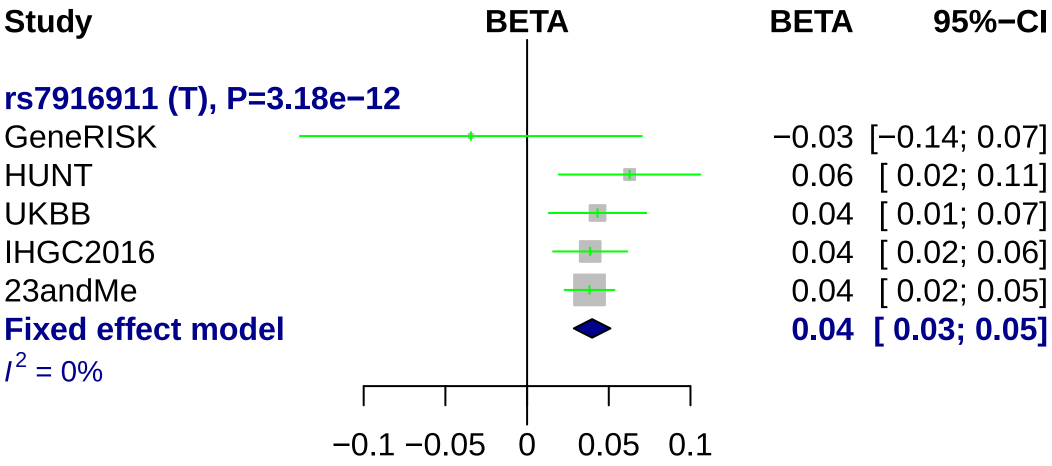
Study
BETA
BETA
95%−CI
rs7916911 (T), P=3.18e−12
GeneRISK
−0.03
[−0.14; 0.07]
HUNT
0.06
[ 0.02; 0.11]
UKBB
0.04
[ 0.01; 0.07]
IHGC2016
0.04
[ 0.02; 0.06]
23andMe
0.04
[ 0.02; 0.05]
Fixed effect model
0.04
[ 0.03; 0.05]
I2 = 0%
−0.1
−0.05
0
0.05
0.1

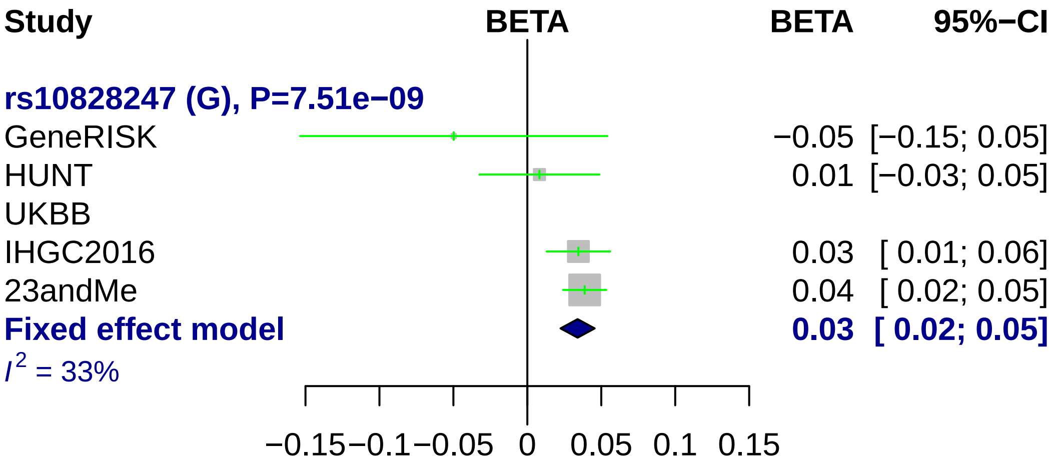

Study
BETA
BETA
95%−CI
rs10828247 (G), P=7.51e−09
GeneRISK
−0.05 [−0.15; 0.05]
HUNT
0.01 [−0.03; 0.05]
UKBB
IHGC2016
0.03 [ 0.01; 0.06]
23andMe
0.04 [ 0.02; 0.05]
Fixed effect model
0.03 [ 0.02; 0.05]
I2 = 33%
−0.15
−0.1
−0.05
0
0.05
0.1
0.15

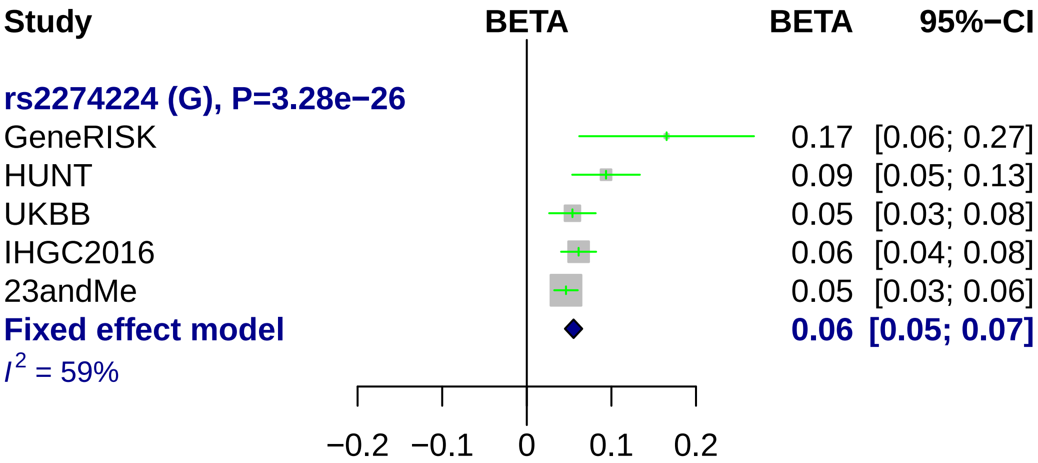
Study
BETA
BETA
95%−CI
rs2274224 (G), P=3.28e−26
GeneRISK 0.17 [0.06; 0.27]
HUNT 0.09 [0.05; 0.13]
UKBB 0.05 [0.03; 0.08]
IHGC2016 0.06 [0.04; 0.08]
23andMe 0.05 [0.03; 0.06]
Fixed effect model 0.06 [0.05; 0.07]
I2 = 59%
−0.2 −0.1 0 0.1 0.2

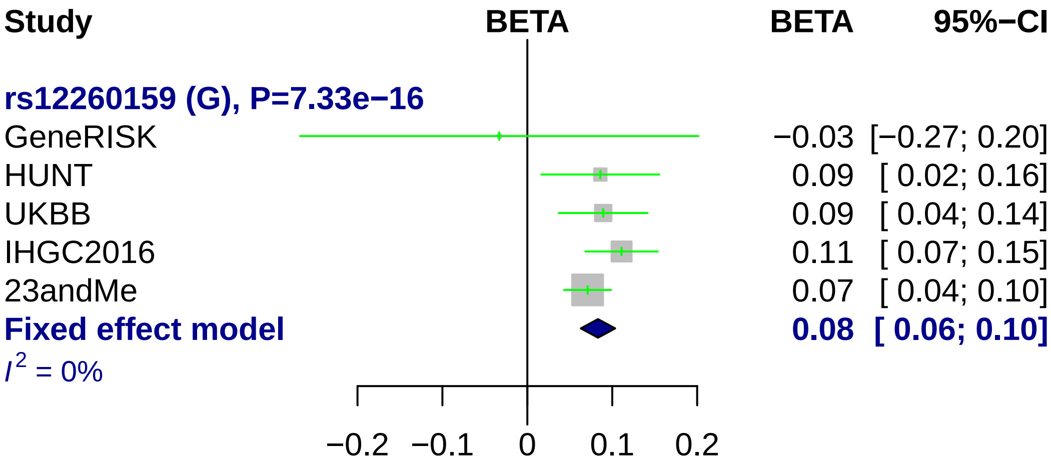

Study
BETA
BETA
95%−CI
rs12260159 (G), P=7.33e−16
GeneRISK −0.03 [−0.27; 0.20]
HUNT 0.09 [ 0.02; 0.16]
UKBB 0.09 [ 0.04; 0.14]
IHGC2016 0.11 [ 0.07; 0.15]
23andMe 0.07 [ 0.04; 0.10]
Fixed effect model 0.08 [ 0.06; 0.10]
I^2 = 0%
−0.2 −0.1 0 0.1 0.2

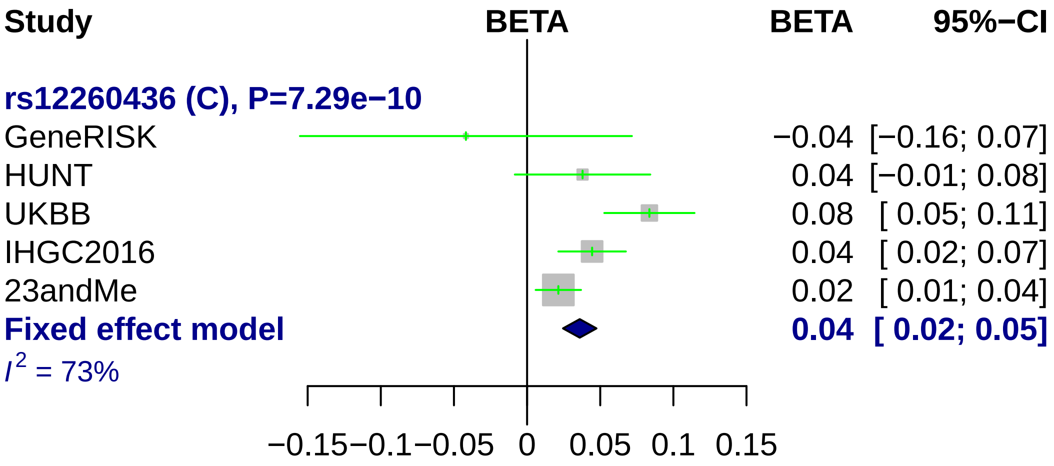

Study
BETA
BETA
95%−CI
rs12260436 (C), P=7.29e−10
GeneRISK
−0.04 [−0.16; 0.07]
HUNT
0.04 [−0.01; 0.08]
UKBB
0.08 [ 0.05; 0.11]
IHGC2016
0.04 [ 0.02; 0.07]
23andMe
0.02 [ 0.01; 0.04]
Fixed effect model
0.04 [ 0.02; 0.05]
$I^2$ = 73%
−0.15
−0.1
−0.05
0
0.05
0.1
0.15

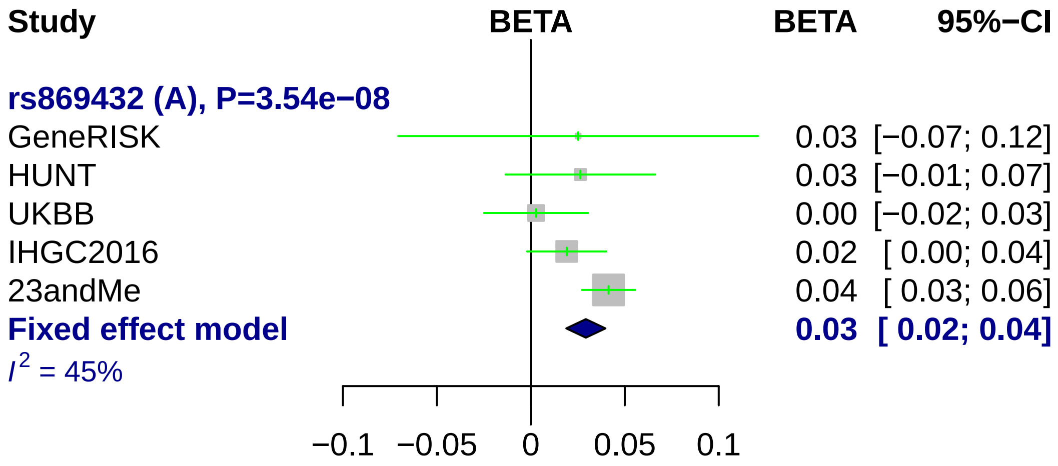
Study
BETA
BETA
95%−CI
rs869432 (A), P=3.54e−08
GeneRISK
0.03 [−0.07; 0.12]
HUNT
0.03 [−0.01; 0.07]
UKBB
0.00 [−0.02; 0.03]
IHGC2016
0.02 [ 0.00; 0.04]
23andMe
0.04 [ 0.03; 0.06]
Fixed effect model
0.03 [ 0.02; 0.04]
$I^2$ = 45%
−0.1
−0.05
0
0.05
0.1

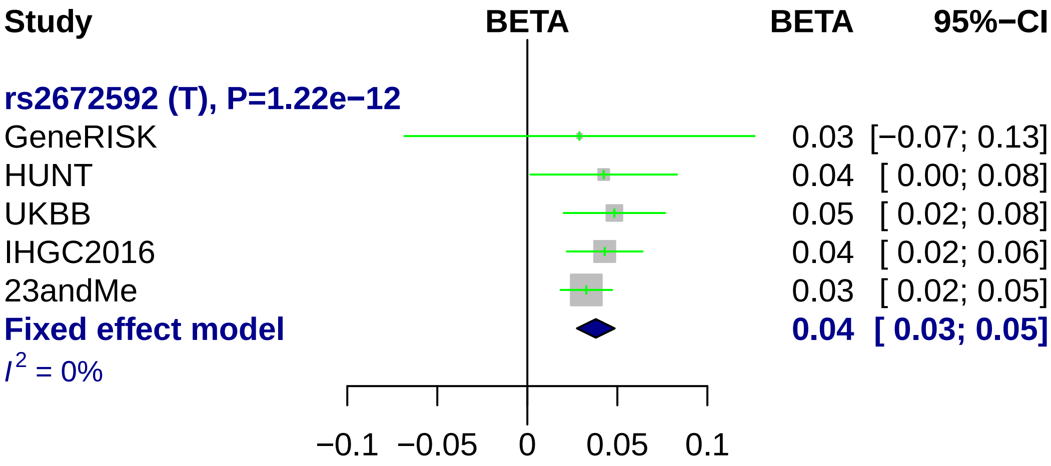
Study
BETA
BETA
95%−CI
rs2672592 (T), P=1.22e−12
GeneRISK 0.03 [−0.07; 0.13]
HUNT 0.04 [ 0.00; 0.08]
UKBB 0.05 [ 0.02; 0.08]
IHGC2016 0.04 [ 0.02; 0.06]
23andMe 0.03 [ 0.02; 0.05]
Fixed effect model 0.04 [ 0.03; 0.05]
I^2 = 0%
−0.1
−0.05
0
0.05
0.1

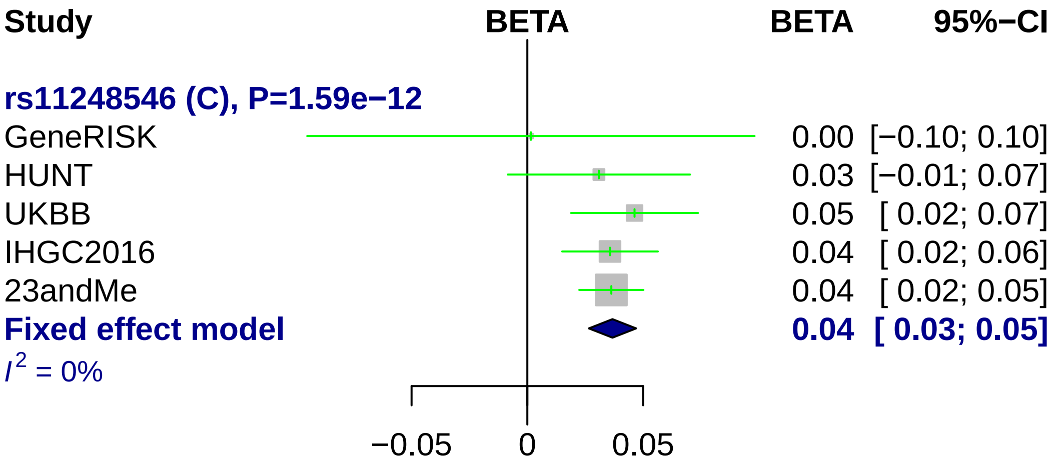

Study
BETA
BETA
95%−CI
rs11248546 (C), P=1.59e−12
GeneRISK 0.00 [−0.10; 0.10]
HUNT 0.03 [−0.01; 0.07]
UKBB 0.05 [ 0.02; 0.07]
IHGC2016 0.04 [ 0.02; 0.06]
23andMe 0.04 [ 0.02; 0.05]
Fixed effect model 0.04 [ 0.03; 0.05]
I^2 = 0%
−0.05
0
0.05

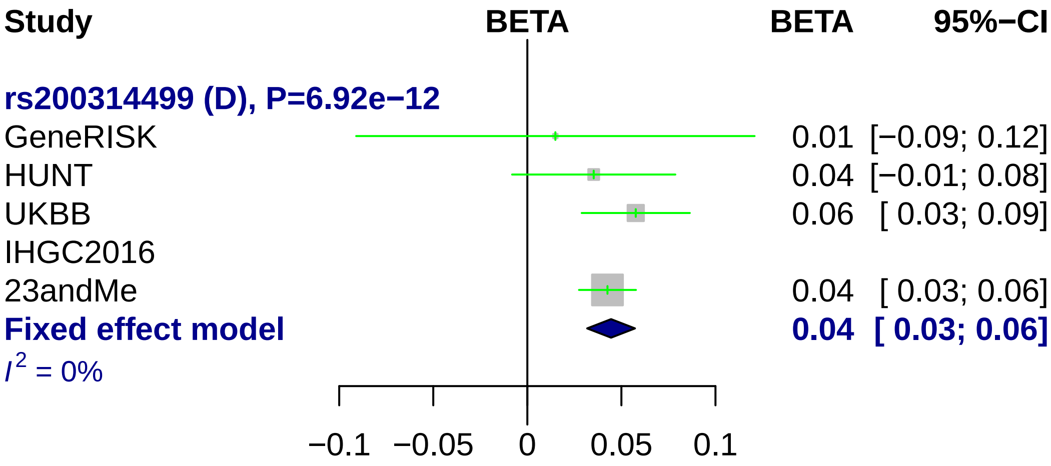
Study
BETA
BETA
95%−CI
rs200314499 (D), P=6.92e−12
GeneRISK
0.01 [−0.09; 0.12]
HUNT
0.04 [−0.01; 0.08]
UKBB
0.06 [ 0.03; 0.09]
IHGC2016
23andMe
0.04 [ 0.03; 0.06]
Fixed effect model
0.04 [ 0.03; 0.06]
I2 = 0%
−0.1
−0.05
0
0.05
0.1

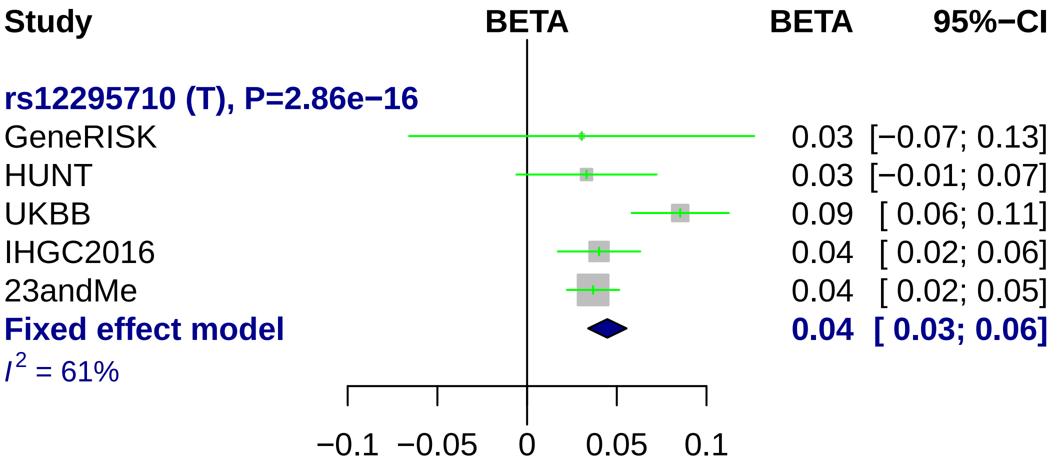

Study
BETA
BETA
95%−CI
rs12295710 (T), P=2.86e−16
GeneRISK 0.03 [−0.07; 0.13]
HUNT 0.03 [−0.01; 0.07]
UKBB 0.09 [ 0.06; 0.11]
IHGC2016 0.04 [ 0.02; 0.06]
23andMe 0.04 [ 0.02; 0.05]
Fixed effect model 0.04 [ 0.03; 0.06]
I2 = 61%
−0.1
−0.05
0
0.05
0.1

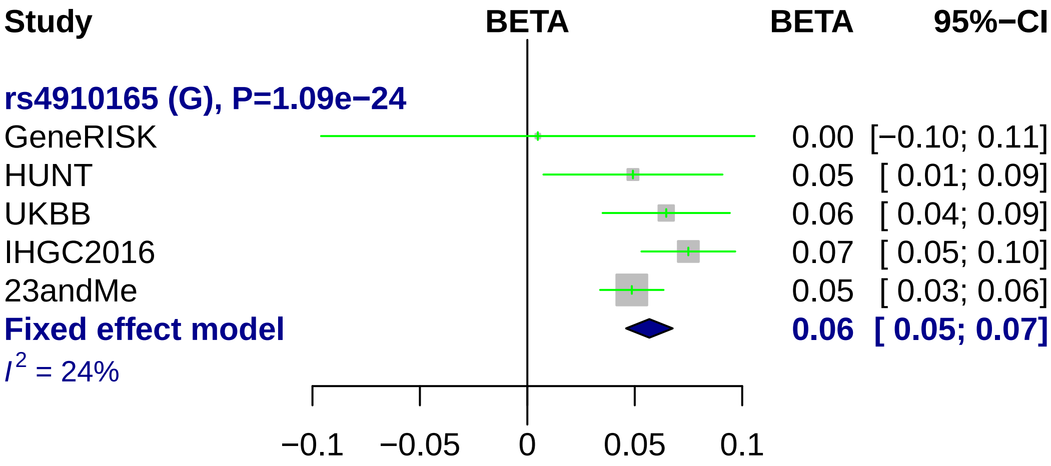

Study
BETA
BETA
95%−CI
rs4910165 (G), P=1.09e−24
GeneRISK 0.00 [−0.10; 0.11]
HUNT 0.05 [ 0.01; 0.09]
UKBB 0.06 [ 0.04; 0.09]
IHGC2016 0.07 [ 0.05; 0.10]
23andMe 0.05 [ 0.03; 0.06]
Fixed effect model 0.06 [ 0.05; 0.07]
I2 = 24%
−0.1
−0.05
0
0.05
0.1

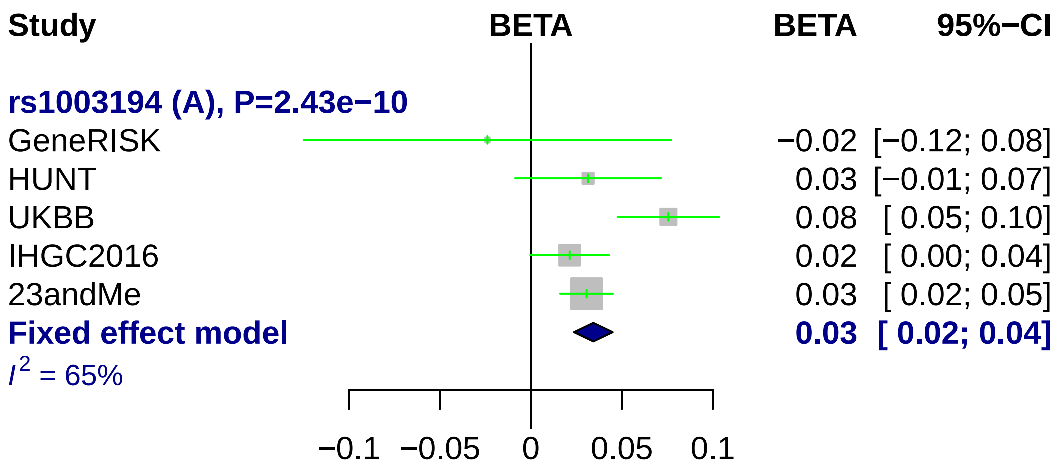
Study
BETA
BETA
95%−CI
rs1003194 (A), P=2.43e−10
GeneRISK
−0.02 [−0.12; 0.08]
HUNT
0.03 [−0.01; 0.07]
UKBB
0.08 [ 0.05; 0.10]
IHGC2016
0.02 [ 0.00; 0.04]
23andMe
0.03 [ 0.02; 0.05]
Fixed effect model
0.03 [ 0.02; 0.04]
I2 = 65%
−0.1
−0.05
0
0.05
0.1

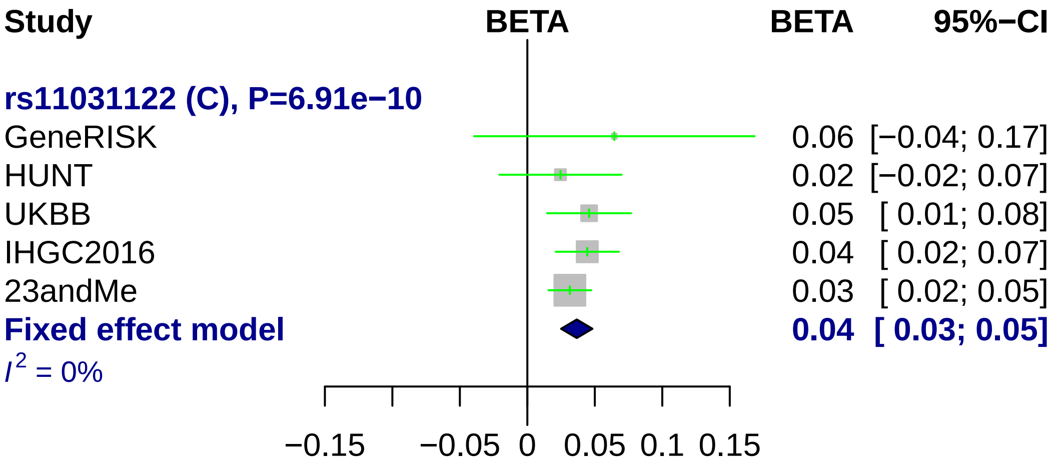

Study
BETA
BETA
95%−CI
rs11031122 (C), P=6.91e−10
GeneRISK
0.06 [−0.04; 0.17]
HUNT
0.02 [−0.02; 0.07]
UKBB
0.05 [ 0.01; 0.08]
IHGC2016
0.04 [ 0.02; 0.07]
23andMe
0.03 [ 0.02; 0.05]
Fixed effect model
0.04 [ 0.03; 0.05]
I2 = 0%
−0.15
−0.05
0
0.05
0.1
0.15

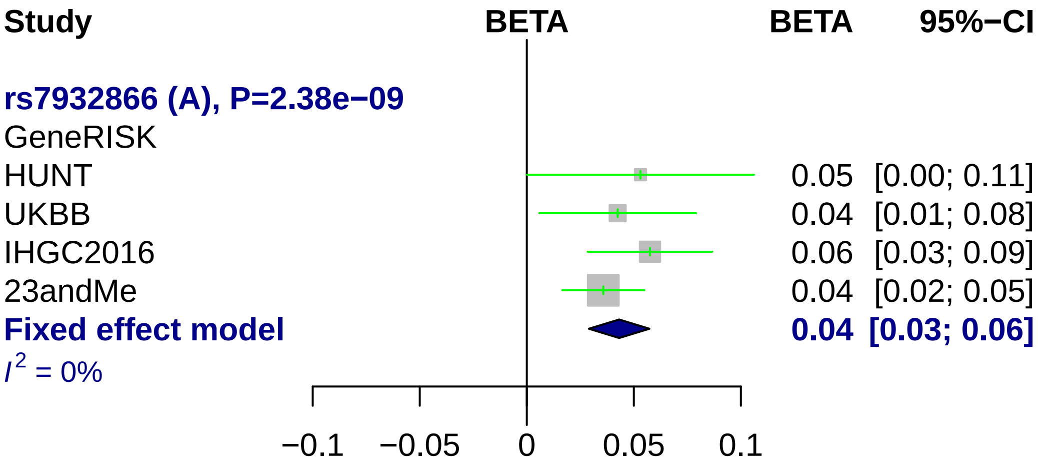
Study
BETA
BETA
95%−CI
rs7932866 (A), P=2.38e−09
GeneRISK
HUNT
0.05 [0.00; 0.11]
UKBB
0.04 [0.01; 0.08]
IHGC2016
0.06 [0.03; 0.09]
23andMe
0.04 [0.02; 0.05]
Fixed effect model
0.04 [0.03; 0.06]
I^2 = 0%
−0.1
−0.05
0
0.05
0.1

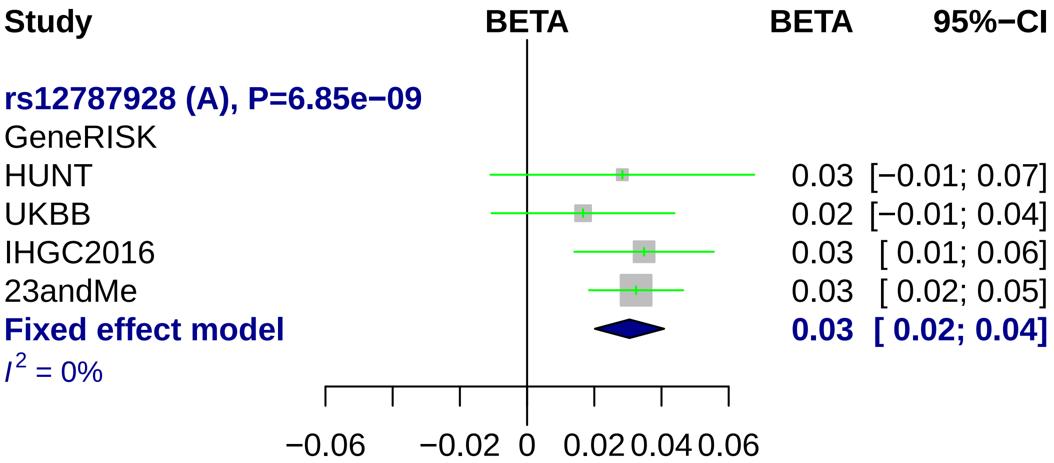
Study
BETA
BETA
95%−CI
rs12787928 (A), P=6.85e−09
GeneRISK
HUNT
0.03 [−0.01; 0.07]
UKBB
0.02 [−0.01; 0.04]
IHGC2016
0.03 [ 0.01; 0.06]
23andMe
0.03 [ 0.02; 0.05]
Fixed effect model
0.03 [ 0.02; 0.04]
I2 = 0%
−0.06
−0.02
0
0.02
0.04
0.06

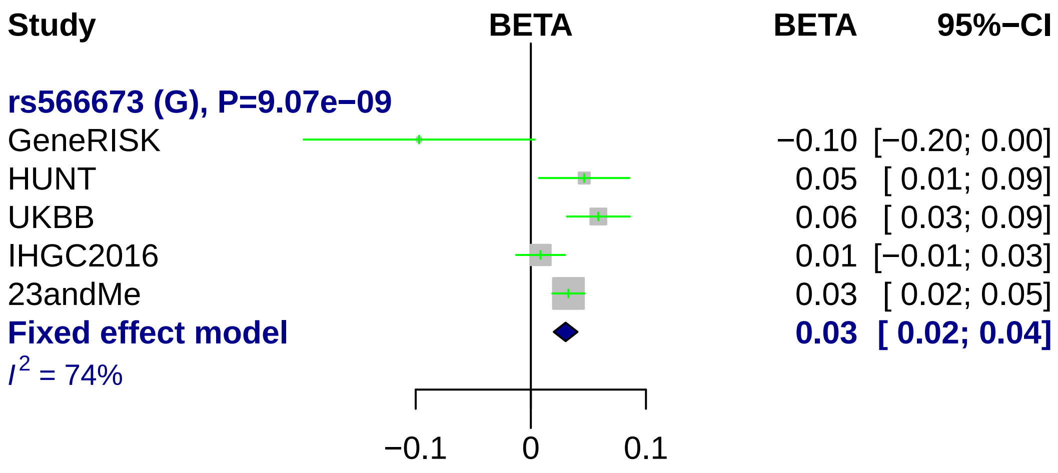
Study
BETA
BETA
95%−CI
rs566673 (G), P=9.07e−09
GeneRISK −0.10 [−0.20; 0.00]
HUNT 0.05 [ 0.01; 0.09]
UKBB 0.06 [ 0.03; 0.09]
IHGC2016 0.01 [−0.01; 0.03]
23andMe 0.03 [ 0.02; 0.05]
Fixed effect model 0.03 [ 0.02; 0.04]
I2 = 74%
−0.1
0
0.1

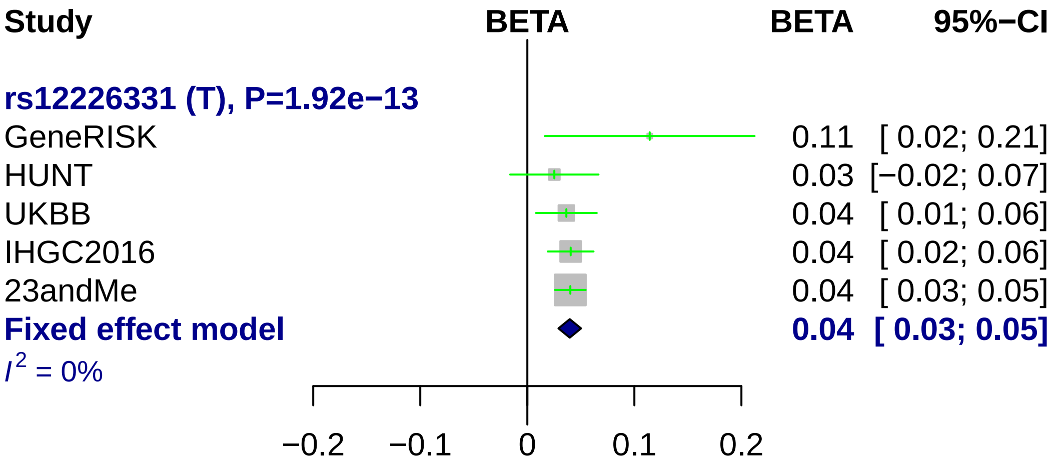
Study
BETA
BETA
95%−CI
rs12226331 (T), P=1.92e−13
GeneRISK
0.11 [ 0.02; 0.21]
HUNT
0.03 [−0.02; 0.07]
UKBB
0.04 [ 0.01; 0.06]
IHGC2016
0.04 [ 0.02; 0.06]
23andMe
0.04 [ 0.03; 0.05]
Fixed effect model
0.04 [ 0.03; 0.05]
I^2 = 0%
−0.2
−0.1
0
0.1
0.2

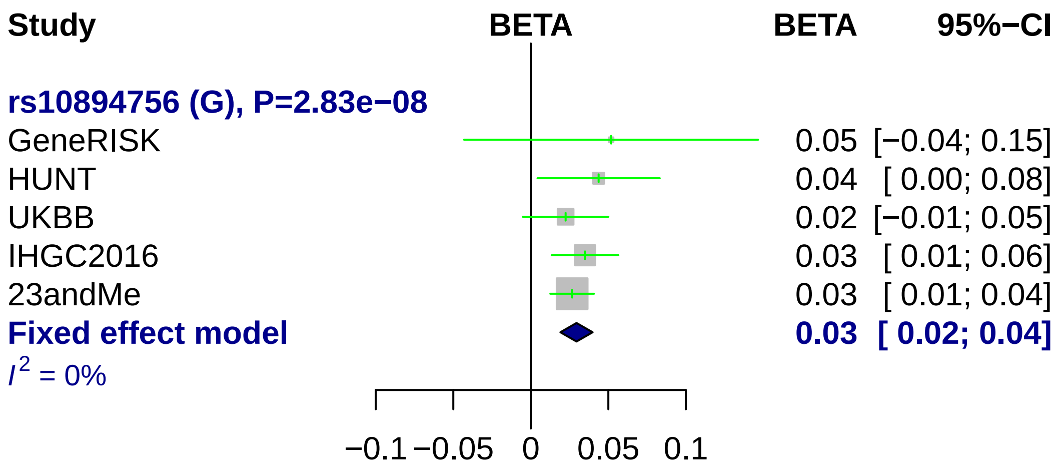
Study
BETA
BETA
95%−CI
rs10894756 (G), P=2.83e−08
GeneRISK
0.05 [−0.04; 0.15]
HUNT
0.04 [ 0.00; 0.08]
UKBB
0.02 [−0.01; 0.05]
IHGC2016
0.03 [ 0.01; 0.06]
23andMe
0.03 [ 0.01; 0.04]
Fixed effect model
0.03 [ 0.02; 0.04]
I2 = 0%
−0.1
−0.05
0
0.05
0.1

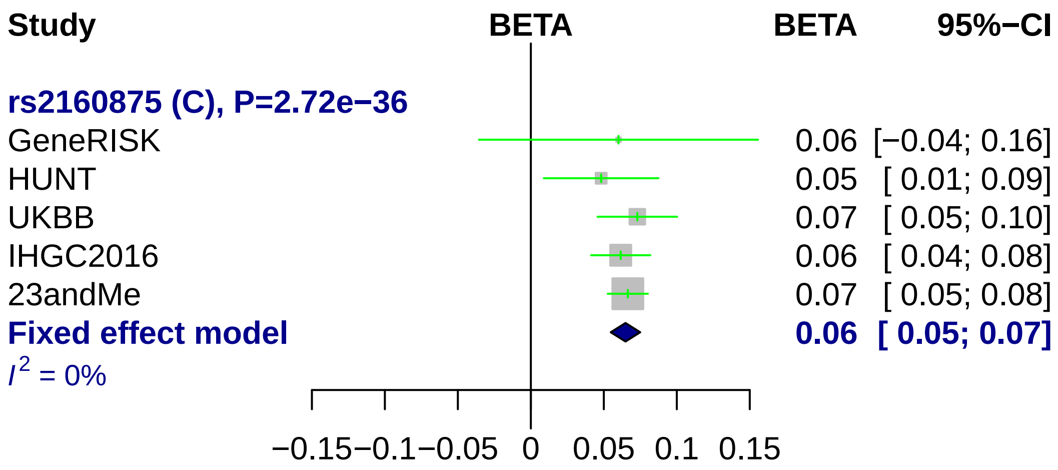
Study
BETA
BETA
95%−CI
rs2160875 (C), P=2.72e−36
GeneRISK 0.06 [−0.04; 0.16]
HUNT 0.05 [ 0.01; 0.09]
UKBB 0.07 [ 0.05; 0.10]
IHGC2016 0.06 [ 0.04; 0.08]
23andMe 0.07 [ 0.05; 0.08]
Fixed effect model 0.06 [ 0.05; 0.07]
I^2 = 0%
−0.15 −0.1 −0.05 0 0.05 0.1 0.15

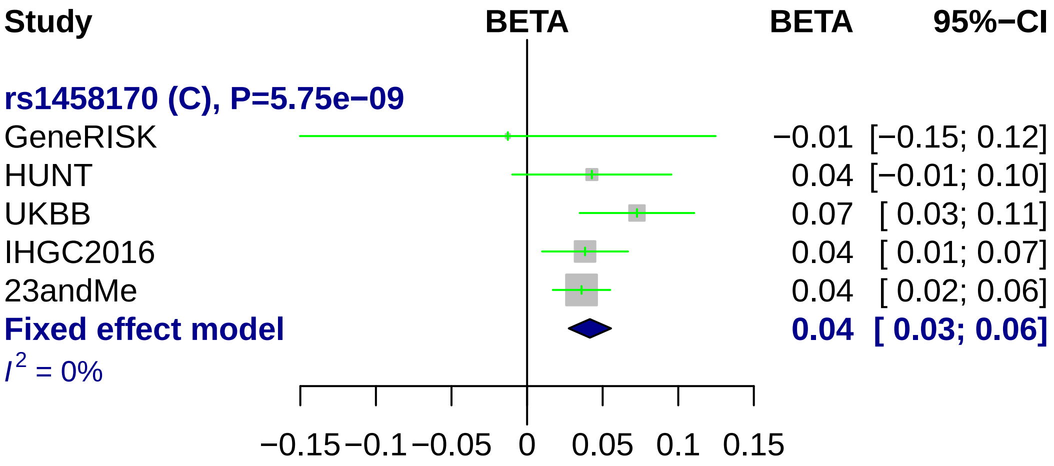
Study
BETA
BETA
95%−CI
rs1458170 (C), P=5.75e−09
GeneRISK −0.01 [−0.15; 0.12]
HUNT 0.04 [−0.01; 0.10]
UKBB 0.07 [ 0.03; 0.11]
IHGC2016 0.04 [ 0.01; 0.07]
23andMe 0.04 [ 0.02; 0.06]
Fixed effect model 0.04 [ 0.03; 0.06]
I2 = 0%
−0.15 −0.1 −0.05 0 0.05 0.1 0.15

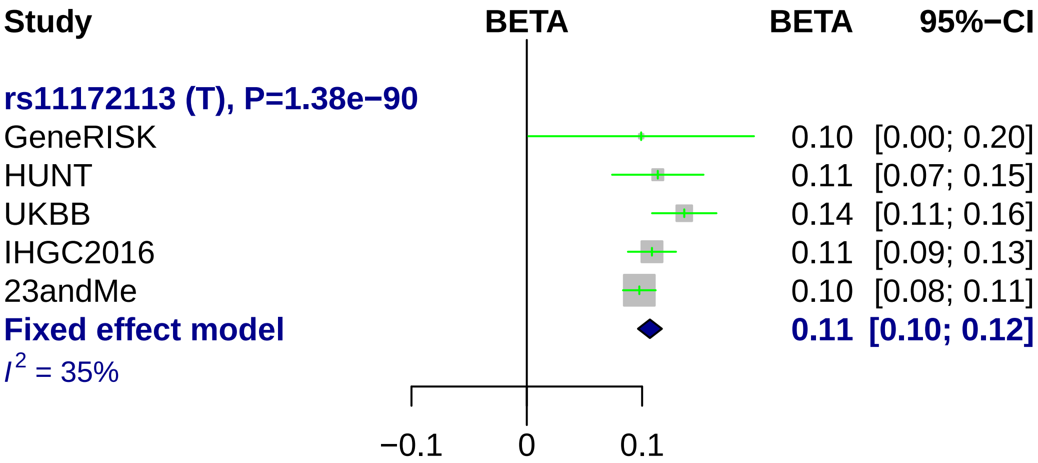
Study
BETA
BETA
95%−CI
rs11172113 (T), P=1.38e−90
GeneRISK
0.10 [0.00; 0.20]
HUNT
0.11 [0.07; 0.15]
UKBB
0.14 [0.11; 0.16]
IHGC2016
0.11 [0.09; 0.13]
23andMe
0.10 [0.08; 0.11]
Fixed effect model
0.11 [0.10; 0.12]
I^2 = 35%
−0.1
0
0.1

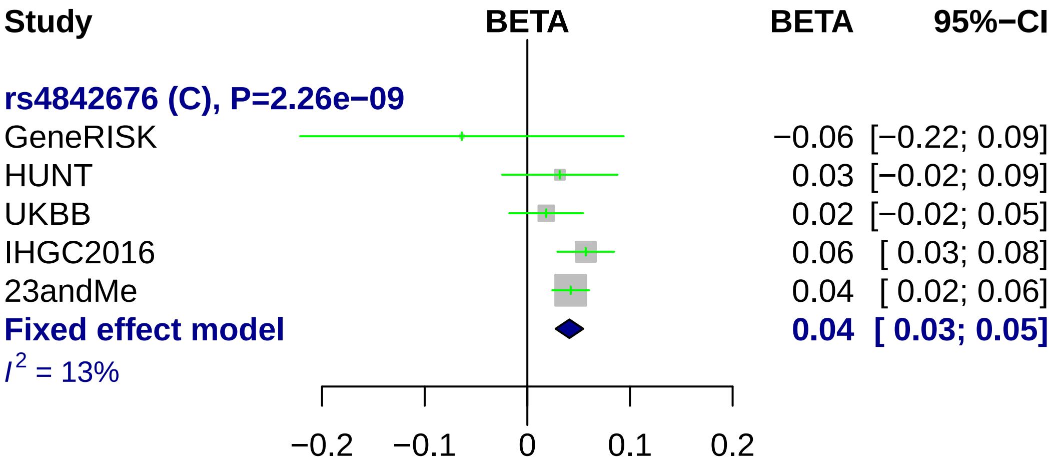
Study
BETA
BETA
95%−CI
rs4842676 (C), P=2.26e−09
GeneRISK −0.06 [−0.22; 0.09]
HUNT 0.03 [−0.02; 0.09]
UKBB 0.02 [−0.02; 0.05]
IHGC2016 0.06 [ 0.03; 0.08]
23andMe 0.04 [ 0.02; 0.06]
Fixed effect model 0.04 [ 0.03; 0.05]
$I^2$ = 13%
−0.2
−0.1
0
0.1
0.2

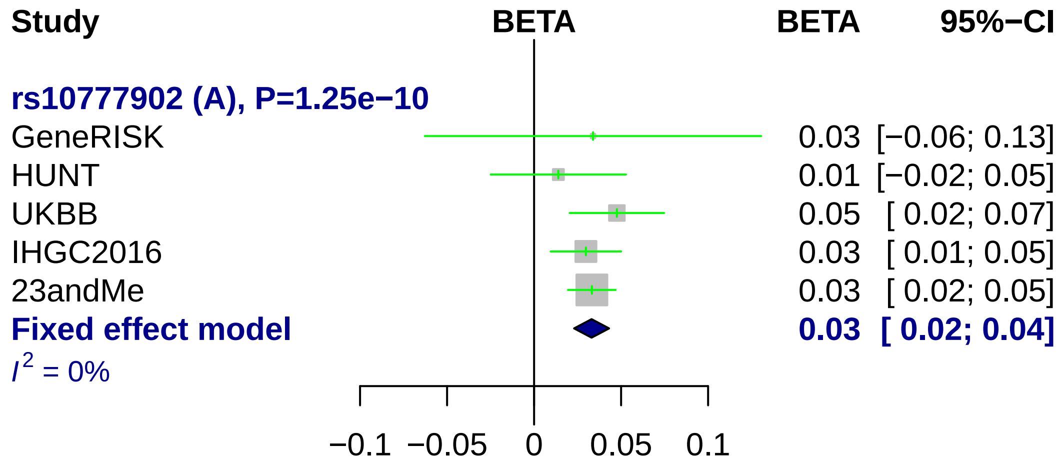
Study
BETA
BETA
95%−CI
rs10777902 (A), P=1.25e−10
GeneRISK
0.03 [−0.06; 0.13]
HUNT
0.01 [−0.02; 0.05]
UKBB
0.05 [ 0.02; 0.07]
IHGC2016
0.03 [ 0.01; 0.05]
23andMe
0.03 [ 0.02; 0.05]
Fixed effect model
0.03 [ 0.02; 0.04]
I2 = 0%
−0.1
−0.05
0
0.05
0.1

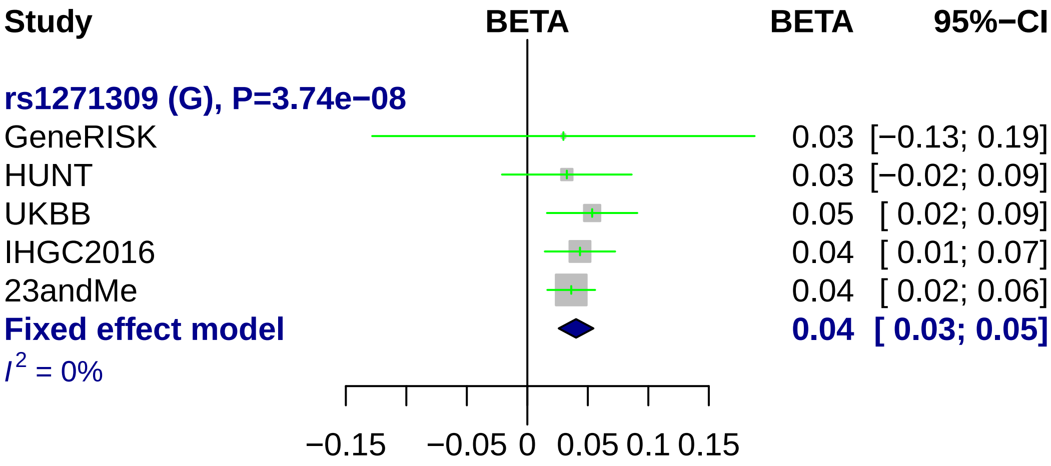
Study
BETA
BETA
95%−CI
rs1271309 (G), P=3.74e−08
GeneRISK
0.03 [−0.13; 0.19]
HUNT
0.03 [−0.02; 0.09]
UKBB
0.05 [ 0.02; 0.09]
IHGC2016
0.04 [ 0.01; 0.07]
23andMe
0.04 [ 0.02; 0.06]
Fixed effect model
0.04 [ 0.03; 0.05]
I^2 = 0%
−0.15
−0.05
0
0.05
0.1
0.15

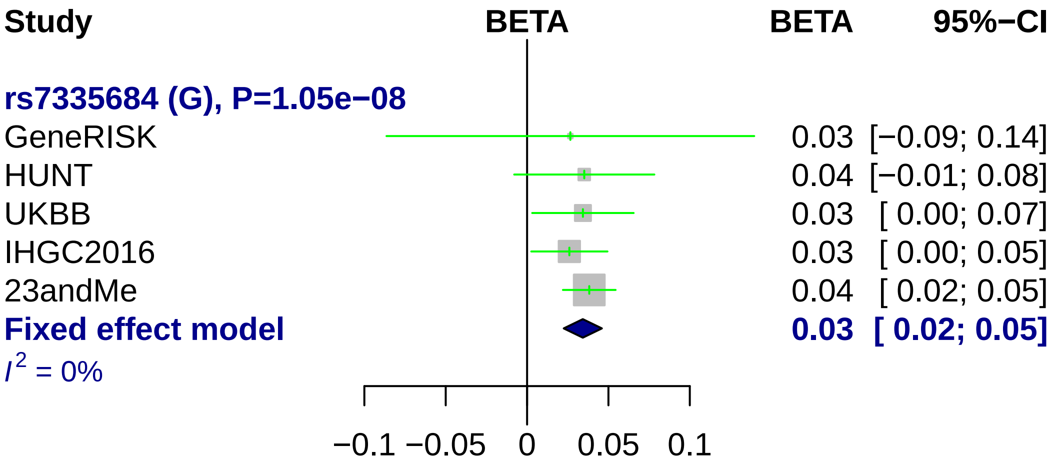
Study
BETA
BETA
95%−CI
rs7335684 (G), P=1.05e−08
GeneRISK
0.03 [−0.09; 0.14]
HUNT
0.04 [−0.01; 0.08]
UKBB
0.03 [ 0.00; 0.07]
IHGC2016
0.03 [ 0.00; 0.05]
23andMe
0.04 [ 0.02; 0.05]
Fixed effect model
0.03 [ 0.02; 0.05]
I 2 = 0%
−0.1
−0.05
0
0.05
0.1

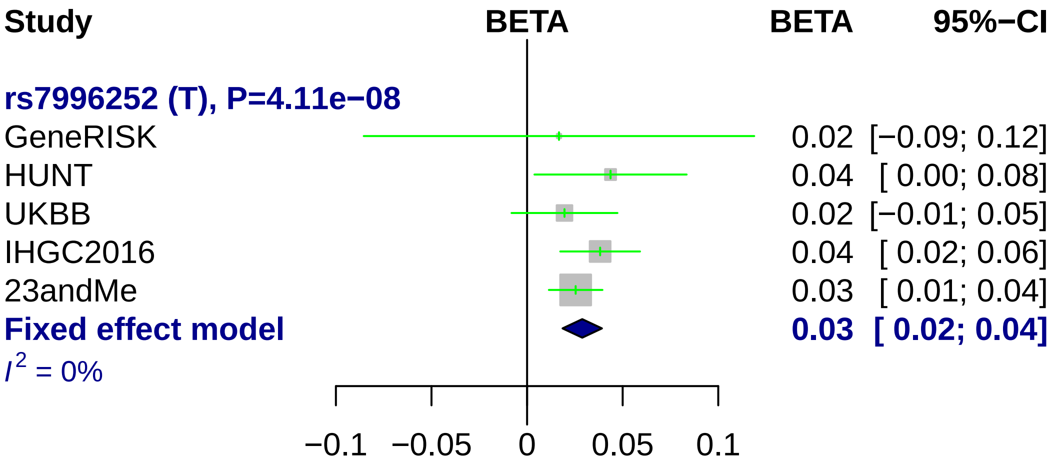
Study
BETA
BETA
95%−CI
rs7996252 (T), P=4.11e−08
GeneRISK
0.02 [−0.09; 0.12]
HUNT
0.04 [ 0.00; 0.08]
UKBB
0.02 [−0.01; 0.05]
IHGC2016
0.04 [ 0.02; 0.06]
23andMe
0.03 [ 0.01; 0.04]
Fixed effect model
0.03 [ 0.02; 0.04]
I2 = 0%
−0.1
−0.05
0
0.05
0.1

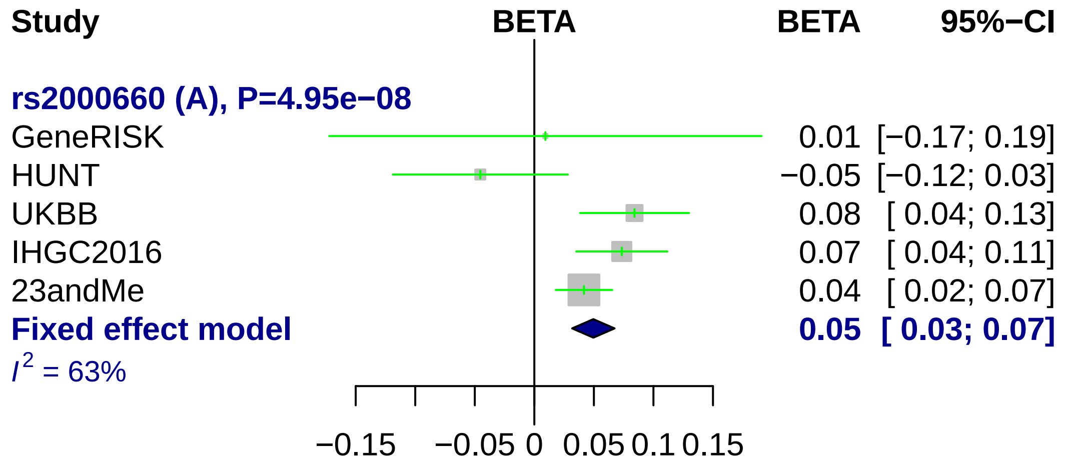
Study
BETA
BETA
95%−CI
rs2000660 (A), P=4.95e−08
GeneRISK
0.01 [−0.17; 0.19]
HUNT
−0.05 [−0.12; 0.03]
UKBB
0.08 [ 0.04; 0.13]
IHGC2016
0.07 [ 0.04; 0.11]
23andMe
0.04 [ 0.02; 0.07]
Fixed effect model
0.05 [ 0.03; 0.07]
I2 = 63%
−0.15
−0.05
0
0.05
0.1
0.15

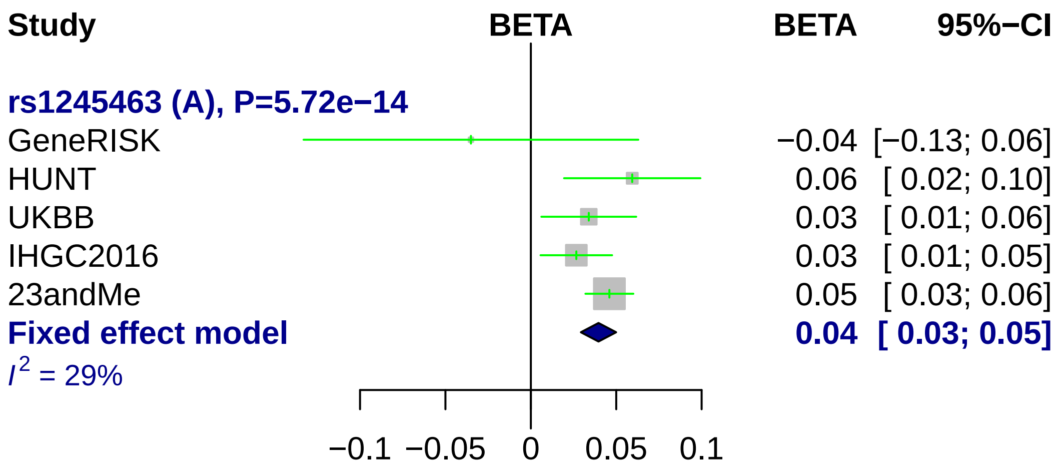
Study
BETA
BETA
95%−CI
rs1245463 (A), P=5.72e−14
GeneRISK −0.04 [−0.13; 0.06]
HUNT 0.06 [ 0.02; 0.10]
UKBB 0.03 [ 0.01; 0.06]
IHGC2016 0.03 [ 0.01; 0.05]
23andMe 0.05 [ 0.03; 0.06]
Fixed effect model 0.04 [ 0.03; 0.05]
I2 = 29%
−0.1 −0.05 0 0.05 0.1

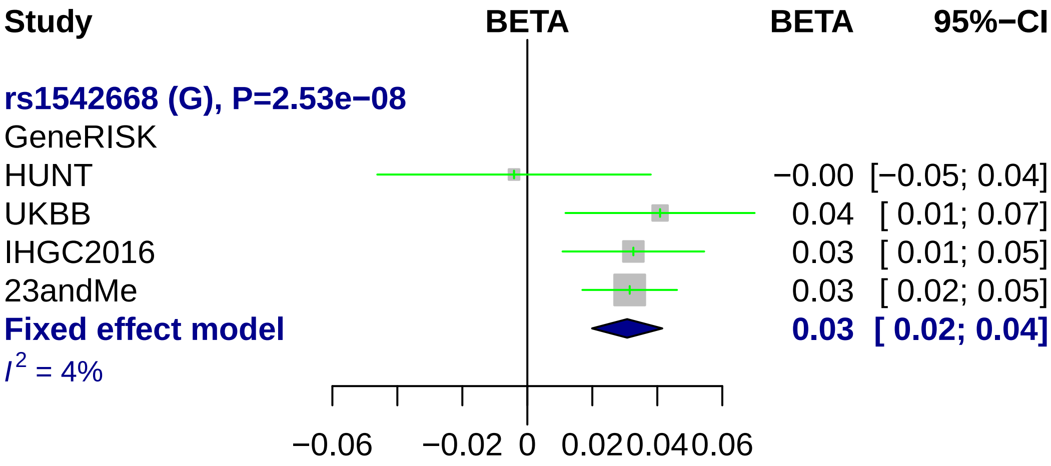

Study
BETA
BETA
95%−CI
rs1542668 (G), P=2.53e−08
GeneRISK
HUNT
−0.00 [−0.05; 0.04]
UKBB
0.04 [ 0.01; 0.07]
IHGC2016
0.03 [ 0.01; 0.05]
23andMe
0.03 [ 0.02; 0.05]
Fixed effect model
0.03 [ 0.02; 0.04]
I2 = 4%
−0.06
−0.02
0
0.02
0.04
0.06

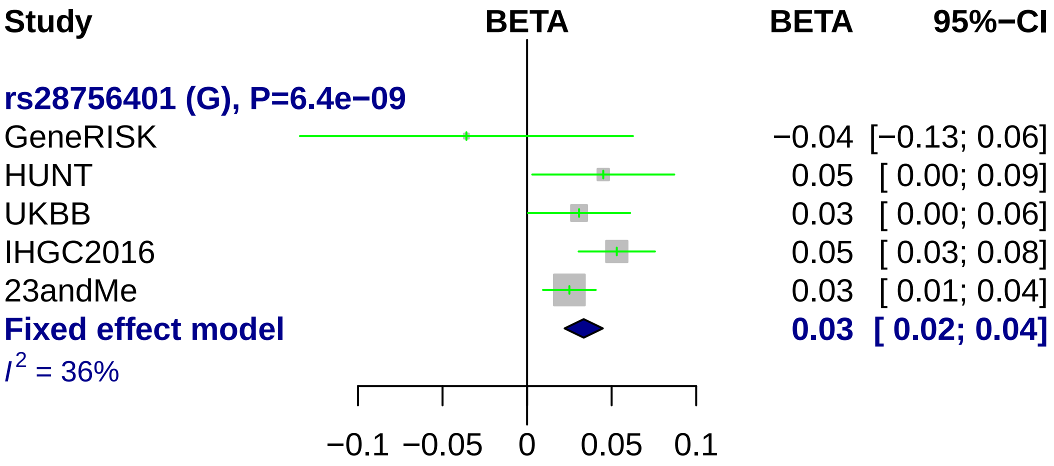
Study
BETA
BETA
95%−CI
rs28756401 (G), P=6.4e−09
GeneRISK −0.04 [−0.13; 0.06]
HUNT 0.05 [ 0.00; 0.09]
UKBB 0.03 [ 0.00; 0.06]
IHGC2016 0.05 [ 0.03; 0.08]
23andMe 0.03 [ 0.01; 0.04]
Fixed effect model 0.03 [ 0.02; 0.04]
I^2 = 36%
−0.1 −0.05 0 0.05 0.1

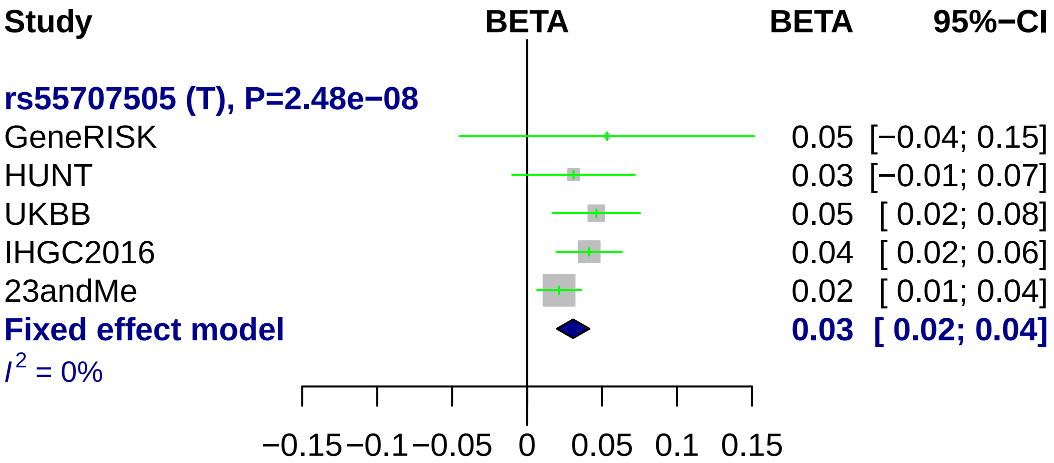
Study
BETA
BETA
95%−CI
rs55707505 (T), P=2.48e−08
GeneRISK
0.05 [−0.04; 0.15]
HUNT
0.03 [−0.01; 0.07]
UKBB
0.05 [ 0.02; 0.08]
IHGC2016
0.04 [ 0.02; 0.06]
23andMe
0.02 [ 0.01; 0.04]
Fixed effect model
0.03 [ 0.02; 0.04]
I2 = 0%
−0.15
−0.1
−0.05
0
0.05
0.1
0.15

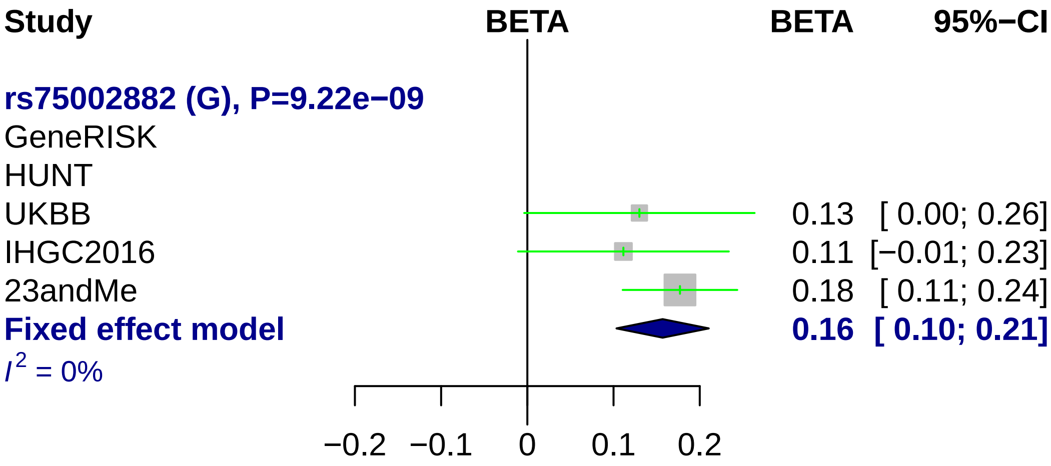
Study
BETA
BETA
95%−CI
rs75002882 (G), P=9.22e−09
GeneRISK
HUNT
UKBB
0.13 [ 0.00; 0.26]
IHGC2016
0.11 [−0.01; 0.23]
23andMe
0.18 [ 0.11; 0.24]
Fixed effect model
0.16 [ 0.10; 0.21]
I2 = 0%
−0.2
−0.1
0
0.1
0.2

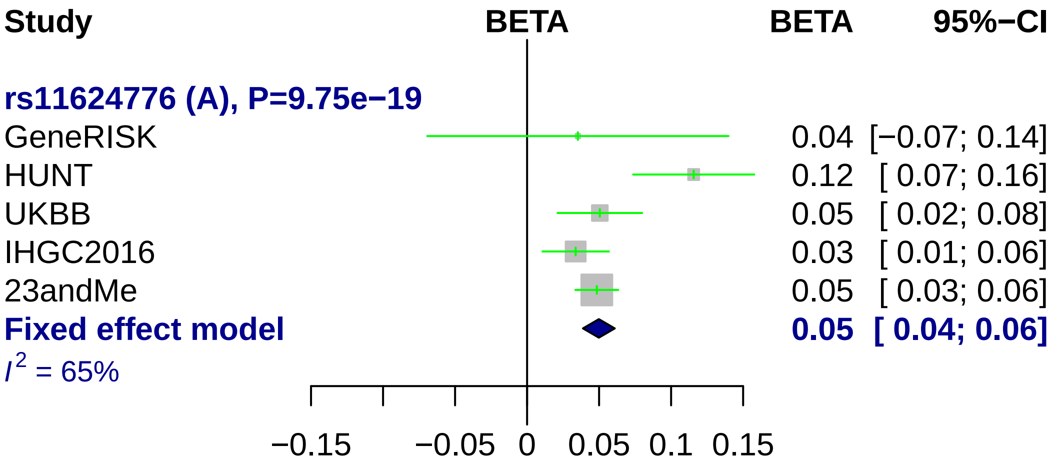
Study
BETA
BETA
95%−CI
rs11624776 (A), P=9.75e−19
GeneRISK 0.04 [−0.07; 0.14]
HUNT 0.12 [ 0.07; 0.16]
UKBB 0.05 [ 0.02; 0.08]
IHGC2016 0.03 [ 0.01; 0.06]
23andMe 0.05 [ 0.03; 0.06]
Fixed effect model 0.05 [ 0.04; 0.06]
I2 = 65%
−0.15
−0.05
0
0.05
0.1
0.15

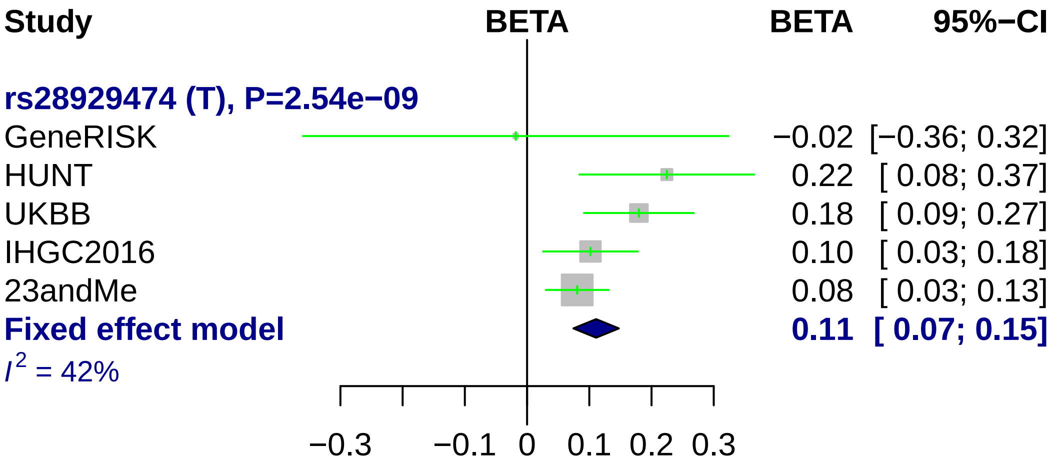
Study
BETA
BETA
95%−CI
rs28929474 (T), P=2.54e−09
GeneRISK −0.02 [−0.36; 0.32]
HUNT 0.22 [ 0.08; 0.37]
UKBB 0.18 [ 0.09; 0.27]
IHGC2016 0.10 [ 0.03; 0.18]
23andMe 0.08 [ 0.03; 0.13]
Fixed effect model 0.11 [ 0.07; 0.15]
I^2 = 42%
−0.3
−0.1
0
0.1
0.2
0.3

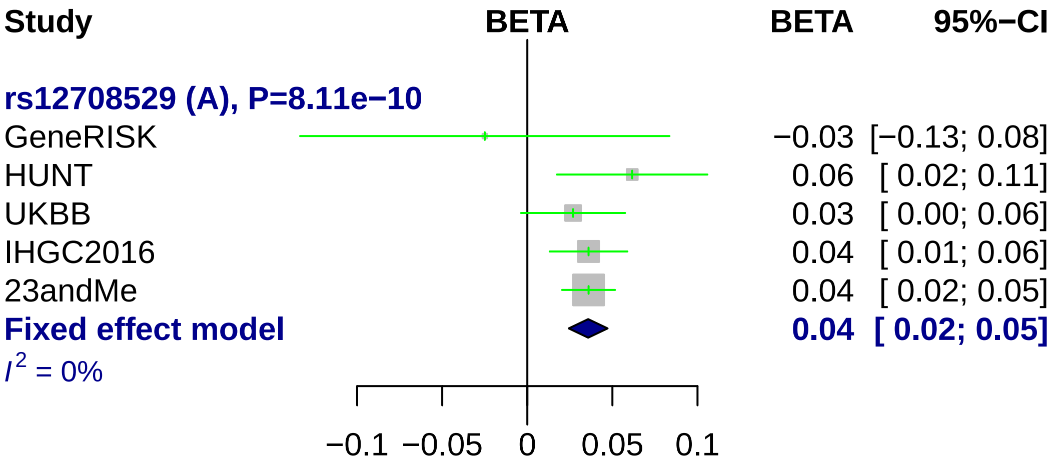
Study
BETA
BETA
95%−CI
rs12708529 (A), P=8.11e−10
GeneRISK
HUNT
UKBB
IHGC2016
23andMe
Fixed effect model
I2 = 0%
−0.03 [−0.13; 0.08]
0.06 [ 0.02; 0.11]
0.03 [ 0.00; 0.06]
0.04 [ 0.01; 0.06]
0.04 [ 0.02; 0.05]
0.04 [ 0.02; 0.05]
−0.1
−0.05
0
0.05
0.1

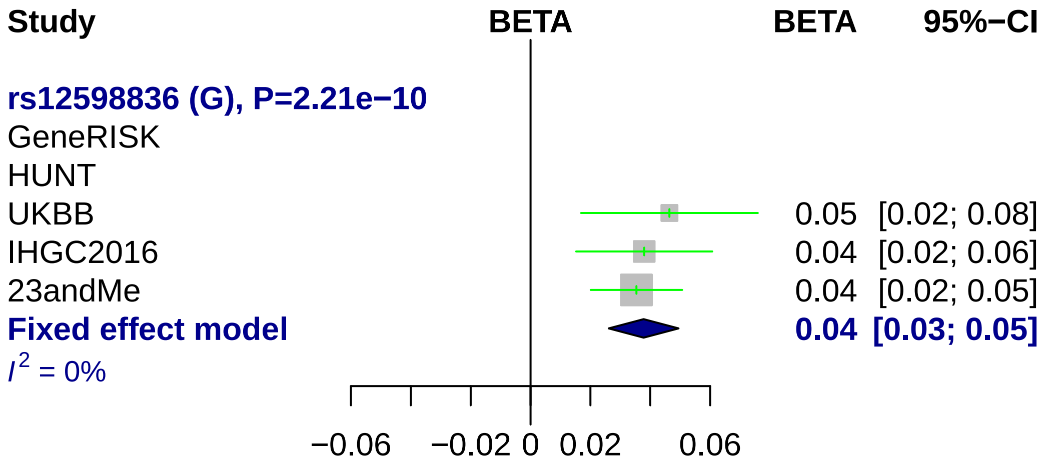

Study
BETA
BETA
95%−CI
rs12598836 (G), P=2.21e−10
GeneRISK
HUNT
UKBB
0.05 [0.02; 0.08]
IHGC2016
0.04 [0.02; 0.06]
23andMe
0.04 [0.02; 0.05]
Fixed effect model
0.04 [0.03; 0.05]
I2 = 0%
−0.06
−0.02
0
0.02
0.06

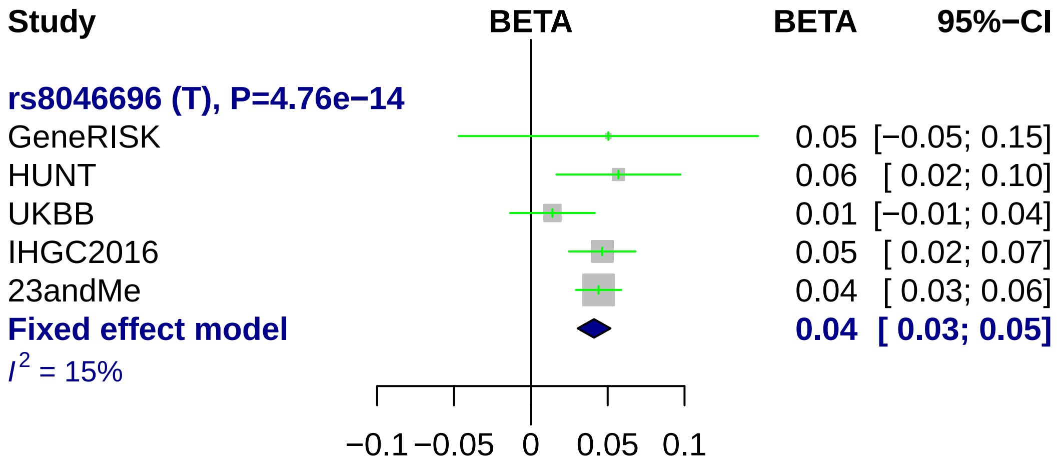

Study
BETA
BETA
95%−CI
rs8046696 (T), P=4.76e−14
GeneRISK 0.05 [−0.05; 0.15]
HUNT 0.06 [ 0.02; 0.10]
UKBB 0.01 [−0.01; 0.04]
IHGC2016 0.05 [ 0.02; 0.07]
23andMe 0.04 [ 0.03; 0.06]
Fixed effect model 0.04 [ 0.03; 0.05]
I2 = 15%
−0.1
−0.05
0
0.05
0.1

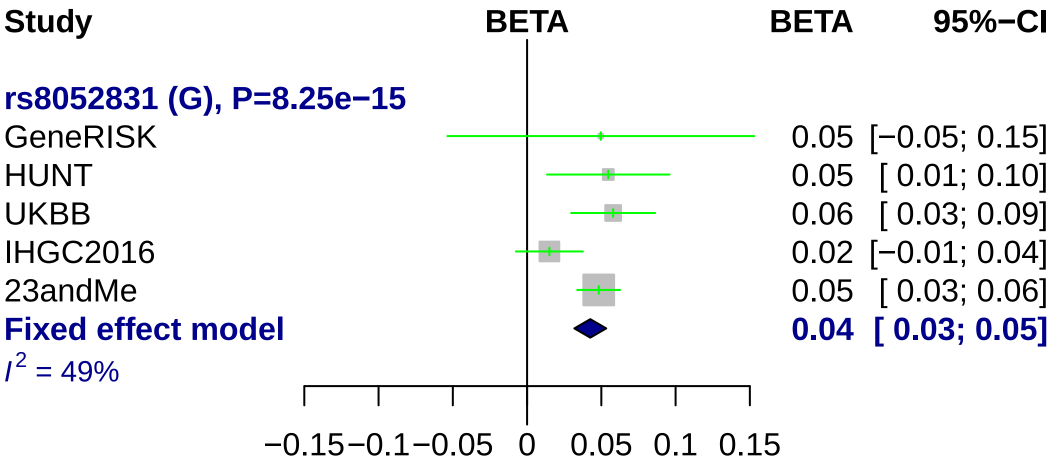

Study
BETA
BETA
95%−CI
rs8052831 (G), P=8.25e−15
GeneRISK 0.05 [−0.05; 0.15]
HUNT 0.05 [ 0.01; 0.10]
UKBB 0.06 [ 0.03; 0.09]
IHGC2016 0.02 [−0.01; 0.04]
23andMe 0.05 [ 0.03; 0.06]
Fixed effect model 0.04 [ 0.03; 0.05]
$I^2$ = 49%
−0.15 −0.1 −0.05 0 0.05 0.1 0.15

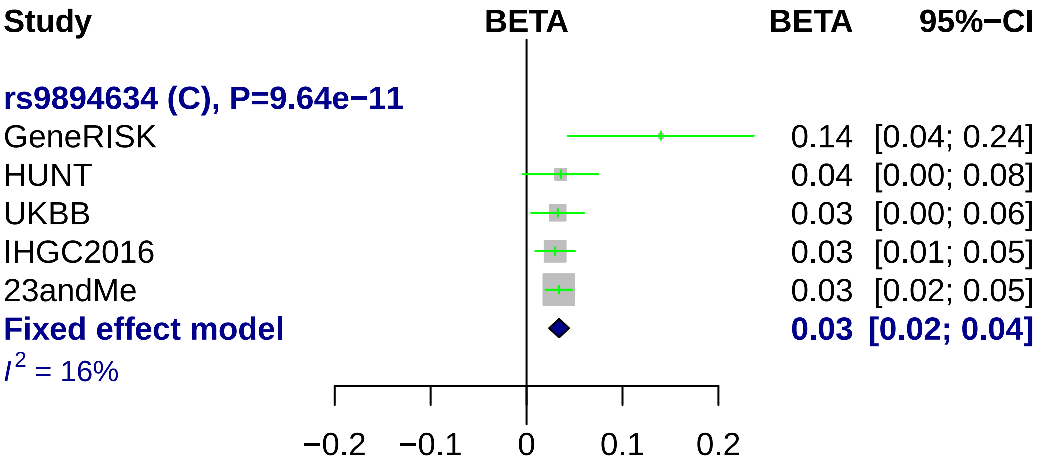
Study
BETA
BETA
95%−CI
rs9894634 (C), P=9.64e−11
GeneRISK 0.14 [0.04; 0.24]
HUNT 0.04 [0.00; 0.08]
UKBB 0.03 [0.00; 0.06]
IHGC2016 0.03 [0.01; 0.05]
23andMe 0.03 [0.02; 0.05]
Fixed effect model 0.03 [0.02; 0.04]
I2 = 16%
−0.2
−0.1
0
0.1
0.2

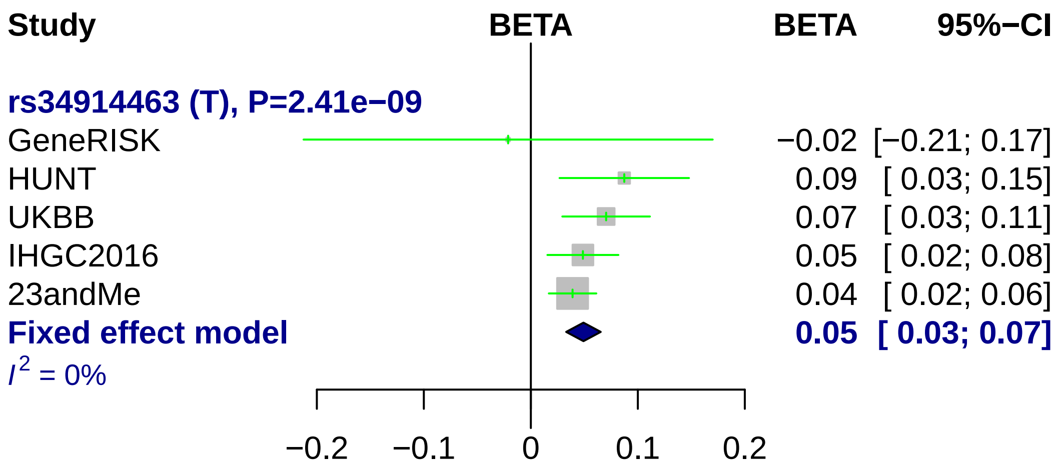
Study
BETA
BETA
95%−CI
rs34914463 (T), P=2.41e−09
GeneRISK
−0.02 [−0.21; 0.17]
HUNT
0.09 [ 0.03; 0.15]
UKBB
0.07 [ 0.03; 0.11]
IHGC2016
0.05 [ 0.02; 0.08]
23andMe
0.04 [ 0.02; 0.06]
Fixed effect model
0.05 [ 0.03; 0.07]
I^2 = 0%
−0.2
−0.1
0
0.1
0.2

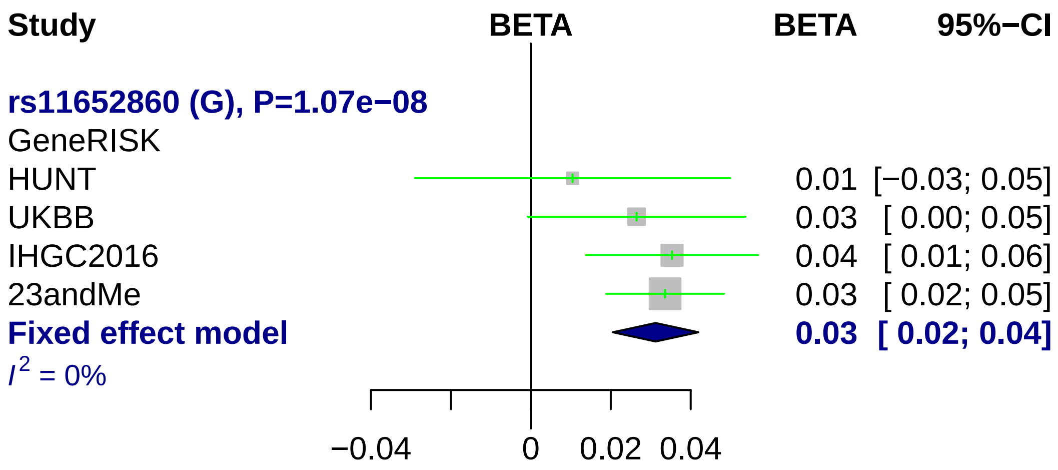
Study
BETA
BETA
95%−CI
rs11652860 (G), P=1.07e−08
GeneRISK
HUNT
0.01 [−0.03; 0.05]
UKBB
0.03 [ 0.00; 0.05]
IHGC2016
0.04 [ 0.01; 0.06]
23andMe
0.03 [ 0.02; 0.05]
Fixed effect model
0.03 [ 0.02; 0.04]
I2 = 0%
−0.04
0
0.02
0.04

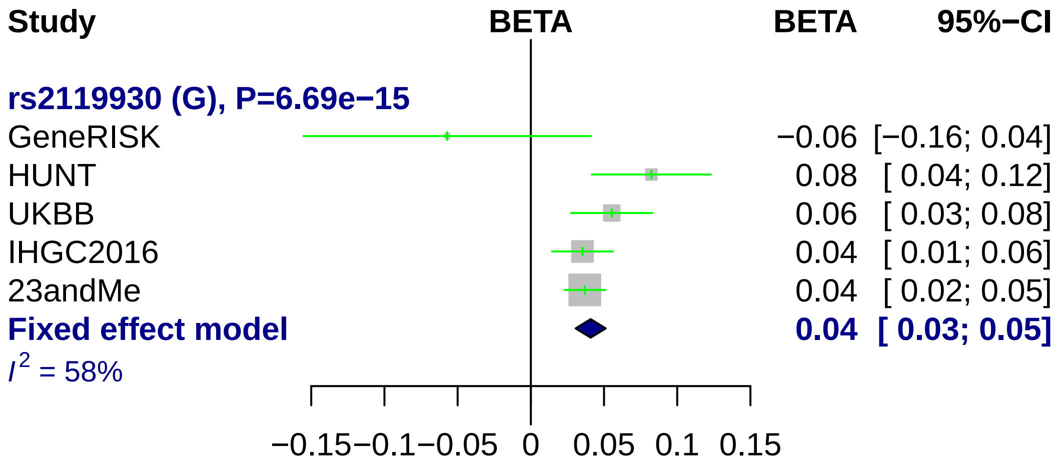
Study
BETA
BETA
95%−CI
rs2119930 (G), P=6.69e−15
GeneRISK −0.06 [−0.16; 0.04]
HUNT 0.08 [ 0.04; 0.12]
UKBB 0.06 [ 0.03; 0.08]
IHGC2016 0.04 [ 0.01; 0.06]
23andMe 0.04 [ 0.02; 0.05]
Fixed effect model 0.04 [ 0.03; 0.05]
$I^2$ = 58%
−0.15 −0.1 −0.05 0 0.05 0.1 0.15

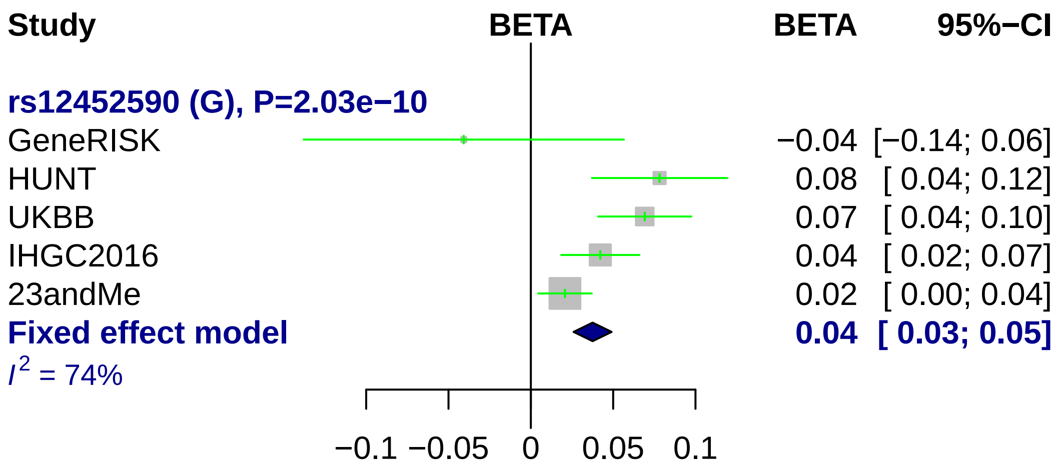

Study
BETA
BETA
95%−CI
rs12452590 (G), P=2.03e−10
GeneRISK −0.04 [−0.14; 0.06]
HUNT 0.08 [ 0.04; 0.12]
UKBB 0.07 [ 0.04; 0.10]
IHGC2016 0.04 [ 0.02; 0.07]
23andMe 0.02 [ 0.00; 0.04]
Fixed effect model 0.04 [ 0.03; 0.05]
$I^2$ = 74%
−0.1 −0.05 0 0.05 0.1

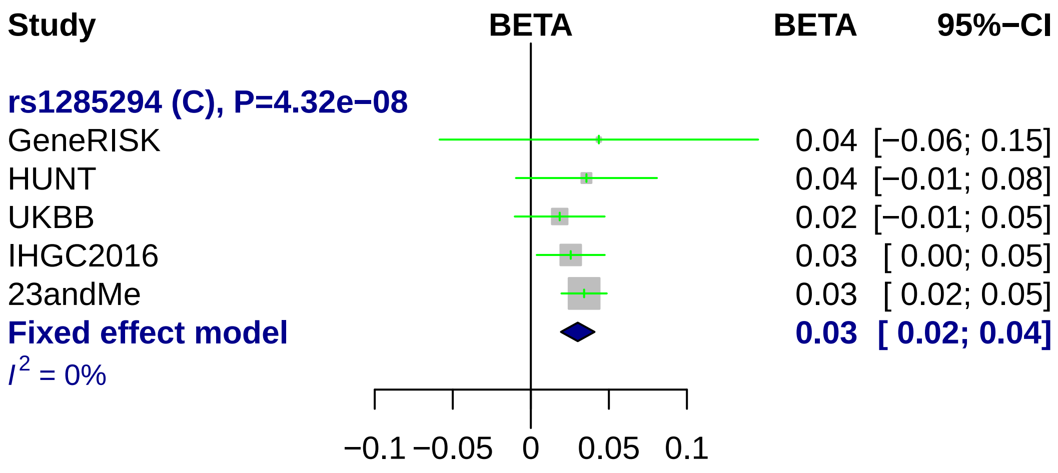

Study
BETA
BETA
95%−CI
rs1285294 (C), P=4.32e−08
GeneRISK 0.04 [−0.06; 0.15]
HUNT 0.04 [−0.01; 0.08]
UKBB 0.02 [−0.01; 0.05]
IHGC2016 0.03 [ 0.00; 0.05]
23andMe 0.03 [ 0.02; 0.05]
Fixed effect model 0.03 [ 0.02; 0.04]
$I^2$ = 0%
−0.1
−0.05
0
0.05
0.1

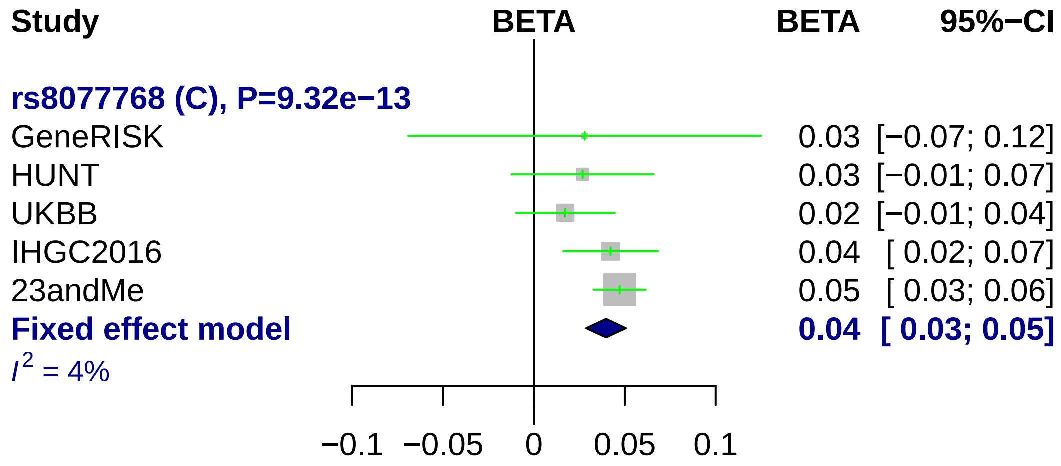
Study
BETA
BETA
95%−CI
rs8077768 (C), P=9.32e−13
GeneRISK
0.03 [−0.07; 0.12]
HUNT
0.03 [−0.01; 0.07]
UKBB
0.02 [−0.01; 0.04]
IHGC2016
0.04 [ 0.02; 0.07]
23andMe
0.05 [ 0.03; 0.06]
Fixed effect model
0.04 [ 0.03; 0.05]
I2 = 4%
−0.1
−0.05
0
0.05
0.1

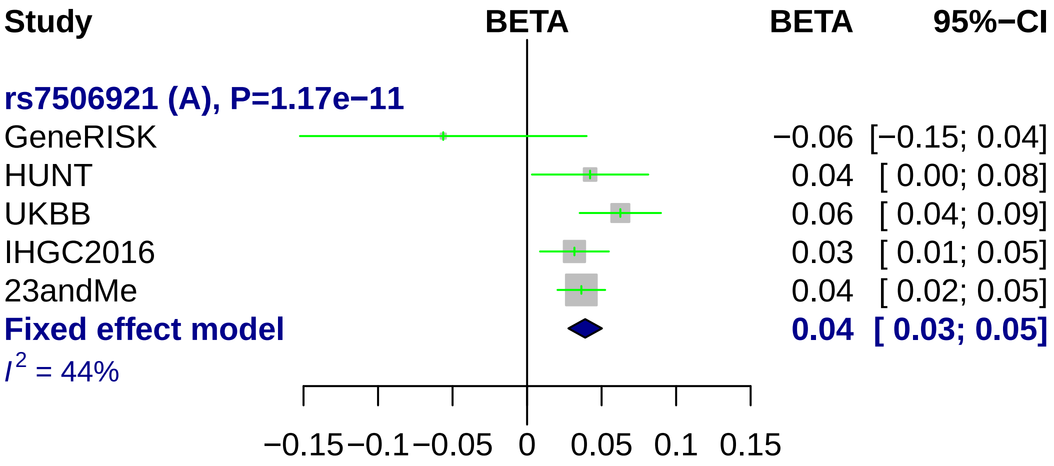

Study
BETA
BETA
95%−CI
rs7506921 (A), P=1.17e−11
GeneRISK
−0.06 [−0.15; 0.04]
HUNT
0.04 [ 0.00; 0.08]
UKBB
0.06 [ 0.04; 0.09]
IHGC2016
0.03 [ 0.01; 0.05]
23andMe
0.04 [ 0.02; 0.05]
Fixed effect model
0.04 [ 0.03; 0.05]
I2 = 44%
−0.15
−0.1
−0.05
0
0.05
0.1
0.15

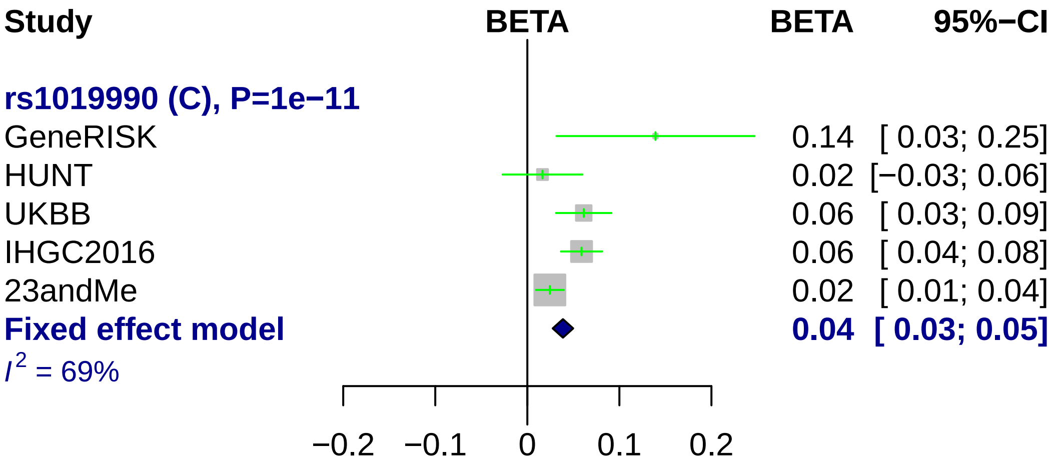

Study
BETA
BETA
95%−CI
rs1019990 (C), P=1e−11
GeneRISK
0.14 [ 0.03; 0.25]
HUNT
0.02 [−0.03; 0.06]
UKBB
0.06 [ 0.03; 0.09]
IHGC2016
0.06 [ 0.04; 0.08]
23andMe
0.02 [ 0.01; 0.04]
Fixed effect model
0.04 [ 0.03; 0.05]
I² = 69%
−0.2
−0.1
0
0.1
0.2

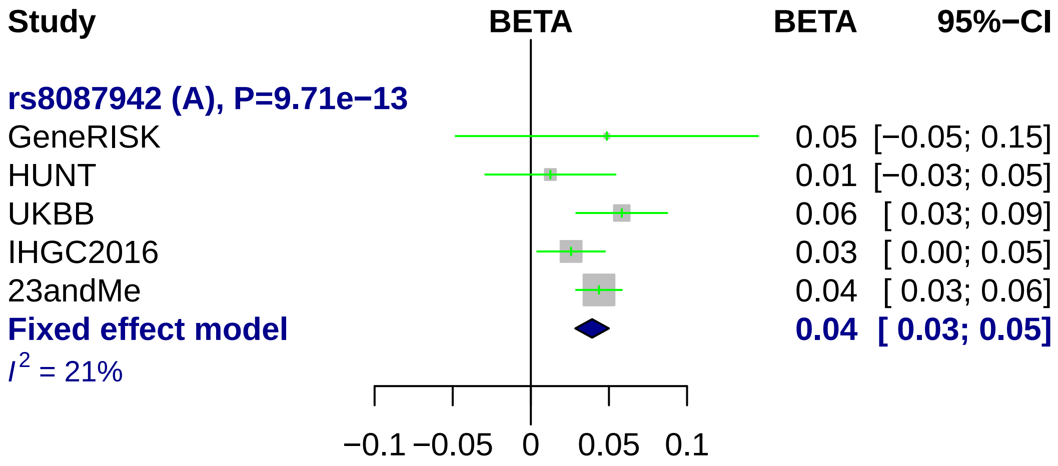
Study
BETA
BETA
95%−CI
rs8087942 (A), P=9.71e−13
GeneRISK 0.05 [−0.05; 0.15]
HUNT 0.01 [−0.03; 0.05]
UKBB 0.06 [ 0.03; 0.09]
IHGC2016 0.03 [ 0.00; 0.05]
23andMe 0.04 [ 0.03; 0.06]
Fixed effect model 0.04 [ 0.03; 0.05]
$I^2$ = 21%
−0.1
−0.05
0
0.05
0.1

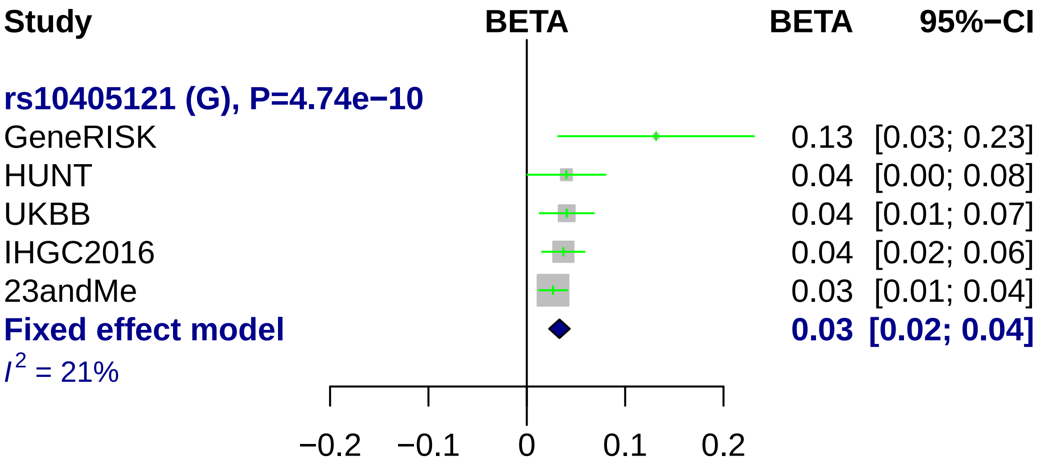
Study
BETA
BETA
95%−CI
rs10405121 (G), P=4.74e−10
GeneRISK
0.13 [0.03; 0.23]
HUNT
0.04 [0.00; 0.08]
UKBB
0.04 [0.01; 0.07]
IHGC2016
0.04 [0.02; 0.06]
23andMe
0.03 [0.01; 0.04]
Fixed effect model
0.03 [0.02; 0.04]
I^2 = 21%
−0.2
−0.1
0
0.1
0.2

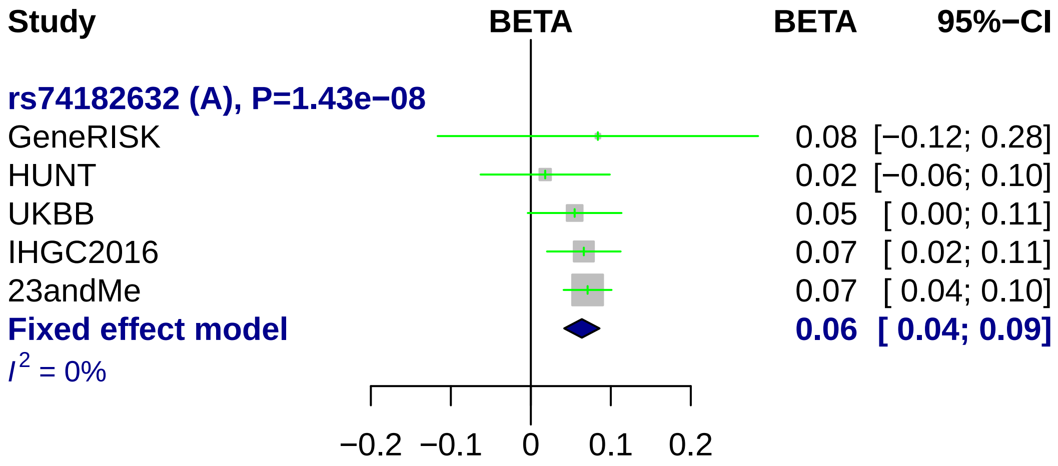
Study
BETA
BETA
95%−CI
rs74182632 (A), P=1.43e−08
GeneRISK
0.08 [−0.12; 0.28]
HUNT
0.02 [−0.06; 0.10]
UKBB
0.05 [ 0.00; 0.11]
IHGC2016
0.07 [ 0.02; 0.11]
23andMe
0.07 [ 0.04; 0.10]
Fixed effect model
0.06 [ 0.04; 0.09]
I2 = 0%
−0.2
−0.1
0
0.1
0.2

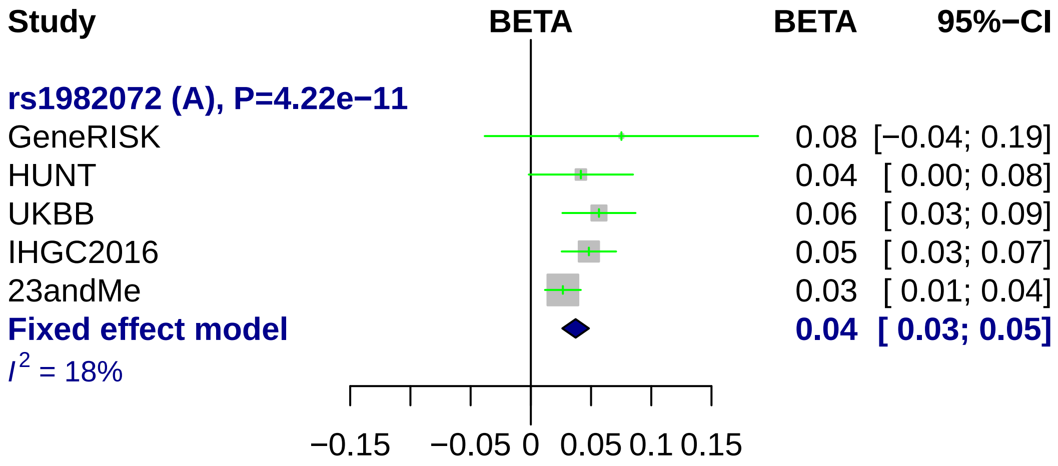
Study
BETA
BETA
95%−CI
rs1982072 (A), P=4.22e−11
GeneRISK
0.08 [−0.04; 0.19]
HUNT
0.04 [ 0.00; 0.08]
UKBB
0.06 [ 0.03; 0.09]
IHGC2016
0.05 [ 0.03; 0.07]
23andMe
0.03 [ 0.01; 0.04]
Fixed effect model
0.04 [ 0.03; 0.05]
I2 = 18%
−0.15
−0.05
0
0.05
0.1
0.15

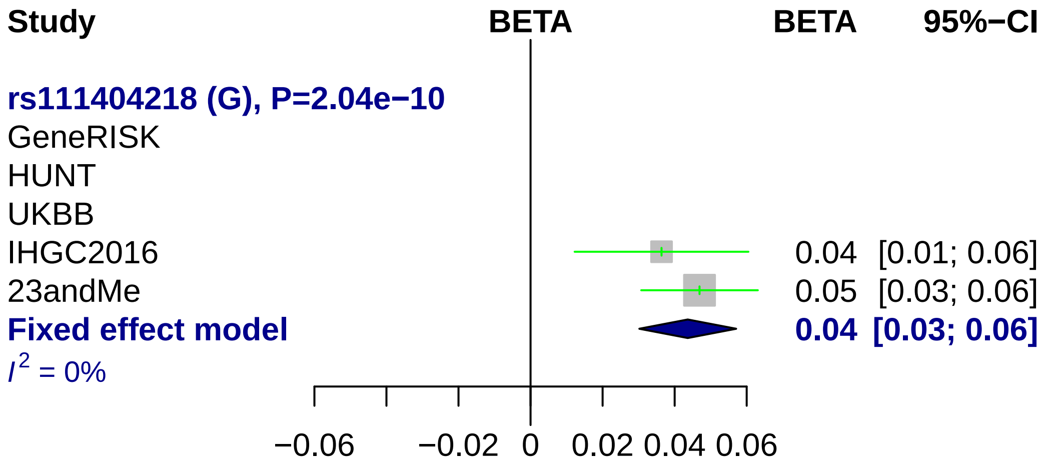
Study
BETA
BETA
95%−CI
rs111404218 (G), P=2.04e−10
GeneRISK
HUNT
UKBB
IHGC2016
0.04
[0.01; 0.06]
23andMe
0.05
[0.03; 0.06]
Fixed effect model
0.04
[0.03; 0.06]
$I^2$ = 0%
−0.06
−0.02
0
0.02
0.04
0.06

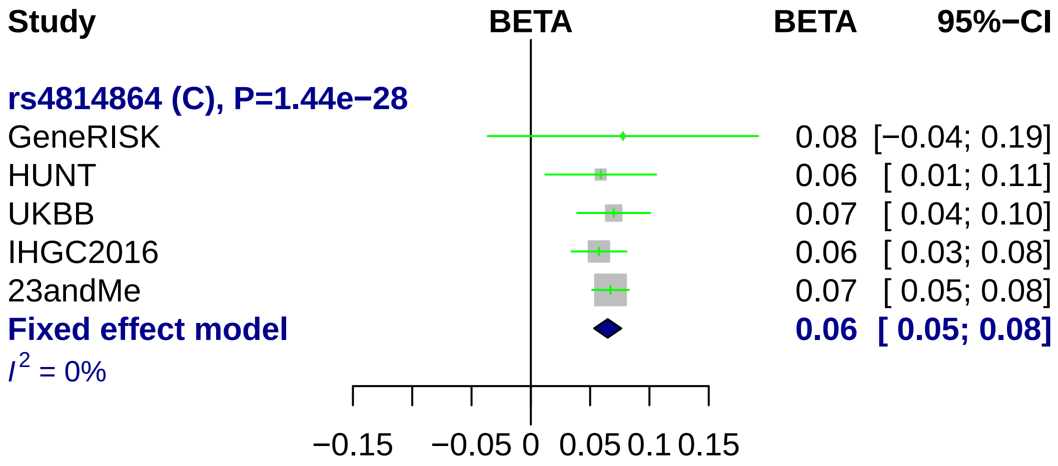
Study
BETA
BETA
95%−CI
rs4814864 (C), P=1.44e−28
GeneRISK
0.08 [−0.04; 0.19]
HUNT
0.06 [ 0.01; 0.11]
UKBB
0.07 [ 0.04; 0.10]
IHGC2016
0.06 [ 0.03; 0.08]
23andMe
0.07 [ 0.05; 0.08]
Fixed effect model
0.06 [ 0.05; 0.08]
I 2 = 0%
−0.15
−0.05
0
0.05
0.1
0.15

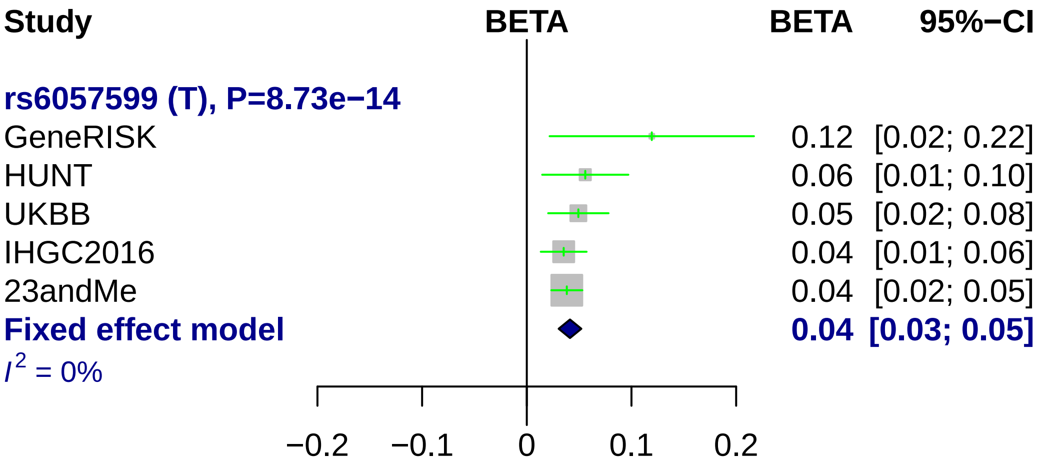
Study
BETA
BETA
95%−CI
rs6057599 (T), P=8.73e−14
GeneRISK 0.12 [0.02; 0.22]
HUNT 0.06 [0.01; 0.10]
UKBB 0.05 [0.02; 0.08]
IHGC2016 0.04 [0.01; 0.06]
23andMe 0.04 [0.02; 0.05]
Fixed effect model 0.04 [0.03; 0.05]
I^2 = 0%
−0.2
−0.1
0
0.1
0.2

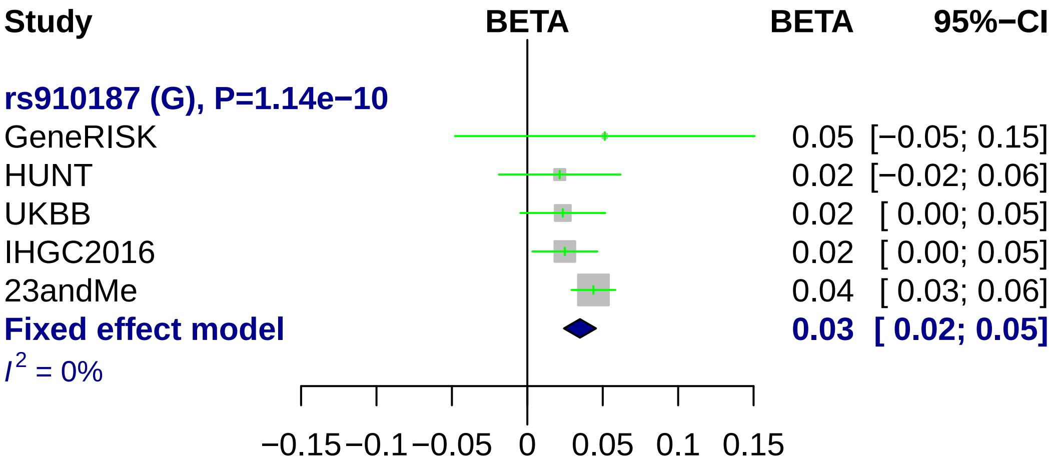
Study
BETA
BETA
95%−CI
rs910187 (G), P=1.14e−10
GeneRISK
0.05 [−0.05; 0.15]
HUNT
0.02 [−0.02; 0.06]
UKBB
0.02 [ 0.00; 0.05]
IHGC2016
0.02 [ 0.00; 0.05]
23andMe
0.04 [ 0.03; 0.06]
Fixed effect model
0.03 [ 0.02; 0.05]
I^2 = 0%
−0.15 −0.1 −0.05 0 0.05 0.1 0.15

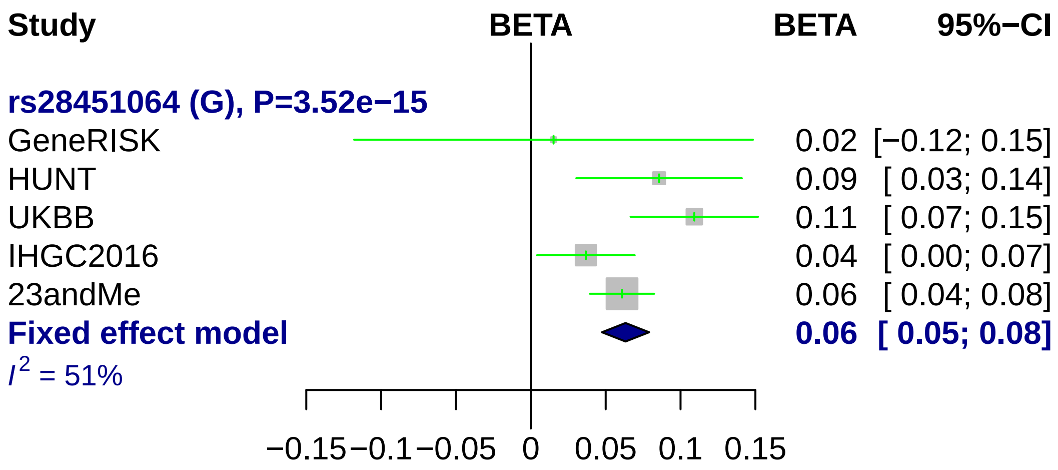
Study
BETA
BETA
95%−CI
rs28451064 (G), P=3.52e−15
GeneRISK
0.02
[−0.12; 0.15]
HUNT
0.09
[ 0.03; 0.14]
UKBB
0.11
[ 0.07; 0.15]
IHGC2016
0.04
[ 0.00; 0.07]
23andMe
0.06
[ 0.04; 0.08]
Fixed effect model
0.06
[ 0.05; 0.08]
I2 = 51%
−0.15
−0.1
−0.05
0
0.05
0.1
0.15

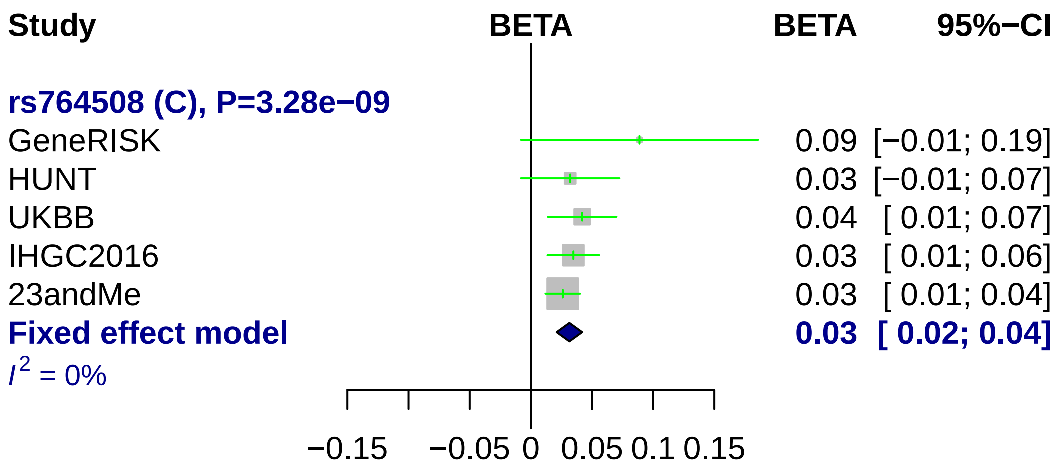

Study
BETA
BETA
95%−CI
rs764508 (C), P=3.28e−09
GeneRISK
0.09 [−0.01; 0.19]
HUNT
0.03 [−0.01; 0.07]
UKBB
0.04 [ 0.01; 0.07]
IHGC2016
0.03 [ 0.01; 0.06]
23andMe
0.03 [ 0.01; 0.04]
Fixed effect model
0.03 [ 0.02; 0.04]
I2 = 0%
−0.15
−0.05
0
0.05
0.1
0.15

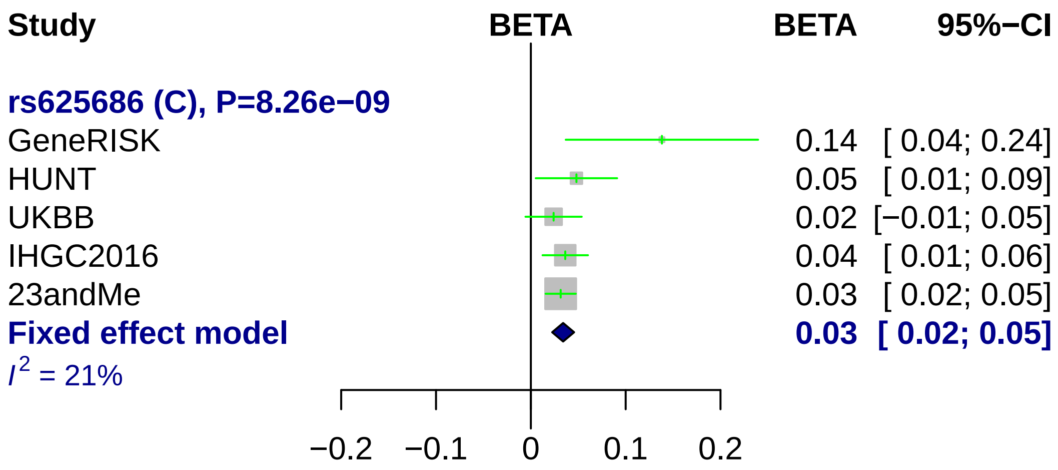
Study
BETA
BETA
95%−CI
rs625686 (C), P=8.26e−09
GeneRISK
0.14 [ 0.04; 0.24]
HUNT
0.05 [ 0.01; 0.09]
UKBB
0.02 [−0.01; 0.05]
IHGC2016
0.04 [ 0.01; 0.06]
23andMe
0.03 [ 0.02; 0.05]
Fixed effect model
0.03 [ 0.02; 0.05]
I2 = 21%
−0.2
−0.1
0
0.1
0.2

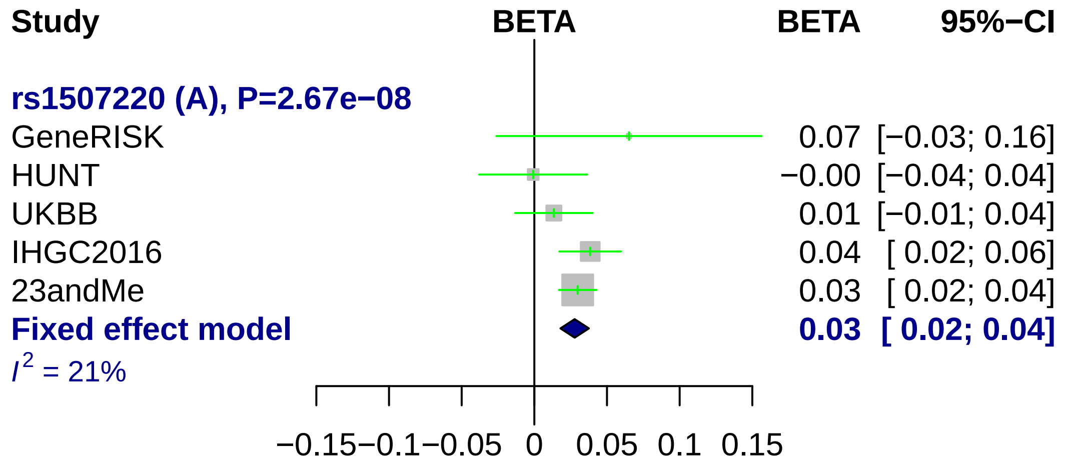
Study
BETA
BETA
95%−CI
rs1507220 (A), P=2.67e−08
GeneRISK
0.07 [−0.03; 0.16]
HUNT
−0.00 [−0.04; 0.04]
UKBB
0.01 [−0.01; 0.04]
IHGC2016
0.04 [ 0.02; 0.06]
23andMe
0.03 [ 0.02; 0.04]
Fixed effect model
0.03 [ 0.02; 0.04]
I2 = 21%
−0.15
−0.1
−0.05
0
0.05
0.1
0.15

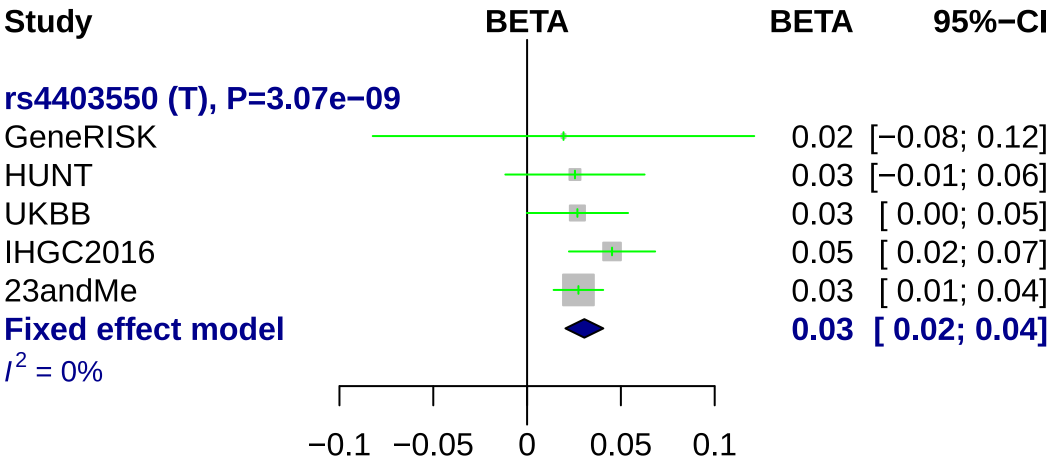

Study
BETA
BETA
95%−CI
rs4403550 (T), P=3.07e−09
GeneRISK
0.02 [−0.08; 0.12]
HUNT
0.03 [−0.01; 0.06]
UKBB
0.03 [ 0.00; 0.05]
IHGC2016
0.05 [ 0.02; 0.07]
23andMe
0.03 [ 0.01; 0.04]
Fixed effect model
0.03 [ 0.02; 0.04]
I2 = 0%
−0.1
−0.05
0
0.05
0.1
