## Supplementary Figure 8. for "Genome-wide analysis of 102,084 migraine cases identifies 123 risk loci and subtype-specific risk alleles"

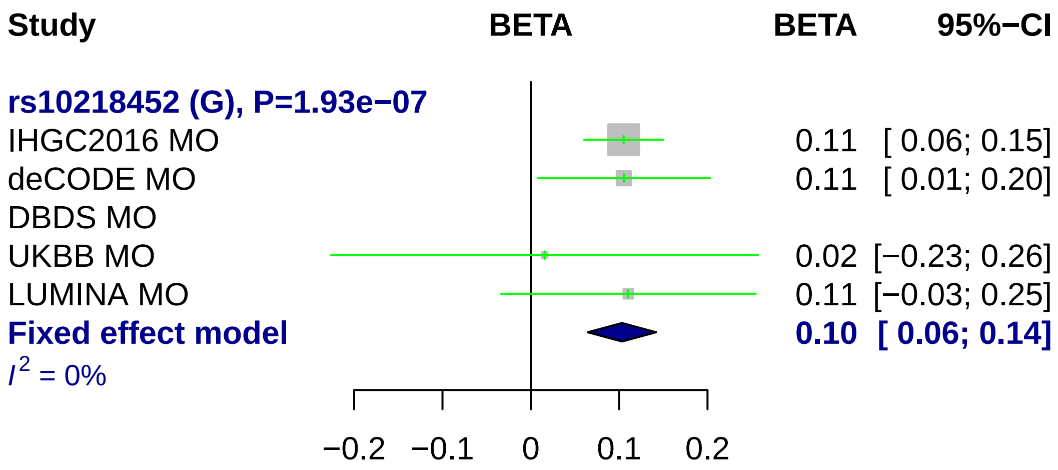
Study
BETA
BETA
95%−CI
rs10218452 (G), P=1.93e−07
IHGC2016 MO
0.11 [ 0.06; 0.15]
deCODE MO
0.11 [ 0.01; 0.20]
DBDS MO
UKBB MO
0.02 [−0.23; 0.26]
LUMINA MO
0.11 [−0.03; 0.25]
Fixed effect model
0.10 [ 0.06; 0.14]
I2 = 0%
−0.2
−0.1
0
0.1
0.2

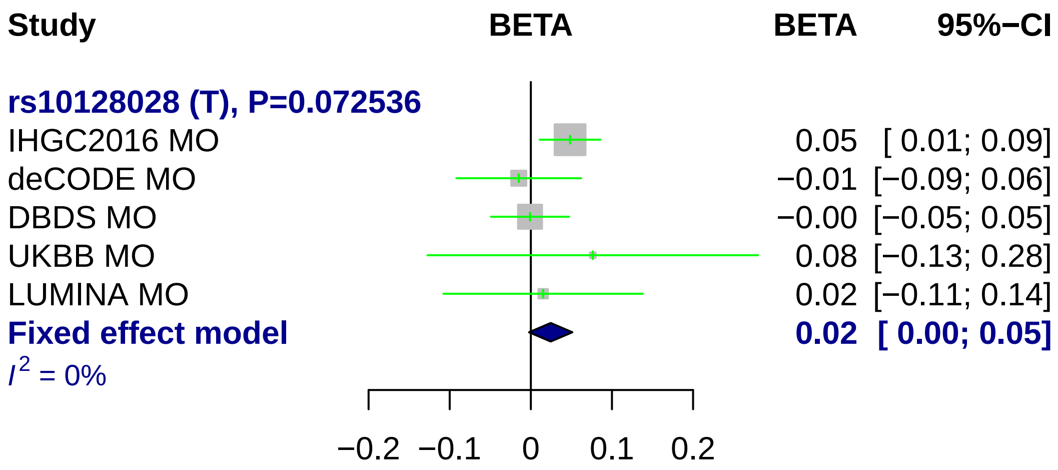

Study
BETA
BETA
95%−CI
rs10128028 (T), P=0.072536
IHGC2016 MO
0.05 [ 0.01; 0.09]
deCODE MO
−0.01 [−0.09; 0.06]
DBDS MO
−0.00 [−0.05; 0.05]
UKBB MO
0.08 [−0.13; 0.28]
LUMINA MO
0.02 [−0.11; 0.14]
Fixed effect model
0.02 [ 0.00; 0.05]
I2 = 0%
−0.2
−0.1
0
0.1
0.2

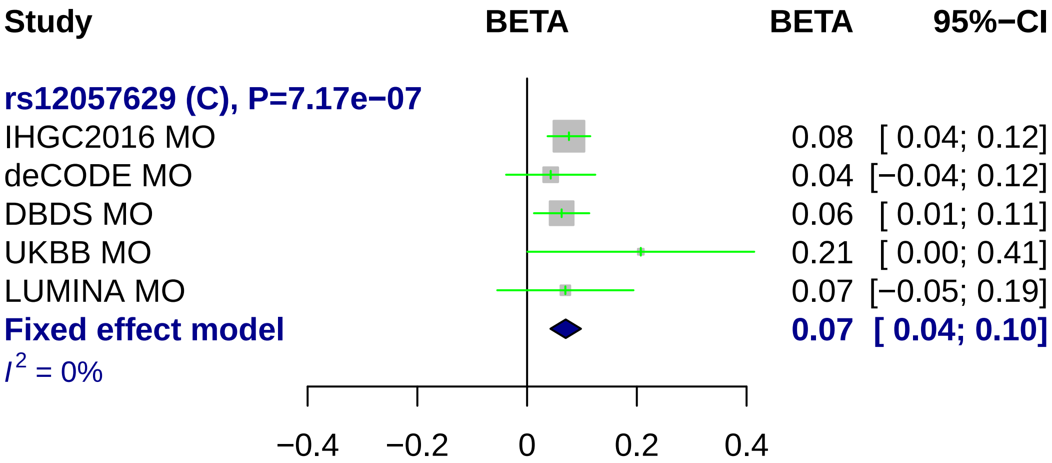
Study
BETA
BETA
95%−CI
rs12057629 (C), P=7.17e−07
IHGC2016 MO
0.08 [ 0.04; 0.12]
deCODE MO
0.04 [−0.04; 0.12]
DBDS MO
0.06 [ 0.01; 0.11]
UKBB MO
0.21 [ 0.00; 0.41]
LUMINA MO
0.07 [−0.05; 0.19]
Fixed effect model
0.07 [ 0.04; 0.10]
I2 = 0%
−0.4
−0.2
0
0.2
0.4

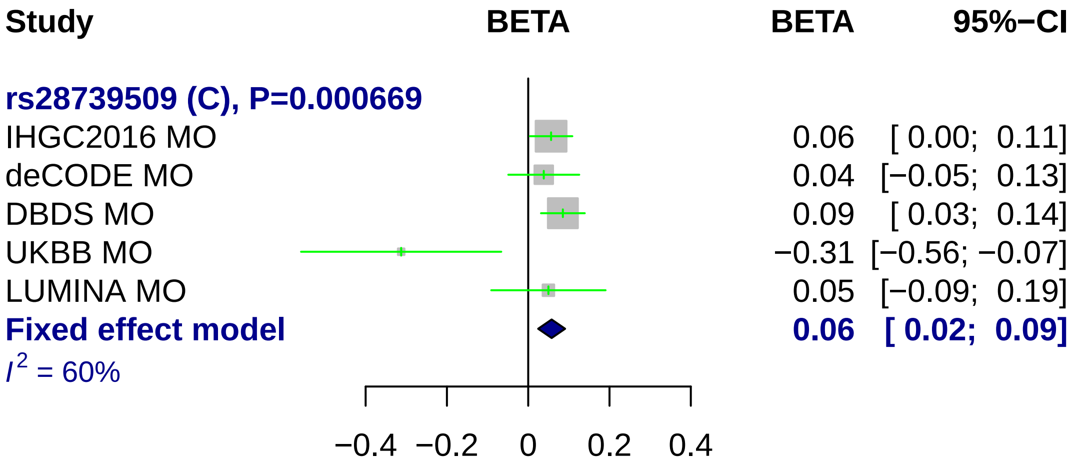
Study
BETA
BETA
95%−CI
rs28739509 (C), P=0.000669
IHGC2016 MO
0.06 [ 0.00; 0.11]
deCODE MO
0.04 [−0.05; 0.13]
DBDS MO
0.09 [ 0.03; 0.14]
UKBB MO
−0.31 [−0.56; −0.07]
LUMINA MO
0.05 [−0.09; 0.19]
Fixed effect model
0.06 [ 0.02; 0.09]
I^2 = 60%
−0.4 −0.2 0 0.2 0.4

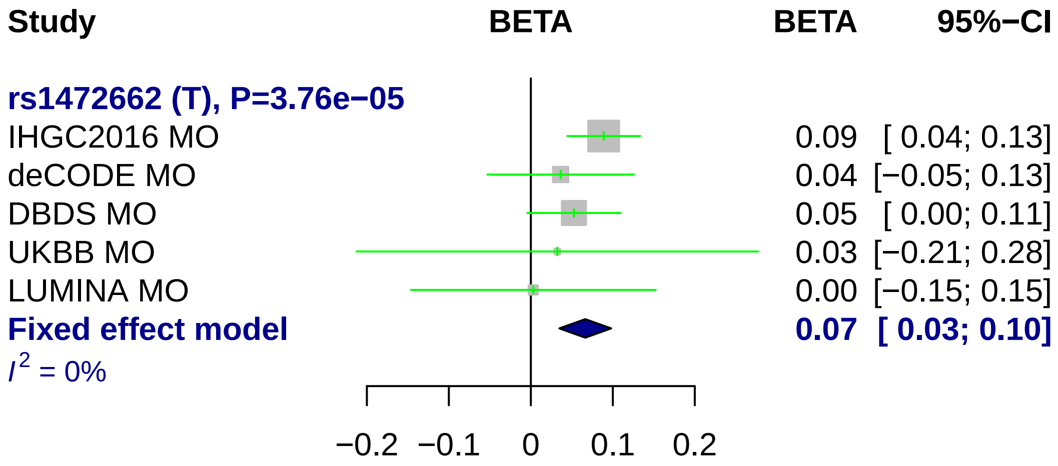

Study
BETA
BETA
95%−CI
rs1472662 (T), P=3.76e−05
IHGC2016 MO
0.09 [ 0.04; 0.13]
deCODE MO
0.04 [−0.05; 0.13]
DBDS MO
0.05 [ 0.00; 0.11]
UKBB MO
0.03 [−0.21; 0.28]
LUMINA MO
0.00 [−0.15; 0.15]
Fixed effect model
0.07 [ 0.03; 0.10]
I2 = 0%
−0.2
−0.1
0
0.1
0.2

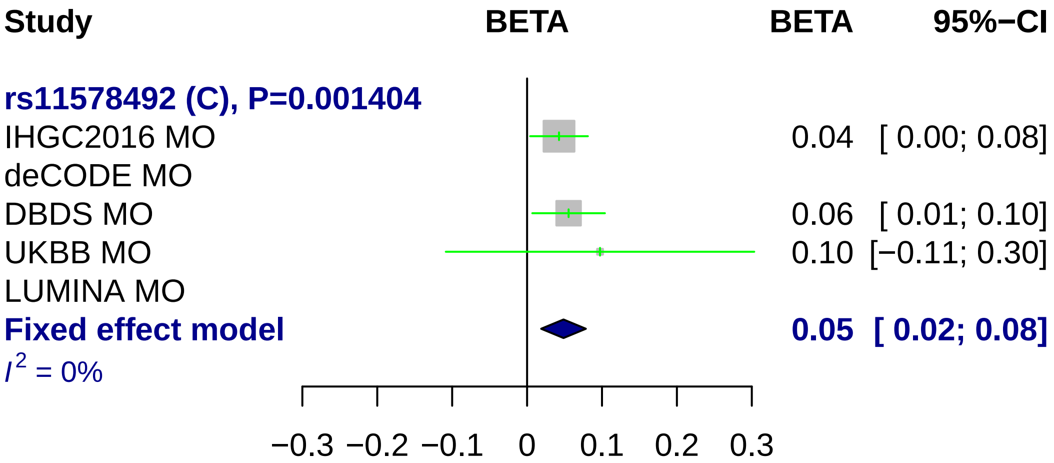

Study
BETA
BETA
95%−CI
rs11578492 (C), P=0.001404
IHGC2016 MO
0.04 [ 0.00; 0.08]
deCODE MO
DBDS MO
0.06 [ 0.01; 0.10]
UKBB MO
0.10 [−0.11; 0.30]
LUMINA MO
Fixed effect model
0.05 [ 0.02; 0.08]
I2 = 0%
−0.3 −0.2 −0.1 0 0.1 0.2 0.3

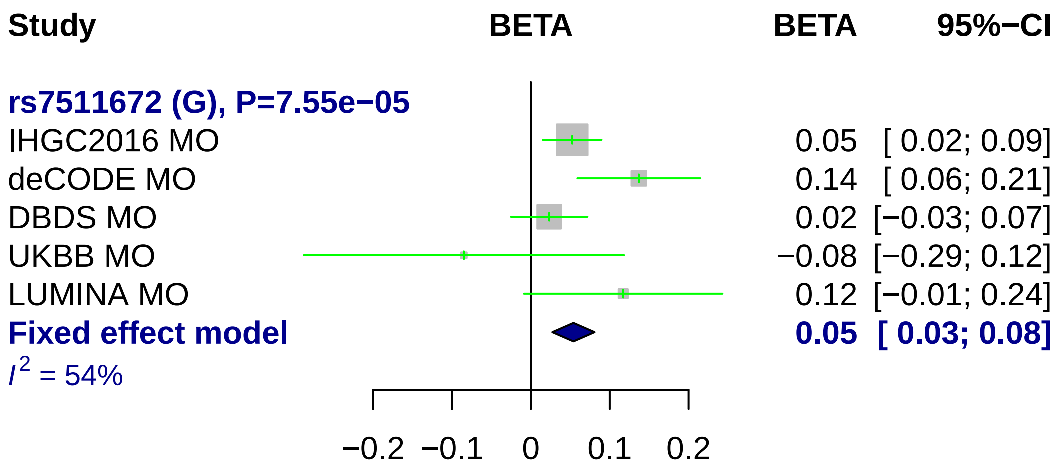
Study
BETA
BETA
95%−CI
rs7511672 (G), P=7.55e−05
IHGC2016 MO 0.05 [ 0.02; 0.09]
deCODE MO 0.14 [ 0.06; 0.21]
DBDS MO 0.02 [−0.03; 0.07]
UKBB MO −0.08 [−0.29; 0.12]
LUMINA MO 0.12 [−0.01; 0.24]
Fixed effect model 0.05 [ 0.03; 0.08]
I^2 = 54%
−0.2
−0.1
0
0.1
0.2

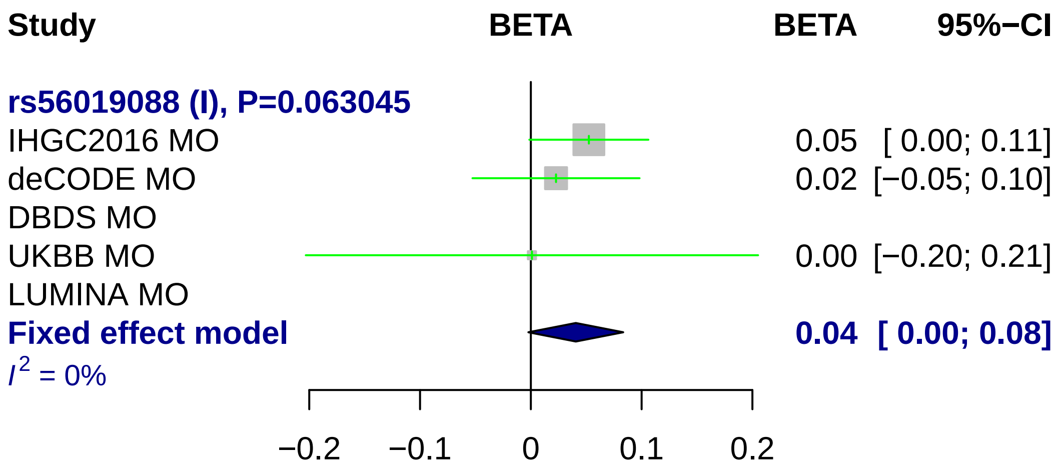
Study
BETA
BETA
95%−CI
rs56019088 (I), P=0.063045
IHGC2016 MO
0.05
[ 0.00; 0.11]
deCODE MO
0.02
[−0.05; 0.10]
DBDS MO
UKBB MO
0.00
[−0.20; 0.21]
LUMINA MO
Fixed effect model
0.04
[ 0.00; 0.08]
I^2 = 0%
−0.2
−0.1
0
0.1
0.2

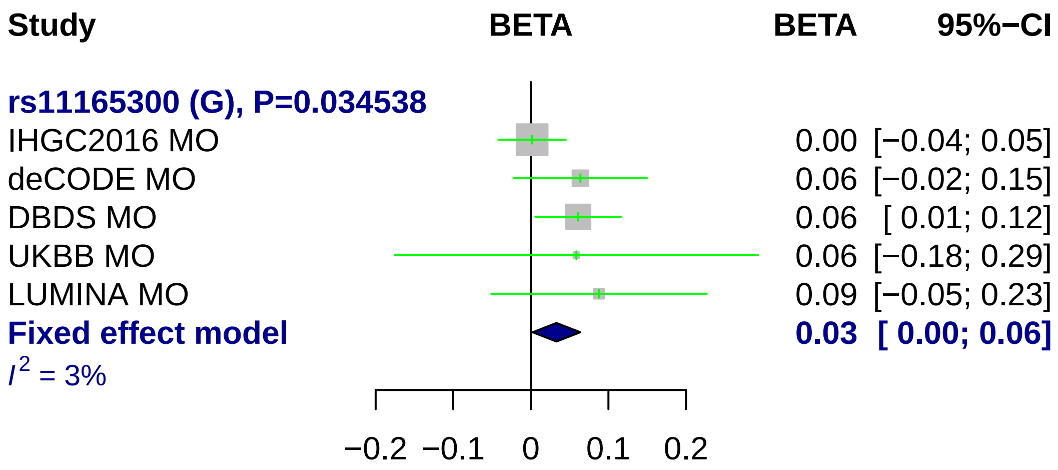
Study
BETA
BETA
95%−CI
rs11165300 (G), P=0.034538
IHGC2016 MO
0.00 [−0.04; 0.05]
deCODE MO
0.06 [−0.02; 0.15]
DBDS MO
0.06 [ 0.01; 0.12]
UKBB MO
0.06 [−0.18; 0.29]
LUMINA MO
0.09 [−0.05; 0.23]
Fixed effect model
0.03 [ 0.00; 0.06]
$I^2$ = 3%
−0.2
−0.1
0
0.1
0.2

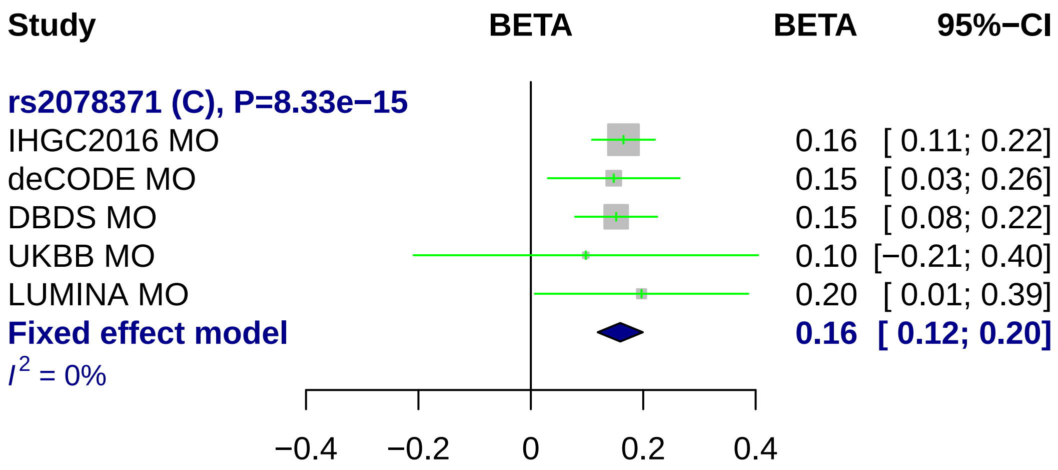

Study
BETA
BETA
95%−CI
rs2078371 (C), P=8.33e−15
IHGC2016 MO
0.16 [ 0.11; 0.22]
deCODE MO
0.15 [ 0.03; 0.26]
DBDS MO
0.15 [ 0.08; 0.22]
UKBB MO
0.10 [−0.21; 0.40]
LUMINA MO
0.20 [ 0.01; 0.39]
Fixed effect model
0.16 [ 0.12; 0.20]
$I^2$ = 0%
−0.4
−0.2
0
0.2
0.4

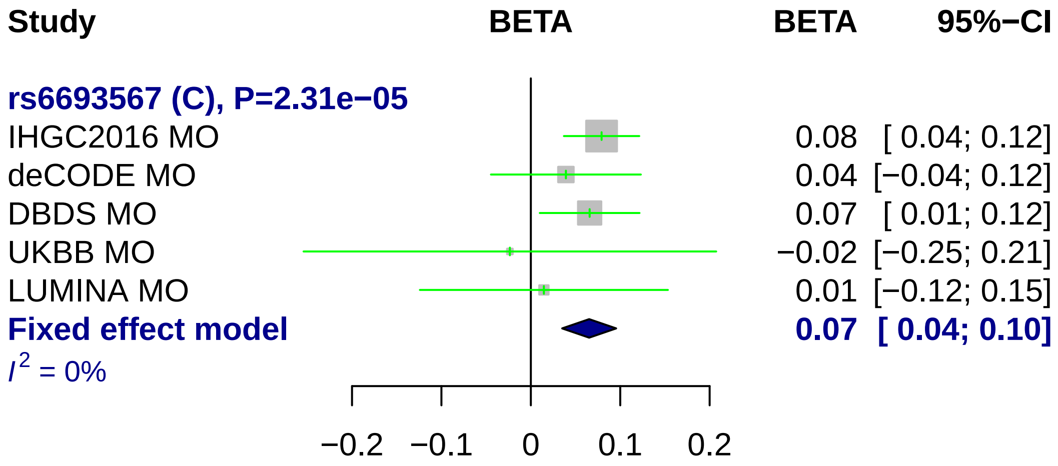

Study
BETA
BETA
95%−CI
rs6693567 (C), P=2.31e−05
IHGC2016 MO
0.08 [ 0.04; 0.12]
deCODE MO
0.04 [−0.04; 0.12]
DBDS MO
0.07 [ 0.01; 0.12]
UKBB MO
−0.02 [−0.25; 0.21]
LUMINA MO
0.01 [−0.12; 0.15]
Fixed effect model
0.07 [ 0.04; 0.10]
I2 = 0%
−0.2
−0.1
0
0.1
0.2

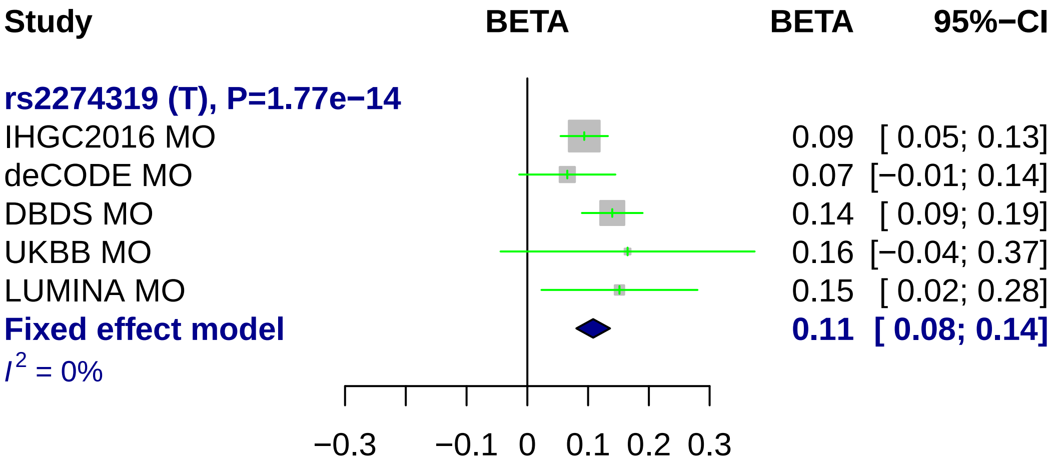
Study
BETA
BETA
95%−CI
rs2274319 (T), P=1.77e−14
IHGC2016 MO
0.09 [ 0.05; 0.13]
deCODE MO
0.07 [−0.01; 0.14]
DBDS MO
0.14 [ 0.09; 0.19]
UKBB MO
0.16 [−0.04; 0.37]
LUMINA MO
0.15 [ 0.02; 0.28]
Fixed effect model
0.11 [ 0.08; 0.14]
I2 = 0%
−0.3
−0.1
0
0.1
0.2
0.3

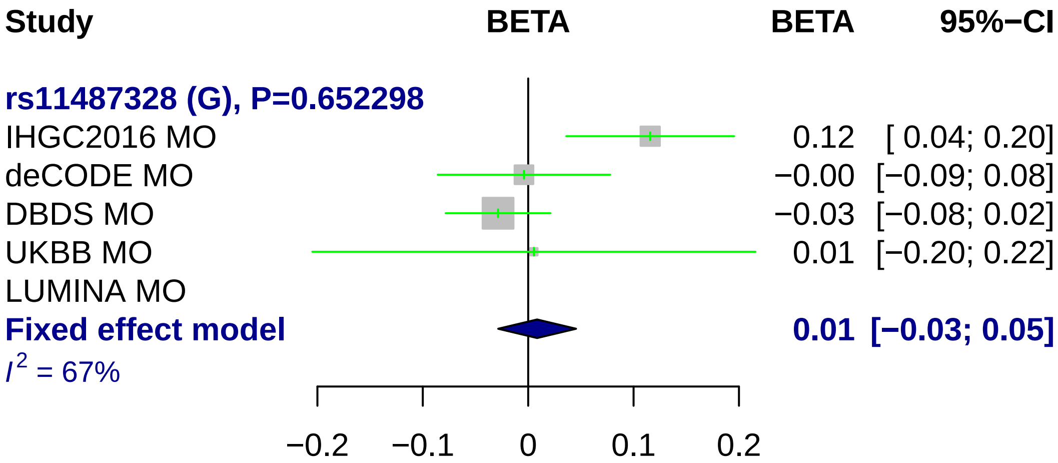

Study
BETA
BETA
95%−CI
rs11487328 (G), P=0.652298
IHGC2016 MO 0.12 [ 0.04; 0.20]
deCODE MO −0.00 [−0.09; 0.08]
DBDS MO −0.03 [−0.08; 0.02]
UKBB MO 0.01 [−0.20; 0.22]
LUMINA MO
Fixed effect model 0.01 [−0.03; 0.05]
I2 = 67%
−0.2
−0.1
0
0.1
0.2

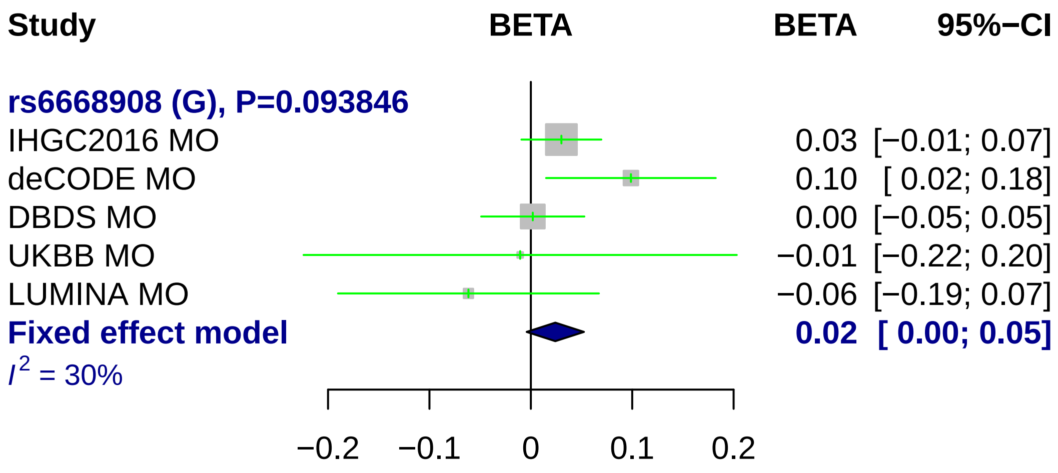
Study
BETA
BETA
95%−CI
rs6668908 (G), P=0.093846
IHGC2016 MO
0.03 [−0.01; 0.07]
deCODE MO
0.10 [ 0.02; 0.18]
DBDS MO
0.00 [−0.05; 0.05]
UKBB MO
−0.01 [−0.22; 0.20]
LUMINA MO
−0.06 [−0.19; 0.07]
Fixed effect model
0.02 [ 0.00; 0.05]
I2 = 30%
−0.2
−0.1
0
0.1
0.2

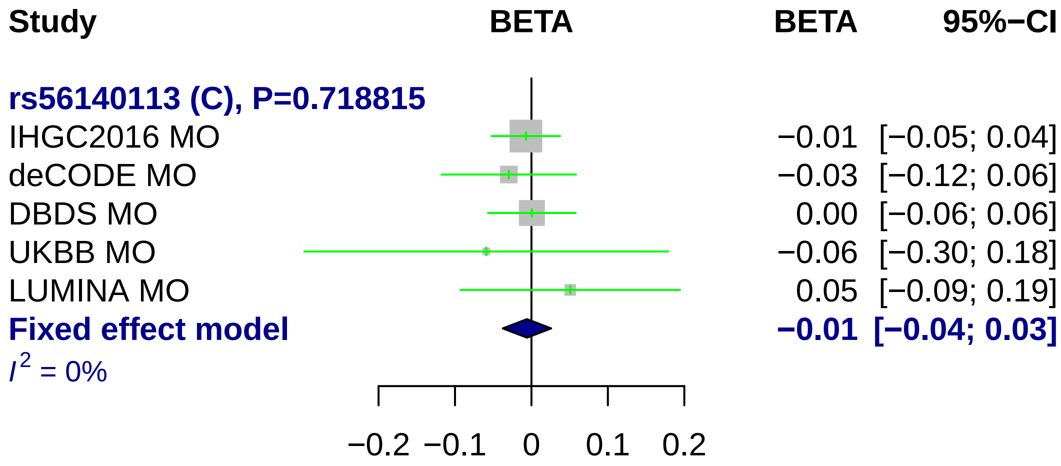
Study
BETA
BETA
95%−CI
rs56140113 (C), P=0.718815
IHGC2016 MO −0.01 [−0.05; 0.04]
deCODE MO −0.03 [−0.12; 0.06]
DBDS MO 0.00 [−0.06; 0.06]
UKBB MO −0.06 [−0.30; 0.18]
LUMINA MO 0.05 [−0.09; 0.19]
Fixed effect model −0.01 [−0.04; 0.03]
I² = 0%
−0.2 −0.1 0 0.1 0.2

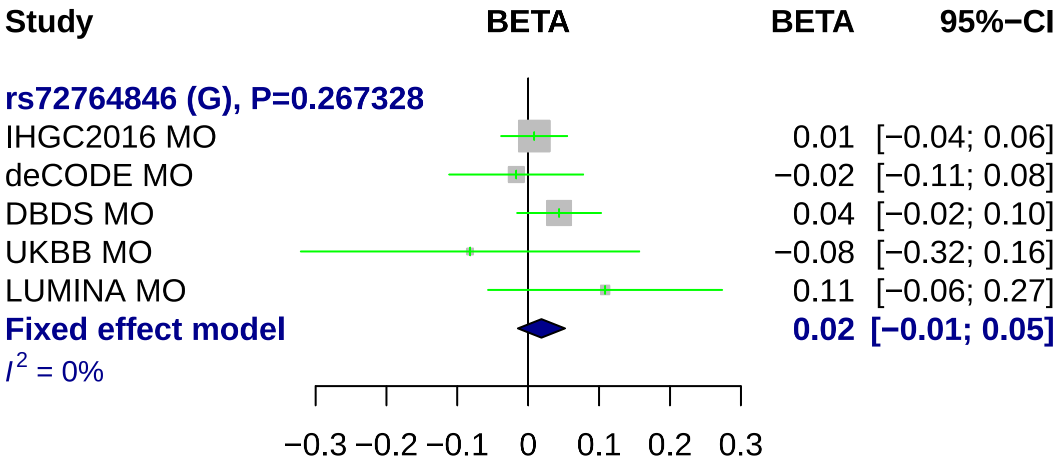
Study
BETA
BETA
95%−CI
rs72764846 (G), P=0.267328
IHGC2016 MO
0.01
[−0.04; 0.06]
deCODE MO
−0.02
[−0.11; 0.08]
DBDS MO
0.04
[−0.02; 0.10]
UKBB MO
−0.08
[−0.32; 0.16]
LUMINA MO
0.11
[−0.06; 0.27]
Fixed effect model
0.02
[−0.01; 0.05]
I2 = 0%
−0.3
−0.2
−0.1
0
0.1
0.2
0.3

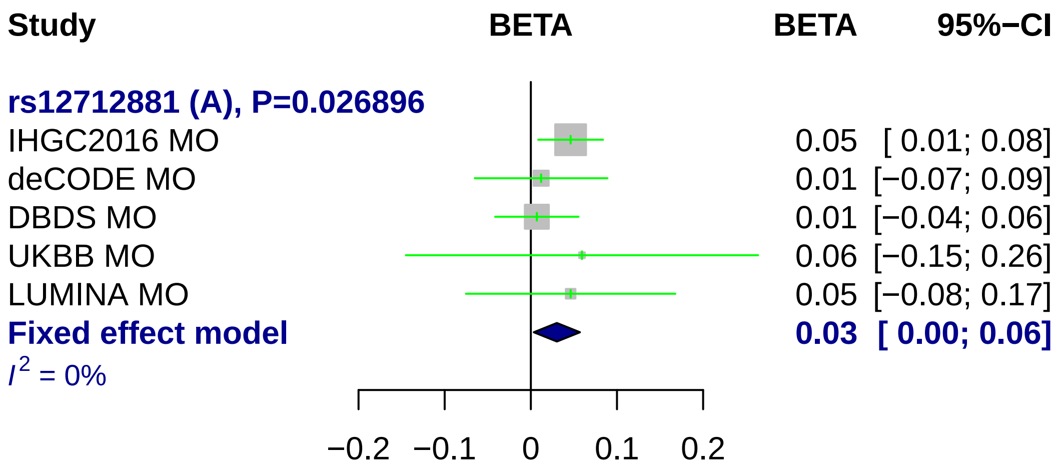
Study
BETA
BETA
95%−CI
rs12712881 (A), P=0.026896
IHGC2016 MO
0.05 [ 0.01; 0.08]
deCODE MO
0.01 [−0.07; 0.09]
DBDS MO
0.01 [−0.04; 0.06]
UKBB MO
0.06 [−0.15; 0.26]
LUMINA MO
0.05 [−0.08; 0.17]
Fixed effect model
0.03 [ 0.00; 0.06]
I² = 0%
−0.2
−0.1
0
0.1
0.2

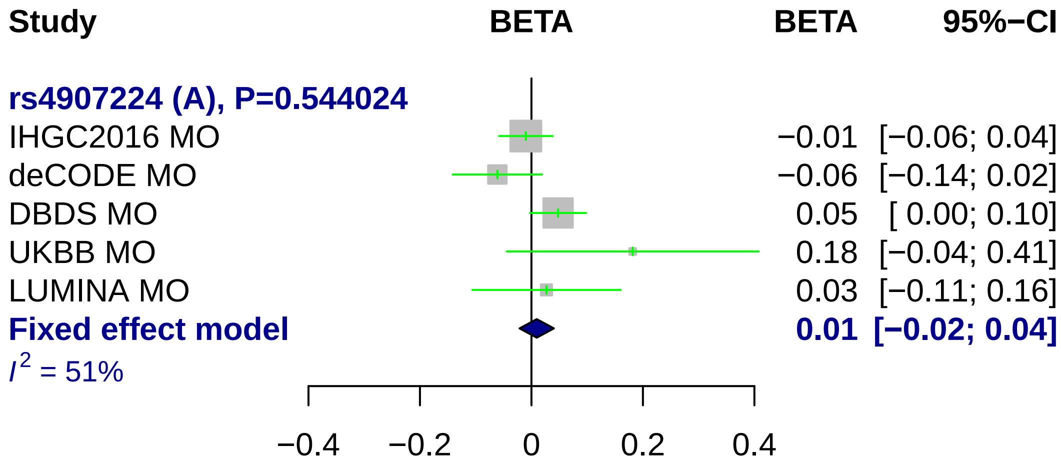
Study
BETA
BETA
95%−CI
rs4907224 (A), P=0.544024
IHGC2016 MO
−0.01 [−0.06; 0.04]
deCODE MO
−0.06 [−0.14; 0.02]
DBDS MO
0.05 [ 0.00; 0.10]
UKBB MO
0.18 [−0.04; 0.41]
LUMINA MO
0.03 [−0.11; 0.16]
Fixed effect model
0.01 [−0.02; 0.04]
$I^2$ = 51%
−0.4
−0.2
0
0.2
0.4

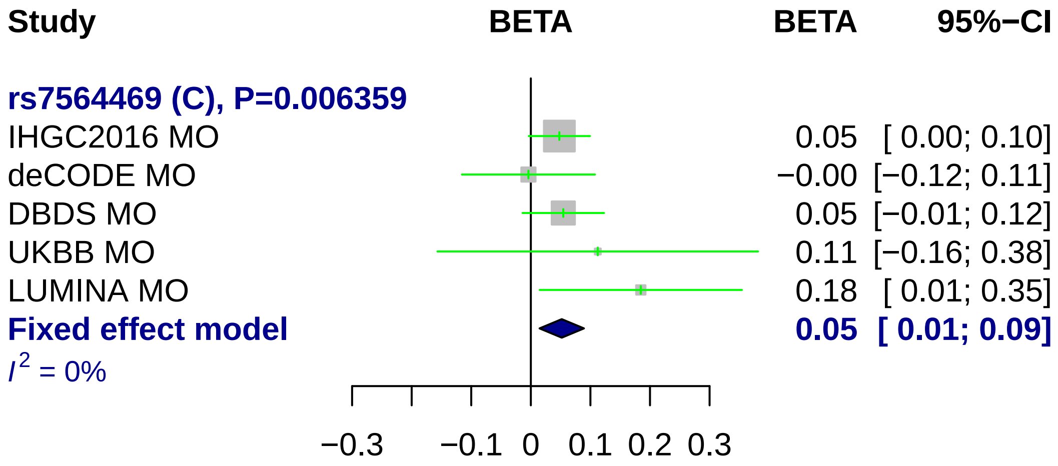

Study
BETA
BETA
95%−CI
rs7564469 (C), P=0.006359
IHGC2016 MO
0.05 [ 0.00; 0.10]
deCODE MO
−0.00 [−0.12; 0.11]
DBDS MO
0.05 [−0.01; 0.12]
UKBB MO
0.11 [−0.16; 0.38]
LUMINA MO
0.18 [ 0.01; 0.35]
Fixed effect model
0.05 [ 0.01; 0.09]
I2 = 0%
−0.3
−0.1
0
0.1
0.2
0.3

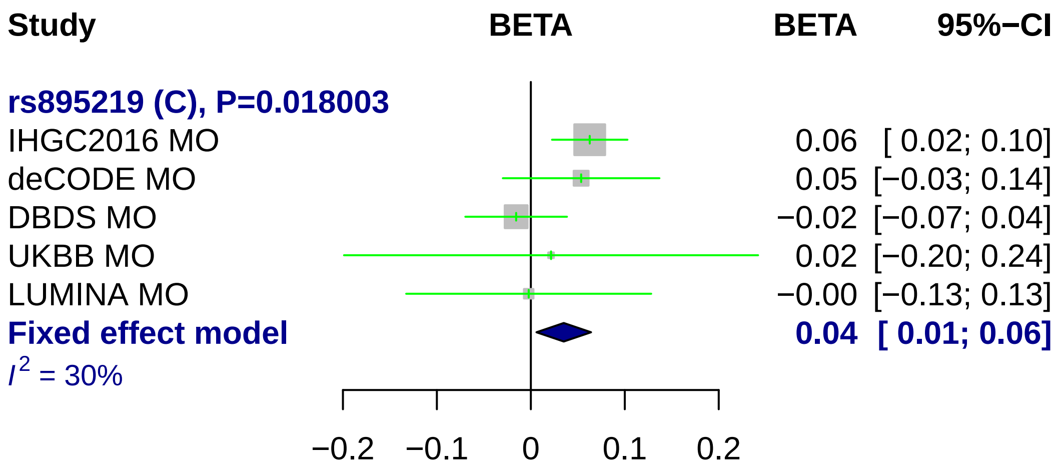
Study
BETA
BETA
95%−CI
rs895219 (C), P=0.018003
IHGC2016 MO
0.06
[ 0.02; 0.10]
deCODE MO
0.05
[−0.03; 0.14]
DBDS MO
−0.02
[−0.07; 0.04]
UKBB MO
0.02
[−0.20; 0.24]
LUMINA MO
−0.00
[−0.13; 0.13]
Fixed effect model
0.04
[ 0.01; 0.06]
I2 = 30%
−0.2
−0.1
0
0.1
0.2

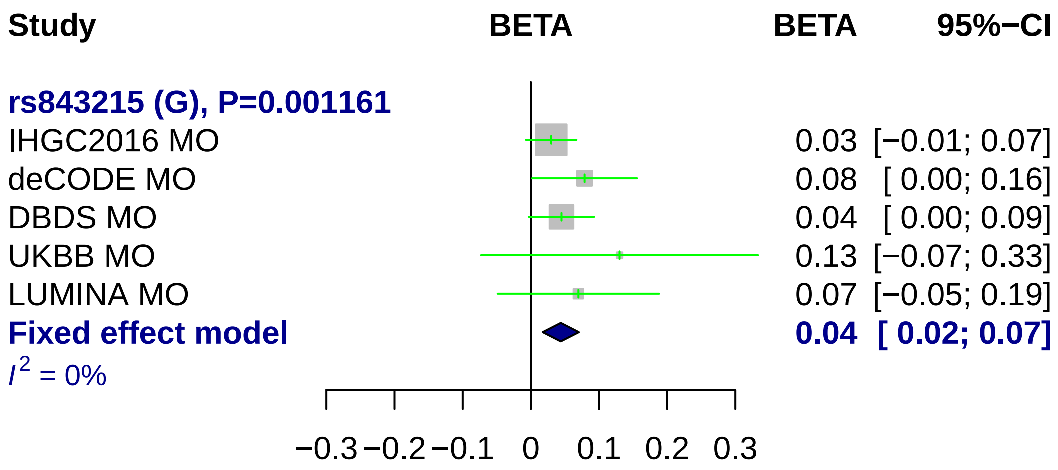
Study
BETA
BETA
95%−CI
rs843215 (G), P=0.001161
IHGC2016 MO 0.03 [−0.01; 0.07]
deCODE MO 0.08 [ 0.00; 0.16]
DBDS MO 0.04 [ 0.00; 0.09]
UKBB MO 0.13 [−0.07; 0.33]
LUMINA MO 0.07 [−0.05; 0.19]
Fixed effect model 0.04 [ 0.02; 0.07]
I² = 0%
−0.3 −0.2 −0.1 0 0.1 0.2 0.3

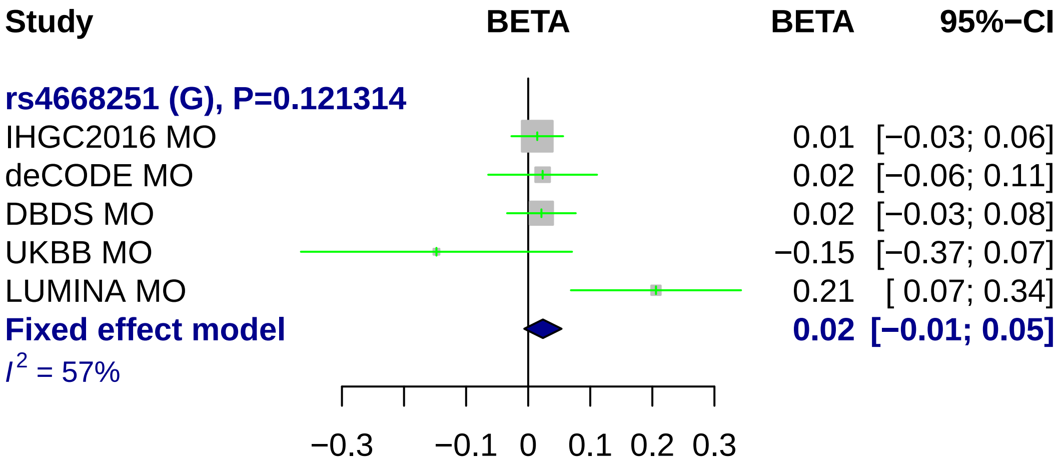
Study
BETA
BETA
95%−CI
rs4668251 (G), P=0.121314
IHGC2016 MO
0.01 [−0.03; 0.06]
deCODE MO
0.02 [−0.06; 0.11]
DBDS MO
0.02 [−0.03; 0.08]
UKBB MO
−0.15 [−0.37; 0.07]
LUMINA MO
0.21 [ 0.07; 0.34]
Fixed effect model
0.02 [−0.01; 0.05]
I2 = 57%
−0.3
−0.1
0
0.1
0.2
0.3

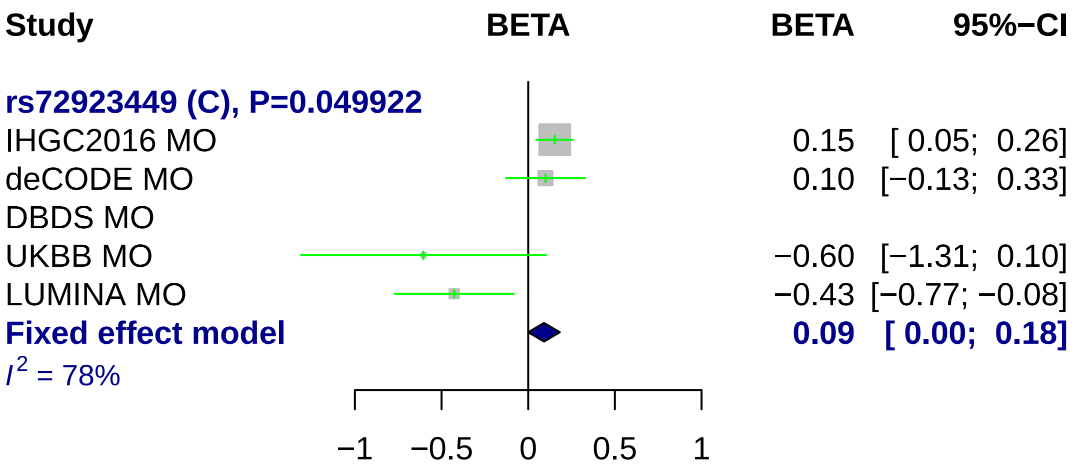
Study
BETA
BETA
95%−CI
rs72923449 (C), P=0.049922
IHGC2016 MO 0.15 [ 0.05; 0.26]
deCODE MO 0.10 [−0.13; 0.33]
DBDS MO
UKBB MO −0.60 [−1.31; 0.10]
LUMINA MO −0.43 [−0.77; −0.08]
Fixed effect model 0.09 [ 0.00; 0.18]
I2 = 78%
−1
−0.5
0
0.5
1

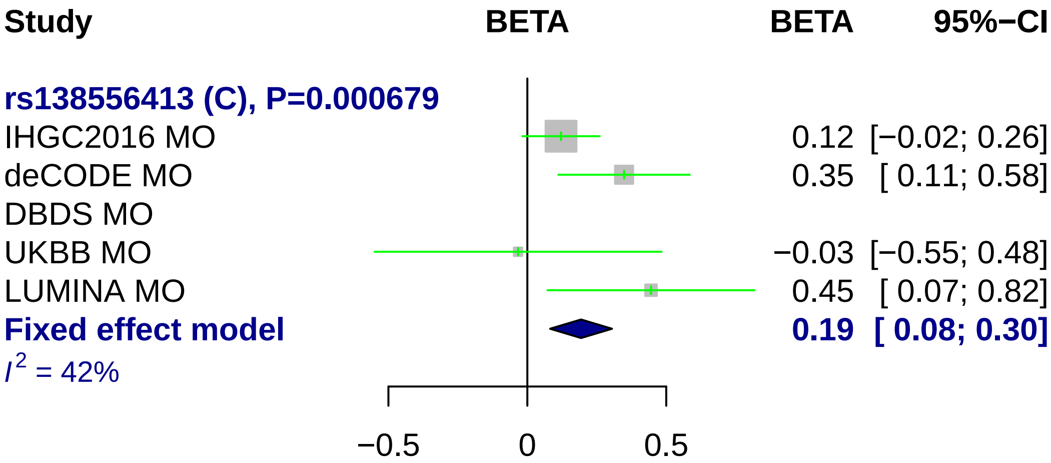

Study
BETA
BETA
95%−CI
rs138556413 (C), P=0.000679
IHGC2016 MO
0.12 [−0.02; 0.26]
deCODE MO
0.35 [ 0.11; 0.58]
DBDS MO
UKBB MO
−0.03 [−0.55; 0.48]
LUMINA MO
0.45 [ 0.07; 0.82]
Fixed effect model
0.19 [ 0.08; 0.30]
I2 = 42%
−0.5
0
0.5

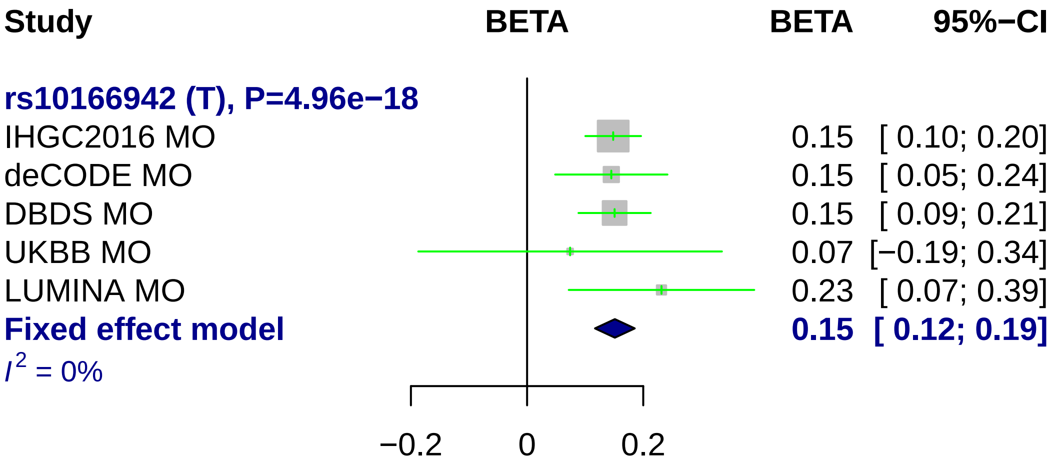
Study
BETA
BETA
95%−CI
rs10166942 (T), P=4.96e−18
IHGC2016 MO
0.15 [ 0.10; 0.20]
deCODE MO
0.15 [ 0.05; 0.24]
DBDS MO
0.15 [ 0.09; 0.21]
UKBB MO
0.07 [−0.19; 0.34]
LUMINA MO
0.23 [ 0.07; 0.39]
Fixed effect model
0.15 [ 0.12; 0.19]
I^2 = 0%
−0.2
0
0.2

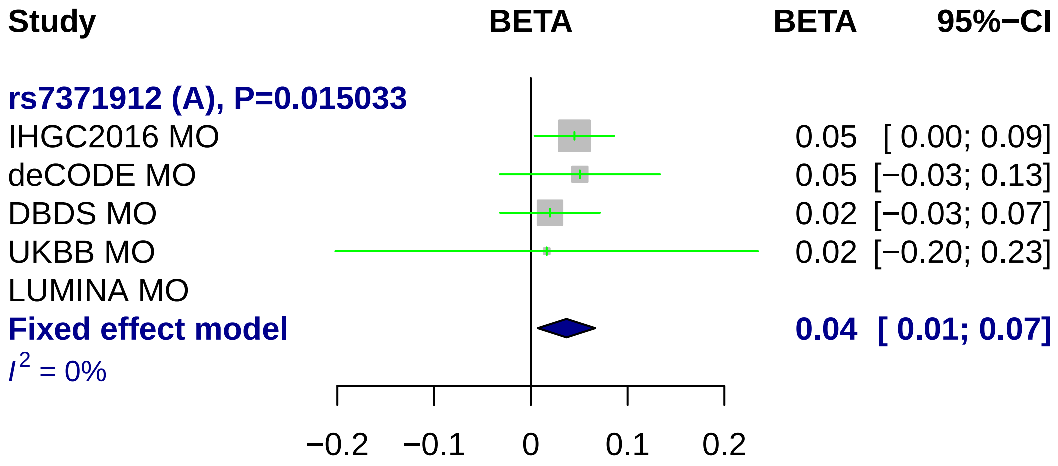

Study
BETA
BETA
95%−CI
rs7371912 (A), P=0.015033
IHGC2016 MO
0.05 [ 0.00; 0.09]
deCODE MO
0.05 [−0.03; 0.13]
DBDS MO
0.02 [−0.03; 0.07]
UKBB MO
0.02 [−0.20; 0.23]
LUMINA MO
Fixed effect model
0.04 [ 0.01; 0.07]
I2 = 0%
−0.2
−0.1
0
0.1
0.2

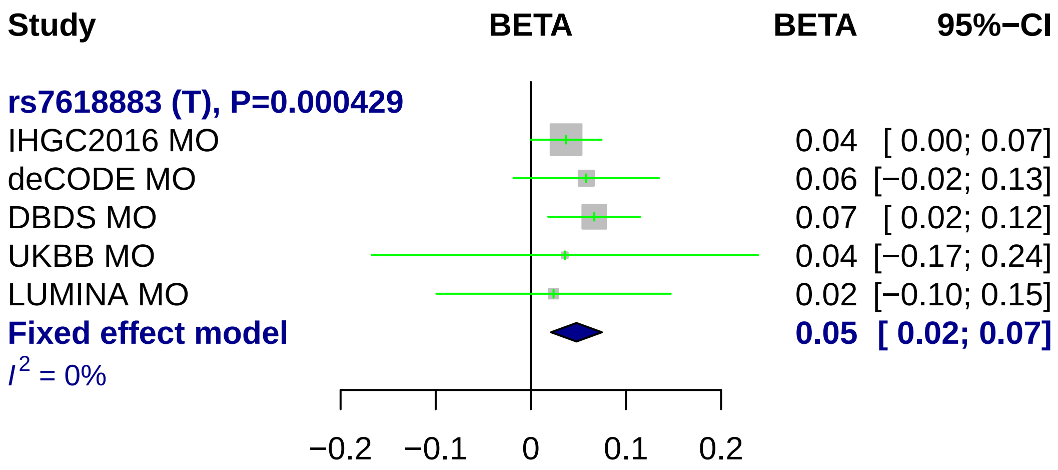
Study
BETA
BETA
95%−CI
rs7618883 (T), P=0.000429
IHGC2016 MO
0.04 [ 0.00; 0.07]
deCODE MO
0.06 [−0.02; 0.13]
DBDS MO
0.07 [ 0.02; 0.12]
UKBB MO
0.04 [−0.17; 0.24]
LUMINA MO
0.02 [−0.10; 0.15]
Fixed effect model
0.05 [ 0.02; 0.07]
I^2 = 0%
−0.2
−0.1
0
0.1
0.2

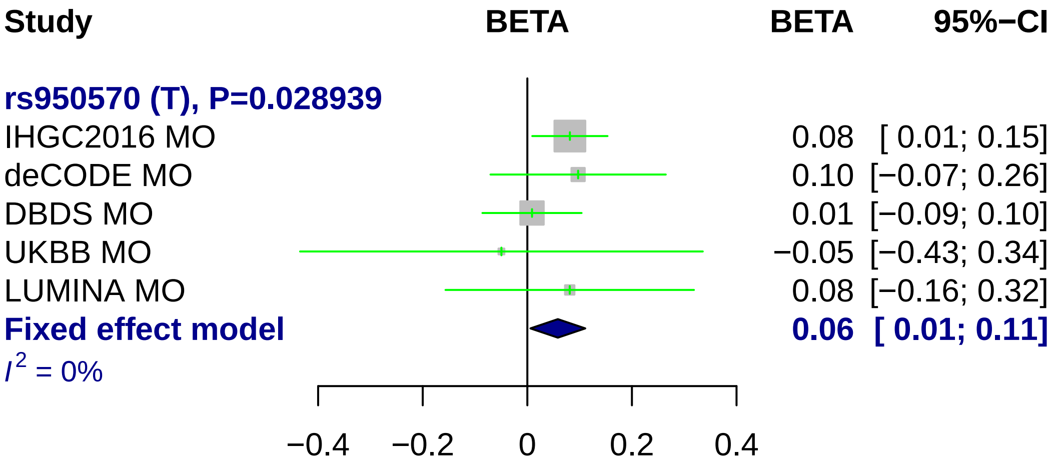
Study
BETA
BETA
95%−CI
rs950570 (T), P=0.028939
IHGC2016 MO
0.08 [ 0.01; 0.15]
deCODE MO
0.10 [−0.07; 0.26]
DBDS MO
0.01 [−0.09; 0.10]
UKBB MO
−0.05 [−0.43; 0.34]
LUMINA MO
0.08 [−0.16; 0.32]
Fixed effect model
0.06 [ 0.01; 0.11]
I2 = 0%
−0.4
−0.2
0
0.2
0.4

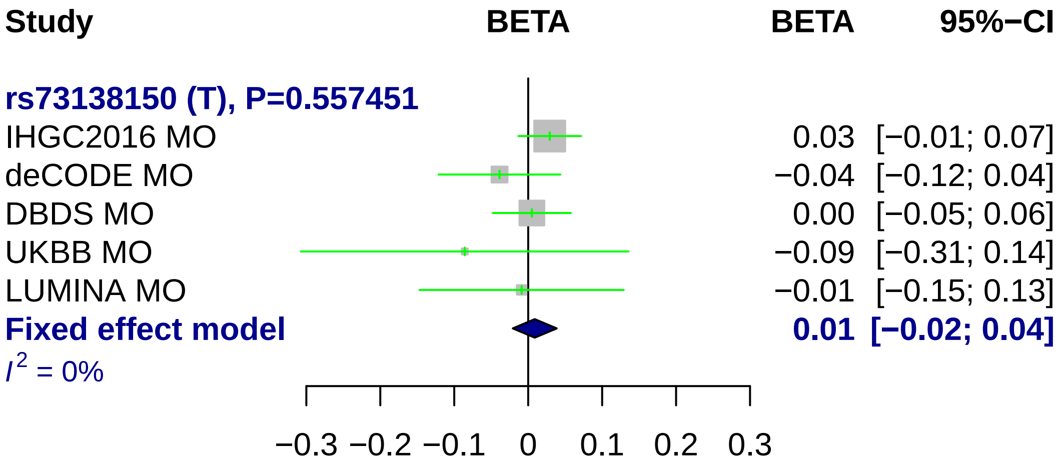

Study
BETA
BETA
95%−CI
rs73138150 (T), P=0.557451
IHGC2016 MO
0.03 [−0.01; 0.07]
deCODE MO
−0.04 [−0.12; 0.04]
DBDS MO
0.00 [−0.05; 0.06]
UKBB MO
−0.09 [−0.31; 0.14]
LUMINA MO
−0.01 [−0.15; 0.13]
Fixed effect model
0.01 [−0.02; 0.04]
$I^2$ = 0%
−0.3 −0.2 −0.1 0 0.1 0.2 0.3

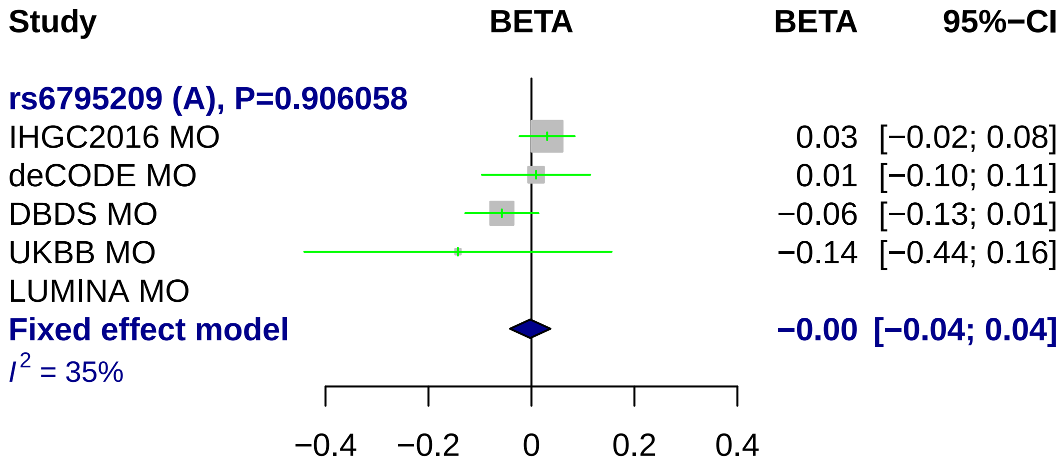
Study
BETA
BETA
95%−CI
rs6795209 (A), P=0.906058
IHGC2016 MO 0.03 [−0.02; 0.08]
deCODE MO 0.01 [−0.10; 0.11]
DBDS MO −0.06 [−0.13; 0.01]
UKBB MO −0.14 [−0.44; 0.16]
LUMINA MO
Fixed effect model −0.00 [−0.04; 0.04]
I2 = 35%
−0.4
−0.2
0
0.2
0.4

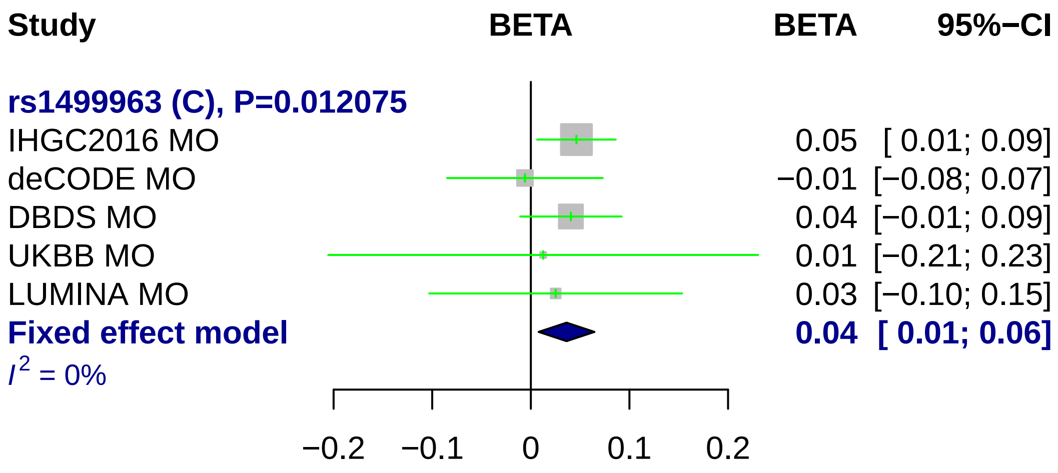
Study
BETA
BETA
95%−CI
rs1499963 (C), P=0.012075
IHGC2016 MO
0.05 [ 0.01; 0.09]
deCODE MO
−0.01 [−0.08; 0.07]
DBDS MO
0.04 [−0.01; 0.09]
UKBB MO
0.01 [−0.21; 0.23]
LUMINA MO
0.03 [−0.10; 0.15]
Fixed effect model
0.04 [ 0.01; 0.06]
I2 = 0%
−0.2
−0.1
0
0.1
0.2

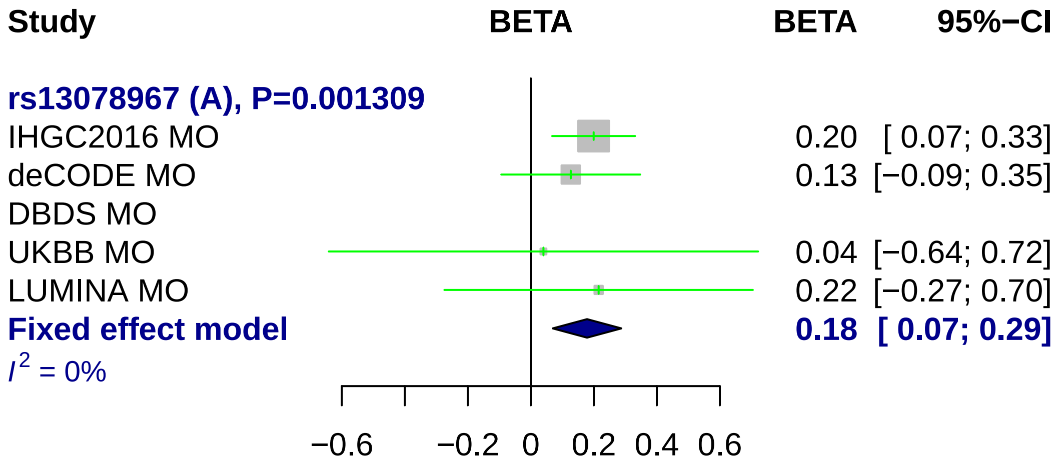
Study
BETA
BETA
95%−CI
rs13078967 (A), P=0.001309
IHGC2016 MO
0.20 [ 0.07; 0.33]
deCODE MO
0.13 [−0.09; 0.35]
DBDS MO
UKBB MO
0.04 [−0.64; 0.72]
LUMINA MO
0.22 [−0.27; 0.70]
Fixed effect model
0.18 [ 0.07; 0.29]
I² = 0%
−0.6
−0.2
0
0.2
0.4
0.6

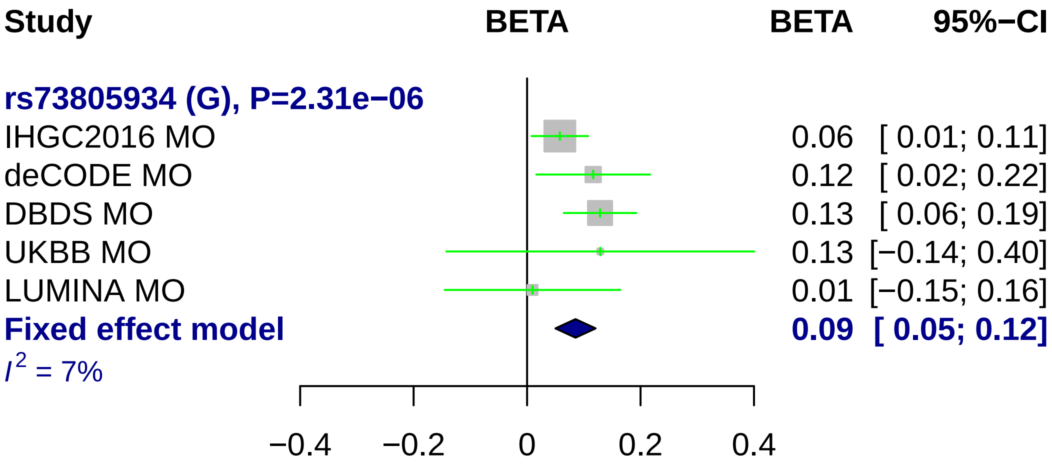

Study
BETA
BETA
95%−CI
rs73805934 (G), P=2.31e−06
IHGC2016 MO
0.06 [ 0.01; 0.11]
deCODE MO
0.12 [ 0.02; 0.22]
DBDS MO
0.13 [ 0.06; 0.19]
UKBB MO
0.13 [−0.14; 0.40]
LUMINA MO
0.01 [−0.15; 0.16]
Fixed effect model
0.09 [ 0.05; 0.12]
I2 = 7%
−0.4
−0.2
0
0.2
0.4

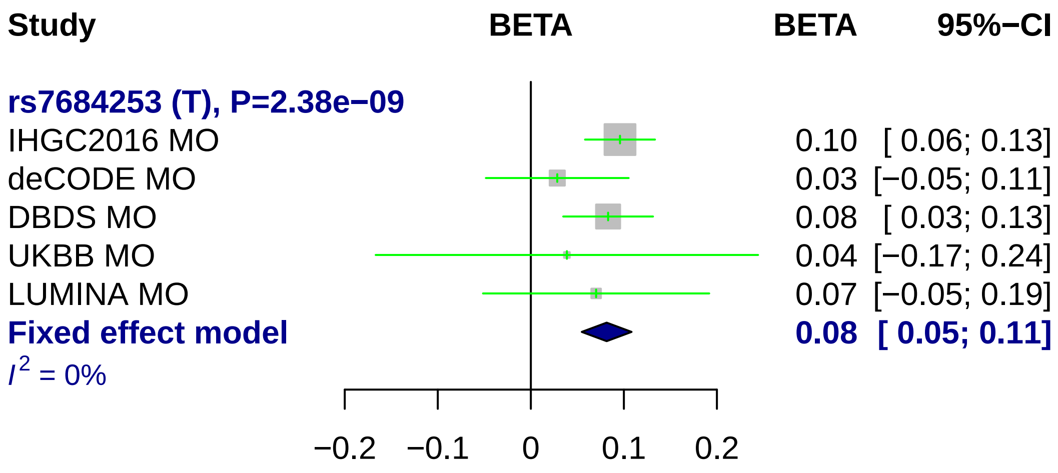
Study
BETA
BETA
95%−CI
rs7684253 (T), P=2.38e−09
IHGC2016 MO
0.10 [ 0.06; 0.13]
deCODE MO
0.03 [−0.05; 0.11]
DBDS MO
0.08 [ 0.03; 0.13]
UKBB MO
0.04 [−0.17; 0.24]
LUMINA MO
0.07 [−0.05; 0.19]
Fixed effect model
0.08 [ 0.05; 0.11]
I2 = 0%
−0.2
−0.1
0
0.1
0.2

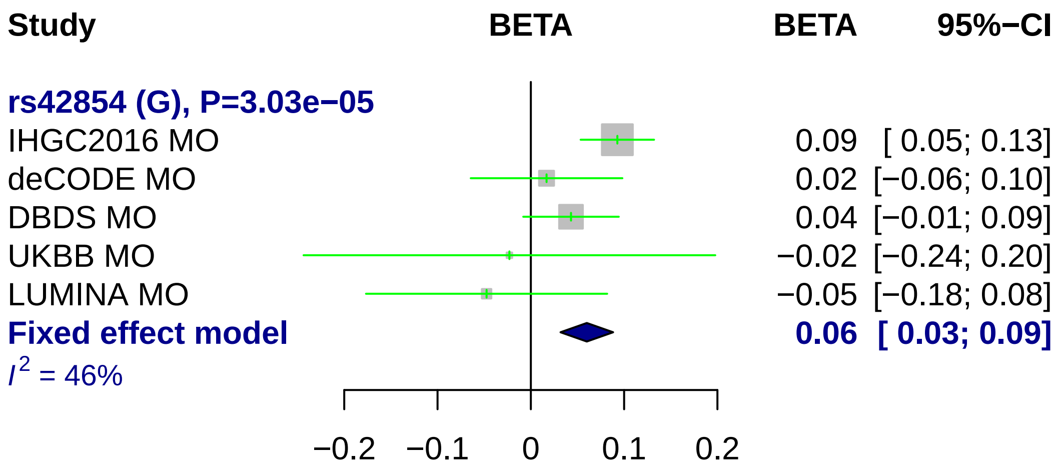

Study
BETA
BETA
95%−CI
rs42854 (G), P=3.03e−05
IHGC2016 MO
0.09 [ 0.05; 0.13]
deCODE MO
0.02 [−0.06; 0.10]
DBDS MO
0.04 [−0.01; 0.09]
UKBB MO
−0.02 [−0.24; 0.20]
LUMINA MO
−0.05 [−0.18; 0.08]
Fixed effect model
0.06 [ 0.03; 0.09]
I2 = 46%
−0.2
−0.1
0
0.1
0.2

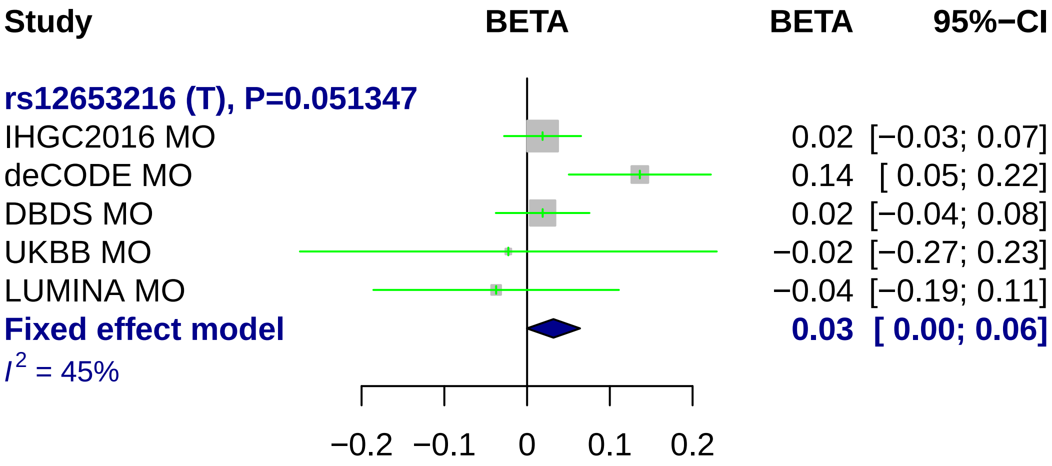
Study
BETA
BETA
95%−CI
rs12653216 (T), P=0.051347
IHGC2016 MO
0.02 [−0.03; 0.07]
deCODE MO
0.14 [ 0.05; 0.22]
DBDS MO
0.02 [−0.04; 0.08]
UKBB MO
−0.02 [−0.27; 0.23]
LUMINA MO
−0.04 [−0.19; 0.11]
Fixed effect model
0.03 [ 0.00; 0.06]
I^2 = 45%
−0.2
−0.1
0
0.1
0.2

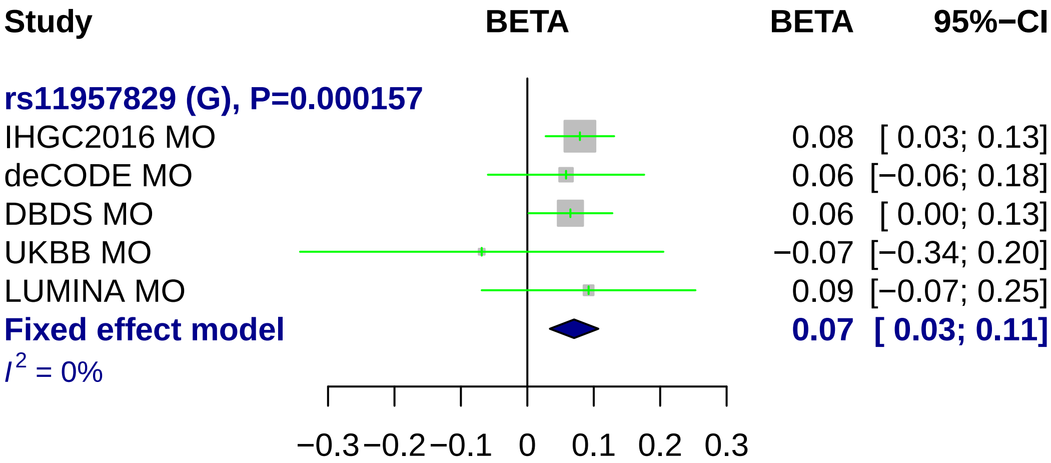

Study
BETA
BETA
95%−CI
rs11957829 (G), P=0.000157
IHGC2016 MO
0.08 [ 0.03; 0.13]
deCODE MO
0.06 [−0.06; 0.18]
DBDS MO
0.06 [ 0.00; 0.13]
UKBB MO
−0.07 [−0.34; 0.20]
LUMINA MO
0.09 [−0.07; 0.25]
Fixed effect model
0.07 [ 0.03; 0.11]
I2 = 0%
−0.3
−0.2
−0.1
0
0.1
0.2
0.3

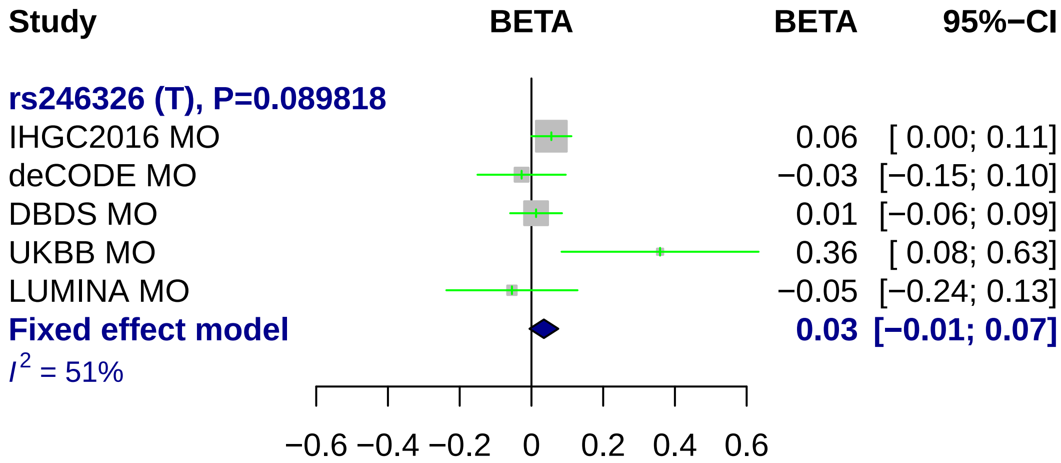

Study
BETA
BETA
95%−CI
rs246326 (T), P=0.089818
IHGC2016 MO 0.06 [ 0.00; 0.11]
deCODE MO −0.03 [−0.15; 0.10]
DBDS MO 0.01 [−0.06; 0.09]
UKBB MO 0.36 [ 0.08; 0.63]
LUMINA MO −0.05 [−0.24; 0.13]
Fixed effect model 0.03 [−0.01; 0.07]
I^2 = 51%
−0.6 −0.4 −0.2 0 0.2 0.4 0.6

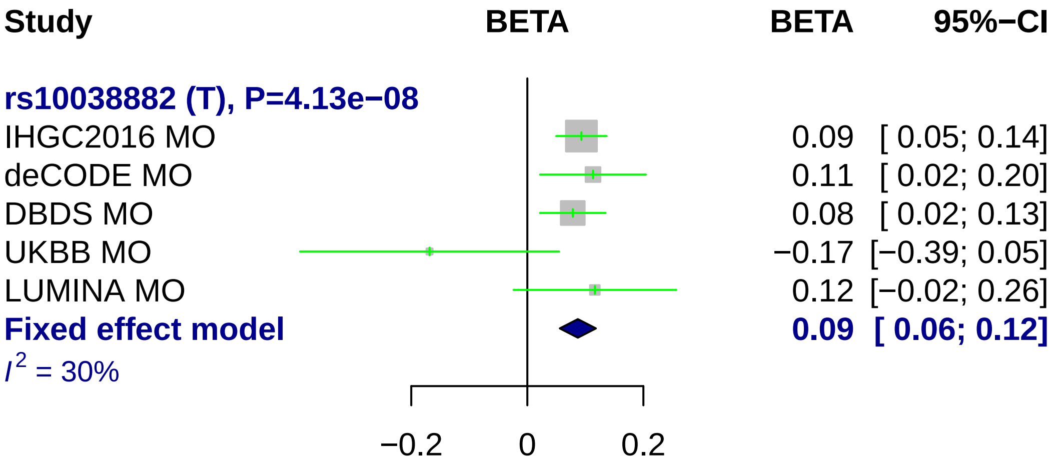
Study
BETA
BETA
95%−CI
rs10038882 (T), P=4.13e−08
IHGC2016 MO
0.09 [ 0.05; 0.14]
deCODE MO
0.11 [ 0.02; 0.20]
DBDS MO
0.08 [ 0.02; 0.13]
UKBB MO
−0.17 [−0.39; 0.05]
LUMINA MO
0.12 [−0.02; 0.26]
Fixed effect model
0.09 [ 0.06; 0.12]
I2 = 30%
−0.2
0
0.2

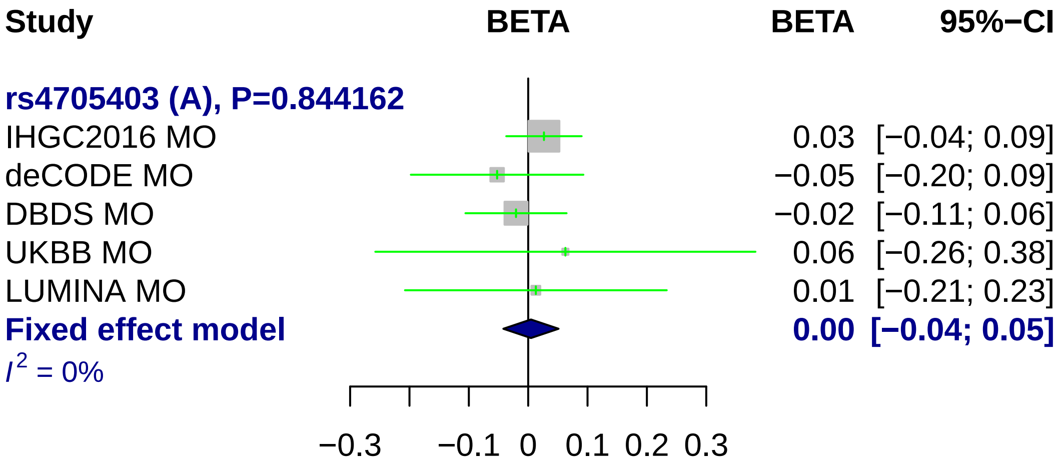
Study
BETA
BETA
95%−CI
rs4705403 (A), P=0.844162
IHGC2016 MO
0.03 [−0.04; 0.09]
deCODE MO
−0.05 [−0.20; 0.09]
DBDS MO
−0.02 [−0.11; 0.06]
UKBB MO
0.06 [−0.26; 0.38]
LUMINA MO
0.01 [−0.21; 0.23]
Fixed effect model
0.00 [−0.04; 0.05]
I2 = 0%
−0.3
−0.1
0
0.1
0.2
0.3

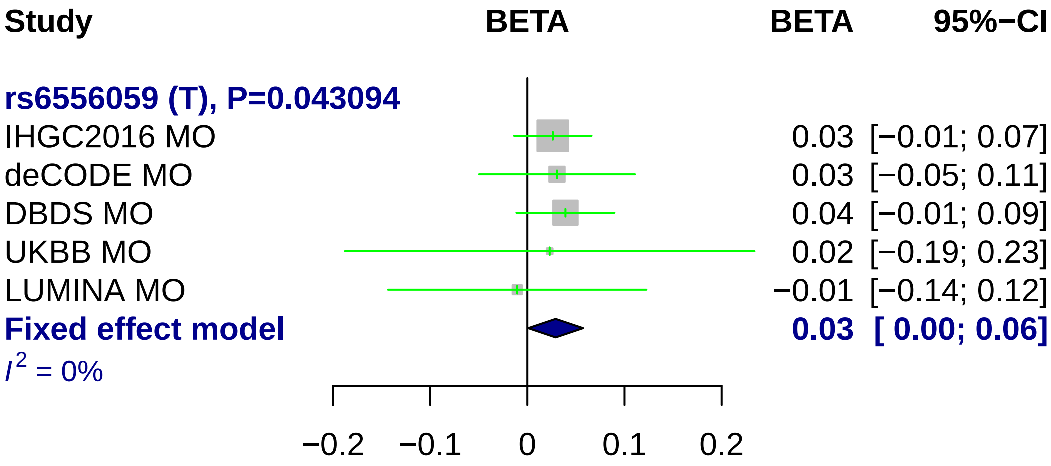
Study
BETA
BETA
95%−CI
rs6556059 (T), P=0.043094
IHGC2016 MO
0.03 [−0.01; 0.07]
deCODE MO
0.03 [−0.05; 0.11]
DBDS MO
0.04 [−0.01; 0.09]
UKBB MO
0.02 [−0.19; 0.23]
LUMINA MO
−0.01 [−0.14; 0.12]
Fixed effect model
0.03 [ 0.00; 0.06]
I2 = 0%
−0.2
−0.1
0
0.1
0.2

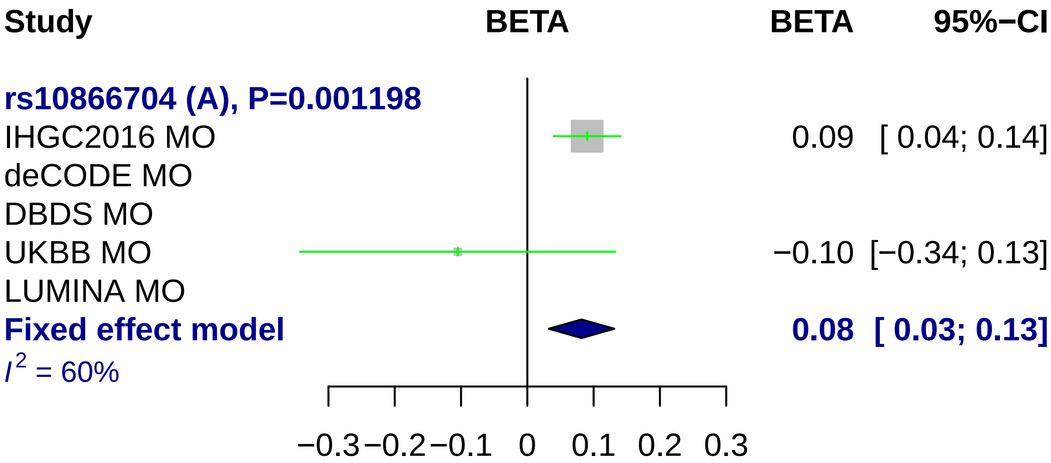
Study
BETA
BETA
95%−CI
rs10866704 (A), P=0.001198
IHGC2016 MO
0.09 [ 0.04; 0.14]
deCODE MO
DBDS MO
UKBB MO
−0.10 [−0.34; 0.13]
LUMINA MO
Fixed effect model
0.08 [ 0.03; 0.13]
$I^2$ = 60%
−0.3 −0.2 −0.1 0 0.1 0.2 0.3

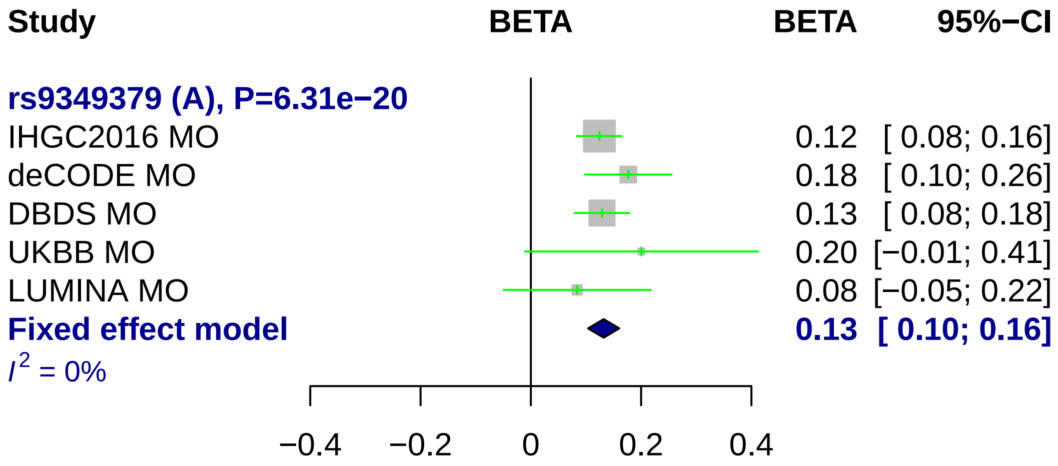
Study
BETA
BETA
95%−CI
rs9349379 (A), P=6.31e−20
IHGC2016 MO
0.12 [ 0.08; 0.16]
deCODE MO
0.18 [ 0.10; 0.26]
DBDS MO
0.13 [ 0.08; 0.18]
UKBB MO
0.20 [−0.01; 0.41]
LUMINA MO
0.08 [−0.05; 0.22]
Fixed effect model
0.13 [ 0.10; 0.16]
I2 = 0%
−0.4
−0.2
0
0.2
0.4

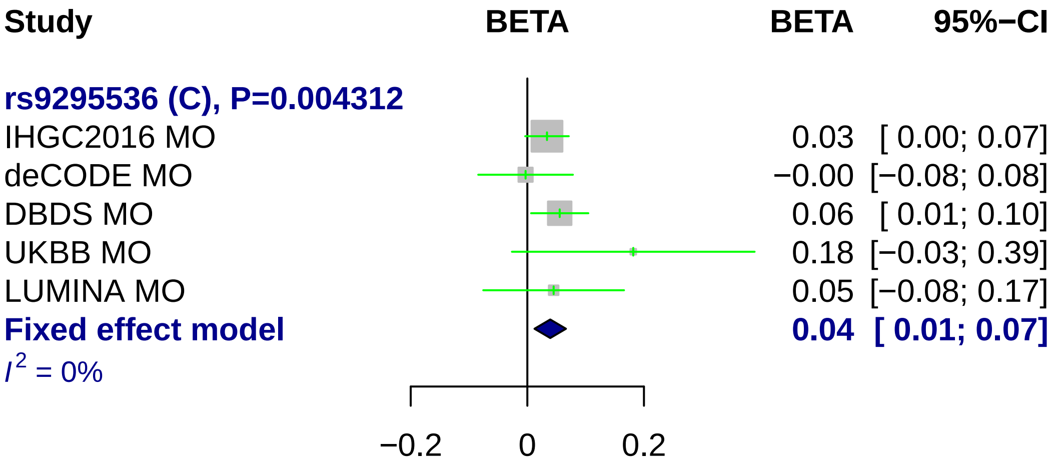
Study
BETA
BETA
95%−CI
rs9295536 (C), P=0.004312
IHGC2016 MO
0.03 [ 0.00; 0.07]
deCODE MO
−0.00 [−0.08; 0.08]
DBDS MO
0.06 [ 0.01; 0.10]
UKBB MO
0.18 [−0.03; 0.39]
LUMINA MO
0.05 [−0.08; 0.17]
Fixed effect model
0.04 [ 0.01; 0.07]
$I^2$ = 0%
−0.2
0
0.2

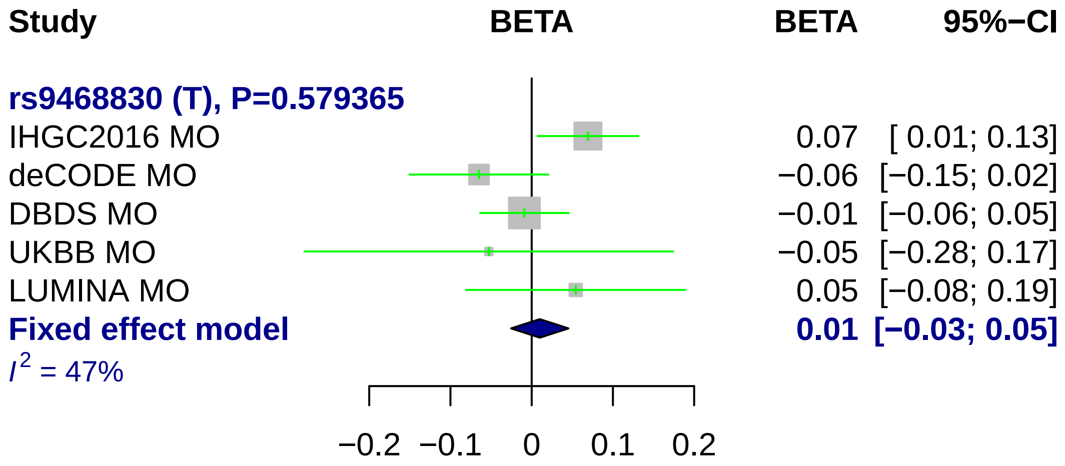

Study
BETA
BETA
95%−CI
rs9468830 (T), P=0.579365
IHGC2016 MO 0.07 [ 0.01; 0.13]
deCODE MO −0.06 [−0.15; 0.02]
DBDS MO −0.01 [−0.06; 0.05]
UKBB MO −0.05 [−0.28; 0.17]
LUMINA MO 0.05 [−0.08; 0.19]
Fixed effect model 0.01 [−0.03; 0.05]
$I^2$ = 47%
−0.2 −0.1 0 0.1 0.2

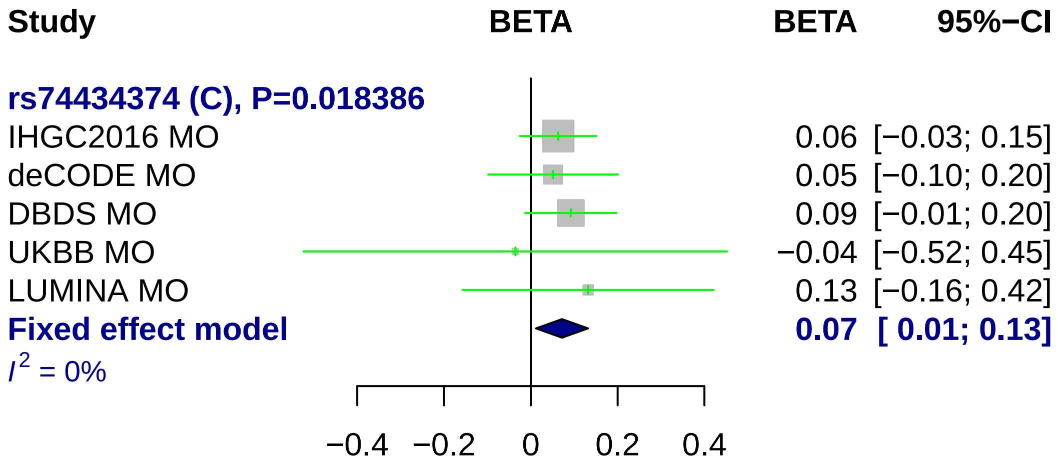
Study
BETA
BETA
95%−CI
rs74434374 (C), P=0.018386
IHGC2016 MO
0.06 [−0.03; 0.15]
deCODE MO
0.05 [−0.10; 0.20]
DBDS MO
0.09 [−0.01; 0.20]
UKBB MO
−0.04 [−0.52; 0.45]
LUMINA MO
0.13 [−0.16; 0.42]
Fixed effect model
0.07 [ 0.01; 0.13]
I2 = 0%
−0.4
−0.2
0
0.2
0.4

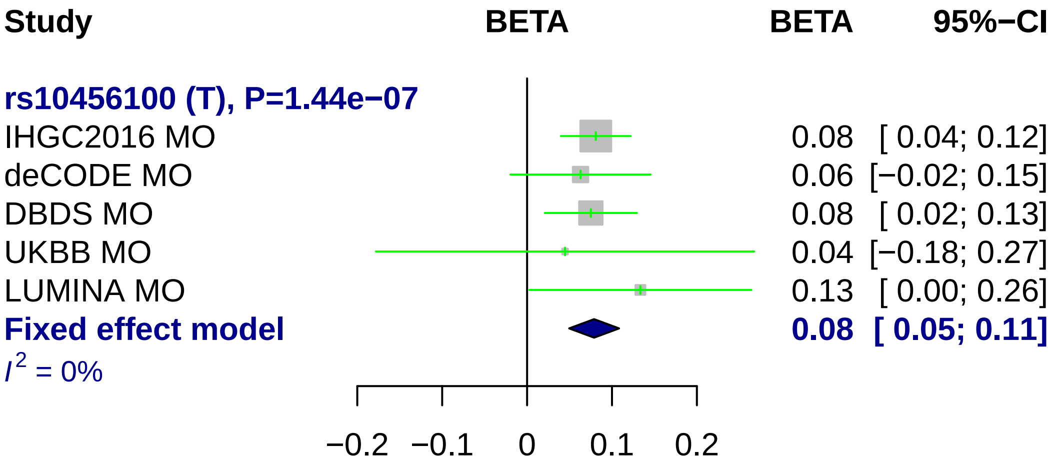
Study
BETA
BETA
95%−CI
rs10456100 (T), P=1.44e−07
IHGC2016 MO
0.08 [ 0.04; 0.12]
deCODE MO
0.06 [−0.02; 0.15]
DBDS MO
0.08 [ 0.02; 0.13]
UKBB MO
0.04 [−0.18; 0.27]
LUMINA MO
0.13 [ 0.00; 0.26]
Fixed effect model
0.08 [ 0.05; 0.11]
I^2 = 0%
−0.2
−0.1
0
0.1
0.2

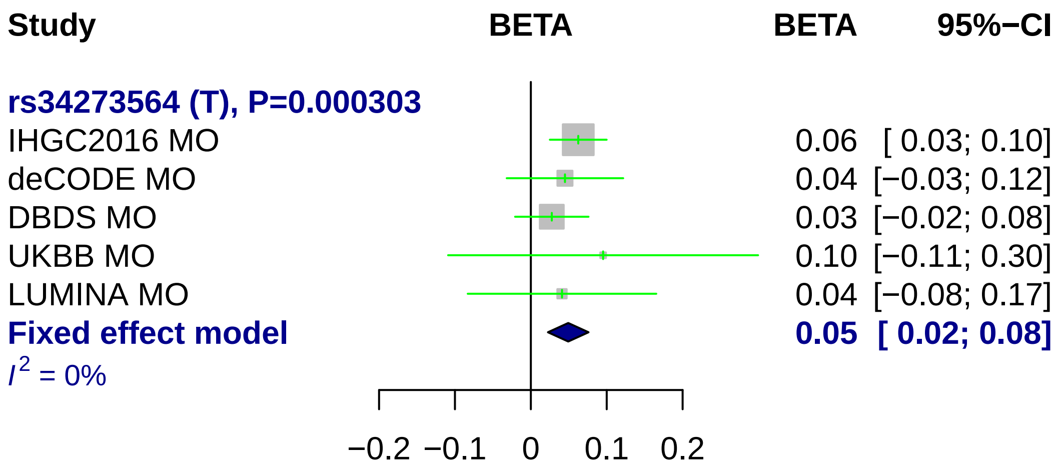
Study
BETA
BETA
95%−CI
rs34273564 (T), P=0.000303
IHGC2016 MO
0.06 [ 0.03; 0.10]
deCODE MO
0.04 [−0.03; 0.12]
DBDS MO
0.03 [−0.02; 0.08]
UKBB MO
0.10 [−0.11; 0.30]
LUMINA MO
0.04 [−0.08; 0.17]
Fixed effect model
0.05 [ 0.02; 0.08]
I² = 0%
−0.2
−0.1
0
0.1
0.2

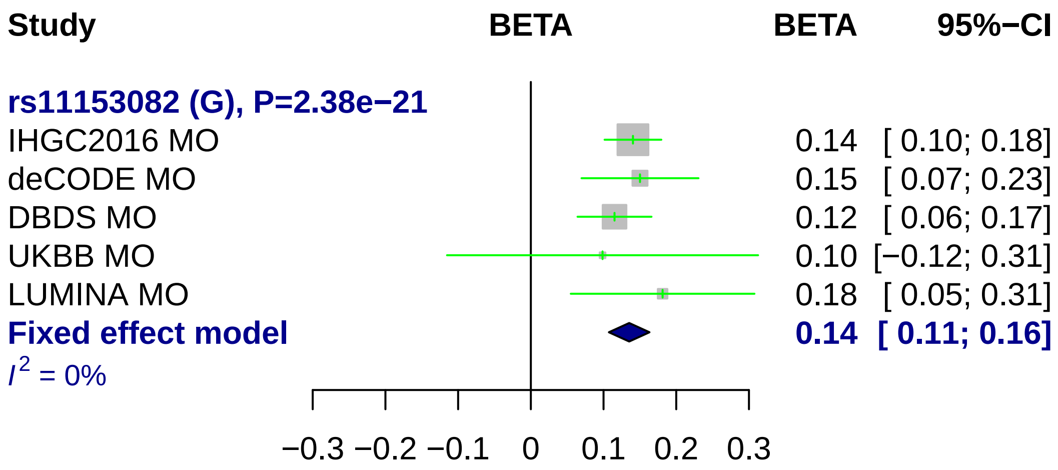

Study
BETA
BETA
95%−CI
rs11153082 (G), P=2.38e−21
IHGC2016 MO
0.14 [ 0.10; 0.18]
deCODE MO
0.15 [ 0.07; 0.23]
DBDS MO
0.12 [ 0.06; 0.17]
UKBB MO
0.10 [−0.12; 0.31]
LUMINA MO
0.18 [ 0.05; 0.31]
Fixed effect model
0.14 [ 0.11; 0.16]
I2 = 0%
−0.3 −0.2 −0.1 0 0.1 0.2 0.3

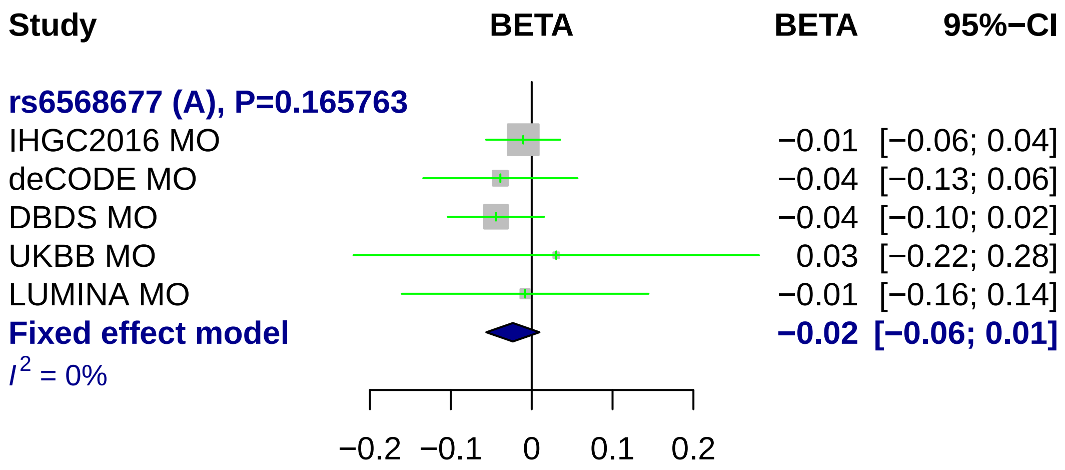
Study
BETA
BETA
95%−CI
rs6568677 (A), P=0.165763
IHGC2016 MO
−0.01 [−0.06; 0.04]
deCODE MO
−0.04 [−0.13; 0.06]
DBDS MO
−0.04 [−0.10; 0.02]
UKBB MO
0.03 [−0.22; 0.28]
LUMINA MO
−0.01 [−0.16; 0.14]
Fixed effect model
−0.02 [−0.06; 0.01]
I2 = 0%
−0.2
−0.1
0
0.1
0.2

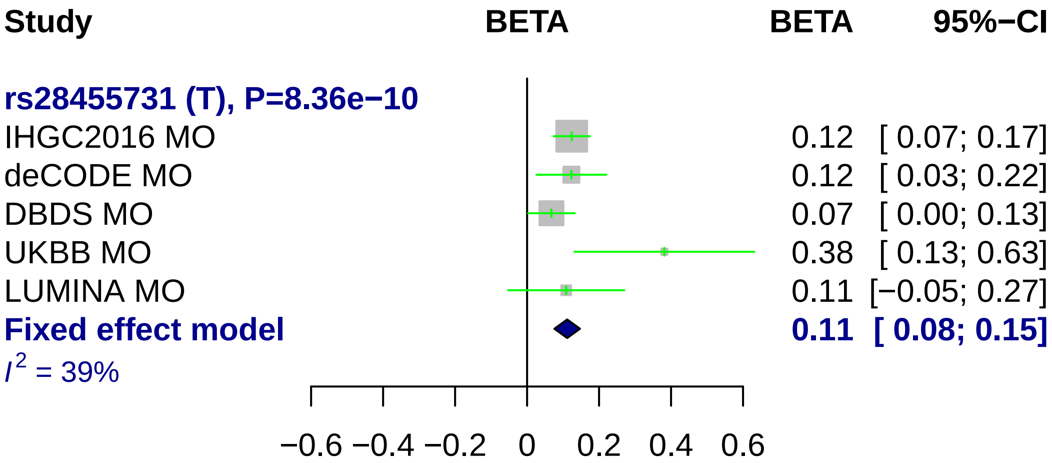

Study
BETA
BETA
95%−CI
rs28455731 (T), P=8.36e−10
IHGC2016 MO
0.12 [ 0.07; 0.17]
deCODE MO
0.12 [ 0.03; 0.22]
DBDS MO
0.07 [ 0.00; 0.13]
UKBB MO
0.38 [ 0.13; 0.63]
LUMINA MO
0.11 [−0.05; 0.27]
Fixed effect model
0.11 [ 0.08; 0.15]
I2 = 39%
−0.6
−0.4
−0.2
0
0.2
0.4
0.6

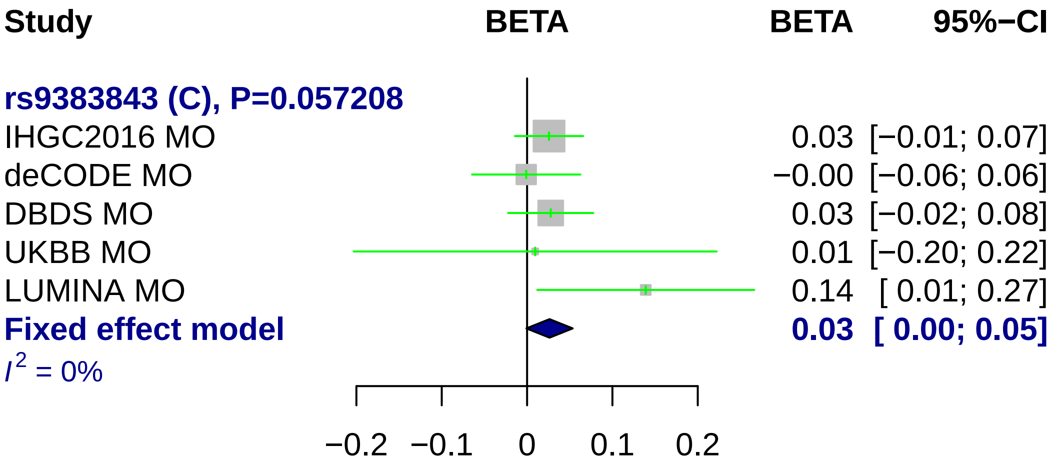
Study
BETA
BETA
95%−CI
rs9383843 (C), P=0.057208
IHGC2016 MO
0.03 [−0.01; 0.07]
deCODE MO
−0.00 [−0.06; 0.06]
DBDS MO
0.03 [−0.02; 0.08]
UKBB MO
0.01 [−0.20; 0.22]
LUMINA MO
0.14 [ 0.01; 0.27]
Fixed effect model
0.03 [ 0.00; 0.05]
I² = 0%
−0.2
−0.1
0
0.1
0.2

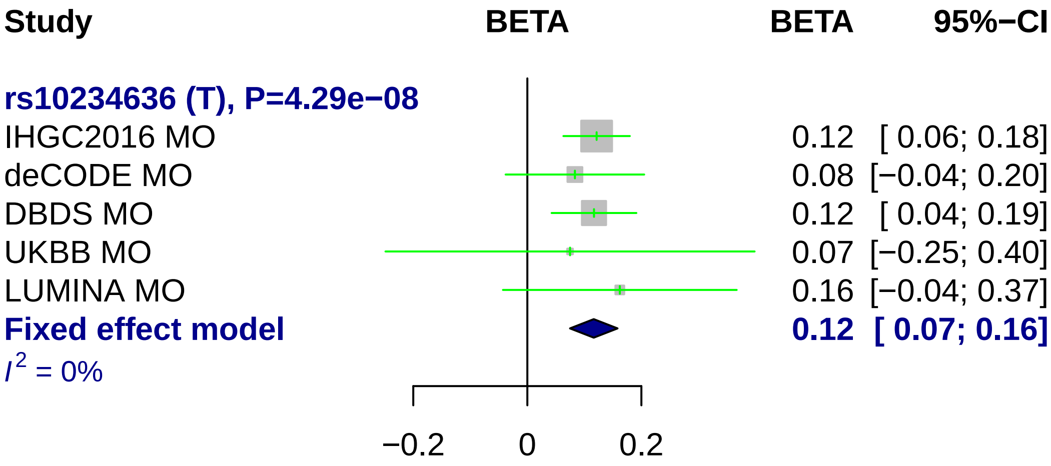
Study
BETA
BETA
95%−CI
rs10234636 (T), P=4.29e−08
IHGC2016 MO
0.12 [ 0.06; 0.18]
deCODE MO
0.08 [−0.04; 0.20]
DBDS MO
0.12 [ 0.04; 0.19]
UKBB MO
0.07 [−0.25; 0.40]
LUMINA MO
0.16 [−0.04; 0.37]
Fixed effect model
0.12 [ 0.07; 0.16]
I2 = 0%
−0.2
0
0.2

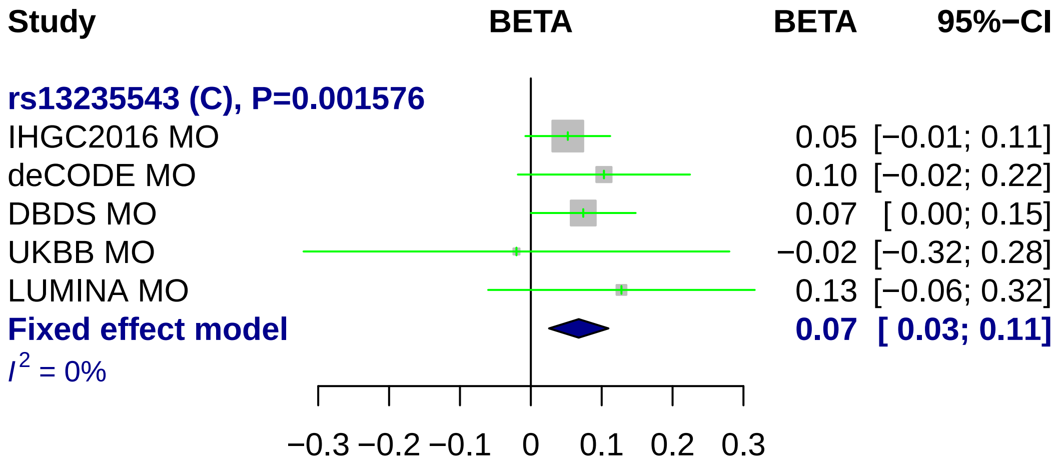
Study
BETA
BETA
95%−CI
rs13235543 (C), P=0.001576
IHGC2016 MO
0.05 [−0.01; 0.11]
deCODE MO
0.10 [−0.02; 0.22]
DBDS MO
0.07 [ 0.00; 0.15]
UKBB MO
−0.02 [−0.32; 0.28]
LUMINA MO
0.13 [−0.06; 0.32]
Fixed effect model
0.07 [ 0.03; 0.11]
I^2 = 0%
−0.3 −0.2 −0.1 0 0.1 0.2 0.3

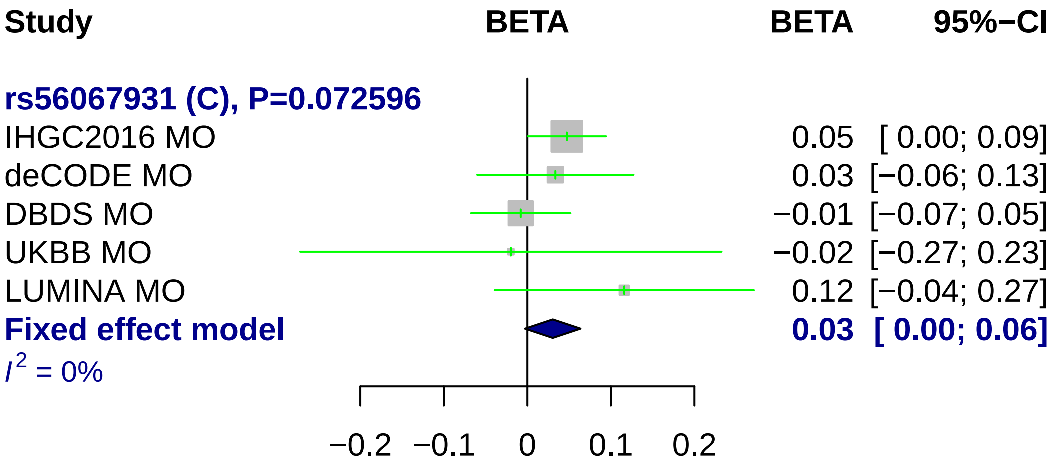
Study
BETA
BETA
95%−CI
rs56067931 (C), P=0.072596
IHGC2016 MO
0.05 [ 0.00; 0.09]
deCODE MO
0.03 [−0.06; 0.13]
DBDS MO
−0.01 [−0.07; 0.05]
UKBB MO
−0.02 [−0.27; 0.23]
LUMINA MO
0.12 [−0.04; 0.27]
Fixed effect model
0.03 [ 0.00; 0.06]
I2 = 0%
−0.2
−0.1
0
0.1
0.2

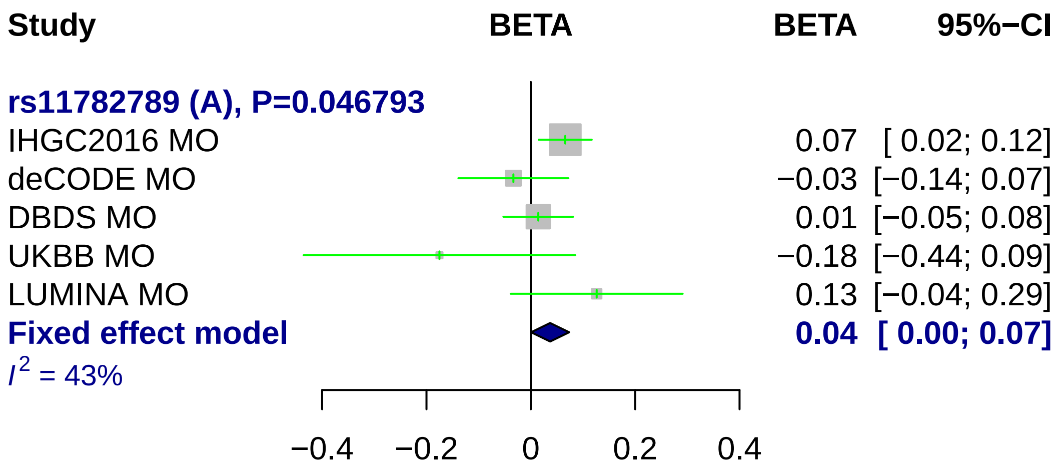
Study
BETA
BETA
95%−CI
rs11782789 (A), P=0.046793
IHGC2016 MO
0.07 [ 0.02; 0.12]
deCODE MO
−0.03 [−0.14; 0.07]
DBDS MO
0.01 [−0.05; 0.08]
UKBB MO
−0.18 [−0.44; 0.09]
LUMINA MO
0.13 [−0.04; 0.29]
Fixed effect model
0.04 [ 0.00; 0.07]
I2 = 43%
−0.4
−0.2
0
0.2
0.4

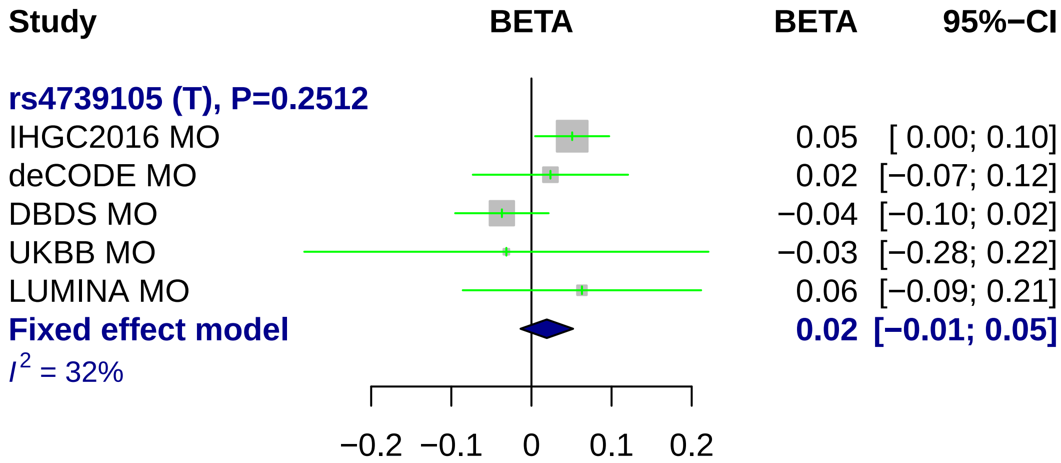
Study
BETA
BETA
95%−CI
rs4739105 (T), P=0.2512
IHGC2016 MO
0.05
[ 0.00; 0.10]
deCODE MO
0.02
[−0.07; 0.12]
DBDS MO
−0.04
[−0.10; 0.02]
UKBB MO
−0.03
[−0.28; 0.22]
LUMINA MO
0.06
[−0.09; 0.21]
Fixed effect model
0.02
[−0.01; 0.05]
I2 = 32%
−0.2
−0.1
0
0.1
0.2

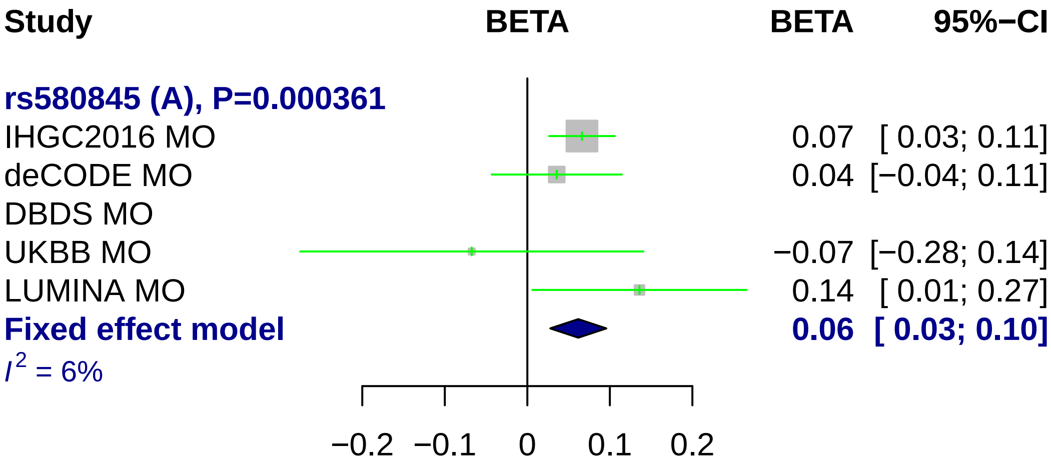
Study
BETA
BETA
95%−CI
rs580845 (A), P=0.000361
IHGC2016 MO 0.07 [ 0.03; 0.11]
deCODE MO 0.04 [−0.04; 0.11]
DBDS MO
UKBB MO −0.07 [−0.28; 0.14]
LUMINA MO 0.14 [ 0.01; 0.27]
Fixed effect model 0.06 [ 0.03; 0.10]
I2 = 6%
−0.2 −0.1 0 0.1 0.2

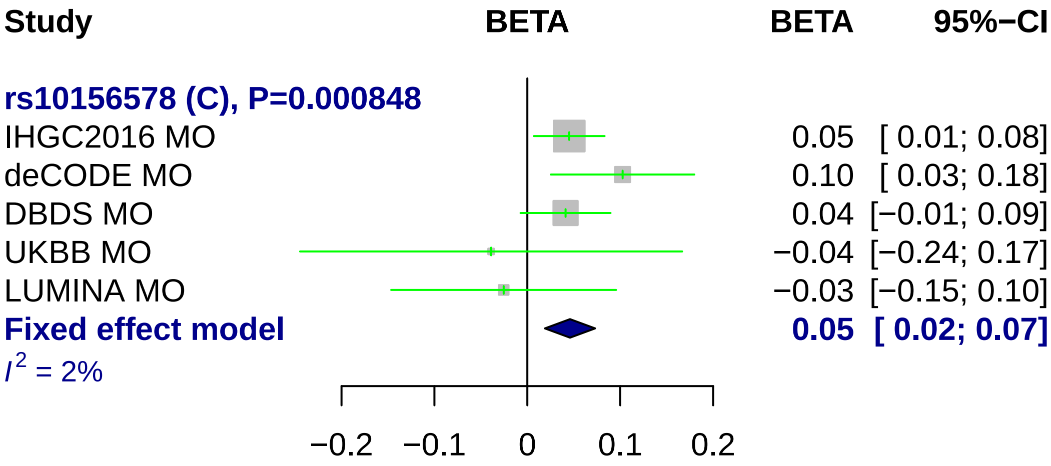

Study
BETA
BETA
95%−CI
rs10156578 (C), P=0.000848
IHGC2016 MO 0.05 [ 0.01; 0.08]
deCODE MO 0.10 [ 0.03; 0.18]
DBDS MO 0.04 [−0.01; 0.09]
UKBB MO −0.04 [−0.24; 0.17]
LUMINA MO −0.03 [−0.15; 0.10]
Fixed effect model 0.05 [ 0.02; 0.07]
I2 = 2%
−0.2
−0.1
0
0.1
0.2

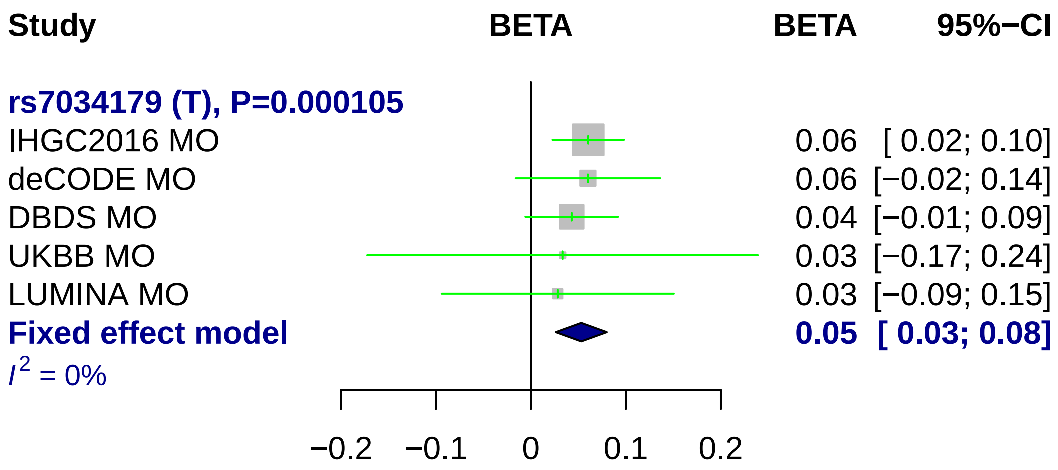
Study
BETA
BETA
95%−CI
rs7034179 (T), P=0.000105
IHGC2016 MO 0.06 [ 0.02; 0.10]
deCODE MO 0.06 [−0.02; 0.14]
DBDS MO 0.04 [−0.01; 0.09]
UKBB MO 0.03 [−0.17; 0.24]
LUMINA MO 0.03 [−0.09; 0.15]
Fixed effect model 0.05 [ 0.03; 0.08]
$I^2$ = 0%
−0.2
−0.1
0
0.1
0.2

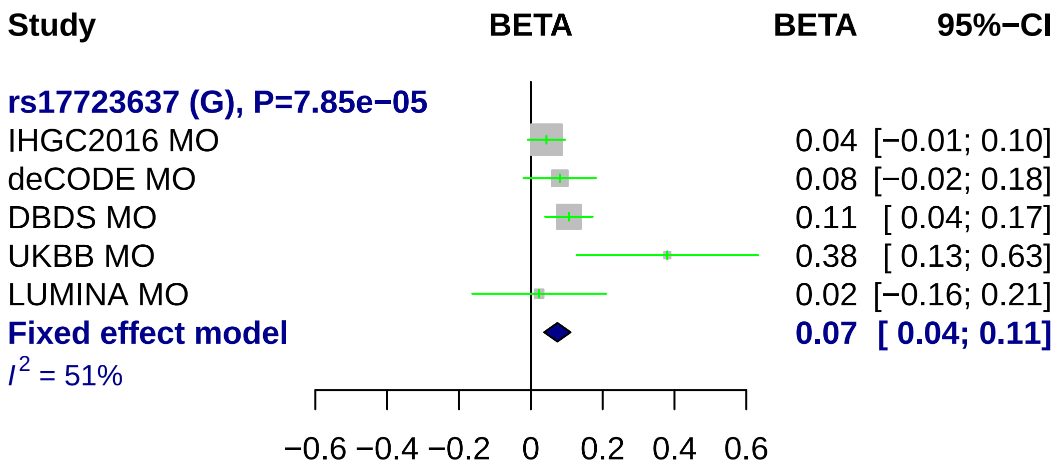
Study
BETA
BETA
95%−CI
rs17723637 (G), P=7.85e−05
IHGC2016 MO 0.04 [−0.01; 0.10]
deCODE MO 0.08 [−0.02; 0.18]
DBDS MO 0.11 [ 0.04; 0.17]
UKBB MO 0.38 [ 0.13; 0.63]
LUMINA MO 0.02 [−0.16; 0.21]
Fixed effect model 0.07 [ 0.04; 0.11]
I2 = 51%
−0.6 −0.4 −0.2 0 0.2 0.4 0.6

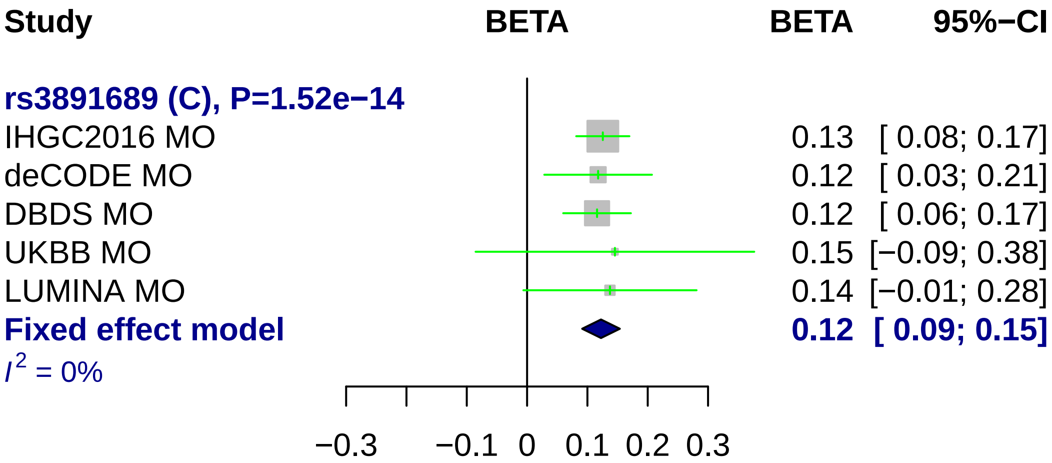
Study
BETA
BETA
95%−CI
rs3891689 (C), P=1.52e−14
IHGC2016 MO 0.13 [ 0.08; 0.17]
deCODE MO 0.12 [ 0.03; 0.21]
DBDS MO 0.12 [ 0.06; 0.17]
UKBB MO 0.15 [−0.09; 0.38]
LUMINA MO 0.14 [−0.01; 0.28]
Fixed effect model 0.12 [ 0.09; 0.15]
I² = 0%
−0.3
−0.1
0
0.1
0.2
0.3

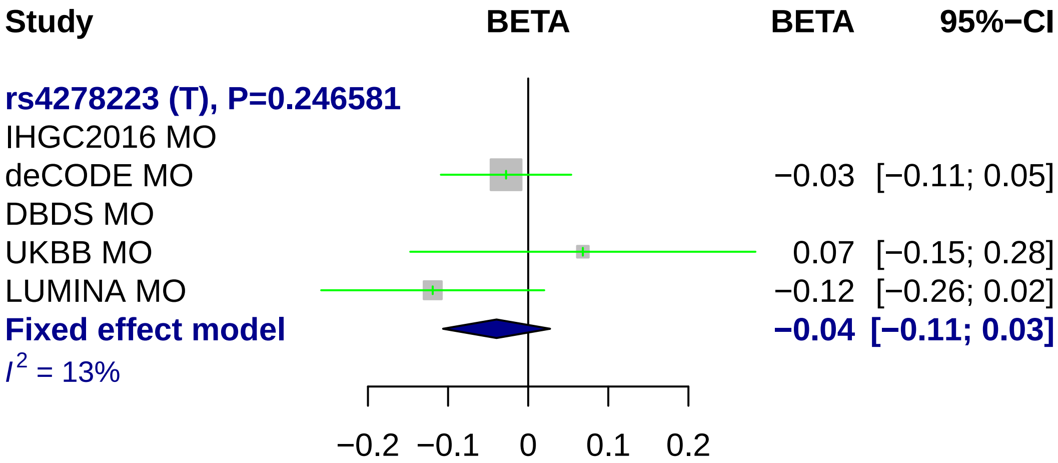

Study
BETA
BETA
95%−CI
rs4278223 (T), P=0.246581
IHGC2016 MO
deCODE MO
−0.03
[−0.11; 0.05]
DBDS MO
UKBB MO
0.07
[−0.15; 0.28]
LUMINA MO
−0.12
[−0.26; 0.02]
Fixed effect model
−0.04
[−0.11; 0.03]
I2 = 13%
−0.2
−0.1
0
0.1
0.2

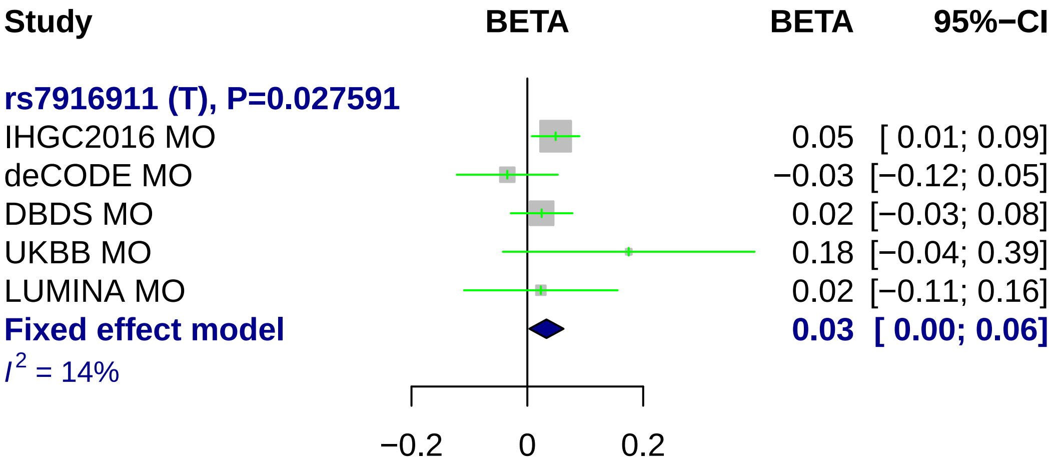
Study
BETA
BETA
95%−CI
rs7916911 (T), P=0.027591
IHGC2016 MO 0.05 [ 0.01; 0.09]
deCODE MO −0.03 [−0.12; 0.05]
DBDS MO 0.02 [−0.03; 0.08]
UKBB MO 0.18 [−0.04; 0.39]
LUMINA MO 0.02 [−0.11; 0.16]
Fixed effect model 0.03 [ 0.00; 0.06]
I2 = 14%
−0.2
0
0.2

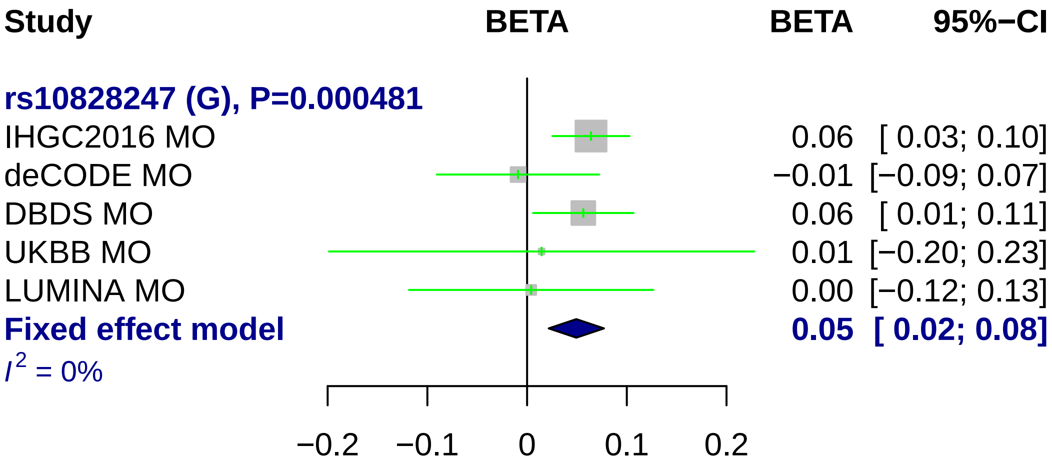
Study
BETA
BETA
95%−CI
rs10828247 (G), P=0.000481
IHGC2016 MO
0.06 [ 0.03; 0.10]
deCODE MO
−0.01 [−0.09; 0.07]
DBDS MO
0.06 [ 0.01; 0.11]
UKBB MO
0.01 [−0.20; 0.23]
LUMINA MO
0.00 [−0.12; 0.13]
Fixed effect model
0.05 [ 0.02; 0.08]
I2 = 0%
−0.2
−0.1
0
0.1
0.2

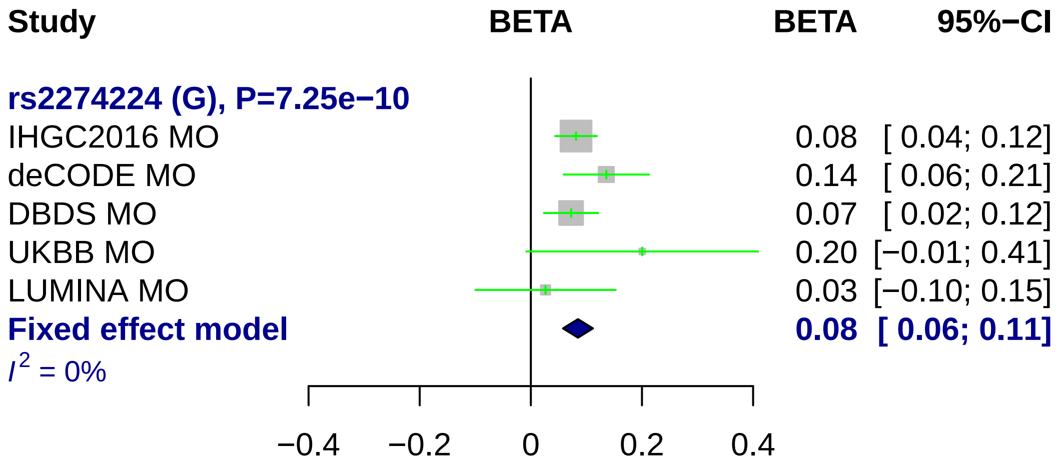

Study
BETA
BETA
95%−CI
rs2274224 (G), P=7.25e−10
IHGC2016 MO
0.08 [ 0.04; 0.12]
deCODE MO
0.14 [ 0.06; 0.21]
DBDS MO
0.07 [ 0.02; 0.12]
UKBB MO
0.20 [−0.01; 0.41]
LUMINA MO
0.03 [−0.10; 0.15]
Fixed effect model
0.08 [ 0.06; 0.11]
I2 = 0%
−0.4
−0.2
0
0.2
0.4

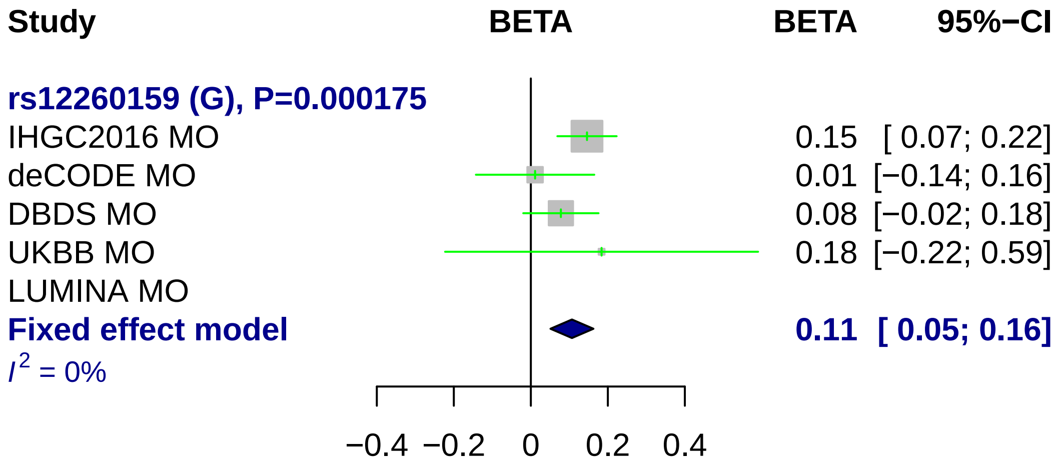
Study
BETA
BETA
95%−CI
rs12260159 (G), P=0.000175
IHGC2016 MO
0.15 [ 0.07; 0.22]
deCODE MO
0.01 [−0.14; 0.16]
DBDS MO
0.08 [−0.02; 0.18]
UKBB MO
0.18 [−0.22; 0.59]
LUMINA MO
Fixed effect model
0.11 [ 0.05; 0.16]
I2 = 0%
−0.4
−0.2
0
0.2
0.4

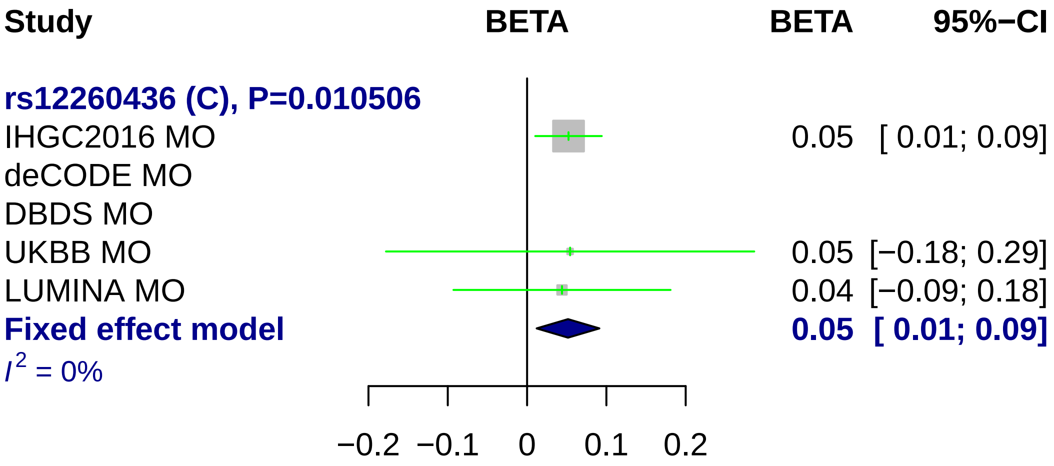
Study
BETA
BETA
95%−CI
rs12260436 (C), P=0.010506
IHGC2016 MO
0.05 [ 0.01; 0.09]
deCODE MO
DBDS MO
UKBB MO
0.05 [−0.18; 0.29]
LUMINA MO
0.04 [−0.09; 0.18]
Fixed effect model
0.05 [ 0.01; 0.09]
I2 = 0%
−0.2
−0.1
0
0.1
0.2

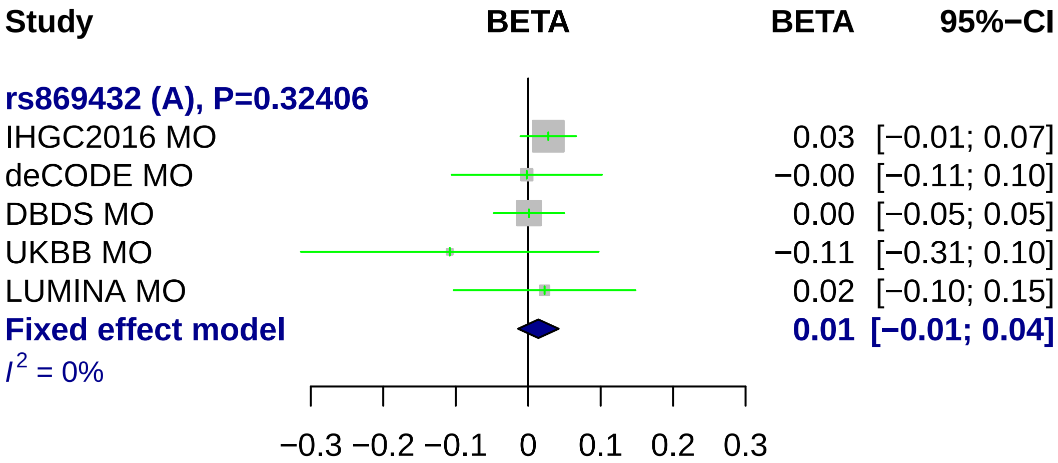
Study
BETA
BETA
95%−CI
rs869432 (A), P=0.32406
IHGC2016 MO
0.03 [−0.01; 0.07]
deCODE MO
−0.00 [−0.11; 0.10]
DBDS MO
0.00 [−0.05; 0.05]
UKBB MO
−0.11 [−0.31; 0.10]
LUMINA MO
0.02 [−0.10; 0.15]
Fixed effect model
0.01 [−0.01; 0.04]
I² = 0%
−0.3
−0.2
−0.1
0
0.1
0.2
0.3

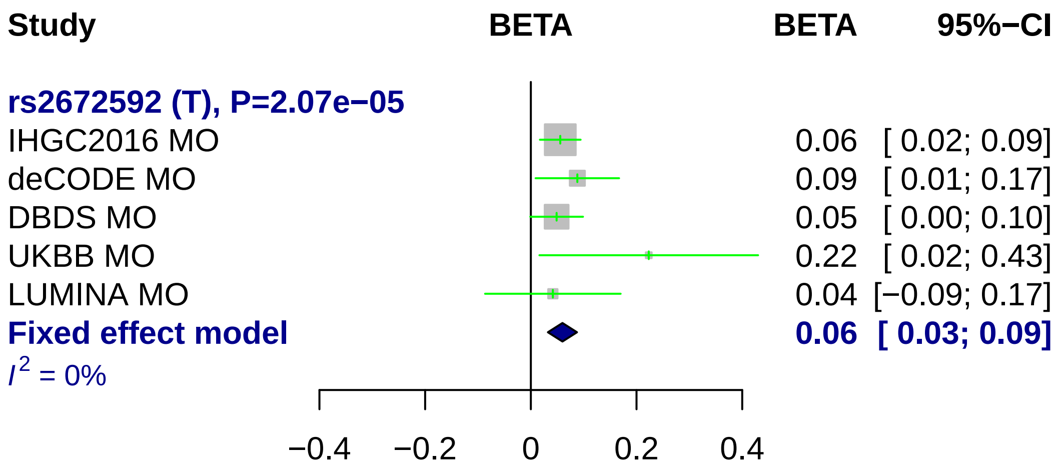
Study
BETA
BETA
95%−CI
rs2672592 (T), P=2.07e−05
IHGC2016 MO 0.06 [ 0.02; 0.09]
deCODE MO 0.09 [ 0.01; 0.17]
DBDS MO 0.05 [ 0.00; 0.10]
UKBB MO 0.22 [ 0.02; 0.43]
LUMINA MO 0.04 [−0.09; 0.17]
Fixed effect model 0.06 [ 0.03; 0.09]
I2 = 0%
−0.4
−0.2
0
0.2
0.4

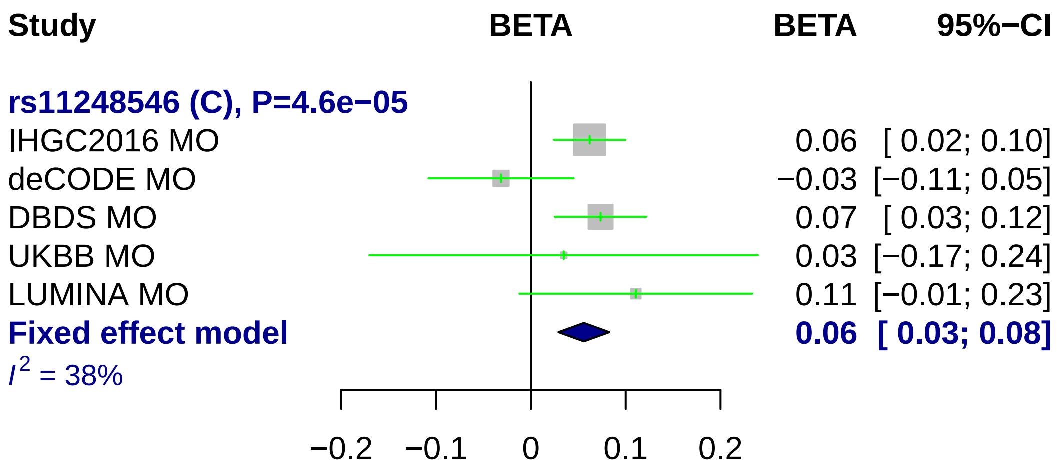
Study
BETA
BETA
95%−CI
rs11248546 (C), P=4.6e−05
IHGC2016 MO
0.06 [ 0.02; 0.10]
deCODE MO
−0.03 [−0.11; 0.05]
DBDS MO
0.07 [ 0.03; 0.12]
UKBB MO
0.03 [−0.17; 0.24]
LUMINA MO
0.11 [−0.01; 0.23]
Fixed effect model
0.06 [ 0.03; 0.08]
I2 = 38%
−0.2
−0.1
0
0.1
0.2

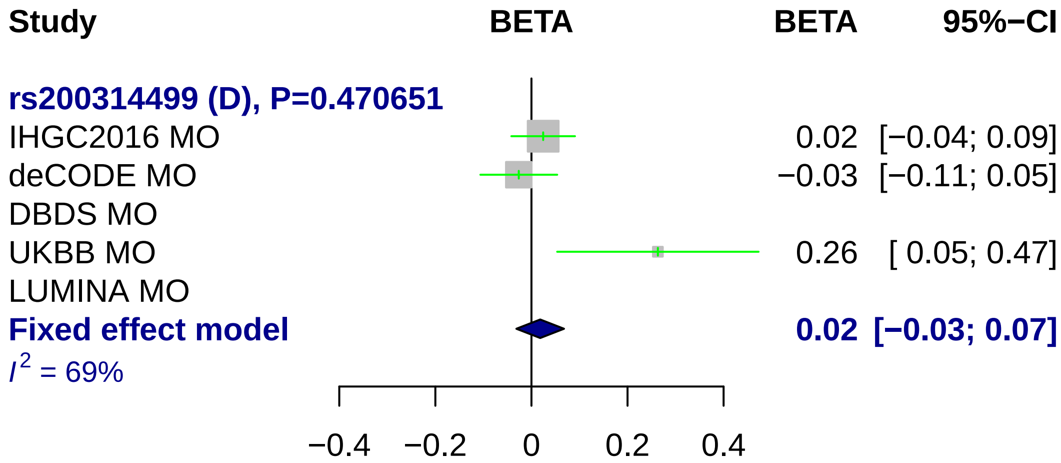
Study
BETA
BETA
95%−CI
rs200314499 (D), P=0.470651
IHGC2016 MO
0.02
[−0.04; 0.09]
deCODE MO
−0.03
[−0.11; 0.05]
DBDS MO
UKBB MO
0.26
[ 0.05; 0.47]
LUMINA MO
Fixed effect model
0.02
[−0.03; 0.07]
I^2 = 69%
−0.4
−0.2
0
0.2
0.4

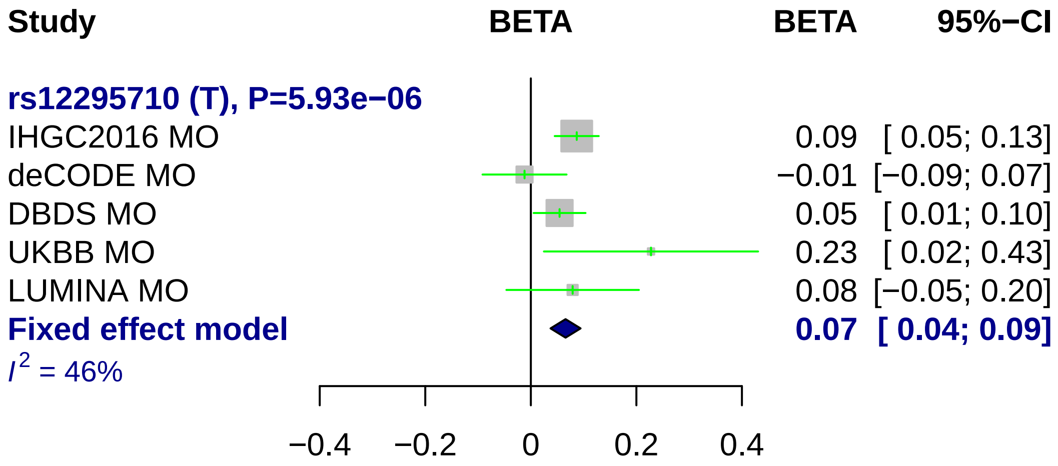
Study
BETA
BETA
95%−CI
rs12295710 (T), P=5.93e−06
IHGC2016 MO
0.09 [ 0.05; 0.13]
deCODE MO
−0.01 [−0.09; 0.07]
DBDS MO
0.05 [ 0.01; 0.10]
UKBB MO
0.23 [ 0.02; 0.43]
LUMINA MO
0.08 [−0.05; 0.20]
Fixed effect model
0.07 [ 0.04; 0.09]
I2 = 46%
−0.4
−0.2
0
0.2
0.4

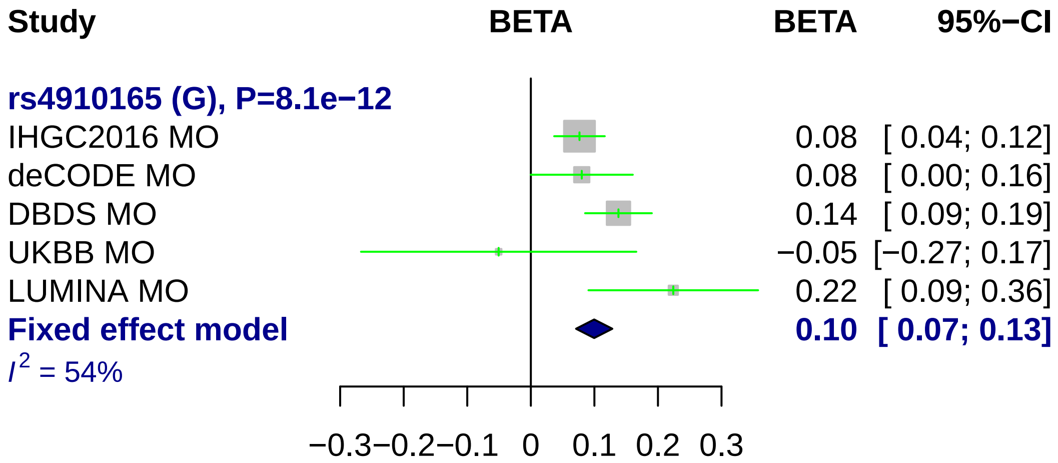
Study
BETA
BETA
95%−CI
rs4910165 (G), P=8.1e−12
IHGC2016 MO 0.08 [ 0.04; 0.12]
deCODE MO 0.08 [ 0.00; 0.16]
DBDS MO 0.14 [ 0.09; 0.19]
UKBB MO −0.05 [−0.27; 0.17]
LUMINA MO 0.22 [ 0.09; 0.36]
Fixed effect model 0.10 [ 0.07; 0.13]
I^2 = 54%
−0.3 −0.2 −0.1 0 0.1 0.2 0.3

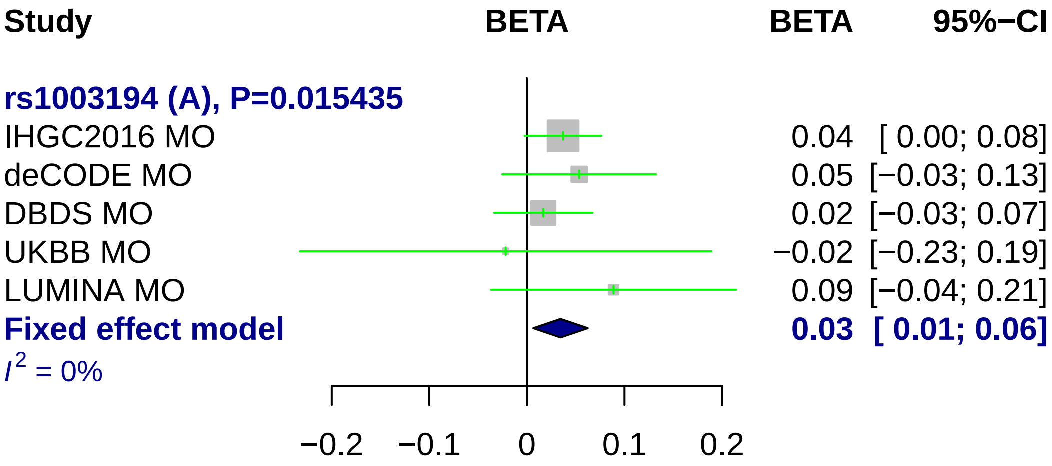
Study
BETA
BETA
95%−CI
rs1003194 (A), P=0.015435
IHGC2016 MO
0.04 [ 0.00; 0.08]
deCODE MO
0.05 [−0.03; 0.13]
DBDS MO
0.02 [−0.03; 0.07]
UKBB MO
−0.02 [−0.23; 0.19]
LUMINA MO
0.09 [−0.04; 0.21]
Fixed effect model
0.03 [ 0.01; 0.06]
$I^2$ = 0%
−0.2
−0.1
0
0.1
0.2

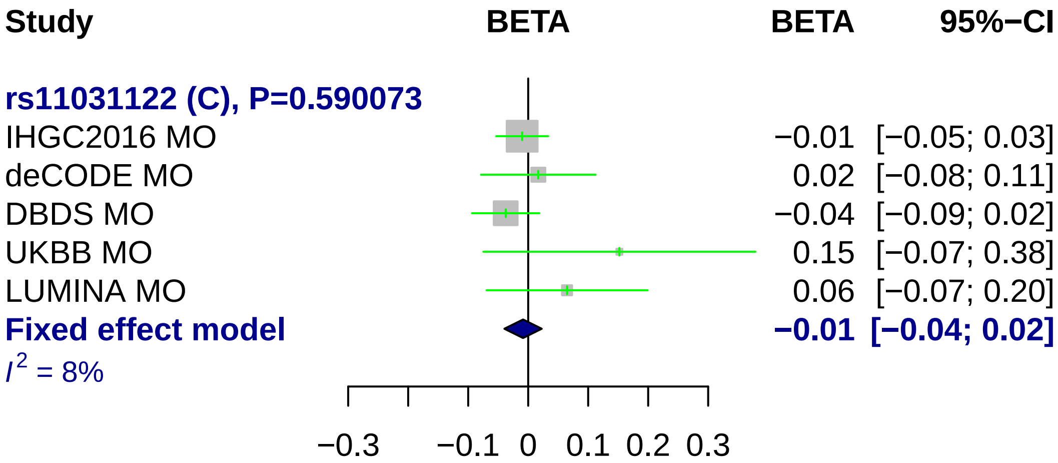
Study
BETA
BETA
95%−CI
rs11031122 (C), P=0.590073
IHGC2016 MO
−0.01 [−0.05; 0.03]
deCODE MO
0.02 [−0.08; 0.11]
DBDS MO
−0.04 [−0.09; 0.02]
UKBB MO
0.15 [−0.07; 0.38]
LUMINA MO
0.06 [−0.07; 0.20]
Fixed effect model
−0.01 [−0.04; 0.02]
I2 = 8%
−0.3
−0.1
0
0.1
0.2
0.3

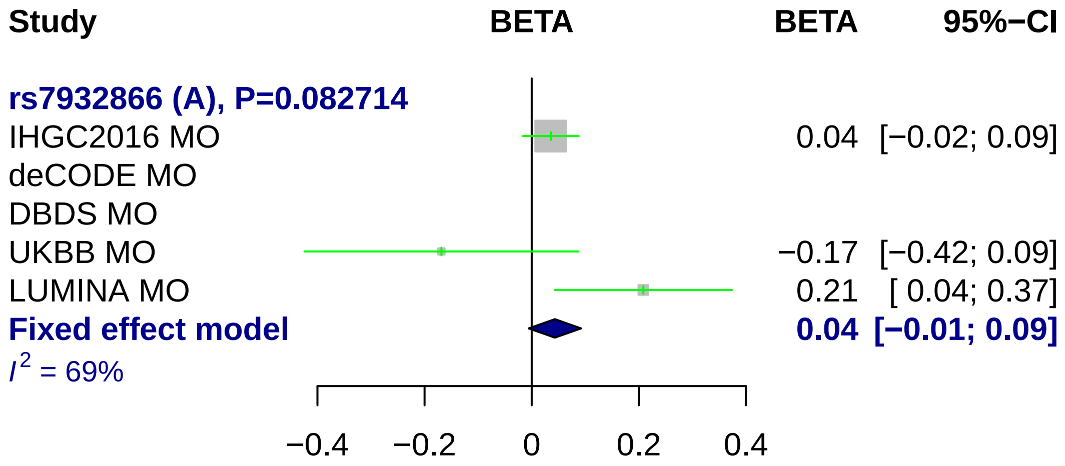

Study
BETA
BETA
95%−CI
rs7932866 (A), P=0.082714
IHGC2016 MO
0.04 [−0.02; 0.09]
deCODE MO
DBDS MO
UKBB MO
−0.17 [−0.42; 0.09]
LUMINA MO
0.21 [ 0.04; 0.37]
Fixed effect model
0.04 [−0.01; 0.09]
$I^2$ = 69%
−0.4
−0.2
0
0.2
0.4

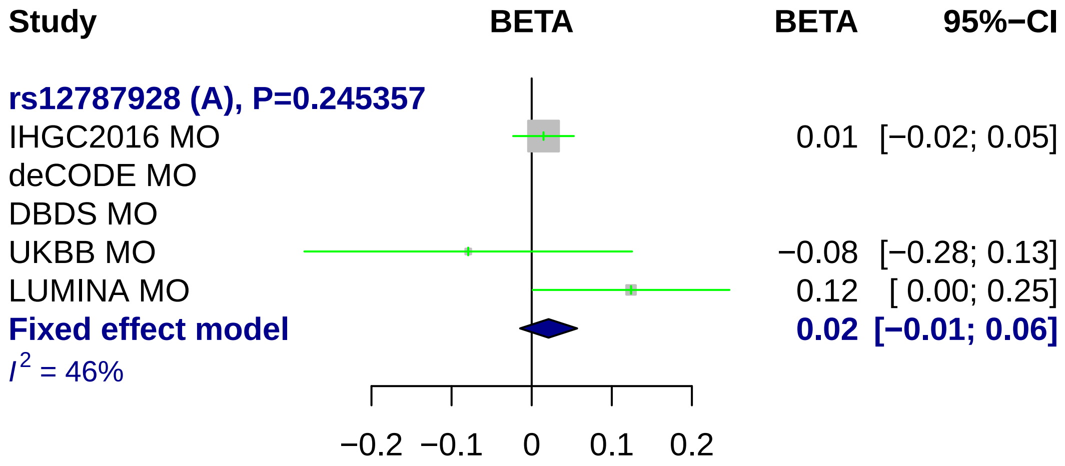

Study
BETA
BETA
95%−CI
rs12787928 (A), P=0.245357
IHGC2016 MO
0.01 [−0.02; 0.05]
deCODE MO
DBDS MO
UKBB MO
−0.08 [−0.28; 0.13]
LUMINA MO
0.12 [ 0.00; 0.25]
Fixed effect model
0.02 [−0.01; 0.06]
I2 = 46%
−0.2
−0.1
0
0.1
0.2

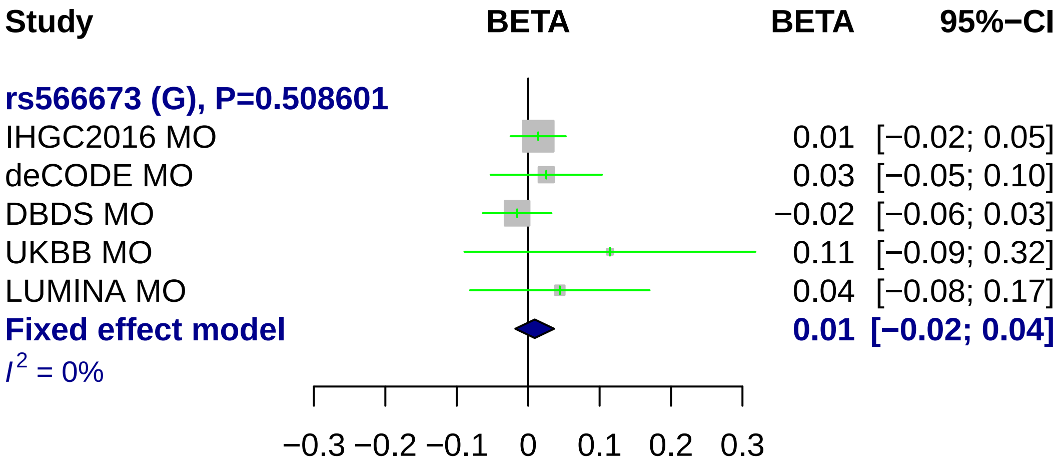
Study
BETA
BETA
95%−CI
rs566673 (G), P=0.508601
IHGC2016 MO 0.01 [−0.02; 0.05]
deCODE MO 0.03 [−0.05; 0.10]
DBDS MO −0.02 [−0.06; 0.03]
UKBB MO 0.11 [−0.09; 0.32]
LUMINA MO 0.04 [−0.08; 0.17]
Fixed effect model 0.01 [−0.02; 0.04]
I2 = 0%
−0.3 −0.2 −0.1 0 0.1 0.2 0.3

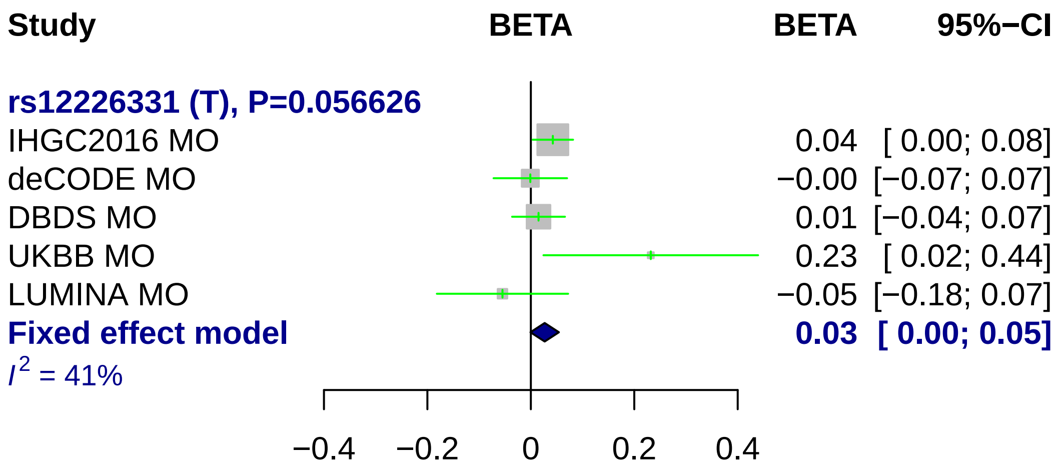

Study
BETA
BETA
95%−CI
rs12226331 (T), P=0.056626
IHGC2016 MO
0.04 [ 0.00; 0.08]
deCODE MO
−0.00 [−0.07; 0.07]
DBDS MO
0.01 [−0.04; 0.07]
UKBB MO
0.23 [ 0.02; 0.44]
LUMINA MO
−0.05 [−0.18; 0.07]
Fixed effect model
0.03 [ 0.00; 0.05]
I2 = 41%
−0.4
−0.2
0
0.2
0.4

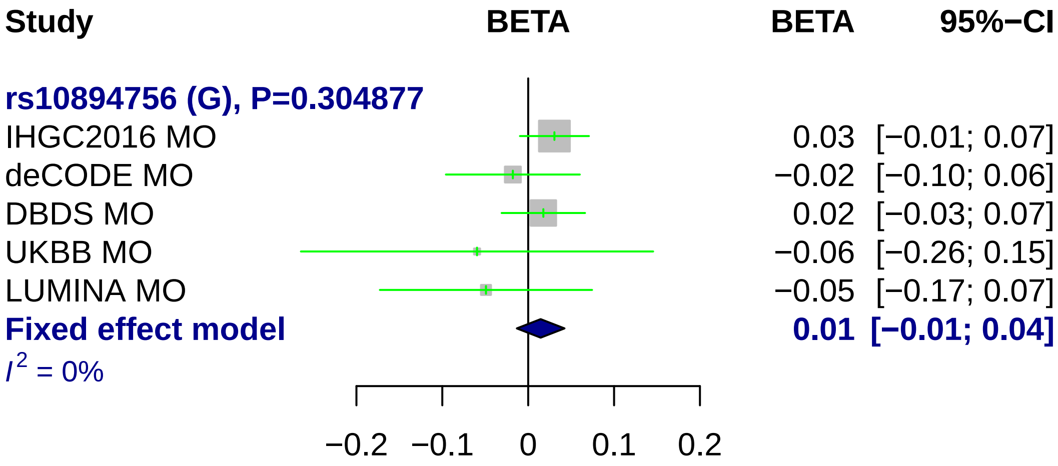
Study
BETA
BETA
95%−CI
rs10894756 (G), P=0.304877
IHGC2016 MO
0.03 [−0.01; 0.07]
deCODE MO
−0.02 [−0.10; 0.06]
DBDS MO
0.02 [−0.03; 0.07]
UKBB MO
−0.06 [−0.26; 0.15]
LUMINA MO
−0.05 [−0.17; 0.07]
Fixed effect model
0.01 [−0.01; 0.04]
I^2 = 0%
−0.2
−0.1
0
0.1
0.2

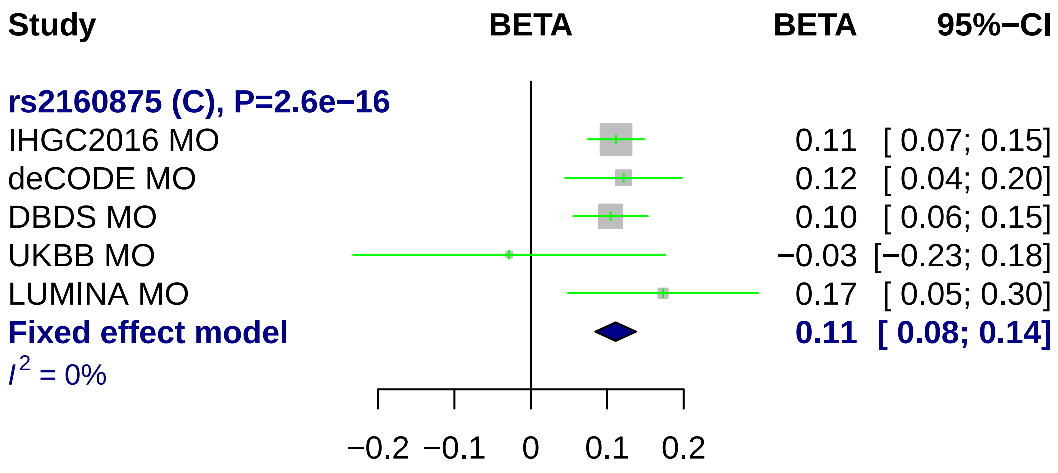
Study
BETA
BETA
95%−CI
rs2160875 (C), P=2.6e−16
IHGC2016 MO
0.11 [ 0.07; 0.15]
deCODE MO
0.12 [ 0.04; 0.20]
DBDS MO
0.10 [ 0.06; 0.15]
UKBB MO
−0.03 [−0.23; 0.18]
LUMINA MO
0.17 [ 0.05; 0.30]
Fixed effect model
0.11 [ 0.08; 0.14]
I² = 0%
−0.2
−0.1
0
0.1
0.2

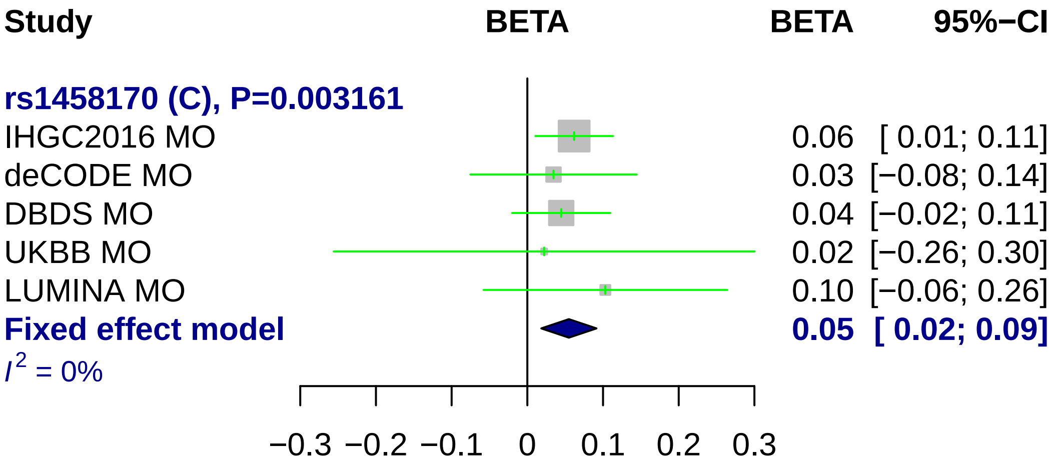

Study
BETA
BETA
95%−CI
rs1458170 (C), P=0.003161
IHGC2016 MO
0.06 [ 0.01; 0.11]
deCODE MO
0.03 [−0.08; 0.14]
DBDS MO
0.04 [−0.02; 0.11]
UKBB MO
0.02 [−0.26; 0.30]
LUMINA MO
0.10 [−0.06; 0.26]
Fixed effect model
0.05 [ 0.02; 0.09]
I2 = 0%
−0.3
−0.2
−0.1
0
0.1
0.2
0.3

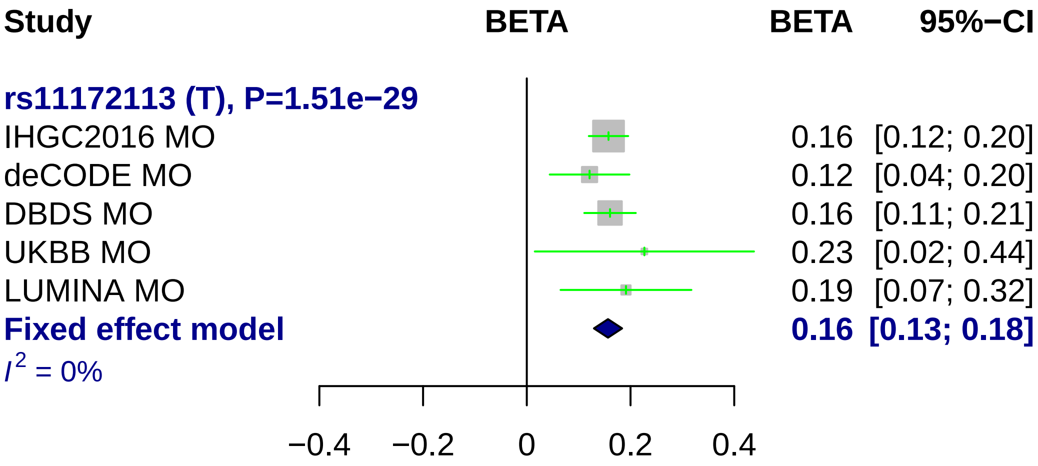
Study
BETA
BETA
95%−CI
rs11172113 (T), P=1.51e−29
IHGC2016 MO
0.16 [0.12; 0.20]
deCODE MO
0.12 [0.04; 0.20]
DBDS MO
0.16 [0.11; 0.21]
UKBB MO
0.23 [0.02; 0.44]
LUMINA MO
0.19 [0.07; 0.32]
Fixed effect model
0.16 [0.13; 0.18]
I2 = 0%
−0.4
−0.2
0
0.2
0.4

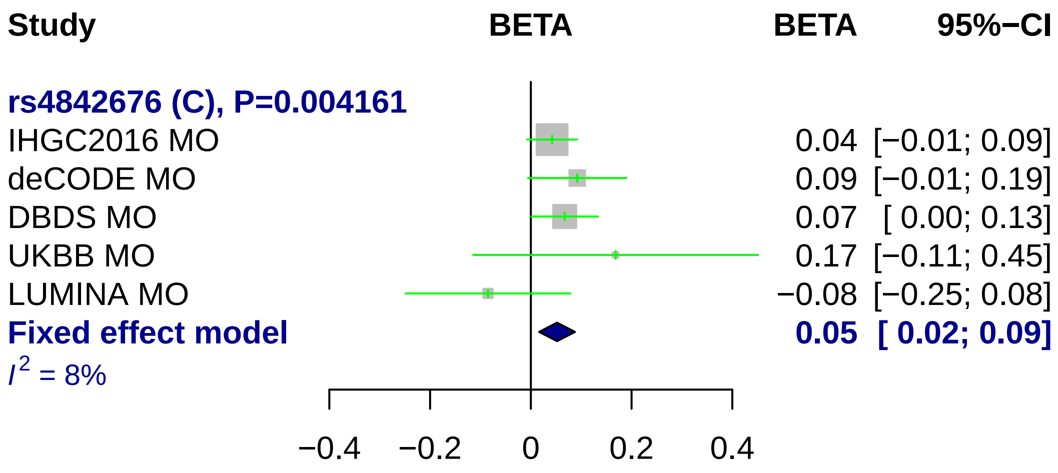
Study
BETA
BETA
95%−CI
rs4842676 (C), P=0.004161
IHGC2016 MO
0.04 [−0.01; 0.09]
deCODE MO
0.09 [−0.01; 0.19]
DBDS MO
0.07 [ 0.00; 0.13]
UKBB MO
0.17 [−0.11; 0.45]
LUMINA MO
−0.08 [−0.25; 0.08]
Fixed effect model
0.05 [ 0.02; 0.09]
I² = 8%
−0.4
−0.2
0
0.2
0.4

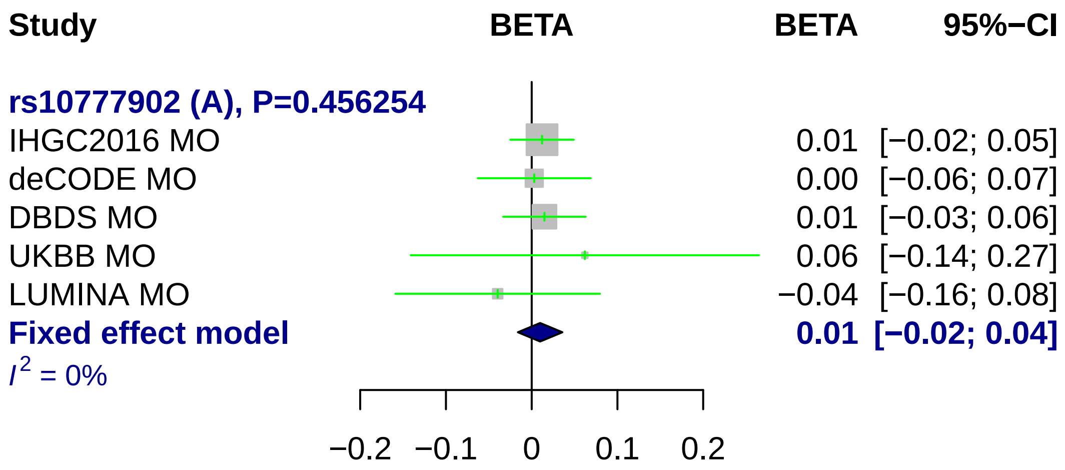
Study
BETA
BETA
95%−CI
rs10777902 (A), P=0.456254
IHGC2016 MO 0.01 [−0.02; 0.05]
deCODE MO 0.00 [−0.06; 0.07]
DBDS MO 0.01 [−0.03; 0.06]
UKBB MO 0.06 [−0.14; 0.27]
LUMINA MO −0.04 [−0.16; 0.08]
Fixed effect model 0.01 [−0.02; 0.04]
I² = 0%
−0.2
−0.1
0
0.1
0.2

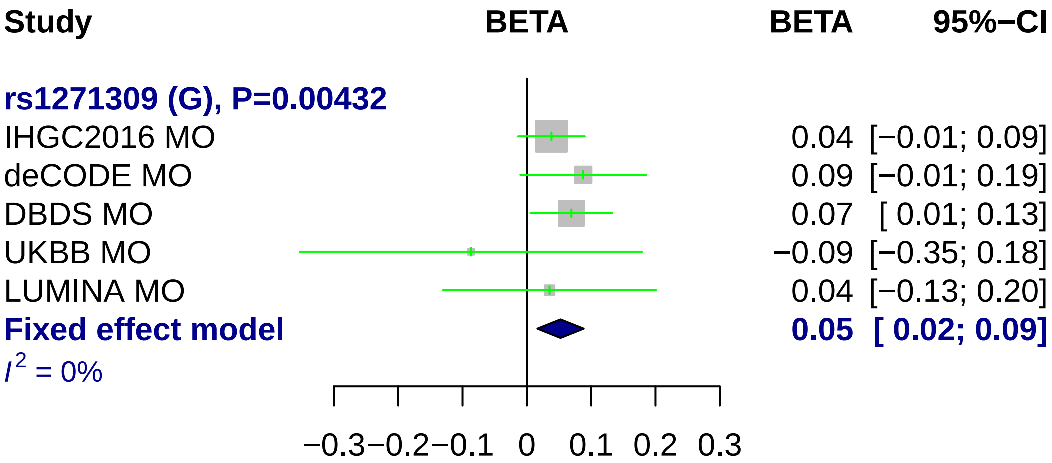
Study
BETA
BETA
95%−CI
rs1271309 (G), P=0.00432
IHGC2016 MO
0.04 [−0.01; 0.09]
deCODE MO
0.09 [−0.01; 0.19]
DBDS MO
0.07 [ 0.01; 0.13]
UKBB MO
−0.09 [−0.35; 0.18]
LUMINA MO
0.04 [−0.13; 0.20]
Fixed effect model
0.05 [ 0.02; 0.09]
I2 = 0%
−0.3 −0.2 −0.1 0 0.1 0.2 0.3

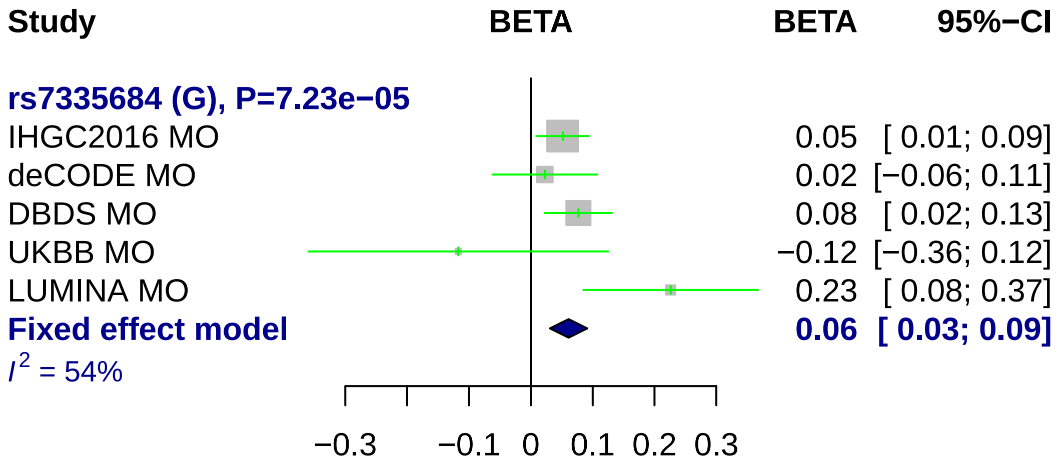
Study
BETA
BETA
95%−CI
rs7335684 (G), P=7.23e−05
IHGC2016 MO
0.05
[ 0.01; 0.09]
deCODE MO
0.02
[−0.06; 0.11]
DBDS MO
0.08
[ 0.02; 0.13]
UKBB MO
−0.12
[−0.36; 0.12]
LUMINA MO
0.23
[ 0.08; 0.37]
Fixed effect model
0.06
[ 0.03; 0.09]
I2 = 54%
−0.3
−0.1
0
0.1
0.2
0.3

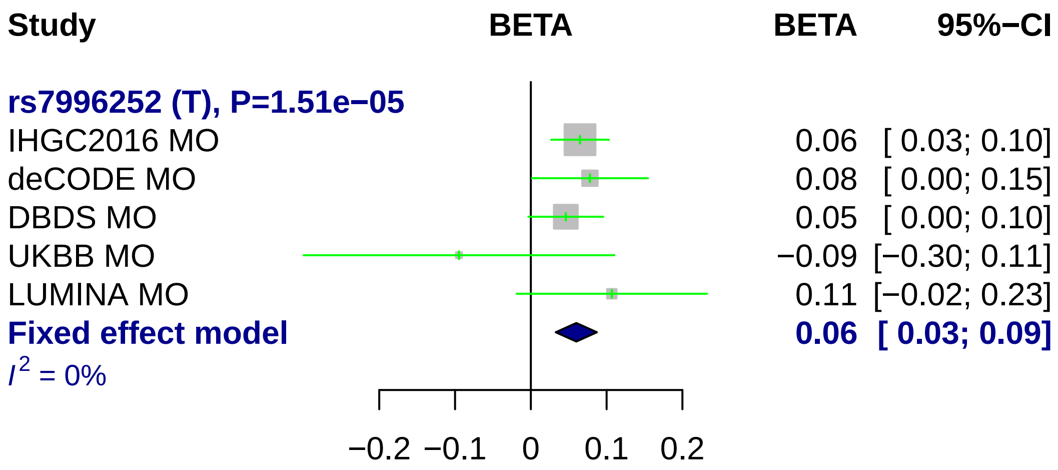
Study
BETA
BETA
95%−CI
rs7996252 (T), P=1.51e−05
IHGC2016 MO 0.06 [ 0.03; 0.10]
deCODE MO 0.08 [ 0.00; 0.15]
DBDS MO 0.05 [ 0.00; 0.10]
UKBB MO −0.09 [−0.30; 0.11]
LUMINA MO 0.11 [−0.02; 0.23]
Fixed effect model 0.06 [ 0.03; 0.09]
I2 = 0%
−0.2 −0.1 0 0.1 0.2

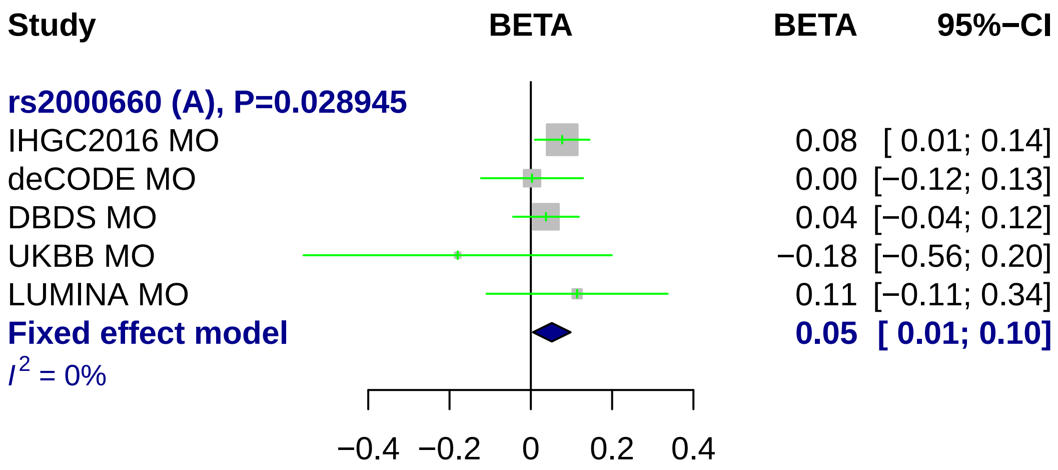
Study
BETA
BETA
95%−CI
rs2000660 (A), P=0.028945
IHGC2016 MO
0.08 [ 0.01; 0.14]
deCODE MO
0.00 [−0.12; 0.13]
DBDS MO
0.04 [−0.04; 0.12]
UKBB MO
−0.18 [−0.56; 0.20]
LUMINA MO
0.11 [−0.11; 0.34]
Fixed effect model
0.05 [ 0.01; 0.10]
I2 = 0%
−0.4
−0.2
0
0.2
0.4

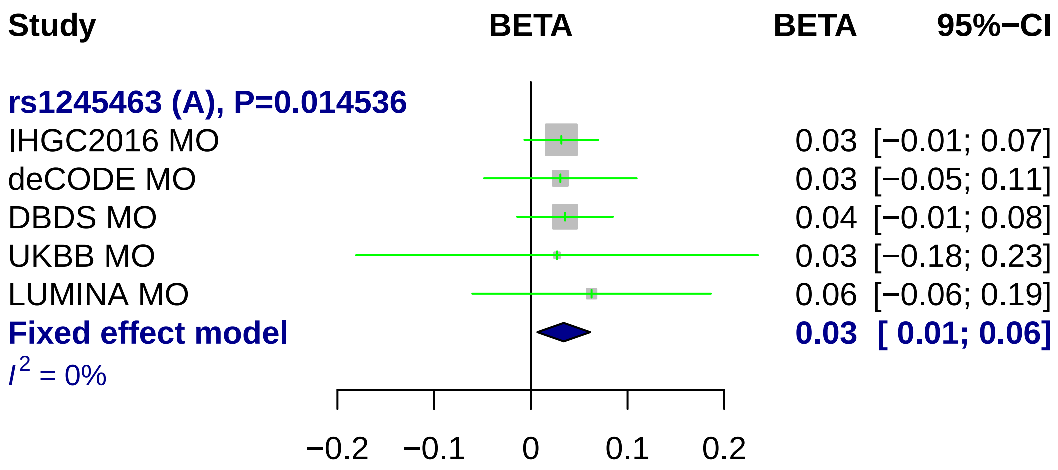
Study
BETA
BETA
95%−CI
rs1245463 (A), P=0.014536
IHGC2016 MO
0.03 [−0.01; 0.07]
deCODE MO
0.03 [−0.05; 0.11]
DBDS MO
0.04 [−0.01; 0.08]
UKBB MO
0.03 [−0.18; 0.23]
LUMINA MO
0.06 [−0.06; 0.19]
Fixed effect model
0.03 [ 0.01; 0.06]
I2 = 0%
−0.2
−0.1
0
0.1
0.2

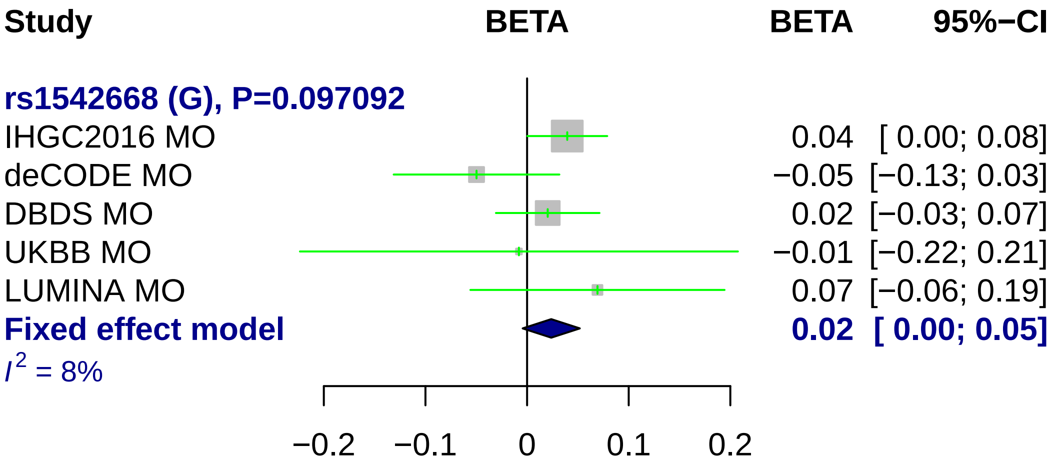

Study
BETA
BETA
95%−CI
rs1542668 (G), P=0.097092
IHGC2016 MO
0.04 [ 0.00; 0.08]
deCODE MO
−0.05 [−0.13; 0.03]
DBDS MO
0.02 [−0.03; 0.07]
UKBB MO
−0.01 [−0.22; 0.21]
LUMINA MO
0.07 [−0.06; 0.19]
Fixed effect model
0.02 [ 0.00; 0.05]
$I^2$ = 8%
−0.2
−0.1
0
0.1
0.2

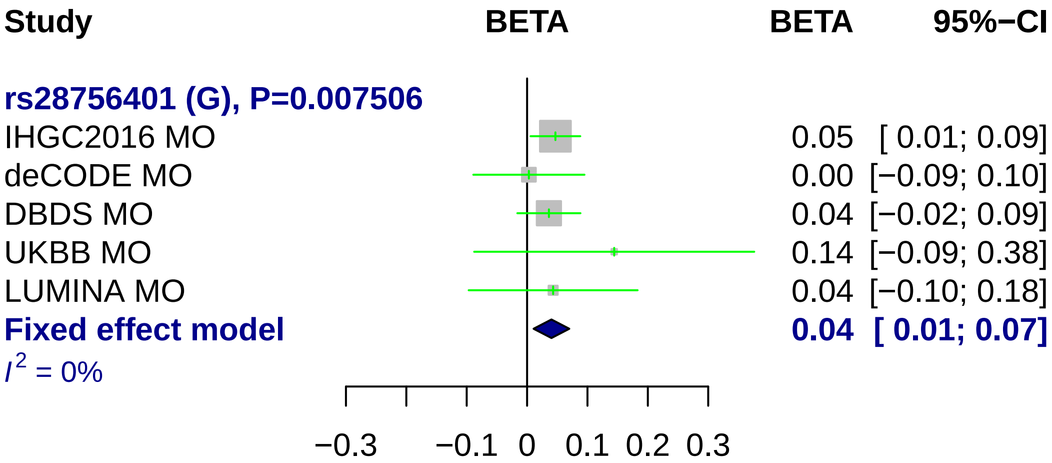
Study
BETA
BETA
95%−CI
rs28756401 (G), P=0.007506
IHGC2016 MO
0.05 [ 0.01; 0.09]
deCODE MO
0.00 [−0.09; 0.10]
DBDS MO
0.04 [−0.02; 0.09]
UKBB MO
0.14 [−0.09; 0.38]
LUMINA MO
0.04 [−0.10; 0.18]
Fixed effect model
0.04 [ 0.01; 0.07]
I2 = 0%
−0.3
−0.1
0
0.1
0.2
0.3

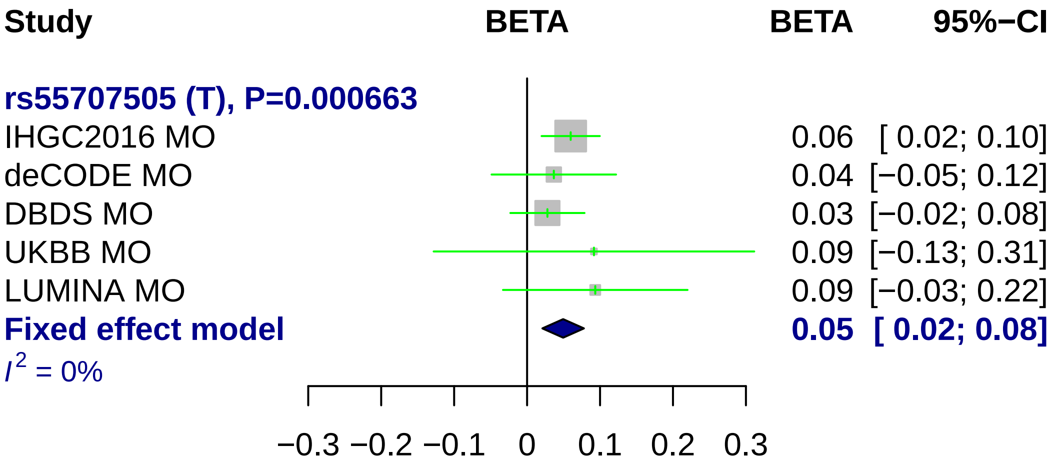
Study
BETA
BETA
95%−CI
rs55707505 (T), P=0.000663
IHGC2016 MO
0.06 [ 0.02; 0.10]
deCODE MO
0.04 [−0.05; 0.12]
DBDS MO
0.03 [−0.02; 0.08]
UKBB MO
0.09 [−0.13; 0.31]
LUMINA MO
0.09 [−0.03; 0.22]
Fixed effect model
0.05 [ 0.02; 0.08]
$I^2$ = 0%
−0.3
−0.2
−0.1
0
0.1
0.2
0.3

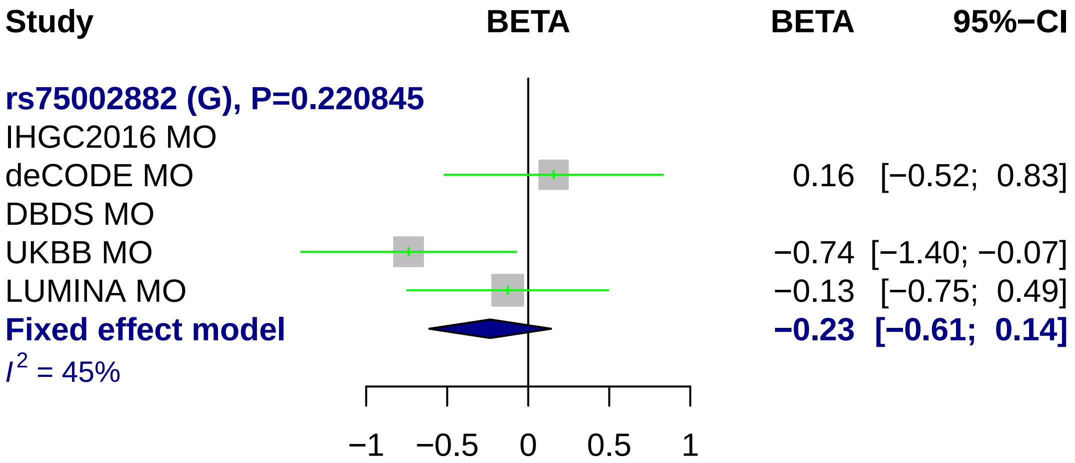
Study
BETA
BETA
95%−CI
rs75002882 (G), P=0.220845
IHGC2016 MO
deCODE MO
0.16 [−0.52; 0.83]
DBDS MO
UKBB MO
−0.74 [−1.40; −0.07]
LUMINA MO
−0.13 [−0.75; 0.49]
Fixed effect model
−0.23 [−0.61; 0.14]
I2 = 45%
−1
−0.5
0
0.5
1

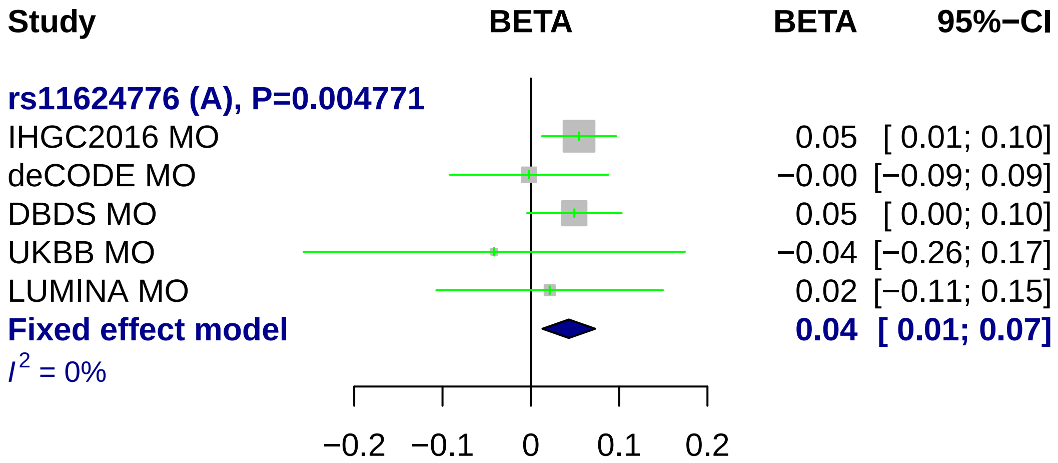
Study
BETA
BETA
95%−CI
rs11624776 (A), P=0.004771
IHGC2016 MO
0.05 [ 0.01; 0.10]
deCODE MO
−0.00 [−0.09; 0.09]
DBDS MO
0.05 [ 0.00; 0.10]
UKBB MO
−0.04 [−0.26; 0.17]
LUMINA MO
0.02 [−0.11; 0.15]
Fixed effect model
0.04 [ 0.01; 0.07]
I2 = 0%
−0.2
−0.1
0
0.1
0.2

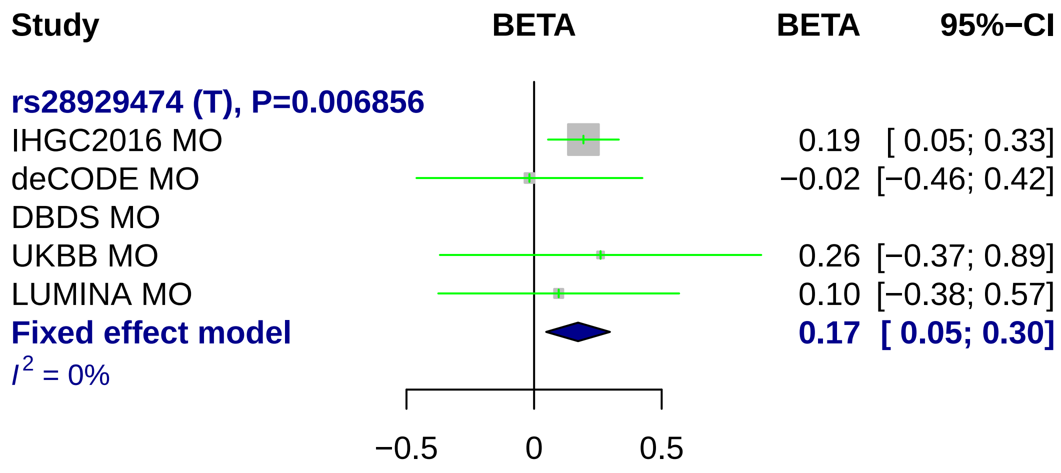
Study
BETA
BETA
95%−CI
rs28929474 (T), P=0.006856
IHGC2016 MO
0.19 [ 0.05; 0.33]
deCODE MO
−0.02 [−0.46; 0.42]
DBDS MO
UKBB MO
0.26 [−0.37; 0.89]
LUMINA MO
0.10 [−0.38; 0.57]
Fixed effect model
0.17 [ 0.05; 0.30]
I2 = 0%
−0.5
0
0.5

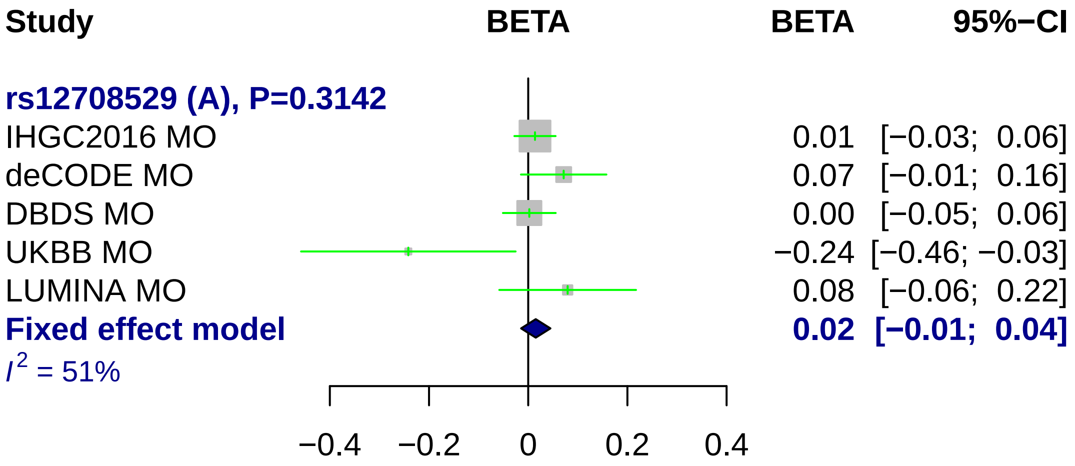

Study
BETA
BETA
95%−CI
rs12708529 (A), P=0.3142
IHGC2016 MO
0.01 [−0.03; 0.06]
deCODE MO
0.07 [−0.01; 0.16]
DBDS MO
0.00 [−0.05; 0.06]
UKBB MO
−0.24 [−0.46; −0.03]
LUMINA MO
0.08 [−0.06; 0.22]
Fixed effect model
0.02 [−0.01; 0.04]
I2 = 51%
−0.4
−0.2
0
0.2
0.4

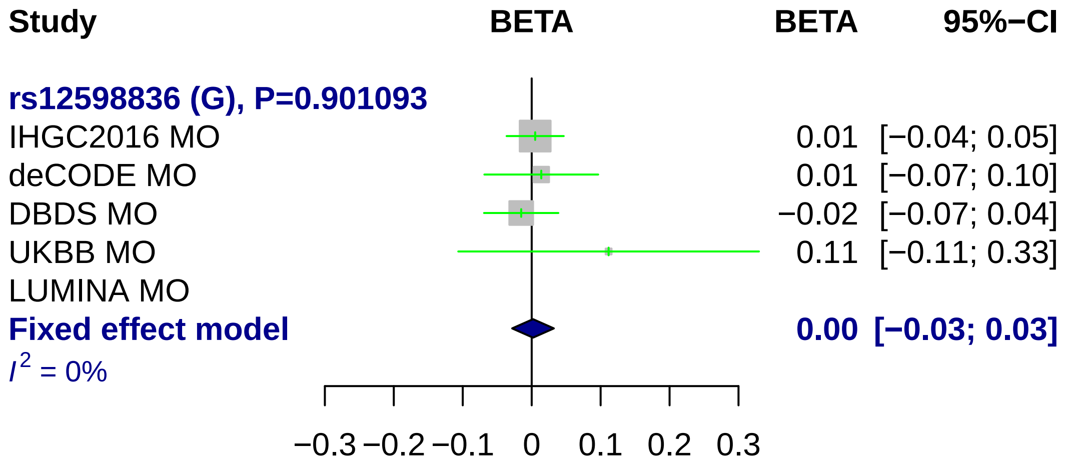

Study
BETA
BETA
95%−CI
rs12598836 (G), P=0.901093
IHGC2016 MO
0.01 [−0.04; 0.05]
deCODE MO
0.01 [−0.07; 0.10]
DBDS MO
−0.02 [−0.07; 0.04]
UKBB MO
0.11 [−0.11; 0.33]
LUMINA MO
Fixed effect model
0.00 [−0.03; 0.03]
I^2 = 0%
−0.3 −0.2 −0.1 0 0.1 0.2 0.3

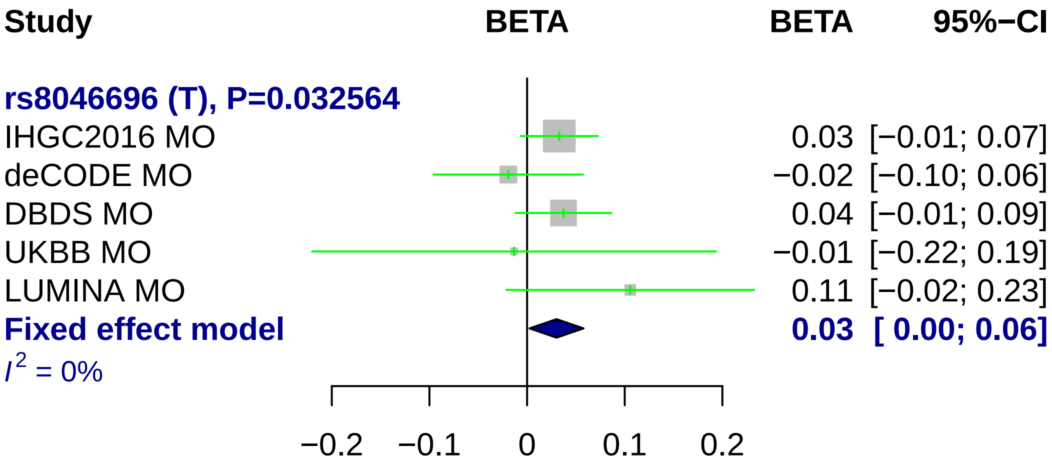
Study
BETA
BETA
95%−CI
rs8046696 (T), P=0.032564
IHGC2016 MO
0.03 [−0.01; 0.07]
deCODE MO
−0.02 [−0.10; 0.06]
DBDS MO
0.04 [−0.01; 0.09]
UKBB MO
−0.01 [−0.22; 0.19]
LUMINA MO
0.11 [−0.02; 0.23]
Fixed effect model
0.03 [ 0.00; 0.06]
I^2 = 0%
−0.2
−0.1
0
0.1
0.2

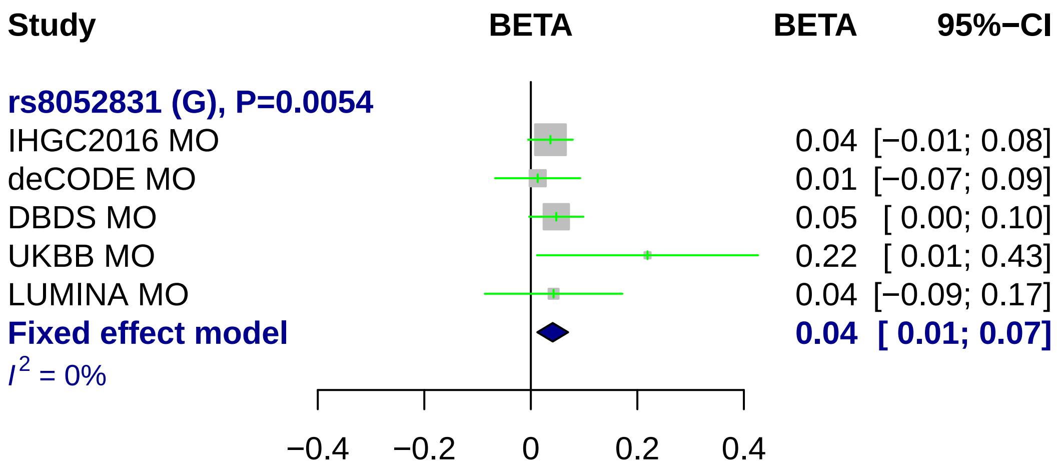
Study
BETA
BETA
95%−CI
rs8052831 (G), P=0.0054
IHGC2016 MO
0.04 [−0.01; 0.08]
deCODE MO
0.01 [−0.07; 0.09]
DBDS MO
0.05 [ 0.00; 0.10]
UKBB MO
0.22 [ 0.01; 0.43]
LUMINA MO
0.04 [−0.09; 0.17]
Fixed effect model
0.04 [ 0.01; 0.07]
I2 = 0%
−0.4
−0.2
0
0.2
0.4

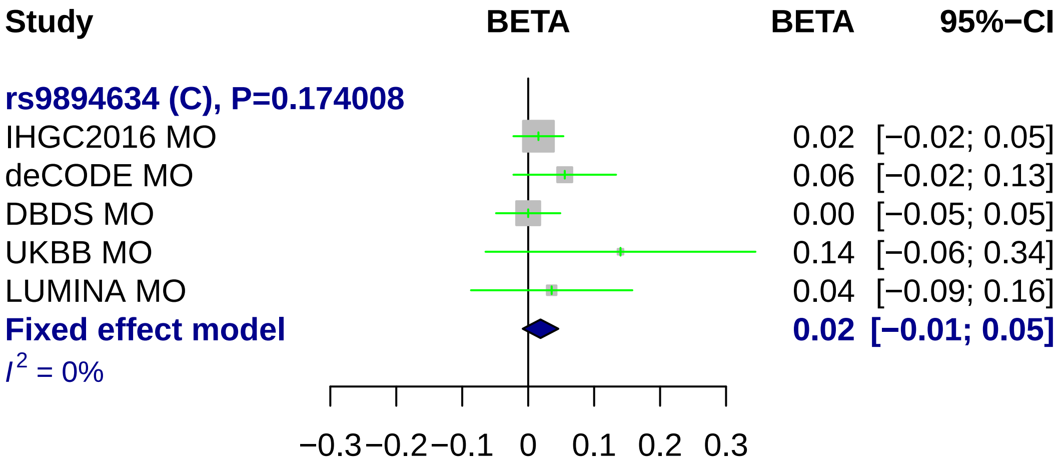
Study
BETA
BETA
95%−CI
rs9894634 (C), P=0.174008
IHGC2016 MO
0.02 [−0.02; 0.05]
deCODE MO
0.06 [−0.02; 0.13]
DBDS MO
0.00 [−0.05; 0.05]
UKBB MO
0.14 [−0.06; 0.34]
LUMINA MO
0.04 [−0.09; 0.16]
Fixed effect model
0.02 [−0.01; 0.05]
I2 = 0%
−0.3
−0.2
−0.1
0
0.1
0.2
0.3

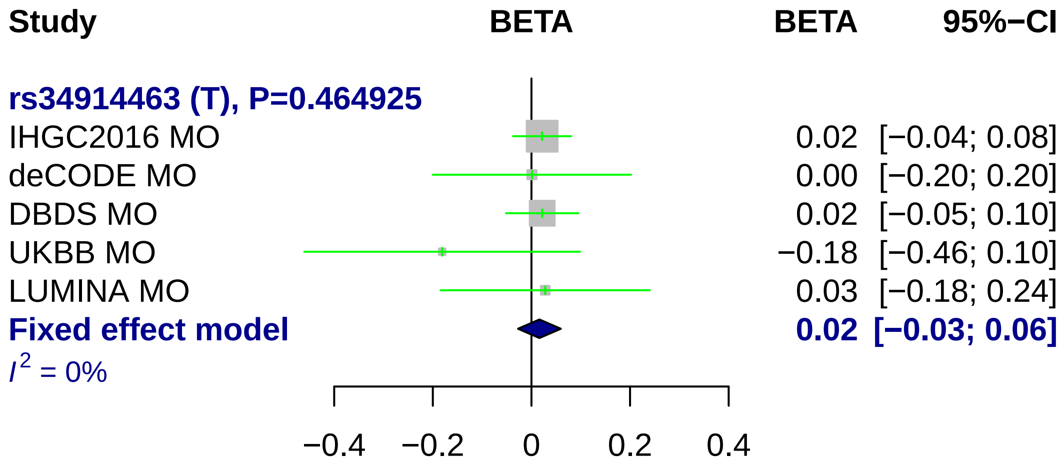
Study
BETA
BETA
95%−CI
rs34914463 (T), P=0.464925
IHGC2016 MO 0.02 [−0.04; 0.08]
deCODE MO 0.00 [−0.20; 0.20]
DBDS MO 0.02 [−0.05; 0.10]
UKBB MO −0.18 [−0.46; 0.10]
LUMINA MO 0.03 [−0.18; 0.24]
Fixed effect model 0.02 [−0.03; 0.06]
I2 = 0%
−0.4
−0.2
0
0.2
0.4

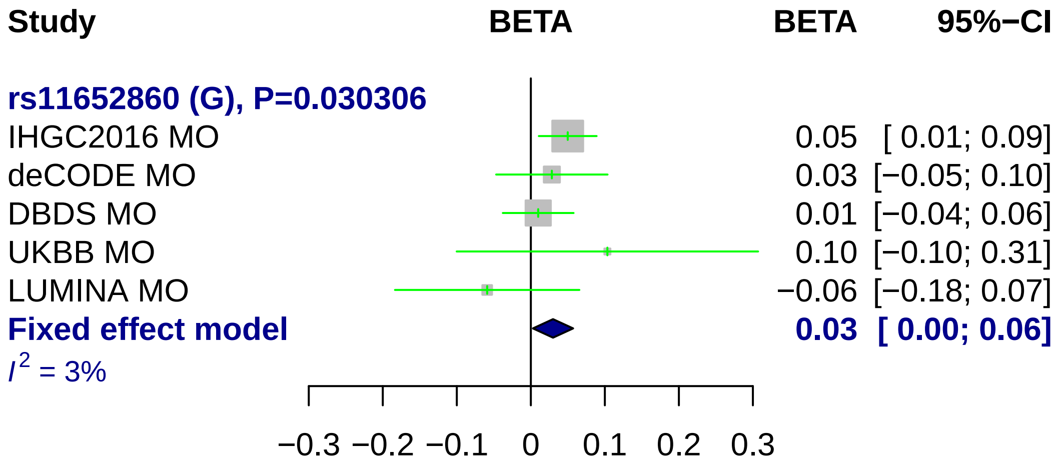
Study
BETA
BETA
95%−CI
rs11652860 (G), P=0.030306
IHGC2016 MO
0.05 [ 0.01; 0.09]
deCODE MO
0.03 [−0.05; 0.10]
DBDS MO
0.01 [−0.04; 0.06]
UKBB MO
0.10 [−0.10; 0.31]
LUMINA MO
−0.06 [−0.18; 0.07]
Fixed effect model
0.03 [ 0.00; 0.06]
I2 = 3%
−0.3
−0.2
−0.1
0
0.1
0.2
0.3

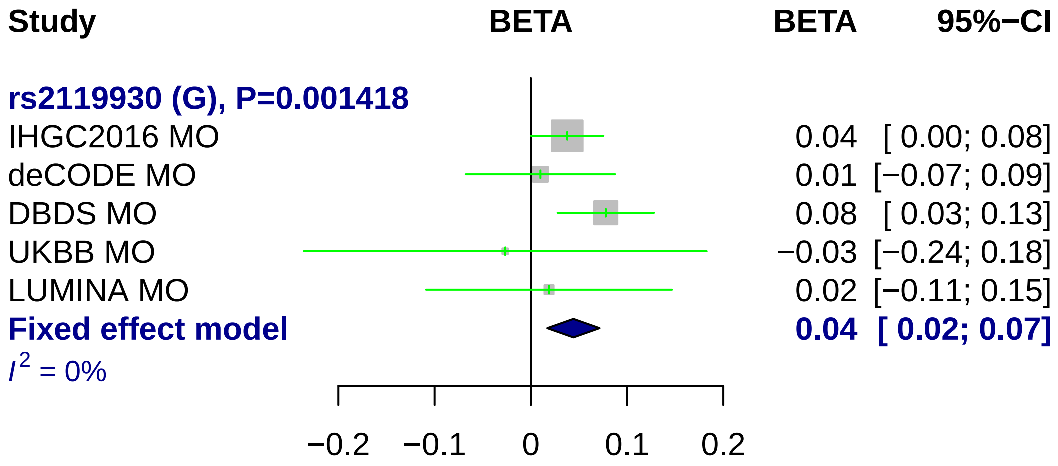
Study
BETA
BETA
95%−CI
rs2119930 (G), P=0.001418
IHGC2016 MO
0.04 [ 0.00; 0.08]
deCODE MO
0.01 [−0.07; 0.09]
DBDS MO
0.08 [ 0.03; 0.13]
UKBB MO
−0.03 [−0.24; 0.18]
LUMINA MO
0.02 [−0.11; 0.15]
Fixed effect model
0.04 [ 0.02; 0.07]
I 2 = 0%
−0.2
−0.1
0
0.1
0.2

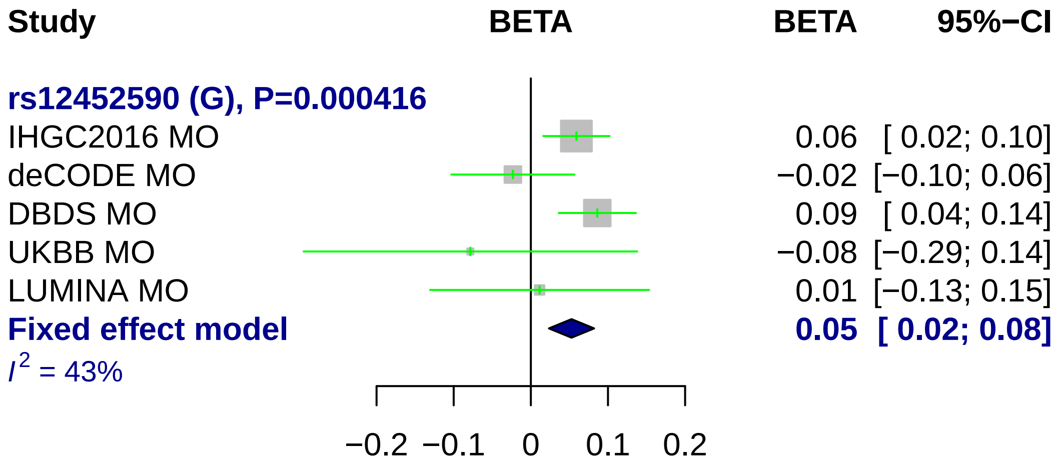
Study
BETA
BETA
95%−CI
rs12452590 (G), P=0.000416
IHGC2016 MO
0.06 [ 0.02; 0.10]
deCODE MO
−0.02 [−0.10; 0.06]
DBDS MO
0.09 [ 0.04; 0.14]
UKBB MO
−0.08 [−0.29; 0.14]
LUMINA MO
0.01 [−0.13; 0.15]
Fixed effect model
0.05 [ 0.02; 0.08]
I2 = 43%
−0.2
−0.1
0
0.1
0.2

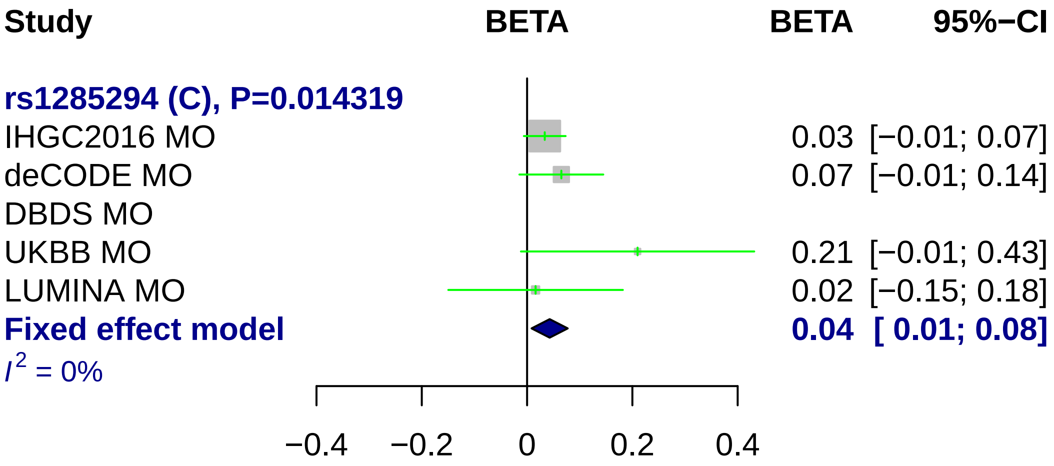
Study
BETA
BETA
95%−CI
rs1285294 (C), P=0.014319
IHGC2016 MO
0.03 [−0.01; 0.07]
deCODE MO
0.07 [−0.01; 0.14]
DBDS MO
UKBB MO
0.21 [−0.01; 0.43]
LUMINA MO
0.02 [−0.15; 0.18]
Fixed effect model
0.04 [ 0.01; 0.08]
I² = 0%
−0.4
−0.2
0
0.2
0.4

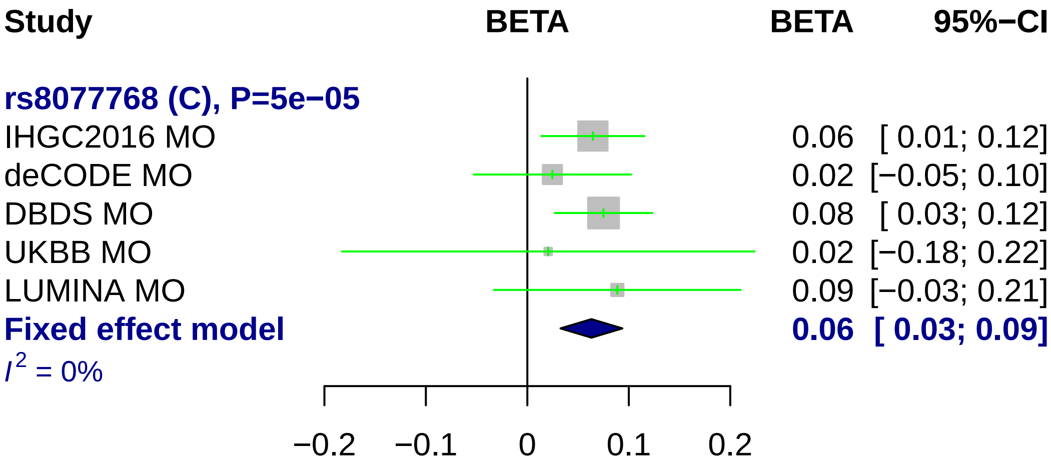
Study
BETA
BETA
95%−CI
rs8077768 (C), P=5e−05
IHGC2016 MO
0.06 [ 0.01; 0.12]
deCODE MO
0.02 [−0.05; 0.10]
DBDS MO
0.08 [ 0.03; 0.12]
UKBB MO
0.02 [−0.18; 0.22]
LUMINA MO
0.09 [−0.03; 0.21]
Fixed effect model
0.06 [ 0.03; 0.09]
I2 = 0%
−0.2
−0.1
0
0.1
0.2

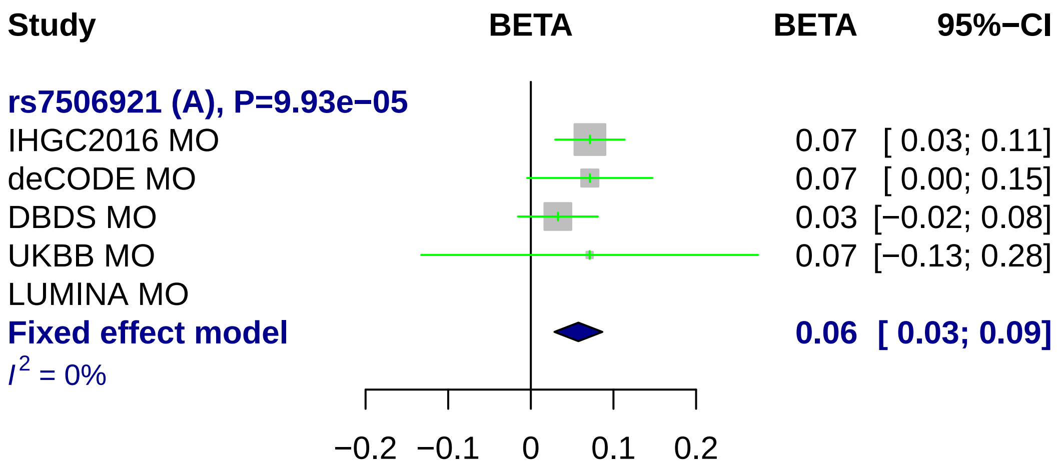
Study
BETA
BETA
95%−CI
rs7506921 (A), P=9.93e−05
IHGC2016 MO 0.07 [ 0.03; 0.11]
deCODE MO 0.07 [ 0.00; 0.15]
DBDS MO 0.03 [−0.02; 0.08]
UKBB MO 0.07 [−0.13; 0.28]
LUMINA MO
Fixed effect model 0.06 [ 0.03; 0.09]
I2 = 0%
−0.2
−0.1
0
0.1
0.2

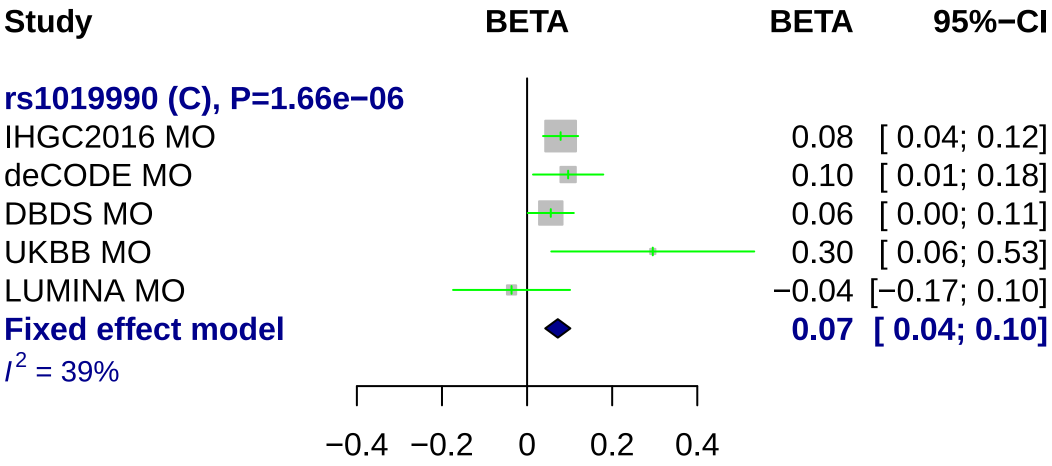
Study
BETA
BETA
95%−CI
rs1019990 (C), P=1.66e−06
IHGC2016 MO
0.08 [ 0.04; 0.12]
deCODE MO
0.10 [ 0.01; 0.18]
DBDS MO
0.06 [ 0.00; 0.11]
UKBB MO
0.30 [ 0.06; 0.53]
LUMINA MO
−0.04 [−0.17; 0.10]
Fixed effect model
0.07 [ 0.04; 0.10]
I^2 = 39%
−0.4
−0.2
0
0.2
0.4

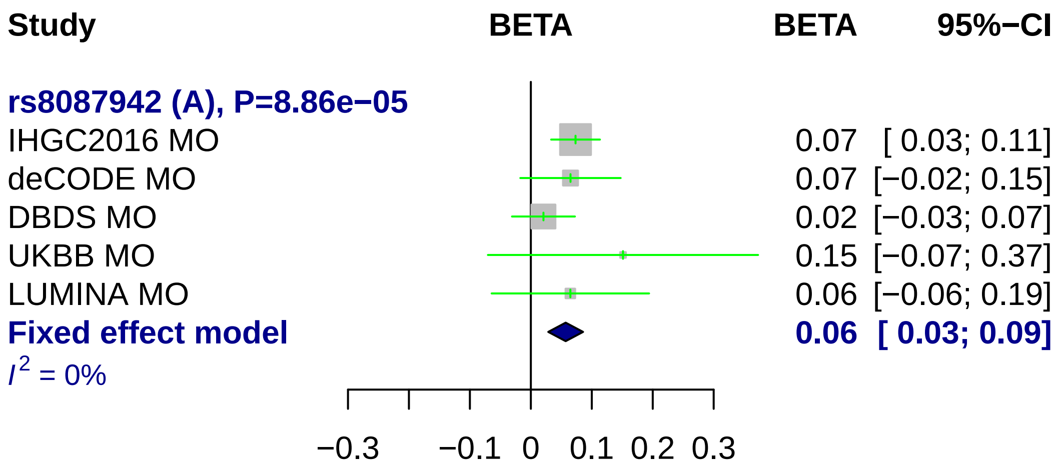
Study
BETA
BETA
95%−CI
rs8087942 (A), P=8.86e−05
IHGC2016 MO
0.07 [ 0.03; 0.11]
deCODE MO
0.07 [−0.02; 0.15]
DBDS MO
0.02 [−0.03; 0.07]
UKBB MO
0.15 [−0.07; 0.37]
LUMINA MO
0.06 [−0.06; 0.19]
Fixed effect model
0.06 [ 0.03; 0.09]
I2 = 0%
−0.3
−0.1
0
0.1
0.2
0.3

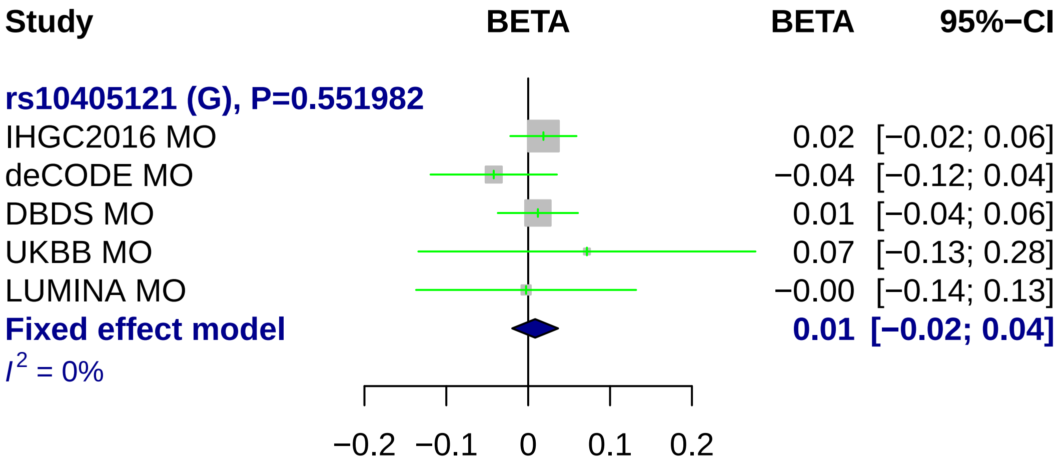

Study
BETA
BETA
95%−CI
rs10405121 (G), P=0.551982
IHGC2016 MO 0.02 [−0.02; 0.06]
deCODE MO −0.04 [−0.12; 0.04]
DBDS MO 0.01 [−0.04; 0.06]
UKBB MO 0.07 [−0.13; 0.28]
LUMINA MO −0.00 [−0.14; 0.13]
Fixed effect model 0.01 [−0.02; 0.04]
I^2 = 0%
−0.2 −0.1 0 0.1 0.2

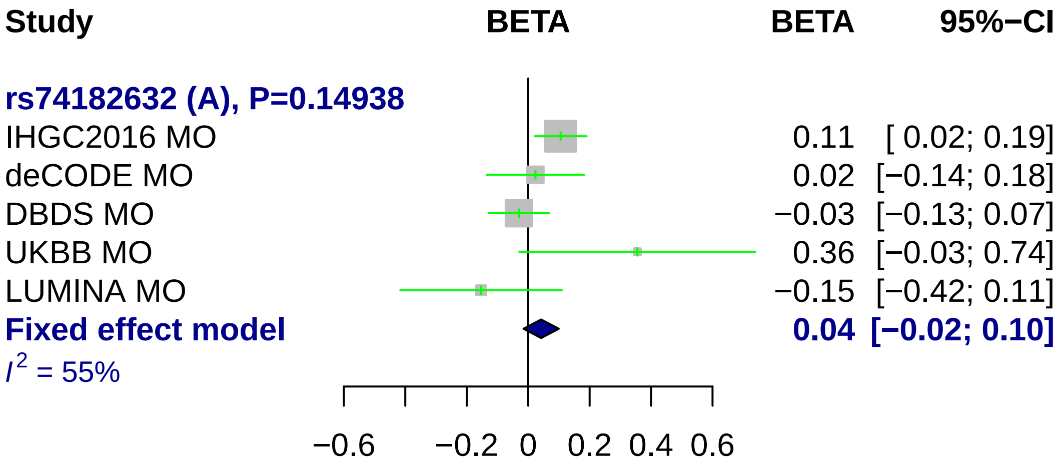
Study
BETA
BETA
95%−CI
rs74182632 (A), P=0.14938
IHGC2016 MO
0.11
[ 0.02; 0.19]
deCODE MO
0.02
[−0.14; 0.18]
DBDS MO
−0.03
[−0.13; 0.07]
UKBB MO
0.36
[−0.03; 0.74]
LUMINA MO
−0.15
[−0.42; 0.11]
Fixed effect model
0.04
[−0.02; 0.10]
I2 = 55%
−0.6
−0.2
0
0.2
0.4
0.6

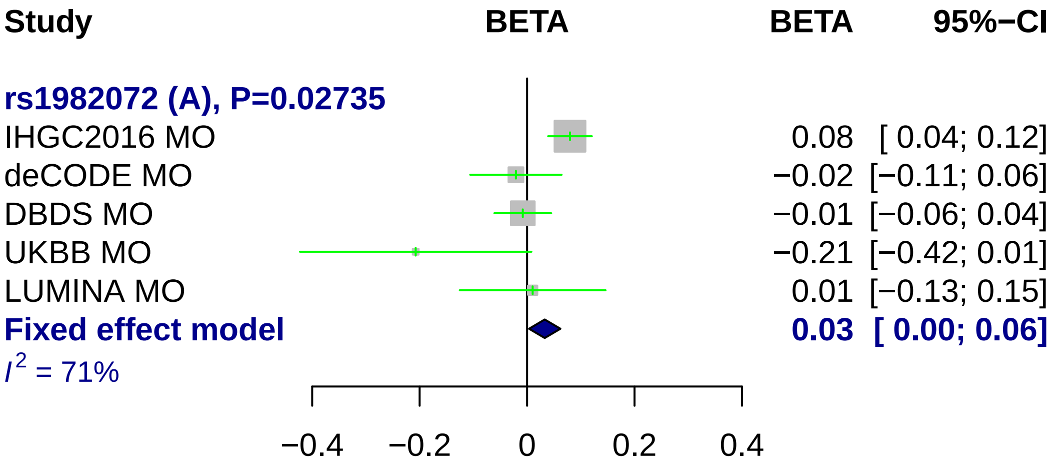
Study
BETA
BETA
95%−CI
rs1982072 (A), P=0.02735
IHGC2016 MO
0.08 [ 0.04; 0.12]
deCODE MO
−0.02 [−0.11; 0.06]
DBDS MO
−0.01 [−0.06; 0.04]
UKBB MO
−0.21 [−0.42; 0.01]
LUMINA MO
0.01 [−0.13; 0.15]
Fixed effect model
0.03 [ 0.00; 0.06]
I2 = 71%
−0.4
−0.2
0
0.2
0.4

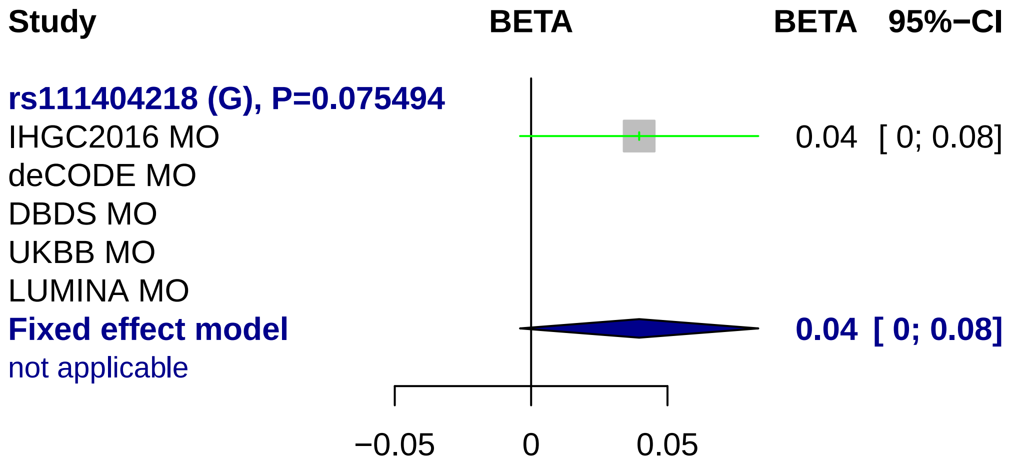
Study
BETA
BETA 95%−CI
rs111404218 (G), P=0.075494
IHGC2016 MO
0.04 [ 0; 0.08]
deCODE MO
DBDS MO
UKBB MO
LUMINA MO
Fixed effect model
0.04 [ 0; 0.08]
not applicable
−0.05
0
0.05

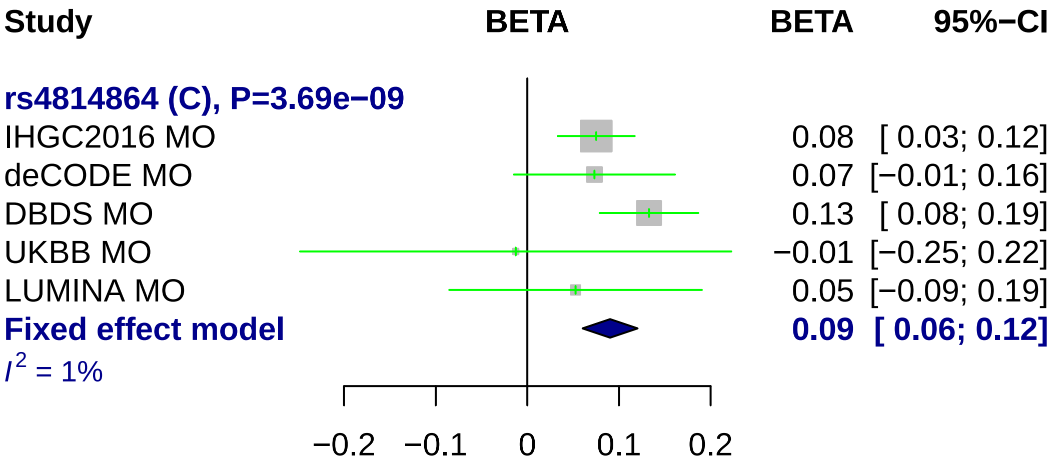
Study
BETA
BETA
95%−CI
rs4814864 (C), P=3.69e−09
IHGC2016 MO
0.08
[ 0.03; 0.12]
deCODE MO
0.07
[−0.01; 0.16]
DBDS MO
0.13
[ 0.08; 0.19]
UKBB MO
−0.01
[−0.25; 0.22]
LUMINA MO
0.05
[−0.09; 0.19]
Fixed effect model
0.09
[ 0.06; 0.12]
I2 = 1%
−0.2
−0.1
0
0.1
0.2

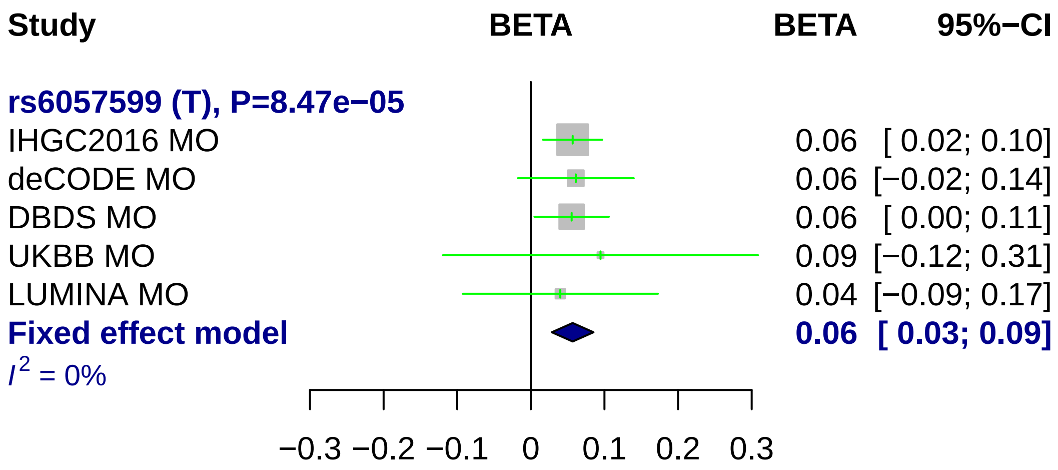

Study
BETA
BETA
95%−CI
rs6057599 (T), P=8.47e−05
IHGC2016 MO 0.06 [ 0.02; 0.10]
deCODE MO 0.06 [−0.02; 0.14]
DBDS MO 0.06 [ 0.00; 0.11]
UKBB MO 0.09 [−0.12; 0.31]
LUMINA MO 0.04 [−0.09; 0.17]
Fixed effect model 0.06 [ 0.03; 0.09]
I^2 = 0%
−0.3 −0.2 −0.1 0 0.1 0.2 0.3

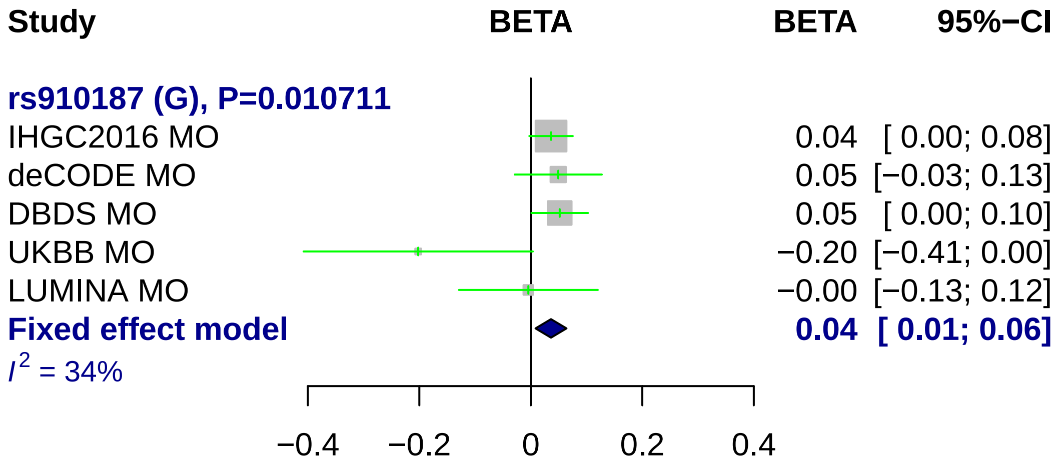
Study
BETA
BETA
95%−CI
rs910187 (G), P=0.010711
IHGC2016 MO 0.04 [ 0.00; 0.08]
deCODE MO 0.05 [−0.03; 0.13]
DBDS MO 0.05 [ 0.00; 0.10]
UKBB MO −0.20 [−0.41; 0.00]
LUMINA MO −0.00 [−0.13; 0.12]
Fixed effect model 0.04 [ 0.01; 0.06]
I2 = 34%
−0.4
−0.2
0
0.2
0.4

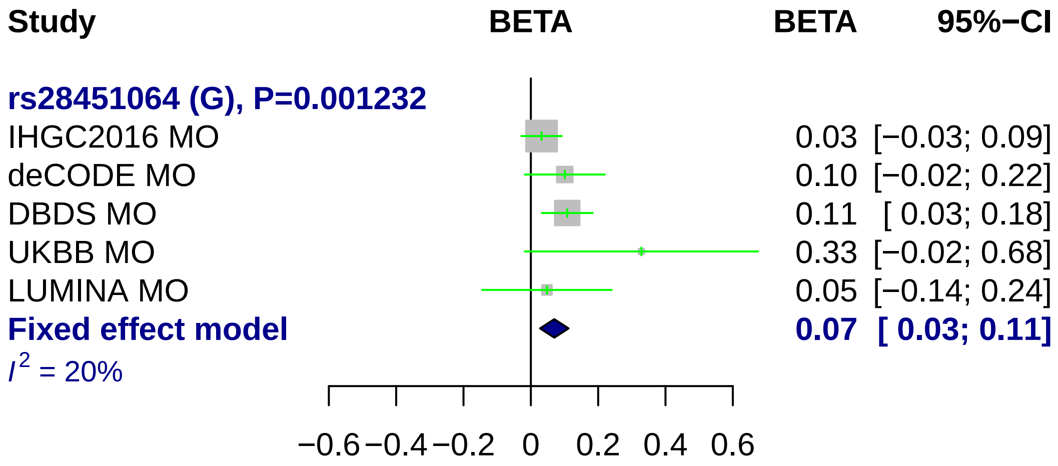
Study
BETA
BETA
95%−CI
rs28451064 (G), P=0.001232
IHGC2016 MO
0.03 [−0.03; 0.09]
deCODE MO
0.10 [−0.02; 0.22]
DBDS MO
0.11 [ 0.03; 0.18]
UKBB MO
0.33 [−0.02; 0.68]
LUMINA MO
0.05 [−0.14; 0.24]
Fixed effect model
0.07 [ 0.03; 0.11]
I2 = 20%
−0.6 −0.4 −0.2 0 0.2 0.4 0.6

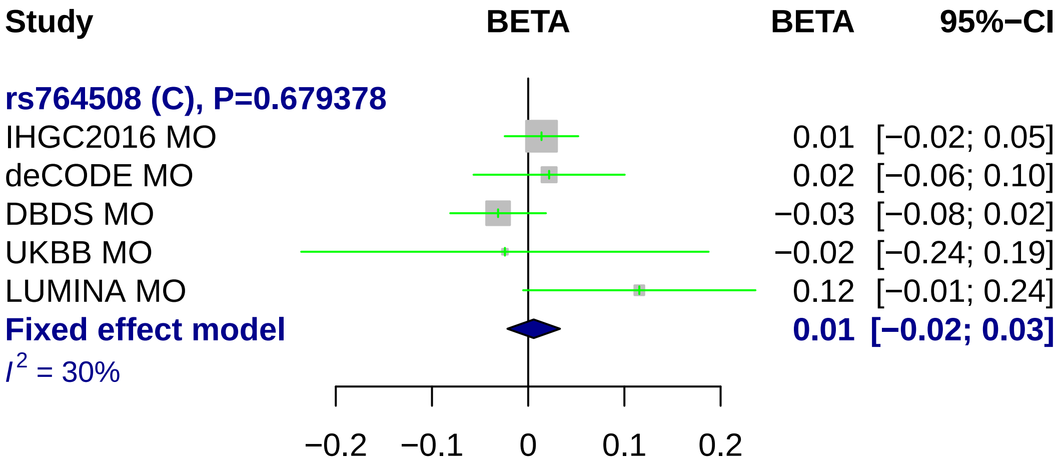
Study
BETA
BETA
95%−CI
rs764508 (C), P=0.679378
IHGC2016 MO
0.01 [−0.02; 0.05]
deCODE MO
0.02 [−0.06; 0.10]
DBDS MO
−0.03 [−0.08; 0.02]
UKBB MO
−0.02 [−0.24; 0.19]
LUMINA MO
0.12 [−0.01; 0.24]
Fixed effect model
0.01 [−0.02; 0.03]
I2 = 30%
−0.2
−0.1
0
0.1
0.2

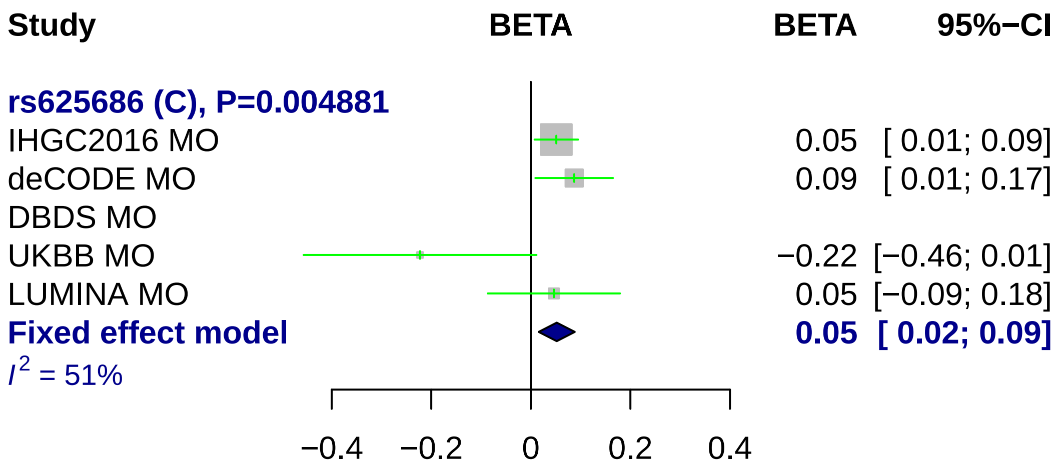
Study
BETA
BETA
95%−CI
rs625686 (C), P=0.004881
IHGC2016 MO
0.05 [ 0.01; 0.09]
deCODE MO
0.09 [ 0.01; 0.17]
DBDS MO
UKBB MO
−0.22 [−0.46; 0.01]
LUMINA MO
0.05 [−0.09; 0.18]
Fixed effect model
0.05 [ 0.02; 0.09]
I2 = 51%
−0.4
−0.2
0
0.2
0.4

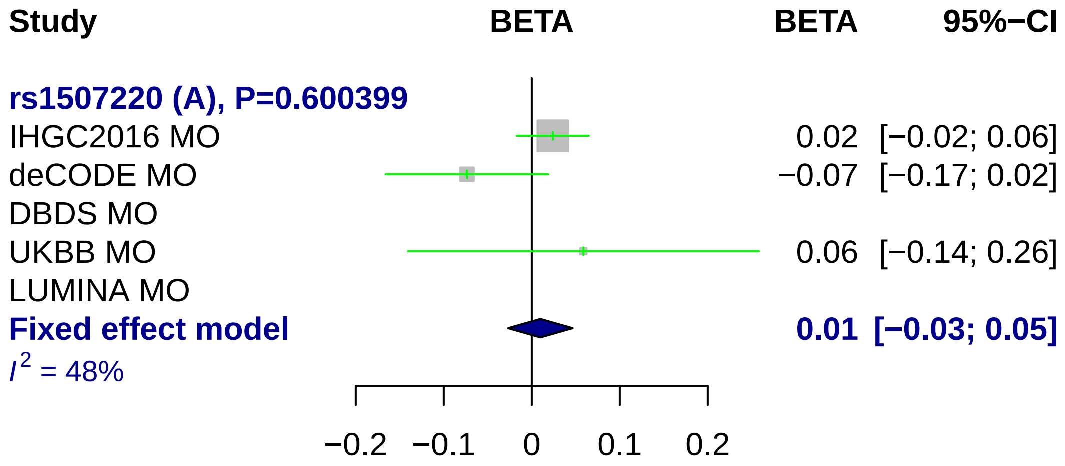

Study
BETA
BETA
95%−CI
rs1507220 (A), P=0.600399
IHGC2016 MO
0.02
[−0.02; 0.06]
deCODE MO
−0.07
[−0.17; 0.02]
DBDS MO
UKBB MO
0.06
[−0.14; 0.26]
LUMINA MO
Fixed effect model
0.01
[−0.03; 0.05]
I2 = 48%
−0.2
−0.1
0
0.1
0.2

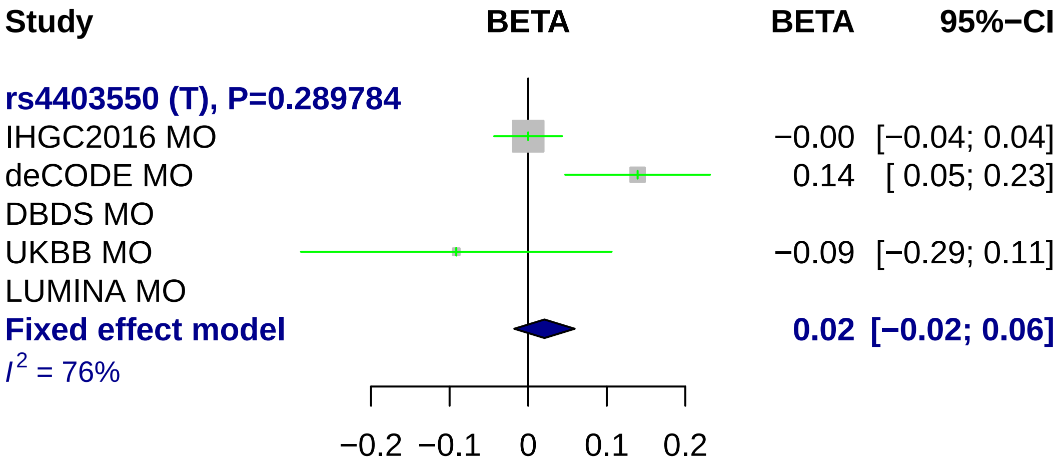
Study
BETA
BETA
95%−CI
rs4403550 (T), P=0.289784
IHGC2016 MO
−0.00
[−0.04; 0.04]
deCODE MO
0.14
[ 0.05; 0.23]
DBDS MO
UKBB MO
−0.09
[−0.29; 0.11]
LUMINA MO
Fixed effect model
0.02
[−0.02; 0.06]
I2 = 76%
−0.2
−0.1
0
0.1
0.2
