## Supplementary Figure 9. for "Genome-wide analysis of 102,084 migraine cases identifies 123 risk loci and subtype-specific risk alleles"

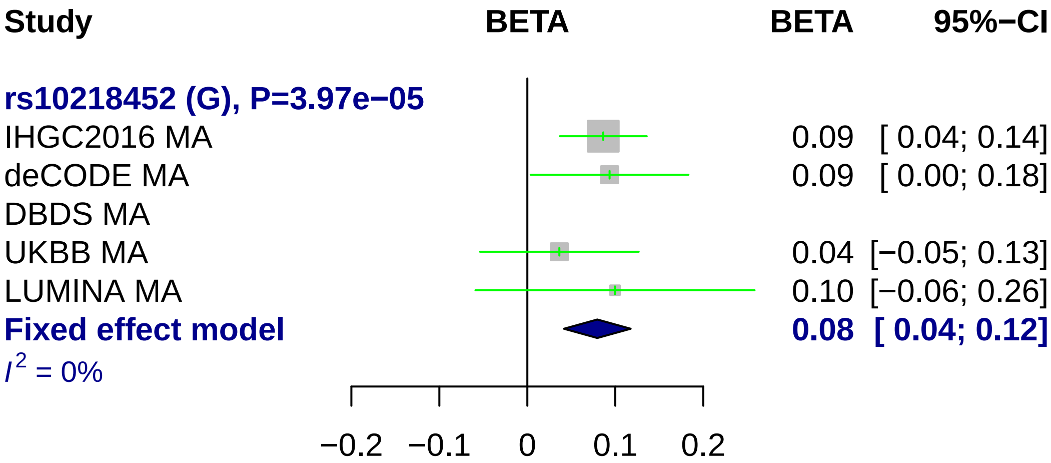
Study
BETA
BETA
95%−CI
rs10218452 (G), P=3.97e−05
IHGC2016 MA
0.09 [ 0.04; 0.14]
deCODE MA
0.09 [ 0.00; 0.18]
DBDS MA
UKBB MA
0.04 [−0.05; 0.13]
LUMINA MA
0.10 [−0.06; 0.26]
Fixed effect model
0.08 [ 0.04; 0.12]
I^2 = 0%
−0.2
−0.1
0
0.1
0.2

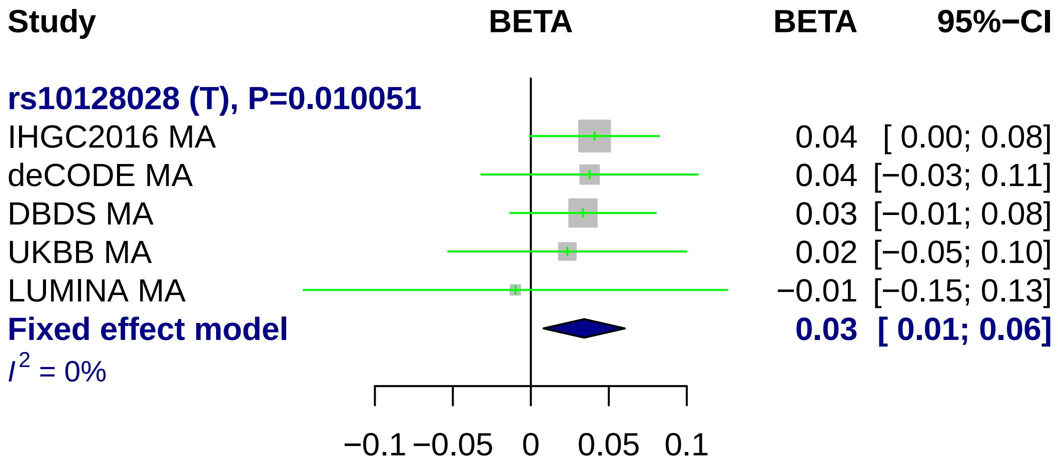

Study
BETA
BETA
95%−CI
rs10128028 (T), P=0.010051
IHGC2016 MA 0.04 [ 0.00; 0.08]
deCODE MA 0.04 [−0.03; 0.11]
DBDS MA 0.03 [−0.01; 0.08]
UKBB MA 0.02 [−0.05; 0.10]
LUMINA MA −0.01 [−0.15; 0.13]
Fixed effect model 0.03 [ 0.01; 0.06]
$I^2$ = 0%
−0.1
−0.05
0
0.05
0.1

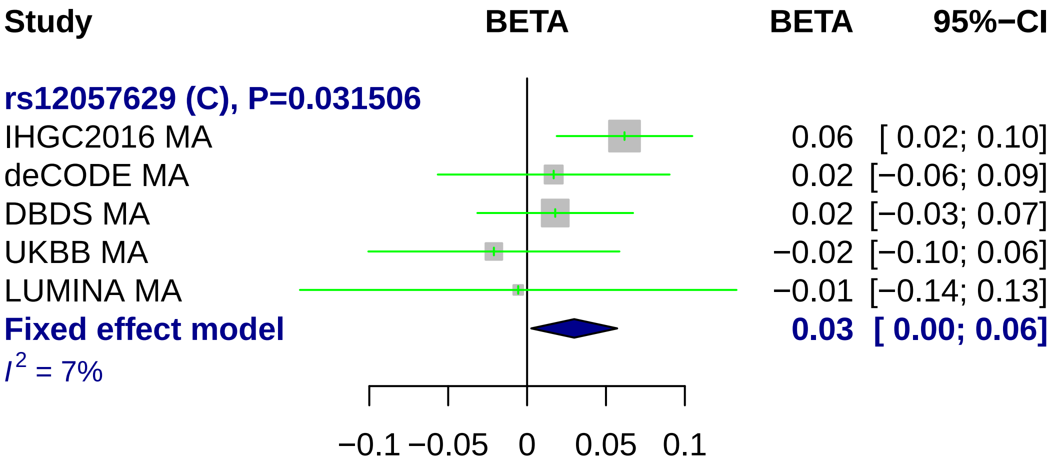
Study
BETA
BETA
95%−CI
rs12057629 (C), P=0.031506
IHGC2016 MA
0.06 [ 0.02; 0.10]
deCODE MA
0.02 [−0.06; 0.09]
DBDS MA
0.02 [−0.03; 0.07]
UKBB MA
−0.02 [−0.10; 0.06]
LUMINA MA
−0.01 [−0.14; 0.13]
Fixed effect model
0.03 [ 0.00; 0.06]
I2 = 7%
−0.1
−0.05
0
0.05
0.1

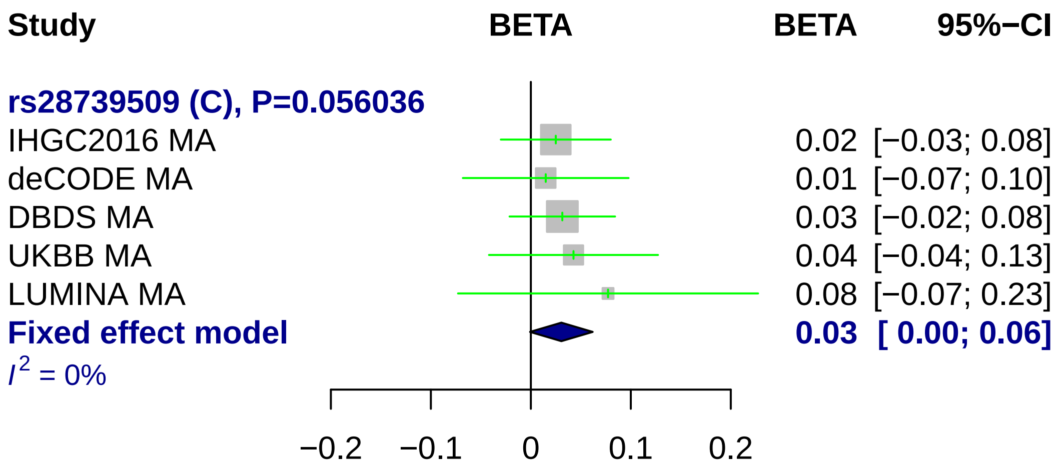
Study
BETA
BETA
95%−CI
rs28739509 (C), P=0.056036
IHGC2016 MA
0.02 [−0.03; 0.08]
deCODE MA
0.01 [−0.07; 0.10]
DBDS MA
0.03 [−0.02; 0.08]
UKBB MA
0.04 [−0.04; 0.13]
LUMINA MA
0.08 [−0.07; 0.23]
Fixed effect model
0.03 [ 0.00; 0.06]
I2 = 0%
−0.2
−0.1
0
0.1
0.2

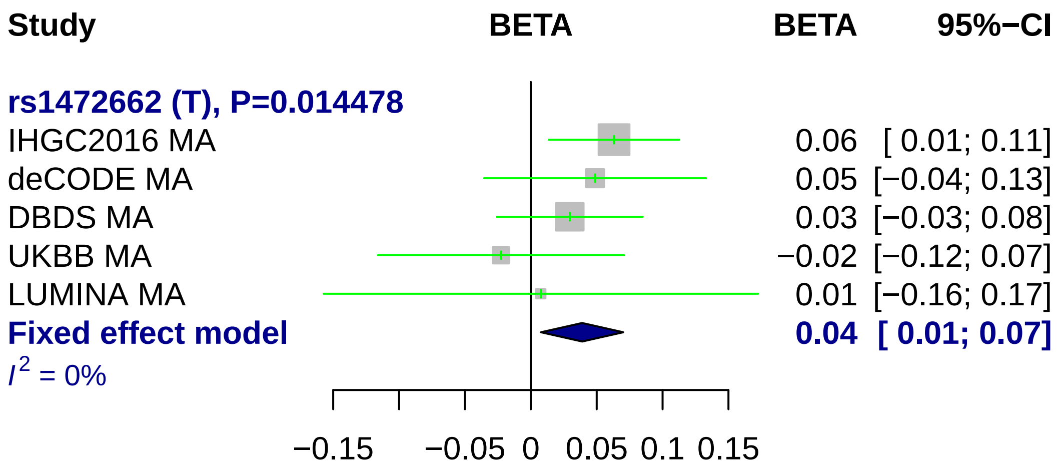
Study
BETA
BETA
95%−CI
rs1472662 (T), P=0.014478
IHGC2016 MA 0.06 [ 0.01; 0.11]
deCODE MA 0.05 [−0.04; 0.13]
DBDS MA 0.03 [−0.03; 0.08]
UKBB MA −0.02 [−0.12; 0.07]
LUMINA MA 0.01 [−0.16; 0.17]
Fixed effect model 0.04 [ 0.01; 0.07]
I2 = 0%
−0.15
−0.05
0
0.05
0.1
0.15

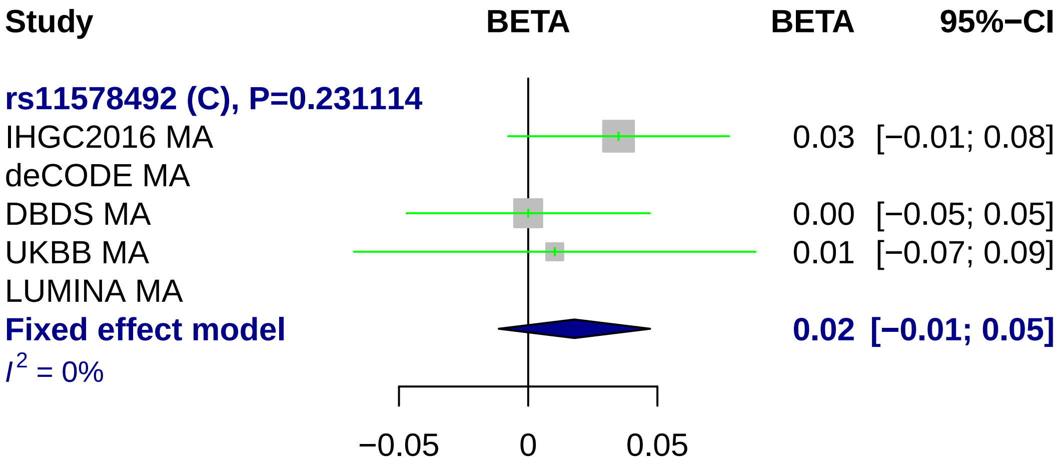
Study
BETA
BETA
95%−CI
rs11578492 (C), P=0.231114
IHGC2016 MA
0.03 [−0.01; 0.08]
deCODE MA
DBDS MA
0.00 [−0.05; 0.05]
UKBB MA
0.01 [−0.07; 0.09]
LUMINA MA
Fixed effect model
0.02 [−0.01; 0.05]
I2 = 0%
−0.05
0
0.05

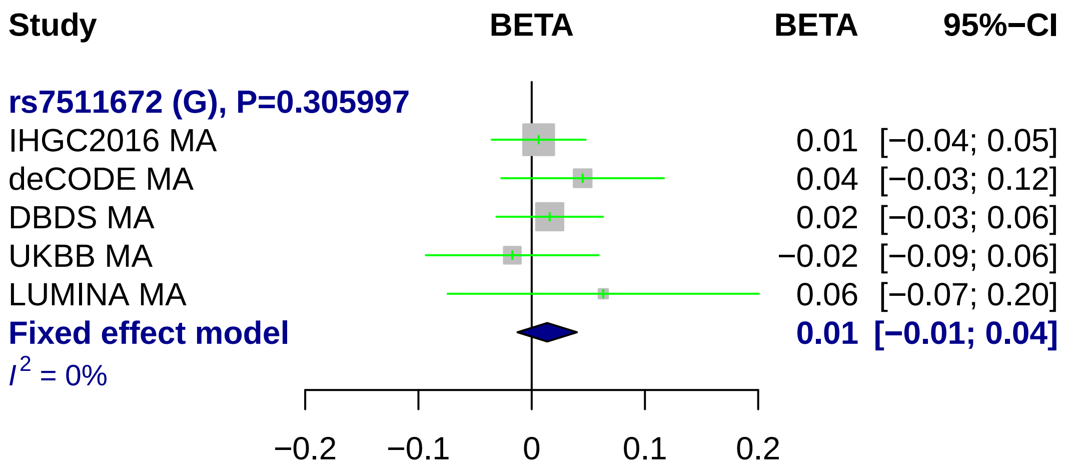
Study
BETA
BETA
95%−CI
rs7511672 (G), P=0.305997
IHGC2016 MA
0.01 [−0.04; 0.05]
deCODE MA
0.04 [−0.03; 0.12]
DBDS MA
0.02 [−0.03; 0.06]
UKBB MA
−0.02 [−0.09; 0.06]
LUMINA MA
0.06 [−0.07; 0.20]
Fixed effect model
0.01 [−0.01; 0.04]
I2 = 0%
−0.2
−0.1
0
0.1
0.2

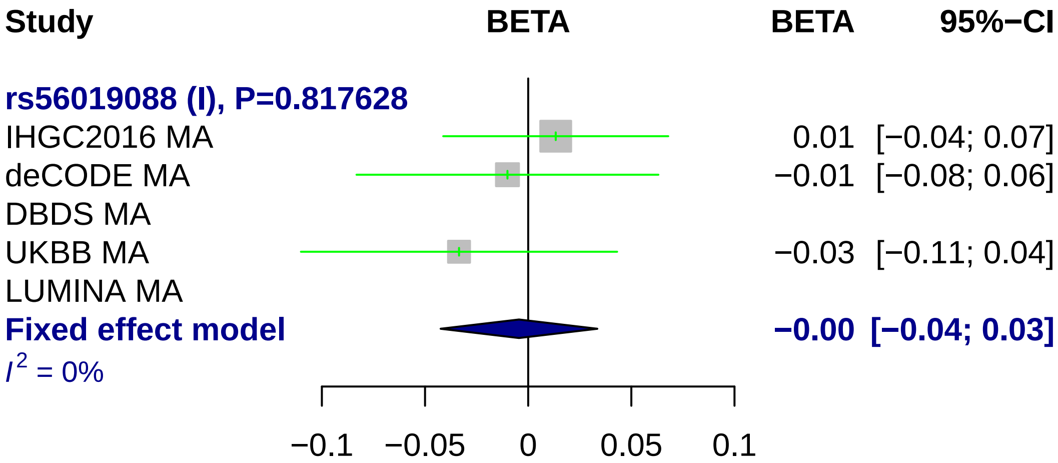
Study
BETA
BETA
95%−CI
rs56019088 (I), P=0.817628
IHGC2016 MA
0.01 [−0.04; 0.07]
deCODE MA
−0.01 [−0.08; 0.06]
DBDS MA
UKBB MA
−0.03 [−0.11; 0.04]
LUMINA MA
Fixed effect model
−0.00 [−0.04; 0.03]
I^2 = 0%
−0.1
−0.05
0
0.05
0.1

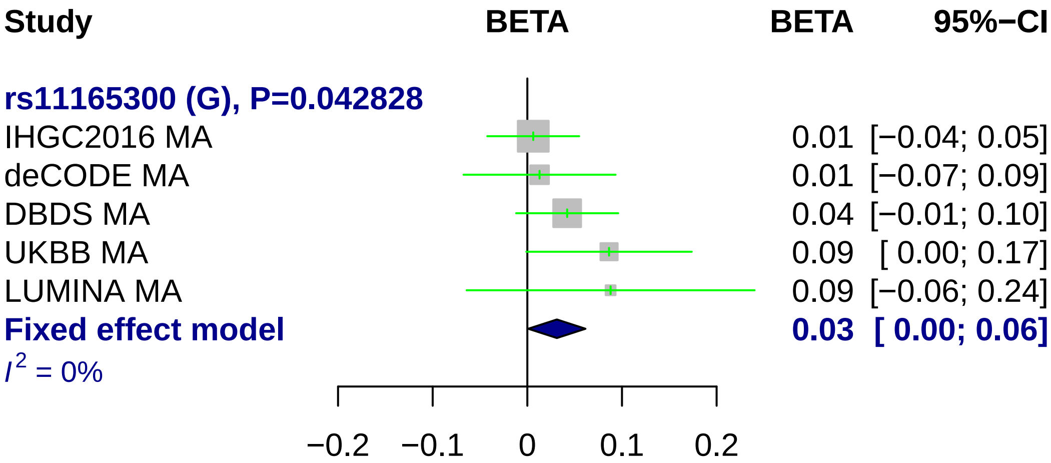

Study
BETA
BETA
95%−CI
rs11165300 (G), P=0.042828
IHGC2016 MA
0.01 [−0.04; 0.05]
deCODE MA
0.01 [−0.07; 0.09]
DBDS MA
0.04 [−0.01; 0.10]
UKBB MA
0.09 [ 0.00; 0.17]
LUMINA MA
0.09 [−0.06; 0.24]
Fixed effect model
0.03 [ 0.00; 0.06]
I2 = 0%
−0.2
−0.1
0
0.1
0.2

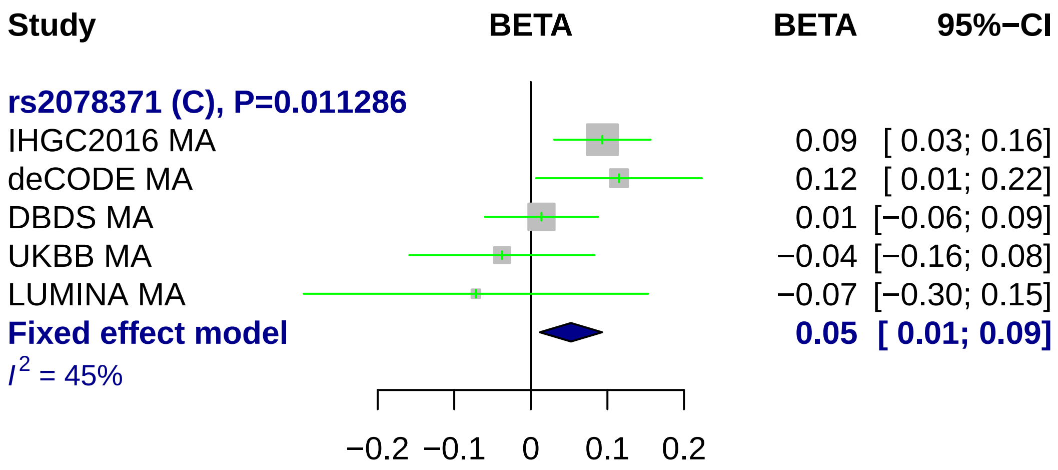
Study
BETA
BETA
95%−CI
rs2078371 (C), P=0.011286
IHGC2016 MA
0.09
[ 0.03; 0.16]
deCODE MA
0.12
[ 0.01; 0.22]
DBDS MA
0.01
[−0.06; 0.09]
UKBB MA
−0.04
[−0.16; 0.08]
LUMINA MA
−0.07
[−0.30; 0.15]
Fixed effect model
0.05
[ 0.01; 0.09]
I^2 = 45%
−0.2
−0.1
0
0.1
0.2

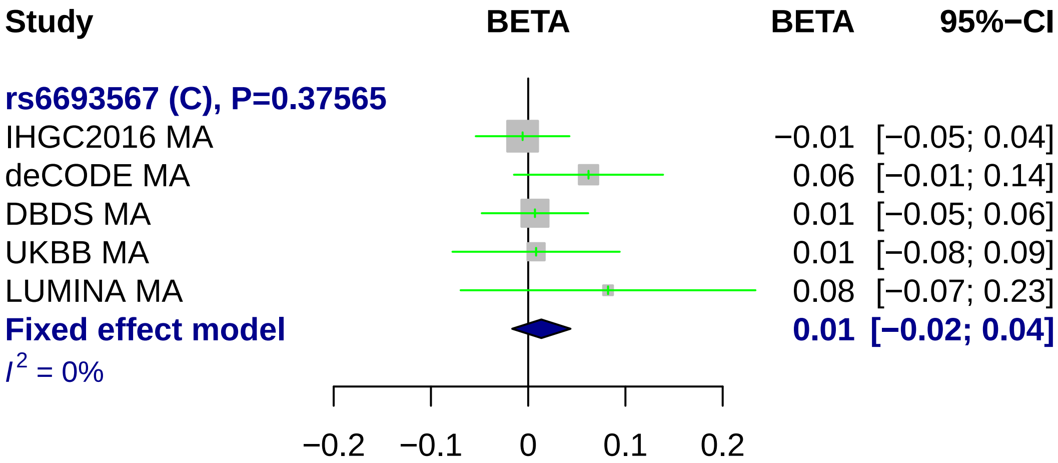
Study
BETA
BETA
95%−CI
rs6693567 (C), P=0.37565
IHGC2016 MA
−0.01 [−0.05; 0.04]
deCODE MA
0.06 [−0.01; 0.14]
DBDS MA
0.01 [−0.05; 0.06]
UKBB MA
0.01 [−0.08; 0.09]
LUMINA MA
0.08 [−0.07; 0.23]
Fixed effect model
0.01 [−0.02; 0.04]
I^2 = 0%
−0.2
−0.1
0
0.1
0.2

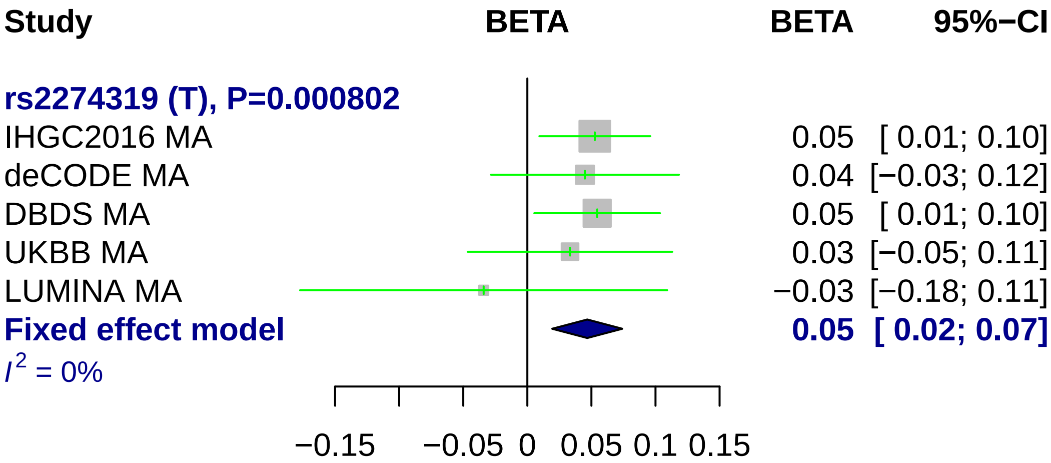
Study
BETA
BETA
95%−CI
rs2274319 (T), P=0.000802
IHGC2016 MA 0.05 [ 0.01; 0.10]
deCODE MA 0.04 [−0.03; 0.12]
DBDS MA 0.05 [ 0.01; 0.10]
UKBB MA 0.03 [−0.05; 0.11]
LUMINA MA −0.03 [−0.18; 0.11]
Fixed effect model 0.05 [ 0.02; 0.07]
I² = 0%
−0.15
−0.05
0
0.05
0.1
0.15

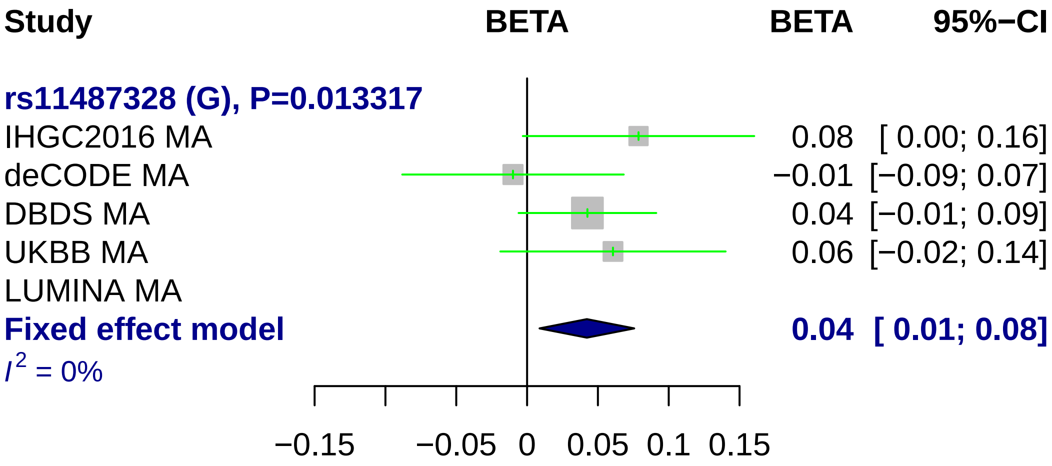
Study
BETA
BETA
95%−CI
rs11487328 (G), P=0.013317
IHGC2016 MA
0.08 [ 0.00; 0.16]
deCODE MA
−0.01 [−0.09; 0.07]
DBDS MA
0.04 [−0.01; 0.09]
UKBB MA
0.06 [−0.02; 0.14]
LUMINA MA
Fixed effect model
0.04 [ 0.01; 0.08]
I2 = 0%
−0.15
−0.05
0
0.05
0.1
0.15

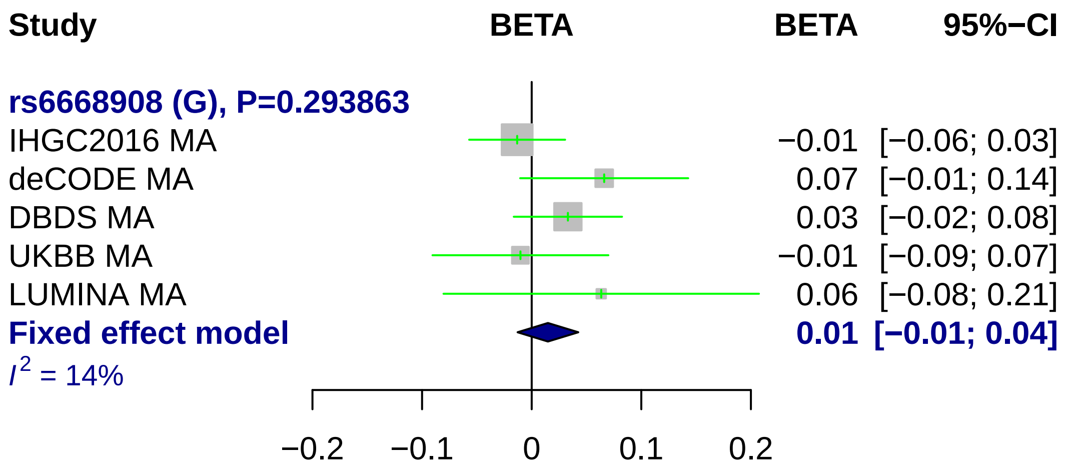
Study
BETA
BETA
95%−CI
rs6668908 (G), P=0.293863
IHGC2016 MA
−0.01 [−0.06; 0.03]
deCODE MA
0.07 [−0.01; 0.14]
DBDS MA
0.03 [−0.02; 0.08]
UKBB MA
−0.01 [−0.09; 0.07]
LUMINA MA
0.06 [−0.08; 0.21]
Fixed effect model
0.01 [−0.01; 0.04]
I2 = 14%
−0.2
−0.1
0
0.1
0.2

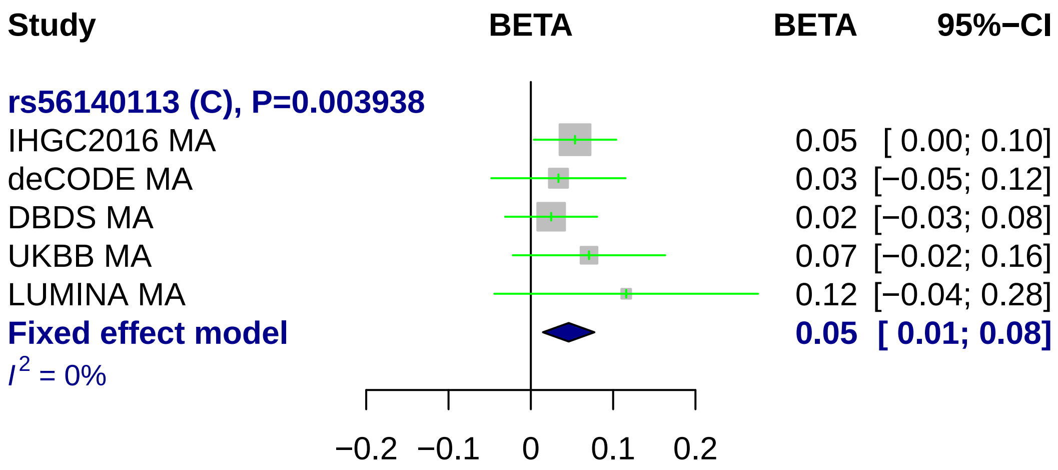
Study
BETA
BETA
95%−CI
rs56140113 (C), P=0.003938
IHGC2016 MA
0.05 [ 0.00; 0.10]
deCODE MA
0.03 [−0.05; 0.12]
DBDS MA
0.02 [−0.03; 0.08]
UKBB MA
0.07 [−0.02; 0.16]
LUMINA MA
0.12 [−0.04; 0.28]
Fixed effect model
0.05 [ 0.01; 0.08]
I2 = 0%
−0.2
−0.1
0
0.1
0.2

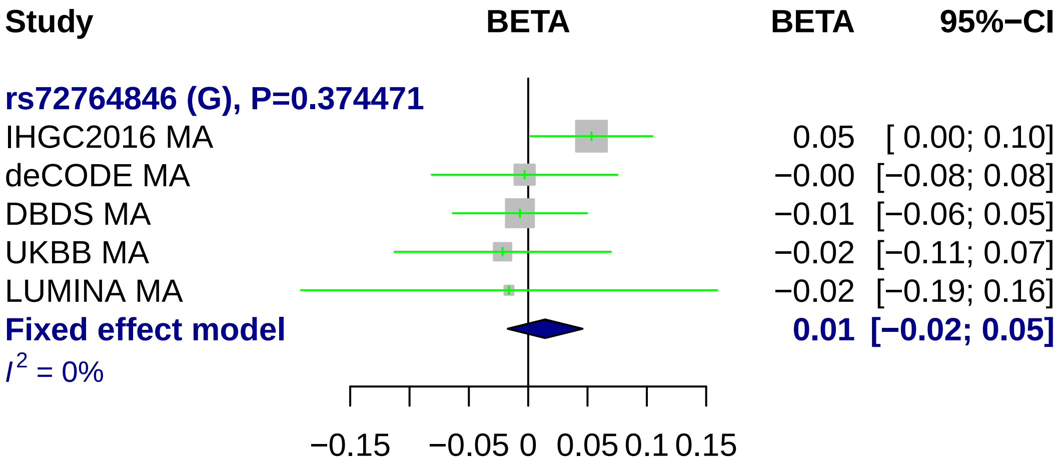
Study
BETA
BETA
95%−CI
rs72764846 (G), P=0.374471
IHGC2016 MA
deCODE MA
DBDS MA
UKBB MA
LUMINA MA
Fixed effect model
$I^2$ = 0%
0.05 [ 0.00; 0.10]
−0.00 [−0.08; 0.08]
−0.01 [−0.06; 0.05]
−0.02 [−0.11; 0.07]
−0.02 [−0.19; 0.16]
0.01 [−0.02; 0.05]
−0.15
−0.05
0
0.05
0.1
0.15

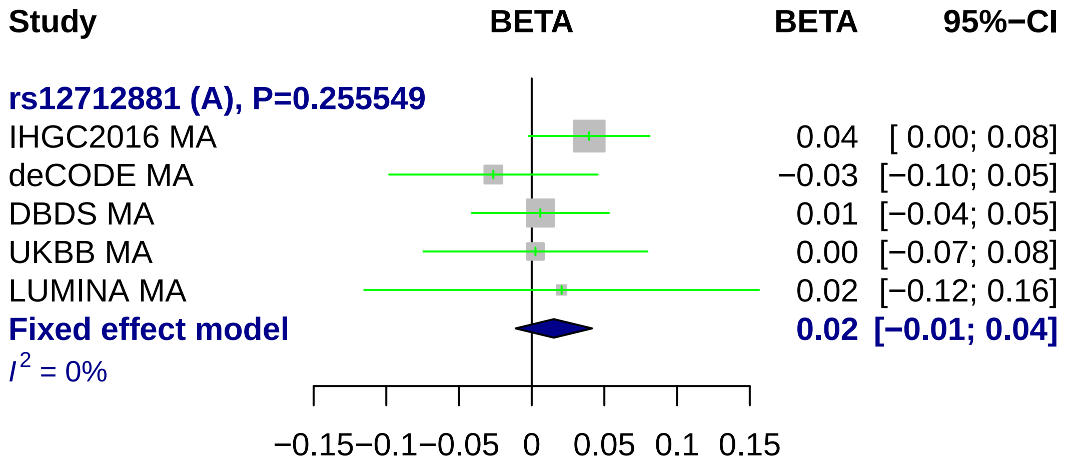
Study
BETA
BETA
95%−CI
rs12712881 (A), P=0.255549
IHGC2016 MA
0.04 [ 0.00; 0.08]
deCODE MA
−0.03 [−0.10; 0.05]
DBDS MA
0.01 [−0.04; 0.05]
UKBB MA
0.00 [−0.07; 0.08]
LUMINA MA
0.02 [−0.12; 0.16]
Fixed effect model
0.02 [−0.01; 0.04]
I 2 = 0%
−0.15
−0.1
−0.05
0
0.05
0.1
0.15

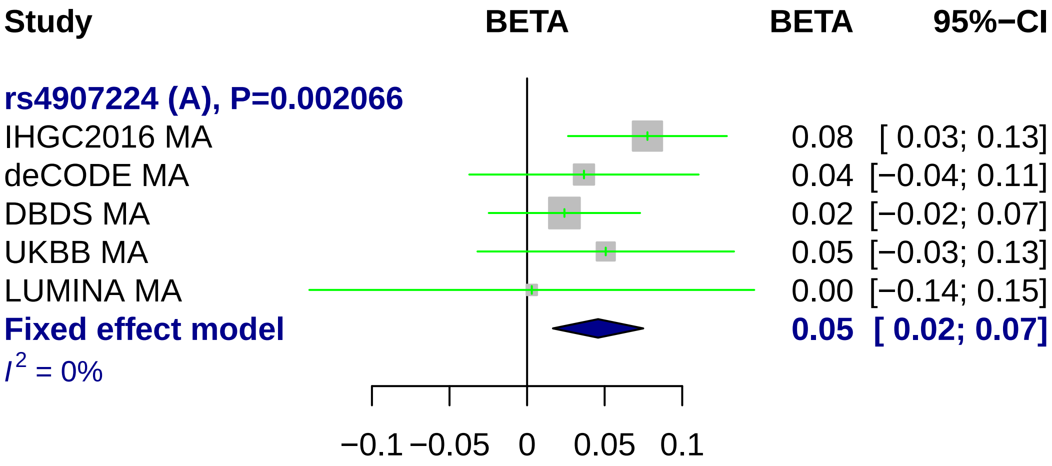
Study
BETA
BETA
95%−CI
rs4907224 (A), P=0.002066
IHGC2016 MA
0.08 [ 0.03; 0.13]
deCODE MA
0.04 [−0.04; 0.11]
DBDS MA
0.02 [−0.02; 0.07]
UKBB MA
0.05 [−0.03; 0.13]
LUMINA MA
0.00 [−0.14; 0.15]
Fixed effect model
0.05 [ 0.02; 0.07]
I2 = 0%
−0.1
−0.05
0
0.05
0.1

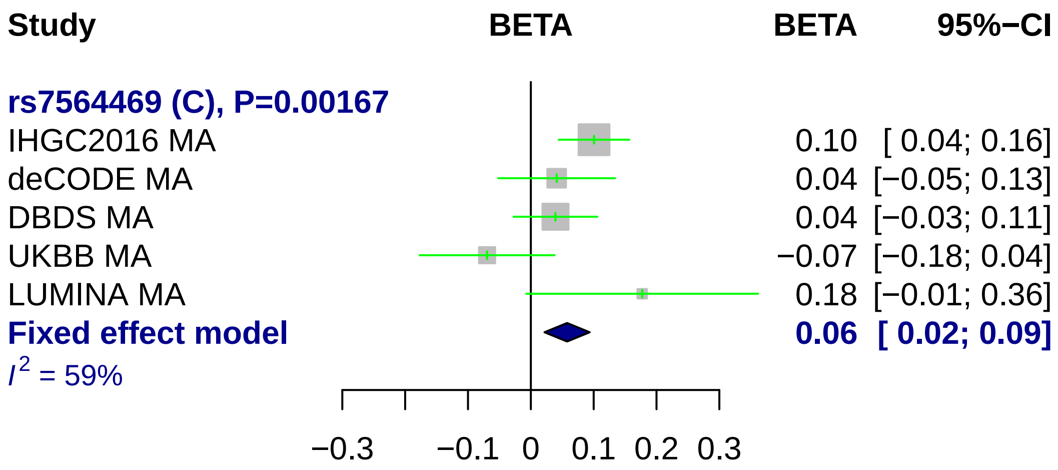
Study
BETA
BETA
95%−CI
rs7564469 (C), P=0.00167
IHGC2016 MA
0.10 [ 0.04; 0.16]
deCODE MA
0.04 [−0.05; 0.13]
DBDS MA
0.04 [−0.03; 0.11]
UKBB MA
−0.07 [−0.18; 0.04]
LUMINA MA
0.18 [−0.01; 0.36]
Fixed effect model
0.06 [ 0.02; 0.09]
$I^2$ = 59%
−0.3
−0.1
0
0.1
0.2
0.3

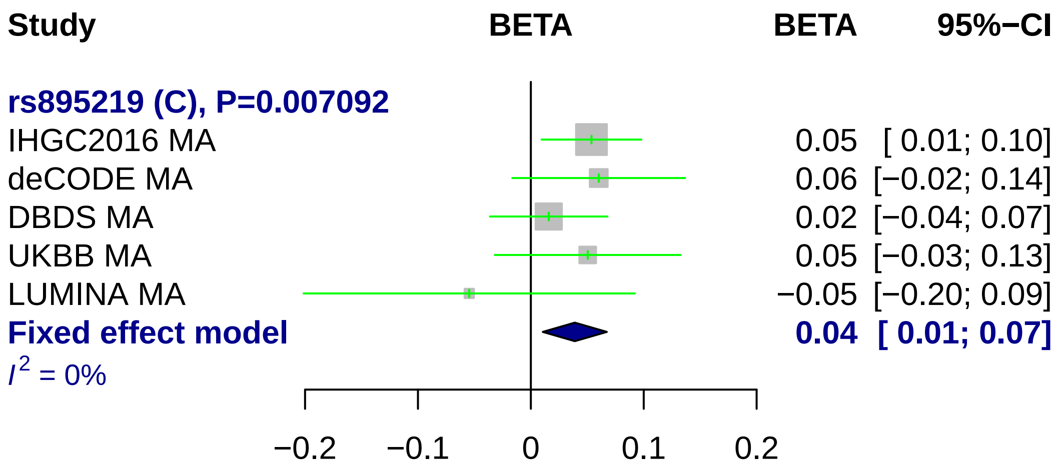
Study
BETA
BETA
95%−CI
rs895219 (C), P=0.007092
IHGC2016 MA
0.05 [ 0.01; 0.10]
deCODE MA
0.06 [−0.02; 0.14]
DBDS MA
0.02 [−0.04; 0.07]
UKBB MA
0.05 [−0.03; 0.13]
LUMINA MA
−0.05 [−0.20; 0.09]
Fixed effect model
0.04 [ 0.01; 0.07]
I2 = 0%
−0.2
−0.1
0
0.1
0.2

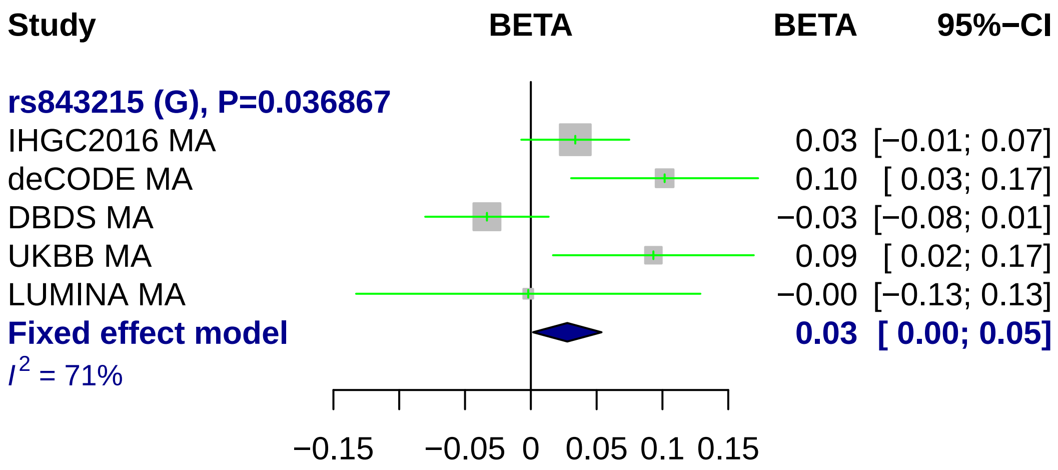
Study
BETA
BETA
95%−CI
rs843215 (G), P=0.036867
IHGC2016 MA
0.03 [−0.01; 0.07]
deCODE MA
0.10 [ 0.03; 0.17]
DBDS MA
−0.03 [−0.08; 0.01]
UKBB MA
0.09 [ 0.02; 0.17]
LUMINA MA
−0.00 [−0.13; 0.13]
Fixed effect model
0.03 [ 0.00; 0.05]
I² = 71%
−0.15
−0.05
0
0.05
0.1
0.15

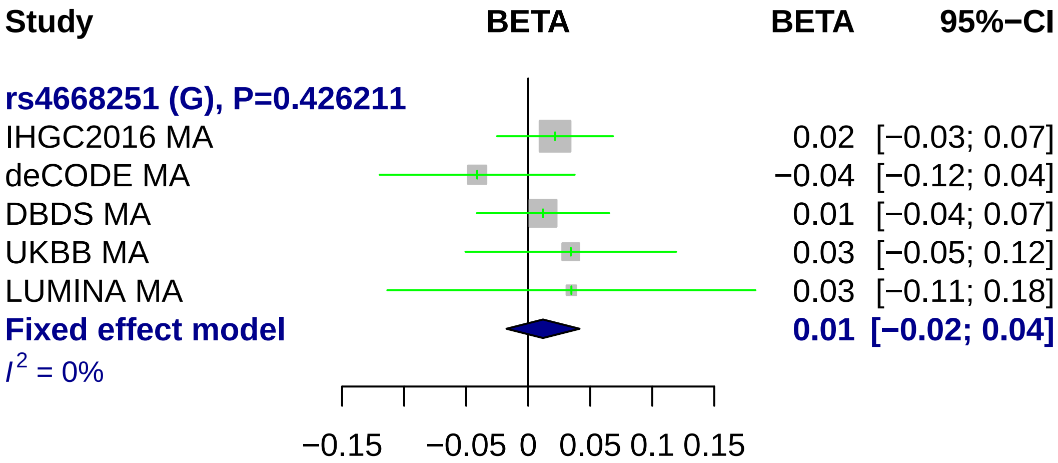
Study
BETA
BETA
95%−CI
rs4668251 (G), P=0.426211
IHGC2016 MA
0.02 [−0.03; 0.07]
deCODE MA
−0.04 [−0.12; 0.04]
DBDS MA
0.01 [−0.04; 0.07]
UKBB MA
0.03 [−0.05; 0.12]
LUMINA MA
0.03 [−0.11; 0.18]
Fixed effect model
0.01 [−0.02; 0.04]
I2 = 0%
−0.15
−0.05
0
0.05
0.1
0.15

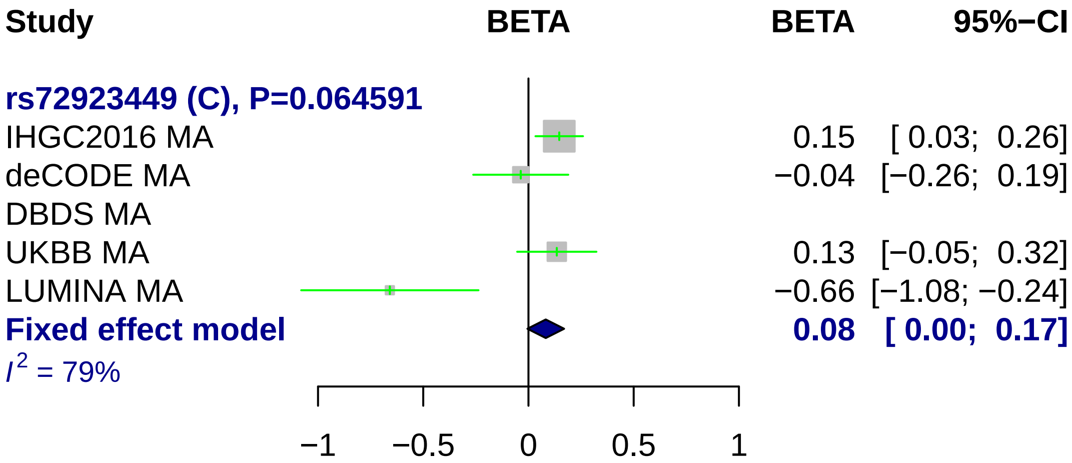
Study
BETA
BETA
95%−CI
rs72923449 (C), P=0.064591
IHGC2016 MA
0.15
[ 0.03; 0.26]
deCODE MA
−0.04
[−0.26; 0.19]
DBDS MA
UKBB MA
0.13
[−0.05; 0.32]
LUMINA MA
−0.66
[−1.08; −0.24]
Fixed effect model
0.08
[ 0.00; 0.17]
$I^2$ = 79%
−1
−0.5
0
0.5
1

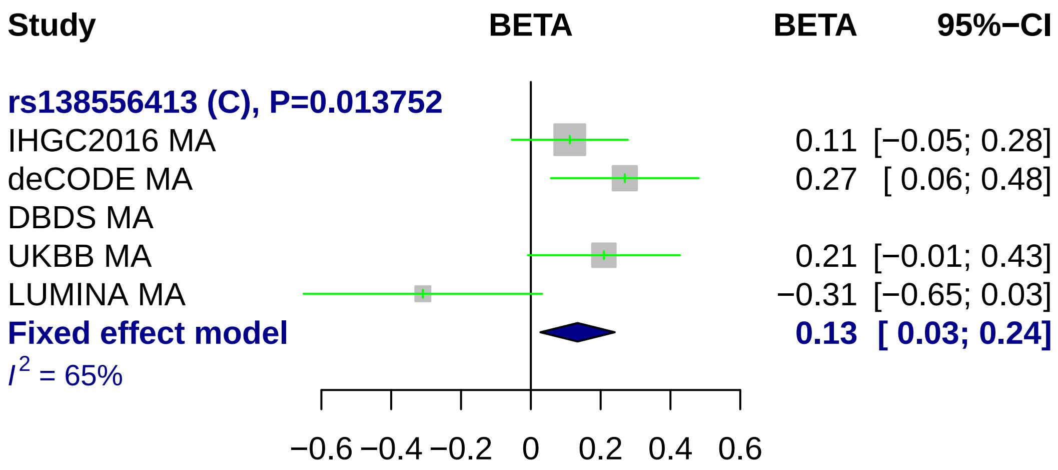
Study
BETA
BETA
95%−CI
rs138556413 (C), P=0.013752
IHGC2016 MA
0.11 [−0.05; 0.28]
deCODE MA
0.27 [ 0.06; 0.48]
DBDS MA
UKBB MA
0.21 [−0.01; 0.43]
LUMINA MA
−0.31 [−0.65; 0.03]
Fixed effect model
0.13 [ 0.03; 0.24]
I² = 65%
−0.6
−0.4
−0.2
0
0.2
0.4
0.6

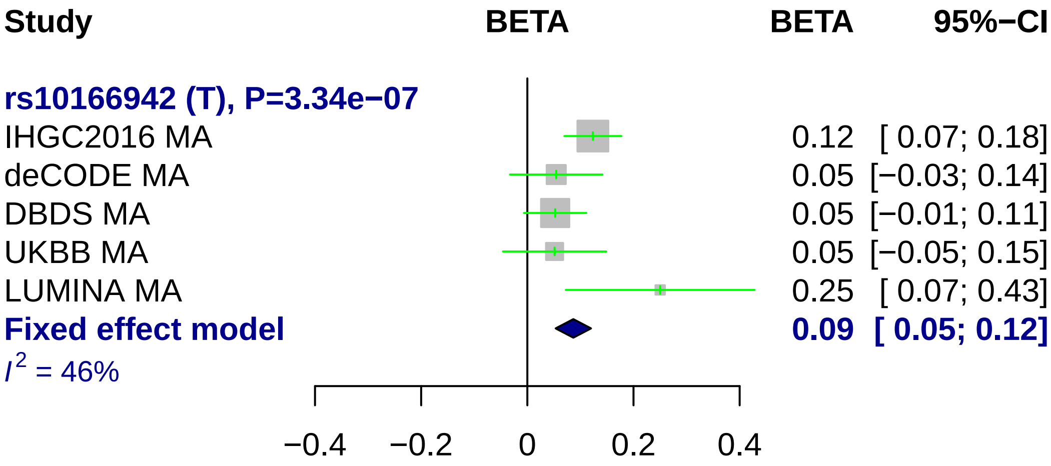
Study
BETA
BETA
95%−CI
rs10166942 (T), P=3.34e−07
IHGC2016 MA
0.12 [ 0.07; 0.18]
deCODE MA
0.05 [−0.03; 0.14]
DBDS MA
0.05 [−0.01; 0.11]
UKBB MA
0.05 [−0.05; 0.15]
LUMINA MA
0.25 [ 0.07; 0.43]
Fixed effect model
0.09 [ 0.05; 0.12]
I2 = 46%
−0.4
−0.2
0
0.2
0.4

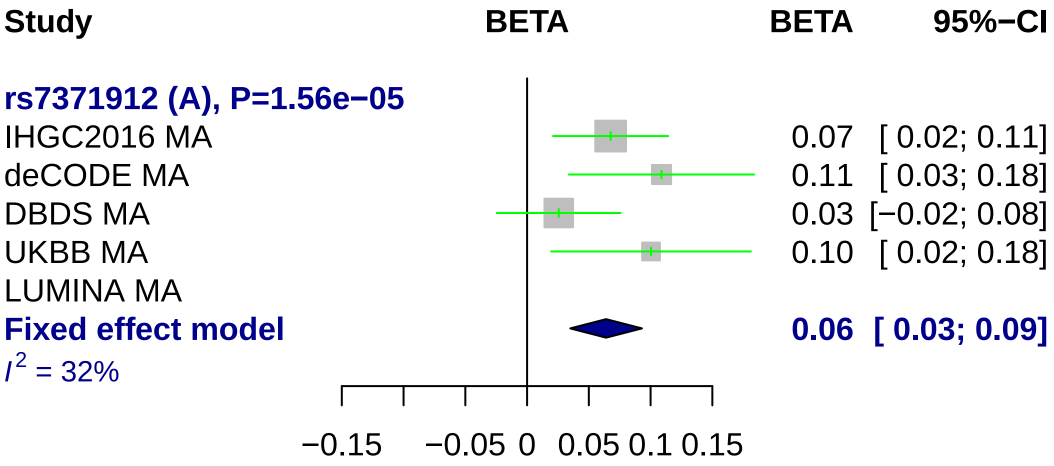
Study
BETA
BETA
95%−CI
rs7371912 (A), P=1.56e−05
IHGC2016 MA
0.07 [ 0.02; 0.11]
deCODE MA
0.11 [ 0.03; 0.18]
DBDS MA
0.03 [−0.02; 0.08]
UKBB MA
0.10 [ 0.02; 0.18]
LUMINA MA
Fixed effect model
0.06 [ 0.03; 0.09]
I2 = 32%
−0.15
−0.05
0
0.05
0.1
0.15

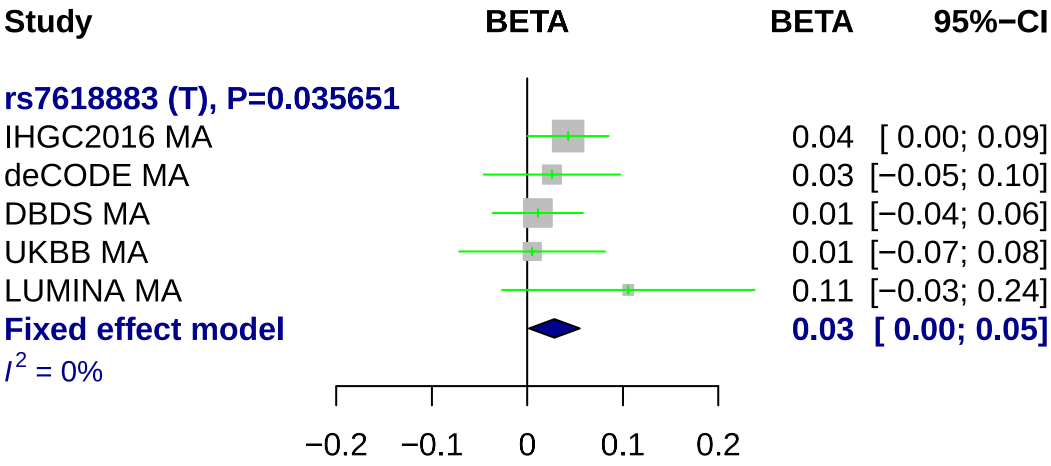

Study
BETA
BETA
95%−CI
rs7618883 (T), P=0.035651
IHGC2016 MA
0.04
[ 0.00; 0.09]
deCODE MA
0.03
[−0.05; 0.10]
DBDS MA
0.01
[−0.04; 0.06]
UKBB MA
0.01
[−0.07; 0.08]
LUMINA MA
0.11
[−0.03; 0.24]
Fixed effect model
0.03
[ 0.00; 0.05]
I2 = 0%
−0.2
−0.1
0
0.1
0.2

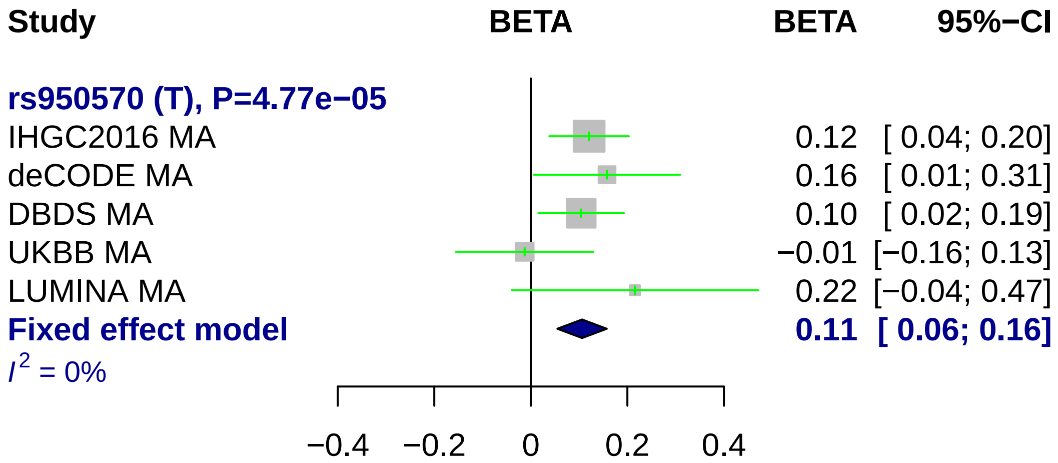
Study
BETA
BETA
95%−CI
rs950570 (T), P=4.77e−05
IHGC2016 MA
0.12 [ 0.04; 0.20]
deCODE MA
0.16 [ 0.01; 0.31]
DBDS MA
0.10 [ 0.02; 0.19]
UKBB MA
−0.01 [−0.16; 0.13]
LUMINA MA
0.22 [−0.04; 0.47]
Fixed effect model
0.11 [ 0.06; 0.16]
I^2 = 0%
−0.4
−0.2
0
0.2
0.4

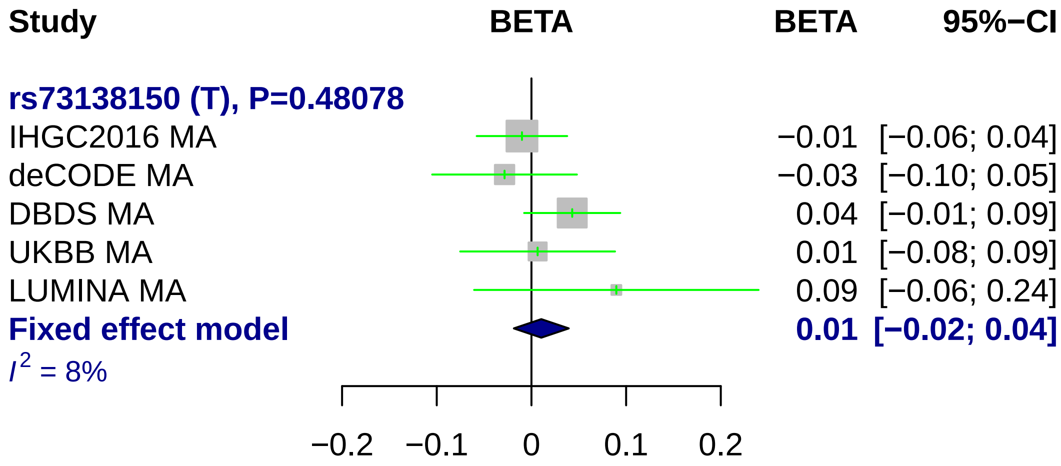
Study
BETA
BETA
95%−CI
rs73138150 (T), P=0.48078
IHGC2016 MA
−0.01 [−0.06; 0.04]
deCODE MA
−0.03 [−0.10; 0.05]
DBDS MA
0.04 [−0.01; 0.09]
UKBB MA
0.01 [−0.08; 0.09]
LUMINA MA
0.09 [−0.06; 0.24]
Fixed effect model
0.01 [−0.02; 0.04]
$I^2$ = 8%
−0.2
−0.1
0
0.1
0.2

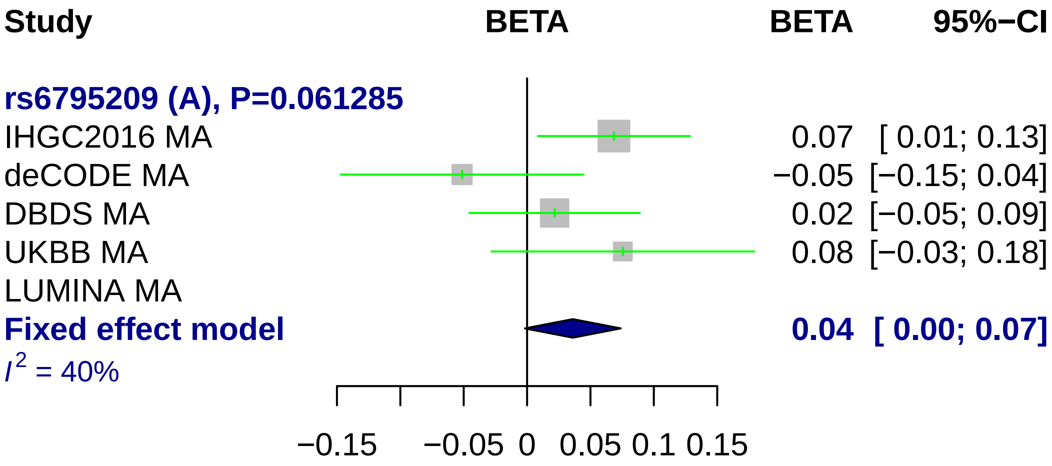
Study
BETA
BETA
95%−CI
rs6795209 (A), P=0.061285
IHGC2016 MA
0.07
[ 0.01; 0.13]
deCODE MA
−0.05
[−0.15; 0.04]
DBDS MA
0.02
[−0.05; 0.09]
UKBB MA
0.08
[−0.03; 0.18]
LUMINA MA
Fixed effect model
0.04
[ 0.00; 0.07]
I2 = 40%
−0.15
−0.05
0
0.05
0.1
0.15

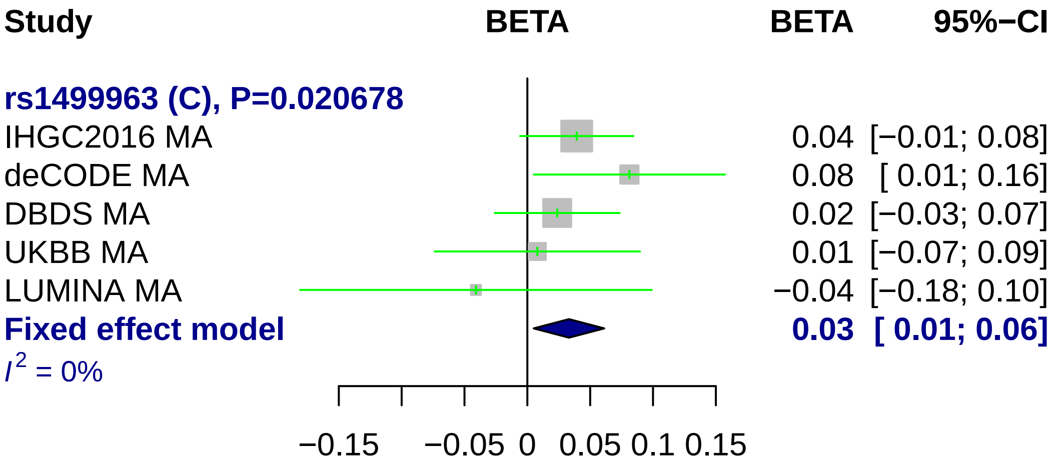
Study
BETA
BETA
95%−CI
rs1499963 (C), P=0.020678
IHGC2016 MA
0.04 [−0.01; 0.08]
deCODE MA
0.08 [ 0.01; 0.16]
DBDS MA
0.02 [−0.03; 0.07]
UKBB MA
0.01 [−0.07; 0.09]
LUMINA MA
−0.04 [−0.18; 0.10]
Fixed effect model
0.03 [ 0.01; 0.06]
I2 = 0%
−0.15
−0.05
0
0.05
0.1
0.15

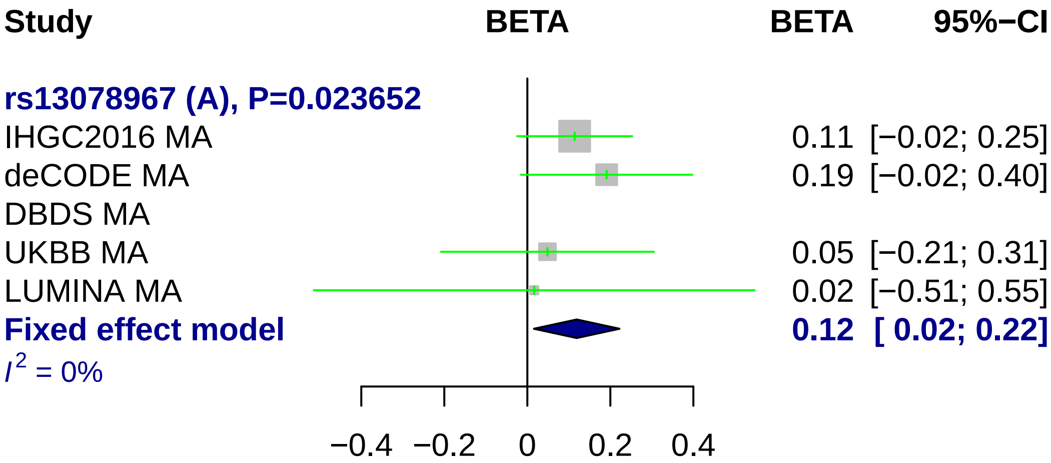
Study
BETA
BETA
95%−CI
rs13078967 (A), P=0.023652
IHGC2016 MA
0.11 [−0.02; 0.25]
deCODE MA
0.19 [−0.02; 0.40]
DBDS MA
UKBB MA
0.05 [−0.21; 0.31]
LUMINA MA
0.02 [−0.51; 0.55]
Fixed effect model
0.12 [ 0.02; 0.22]
I2 = 0%
−0.4
−0.2
0
0.2
0.4

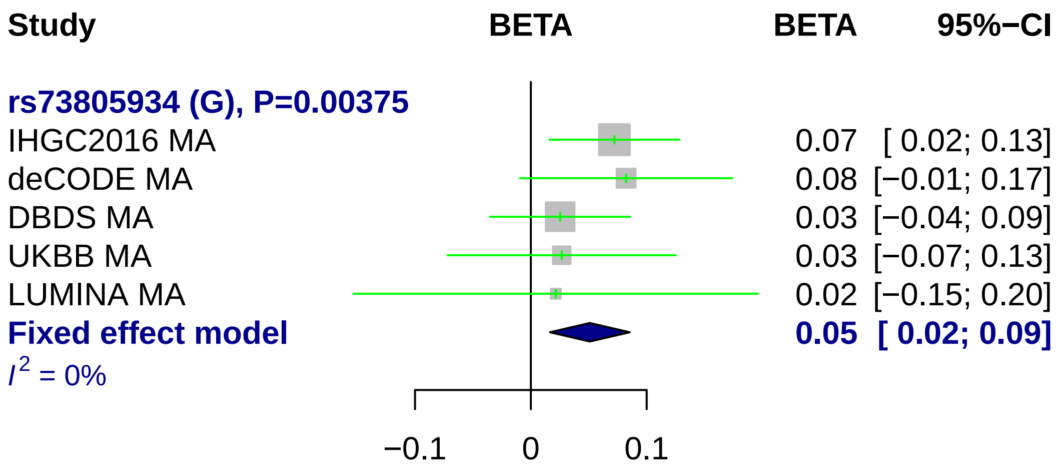
Study
BETA
BETA
95%−CI
rs73805934 (G), P=0.00375
IHGC2016 MA 0.07 [ 0.02; 0.13]
deCODE MA 0.08 [−0.01; 0.17]
DBDS MA 0.03 [−0.04; 0.09]
UKBB MA 0.03 [−0.07; 0.13]
LUMINA MA 0.02 [−0.15; 0.20]
Fixed effect model 0.05 [ 0.02; 0.09]
I2 = 0%
−0.1
0
0.1

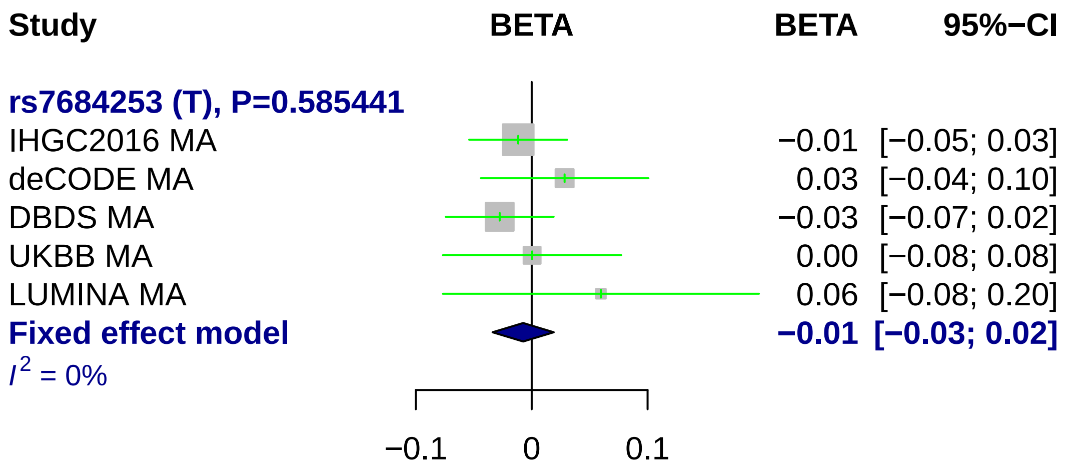

Study
BETA
BETA
95%−CI
rs7684253 (T), P=0.585441
IHGC2016 MA −0.01 [−0.05; 0.03]
deCODE MA 0.03 [−0.04; 0.10]
DBDS MA −0.03 [−0.07; 0.02]
UKBB MA 0.00 [−0.08; 0.08]
LUMINA MA 0.06 [−0.08; 0.20]
Fixed effect model −0.01 [−0.03; 0.02]
$I^2$ = 0%
−0.1
0
0.1

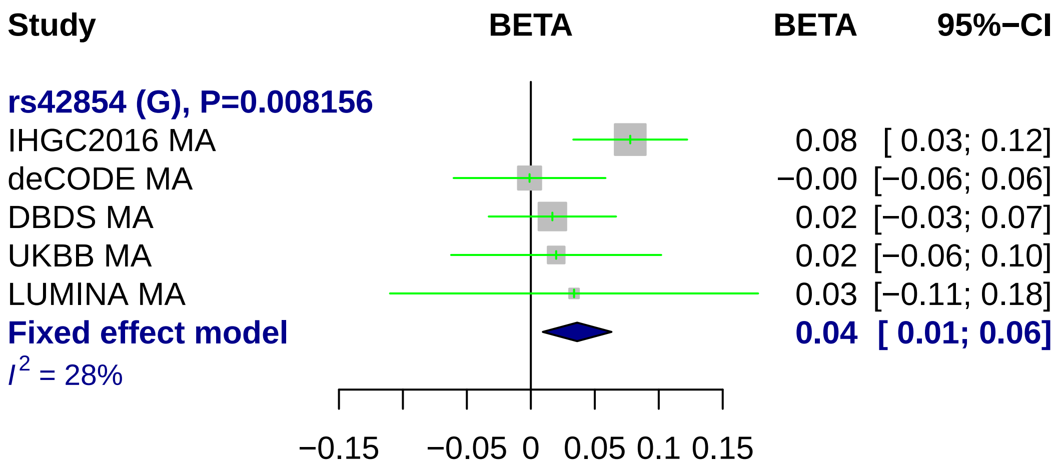
Study
BETA
BETA
95%−CI
rs42854 (G), P=0.008156
IHGC2016 MA
0.08 [ 0.03; 0.12]
deCODE MA
−0.00 [−0.06; 0.06]
DBDS MA
0.02 [−0.03; 0.07]
UKBB MA
0.02 [−0.06; 0.10]
LUMINA MA
0.03 [−0.11; 0.18]
Fixed effect model
0.04 [ 0.01; 0.06]
I2 = 28%
−0.15
−0.05
0
0.05
0.1
0.15

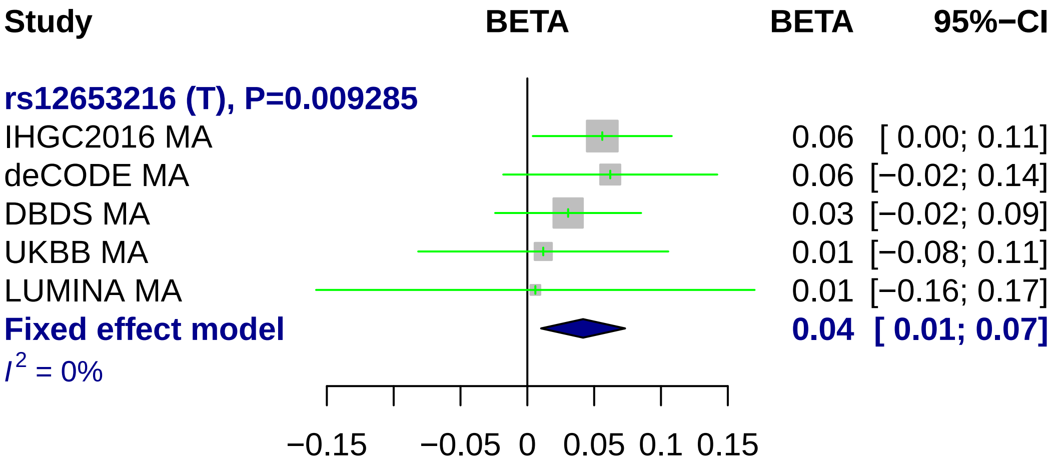
Study
BETA
BETA
95%−CI
rs12653216 (T), P=0.009285
IHGC2016 MA
0.06 [ 0.00; 0.11]
deCODE MA
0.06 [−0.02; 0.14]
DBDS MA
0.03 [−0.02; 0.09]
UKBB MA
0.01 [−0.08; 0.11]
LUMINA MA
0.01 [−0.16; 0.17]
Fixed effect model
0.04 [ 0.01; 0.07]
$I^2$ = 0%
−0.15
−0.05
0
0.05
0.1
0.15

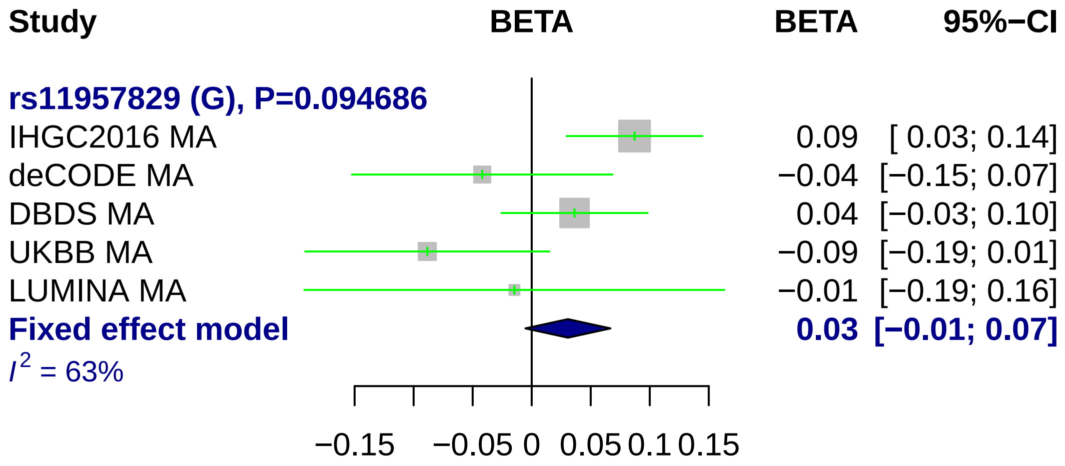
Study
BETA
BETA
95%−CI
rs11957829 (G), P=0.094686
IHGC2016 MA
0.09
[ 0.03; 0.14]
deCODE MA
−0.04
[−0.15; 0.07]
DBDS MA
0.04
[−0.03; 0.10]
UKBB MA
−0.09
[−0.19; 0.01]
LUMINA MA
−0.01
[−0.19; 0.16]
Fixed effect model
0.03
[−0.01; 0.07]
I 2 = 63%
−0.15
−0.05
0
0.05
0.1
0.15

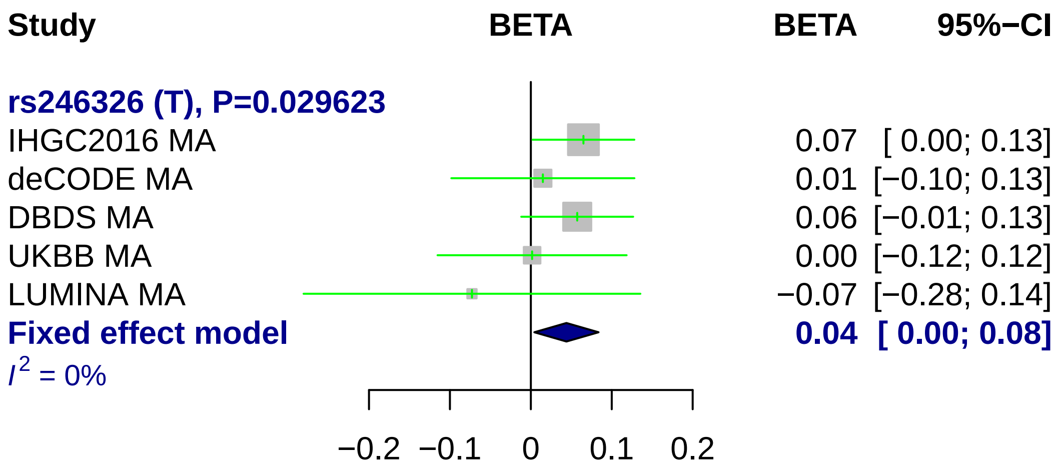
Study
BETA
BETA
95%−CI
rs246326 (T), P=0.029623
IHGC2016 MA
0.07 [ 0.00; 0.13]
deCODE MA
0.01 [−0.10; 0.13]
DBDS MA
0.06 [−0.01; 0.13]
UKBB MA
0.00 [−0.12; 0.12]
LUMINA MA
−0.07 [−0.28; 0.14]
Fixed effect model
0.04 [ 0.00; 0.08]
I2 = 0%
−0.2
−0.1
0
0.1
0.2

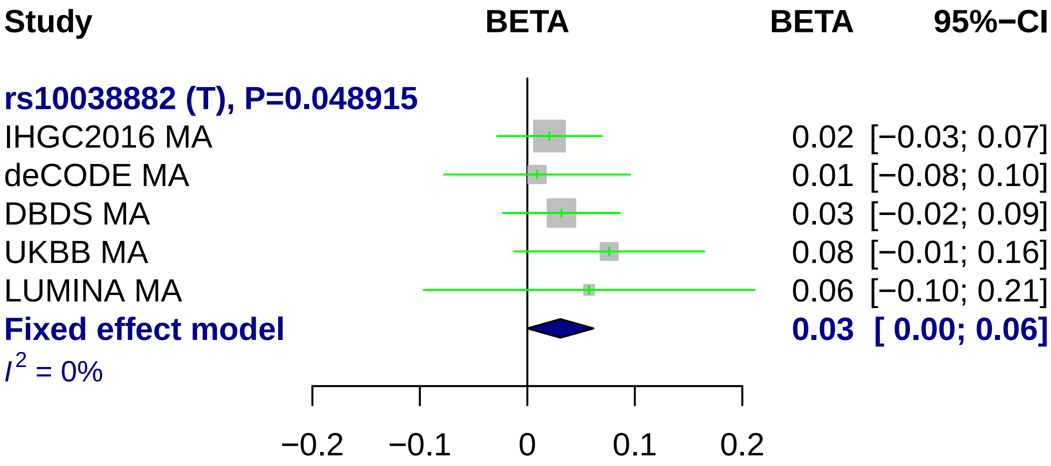
Study
BETA
BETA
95%−CI
rs10038882 (T), P=0.048915
IHGC2016 MA
0.02 [−0.03; 0.07]
deCODE MA
0.01 [−0.08; 0.10]
DBDS MA
0.03 [−0.02; 0.09]
UKBB MA
0.08 [−0.01; 0.16]
LUMINA MA
0.06 [−0.10; 0.21]
Fixed effect model
0.03 [ 0.00; 0.06]
I2 = 0%
−0.2
−0.1
0
0.1
0.2

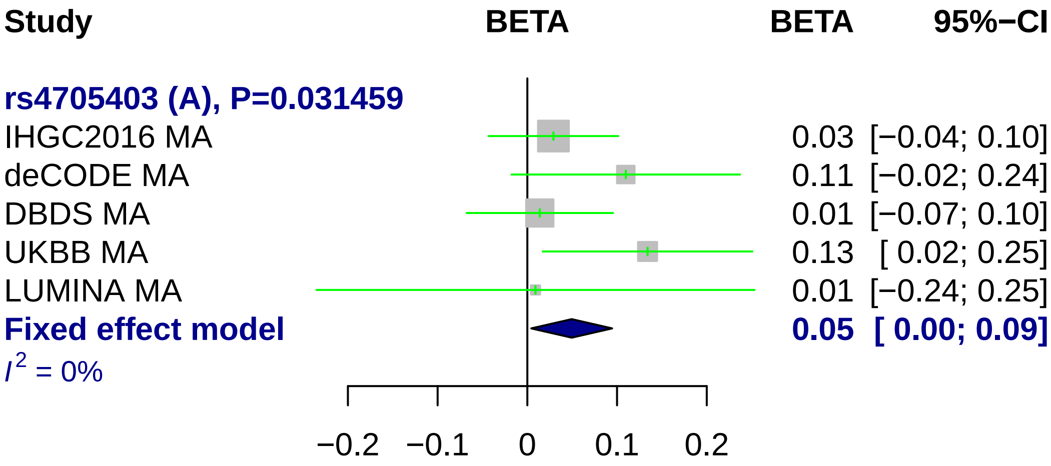
Study
BETA
BETA
95%−CI
rs4705403 (A), P=0.031459
IHGC2016 MA
0.03 [−0.04; 0.10]
deCODE MA
0.11 [−0.02; 0.24]
DBDS MA
0.01 [−0.07; 0.10]
UKBB MA
0.13 [ 0.02; 0.25]
LUMINA MA
0.01 [−0.24; 0.25]
Fixed effect model
0.05 [ 0.00; 0.09]
I^2 = 0%
−0.2
−0.1
0
0.1
0.2

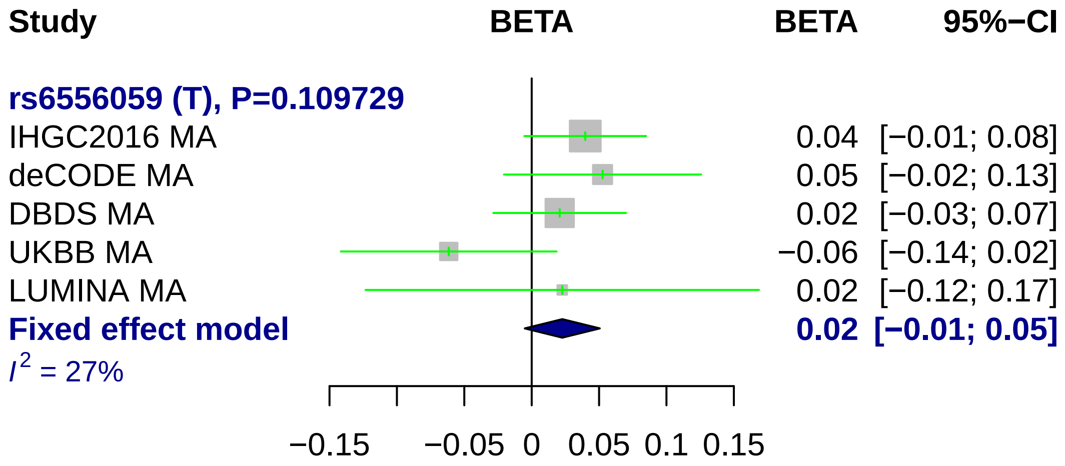
Study
BETA
BETA
95%−CI
rs6556059 (T), P=0.109729
IHGC2016 MA
0.04
[−0.01; 0.08]
deCODE MA
0.05
[−0.02; 0.13]
DBDS MA
0.02
[−0.03; 0.07]
UKBB MA
−0.06
[−0.14; 0.02]
LUMINA MA
0.02
[−0.12; 0.17]
Fixed effect model
0.02
[−0.01; 0.05]
I^2 = 27%
−0.15
−0.05
0
0.05
0.1
0.15

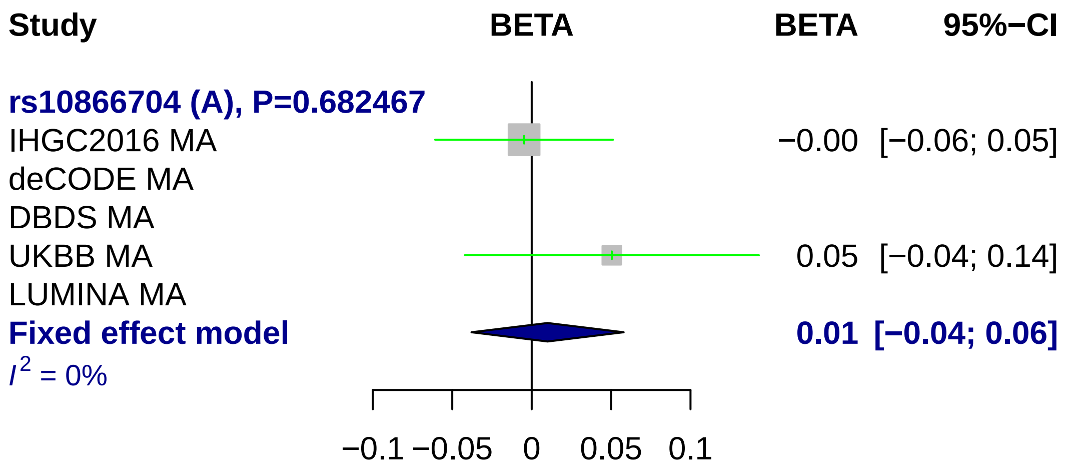
Study
BETA
BETA
95%−CI
rs10866704 (A), P=0.682467
IHGC2016 MA
−0.00 [−0.06; 0.05]
deCODE MA
DBDS MA
UKBB MA
0.05 [−0.04; 0.14]
LUMINA MA
Fixed effect model
0.01 [−0.04; 0.06]
I2 = 0%
−0.1
−0.05
0
0.05
0.1

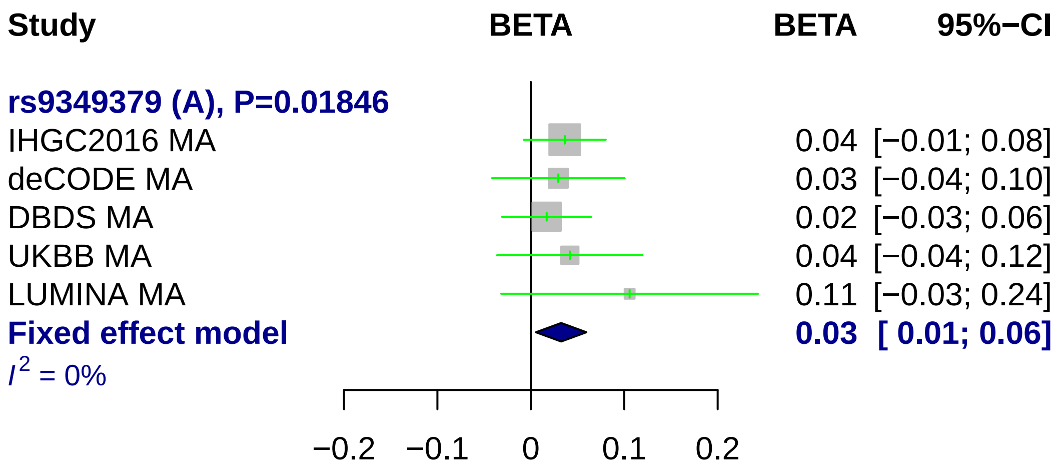
Study
BETA
BETA
95%−CI
rs9349379 (A), P=0.01846
IHGC2016 MA 0.04 [−0.01; 0.08]
deCODE MA 0.03 [−0.04; 0.10]
DBDS MA 0.02 [−0.03; 0.06]
UKBB MA 0.04 [−0.04; 0.12]
LUMINA MA 0.11 [−0.03; 0.24]
Fixed effect model 0.03 [ 0.01; 0.06]
I² = 0%
−0.2
−0.1
0
0.1
0.2

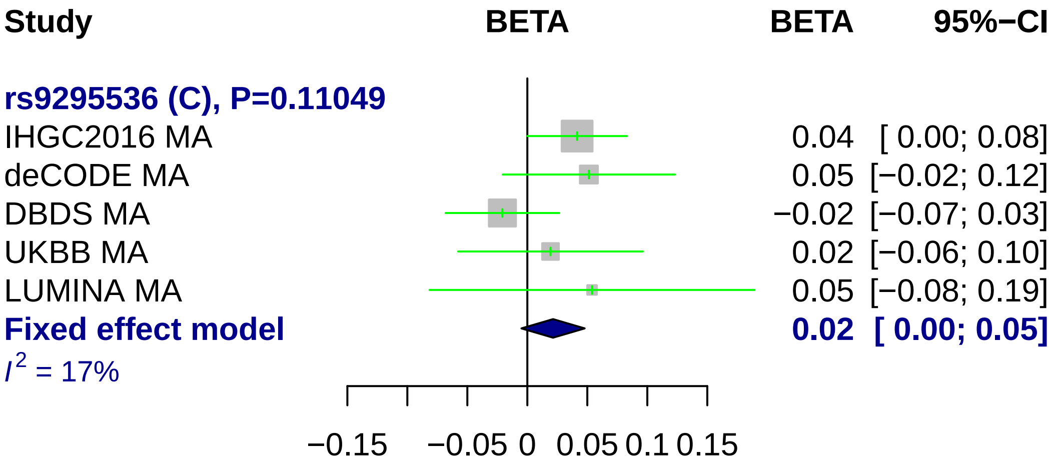
Study
BETA
BETA
95%−CI
rs9295536 (C), P=0.11049
IHGC2016 MA 0.04 [ 0.00; 0.08]
deCODE MA 0.05 [−0.02; 0.12]
DBDS MA −0.02 [−0.07; 0.03]
UKBB MA 0.02 [−0.06; 0.10]
LUMINA MA 0.05 [−0.08; 0.19]
Fixed effect model 0.02 [ 0.00; 0.05]
I^2 = 17%
−0.15
−0.05
0
0.05
0.1
0.15

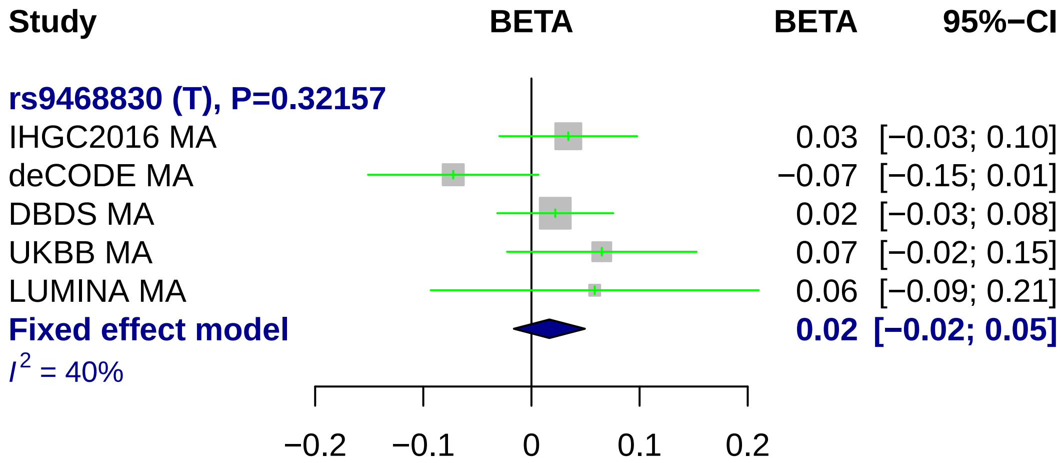
Study
BETA
BETA
95%−CI
rs9468830 (T), P=0.32157
IHGC2016 MA
0.03 [−0.03; 0.10]
deCODE MA
−0.07 [−0.15; 0.01]
DBDS MA
0.02 [−0.03; 0.08]
UKBB MA
0.07 [−0.02; 0.15]
LUMINA MA
0.06 [−0.09; 0.21]
Fixed effect model
0.02 [−0.02; 0.05]
I2 = 40%
−0.2
−0.1
0
0.1
0.2

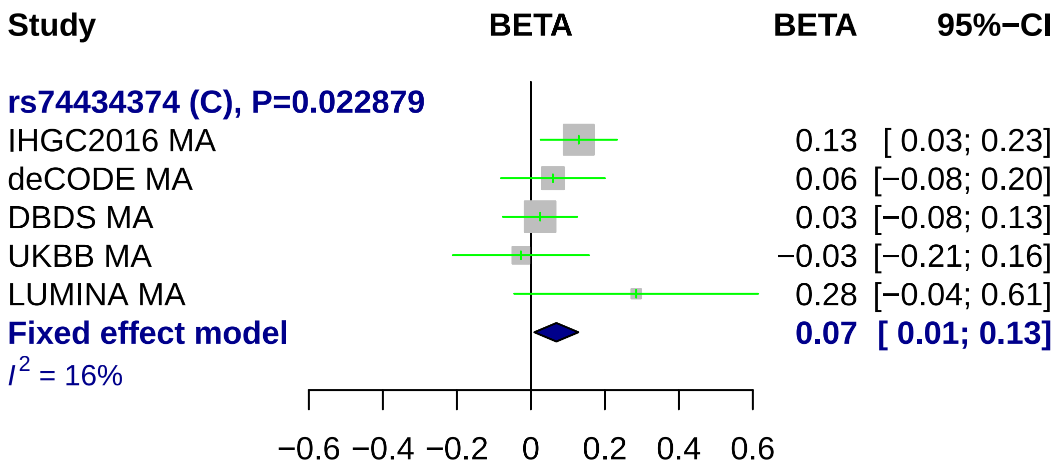
Study
BETA
BETA
95%−CI
rs74434374 (C), P=0.022879
IHGC2016 MA
deCODE MA
DBDS MA
UKBB MA
LUMINA MA
Fixed effect model
$I^2$ = 16%
0.13 [ 0.03; 0.23]
0.06 [−0.08; 0.20]
0.03 [−0.08; 0.13]
−0.03 [−0.21; 0.16]
0.28 [−0.04; 0.61]
0.07 [ 0.01; 0.13]
−0.6 −0.4 −0.2 0 0.2 0.4 0.6

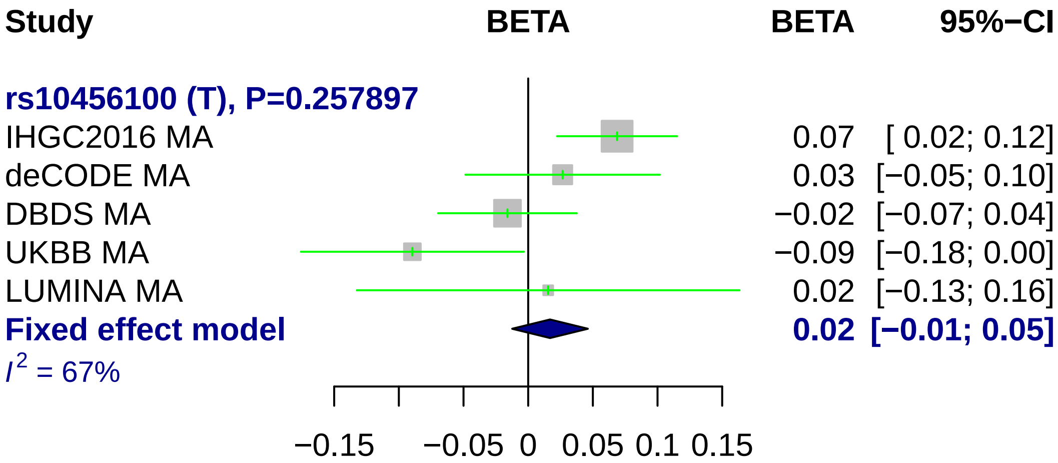

Study
BETA
BETA
95%−CI
rs10456100 (T), P=0.257897
IHGC2016 MA
0.07
[ 0.02; 0.12]
deCODE MA
0.03
[−0.05; 0.10]
DBDS MA
−0.02
[−0.07; 0.04]
UKBB MA
−0.09
[−0.18; 0.00]
LUMINA MA
0.02
[−0.13; 0.16]
Fixed effect model
0.02
[−0.01; 0.05]
I2 = 67%
−0.15
−0.05
0
0.05
0.1
0.15

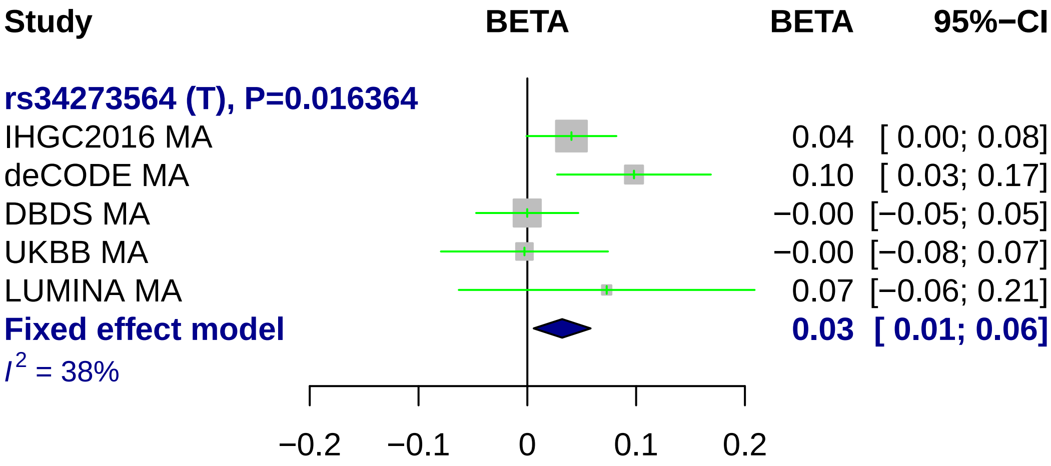
Study
BETA
BETA
95%−CI
rs34273564 (T), P=0.016364
IHGC2016 MA
0.04
[ 0.00; 0.08]
deCODE MA
0.10
[ 0.03; 0.17]
DBDS MA
−0.00
[−0.05; 0.05]
UKBB MA
−0.00
[−0.08; 0.07]
LUMINA MA
0.07
[−0.06; 0.21]
Fixed effect model
0.03
[ 0.01; 0.06]
I2 = 38%
−0.2
−0.1
0
0.1
0.2

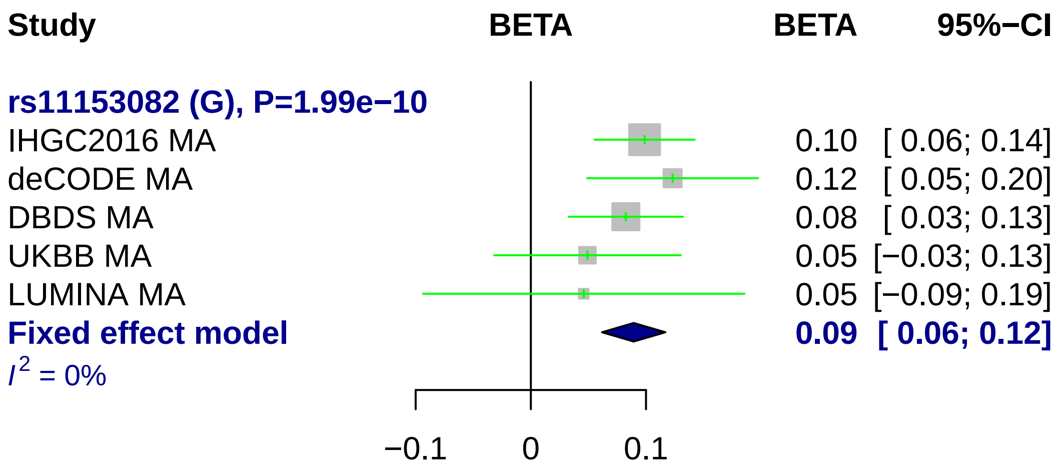
Study
BETA
BETA
95%−CI
rs11153082 (G), P=1.99e−10
IHGC2016 MA 0.10 [ 0.06; 0.14]
deCODE MA 0.12 [ 0.05; 0.20]
DBDS MA 0.08 [ 0.03; 0.13]
UKBB MA 0.05 [−0.03; 0.13]
LUMINA MA 0.05 [−0.09; 0.19]
Fixed effect model 0.09 [ 0.06; 0.12]
I2 = 0%
−0.1
0
0.1

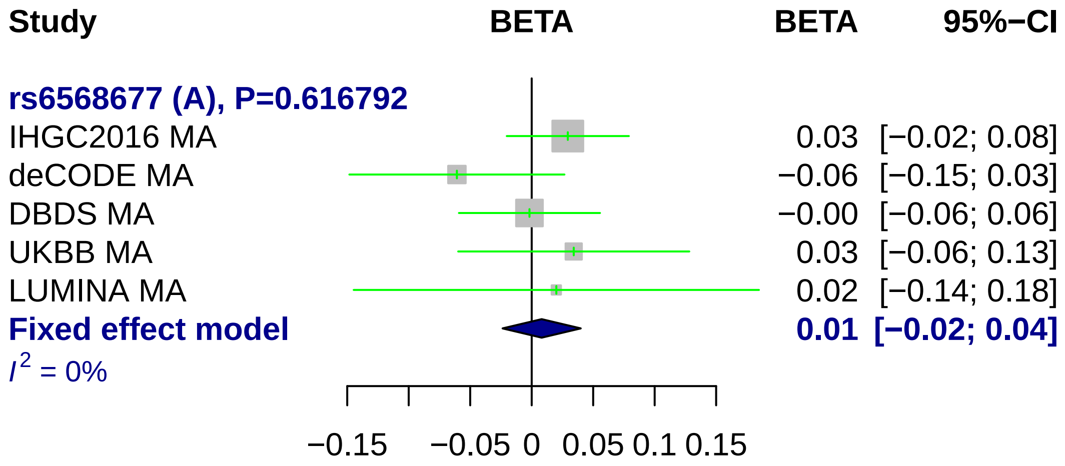
Study
BETA
BETA
95%−CI
rs6568677 (A), P=0.616792
IHGC2016 MA
0.03 [−0.02; 0.08]
deCODE MA
−0.06 [−0.15; 0.03]
DBDS MA
−0.00 [−0.06; 0.06]
UKBB MA
0.03 [−0.06; 0.13]
LUMINA MA
0.02 [−0.14; 0.18]
Fixed effect model
0.01 [−0.02; 0.04]
I2 = 0%
−0.15
−0.05
0
0.05
0.1
0.15

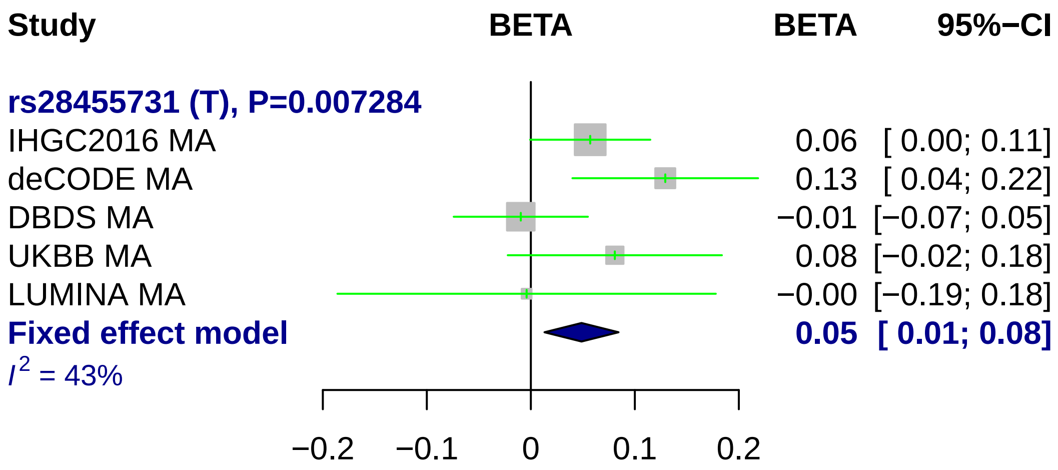
Study
BETA
BETA
95%−CI
rs28455731 (T), P=0.007284
IHGC2016 MA 0.06 [ 0.00; 0.11]
deCODE MA 0.13 [ 0.04; 0.22]
DBDS MA −0.01 [−0.07; 0.05]
UKBB MA 0.08 [−0.02; 0.18]
LUMINA MA −0.00 [−0.19; 0.18]
Fixed effect model 0.05 [ 0.01; 0.08]
I^2 = 43%
−0.2
−0.1
0
0.1
0.2

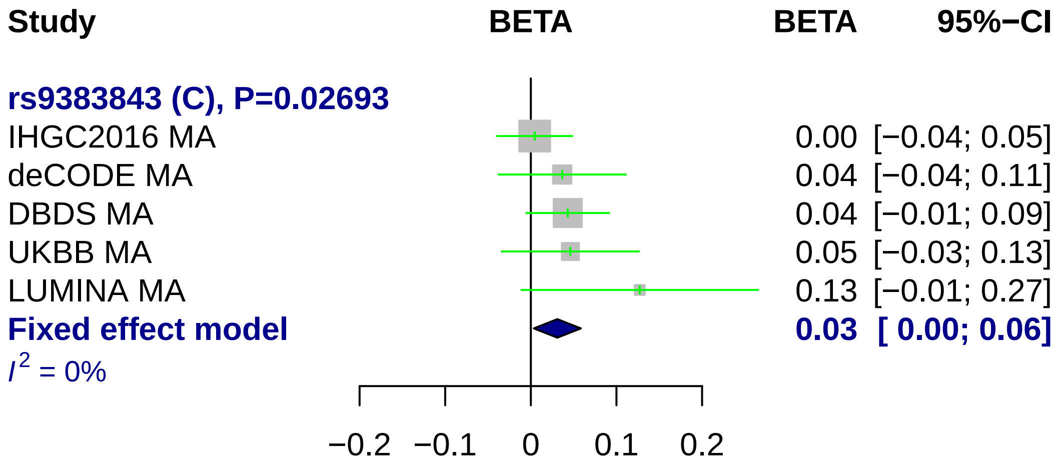
Study
BETA
BETA
95%−CI
rs9383843 (C), P=0.02693
IHGC2016 MA
0.00 [−0.04; 0.05]
deCODE MA
0.04 [−0.04; 0.11]
DBDS MA
0.04 [−0.01; 0.09]
UKBB MA
0.05 [−0.03; 0.13]
LUMINA MA
0.13 [−0.01; 0.27]
Fixed effect model
0.03 [ 0.00; 0.06]
I2 = 0%
−0.2
−0.1
0
0.1
0.2

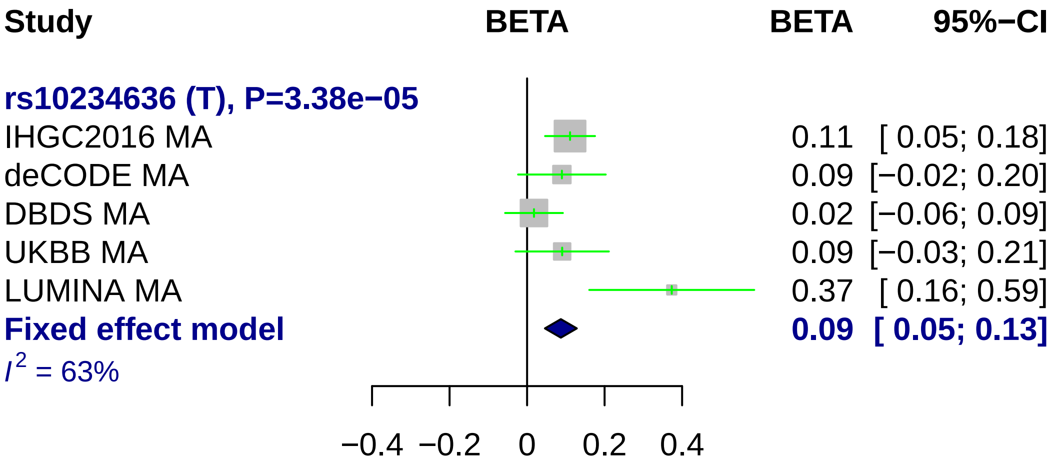
Study
BETA
BETA
95%−CI
rs10234636 (T), P=3.38e−05
IHGC2016 MA
0.11 [ 0.05; 0.18]
deCODE MA
0.09 [−0.02; 0.20]
DBDS MA
0.02 [−0.06; 0.09]
UKBB MA
0.09 [−0.03; 0.21]
LUMINA MA
0.37 [ 0.16; 0.59]
Fixed effect model
0.09 [ 0.05; 0.13]
I2 = 63%
−0.4
−0.2
0
0.2
0.4

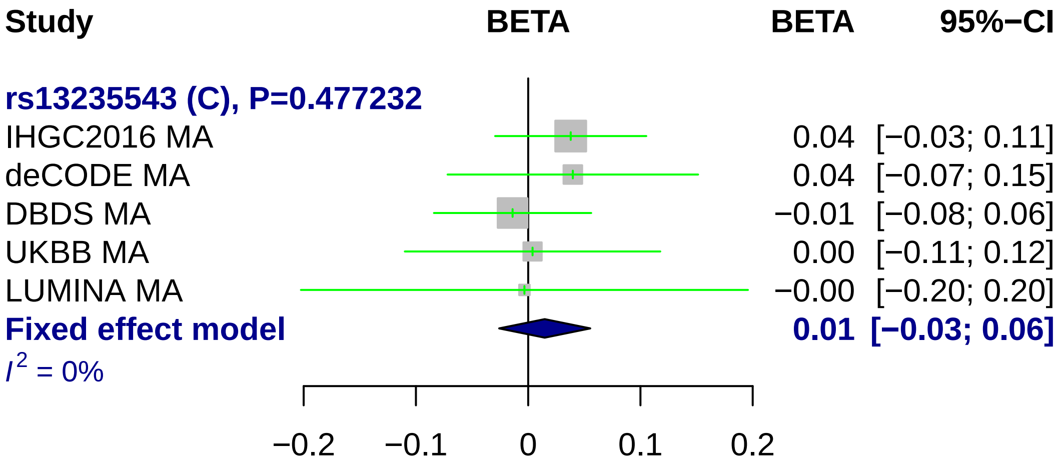
Study
BETA
BETA
95%−CI
rs13235543 (C), P=0.477232
IHGC2016 MA
0.04 [−0.03; 0.11]
deCODE MA
0.04 [−0.07; 0.15]
DBDS MA
−0.01 [−0.08; 0.06]
UKBB MA
0.00 [−0.11; 0.12]
LUMINA MA
−0.00 [−0.20; 0.20]
Fixed effect model
0.01 [−0.03; 0.06]
I2 = 0%
−0.2
−0.1
0
0.1
0.2

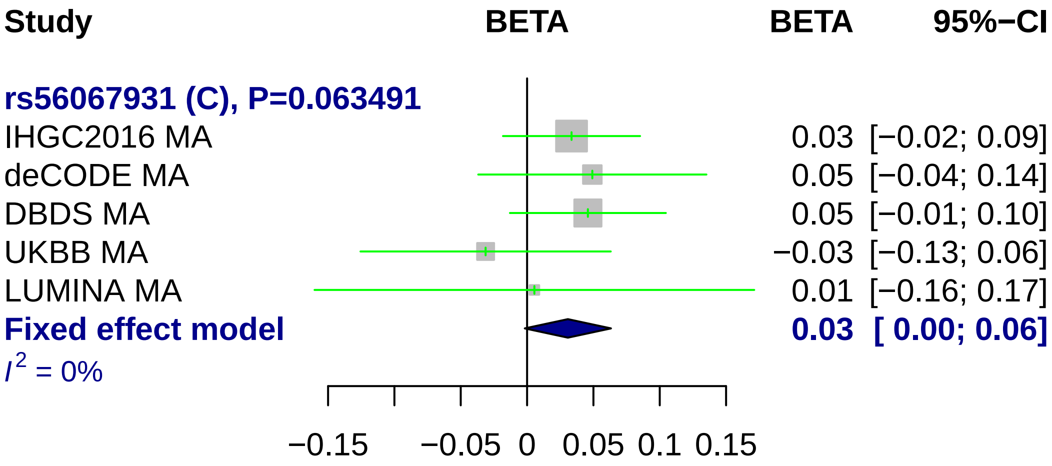
Study
BETA
BETA
95%−CI
rs56067931 (C), P=0.063491
IHGC2016 MA 0.03 [−0.02; 0.09]
deCODE MA 0.05 [−0.04; 0.14]
DBDS MA 0.05 [−0.01; 0.10]
UKBB MA −0.03 [−0.13; 0.06]
LUMINA MA 0.01 [−0.16; 0.17]
Fixed effect model 0.03 [ 0.00; 0.06]
I2 = 0%
−0.15
−0.05
0
0.05
0.1
0.15

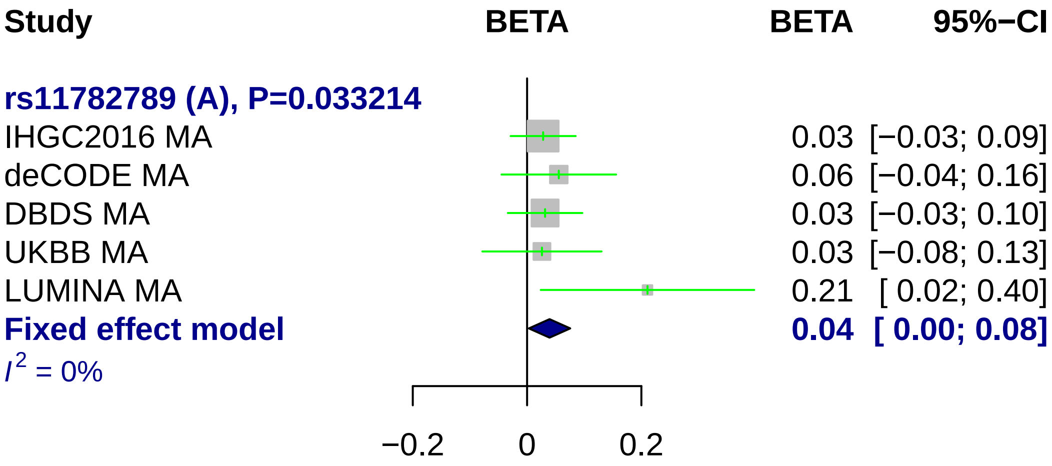
Study
BETA
BETA
95%−CI
rs11782789 (A), P=0.033214
IHGC2016 MA 0.03 [−0.03; 0.09]
deCODE MA 0.06 [−0.04; 0.16]
DBDS MA 0.03 [−0.03; 0.10]
UKBB MA 0.03 [−0.08; 0.13]
LUMINA MA 0.21 [ 0.02; 0.40]
Fixed effect model 0.04 [ 0.00; 0.08]
I2 = 0%
−0.2
0
0.2

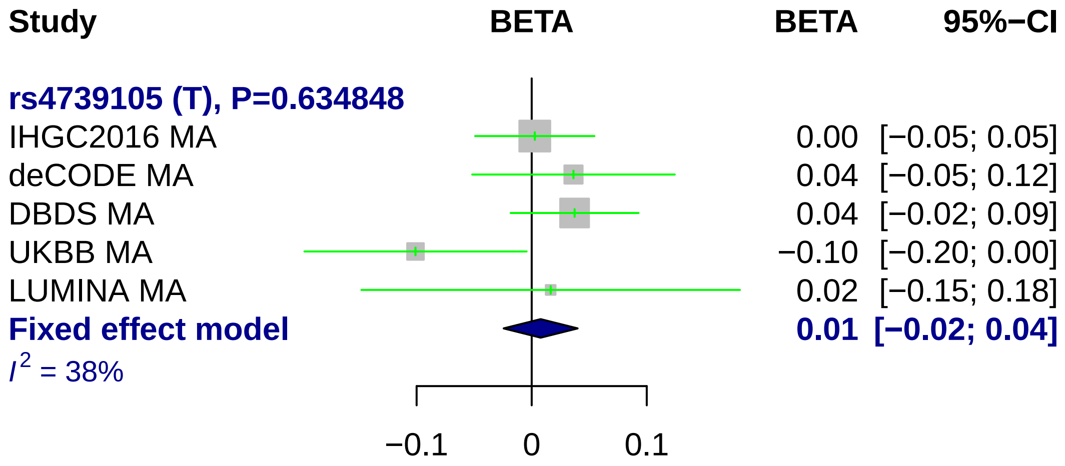
Study
BETA
BETA
95%−CI
rs4739105 (T), P=0.634848
IHGC2016 MA 0.00 [−0.05; 0.05]
deCODE MA 0.04 [−0.05; 0.12]
DBDS MA 0.04 [−0.02; 0.09]
UKBB MA −0.10 [−0.20; 0.00]
LUMINA MA 0.02 [−0.15; 0.18]
Fixed effect model 0.01 [−0.02; 0.04]
I2 = 38%
−0.1
0
0.1

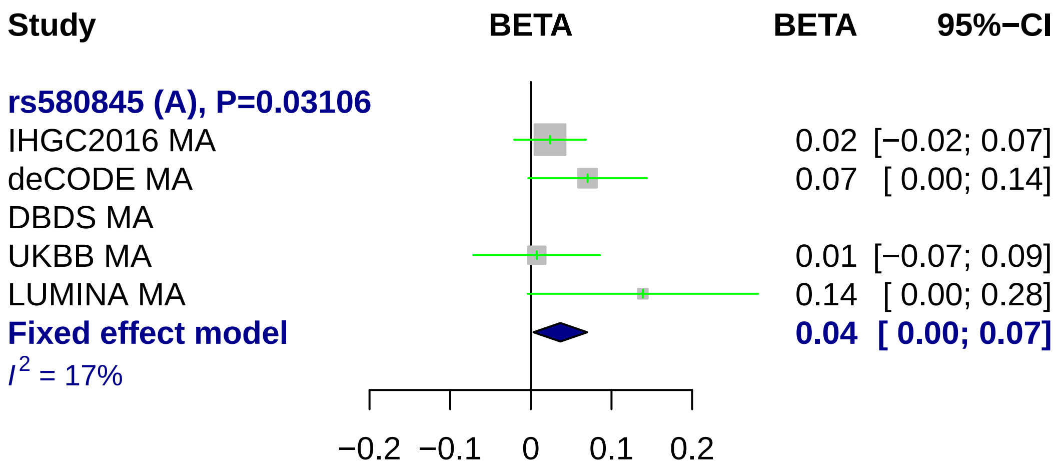
Study
BETA
BETA
95%−CI
rs580845 (A), P=0.03106
IHGC2016 MA
0.02 [−0.02; 0.07]
deCODE MA
0.07 [ 0.00; 0.14]
DBDS MA
UKBB MA
0.01 [−0.07; 0.09]
LUMINA MA
0.14 [ 0.00; 0.28]
Fixed effect model
0.04 [ 0.00; 0.07]
I^2 = 17%
−0.2
−0.1
0
0.1
0.2

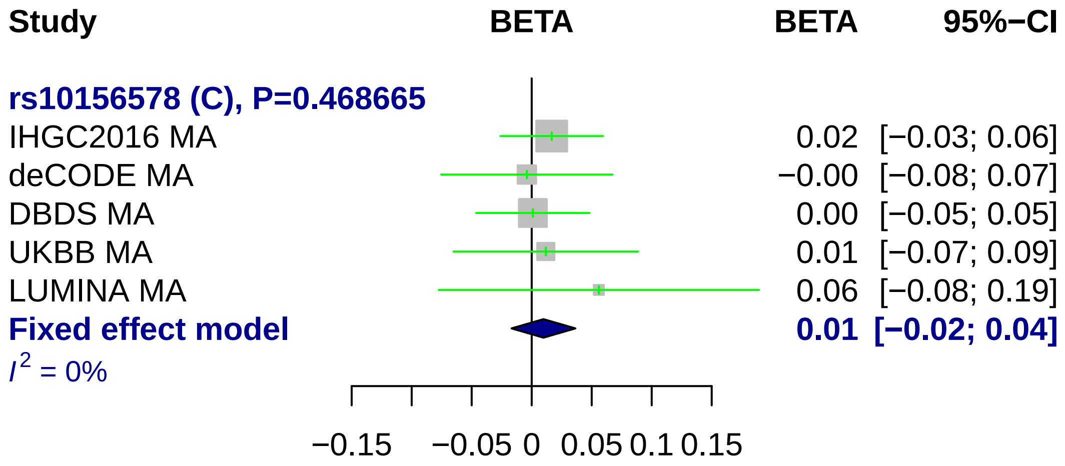
Study
BETA
BETA
95%−CI
rs10156578 (C), P=0.468665
IHGC2016 MA
0.02 [−0.03; 0.06]
deCODE MA
−0.00 [−0.08; 0.07]
DBDS MA
0.00 [−0.05; 0.05]
UKBB MA
0.01 [−0.07; 0.09]
LUMINA MA
0.06 [−0.08; 0.19]
Fixed effect model
0.01 [−0.02; 0.04]
I2 = 0%
−0.15
−0.05
0
0.05
0.1
0.15

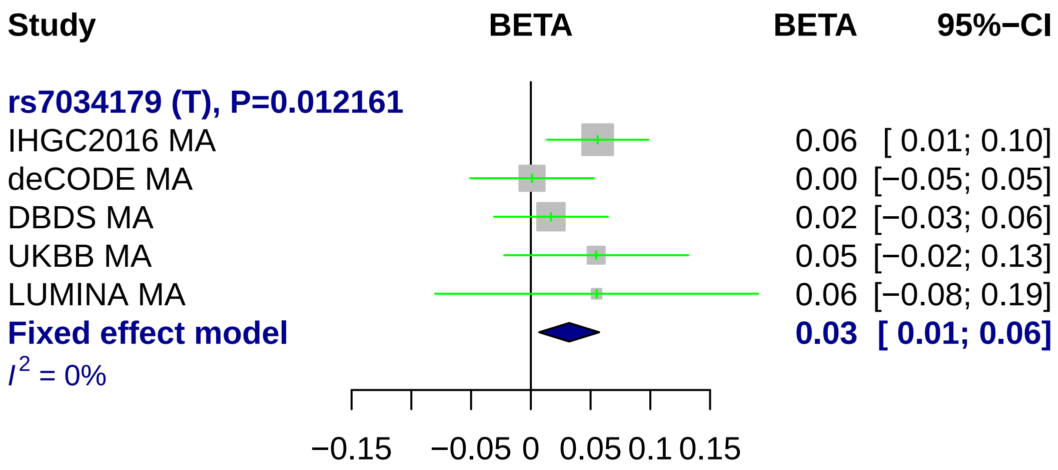
Study
BETA
BETA
95%−CI
rs7034179 (T), P=0.012161
IHGC2016 MA
0.06 [ 0.01; 0.10]
deCODE MA
0.00 [−0.05; 0.05]
DBDS MA
0.02 [−0.03; 0.06]
UKBB MA
0.05 [−0.02; 0.13]
LUMINA MA
0.06 [−0.08; 0.19]
Fixed effect model
0.03 [ 0.01; 0.06]
I2 = 0%
−0.15
−0.05
0
0.05
0.1
0.15

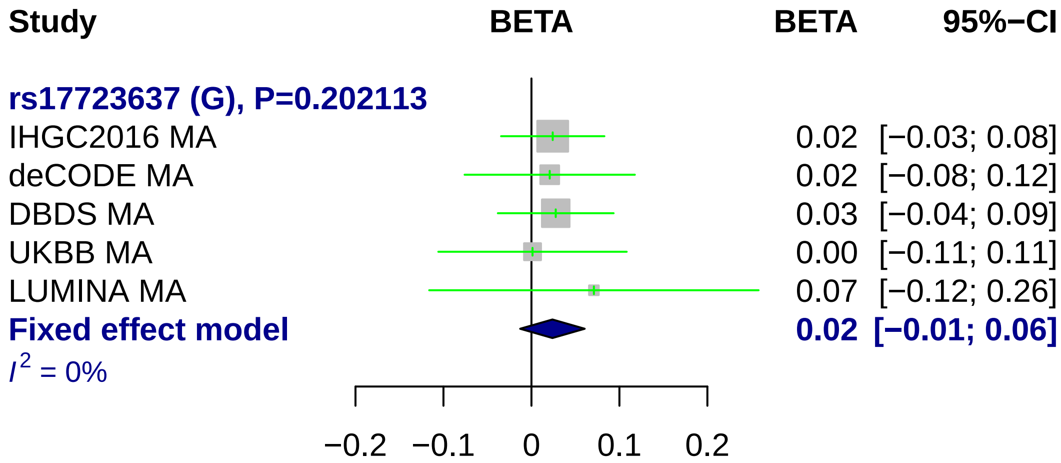
Study
BETA
BETA
95%−CI
rs17723637 (G), P=0.202113
IHGC2016 MA 0.02 [−0.03; 0.08]
deCODE MA 0.02 [−0.08; 0.12]
DBDS MA 0.03 [−0.04; 0.09]
UKBB MA 0.00 [−0.11; 0.11]
LUMINA MA 0.07 [−0.12; 0.26]
Fixed effect model 0.02 [−0.01; 0.06]
$I^2$ = 0%
−0.2
−0.1
0
0.1
0.2

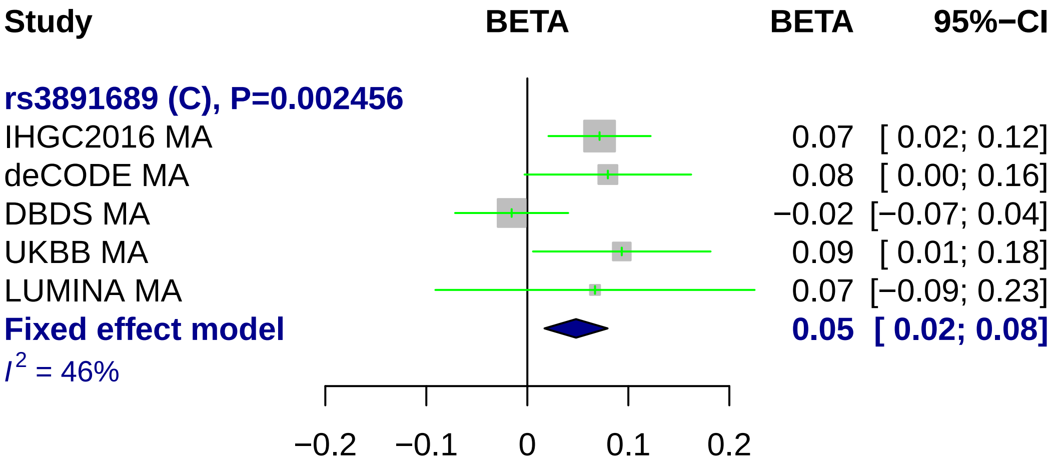
Study
BETA
BETA
95%−CI
rs3891689 (C), P=0.002456
IHGC2016 MA
0.07 [ 0.02; 0.12]
deCODE MA
0.08 [ 0.00; 0.16]
DBDS MA
−0.02 [−0.07; 0.04]
UKBB MA
0.09 [ 0.01; 0.18]
LUMINA MA
0.07 [−0.09; 0.23]
Fixed effect model
0.05 [ 0.02; 0.08]
I2 = 46%
−0.2
−0.1
0
0.1
0.2

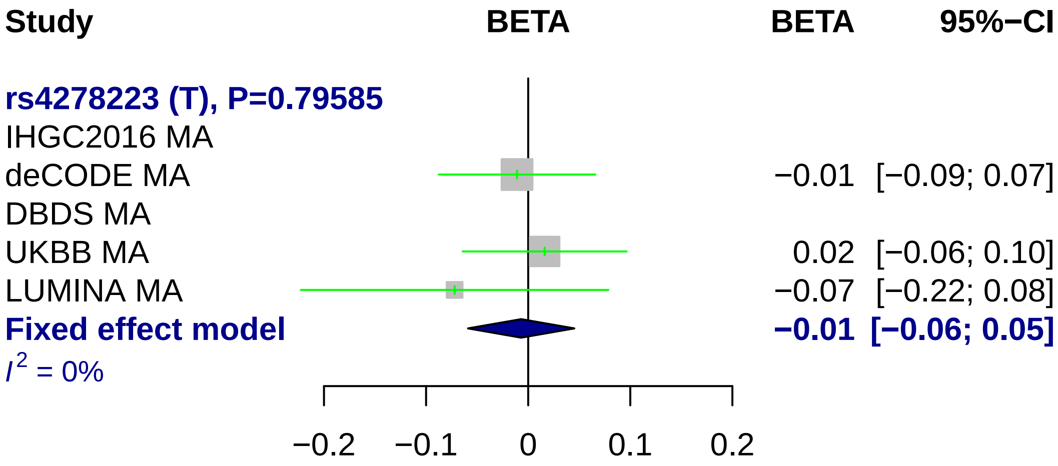
Study
BETA
BETA
95%−CI
rs4278223 (T), P=0.79585
IHGC2016 MA
deCODE MA −0.01 [−0.09; 0.07]
DBDS MA
UKBB MA 0.02 [−0.06; 0.10]
LUMINA MA −0.07 [−0.22; 0.08]
Fixed effect model −0.01 [−0.06; 0.05]
I² = 0%
−0.2
−0.1
0
0.1
0.2

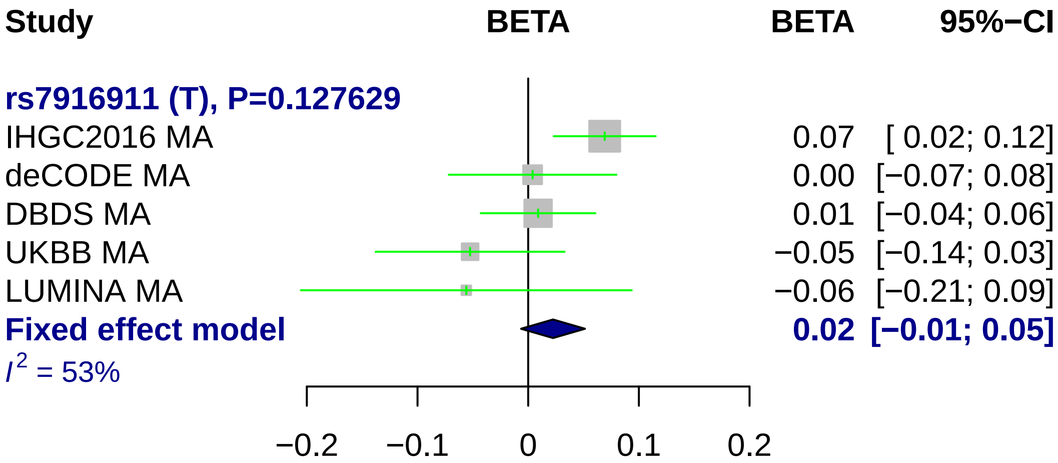
Study
BETA
BETA
95%−CI
rs7916911 (T), P=0.127629
IHGC2016 MA
0.07
[ 0.02; 0.12]
deCODE MA
0.00
[−0.07; 0.08]
DBDS MA
0.01
[−0.04; 0.06]
UKBB MA
−0.05
[−0.14; 0.03]
LUMINA MA
−0.06
[−0.21; 0.09]
Fixed effect model
0.02
[−0.01; 0.05]
I² = 53%
−0.2
−0.1
0
0.1
0.2

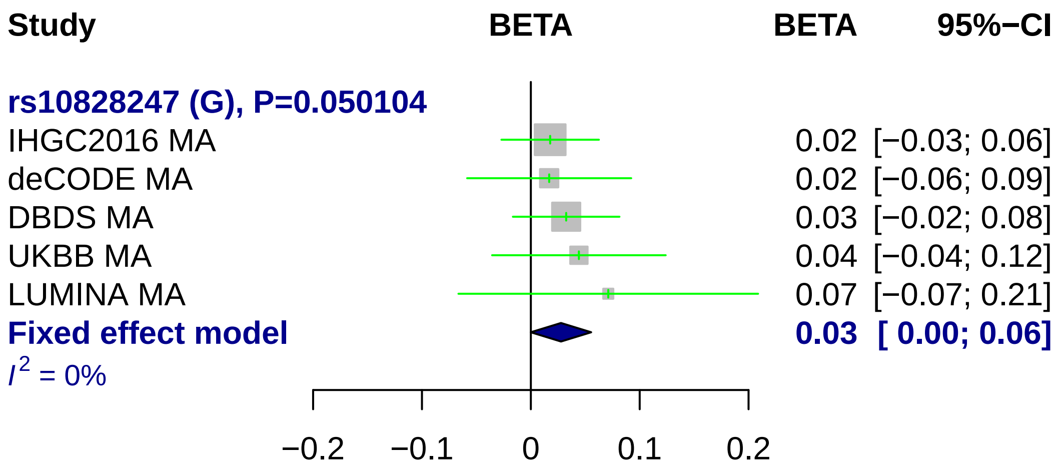
Study
BETA
BETA
95%−CI
rs10828247 (G), P=0.050104
IHGC2016 MA
0.02 [−0.03; 0.06]
deCODE MA
0.02 [−0.06; 0.09]
DBDS MA
0.03 [−0.02; 0.08]
UKBB MA
0.04 [−0.04; 0.12]
LUMINA MA
0.07 [−0.07; 0.21]
Fixed effect model
0.03 [ 0.00; 0.06]
I² = 0%
−0.2
−0.1
0
0.1
0.2

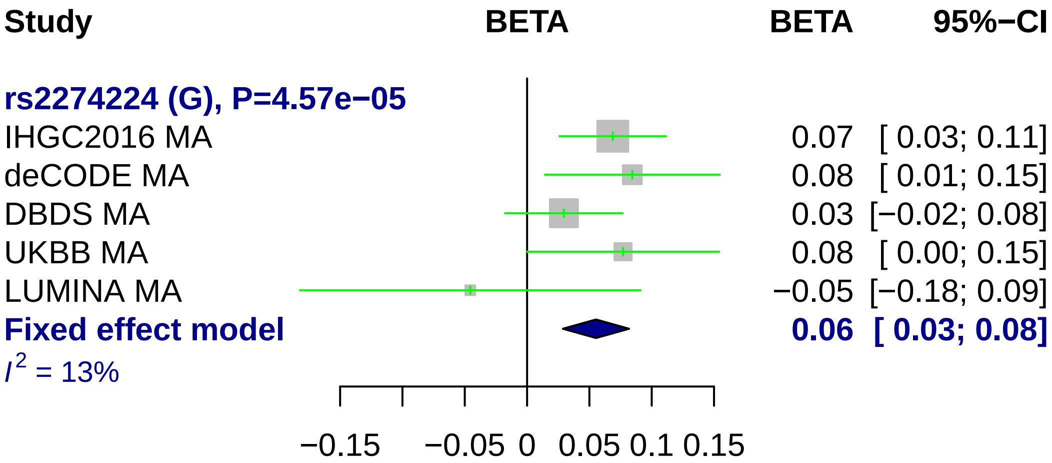
Study
BETA
BETA
95%−CI
rs2274224 (G), P=4.57e−05
IHGC2016 MA
0.07
[ 0.03; 0.11]
deCODE MA
0.08
[ 0.01; 0.15]
DBDS MA
0.03
[−0.02; 0.08]
UKBB MA
0.08
[ 0.00; 0.15]
LUMINA MA
−0.05
[−0.18; 0.09]
Fixed effect model
0.06
[ 0.03; 0.08]
$I^2$ = 13%
−0.15
−0.05
0
0.05
0.1
0.15

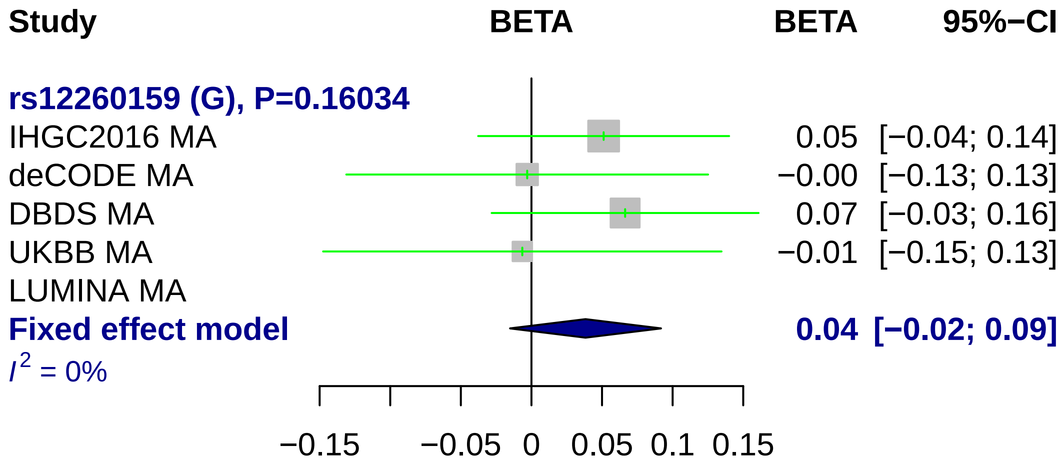
Study
BETA
BETA
95%−CI
rs12260159 (G), P=0.16034
IHGC2016 MA
0.05 [−0.04; 0.14]
deCODE MA
−0.00 [−0.13; 0.13]
DBDS MA
0.07 [−0.03; 0.16]
UKBB MA
−0.01 [−0.15; 0.13]
LUMINA MA
Fixed effect model
0.04 [−0.02; 0.09]
I^2 = 0%
−0.15
−0.05
0
0.05
0.1
0.15

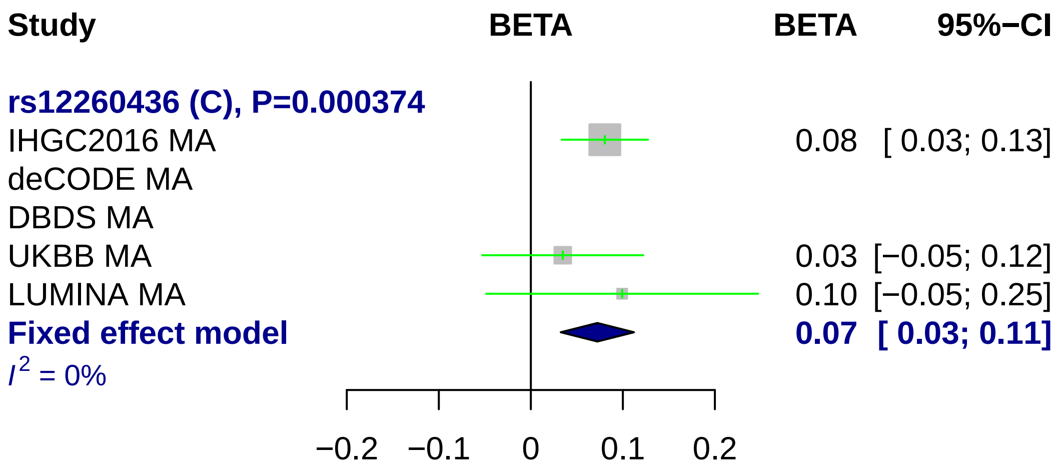
Study
BETA
BETA
95%−CI
rs12260436 (C), P=0.000374
IHGC2016 MA
0.08 [ 0.03; 0.13]
deCODE MA
DBDS MA
UKBB MA
0.03 [−0.05; 0.12]
LUMINA MA
0.10 [−0.05; 0.25]
Fixed effect model
0.07 [ 0.03; 0.11]
I2 = 0%
−0.2
−0.1
0
0.1
0.2

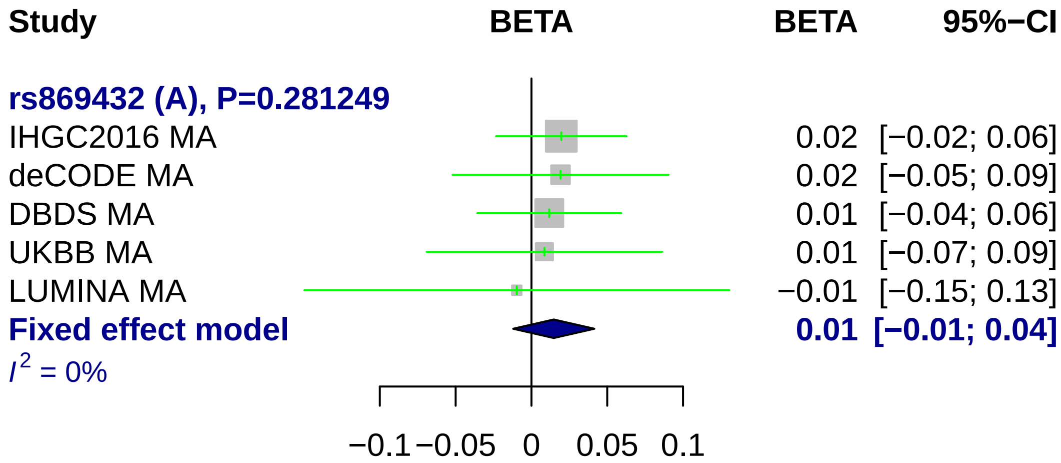
Study
BETA
BETA
95%−CI
rs869432 (A), P=0.281249
IHGC2016 MA
0.02
[−0.02; 0.06]
deCODE MA
0.02
[−0.05; 0.09]
DBDS MA
0.01
[−0.04; 0.06]
UKBB MA
0.01
[−0.07; 0.09]
LUMINA MA
−0.01
[−0.15; 0.13]
Fixed effect model
0.01
[−0.01; 0.04]
I2 = 0%
−0.1
−0.05
0
0.05
0.1

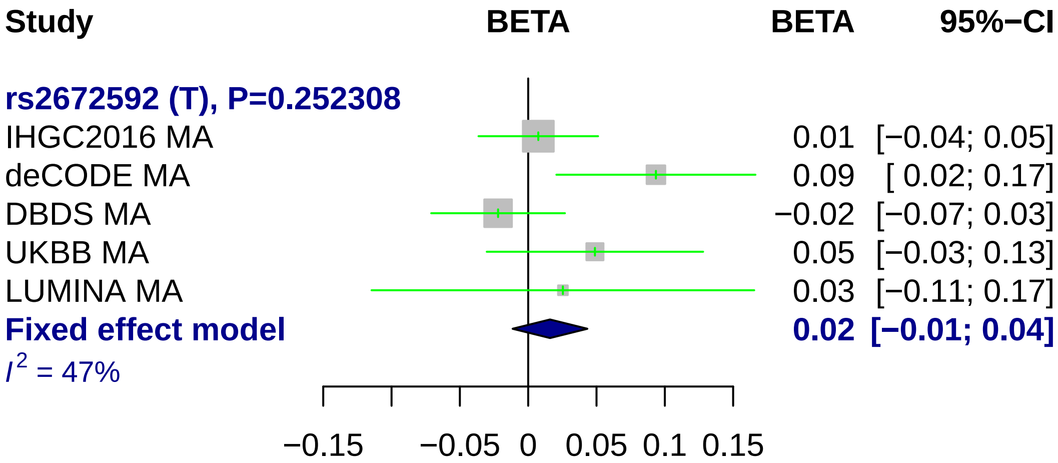
Study
BETA
BETA
95%−CI
rs2672592 (T), P=0.252308
IHGC2016 MA
0.01 [−0.04; 0.05]
deCODE MA
0.09 [ 0.02; 0.17]
DBDS MA
−0.02 [−0.07; 0.03]
UKBB MA
0.05 [−0.03; 0.13]
LUMINA MA
0.03 [−0.11; 0.17]
Fixed effect model
0.02 [−0.01; 0.04]
I^2 = 47%
−0.15
−0.05
0
0.05
0.1
0.15

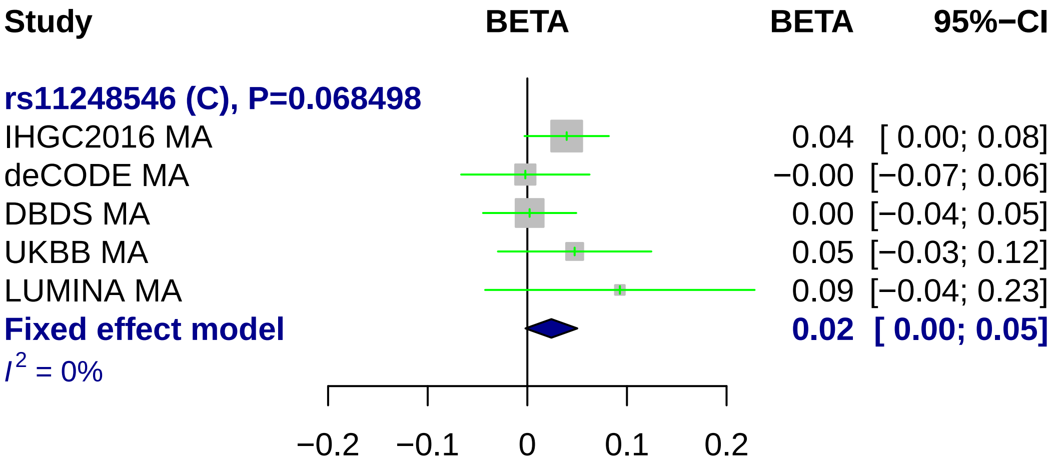
Study
BETA
BETA
95%−CI
rs11248546 (C), P=0.068498
IHGC2016 MA
0.04 [ 0.00; 0.08]
deCODE MA
−0.00 [−0.07; 0.06]
DBDS MA
0.00 [−0.04; 0.05]
UKBB MA
0.05 [−0.03; 0.12]
LUMINA MA
0.09 [−0.04; 0.23]
Fixed effect model
0.02 [ 0.00; 0.05]
I2 = 0%
−0.2
−0.1
0
0.1
0.2

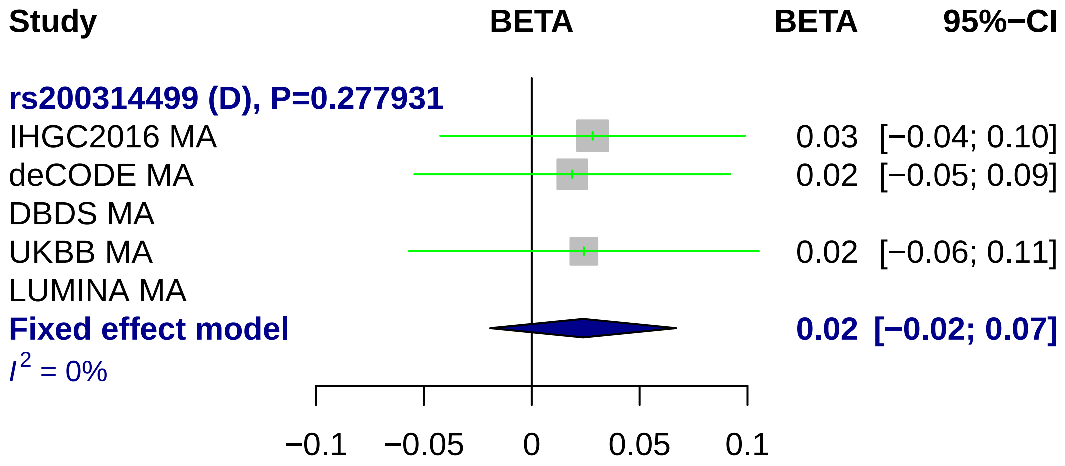
Study
BETA
BETA
95%−CI
rs200314499 (D), P=0.277931
IHGC2016 MA
0.03 [−0.04; 0.10]
deCODE MA
0.02 [−0.05; 0.09]
DBDS MA
UKBB MA
0.02 [−0.06; 0.11]
LUMINA MA
Fixed effect model
0.02 [−0.02; 0.07]
I2 = 0%
−0.1
−0.05
0
0.05
0.1

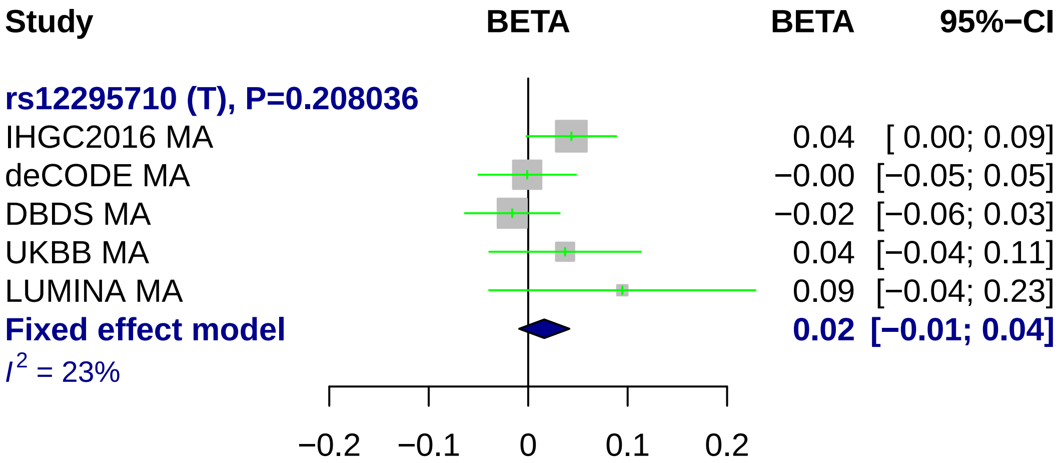
Study
BETA
BETA
95%−CI
rs12295710 (T), P=0.208036
IHGC2016 MA
0.04
[ 0.00; 0.09]
deCODE MA
−0.00
[−0.05; 0.05]
DBDS MA
−0.02
[−0.06; 0.03]
UKBB MA
0.04
[−0.04; 0.11]
LUMINA MA
0.09
[−0.04; 0.23]
Fixed effect model
0.02
[−0.01; 0.04]
I^2 = 23%
−0.2
−0.1
0
0.1
0.2

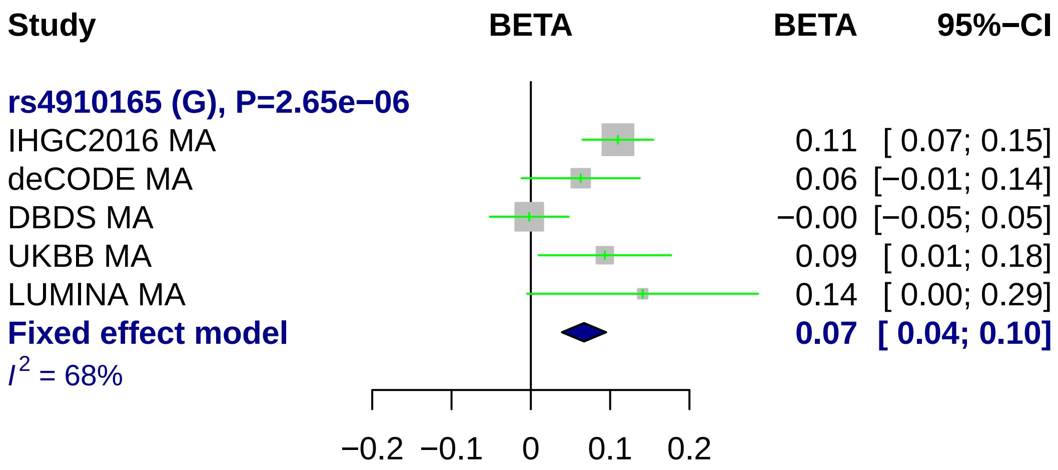
Study
BETA
BETA
95%−CI
rs4910165 (G), P=2.65e−06
IHGC2016 MA
0.11 [ 0.07; 0.15]
deCODE MA
0.06 [−0.01; 0.14]
DBDS MA
−0.00 [−0.05; 0.05]
UKBB MA
0.09 [ 0.01; 0.18]
LUMINA MA
0.14 [ 0.00; 0.29]
Fixed effect model
0.07 [ 0.04; 0.10]
I² = 68%
−0.2
−0.1
0
0.1
0.2

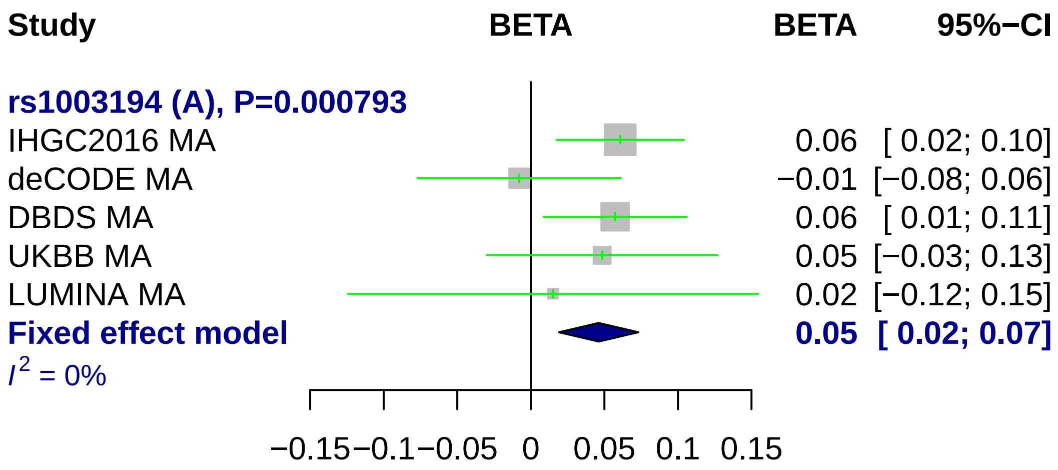
Study
BETA
BETA
95%−CI
rs1003194 (A), P=0.000793
IHGC2016 MA
0.06 [ 0.02; 0.10]
deCODE MA
−0.01 [−0.08; 0.06]
DBDS MA
0.06 [ 0.01; 0.11]
UKBB MA
0.05 [−0.03; 0.13]
LUMINA MA
0.02 [−0.12; 0.15]
Fixed effect model
0.05 [ 0.02; 0.07]
$I^2$ = 0%
−0.15 −0.1 −0.05 0 0.05 0.1 0.15

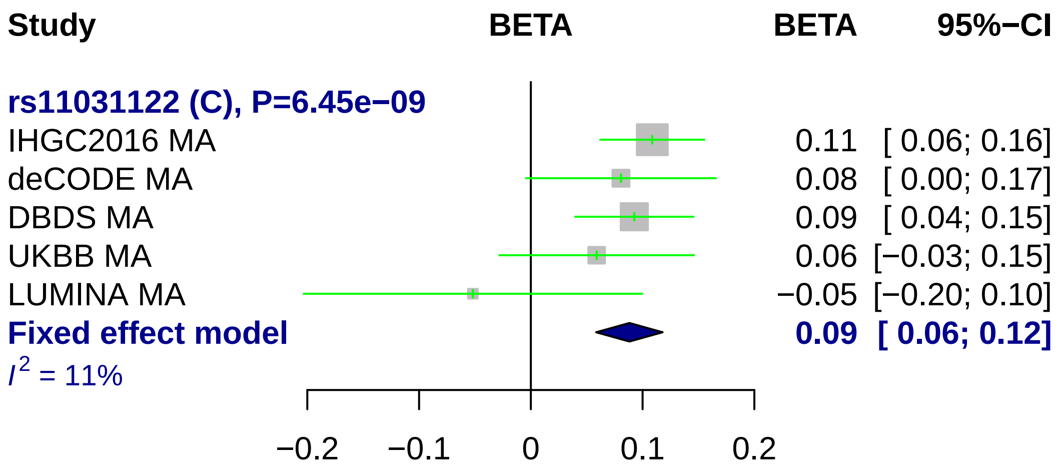
Study
BETA
BETA
95%−CI
rs11031122 (C), P=6.45e−09
IHGC2016 MA
0.11 [ 0.06; 0.16]
deCODE MA
0.08 [ 0.00; 0.17]
DBDS MA
0.09 [ 0.04; 0.15]
UKBB MA
0.06 [−0.03; 0.15]
LUMINA MA
−0.05 [−0.20; 0.10]
Fixed effect model
0.09 [ 0.06; 0.12]
$I^2$ = 11%
−0.2
−0.1
0
0.1
0.2

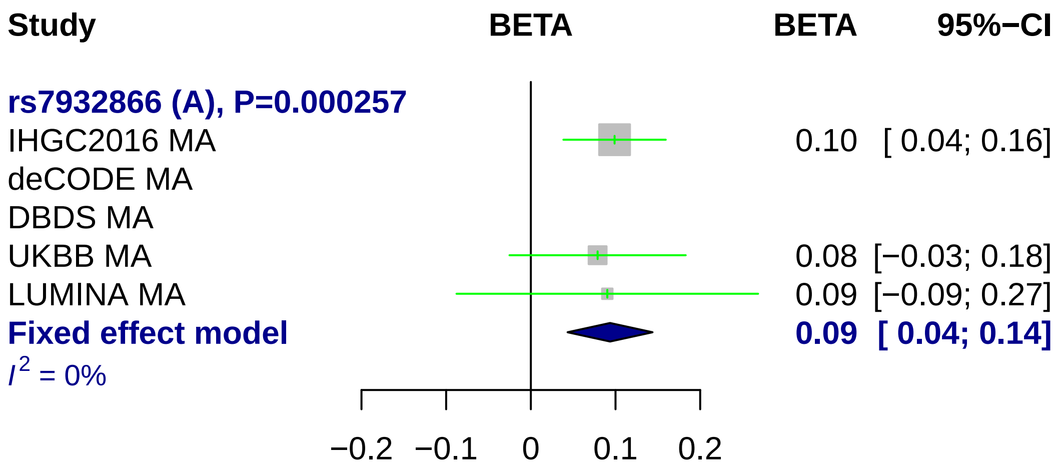

Study
BETA
BETA
95%−CI
rs7932866 (A), P=0.000257
IHGC2016 MA
0.10 [ 0.04; 0.16]
deCODE MA
DBDS MA
UKBB MA
0.08 [−0.03; 0.18]
LUMINA MA
0.09 [−0.09; 0.27]
Fixed effect model
0.09 [ 0.04; 0.14]
I2 = 0%
−0.2
−0.1
0
0.1
0.2

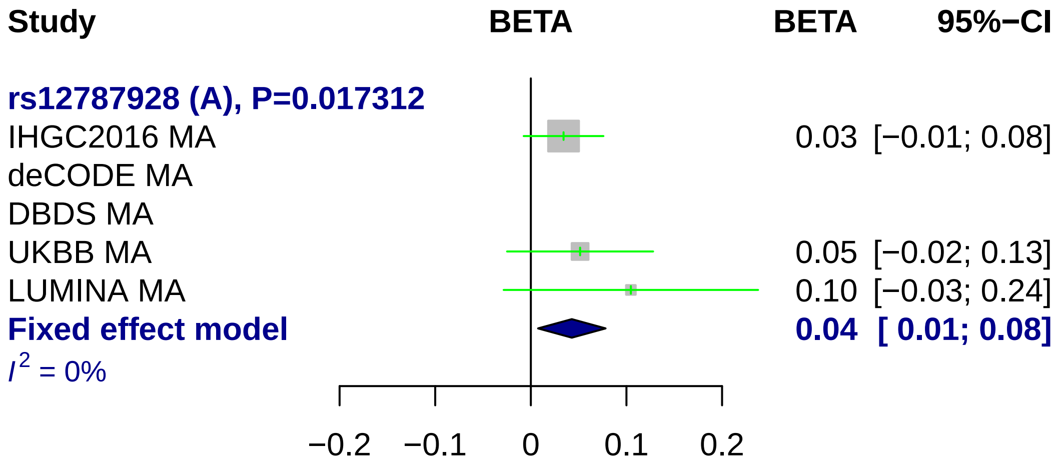
Study
BETA
BETA
95%−CI
rs12787928 (A), P=0.017312
IHGC2016 MA
0.03 [−0.01; 0.08]
deCODE MA
DBDS MA
UKBB MA
0.05 [−0.02; 0.13]
LUMINA MA
0.10 [−0.03; 0.24]
Fixed effect model
0.04 [ 0.01; 0.08]
I^2 = 0%
−0.2
−0.1
0
0.1
0.2

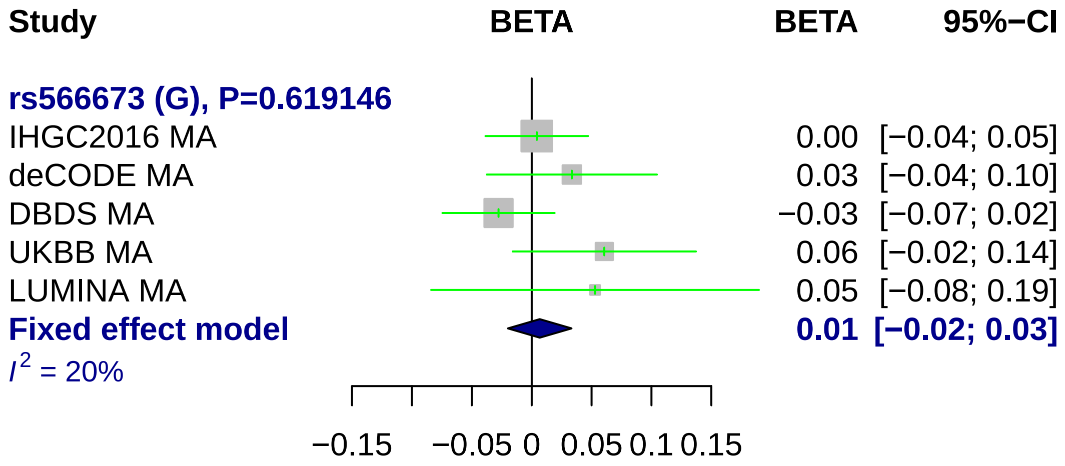

Study
BETA
BETA
95%−CI
rs566673 (G), P=0.619146
IHGC2016 MA
0.00 [−0.04; 0.05]
deCODE MA
0.03 [−0.04; 0.10]
DBDS MA
−0.03 [−0.07; 0.02]
UKBB MA
0.06 [−0.02; 0.14]
LUMINA MA
0.05 [−0.08; 0.19]
Fixed effect model
0.01 [−0.02; 0.03]
I2 = 20%
−0.15
−0.05
0
0.05
0.1
0.15

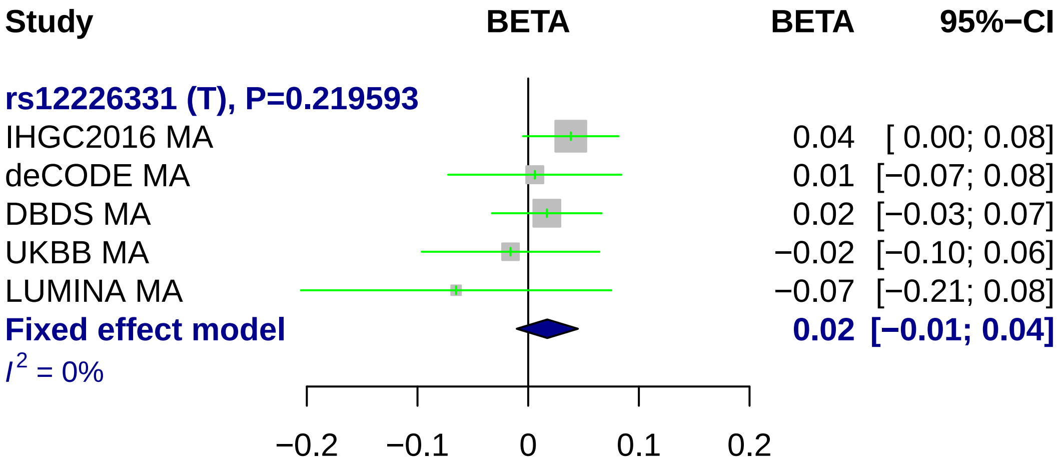

Study
BETA
BETA
95%−CI
rs12226331 (T), P=0.219593
IHGC2016 MA
0.04
[ 0.00; 0.08]
deCODE MA
0.01
[−0.07; 0.08]
DBDS MA
0.02
[−0.03; 0.07]
UKBB MA
−0.02
[−0.10; 0.06]
LUMINA MA
−0.07
[−0.21; 0.08]
Fixed effect model
0.02
[−0.01; 0.04]
I2 = 0%
−0.2
−0.1
0
0.1
0.2

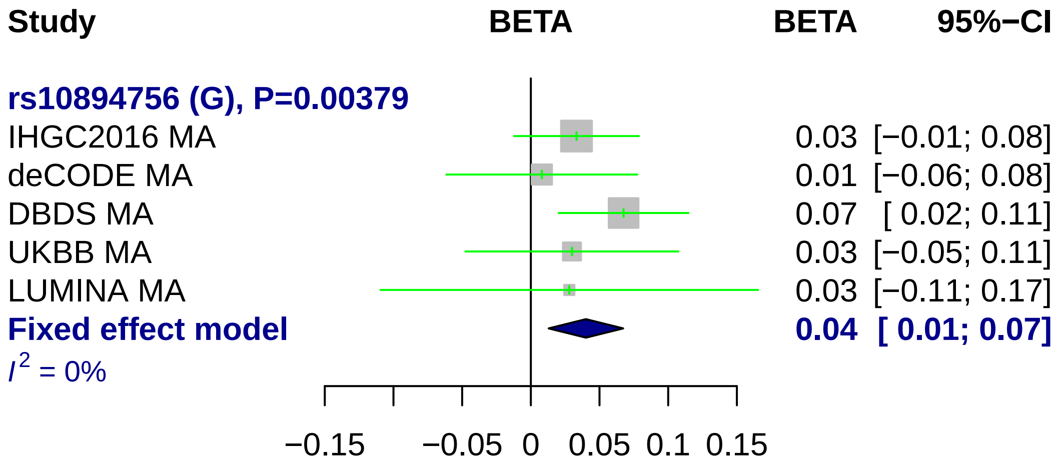

Study
BETA
BETA
95%−CI
rs10894756 (G), P=0.00379
IHGC2016 MA
0.03 [−0.01; 0.08]
deCODE MA
0.01 [−0.06; 0.08]
DBDS MA
0.07 [ 0.02; 0.11]
UKBB MA
0.03 [−0.05; 0.11]
LUMINA MA
0.03 [−0.11; 0.17]
Fixed effect model
0.04 [ 0.01; 0.07]
I2 = 0%
−0.15
−0.05
0
0.05
0.1
0.15

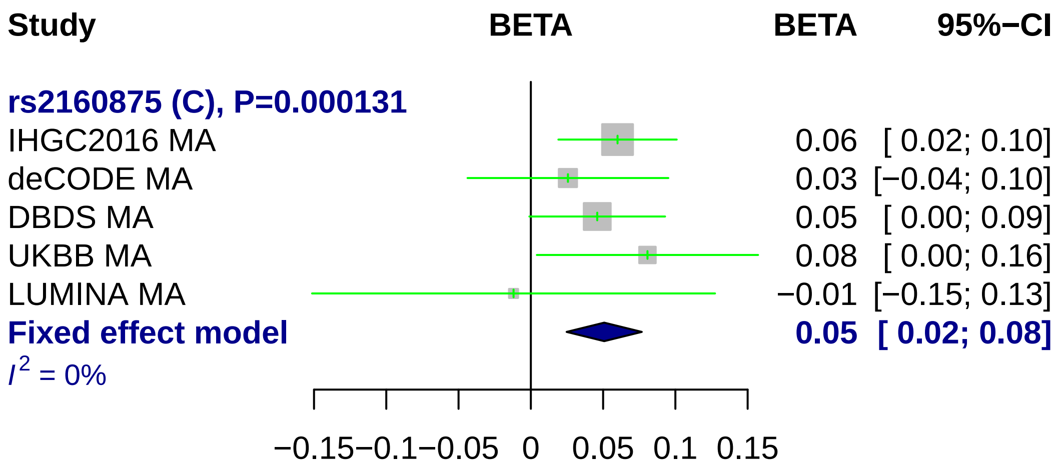
Study
BETA
BETA
95%−CI
rs2160875 (C), P=0.000131
IHGC2016 MA
0.06
[ 0.02; 0.10]
deCODE MA
0.03
[−0.04; 0.10]
DBDS MA
0.05
[ 0.00; 0.09]
UKBB MA
0.08
[ 0.00; 0.16]
LUMINA MA
−0.01
[−0.15; 0.13]
Fixed effect model
0.05
[ 0.02; 0.08]
I2 = 0%
−0.15
−0.1
−0.05
0
0.05
0.1
0.15

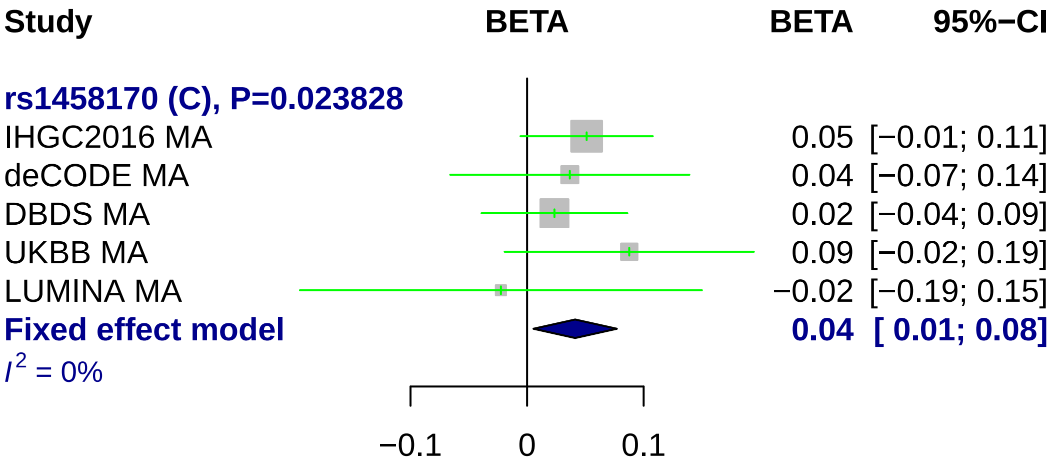
Study
BETA
BETA
95%−CI
rs1458170 (C), P=0.023828
IHGC2016 MA
0.05 [−0.01; 0.11]
deCODE MA
0.04 [−0.07; 0.14]
DBDS MA
0.02 [−0.04; 0.09]
UKBB MA
0.09 [−0.02; 0.19]
LUMINA MA
−0.02 [−0.19; 0.15]
Fixed effect model
0.04 [ 0.01; 0.08]
I² = 0%
−0.1
0
0.1

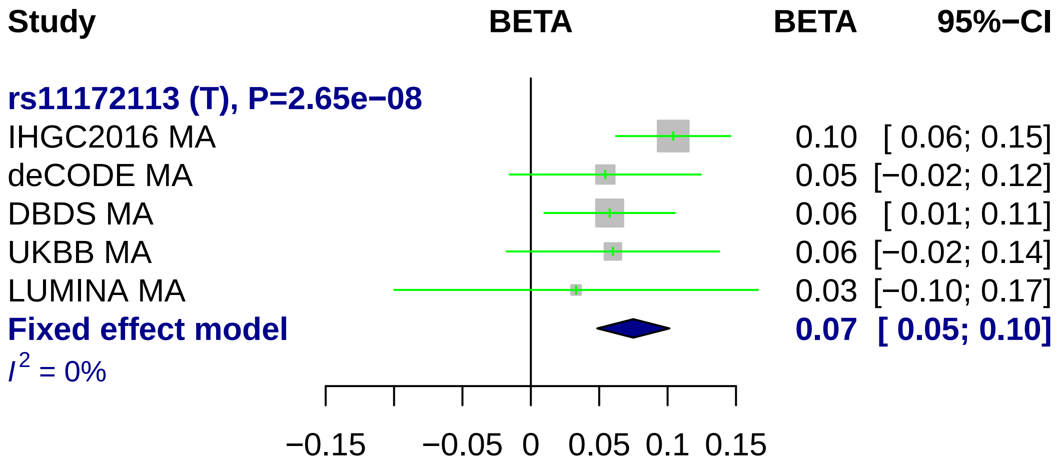

Study
BETA
BETA
95%−CI
rs11172113 (T), P=2.65e−08
IHGC2016 MA
0.10 [ 0.06; 0.15]
deCODE MA
0.05 [−0.02; 0.12]
DBDS MA
0.06 [ 0.01; 0.11]
UKBB MA
0.06 [−0.02; 0.14]
LUMINA MA
0.03 [−0.10; 0.17]
Fixed effect model
0.07 [ 0.05; 0.10]
I2 = 0%
−0.15
−0.05
0
0.05
0.1
0.15

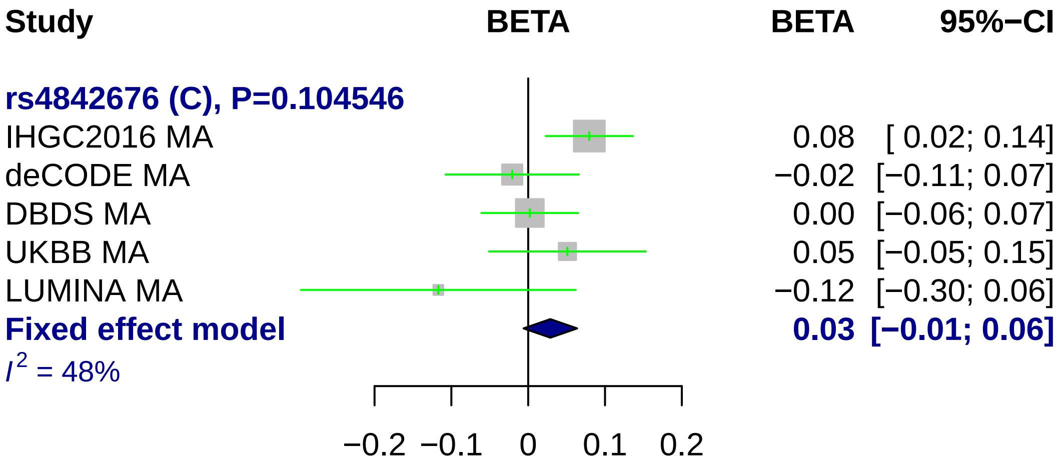
Study
BETA
BETA
95%−CI
rs4842676 (C), P=0.104546
IHGC2016 MA
0.08
[ 0.02; 0.14]
deCODE MA
−0.02
[−0.11; 0.07]
DBDS MA
0.00
[−0.06; 0.07]
UKBB MA
0.05
[−0.05; 0.15]
LUMINA MA
−0.12
[−0.30; 0.06]
Fixed effect model
0.03
[−0.01; 0.06]
I² = 48%
−0.2
−0.1
0
0.1
0.2

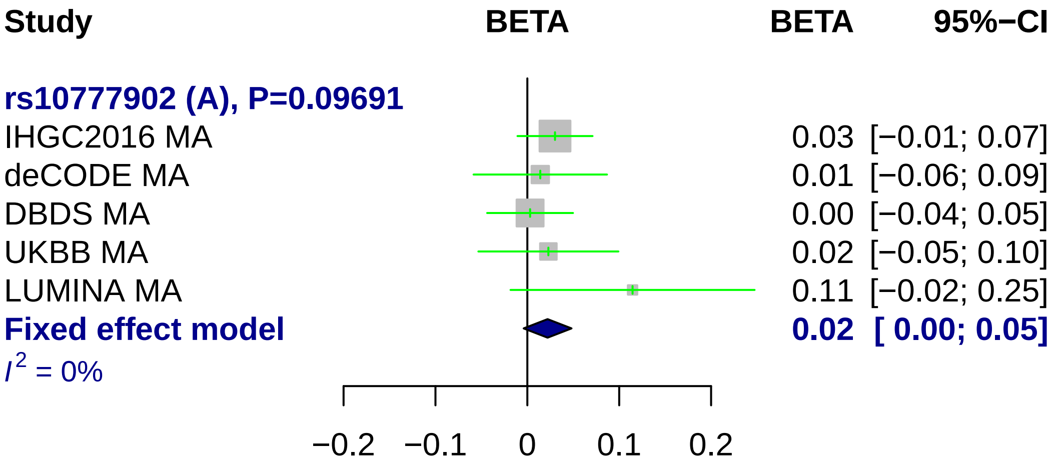
Study
BETA
BETA
95%−CI
rs10777902 (A), P=0.09691
IHGC2016 MA
0.03 [−0.01; 0.07]
deCODE MA
0.01 [−0.06; 0.09]
DBDS MA
0.00 [−0.04; 0.05]
UKBB MA
0.02 [−0.05; 0.10]
LUMINA MA
0.11 [−0.02; 0.25]
Fixed effect model
0.02 [ 0.00; 0.05]
I2 = 0%
−0.2
−0.1
0
0.1
0.2

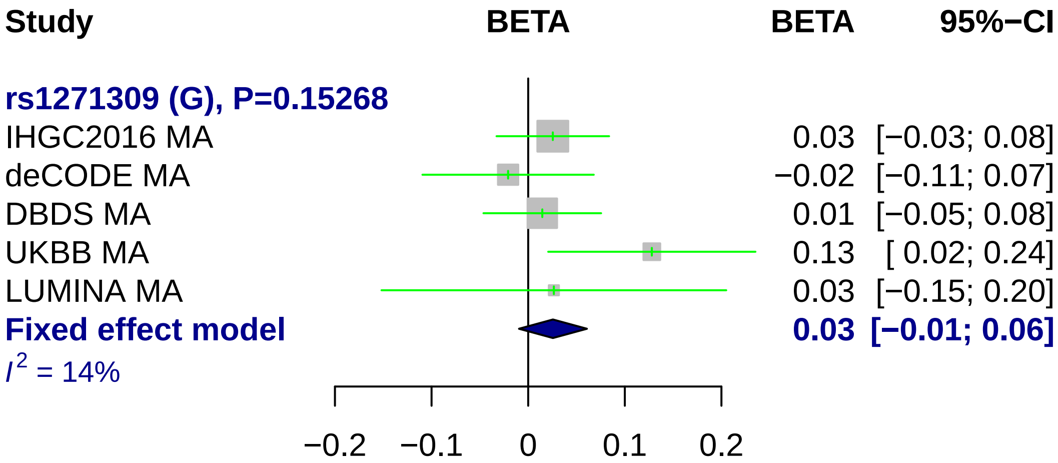

Study
BETA
BETA
95%−CI
rs1271309 (G), P=0.15268
IHGC2016 MA
0.03 [−0.03; 0.08]
deCODE MA
−0.02 [−0.11; 0.07]
DBDS MA
0.01 [−0.05; 0.08]
UKBB MA
0.13 [ 0.02; 0.24]
LUMINA MA
0.03 [−0.15; 0.20]
Fixed effect model
0.03 [−0.01; 0.06]
I2 = 14%
−0.2
−0.1
0
0.1
0.2

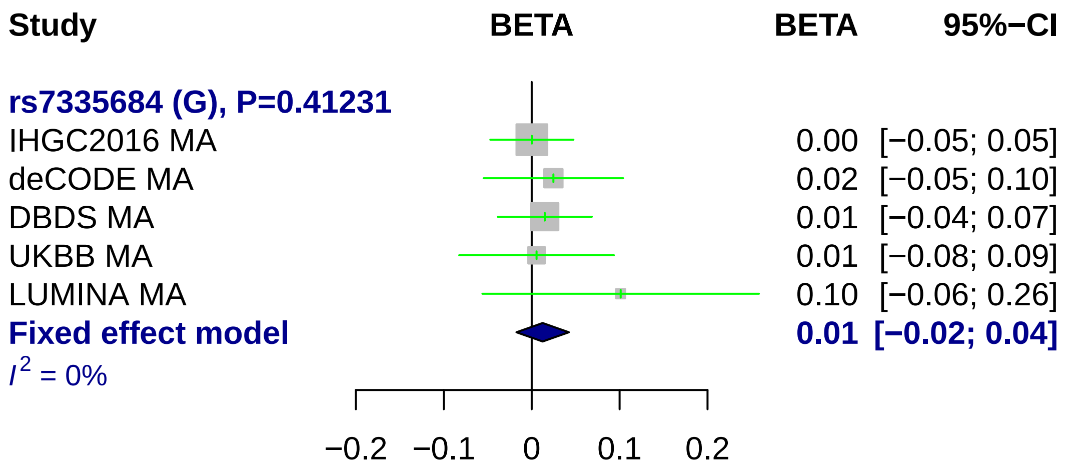
Study
BETA
BETA
95%−CI
rs7335684 (G), P=0.41231
IHGC2016 MA
0.00 [−0.05; 0.05]
deCODE MA
0.02 [−0.05; 0.10]
DBDS MA
0.01 [−0.04; 0.07]
UKBB MA
0.01 [−0.08; 0.09]
LUMINA MA
0.10 [−0.06; 0.26]
Fixed effect model
0.01 [−0.02; 0.04]
I2 = 0%
−0.2
−0.1
0
0.1
0.2

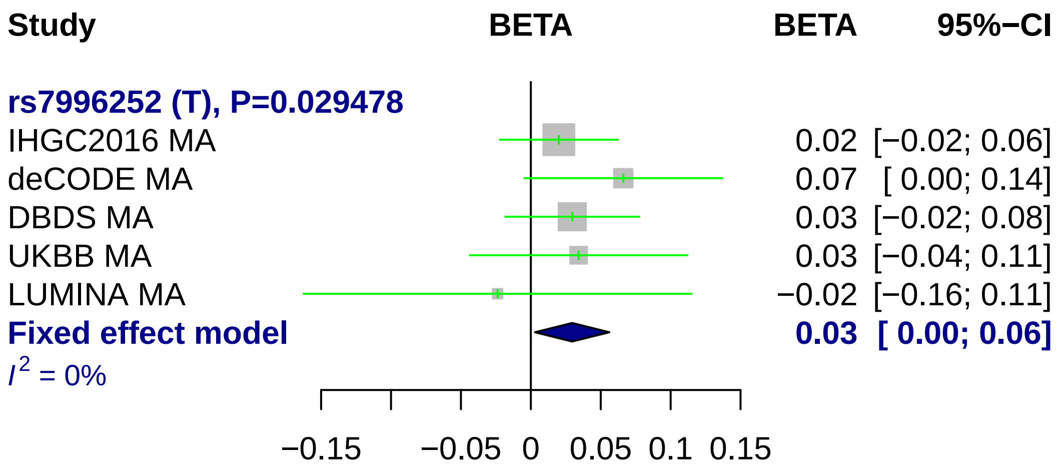

Study
BETA
BETA
95%−CI
rs7996252 (T), P=0.029478
IHGC2016 MA
0.02 [−0.02; 0.06]
deCODE MA
0.07 [ 0.00; 0.14]
DBDS MA
0.03 [−0.02; 0.08]
UKBB MA
0.03 [−0.04; 0.11]
LUMINA MA
−0.02 [−0.16; 0.11]
Fixed effect model
0.03 [ 0.00; 0.06]
I2 = 0%
−0.15
−0.05
0
0.05
0.1
0.15

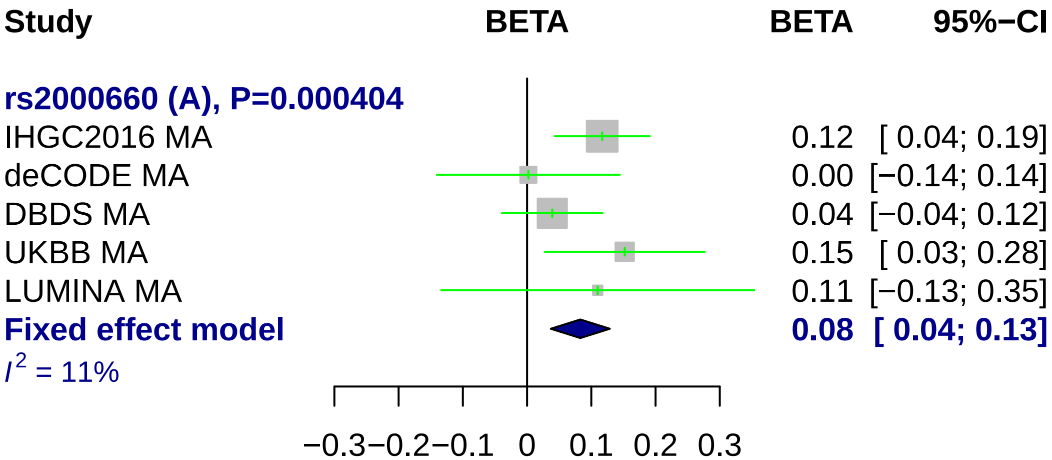
Study
BETA
BETA
95%−CI
rs2000660 (A), P=0.000404
IHGC2016 MA
0.12 [ 0.04; 0.19]
deCODE MA
0.00 [−0.14; 0.14]
DBDS MA
0.04 [−0.04; 0.12]
UKBB MA
0.15 [ 0.03; 0.28]
LUMINA MA
0.11 [−0.13; 0.35]
Fixed effect model
0.08 [ 0.04; 0.13]
I2 = 11%
−0.3 −0.2 −0.1 0 0.1 0.2 0.3

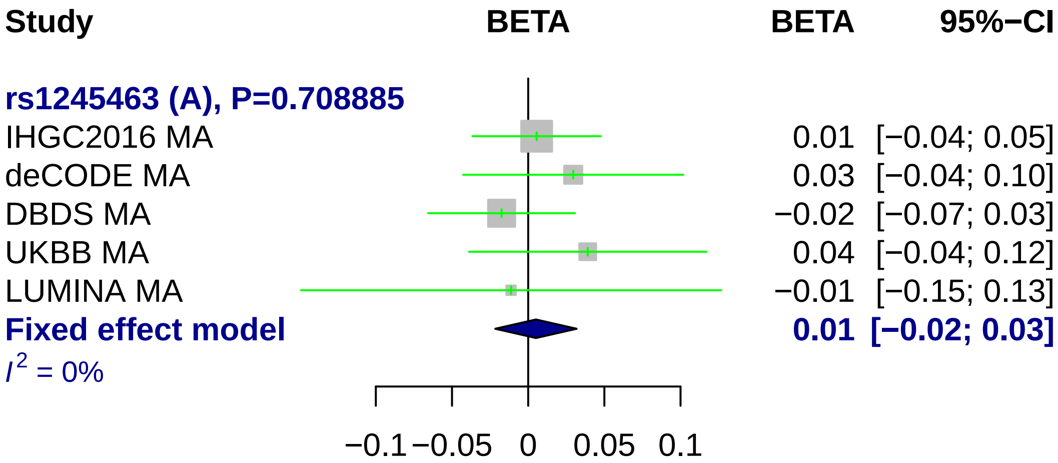
Study
BETA
BETA
95%−CI
rs1245463 (A), P=0.708885
IHGC2016 MA 0.01 [−0.04; 0.05]
deCODE MA 0.03 [−0.04; 0.10]
DBDS MA −0.02 [−0.07; 0.03]
UKBB MA 0.04 [−0.04; 0.12]
LUMINA MA −0.01 [−0.15; 0.13]
Fixed effect model 0.01 [−0.02; 0.03]
I2 = 0%
−0.1
−0.05
0
0.05
0.1

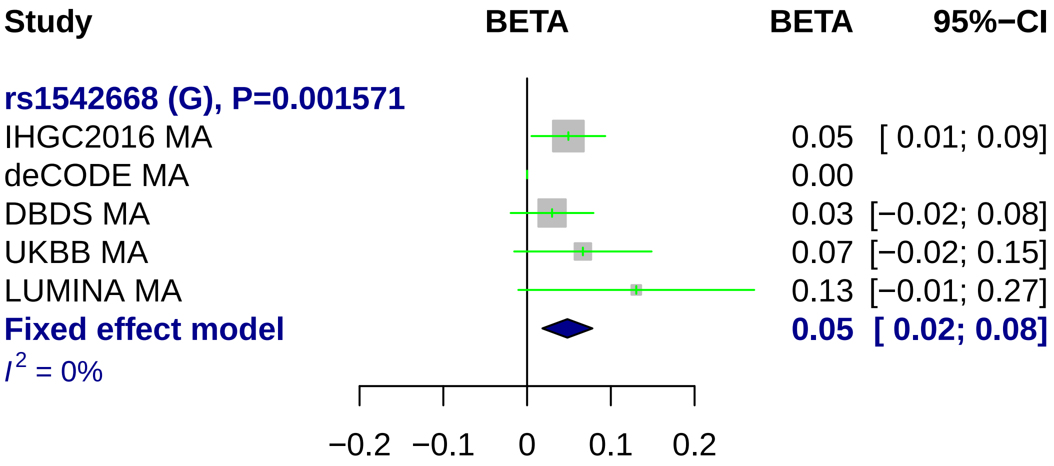
Study
BETA
BETA
95%−CI
rs1542668 (G), P=0.001571
IHGC2016 MA
0.05
[ 0.01; 0.09]
deCODE MA
0.00
DBDS MA
0.03
[−0.02; 0.08]
UKBB MA
0.07
[−0.02; 0.15]
LUMINA MA
0.13
[−0.01; 0.27]
Fixed effect model
0.05
[ 0.02; 0.08]
I^2 = 0%
−0.2
−0.1
0
0.1
0.2

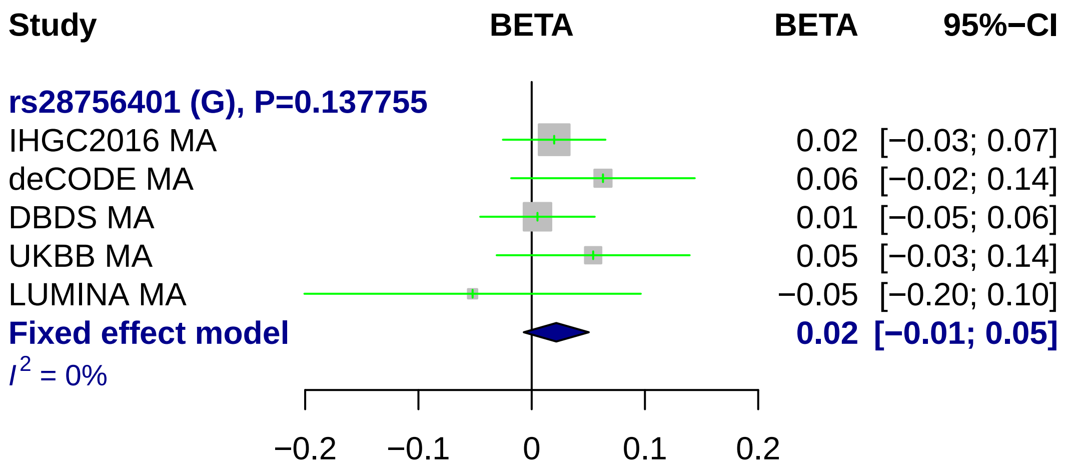

Study
BETA
BETA
95%−CI
rs28756401 (G), P=0.137755
IHGC2016 MA 0.02 [−0.03; 0.07]
deCODE MA 0.06 [−0.02; 0.14]
DBDS MA 0.01 [−0.05; 0.06]
UKBB MA 0.05 [−0.03; 0.14]
LUMINA MA −0.05 [−0.20; 0.10]
Fixed effect model 0.02 [−0.01; 0.05]
I2 = 0%
−0.2
−0.1
0
0.1
0.2

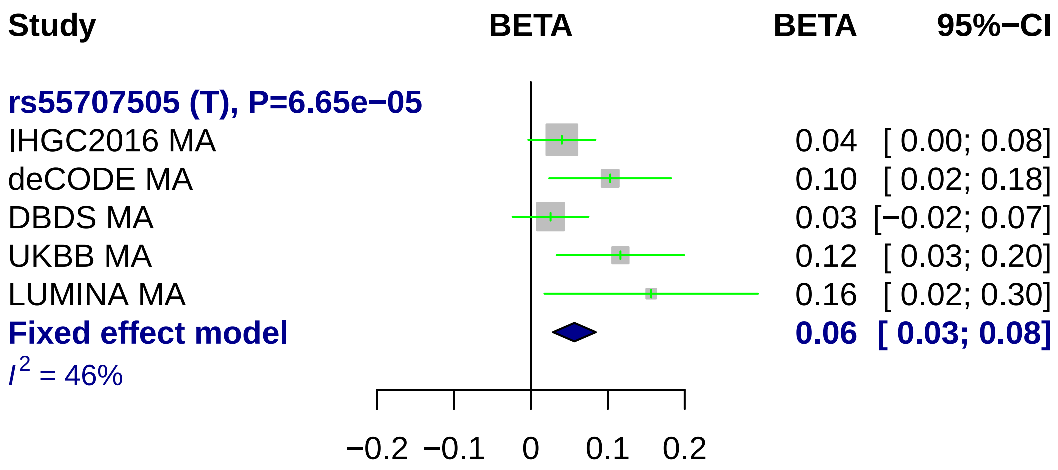
Study
BETA
BETA
95%−CI
rs55707505 (T), P=6.65e−05
IHGC2016 MA
0.04 [ 0.00; 0.08]
deCODE MA
0.10 [ 0.02; 0.18]
DBDS MA
0.03 [−0.02; 0.07]
UKBB MA
0.12 [ 0.03; 0.20]
LUMINA MA
0.16 [ 0.02; 0.30]
Fixed effect model
0.06 [ 0.03; 0.08]
I2 = 46%
−0.2
−0.1
0
0.1
0.2

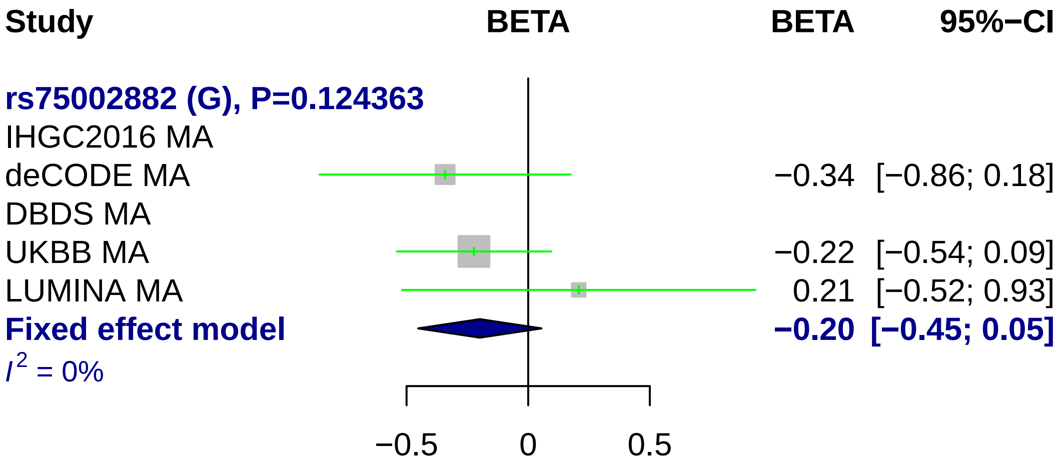
Study
BETA
BETA
95%−CI
rs75002882 (G), P=0.124363
IHGC2016 MA
deCODE MA
−0.34
[−0.86; 0.18]
DBDS MA
UKBB MA
−0.22
[−0.54; 0.09]
LUMINA MA
0.21
[−0.52; 0.93]
Fixed effect model
−0.20
[−0.45; 0.05]
I2 = 0%
−0.5
0
0.5

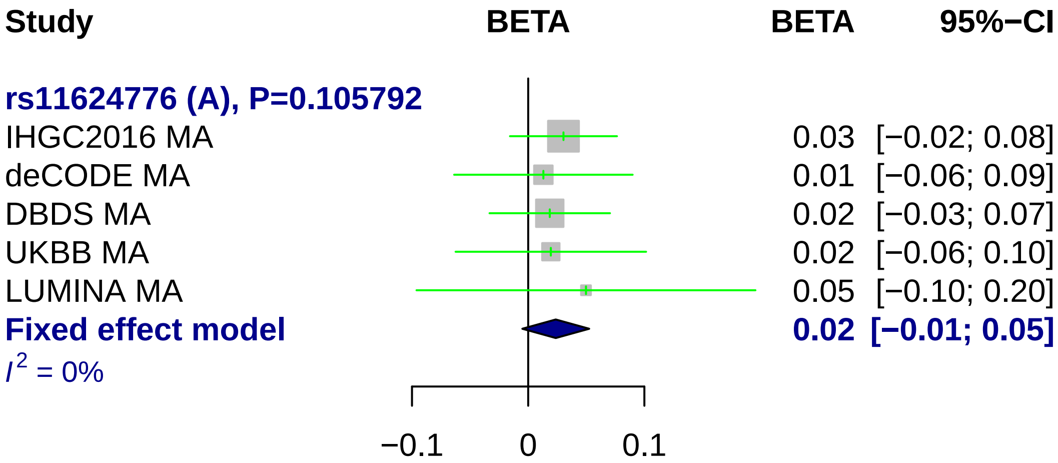

Study
BETA
BETA
95%−CI
rs11624776 (A), P=0.105792
IHGC2016 MA 0.03 [−0.02; 0.08]
deCODE MA 0.01 [−0.06; 0.09]
DBDS MA 0.02 [−0.03; 0.07]
UKBB MA 0.02 [−0.06; 0.10]
LUMINA MA 0.05 [−0.10; 0.20]
Fixed effect model 0.02 [−0.01; 0.05]
I² = 0%
−0.1
0
0.1

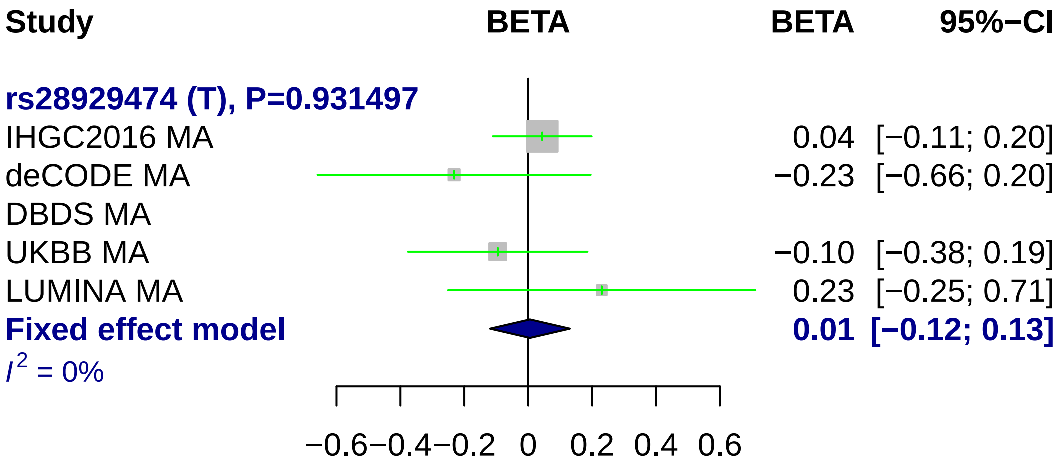
Study
BETA
BETA
95%−CI
rs28929474 (T), P=0.931497
IHGC2016 MA 0.04 [−0.11; 0.20]
deCODE MA −0.23 [−0.66; 0.20]
DBDS MA
UKBB MA −0.10 [−0.38; 0.19]
LUMINA MA 0.23 [−0.25; 0.71]
Fixed effect model 0.01 [−0.12; 0.13]
I2 = 0%
−0.6 −0.4 −0.2 0 0.2 0.4 0.6

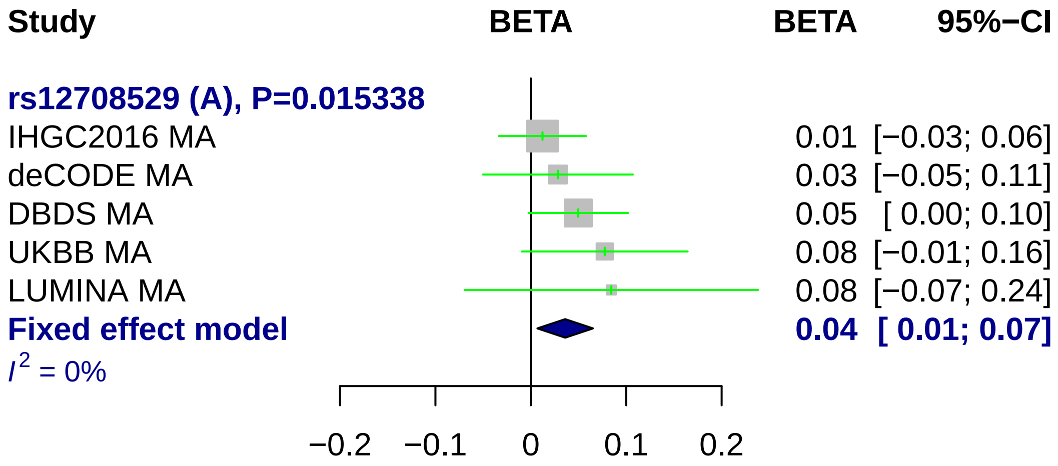
Study
BETA
BETA
95%−CI
rs12708529 (A), P=0.015338
IHGC2016 MA
0.01 [−0.03; 0.06]
deCODE MA
0.03 [−0.05; 0.11]
DBDS MA
0.05 [ 0.00; 0.10]
UKBB MA
0.08 [−0.01; 0.16]
LUMINA MA
0.08 [−0.07; 0.24]
Fixed effect model
0.04 [ 0.01; 0.07]
I^2 = 0%
−0.2
−0.1
0
0.1
0.2

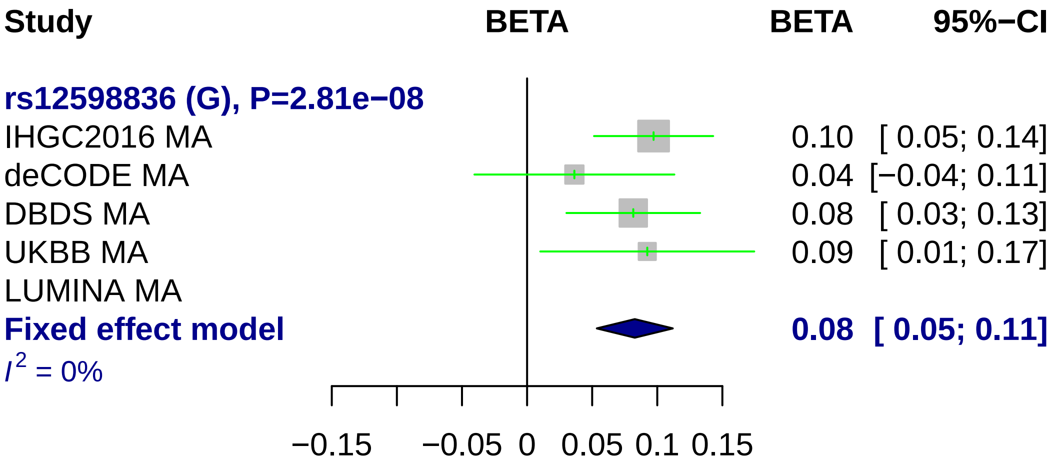
Study
BETA
BETA
95%−CI
rs12598836 (G), P=2.81e−08
IHGC2016 MA
0.10 [ 0.05; 0.14]
deCODE MA
0.04 [−0.04; 0.11]
DBDS MA
0.08 [ 0.03; 0.13]
UKBB MA
0.09 [ 0.01; 0.17]
LUMINA MA
Fixed effect model
0.08 [ 0.05; 0.11]
I^2 = 0%
−0.15
−0.05
0
0.05
0.1
0.15

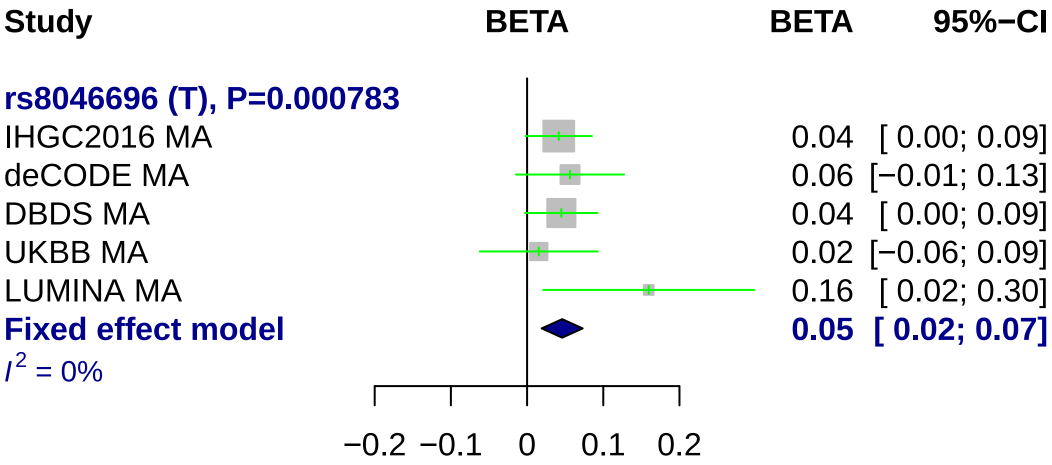
Study
BETA
BETA
95%−CI
rs8046696 (T), P=0.000783
IHGC2016 MA
0.04 [ 0.00; 0.09]
deCODE MA
0.06 [−0.01; 0.13]
DBDS MA
0.04 [ 0.00; 0.09]
UKBB MA
0.02 [−0.06; 0.09]
LUMINA MA
0.16 [ 0.02; 0.30]
Fixed effect model
0.05 [ 0.02; 0.07]
I2 = 0%
−0.2 −0.1 0 0.1 0.2

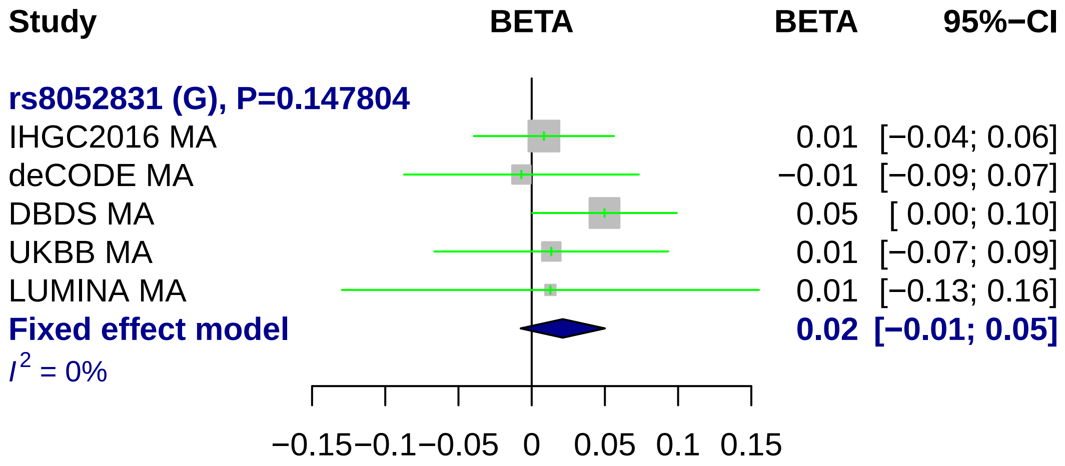
Study
BETA
BETA
95%−CI
rs8052831 (G), P=0.147804
IHGC2016 MA
0.01 [−0.04; 0.06]
deCODE MA
−0.01 [−0.09; 0.07]
DBDS MA
0.05 [ 0.00; 0.10]
UKBB MA
0.01 [−0.07; 0.09]
LUMINA MA
0.01 [−0.13; 0.16]
Fixed effect model
0.02 [−0.01; 0.05]
I2 = 0%
−0.15 −0.1 −0.05 0 0.05 0.1 0.15

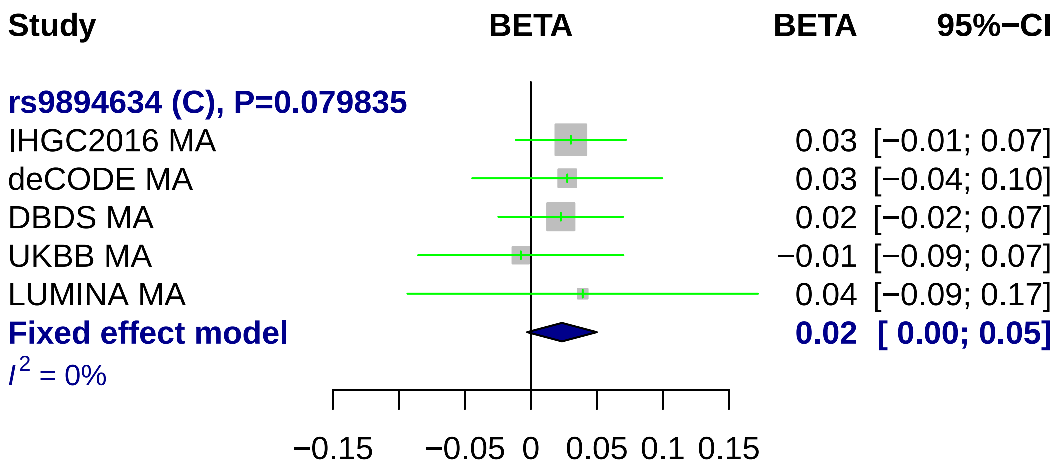
Study
BETA
BETA
95%−CI
rs9894634 (C), P=0.079835
IHGC2016 MA
0.03 [−0.01; 0.07]
deCODE MA
0.03 [−0.04; 0.10]
DBDS MA
0.02 [−0.02; 0.07]
UKBB MA
−0.01 [−0.09; 0.07]
LUMINA MA
0.04 [−0.09; 0.17]
Fixed effect model
0.02 [ 0.00; 0.05]
I2 = 0%
−0.15
−0.05
0
0.05
0.1
0.15

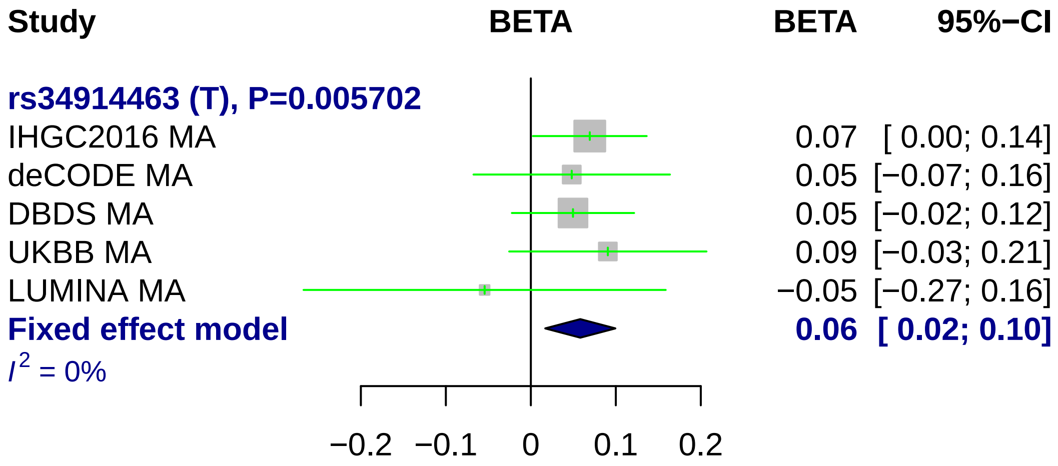
Study
BETA
BETA
95%−CI
rs34914463 (T), P=0.005702
IHGC2016 MA
0.07 [ 0.00; 0.14]
deCODE MA
0.05 [−0.07; 0.16]
DBDS MA
0.05 [−0.02; 0.12]
UKBB MA
0.09 [−0.03; 0.21]
LUMINA MA
−0.05 [−0.27; 0.16]
Fixed effect model
0.06 [ 0.02; 0.10]
I2 = 0%
−0.2
−0.1
0
0.1
0.2

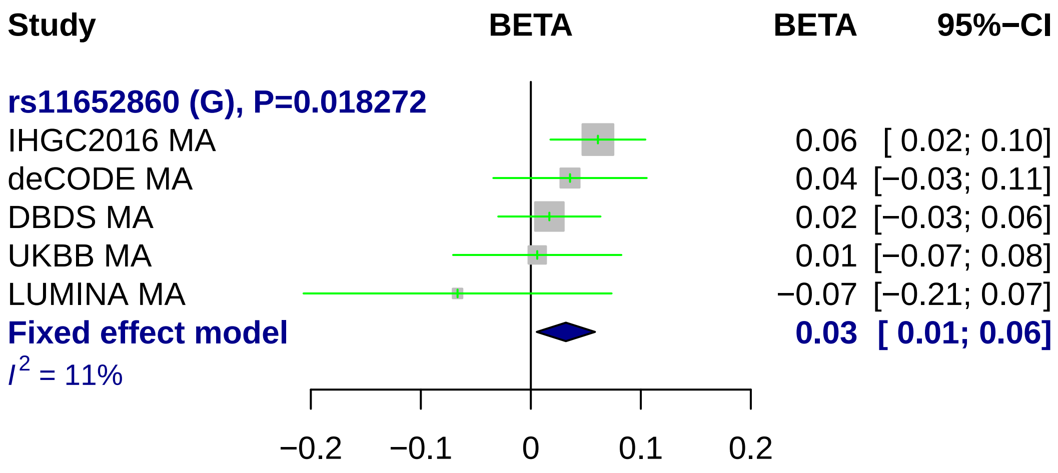
Study
BETA
BETA
95%−CI
rs11652860 (G), P=0.018272
IHGC2016 MA
0.06 [ 0.02; 0.10]
deCODE MA
0.04 [−0.03; 0.11]
DBDS MA
0.02 [−0.03; 0.06]
UKBB MA
0.01 [−0.07; 0.08]
LUMINA MA
−0.07 [−0.21; 0.07]
Fixed effect model
0.03 [ 0.01; 0.06]
I2 = 11%
−0.2
−0.1
0
0.1
0.2

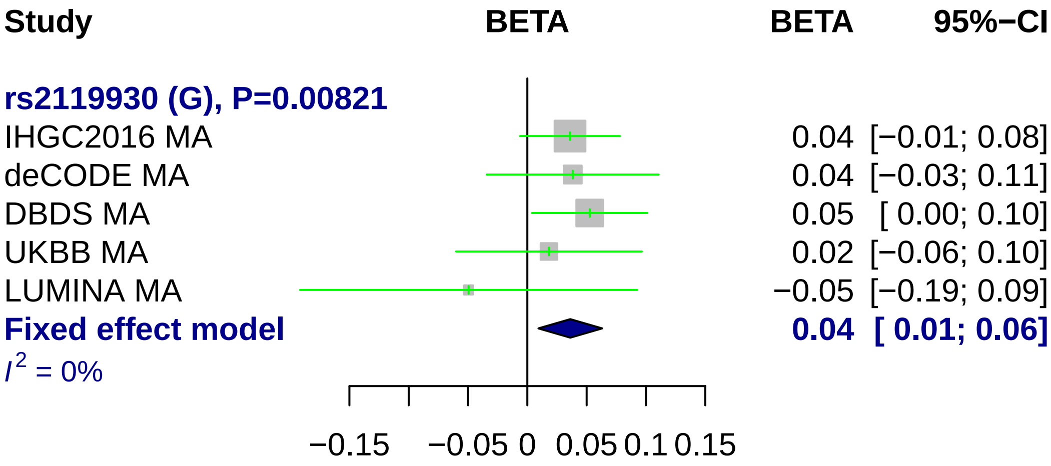
Study
BETA
BETA
95%−CI
rs2119930 (G), P=0.00821
IHGC2016 MA
0.04 [−0.01; 0.08]
deCODE MA
0.04 [−0.03; 0.11]
DBDS MA
0.05 [ 0.00; 0.10]
UKBB MA
0.02 [−0.06; 0.10]
LUMINA MA
−0.05 [−0.19; 0.09]
Fixed effect model
0.04 [ 0.01; 0.06]
I² = 0%
−0.15
−0.05
0
0.05
0.1
0.15

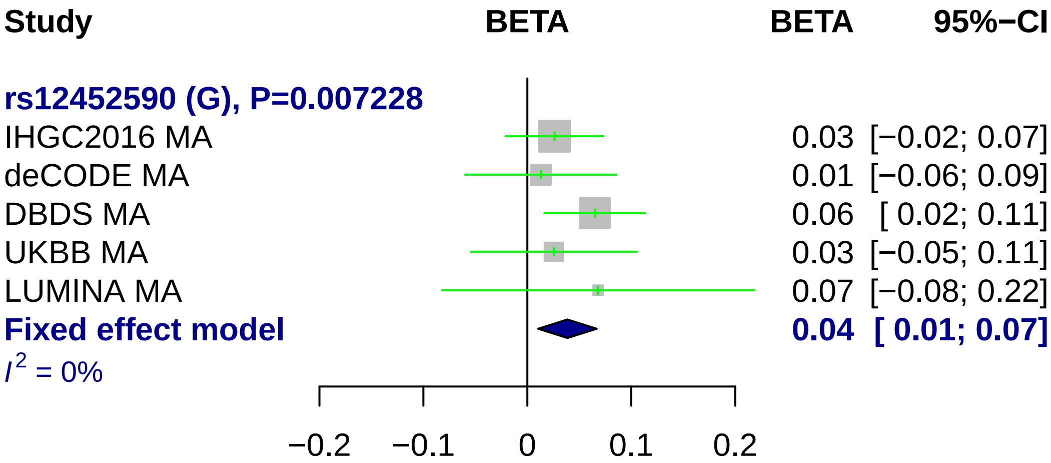
Study
BETA
BETA
95%−CI
rs12452590 (G), P=0.007228
IHGC2016 MA
0.03 [−0.02; 0.07]
deCODE MA
0.01 [−0.06; 0.09]
DBDS MA
0.06 [ 0.02; 0.11]
UKBB MA
0.03 [−0.05; 0.11]
LUMINA MA
0.07 [−0.08; 0.22]
Fixed effect model
0.04 [ 0.01; 0.07]
I^2 = 0%
−0.2
−0.1
0
0.1
0.2

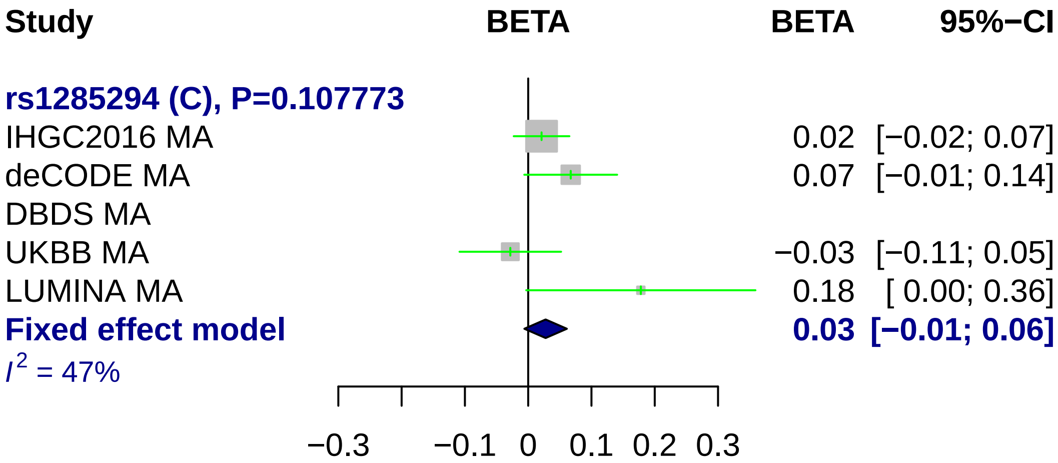
Study
BETA
BETA
95%−CI
rs1285294 (C), P=0.107773
IHGC2016 MA
0.02
[−0.02; 0.07]
deCODE MA
0.07
[−0.01; 0.14]
DBDS MA
UKBB MA
−0.03
[−0.11; 0.05]
LUMINA MA
0.18
[ 0.00; 0.36]
Fixed effect model
0.03
[−0.01; 0.06]
I2 = 47%
−0.3
−0.1
0
0.1
0.2
0.3

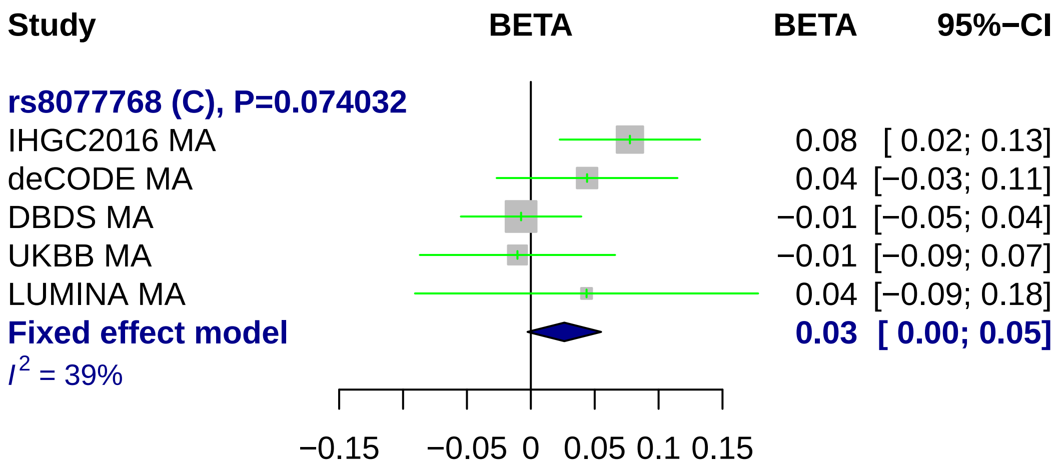
Study
BETA
BETA
95%−CI
rs8077768 (C), P=0.074032
IHGC2016 MA
0.08 [ 0.02; 0.13]
deCODE MA
0.04 [−0.03; 0.11]
DBDS MA
−0.01 [−0.05; 0.04]
UKBB MA
−0.01 [−0.09; 0.07]
LUMINA MA
0.04 [−0.09; 0.18]
Fixed effect model
0.03 [ 0.00; 0.05]
I2 = 39%
−0.15
−0.05
0
0.05
0.1
0.15

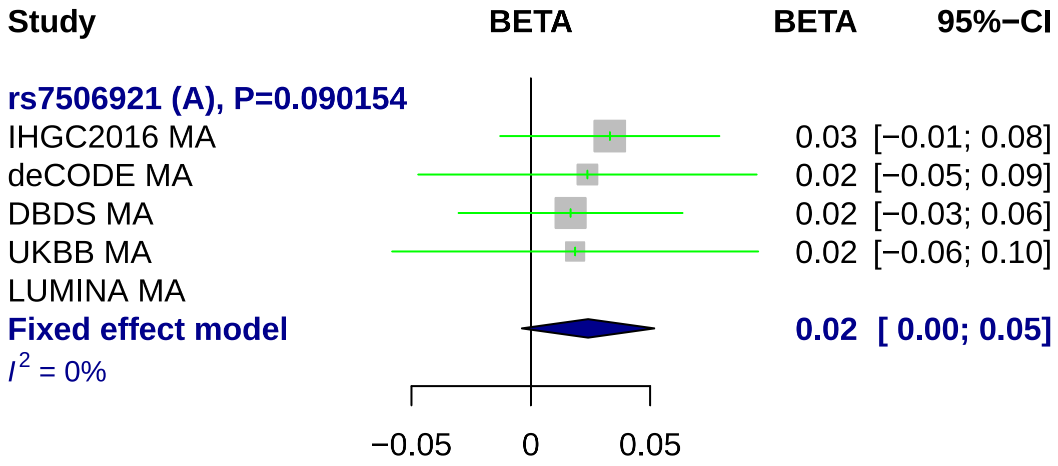
Study
BETA
BETA
95%−CI
rs7506921 (A), P=0.090154
IHGC2016 MA
0.03 [−0.01; 0.08]
deCODE MA
0.02 [−0.05; 0.09]
DBDS MA
0.02 [−0.03; 0.06]
UKBB MA
0.02 [−0.06; 0.10]
LUMINA MA
Fixed effect model
0.02 [ 0.00; 0.05]
I2 = 0%
−0.05
0
0.05

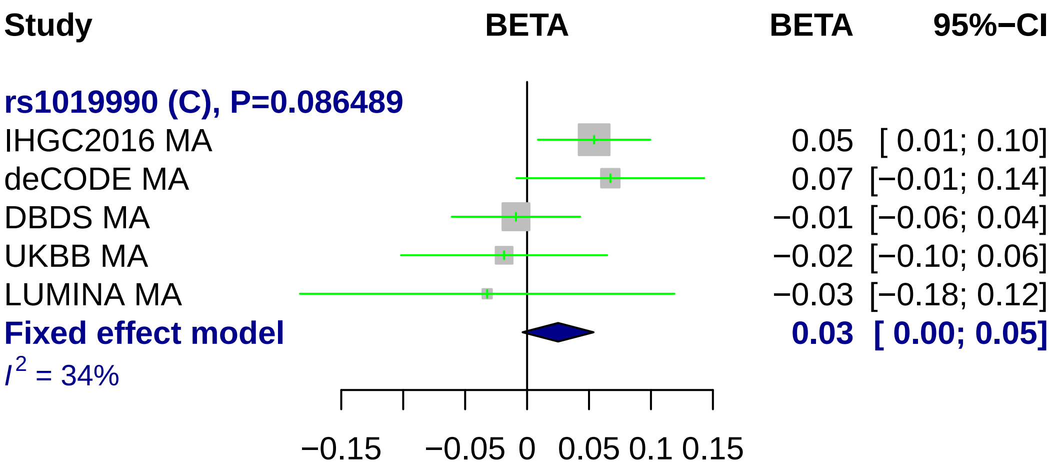
Study
BETA
BETA
95%−CI
rs1019990 (C), P=0.086489
IHGC2016 MA
0.05 [ 0.01; 0.10]
deCODE MA
0.07 [−0.01; 0.14]
DBDS MA
−0.01 [−0.06; 0.04]
UKBB MA
−0.02 [−0.10; 0.06]
LUMINA MA
−0.03 [−0.18; 0.12]
Fixed effect model
0.03 [ 0.00; 0.05]
I2 = 34%
−0.15
−0.05
0
0.05
0.1
0.15

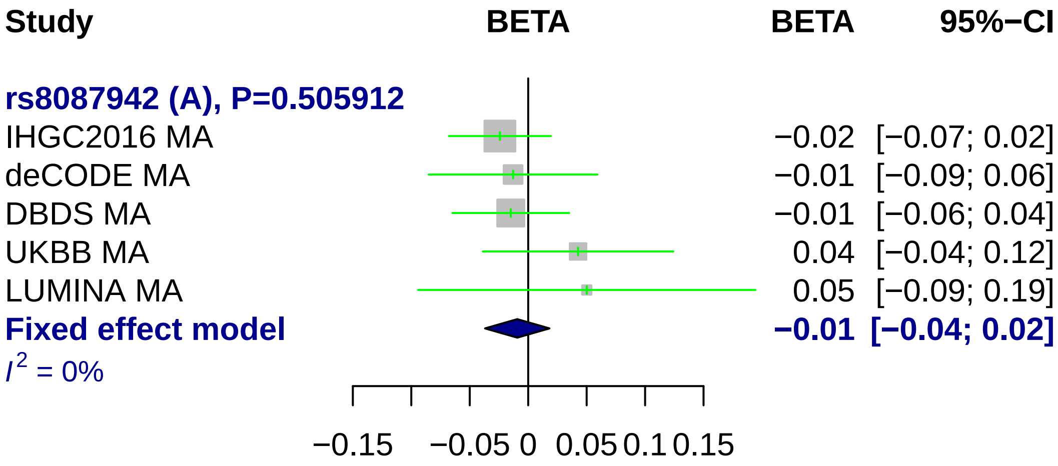
Study
BETA
BETA
95%−CI
rs8087942 (A), P=0.505912
IHGC2016 MA −0.02 [−0.07; 0.02]
deCODE MA −0.01 [−0.09; 0.06]
DBDS MA −0.01 [−0.06; 0.04]
UKBB MA 0.04 [−0.04; 0.12]
LUMINA MA 0.05 [−0.09; 0.19]
Fixed effect model −0.01 [−0.04; 0.02]
I2 = 0%
−0.15 −0.05 0 0.05 0.1 0.15

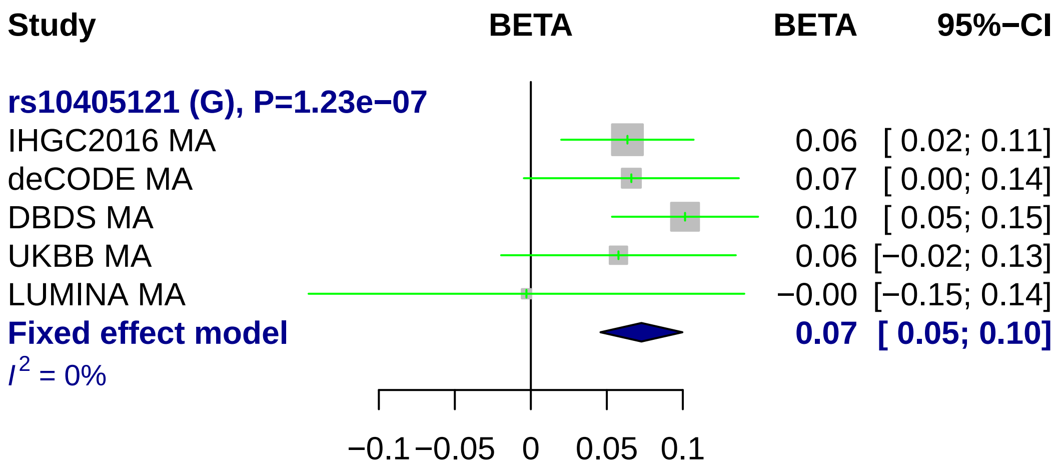
Study
BETA
BETA
95%−CI
rs10405121 (G), P=1.23e−07
IHGC2016 MA
0.06 [ 0.02; 0.11]
deCODE MA
0.07 [ 0.00; 0.14]
DBDS MA
0.10 [ 0.05; 0.15]
UKBB MA
0.06 [−0.02; 0.13]
LUMINA MA
−0.00 [−0.15; 0.14]
Fixed effect model
0.07 [ 0.05; 0.10]
I² = 0%
−0.1
−0.05
0
0.05
0.1

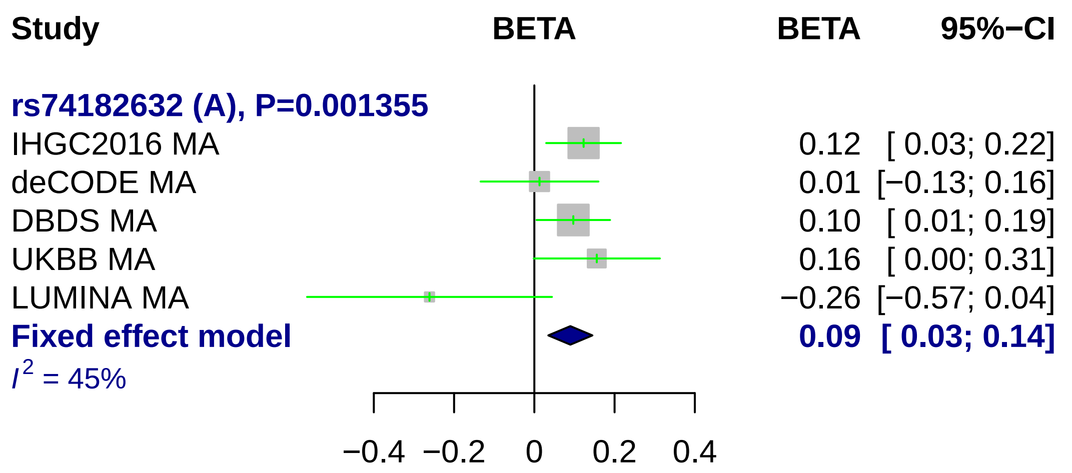
Study
BETA
BETA
95%−CI
rs74182632 (A), P=0.001355
IHGC2016 MA
0.12 [ 0.03; 0.22]
deCODE MA
0.01 [−0.13; 0.16]
DBDS MA
0.10 [ 0.01; 0.19]
UKBB MA
0.16 [ 0.00; 0.31]
LUMINA MA
−0.26 [−0.57; 0.04]
Fixed effect model
0.09 [ 0.03; 0.14]
I2 = 45%
−0.4
−0.2
0
0.2
0.4

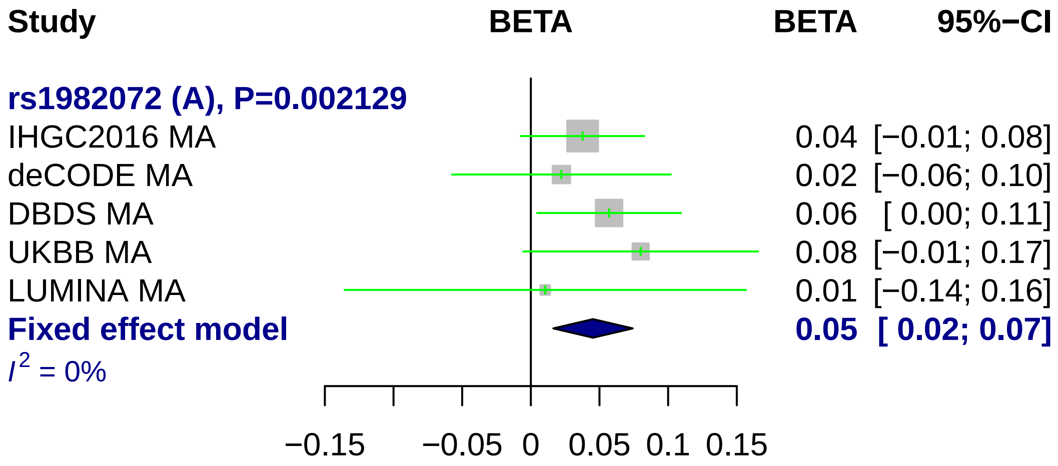

Study
BETA
BETA
95%−CI
rs1982072 (A), P=0.002129
IHGC2016 MA
0.04 [−0.01; 0.08]
deCODE MA
0.02 [−0.06; 0.10]
DBDS MA
0.06 [ 0.00; 0.11]
UKBB MA
0.08 [−0.01; 0.17]
LUMINA MA
0.01 [−0.14; 0.16]
Fixed effect model
0.05 [ 0.02; 0.07]
I2 = 0%
−0.15
−0.05
0
0.05
0.1
0.15

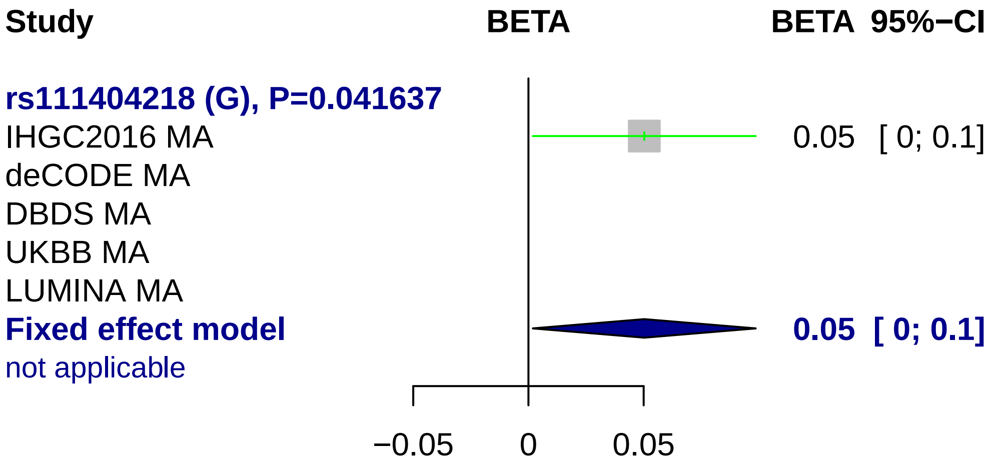
Study
BETA
BETA 95%−CI
rs111404218 (G), P=0.041637
IHGC2016 MA
0.05 [ 0; 0.1]
deCODE MA
DBDS MA
UKBB MA
LUMINA MA
Fixed effect model
0.05 [ 0; 0.1]
not applicable
−0.05
0
0.05

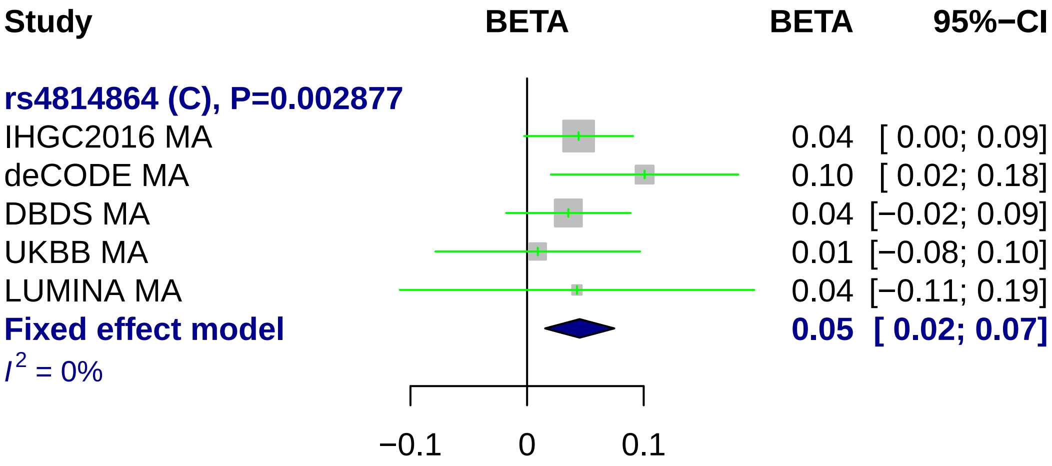

Study
BETA
BETA
95%−CI
rs4814864 (C), P=0.002877
IHGC2016 MA
0.04 [ 0.00; 0.09]
deCODE MA
0.10 [ 0.02; 0.18]
DBDS MA
0.04 [−0.02; 0.09]
UKBB MA
0.01 [−0.08; 0.10]
LUMINA MA
0.04 [−0.11; 0.19]
Fixed effect model
0.05 [ 0.02; 0.07]
I² = 0%
−0.1
0
0.1

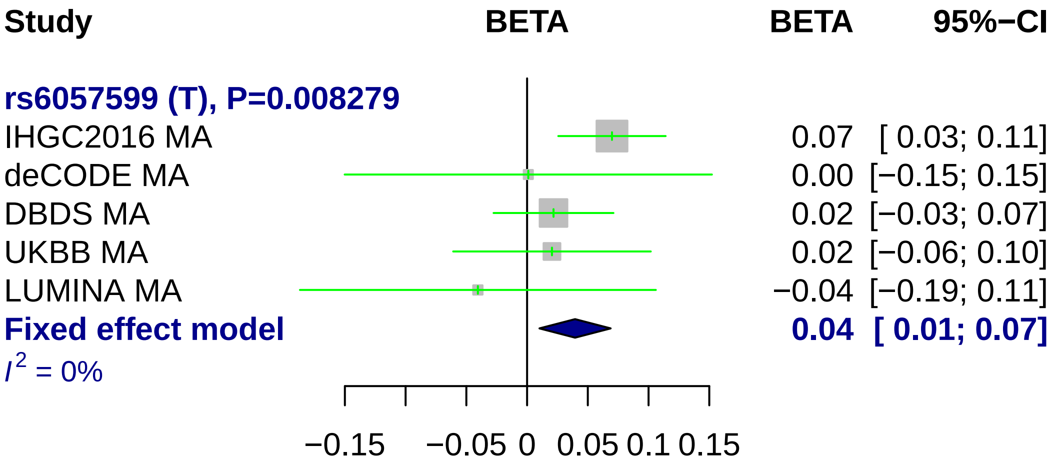

Study
BETA
BETA
95%−CI
rs6057599 (T), P=0.008279
IHGC2016 MA
0.07 [ 0.03; 0.11]
deCODE MA
0.00 [−0.15; 0.15]
DBDS MA
0.02 [−0.03; 0.07]
UKBB MA
0.02 [−0.06; 0.10]
LUMINA MA
−0.04 [−0.19; 0.11]
Fixed effect model
0.04 [ 0.01; 0.07]
I2 = 0%
−0.15
−0.05
0
0.05
0.1
0.15

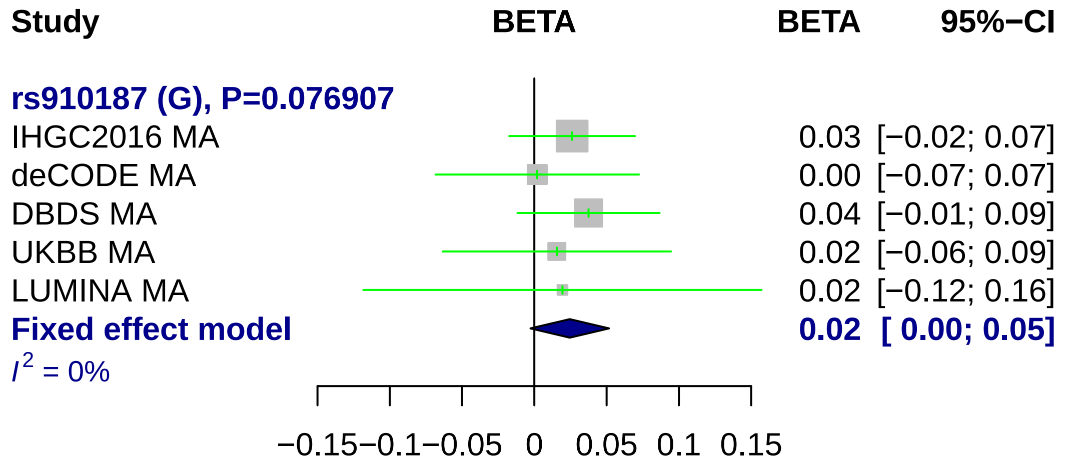

Study
BETA
BETA
95%−CI
rs910187 (G), P=0.076907
IHGC2016 MA
0.03 [−0.02; 0.07]
deCODE MA
0.00 [−0.07; 0.07]
DBDS MA
0.04 [−0.01; 0.09]
UKBB MA
0.02 [−0.06; 0.09]
LUMINA MA
0.02 [−0.12; 0.16]
Fixed effect model
0.02 [ 0.00; 0.05]
I2 = 0%
−0.15
−0.1
−0.05
0
0.05
0.1
0.15

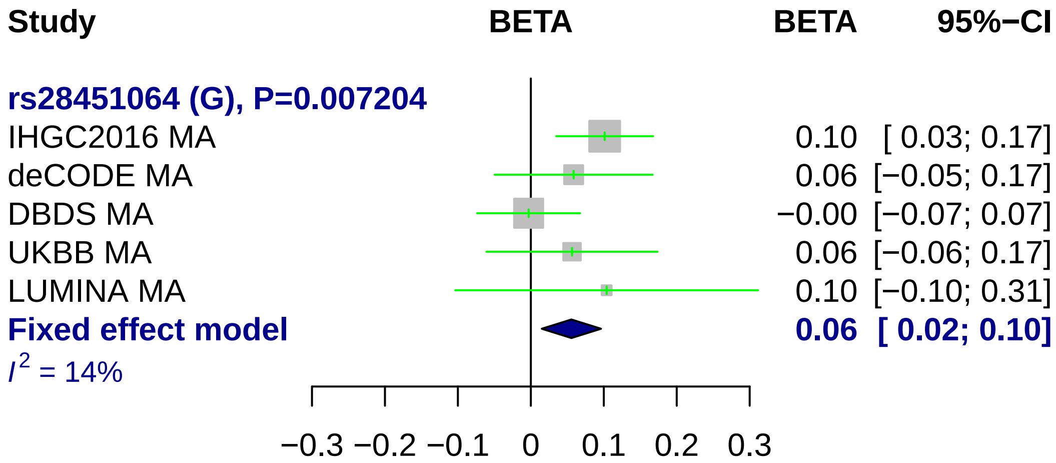
Study
BETA
BETA
95%−CI
rs28451064 (G), P=0.007204
IHGC2016 MA 0.10 [ 0.03; 0.17]
deCODE MA 0.06 [−0.05; 0.17]
DBDS MA −0.00 [−0.07; 0.07]
UKBB MA 0.06 [−0.06; 0.17]
LUMINA MA 0.10 [−0.10; 0.31]
Fixed effect model 0.06 [ 0.02; 0.10]
$I^2$ = 14%
−0.3 −0.2 −0.1 0 0.1 0.2 0.3

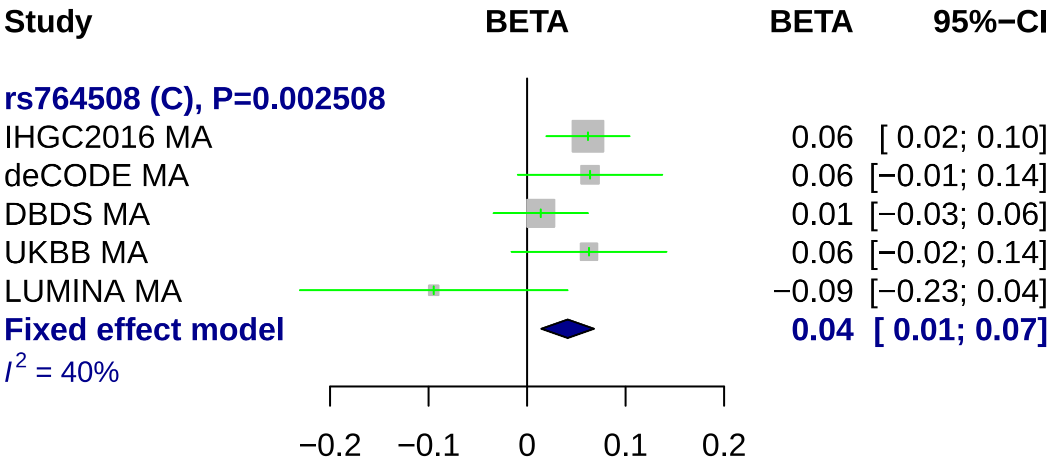
Study
BETA
BETA
95%−CI
rs764508 (C), P=0.002508
IHGC2016 MA
0.06 [ 0.02; 0.10]
deCODE MA
0.06 [−0.01; 0.14]
DBDS MA
0.01 [−0.03; 0.06]
UKBB MA
0.06 [−0.02; 0.14]
LUMINA MA
−0.09 [−0.23; 0.04]
Fixed effect model
0.04 [ 0.01; 0.07]
I2 = 40%
−0.2
−0.1
0
0.1
0.2

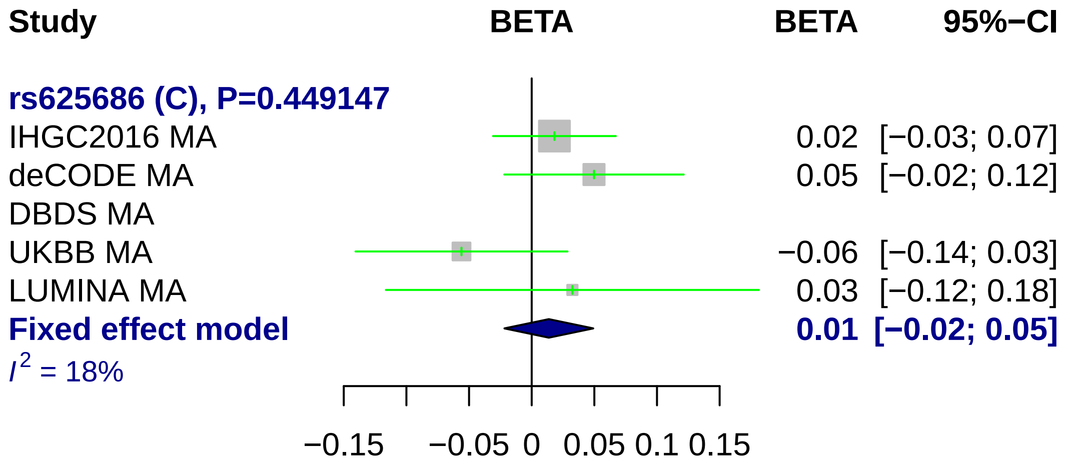
Study
BETA
BETA
95%−CI
rs625686 (C), P=0.449147
IHGC2016 MA 0.02 [−0.03; 0.07]
deCODE MA 0.05 [−0.02; 0.12]
DBDS MA
UKBB MA −0.06 [−0.14; 0.03]
LUMINA MA 0.03 [−0.12; 0.18]
Fixed effect model 0.01 [−0.02; 0.05]
$I^2$ = 18%
−0.15
−0.05
0
0.05
0.1
0.15

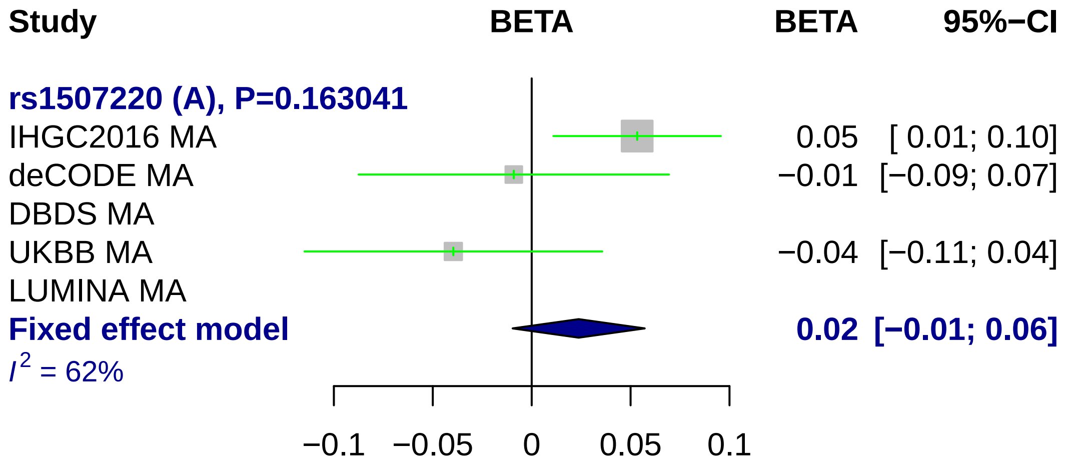
Study
BETA
BETA
95%−CI
rs1507220 (A), P=0.163041
IHGC2016 MA
0.05 [ 0.01; 0.10]
deCODE MA
−0.01 [−0.09; 0.07]
DBDS MA
UKBB MA
−0.04 [−0.11; 0.04]
LUMINA MA
Fixed effect model
0.02 [−0.01; 0.06]
I2 = 62%
−0.1
−0.05
0
0.05
0.1

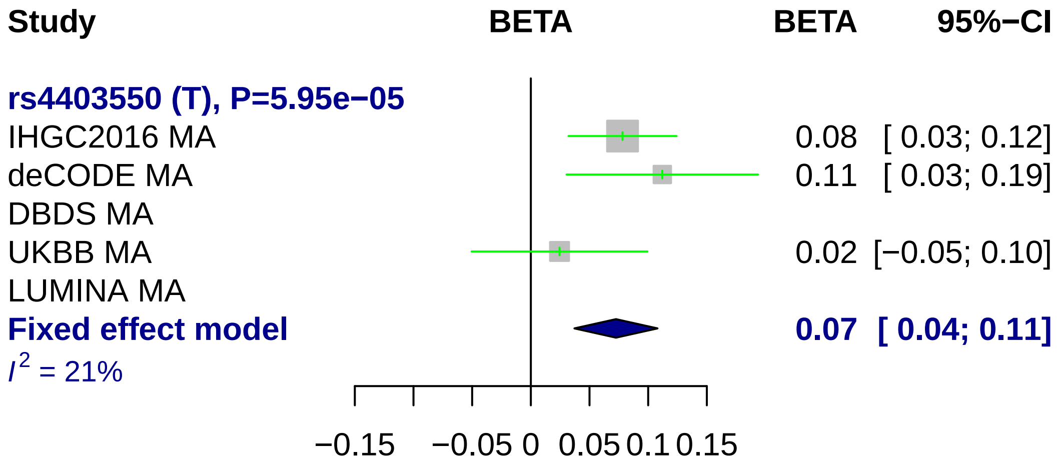
Study
BETA
BETA
95%−CI
rs4403550 (T), P=5.95e−05
IHGC2016 MA
0.08 [ 0.03; 0.12]
deCODE MA
0.11 [ 0.03; 0.19]
DBDS MA
UKBB MA
0.02 [−0.05; 0.10]
LUMINA MA
Fixed effect model
0.07 [ 0.04; 0.11]
$I^2$ = 21%
−0.15
−0.05
0
0.05
0.1
0.15
