## Supplementary Figure 10. for "Genome-wide analysis of 102,084 migraine cases identifies 123 risk loci and subtype-specific risk alleles"

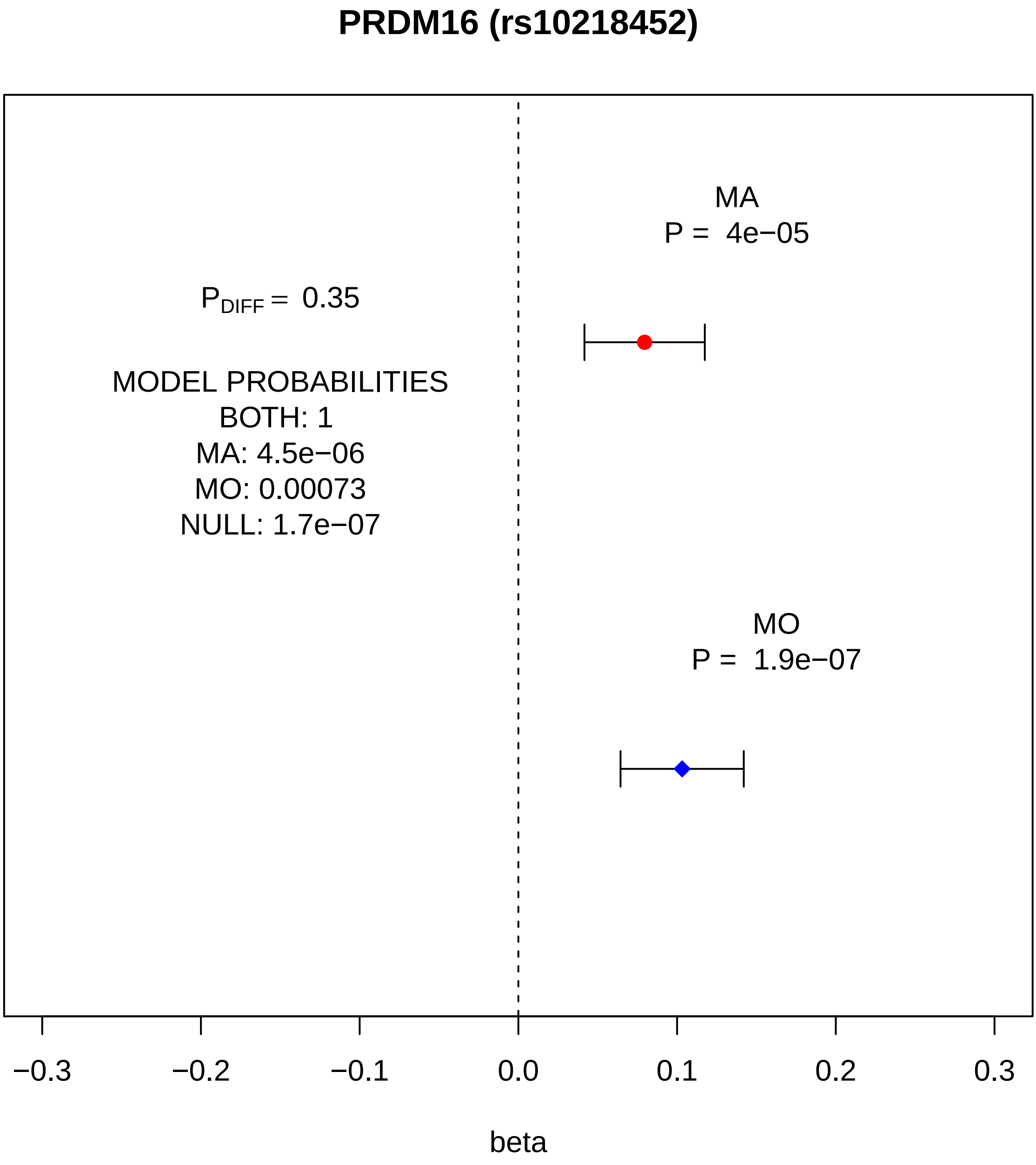
PRDM16 (rs10218452)
MA
P = 4e−05
$P_{DIFF}$ = 0.35
MODEL PROBABILITIES
BOTH: 1
MA: 4.5e−06
MO: 0.00073
NULL: 1.7e−07
MO
P = 1.9e−07
−0.3
−0.2
−0.1
0.0
0.1
0.2
0.3
beta

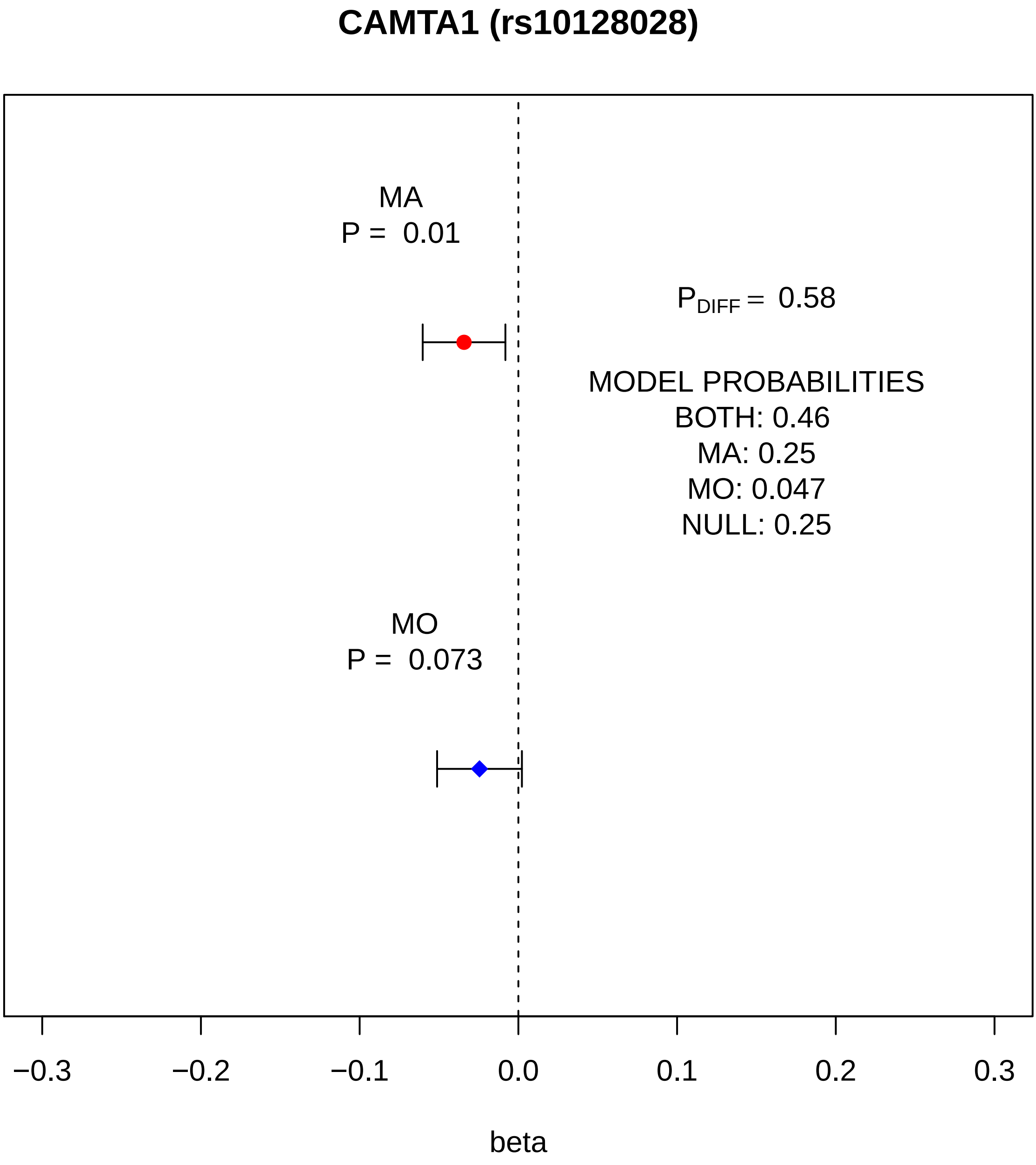
CAMTA1 (rs10128028)
MA
P = 0.01
$P_{DIFF}$ = 0.58
MODEL PROBABILITIES
BOTH: 0.46
MA: 0.25
MO: 0.047
NULL: 0.25
MO
P = 0.073
−0.3
−0.2
−0.1
0.0
0.1
0.2
0.3
beta

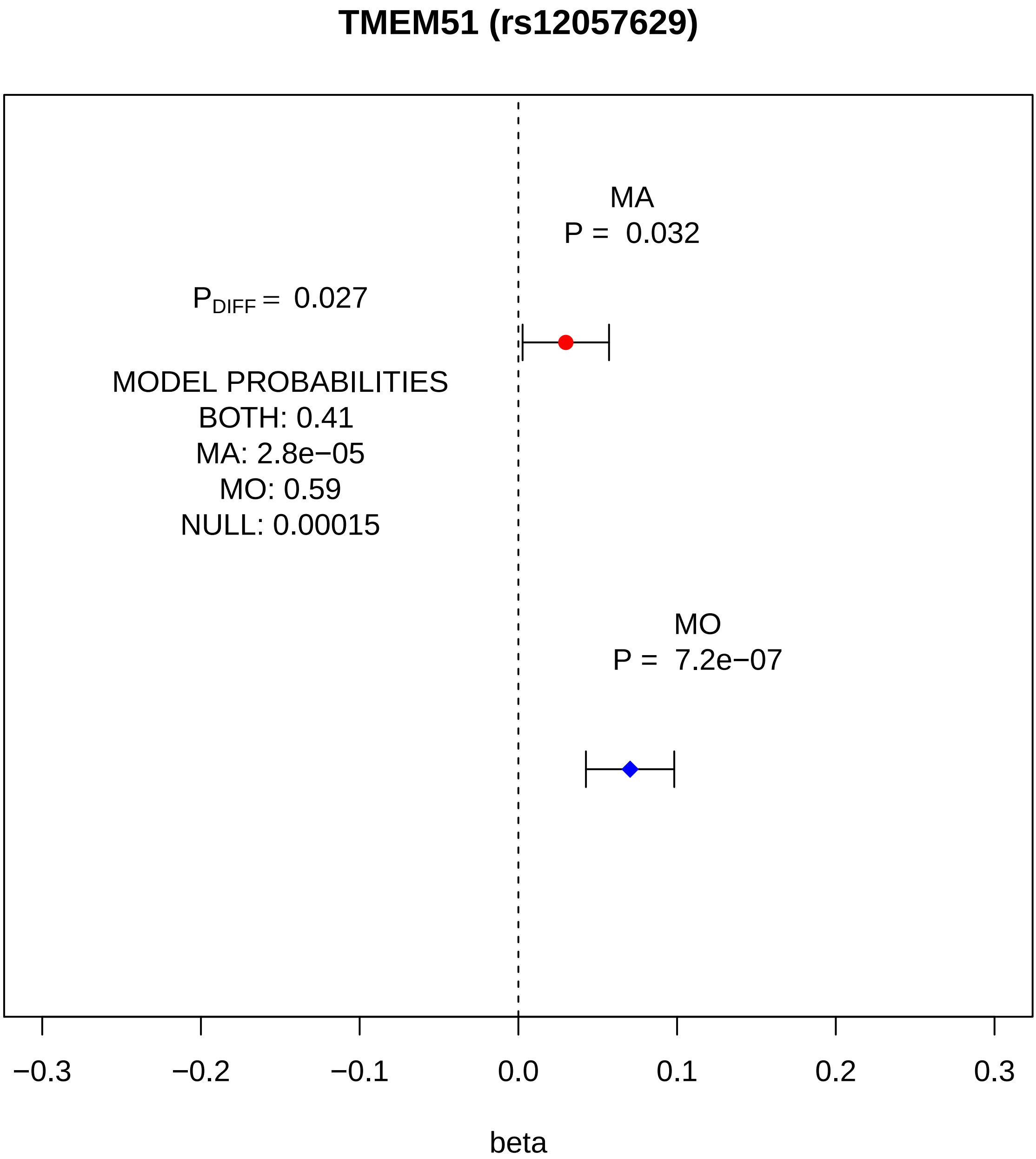
TMEM51 (rs12057629)
MA
P = 0.032
$P_{DIFF}$ = 0.027
MODEL PROBABILITIES
BOTH: 0.41
MA: 2.8e−05
MO: 0.59
NULL: 0.00015
MO
P = 7.2e−07
−0.3
−0.2
−0.1
0.0
0.1
0.2
0.3
beta

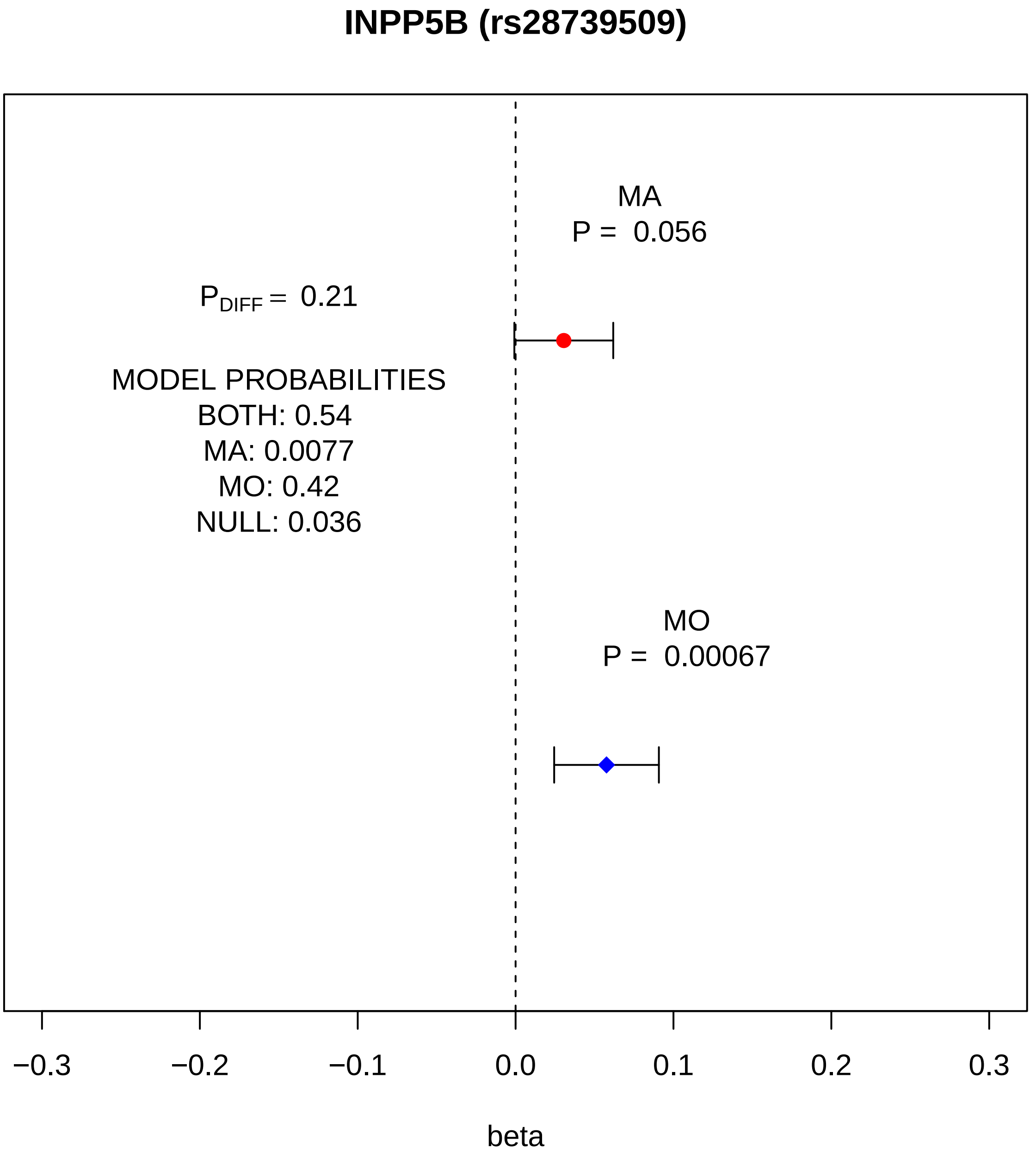

INPP5B (rs28739509)
MA
P = 0.056
$P_{DIFF}$ = 0.21
MODEL PROBABILITIES
BOTH: 0.54
MA: 0.0077
MO: 0.42
NULL: 0.036
MO
P = 0.00067
−0.3
−0.2
−0.1
0.0
0.1
0.2
0.3
beta

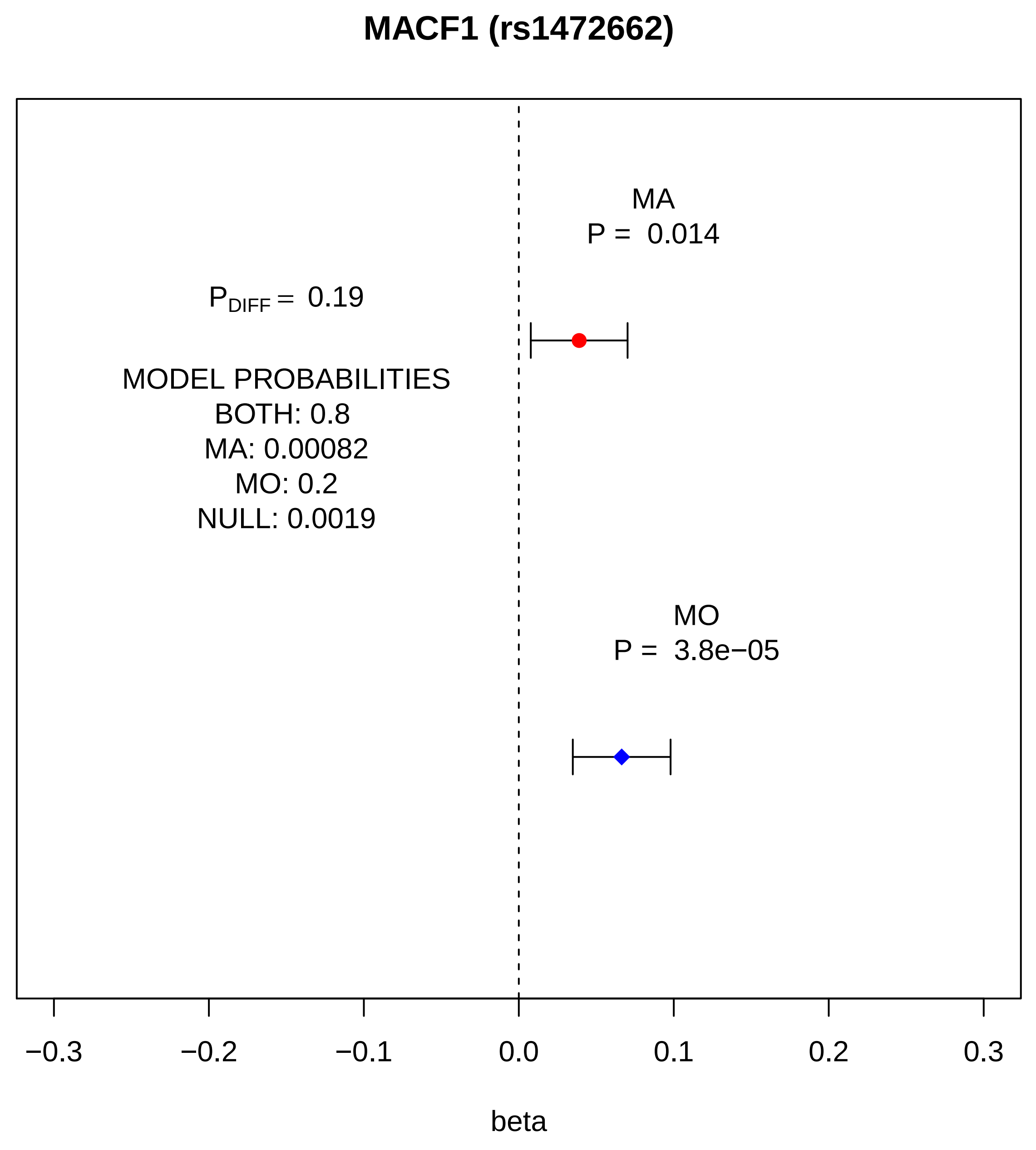
MACF1 (rs1472662)
MA
P = 0.014
$P_{DIFF}$ = 0.19
MODEL PROBABILITIES
BOTH: 0.8
MA: 0.00082
MO: 0.2
NULL: 0.0019
MO
P = 3.8e−05
−0.3
−0.2
−0.1
0.0
0.1
0.2
0.3
beta

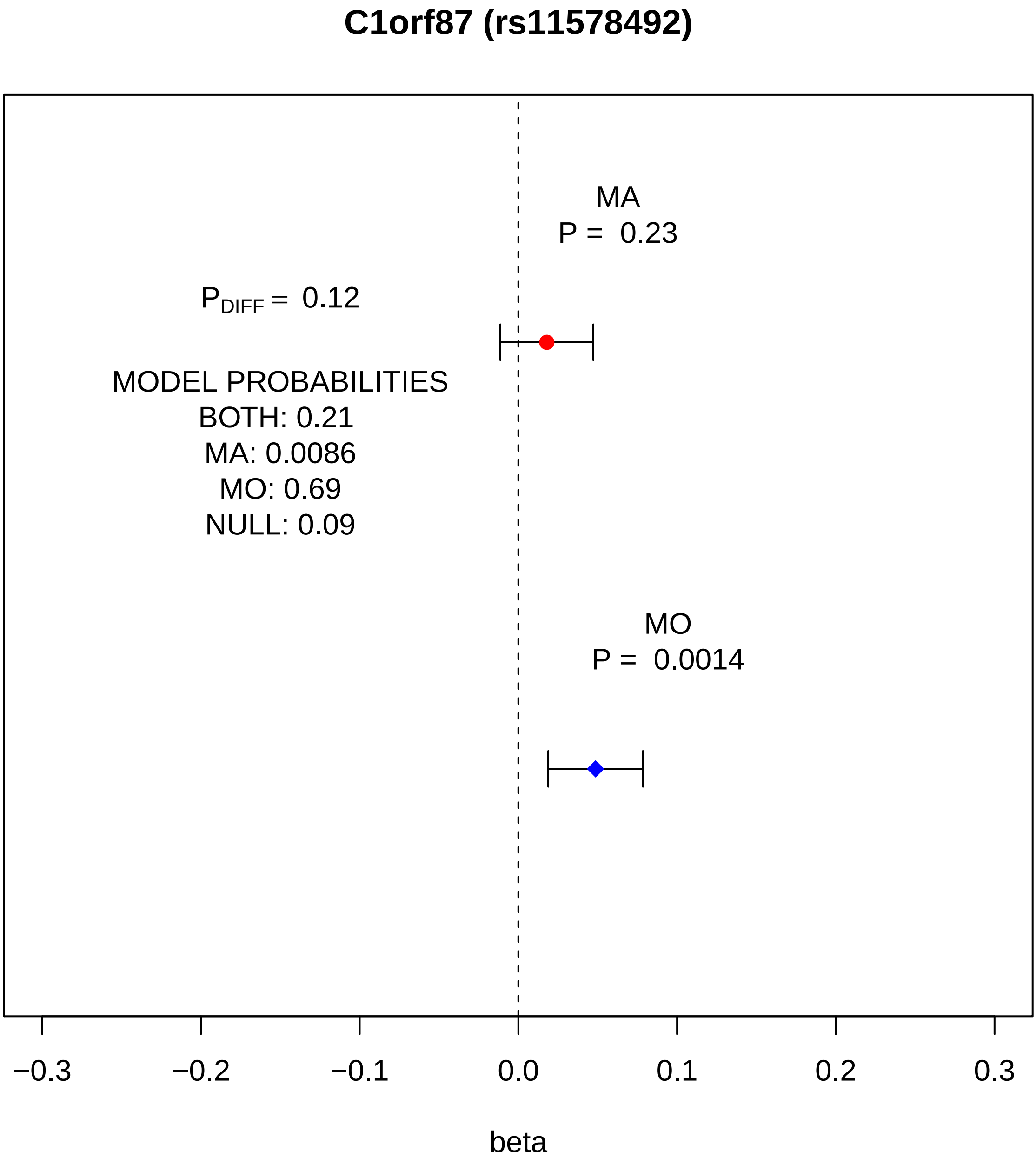
C1orf87 (rs11578492)
MA
P = 0.23
$P_{DIFF}$ = 0.12
MODEL PROBABILITIES
BOTH: 0.21
MA: 0.0086
MO: 0.69
NULL: 0.09
MO
P = 0.0014
−0.3
−0.2
−0.1
0.0
0.1
0.2
0.3
beta

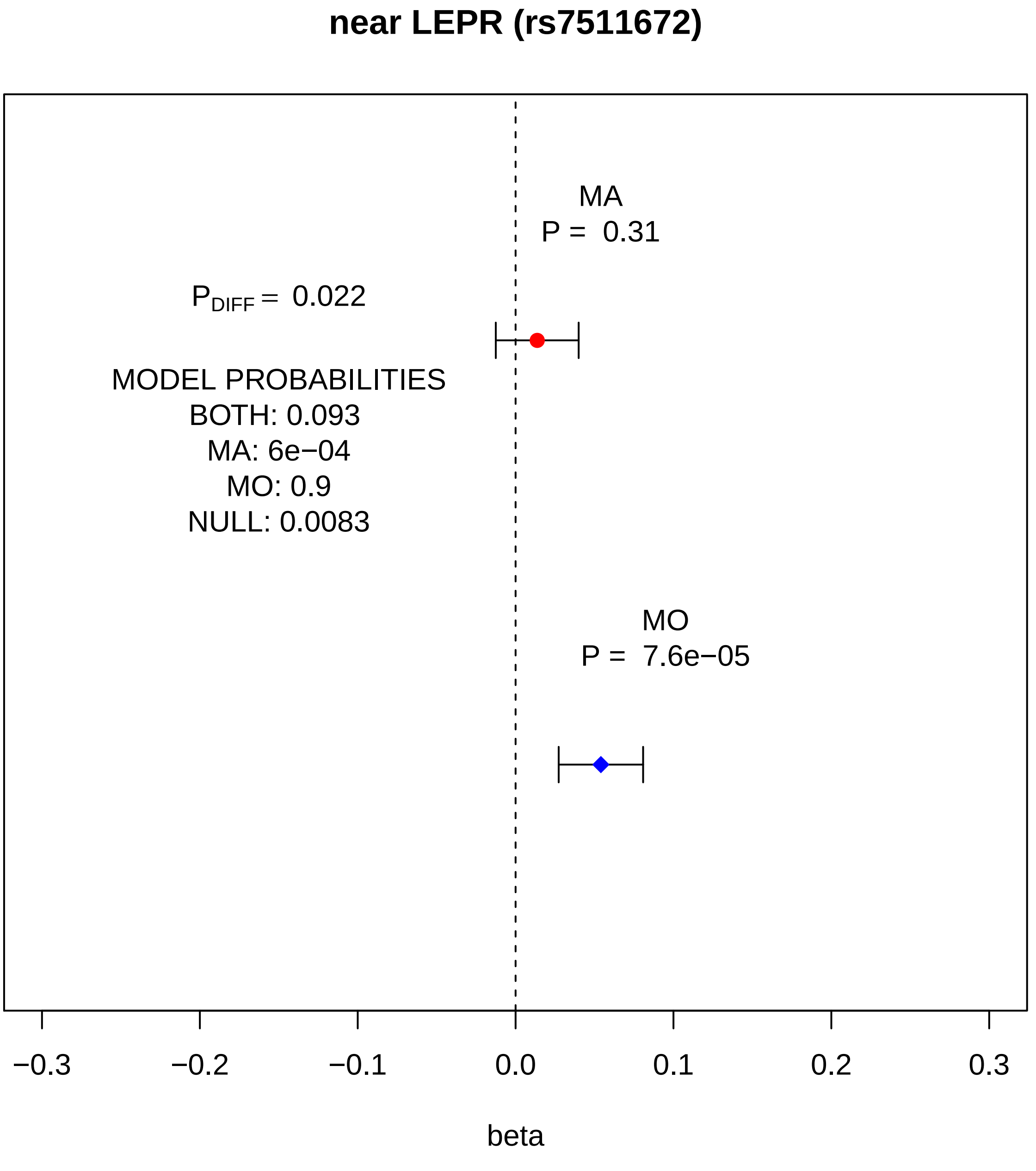
near LEPR (rs7511672)
MA
P = 0.31
P_DIFF = 0.022
MODEL PROBABILITIES
BOTH: 0.093
MA: 6e−04
MO: 0.9
NULL: 0.0083
MO
P = 7.6e−05
−0.3
−0.2
−0.1
0.0
0.1
0.2
0.3
beta

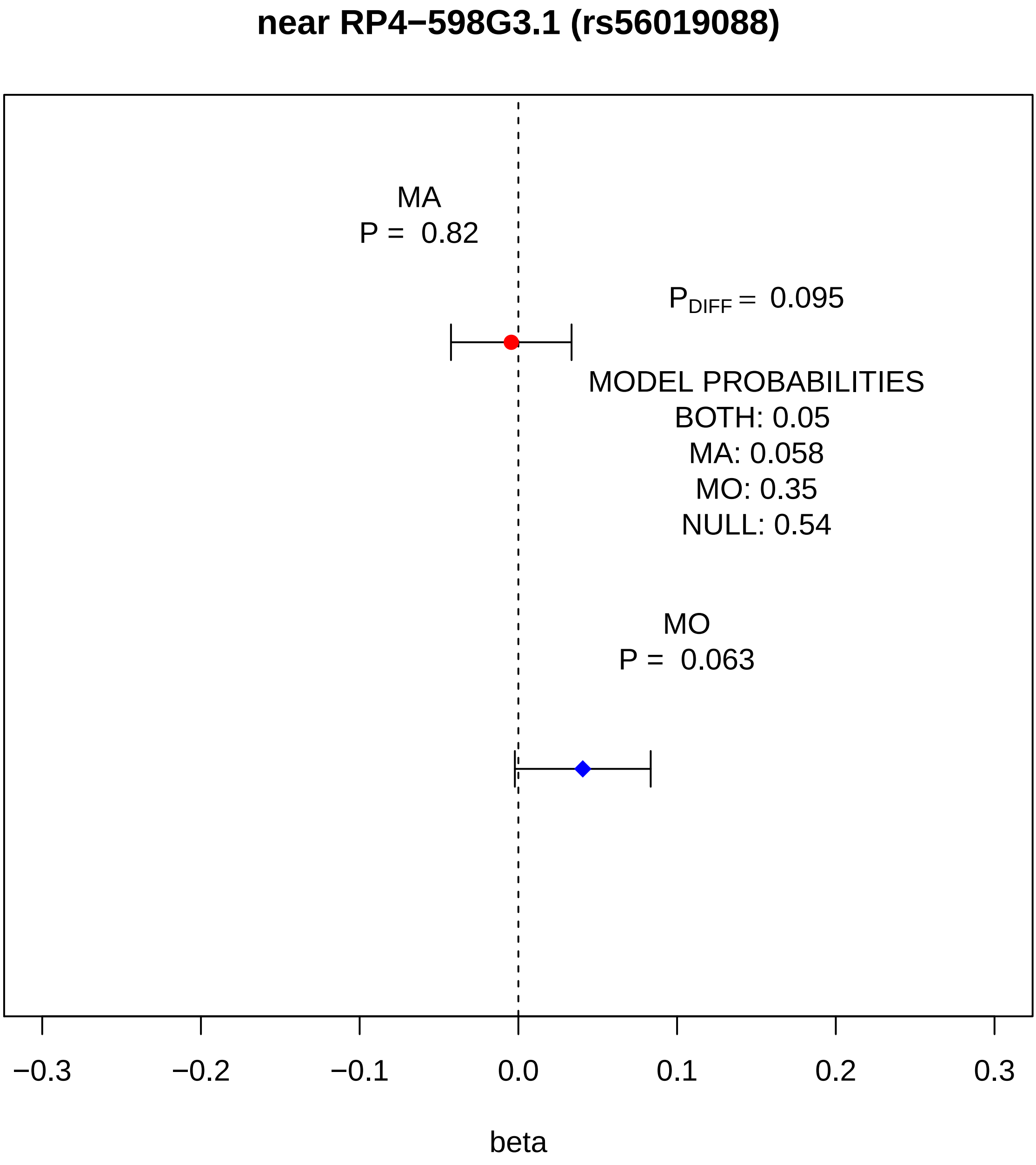
near RP4−598G3.1 (rs56019088)
MA
P = 0.82
$P_{DIFF}$ = 0.095
MODEL PROBABILITIES
BOTH: 0.05
MA: 0.058
MO: 0.35
NULL: 0.54
MO
P = 0.063
−0.3
−0.2
−0.1
0.0
0.1
0.2
0.3
beta

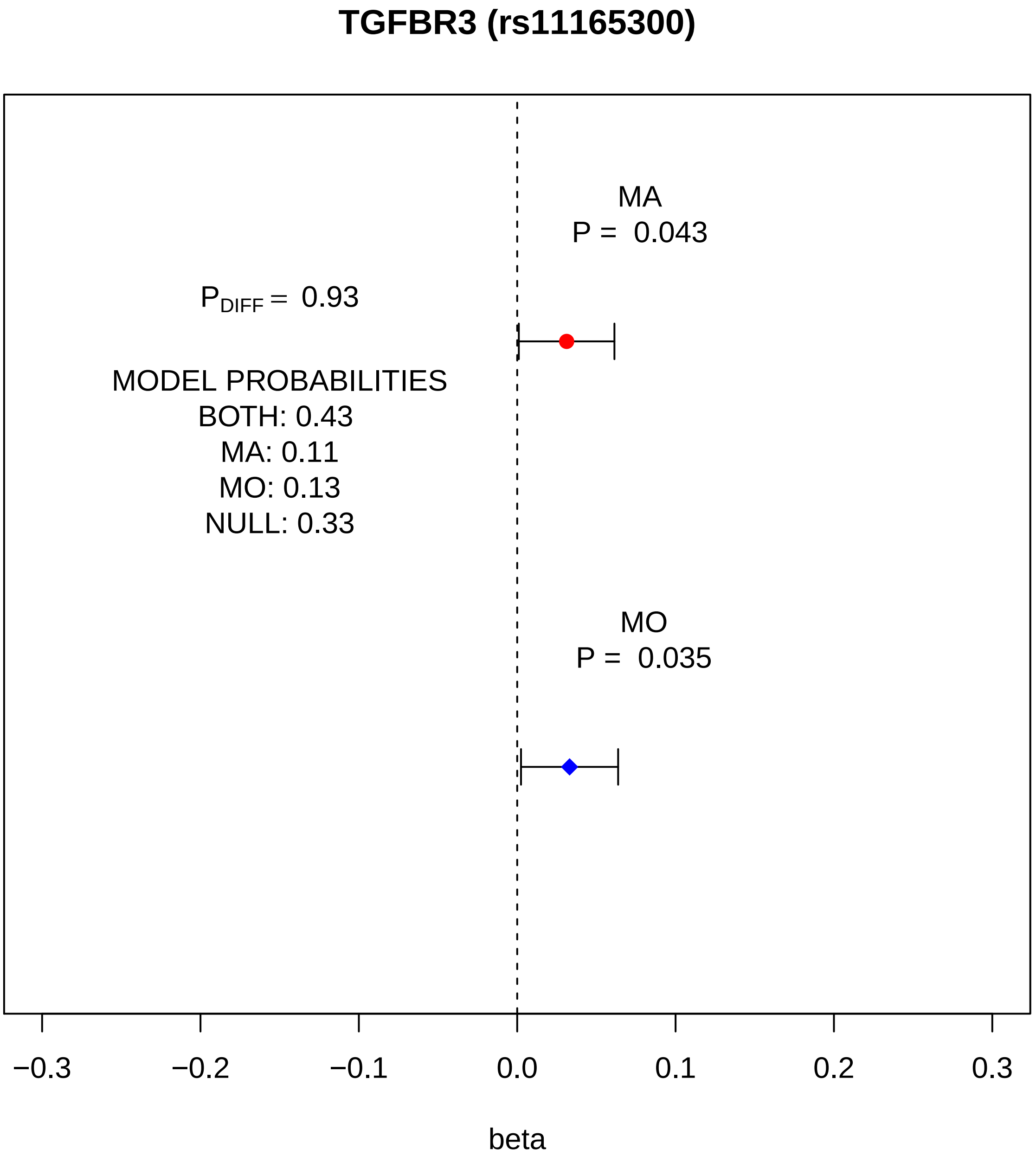
TGFBR3 (rs11165300)
MA
P = 0.043
$P_{DIFF}$ = 0.93
MODEL PROBABILITIES
BOTH: 0.43
MA: 0.11
MO: 0.13
NULL: 0.33
MO
P = 0.035
−0.3
−0.2
−0.1
0.0
0.1
0.2
0.3
beta

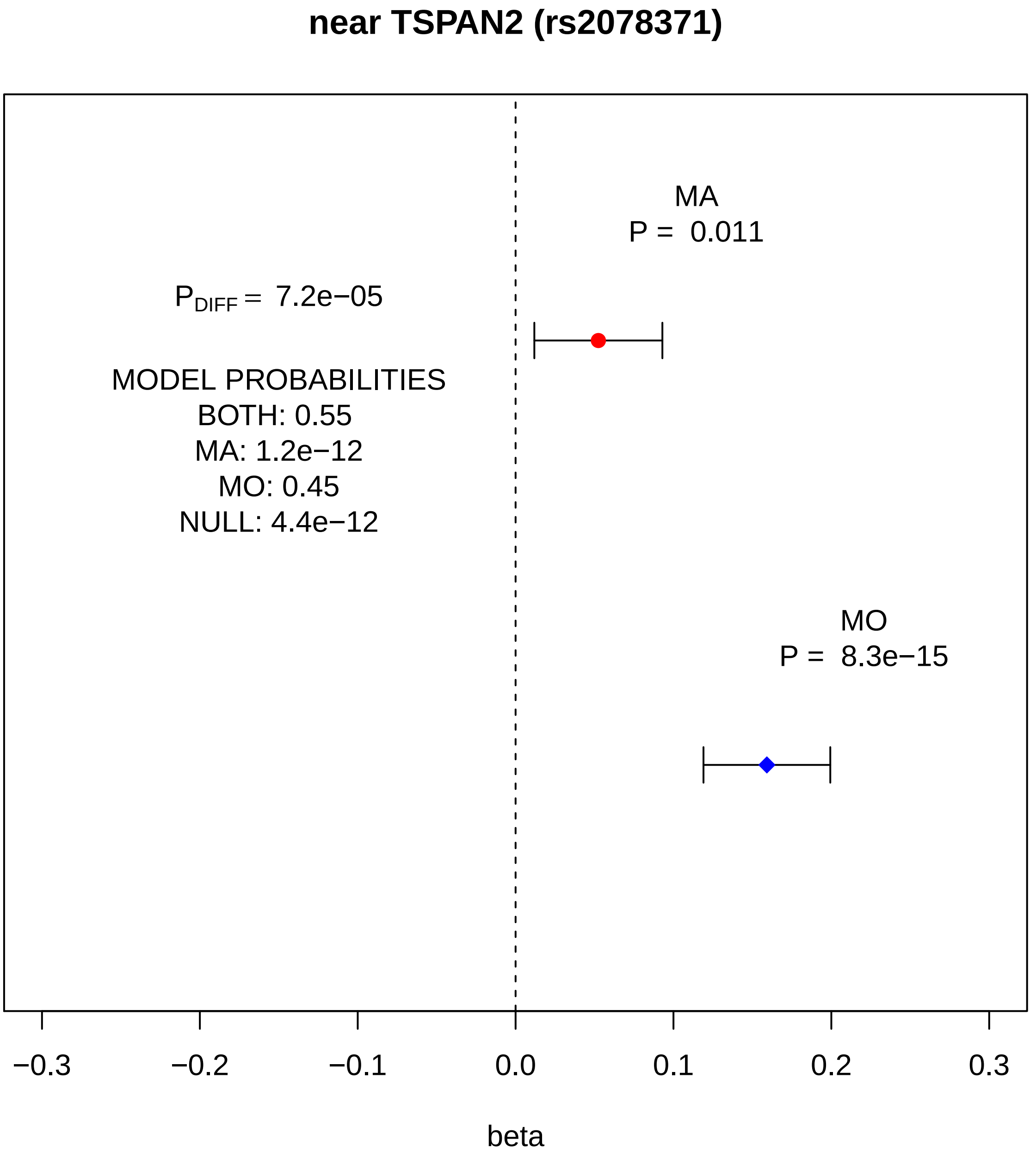
near TSPAN2 (rs2078371)
MA
P = 0.011
P_DIFF = 7.2e−05
MODEL PROBABILITIES
BOTH: 0.55
MA: 1.2e−12
MO: 0.45
NULL: 4.4e−12
MO
P = 8.3e−15
−0.3
−0.2
−0.1
0.0
0.1
0.2
0.3
beta

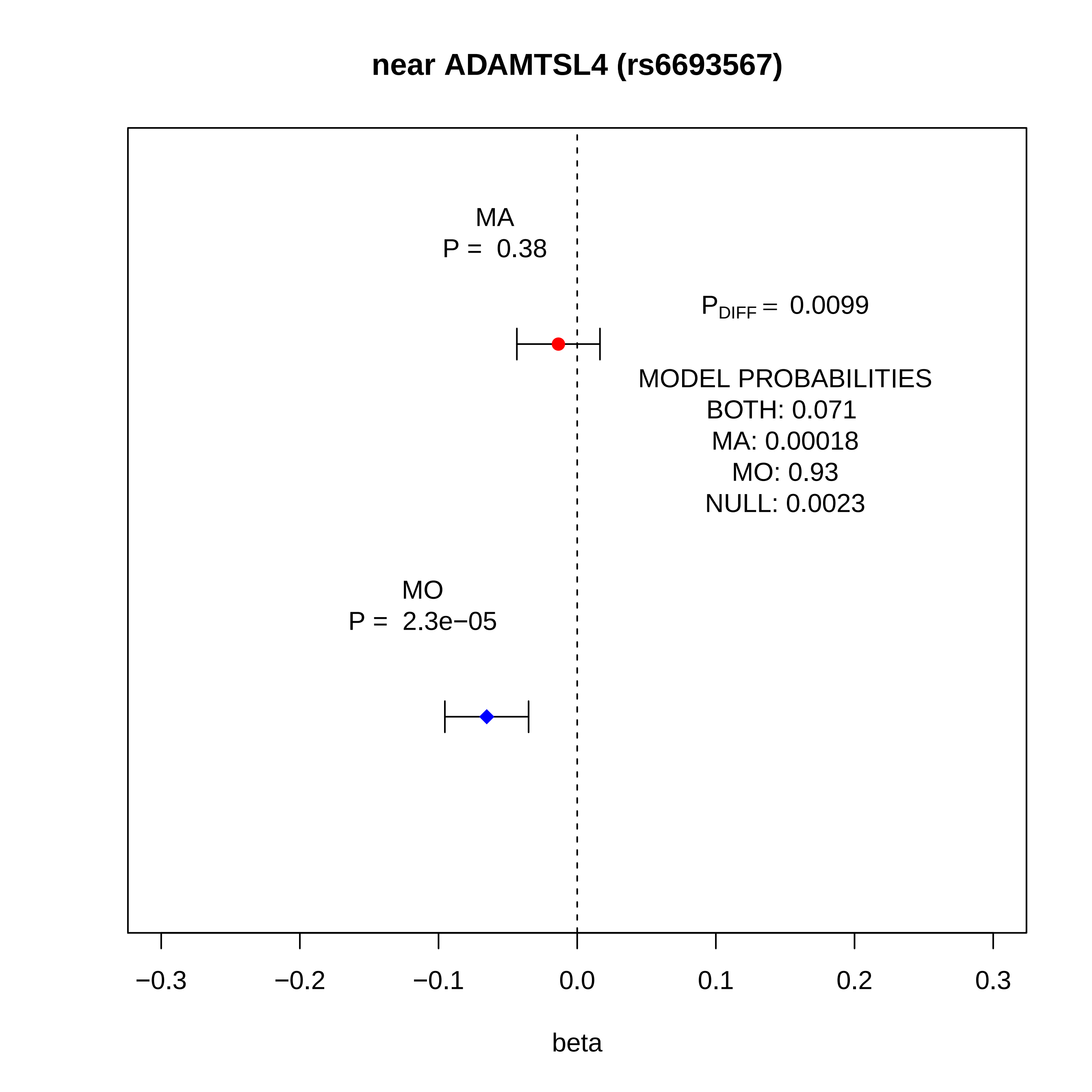
near ADAMTSL4 (rs6693567)
MA
P = 0.38
P_DIFF = 0.0099
MODEL PROBABILITIES
BOTH: 0.071
MA: 0.00018
MO: 0.93
NULL: 0.0023
MO
P = 2.3e−05
−0.3
−0.2
−0.1
0.0
0.1
0.2
0.3
beta

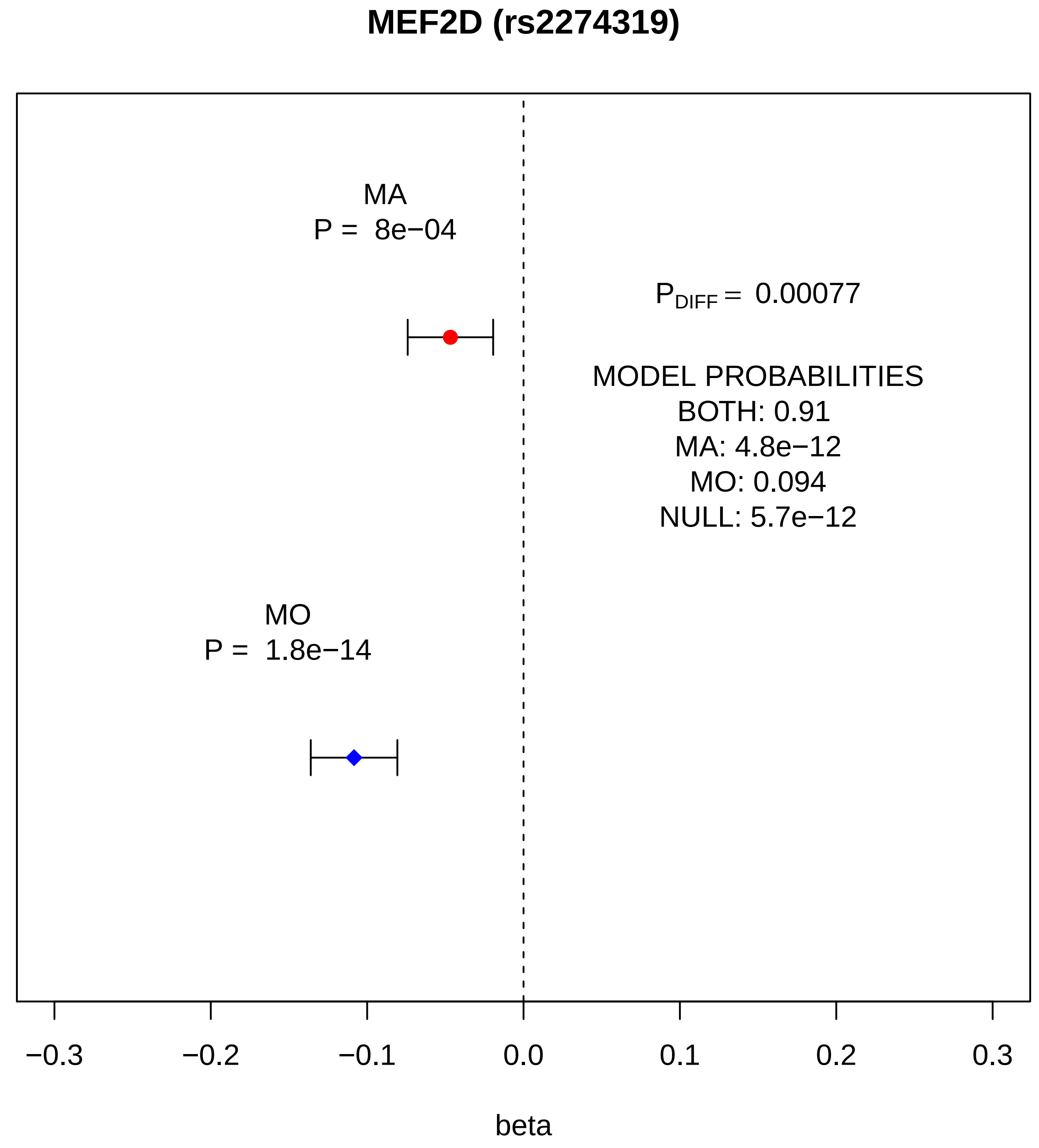
MEF2D (rs2274319)
MA
P = 8e−04
P_DIFF = 0.00077
MODEL PROBABILITIES
BOTH: 0.91
MA: 4.8e−12
MO: 0.094
NULL: 5.7e−12
MO
P = 1.8e−14
−0.3
−0.2
−0.1
0.0
0.1
0.2
0.3
beta

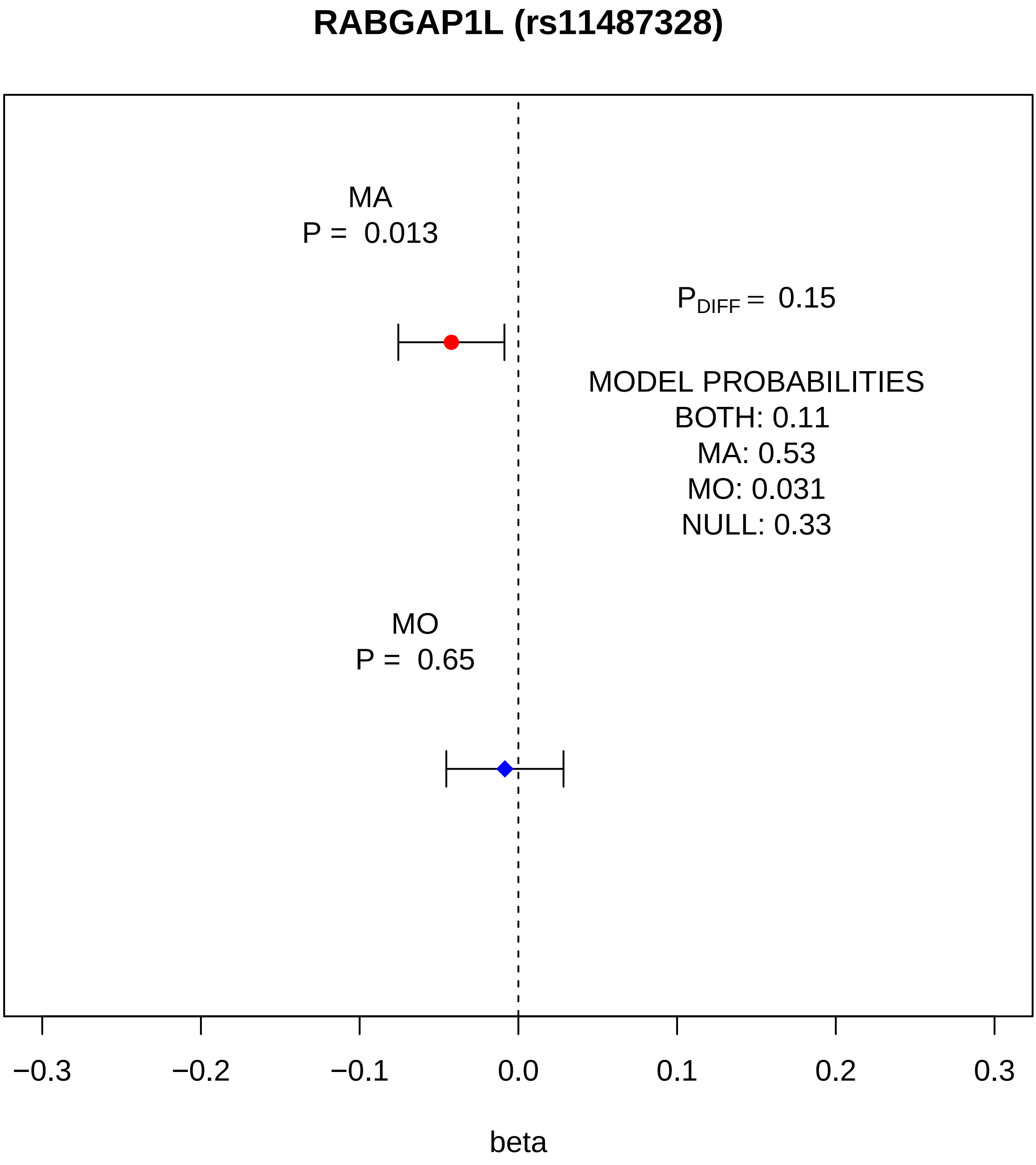
RABGAP1L (rs11487328)
MA
P = 0.013
$P_{DIFF}$ = 0.15
MODEL PROBABILITIES
BOTH: 0.11
MA: 0.53
MO: 0.031
NULL: 0.33
MO
P = 0.65
−0.3
−0.2
−0.1
0.0
0.1
0.2
0.3
beta

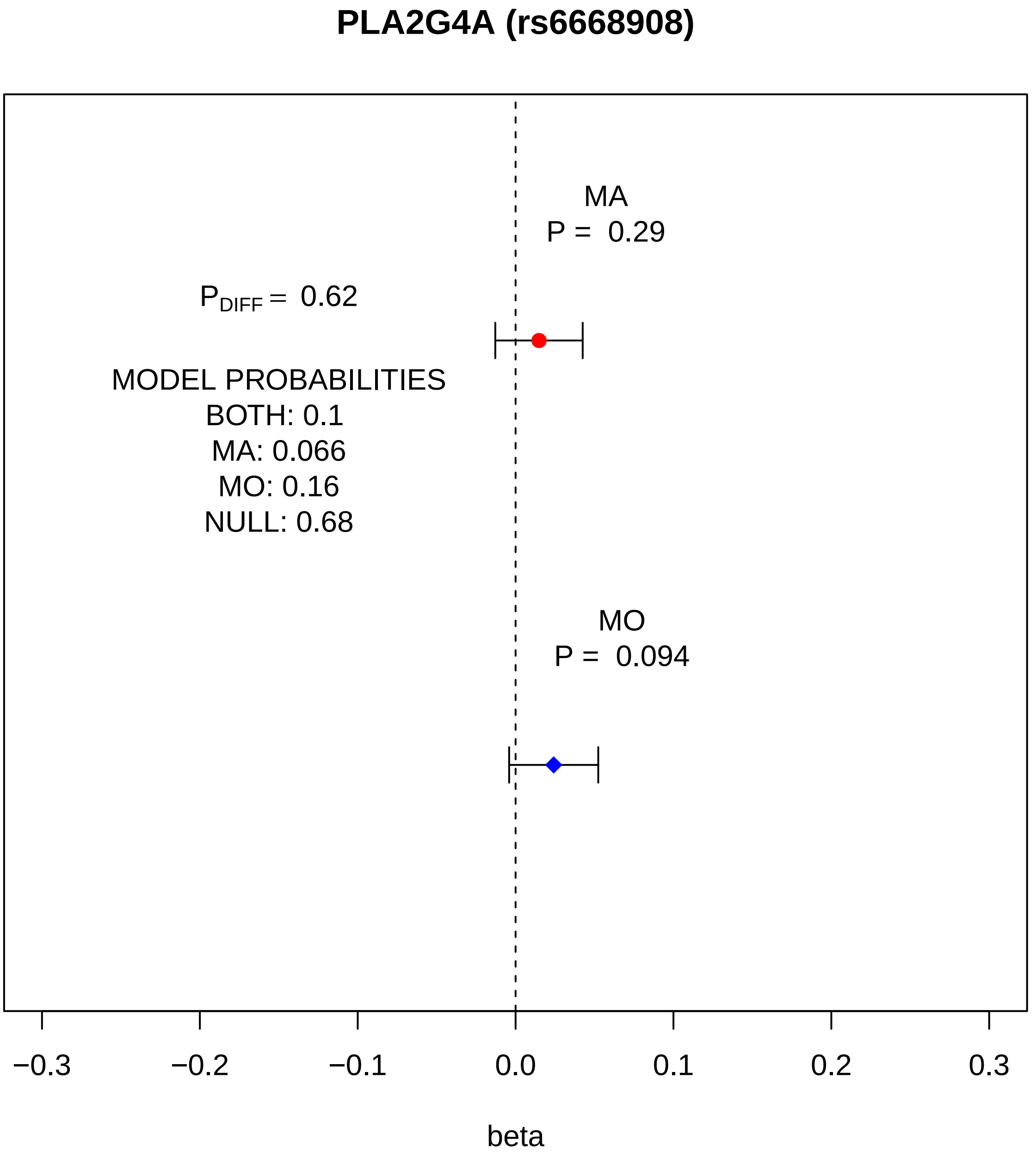
PLA2G4A (rs6668908)
MA
P = 0.29
P_DIFF = 0.62
MODEL PROBABILITIES
BOTH: 0.1
MA: 0.066
MO: 0.16
NULL: 0.68
MO
P = 0.094
−0.3
−0.2
−0.1
0.0
0.1
0.2
0.3
beta

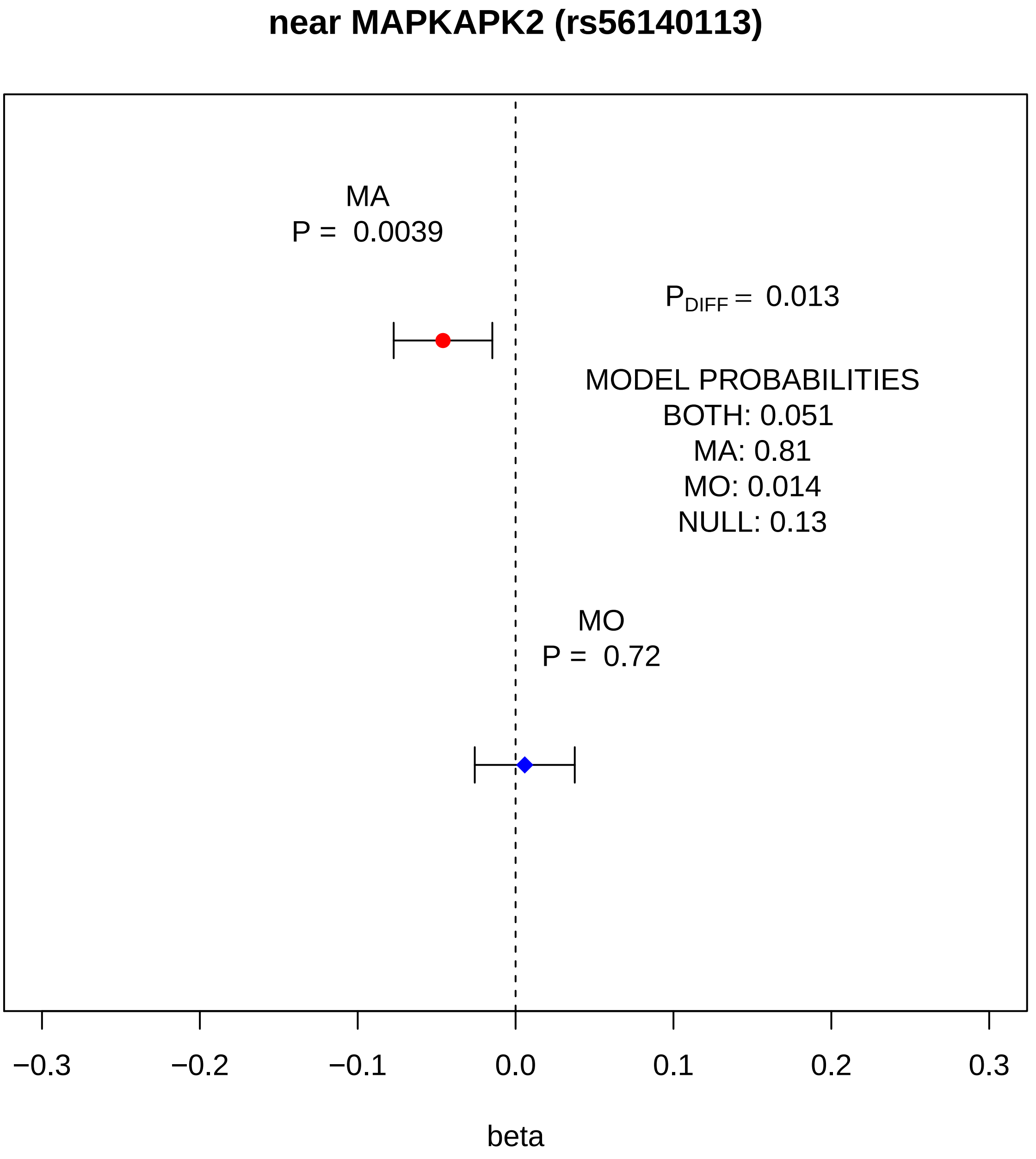
near MAPKAPK2 (rs56140113)
MA
P = 0.0039
P_DIFF = 0.013
MODEL PROBABILITIES
BOTH: 0.051
MA: 0.81
MO: 0.014
NULL: 0.13
MO
P = 0.72
−0.3
−0.2
−0.1
0.0
0.1
0.2
0.3
beta

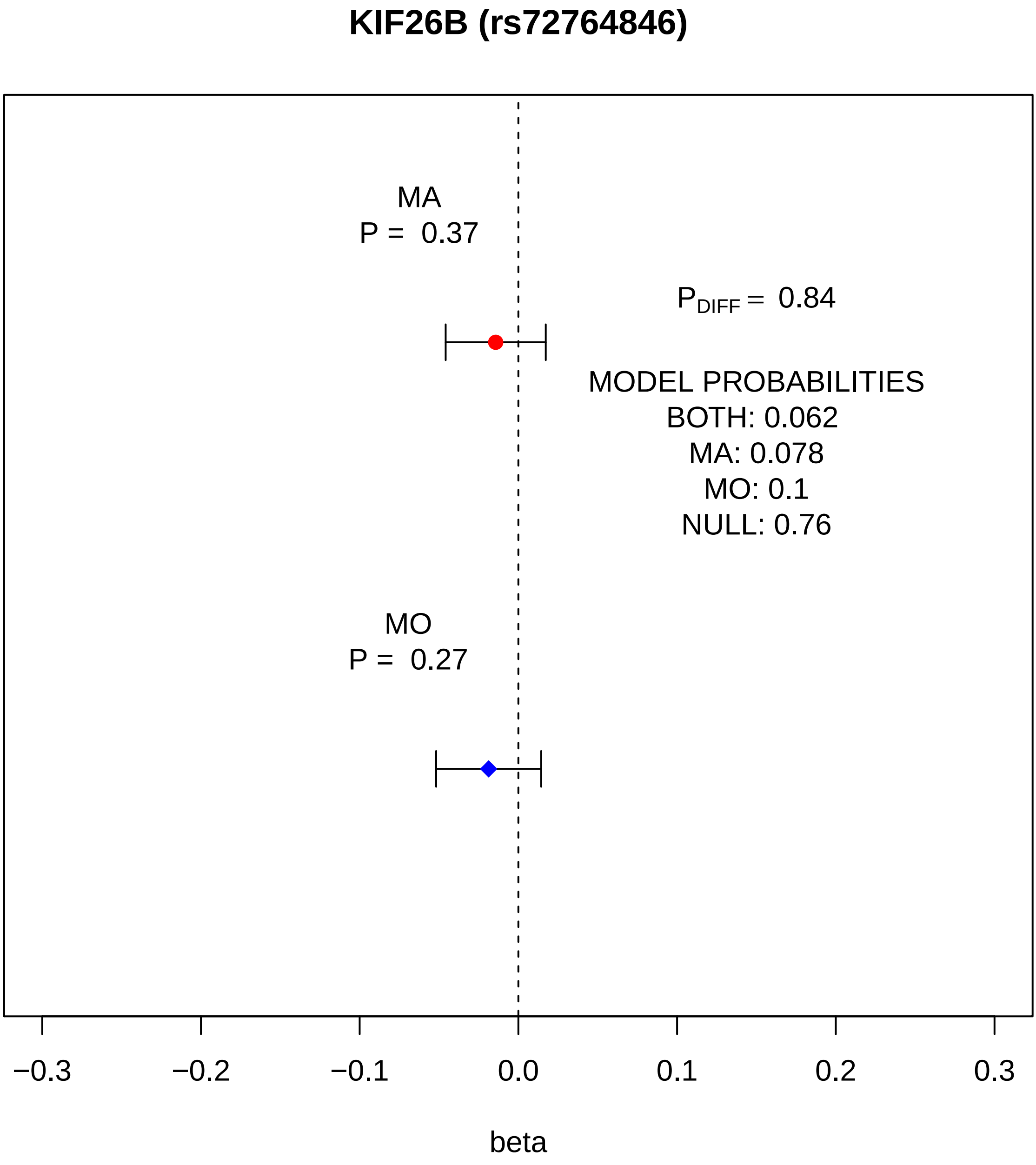
KIF26B (rs72764846)
MA
P = 0.37
P_DIFF = 0.84
MODEL PROBABILITIES
BOTH: 0.062
MA: 0.078
MO: 0.1
NULL: 0.76
MO
P = 0.27
−0.3
−0.2
−0.1
0.0
0.1
0.2
0.3
beta

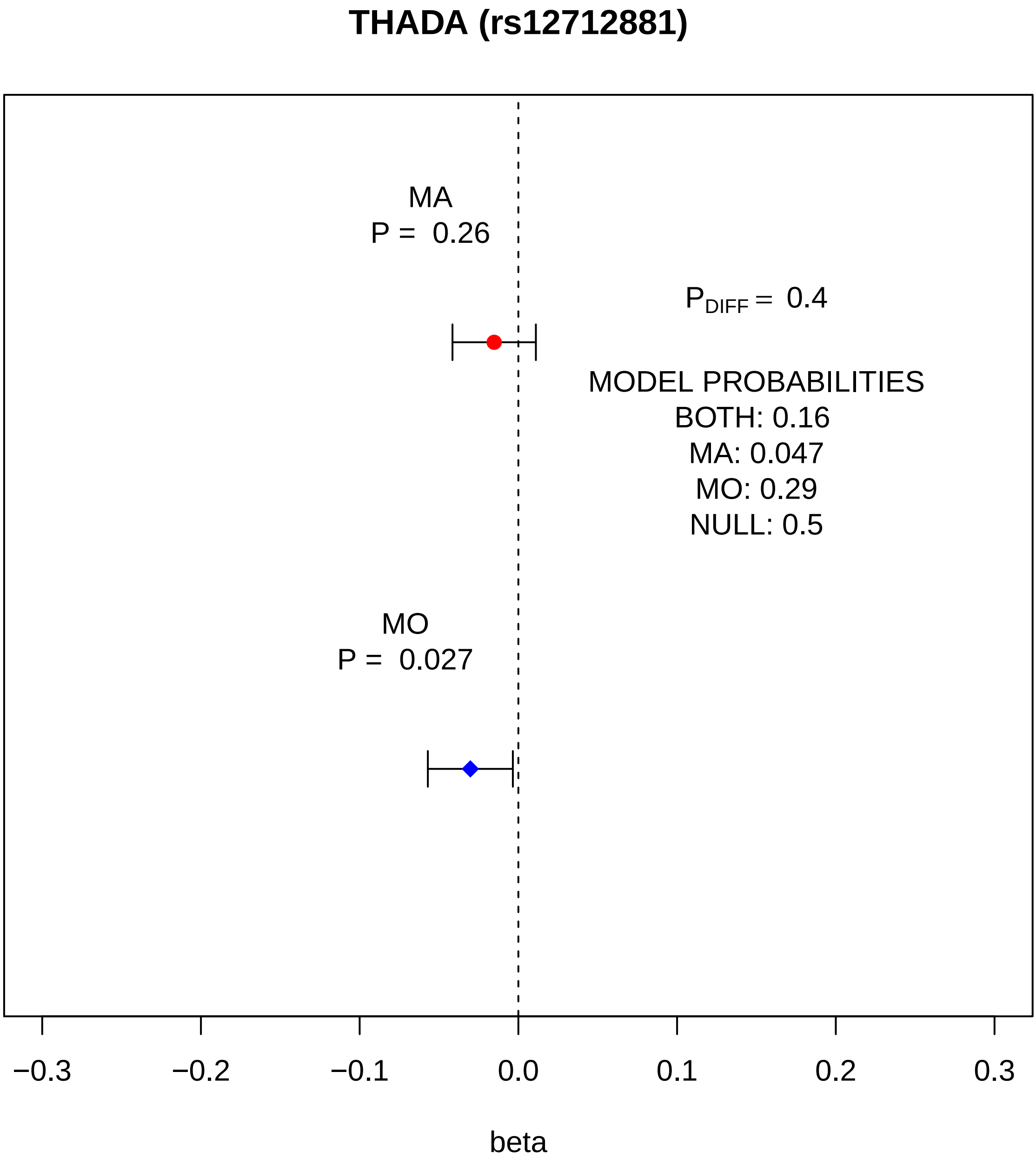
THADA (rs12712881)
MA
P = 0.26
$P_{DIFF}$ = 0.4
MODEL PROBABILITIES
BOTH: 0.16
MA: 0.047
MO: 0.29
NULL: 0.5
MO
P = 0.027
−0.3
−0.2
−0.1
0.0
0.1
0.2
0.3
beta

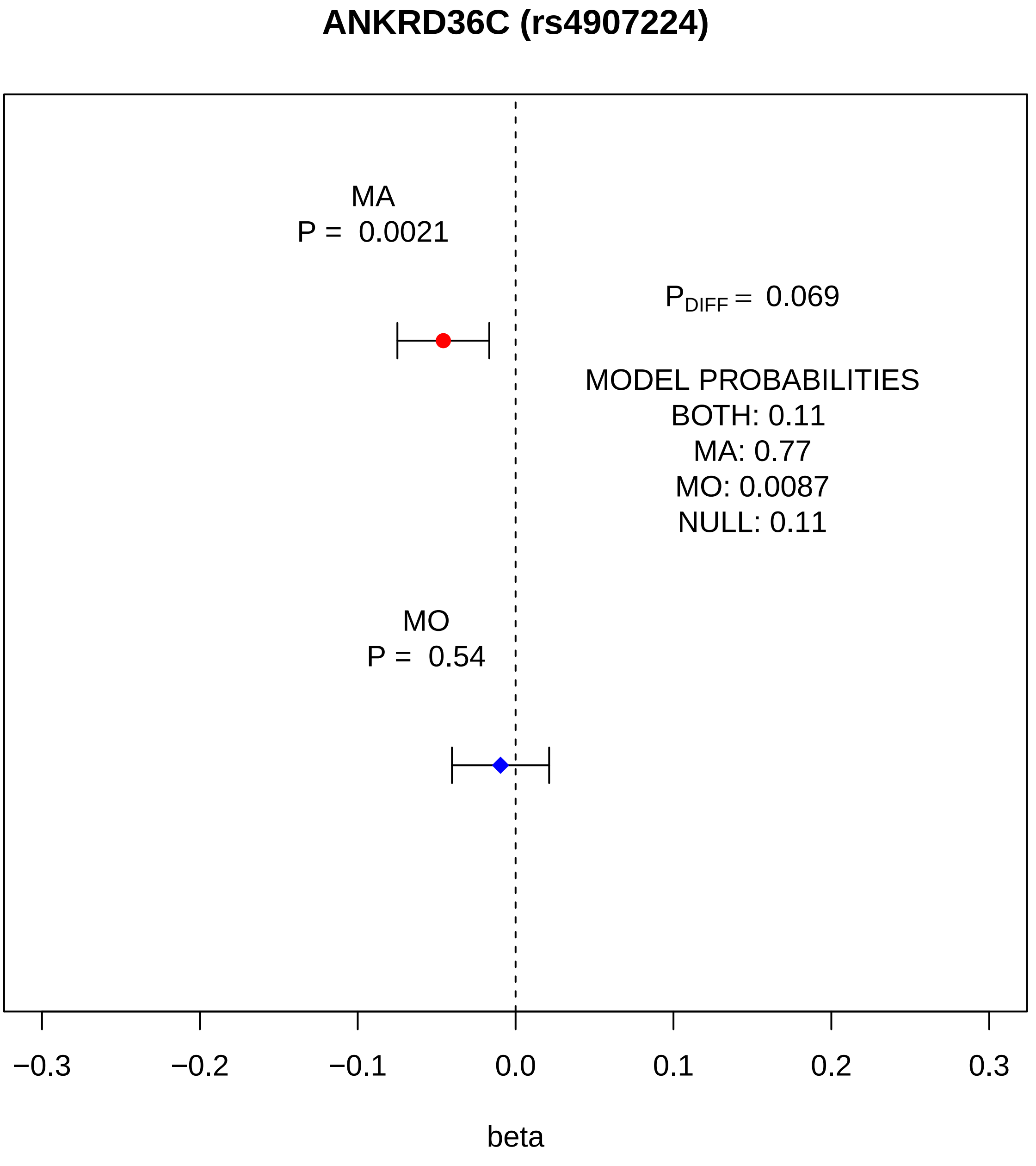
ANKRD36C (rs4907224)
MA
P = 0.0021
P DIFF = 0.069
MODEL PROBABILITIES
BOTH: 0.11
MA: 0.77
MO: 0.0087
NULL: 0.11
MO
P = 0.54
−0.3
−0.2
−0.1
0.0
0.1
0.2
0.3
beta

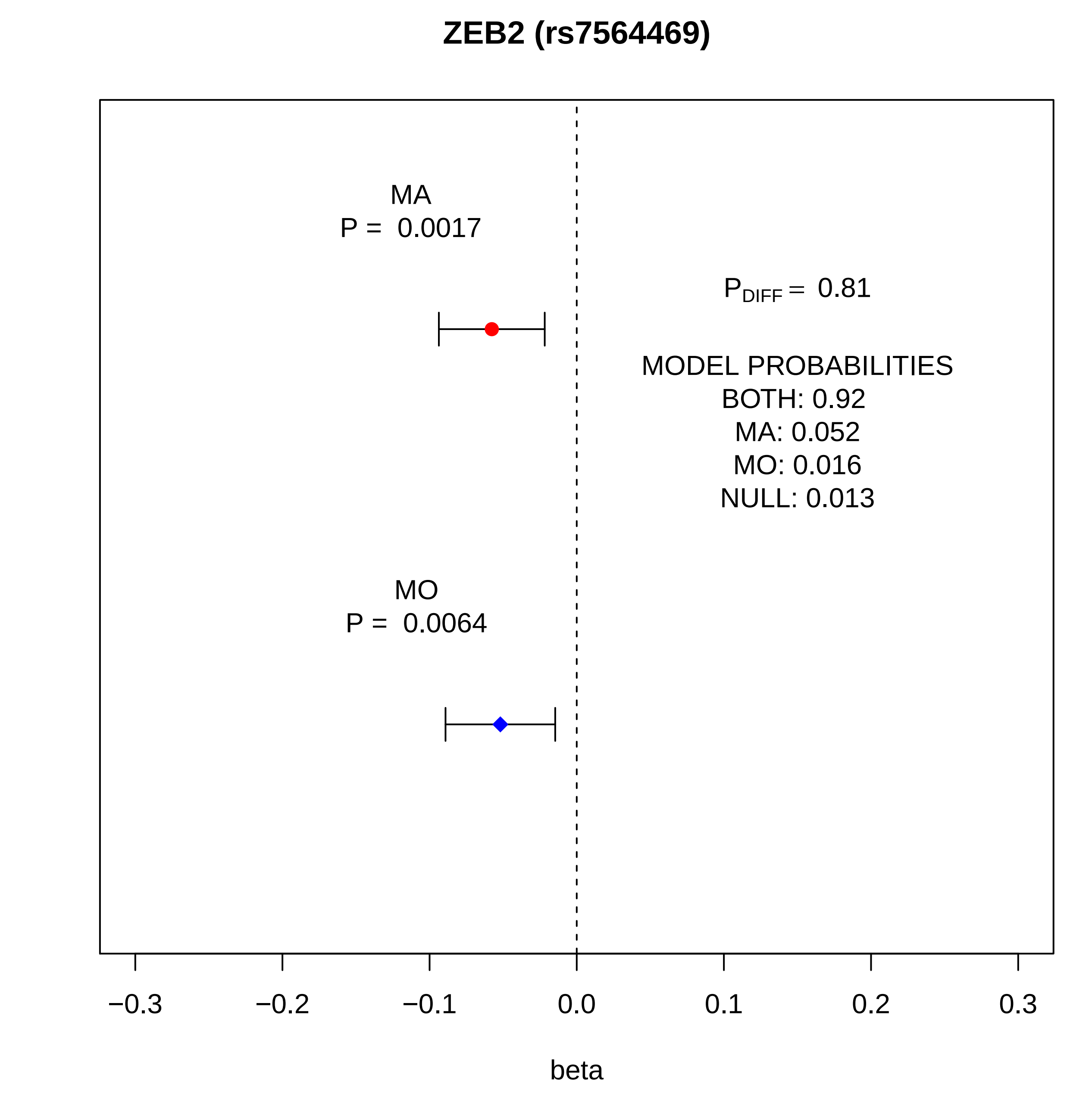
ZEB2 (rs7564469)
MA
P = 0.0017
$P_{DIFF}$ = 0.81
MODEL PROBABILITIES
BOTH: 0.92
MA: 0.052
MO: 0.016
NULL: 0.013
MO
P = 0.0064
−0.3
−0.2
−0.1
0.0
0.1
0.2
0.3
beta

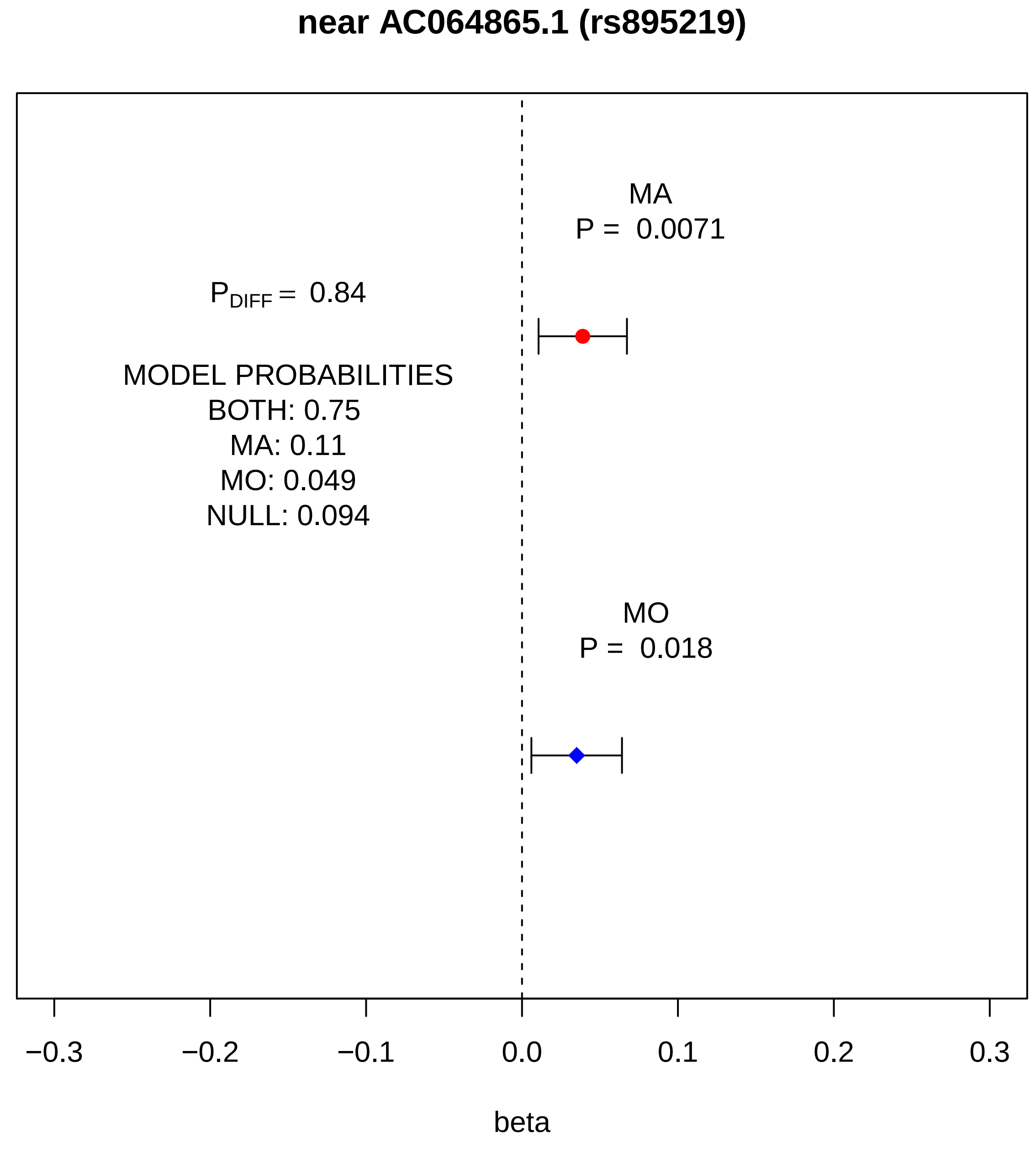

near AC064865.1 (rs895219)
MA
P = 0.0071
P_DIFF = 0.84
MODEL PROBABILITIES
BOTH: 0.75
MA: 0.11
MO: 0.049
NULL: 0.094
MO
P = 0.018
−0.3
−0.2
−0.1
0.0
0.1
0.2
0.3
beta

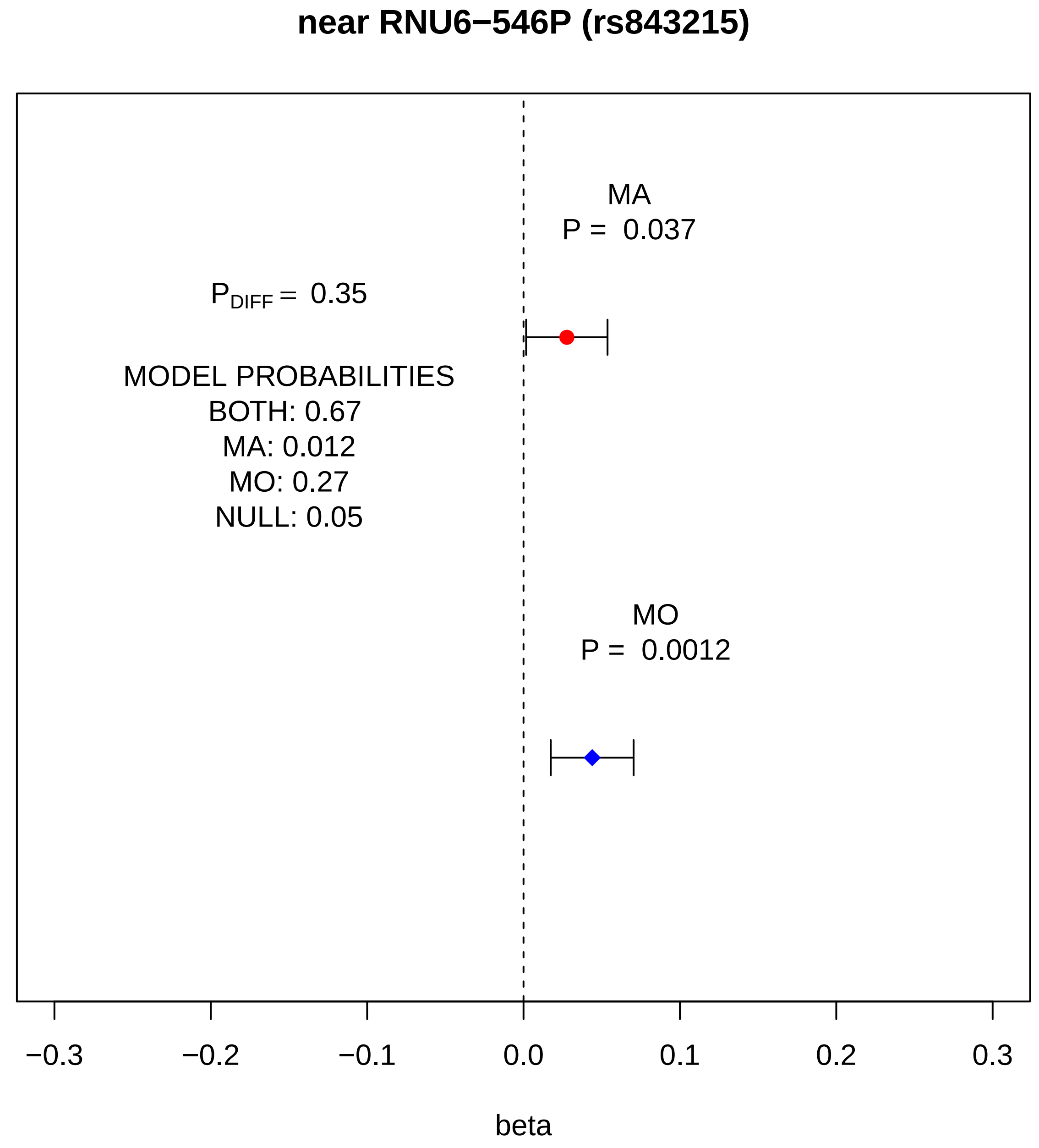

near RNU6−546P (rs843215)
MA
P = 0.037
P_DIFF = 0.35
MODEL PROBABILITIES
BOTH: 0.67
MA: 0.012
MO: 0.27
NULL: 0.05
MO
P = 0.0012
−0.3
−0.2
−0.1
0.0
0.1
0.2
0.3
beta

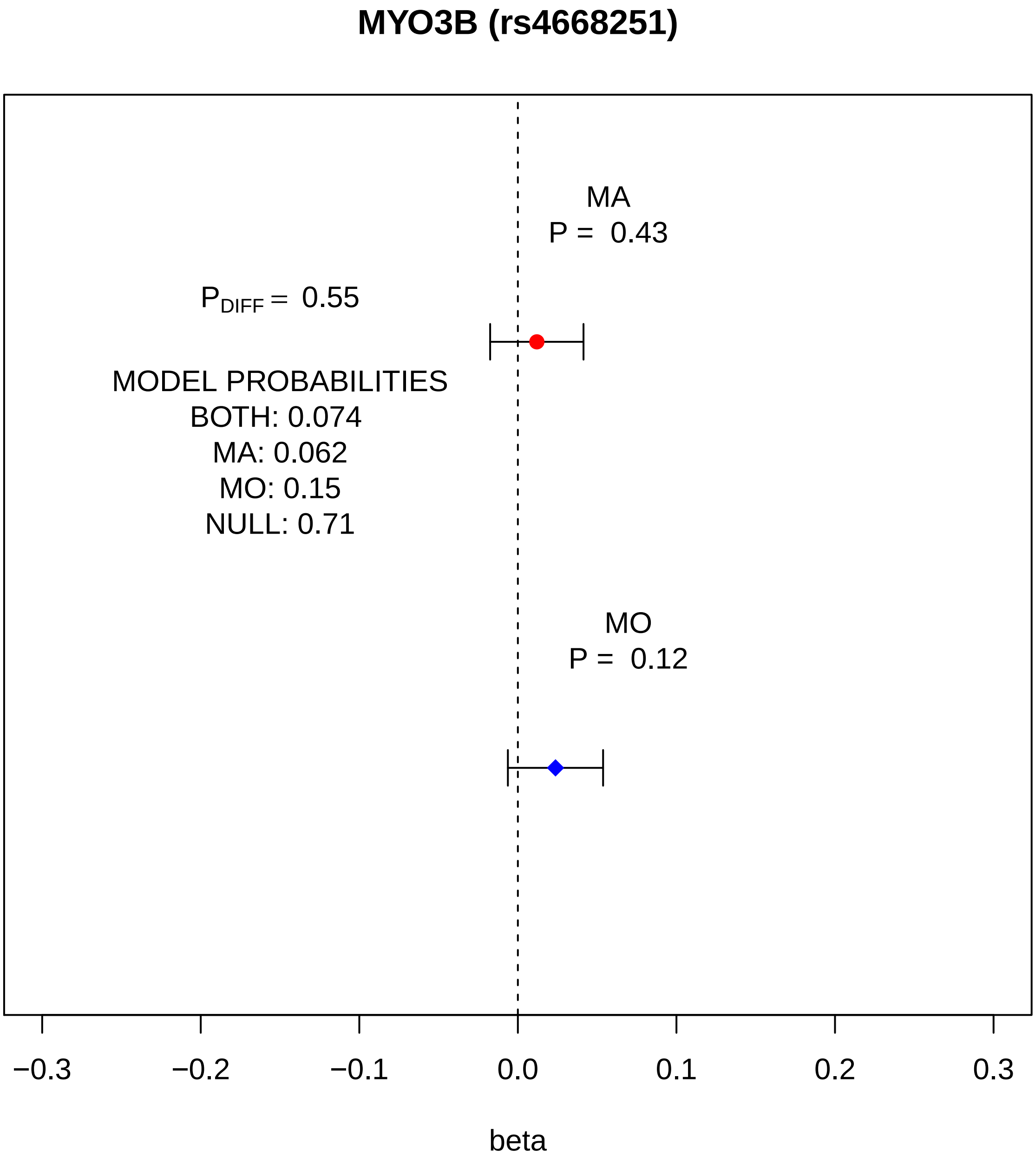

MYO3B (rs4668251)
MA
P = 0.43
P_DIFF = 0.55
MODEL PROBABILITIES
BOTH: 0.074
MA: 0.062
MO: 0.15
NULL: 0.71
MO
P = 0.12
−0.3
−0.2
−0.1
0.0
0.1
0.2
0.3
beta

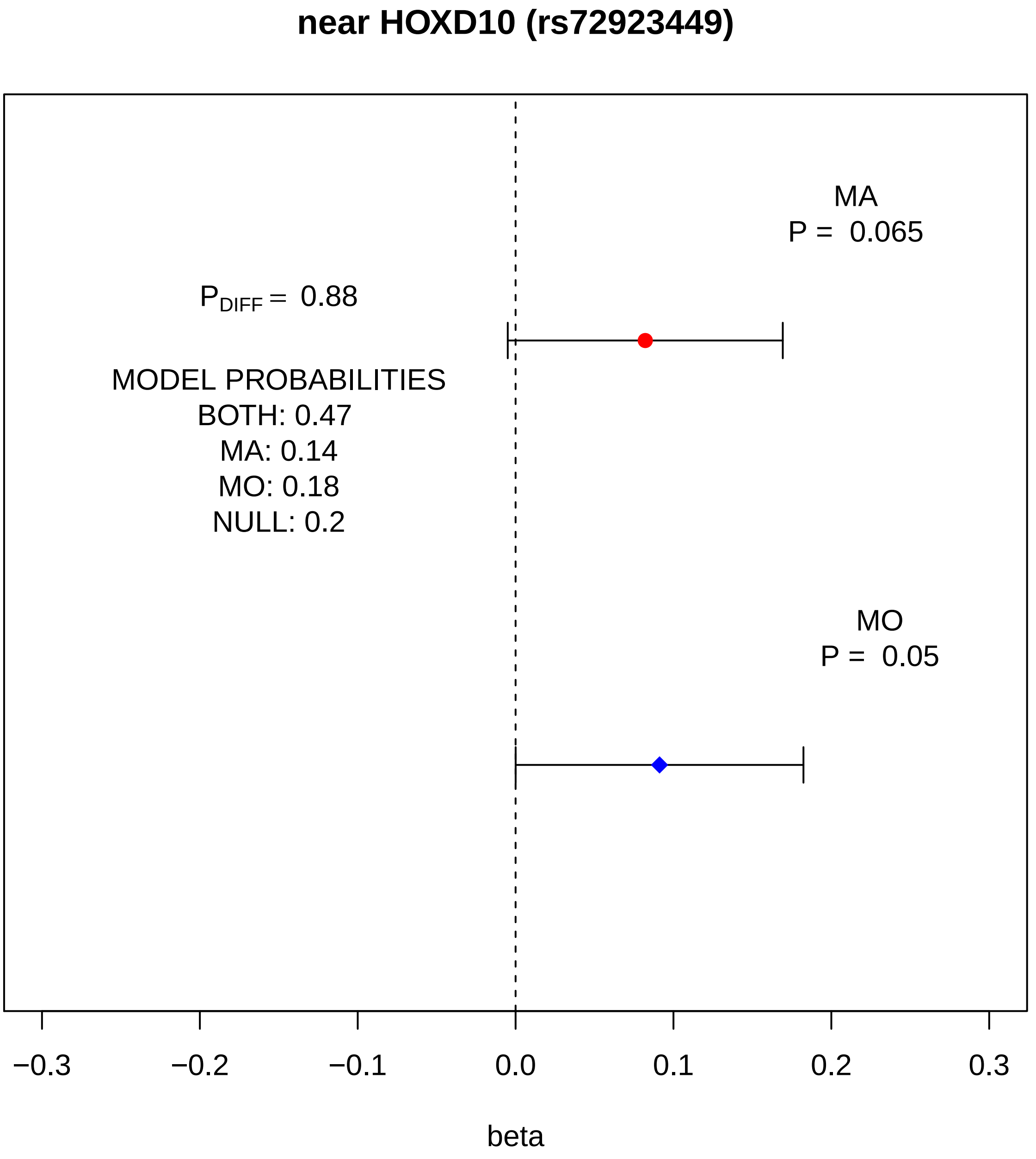
near HOXD10 (rs72923449)
MA
P = 0.065
P_DIFF = 0.88
MODEL PROBABILITIES
BOTH: 0.47
MA: 0.14
MO: 0.18
NULL: 0.2
MO
P = 0.05
−0.3
−0.2
−0.1
0.0
0.1
0.2
0.3
beta

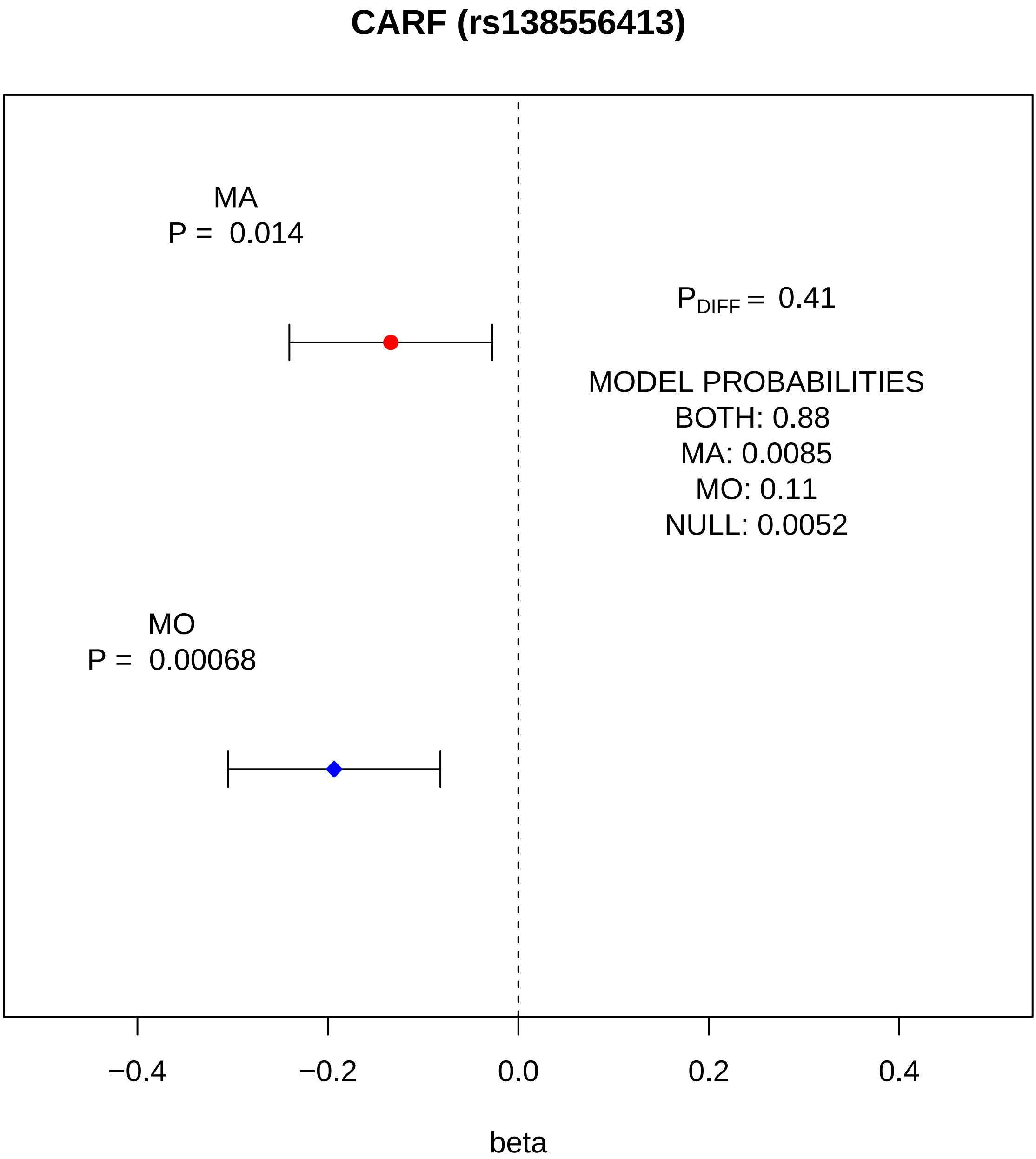
CARF (rs138556413)
MA
P = 0.014
$P_{DIFF}$ = 0.41
MODEL PROBABILITIES
BOTH: 0.88
MA: 0.0085
MO: 0.11
NULL: 0.0052
MO
P = 0.00068
−0.4
−0.2
0.0
0.2
0.4
beta

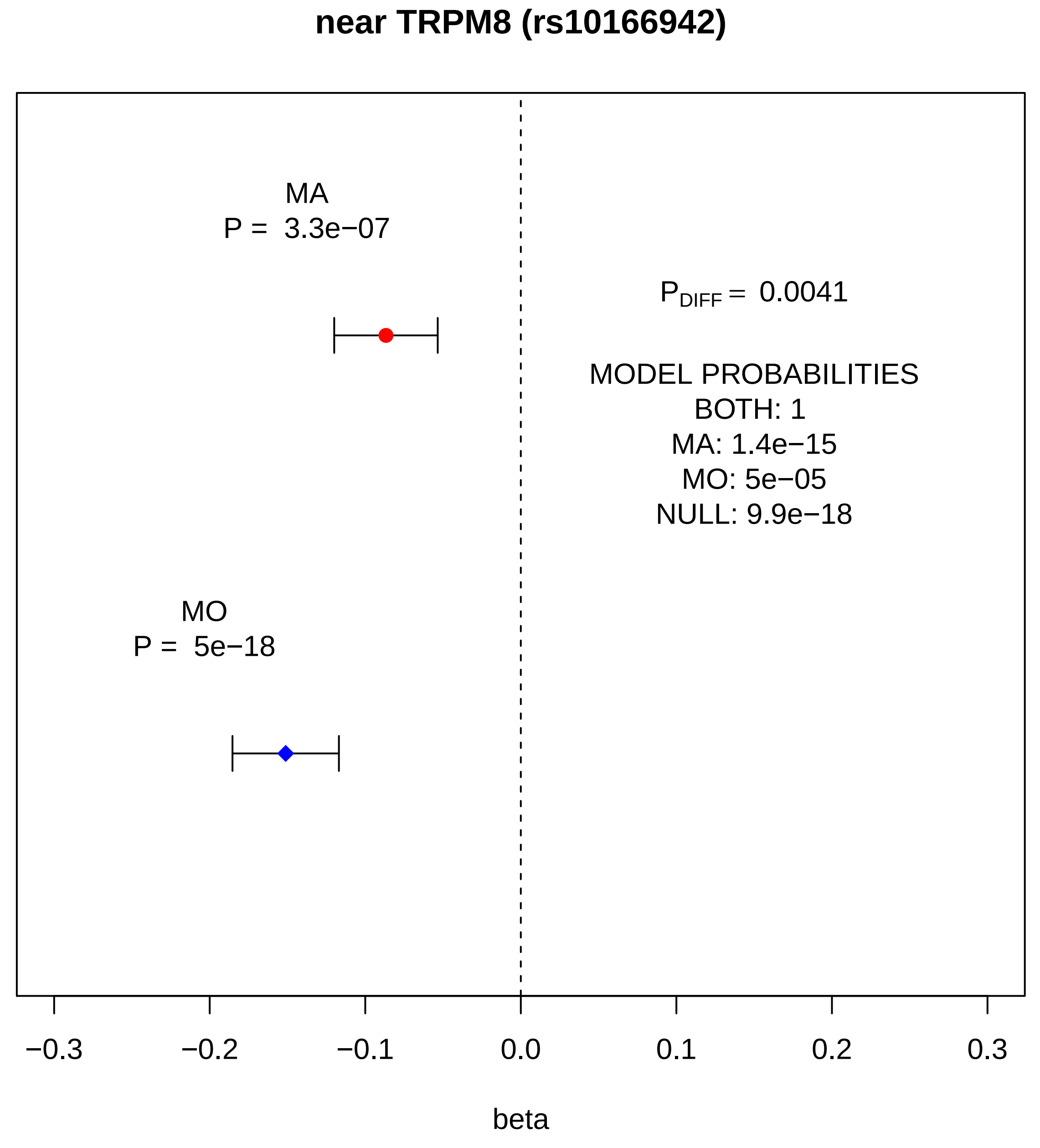
near TRPM8 (rs10166942)
MA
P = 3.3e−07
$P_{DIFF}$ = 0.0041
MODEL PROBABILITIES
BOTH: 1
MA: 1.4e−15
MO: 5e−05
NULL: 9.9e−18
MO
P = 5e−18
−0.3
−0.2
−0.1
0.0
0.1
0.2
0.3
beta

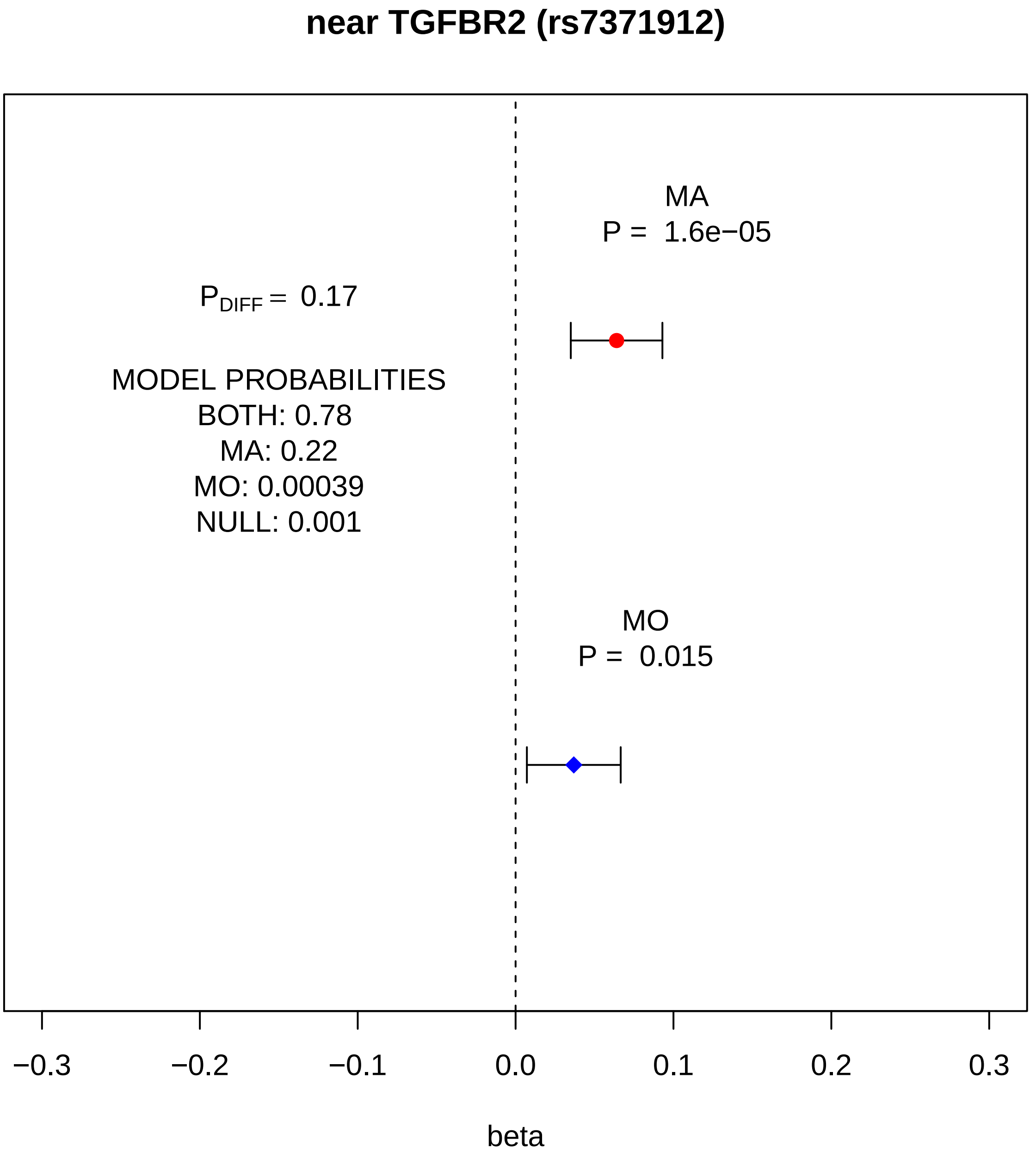
near TGFBR2 (rs7371912)
MA
P = 1.6e−05
P_DIFF = 0.17
MODEL PROBABILITIES
BOTH: 0.78
MA: 0.22
MO: 0.00039
NULL: 0.001
MO
P = 0.015
−0.3
−0.2
−0.1
0.0
0.1
0.2
0.3
beta

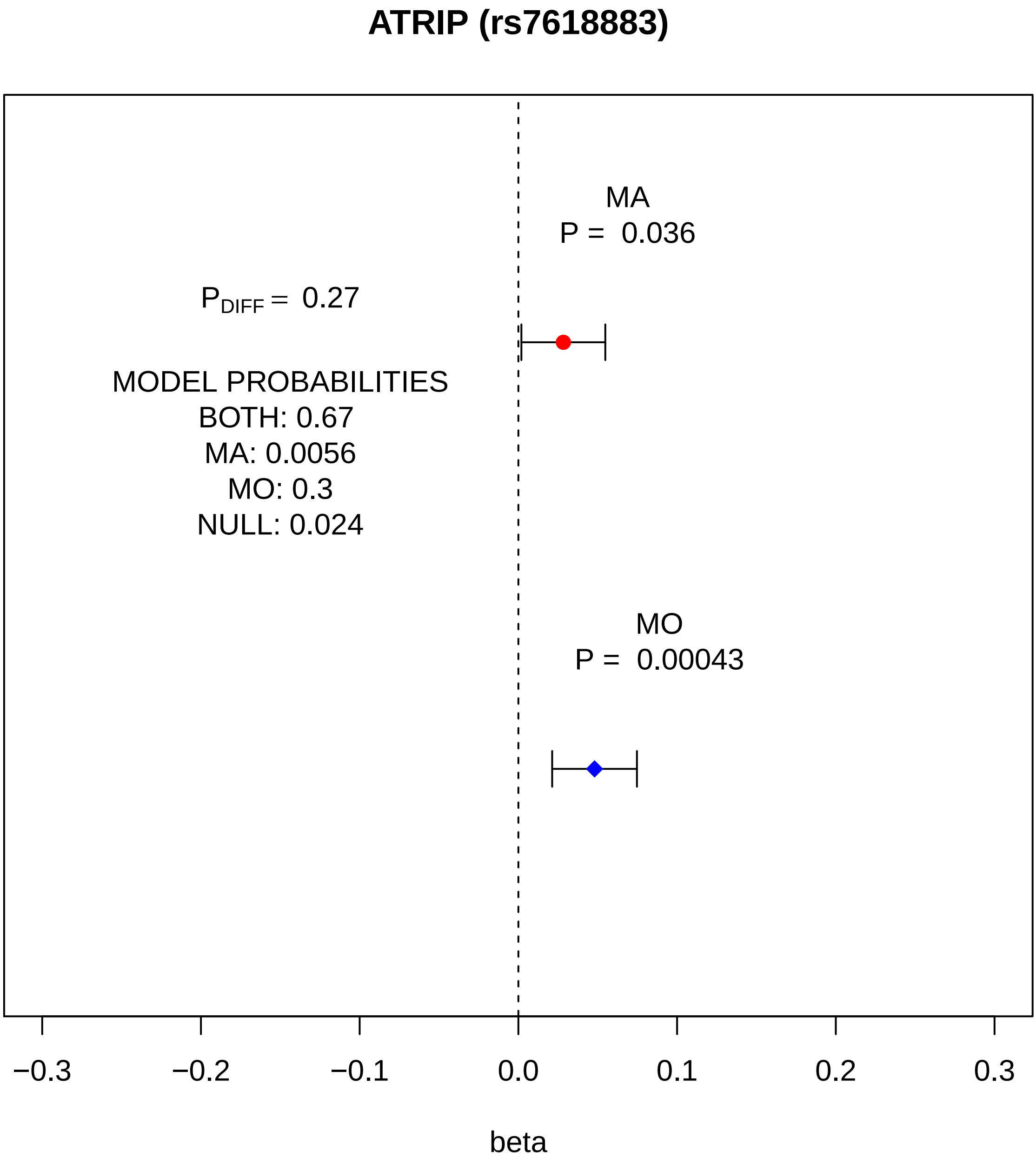

ATRIP (rs7618883)
MA
P = 0.036
P_DIFF = 0.27
MODEL PROBABILITIES
BOTH: 0.67
MA: 0.0056
MO: 0.3
NULL: 0.024
MO
P = 0.00043
−0.3
−0.2
−0.1
0.0
0.1
0.2
0.3
beta

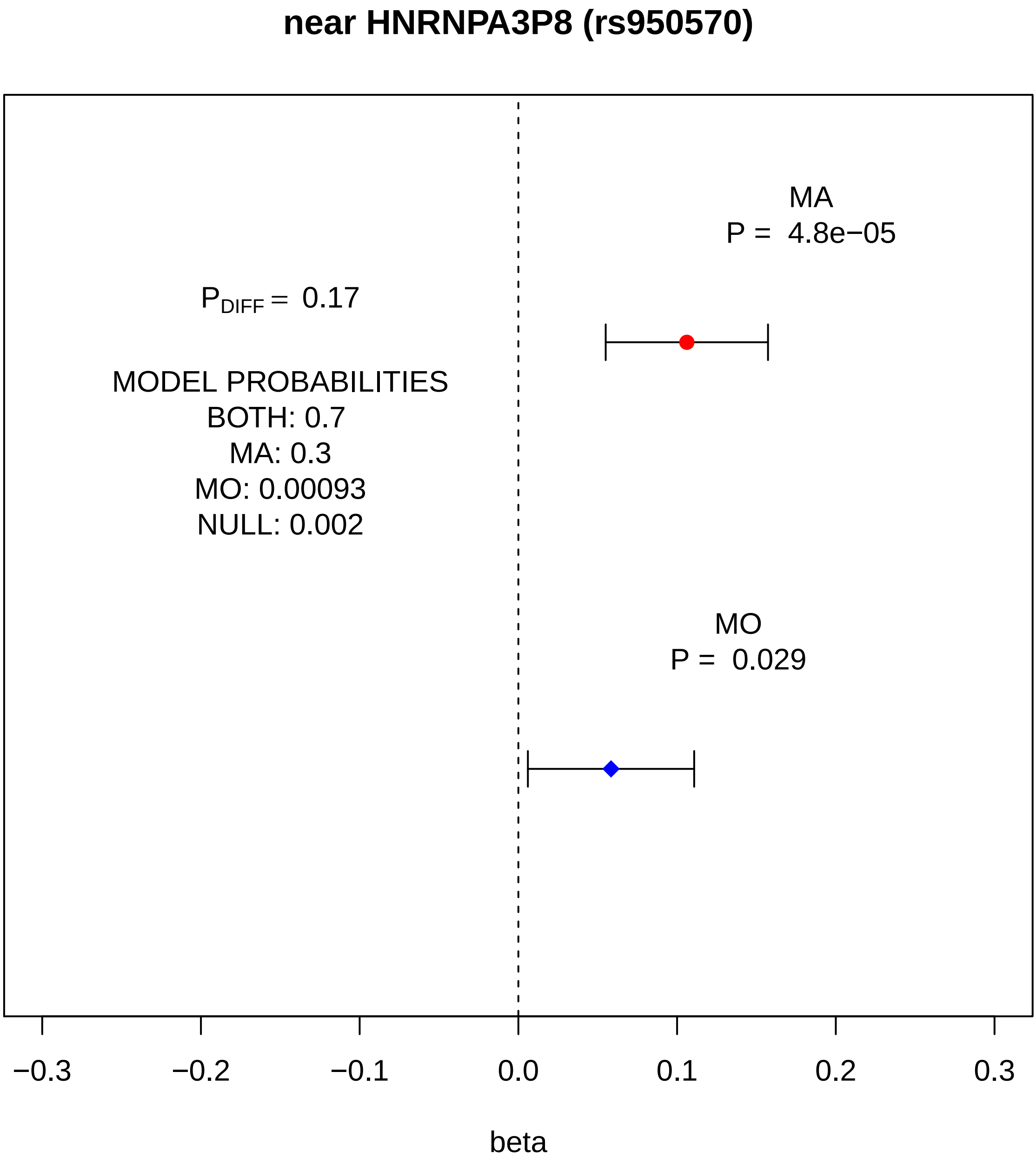
near HNRNPA3P8 (rs950570)
MA
P = 4.8e−05
P_DIFF = 0.17
MODEL PROBABILITIES
BOTH: 0.7
MA: 0.3
MO: 0.00093
NULL: 0.002
MO
P = 0.029
−0.3
−0.2
−0.1
0.0
0.1
0.2
0.3
beta

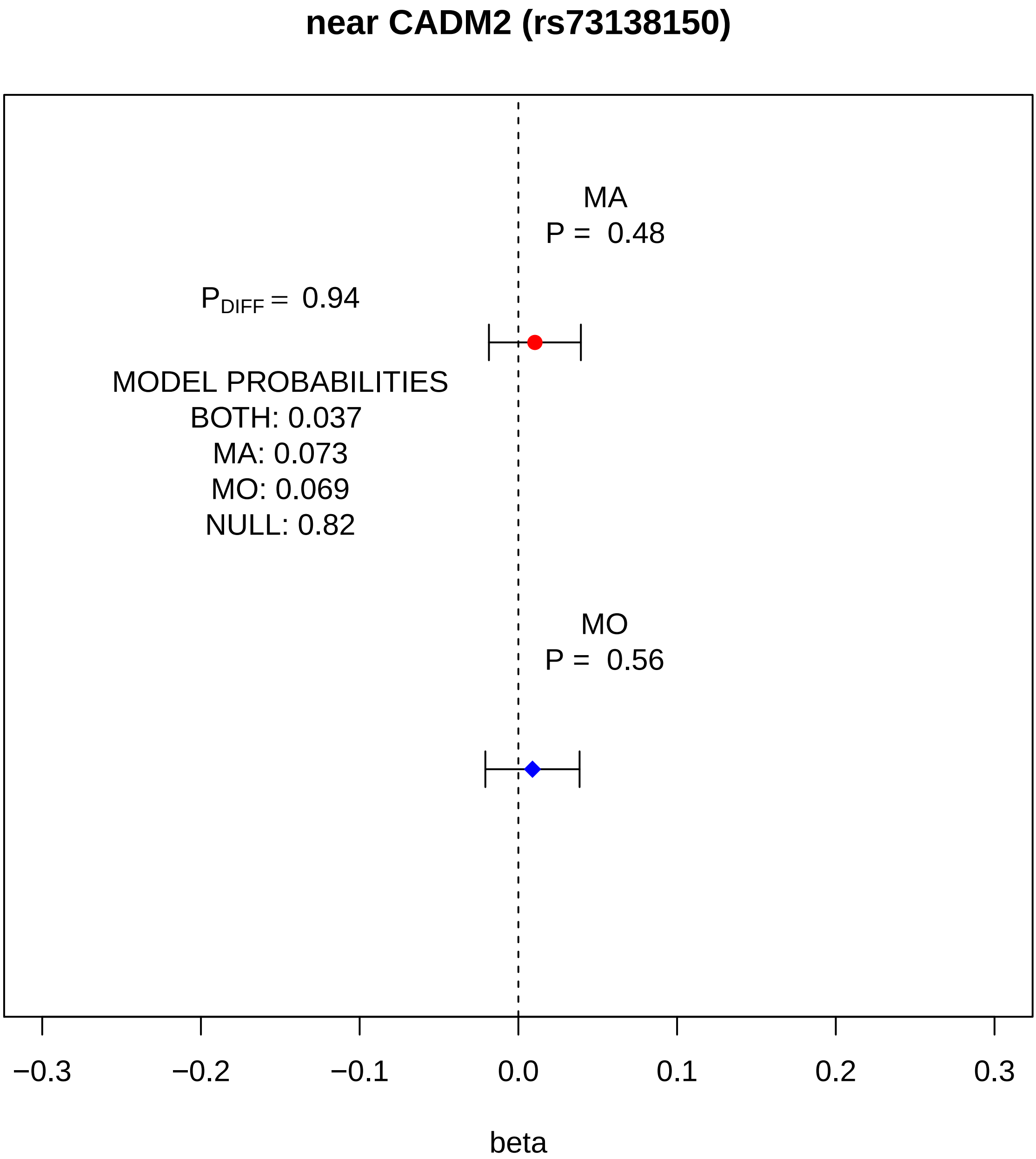
near CADM2 (rs73138150)
MA
P = 0.48
$P_{DIFF}$ = 0.94
MODEL PROBABILITIES
BOTH: 0.037
MA: 0.073
MO: 0.069
NULL: 0.82
MO
P = 0.56
−0.3
−0.2
−0.1
0.0
0.1
0.2
0.3
beta

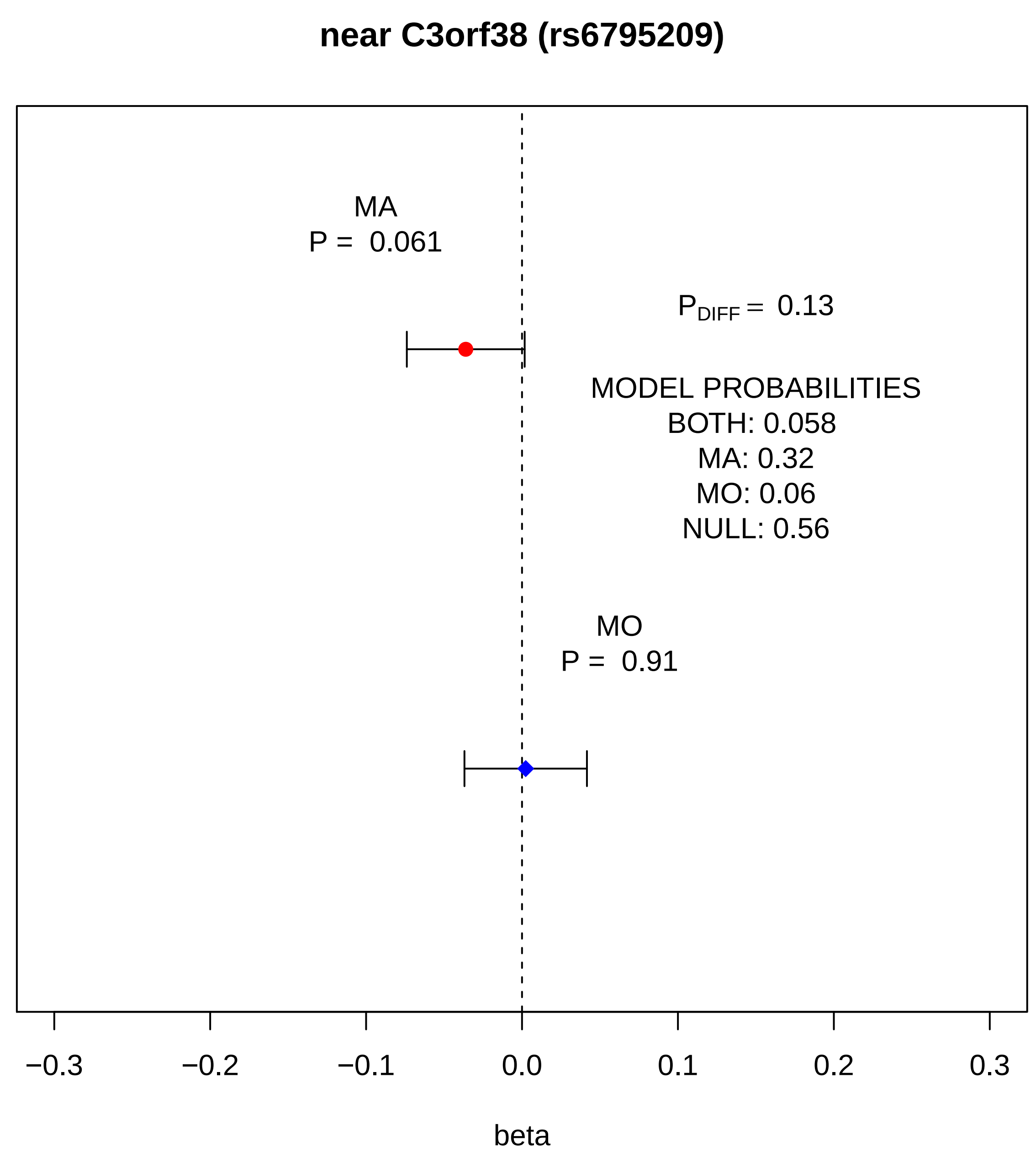
near C3orf38 (rs6795209)
MA
P = 0.061
P_DIFF = 0.13
MODEL PROBABILITIES
BOTH: 0.058
MA: 0.32
MO: 0.06
NULL: 0.56
MO
P = 0.91
−0.3
−0.2
−0.1
0.0
0.1
0.2
0.3
beta

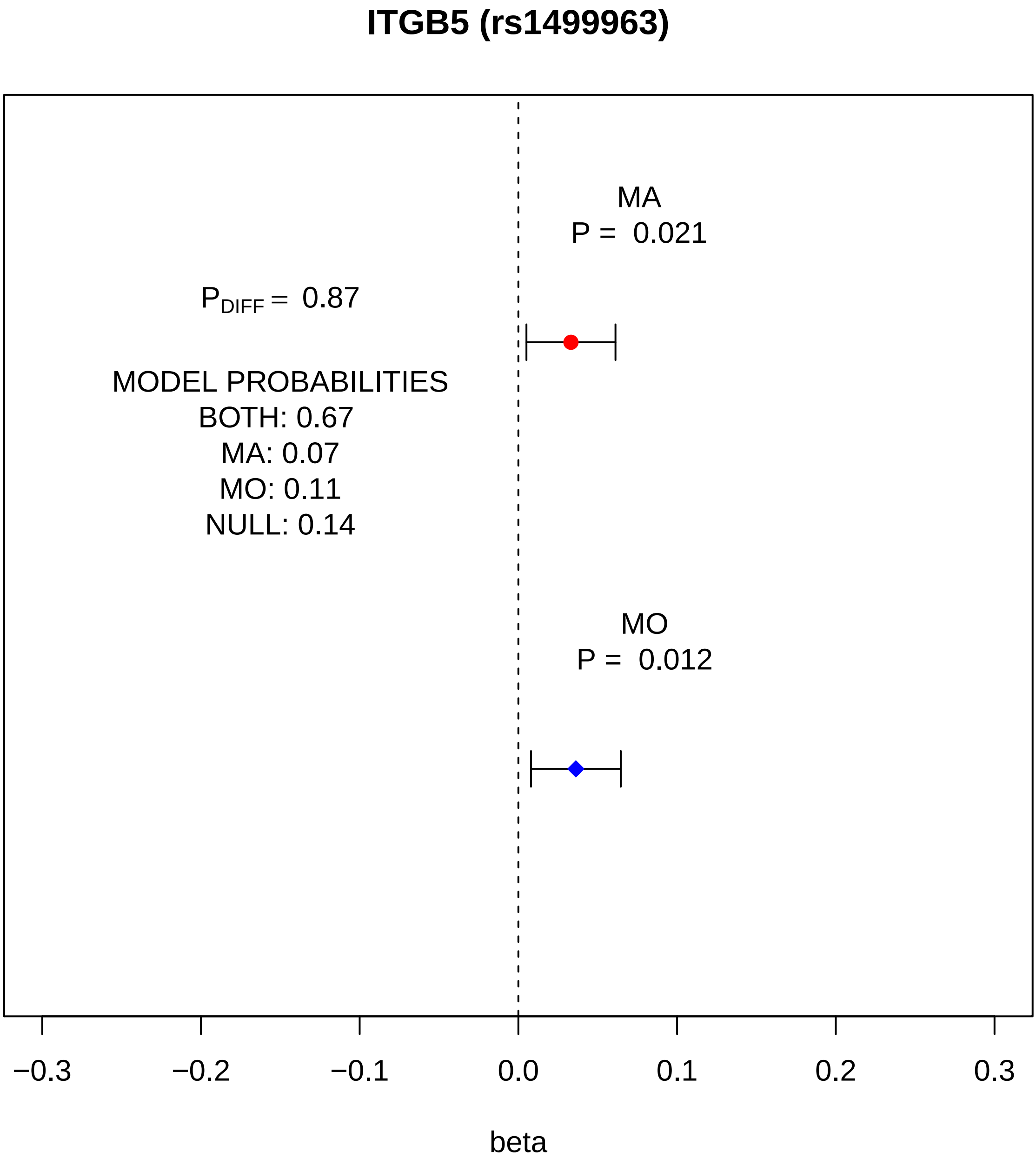
ITGB5 (rs1499963)
MA
P = 0.021
P_DIFF = 0.87
MODEL PROBABILITIES
BOTH: 0.67
MA: 0.07
MO: 0.11
NULL: 0.14
MO
P = 0.012
−0.3
−0.2
−0.1
0.0
0.1
0.2
0.3
beta

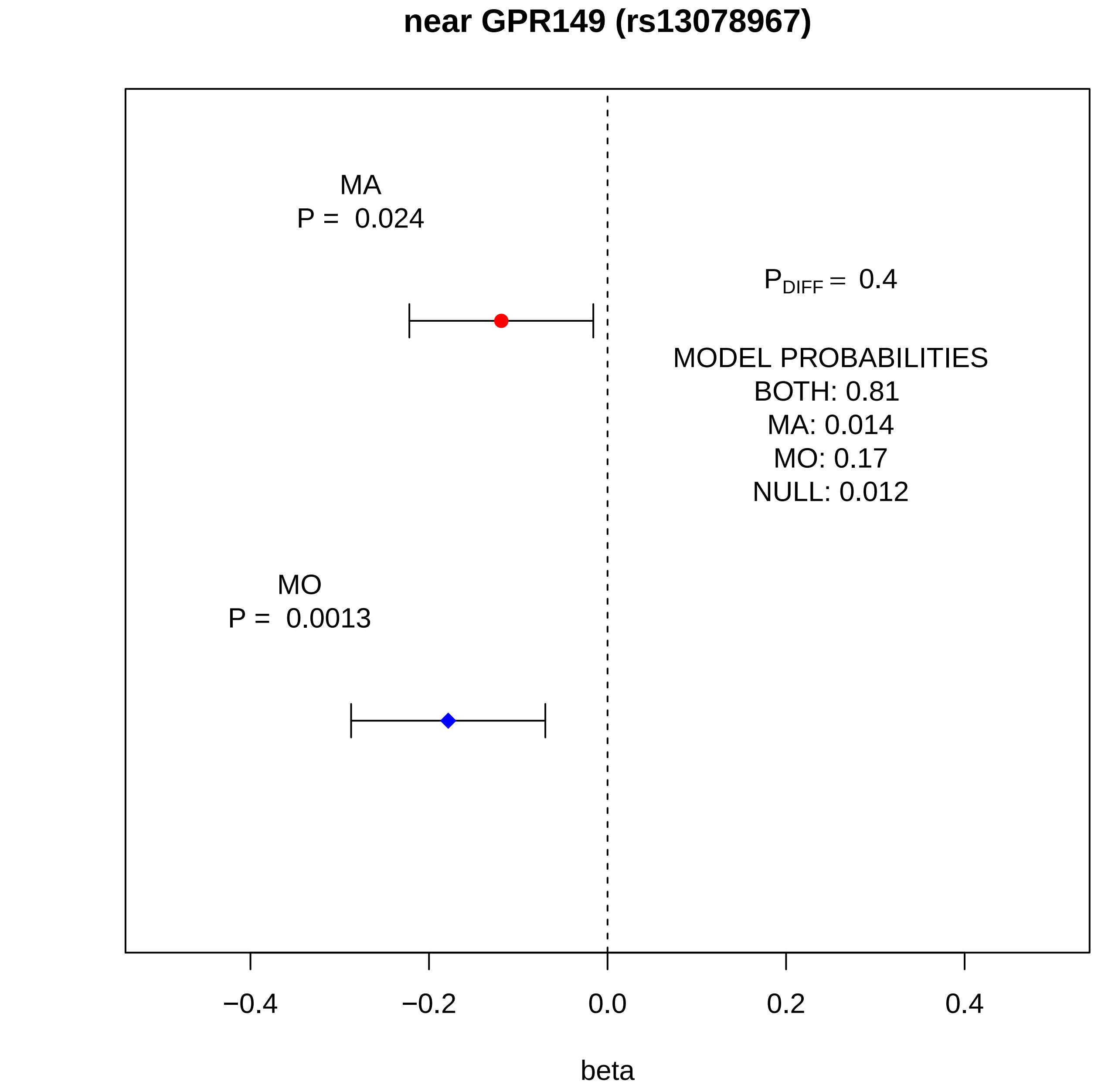

near GPR149 (rs13078967)
MA
P = 0.024
$P_{DIFF}$ = 0.4
MODEL PROBABILITIES
BOTH: 0.81
MA: 0.014
MO: 0.17
NULL: 0.012
MO
P = 0.0013
−0.4
−0.2
0.0
0.2
0.4
beta

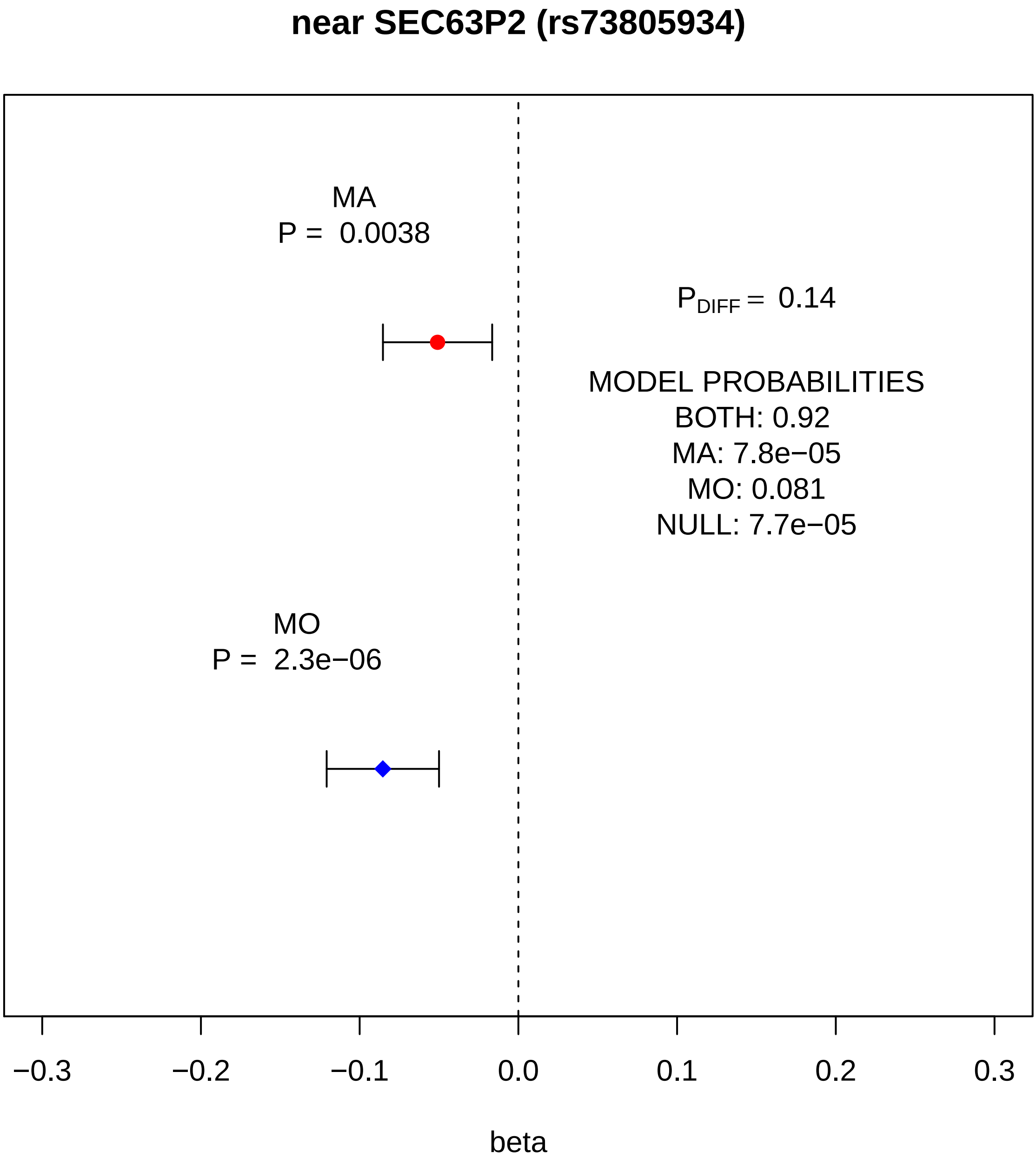
near SEC63P2 (rs73805934)
MA
P = 0.0038
$P_{DIFF}$ = 0.14
MODEL PROBABILITIES
BOTH: 0.92
MA: 7.8e−05
MO: 0.081
NULL: 7.7e−05
MO
P = 2.3e−06
−0.3
−0.2
−0.1
0.0
0.1
0.2
0.3
beta

#### near SPINK2 (rs7684253)

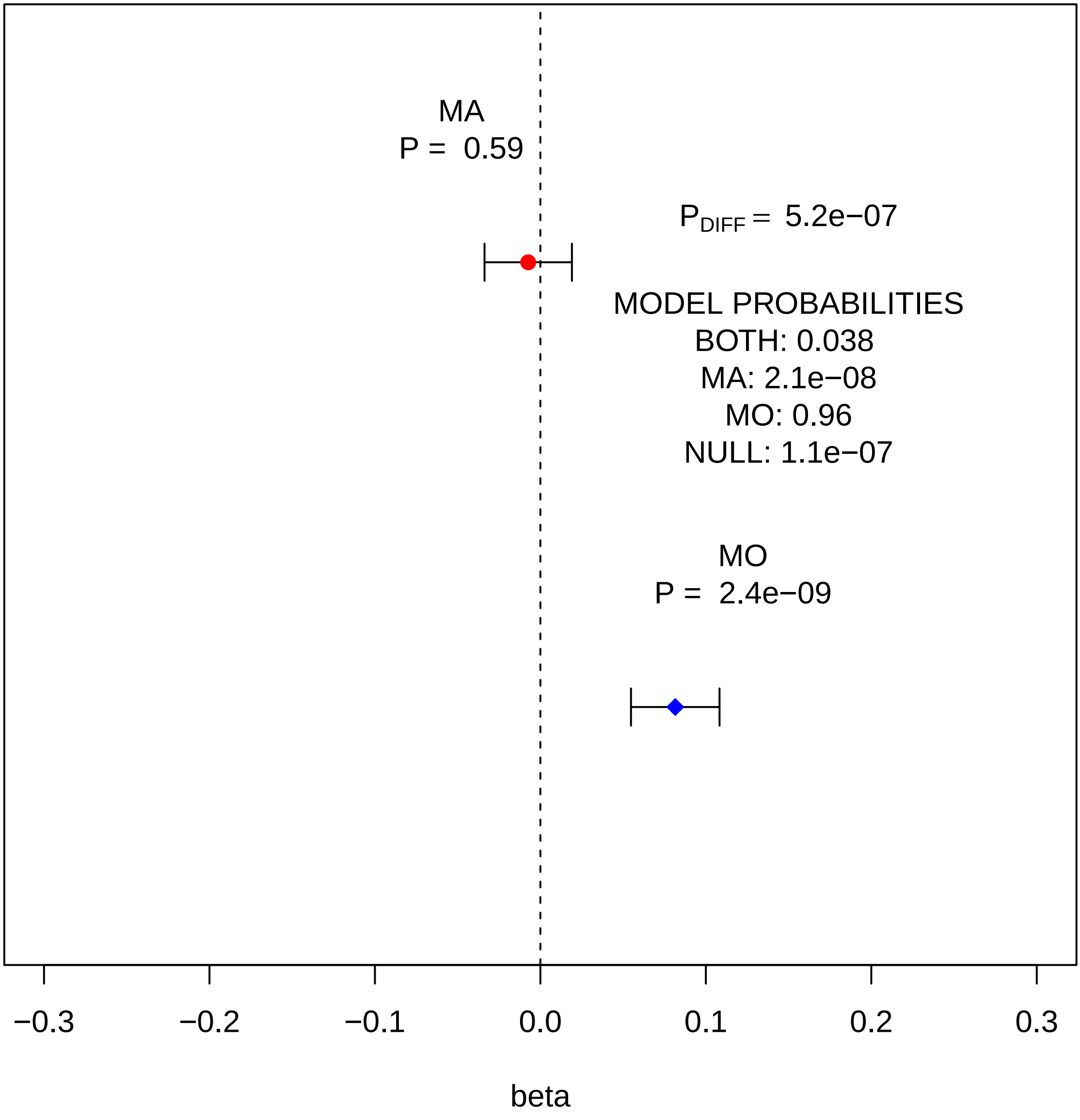

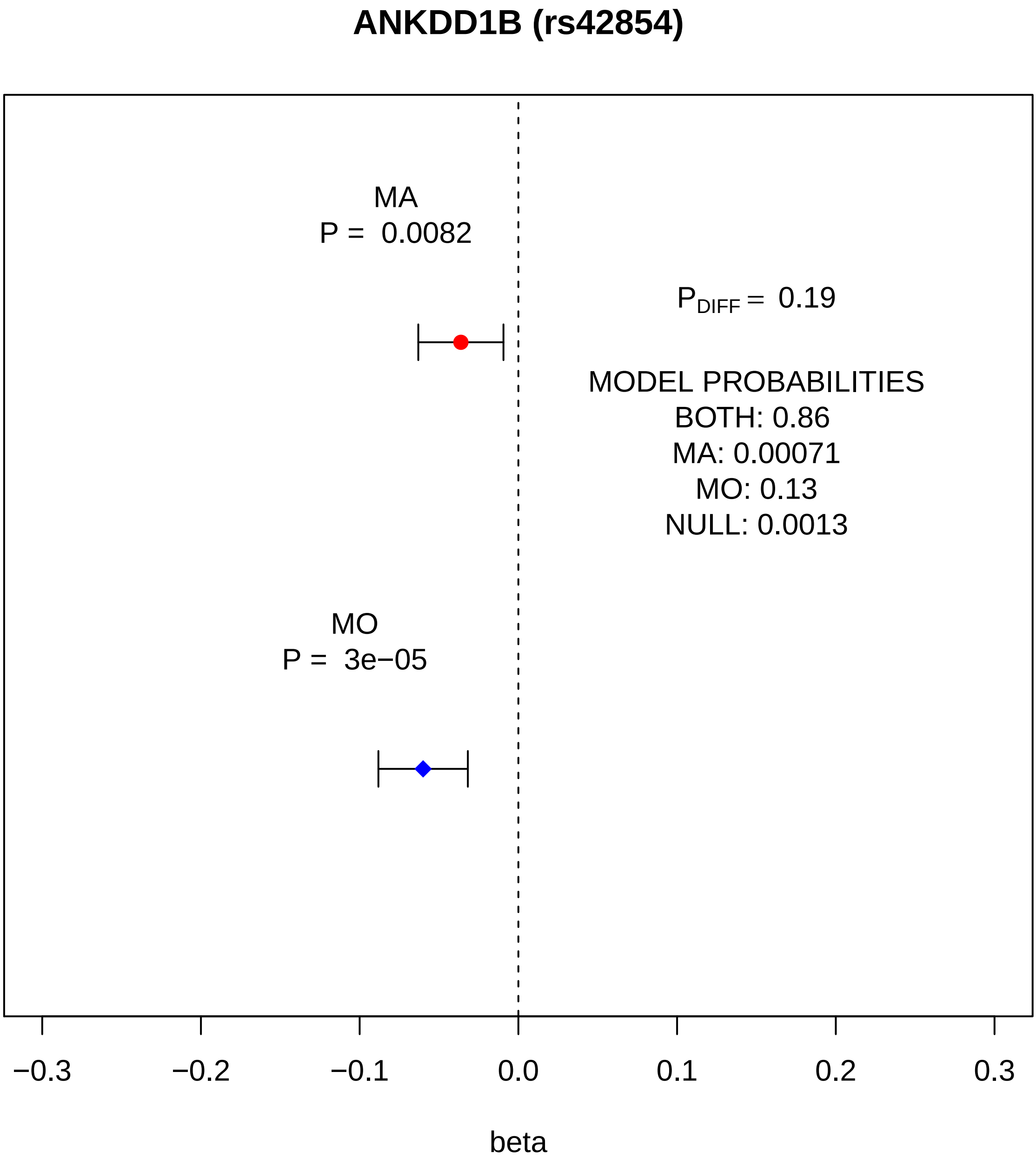
ANKDD1B (rs42854)
MA
P = 0.0082
$P_{DIFF}$ = 0.19
MODEL PROBABILITIES
BOTH: 0.86
MA: 0.00071
MO: 0.13
NULL: 0.0013
MO
P = 3e−05
−0.3
−0.2
−0.1
0.0
0.1
0.2
0.3
beta

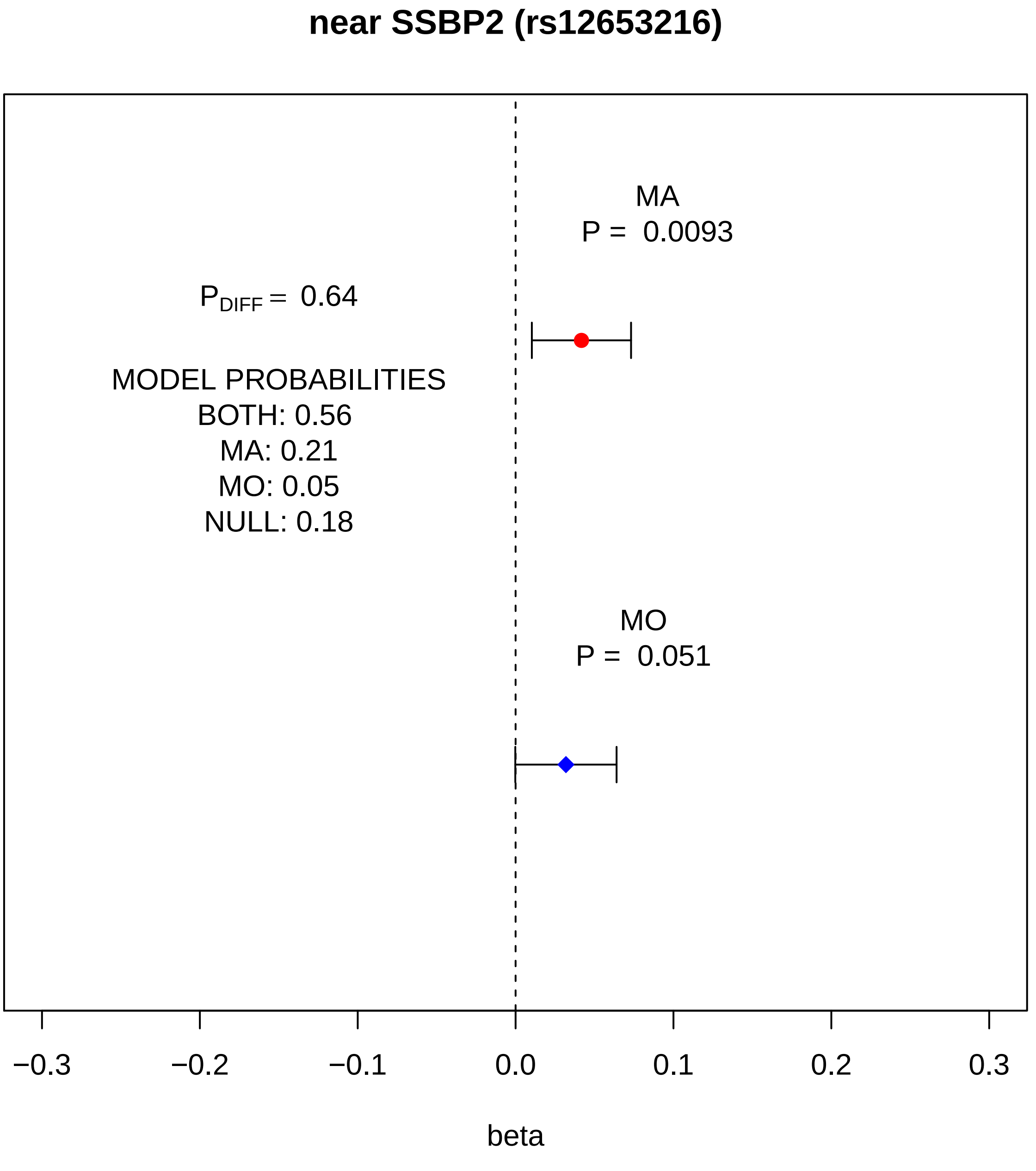
near SSBP2 (rs12653216)
MA
P = 0.0093
$P_{DIFF}$ = 0.64
MODEL PROBABILITIES
BOTH: 0.56
MA: 0.21
MO: 0.05
NULL: 0.18
MO
P = 0.051
−0.3
−0.2
−0.1
0.0
0.1
0.2
0.3
beta

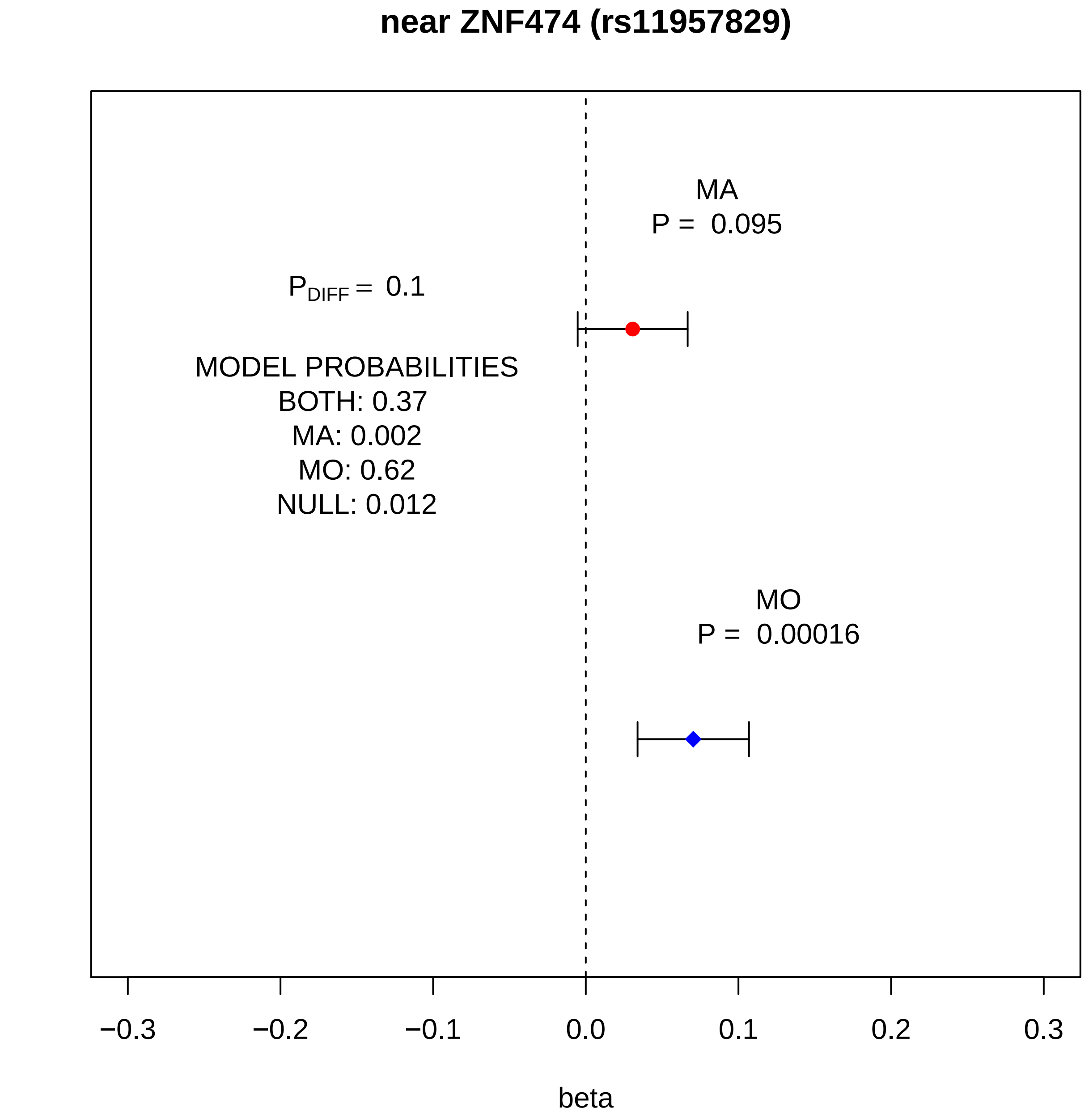
near ZNF474 (rs11957829)
MA
P = 0.095
P_DIFF = 0.1
MODEL PROBABILITIES
BOTH: 0.37
MA: 0.002
MO: 0.62
NULL: 0.012
MO
P = 0.00016
−0.3
−0.2
−0.1
0.0
0.1
0.2
0.3
beta

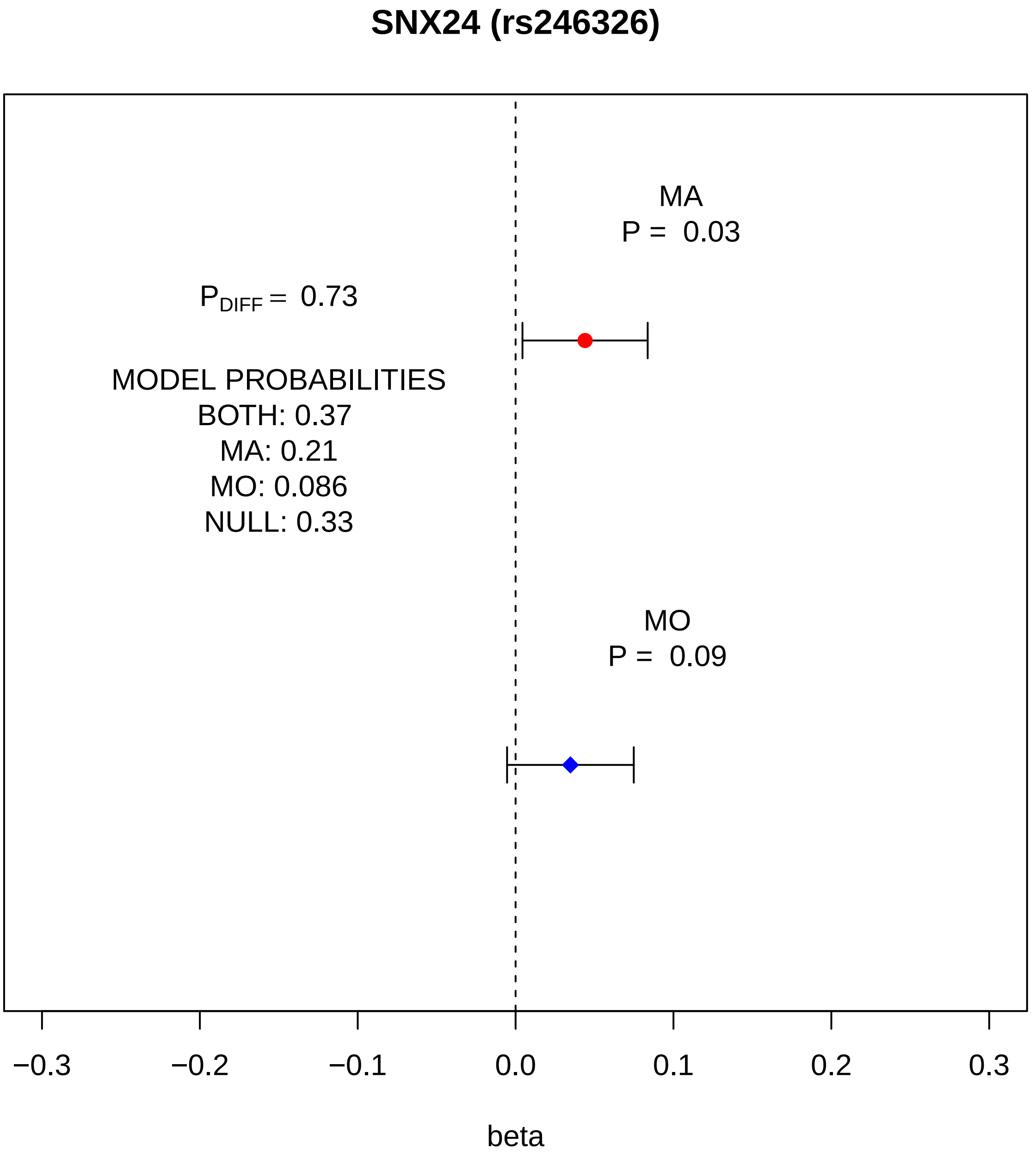
SNX24 (rs246326)
MA
P = 0.03
P_DIFF = 0.73
MODEL PROBABILITIES
BOTH: 0.37
MA: 0.21
MO: 0.086
NULL: 0.33
MO
P = 0.09
−0.3
−0.2
−0.1
0.0
0.1
0.2
0.3
beta

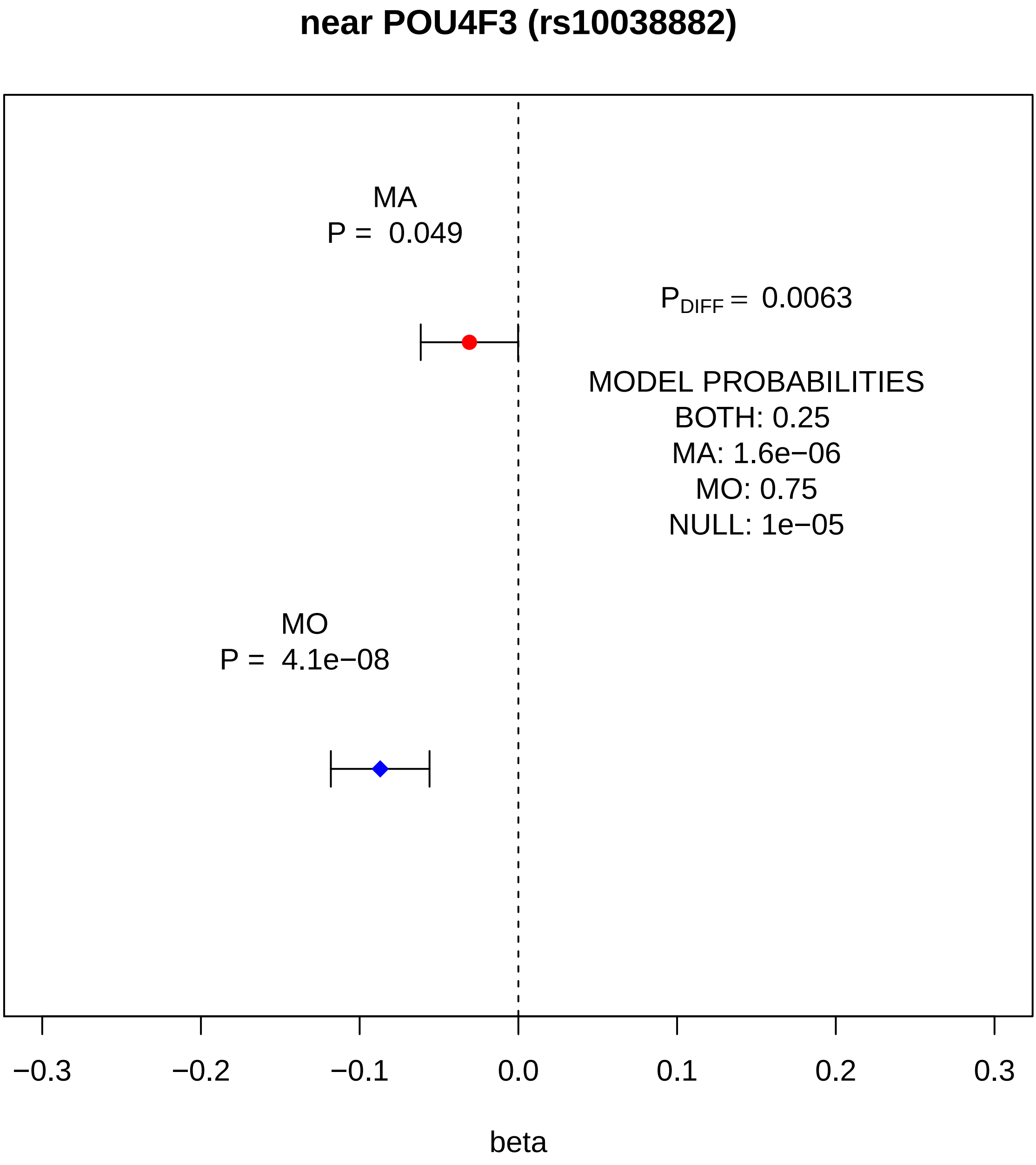
near POU4F3 (rs10038882)
MA
P = 0.049
$P_{DIFF}$ = 0.0063
MODEL PROBABILITIES
BOTH: 0.25
MA: 1.6e−06
MO: 0.75
NULL: 1e−05
MO
P = 4.1e−08
−0.3
−0.2
−0.1
0.0
0.1
0.2
0.3
beta

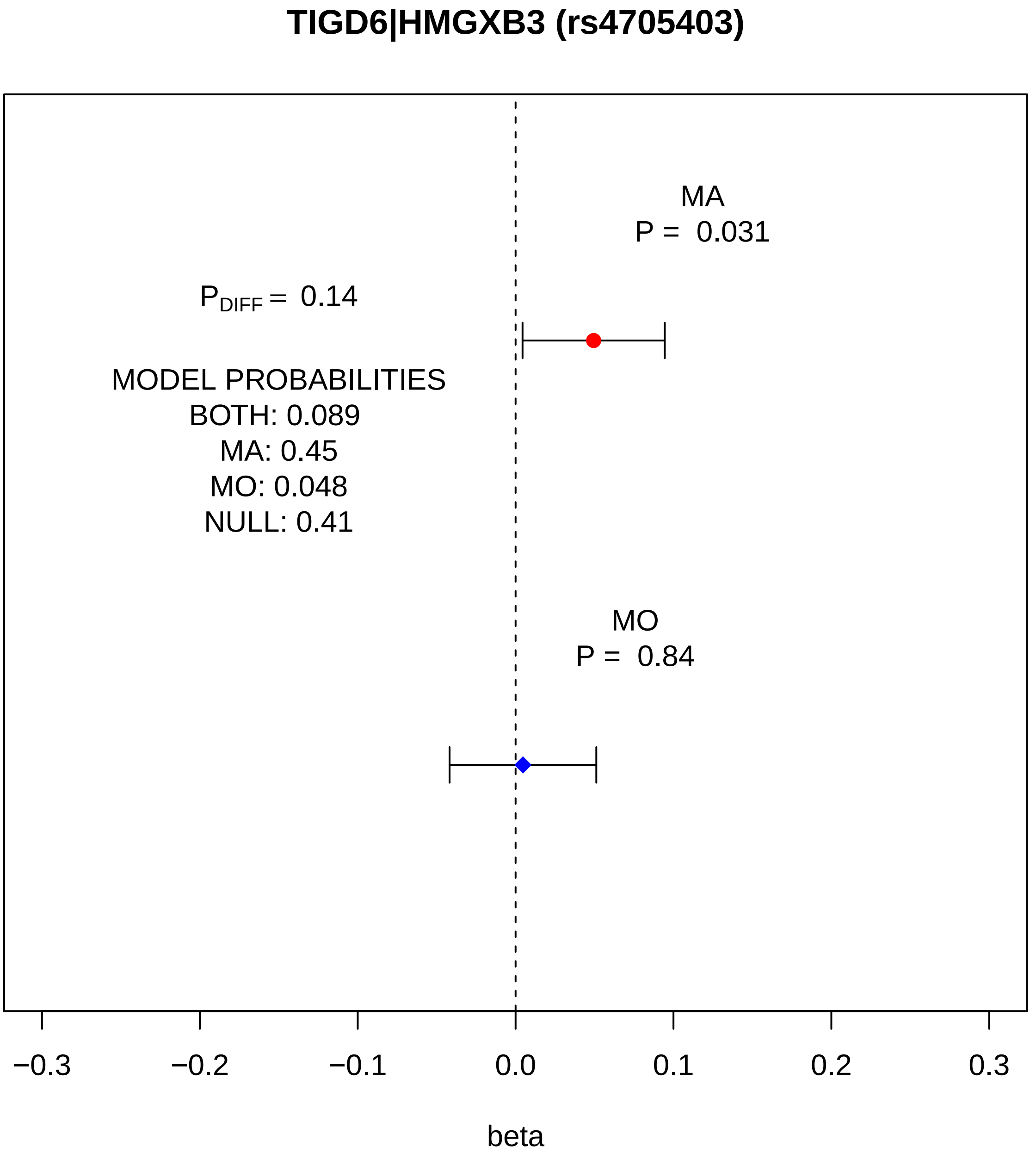
TIGD6|HMGXB3 (rs4705403)
MA
P = 0.031
P DIFF = 0.14
MODEL PROBABILITIES
BOTH: 0.089
MA: 0.45
MO: 0.048
NULL: 0.41
MO
P = 0.84
−0.3
−0.2
−0.1
0.0
0.1
0.2
0.3
beta

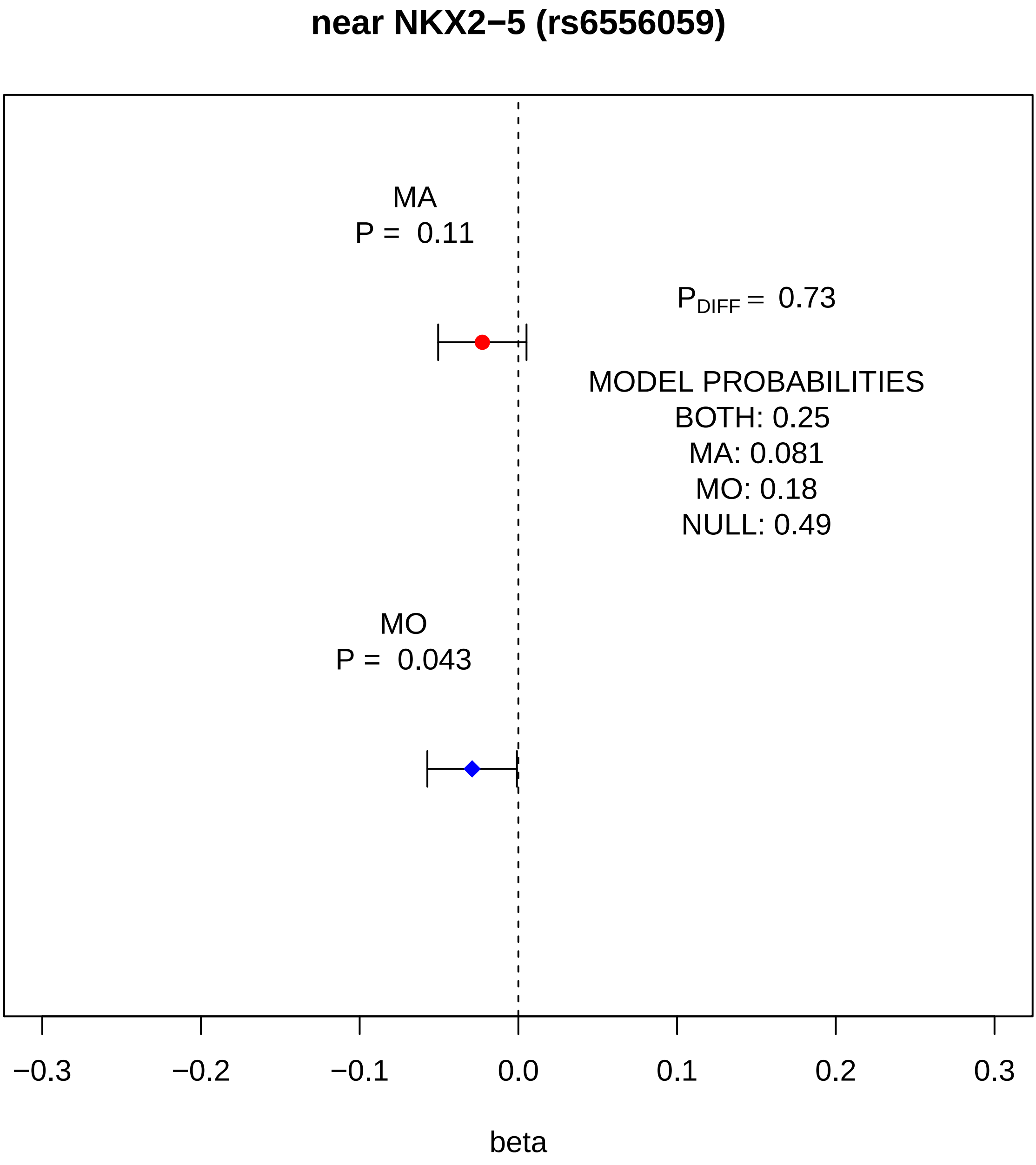

near NKX2−5 (rs6556059)
MA
P = 0.11
P_DIFF = 0.73
MODEL PROBABILITIES
BOTH: 0.25
MA: 0.081
MO: 0.18
NULL: 0.49
MO
P = 0.043
−0.3
−0.2
−0.1
0.0
0.1
0.2
0.3
beta

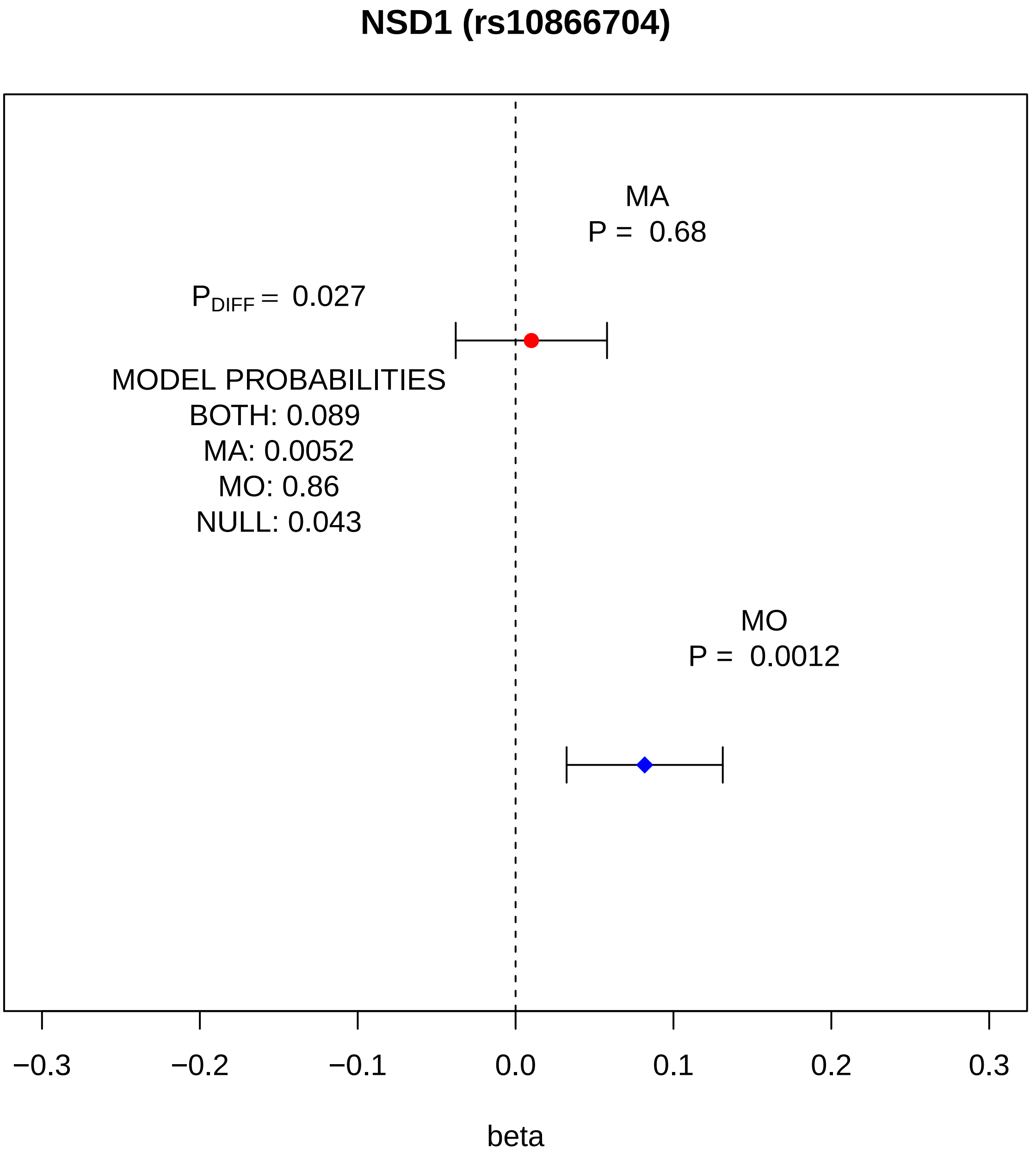
NSD1 (rs10866704)
MA
P = 0.68
$P_{DIFF}$ = 0.027
MODEL PROBABILITIES
BOTH: 0.089
MA: 0.0052
MO: 0.86
NULL: 0.043
MO
P = 0.0012
−0.3
−0.2
−0.1
0.0
0.1
0.2
0.3
beta

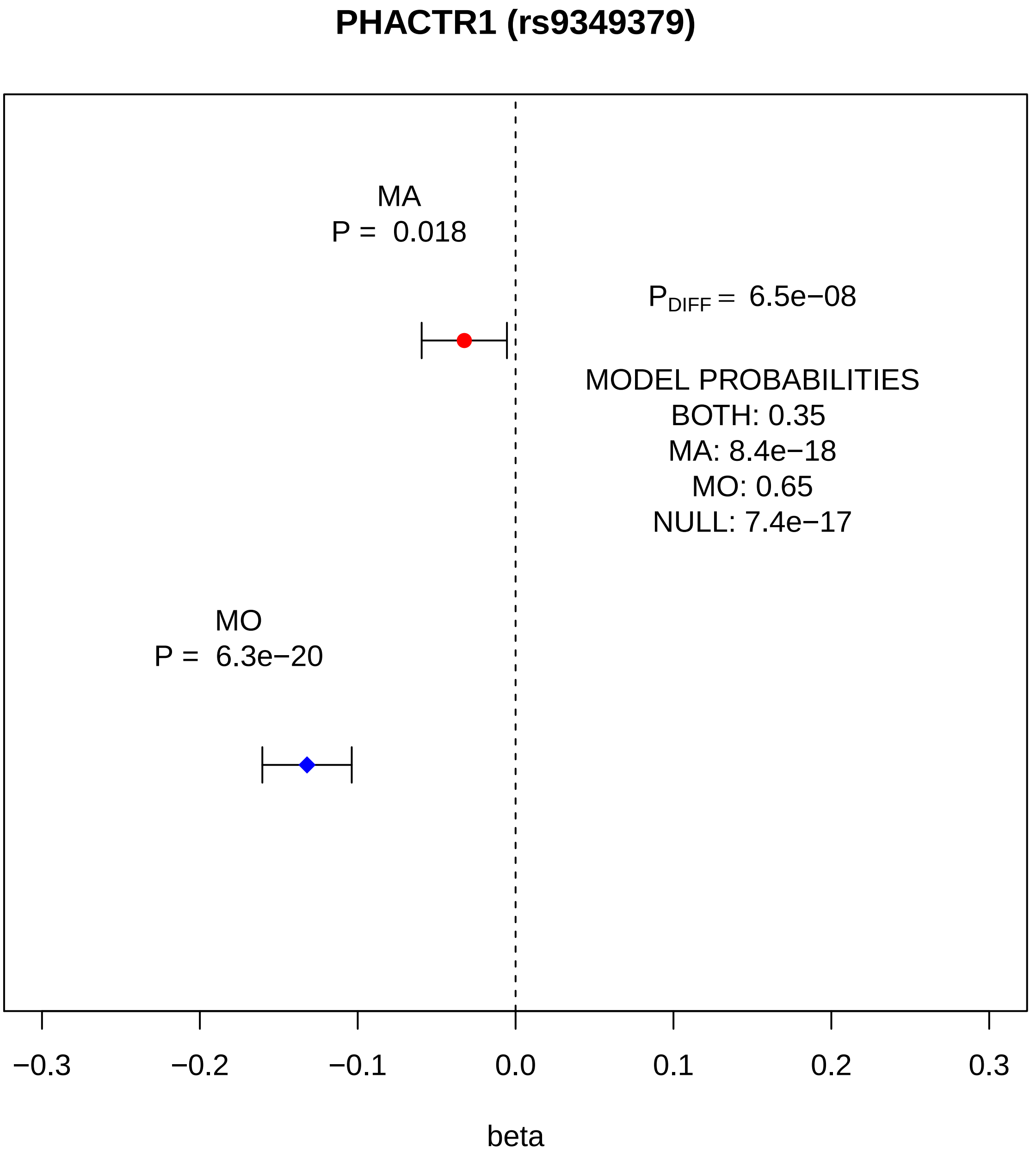
PHACTR1 (rs9349379)
MA
P = 0.018
P_DIFF = 6.5e−08
MODEL PROBABILITIES
BOTH: 0.35
MA: 8.4e−18
MO: 0.65
NULL: 7.4e−17
MO
P = 6.3e−20
−0.3
−0.2
−0.1
0.0
0.1
0.2
0.3
beta

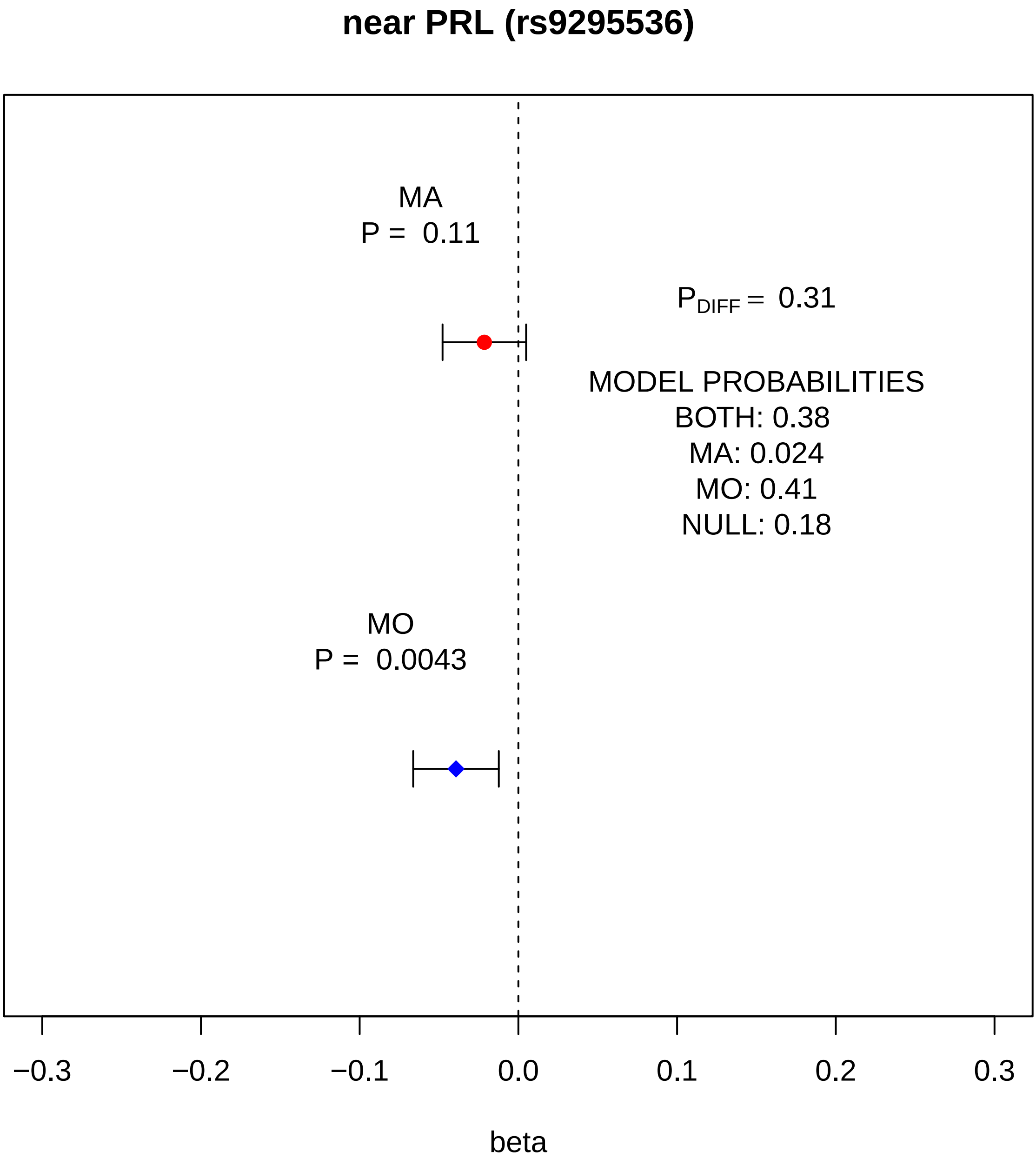
near PRL (rs9295536)
MA
P = 0.11
P DIFF = 0.31
MODEL PROBABILITIES
BOTH: 0.38
MA: 0.024
MO: 0.41
NULL: 0.18
MO
P = 0.0043
−0.3
−0.2
−0.1
0.0
0.1
0.2
0.3
beta

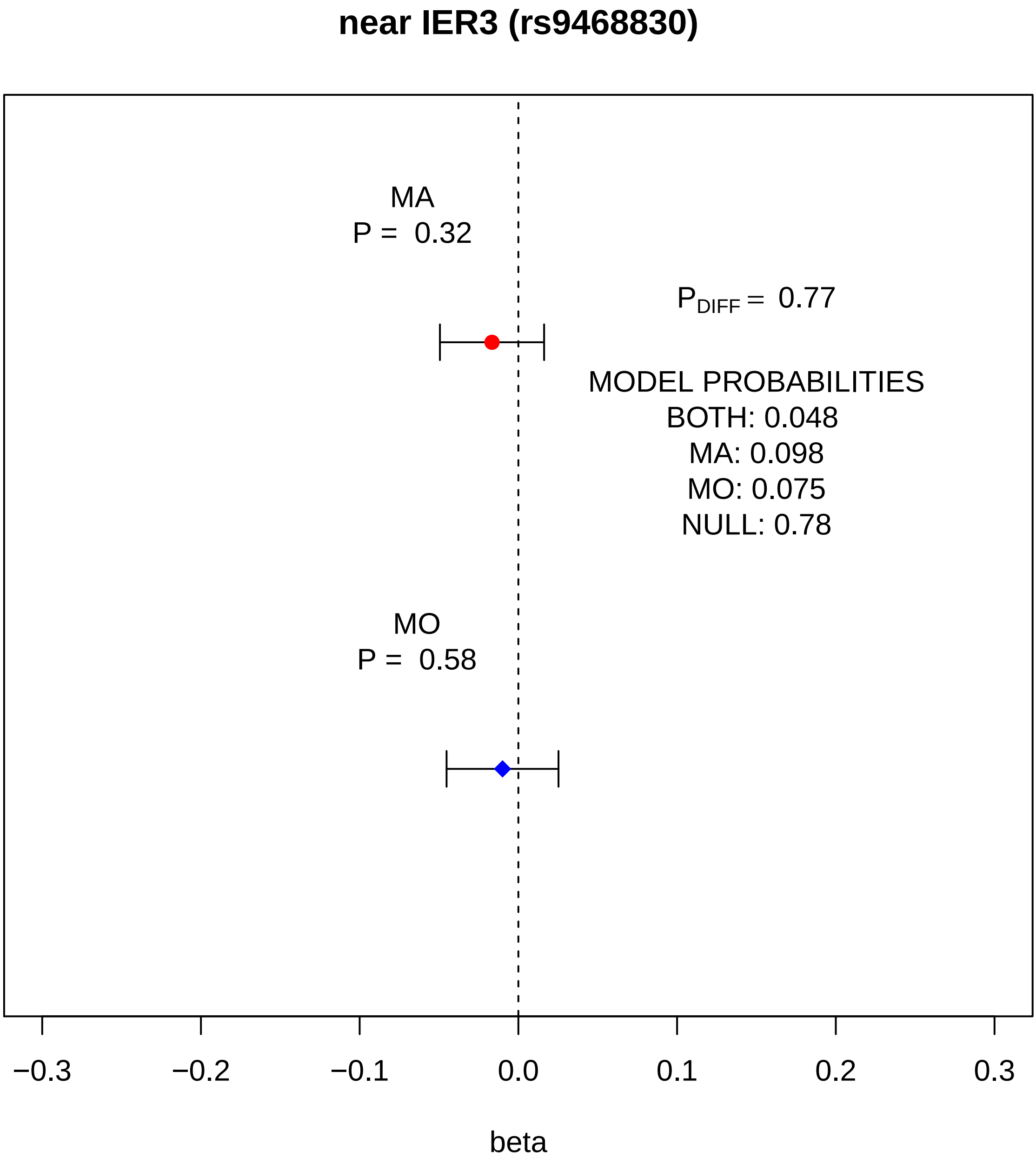
near IER3 (rs9468830)
MA
P = 0.32
$P_{DIFF}$ = 0.77
MODEL PROBABILITIES
BOTH: 0.048
MA: 0.098
MO: 0.075
NULL: 0.78
MO
P = 0.58
−0.3
−0.2
−0.1
0.0
0.1
0.2
0.3
beta

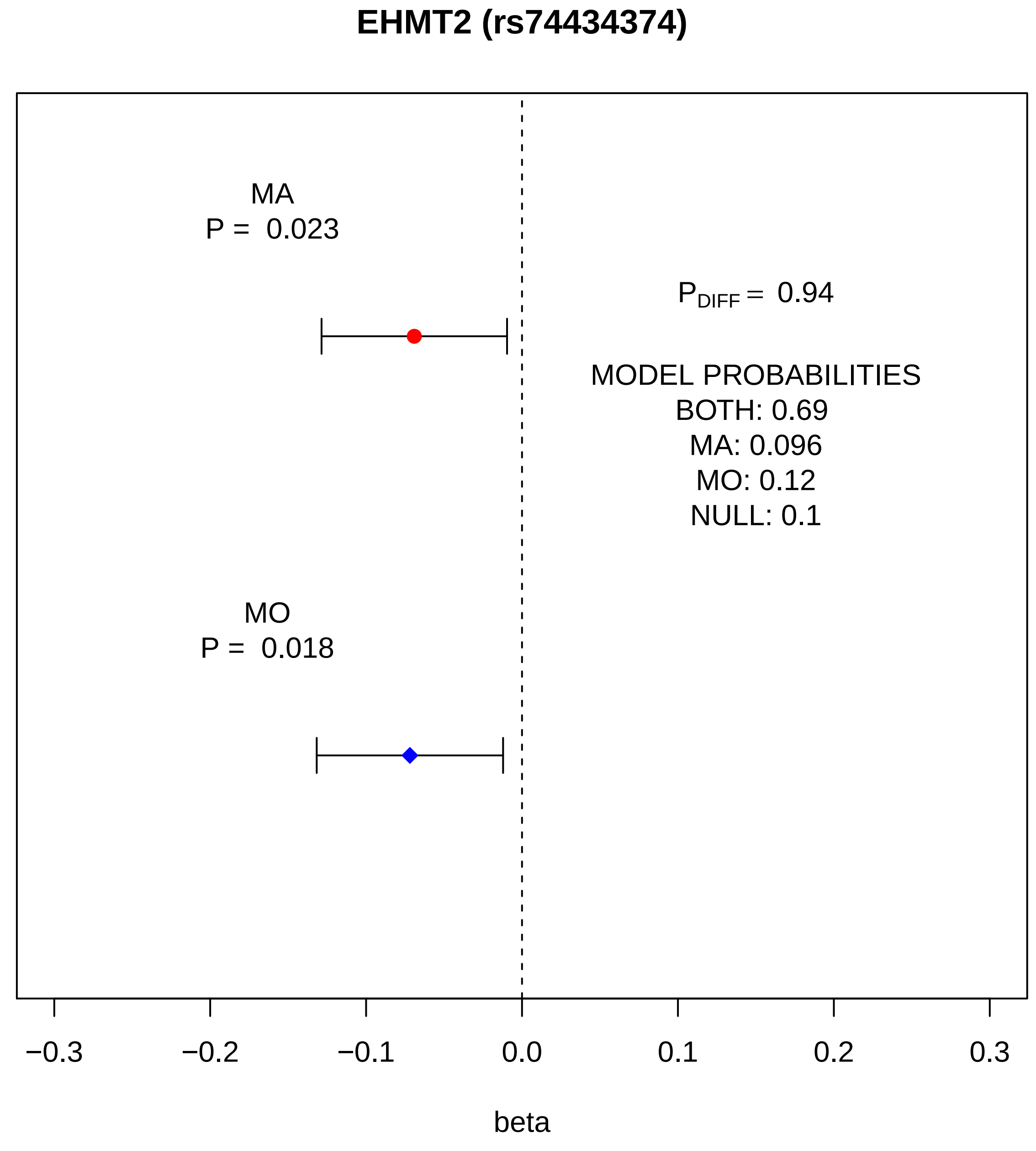
EHMT2 (rs74434374)
MA
P = 0.023
$P_{DIFF}$ = 0.94
MODEL PROBABILITIES
BOTH: 0.69
MA: 0.096
MO: 0.12
NULL: 0.1
MO
P = 0.018
−0.3
−0.2
−0.1
0.0
0.1
0.2
0.3
beta

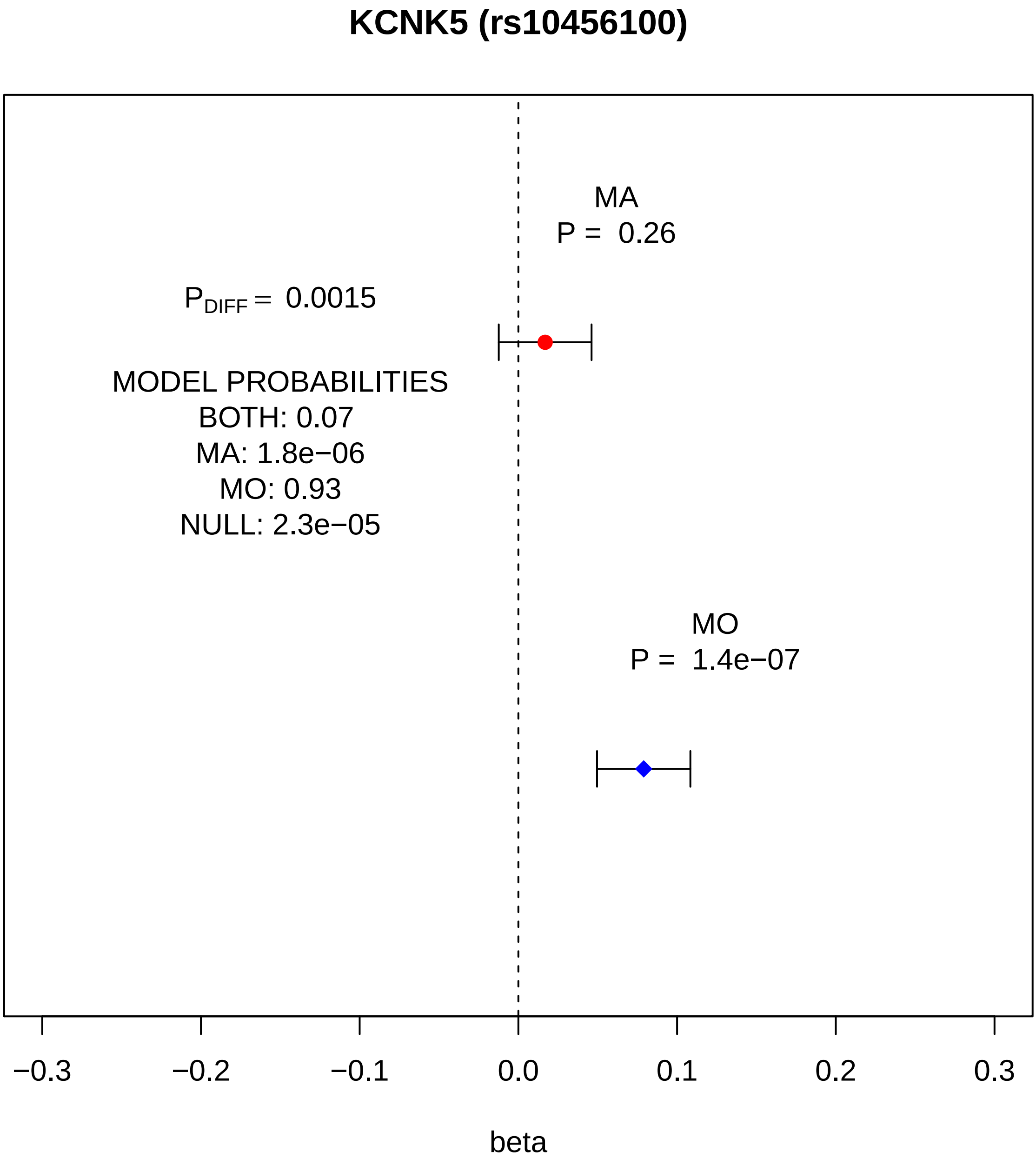
KCNK5 (rs10456100)
MA
P = 0.26
P_DIFF = 0.0015
MODEL PROBABILITIES
BOTH: 0.07
MA: 1.8e−06
MO: 0.93
NULL: 2.3e−05
MO
P = 1.4e−07
−0.3
−0.2
−0.1
0.0
0.1
0.2
0.3
beta

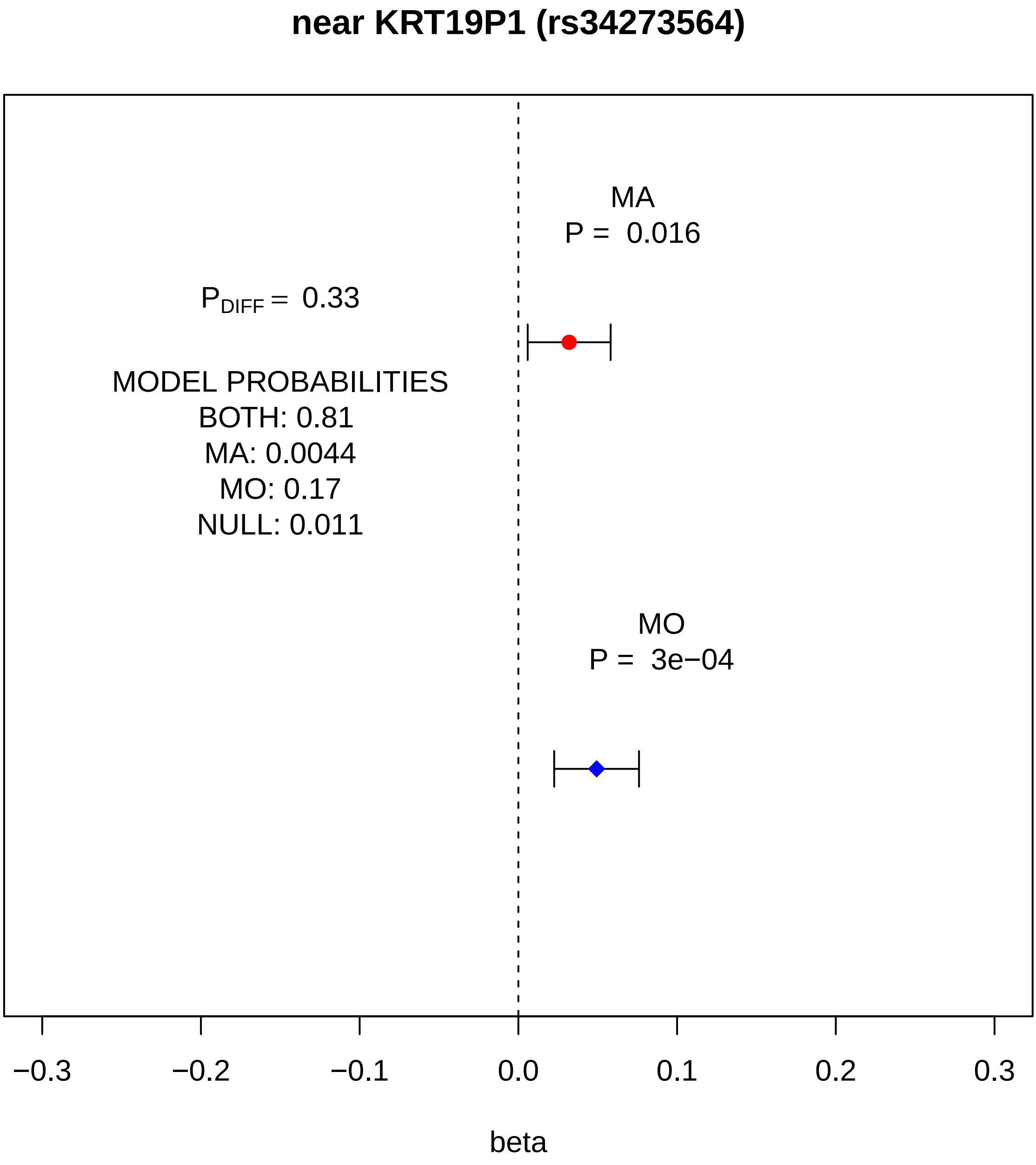
near KRT19P1 (rs34273564)
MA
P = 0.016
$P_{DIFF}$ = 0.33
MODEL PROBABILITIES
BOTH: 0.81
MA: 0.0044
MO: 0.17
NULL: 0.011
MO
P = 3e−04
−0.3
−0.2
−0.1
0.0
0.1
0.2
0.3
beta

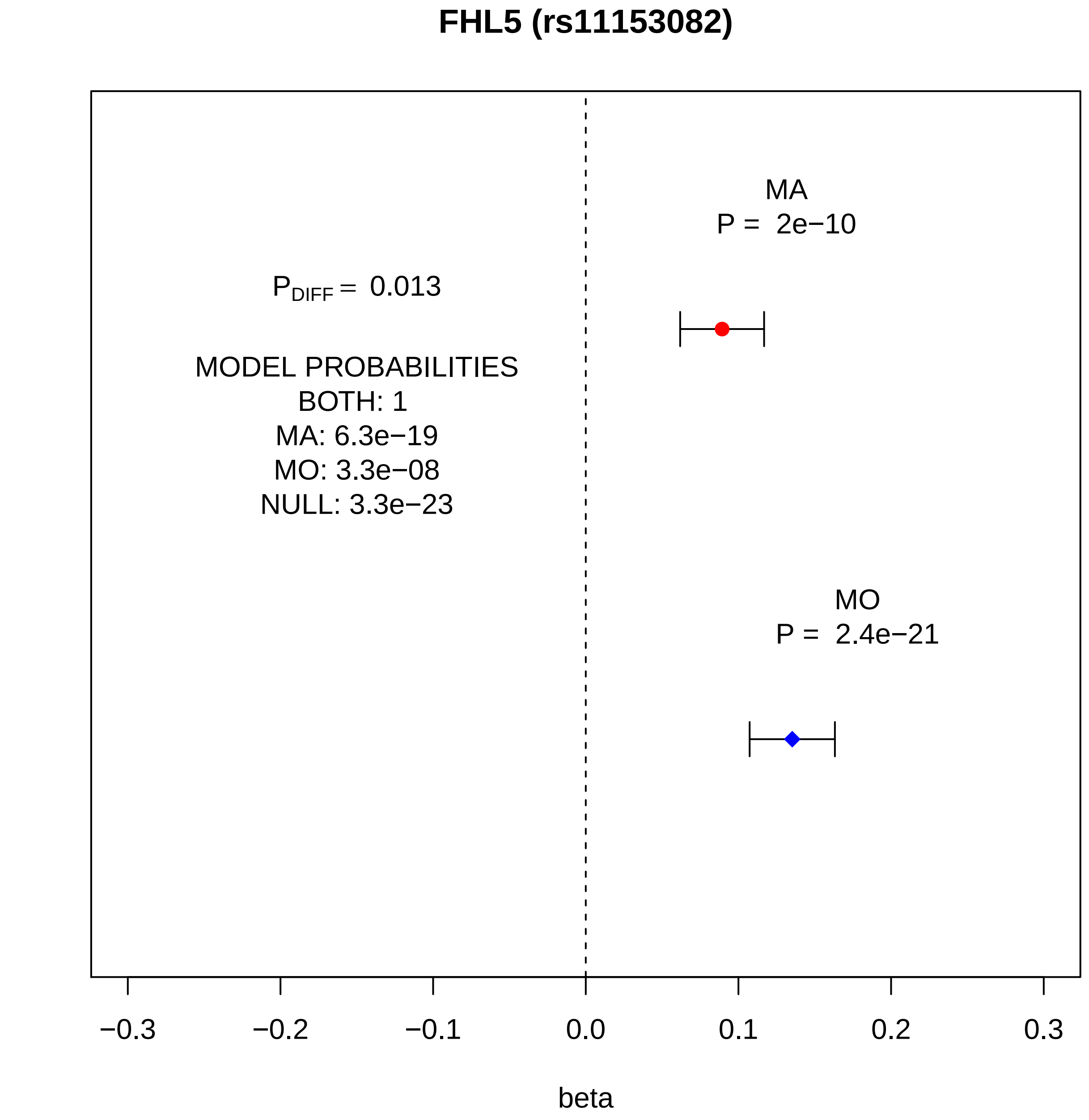
FHL5 (rs11153082)
MA
P = 2e−10
$P_{DIFF}$ = 0.013
MODEL PROBABILITIES
BOTH: 1
MA: 6.3e−19
MO: 3.3e−08
NULL: 3.3e−23
MO
P = 2.4e−21
−0.3
−0.2
−0.1
0.0
0.1
0.2
0.3
beta

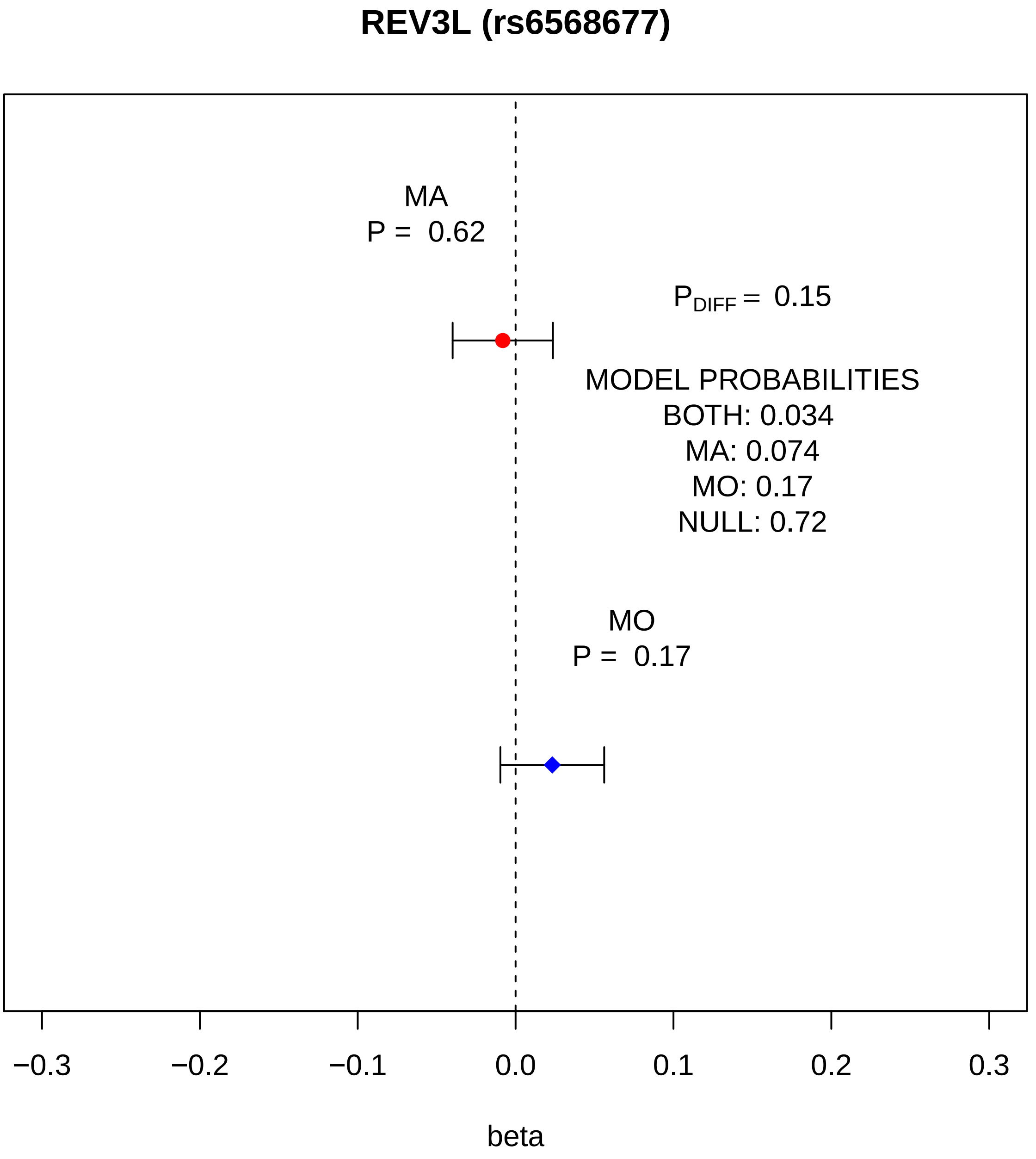
REV3L (rs6568677)
MA
P = 0.62
$P_{DIFF}$ = 0.15
MODEL PROBABILITIES
BOTH: 0.034
MA: 0.074
MO: 0.17
NULL: 0.72
MO
P = 0.17
−0.3
−0.2
−0.1
0.0
0.1
0.2
0.3
beta

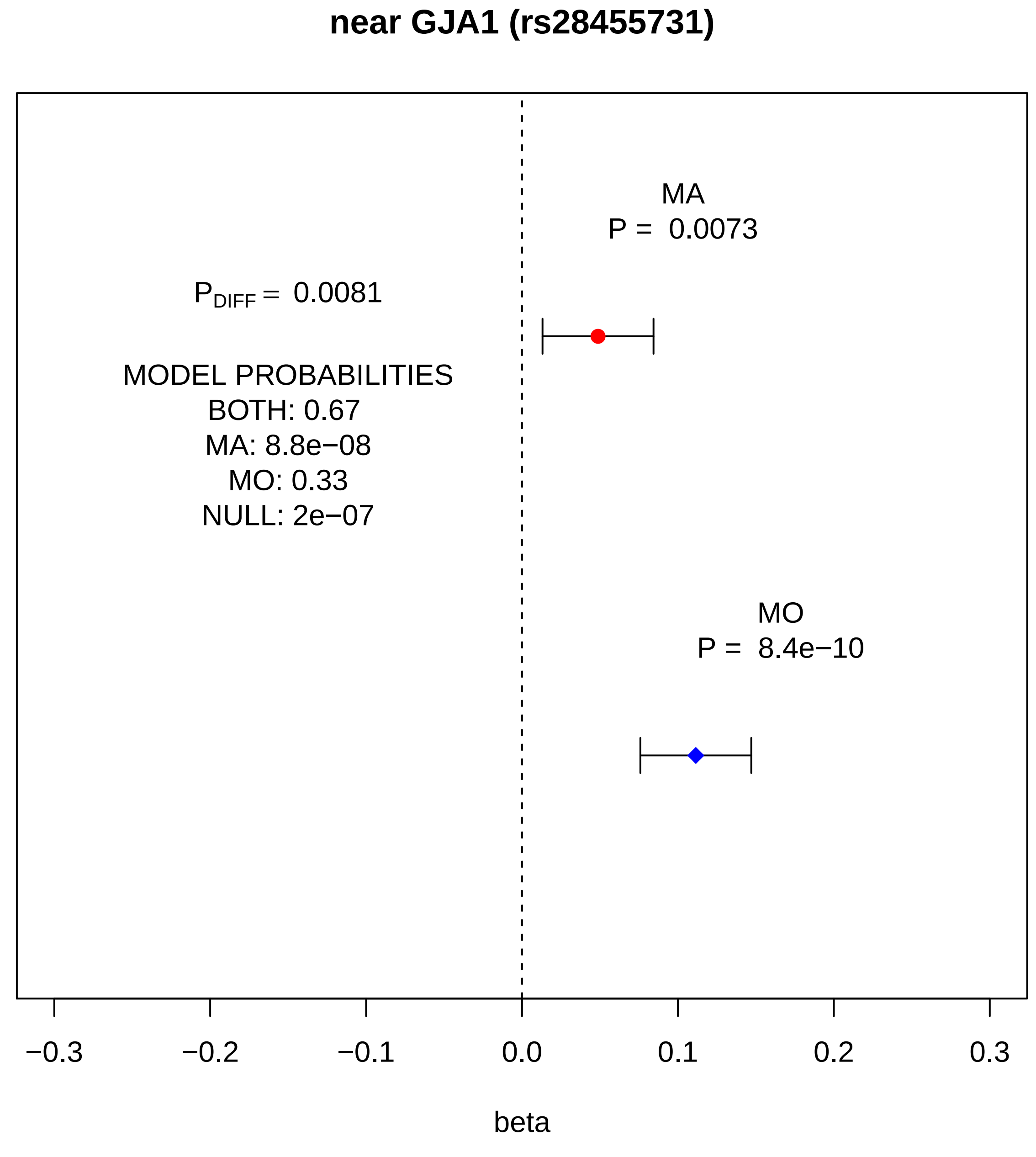
near GJA1 (rs28455731)
MA
P = 0.0073
P_DIFF = 0.0081
MODEL PROBABILITIES
BOTH: 0.67
MA: 8.8e−08
MO: 0.33
NULL: 2e−07
MO
P = 8.4e−10
−0.3
−0.2
−0.1
0.0
0.1
0.2
0.3
beta

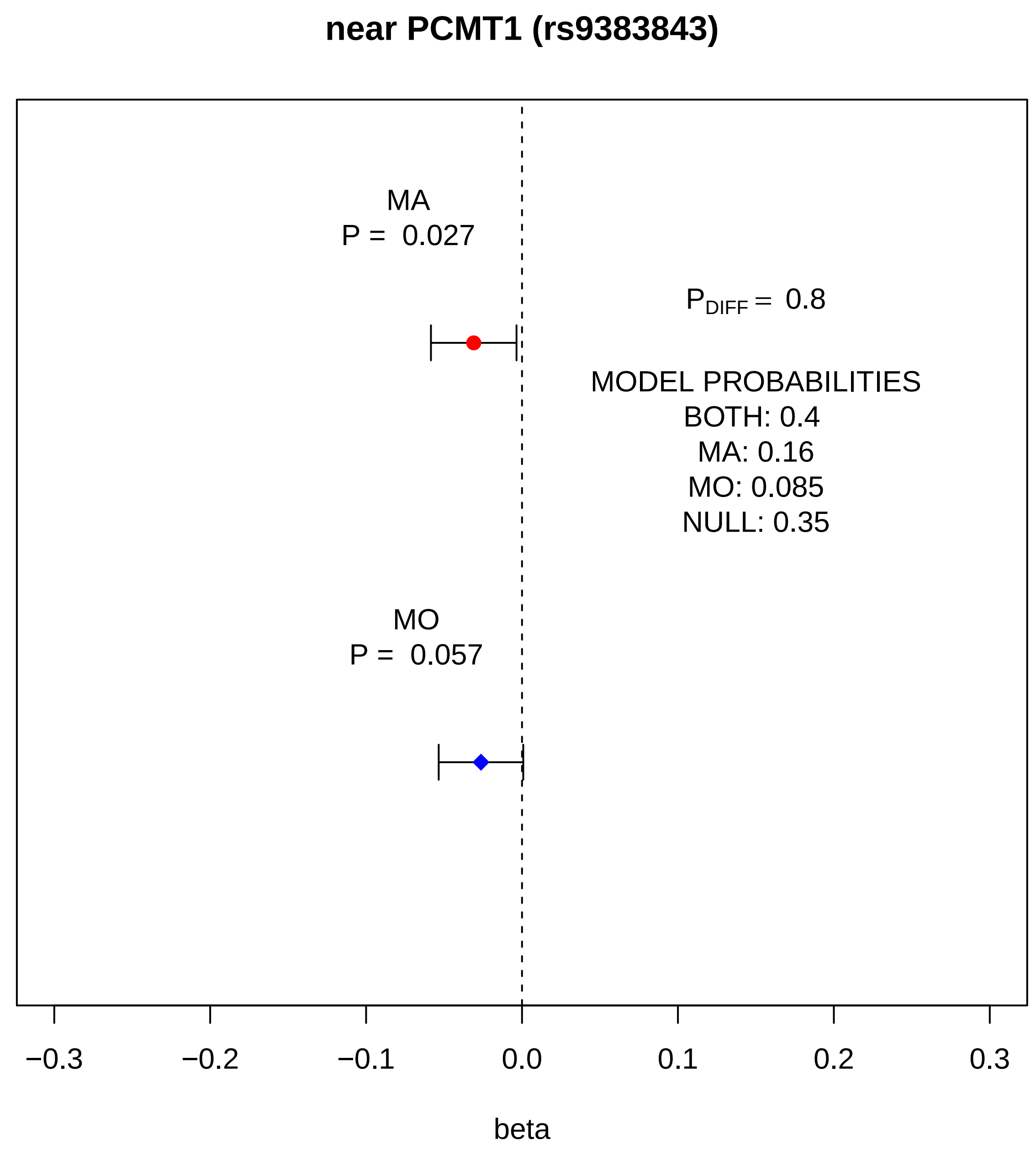
near PCMT1 (rs9383843)
MA
P = 0.027
MO
P = 0.057
P_DIFF = 0.8
MODEL PROBABILITIES
BOTH: 0.4
MA: 0.16
MO: 0.085
NULL: 0.35
−0.3
−0.2
−0.1
0.0
0.1
0.2
0.3
beta

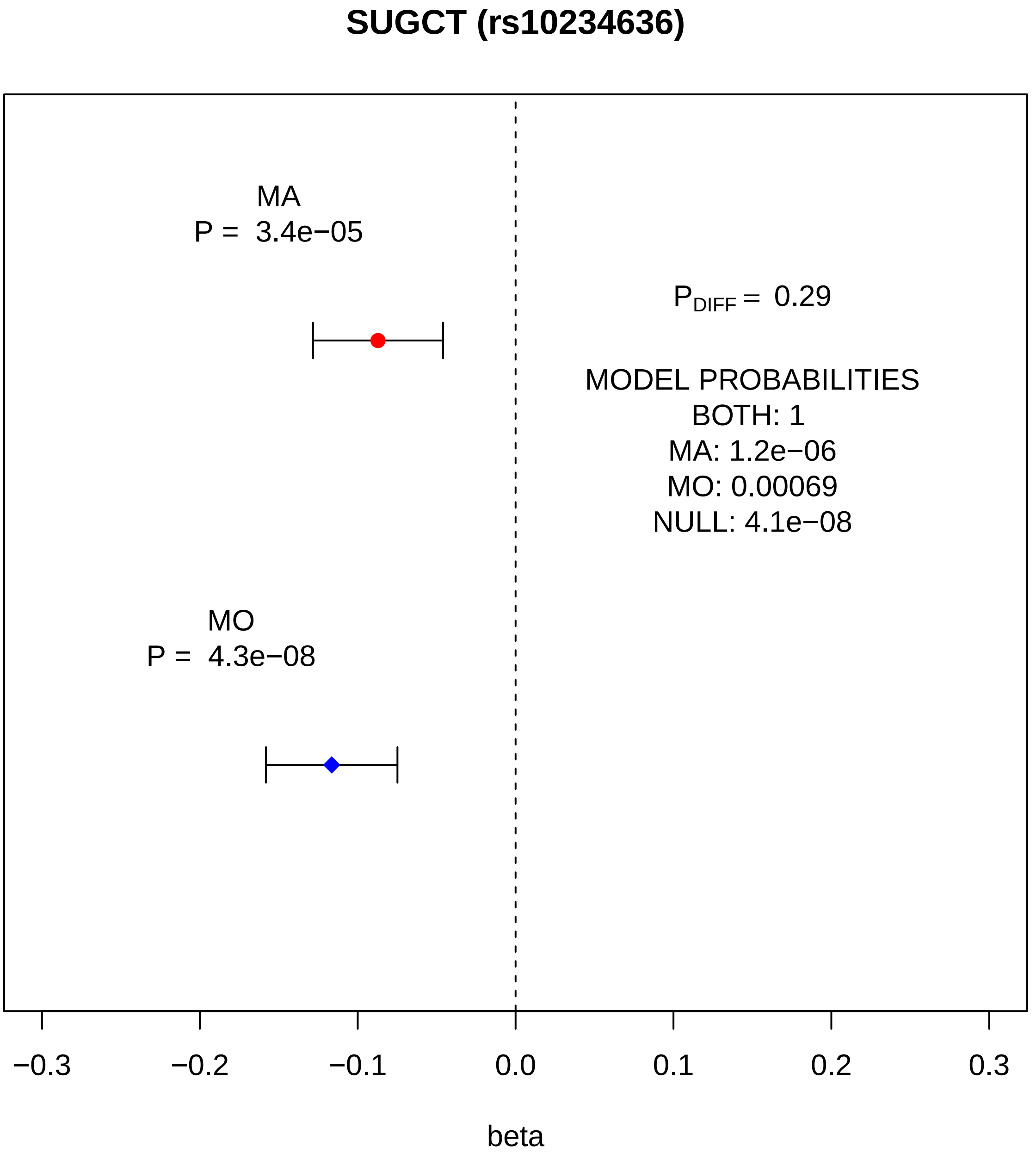
SUGCT (rs10234636)
MA
P = 3.4e−05
$P_{DIFF}$ = 0.29
MODEL PROBABILITIES
BOTH: 1
MA: 1.2e−06
MO: 0.00069
NULL: 4.1e−08
MO
P = 4.3e−08
−0.3
−0.2
−0.1
0.0
0.1
0.2
0.3
beta

### MLXIPL (rs13235543)

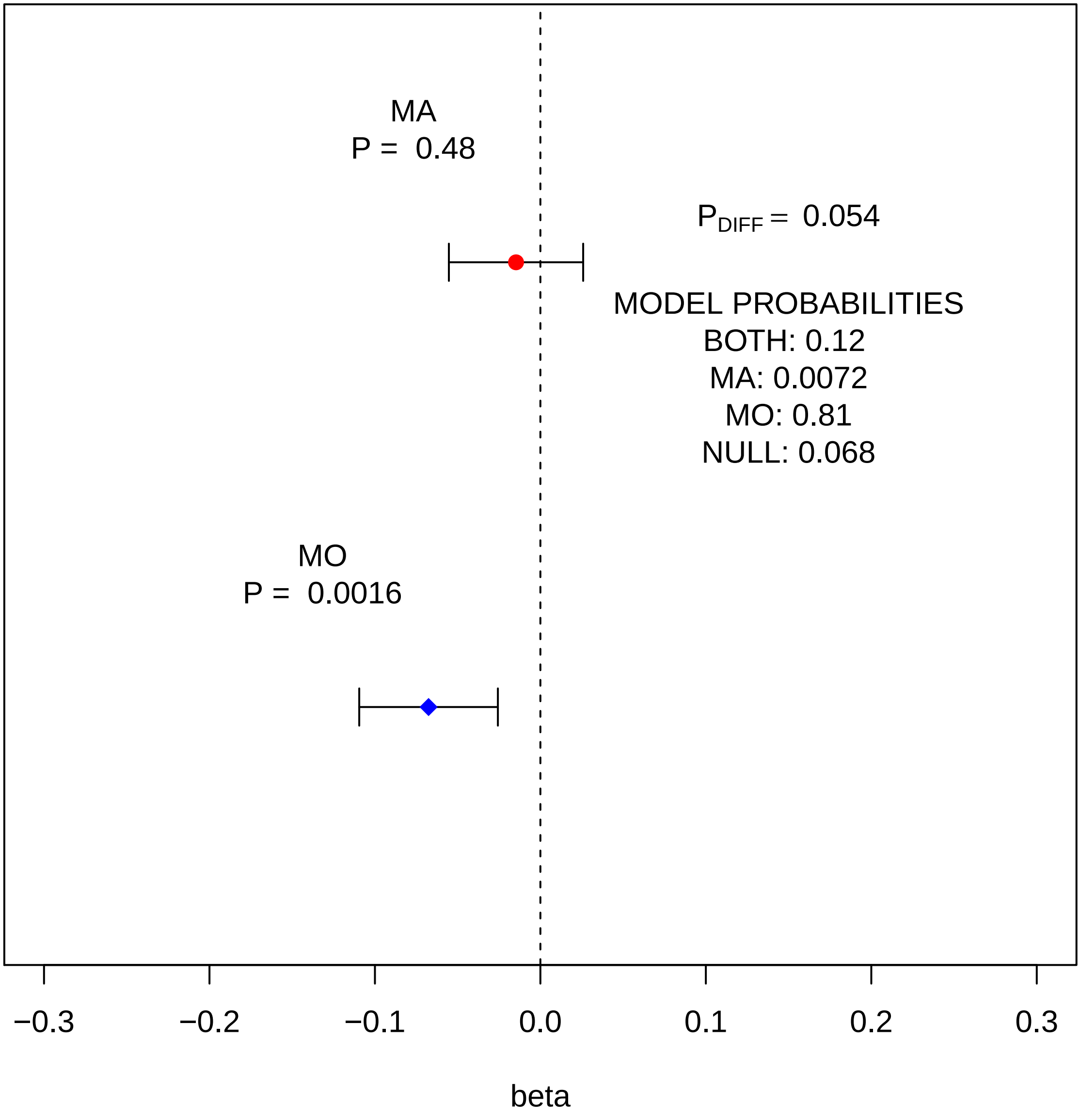

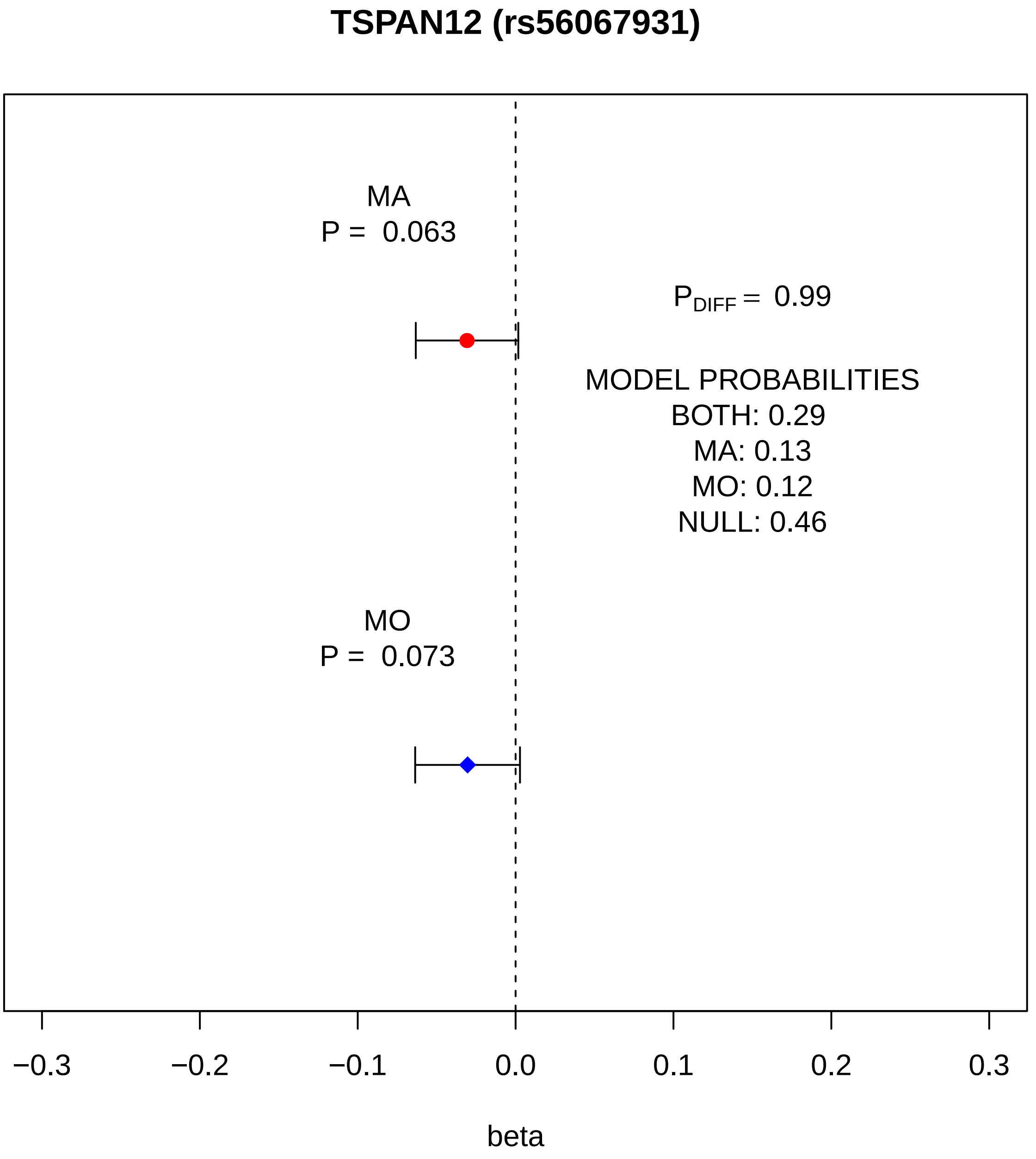
TSPAN12 (rs56067931)
MA
P = 0.063
$P_{DIFF}$ = 0.99
MODEL PROBABILITIES
BOTH: 0.29
MA: 0.13
MO: 0.12
NULL: 0.46
MO
P = 0.073
−0.3
−0.2
−0.1
0.0
0.1
0.2
0.3
beta

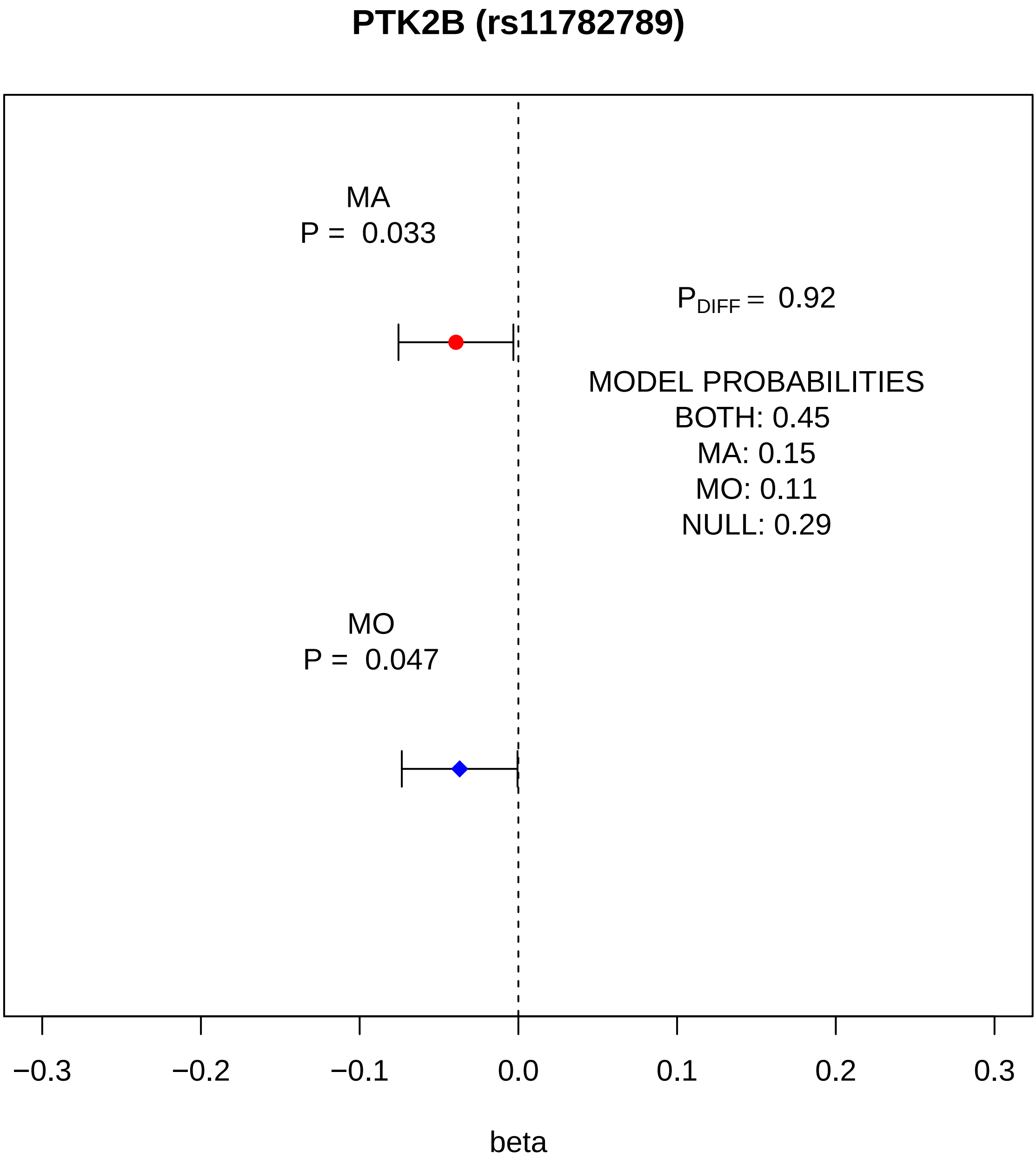
PTK2B (rs11782789)
MA
P = 0.033
$P_{DIFF}$ = 0.92
MODEL PROBABILITIES
BOTH: 0.45
MA: 0.15
MO: 0.11
NULL: 0.29
MO
P = 0.047
−0.3
−0.2
−0.1
0.0
0.1
0.2
0.3
beta

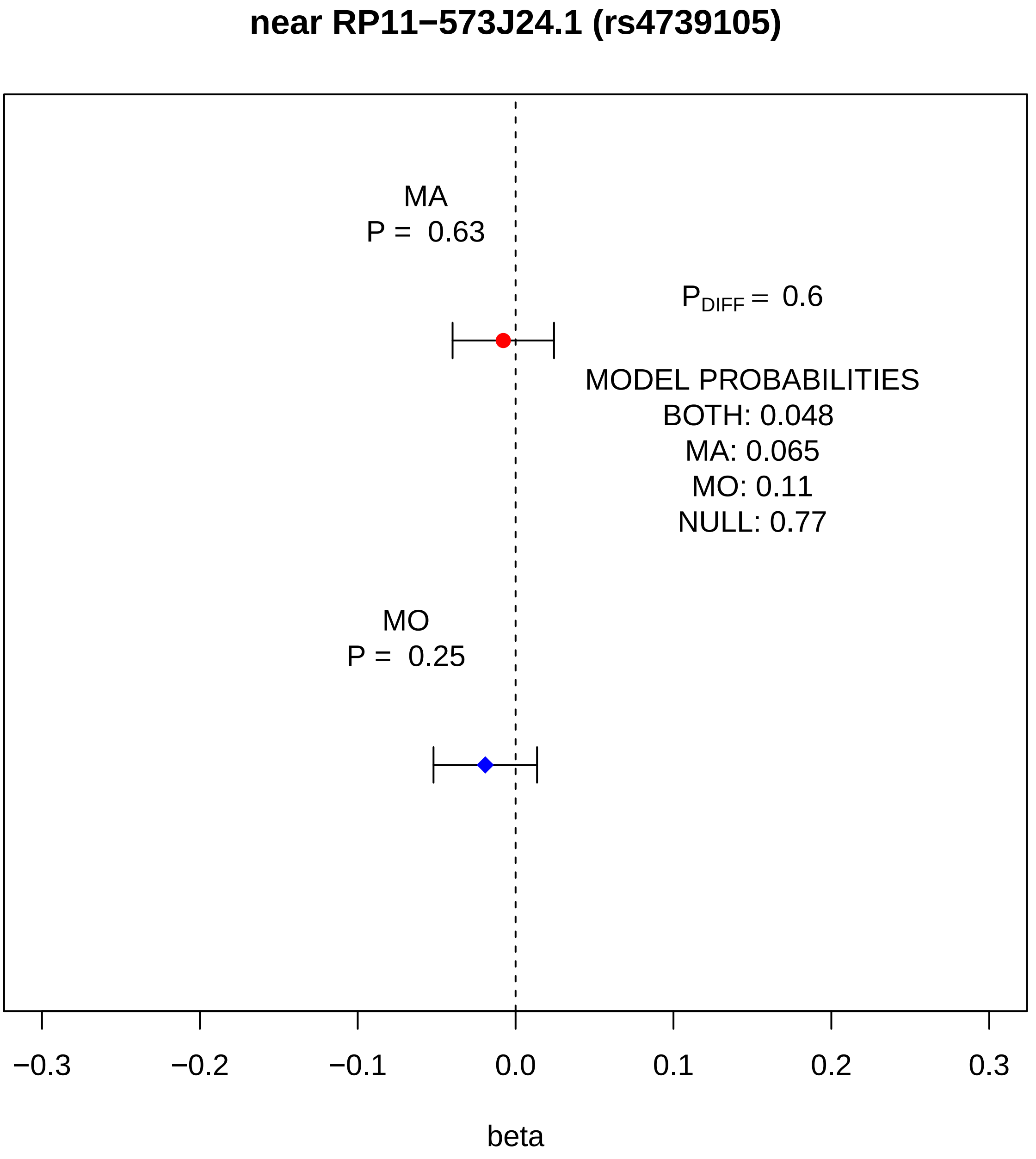
near RP11−573J24.1 (rs4739105)
MA
P = 0.63
$P_{DIFF}$ = 0.6
MODEL PROBABILITIES
BOTH: 0.048
MA: 0.065
MO: 0.11
NULL: 0.77
MO
P = 0.25
−0.3
−0.2
−0.1
0.0
0.1
0.2
0.3
beta

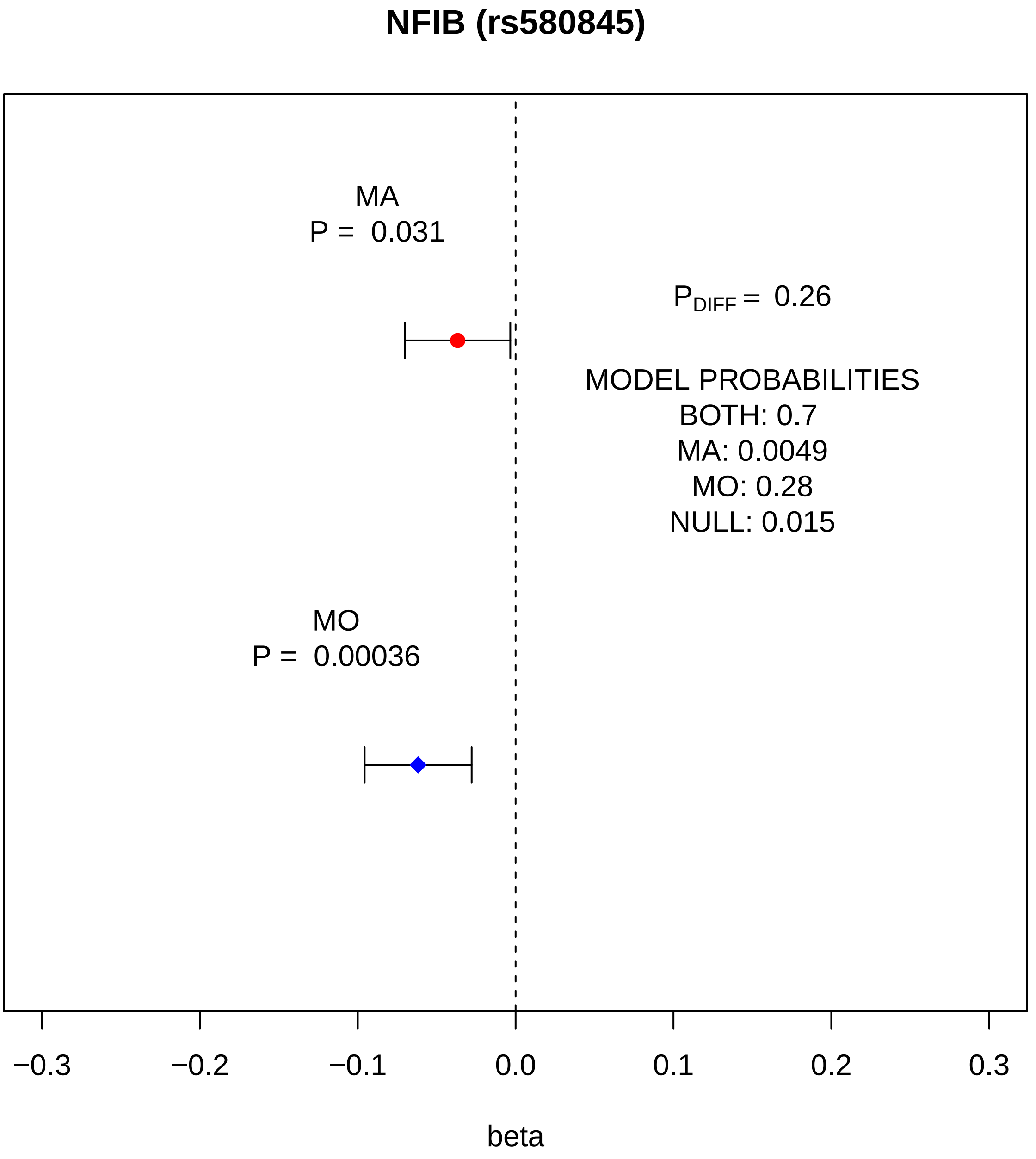
NFIB (rs580845)
MA
P = 0.031
P_DIFF = 0.26
MODEL PROBABILITIES
BOTH: 0.7
MA: 0.0049
MO: 0.28
NULL: 0.015
MO
P = 0.00036
−0.3
−0.2
−0.1
0.0
0.1
0.2
0.3
beta

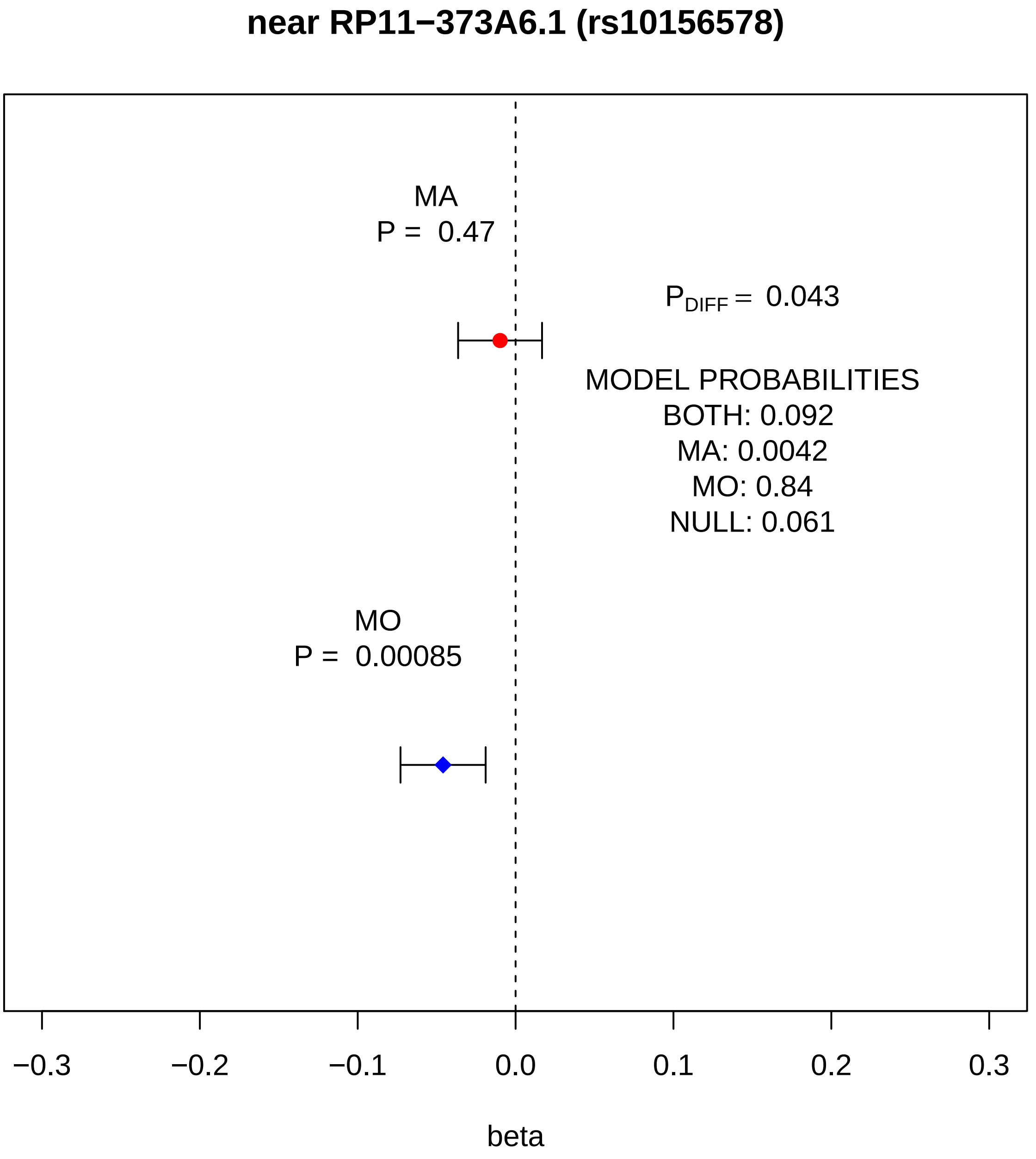
near RP11−373A6.1 (rs10156578)
MA
P = 0.47
P_DIFF = 0.043
MODEL PROBABILITIES
BOTH: 0.092
MA: 0.0042
MO: 0.84
NULL: 0.061
MO
P = 0.00085
−0.3
−0.2
−0.1
0.0
0.1
0.2
0.3
beta

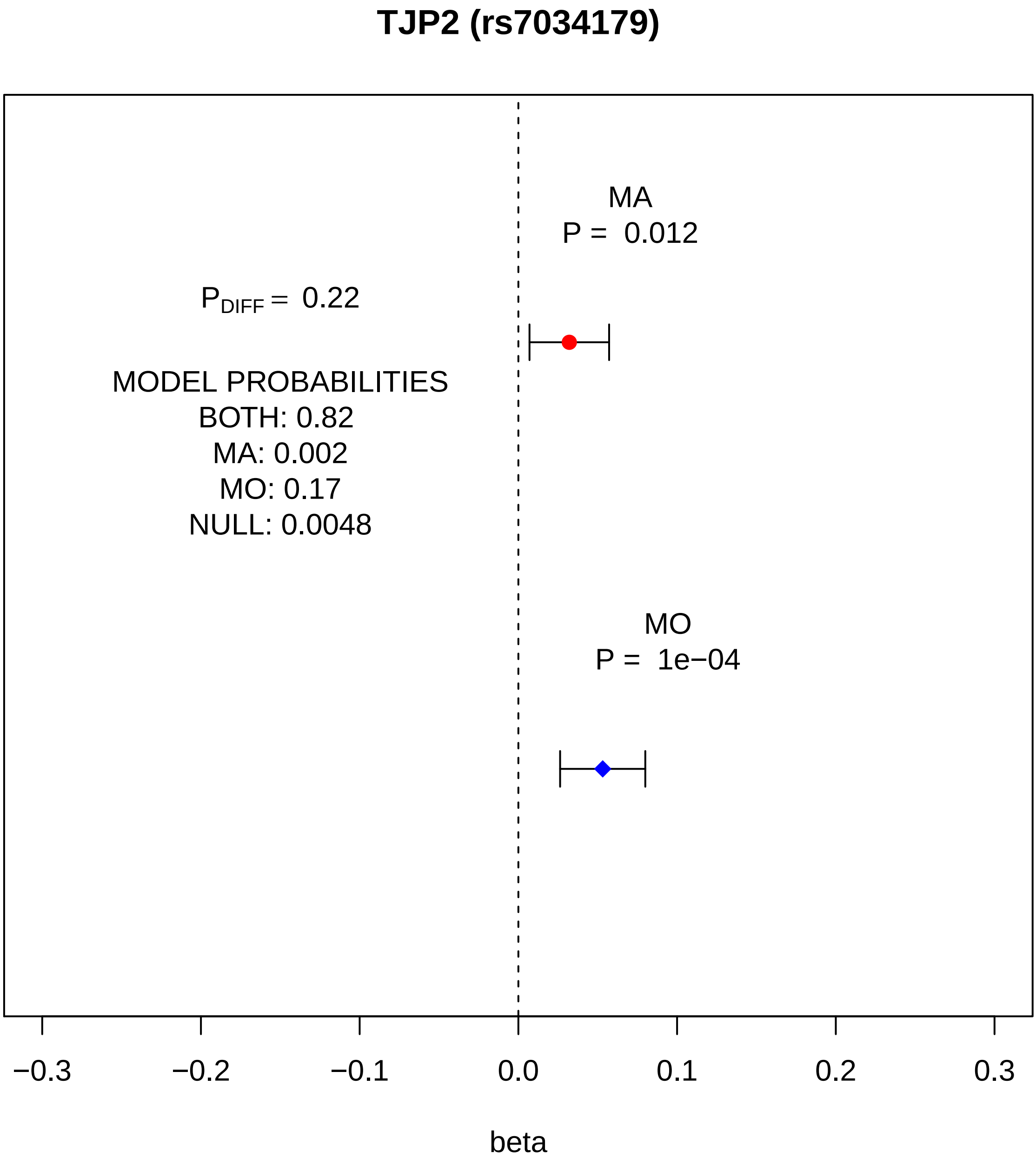
TJP2 (rs7034179)
MA
P = 0.012
$P_{DIFF}$ = 0.22
MODEL PROBABILITIES
BOTH: 0.82
MA: 0.002
MO: 0.17
NULL: 0.0048
MO
P = 1e−04
−0.3
−0.2
−0.1
0.0
0.1
0.2
0.3
beta

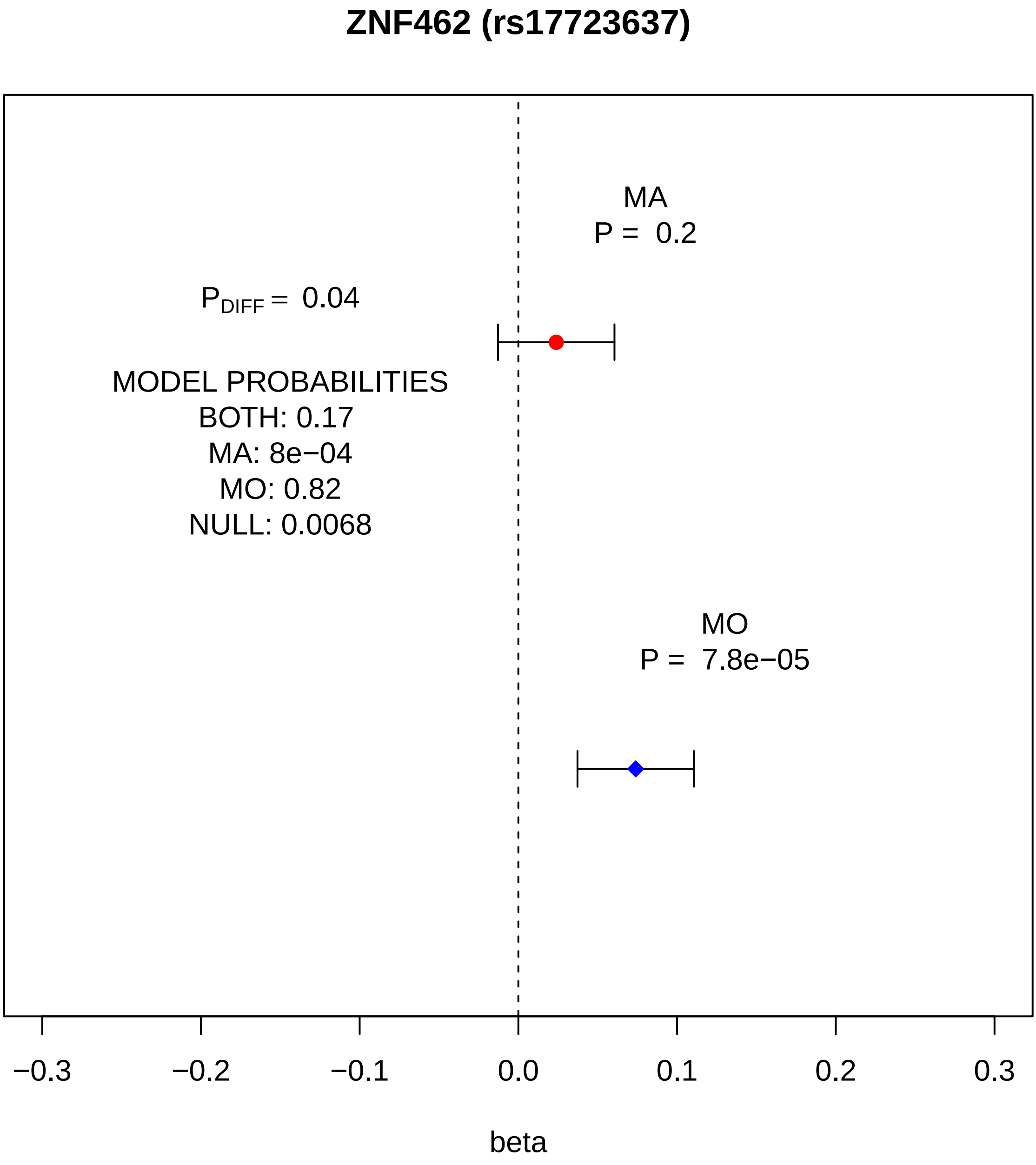

ZNF462 (rs17723637)
MA
P = 0.2
$P_{DIFF}$ = 0.04
MODEL PROBABILITIES
BOTH: 0.17
MA: 8e−04
MO: 0.82
NULL: 0.0068
MO
P = 7.8e−05
−0.3
−0.2
−0.1
0.0
0.1
0.2
0.3
beta

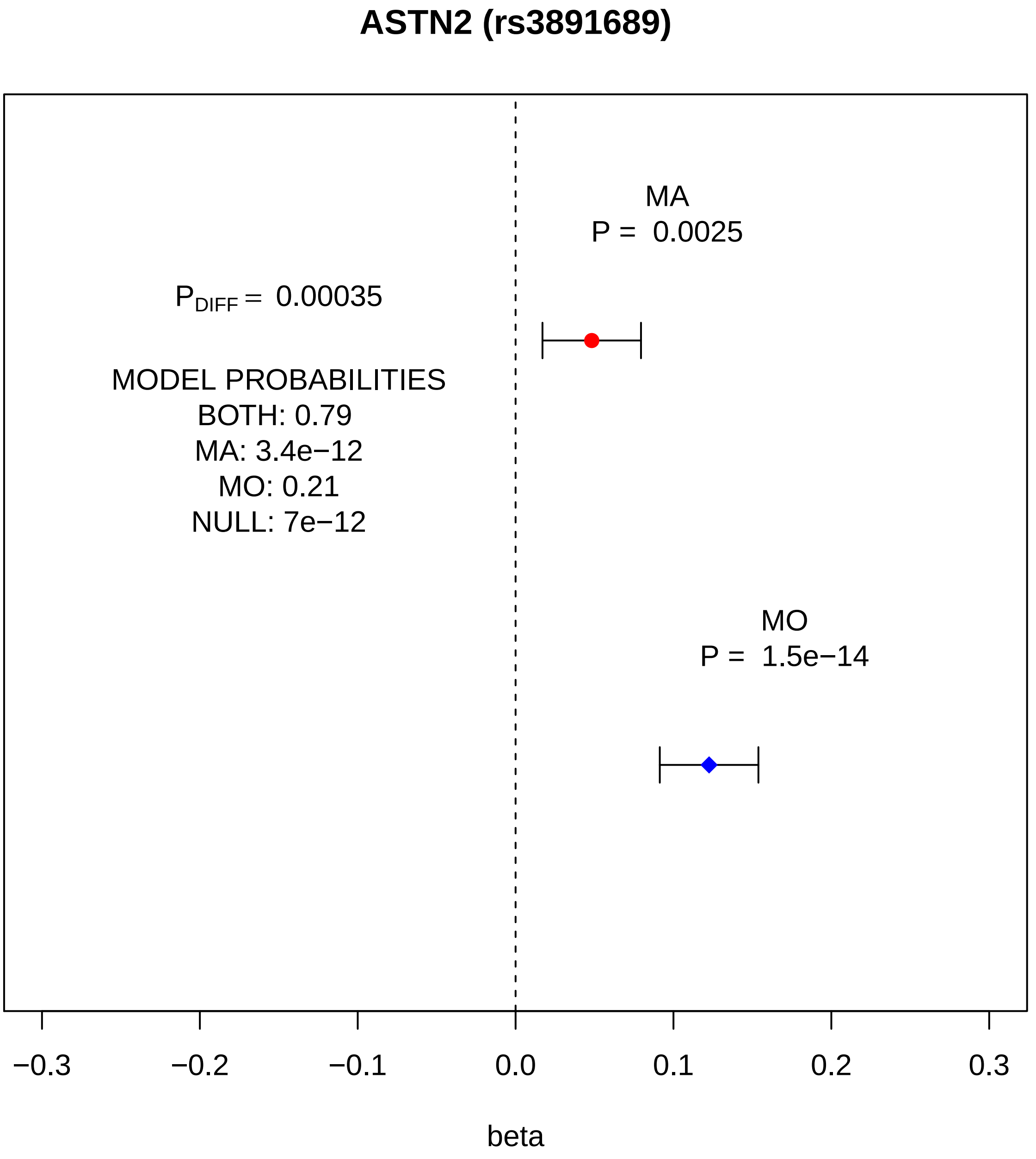
ASTN2 (rs3891689)
MA
P = 0.0025
$P_{DIFF}$ = 0.00035
MODEL PROBABILITIES
BOTH: 0.79
MA: 3.4e−12
MO: 0.21
NULL: 7e−12
MO
P = 1.5e−14
−0.3
−0.2
−0.1
0.0
0.1
0.2
0.3
beta

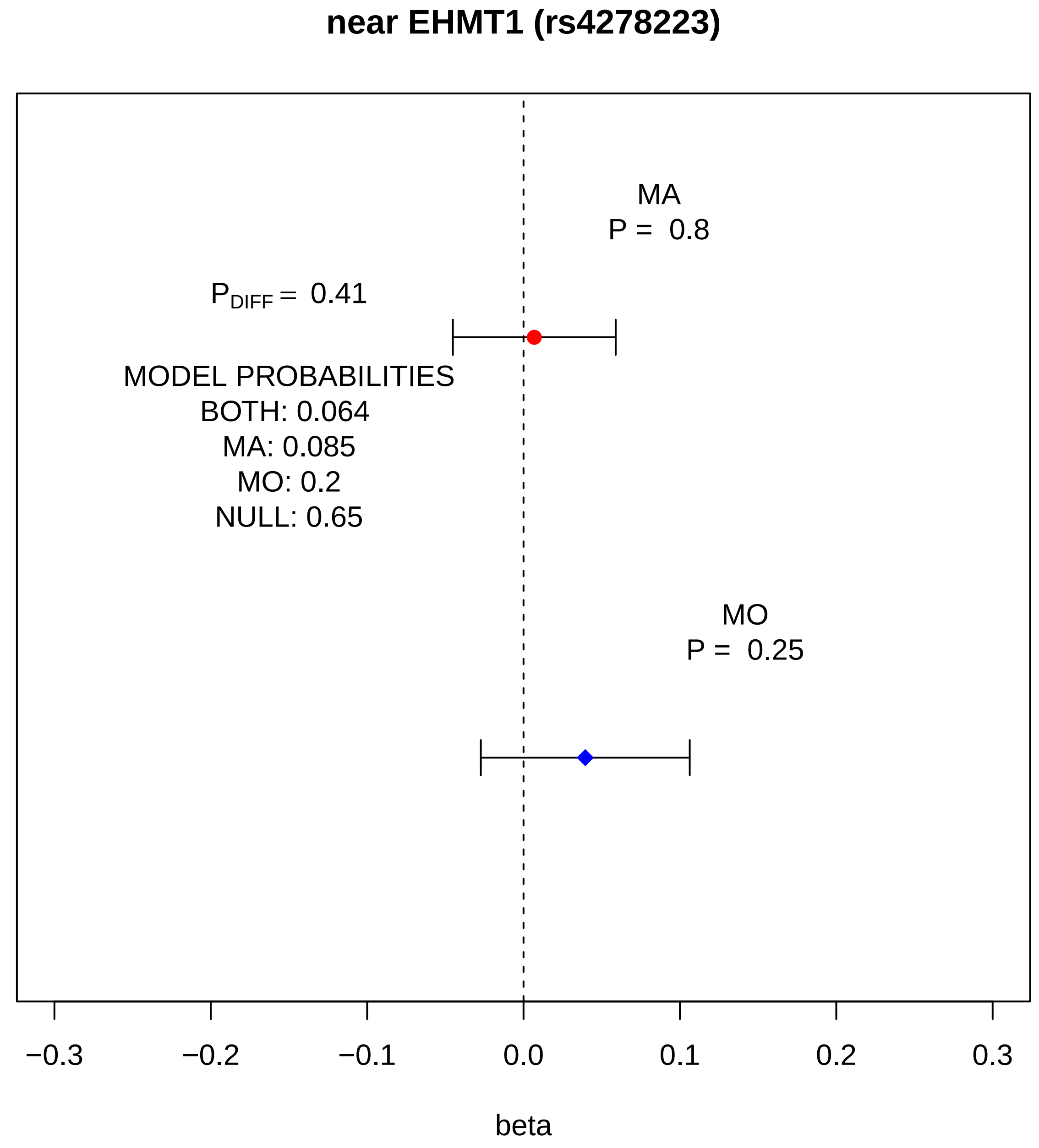
near EHMT1 (rs4278223)
MA
P = 0.8
$P_{DIFF}$ = 0.41
MODEL PROBABILITIES
BOTH: 0.064
MA: 0.085
MO: 0.2
NULL: 0.65
MO
P = 0.25
−0.3
−0.2
−0.1
0.0
0.1
0.2
0.3
beta

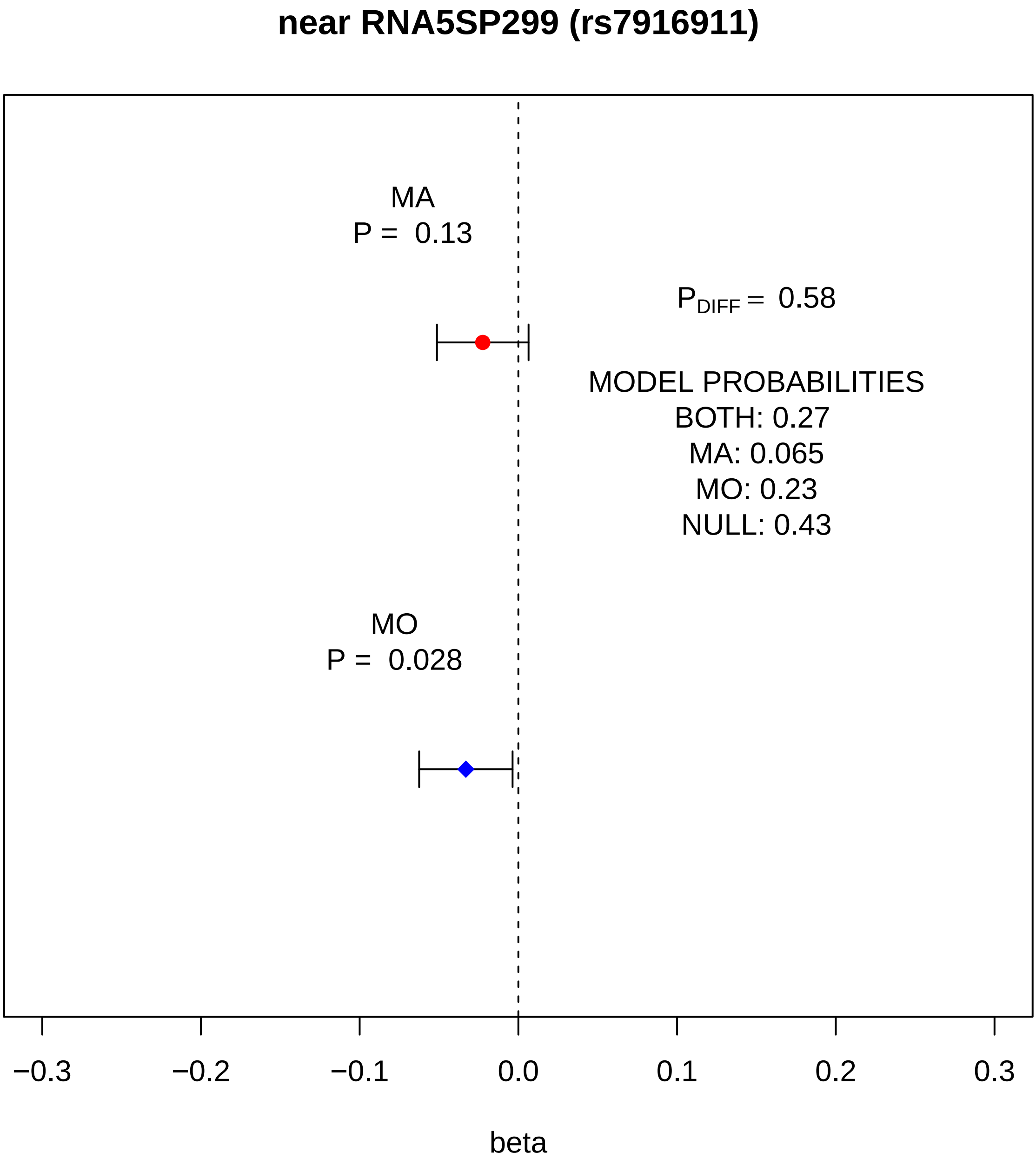
near RNA5SP299 (rs7916911)
MA
P = 0.13
$P_{DIFF}$ = 0.58
MODEL PROBABILITIES
BOTH: 0.27
MA: 0.065
MO: 0.23
NULL: 0.43
MO
P = 0.028
−0.3
−0.2
−0.1
0.0
0.1
0.2
0.3
beta

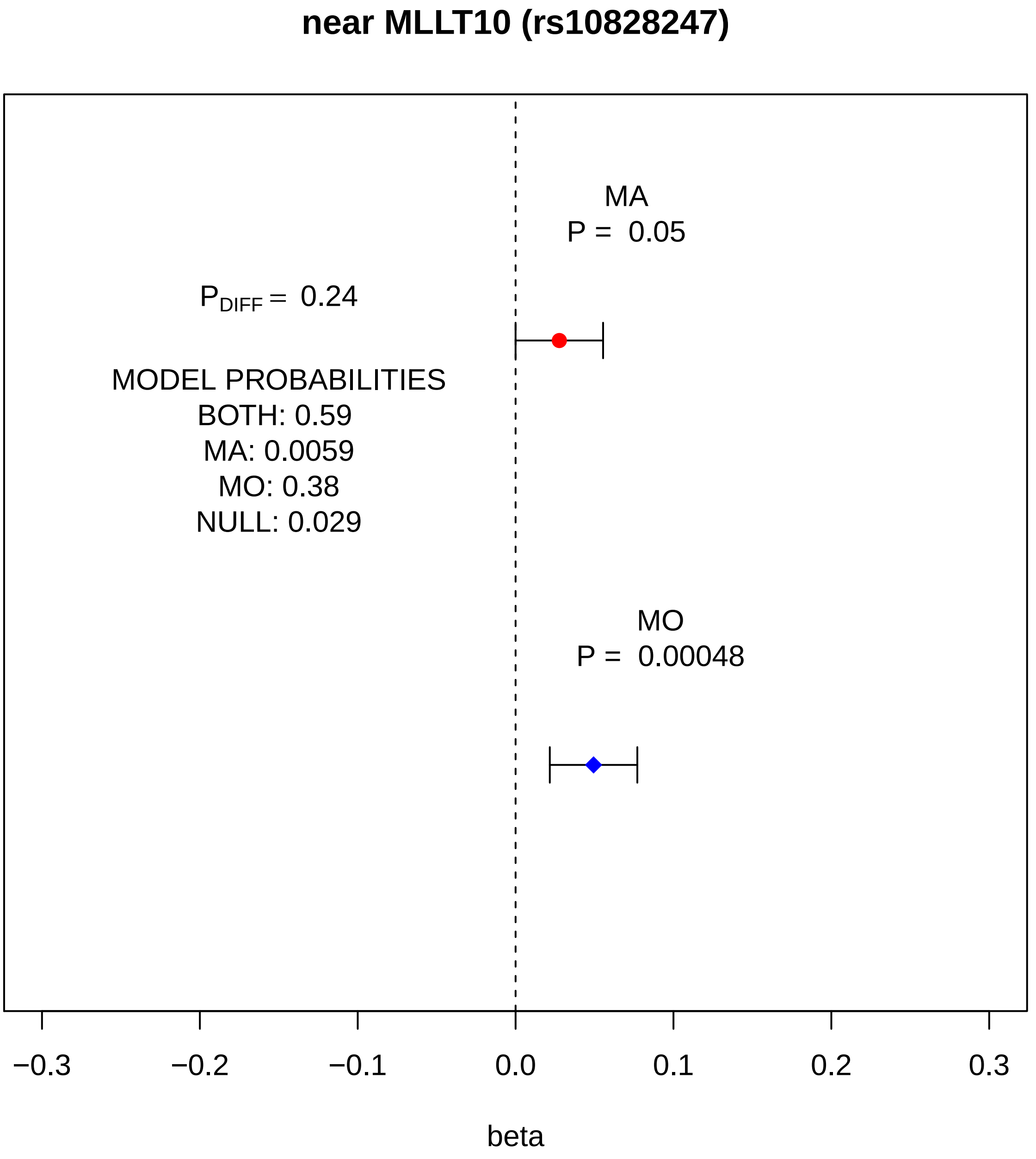
near MLLT10 (rs10828247)
MA
P = 0.05
P$_{DIFF}$ = 0.24
MODEL PROBABILITIES
BOTH: 0.59
MA: 0.0059
MO: 0.38
NULL: 0.029
MO
P = 0.00048
−0.3
−0.2
−0.1
0.0
0.1
0.2
0.3
beta

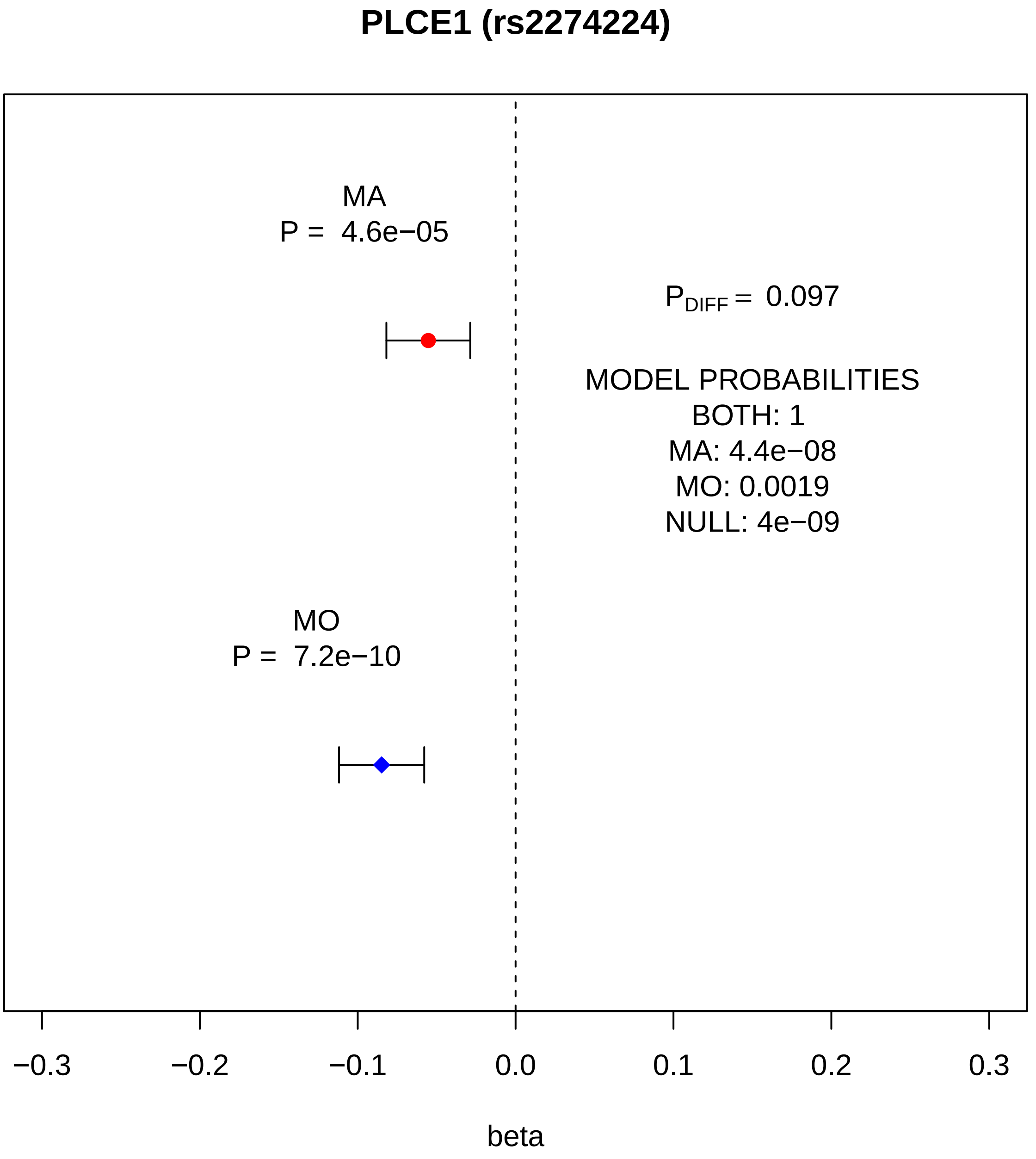
PLCE1 (rs2274224)
MA
P = 4.6e−05
P_DIFF = 0.097
MODEL PROBABILITIES
BOTH: 1
MA: 4.4e−08
MO: 0.0019
NULL: 4e−09
MO
P = 7.2e−10
−0.3
−0.2
−0.1
0.0
0.1
0.2
0.3
beta

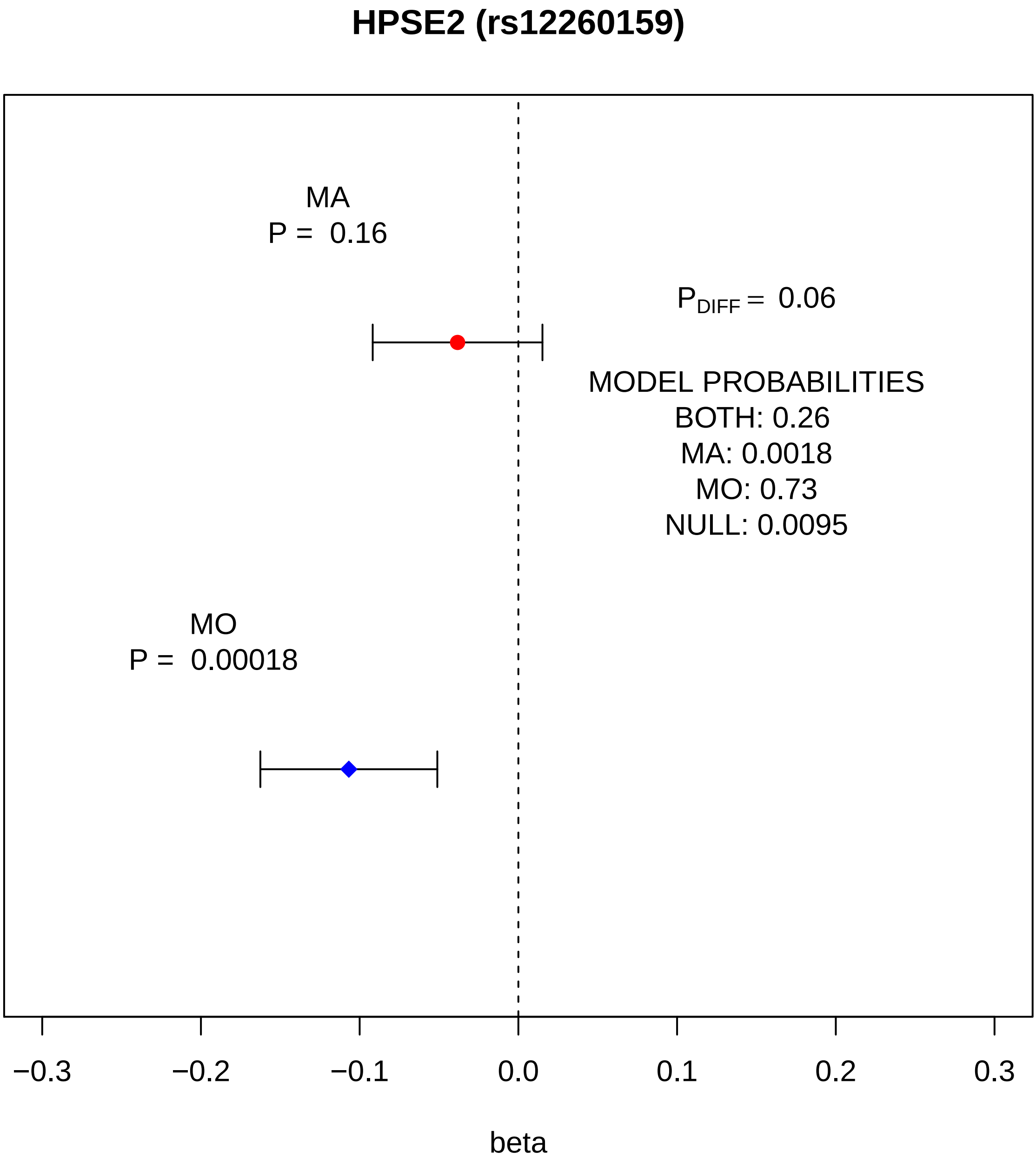

HPSE2 (rs12260159)
MA
P = 0.16
P$_{DIFF}$ = 0.06
MODEL PROBABILITIES
BOTH: 0.26
MA: 0.0018
MO: 0.73
NULL: 0.0095
MO
P = 0.00018
−0.3
−0.2
−0.1
0.0
0.1
0.2
0.3
beta

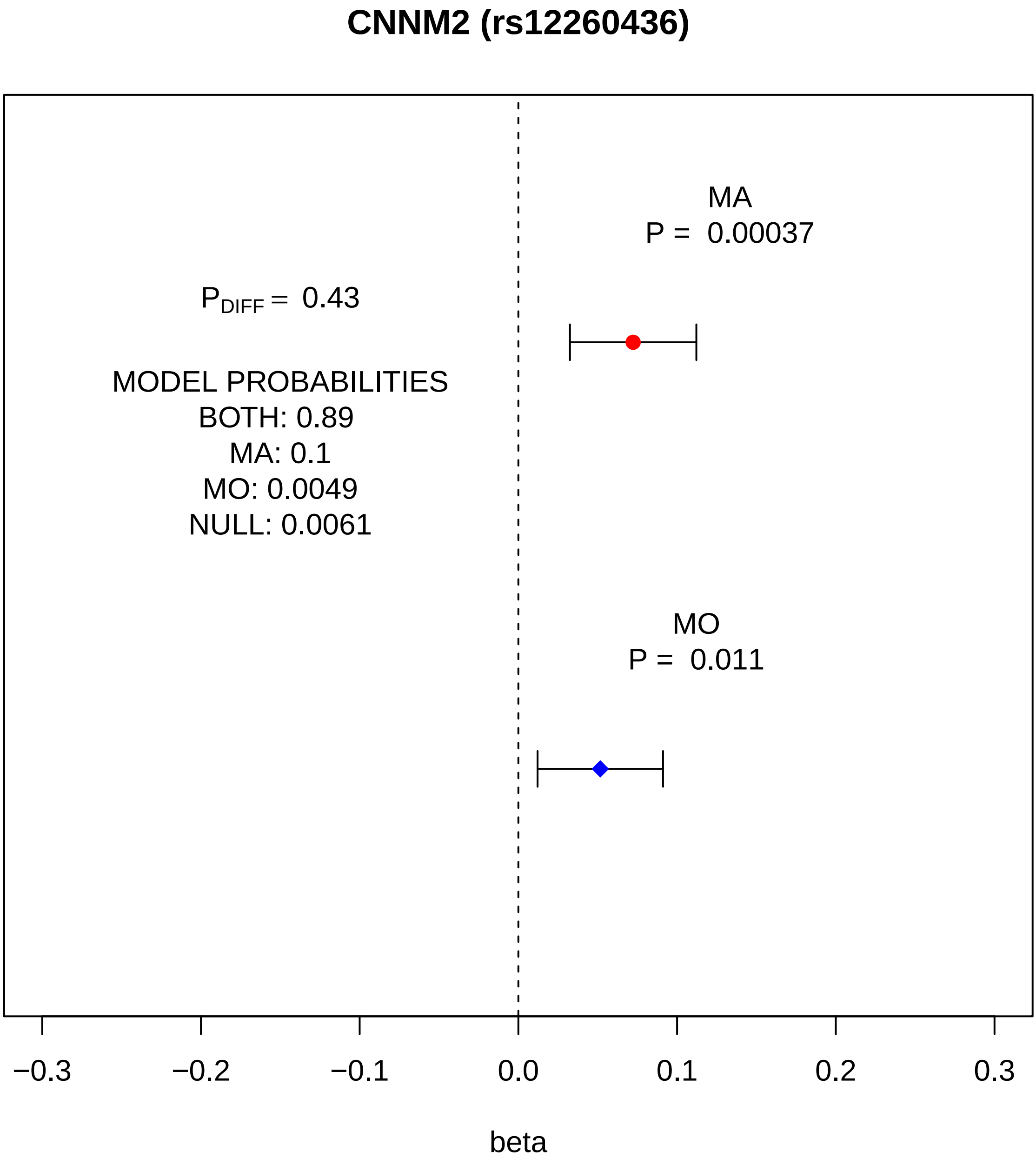
CNNM2 (rs12260436)
MA
P = 0.00037
P_DIFF = 0.43
MODEL PROBABILITIES
BOTH: 0.89
MA: 0.1
MO: 0.0049
NULL: 0.0061
MO
P = 0.011
−0.3
−0.2
−0.1
0.0
0.1
0.2
0.3
beta

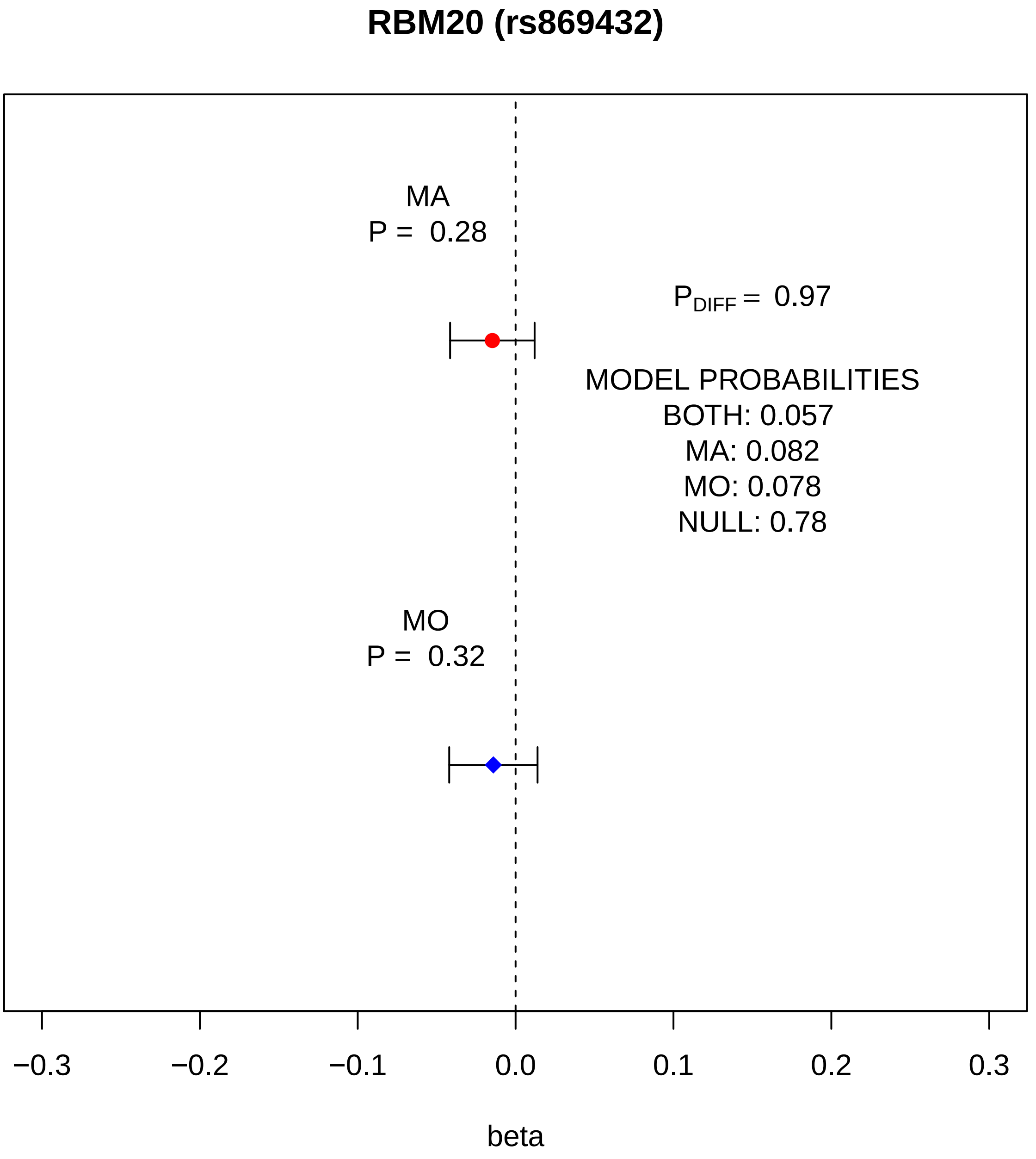

RBM20 (rs869432)
MA
P = 0.28
P_DIFF = 0.97
MODEL PROBABILITIES
BOTH: 0.057
MA: 0.082
MO: 0.078
NULL: 0.78
MO
P = 0.32
−0.3
−0.2
−0.1
0.0
0.1
0.2
0.3
beta

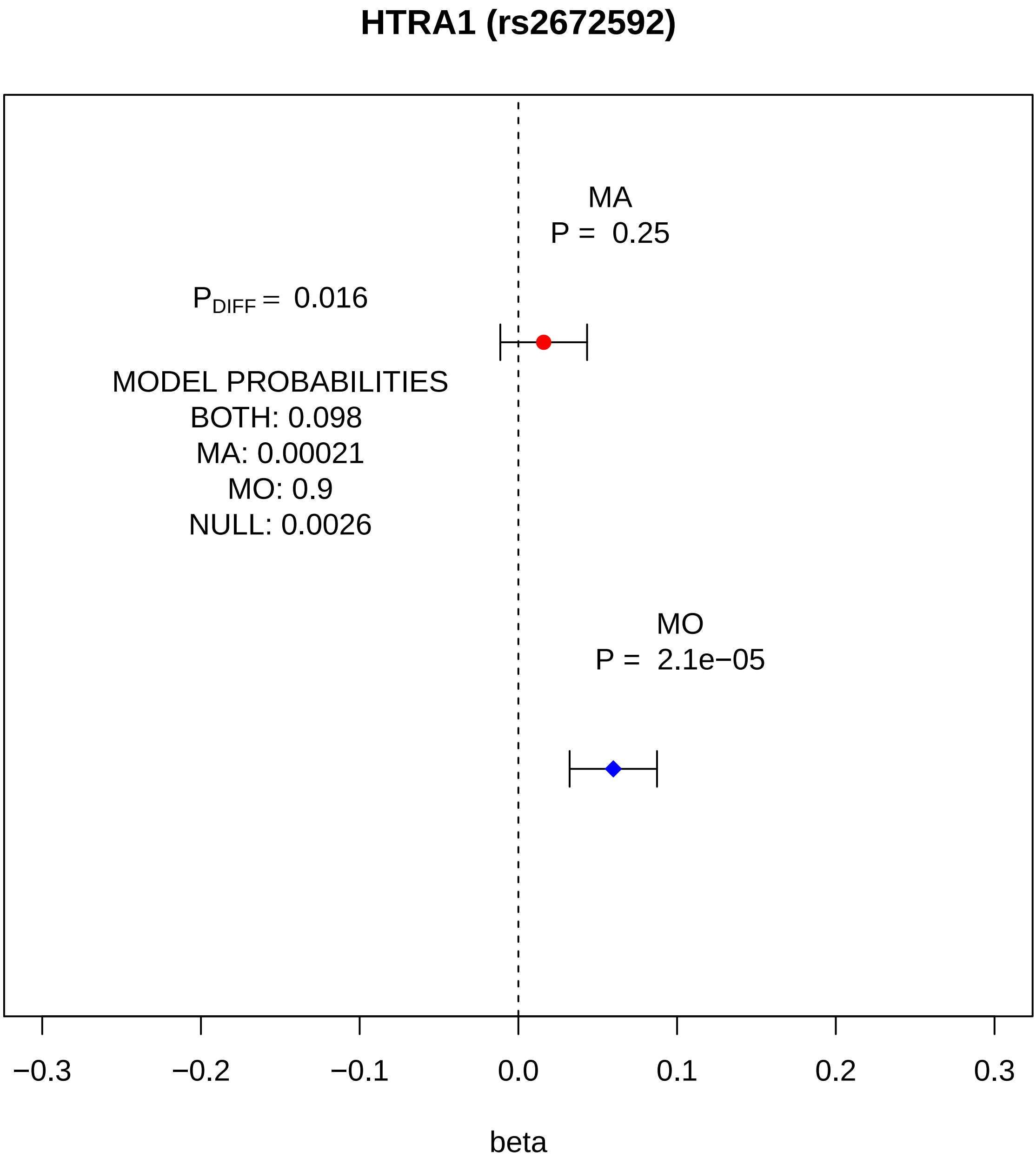
HTRA1 (rs2672592)
MA
P = 0.25
P_DIFF = 0.016
MODEL PROBABILITIES
BOTH: 0.098
MA: 0.00021
MO: 0.9
NULL: 0.0026
MO
P = 2.1e−05
−0.3
−0.2
−0.1
0.0
0.1
0.2
0.3
beta

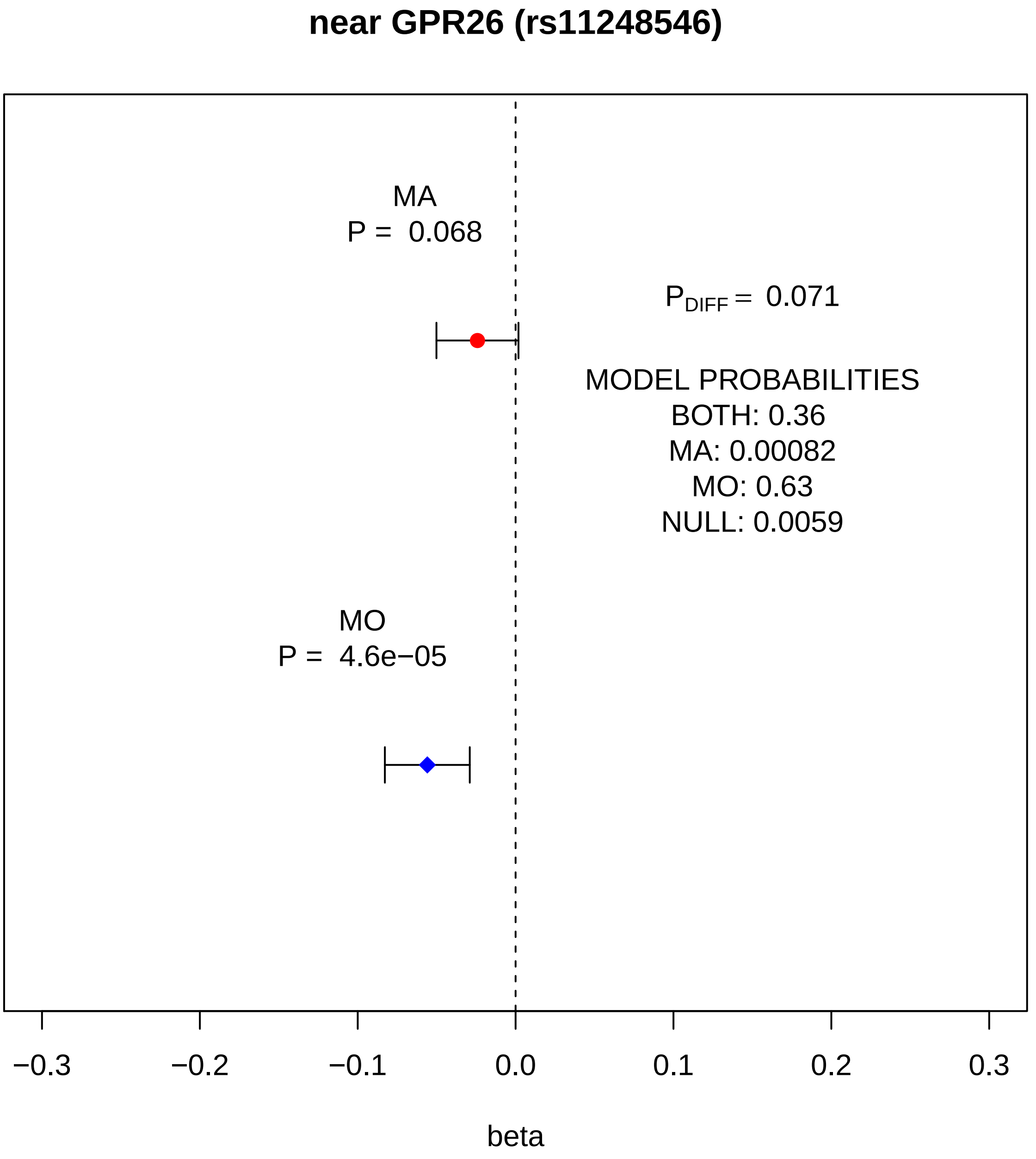
near GPR26 (rs11248546)
MA
P = 0.068
$P_{DIFF}$ = 0.071
MODEL PROBABILITIES
BOTH: 0.36
MA: 0.00082
MO: 0.63
NULL: 0.0059
MO
P = 4.6e−05
−0.3
−0.2
−0.1
0.0
0.1
0.2
0.3
beta

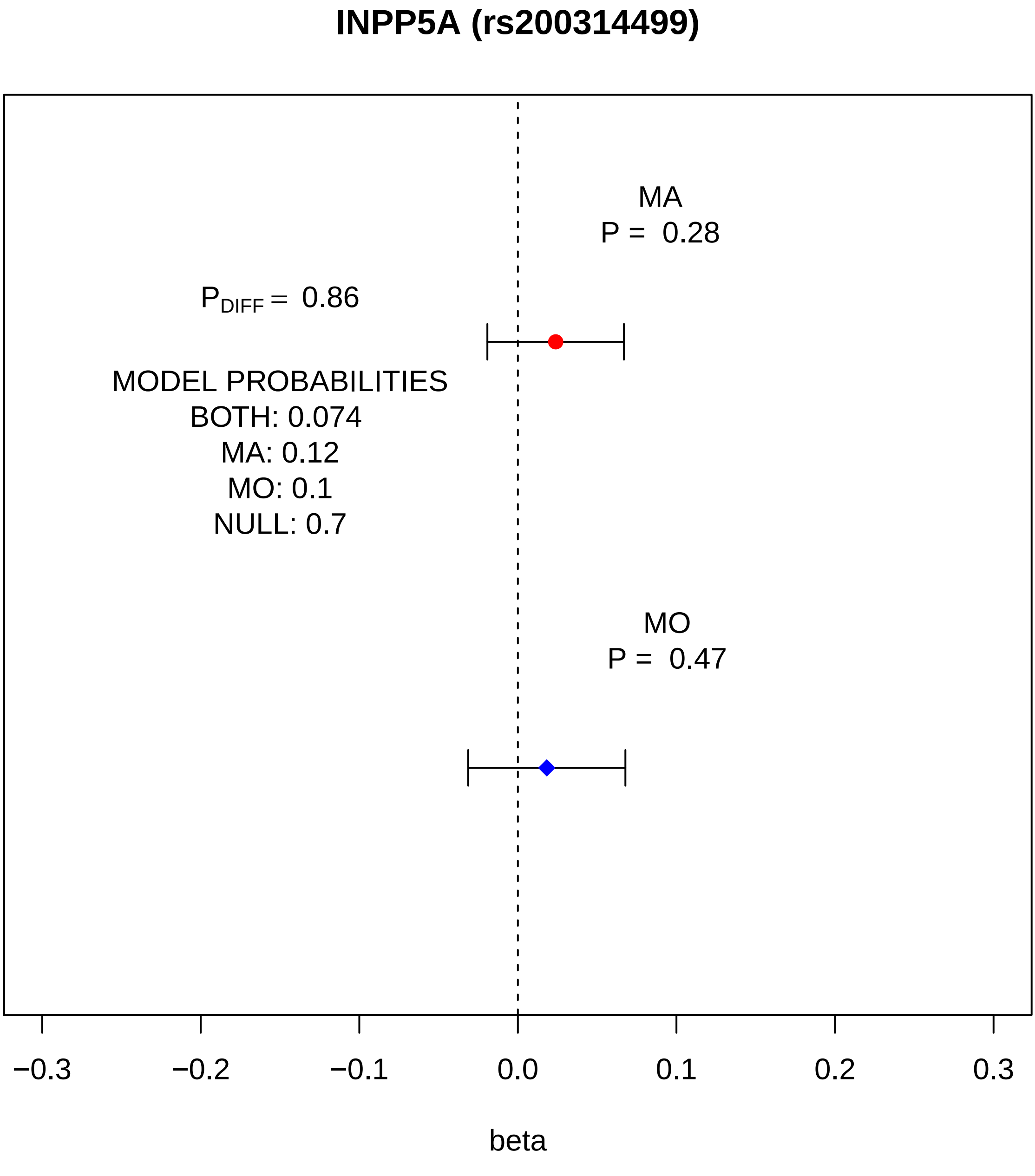
INPP5A (rs200314499)
MA
P = 0.28
$P_{DIFF}$ = 0.86
MODEL PROBABILITIES
BOTH: 0.074
MA: 0.12
MO: 0.1
NULL: 0.7
MO
P = 0.47
−0.3
−0.2
−0.1
0.0
0.1
0.2
0.3
beta

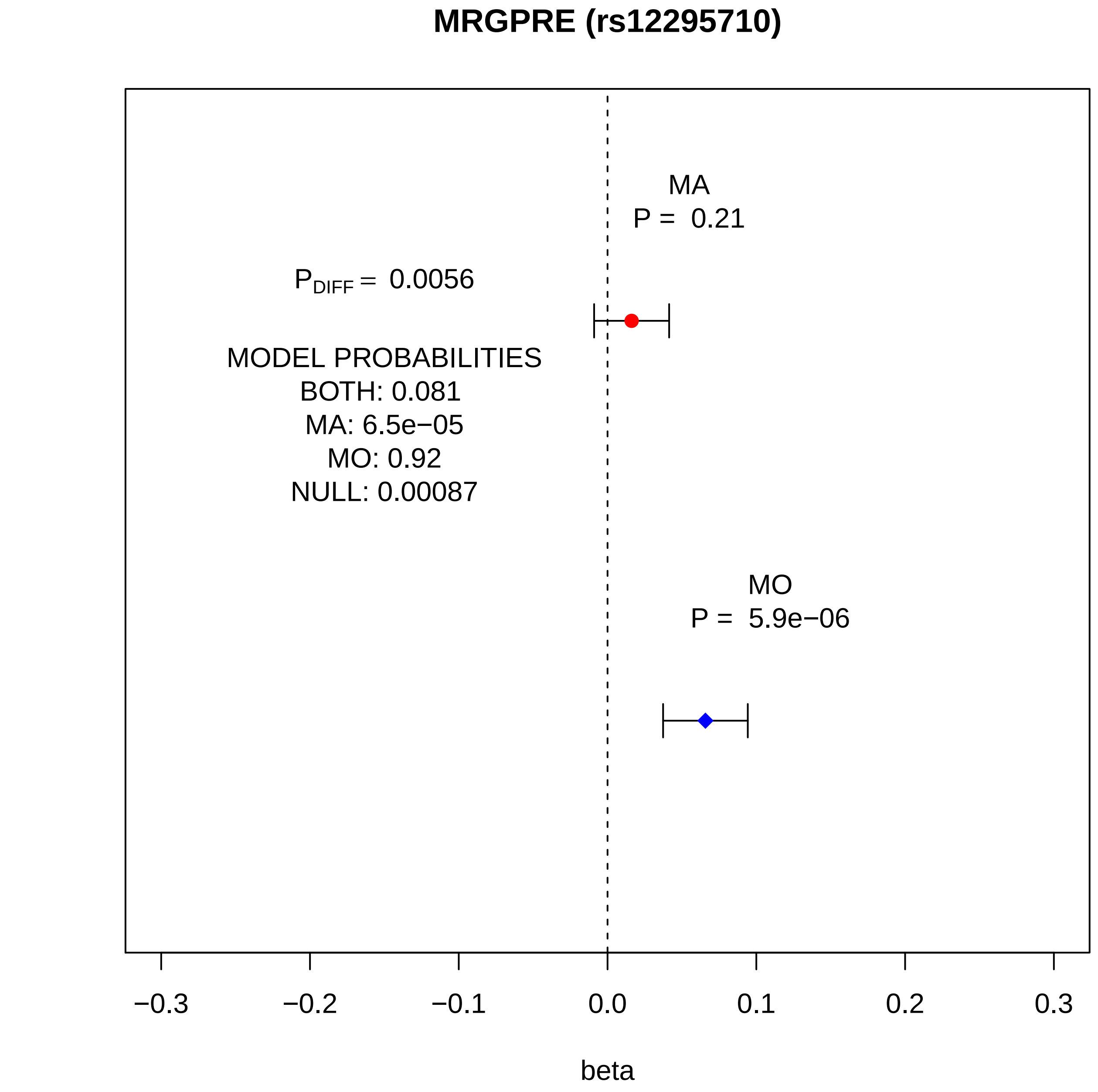
MRGPRE (rs12295710)
MA
P = 0.21
$P_{DIFF}$ = 0.0056
MODEL PROBABILITIES
BOTH: 0.081
MA: 6.5e−05
MO: 0.92
NULL: 0.00087
MO
P = 5.9e−06
−0.3
−0.2
−0.1
0.0
0.1
0.2
0.3
beta

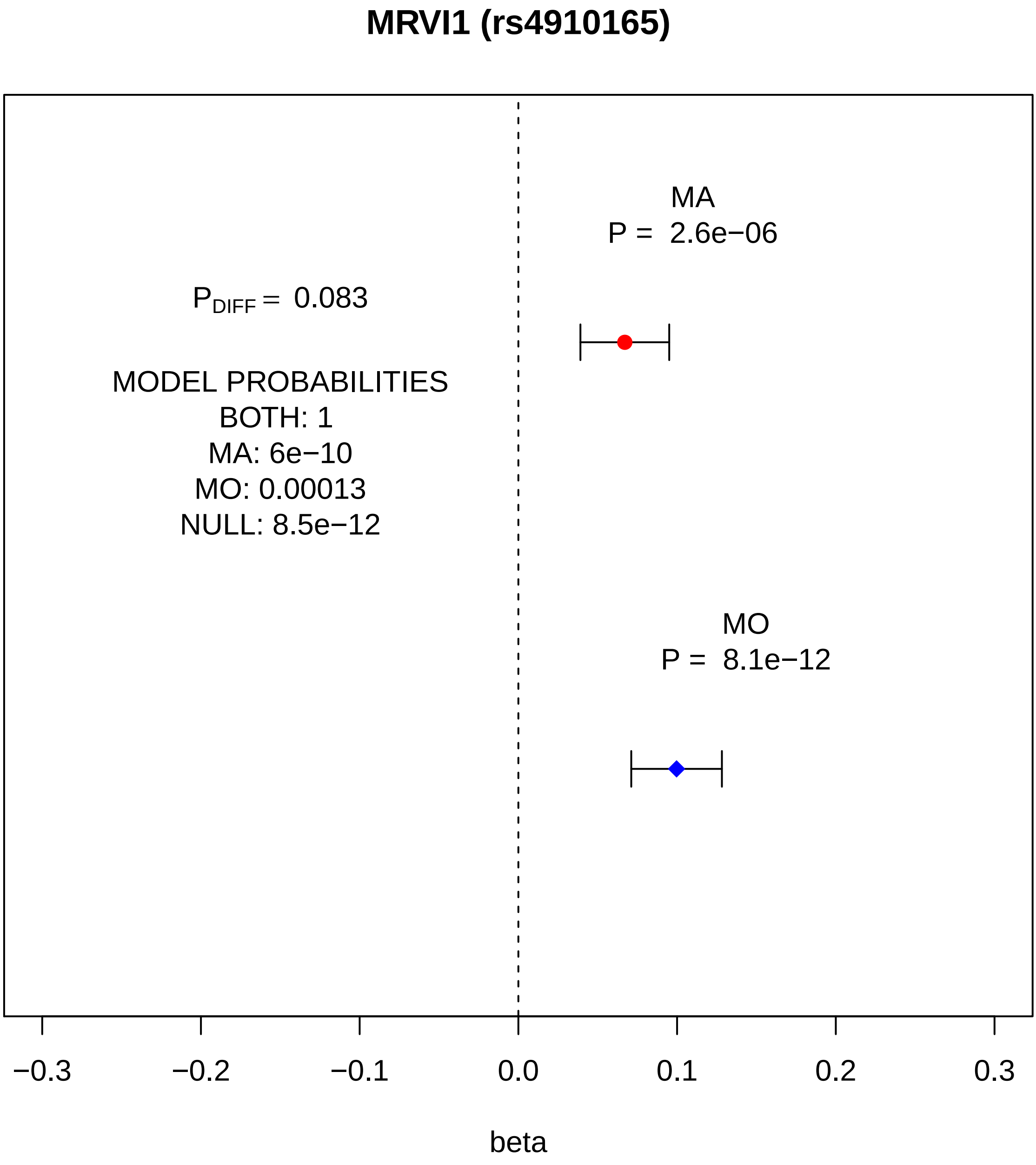
MRVI1 (rs4910165)
MA
P = 2.6e−06
$P_{DIFF}$ = 0.083
MODEL PROBABILITIES
BOTH: 1
MA: 6e−10
MO: 0.00013
NULL: 8.5e−12
MO
P = 8.1e−12
−0.3
−0.2
−0.1
0.0
0.1
0.2
0.3
beta

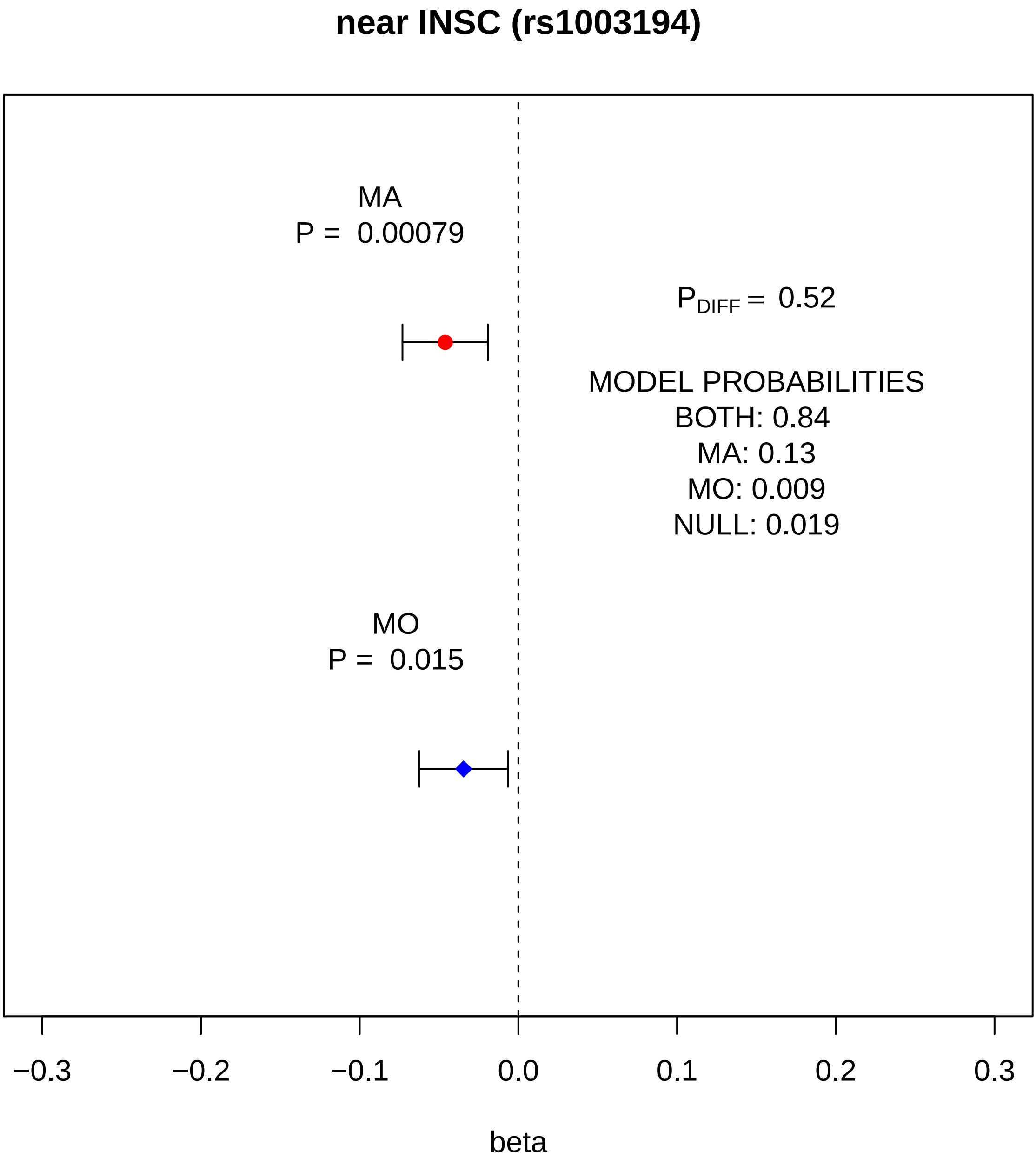
near INSC (rs1003194)
MA
P = 0.00079
$P_{DIFF}$ = 0.52
MODEL PROBABILITIES
BOTH: 0.84
MA: 0.13
MO: 0.009
NULL: 0.019
MO
P = 0.015
−0.3
−0.2
−0.1
0.0
0.1
0.2
0.3
beta

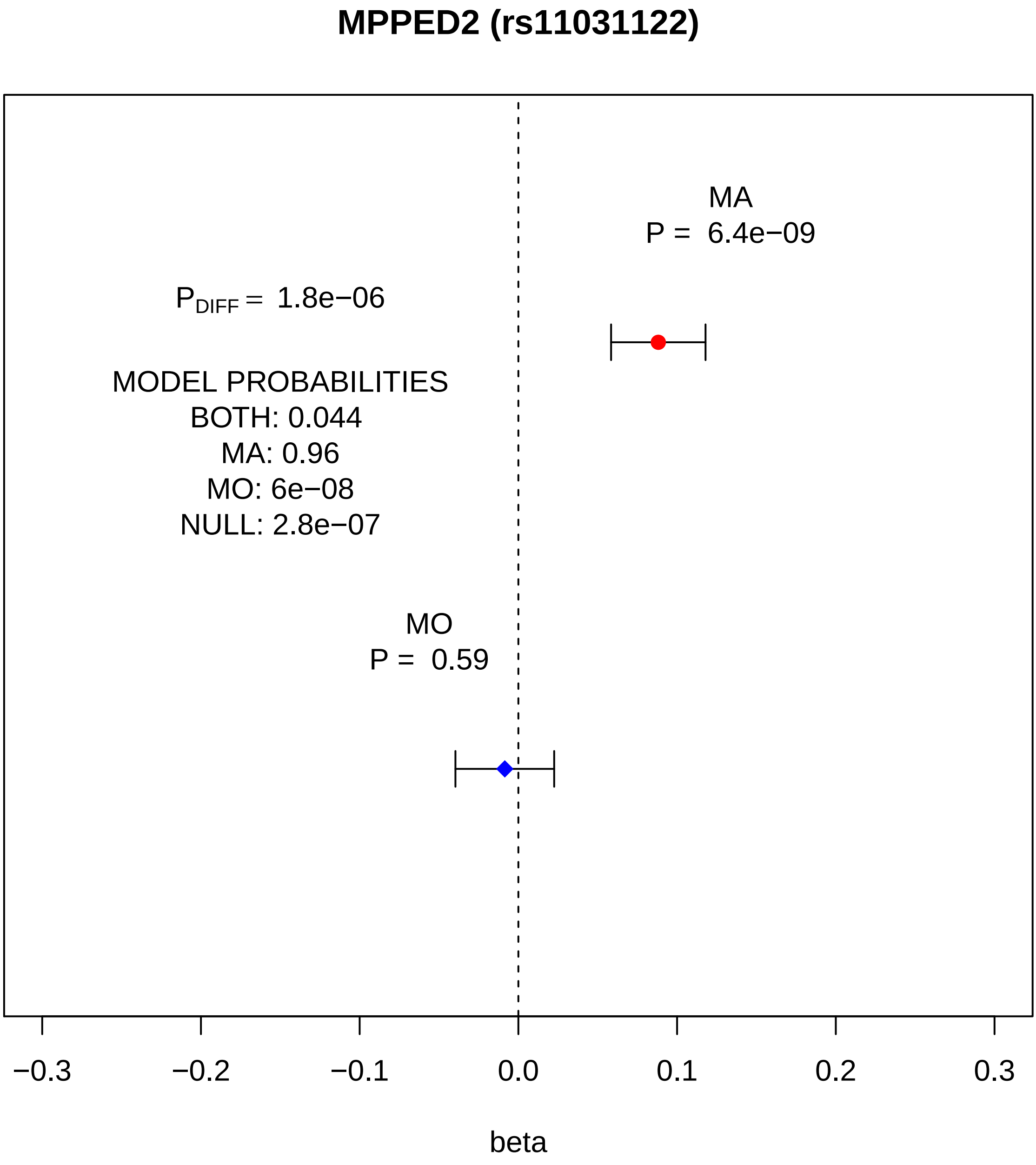
MPPED2 (rs11031122)
MA
P = 6.4e−09
P_DIFF = 1.8e−06
MODEL PROBABILITIES
BOTH: 0.044
MA: 0.96
MO: 6e−08
NULL: 2.8e−07
MO
P = 0.59
−0.3
−0.2
−0.1
0.0
0.1
0.2
0.3
beta

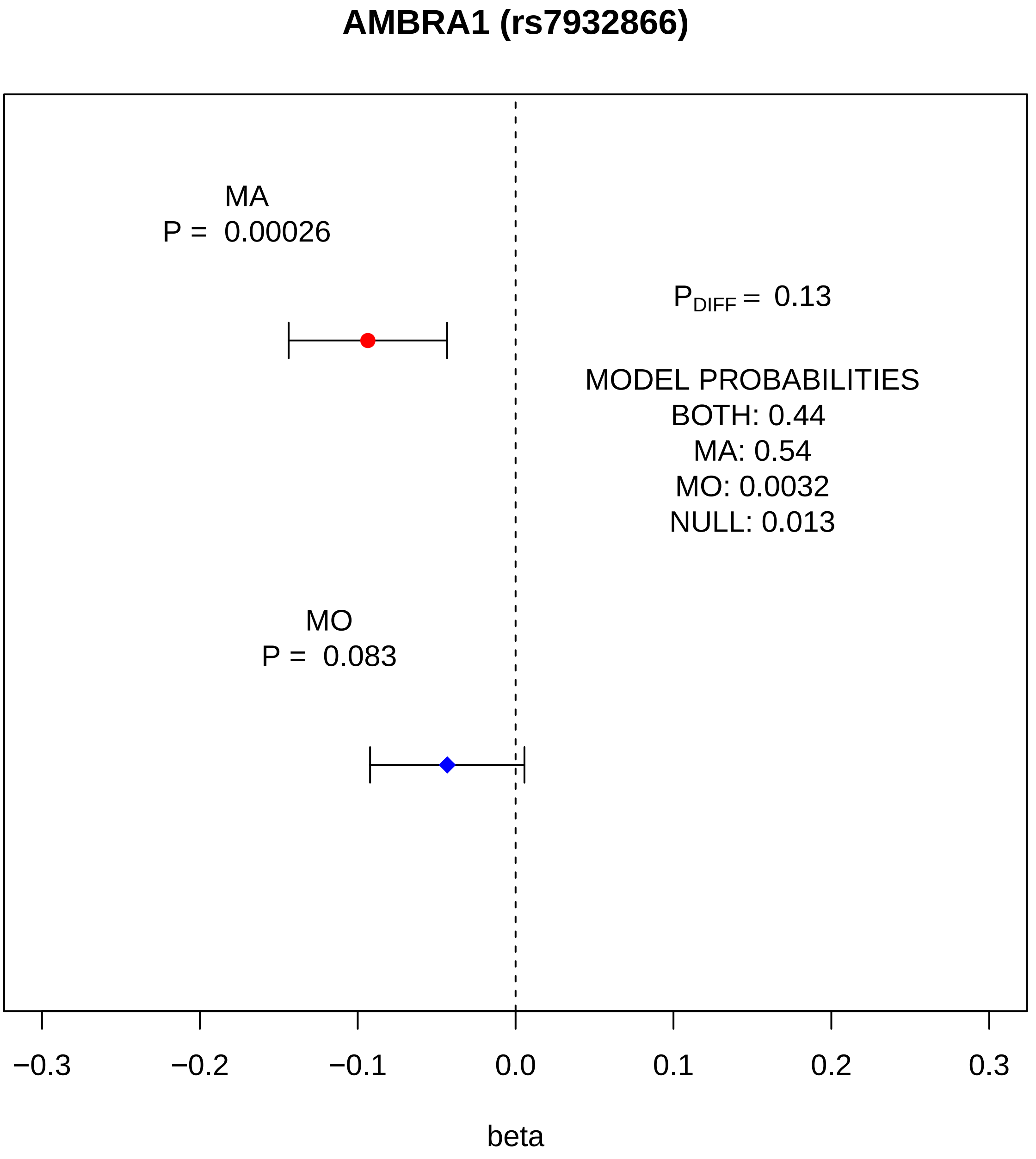
AMBRA1 (rs7932866)
MA
P = 0.00026
P_DIFF = 0.13
MODEL PROBABILITIES
BOTH: 0.44
MA: 0.54
MO: 0.0032
NULL: 0.013
MO
P = 0.083
−0.3
−0.2
−0.1
0.0
0.1
0.2
0.3
beta

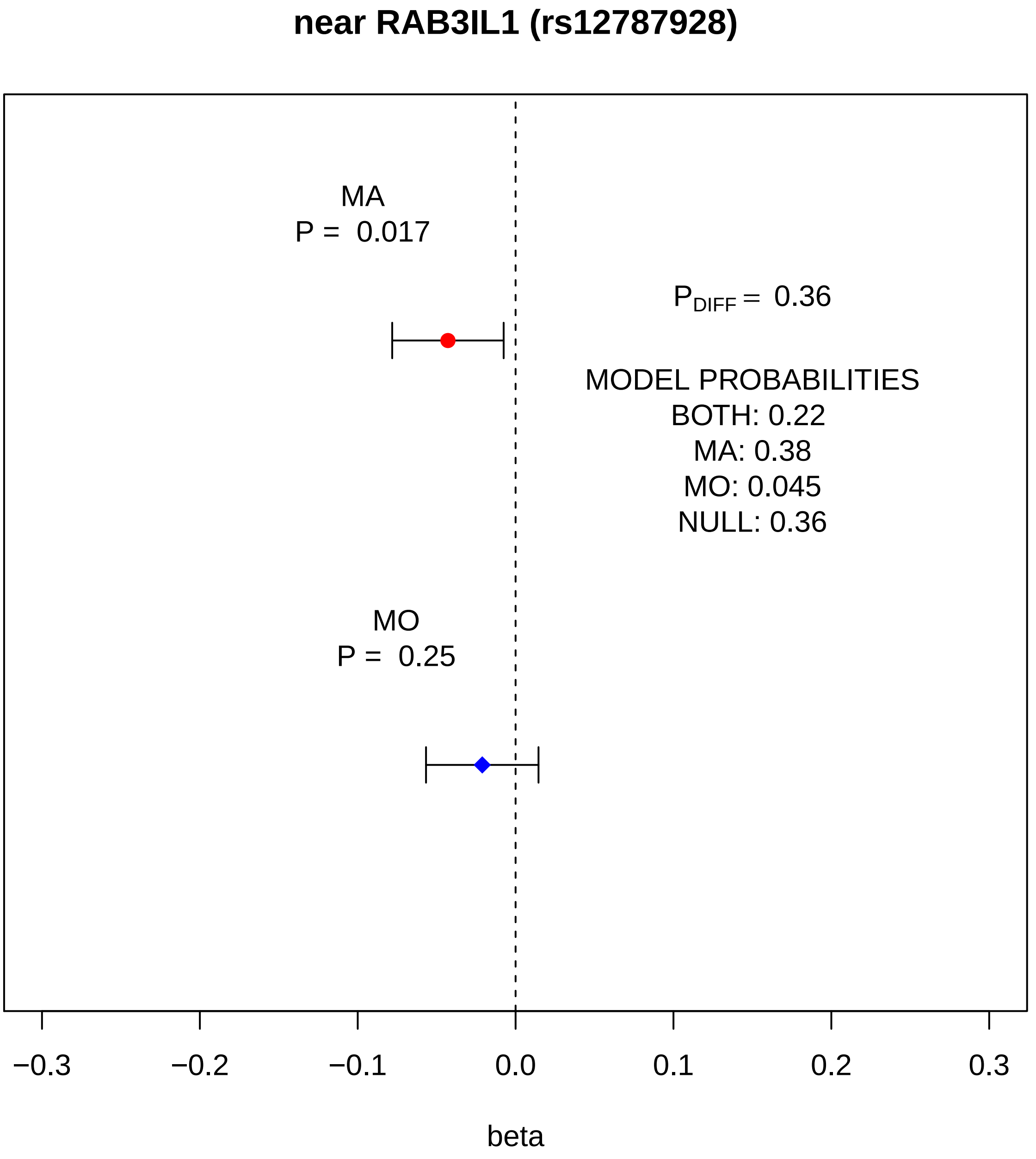

near RAB3IL1 (rs12787928)
MA
P = 0.017
P DIFF = 0.36
MODEL PROBABILITIES
BOTH: 0.22
MA: 0.38
MO: 0.045
NULL: 0.36
MO
P = 0.25
−0.3
−0.2
−0.1
0.0
0.1
0.2
0.3
beta

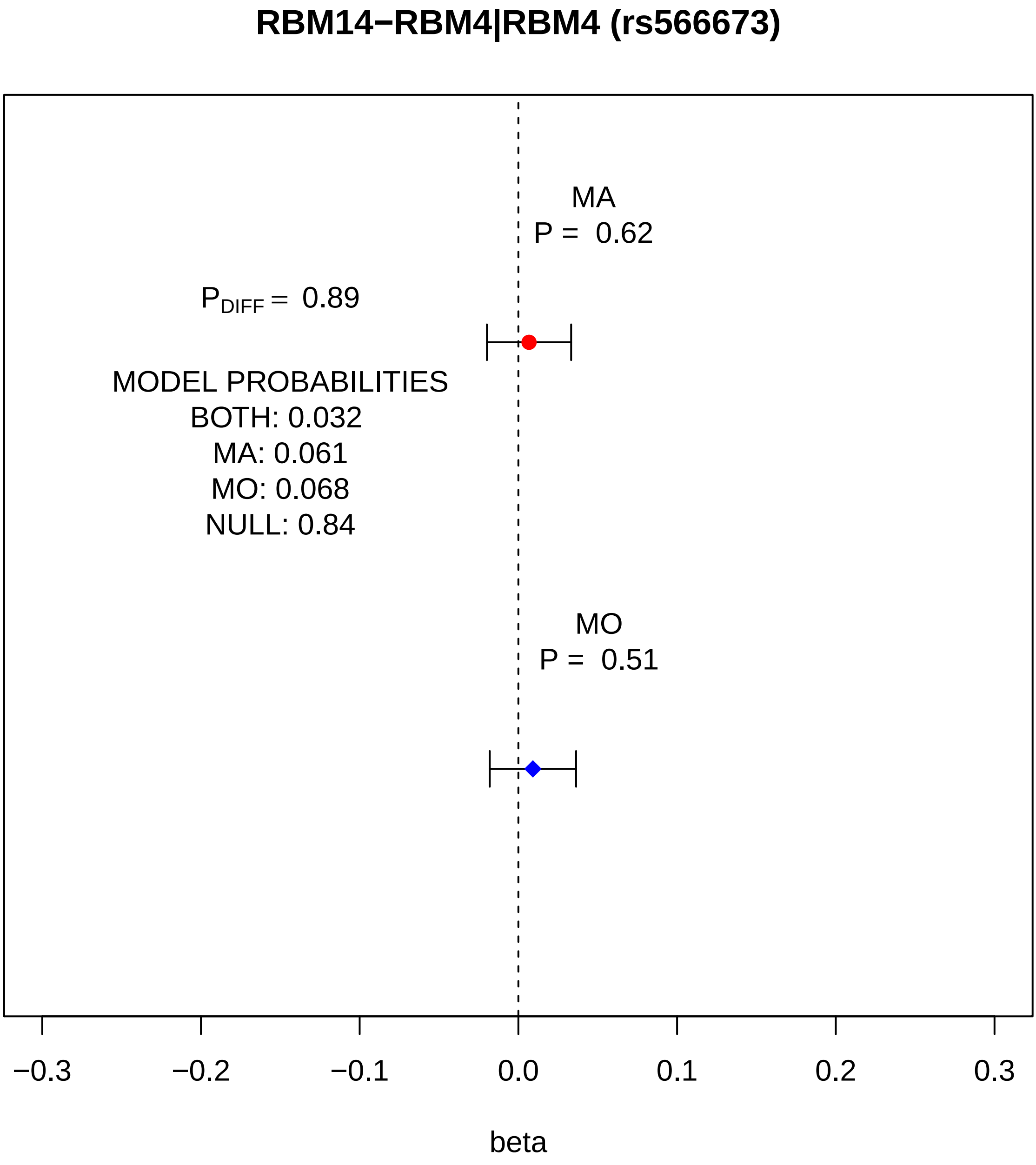
RBM14−RBM4|RBM4 (rs566673)
MA
P = 0.62
P_DIFF = 0.89
MODEL PROBABILITIES
BOTH: 0.032
MA: 0.061
MO: 0.068
NULL: 0.84
MO
P = 0.51
−0.3
−0.2
−0.1
0.0
0.1
0.2
0.3
beta

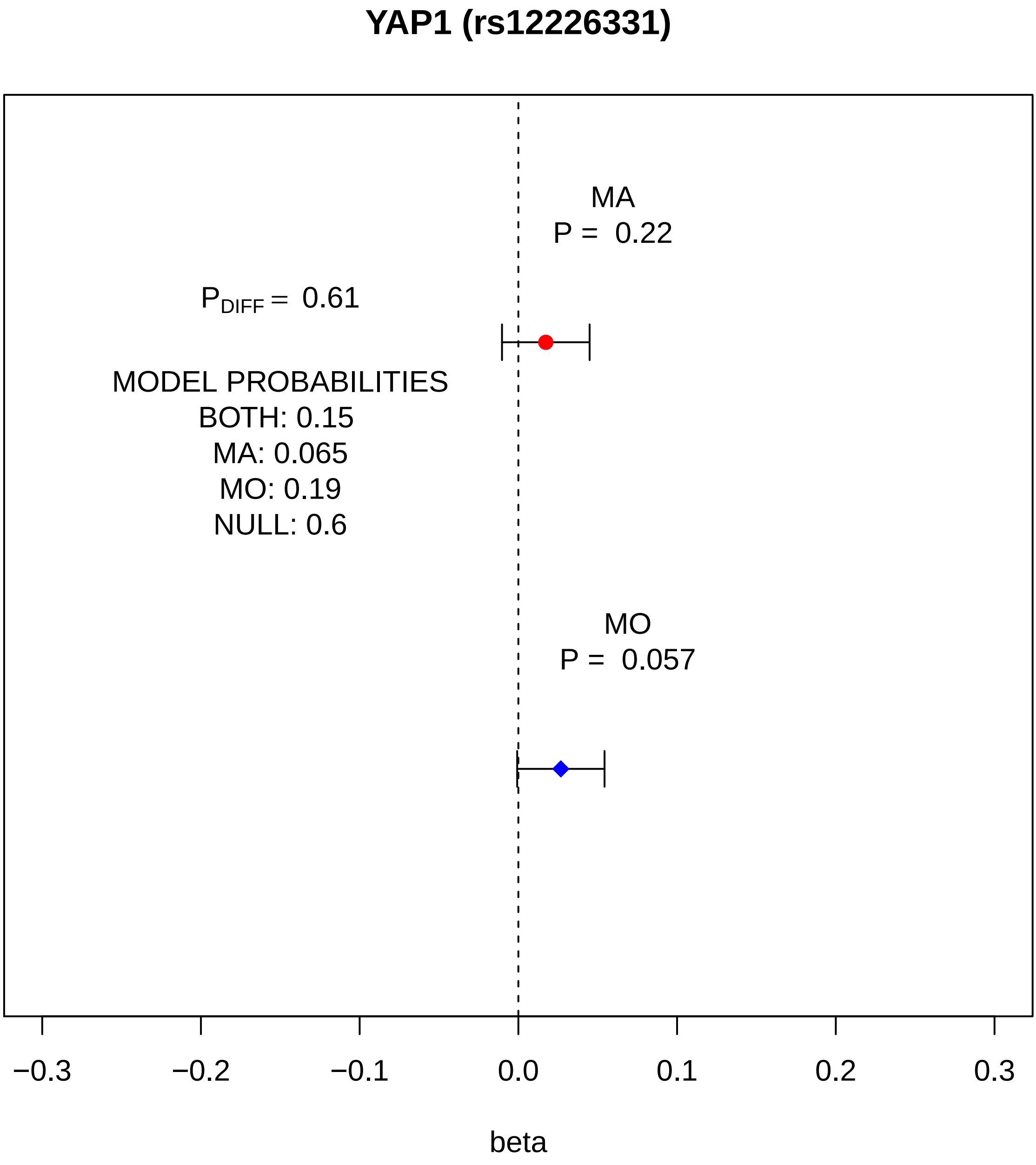

YAP1 (rs12226331)
MA
P = 0.22
P DIFF = 0.61
MODEL PROBABILITIES
BOTH: 0.15
MA: 0.065
MO: 0.19
NULL: 0.6
MO
P = 0.057
−0.3
−0.2
−0.1
0.0
0.1
0.2
0.3
beta

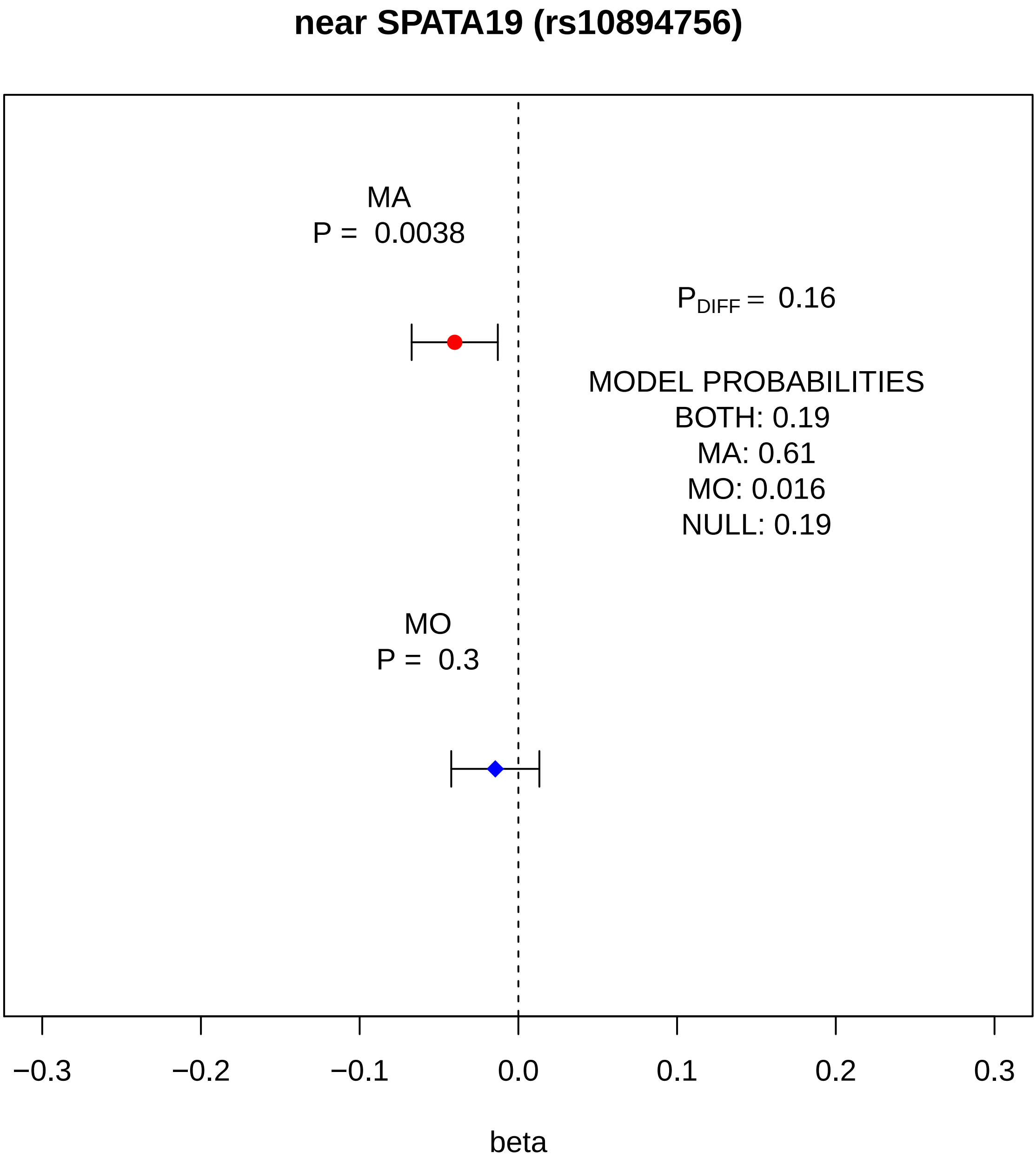
near SPATA19 (rs10894756)
MA
P = 0.0038
$P_{DIFF}$ = 0.16
MODEL PROBABILITIES
BOTH: 0.19
MA: 0.61
MO: 0.016
NULL: 0.19
MO
P = 0.3
−0.3
−0.2
−0.1
0.0
0.1
0.2
0.3
beta

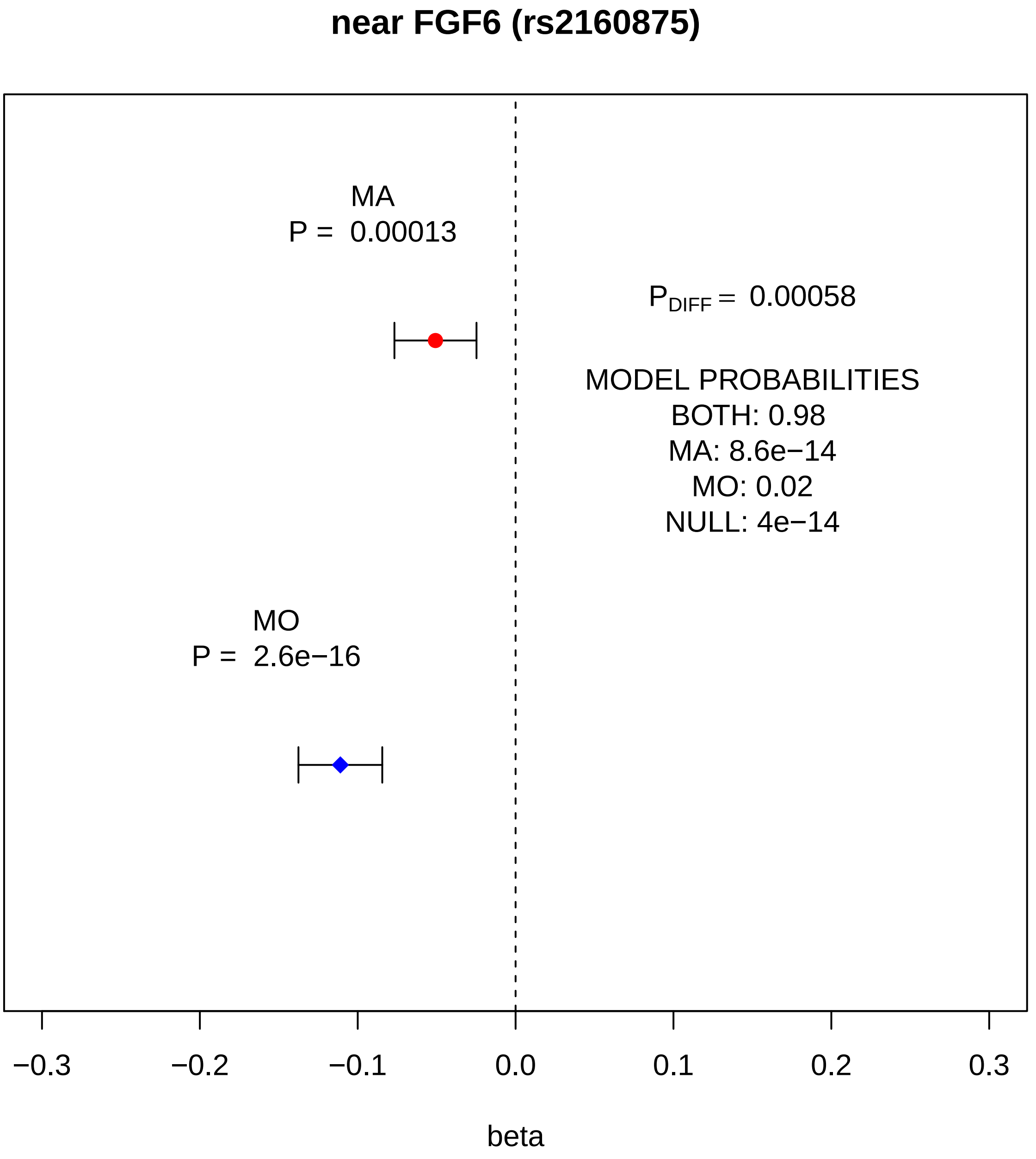
near FGF6 (rs2160875)
MA
P = 0.00013
MO
P = 2.6e−16
P_DIFF = 0.00058
MODEL PROBABILITIES
BOTH: 0.98
MA: 8.6e−14
MO: 0.02
NULL: 4e−14
−0.3
−0.2
−0.1
0.0
0.1
0.2
0.3
beta

#### PDZRN4 (rs1458170)

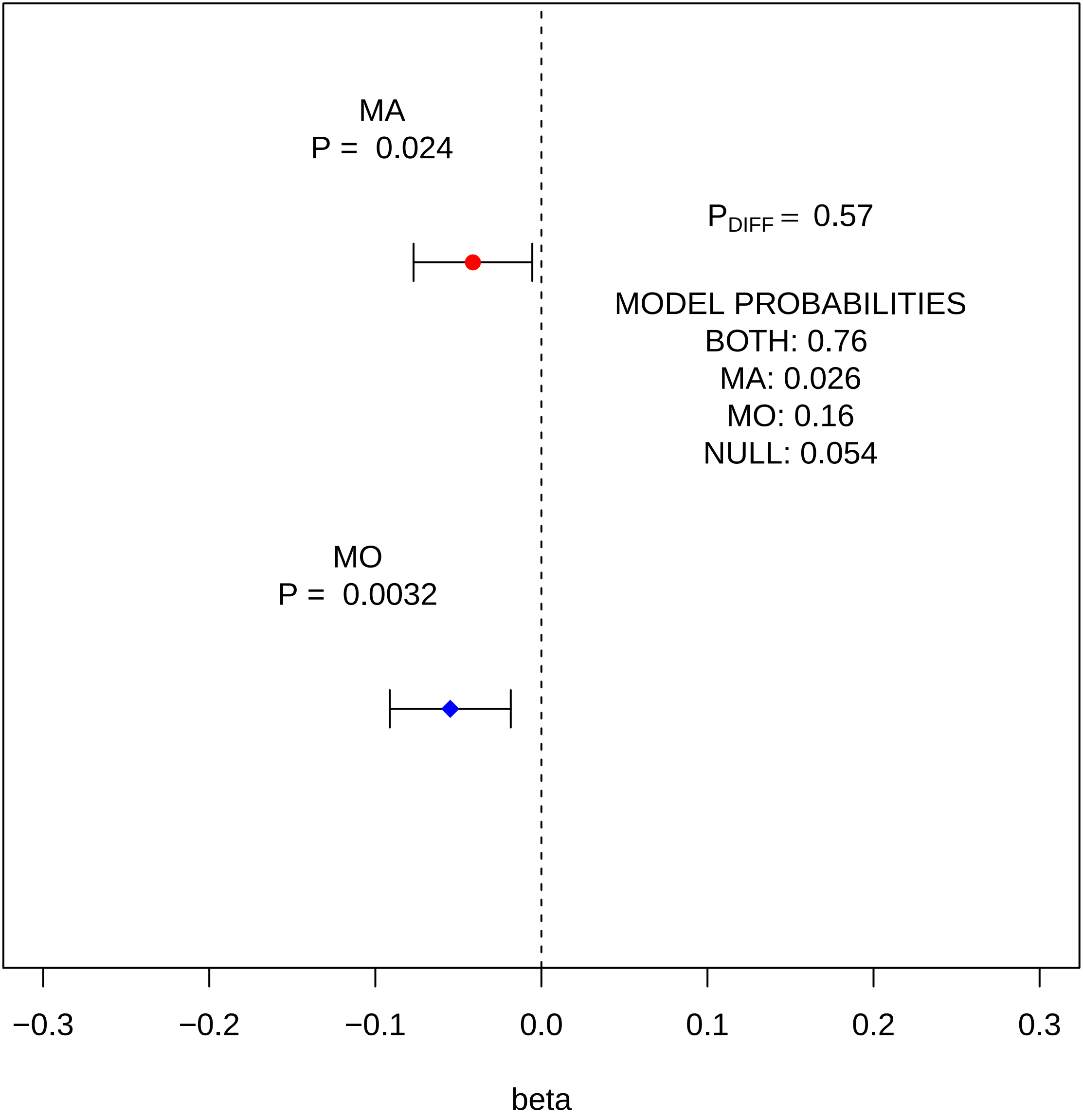

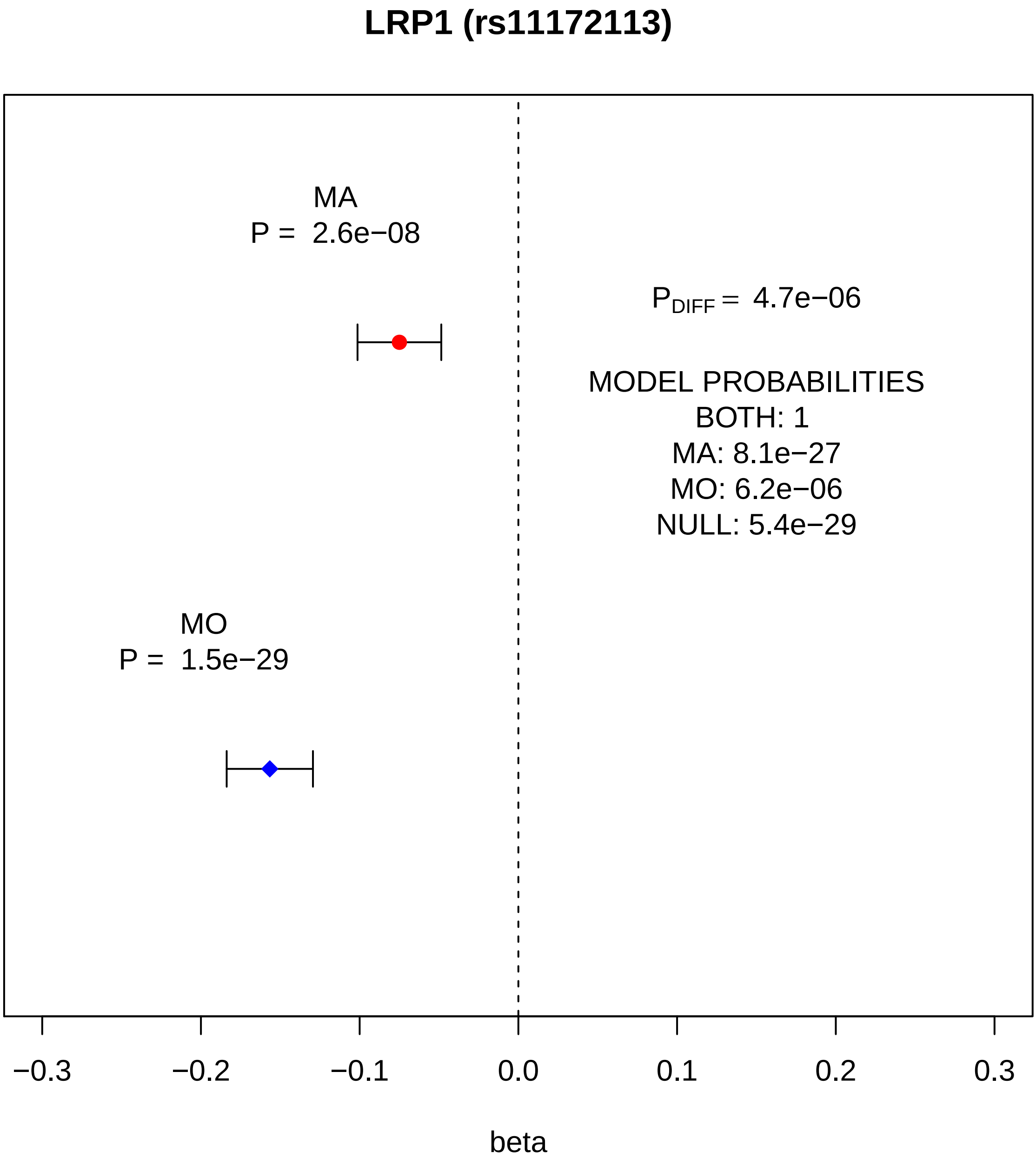
LRP1 (rs11172113)
MA
P = 2.6e−08
$P_{DIFF}$ = 4.7e−06
MODEL PROBABILITIES
BOTH: 1
MA: 8.1e−27
MO: 6.2e−06
NULL: 5.4e−29
MO
P = 1.5e−29
−0.3
−0.2
−0.1
0.0
0.1
0.2
0.3
beta

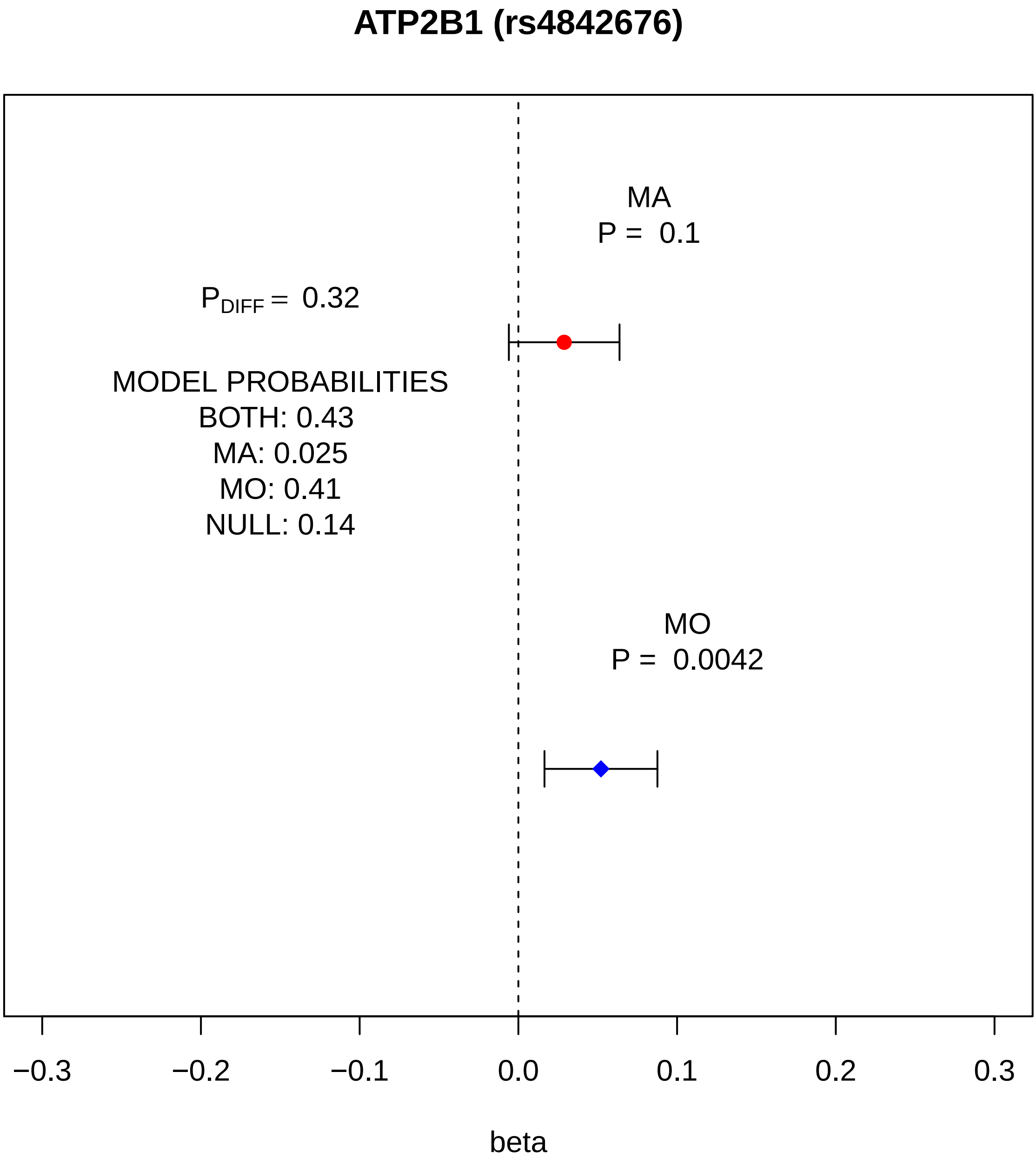
ATP2B1 (rs4842676)
MA
P = 0.1
P_DIFF = 0.32
MODEL PROBABILITIES
BOTH: 0.43
MA: 0.025
MO: 0.41
NULL: 0.14
MO
P = 0.0042
−0.3
−0.2
−0.1
0.0
0.1
0.2
0.3
beta

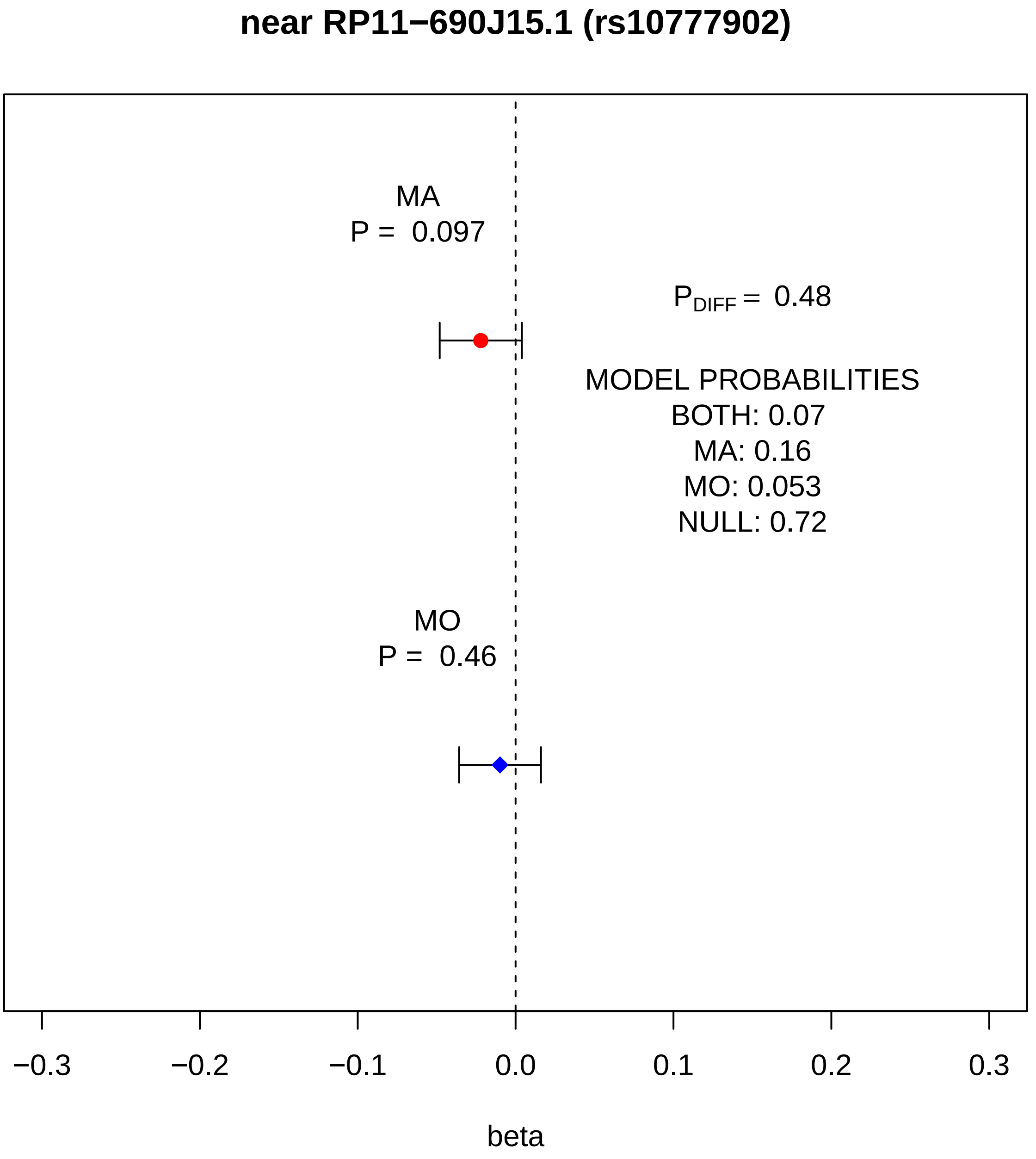
near RP11−690J15.1 (rs10777902)
MA
P = 0.097
P_DIFF = 0.48
MODEL PROBABILITIES
BOTH: 0.07
MA: 0.16
MO: 0.053
NULL: 0.72
MO
P = 0.46
−0.3
−0.2
−0.1
0.0
0.1
0.2
0.3
beta

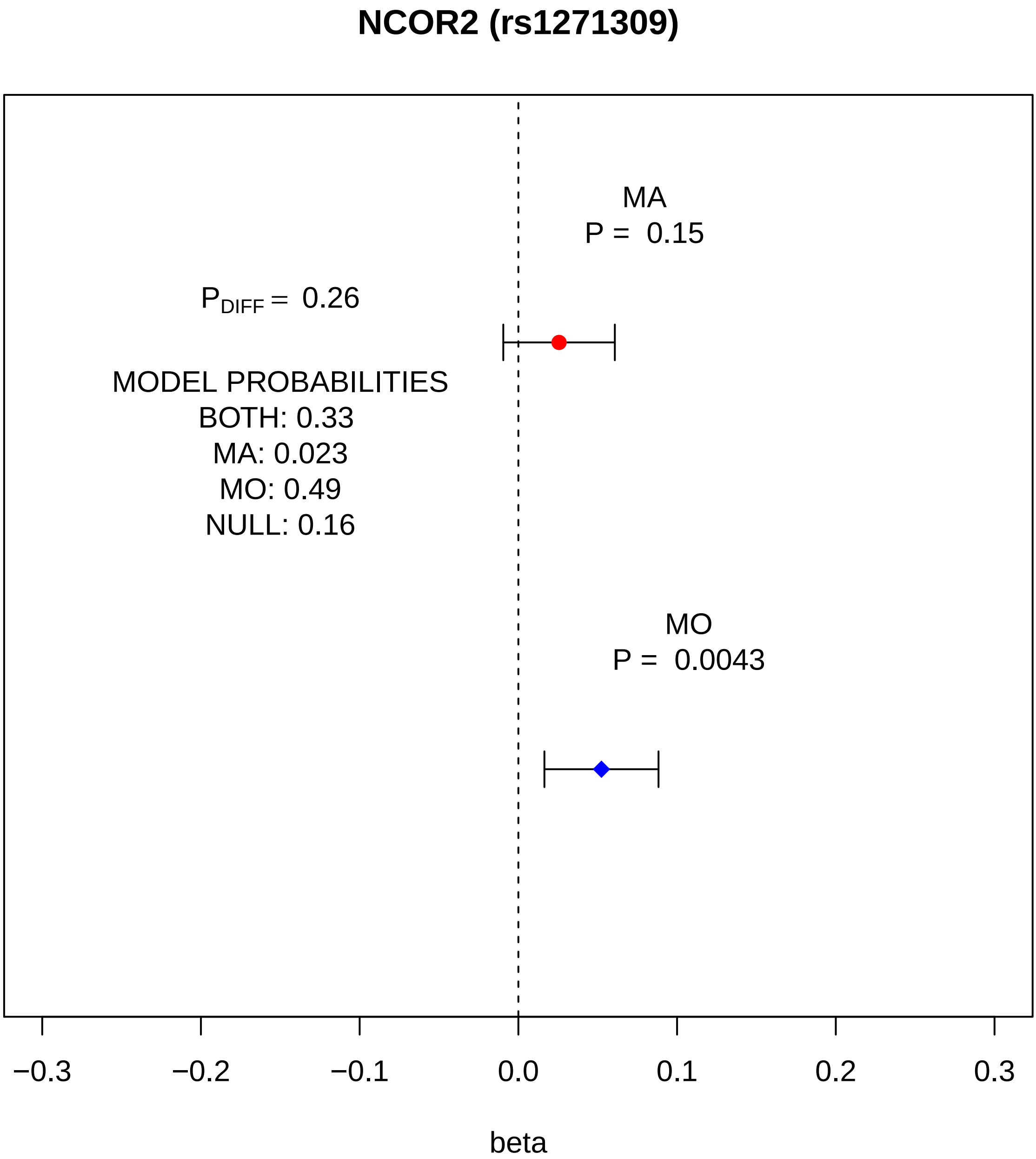

NCOR2 (rs1271309)
MA
P = 0.15
P_DIFF = 0.26
MODEL PROBABILITIES
BOTH: 0.33
MA: 0.023
MO: 0.49
NULL: 0.16
MO
P = 0.0043
−0.3
−0.2
−0.1
0.0
0.1
0.2
0.3
beta

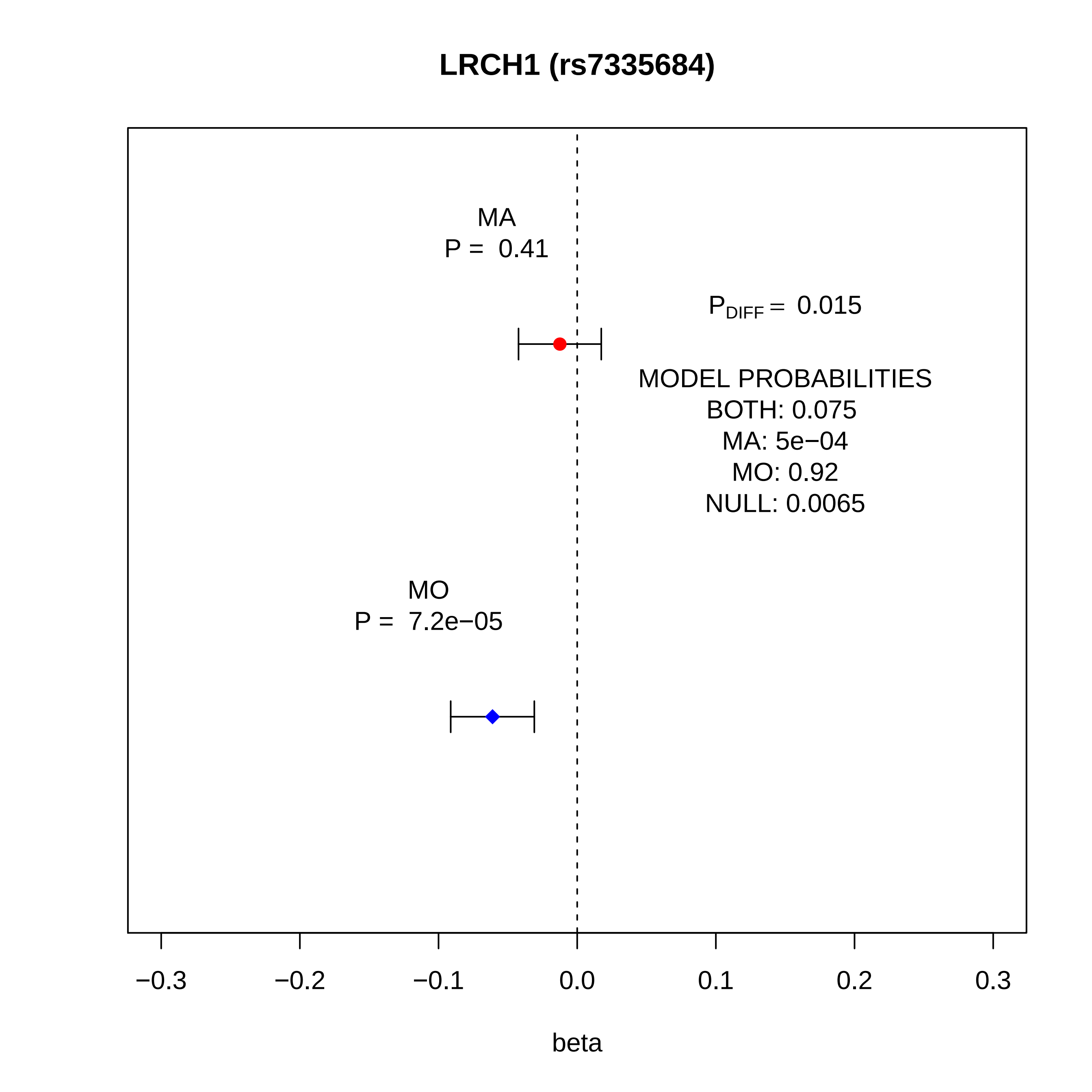

LRCH1 (rs7335684)
MA
P = 0.41
$P_{DIFF}$ = 0.015
MODEL PROBABILITIES
BOTH: 0.075
MA: 5e−04
MO: 0.92
NULL: 0.0065
MO
P = 7.2e−05
−0.3
−0.2
−0.1
0.0
0.1
0.2
0.3
beta

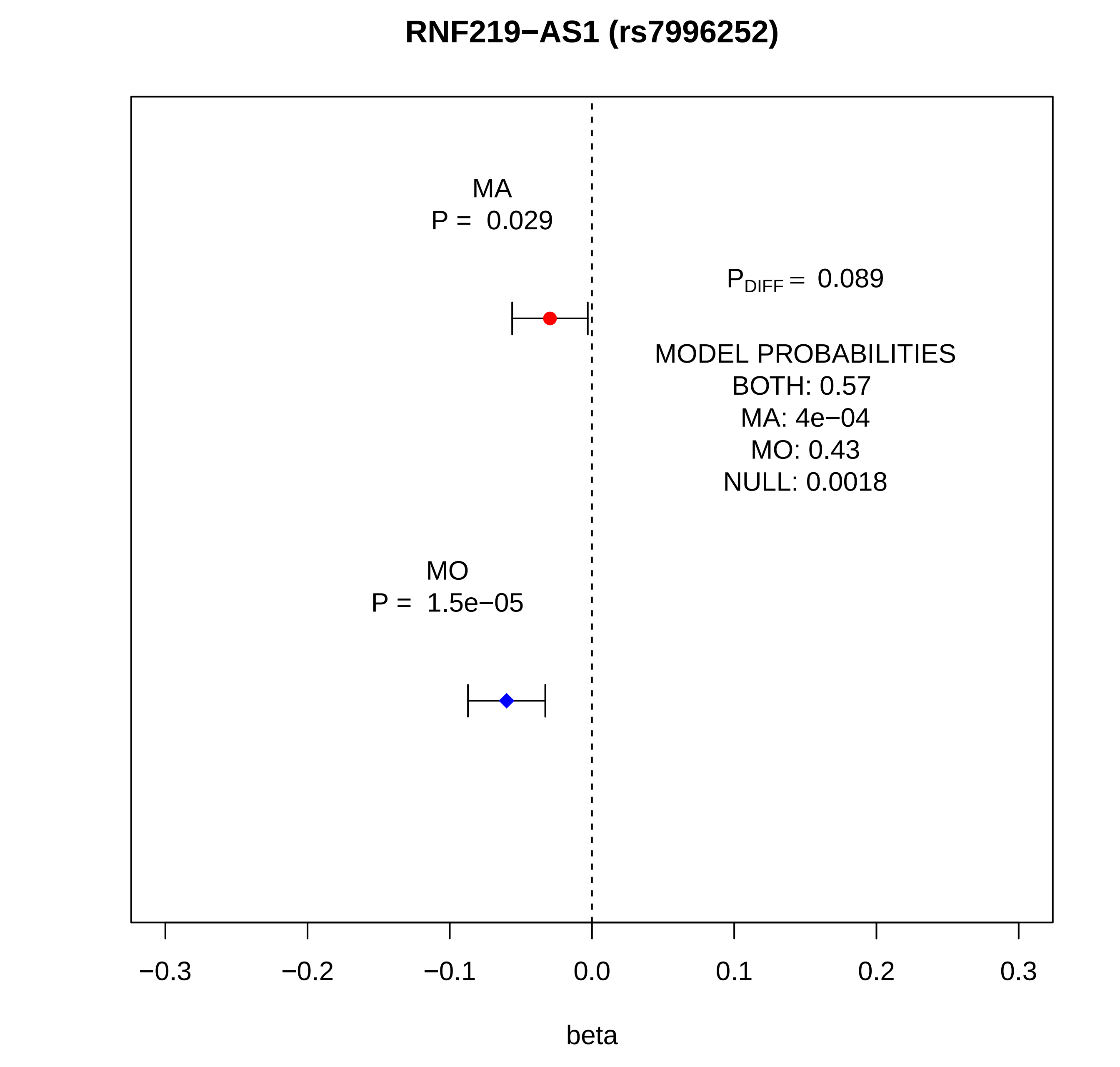
RNF219−AS1 (rs7996252)
MA
P = 0.029
$P_{DIFF}$ = 0.089
MODEL PROBABILITIES
BOTH: 0.57
MA: 4e−04
MO: 0.43
NULL: 0.0018
MO
P = 1.5e−05
−0.3
−0.2
−0.1
0.0
0.1
0.2
0.3
beta

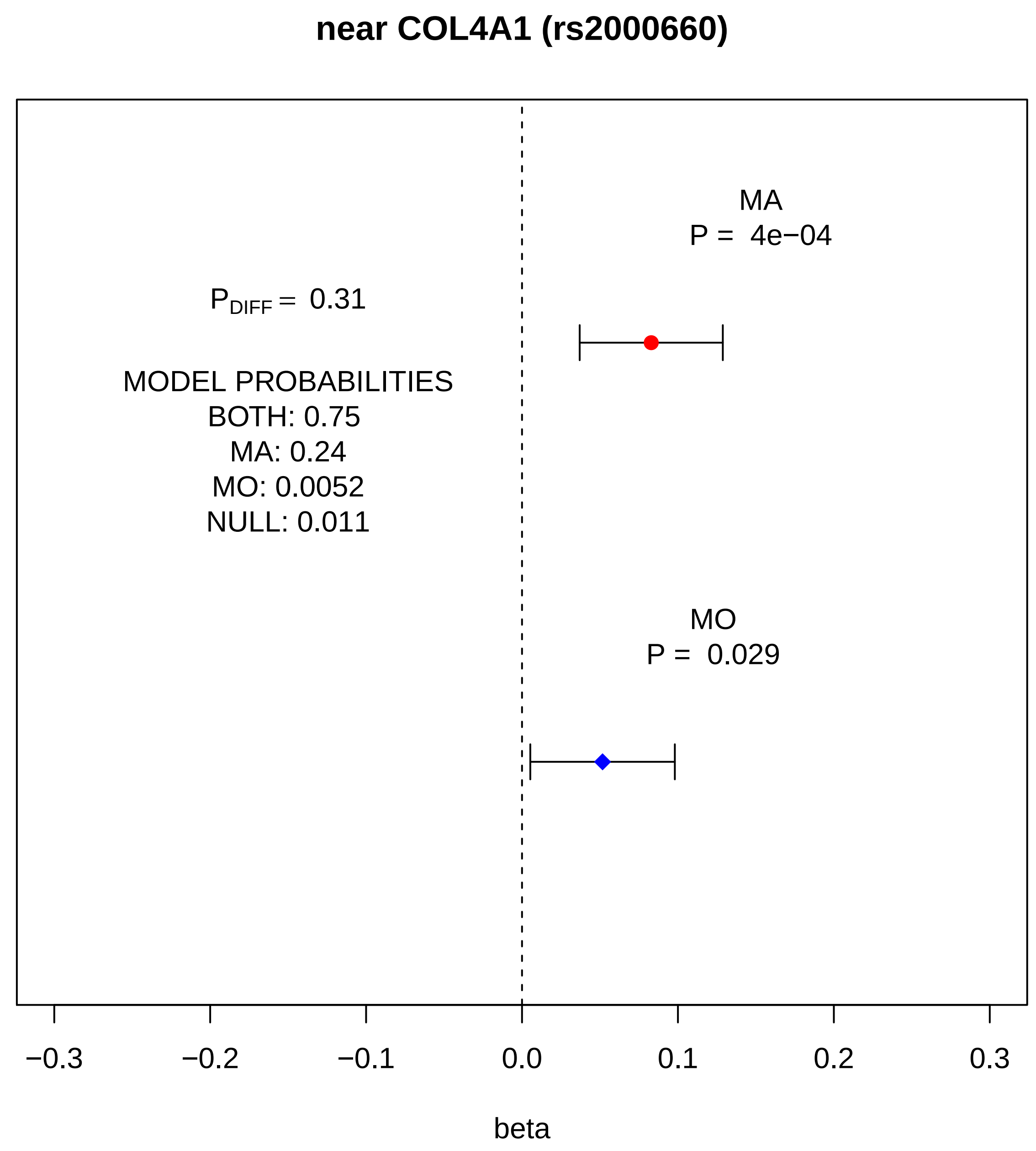
near COL4A1 (rs2000660)
MA
P = 4e−04
$P_{DIFF}$ = 0.31
MODEL PROBABILITIES
BOTH: 0.75
MA: 0.24
MO: 0.0052
NULL: 0.011
MO
P = 0.029
−0.3
−0.2
−0.1
0.0
0.1
0.2
0.3
beta

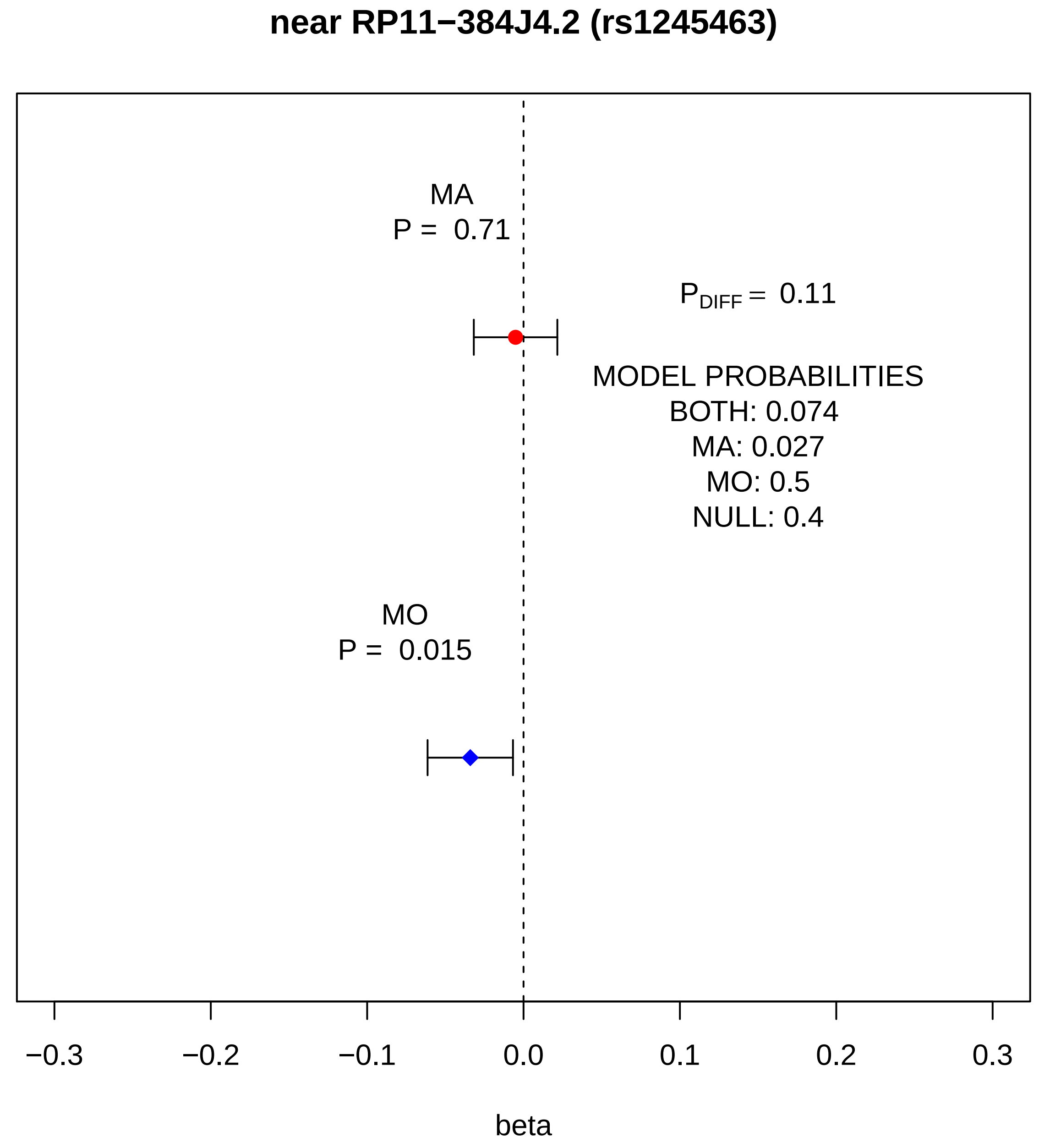
near RP11−384J4.2 (rs1245463)
MA
P = 0.71
$P_{DIFF}$ = 0.11
MODEL PROBABILITIES
BOTH: 0.074
MA: 0.027
MO: 0.5
NULL: 0.4
MO
P = 0.015
−0.3
−0.2
−0.1
0.0
0.1
0.2
0.3
beta

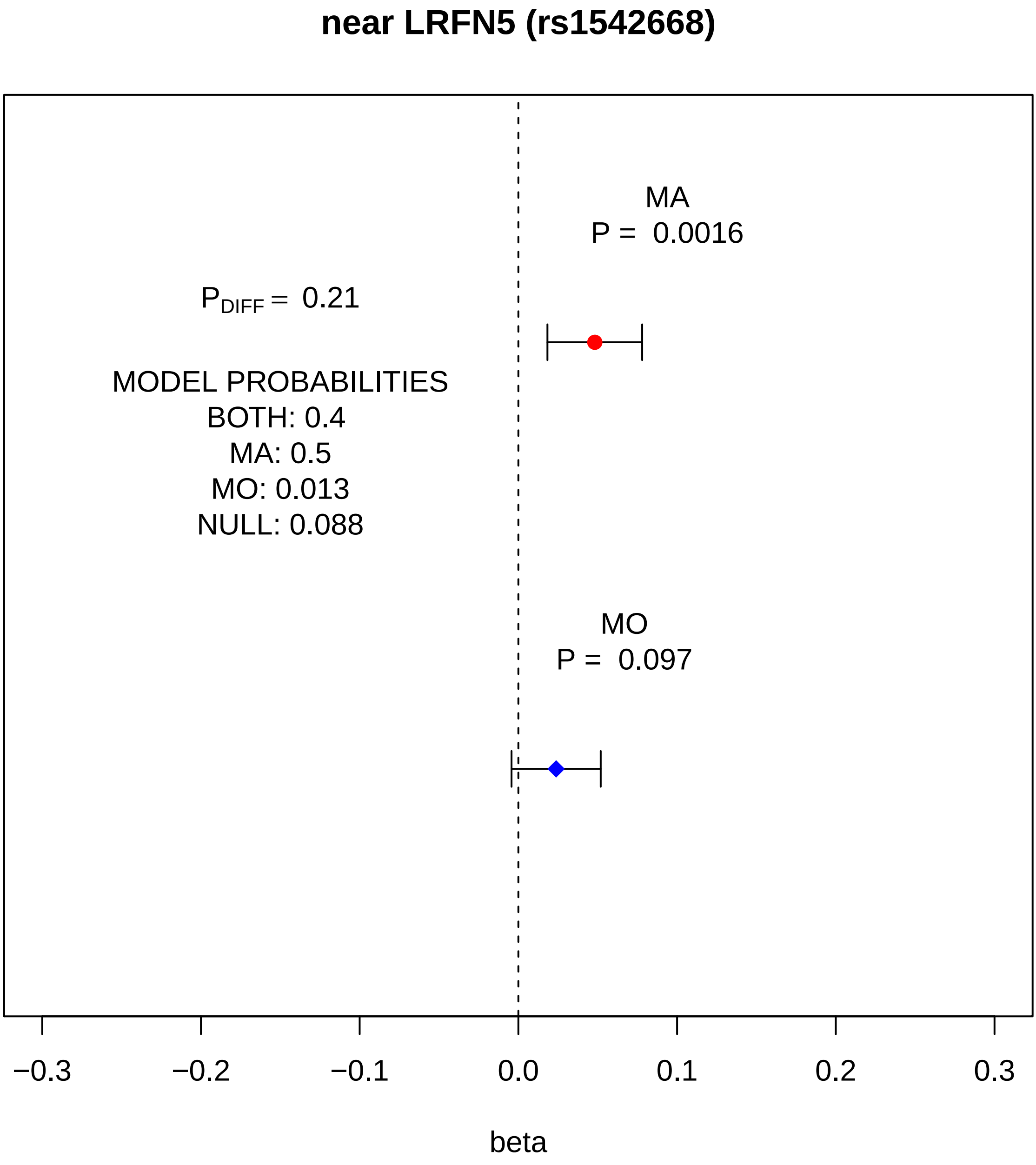

near LRFN5 (rs1542668)
MA
P = 0.0016
P_DIFF = 0.21
MODEL PROBABILITIES
BOTH: 0.4
MA: 0.5
MO: 0.013
NULL: 0.088
MO
P = 0.097
−0.3
−0.2
−0.1
0.0
0.1
0.2
0.3
beta

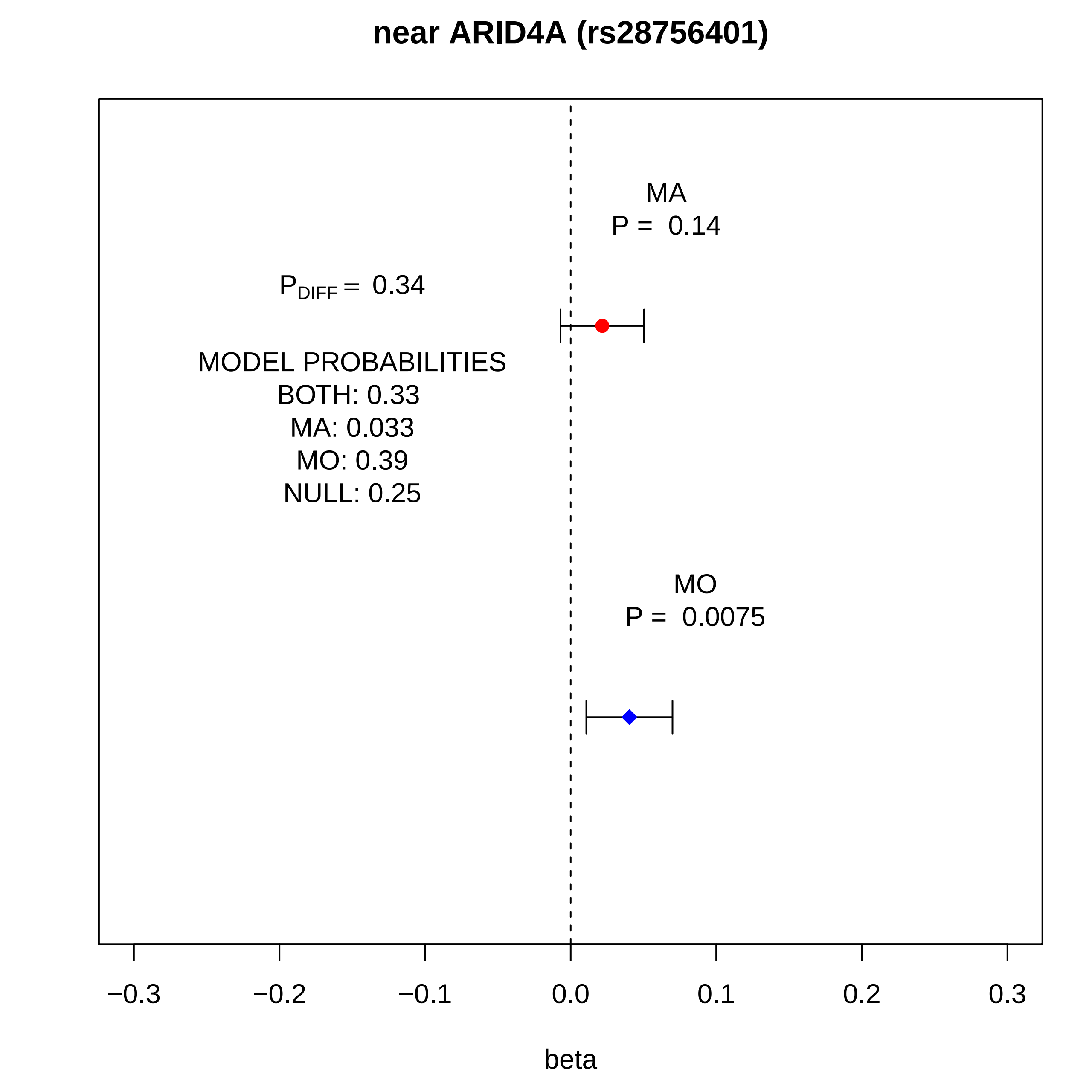
near ARID4A (rs28756401)
MA
P = 0.14
$P_{DIFF}$ = 0.34
MODEL PROBABILITIES
BOTH: 0.33
MA: 0.033
MO: 0.39
NULL: 0.25
MO
P = 0.0075
−0.3
−0.2
−0.1
0.0
0.1
0.2
0.3
beta

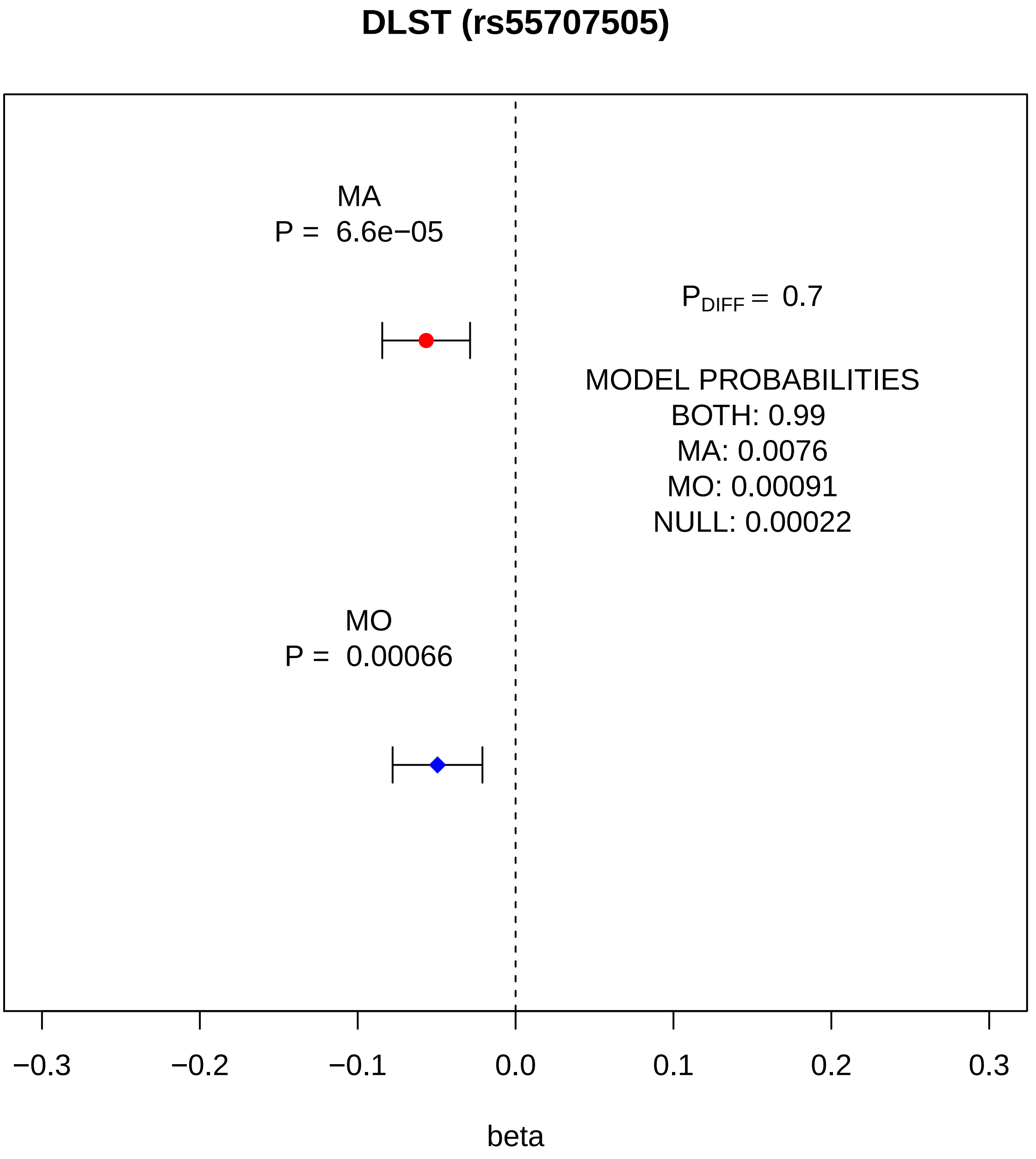
DLST (rs55707505)
MA
P = 6.6e−05
P_DIFF = 0.7
MODEL PROBABILITIES
BOTH: 0.99
MA: 0.0076
MO: 0.00091
NULL: 0.00022
MO
P = 0.00066
−0.3
−0.2
−0.1
0.0
0.1
0.2
0.3
beta

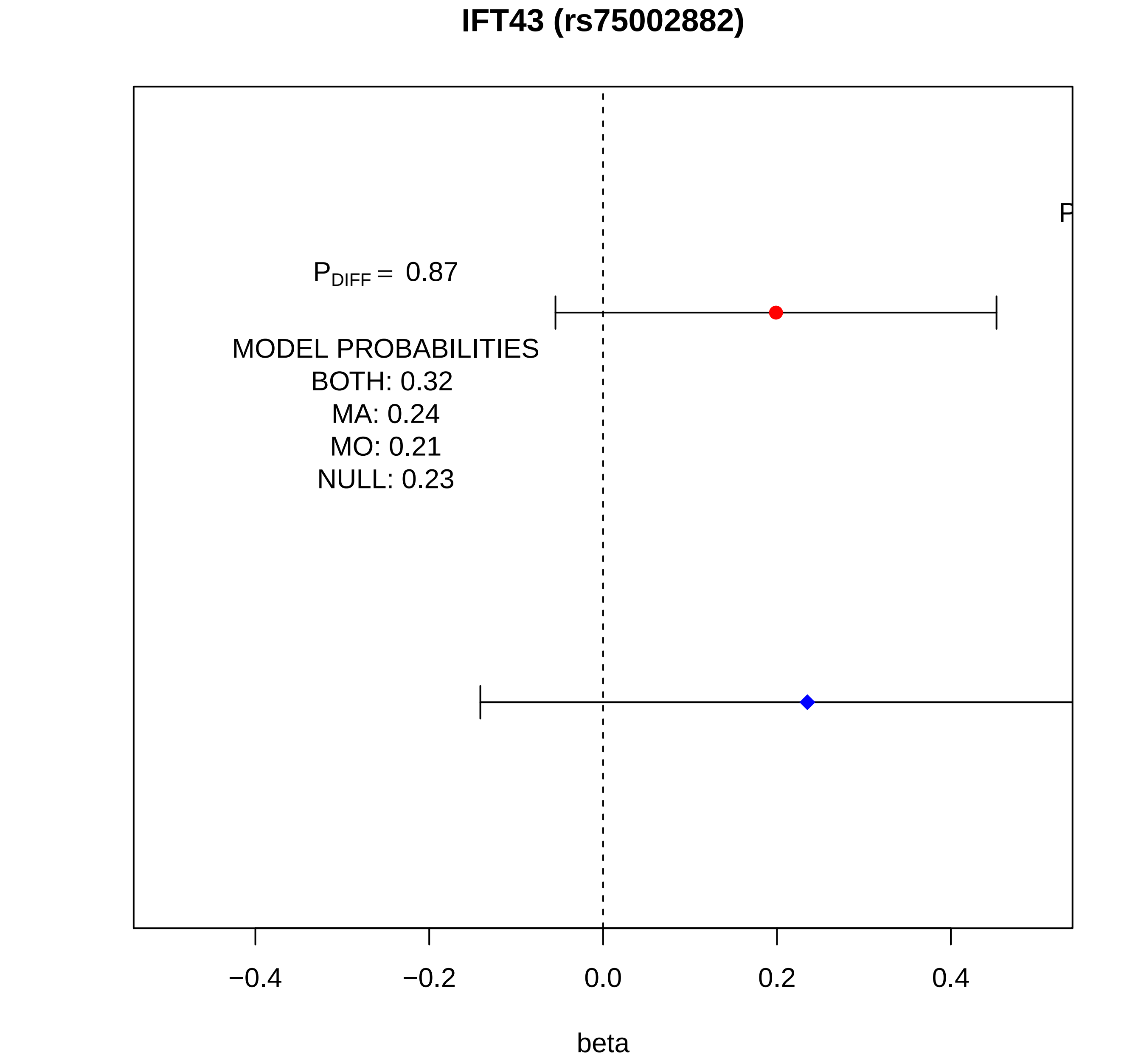
IFT43 (rs75002882)
P
$P_{DIFF}$ = 0.87
MODEL PROBABILITIES
BOTH: 0.32
MA: 0.24
MO: 0.21
NULL: 0.23
−0.4
−0.2
0.0
0.2
0.4
beta

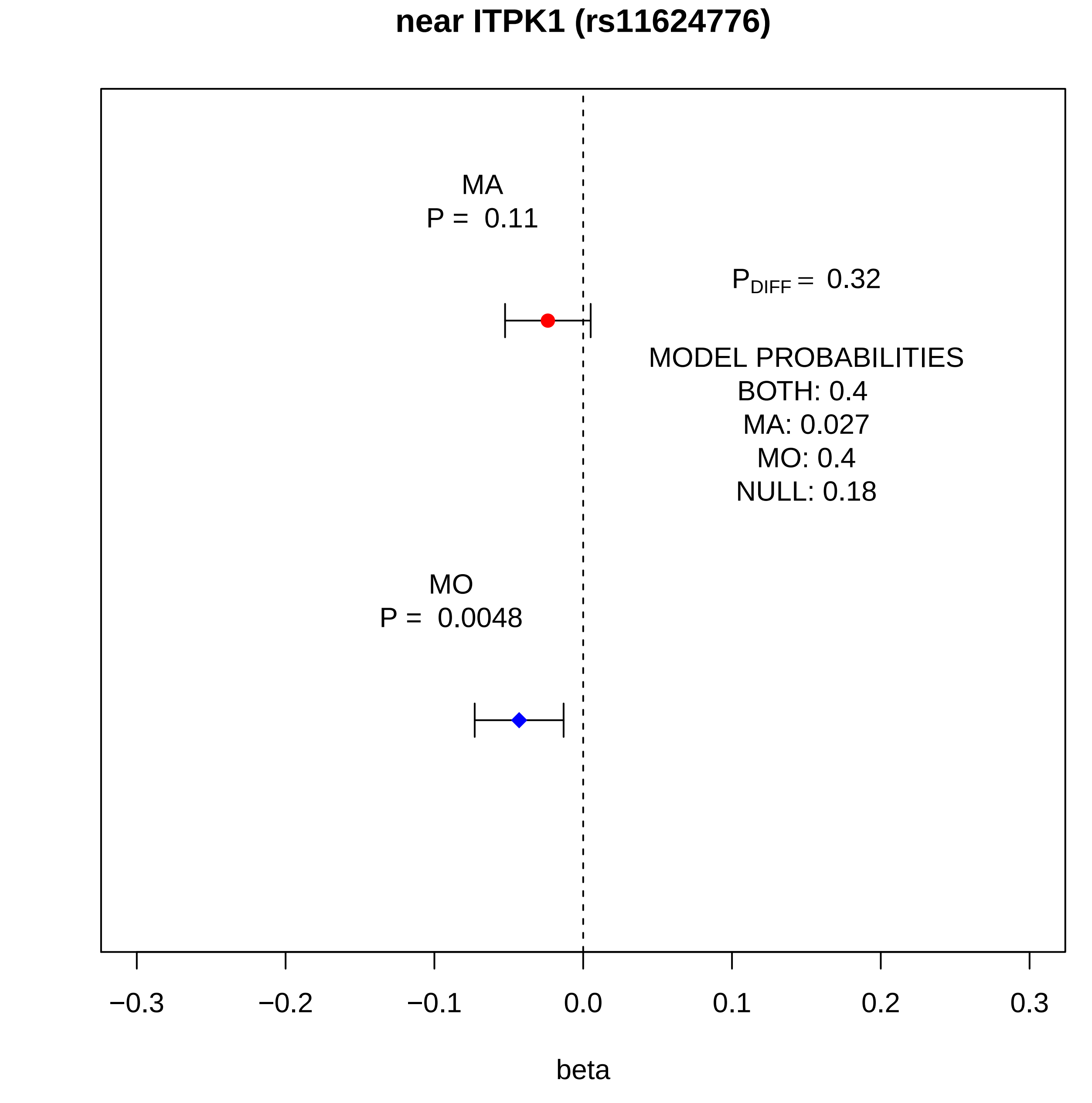
near ITPK1 (rs11624776)
MA
P = 0.11
P_DIFF = 0.32
MODEL PROBABILITIES
BOTH: 0.4
MA: 0.027
MO: 0.4
NULL: 0.18
MO
P = 0.0048
−0.3
−0.2
−0.1
0.0
0.1
0.2
0.3
beta

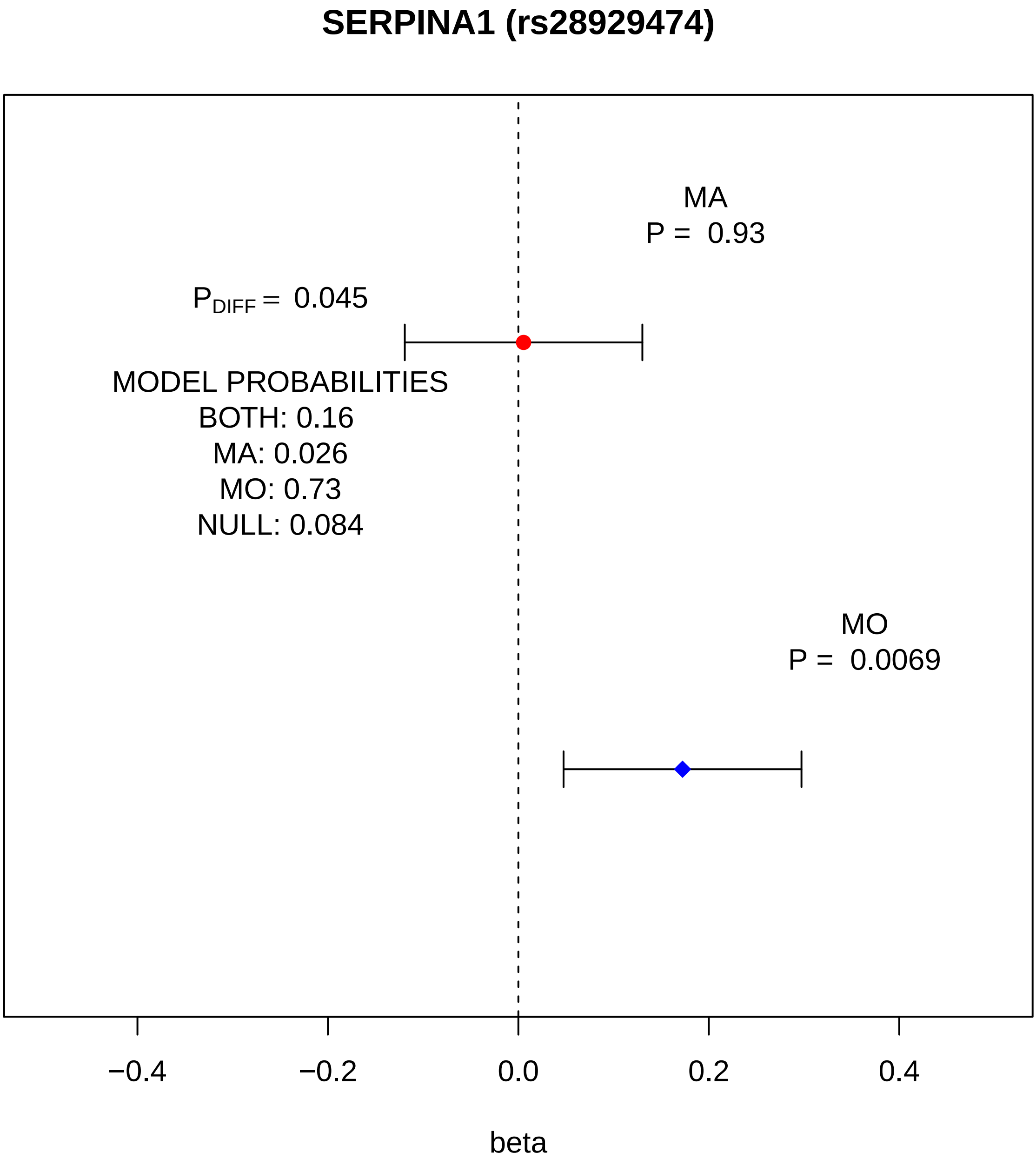
SERPINA1 (rs28929474)
MA
P = 0.93
P_DIFF = 0.045
MODEL PROBABILITIES
BOTH: 0.16
MA: 0.026
MO: 0.73
NULL: 0.084
MO
P = 0.0069
−0.4
−0.2
0.0
0.2
0.4
beta

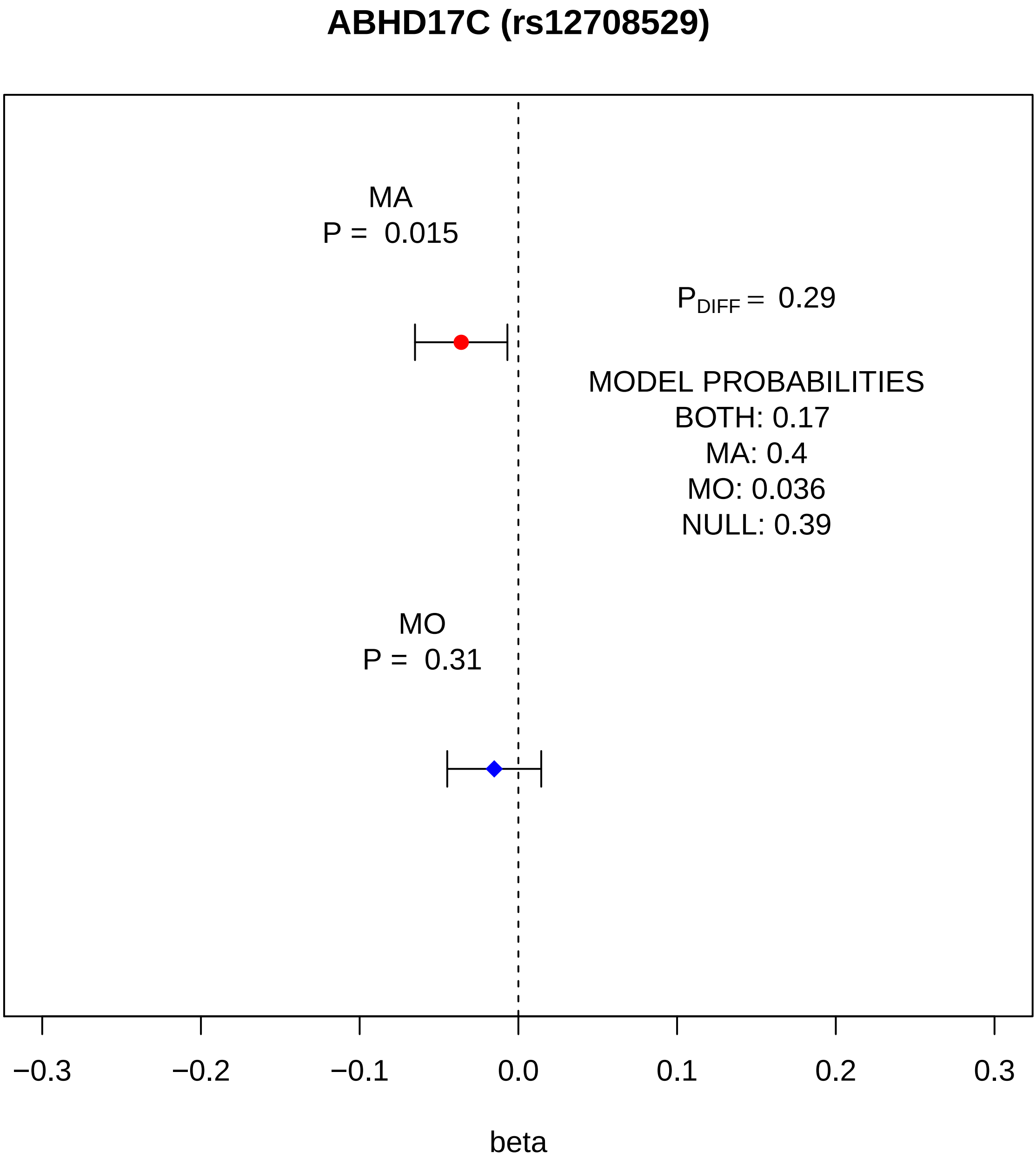
ABHD17C (rs12708529)
MA
P = 0.015
$P_{DIFF}$ = 0.29
MODEL PROBABILITIES
BOTH: 0.17
MA: 0.4
MO: 0.036
NULL: 0.39
MO
P = 0.31
−0.3
−0.2
−0.1
0.0
0.1
0.2
0.3
beta

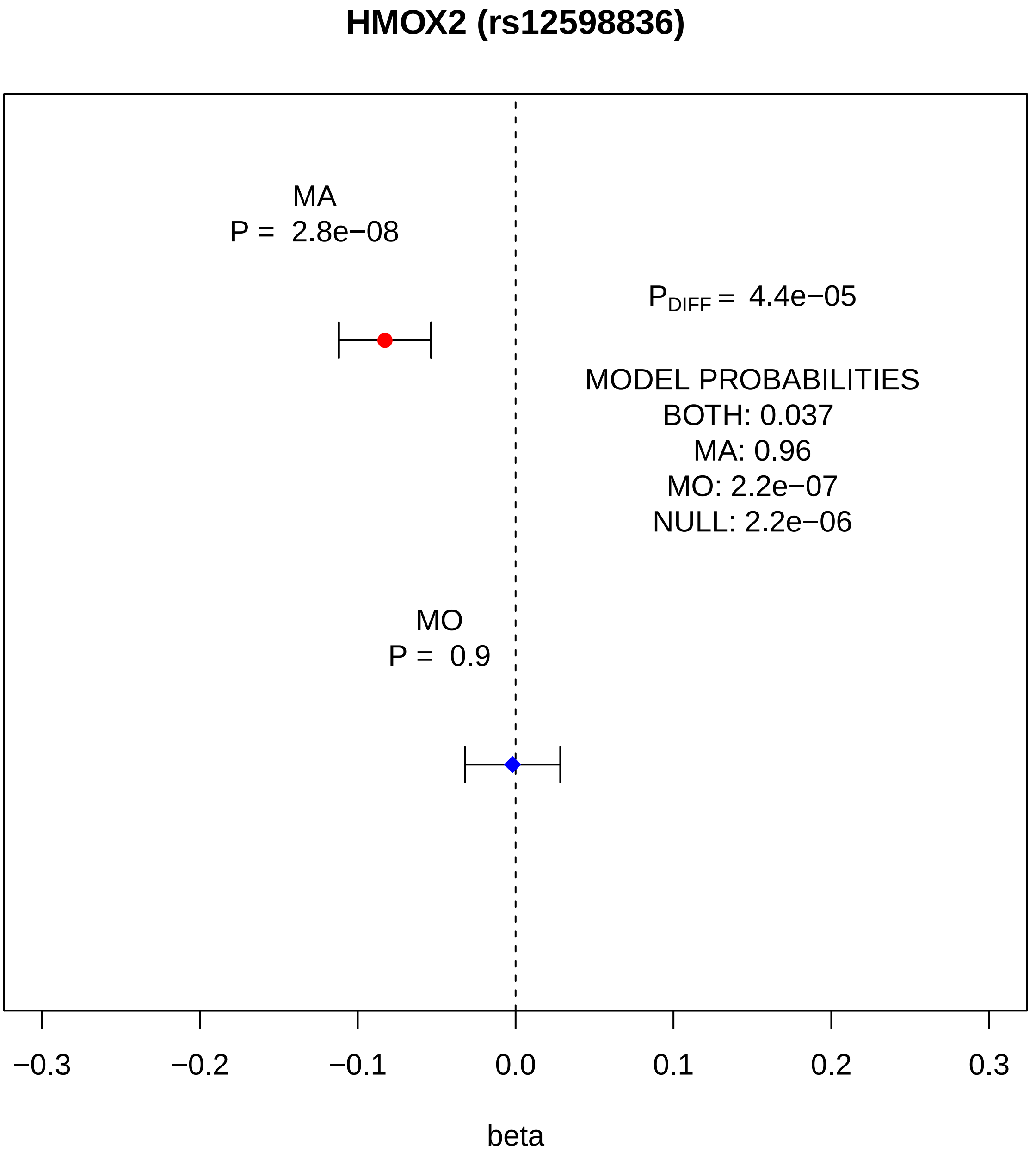
HMOX2 (rs12598836)
MA
P = 2.8e−08
PDIFF = 4.4e−05
MODEL PROBABILITIES
BOTH: 0.037
MA: 0.96
MO: 2.2e−07
NULL: 2.2e−06
MO
P = 0.9
−0.3
−0.2
−0.1
0.0
0.1
0.2
0.3
beta

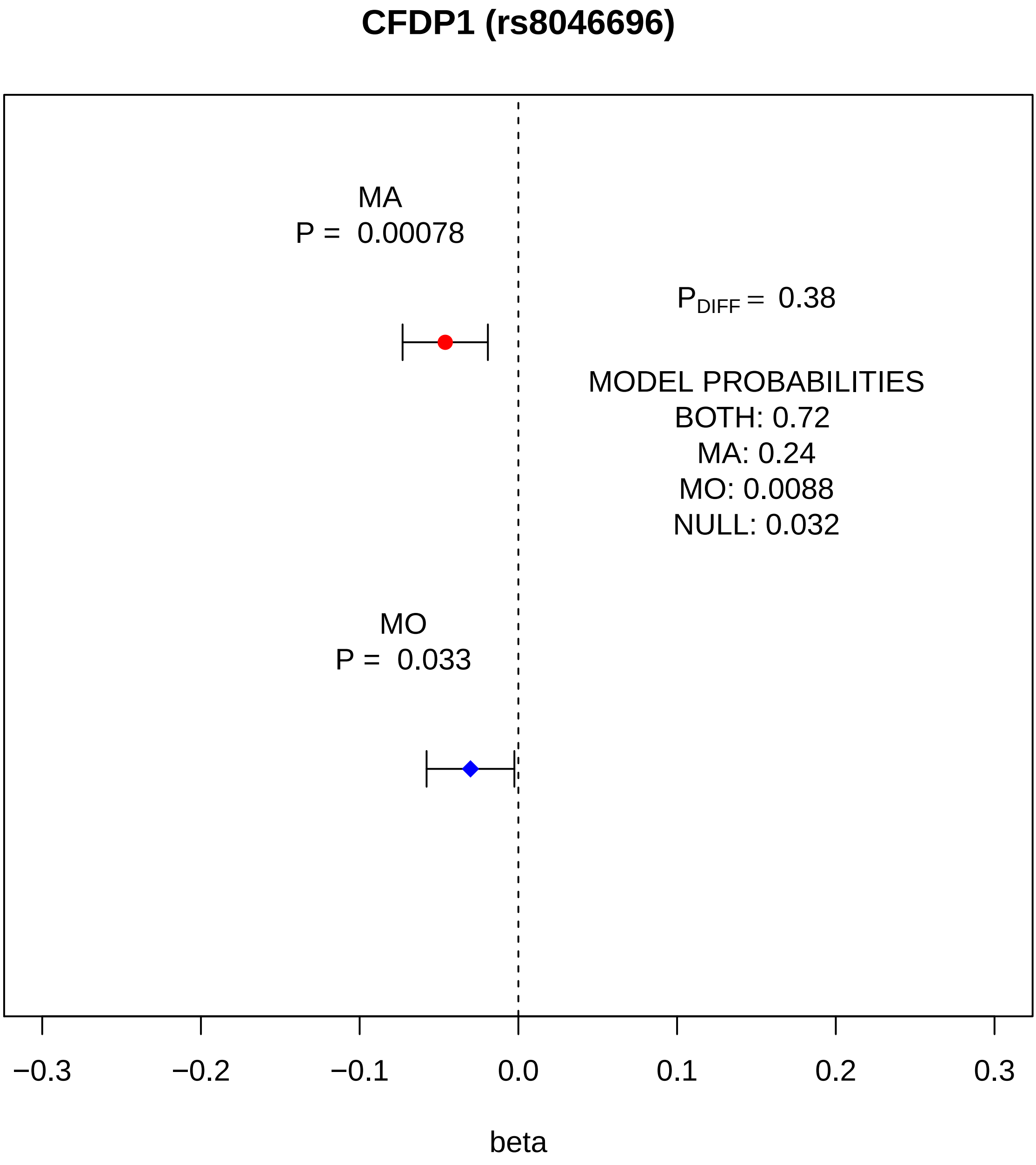
CFDP1 (rs8046696)
MA
P = 0.00078
$P_{DIFF}$ = 0.38
MODEL PROBABILITIES
BOTH: 0.72
MA: 0.24
MO: 0.0088
NULL: 0.032
MO
P = 0.033
−0.3
−0.2
−0.1
0.0
0.1
0.2
0.3
beta

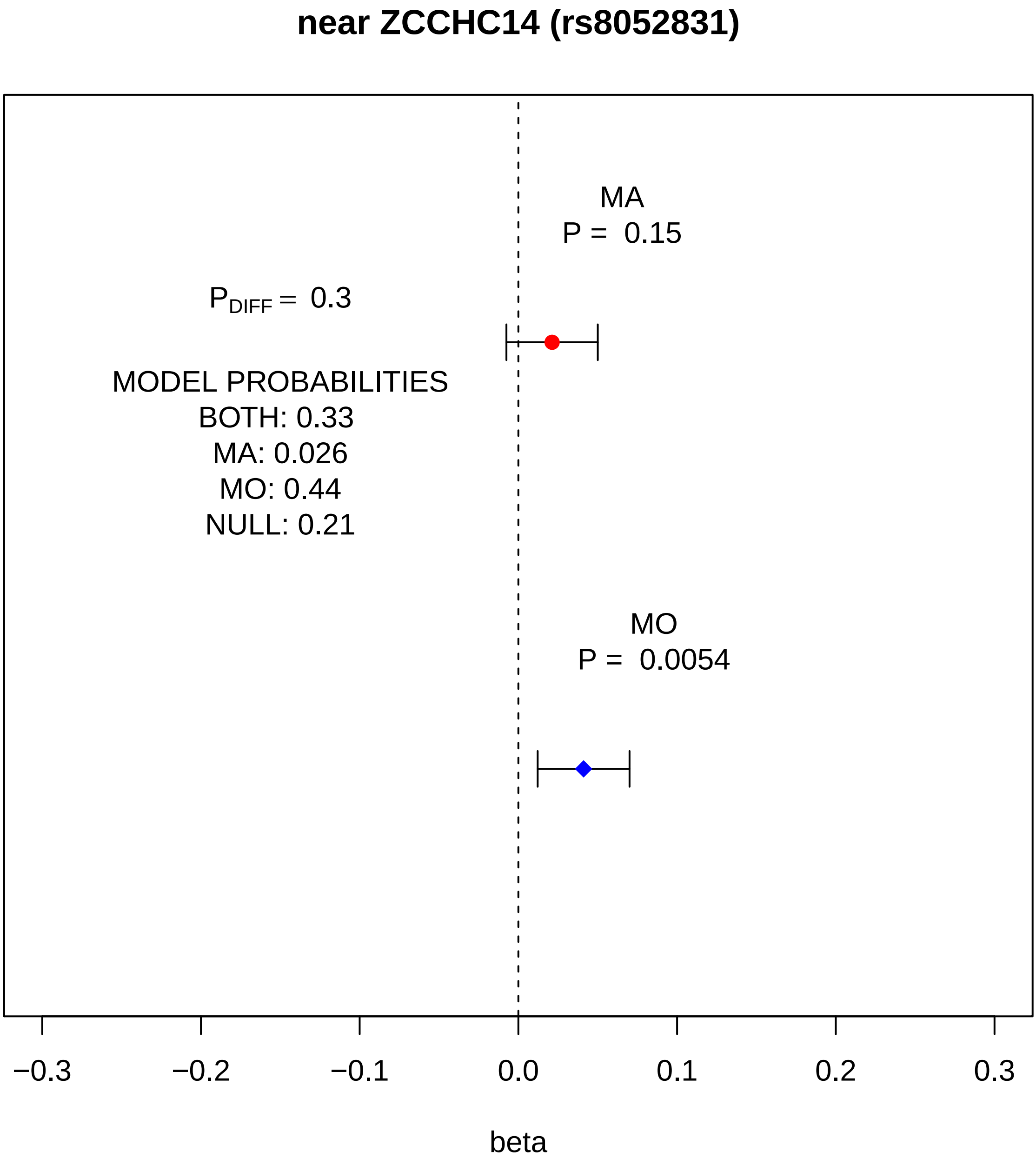
near ZCCHC14 (rs8052831)
MA
P = 0.15
$P_{DIFF}$ = 0.3
MODEL PROBABILITIES
BOTH: 0.33
MA: 0.026
MO: 0.44
NULL: 0.21
MO
P = 0.0054
−0.3
−0.2
−0.1
0.0
0.1
0.2
0.3
beta

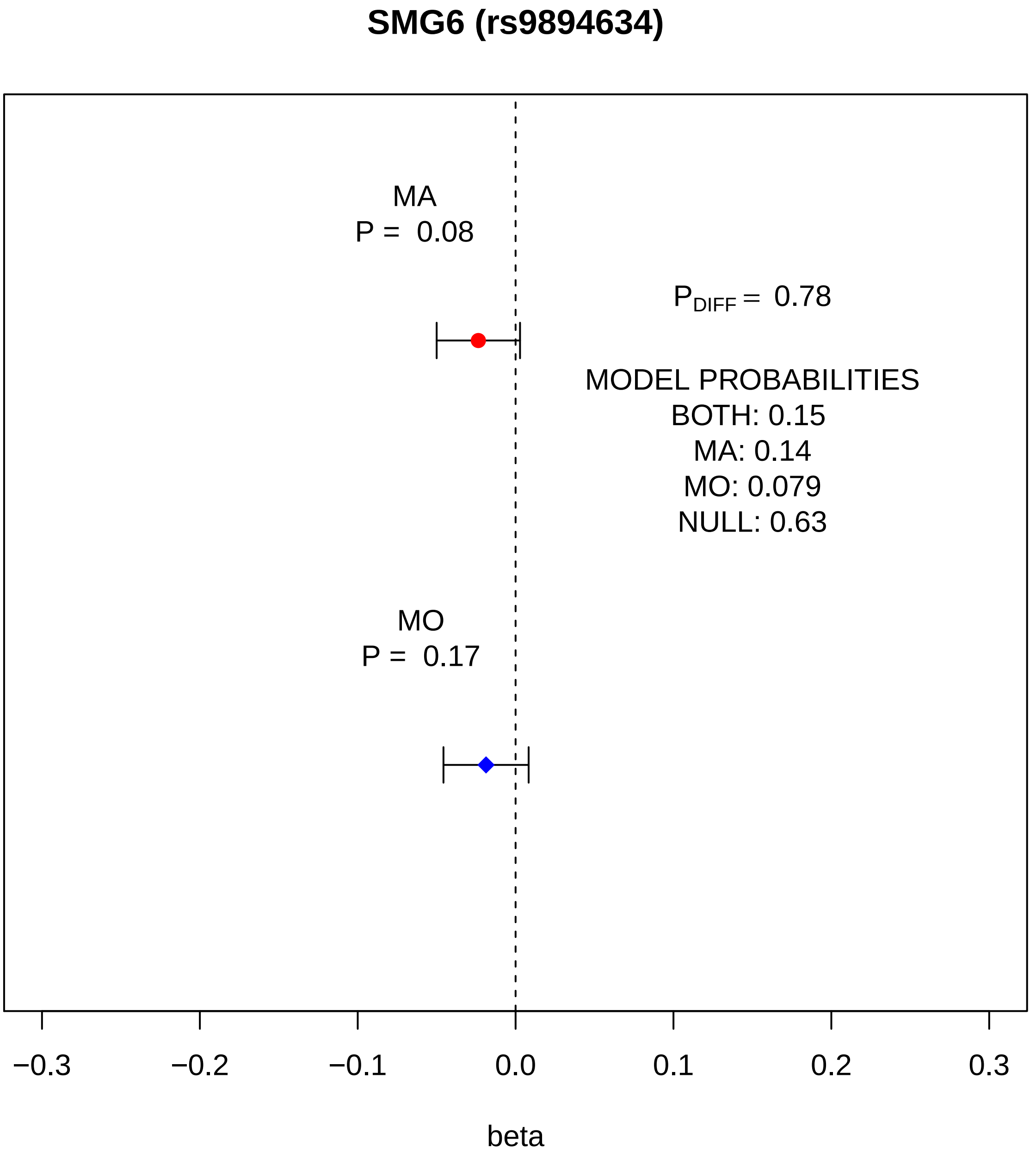
SMG6 (rs9894634)
MA
P = 0.08
P_DIFF = 0.78
MODEL PROBABILITIES
BOTH: 0.15
MA: 0.14
MO: 0.079
NULL: 0.63
MO
P = 0.17
−0.3
−0.2
−0.1
0.0
0.1
0.2
0.3
beta

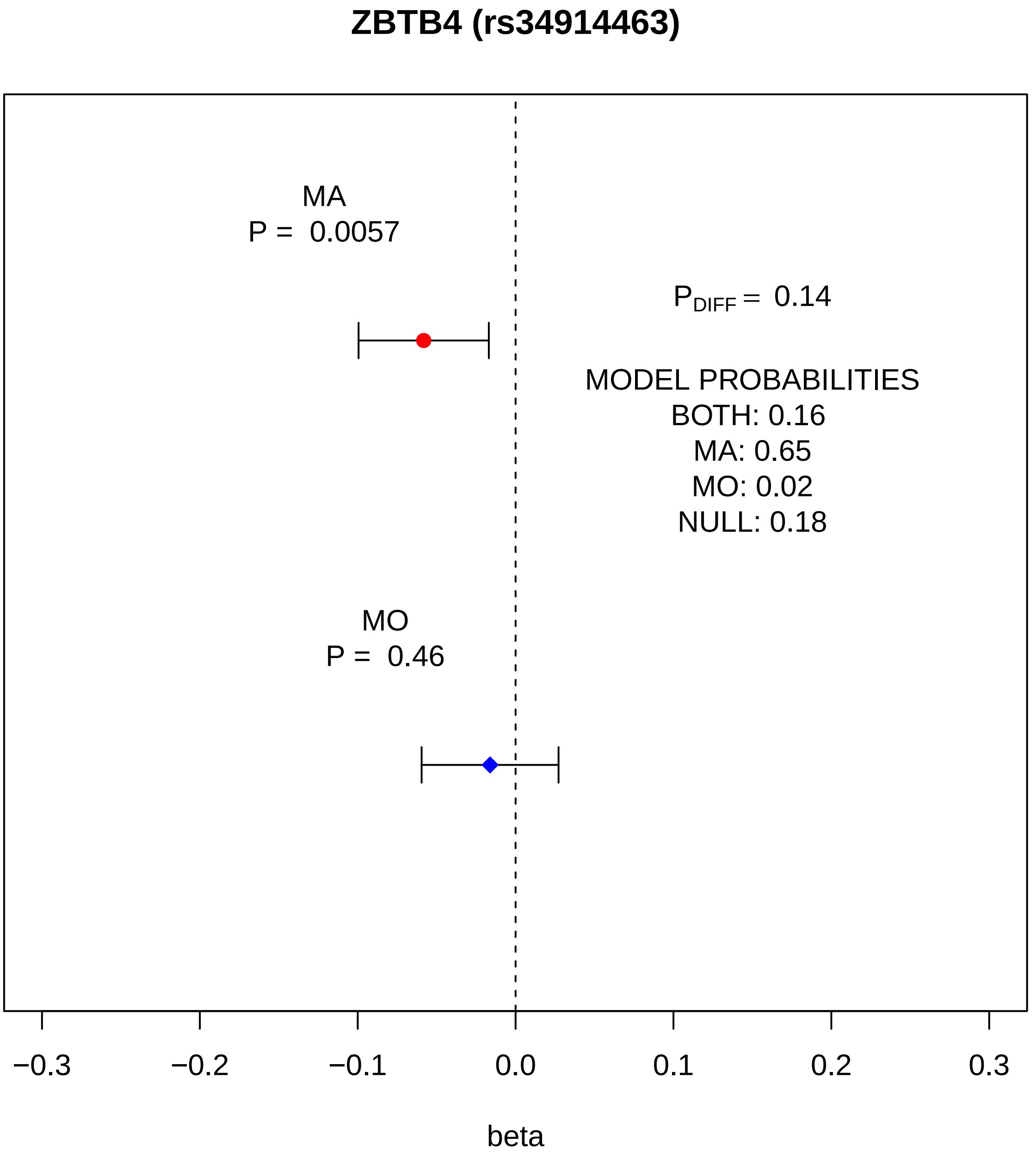
ZBTB4 (rs34914463)
MA
P = 0.0057
$P_{DIFF}$ = 0.14
MODEL PROBABILITIES
BOTH: 0.16
MA: 0.65
MO: 0.02
NULL: 0.18
MO
P = 0.46
−0.3
−0.2
−0.1
0.0
0.1
0.2
0.3
beta

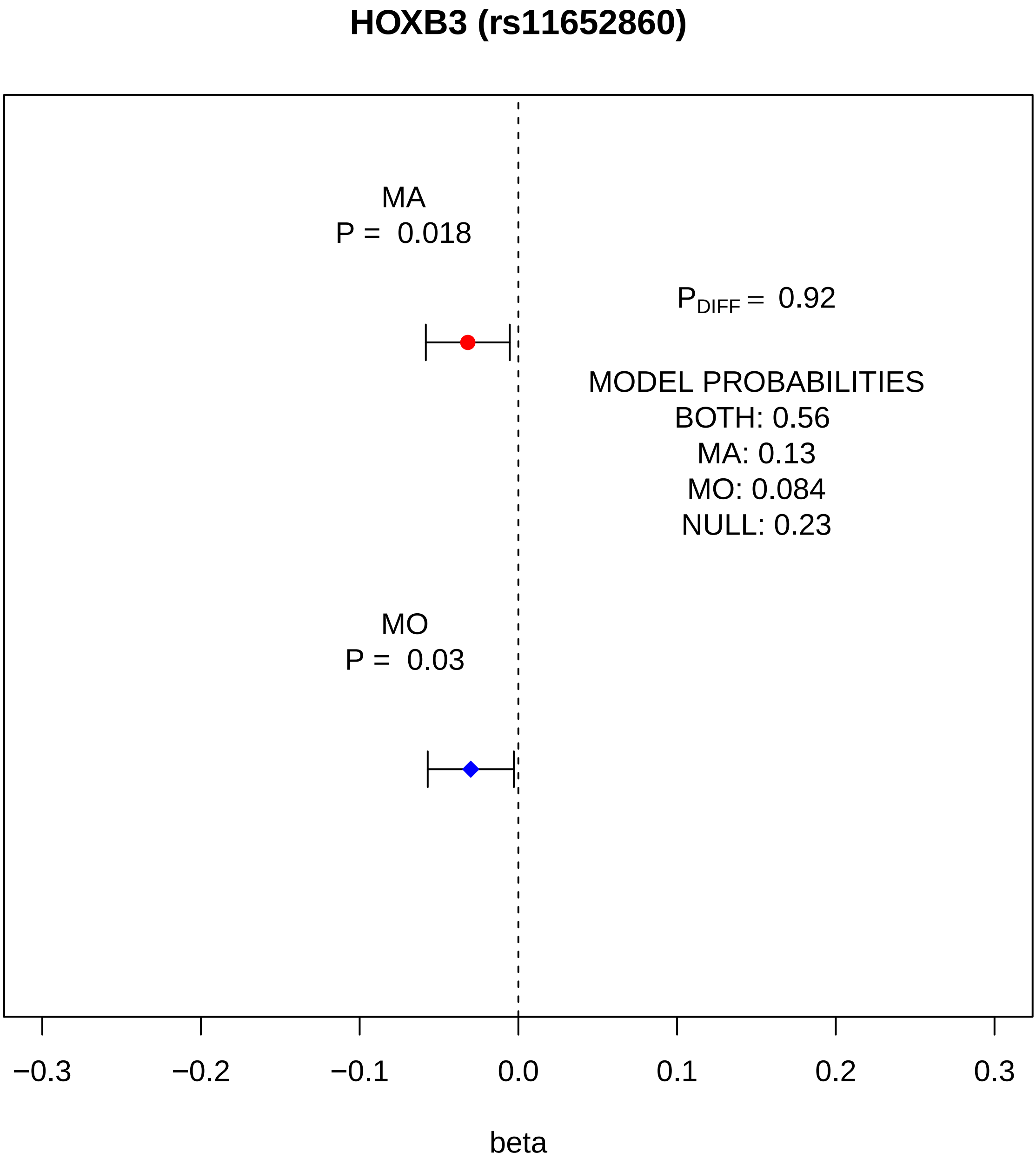
HOXB3 (rs11652860)
MA
P = 0.018
$P_{DIFF}$ = 0.92
MODEL PROBABILITIES
BOTH: 0.56
MA: 0.13
MO: 0.084
NULL: 0.23
MO
P = 0.03
−0.3
−0.2
−0.1
0.0
0.1
0.2
0.3
beta

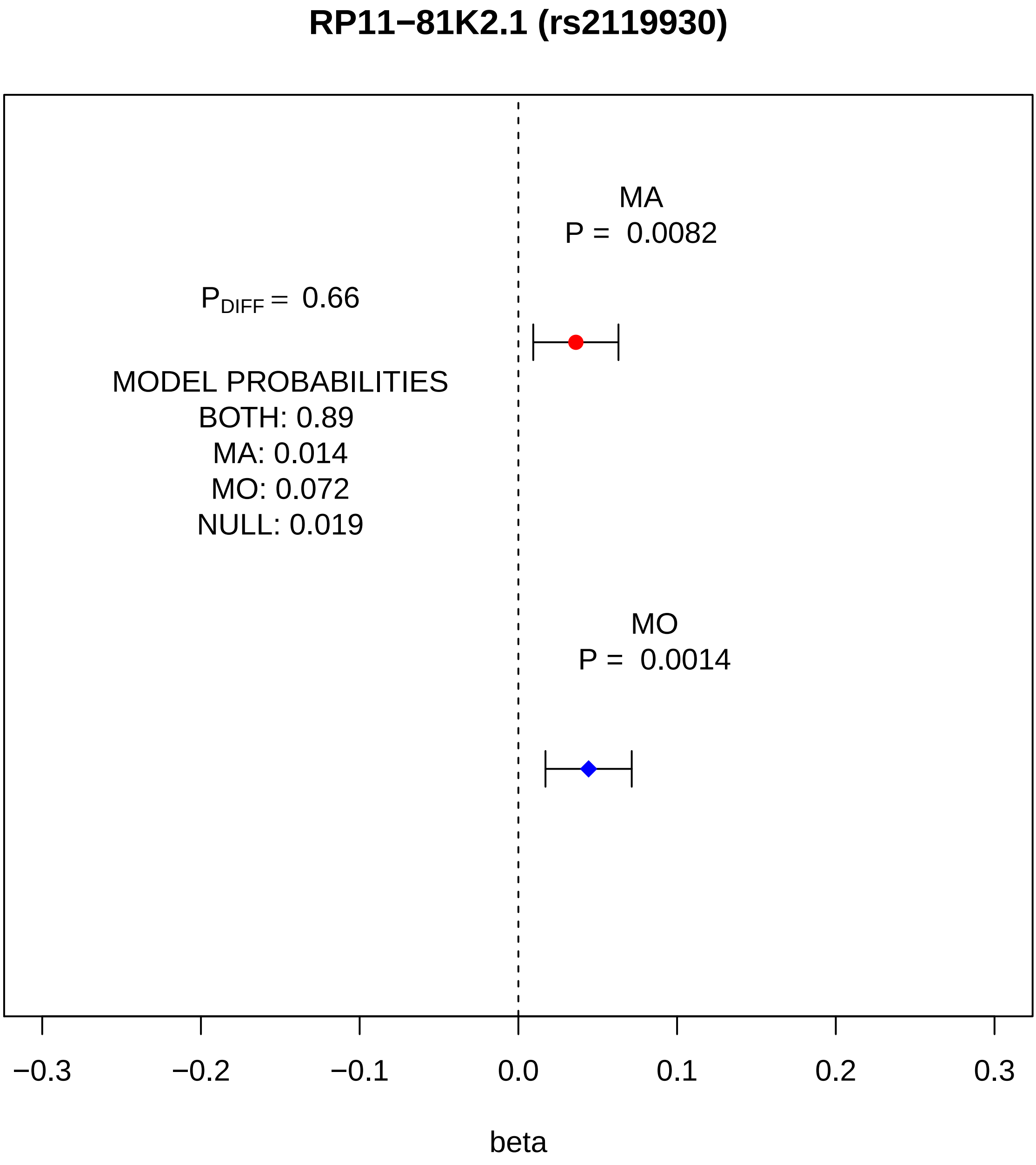
RP11−81K2.1 (rs2119930)
MA
P = 0.0082
$P_{DIFF}$ = 0.66
MODEL PROBABILITIES
BOTH: 0.89
MA: 0.014
MO: 0.072
NULL: 0.019
MO
P = 0.0014
−0.3
−0.2
−0.1
0.0
0.1
0.2
0.3
beta

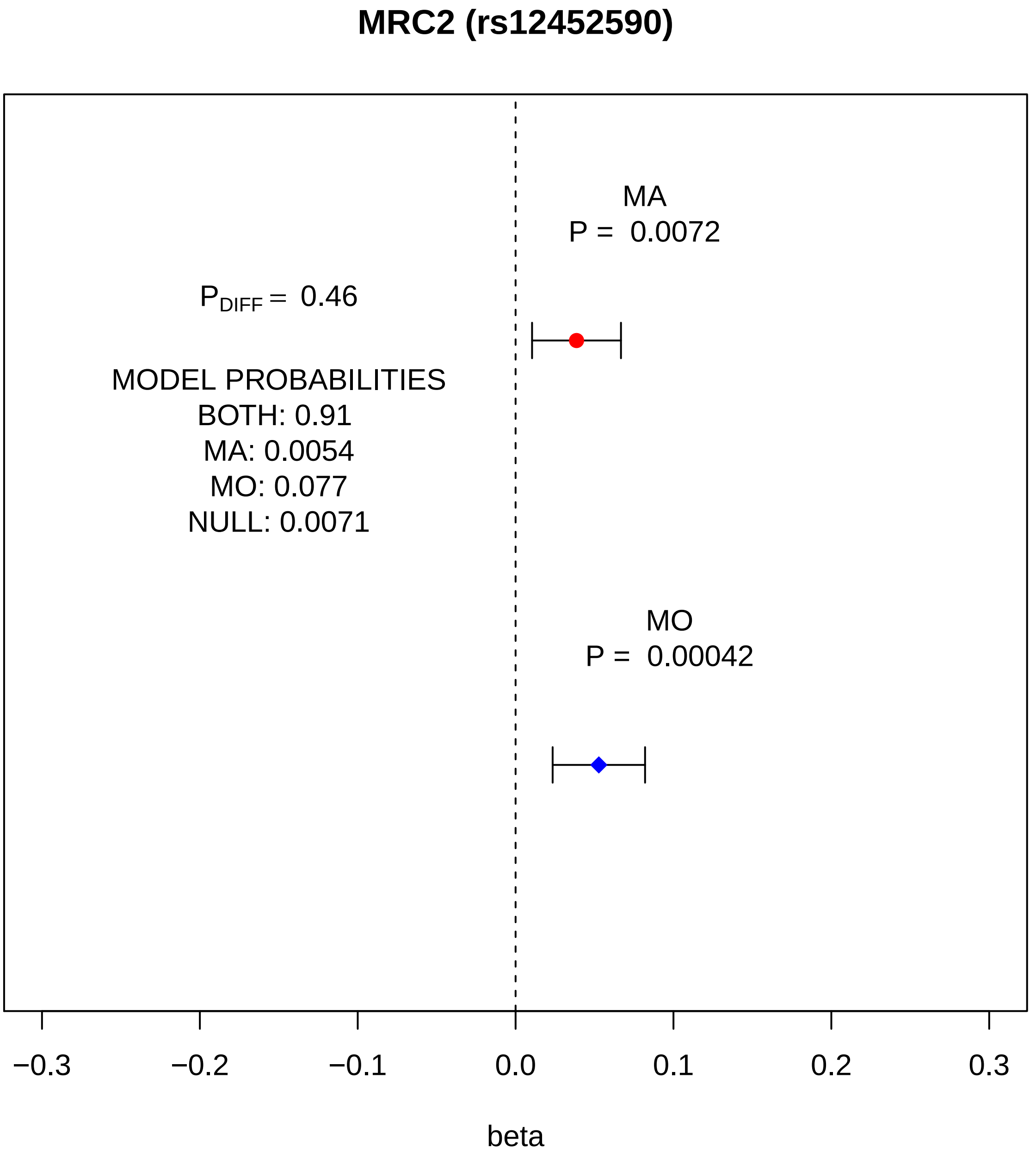
MRC2 (rs12452590)
MA
P = 0.0072
P DIFF = 0.46
MODEL PROBABILITIES
BOTH: 0.91
MA: 0.0054
MO: 0.077
NULL: 0.0071
MO
P = 0.00042
−0.3
−0.2
−0.1
0.0
0.1
0.2
0.3
beta

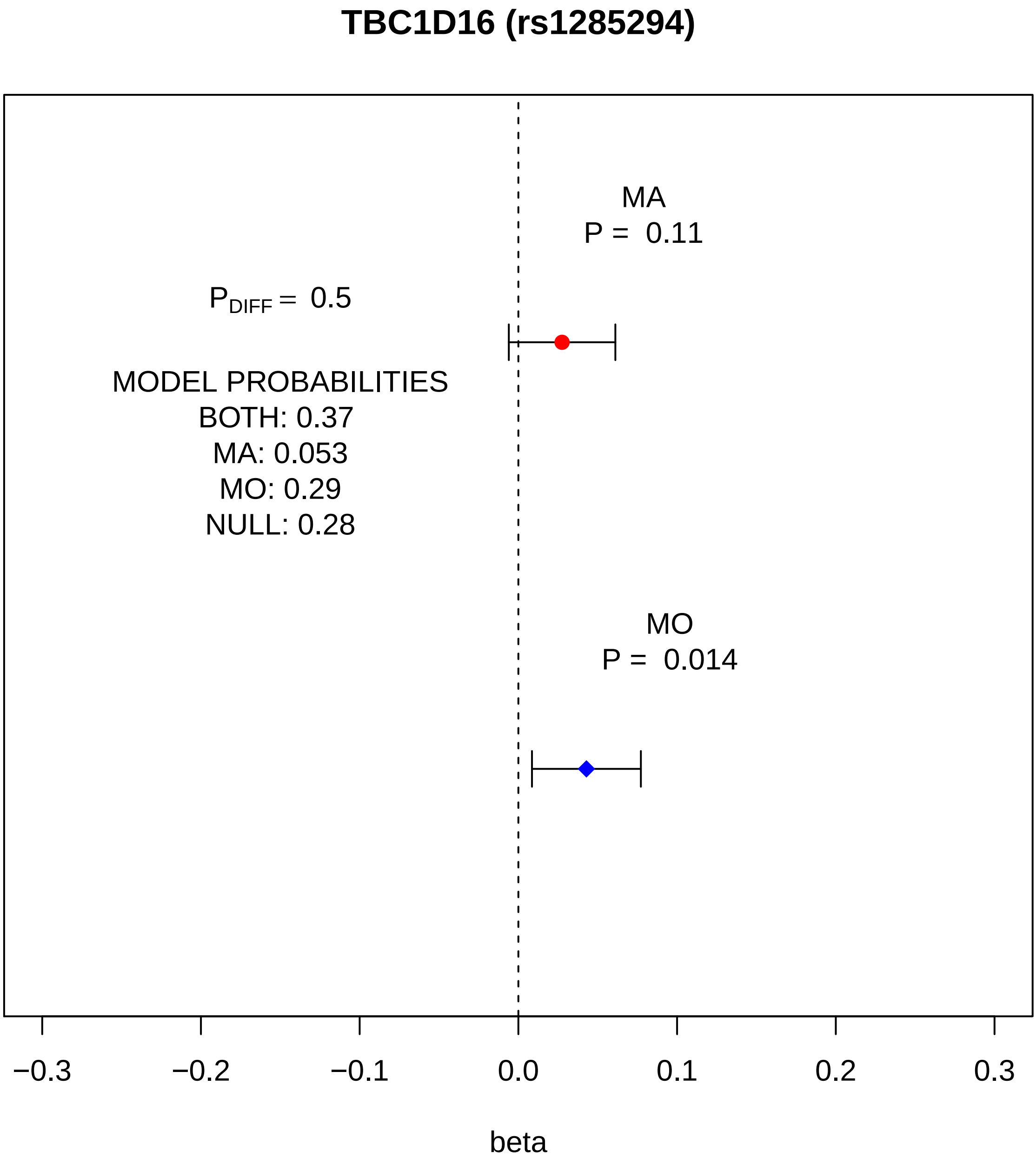

TBC1D16 (rs1285294)
MA
P = 0.11
P_DIFF = 0.5
MODEL PROBABILITIES
BOTH: 0.37
MA: 0.053
MO: 0.29
NULL: 0.28
MO
P = 0.014
−0.3
−0.2
−0.1
0.0
0.1
0.2
0.3
beta

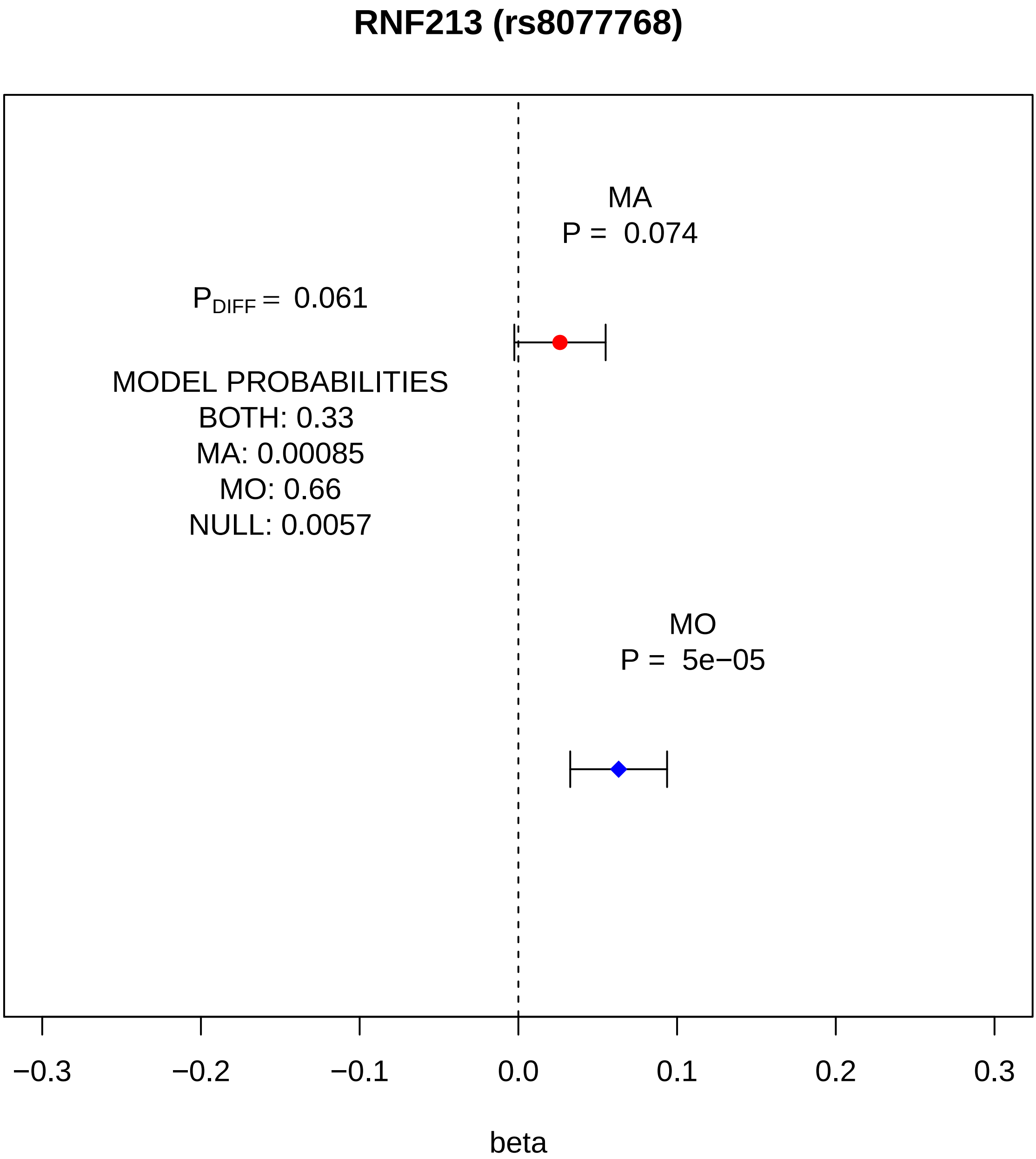
RNF213 (rs8077768)
MA
P = 0.074
P_DIFF = 0.061
MODEL PROBABILITIES
BOTH: 0.33
MA: 0.00085
MO: 0.66
NULL: 0.0057
MO
P = 5e−05
−0.3
−0.2
−0.1
0.0
0.1
0.2
0.3
beta

#### near RBBP8 (rs7506921)

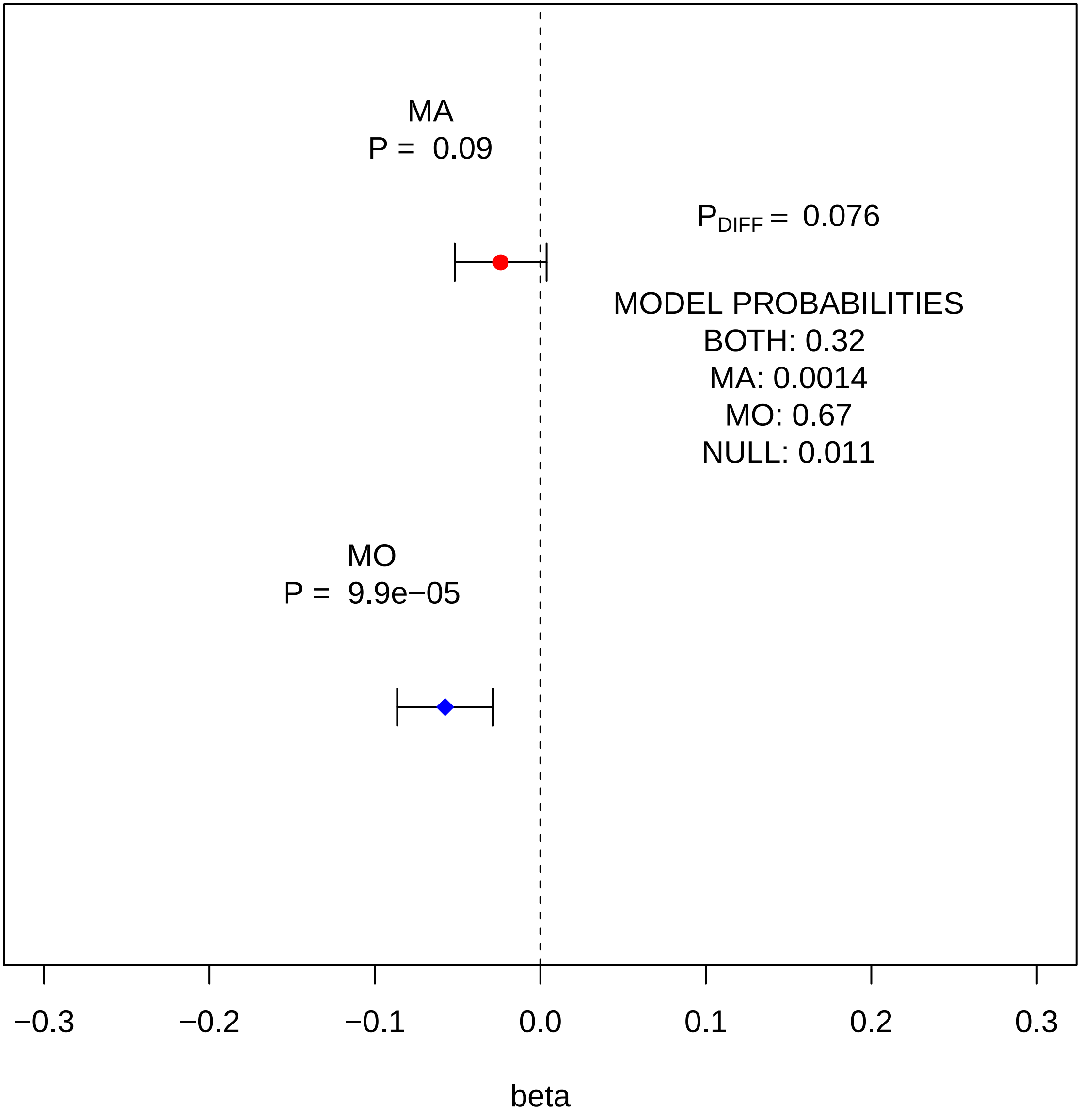

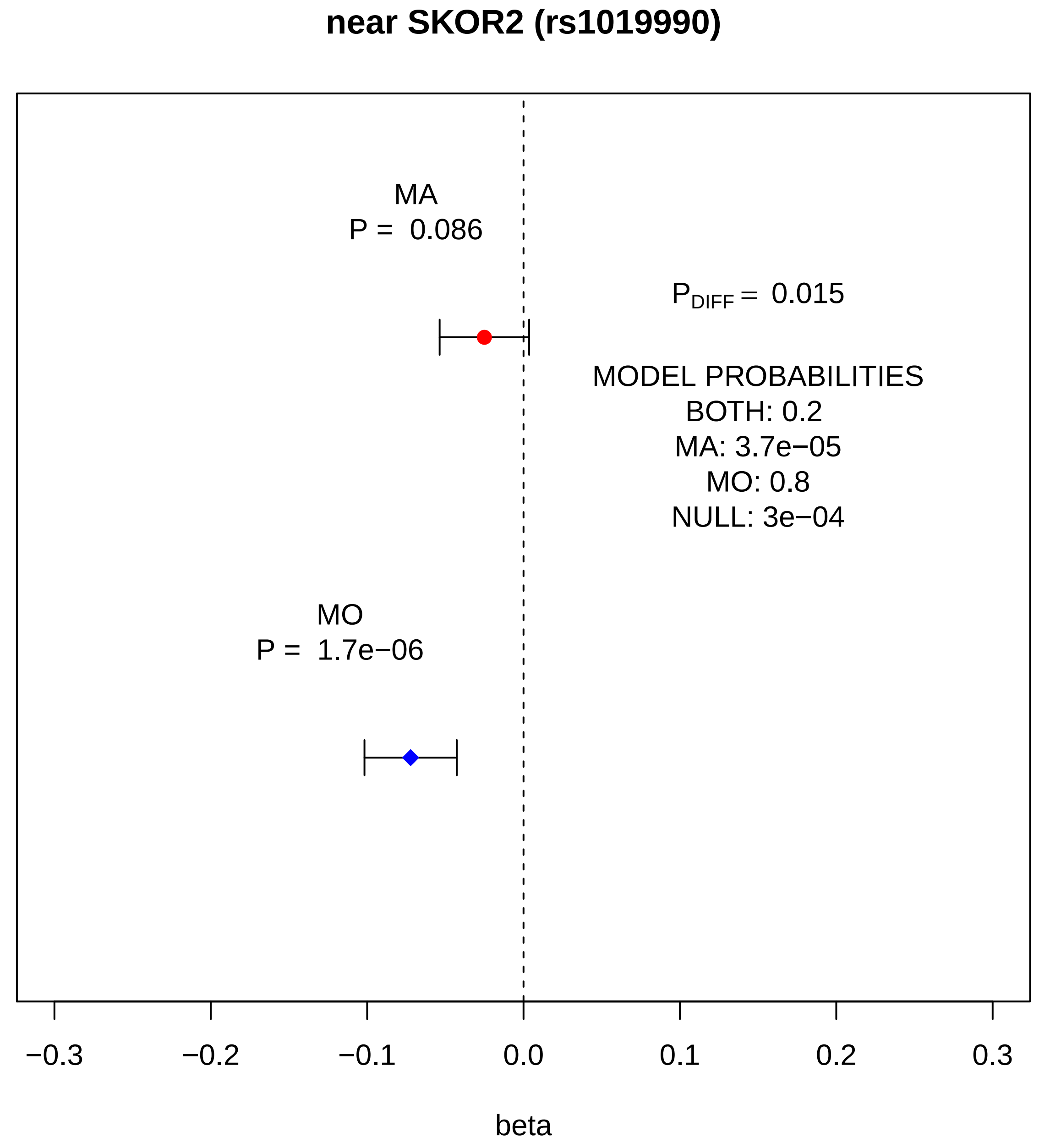

near SKOR2 (rs1019990)
MA
P = 0.086
P_DIFF = 0.015
MODEL PROBABILITIES
BOTH: 0.2
MA: 3.7e−05
MO: 0.8
NULL: 3e−04
MO
P = 1.7e−06
−0.3
−0.2
−0.1
0.0
0.1
0.2
0.3
beta

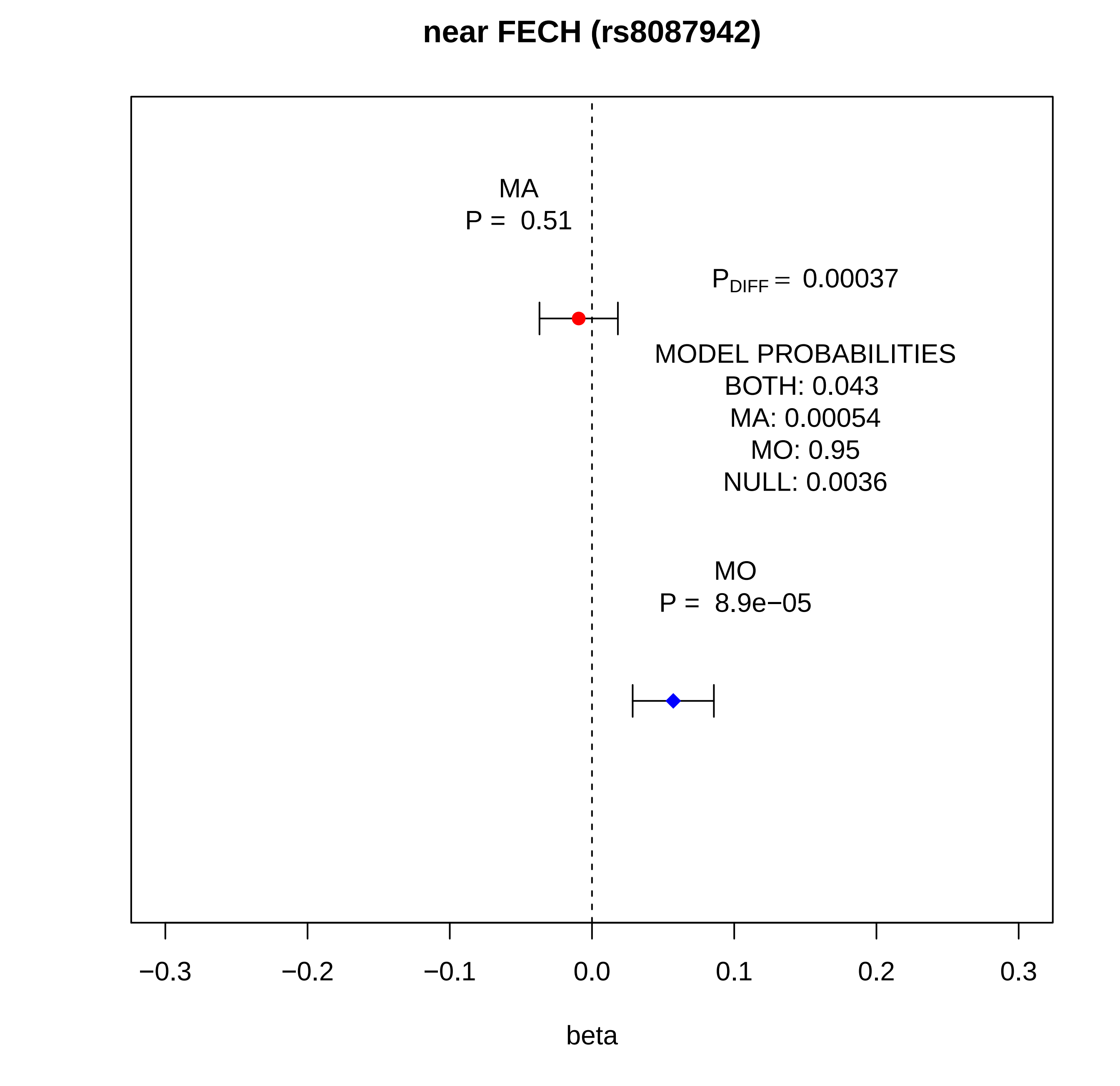

near FECH (rs8087942)
MA
P = 0.51
P_DIFF = 0.00037
MODEL PROBABILITIES
BOTH: 0.043
MA: 0.00054
MO: 0.95
NULL: 0.0036
MO
P = 8.9e−05
−0.3
−0.2
−0.1
0.0
0.1
0.2
0.3
beta

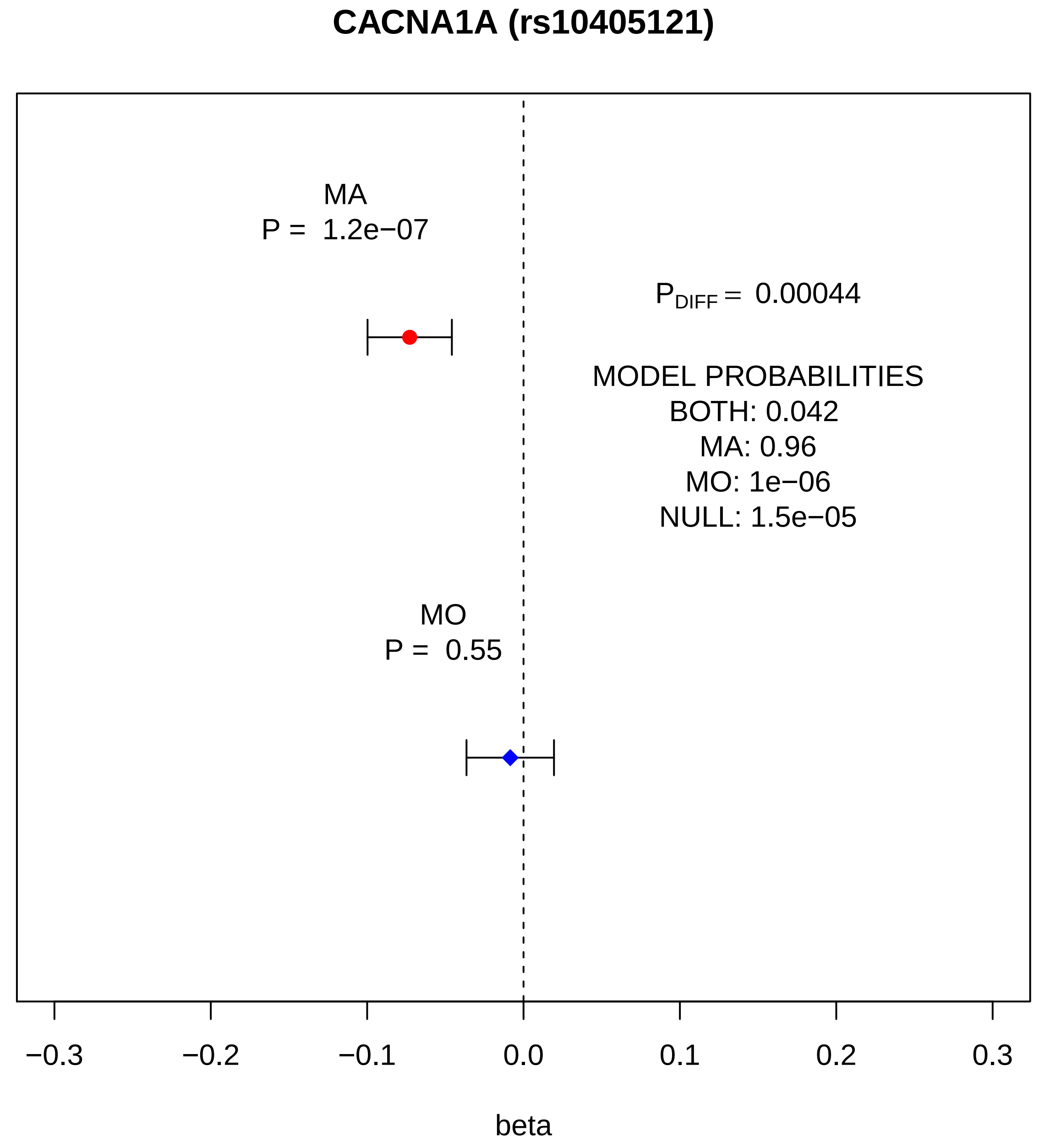
CACNA1A (rs10405121)
MA
P = 1.2e−07
P_DIFF = 0.00044
MODEL PROBABILITIES
BOTH: 0.042
MA: 0.96
MO: 1e−06
NULL: 1.5e−05
MO
P = 0.55
−0.3
−0.2
−0.1
0.0
0.1
0.2
0.3
beta

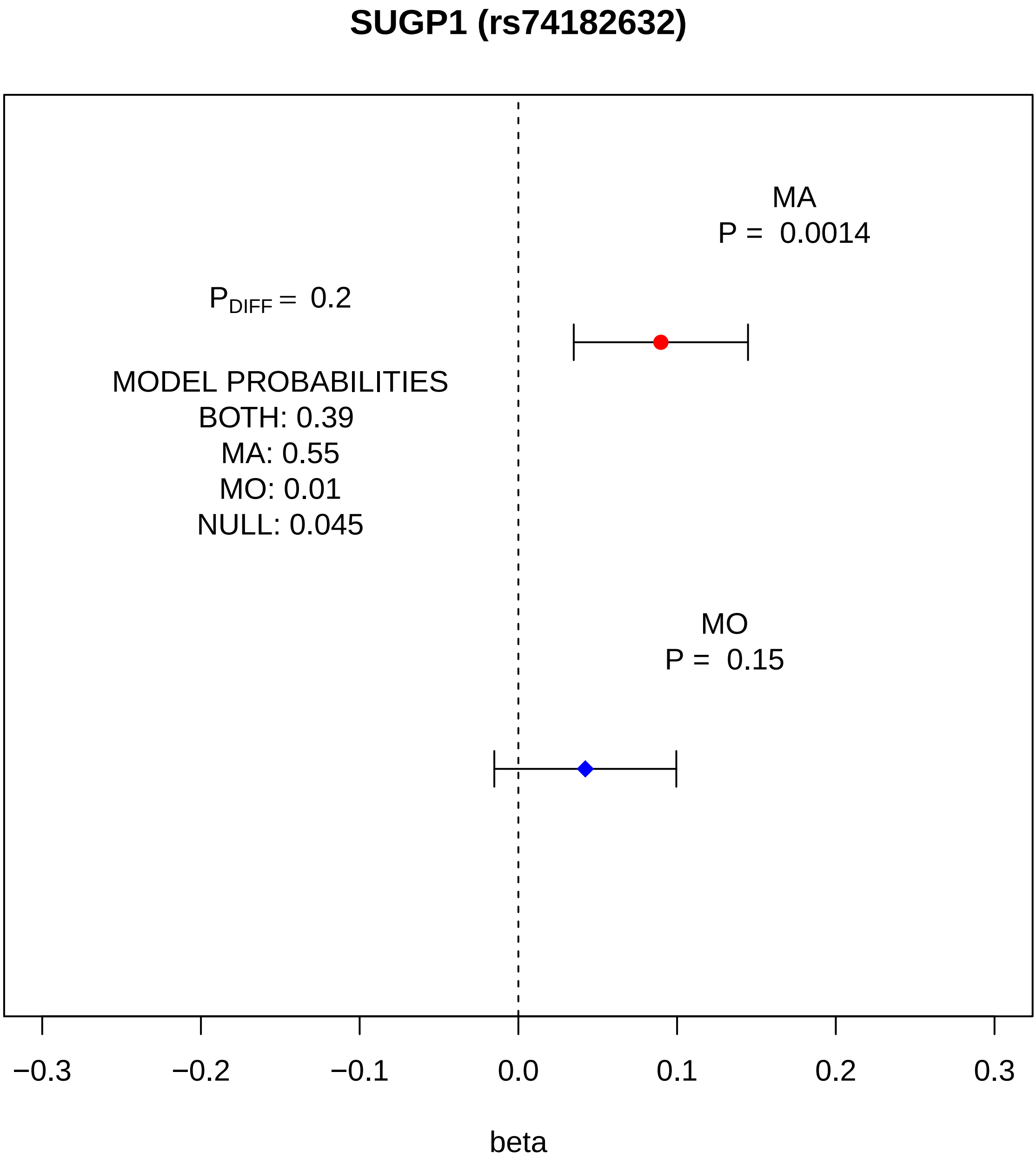
SUGP1 (rs74182632)
MA
P = 0.0014
$P_{DIFF}$ = 0.2
MODEL PROBABILITIES
BOTH: 0.39
MA: 0.55
MO: 0.01
NULL: 0.045
MO
P = 0.15
−0.3
−0.2
−0.1
0.0
0.1
0.2
0.3
beta

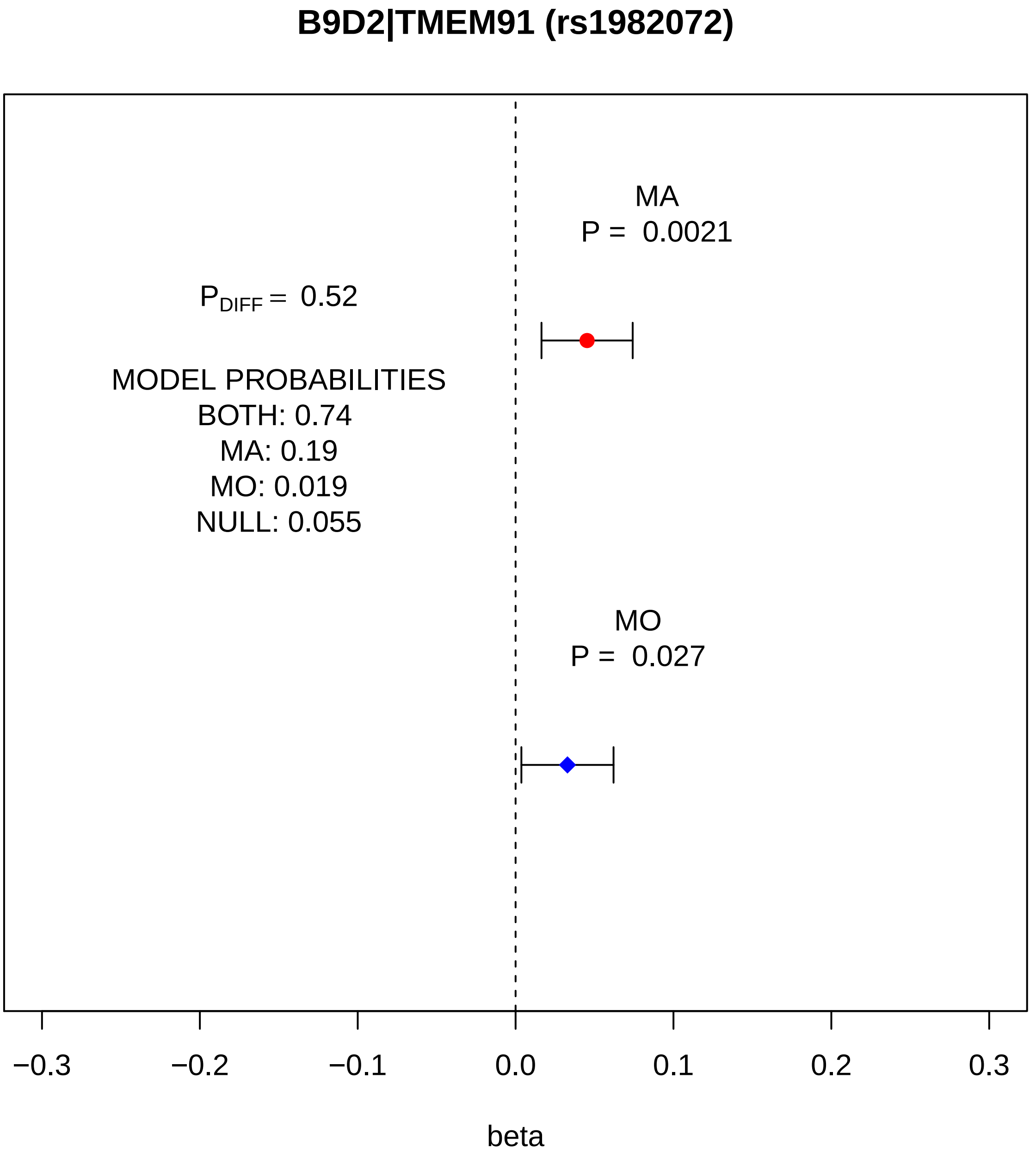
B9D2|TMEM91 (rs1982072)
MA
P = 0.0021
$P_{DIFF}$ = 0.52
MODEL PROBABILITIES
BOTH: 0.74
MA: 0.19
MO: 0.019
NULL: 0.055
MO
P = 0.027
−0.3
−0.2
−0.1
0.0
0.1
0.2
0.3
beta

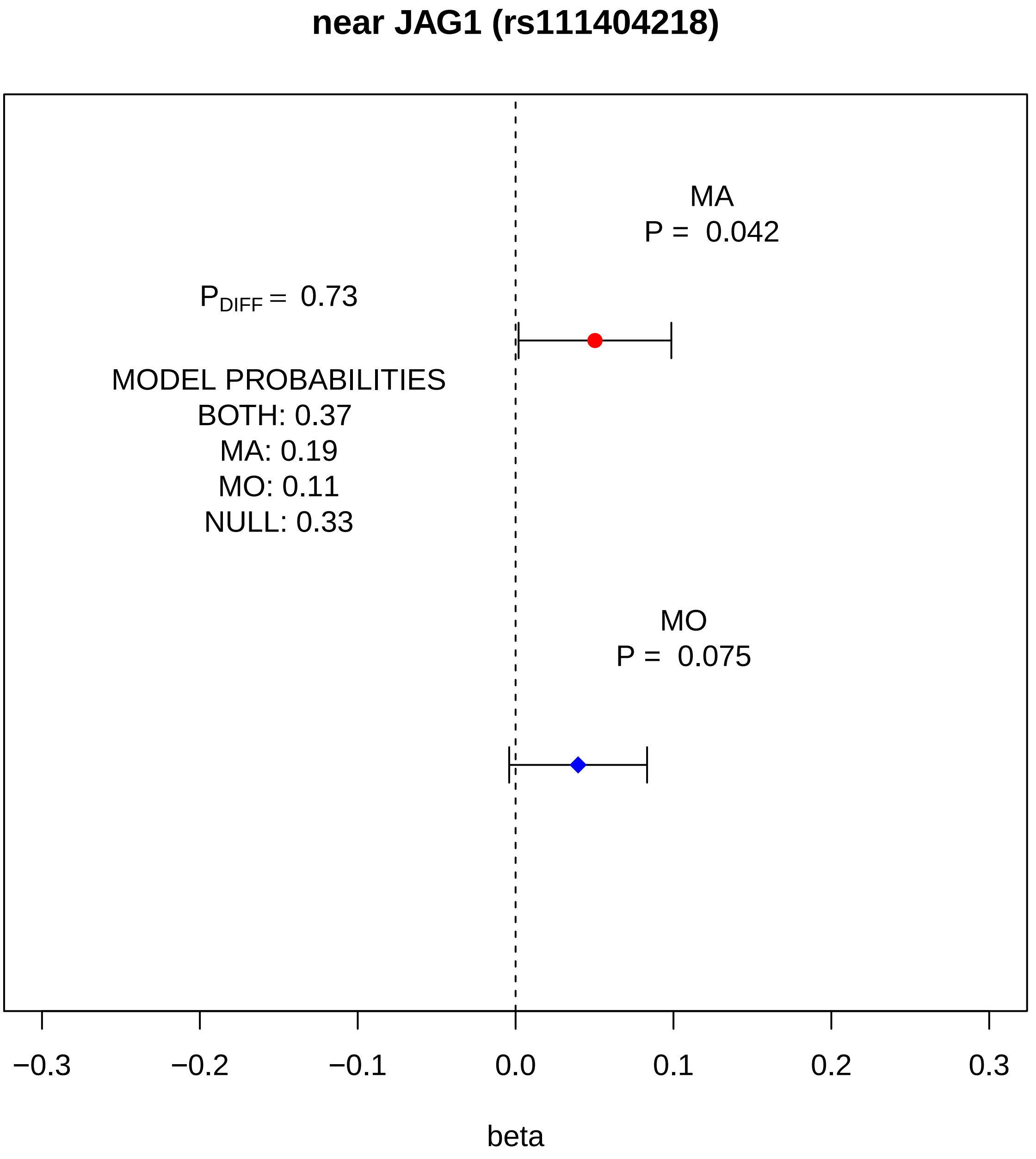
near JAG1 (rs111404218)
MA
P = 0.042
P_DIFF = 0.73
MODEL PROBABILITIES
BOTH: 0.37
MA: 0.19
MO: 0.11
NULL: 0.33
MO
P = 0.075
−0.3
−0.2
−0.1
0.0
0.1
0.2
0.3
beta

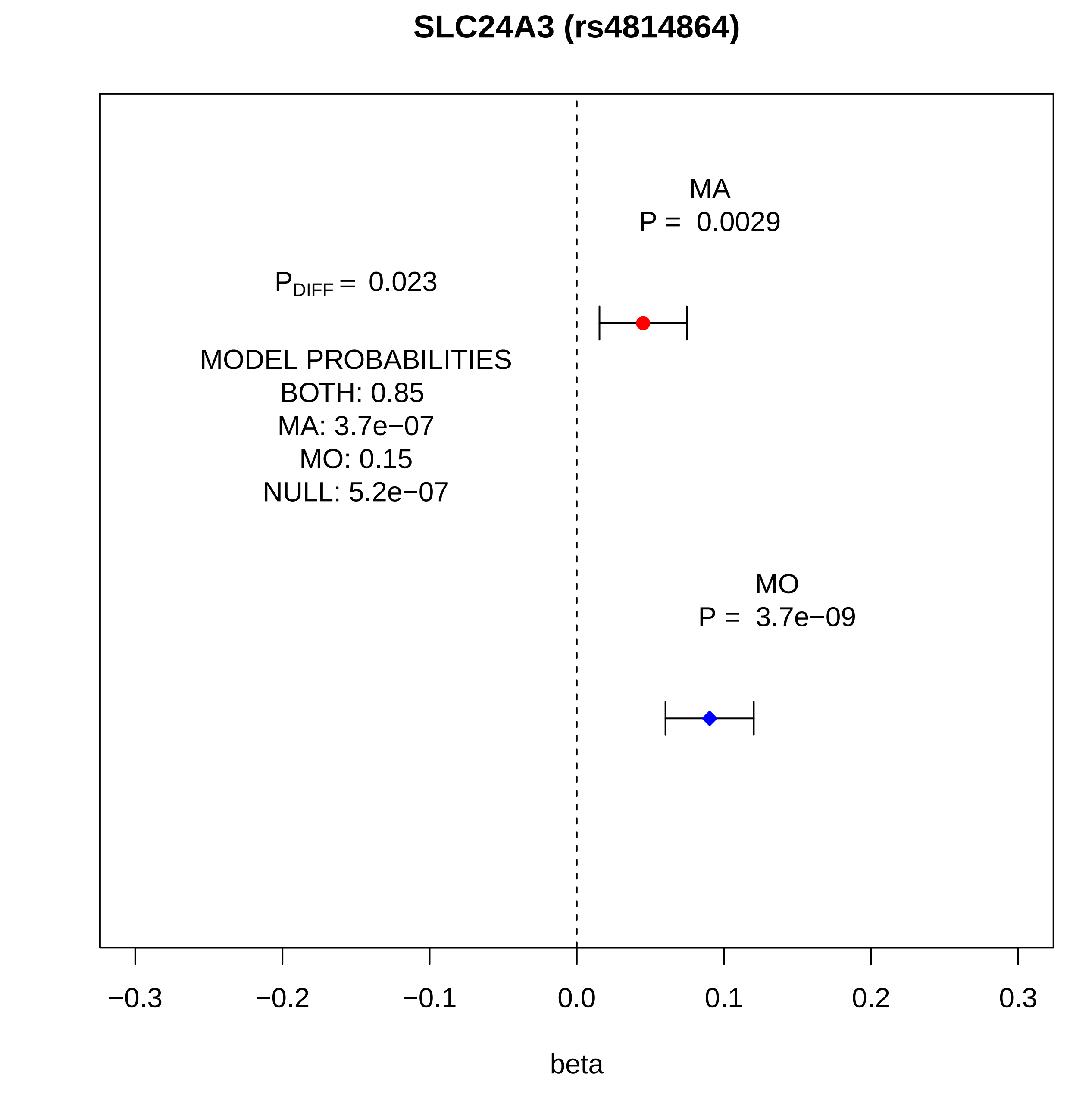
SLC24A3 (rs4814864)
MA
P = 0.0029
$P_{DIFF}$ = 0.023
MODEL PROBABILITIES
BOTH: 0.85
MA: 3.7e−07
MO: 0.15
NULL: 5.2e−07
MO
P = 3.7e−09
−0.3
−0.2
−0.1
0.0
0.1
0.2
0.3
beta

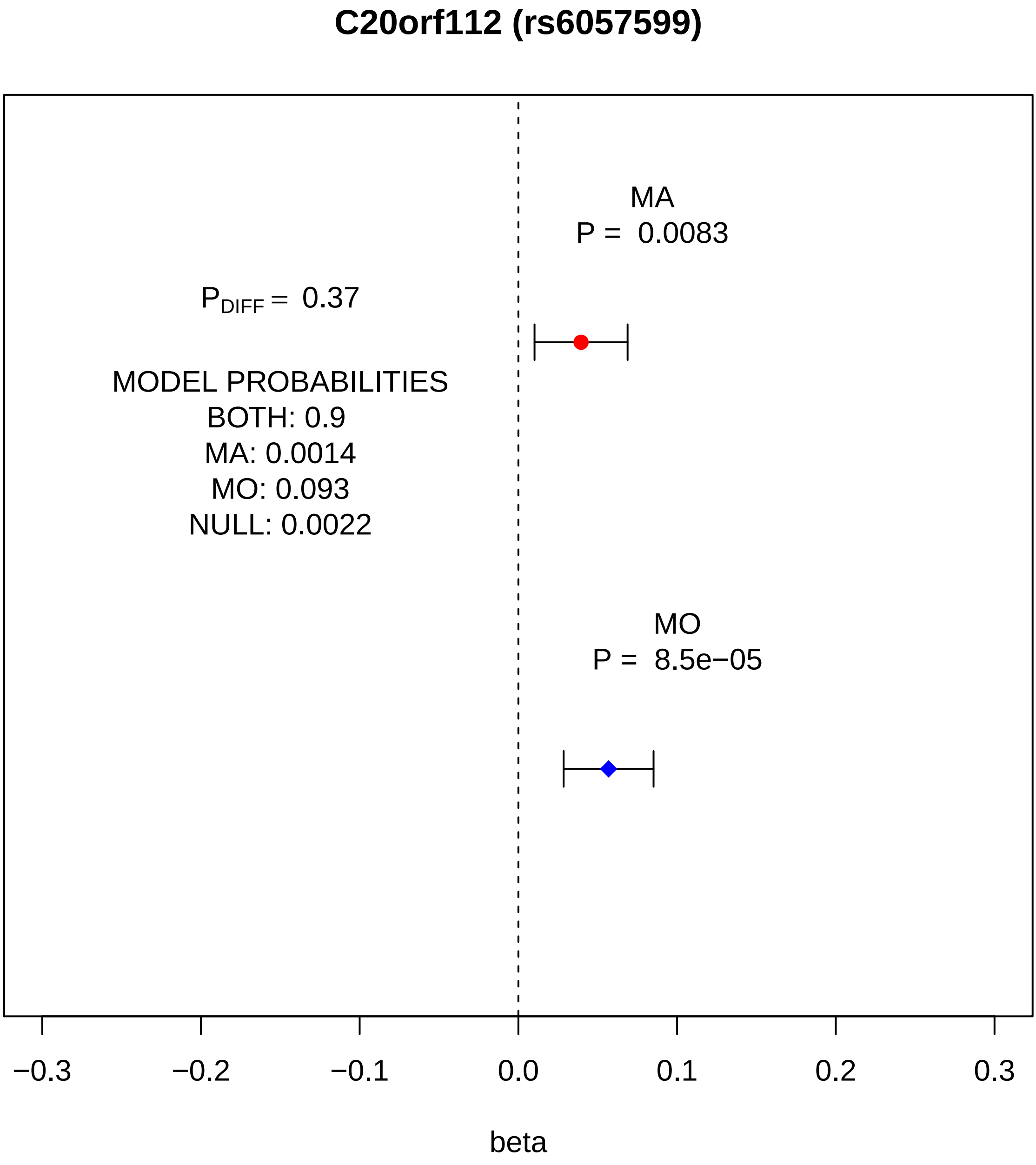
C20orf112 (rs6057599)
MA
P = 0.0083
P_DIFF = 0.37
MODEL PROBABILITIES
BOTH: 0.9
MA: 0.0014
MO: 0.093
NULL: 0.0022
MO
P = 8.5e−05
−0.3
−0.2
−0.1
0.0
0.1
0.2
0.3
beta

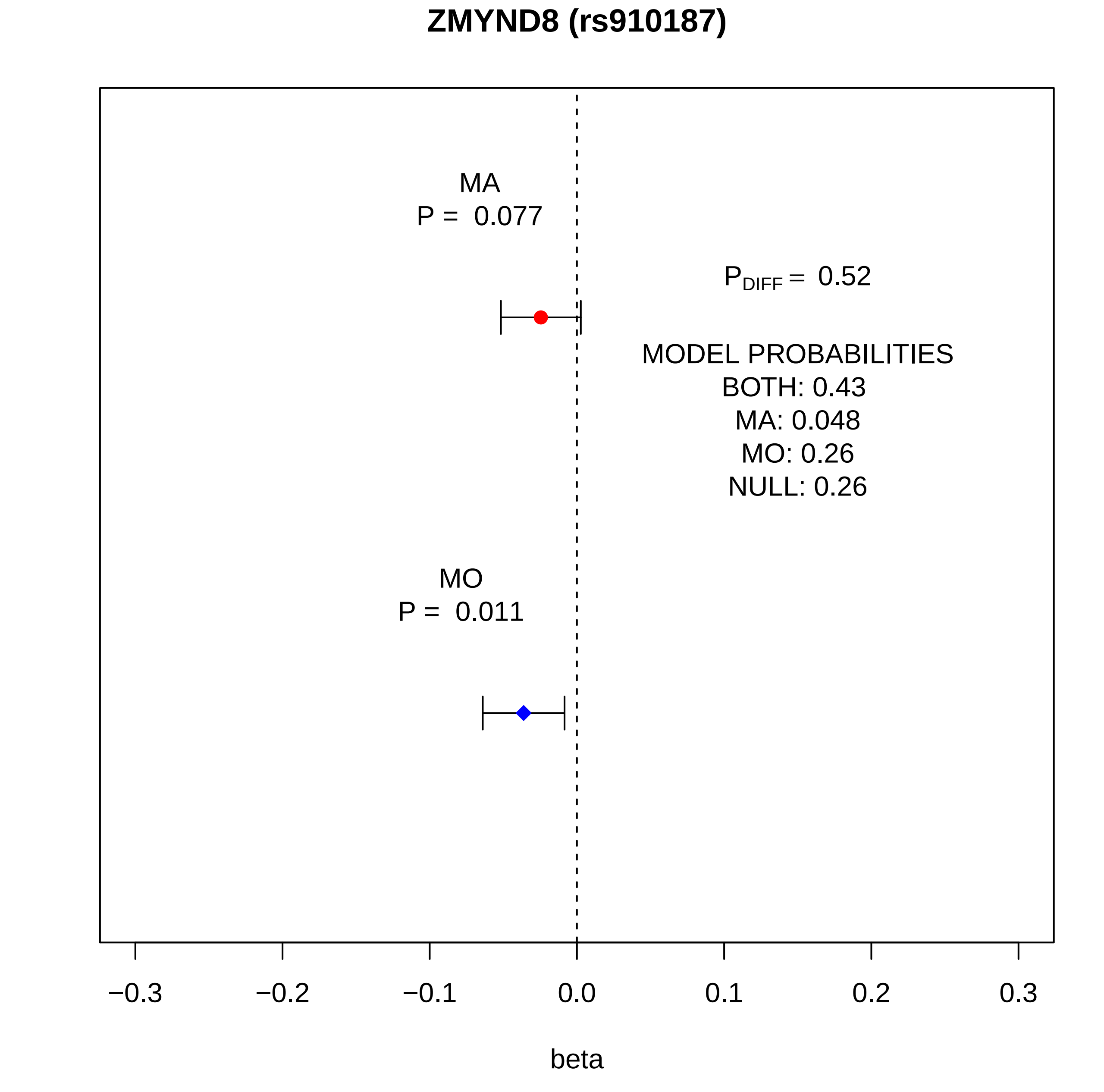
ZMYND8 (rs910187)
MA
P = 0.077
P_DIFF = 0.52
MODEL PROBABILITIES
BOTH: 0.43
MA: 0.048
MO: 0.26
NULL: 0.26
MO
P = 0.011
−0.3
−0.2
−0.1
0.0
0.1
0.2
0.3
beta

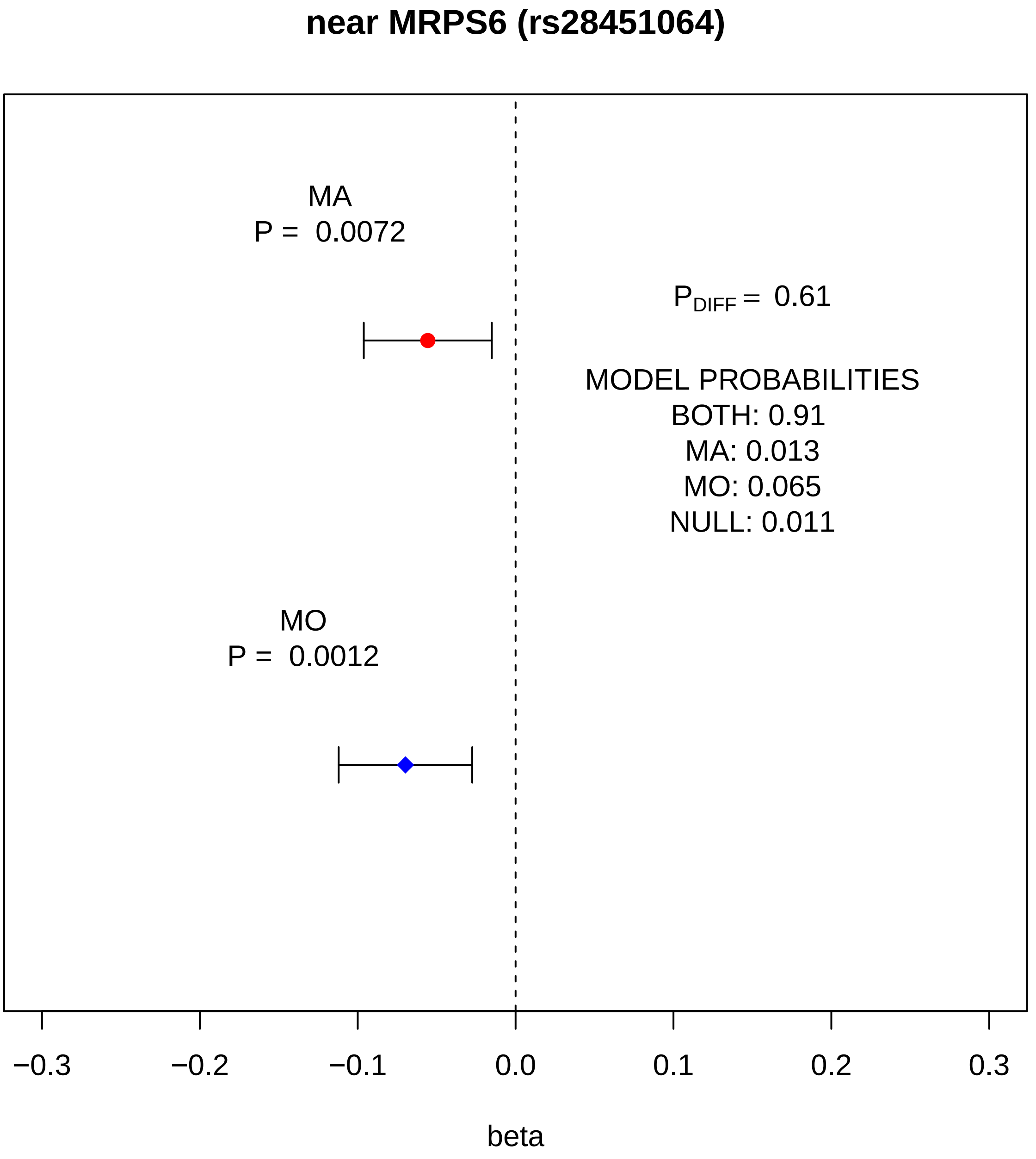

near MRPS6 (rs28451064)
MA
P = 0.0072
P_DIFF = 0.61
MODEL PROBABILITIES
BOTH: 0.91
MA: 0.013
MO: 0.065
NULL: 0.011
MO
P = 0.0012
−0.3
−0.2
−0.1
0.0
0.1
0.2
0.3
beta

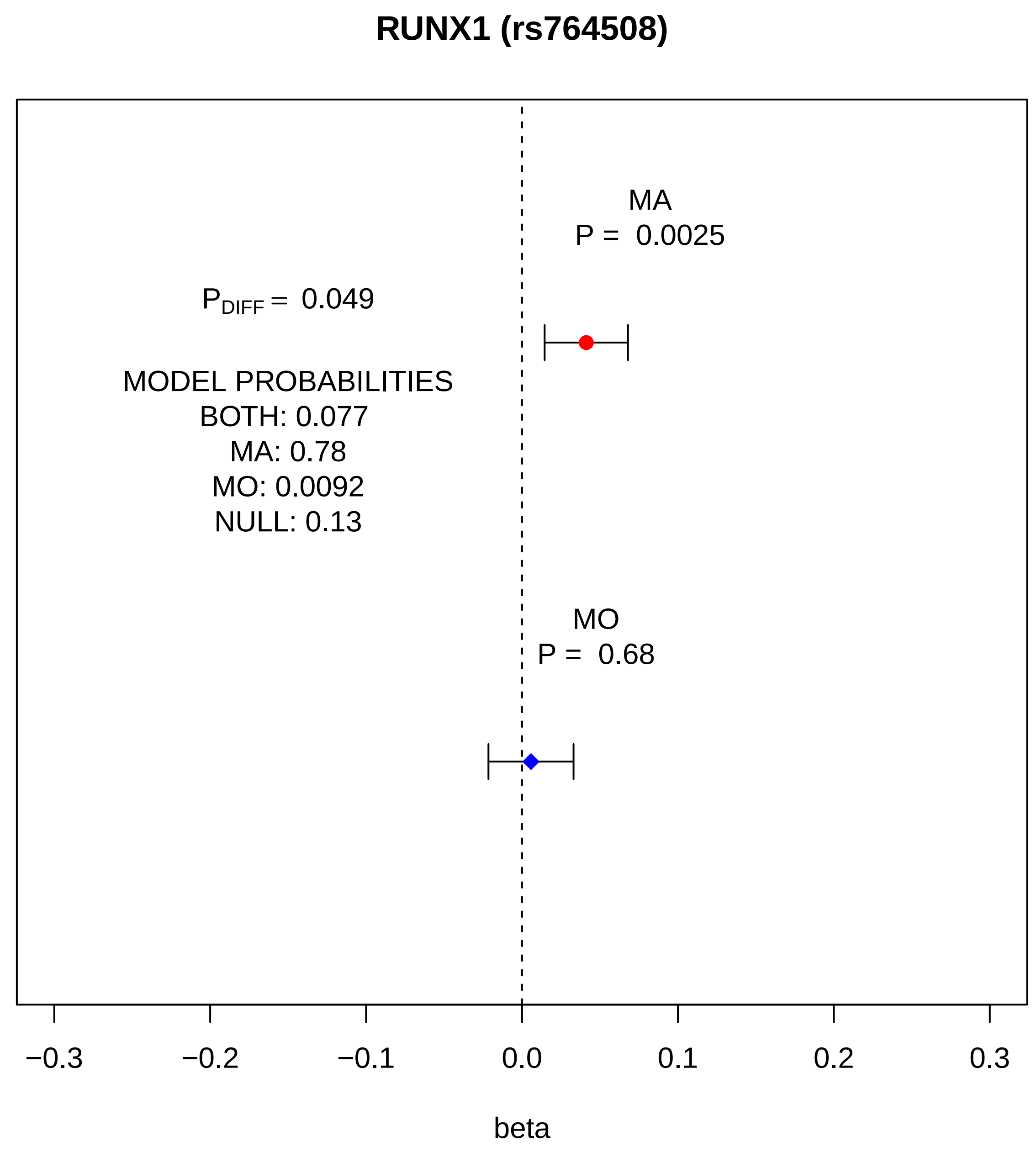
RUNX1 (rs764508)
MA
P = 0.0025
P_DIFF = 0.049
MODEL PROBABILITIES
BOTH: 0.077
MA: 0.78
MO: 0.0092
NULL: 0.13
MO
P = 0.68
−0.3
−0.2
−0.1
0.0
0.1
0.2
0.3
beta

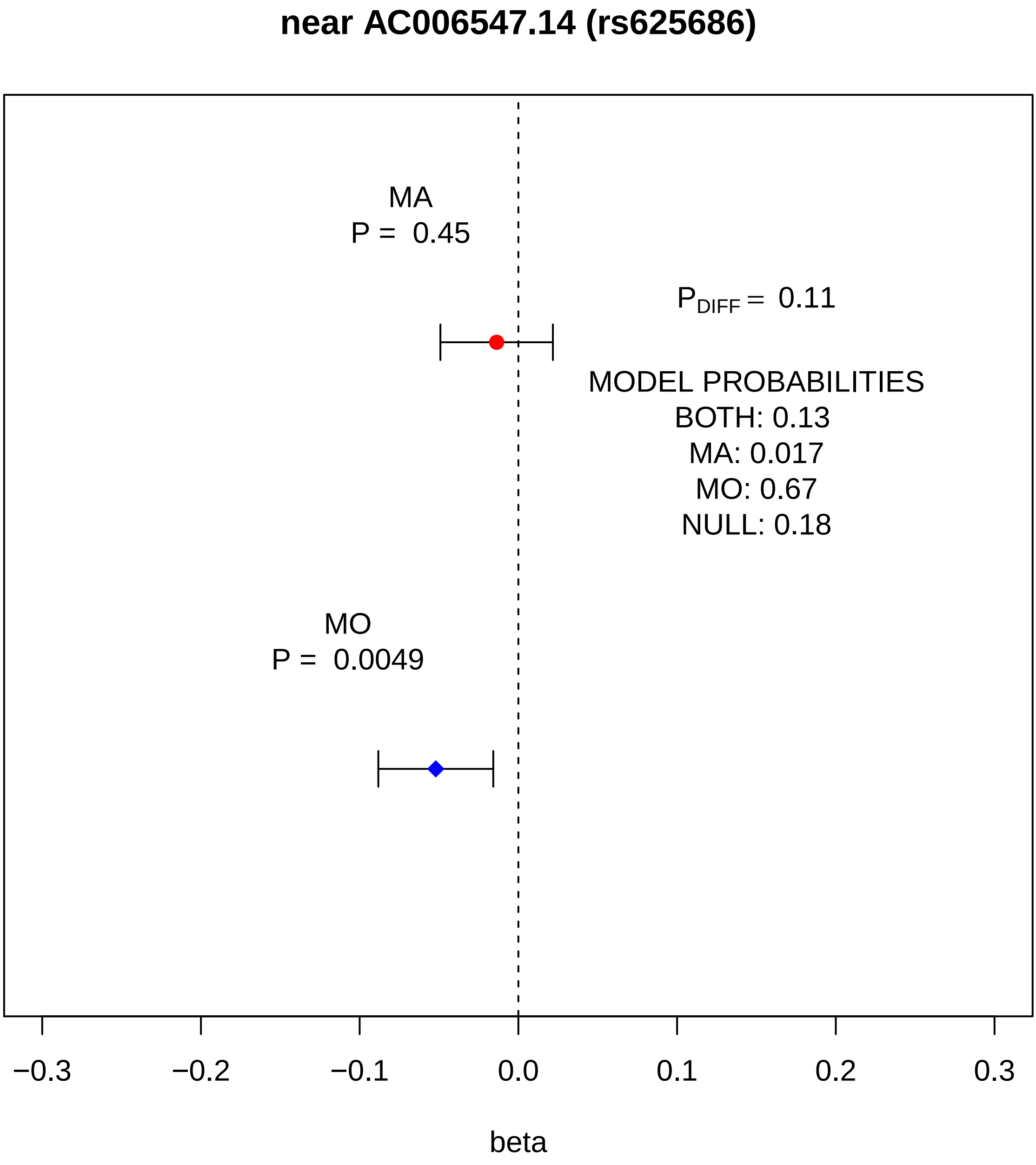

near AC006547.14 (rs625686)
MA
P = 0.45
$P_{DIFF}$ = 0.11
MODEL PROBABILITIES
BOTH: 0.13
MA: 0.017
MO: 0.67
NULL: 0.18
MO
P = 0.0049
−0.3
−0.2
−0.1
0.0
0.1
0.2
0.3
beta

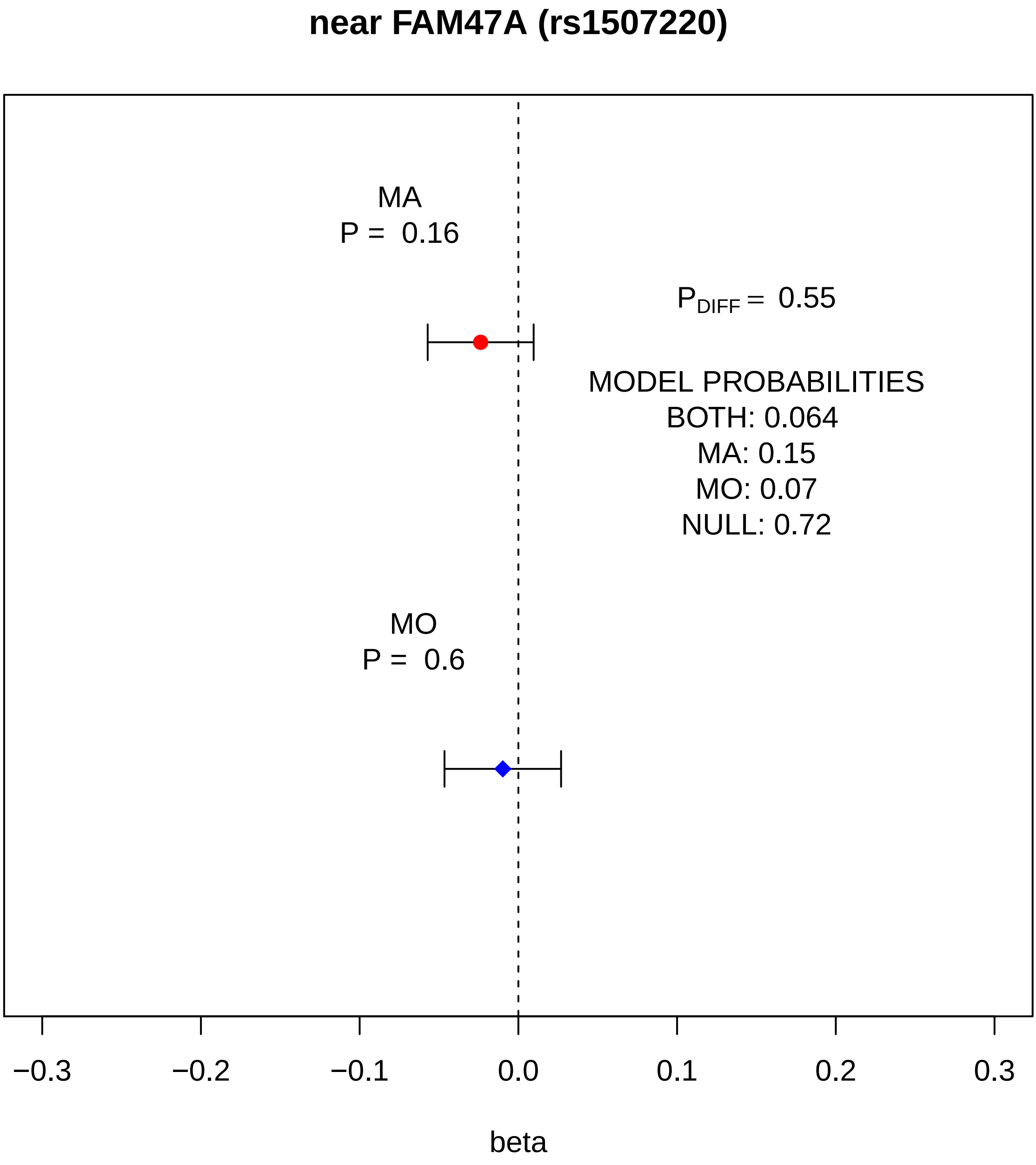

near FAM47A (rs1507220)
MA
P = 0.16
P_DIFF = 0.55
MODEL PROBABILITIES
BOTH: 0.064
MA: 0.15
MO: 0.07
NULL: 0.72
MO
P = 0.6
−0.3
−0.2
−0.1
0.0
0.1
0.2
0.3
beta

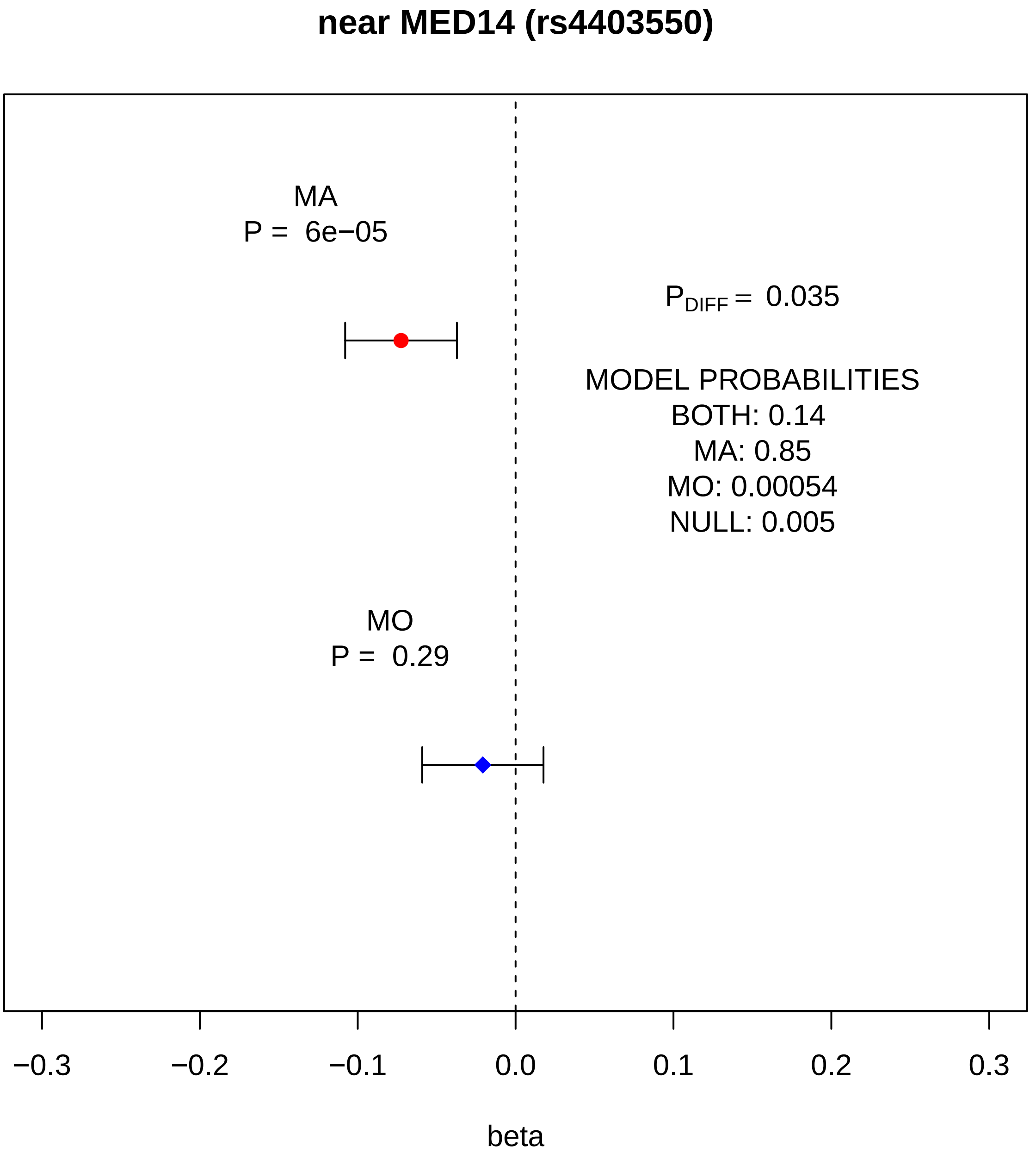
near MED14 (rs4403550)
MA
P = 6e−05
P_DIFF = 0.035
MODEL PROBABILITIES
BOTH: 0.14
MA: 0.85
MO: 0.00054
NULL: 0.005
MO
P = 0.29
−0.3
−0.2
−0.1
0.0
0.1
0.2
0.3
beta
