## Supplementary Figure 12. for "Genome-wide analysis of 102,084 migraine cases identifies 123 risk loci and subtype-specific risk alleles"

**Supplementary Figure 12. Pairwise EAF and MAF plots against the reference cohort (UKBB).** We plot four plots per cohort pair and separately for autosomal SNPs, autosomal indels, SNPs in chromosome X and indels in chromosome X. The first plot shows EAFs of overlapping variants before flipping alleles to match reference data and without exclusions. Second plot shows EAFs after allele flipping and after removing variants with non-matching alleles and variants with EAF discrepancy over 0.3 (SNPs) or over 0.2 (indels). Third plot shows MAFs of overlapping variants before flipping alleles to match reference data and without exclusions. Fourth plot shows MAFs after allele flipping and after removing variants with non-matching alleles and variants with EAF discrepancy over 0.3 (SNPs) or over 0.2 (indels). EAF = effect allele frequency, MAF = minor allele frequency.

Autosomal biallelic SNPs

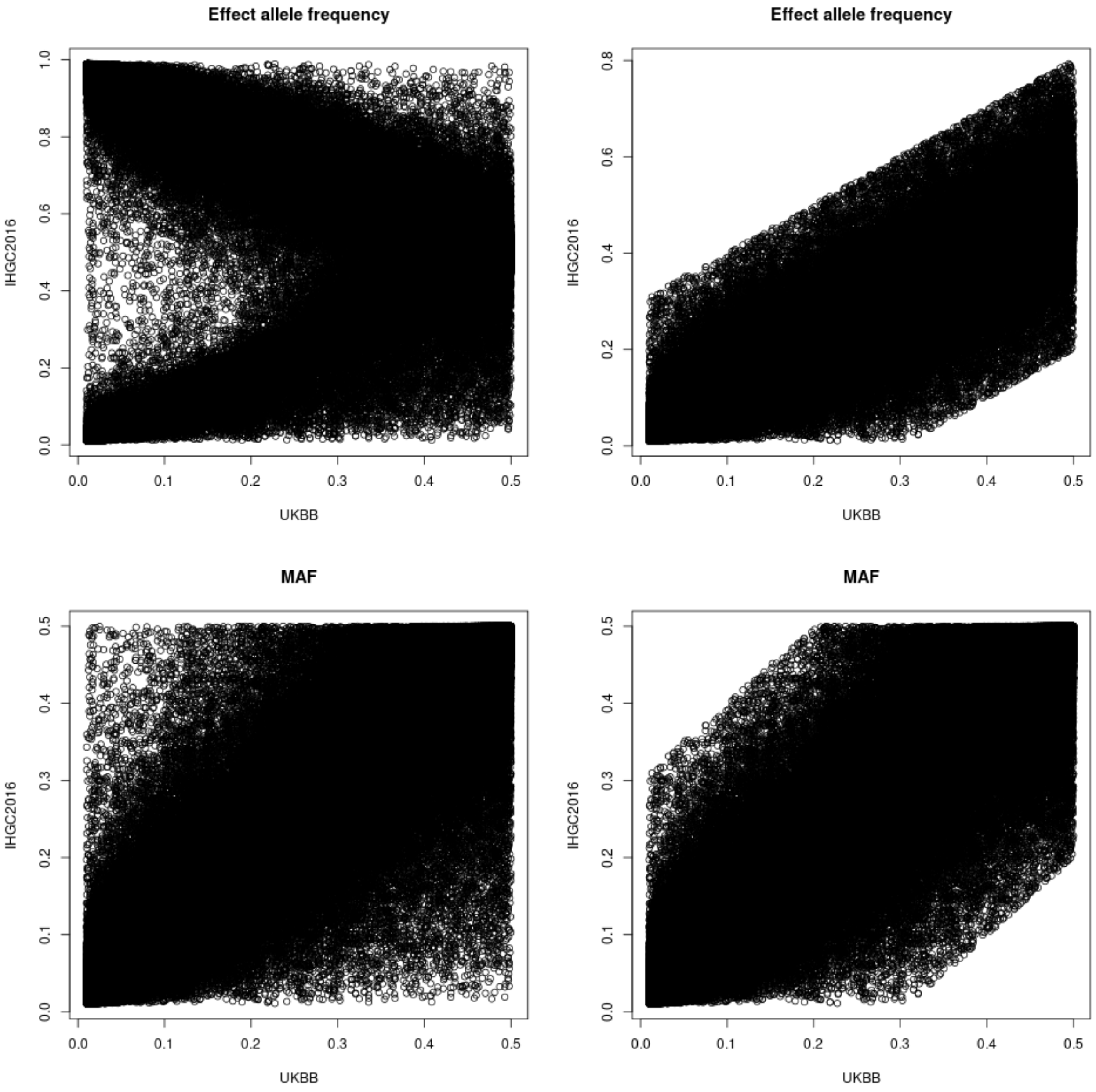


UKBB vs. IHGC2016: 7,525,014 overlapping autosomal SNPs, 7,524,675 with matching alleles and 3,617 variants with EAF discrepancy > 0.3.

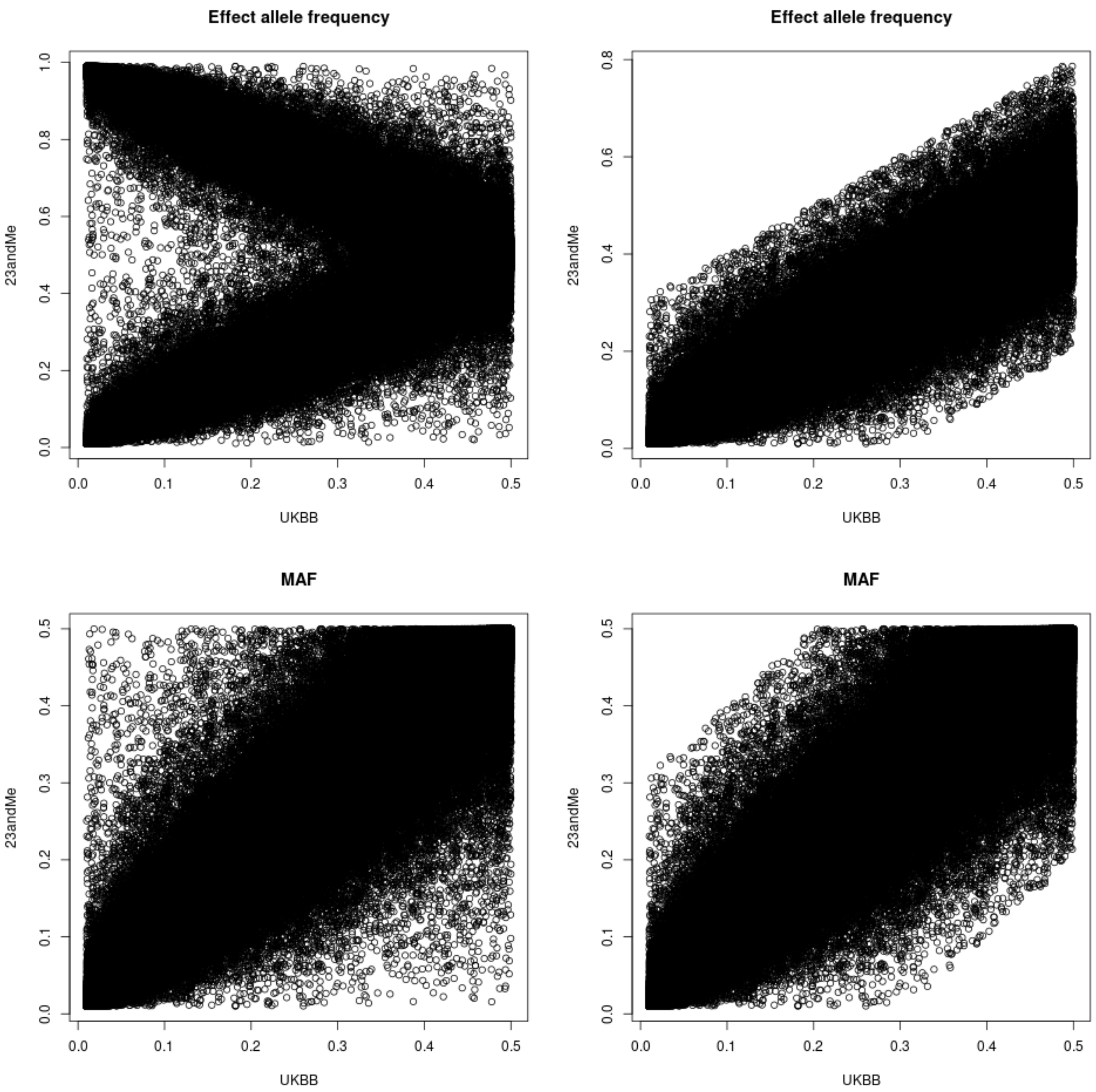


UKBB vs. 23andMe: 7,396,129 overlapping autosomal SNPs, 7,395,895 with matching alleles and 1,209 variants with EAF discrepancy > 0.3.

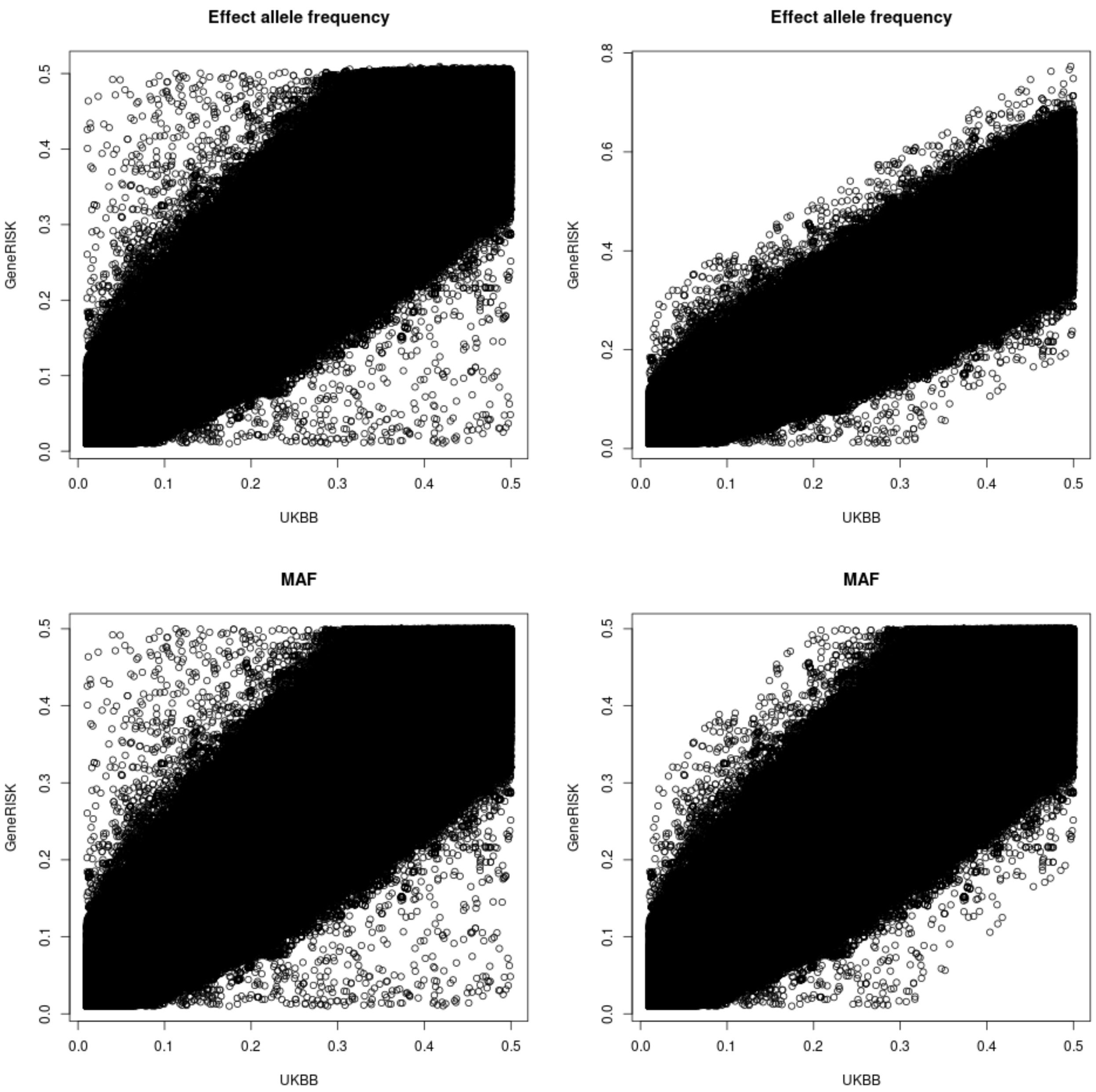


UKBB vs. GeneRISK: 7,093,034 overlapping autosomal SNPs, 7,092,857 with matching alleles and 470 variants with EAF discrepancy > 0.3.

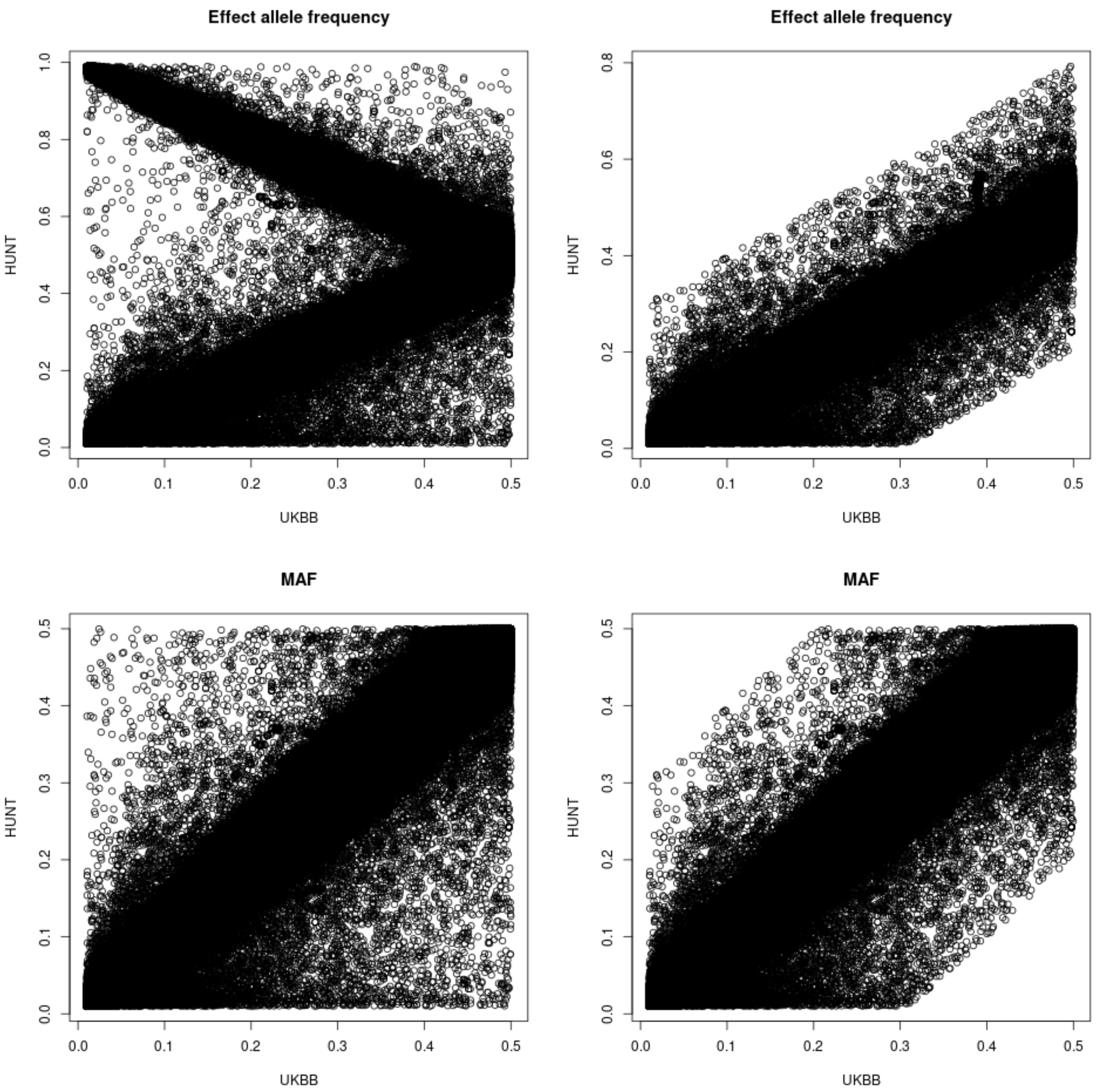


UKBB vs. HUNT: 7,754,925 overlapping autosomal SNPs, 7,754,843 with matching alleles, 1,082 variants with EAF discrepancy > 0.3.

Biallelic SNPs (chromosome X)

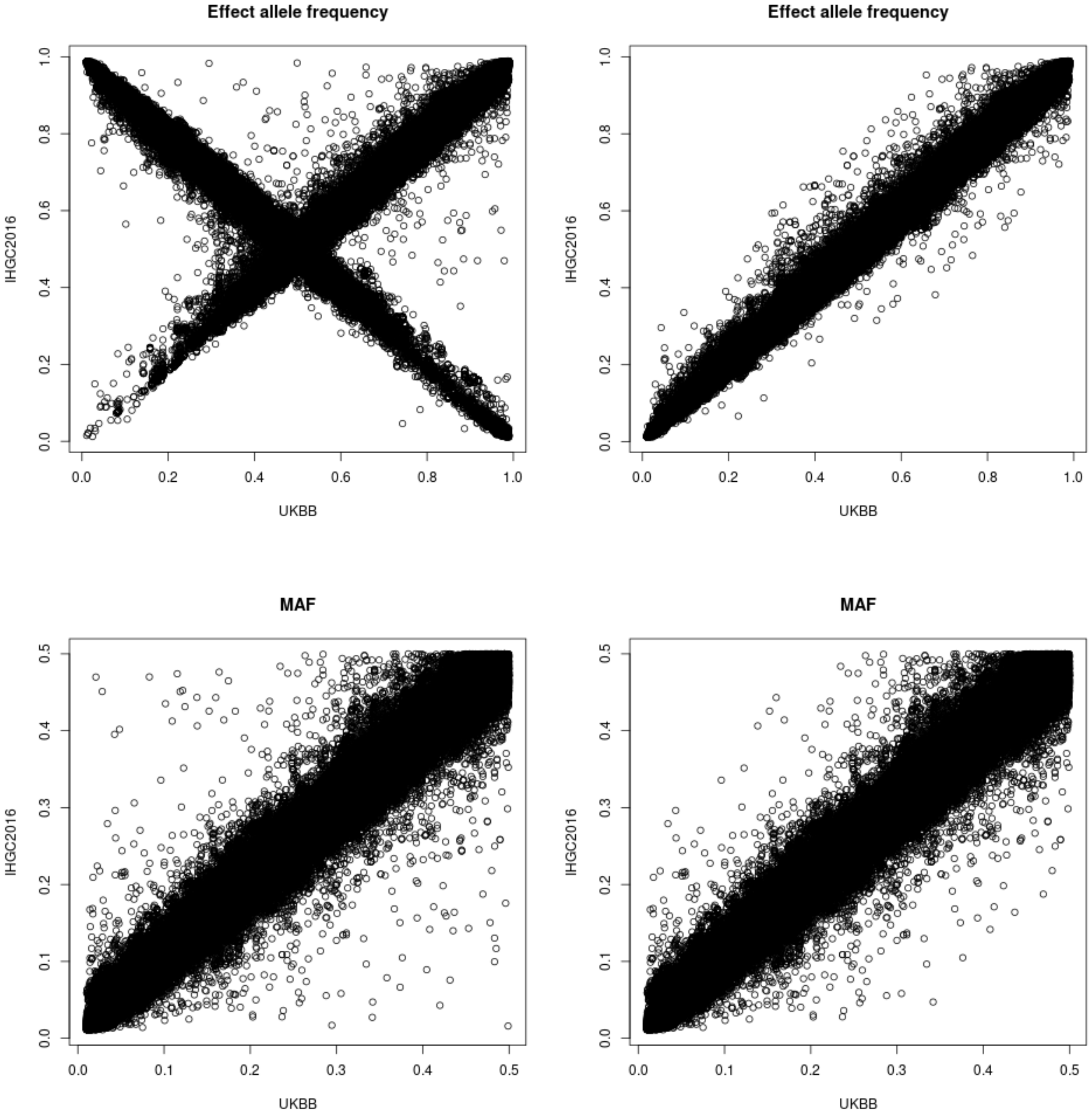


UKBB vs. IHGC2016:194,605 overlapping SNPs in chromosome X, 194,589 with matching alleles, 45 variants with EAF discrepancy > 0.3.

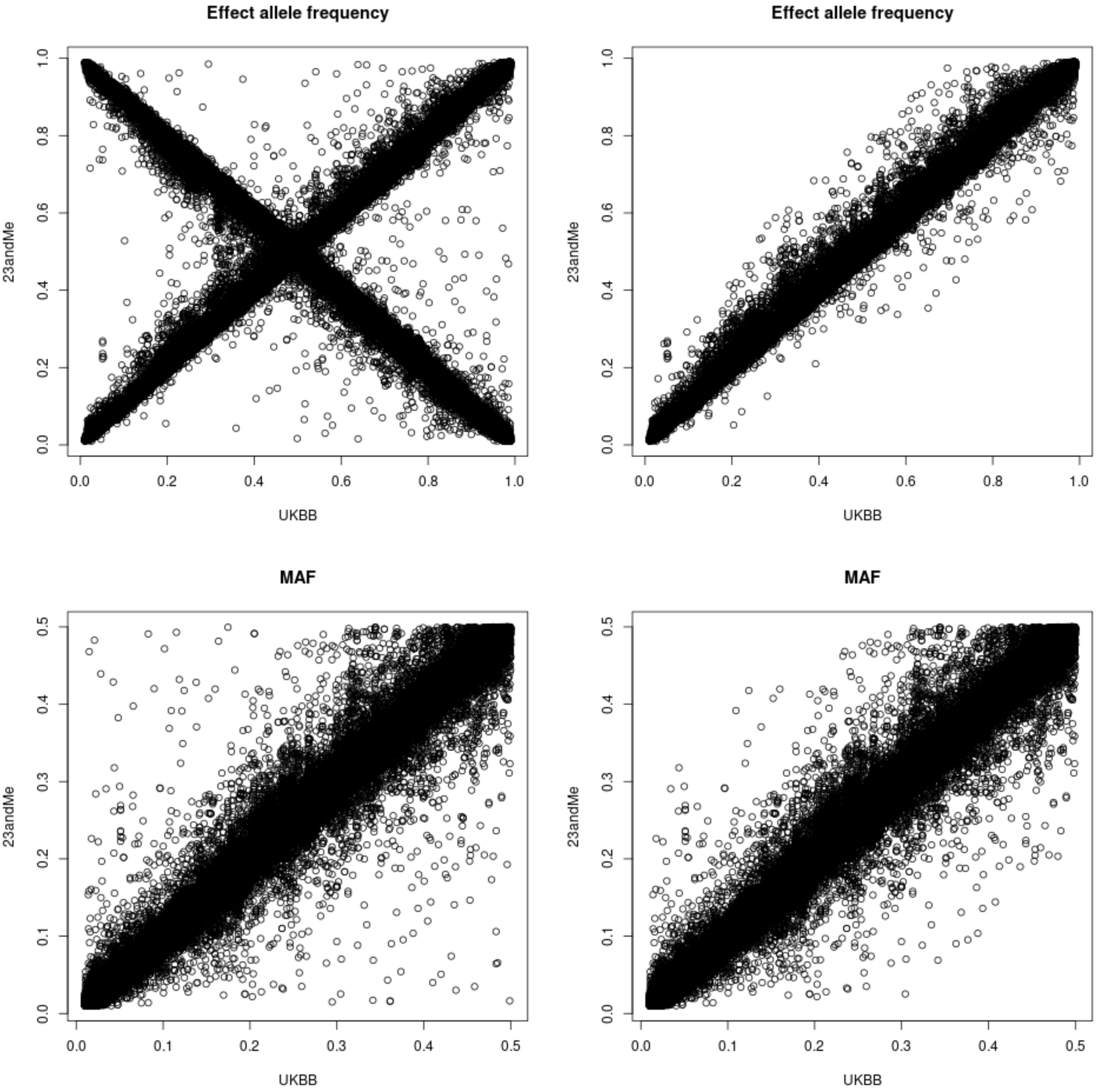


UKBB vs. 23andMe: 210,128 overlapping SNPs in chromosome X, 210,108 with matching alleles, 75 variants with EAF discrepancy > 0.3.

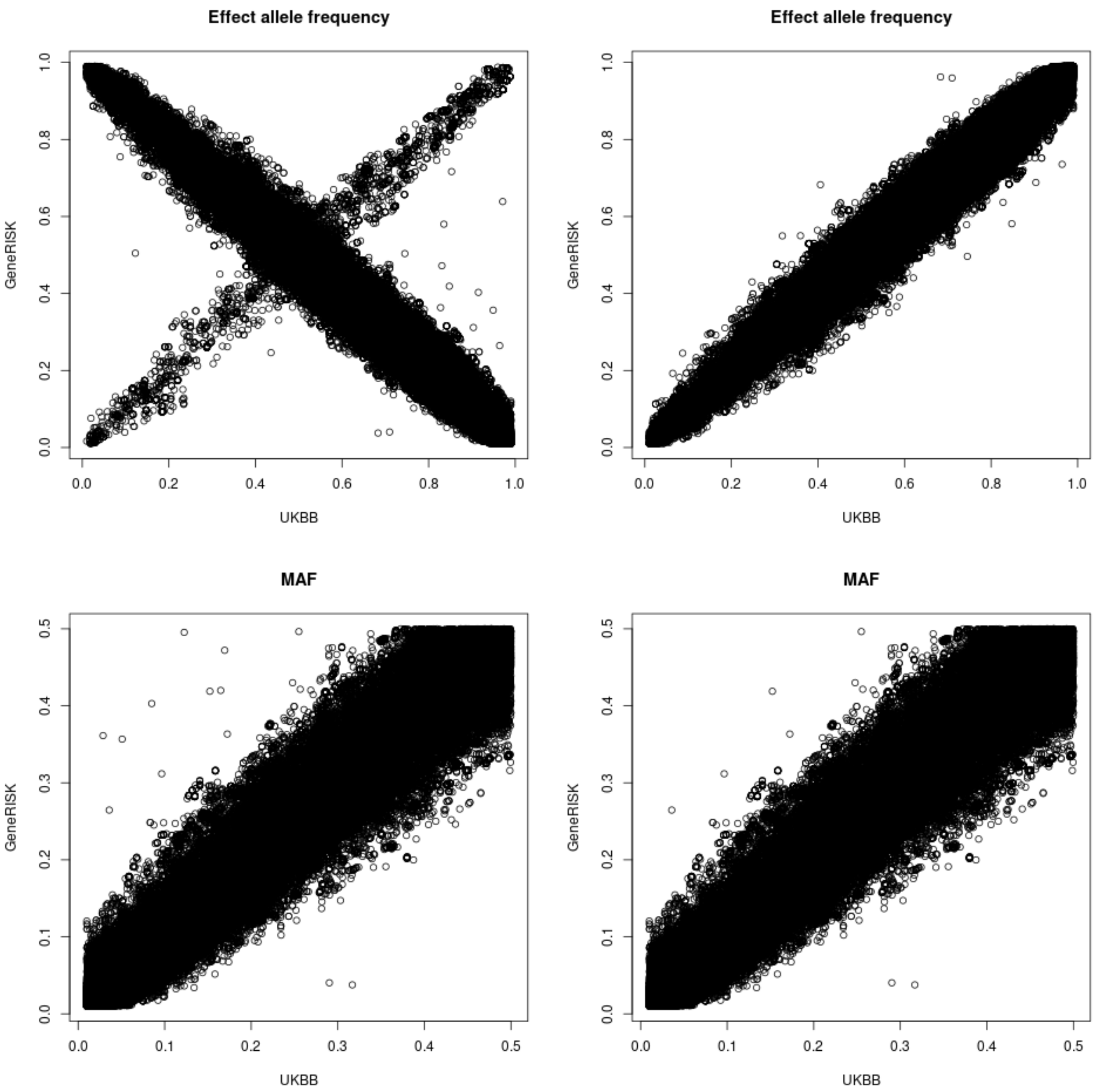


UKBB vs. GeneRISK: 193,169 overlapping SNPs in chromosome X, 193,166 with matching alleles, 11 variants with EAF discrepancy > 0.3.

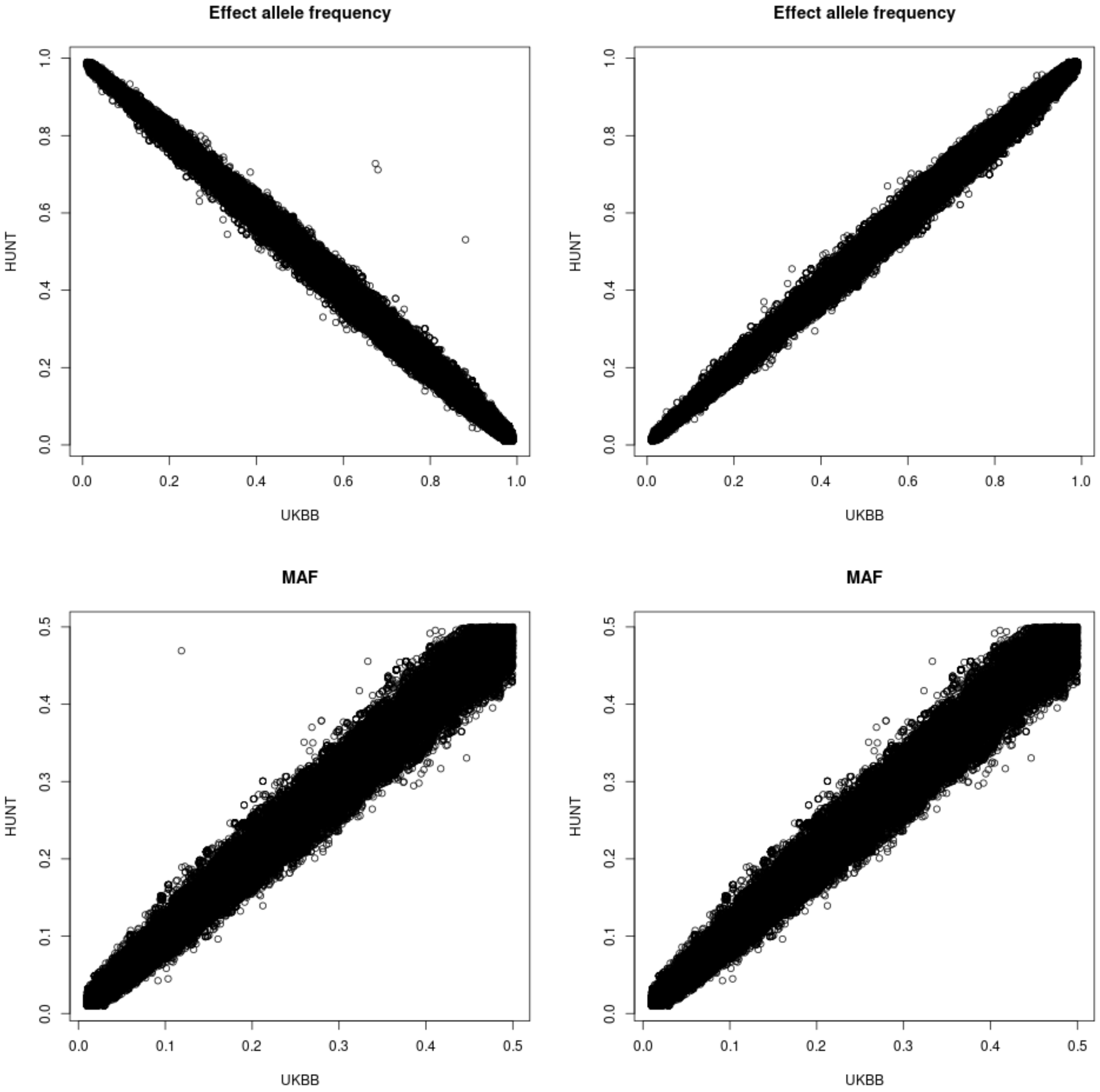


UKBB vs. HUNT: 201,225 overlapping SNPs in chromosome X, 201,225 with matching alleles, 3 variants with EAF discrepancy > 0.3.

Autosomal biallelic indels

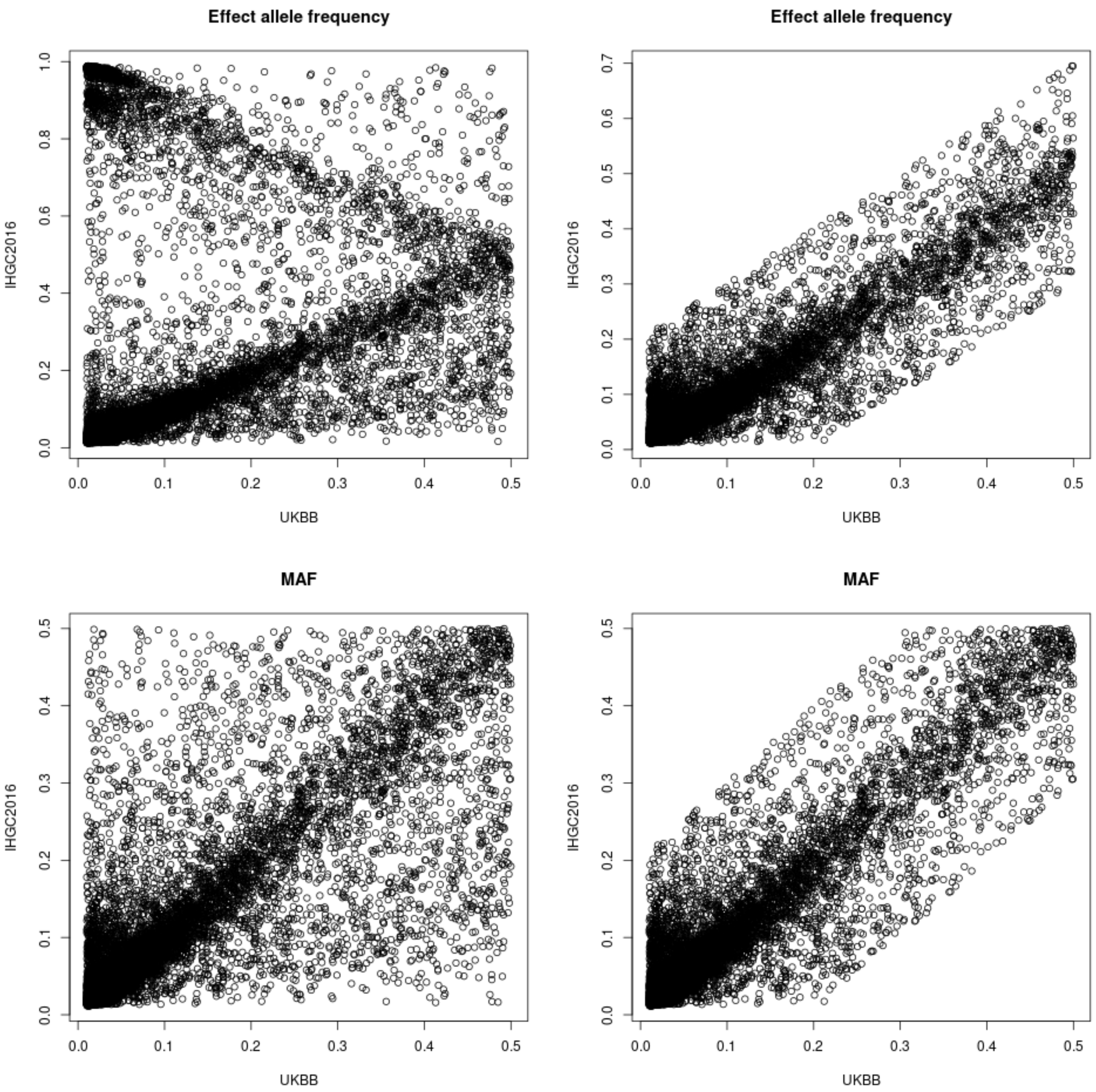


UKBB vs. IHGC2016: 14,008 overlapping autosomal indels, 4,262 indels with EAF discrepancy > 0.2.

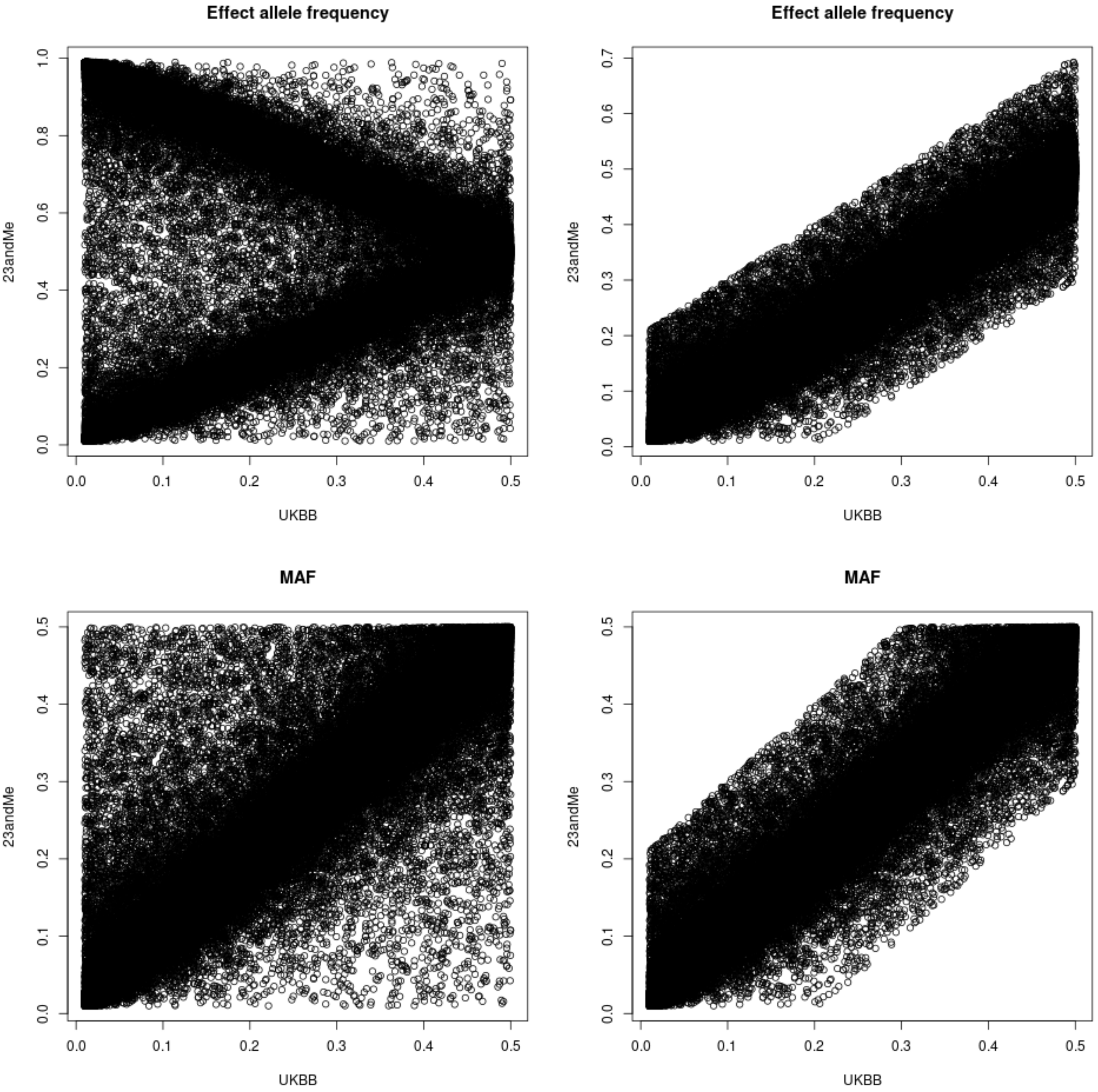


UKBB vs. 23andMe: 490,083 overlapping autosomal indels, 1,042 indels with EAF discrepancy > 0.2.

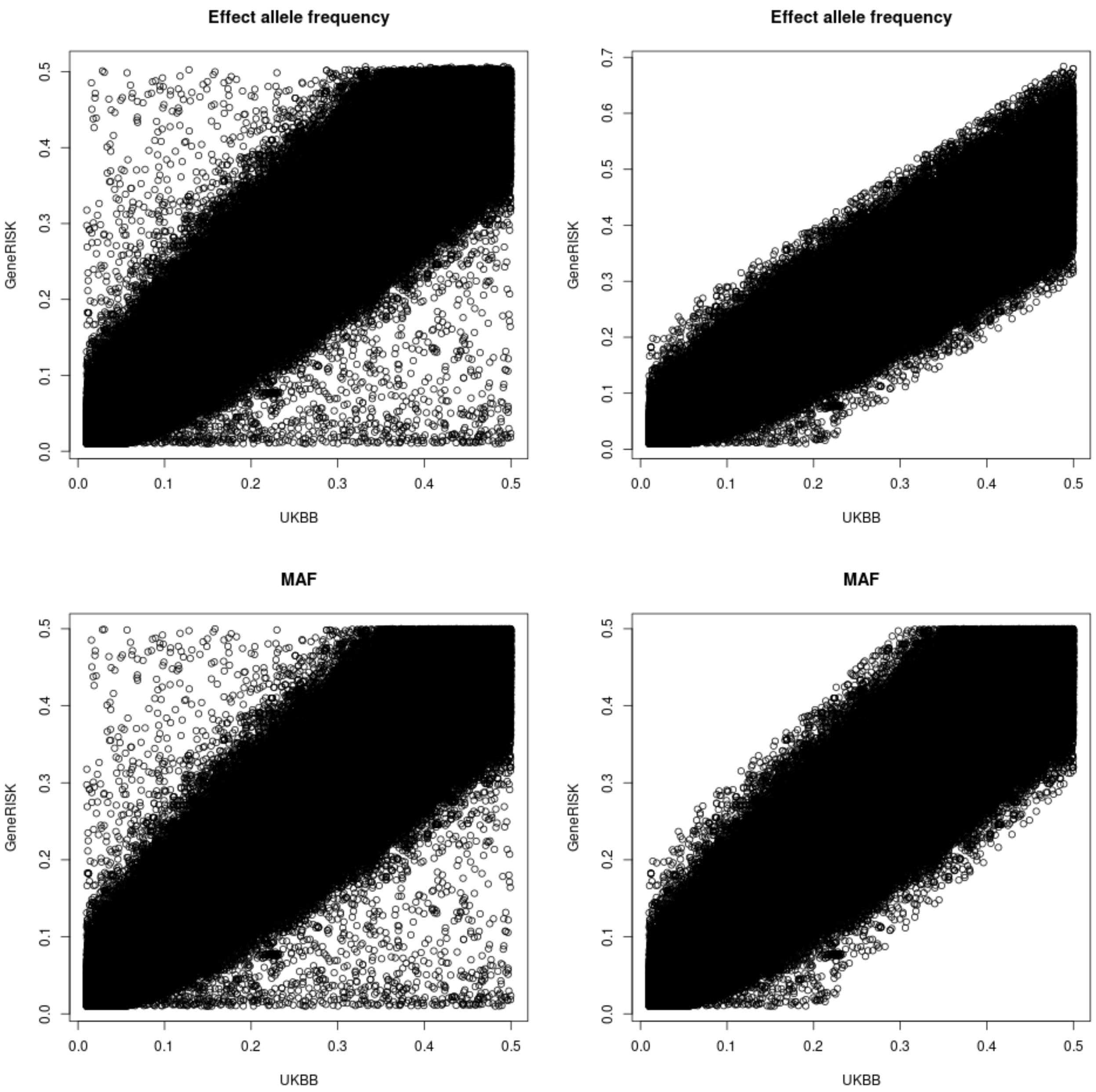


UKBB vs. GeneRISK: 478,926 overlapping autosomal indels, 1,639 indels with EAF discrepancy > 0.2.

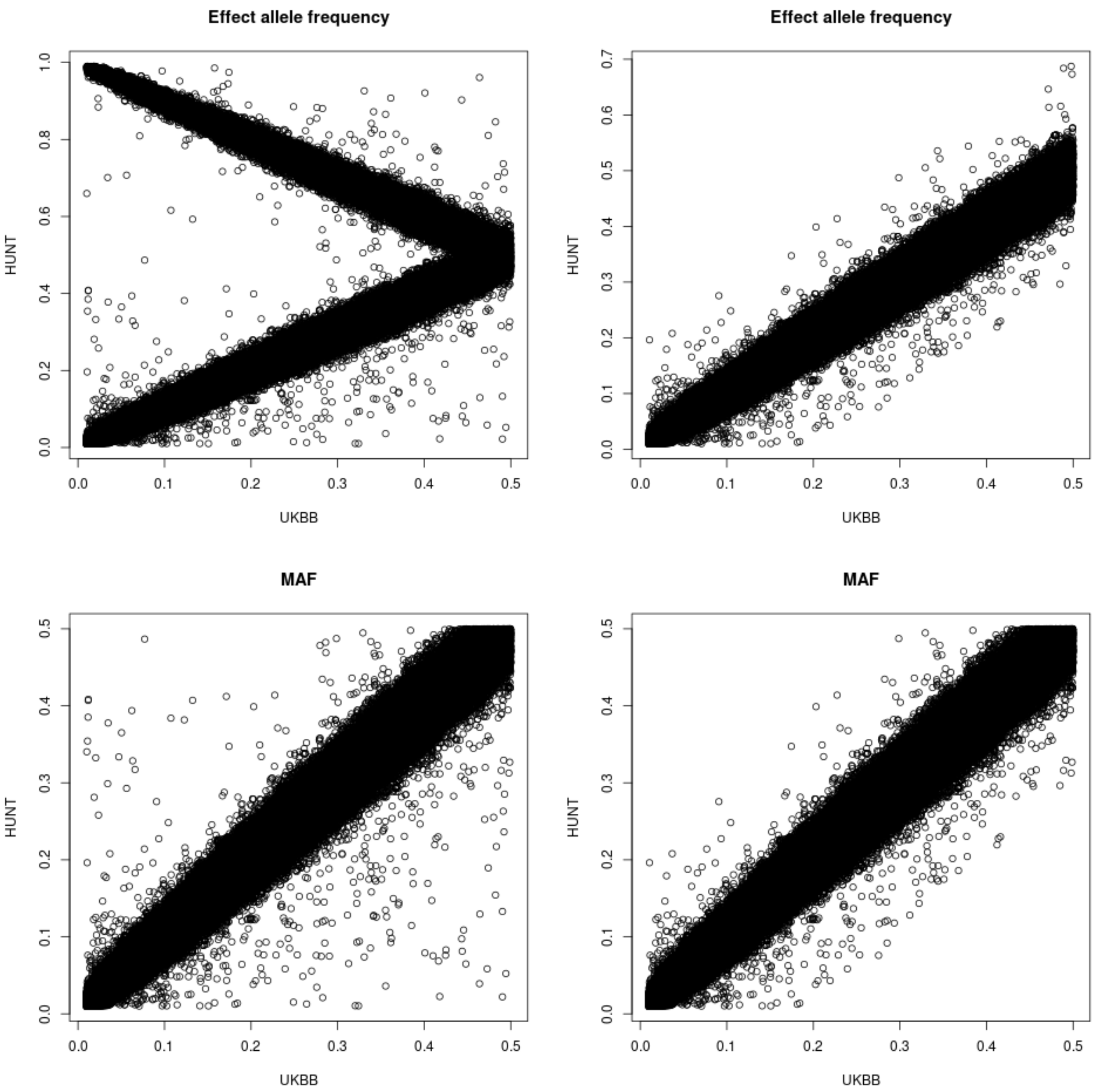


UKBB vs. HUNT: 288,013 overlapping autosomal indels, 127 indels with EAF discrepancy > 0.2.

Biallelic indels (chromosome X)

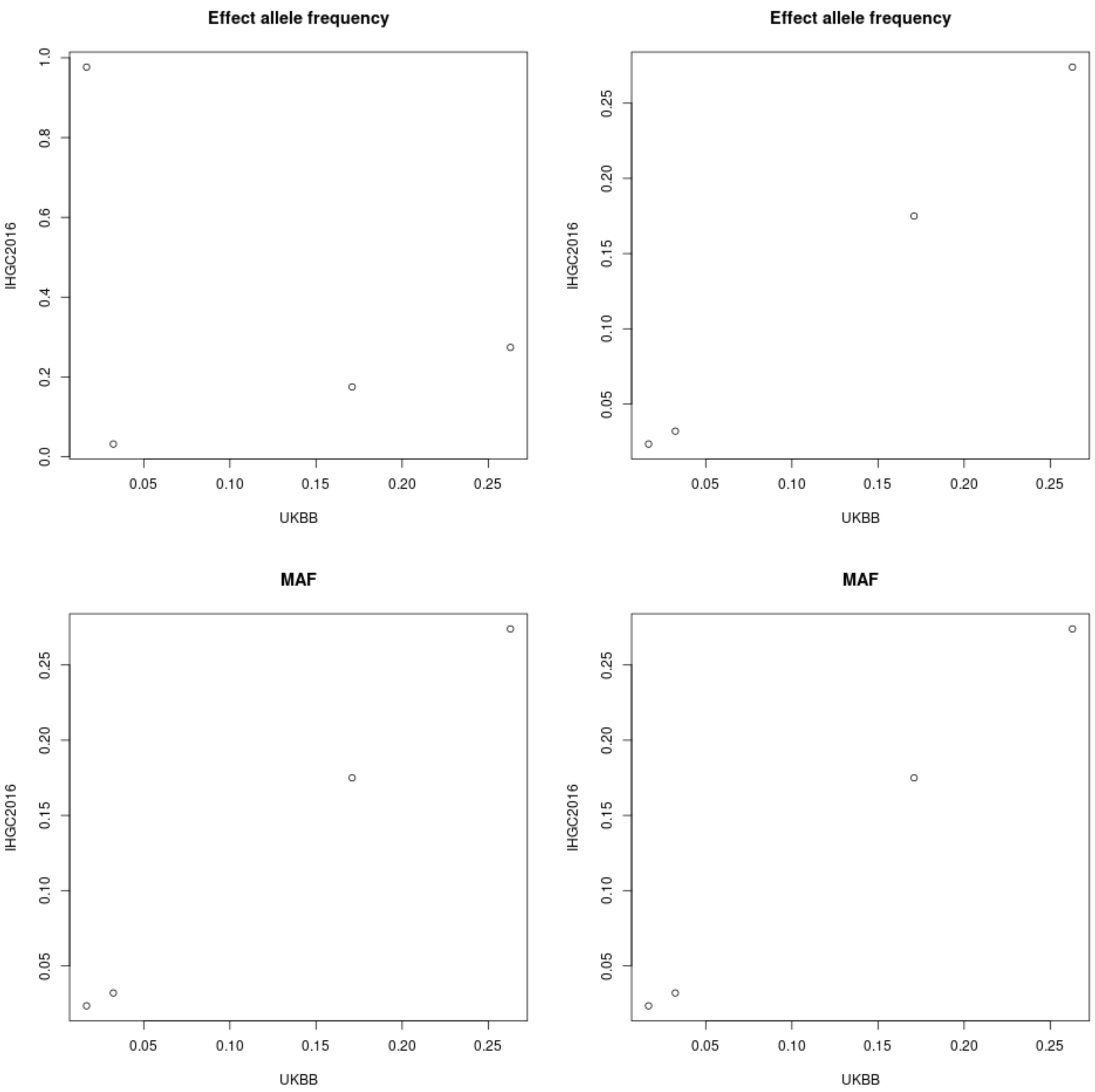


UKBB vs. IHGC2016: 4 overlapping indels in chromosome X, 0 variants with EAF discrepancy > 0.2.

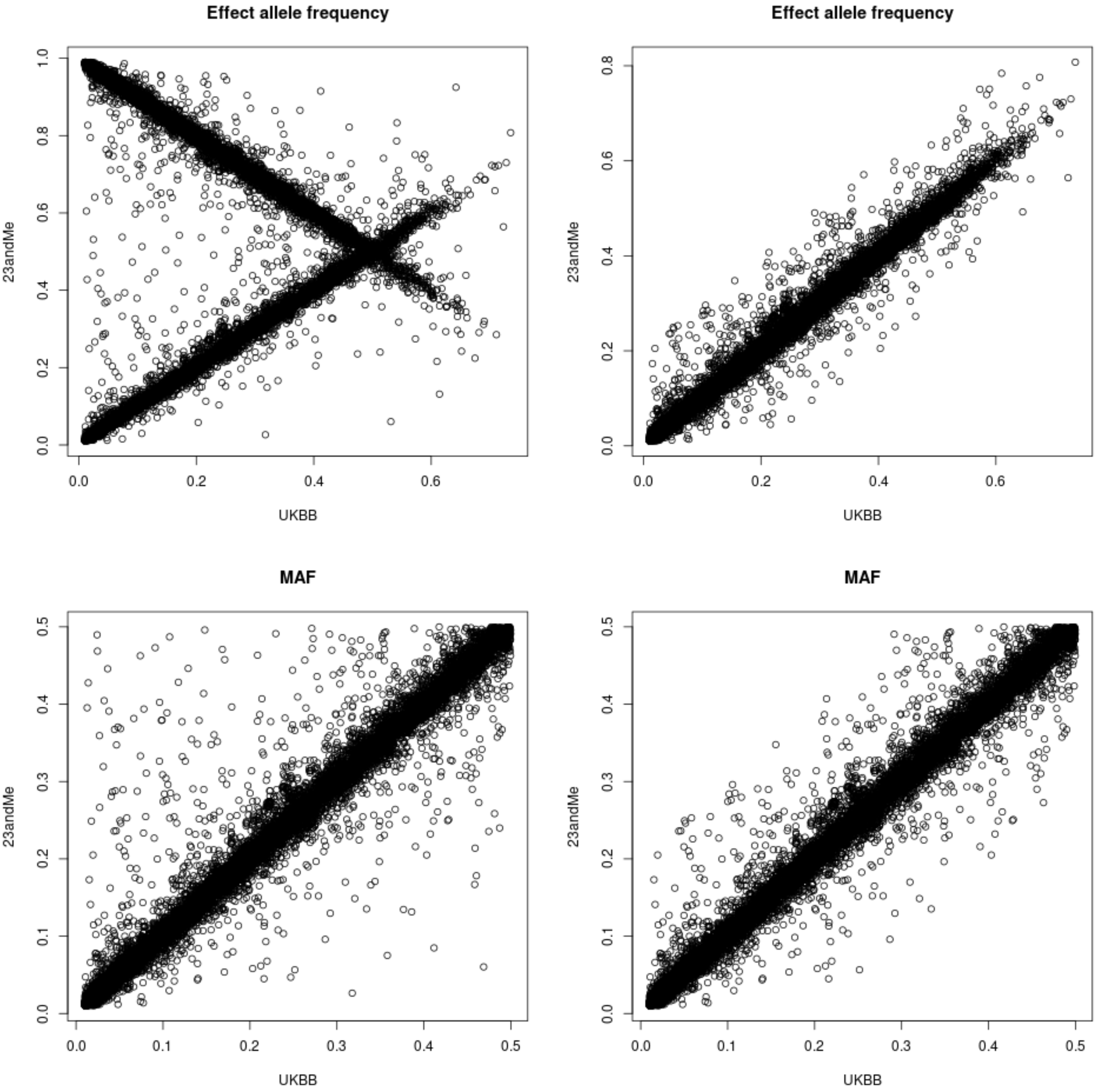


UKBB vs. 23andMe: 14,625 overlapping indels in chromosome X, 92 variants with EAF discrepancy > 0.2.

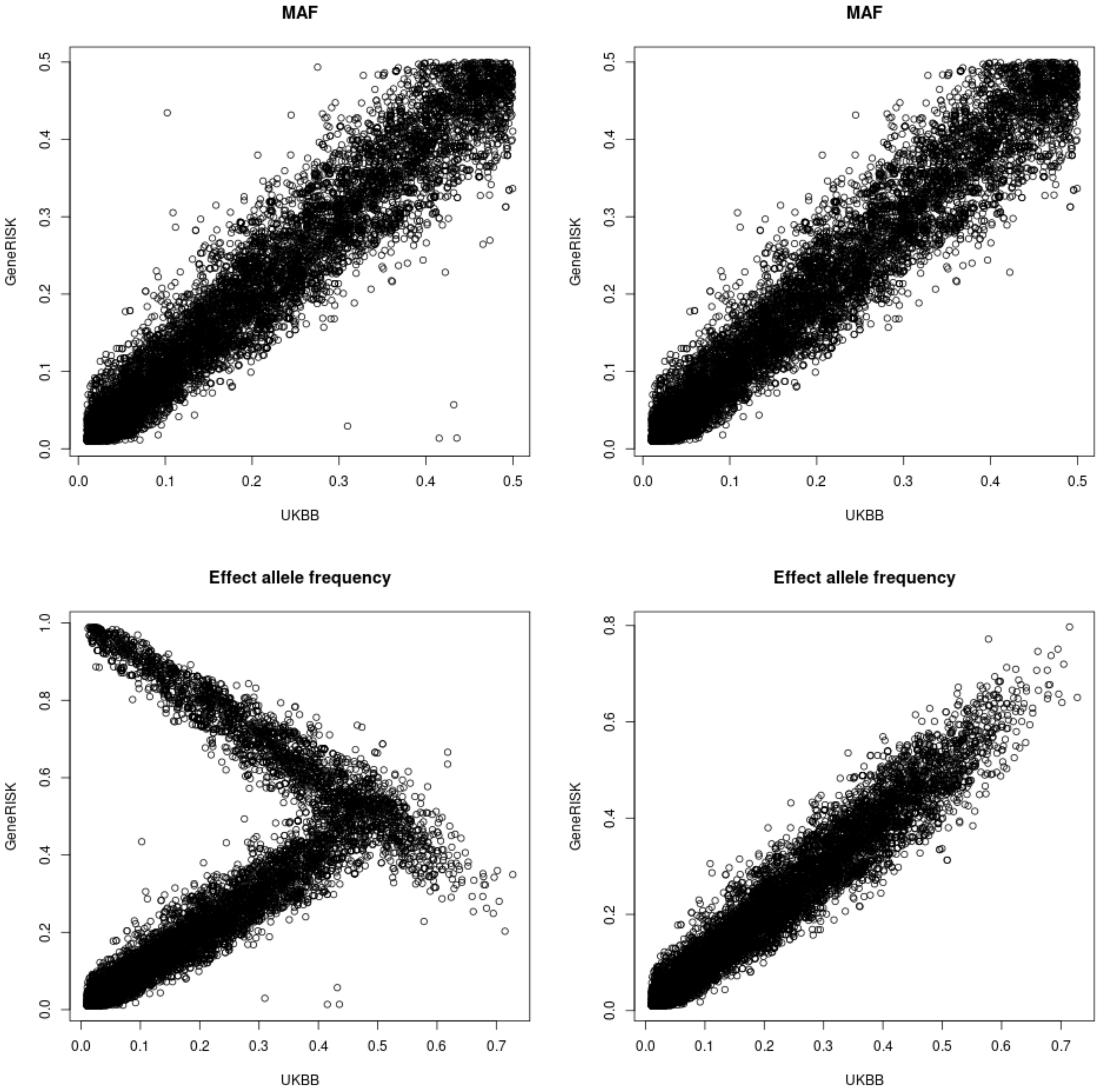


UKBB vs. GeneRISK: 11,518 overlapping indels in chromosome X, 20 variants with EAF discrepancy > 0.2.
