## Supplementary figures and images for "Genome-wide analysis of 102,084 migraine cases identifies 123 risk loci and subtype-specific risk alleles"

### Supplementary Figure 13.

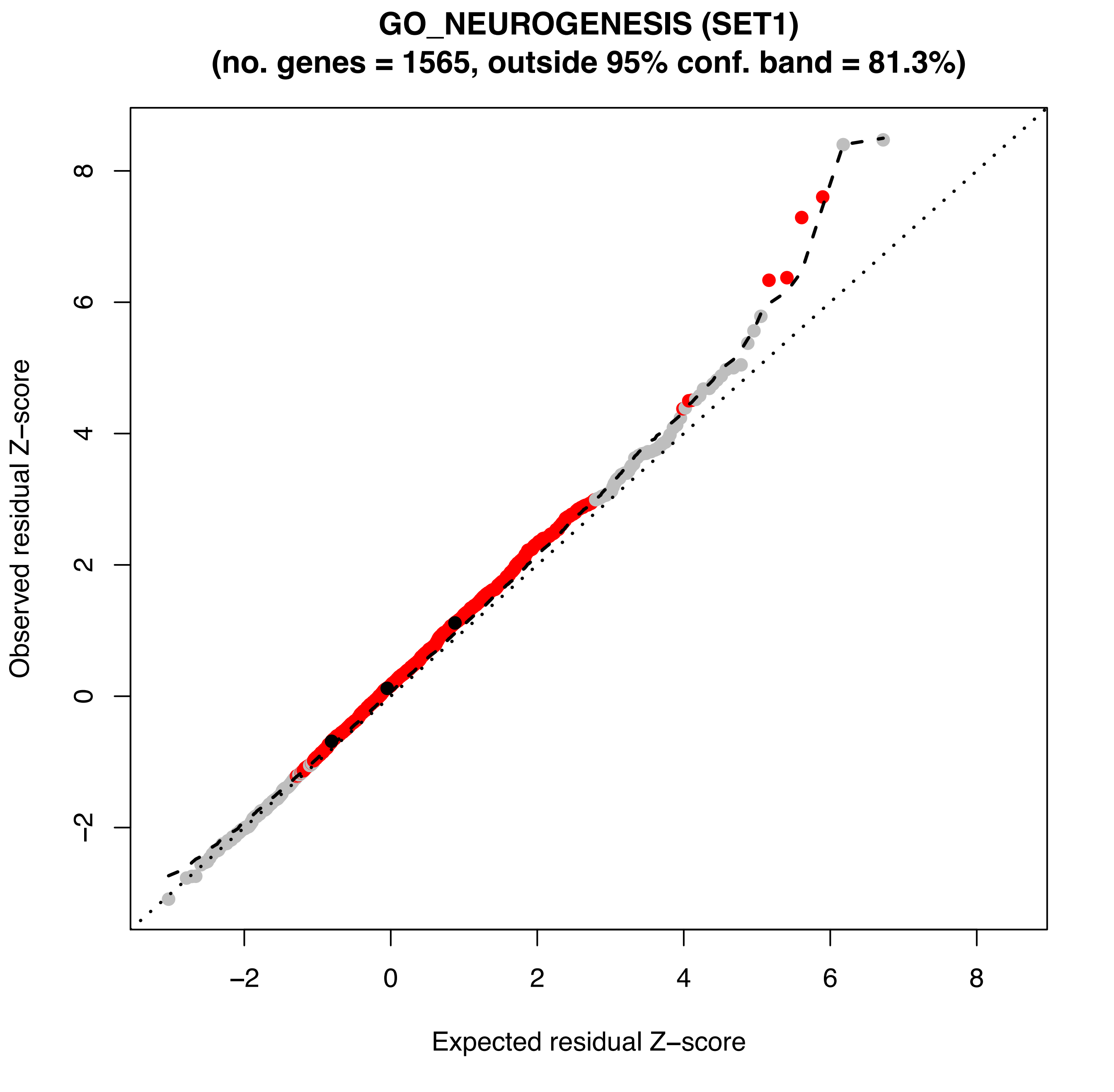

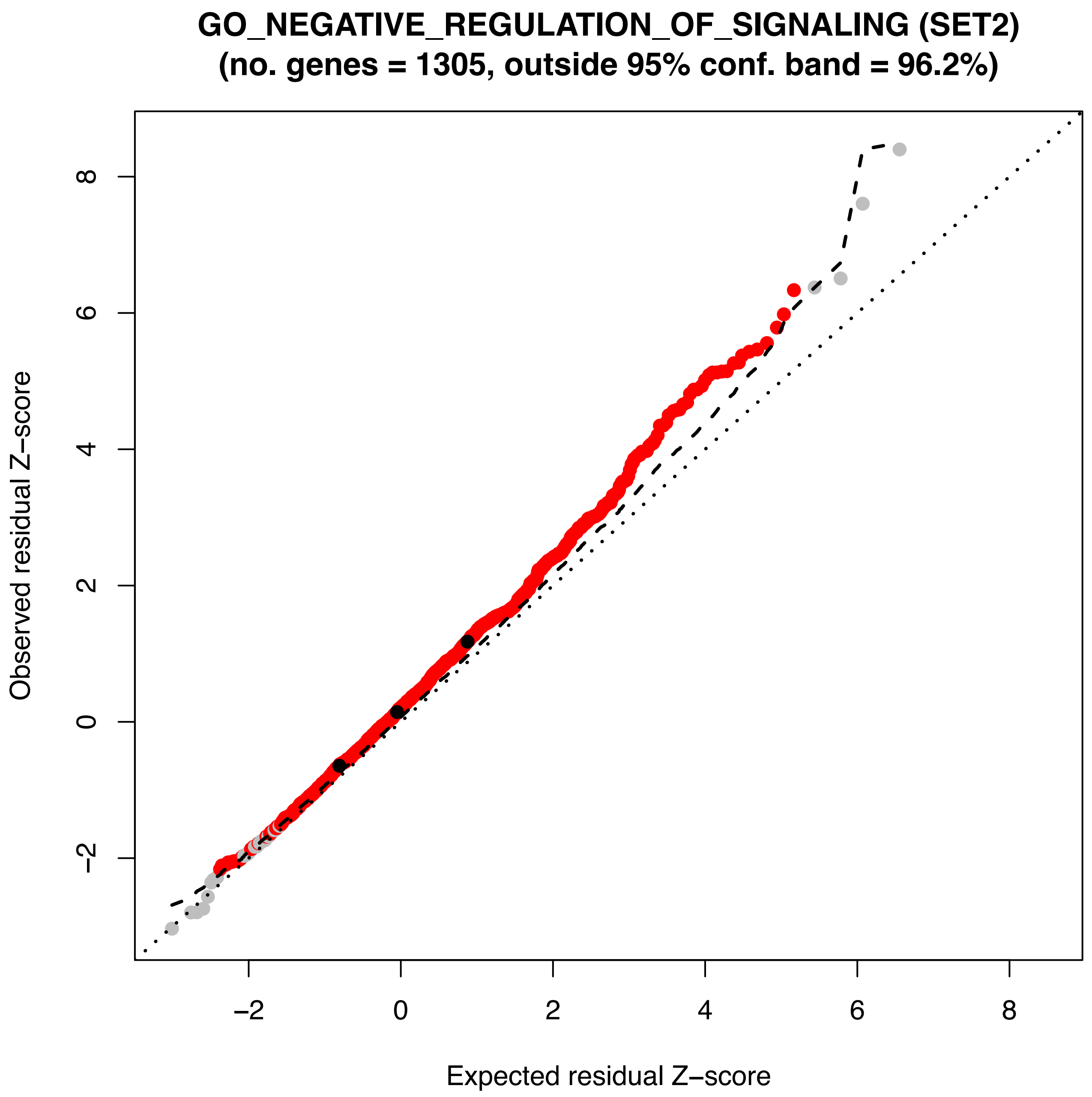

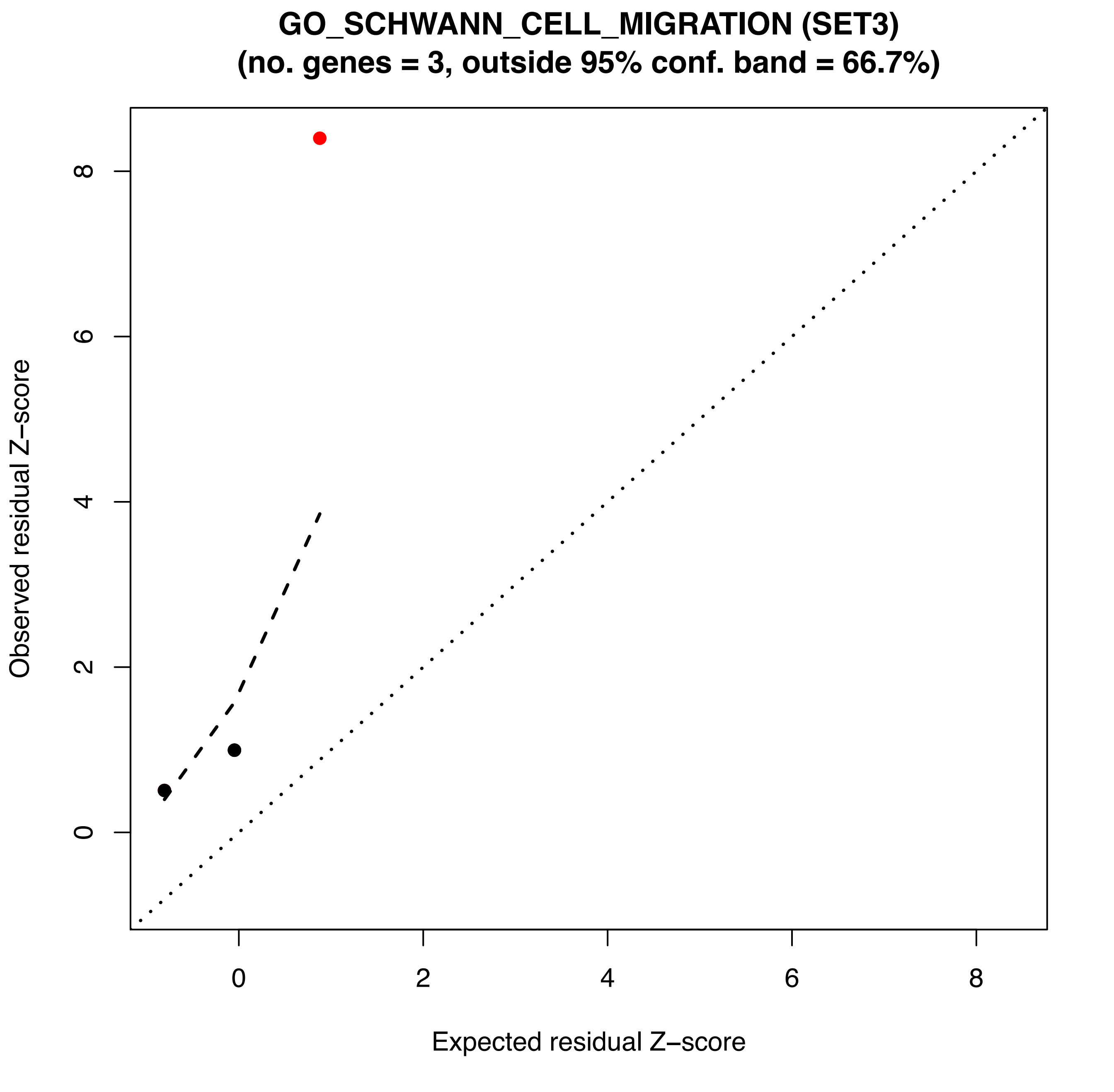

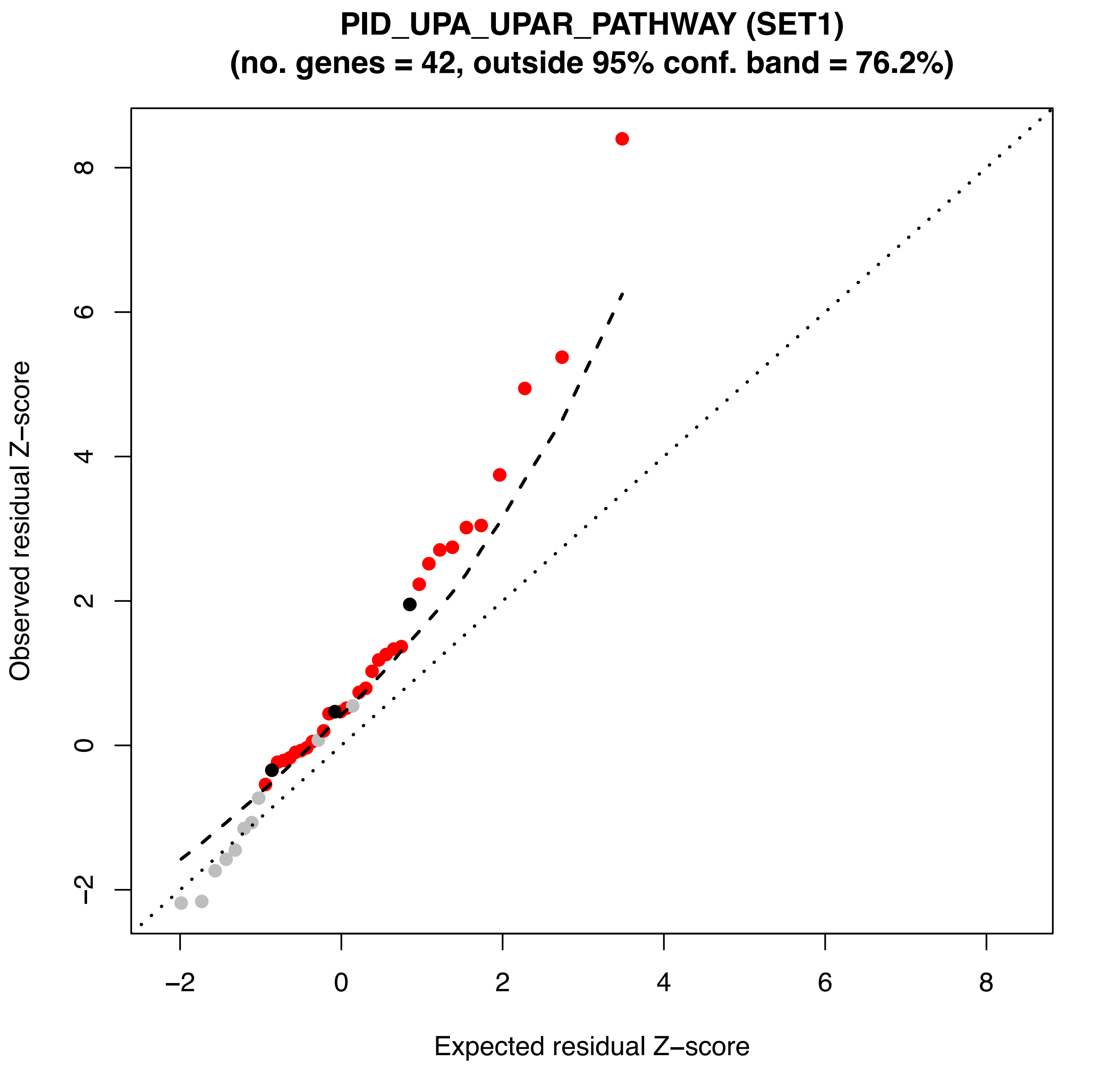

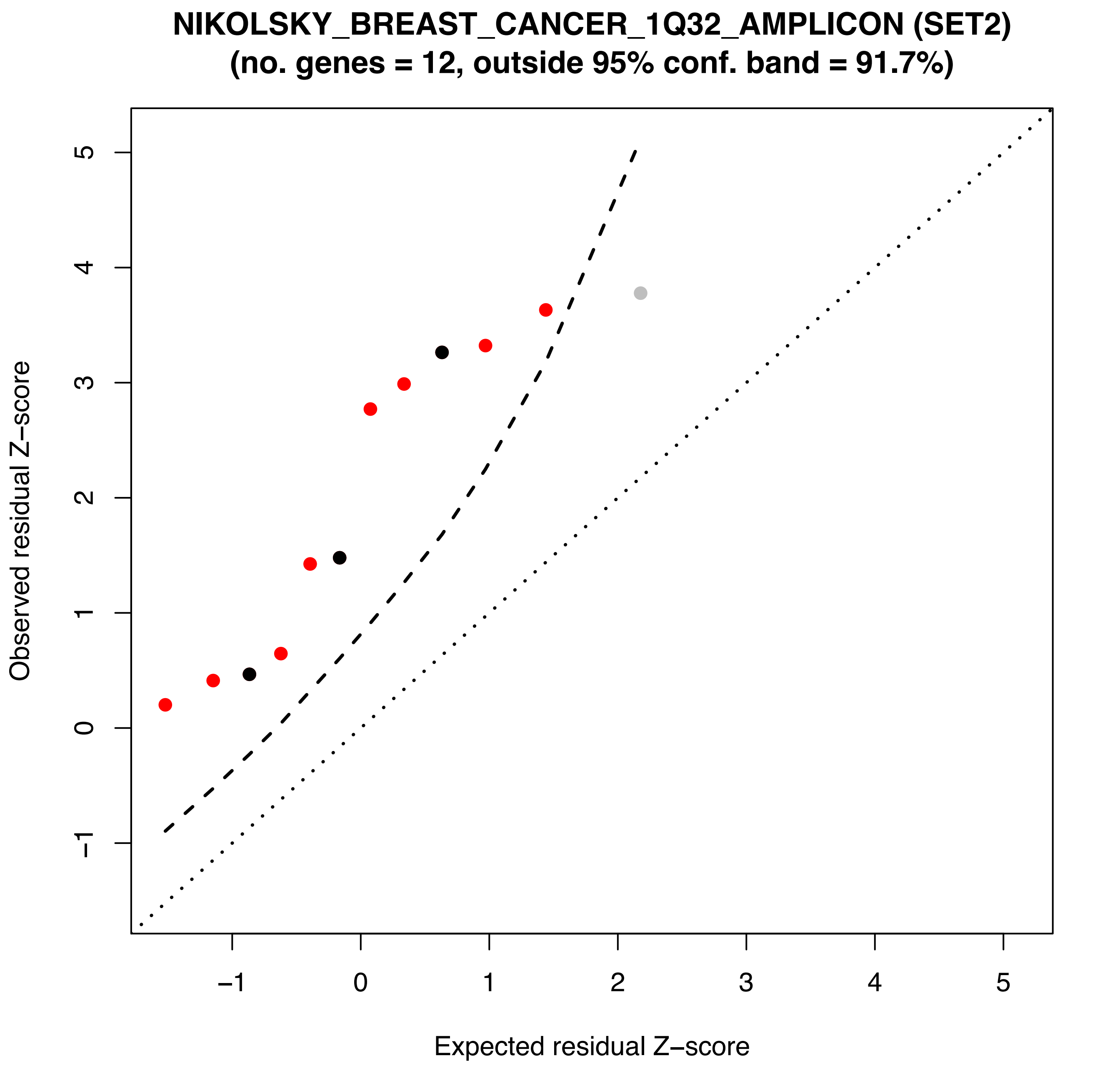
